# Glomerular spatial transcriptomics of IgA nephropathy according to the presence of mesangial proliferation

**DOI:** 10.1101/2022.11.17.22282435

**Authors:** Sehoon Park, Minji Kang, Yong Chul Kim, Dong Ki Kim, Kook-Hwan Oh, Kwon Wook Joo, Yon Su Kim, Hyunje Kim, Kyung Chul Moon, Hajeong Lee

**Author notes:** **Corresponding author** Hajeong Lee, MD, PhD, Associate Professor, Department of Internal Medicine, Seoul National University Hospital, 101 Daehak-ro, Jongno-gu, Seoul, 03080, Korea.

## Abstract

**Background:** Mesangial proliferation is a key pathologic feature of diagnosis and also a risk factor for progression in immunoglobulin A nephropathy (IgAN). We aimed to investigate the gene expression profiles of IgAN glomerulus according to the presence of mesangial proliferation.

**Methods:** We performed spatial-specific transcriptomic profiling for formalin-fixed-paraffin-embedded kidney biopsy tissues using the GeoMX Digital Spatial Profiler. A total of 3 glomeruli with direct pathologic classification for each case were profiled, including 12 cases (4 M1-IgAN, 4 M0-IgAN, and 4 donor controls). The differentially expressed genes (DEGs) were analyzed between the groups with gene ontology (GO) term annotation.

**Results:** In the result which successfully enriched glom-specific genes, M1-IgAN were distinctively separated from controls (77 upregulated and 55 downregulated DEGs), while certain DEGs were identified between M1-IgAN and M0-IgAN cases (24 upregulated and 8 downregulated DEGs) or between M0 and controls (1 upregulated and 16 downregulated DEGs). The upregulated DEG that was consistent in M1-IgAN and M0-IgAN cases was TCF21 which is known as early podocyte injury marker while the DEGs uniformly downregulated in IgAN cases included ATF3, EGR1, DUSP1, FOS, JUNB, KLF2, KLF4, NR4A1, RHOB, and ZFP36. Glomeruli from M1-IgAN cases were significantly enriched for cell surface/adhesion molecules and the gene expressions that consist vascular development or extracellular matrix.

**Conclusion:** We found certain transcriptomic differences according to the histologic changes in IgAN which was present even in the early stage of the disease. Spatial transcriptomic analysis may contribute to dissecting structure-specific pathogenesis and molecular changes in IgAN.

**Significance Statement:** Immunoglobulin A nephropathy (IgAN) is the most common primary glomerulonephritis with distinct pathologic features of mesangial proliferation with IgA deposition. The authors used spatial transcriptomic analysis to investigate the gene expression profiles from glomeruli with direct pathology annotation. The analysis revealed notable differentially expressed genes in IgAN with and without mesangial proliferation including early podocyte changes, expressions of cell adhesion molecules, alteration in transcription factor pathway, vascular development and extracellular matrix production. The current spatial transcriptomic analysis suggests notable gene expression profiles that may contribute to pathogenesis and progression of IgAN.

## Introduction

Immunoglobulin A nephropathy (IgAN) is one of the most common primary glomerular diseases in adults.^1^ Approximately 20% of IgAN patients progress to end-stage kidney disease, ranking the disease as a major cause of medical and socioeconomic burden related to kidney dysfunction.^2^ Most of these patients show initial kidney dysfunction, high blood pressure, a large amount of proteinuria, or advanced histologic damage. On the other hand, IgAN patients without clinical or pathologic risk factors usually show benign courses. Such heterogeneity in clinical prognosis led to research focusing on prognostic prediction and stratification of IgAN,^3^ and treatment strategies may be differently applied according to the risk profiles.

The most common clinical features of IgAN are acute glomerulonephritis and asymptomatic urinary abnormalities including microscopic hematuria or incidentally found proteinuria. With a recent increase in routine urinalysis surveillance in school or health screening, a significant proportion of IgAN patients is presented with asymptomatic urinary abnormalities. This makes more IgAN patients diagnosed earlier with preserved kidney function, and therefore pathologic features play a key role in both diagnosis and risk stratification of IgAN. After the pathologic diagnosis by confirmation of glomerular IgA disposition and mesangial changes, the Oxford classification is widely used to subclassify the pathologic findings of IgAN because of their clinical utility in predicting long-term prognosis.^4^

As the pathologic features reflect the mechanism and severity of IgAN, a transcriptomic profiling targeting a specific pathologic change of IgAN may reveal a potential therapeutic or prognostic biomarker related to IgAN pathophysiology.^5, 6^ Particularly, as a glomerular mesangium is a key region where pathogenetic galactose-deficient IgA1 accumulates and the inflammatory process initiates, mRNA profiles related to the degree of mesangial proliferation in IgAN glomerulus may reflect the underlying mechanism of early disease development. Such investigation would be possible by spatial transcriptomic analysis tools which were recently introduced to nephrology area and reported glom-or tubule-specific changes of certain kidney diseases.^7, 8^

In this study, we aimed to profile the transcriptomic findings of IgAN glomerulus by spatial transcriptomic analysis. To reveal mesangial proliferation-specific transcriptomic changes, we carefully reviewed the pathologic parameters and dissected the patients according to the severity of mesangial proliferation. We hypothesized that the glomerular IgAN transcriptome would reveal a list of biomarkers that reflects the early pathophysiology of the disease.

## Methods

### Ethical considerations

This study was approved by the Institutional Reviewer Boards of Seoul National University Hospital (IRB No. H-2205-104-1325). All biospecimens and clinical data were collected after appropriate informed consent by the patients. This study was performed in accordance to the declaration of Helsinki.

### Study subjects – IgAN cases

In the study hospital, when informed consent was acquired, kidney biopsy tissues were collected and the remnant specimen after the establishment of diagnosis are stored in formalin-fixed paraffin-embedded (FFPE) blocks for future research or look-back diagnosis. We reviewed the IgAN cases with available remnant FFPE tissue blocks for their initial diagnostic biopsy and selected the target population according to the certain criteria. The criteria were defined to include IgAN appropriate for the spatial transcriptomic analysis in relatively early development courses without profound kidney dysfunction, including cases 1) with a sufficient number of glomeruli (> 10 per section), 2) biopsy performed in the recent period (≥ year 2018), 3) without immunosuppressive treatment history, 4) without comorbid diabetes mellitus, 5) with preserved estimated glomerular filtration rate (eGFR, ≥ 60 mL/min/1.73 m2), 6) and with established Oxford classification.^4^ We also limited the age range to 20 to 59 years old to exclude elderly cases to minimize the aging effect on glomerular changes. Lastly, we did not include cases with cellular or fibrocellular crescents, as such active immune status may represent fundamentally different characteristics and minimized the inclusion of the cases with tubular atrophy/interstitial fibrosis or endocapillary proliferation.^4, 9^ Some cases with segmental sclerosis or other findings than mesangial proliferation were included but we selected the glomerulus without such finding when selecting the target area-of-interest (AOI) for the spatial transcriptomic analysis.

Finally, the biopsy specimens were reviewed by a kidney pathology specialist for the degree of mesangial proliferation. IgAN cases with notable mesangial proliferation (M1) and mild changes (M0) were selected for 4 cases in each group. The glomeruli with certain M1 and M0 findings were marked by the pathologist and went through the spatial transcriptomic analysis.

### Study subjects – control group

We included 4 donor kidney biopsy samples as the control group. The donor kidney biopsy samples were collected immediate after implantation during transplantation surgery. We selected the cases in which the biopsies were performed in the same period as the IgAN cases (from 2018 to 2021) and considered balanced inclusion for age (1 case each for the 20s, 30s, 40s, and 50s) and sex (2 male and 2 female cases). The included donors had all preserved kidney function (eGFR ≥ 80 mL/min/1.73 m^2^) and no hematuria or proteinuria in their urinalysis results. In addition, pathology diagnosis including Banff classification 2017 ensured the absence of any glomerular disease in the donor control cases.^10^

### Spatial transcriptomic analysis

GeoMX Digital Spatial Profiler (DSP, Nanostring, WA, USA) is a spatial transcriptomic profiling tool that quantifies mRNA transcripts within designated AOIs. The method has been implemented in previous nephrology research to report spatially resolved transcriptomes of glomerulus and tubules.

Slide preparation was performed following the manufacturer’s guidelines (NanoString GeoMx DSP Manual Slide Preparation, MAN-10150-01). To avoid batch effects, three slides contained a mixture of groups including 4 biopsy specimens uploaded to each slide. For FFPE blocks, 5 *μ*m-thick tissue sections were mounted on charged slides (Leica BOND Plus slides) ensuring that they fit inside the Scan Area of the slide. The slides were baked at 60C for 30min, and then deparaffinized with CitriSolv and rehydrated with ethanol and 1X phosphate-buffered saline. Target epitopes were retrieved and exposed by incubating in 1X Tris-EDTA, pH 9, and preheated 1 *μ*g/ml proteinase K, respectively. After washing in 1X PBS, the slides were incubated with RNA detection probes that are linked to the DNA oligo barcode with UV photocleavable linker for NGS readout WTA (Whole Transcriptome Atlas) to hybridize overnight at 37C. On the following day, the slides were stained with SYTO 13 (NanoString, 121300303), CD68 (Novus, NBP2-34587AF532), PanCK (Novus, NBP2-33200AF594), and alpha Smooth Muscle Actin (Abcam, ab202296) for 1 h at room temperature, then loaded into the GeoMx DSP instrument for scanning and selecting AOI. Oligonucleotides encoding their target gene were cleaved and released from the selected AOI, and an average of 173.3125 cells/AOI were collected into DSP collection plate. Primer pairs, i5, and i7 dual-indexing sequences were used to amplify and index the oligonucleotides during polymerase chain reaction (PCR). Libraries were pooled into a single microtube and cleaned up with AMPure beads. Using the Agilent 4200 TapeStation system, the libraries appear around 174 base pairs and these were sequenced on Illumina Counting Platform, NextSeq 6000 with 27×27 paired end reads.

### Bioinformatics and statistical analysis

With GeoMx NGS Pipeline software (v2.0), the raw fastq files were converted into Digital Count Conversion files. We used the DSP Data Analysis Suite (v2.4) and quality-controlled the results. Gene expressions with lower than the limit of quantitation and user-defined values set to 2 were initially disregarded. The limit of quantitation is set to be the negative probe geomean multiplied by the geometric standard deviations of the negative probes. In addition, the genes without any expression in more than half of the included samples were also excluded.

Unnormalized read counts of the filtered genes were used to draw relative log expression plots to inspect technical outliers. Next, principal component analysis was used to plot the similarities and differences between the read counts of the included AOIs.

To identify differentially expressed genes (DEGs), we used the DESeq2 package in R (version 3.6.2).^11^ We compared the differences between the AOIs group IgAN and controls, and additional comparison was compared between the M0, M1, and control groups. The genes with a false-discovery rate < 0.1 by the Benjamini–Hochberg correction were considered as the DEGs. The identified DEGs were analyzed by the Toppgene suite mainly for gene ontology annotation.^12^ To investigate the protein-protein interaction network of the identified DEGs, we used the STRING database and the DEGs with at least one significant connection with another DEG were presented.^13^

## Results

### Characteristics of the study subjects

The characteristics of the study subjects are summarized in Table 1. IgAN cases had age range from 20 to 54 years old. None of them were on immunosuppressive drugs or had underlying diabetes mellitus. With relatively large numbers of glomerulus included in their biopsies, 1 IgAN case with M1 finding had E1, and 4 IgAN cases (1 from M0 and 3 from M1 group) had segmental sclerosis. All of the IgAN cases had eGFR > 60 mL/min/1.73 m^2^ and those with M1 findings showed higher amounts of baseline proteinuria. All IgAN with M1 glomerulus exhibit podocyte foot process effacements, which was present in 2 of IgAN with M0 glomerulus and 2 of control cases. Regarding the zero-time allograft biopsy controls, none of the cases had significant pathologic changes in their glomerulus, and only one case had a low grade changes of tubular atrophy and interstitial fibrosis. No significant proteinuria was detected in the control cases.

**Table 1.** Baseline characteristics of the study subjects.

| Group | Age * | Sex | RAAS B | ISD | Pathologic classification | Glo m N | Global sclerosis N (%) | Segmental sclerosis N (%) | Crescent N (%) | Diabetes | Hypertension | eGFR (mL/min/1.73 m<sup>2</sup>) | proteinuria (g/g or g/24h) | Electron microscopy |  |  |
| --- | --- | --- | --- | --- | --- | --- | --- | --- | --- | --- | --- | --- | --- | --- | --- | --- |
|  |  |  |  |  |  |  |  |  |  |  |  |  |  | Electron-dense deposit | GBM | Foot process effacement |
| M0 | 30s | F | (-) | (-) | M0, E0, S0, T0, C0 | 12 | 0 (0%) | 0 (0%) | 0 (0%) | (-) | (-) | 110.3 | 0.34 | Small mesangial | (-) | Focal slight |
| M0 | 50s | F | (+) | (-) | M0, E0, S1, T1, C0 | 25 | 2 (8%) | 1 (4%) | 0 (0%) | (-) | (+) | 86.9 | 0.51 | Small mesangial | (-) | (-) |
| M0 | 30s | M | (-) | (-) | M0, E0, S0, T0, C0 | 25 | 0 (0%) | 0 (0%) | 0 (0%) | (-) | (-) | 110.3 | 0.14 | Small mesangial | (-) | Focal mild |
| M0 | 20s | M | (-) | (-) | M0, E0, S0, T0, C0 | 27 | 0 (0%) | 0 (0%) | 0 (0%) | (-) | (-) | 134.70 | 0.18 | Small mesangial | (-) | (-) |
| M1 | 30s | F | (-) | (-) | M1, E0, S1, T0, C0 | 15 | 2 (13%) | 3 (20%) | 0 (0%) | (-) | (-) | 105.8 | 2.44 | Small mesangial | (-) | Focal mild |
| M1 | 20s | F | (-) | (-) | M1, E0, S1, T0, C0 | 59 | 8 (14%) | 2 (3%) | 0 (0%) | (-) | (-) | 136.2 | 3.13 | Moderate mesangial | (-) | Focal slight |
| M1 | 50s | M | (+) | (-) | M1, E0, S1, T0, C0 | 34 | 7 (21%) | 4 (12%) | 0 (0%) | (-) | (+) | 67.50 | 1.66 | Small mesangial | Thickening | Focal mild |
| M1 | 30s | F | (+) | (-) | M1, E1, S0, T0, C0 | 43 | 0 (0%) | 0 (0%) | 0 (0%) | (-) | (-) | 120.60 | 1.81 | Moderate mesangial, small subendothelial | (-) | Focal moderate |
| Control | 20s | M | (-) | (-) | Banff: no positive findings | 75 | 0 (0%) | 0 (0%) | 0 (0%) | (-) | (-) | 112.6 | Not detected | Small mesangial | (-) | Focal mild |
| Control | 30s | F | (-) | (-) | Banff: no positive findings | 76 | 8 (11%) | 0 (0%) | 0 (0%) | (-) | (-) | 113.2 | Not detected | (-) | (-) | Focal slight |
| Control | 40s | M | (-) | (-) | Banff: no positive findings | 24 | 0 (0%) | 0 (0%) | 0 (0%) | (-) | (-) | 111.9 | Not detected | (-) | (-) | (-) |
| Control | 50s | F | (-) | (-) | Banff: interstitial fibrosis/tubular atrophy grade 1 (ci1, ct1, i-IFTA2, t-IFTA1) | 42 | 3 (7%) | 0 (0%) | 0 (0%) | (-) | (-) | 90.2 | Not detected | (-) | (-) | (-) |

RAASB = renin-angiotensin-aldosterone system blockades, ISD = immunosuppressive drugs, eGFR = estimated glomerular filtration rate, GBM = glomerular basement membrane
\* Age was presented as age ranges (10-years range) for deidentification.

The representative light microscope biopsy slide scan results with the Periodic acid–Schiff staining are presented in Figure 1. We carefully selected the target AOI glomerulus to include prominent mesangial changes in the M1 group and to include mild changes in the M0 group. The control cases showed normal findings in the results.

**Figure 1.**
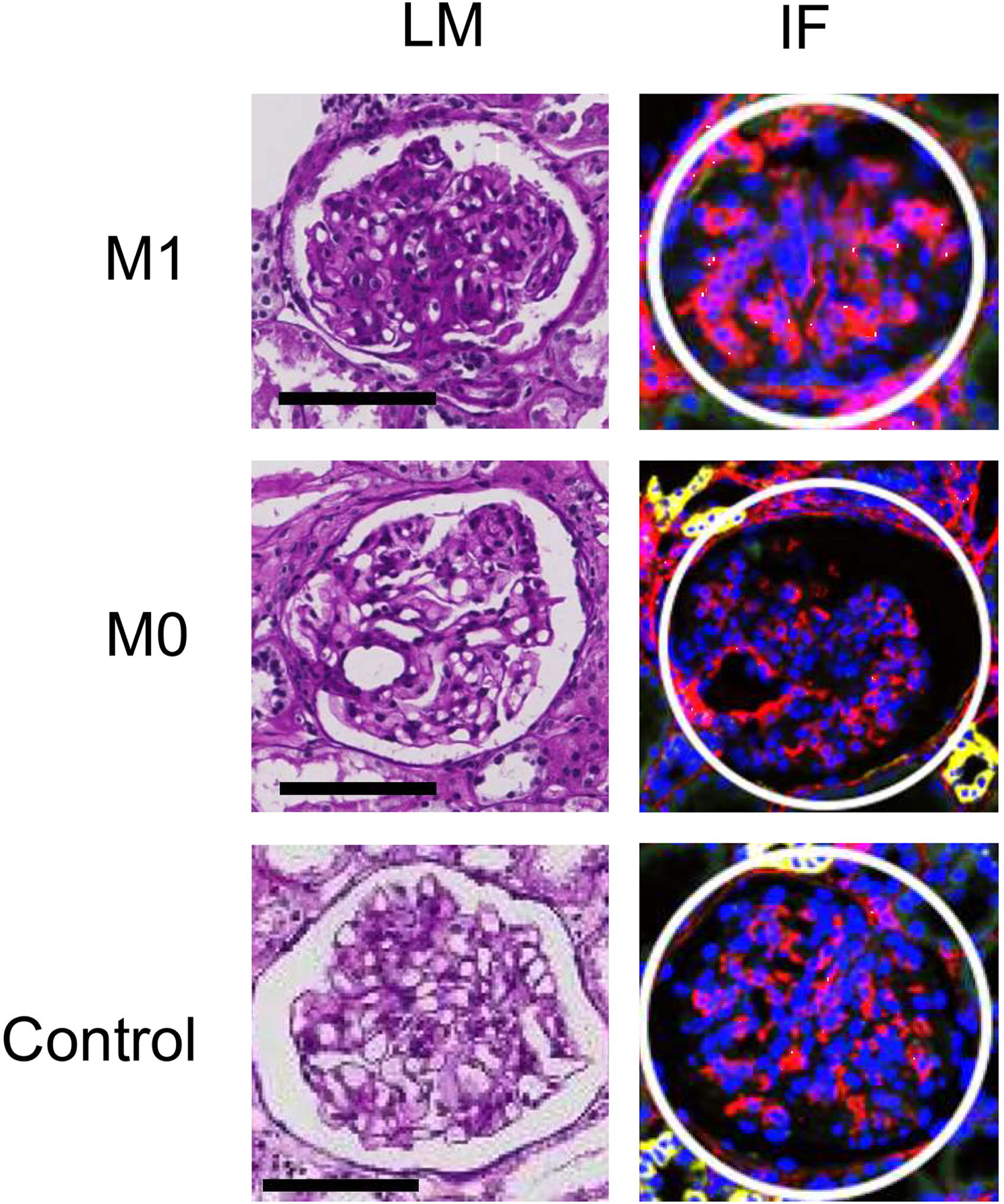
Representative images showing the glomerulus targeted in the study. Left graphs show the light microscopy images (bars = 100 micrometers) and the right graphs show the immunofluorescence images (blue, SYTO13; yellow, panCK; red, alpha Smooth Muscle Actin; green, CD68). White circles show the designated area-of-interest in the GeoMX Digital Spatial Profiler. Each glomeruli included in the spatial transcriptomic profiling showed distinct degree of mesangial proliferation directly annotated from the attending pathologist.

### Spatial transcriptomic profiling results

Round AOIs with a median of 205.8 (interquartile ranges 192.0, 222.9) um were drawn to include median 185 (139, 211) cells per AOIs (Figure 1). Median 1857156 reads were aligned from the RNA-seq with all cases reaching > 80% of Q30 bases, with a median mean quality score of 35 (interquartile range 34.9, 35.4). When we filtered the genes with lower expressions than the limit of quantitation or lower prevalence than 50% of the AOIs, a total of 14888 genes were remaining for the downstream analysis.

The relative log expression plots supported low likelihood of extreme outliers (Figure 2A). When gene expression profiles were initially investigated for kidney substructure-specific genes, the counts per million expression ranks for well-known glomerulus-specific genes were prominently higher (PODXL, rank 4; NPHS2, rank 108; NPHS1, rank 331) than those for proximal convoluted tubule (SLC34A1, rank 3005), distal convoluted tubule (SLC12A3, rank 2402) or those for collecting duct (AQP2, rank 2878) (Figure 2B).^14^ Considering the large amounts of tubulointerstitial tissue included in kidney biopsy samples, the findings supported that the GeoMX DSP successfully captured the enriched genes within kidney glomerulus.

**Figure 2.**
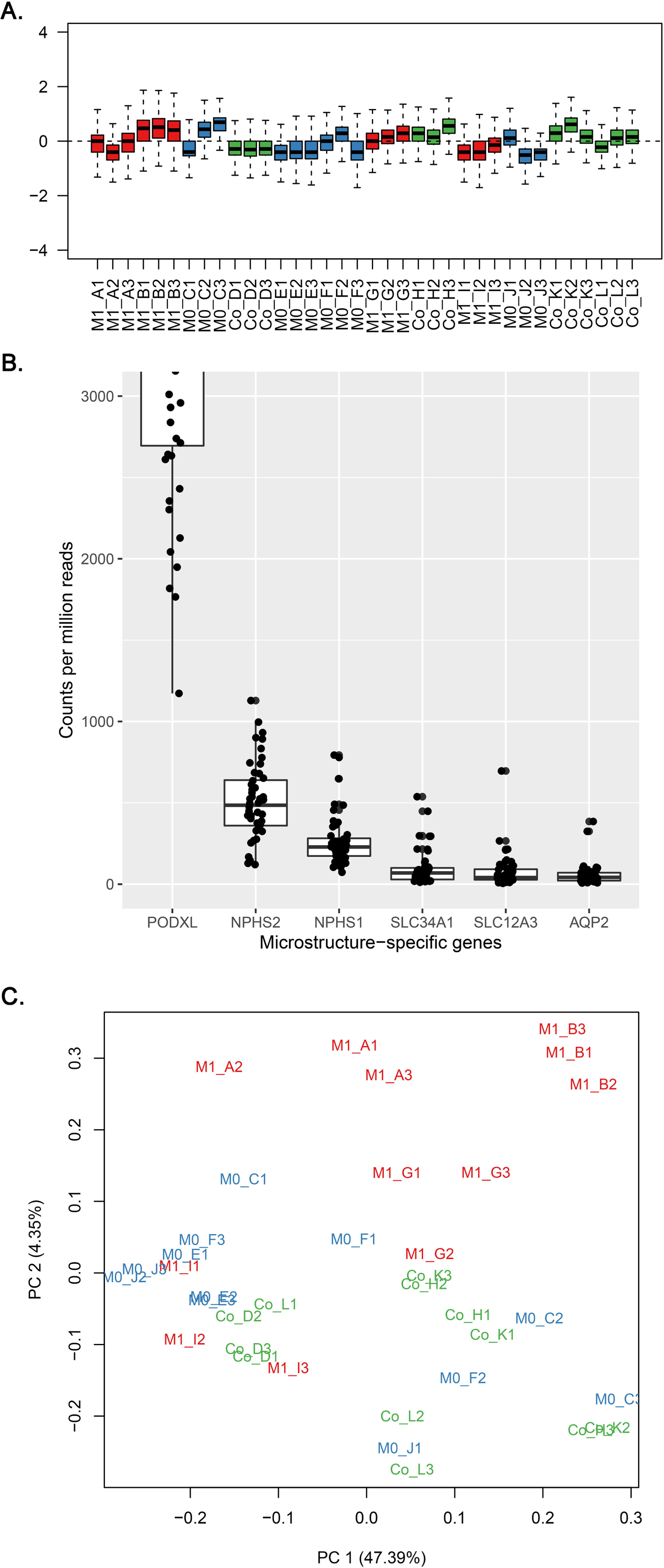
Initial quality control process of the read count results. A) Relative log expression plot demonstrated absence of notable batch effects. Each read counts from a area-of-interest is shown as a box plot, with the same order as the samples mounted on the experiment slides. Each slide included 12 area-of-interests from 4 cases, 3 glomeruli each (red, M1 cases; blue, M0 cases; green, control donor-kidney). B) Kidney microstructure-specific gene expression profiles indicated relative enrichment of glomerulus-specific genes (PODXL, NPHS2, and NPHS1) when compared to proximal convoluted tubule (SLC34A1), distal convoluted tubule (SLC12A3), and collecting duct (AQP2)-specific genes. C) Principal component analysis showed certain difference between the study groups (red, M1 cases; blue, M0 cases; green, control donor-kidney) which was particularly prominent for the M1 cases.

When we performed the principal component analysis, the results showed that, along with certain overlap between the M0, M1, and control AOIs, there was some discrimination for the M1 group (Figure 2C).

### Differentially expressed genes between IgAN and controls

The total list of differences between gene expression profiles of the study groups is presented in Supplemental Table 1. When we applied the false-discovery rate < 0.1 thresholds, between M0 and control, 16 downregulated DEGs [were identified with 1 upregulated DEG (Transcription factor 21, TCF21) (Table 2 and Figure 3A). In the GO term annotation (Table 3 and Supplemental Table 2), 5 among 16 downregulated DEGs between M0 and controls were annotated to the same molecular function GO term related to DNA-binding transcription activator activity and the genes included in the GO term were Fos proto-oncogene (FOS), KLF transcription factor 2 (KLF2), KLF4, early growth response 1 (EGR1), and JunB proto-oncogene (JUNB). FOS and JUNB were annotated to transcription factor AP-1 complex. Regarding the biological process GO term, 7 downregulated genes were annotated to the cellular response to endogenous stimulus including cytokine responses, hormonal stimulus, or oxidative stress. In protein-protein interaction analysis, the JUNB/FOS-related molecules interacted with each other (Figure 4A).

**Table 2.**
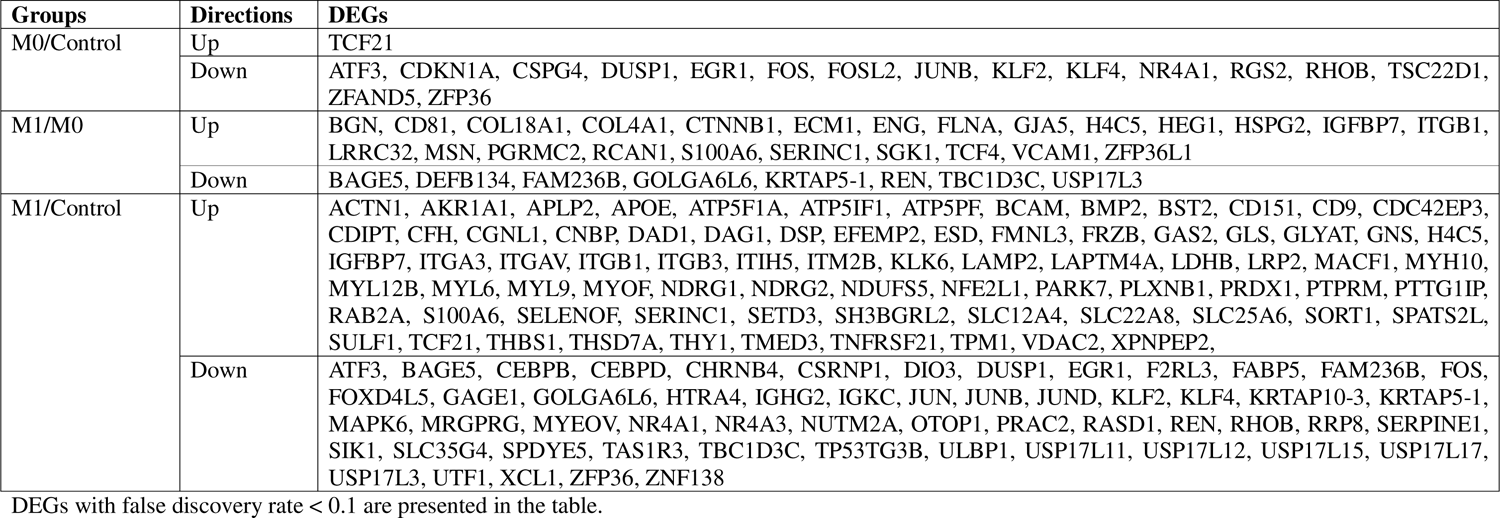
List of identified DEGs between the study groups.

**Table 3.** Top gene ontologies in each annotation results

| Comparison | Direction | Domain | Gene ontology terms | False discovery rate | Annotated gene lists |
| --- | --- | --- | --- | --- | --- |
| M0/Control | Up | Biological process | GO:0072275 (metanephric glomerulus morphogenesis) | 0.004 | TCF21 |
|  |  | Molecular function | GO:0050681 (nuclear androgen receptor binding) | 0.019 | TCF21 |
|  |  | Cellular component | NA | NA | NA |
|  | Down | Biological process | GO:0034097 (response to cytokine) | < 0.001 | FOS, ZFP36, KLF4, DUSP1, EGR1, KLF2, JUNB |
|  |  | Molecular function | GO:0000978 (RNA polymerase II cis-regulatory region sequence-specific DNA binding) | < 0.001 | FOS, KLF4, EGR1, KLF2, JUNB |
|  |  | Cellular component | GO:0000785 (chromatin) | 0.002 | FOS, KLF4, EGR1, KLF2, JUNB |
| M1/Control | Up | Biological process | GO:0048646 (anatomical structure formation involved in morphogenesis) | < 0.001 | TPM1, CFH, FMNL3, CNBP, BMP2, MYOF, THBS1, MYH10, ATP5IF1, THY1, MYL9, THSD7A, TCF21, CD9, IGFBP7, PTPRM, SULF1, CDIPT, LRP2, DAG1, MACF1, ACTN1, ITGA3, ITGAV, ITGB1, ITGB3, TNFRSF21 |
|  |  | Molecular function | GO:0050839 (cell adhesion) | < 0.001 | THBS1, NDRG1, THY1, CD9, PTPRM, DSP, PARK7, PRDX1, MACF1, CD151, ACTN1, BCAM, ITGA3, ITGAV, ITGB1, ITGB3 |
|  |  | Cellular component | GO:0009986 (cell surface) | < 0.001 | SORT1, CFH, ATP5PF, BMP2, THBS1, ATP5IF1, THY1, CD9, BST2, SULF1, LRP2, DAG1, CD151, BCAM, ITGA3, APOE, ITGAV, ITGB1, ITGB3, ATP5F1A |
|  | Down | Biological process | GO:0033993 (response to lipid) | 0.002 | EGR1, FOS, ZFP36, DUSP1, REN, CEBPB, NR4A1 |
|  |  | Molecular function | GO:0001216 (DNA-binding transcription activator activity) | < 0.001 | EGR1, FOS, ATF3, CEBPB, NR4A1 |
|  |  | Cellular component | GO:0005667 (transcription regulator complex) | 0.015 | FOS, ATF3, CEBPB, NR4A1 |
| M1/M0 | Up | Biological process | GO:0001944 (vascular development) | < 0.001 | COL4A1, HSPG2, FLNA, COL18A1, TCF4, GJA5, HEG1, CTNNB1, IGFBP7, RCAN1, ZFP36L1, ECM1, ENG, ITGB1, VCAM1, MSN |
|  |  | Molecular function | GO:0005201 (extracellular matrix constituent) | < 0.001 | COL4A1, HSPG2, COL18A1, IGFBP7, ECM1, ENG, BGN |
|  |  | Cellular component | GO:0030312 (extracellular matrix) | < 0.001 | COL4A1, S100A6, HSPG2, COL18A1, IGFBP7, ECM1, ITGB1, LRRC32, BGN |
|  | Down | Biological process | NA | NA | NA |
|  |  | Molecular function | NA | NA | NA |
|  |  | Cellular component | NA | NA | NA |

The gene ontology terms with the lowest false discovery rate in each annotation analysis are presented in the table.

**Figure 3.**
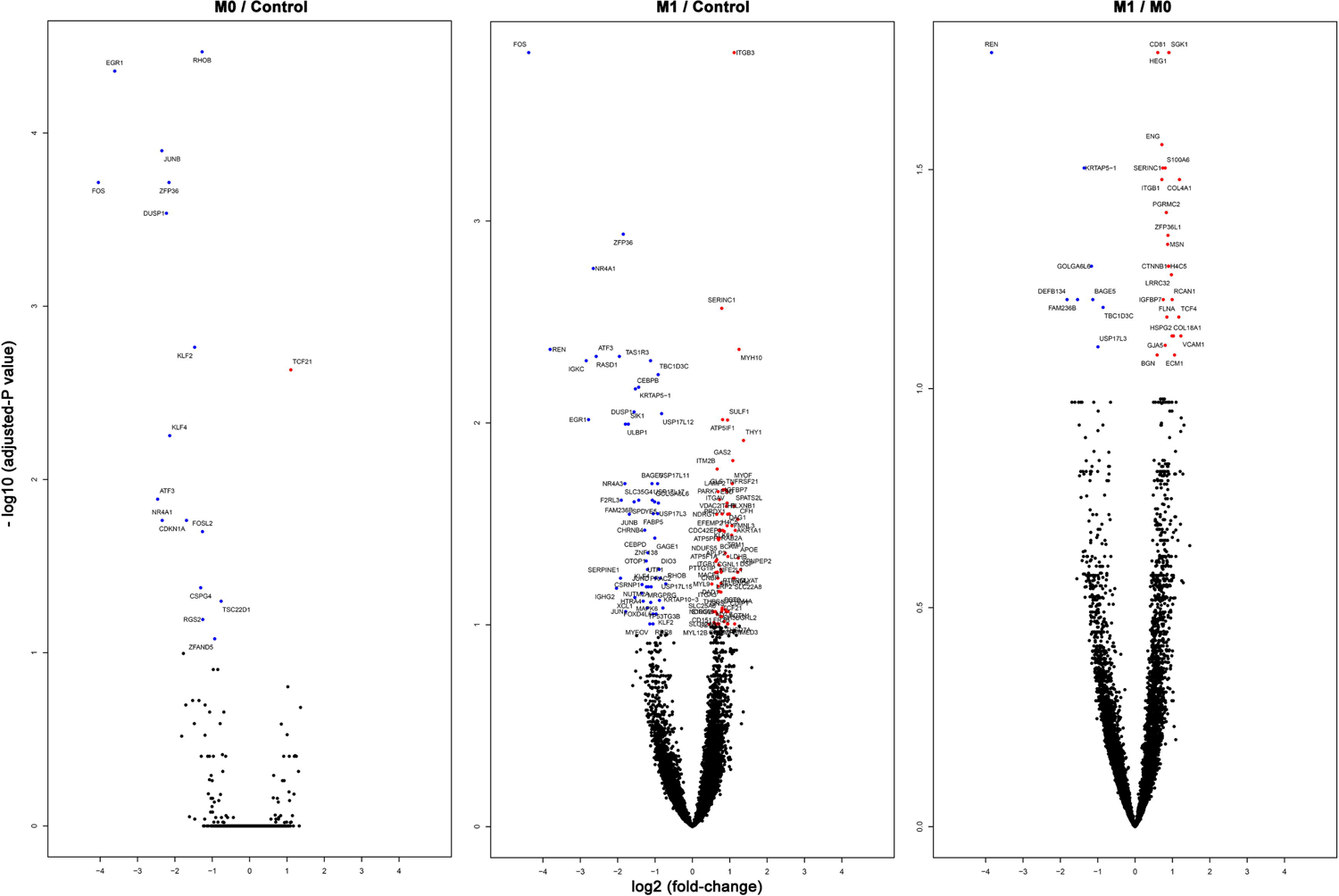
Volcano plots showing the differentially expressed genes (DEGs). X-axes indicate the log2 (fold-change) and Y-axes indicate -log10 (adjusted-P value). Significant (adjusted-P value < 0.1) downregulated genes are marked with blue color and upregulated genes with red. Left graph shows the DEGs between IgAN M0 cases and controls, middle between IgAN M0 cases and controls, and right between IgAN M1 and IgAN M0 cases.

**Figure 4.**
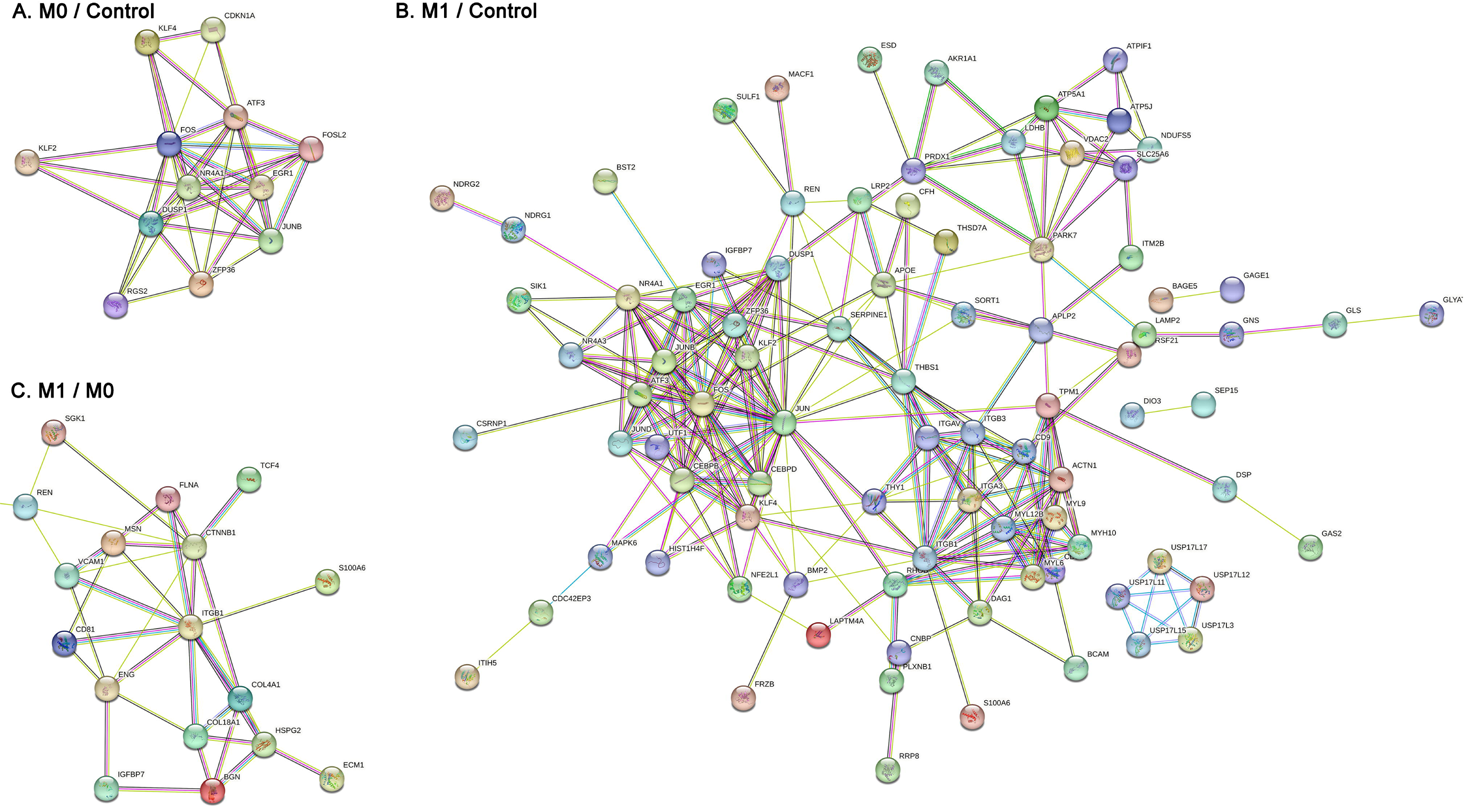
Protein-protein interaction network analysis. We used the STRING database and the DEGs with at least one significant connection with another DEG are presented in each comparison which was performed between A) IgAN M0 cases and controls, B) IgAN M1 cases and controls, and C) IgAN M1 cases and M0 cases.

Between M1 and control, the number of DEGs markedly increased, supporting the prominent difference between the glomerulus from M1 cases and controls, and we could identify 77 upregulated DEGs and 14 downregulated ones (Table 2 and Figure 3B). TCF21 was again included in the significant upregulated DEGs as the comparison between M0 and control. The consistently downregulated DEGs in both M1 and M0 IgAN were FOS, DUSP1, EGR1, and ZFP36. When we annotated the DEGs identified between M1 and controls, again, DNA-binding transcription factor activity was the frequently reported molecular function GO term that was annotated by downregulated genes, and 4 genes were annotated to transcription regulator complex cellular component (Table 3). Next, we annotated the 77 upregulated DEGs identified in M1 when compared to controls, and the most frequently annotated molecular function domain included cell adhesion molecule binding including extracellular matrix binding or integrin binding related GO terms. Regarding biological process GO terms, cell adhesion, endothelial cell proliferation, cell-matrix adhesion were notable domains with significantly enriched results. The functional network analysis also reported similar findings, as the JUN/FOS-related molecules were clustered, whilst, the integrin-or myosin-related molecules were also strongly interconnected (Figure 4B). Notably, ubiquitin-specific peptidase 17-like family member (USP17LX) molecules were also identified to be included in the upregulated DEGs.

The heatmap demonstrating the expression profiles of the genes included in notable GO terms is presented in Supplementary Figure 1.

### Differentially expressed genes between M1 and M0 among IgAN

We identified 24 upregulated and 8 downregulated DEGs between M1 and M0 glomerulus (Table 2, Figure 3C, and Supplemental Table 1). When the DEGs were annotated (Table 3 and Supplemental Table 3), total of 7/24 upregulated DEGs were annotated to extracellular matrix structural constituent (GO:0005201), including collagen alpha-1(IV) chain (COL4A1), COL18A1, heparin sulfate proteoglycan 2 (HSPG2), insulin-like growth factor binding protein 7 (IGFBP7), extracellular matrix protein 1 (ECM1), endoglin (ENG), and biglycan (BGN) (Table 3). In addition, cell adhesion molecule binding (GO:0050839) molecular function GO terms was another notable GO term with 7/24 upregulated DEGs were annotated. In addition, a total of 9 genes were annotated to signal receptor binding (GO:0005102) molecular function GO term, including HSPG2, ECM1, ENG, filamin A (FLNA), CD81, catenin beta 1 (CTNNB1), integrin subunit beta 1 (ITGB1), vascular cell adhesion molecule 1 (VCAM1), and moesin (MSN). For biological process GO terms, most of the upregulated DEGs (up to 16/24) were annotated to vascular development-related GO terms such as collagens (e.g. COL4A1, COL18A1) or cell adhesion-related molecules (e.g. ITGB1, VCAM1). The most commonly annotated cellular component GO terms were related to the plasma membrane, including integral component of plasma membrane (GO:0005887), membrane protein complex (GO:0098796), intrinsic component of plasma membrane (GO:0031226), and plasma membrane region (GO:0098590). In the functional network analysis, cell adhesion molecules and extracellular matrix components clustered together (Figure 4C). The heatmap showing the expression counts of the genes included in notable pathways is presented as Supplementary Figure 1.

## Discussion

By the current spatial transcriptomic profiling analysis, we revealed the DEGs that those expressions were altered in the glomerulus of IgAN. Related to the degree of mesangial proliferation, M1 and M0 glomerulus showed significant differences in their transcriptomic profiles, while a commonly altered signature even from the early IgAN without predominant mesangial proliferation was identified. The current results provide insights to the pathophysiologic changes that occur during the development and progression of IgAN.

The pathophysiologic mechanism of IgAN is explained by the glomerular accumulation of IgA1 and associated immune-complex with aberrant galactosylation.^15^ Such pathogenic IgA1 and immune-compex deposition is a pathologic hallmark of IgAN along with the consequent mesangial expansion of the glomerulus. The degree of mesangial proliferation reflects the disease activity of IgAN and those with M1 show a worse prognosis than those with M0.^4, 16^ The current literature suggests that the progression of mesangial expansion and related inflammatory signals lead to pathologic changes in tubulointerstitial tissue of IgAN.^17, 18^ In together, disrupted glomerular and tubular structure leads to proteinuria, which is a representative clinical parameter related to disease activity,^3^ and may result in non-reversible impairments in kidney function. Considering the above cascade of IgAN development and progression, the glomerulus having IgAN-specific pathologic changes of IgAN patients without clinical risk factors for progression may represent the early pathophysiologic mechanism of IgAN. Thus, there were efforts for targeted profiling of gene expression signatures in the glomerulus of clinically early IgAN patients.^5, 6^ However, because it was difficult in the past to identify the gene expression profiles of the glomerulus with directly annotated pathologic readings, the dissection of transcriptomic profiles of IgAN glomerulus regarding the degree of mesangial expansion was yet performed. We implemented a spatial transcriptomic profiling strategy, which has been recently introduced to the nephrology field with the utility to dissect kidney substructures,^7^ to investigate the transcriptomic profiles of IgAN regarding the degree of mesangial changes. Our study has main strengths in that 1) we profiled the transcriptome from pathology-annotated glomerulus of IgAN, 2) included early IgAN patients without established reduction in eGFR to reflect the early pathophysiologic signatures, 3) included allograft implantation biopsy controls, which the specimen would have certain power as clean-controls when compared to other types of control tissues commonly used in the field (e.g. tumor nephrectomy tissues), 4) and the profiles successfully enriched the gene expression profiles from the glomerulus. Our study revealed notable gene expression profiles of IgAN glomerulus, which were significantly different according to the degree of mesangial proliferation.

In our study, TCF21 was the single upregulated DEG commonly identified in M1 and M0 glomerulus, when compared to the controls. Interestingly, TCF21 is an essential mesoderm-specific gene and plays a key role in the embryonic development of glomerulus epithelial cells in the kidney, mainly expressed in podocytes. Previous studies demonstrated that lack of TCF21 resulted in defective differentiation of mesangial cells.^19, 20^ In addition, the expression of TCF21 in podocytes were markedly increased in injured glomeruli of IgAN, which was suggested to protect podocyte function depending on the podocyte damage.^21^ Also, a previous study reported the degree of proteinuria in IgAN patients showed correlation with TCF21 protein expressions.^22^ Considering the suggested importance of TCF21 in proteinuric glomerulonephritis and IgAN, we could identify the evidence of podocyte injury even in the early IgAN cases without prominent glomerular pathologic changes.

FOS and JUN-related gene expression profiles, related to transcription factor AP-2 complex, were downregulated in IgAN glomerulus. Although JUN/FOS pathway is considered a pro-inflammatory mechanism, the reduction in JUN/FOS in IgAN has been repetitively reported. In our previous transcriptomic investigation for microdissected IgAN glomerulus, these genes were prominently reduced in IgAN patients.^6^ In addition, a bioinformatic analysis including another previous transcriptomic profiling study for IgAN glomerulus by microarray addressed a significant reduction of FOS, JUN, FOSB, EGR1, and DUSP1 in IgAN, similar to the current results.^23^ Therefore, the early downregulation of JUN/FOS pathway may have particular importance in IgAN pathophysiology although the direct mechanism should be revealed in future experimental studies. Another notable gene that was downregulated in IgAN glomerulus was DUSP1. DUSP1 is an inhibitor of the mitogen-activated protein kinase (MAPK)-pathway, and our previous transcriptomic investigation demonstrated the DUSP1 expression was markedly low in IgAN glomerulus or even in tubules.^24^ Additional investigation hypothesizing that MAPK-pathway may play a universal role in pathogenesis or progression of IgAN in kidney glomerulus and tubules.

Glomerulus from M1 cases showed even marked differences in transcriptomic profiles with control glomerulus which would be reasonable considering the disease activity. THY1 is a representative marker for mesangial proliferation,^25^ and that the expression was increased in our results supports the validity of the current results. Considering that the extracellular matrix largely consists of the expanded mesangium, that extracellular matrix component-related molecules were the majority of upregulated gene expressions in M1 glomerulus was also reasonable. The current profiles suggests that with the progression of mesangial proliferation, inflammatory cell adhesion molecules express higher in IgAN glomerulus and the process related to endothelial proliferation may take place in the active stage of IgAN. We believe the current findings reflect the pathophysiologic mechanism of IgAN dissected by the degree of mesangial proliferation.

Future usage of the current study results would be further investigation of the clinical utility of the reported DEG lists. A further study may target TCF21, DUSP1, or JUN/FOS-related pathway to develop a medical strategy to block or reverse the pathophysiologic development of IgAN. In addition, the molecules related to mesangial proliferation are now noted in the transcriptomic profiles of M1 glomerulus, thus, such targets may be considered when assessing the disease severity of IgAN. In addition, a mechanism study which may reveal a therapeutic biomarker in IgAN to reduce the disease activity and mesangial proliferation.

The current study has several limitations. First, the sequencing depth of the transcriptomic profiles was hard to be deep considering the included number of cells, limiting the sensitivity of the current study. Namely, nominal changes from genes with relatively low absolute expression might not have been captured as a significant DEG. Thus, besides the reported DEGs, the total list may need to be reviewed with a less stringent interpretation for the P values, and the genes not reported as DEGs may still have clinical importance. Second, the gene expression profiles might have been affected by AOI selection. We noted an intra-individual variation of certain genes was prominent. This means that unintended differences in inclusion of juxtaglomerular cells or tubular cells might have resulted in the finding and that spatial transcriptomic profiling from glomerulus with small amounts of mRNA would be easily affected by such factors. Lastly, the current study only included East Asian cases from a single university hospital, thus, the generalizability of the current finding should be expanded by future research.

In conclusion, spatial transcriptomic analysis of IgAN glomerulus was a useful way to profile the gene expression signature dissected by the degree of mesangial proliferation. Future studies may use the results and the methodology to further expand the understanding of the IgAN pathophysiology and reveal a disease-specific biomarker.

## Data Availability

Data for this study is available by the corresponding author upon reasonable request.

## Author contribution statement

The corresponding author attests that all of the listed authors meet the authorship criteria and that no others meeting the criteria have been omitted. SP, HC, and HL contributed to the conception and design of the study. SP, MK, HC advised on statistical aspects and interpreted the data. SP, and MK performed the main statistical analysis. KCM was the main pathologist contributed to the pathologic annotation and case selection. SP and HL obtained funding and supervised the overall project. All of the authors participated in drafting the manuscript. All of the authors reviewed the manuscript and approved the final version to be published.

## Acknowlegment

The biospecimens for this study were provided by the Seoul National University Hospital Human Biobank, a member of the Korea Biobank Network, which is supported by the Ministry of Health and Welfare. This research was supported by a grant from National Research Fund of Republic of Korea (2022R1A2C2011190). This research was supported by a grant of the MD-Phd/Medical Scientist Training Program through the Korea Health Industry Development Institute (KHIDI), funded by the Ministry of Health & Welfare, Republic of Korea. This study was supported by the 2022 Young Investigator Research Grant from the KOREAN NEPHROLOGY RESEARCH FOUNDATION.

