## Supplemental Table 1 for "Glomerular spatial transcriptomics of IgA nephropathy according to the presence of mesangial proliferation"

| Gene | M0/Control | | | | M1/Control | | | | M1/M0 | | | |
|---|---|---|---|---|---|---|---|---|---|---|---|---|
| | baseMean | log2FoldC | padj | DEG | baseMean | log2FoldC | padj | DEG | baseMean | log2FoldC | padj | DEG |
| FOS | 45.15398 | -4.04808 | 0.000193 | DOWN | 50.66936 | -4.38146 | 0.000146 | DOWN | 4.46331 | -0.17973 | 0.909084 | |
| ITGB3 | 11.41616 | 0.54869 | 0.999532 | | 16.30643 | 1.118569 | 0.000146 | UP | 15.78528 | 0.5781 | 0.298614 | |
| ZFP36 | 23.99925 | -2.15674 | 0.000193 | DOWN | 28.21874 | -1.85095 | 0.001162 | DOWN | 9.243578 | 0.368711 | 0.632113 | |
| NR4A1 | 21.09421 | -2.33953 | 0.017231 | DOWN | 23.05969 | -2.65118 | 0.001718 | DOWN | 6.018148 | -0.16356 | 0.89433 | |
| SERINC1 | 25.94273 | 0.062079 | 0.999532 | | 38.20737 | 0.784837 | 0.002706 | UP | 33.35001 | 0.739632 | 0.031356 | UP |
| REN | 46.98585 | 0.045436 | 0.999532 | | 27.9355 | -3.80818 | 0.004322 | DOWN | 24.47307 | -3.84166 | 0.017097 | DOWN |
| MYH10 | 5.271527 | 0.387987 | 0.999532 | | 8.925281 | 1.248636 | 0.004322 | UP | 8.239981 | 0.885146 | 0.154583 | |
| ATF3 | 10.28229 | -2.46142 | 0.012998 | DOWN | 11.65136 | -2.57518 | 0.004682 | DOWN | 2.977464 | -0.18529 | 0.914367 | |
| RASD1 | 4.725067 | -1.29355 | 0.396029 | | 4.736647 | -1.95433 | 0.004682 | DOWN | 2.256862 | -0.6293 | 0.676565 | |
| IGKC | 14.03416 | -1.01673 | 0.999532 | | 11.96886 | -2.84168 | 0.004917 | DOWN | 5.090385 | -1.55785 | 0.271669 | |
| TAS1R3 | 11.44212 | -0.15025 | 0.999532 | | 9.708588 | -1.12014 | 0.004917 | DOWN | 8.17883 | -0.96028 | 0.145899 | |
| TBC1D3C | 19.93716 | -0.03647 | 0.999532 | | 17.31363 | -0.916 | 0.005763 | DOWN | 14.50302 | -0.85552 | 0.065256 | DOWN |
| KRTAP5- | 10.1279 | -0.05722 | 0.999532 | | 8.162164 | -1.43864 | 0.006655 | DOWN | 6.77966 | -1.36248 | 0.031356 | DOWN |
| CEBPB | 6.02566 | -0.65626 | 0.999532 | | 5.557653 | -1.52489 | 0.006781 | DOWN | 3.353563 | -0.84268 | 0.394452 | |
| DUSP1 | 21.95329 | -2.22642 | 0.000291 | DOWN | 27.27386 | -1.56124 | 0.00882 | DOWN | 9.481322 | 0.736326 | 0.183436 | |
| USP17L1 | 38.44055 | -0.0835 | 0.999532 | | 34.79032 | -0.82482 | 0.008994 | DOWN | 28.7123 | -0.7135 | 0.162261 | |
| EGR1 | 46.09707 | -3.61074 | 4.39E-05 | DOWN | 55.33649 | -2.78061 | 0.00963 | DOWN | 9.224201 | 0.894092 | 0.37404 | |
| ATP5IF1 | 17.98074 | 0.183256 | 0.999532 | | 26.58421 | 0.808986 | 0.00963 | UP | 23.21328 | 0.662636 | 0.241605 | |
| SULF1 | 10.71168 | 0.445634 | 0.999532 | | 14.68744 | 0.940604 | 0.00967 | UP | 14.16978 | 0.516444 | 0.336384 | |
| SIK1 | 7.491922 | -1.35539 | 0.18862 | | 8.08327 | -1.714 | 0.010137 | DOWN | 3.638021 | -0.36264 | 0.779652 | |
| ULBP1 | 4.759949 | -0.26104 | 0.999532 | | 3.828919 | -1.7929 | 0.010137 | DOWN | 2.898396 | -1.51201 | 0.107433 | |
| THY1 | 7.389435 | 0.160621 | 0.999532 | | 13.66589 | 1.367814 | 0.012216 | UP | 12.02715 | 1.222298 | 0.125254 | |
| GAS2 | 5.176974 | 0.367707 | 0.999532 | | 7.934236 | 1.081846 | 0.015368 | UP | 7.550887 | 0.736042 | 0.266797 | |
| ITM2B | 123.4679 | 0.171867 | 0.999532 | | 168.2668 | 0.663118 | 0.016925 | UP | 149.9947 | 0.516655 | 0.162189 | |
| MYOF | 5.463193 | 0.57562 | 0.999532 | | 7.534489 | 1.066999 | 0.019988 | UP | 7.47772 | 0.509162 | 0.532088 | |
| USP17L1 | 40.52232 | -0.32397 | 0.999532 | | 38.81517 | -0.93166 | 0.019988 | DOWN | 28.94413 | -0.5812 | 0.380614 | |
| NR4A3 | 6.252323 | -1.52417 | 0.18862 | | 6.67437 | -1.80717 | 0.019988 | DOWN | 2.808168 | -0.21867 | 0.899852 | |
| BAGE5 | 10.42416 | 0.063796 | 0.999532 | | 8.331113 | -1.08173 | 0.019988 | DOWN | 7.508725 | -1.13163 | 0.062627 | DOWN |
| GLS | 8.984493 | 0.264078 | 0.999532 | | 12.36483 | 0.880974 | 0.021418 | UP | 11.36805 | 0.619866 | 0.282041 | |
| TNFRSF2 | 7.445565 | 0.02449 | 0.999532 | | 12.33615 | 0.869845 | 0.021418 | UP | 10.30122 | 0.864179 | 0.148529 | |
| IGFBP7 | 159.7731 | 0.092847 | 0.999532 | | 240.7715 | 0.82183 | 0.021418 | UP | 209.8945 | 0.754537 | 0.062627 | UP |
| ESD | 13.84631 | 0.111199 | 0.999532 | | 19.77858 | 0.689209 | 0.021892 | UP | 17.08564 | 0.598906 | 0.154583 | |
| LAMP2 | 7.857938 | 0.216875 | 0.999532 | | 11.59606 | 0.931279 | 0.021892 | UP | 10.47734 | 0.728726 | 0.212469 | |
| PARK7 | 13.66057 | 0.08171 | 0.999532 | | 19.54929 | 0.727071 | 0.023952 | UP | 17.22999 | 0.662133 | 0.157378 | |
| USP17L1 | 36.31946 | -0.06123 | 0.999532 | | 30.91451 | -1.07264 | 0.02414 | DOWN | 25.82049 | -0.99306 | 0.124736 | |
| F2RL3 | 4.33548 | -1.70723 | 0.20025 | | 4.725096 | -1.90417 | 0.02414 | DOWN | 1.811271 | -0.14924 | 0.948865 | |
| SLC35G4 | 4.77518 | -0.22695 | 0.999532 | | 4.373398 | -1.43715 | 0.02414 | DOWN | 3.070681 | -1.14703 | 0.241605 | |
| FAM236B | 4.948682 | -0.00635 | 0.999532 | | 3.771326 | -1.55962 | 0.024598 | DOWN | 3.26555 | -1.54079 | 0.062627 | DOWN |
| GOLGA6 | 13.69911 | 0.180554 | 0.999532 | | 10.70026 | -1.01116 | 0.024598 | DOWN | 10.12067 | -1.16735 | 0.052543 | DOWN |
| PRDX1 | 8.843657 | 0.204064 | 0.999532 | | 13.25098 | 0.928888 | 0.024935 | UP | 11.99095 | 0.743792 | 0.154583 | |
| SPDYE5 | 15.36499 | -0.13853 | 0.999532 | | 14.12361 | -0.91043 | 0.024935 | DOWN | 11.30292 | -0.75273 | 0.162189 | |
| SPATS2L | 6.384793 | 1.022852 | 0.157178 | | 7.506561 | 1.123652 | 0.025845 | UP | 8.494865 | 0.123358 | 0.907968 | |
| ITGAV | 23.32666 | 0.281558 | 0.999532 | | 33.91771 | 0.910082 | 0.025875 | UP | 31.01984 | 0.652591 | 0.24893 | |
| FABP5 | 7.586436 | -0.71413 | 0.999532 | | 7.805946 | -1.04848 | 0.028135 | DOWN | 4.956438 | -0.33085 | 0.791576 | |
| USP17L3 | 48.95356 | 0.09678 | 0.999532 | | 40.87839 | -0.9288 | 0.028135 | DOWN | 36.47043 | -0.99299 | 0.080292 | DOWN |
| VDAC2 | 9.919346 | 0.238109 | 0.999532 | | 14.20905 | 0.795232 | 0.028232 | UP | 12.59844 | 0.567465 | 0.266933 | |
| PLXNB1 | 6.935142 | 0.779622 | 0.893099 | | 8.381031 | 0.987664 | 0.028232 | UP | 8.533797 | 0.215055 | 0.824012 | |
| JUNB | 15.977 | -2.34874 | 0.000127 | DOWN | 19.72512 | -1.68842 | 0.028325 | DOWN | 6.329087 | 0.723561 | 0.419745 | |
| ITIH5 | 10.0566 | 0.420968 | 0.999532 | | 14.10647 | 0.944416 | 0.028325 | UP | 13.31889 | 0.548428 | 0.300852 | |
| NDRG1 | 28.58149 | 0.114263 | 0.999532 | | 39.81053 | 0.650051 | 0.028325 | UP | 34.42685 | 0.545179 | 0.348525 | |
| CFH | 5.921191 | 0.487923 | 0.999532 | | 9.251085 | 1.213037 | 0.030034 | UP | 8.742599 | 0.751178 | 0.297055 | |
| DAG1 | 12.44096 | 0.443361 | 0.999532 | | 17.17361 | 0.929524 | 0.032285 | UP | 16.403 | 0.50896 | 0.451915 | |
| FMNL3 | 3.795505 | 0.253019 | 0.999532 | | 6.053434 | 1.058404 | 0.032285 | UP | 5.372818 | 0.822439 | 0.242703 | |
| CHRNB4 | 6.131856 | -0.45376 | 0.999532 | | 5.773786 | -1.2766 | 0.033967 | DOWN | 4.083485 | -0.80325 | 0.421517 | |
| ATP5PF | 12.26092 | 0.327815 | 0.999532 | | 15.91453 | 0.708823 | 0.034063 | UP | 14.81419 | 0.397166 | 0.515762 | |
| CDC42EP | 19.87574 | 0.633448 | 0.687916 | | 24.31907 | 0.806527 | 0.034063 | UP | 24.74954 | 0.190424 | 0.779299 | |
| H4C5 | 11.84408 | -0.14983 | 0.999532 | | 18.41438 | 0.729004 | 0.034063 | UP | 15.24594 | 0.891176 | 0.052543 | UP |
| AKR1A1 | 3.999341 | 0.26929 | 0.999532 | | 6.414965 | 1.148444 | 0.034063 | UP | 5.864182 | 0.896672 | 0.254225 | |
| EFEMP2 | 8.474418 | 0.429209 | 0.999532 | | 10.99337 | 0.85109 | 0.034436 | UP | 10.74011 | 0.442257 | 0.486743 | |
| KLK6 | 6.311367 | 0.375744 | 0.999532 | | 9.648788 | 1.050785 | 0.035966 | UP | 8.825381 | 0.686531 | 0.363418 | |
| RAB2A | 11.44574 | 0.09987 | 0.999532 | | 16.55448 | 0.724291 | 0.037196 | UP | 14.49223 | 0.643692 | 0.148529 | |
| GAGE1 | 10.26611 | 0.003879 | 0.999532 | | 8.70211 | -1.00614 | 0.037196 | DOWN | 7.42348 | -0.98794 | 0.11254 | |
| BCAM | 68.34534 | 0.372352 | 0.999532 | | 87.02752 | 0.68557 | 0.037196 | UP | 81.94999 | 0.343222 | 0.520848 | |
| NDUFS5 | 9.278089 | -0.0359 | 0.999532 | | 14.44091 | 0.701453 | 0.037897 | UP | 11.86641 | 0.761341 | 0.154583 | |
| CEBPD | 17.42393 | -1.19073 | 0.20025 | | 19.65277 | -1.19179 | 0.043968 | DOWN | 9.799318 | 0.027084 | 0.987491 | |
| TPM1 | 10.16308 | 0.276532 | 0.999532 | | 14.76523 | 0.880524 | 0.044101 | UP | 13.60026 | 0.62813 | 0.350296 | |
| LDHB | 15.0579 | 0.285472 | 0.999532 | | 22.47121 | 0.949886 | 0.045898 | UP | 20.26189 | 0.690525 | 0.438594 | |
| APOE | 9.634739 | 0.538173 | 0.999532 | | 14.76649 | 1.22858 | 0.046654 | UP | 13.91878 | 0.728104 | 0.566812 | |
| PTTG1IP | 17.3862 | 0.212182 | 0.999532 | | 23.06193 | 0.652414 | 0.047597 | UP | 20.8182 | 0.45893 | 0.363418 | |
| ZNF138 | 5.192446 | -0.04681 | 0.999532 | | 4.350568 | -1.23658 | 0.048285 | DOWN | 3.601101 | -1.16982 | 0.154583 | |
| APLP2 | 20.20225 | 0.032125 | 0.999532 | | 28.77273 | 0.631326 | 0.048517 | UP | 24.74639 | 0.61998 | 0.107433 | |
| ATP5F1A | 16.12915 | 0.080738 | 0.999532 | | 23.53355 | 0.706361 | 0.050496 | UP | 20.10569 | 0.658854 | 0.240311 | |
| CNBP | 8.8329 | 0.01738 | 0.999532 | | 13.299 | 0.76291 | 0.053212 | UP | 11.18594 | 0.766329 | 0.154583 | |
| OTOP1 | 6.05438 | -0.25221 | 0.999532 | | 5.548866 | -1.20053 | 0.053212 | DOWN | 3.855071 | -0.93042 | 0.263754 | |
| DIO3 | 17.96999 | -0.44308 | 0.999532 | | 18.1447 | -0.89164 | 0.053212 | DOWN | 12.54684 | -0.42305 | 0.542124 | |
| XPNPEP2 | 3.923895 | 0.625308 | 0.999532 | | 5.751266 | 1.291714 | 0.053212 | UP | 5.712068 | 0.685876 | 0.520848 | |
| ITGB1 | 28.77322 | -0.02163 | 0.999532 | | 41.61921 | 0.668988 | 0.054848 | UP | 35.74094 | 0.715794 | 0.033324 | UP |
| GLYAT | 3.400678 | 0.264239 | 0.999532 | | 5.713366 | 1.209298 | 0.054848 | UP | 5.012555 | 0.978715 | 0.274496 | |
| PTPRM | 7.001842 | -0.08856 | 0.999532 | | 11.02468 | 0.769447 | 0.054848 | UP | 9.109467 | 0.875736 | 0.107433 | |
| DSP | 3.352516 | 0.950301 | 0.999532 | | 4.237212 | 1.215211 | 0.054848 | UP | 4.476609 | 0.264829 | 0.84579 | |
| CGNL1 | 16.00972 | 0.525169 | 0.999532 | | 18.65574 | 0.623437 | 0.055119 | UP | 18.284 | 0.109419 | 0.888637 | |
| UTF1 | 16.1361 | -0.37026 | 0.999532 | | 16.08383 | -0.98656 | 0.058755 | DOWN | 11.25805 | -0.55297 | 0.550947 | |
| SERPINE | 5.929583 | -1.61036 | 0.8846 | | 6.367282 | -1.92457 | 0.058755 | DOWN | 2.484419 | -0.26955 | 0.877943 | |
| USP17L1 | 21.10335 | -0.11133 | 0.999532 | | 19.41393 | -0.86333 | 0.058755 | DOWN | 15.83103 | -0.72726 | 0.254225 | |
| LRP2 | 6.756034 | 0.247505 | 0.999532 | | 10.79911 | 1.120351 | 0.058755 | UP | 9.709084 | 0.885211 | 0.329409 | |
| NFE2L1 | 24.90544 | 0.375464 | 0.999532 | | 31.29487 | 0.680539 | 0.058755 | UP | 29.64361 | 0.327886 | 0.578798 | |
| SLC22A8 | 7.422883 | 0.068931 | 0.999532 | | 12.3063 | 1.089234 | 0.058924 | UP | 10.64665 | 1.001541 | 0.397004 | |
| MACF1 | 15.23822 | 0.127953 | 0.999532 | | 22.15302 | 0.769173 | 0.062239 | UP | 19.50469 | 0.661631 | 0.274496 | |
| RHOB | 24.51093 | -1.27152 | 3.40E-05 | DOWN | 31.3409 | -0.71335 | 0.062682 | DOWN | 16.75045 | 0.588077 | 0.193201 | |
| MYL9 | 101.8636 | 0.136327 | 0.999532 | | 133.0572 | 0.519412 | 0.062801 | UP | 117.5806 | 0.410163 | 0.313753 | |
| KLF4 | 8.867857 | -2.13892 | 0.005583 | DOWN | 11.42878 | -1.34877 | 0.063164 | DOWN | 4.146823 | 0.771789 | 0.405918 | |
| ITGA3 | 35.35248 | 0.242763 | 0.999532 | | 47.18505 | 0.696077 | 0.064449 | UP | 43.20883 | 0.478531 | 0.266933 | |
| JUND | 9.354854 | -1.07309 | 0.396029 | | 10.4873 | -1.10426 | 0.064836 | DOWN | 5.391999 | -0.00889 | 0.996891 | |
| MRGPRG | 4.711102 | -0.22254 | 0.999532 | | 4.181317 | -1.23127 | 0.064836 | DOWN | 3.242691 | -0.98646 | 0.300852 | |
| PRAC2 | 4.479232 | -0.46119 | 0.999532 | | 4.221527 | -1.17947 | 0.064836 | DOWN | 2.954826 | -0.70241 | 0.536706 | |
| IGHG2 | 10.19932 | -1.23405 | 0.999532 | | 10.04028 | -2.03197 | 0.065895 | DOWN | 4.326625 | -0.59866 | 0.617899 | |
| SELENOI | 8.196129 | 0.199787 | 0.999532 | | 11.57848 | 0.712509 | 0.068404 | UP | 10.51901 | 0.534964 | 0.342845 | |
| DAD1 | 9.654884 | -0.04078 | 0.999532 | | 14.55866 | 0.762081 | 0.068829 | UP | 12.33297 | 0.821395 | 0.107433 | |
| CSRNP1 | 8.108273 | -1.77209 | 0.100961 | | 9.991105 | -1.35534 | 0.069691 | DOWN | 4.028642 | 0.367081 | 0.755301 | |
| XCL1 | 3.978291 | -0.06428 | 0.999532 | | 3.162743 | -1.54098 | 0.073237 | DOWN | 2.503522 | -1.45481 | 0.154583 | |
| MAPK6 | 10.40835 | -0.38272 | 0.999532 | | 10.80553 | -0.88225 | 0.075671 | DOWN | 7.529993 | -0.45053 | 0.630705 | |
| HTRA4 | 4.245193 | 0.008124 | 0.999532 | | 3.311063 | -1.31662 | 0.076388 | DOWN | 2.849977 | -1.30923 | 0.154583 | |
| NUTM2A | 6.422207 | -0.26537 | 0.999532 | | 5.818047 | -1.11492 | 0.077473 | DOWN | 4.416198 | -0.83168 | 0.348787 | |
| TP53TG3 | 4.333082 | -0.64772 | 0.999532 | | 4.659771 | -1.2109 | 0.082536 | DOWN | 2.696238 | -0.50014 | 0.717627 | |
| KRTAP10 | 25.65382 | -0.2193 | 0.999532 | | 24.86947 | -0.79025 | 0.082536 | DOWN | 19.24772 | -0.54467 | 0.397004 | |
| BST2 | 30.84049 | 0.263369 | 0.999532 | | 42.84319 | 0.799883 | 0.082536 | UP | 39.20248 | 0.557434 | 0.323635 | |
| SH3BGRL | 6.499776 | 0.429746 | 0.999532 | | 8.759883 | 0.79687 | 0.083731 | UP | 8.18958 | 0.38287 | 0.617899 | |
| LAPTM4A | 16.8447 | 0.202203 | 0.999532 | | 24.01386 | 0.791959 | 0.084009 | UP | 21.6422 | 0.609355 | 0.339543 | |
| THBS1 | 13.03648 | 0.464678 | 0.999532 | | 17.93968 | 0.884765 | 0.084298 | UP | 16.66072 | 0.496286 | 0.566578 | |
| JUN | 8.223398 | -1.81784 | 0.303135 | | 9.370047 | -1.78547 | 0.086025 | DOWN | 3.461644 | 0.067893 | 0.972221 | |
| S100A6 | 75.30737 | -0.16286 | 0.999532 | | 113.3327 | 0.619909 | 0.086025 | UP | 92.50371 | 0.80661 | 0.031356 | UP |
| CDIPT | 4.864318 | 0.700602 | 0.999532 | | 6.011597 | 0.955596 | 0.086025 | UP | 6.247512 | 0.272395 | 0.789815 | |
| NDRG2 | 21.90798 | 0.025117 | 0.999532 | | 30.29008 | 0.554475 | 0.086085 | UP | 25.85489 | 0.550568 | 0.204889 | |
| FRZB | 10.83158 | 0.140707 | 0.999532 | | 15.51357 | 0.781207 | 0.086085 | UP | 13.89865 | 0.661109 | 0.241605 | |
| CD151 | 50.85066 | 0.048055 | 0.999532 | | 69.72091 | 0.568274 | 0.086085 | UP | 60.29593 | 0.54347 | 0.107433 | |
| GNS | 4.012641 | -0.17576 | 0.999532 | | 6.889161 | 0.906145 | 0.086085 | UP | 5.65386 | 1.097733 | 0.107433 | |
| KLF2 | 30.55646 | -1.47131 | 0.001726 | DOWN | 38.23643 | -0.97178 | 0.088391 | DOWN | 18.603 | 0.542867 | 0.19891 | |
| FOXD4L5 | 11.40375 | -0.04442 | 0.999532 | | 9.7456 | -1.05413 | 0.088391 | DOWN | 8.261332 | -0.98912 | 0.209531 | |
| SLC25A6 | 11.16076 | 0.195615 | 0.999532 | | 14.88229 | 0.662418 | 0.088475 | UP | 13.46885 | 0.487919 | 0.385915 | |
| TCF21 | 25.0857 | 1.102807 | 0.00233 | UP | 24.1009 | 0.760861 | 0.090768 | UP | 29.1011 | -0.32017 | 0.672452 | |
| SETD3 | 5.820684 | 0.047004 | 0.999532 | | 8.433021 | 0.785323 | 0.091019 | UP | 7.511843 | 0.765005 | 0.226915 | |
| THSD7A | 5.975031 | 0.312943 | 0.999532 | | 7.951656 | 0.850406 | 0.096387 | UP | 7.441239 | 0.548043 | 0.481527 | |
| MYL6 | 41.86824 | 0.152489 | 0.999532 | | 55.38575 | 0.578336 | 0.097416 | UP | 49.28192 | 0.449292 | 0.277462 | |
| RRP8 | 5.415331 | -0.24818 | 0.999532 | | 4.724368 | -1.05468 | 0.099003 | DOWN | 3.730665 | -0.79871 | 0.426561 | |
| TMED3 | 3.470553 | 0.6915 | 0.999532 | | 4.969733 | 1.130695 | 0.099003 | UP | 4.923299 | 0.461836 | 0.659727 | |
| ACTN1 | 6.475749 | 0.283176 | 0.999532 | | 9.516108 | 0.937324 | 0.099003 | UP | 8.899966 | 0.676977 | 0.39437 | |
| SORT1 | 13.68072 | 0.491194 | 0.999532 | | 16.42545 | 0.706943 | 0.099003 | UP | 16.05927 | 0.240402 | 0.75311 | |
| BMP2 | 5.169603 | 0.640782 | 0.999532 | | 6.41081 | 0.947847 | 0.099003 | UP | 6.674123 | 0.313749 | 0.771685 | |
| CD9 | 17.03939 | -0.09797 | 0.999532 | | 24.95878 | 0.611848 | 0.099003 | UP | 20.53546 | 0.734197 | 0.105617 | |
| MYEOV | 4.399552 | -0.54641 | 0.999532 | | 4.382541 | -1.13922 | 0.099003 | DOWN | 2.995243 | -0.56485 | 0.666906 | |
| SLC12A4 | 11.40944 | 0.061311 | 0.999532 | | 16.57158 | 0.671631 | 0.099003 | UP | 14.40433 | 0.636155 | 0.266933 | |
| MYL12B | 25.11272 | -0.01361 | 0.999532 | | 32.93367 | 0.446634 | 0.099392 | UP | 28.263 | 0.466676 | 0.175031 | |
| RBP5 | 5.650746 | 0.537122 | 0.999532 | | 8.637348 | 1.257074 | 0.101579 | | 8.417787 | 0.728729 | 0.515762 | |
| TTC14 | 6.128804 | 0.912578 | 0.396029 | | 6.723009 | 0.894483 | 0.102526 | | 7.574663 | -0.00187 | 0.999385 | |
| APP | 28.86336 | 0.043183 | 0.999532 | | 38.91862 | 0.49541 | 0.102526 | | 33.26089 | 0.474163 | 0.266933 | |
| NDUFA1 | 20.78724 | 0.192742 | 0.999532 | | 26.82531 | 0.558051 | 0.102526 | | 24.2316 | 0.384835 | 0.515762 | |
| CDC37 | 18.16597 | 0.160179 | 0.999532 | | 24.26915 | 0.614121 | 0.102526 | | 21.78516 | 0.470965 | 0.342845 | |
| PPIB | 11.87471 | 0.112785 | 0.999532 | | 17.17227 | 0.726158 | 0.102526 | | 14.81438 | 0.634385 | 0.266933 | |
| NDUFB4 | 11.23961 | -0.02259 | 0.999532 | | 16.5867 | 0.634781 | 0.102526 | | 13.96321 | 0.676385 | 0.162189 | |
| ITGB5 | 15.21623 | 0.333842 | 0.999532 | | 19.25578 | 0.638988 | 0.102526 | | 18.17827 | 0.32715 | 0.603483 | |
| ST6GALN | 12.44031 | 0.043771 | 0.999532 | | 18.98452 | 0.779009 | 0.103912 | | 16.10662 | 0.766114 | 0.158141 | |
| PCBP2 | 47.44132 | 0.054411 | 0.999532 | | 64.47456 | 0.570805 | 0.103912 | | 55.77591 | 0.539291 | 0.2006 | |
| GABARA | 18.36844 | 0.286219 | 0.999532 | | 23.07763 | 0.590608 | 0.103912 | | 21.4541 | 0.321801 | 0.59686 | |
| TRIR | 8.771545 | 0.166786 | 0.999532 | | 12.75108 | 0.754322 | 0.10586 | | 11.128 | 0.613058 | 0.283071 | |
| SLC16A9 | 4.583017 | 0.316849 | 0.999532 | | 7.415737 | 1.130343 | 0.10678 | | 6.568056 | 0.857802 | 0.386771 | |
| SUMF2 | 5.921307 | -0.04609 | 0.999532 | | 9.409494 | 0.7977 | 0.10823 | | 7.860048 | 0.865544 | 0.178337 | |
| CUX1 | 14.20391 | 0.462993 | 0.999532 | | 17.6174 | 0.667076 | 0.10859 | | 16.85788 | 0.231651 | 0.744708 | |
| CCL3 | 18.76183 | -0.29266 | 0.999532 | | 18.21517 | -0.81223 | 0.10859 | | 14.00768 | -0.50222 | 0.49299 | |
| SPOCK1 | 37.31155 | 0.643955 | 0.51089 | | 43.094 | 0.708143 | 0.10859 | | 44.1658 | 0.089679 | 0.92697 | |
| RAB1A | 11.29542 | 0.028996 | 0.999532 | | 15.68174 | 0.6011 | 0.10859 | | 13.17519 | 0.592784 | 0.226915 | |
| RPS28 | 47.69773 | 0.06753 | 0.999532 | | 67.34382 | 0.670748 | 0.10892 | | 58.7545 | 0.620015 | 0.246926 | |
| CCNI | 19.13727 | 0.062177 | 0.999532 | | 26.42522 | 0.596716 | 0.10892 | | 22.91307 | 0.556341 | 0.254225 | |
| FABP1 | 5.98675 | 0.401755 | 0.999532 | | 8.598657 | 1.094124 | 0.10892 | | 8.065425 | 0.718279 | 0.455747 | |
| ARF1 | 35.85424 | 0.055349 | 0.999532 | | 48.1401 | 0.498615 | 0.109477 | | 41.4062 | 0.467851 | 0.388809 | |
| GADD45A | 11.11444 | 0.145153 | 0.999532 | | 15.82894 | 0.733052 | 0.111402 | | 13.90956 | 0.608649 | 0.424976 | |
| POU5F1 | 12.10333 | -0.32007 | 0.999532 | | 12.20479 | -0.68523 | 0.111604 | | 9.54948 | -0.35136 | 0.673405 | |
| TCEAL9 | 8.956837 | 0.303613 | 0.999532 | | 11.73526 | 0.674762 | 0.111604 | | 10.66625 | 0.392824 | 0.561068 | |
| ARF5 | 5.442769 | 0.048642 | 0.999532 | | 8.207134 | 0.775709 | 0.111604 | | 7.098436 | 0.746093 | 0.258391 | |
| PSMA7 | 8.229446 | 0.393535 | 0.999532 | | 10.13875 | 0.774797 | 0.111604 | | 9.920673 | 0.403109 | 0.573903 | |
| PTBP3 | 10.48044 | 0.227646 | 0.999532 | | 13.65853 | 0.668659 | 0.111604 | | 12.44632 | 0.453584 | 0.496445 | |
| PSG1 | 13.04873 | -0.011 | 0.999532 | | 11.71298 | -0.84239 | 0.11259 | | 9.874367 | -0.8154 | 0.175031 | |
| CCDC93 | 2.588417 | 0.902109 | 0.999532 | | 3.342679 | 1.176428 | 0.112726 | | 3.639361 | 0.292634 | 0.816775 | |
| FCF1 | 136.1564 | -0.04598 | 0.999532 | | 125.2897 | -0.72057 | 0.112726 | | 104.5542 | -0.64784 | 0.241605 | |
| FAM71E2 | 2.642283 | -0.69886 | 0.999532 | | 2.481658 | -1.49303 | 0.113181 | | 1.626088 | -0.78342 | 0.655324 | |
| PBLD | 3.781974 | 0.243387 | 0.999532 | | 5.486895 | 0.97313 | 0.113572 | | 4.977276 | 0.745807 | 0.375483 | |
| MXRA8 | 23.26756 | 0.38791 | 0.999532 | | 27.90811 | 0.542749 | 0.114317 | | 26.46857 | 0.167555 | 0.836453 | |
| LAMP1 | 17.00419 | 0.144813 | 0.999532 | | 23.17474 | 0.647839 | 0.115598 | | 20.32066 | 0.530927 | 0.335411 | |
| MATR3 | 7.925331 | 0.122131 | 0.999532 | | 11.19834 | 0.729385 | 0.115598 | | 9.728792 | 0.632145 | 0.281853 | |
| NFE2L3 | 7.260797 | -0.33173 | 0.999532 | | 7.122862 | -0.91284 | 0.115598 | | 5.166978 | -0.55134 | 0.619775 | |
| EMCN | 59.6195 | 0.033559 | 0.999532 | | 82.88481 | 0.597146 | 0.116884 | | 71.69075 | 0.597417 | 0.161146 | |
| ASAH1 | 13.22591 | 0.156012 | 0.999532 | | 17.50458 | 0.63723 | 0.118931 | | 15.50548 | 0.497891 | 0.386321 | |
| FKBP2 | 14.84573 | 0.345786 | 0.999532 | | 18.63679 | 0.598242 | 0.11945 | | 17.52721 | 0.275346 | 0.67176 | |
| PGRMC2 | 15.07187 | -0.2576 | 0.999532 | | 22.82182 | 0.561458 | 0.122725 | | 18.15793 | 0.836495 | 0.039624 | UP |
| CCDC144 | 5.865223 | -0.15474 | 0.999532 | | 5.449247 | -0.97315 | 0.122891 | | 4.359279 | -0.78844 | 0.397004 | |
| CALM2 | 12.92935 | 0.012645 | 0.999532 | | 18.48524 | 0.641452 | 0.122891 | | 15.7603 | 0.648317 | 0.204615 | |
| EEF2KMT | 5.764573 | -0.10778 | 0.999532 | | 5.081421 | -0.99602 | 0.122891 | | 4.287112 | -0.87432 | 0.299644 | |
| TIMP2 | 33.72206 | 0.088411 | 0.999532 | | 43.48271 | 0.495597 | 0.122891 | | 38.71118 | 0.432407 | 0.266933 | |
| CYB5A | 19.88339 | 0.360338 | 0.999532 | | 26.67271 | 0.770365 | 0.122891 | | 24.68638 | 0.452683 | 0.566812 | |
| FLNA | 37.55502 | -0.14471 | 0.999532 | | 57.93291 | 0.680604 | 0.122891 | | 47.58279 | 0.850828 | 0.068639 | UP |
| RPS26 | 27.58472 | -0.09147 | 0.999532 | | 27.35899 | -0.49657 | 0.122891 | | 22.17869 | -0.38204 | 0.404063 | |
| DPYSL3 | 7.122271 | 0.767933 | 0.727584 | | 8.262662 | 0.828696 | 0.123087 | | 8.765259 | 0.081327 | 0.947322 | |
| PEX6 | 3.953452 | -0.24763 | 0.999532 | | 3.639327 | -1.14004 | 0.123513 | | 2.692587 | -0.86894 | 0.443563 | |
| FAM43B | 3.99168 | -0.13998 | 0.999532 | | 3.478135 | -1.20203 | 0.123513 | | 2.686993 | -1.04029 | 0.314028 | |
| KRTAP9- | 8.311436 | 0.263254 | 0.999532 | | 6.390546 | -0.86416 | 0.123525 | | 6.63084 | -1.10508 | 0.107433 | |
| DDIT4 | 5.994038 | -0.9064 | 0.893099 | | 6.900781 | -0.97931 | 0.126523 | | 3.73242 | -0.06765 | 0.971732 | |
| FOSL2 | 26.79492 | -1.26144 | 0.019997 | DOWN | 32.21072 | -0.94191 | 0.126523 | | 16.65606 | 0.370697 | 0.652925 | |
| SUPT7L | 10.74373 | 0.203398 | 0.999532 | | 16.43552 | 1.020948 | 0.131387 | | 14.92752 | 0.793039 | 0.566812 | |
| ITGB8 | 15.77533 | 0.262163 | 0.999532 | | 20.35099 | 0.639119 | 0.132137 | | 18.72221 | 0.399446 | 0.491726 | |
| OR2L2 | 2.946846 | -0.28781 | 0.999532 | | 2.565107 | -1.39094 | 0.132741 | | 1.874847 | -1.08368 | 0.414154 | |
| CDH11 | 4.969074 | -0.05514 | 0.999532 | | 7.982151 | 0.801849 | 0.133546 | | 6.725115 | 0.875702 | 0.169993 | |
| GAGE10 | 31.59665 | 0.019707 | 0.999532 | | 27.44099 | -0.8602 | 0.133546 | | 23.67925 | -0.86029 | 0.121098 | |
| HR | 4.82464 | -0.33422 | 0.999532 | | 4.461664 | -1.00349 | 0.133546 | | 3.308446 | -0.65305 | 0.552672 | |
| CUBN | 2.951217 | 0.317375 | 0.999532 | | 4.595686 | 1.179071 | 0.133546 | | 4.157128 | 0.894946 | 0.421517 | |
| PLCG2 | 5.973375 | 1.060848 | 0.396029 | | 6.987085 | 1.11862 | 0.133546 | | 7.756328 | 0.103583 | 0.94861 | |
| KIAA031 | 2.718212 | -0.06018 | 0.999532 | | 4.419658 | 1.021266 | 0.133546 | | 3.775967 | 1.090715 | 0.210282 | |
| KCNJ16 | 3.309364 | 0.9634 | 0.999532 | | 3.815565 | 1.054642 | 0.133546 | | 4.189835 | 0.09605 | 0.955834 | |
| KMT2E | 4.85368 | -0.05289 | 0.999532 | | 7.328511 | 0.775429 | 0.135909 | | 6.312171 | 0.841922 | 0.235624 | |
| HSDL2 | 2.89757 | 0.549567 | 0.999532 | | 4.089265 | 1.012003 | 0.135909 | | 3.865344 | 0.474411 | 0.657538 | |
| GIPC1 | 4.851999 | 0.549845 | 0.999532 | | 6.063546 | 0.840576 | 0.135909 | | 6.140749 | 0.308764 | 0.750371 | |
| EIF4G2 | 17.41932 | 0.018174 | 0.999532 | | 23.84617 | 0.583091 | 0.135909 | | 20.84102 | 0.585524 | 0.203949 | |
| COL4A1 | 25.4139 | -0.34052 | 0.999532 | | 44.10261 | 0.832095 | 0.135909 | | 34.96252 | 1.188397 | 0.033324 | UP |
| DPYS | 3.337999 | 1.068676 | 0.713279 | | 3.816905 | 1.104062 | 0.135909 | | 4.299662 | 0.054982 | 0.976603 | |
| EVX1 | 6.577462 | -0.1704 | 0.999532 | | 6.016602 | -0.95424 | 0.135909 | | 4.71847 | -0.77039 | 0.344878 | |
| POLR2A | 4.701293 | 0.341537 | 0.999532 | | 6.781803 | 0.887089 | 0.135909 | | 5.993582 | 0.563104 | 0.542124 | |
| CTDSP2 | 11.20814 | 0.110926 | 0.999532 | | 15.49238 | 0.575786 | 0.136394 | | 13.38496 | 0.485072 | 0.392322 | |
| NFKBIL1 | 6.515577 | -0.42163 | 0.999532 | | 6.445318 | -0.90659 | 0.136394 | | 4.51815 | -0.46352 | 0.657538 | |
| UQCRB | 13.01349 | 0.166511 | 0.999532 | | 17.41827 | 0.603955 | 0.136394 | | 15.30014 | 0.462114 | 0.404063 | |
| SLC26A1 | 2.57699 | -0.42486 | 0.999532 | | 2.113409 | -1.49065 | 0.136394 | | 1.692011 | -1.04845 | 0.515762 | |
| ATAD3C | 4.294707 | -0.22513 | 0.999532 | | 3.816635 | -1.08567 | 0.136394 | | 2.925675 | -0.84231 | 0.424976 | |
| PDZK1IP | 11.70264 | 0.457277 | 0.999532 | | 15.57426 | 0.886949 | 0.136394 | | 14.77254 | 0.456947 | 0.663009 | |
| CD5 | 3.509192 | -0.65155 | 0.999532 | | 3.308733 | -1.15385 | 0.136394 | | 2.383942 | -0.50768 | 0.747457 | |
| HPN | 2.776568 | 0.511065 | 0.999532 | | 3.97882 | 1.067128 | 0.136394 | | 3.765599 | 0.58295 | 0.583371 | |
| HOXD4 | 5.503521 | 0.773049 | 0.912432 | | 6.664152 | 0.832656 | 0.136394 | | 6.816055 | 0.08425 | 0.946512 | |
| GINM1 | 8.877169 | 0.131043 | 0.999532 | | 12.11065 | 0.61088 | 0.136394 | | 10.70079 | 0.499185 | 0.366512 | |
| CTNNB1 | 14.15535 | -0.28165 | 0.999532 | | 21.44886 | 0.613141 | 0.136527 | | 17.32572 | 0.901201 | 0.052543 | UP |
| VGLL4 | 4.84048 | 0.211514 | 0.999532 | | 6.88131 | 0.805579 | 0.136527 | | 6.185499 | 0.613097 | 0.408514 | |
| ATP5F1B | 16.75942 | -0.01961 | 0.999532 | | 25.64881 | 0.747593 | 0.136888 | | 21.36647 | 0.800108 | 0.212897 | |
| ZC3H7B | 10.00539 | 0.484222 | 0.999532 | | 11.90948 | 0.645894 | 0.136888 | | 11.67047 | 0.178492 | 0.837256 | |
| RPN1 | 10.50327 | -0.01461 | 0.999532 | | 15.17667 | 0.658628 | 0.137988 | | 12.87368 | 0.692656 | 0.216108 | |
| ANXA2 | 35.75248 | 0.237216 | 0.999532 | | 46.2836 | 0.607984 | 0.137988 | | 41.90674 | 0.394915 | 0.546456 | |
| EID3 | 3.832213 | -0.18385 | 0.999532 | | 3.371754 | -1.10659 | 0.137988 | | 2.733196 | -0.90684 | 0.389782 | |
| OAS3 | 3.209097 | 0.817704 | 0.999532 | | 3.786195 | 1.060577 | 0.139027 | | 4.189145 | 0.254462 | 0.844873 | |
| GPS1 | 6.439739 | 0.188106 | 0.999532 | | 9.069401 | 0.718528 | 0.142459 | | 7.806352 | 0.534791 | 0.49299 | |
| RPS12 | 31.96171 | 0.20921 | 0.999532 | | 42.10099 | 0.625404 | 0.145349 | | 38.2099 | 0.432278 | 0.464519 | |
| DYNC1H1 | 17.83546 | 0.148163 | 0.999532 | | 24.11535 | 0.586939 | 0.145642 | | 21.02273 | 0.468674 | 0.396754 | |
| SGCA | 4.693362 | -0.2462 | 0.999532 | | 4.24141 | -1.0755 | 0.145934 | | 3.390352 | -0.80942 | 0.486743 | |
| CCDC86 | 4.702188 | 0.020797 | 0.999532 | | 4.031414 | -1.06682 | 0.146205 | | 3.508989 | -1.06562 | 0.243833 | |
| ZFP36L1 | 26.05434 | -0.24904 | 0.999532 | | 39.8467 | 0.619501 | 0.146205 | | 32.08029 | 0.880831 | 0.044685 | UP |
| IGFBP2 | 12.75398 | 0.328158 | 0.999532 | | 16.41133 | 0.696631 | 0.146205 | | 15.46673 | 0.378041 | 0.64142 | |
| ICMT | 4.894093 | 0.419586 | 0.999532 | | 6.455208 | 0.804299 | 0.146751 | | 6.101326 | 0.402066 | 0.652925 | |
| IFNA4 | 4.374086 | -0.12366 | 0.999532 | | 3.945078 | -1.05219 | 0.14745 | | 3.078138 | -0.90576 | 0.363418 | |
| TMED4 | 12.3087 | 0.075105 | 0.999532 | | 17.14872 | 0.635752 | 0.148351 | | 14.93522 | 0.581243 | 0.274496 | |
| ATP5MK | 7.177936 | 0.326523 | 0.999532 | | 9.505741 | 0.766949 | 0.149216 | | 8.959637 | 0.457654 | 0.563729 | |
| RNF138 | 9.170188 | -0.77859 | 0.657641 | | 10.60981 | -0.745 | 0.149216 | | 6.481409 | 0.048243 | 0.974421 | |
| SPRED2 | 5.30587 | 0.28634 | 0.999532 | | 7.307795 | 0.780691 | 0.151494 | | 6.724511 | 0.513872 | 0.515762 | |
| VIM | 91.10746 | 0.018411 | 0.999532 | | 131.8897 | 0.669988 | 0.151494 | | 112.8247 | 0.673253 | 0.15668 | |
| KLF9 | 5.977769 | -0.7507 | 0.999532 | | 6.170006 | -1.06491 | 0.156918 | | 3.611924 | -0.26514 | 0.83987 | |
| C19orf81 | 4.600282 | 0.183854 | 0.999532 | | 3.365824 | -1.13168 | 0.158378 | | 3.254108 | -1.30693 | 0.146093 | |
| MTMR2 | 3.448482 | 0.525354 | 0.999532 | | 4.661917 | 0.986795 | 0.158451 | | 4.486318 | 0.484916 | 0.640776 | |
| NDNF | 62.46507 | 0.233335 | 0.999532 | | 84.90106 | 0.706583 | 0.159502 | | 76.81978 | 0.50321 | 0.348787 | |
| SLC34A1 | 4.633217 | 0.525962 | 0.999532 | | 6.840122 | 1.109395 | 0.159502 | | 6.457537 | 0.628766 | 0.577942 | |
| SUCLG1 | 6.429924 | -0.08663 | 0.999532 | | 9.459137 | 0.722213 | 0.159763 | | 7.916593 | 0.812709 | 0.254511 | |
| BTG2 | 15.27962 | -0.99092 | 0.69276 | | 17.18425 | -0.96716 | 0.159763 | | 9.607049 | 0.119638 | 0.90113 | |
| RTN4 | 17.30043 | -0.03131 | 0.999532 | | 24.87595 | 0.62557 | 0.159763 | | 21.06924 | 0.674277 | 0.19389 | |
| NDUFB2 | 12.69764 | 0.350872 | 0.999532 | | 16.41879 | 0.662246 | 0.159763 | | 15.17404 | 0.330228 | 0.647218 | |
| MYLK | 15.1095 | 0.204171 | 0.999532 | | 19.76823 | 0.593433 | 0.159763 | | 17.8678 | 0.408752 | 0.524206 | |
| YPEL3 | 5.636199 | 0.182063 | 0.999532 | | 7.827587 | 0.700566 | 0.159763 | | 7.013841 | 0.538169 | 0.445382 | |
| MCTP1 | 3.373027 | -0.50337 | 0.999532 | | 3.366243 | -1.10847 | 0.160226 | | 2.197302 | -0.58321 | 0.673132 | |
| DST | 12.29088 | 0.394317 | 0.999532 | | 15.01964 | 0.567537 | 0.160226 | | 14.15509 | 0.194172 | 0.809832 | |
| CD6 | 4.022405 | -0.08725 | 0.999532 | | 3.415069 | -1.07122 | 0.160226 | | 2.781522 | -0.96765 | 0.333968 | |
| PRSS3 | 3.943408 | -0.56531 | 0.999532 | | 4.091892 | -1.06583 | 0.160226 | | 2.661294 | -0.4745 | 0.725266 | |
| DCN | 35.04455 | 0.523102 | 0.999532 | | 41.9003 | 0.658735 | 0.160226 | | 41.05872 | 0.145862 | 0.889853 | |
| PIGR | 5.223604 | 0.19181 | 0.999532 | | 8.738258 | 1.108863 | 0.160789 | | 7.697653 | 0.944964 | 0.323635 | |
| OS9 | 9.302662 | 0.341272 | 0.999532 | | 12.22359 | 0.729261 | 0.161025 | | 11.4294 | 0.410236 | 0.596177 | |
| THAP12 | 9.277631 | 0.084918 | 0.999532 | | 7.805284 | -0.92167 | 0.161025 | | 7.02989 | -1.00029 | 0.154583 | |
| GPX8 | 2.960041 | 0.752409 | 0.999532 | | 3.529672 | 1.05178 | 0.161025 | | 3.769928 | 0.308355 | 0.810753 | |
| MEIS2 | 11.9876 | 0.029785 | 0.999532 | | 16.86422 | 0.657576 | 0.161025 | | 14.84382 | 0.660711 | 0.154583 | |
| WWP1 | 3.938454 | 0.68961 | 0.999532 | | 5.124907 | 0.905077 | 0.161025 | | 5.218004 | 0.236493 | 0.838179 | |
| ARHGEF | 10.82219 | 0.276038 | 0.999532 | | 14.308 | 0.68587 | 0.161059 | | 13.09171 | 0.436006 | 0.49299 | |
| EPHX2 | 3.822641 | 0.955705 | 0.999532 | | 4.373791 | 0.977425 | 0.161059 | | 4.767032 | 0.032268 | 0.988853 | |
| PDK4 | 7.66595 | -0.96179 | 0.999532 | | 8.660373 | -1.04184 | 0.162571 | | 4.600616 | -0.10845 | 0.940744 | |
| CCL21 | 3.179223 | 0.931426 | 0.999532 | | 4.602541 | 1.588146 | 0.162914 | | 5.066516 | 0.590963 | 0.743246 | |
| TUBB | 14.50231 | -0.02783 | 0.999532 | | 22.00851 | 0.712586 | 0.162914 | | 18.44256 | 0.773466 | 0.183855 | |
| NUTM2F | 6.707532 | -0.0565 | 0.999532 | | 5.734283 | -0.93757 | 0.162914 | | 4.823057 | -0.85317 | 0.300852 | |
| PICALM | 7.84413 | -0.05325 | 0.999532 | | 11.9409 | 0.688366 | 0.16414 | | 9.884485 | 0.753191 | 0.199116 | |
| FAM227B | 4.548764 | -0.63177 | 0.999532 | | 4.620631 | -0.95656 | 0.166112 | | 2.956271 | -0.30652 | 0.828869 | |
| RPS13 | 12.98545 | 0.146291 | 0.999532 | | 17.19653 | 0.569152 | 0.166112 | | 15.08542 | 0.449083 | 0.442674 | |
| CCL4L2 | 12.18309 | -0.85583 | 0.999532 | | 12.61175 | -1.09172 | 0.166381 | | 7.68391 | -0.19961 | 0.894612 | |
| DCUN1D1 | 6.525418 | 0.420431 | 0.999532 | | 7.923998 | 0.714629 | 0.168331 | | 7.861127 | 0.317307 | 0.702463 | |
| ARFGEF | 3.873782 | 0.761623 | 0.999532 | | 4.57785 | 0.892017 | 0.168893 | | 4.887968 | 0.148325 | 0.903339 | |
| ZBTB16 | 12.00186 | -1.47946 | 0.256559 | | 13.44327 | -1.51765 | 0.169568 | | 5.877788 | 0.014986 | 0.996681 | |
| GOLGA8 | 6.37235 | -0.11797 | 0.999532 | | 6.129913 | -0.83652 | 0.170125 | | 4.773197 | -0.68382 | 0.434766 | |
| DPP4 | 3.500096 | 0.266145 | 0.999532 | | 5.193513 | 0.993602 | 0.170746 | | 4.584728 | 0.758684 | 0.506666 | |
| CTTN | 11.67931 | -0.02484 | 0.999532 | | 16.99584 | 0.635149 | 0.170746 | | 14.24372 | 0.685322 | 0.15668 | |
| CSH2 | 6.840027 | 0.048037 | 0.999532 | | 5.941753 | -1.06771 | 0.170746 | | 4.892783 | -1.10009 | 0.139095 | |
| USF2 | 7.057421 | -0.10916 | 0.999532 | | 10.72152 | 0.653768 | 0.170835 | | 8.991352 | 0.784797 | 0.145899 | |
| NQO2 | 3.466268 | 0.27515 | 0.999532 | | 5.029872 | 0.859452 | 0.174818 | | 4.616198 | 0.607469 | 0.531575 | |
| GOLM2 | 9.106866 | 0.155905 | 0.999532 | | 11.63379 | 0.606578 | 0.176244 | | 10.46608 | 0.476733 | 0.421517 | |
| CNN2 | 17.18225 | 0.187185 | 0.999532 | | 22.58494 | 0.56923 | 0.1766 | | 20.42935 | 0.3993 | 0.442693 | |
| MYL12A | 28.94559 | 0.095375 | 0.999532 | | 40.85993 | 0.676767 | 0.1766 | | 35.84254 | 0.602235 | 0.310637 | |
| MMP9 | 3.346173 | -0.92257 | 0.999532 | | 3.367984 | -1.12021 | 0.1766 | | 2.202694 | -0.15407 | 0.941197 | |
| REEP5 | 10.81515 | 0.145133 | 0.999532 | | 15.6699 | 0.733743 | 0.177469 | | 13.52459 | 0.617977 | 0.436389 | |
| TIMP1 | 15.2975 | 0.110138 | 0.999532 | | 24.62779 | 0.999708 | 0.17766 | | 21.69011 | 0.90774 | 0.15668 | |
| PCK1 | 5.587372 | 0.616991 | 0.999532 | | 7.591339 | 1.036105 | 0.17802 | | 7.511358 | 0.449518 | 0.718643 | |
| SP7 | 7.875386 | 0.006043 | 0.999532 | | 6.687558 | -0.83311 | 0.178395 | | 6.250064 | -0.82449 | 0.335411 | |
| GNLY | 3.946997 | -0.82789 | 0.999532 | | 4.153453 | -1.05127 | 0.178618 | | 2.803314 | -0.20009 | 0.915533 | |
| ITPRIP | 4.723146 | -0.83487 | 0.999532 | | 5.014621 | -1.12497 | 0.178618 | | 2.757403 | -0.2826 | 0.855088 | |
| ZNF66 | 13.85256 | 0.011269 | 0.999532 | | 12.85457 | -0.77708 | 0.178618 | | 10.86782 | -0.75737 | 0.266797 | |
| ELOVL5 | 28.3892 | -0.28316 | 0.999532 | | 29.27981 | -0.64655 | 0.178618 | | 22.04998 | -0.32807 | 0.67937 | |
| NPY1R | 4.159813 | 0.575685 | 0.999532 | | 5.298307 | 0.912685 | 0.178618 | | 5.284197 | 0.353725 | 0.737952 | |
| LYRM2 | 106.0797 | 0.128511 | 0.999532 | | 97.05871 | -0.5861 | 0.178618 | | 87.27439 | -0.68688 | 0.150518 | |
| CLEC18A | 3.806554 | -0.6137 | 0.999532 | | 3.903739 | -1.01863 | 0.178618 | | 2.476309 | -0.38265 | 0.79686 | |
| TMEM248 | 9.44112 | 0.08349 | 0.999532 | | 12.97237 | 0.606933 | 0.178618 | | 11.33445 | 0.544804 | 0.320564 | |
| SZRD1 | 4.056999 | -0.02149 | 0.999532 | | 5.973171 | 0.821267 | 0.178618 | | 5.166902 | 0.853221 | 0.271669 | |
| SMARCC | 7.870171 | 0.321118 | 0.999532 | | 10.27868 | 0.664512 | 0.178618 | | 9.18591 | 0.338752 | 0.658435 | |
| EIF3I | 5.201035 | -0.04898 | 0.999532 | | 7.950115 | 0.7262 | 0.179421 | | 6.608308 | 0.794638 | 0.233094 | |
| SPDYE16 | 43.13038 | -0.23234 | 0.999532 | | 42.82819 | -0.70696 | 0.179421 | | 33.09067 | -0.44412 | 0.560424 | |
| NUDT21 | 4.284043 | 0.469549 | 0.999532 | | 5.660757 | 0.858738 | 0.179421 | | 5.328064 | 0.402851 | 0.699971 | |
| CLIC4 | 13.87089 | -0.17913 | 0.999532 | | 20.57841 | 0.596426 | 0.179421 | | 17.02857 | 0.785981 | 0.156457 | |
| GALNT11 | 3.836625 | 0.131722 | 0.999532 | | 5.781938 | 0.839412 | 0.179421 | | 4.969135 | 0.718368 | 0.397004 | |
| PIMREG | 3.689036 | -0.10814 | 0.999532 | | 3.146288 | -1.1577 | 0.179421 | | 2.611648 | -1.03374 | 0.325475 | |
| C7 | 19.59769 | 0.371925 | 0.999532 | | 27.2949 | 0.88866 | 0.179421 | | 25.52063 | 0.533149 | 0.707142 | |
| UBE2Q2L | 3.897143 | -0.64321 | 0.999532 | | 4.137759 | -0.95838 | 0.179421 | | 2.615108 | -0.29015 | 0.859898 | |
| KIR2DS1 | 5.149706 | -0.31672 | 0.999532 | | 4.849438 | -0.94464 | 0.179421 | | 3.756162 | -0.61388 | 0.577942 | |
| SLC7A8 | 7.110056 | 0.451762 | 0.999532 | | 9.163274 | 0.720261 | 0.179421 | | 8.514239 | 0.287112 | 0.74256 | |
| NFASC | 12.26003 | 0.574845 | 0.999532 | | 14.62352 | 0.719783 | 0.179421 | | 14.37553 | 0.171351 | 0.864268 | |
| EIF2S3B | 11.21582 | 0.180479 | 0.999532 | | 15.10766 | 0.714741 | 0.179421 | | 13.60405 | 0.541038 | 0.49299 | |
| USP22 | 10.37842 | -0.03689 | 0.999532 | | 14.15088 | 0.555254 | 0.179421 | | 12.0589 | 0.603228 | 0.271669 | |
| HINT1 | 15.01971 | 0.123991 | 0.999532 | | 20.13161 | 0.604709 | 0.179421 | | 17.80797 | 0.498586 | 0.374949 | |
| RRP36 | 3.218033 | 0.023626 | 0.999532 | | 5.04004 | 0.871842 | 0.179421 | | 4.320451 | 0.866512 | 0.283338 | |
| TMPRSS1 | 2.844302 | -0.48212 | 0.999532 | | 2.771222 | -1.21253 | 0.179421 | | 1.90416 | -0.70105 | 0.652925 | |
| DPEP1 | 14.7638 | 0.393025 | 0.999532 | | 19.52194 | 0.792917 | 0.179421 | | 18.1532 | 0.408791 | 0.723933 | |
| NECTIN2 | 13.53482 | 0.216316 | 0.999532 | | 17.79983 | 0.506438 | 0.179421 | | 15.53658 | 0.321045 | 0.629006 | |
| COL1A2 | 9.263304 | -0.05677 | 0.999532 | | 14.57655 | 0.818532 | 0.179421 | | 12.24679 | 0.899663 | 0.148529 | |
| PDZK1 | 7.075075 | 0.565063 | 0.999532 | | 9.829079 | 1.026324 | 0.180486 | | 9.530393 | 0.507292 | 0.737952 | |
| PTTG1 | 5.609386 | -0.13775 | 0.999532 | | 4.96332 | -0.87029 | 0.180838 | | 4.117455 | -0.71806 | 0.442315 | |
| CD81 | 83.89111 | -0.17124 | 0.999532 | | 116.5903 | 0.412696 | 0.180944 | | 94.41092 | 0.607233 | 0.017097 | UP |
| MMP2 | 6.415413 | 0.025369 | 0.999532 | | 9.305175 | 0.752729 | 0.180944 | | 8.030805 | 0.751935 | 0.249649 | |
| LIMS3 | 13.00189 | -0.19789 | 0.999532 | | 12.8354 | -0.61296 | 0.182486 | | 10.03842 | -0.38972 | 0.563285 | |
| C17orf67 | 2.41771 | -0.35202 | 0.999532 | | 2.224485 | -1.38879 | 0.182867 | | 1.542213 | -1.01334 | 0.515762 | |
| ACAT1 | 5.247289 | 0.831617 | 0.871349 | | 5.904415 | 0.890368 | 0.185393 | | 6.574794 | 0.065689 | 0.965649 | |
| ECHS1 | 10.44586 | 0.111743 | 0.999532 | | 14.75629 | 0.755418 | 0.185623 | | 13.14683 | 0.649626 | 0.414073 | |
| LAT | 15.72989 | 0.088177 | 0.999532 | | 14.20279 | -0.64692 | 0.185623 | | 12.35517 | -0.71076 | 0.150518 | |
| C18orf32 | 5.407484 | 0.105787 | 0.999532 | | 7.834916 | 0.676582 | 0.185623 | | 6.61477 | 0.591721 | 0.421517 | |
| TMEM176 | 3.735847 | 0.433346 | 0.999532 | | 4.96349 | 0.991425 | 0.186461 | | 4.703169 | 0.587934 | 0.570037 | |
| SPTAN1 | 34.60817 | 0.187211 | 0.999532 | | 42.80461 | 0.455985 | 0.186461 | | 38.54781 | 0.285732 | 0.632113 | |
| WASL | 12.07461 | 0.114773 | 0.999532 | | 16.37309 | 0.612865 | 0.187689 | | 14.22089 | 0.521414 | 0.453011 | |
| MTHFS | 3.034374 | 0.670057 | 0.999532 | | 3.722144 | 1.008058 | 0.189346 | | 3.868613 | 0.353449 | 0.783005 | |
| YWHAB | 5.365755 | 0.014453 | 0.999532 | | 8.178867 | 0.668701 | 0.189605 | | 6.894732 | 0.676507 | 0.362447 | |
| PTPRCAP | 4.489727 | -0.04206 | 0.999532 | | 3.927436 | -0.96961 | 0.190626 | | 3.318243 | -0.90747 | 0.359441 | |
| S100A16 | 5.608442 | 0.2279 | 0.999532 | | 7.955472 | 0.845456 | 0.190714 | | 7.215459 | 0.637942 | 0.462479 | |
| TMEM154 | 4.392322 | -0.05532 | 0.999532 | | 3.771324 | -1.06344 | 0.190714 | | 3.040299 | -0.98978 | 0.319186 | |
| BNIP2 | 12.89637 | -0.09046 | 0.999532 | | 17.85313 | 0.48156 | 0.190714 | | 14.63588 | 0.589261 | 0.209531 | |
| RTP5 | 3.605123 | -0.34581 | 0.999532 | | 3.297233 | -1.06118 | 0.190714 | | 2.475507 | -0.70193 | 0.584522 | |
| USP33 | 3.676431 | 0.410857 | 0.999532 | | 4.728586 | 0.876694 | 0.190714 | | 4.724608 | 0.500394 | 0.632113 | |
| SPATA31 | 3.252747 | -0.53227 | 0.999532 | | 3.239238 | -1.20037 | 0.190714 | | 2.065452 | -0.65162 | 0.660097 | |
| NPHS2 | 38.00758 | 0.414946 | 0.999532 | | 48.0907 | 0.700929 | 0.190714 | | 46.03651 | 0.309055 | 0.652114 | |
| WT1 | 5.476644 | 0.538369 | 0.999532 | | 6.581368 | 0.787586 | 0.190714 | | 6.565074 | 0.265199 | 0.800239 | |
| FAM153B | 4.347038 | -0.48421 | 0.999532 | | 4.281177 | -0.99592 | 0.190796 | | 3.09769 | -0.49969 | 0.711733 | |
| TPM2 | 14.38449 | 0.222341 | 0.999532 | | 19.08701 | 0.647541 | 0.192151 | | 17.57679 | 0.436244 | 0.515762 | |
| GNG14 | 2.459368 | -0.95755 | 0.999532 | | 2.505903 | -1.25567 | 0.192151 | | 1.489516 | -0.28076 | 0.900074 | |
| BCKDHB | 5.117566 | -0.12198 | 0.999532 | | 4.644622 | -0.90604 | 0.192151 | | 3.488021 | -0.76874 | 0.421517 | |
| ZCCHC10 | 7.606128 | -0.054 | 0.999532 | | 7.083275 | -0.74307 | 0.192151 | | 5.735448 | -0.67058 | 0.366512 | |
| ZNF699 | 2.20001 | 0.45471 | 0.999532 | | 3.469026 | 1.025447 | 0.192151 | | 3.204439 | 0.590912 | 0.617953 | |
| ZNF705A | 11.12554 | -0.34323 | 0.999532 | | 11.49411 | -0.79419 | 0.193658 | | 8.400791 | -0.4079 | 0.686704 | |
| INTS3 | 9.379457 | 0.300926 | 0.999532 | | 11.72169 | 0.620263 | 0.193697 | | 10.96837 | 0.335821 | 0.660958 | |
| FOSL1 | 5.025799 | 0.080516 | 0.999532 | | 4.114589 | -0.97821 | 0.195072 | | 3.714932 | -1.03685 | 0.254225 | |
| COX7B | 8.327046 | -0.00758 | 0.999532 | | 11.52684 | 0.594434 | 0.196365 | | 9.843414 | 0.615336 | 0.300852 | |
| SEC31A | 6.797165 | -0.08928 | 0.999532 | | 9.682003 | 0.713247 | 0.196365 | | 8.389449 | 0.781218 | 0.274496 | |
| GOLGA6 | 10.06029 | 0.081281 | 0.999532 | | 8.824386 | -0.71866 | 0.198982 | | 7.813882 | -0.77952 | 0.226707 | |
| CTBP1 | 10.7049 | 0.229582 | 0.999532 | | 13.4931 | 0.553563 | 0.199079 | | 12.28757 | 0.341482 | 0.614824 | |
| MFSD2A | 2.792037 | -0.47539 | 0.999532 | | 2.859124 | -1.1659 | 0.199237 | | 1.913502 | -0.66043 | 0.662584 | |
| KIR3DL1 | 26.73357 | -0.06074 | 0.999532 | | 24.91622 | -0.71301 | 0.20012 | | 20.70557 | -0.62881 | 0.342845 | |
| RCN2 | 6.840583 | 0.257538 | 0.999532 | | 9.198182 | 0.636924 | 0.20012 | | 8.447771 | 0.397756 | 0.578893 | |
| IGHG3 | 8.36231 | 0.169047 | 0.999532 | | 10.01776 | -1.59654 | 0.20012 | | 6.128343 | -0.86797 | 0.286239 | |
| SFPQ | 18.57682 | -0.05656 | 0.999532 | | 24.99399 | 0.515553 | 0.202044 | | 21.10733 | 0.580927 | 0.253256 | |
| LRRC32 | 8.210108 | -0.32475 | 0.999532 | | 12.67219 | 0.61443 | 0.202597 | | 10.39622 | 0.972551 | 0.054952 | UP |
| COX6B1 | 19.51088 | 0.207558 | 0.999532 | | 24.94164 | 0.546039 | 0.203839 | | 22.49398 | 0.362142 | 0.60629 | |
| HSPG2 | 8.160688 | -0.21383 | 0.999532 | | 13.55754 | 0.797547 | 0.204306 | | 11.1955 | 1.025404 | 0.075829 | UP |
| ORSL1 | 8.13322 | -0.04387 | 0.999532 | | 7.54232 | -0.79849 | 0.204306 | | 6.474765 | -0.74729 | 0.348004 | |
| ZFAND3 | 8.337884 | 0.005731 | 0.999532 | | 12.10919 | 0.614031 | 0.204306 | | 10.32031 | 0.622912 | 0.274496 | |
| TCP10L3 | 3.053609 | -0.48966 | 0.999532 | | 2.845195 | -1.11544 | 0.204306 | | 2.210603 | -0.61334 | 0.679818 | |
| SLC39A1 | 5.603889 | -0.05105 | 0.999532 | | 8.134339 | 0.686284 | 0.204306 | | 6.918768 | 0.756383 | 0.241605 | |
| SSX3 | 6.816743 | 0.154054 | 0.999532 | | 5.652195 | -0.9027 | 0.204306 | | 5.080809 | -1.0305 | 0.19389 | |
| ENG | 37.54925 | -0.14672 | 0.999532 | | 54.19302 | 0.540214 | 0.204306 | | 44.75431 | 0.717589 | 0.027734 | UP |
| OR1E1 | 3.753851 | -0.73835 | 0.999532 | | 3.870942 | -1.08598 | 0.204306 | | 2.436707 | -0.33172 | 0.858705 | |
| RPS16 | 16.45366 | 0.158102 | 0.999532 | | 20.71597 | 0.556877 | 0.204306 | | 18.88745 | 0.403664 | 0.499025 | |
| DAB2 | 5.905926 | 0.09391 | 0.999532 | | 8.594284 | 0.784193 | 0.204306 | | 7.370624 | 0.720703 | 0.344798 | |
| SPINT2 | 7.814522 | 0.577798 | 0.999532 | | 9.220638 | 0.654692 | 0.204306 | | 9.085481 | 0.094317 | 0.931155 | |
| HSPB1 | 9.080484 | 0.083947 | 0.999532 | | 12.6146 | 0.675876 | 0.204306 | | 11.11162 | 0.605813 | 0.348787 | |
| ZNF207 | 12.89551 | 0.065137 | 0.999532 | | 17.21691 | 0.538541 | 0.204306 | | 14.92723 | 0.490217 | 0.439373 | |
| TRAM1 | 10.98356 | -0.04658 | 0.999532 | | 15.86181 | 0.598587 | 0.204306 | | 13.38665 | 0.665024 | 0.209531 | |
| MYO1D | 8.766532 | -0.01344 | 0.999532 | | 12.24177 | 0.555386 | 0.204306 | | 10.28235 | 0.586442 | 0.325475 | |
| RPS23 | 36.44922 | 0.056676 | 0.999532 | | 48.67841 | 0.521962 | 0.204306 | | 42.29757 | 0.484629 | 0.340167 | |
| NRSN2 | 8.853944 | 0.080734 | 0.999532 | | 7.931267 | -0.69958 | 0.204306 | | 6.934781 | -0.75692 | 0.266797 | |
| NOTCH1 | 7.021491 | 0.12169 | 0.999532 | | 9.805924 | 0.682466 | 0.204306 | | 8.75289 | 0.578473 | 0.399498 | |
| ACTN4 | 96.84768 | 0.31643 | 0.999532 | | 115.8722 | 0.475867 | 0.204306 | | 108.7255 | 0.185221 | 0.714657 | |
| NHLRC1 | 3.521631 | -0.42673 | 0.999532 | | 3.515944 | -0.99068 | 0.204306 | | 2.491861 | -0.54235 | 0.67937 | |
| DEPDC1 | 3.796242 | -0.03171 | 0.999532 | | 3.408728 | -1.05993 | 0.204516 | | 2.82594 | -0.99401 | 0.359638 | |
| DXO | 3.296876 | 0.769429 | 0.999532 | | 3.785285 | 0.93725 | 0.205544 | | 4.080648 | 0.185692 | 0.89142 | |
| LIMCH1 | 6.513546 | 0.532898 | 0.999532 | | 7.417165 | 0.678623 | 0.205613 | | 7.627545 | 0.165021 | 0.864923 | |
| MYC | 4.21485 | -0.56018 | 0.999532 | | 4.251836 | -0.93985 | 0.205828 | | 2.898816 | -0.36286 | 0.791576 | |
| CCL3L3 | 4.044312 | -0.98381 | 0.999532 | | 4.280463 | -1.07801 | 0.206568 | | 2.403627 | -0.04094 | 0.988853 | |
| CCDC198 | 10.31816 | 0.402776 | 0.999532 | | 13.66762 | 0.813385 | 0.206568 | | 12.70922 | 0.457537 | 0.632113 | |
| HNRNPK | 19.65502 | 0.163576 | 0.999532 | | 25.68805 | 0.553157 | 0.206568 | | 22.74085 | 0.415662 | 0.49299 | |
| RNF130 | 4.610791 | 0.241845 | 0.999532 | | 6.297742 | 0.773111 | 0.206568 | | 5.691561 | 0.547638 | 0.530145 | |
| MDH1 | 5.558231 | 0.362316 | 0.999532 | | 7.528676 | 0.831557 | 0.20909 | | 6.950261 | 0.497467 | 0.579064 | |
| SPCS1 | 10.57052 | -0.0115 | 0.999532 | | 14.72061 | 0.544377 | 0.20909 | | 12.33502 | 0.575755 | 0.247358 | |
| WDR72 | 3.852472 | 0.548426 | 0.999532 | | 4.894623 | 0.922431 | 0.212902 | | 4.822914 | 0.380913 | 0.765852 | |
| C1orf198 | 5.170568 | 0.498198 | 0.999532 | | 6.321254 | 0.758075 | 0.212939 | | 6.152886 | 0.271727 | 0.803337 | |
| VAMP8 | 8.773788 | -0.12945 | 0.999532 | | 12.98491 | 0.5498 | 0.212939 | | 10.54655 | 0.699478 | 0.168532 | |
| COL4A2 | 39.65115 | -0.19844 | 0.999532 | | 61.62451 | 0.687537 | 0.212939 | | 50.31459 | 0.899709 | 0.107433 | |
| CPNE5 | 4.689861 | -0.28744 | 0.999532 | | 4.457785 | -0.86804 | 0.212939 | | 3.4599 | -0.56003 | 0.647581 | |
| LRRC14 | 5.794806 | -0.4013 | 0.999532 | | 5.722982 | -0.83219 | 0.212939 | | 4.109119 | -0.41077 | 0.708936 | |
| ATP6V0C | 9.760077 | 0.29865 | 0.999532 | | 12.39935 | 0.600129 | 0.212939 | | 11.10675 | 0.33112 | 0.668158 | |
| VIPR1 | 6.875723 | -0.51876 | 0.999532 | | 7.15028 | -0.81819 | 0.212939 | | 5.138346 | -0.27818 | 0.830822 | |
| GATM | 13.67184 | 0.110726 | 0.999532 | | 23.28898 | 1.105507 | 0.212939 | | 20.21561 | 1.014995 | 0.49299 | |
| LMO1 | 7.25266 | -0.27991 | 0.999532 | | 6.803598 | -0.92256 | 0.212939 | | 5.179408 | -0.61754 | 0.543211 | |
| STK38 | 2.915546 | 0.338553 | 0.999532 | | 4.333379 | 0.89259 | 0.212939 | | 3.890005 | 0.579556 | 0.578798 | |
| BSG | 25.48112 | 0.152109 | 0.999532 | | 34.13064 | 0.591394 | 0.213335 | | 30.03068 | 0.468312 | 0.49299 | |
| CCNY | 5.42538 | -0.17164 | 0.999532 | | 8.692098 | 0.693948 | 0.213604 | | 7.133508 | 0.881079 | 0.150518 | |
| FBXL3 | 7.062141 | 0.351101 | 0.999532 | | 9.010085 | 0.665203 | 0.214958 | | 8.532704 | 0.335003 | 0.695252 | |
| CDKN1A | 5.100049 | -1.68975 | 0.017231 | DOWN | 6.884637 | -0.90647 | 0.215644 | | 3.129922 | 0.790854 | 0.49299 | |
| GPHN | 5.3889 | 0.528936 | 0.999532 | | 6.587918 | 0.756692 | 0.216754 | | 6.388899 | 0.240377 | 0.826947 | |
| GABARA | 6.54171 | 0.104728 | 0.999532 | | 9.307407 | 0.633045 | 0.216754 | | 8.083402 | 0.550485 | 0.426561 | |
| NDUFB6 | 7.724698 | 0.318535 | 0.999532 | | 9.56688 | 0.610818 | 0.216754 | | 8.99313 | 0.309859 | 0.68597 | |
| RRAGA | 6.178601 | 0.570066 | 0.999532 | | 7.554306 | 0.700452 | 0.219221 | | 7.65918 | 0.146616 | 0.88885 | |
| PRCP | 11.12066 | 0.146001 | 0.999532 | | 14.83302 | 0.591479 | 0.21965 | | 13.20519 | 0.464679 | 0.523065 | |
| RPL19 | 39.42084 | 0.149245 | 0.999532 | | 50.38006 | 0.488598 | 0.220224 | | 44.88315 | 0.358919 | 0.541136 | |
| RPS5 | 15.67254 | -0.07519 | 0.999532 | | 22.71489 | 0.576409 | 0.220224 | | 18.74828 | 0.683136 | 0.204615 | |
| UQCRHL | 10.53623 | 0.529131 | 0.999532 | | 11.99459 | 0.603496 | 0.220224 | | 12.03159 | 0.098666 | 0.913136 | |
| NCAN | 2.893905 | 0.11695 | 0.999532 | | 2.271398 | -1.25675 | 0.220224 | | 2.155703 | -1.34015 | 0.283785 | |
| SKIL | 6.875416 | -0.20427 | 0.999532 | | 10.16543 | 0.589627 | 0.22044 | | 8.267734 | 0.811275 | 0.148529 | |
| CHRDL1 | 5.044402 | 0.381608 | 0.999532 | | 6.953592 | 0.815269 | 0.220763 | | 6.658884 | 0.459181 | 0.666566 | |
| B3GNT3 | 2.871339 | -0.22838 | 0.999532 | | 2.606398 | -1.16739 | 0.221077 | | 1.908362 | -0.9182 | 0.512069 | |
| KRTAP5- | 2.698625 | -0.45072 | 0.999532 | | 2.53047 | -1.2329 | 0.221077 | | 1.896803 | -0.76129 | 0.64142 | |
| IFNAR1 | 8.844978 | 0.289463 | 0.999532 | | 10.86775 | 0.611869 | 0.221307 | | 10.4701 | 0.330647 | 0.682289 | |
| FGF1 | 75.31568 | 0.476262 | 0.999532 | | 86.12674 | 0.523778 | 0.22262 | | 84.90146 | 0.074262 | 0.932589 | |
| MRC2 | 3.82172 | 0.048338 | 0.999532 | | 5.523877 | 0.790165 | 0.223021 | | 4.967591 | 0.762253 | 0.344633 | |
| SUPT4H1 | 5.603981 | 0.412211 | 0.999532 | | 7.263069 | 0.725223 | 0.223021 | | 6.902931 | 0.334194 | 0.728126 | |
| TTC3 | 3.941586 | 0.403347 | 0.999532 | | 5.136081 | 0.825616 | 0.223021 | | 4.94242 | 0.443505 | 0.647581 | |
| ZNF720 | 3.849685 | -0.10293 | 0.999532 | | 3.318952 | -1.06296 | 0.223021 | | 2.8803 | -0.94027 | 0.442274 | |
| DRAXIN | 4.828558 | -0.09592 | 0.999532 | | 4.471147 | -0.94888 | 0.223021 | | 3.657294 | -0.81534 | 0.479746 | |
| MGMT | 3.377564 | 0.785667 | 0.999532 | | 4.035238 | 0.872409 | 0.223021 | | 4.21365 | 0.103915 | 0.94366 | |
| PDXK | 6.21169 | 0.1751 | 0.999532 | | 8.48672 | 0.732261 | 0.223021 | | 7.704791 | 0.565298 | 0.508751 | |
| MMP14 | 6.057077 | -0.00261 | 0.999532 | | 9.003765 | 0.640318 | 0.223235 | | 7.543633 | 0.663722 | 0.32832 | |
| MRPS18B | 6.827324 | 0.345645 | 0.999532 | | 8.54179 | 0.648949 | 0.223235 | | 8.169631 | 0.32692 | 0.699971 | |
| ARL6IP4 | 23.59703 | 0.170918 | 0.999532 | | 29.76836 | 0.432173 | 0.223707 | | 26.56754 | 0.287489 | 0.619775 | |
| UPK3A | 3.420479 | 0.04704 | 0.999532 | | 2.793457 | -1.18446 | 0.224776 | | 2.610527 | -1.22638 | 0.266933 | |
| GUK1 | 7.8436 | 0.214288 | 0.999532 | | 10.23102 | 0.589538 | 0.224776 | | 9.179044 | 0.394817 | 0.610507 | |
| GRN | 8.269883 | -0.0519 | 0.999532 | | 12.77141 | 0.685784 | 0.226263 | | 10.51364 | 0.763233 | 0.266933 | |
| EOMES | 3.035619 | -0.00295 | 0.999532 | | 4.728924 | 0.853293 | 0.22634 | | 3.961186 | 0.874191 | 0.312795 | |
| TLR9 | 3.64226 | 0.149741 | 0.999532 | | 3.09452 | -1.06261 | 0.22634 | | 2.674274 | -1.18769 | 0.224727 | |
| TMBIM6 | 30.23512 | 0.093755 | 0.999532 | | 40.37144 | 0.537742 | 0.226666 | | 35.07899 | 0.469102 | 0.515762 | |
| TPI1 | 11.47693 | 0.226613 | 0.999532 | | 14.42334 | 0.542134 | 0.227686 | | 13.27239 | 0.336853 | 0.595783 | |
| BGN | 90.48458 | -0.13895 | 0.999532 | | 124.2246 | 0.422122 | 0.227765 | | 102.2513 | 0.5911 | 0.083797 | UP |
| PRKCSH | 13.84582 | 0.166891 | 0.999532 | | 18.29785 | 0.620367 | 0.227765 | | 16.44622 | 0.469196 | 0.515762 | |
| KIAA110 | 2.775903 | 0.693093 | 0.999532 | | 3.32716 | 0.950789 | 0.2281 | | 3.544943 | 0.277318 | 0.837927 | |
| ITGAL | 3.489988 | -0.17949 | 0.999532 | | 3.155209 | -0.98662 | 0.229782 | | 2.557139 | -0.7893 | 0.512069 | |
| MAOB | 3.837786 | 0.735439 | 0.999532 | | 4.748952 | 0.885327 | 0.229782 | | 4.816003 | 0.185287 | 0.892646 | |
| UBE2E2 | 6.335464 | 0.395292 | 0.999532 | | 8.053354 | 0.754811 | 0.229782 | | 7.698512 | 0.388128 | 0.647581 | |
| MRPS21 | 5.922858 | 0.581574 | 0.999532 | | 7.257706 | 0.693502 | 0.229782 | | 7.244225 | 0.131324 | 0.902398 | |
| BCL2L12 | 4.685389 | -0.22193 | 0.999532 | | 4.566744 | -0.89677 | 0.230206 | | 3.348136 | -0.65204 | 0.542124 | |
| IER3IP1 | 5.544044 | 0.463213 | 0.999532 | | 6.806377 | 0.733252 | 0.230206 | | 6.63292 | 0.287636 | 0.765852 | |
| DUS3L | 4.949232 | 0.26352 | 0.999532 | | 3.694074 | -0.94015 | 0.232857 | | 3.697468 | -1.19427 | 0.15668 | |
| VPS29 | 8.456484 | 0.275669 | 0.999532 | | 10.39673 | 0.586955 | 0.234513 | | 9.76597 | 0.328107 | 0.659545 | |
| METAP2 | 2.551597 | 0.554251 | 0.999532 | | 3.273758 | 0.961227 | 0.234588 | | 3.283353 | 0.42474 | 0.72271 | |
| CAPN2 | 13.99408 | 0.169371 | 0.999532 | | 18.76268 | 0.633373 | 0.236608 | | 16.62932 | 0.499075 | 0.396743 | |
| SHTN1 | 2.835358 | 0.296209 | 0.999532 | | 3.865581 | 0.866802 | 0.236665 | | 3.714953 | 0.589223 | 0.560699 | |
| GEM | 4.065152 | -0.42664 | 0.999532 | | 7.390224 | 0.865916 | 0.236665 | | 5.762796 | 1.308391 | 0.121156 | |
| IGHG4 | 14.08738 | -0.21839 | 0.999532 | | 11.77698 | -1.25048 | 0.236678 | | 8.613113 | -0.9141 | 0.377051 | |
| NPC2 | 16.89458 | 0.028154 | 0.999532 | | 22.41841 | 0.505965 | 0.236678 | | 19.093 | 0.494959 | 0.401561 | |
| HBP1 | 4.088725 | 0.122751 | 0.999532 | | 6.234631 | 0.776093 | 0.238431 | | 5.274259 | 0.677865 | 0.422058 | |
| DDX17 | 33.34717 | 0.332781 | 0.999532 | | 39.43234 | 0.472877 | 0.238558 | | 37.2032 | 0.164878 | 0.795612 | |
| EIF2AK2 | 4.740275 | 0.599566 | 0.999532 | | 5.310267 | 0.808314 | 0.238558 | | 5.714505 | 0.221966 | 0.849682 | |
| DAW1 | 2.852781 | -0.2728 | 0.999532 | | 2.577631 | -1.12866 | 0.238796 | | 1.985885 | -0.83803 | 0.540342 | |
| SDC2 | 15.47511 | 0.3321 | 0.999532 | | 19.72224 | 0.623989 | 0.238796 | | 18.32274 | 0.325224 | 0.673405 | |
| DUX4 | 75.48023 | -0.18574 | 0.999532 | | 77.1695 | -0.53516 | 0.239322 | | 60.59597 | -0.31368 | 0.647581 | |
| YPEL5 | 9.351276 | -0.22494 | 0.999532 | | 13.85713 | 0.530443 | 0.239322 | | 11.29554 | 0.774157 | 0.107433 | |
| TEX28 | 3.547547 | -0.0652 | 0.999532 | | 3.098316 | -1.05825 | 0.239322 | | 2.720236 | -0.96835 | 0.423641 | |
| MGP | 57.58084 | -0.17482 | 0.999532 | | 91.1655 | 0.717613 | 0.239322 | | 74.1734 | 0.9241 | 0.129363 | |
| ACIN1 | 12.35642 | 0.410162 | 0.999532 | | 15.5938 | 0.645755 | 0.239498 | | 14.87317 | 0.244951 | 0.750371 | |
| SLC3A1 | 4.108019 | 0.340934 | 0.999532 | | 5.860293 | 0.933369 | 0.239498 | | 5.484595 | 0.597514 | 0.571483 | |
| ANKH | 3.125767 | -0.11928 | 0.999532 | | 4.808939 | 0.832925 | 0.239498 | | 3.933073 | 0.971589 | 0.241605 | |
| GOLGA8 | 3.980349 | 0.211732 | 0.999532 | | 3.264158 | -1.00464 | 0.239498 | | 3.043052 | -1.18636 | 0.212897 | |
| SLC25A3 | 11.04962 | 0.439491 | 0.999532 | | 13.13359 | 0.546068 | 0.239498 | | 12.61474 | 0.128334 | 0.88885 | |
| GADD45B | 9.267924 | -0.95708 | 0.950261 | | 10.81312 | -0.88154 | 0.239528 | | 6.074087 | 0.062089 | 0.965649 | |

| | | | | | | | | | | | | |
|---|---|---|---|---|---|---|---|---|---|---|---|---|
| CSRP1 | 39.67468 | 0.064481 | 0.999532 | | 50.49259 | 0.415739 | 0.244226 | | 43.95535 | 0.371667 | 0.415282 | |
| PMPCB | 4.932796 | -0.09071 | 0.999532 | | 6.940723 | 0.703812 | 0.244226 | | 6.036725 | 0.801113 | 0.274496 | |
| KRTAP10 | 3.029483 | -0.08892 | 0.999532 | | 2.623116 | -1.17415 | 0.24622 | | 2.043478 | -1.06867 | 0.412398 | |
| SLC25A5 | 13.26047 | 0.154877 | 0.999532 | | 17.66451 | 0.573943 | 0.24622 | | 15.62114 | 0.439823 | 0.512069 | |
| LONP2 | 4.424921 | 0.263577 | 0.999532 | | 5.634753 | 0.750809 | 0.24622 | | 5.210946 | 0.499781 | 0.61958 | |
| CFL1 | 25.13026 | -0.03828 | 0.999532 | | 35.04867 | 0.538886 | 0.24622 | | 29.6045 | 0.599021 | 0.24893 | |
| URI1 | 6.271762 | 0.359393 | 0.999532 | | 7.753473 | 0.639936 | 0.246377 | | 7.422202 | 0.299008 | 0.720526 | |
| VKORC1 | 8.257893 | 0.578789 | 0.999532 | | 9.613077 | 0.602056 | 0.246377 | | 9.564711 | 0.037377 | 0.978229 | |
| WDFY3 | 3.757708 | 0.244781 | 0.999532 | | 5.222656 | 0.762414 | 0.246377 | | 4.789445 | 0.53685 | 0.552672 | |
| NUMA1 | 8.555095 | 0.143488 | 0.999532 | | 11.40811 | 0.601212 | 0.246377 | | 9.952102 | 0.471261 | 0.536289 | |
| EIF3G | 14.44764 | 0.167108 | 0.999532 | | 18.52129 | 0.508837 | 0.246377 | | 16.73327 | 0.36013 | 0.536289 | |
| EPS8L2 | 10.92197 | 0.532332 | 0.999532 | | 13.27966 | 0.725517 | 0.246377 | | 12.80273 | 0.229197 | 0.816775 | |
| CAVIN2 | 8.978999 | -1.00199 | 0.547204 | | 10.33457 | -0.89003 | 0.246981 | | 5.655996 | 0.175015 | 0.889853 | |
| MRPS15 | 4.43496 | 0.397362 | 0.999532 | | 5.416718 | 0.7681 | 0.248583 | | 5.390421 | 0.391013 | 0.688542 | |
| MED21 | 6.455618 | 0.399047 | 0.999532 | | 7.896863 | 0.66281 | 0.248583 | | 7.487747 | 0.278723 | 0.760572 | |
| IFIT1 | 3.64881 | 0.614607 | 0.999532 | | 4.053405 | 0.861356 | 0.248583 | | 4.38592 | 0.260655 | 0.839261 | |
| ARHGAP | 6.099387 | 0.185885 | 0.999532 | | 8.11034 | 0.648282 | 0.248583 | | 7.28569 | 0.48149 | 0.531732 | |
| SLC6A13 | 4.090984 | 0.517799 | 0.999532 | | 5.554161 | 1.033225 | 0.248583 | | 5.479551 | 0.522595 | 0.660958 | |
| CPNE2 | 5.907626 | 0.102204 | 0.999532 | | 8.559553 | 0.693386 | 0.249025 | | 7.532288 | 0.607088 | 0.397004 | |
| SERBP1 | 8.459059 | 0.294582 | 0.999532 | | 10.89767 | 0.588709 | 0.249107 | | 9.903069 | 0.313895 | 0.699971 | |
| PITX3 | 4.372975 | 0.165395 | 0.999532 | | 3.522257 | -1.04338 | 0.249107 | | 3.16399 | -1.185 | 0.222058 | |
| KIAA0040 | 6.103477 | 0.175633 | 0.999532 | | 8.823416 | 0.747566 | 0.249107 | | 8.035947 | 0.586795 | 0.421517 | |
| CTSB | 19.68077 | 0.085114 | 0.999532 | | 28.48227 | 0.707926 | 0.249107 | | 24.48843 | 0.654601 | 0.421517 | |
| OGDHL | 3.620172 | 0.309332 | 0.999532 | | 5.201259 | 0.865536 | 0.249107 | | 4.667911 | 0.5905 | 0.625223 | |
| IFNA16 | 3.516632 | -0.22089 | 0.999532 | | 3.131858 | -1.0604 | 0.249107 | | 2.714383 | -0.82149 | 0.556866 | |
| NR2F1 | 6.436824 | 0.358955 | 0.999532 | | 8.178523 | 0.684311 | 0.249107 | | 7.68594 | 0.3404 | 0.754884 | |
| TMEM59 | 14.04669 | -0.03554 | 0.999532 | | 20.68014 | 0.645713 | 0.249107 | | 17.23863 | 0.717547 | 0.266933 | |
| PGLS | 6.448973 | 0.138224 | 0.999532 | | 8.521559 | 0.67342 | 0.249107 | | 7.84381 | 0.539598 | 0.515762 | |
| KLF6 | 10.02094 | -0.82526 | 0.713279 | | 11.4766 | -0.74597 | 0.249107 | | 6.770427 | 0.120685 | 0.923938 | |
| NPIPB6 | 25.48788 | 0.37306 | 0.999532 | | 29.71228 | 0.455129 | 0.249464 | | 28.10428 | 0.106678 | 0.899852 | |
| PSMA3 | 5.076969 | -0.0688 | 0.999532 | | 7.679827 | 0.727211 | 0.249464 | | 6.41223 | 0.820855 | 0.222058 | |
| GATD3A | 7.56919 | 0.213106 | 0.999532 | | 10.22684 | 0.604358 | 0.24956 | | 8.843243 | 0.389306 | 0.652925 | |
| SPOCK2 | 36.75441 | 0.409622 | 0.999532 | | 45.72568 | 0.658387 | 0.249773 | | 43.79501 | 0.276097 | 0.688542 | |
| HNRNPUL | 9.303268 | 0.032209 | 0.999532 | | 12.64984 | 0.584914 | 0.250347 | | 11.26339 | 0.564627 | 0.359441 | |
| CLDN4 | 3.467199 | -0.76427 | 0.999532 | | 3.805652 | -0.93247 | 0.251663 | | 2.164159 | -0.15622 | 0.934354 | |
| UBA1 | 10.63771 | 0.40398 | 0.999532 | | 12.77869 | 0.542923 | 0.251663 | | 12.17871 | 0.156856 | 0.85585 | |
| PRDX2 | 6.823963 | 0.407448 | 0.999532 | | 8.730082 | 0.695594 | 0.251798 | | 8.312094 | 0.3049 | 0.723933 | |
| PHLDA2 | 4.628423 | -0.36883 | 0.999532 | | 4.510685 | -0.84614 | 0.251798 | | 3.396551 | -0.4607 | 0.718633 | |
| FAM185A | 5.058208 | -0.00945 | 0.999532 | | 4.215082 | -0.86269 | 0.251966 | | 3.782996 | -0.84225 | 0.389965 | |
| IER3 | 12.02505 | -0.45366 | 0.999532 | | 12.64945 | -0.67145 | 0.254647 | | 8.76291 | -0.18945 | 0.862823 | |
| CDK5RAP | 5.473945 | 0.291054 | 0.999532 | | 7.344129 | 0.69634 | 0.254647 | | 6.603206 | 0.425025 | 0.66549 | |
| EFEMP1 | 15.29842 | 0.25002 | 0.999532 | | 19.88367 | 0.652462 | 0.254647 | | 18.28168 | 0.417968 | 0.578798 | |
| FAM171A | 3.567779 | -0.23069 | 0.999532 | | 3.266565 | -0.96224 | 0.255023 | | 2.410913 | -0.71702 | 0.565022 | |
| ANKRD18 | 5.590653 | 0.185471 | 0.999532 | | 4.674795 | -0.98257 | 0.255023 | | 4.150942 | -1.15218 | 0.150518 | |
| FZD6 | 2.371674 | 0.805317 | 0.999532 | | 2.846462 | 1.004977 | 0.255023 | | 2.899483 | 0.214924 | 0.885783 | |
| GPKOW | 3.042791 | 1.008438 | 0.999532 | | 3.427329 | 0.956331 | 0.255023 | | 3.748666 | -0.03284 | 0.989148 | |
| ATP5F1C | 19.22435 | -0.08742 | 0.999532 | | 26.63098 | 0.50359 | 0.255023 | | 22.30522 | 0.604587 | 0.24893 | |
| FXYD6 | 11.66964 | 0.018441 | 0.999532 | | 16.00339 | 0.573936 | 0.255023 | | 14.00555 | 0.569069 | 0.444687 | |
| VRK3 | 4.661556 | -0.64076 | 0.999532 | | 5.02418 | -0.8072 | 0.255179 | | 3.163509 | -0.14778 | 0.926506 | |
| SARAF | 13.29601 | 0.161859 | 0.999532 | | 16.86372 | 0.540537 | 0.255179 | | 15.24035 | 0.393827 | 0.536289 | |
| TMEM200 | 2.382009 | 0.084711 | 0.999532 | | 3.689005 | 0.934795 | 0.255262 | | 3.232573 | 0.863395 | 0.421517 | |
| PGA3 | 3.668262 | -0.22889 | 0.999532 | | 3.394822 | -1.11933 | 0.255269 | | 2.673697 | -0.87745 | 0.515762 | |
| GABARA | 10.30406 | -0.06612 | 0.999532 | | 14.22403 | 0.500175 | 0.255542 | | 11.7174 | 0.57682 | 0.355692 | |
| PXDN | 5.380506 | 0.128058 | 0.999532 | | 7.7567 | 0.69314 | 0.256511 | | 6.902984 | 0.587121 | 0.484303 | |
| ST3GAL6 | 6.316954 | 0.611369 | 0.999532 | | 7.652194 | 0.738116 | 0.258978 | | 7.656338 | 0.14754 | 0.902619 | |
| BAG1 | 2.733588 | -0.33467 | 0.999532 | | 2.539749 | -1.08143 | 0.259182 | | 1.819334 | -0.72747 | 0.623462 | |
| PSMB7 | 5.631886 | 0.152623 | 0.999532 | | 7.786442 | 0.693449 | 0.259392 | | 6.879085 | 0.561822 | 0.49299 | |
| PTK2 | 6.067544 | 0.26286 | 0.999532 | | 7.619138 | 0.626788 | 0.260801 | | 7.081545 | 0.376883 | 0.654772 | |
| ITM2C | 17.00153 | 0.524763 | 0.999532 | | 19.62036 | 0.579986 | 0.261567 | | 19.42954 | 0.084142 | 0.938929 | |
| TNFAIP8L | 3.609034 | 0.436939 | 0.999532 | | 4.389228 | 0.810819 | 0.261567 | | 4.459148 | 0.393158 | 0.730921 | |
| CELF1 | 4.287518 | 0.057678 | 0.999532 | | 6.194831 | 0.68876 | 0.261567 | | 5.255121 | 0.642392 | 0.442674 | |
| ANKRD17 | 5.172005 | 0.295002 | 0.999532 | | 6.796802 | 0.669304 | 0.262274 | | 6.203296 | 0.391713 | 0.647581 | |
| SRP14 | 19.25785 | 0.260168 | 0.999532 | | 24.00091 | 0.533228 | 0.262274 | | 21.98648 | 0.295818 | 0.682584 | |
| MYO1F | 3.714531 | -0.26606 | 0.999532 | | 3.340167 | -0.97882 | 0.263507 | | 2.697826 | -0.68491 | 0.60629 | |
| CXCL2 | 2.379546 | -0.91045 | 0.999532 | | 2.585484 | -1.10419 | 0.264515 | | 1.539281 | -0.17338 | 0.943059 | |
| CXorf58 | 2.89795 | 0.196198 | 0.999532 | | 2.055346 | -1.23965 | 0.264709 | | 2.137726 | -1.41735 | 0.246926 | |
| POU2F3 | 2.450481 | -0.23831 | 0.999532 | | 2.25416 | -1.20431 | 0.264709 | | 1.738313 | -0.94807 | 0.527774 | |
| EIF5A | 15.31294 | 0.100306 | 0.999532 | | 19.30244 | 0.472778 | 0.266763 | | 17.1723 | 0.386579 | 0.49299 | |
| GRB2 | 6.115638 | 0.252029 | 0.999532 | | 8.690636 | 0.8208 | 0.266974 | | 7.987423 | 0.582083 | 0.445382 | |
| COL1A1 | 2.387744 | 0.168553 | 0.999532 | | 3.536354 | 0.997121 | 0.267673 | | 3.226352 | 0.84626 | 0.436389 | |
| NIP7 | 4.730871 | -0.56172 | 0.999532 | | 4.909583 | -0.75609 | 0.268134 | | 3.427379 | -0.17967 | 0.901033 | |
| PLSCR4 | 2.208838 | -0.20254 | 0.999532 | | 3.770082 | 0.853252 | 0.268134 | | 3.210599 | 1.091436 | 0.241605 | |
| S100A13 | 8.592272 | 0.260291 | 0.999532 | | 10.96505 | 0.607634 | 0.269292 | | 10.09487 | 0.370476 | 0.644077 | |
| GJA5 | 17.53443 | -0.28209 | 0.999532 | | 26.37954 | 0.522493 | 0.269292 | | 21.26744 | 0.806223 | 0.079641 | UP |
| PRAMEF | 7.338449 | -0.36084 | 0.999532 | | 7.335141 | -0.77503 | 0.269292 | | 5.734096 | -0.42166 | 0.720526 | |
| DIRAS3 | 2.869874 | -0.86237 | 0.999532 | | 2.998604 | -0.99011 | 0.269292 | | 1.855302 | -0.12117 | 0.960476 | |
| ID3 | 17.27077 | -0.00658 | 0.999532 | | 23.24234 | 0.416692 | 0.269292 | | 19.55175 | 0.4437 | 0.30352 | |
| INF2 | 8.63307 | 0.113119 | 0.999532 | | 11.82332 | 0.523429 | 0.269292 | | 10.29824 | 0.428802 | 0.512866 | |
| PAX8 | 6.736051 | 0.364449 | 0.999532 | | 9.803957 | 0.939272 | 0.269292 | | 8.878021 | 0.636371 | 0.546319 | |
| SERPINA | 7.330575 | 0.186342 | 0.999532 | | 10.10176 | 0.705371 | 0.269471 | | 8.907945 | 0.545638 | 0.515762 | |
| RTRAF | 5.857948 | 0.253286 | 0.999532 | | 7.844269 | 0.709378 | 0.269471 | | 7.277865 | 0.47144 | 0.584522 | |
| C1S | 6.851189 | 1.004657 | 0.297305 | | 6.839781 | 0.82237 | 0.269704 | | 8.112217 | -0.17106 | 0.895345 | |
| LENG8 | 24.45884 | 0.194442 | 0.999532 | | 30.75425 | 0.468769 | 0.269704 | | 27.37446 | 0.299567 | 0.666906 | |
| WASHC2 | 4.565482 | -0.25531 | 0.999532 | | 4.315227 | -0.81866 | 0.269704 | | 3.382237 | -0.54476 | 0.655993 | |
| C8orf74 | 6.738682 | -0.16006 | 0.999532 | | 6.450842 | -0.74119 | 0.269704 | | 5.084494 | -0.5599 | 0.540342 | |
| GP6 | 5.896082 | -0.1227 | 0.999532 | | 5.24047 | -0.74057 | 0.269704 | | 4.675013 | -0.60772 | 0.566812 | |
| ACSM2B | 10.62683 | 0.10529 | 0.999532 | | 16.29285 | 0.889554 | 0.269704 | | 14.08195 | 0.822596 | 0.363418 | |
| CD59 | 14.63651 | 0.152654 | 0.999532 | | 19.21845 | 0.567419 | 0.269704 | | 16.99717 | 0.442416 | 0.515762 | |
| RBX1 | 5.786054 | -0.20212 | 0.999532 | | 8.571235 | 0.602419 | 0.269704 | | 6.731183 | 0.818824 | 0.178337 | |
| NES | 9.105999 | 0.154695 | 0.999532 | | 12.31159 | 0.6818 | 0.269704 | | 10.92421 | 0.554271 | 0.444687 | |
| RPS20 | 41.5546 | -0.11492 | 0.999532 | | 56.66345 | 0.403665 | 0.270139 | | 46.32792 | 0.547904 | 0.188098 | |
| SCTR | 19.20661 | 0.033126 | 0.999532 | | 17.8361 | -0.53528 | 0.270139 | | 15.20872 | -0.5423 | 0.286063 | |
| COL3A1 | 12.27057 | 0.129164 | 0.999532 | | 23.61988 | 1.361495 | 0.270139 | | 20.68885 | 1.248885 | 0.274496 | |
| ZNF572 | 3.800236 | 0.132301 | 0.999532 | | 3.018775 | -1.05438 | 0.270139 | | 2.944257 | -1.16775 | 0.314028 | |
| C8orf58 | 7.875577 | 0.067384 | 0.999532 | | 11.00765 | 0.588191 | 0.273559 | | 9.610862 | 0.538749 | 0.386321 | |
| XAGE1A | 3.137755 | 0.031543 | 0.999532 | | 2.576048 | -1.18107 | 0.275453 | | 2.356956 | -1.20584 | 0.298628 | |
| EIF4A2 | 16.06608 | 0.254104 | 0.999532 | | 19.48348 | 0.45122 | 0.275511 | | 17.73513 | 0.221626 | 0.736481 | |
| SSX2 | 3.510314 | 0.051569 | 0.999532 | | 2.872111 | -1.03466 | 0.275511 | | 2.63126 | -1.06812 | 0.359441 | |
| MSRA | 3.675379 | 0.36906 | 0.999532 | | 4.655491 | 0.81452 | 0.276996 | | 4.49341 | 0.461289 | 0.66232 | |
| ZNF764 | 6.456126 | -0.1649 | 0.999532 | | 6.260883 | -0.7681 | 0.276996 | | 5.000804 | -0.58349 | 0.543177 | |
| TRIM52 | 3.765103 | 0.466814 | 0.999532 | | 4.958336 | 0.796439 | 0.276996 | | 4.646617 | 0.34939 | 0.740093 | |
| JUP | 7.07995 | 0.037029 | 0.999532 | | 10.12868 | 0.649094 | 0.277906 | | 8.462591 | 0.634839 | 0.412558 | |
| KRTAP4- | 6.269464 | -0.0232 | 0.999532 | | 5.63796 | -0.82275 | 0.277906 | | 4.920747 | -0.78843 | 0.423641 | |
| ZNF776 | 15.92798 | 0.048742 | 0.999532 | | 14.86803 | -0.54065 | 0.277906 | | 13.2643 | -0.56344 | 0.431254 | |
| MRGPRF | 7.983588 | 0.651706 | 0.999532 | | 8.957337 | 0.71812 | 0.277906 | | 9.551398 | 0.078538 | 0.950895 | |
| OR11H1 | 3.057687 | 0.333668 | 0.999532 | | 2.179318 | -1.17156 | 0.277906 | | 2.263554 | -1.48676 | 0.158141 | |
| ARRDC4 | 17.59409 | 0.004178 | 0.999532 | | 23.4891 | 0.51242 | 0.277906 | | 20.26222 | 0.523564 | 0.414073 | |
| SCARB2 | 16.82382 | 0.192332 | 0.999532 | | 21.9361 | 0.592267 | 0.277906 | | 19.86205 | 0.421897 | 0.475974 | |
| SPATA31 | 5.055041 | 0.007915 | 0.999532 | | 4.504772 | -0.90343 | 0.277906 | | 3.852594 | -0.88944 | 0.35437 | |
| SLC17A3 | 2.612276 | 0.816332 | 0.999532 | | 3.300864 | 1.042685 | 0.277906 | | 3.425146 | 0.250618 | 0.877238 | |
| QKI | 11.10376 | 0.144763 | 0.999532 | | 14.36373 | 0.516197 | 0.277906 | | 12.65581 | 0.390972 | 0.531935 | |
| ESYT1 | 10.02097 | 0.193446 | 0.999532 | | 13.13849 | 0.558246 | 0.277906 | | 11.84956 | 0.386367 | 0.570037 | |
| SCARA5 | 2.692214 | -0.3935 | 0.999532 | | 2.533034 | -1.09916 | 0.277906 | | 1.922968 | -0.69267 | 0.652925 | |
| PPP3CB | 6.452448 | 0.251163 | 0.999532 | | 8.263233 | 0.586762 | 0.277906 | | 7.556222 | 0.353973 | 0.658435 | |
| NKG7 | 3.364636 | -0.33178 | 0.999532 | | 3.074209 | -0.95487 | 0.277906 | | 2.49277 | -0.5936 | 0.664512 | |
| PRDX5 | 7.448923 | 0.137671 | 0.999532 | | 9.994885 | 0.635175 | 0.277906 | | 8.876193 | 0.512467 | 0.536289 | |
| CNDP2 | 6.401328 | 0.095999 | 0.999532 | | 9.059809 | 0.63596 | 0.277906 | | 7.831855 | 0.553535 | 0.475206 | |
| HEG1 | 15.91254 | -0.42632 | 0.999532 | | 24.64709 | 0.458035 | 0.277906 | | 18.71735 | 0.902941 | 0.017097 | UP |
| HSPA1A | 13.40269 | 0.160929 | 0.999532 | | 17.20496 | 0.509538 | 0.277906 | | 15.23913 | 0.37406 | 0.532828 | |
| AMOTL2 | 12.63564 | 0.194726 | 0.999532 | | 15.79409 | 0.516462 | 0.279071 | | 14.3257 | 0.341882 | 0.623462 | |
| STMN1 | 3.915744 | 0.366101 | 0.999532 | | 5.040059 | 0.797955 | 0.279071 | | 4.599939 | 0.430139 | 0.702061 | |
| CYP2D7 | 6.121709 | 0.123308 | 0.999532 | | 5.447506 | -0.76819 | 0.279071 | | 4.66798 | -0.86071 | 0.269743 | |
| TLN1 | 17.25569 | -0.20745 | 0.999532 | | 25.46579 | 0.484624 | 0.279071 | | 20.26051 | 0.713168 | 0.108014 | |
| RCAN1 | 19.03416 | -0.36772 | 0.999532 | | 30.44486 | 0.595556 | 0.279071 | | 23.54354 | 0.992688 | 0.062627 | UP |
| PNOC | 3.587475 | -0.04638 | 0.999532 | | 3.015861 | -0.99134 | 0.279071 | | 2.579503 | -0.92991 | 0.424976 | |
| KCTD6 | 2.721008 | 0.708766 | 0.999532 | | 3.135131 | 0.923829 | 0.279071 | | 3.329544 | 0.227936 | 0.877586 | |
| TRIM48 | 2.692948 | -0.27409 | 0.999532 | | 2.638935 | -1.1136 | 0.279071 | | 1.885275 | -0.8098 | 0.581287 | |
| SAT2 | 9.043167 | 0.643749 | 0.999532 | | 9.993731 | 0.640411 | 0.279071 | | 10.27984 | 0.0171 | 0.993618 | |
| ATAD1 | 3.809857 | 0.146132 | 0.999532 | | 5.811028 | 0.709283 | 0.279071 | | 4.885855 | 0.572769 | 0.532088 | |
| MRPL16 | 4.457734 | 0.435249 | 0.999532 | | 5.499779 | 0.710683 | 0.279071 | | 5.322946 | 0.291328 | 0.78258 | |
| PARVA | 23.5638 | 0.19803 | 0.999532 | | 29.81369 | 0.510543 | 0.279071 | | 26.81432 | 0.340538 | 0.563885 | |
| CHD3 | 8.108028 | -0.03577 | 0.999532 | | 11.14058 | 0.547798 | 0.279071 | | 9.282486 | 0.602376 | 0.283785 | |
| ODF3 | 3.379622 | -0.32238 | 0.999532 | | 3.160777 | -0.95339 | 0.279071 | | 2.416142 | -0.61279 | 0.66232 | |
| TSPAN1 | 4.032902 | 0.308655 | 0.999532 | | 5.869713 | 0.906411 | 0.279071 | | 5.247525 | 0.642441 | 0.589729 | |
| RHOC | 16.6254 | -0.04655 | 0.999532 | | 22.79682 | 0.535771 | 0.279071 | | 19.49059 | 0.598173 | 0.205283 | |
| NFKBIZ | 4.347841 | -0.81861 | 0.999532 | | 4.685403 | -0.88908 | 0.279071 | | 2.931998 | -0.03764 | 0.989045 | |
| NDUFB8 | 16.06105 | -0.02802 | 0.999532 | | 21.39093 | 0.49998 | 0.279071 | | 18.21233 | 0.544823 | 0.304062 | |
| FMO1 | 4.809637 | 0.186472 | 0.999532 | | 7.026105 | 0.834283 | 0.279564 | | 6.246134 | 0.671074 | 0.547092 | |
| TAS2R38 | 2.659519 | -0.12341 | 0.999532 | | 2.276577 | -1.13276 | 0.279743 | | 1.921399 | -0.9937 | 0.488953 | |
| PSME3 | 4.829351 | 0.2857 | 0.999532 | | 6.465386 | 0.688262 | 0.280676 | | 5.826052 | 0.423507 | 0.646821 | |
| TRIM28 | 7.841318 | -0.20213 | 0.999532 | | 11.99293 | 0.573183 | 0.281266 | | 9.470632 | 0.795616 | 0.154583 | |
| PPP1CA | 9.321883 | 0.079705 | 0.999532 | | 12.65236 | 0.603786 | 0.281266 | | 11.31942 | 0.547763 | 0.37596 | |
| ESAM | 16.84501 | -0.17844 | 0.999532 | | 24.25474 | 0.499907 | 0.281266 | | 19.70578 | 0.699823 | 0.154583 | |
| DEFB108 | 3.712693 | 0.03569 | 0.999532 | | 3.319817 | -0.97049 | 0.281266 | | 2.838567 | -0.98833 | 0.342845 | |
| PARM1 | 8.409203 | -0.0697 | 0.999532 | | 7.888089 | -0.61442 | 0.281266 | | 6.530734 | -0.52882 | 0.510412 | |
| SNCA | 5.792086 | 0.17579 | 0.999532 | | 7.913637 | 0.649813 | 0.281266 | | 7.143172 | 0.49074 | 0.542124 | |
| SLC36A4 | 4.089847 | 0.240186 | 0.999532 | | 6.347342 | 0.945282 | 0.282217 | | 5.804528 | 0.720463 | 0.527774 | |
| CAPRIN1 | 6.157102 | -0.1781 | 0.999532 | | 9.229392 | 0.619767 | 0.282349 | | 7.464032 | 0.809681 | 0.241605 | |
| EZHIP | 2.885319 | 0.155009 | 0.999532 | | 2.044 | -1.21055 | 0.282349 | | 2.12685 | -1.33768 | 0.299463 | |
| RPL5 | 27.92381 | 0.088096 | 0.999532 | | 35.17101 | 0.402333 | 0.282349 | | 30.59514 | 0.336185 | 0.540342 | |
| PPP2CB | 8.417529 | -0.15665 | 0.999532 | | 12.14502 | 0.614656 | 0.282349 | | 9.929529 | 0.790876 | 0.199116 | |
| USP9X | 5.047873 | 0.563816 | 0.999532 | | 6.163453 | 0.667059 | 0.282427 | | 6.12697 | 0.127395 | 0.920025 | |
| PCM1 | 4.393976 | 0.669261 | 0.999532 | | 5.140272 | 0.754541 | 0.282431 | | 5.342306 | 0.10464 | 0.939913 | |
| ATP5PB | 10.14671 | 0.322833 | 0.999532 | | 11.84971 | 0.507023 | 0.283559 | | 11.21172 | 0.195027 | 0.816372 | |
| CMTM7 | 4.045126 | 0.076837 | 0.999532 | | 3.131493 | -0.94189 | 0.284054 | | 3.013897 | -1.01189 | 0.311486 | |
| VTA1 | 2.947825 | 0.799641 | 0.999532 | | 3.710611 | 0.888979 | 0.284987 | | 3.738462 | 0.117583 | 0.941559 | |
| RAB11A | 12.69705 | -0.30968 | 0.999532 | | 18.02566 | 0.463992 | 0.284987 | | 14.20906 | 0.78445 | 0.105617 | |
| RPL4 | 28.54782 | 0.000387 | 0.999532 | | 39.79322 | 0.56299 | 0.284987 | | 33.7705 | 0.587001 | 0.359638 | |
| SETD5 | 5.797896 | 0.21547 | 0.999532 | | 7.643359 | 0.653729 | 0.285161 | | 6.780889 | 0.455429 | 0.57215 | |
| FCMR | 3.960228 | -0.18 | 0.999532 | | 3.815767 | -0.87058 | 0.285552 | | 3.034955 | -0.66316 | 0.575547 | |
| MIOX | 5.480564 | 0.713434 | 0.999532 | | 6.900425 | 1.011974 | 0.285722 | | 6.944498 | 0.326229 | 0.861737 | |
| CLDN5 | 10.58066 | 0.349214 | 0.999532 | | 13.03913 | 0.587655 | 0.285722 | | 12.34714 | 0.260345 | 0.75038 | |
| GBP5 | 2.488036 | -0.35625 | 0.999532 | | 2.23321 | -1.17729 | 0.285722 | | 1.822624 | -0.80214 | 0.600081 | |
| SMARCA | 3.694375 | -0.36048 | 0.999532 | | 5.897749 | 0.687591 | 0.285722 | | 4.499501 | 1.059469 | 0.15668 | |
| XAF1 | 8.084932 | 0.620458 | 0.999532 | | 8.42318 | 0.581755 | 0.285722 | | 9.138008 | -0.01346 | 0.996681 | |
| ENO1 | 7.125086 | 0.40756 | 0.999532 | | 9.098858 | 0.725312 | 0.285722 | | 8.517341 | 0.352313 | 0.723933 | |
| TK1 | 4.517245 | -0.37118 | 0.999532 | | 4.344301 | -0.87304 | 0.285722 | | 3.18994 | -0.47094 | 0.699971 | |
| CANX | 25.80088 | 0.011936 | 0.999532 | | 34.58081 | 0.467155 | 0.285722 | | 29.36629 | 0.479578 | 0.398124 | |
| ATP5ME | 10.53073 | -0.10516 | 0.999532 | | 15.03862 | 0.529931 | 0.285722 | | 12.43348 | 0.656468 | 0.278559 | |
| TTC26 | 2.791055 | -0.45989 | 0.999532 | | 2.581565 | -1.03298 | 0.285722 | | 1.939675 | -0.56103 | 0.735469 | |
| AP3D1 | 5.244549 | 0.042718 | 0.999532 | | 7.370514 | 0.603755 | 0.287248 | | 6.280761 | 0.575919 | 0.451915 | |
| KRTAP3- | 2.969402 | 0.224641 | 0.999532 | | 2.154336 | -1.17759 | 0.287639 | | 2.142115 | -1.38016 | 0.241605 | |
| IFI6 | 11.79068 | 0.411241 | 0.999532 | | 14.05758 | 0.677215 | 0.289698 | | 13.97626 | 0.288747 | 0.76302 | |
| RPLP2 | 27.91846 | -0.01135 | 0.999532 | | 37.81748 | 0.520512 | 0.290751 | | 32.35383 | 0.546315 | 0.274496 | |
| RSU1 | 13.01586 | 0.204644 | 0.999532 | | 16.57097 | 0.567775 | 0.292608 | | 15.27546 | 0.376912 | 0.570037 | |
| H3-3A | 36.90806 | 0.008036 | 0.999532 | | 47.28347 | 0.396208 | 0.292608 | | 40.65342 | 0.410598 | 0.283071 | |
| MORF4L1 | 37.41318 | -0.04331 | 0.999532 | | 50.17854 | 0.414491 | 0.292608 | | 41.9846 | 0.484715 | 0.282041 | |
| FOXD4L4 | 5.485583 | 0.185011 | 0.999532 | | 4.570423 | -0.83791 | 0.292608 | | 4.150323 | -1.00173 | 0.216015 | |
| CDK2AP2 | 6.092622 | -0.23817 | 0.999532 | | 5.932878 | -0.78557 | 0.292608 | | 4.547533 | -0.5277 | 0.619423 | |
| SHROOM | 5.195596 | 0.401444 | 0.999532 | | 6.428367 | 0.667896 | 0.294412 | | 6.096142 | 0.278843 | 0.774044 | |
| GDF15 | 4.657225 | -1.2029 | 0.912432 | | 5.550378 | -1.05628 | 0.294713 | | 2.833051 | 0.129403 | 0.944635 | |
| SERF2 | 21.439 | -0.04068 | 0.999532 | | 28.25623 | 0.402177 | 0.295363 | | 23.64401 | 0.460405 | 0.340167 | |
| SORBS2 | 5.968759 | 0.267382 | 0.999532 | | 7.21379 | 0.632451 | 0.296724 | | 6.972088 | 0.379522 | 0.666492 | |
| BCAP31 | 7.568649 | 0.049731 | 0.999532 | | 10.33154 | 0.529685 | 0.298283 | | 8.830603 | 0.4988 | 0.486743 | |
| KCNE2 | 3.661512 | 0.158539 | 0.999532 | | 2.945133 | -0.94836 | 0.298283 | | 2.842923 | -1.09416 | 0.273545 | |
| PGK1 | 8.898005 | -0.08182 | 0.999532 | | 12.90322 | 0.544889 | 0.298283 | | 10.27947 | 0.662292 | 0.300261 | |
| CDC73 | 9.503178 | 0.019658 | 0.999532 | | 12.70878 | 0.499124 | 0.299666 | | 10.76344 | 0.496698 | 0.37596 | |
| EHD4 | 3.757136 | -0.10408 | 0.999532 | | 5.591493 | 0.712373 | 0.299666 | | 4.722678 | 0.829368 | 0.298614 | |
| DSTN | 12.12692 | -0.09415 | 0.999532 | | 16.98627 | 0.554675 | 0.300258 | | 14.3003 | 0.672245 | 0.196145 | |
| RSPH10B | 2.943615 | -0.07945 | 0.999532 | | 2.6042 | -1.04362 | 0.300258 | | 2.124033 | -0.9456 | 0.484303 | |
| EIF3H | 8.00298 | -0.10921 | 0.999532 | | 11.44816 | 0.531956 | 0.300258 | | 9.48786 | 0.660601 | 0.241605 | |
| COX7C | 5.840257 | 0.372082 | 0.999532 | | 7.379446 | 0.698619 | 0.300258 | | 6.933919 | 0.344868 | 0.738316 | |
| SERINC5 | 4.966729 | 0.608301 | 0.999532 | | 5.674046 | 0.702777 | 0.300258 | | 5.800494 | 0.111603 | 0.932589 | |
| EEF1AKN | 2.898776 | -0.30346 | 0.999532 | | 2.534944 | -1.05362 | 0.300258 | | 2.027313 | -0.73732 | 0.638015 | |
| FARP1 | 9.891904 | 0.098312 | 0.999532 | | 12.96361 | 0.517163 | 0.300258 | | 11.1601 | 0.43631 | 0.531935 | |
| SQSTM1 | 13.97532 | 0.040942 | 0.999532 | | 18.06387 | 0.45167 | 0.300258 | | 15.28958 | 0.425655 | 0.49299 | |
| OR2H2 | 3.298819 | -0.37592 | 0.999532 | | 3.310153 | -0.93569 | 0.300258 | | 2.323048 | -0.53797 | 0.709633 | |
| TAMALIN | 8.284204 | -1.02879 | 0.51089 | | 10.01274 | -0.75067 | 0.300258 | | 5.669734 | 0.293134 | 0.760473 | |
| SYNJ2BP | 2.643029 | 0.488469 | 0.999532 | | 3.64611 | 0.848243 | 0.300258 | | 3.425555 | 0.377406 | 0.764149 | |
| HADHA | 4.235353 | 0.311082 | 0.999532 | | 5.405922 | 0.700018 | 0.300258 | | 5.197332 | 0.405269 | 0.679818 | |
| EFCAB13 | 5.016662 | -0.07971 | 0.999532 | | 7.6966 | 0.800489 | 0.300258 | | 6.289621 | 0.887538 | 0.332351 | |
| RPL28 | 28.32727 | 0.090332 | 0.999532 | | 37.86608 | 0.566297 | 0.300258 | | 33.13719 | 0.495342 | 0.480252 | |
| MAGED1 | 5.820708 | 0.125487 | 0.999532 | | 7.869099 | 0.632096 | 0.300258 | | 6.941383 | 0.521988 | 0.540342 | |
| CSNK2A3 | 5.858337 | 0.209136 | 0.999532 | | 8.055531 | 0.676055 | 0.300258 | | 7.030276 | 0.502383 | 0.566812 | |
| MIF | 12.76382 | 0.297713 | 0.999532 | | 15.61813 | 0.549455 | 0.300258 | | 14.60155 | 0.2704 | 0.745881 | |
| TDRD9 | 2.774304 | -0.85258 | 0.999532 | | 3.14445 | -0.93159 | 0.300258 | | 1.816439 | -0.05092 | 0.986811 | |
| ARL6IP5 | 10.48627 | -0.0225 | 0.999532 | | 14.67959 | 0.54655 | 0.300258 | | 12.42641 | 0.589151 | 0.342845 | |
| SEL1L | 16.91083 | 0.121396 | 0.999532 | | 21.85593 | 0.409385 | 0.300258 | | 19.12876 | 0.308343 | 0.590692 | |
| PRPF38A | 6.263182 | -0.04134 | 0.999532 | | 5.669267 | -0.69488 | 0.300258 | | 4.837627 | -0.63559 | 0.464732 | |
| PLK3 | 7.164157 | -0.79919 | 0.999532 | | 8.375695 | -0.73845 | 0.300258 | | 4.924352 | 0.075719 | 0.962716 | |
| WDR83OS | 4.323131 | 0.745998 | 0.999532 | | 4.628927 | 0.73655 | 0.300258 | | 5.109176 | 0.007463 | 0.996891 | |
| TMED9 | 9.439767 | 0.179771 | 0.999532 | | 12.55287 | 0.645195 | 0.300258 | | 11.28264 | 0.487397 | 0.520499 | |
| TXN2 | 6.812961 | -0.16482 | 0.999532 | | 9.939243 | 0.530892 | 0.30037 | | 8.35134 | 0.720851 | 0.222057 | |
| SLFN12L | 3.877097 | 0.073226 | 0.999532 | | 3.167647 | -0.9589 | 0.300835 | | 3.00394 | -1.00749 | 0.369628 | |
| OR5Q2 | 4.028598 | 0.061267 | 0.999532 | | 3.351924 | -0.93437 | 0.300835 | | 3.046538 | -0.98004 | 0.324867 | |
| RNASEK | 9.701429 | 0.1785 | 0.999532 | | 12.24242 | 0.510581 | 0.300957 | | 10.89876 | 0.34948 | 0.628895 | |
| DNM2 | 9.89932 | 0.215043 | 0.999532 | | 12.87785 | 0.567909 | 0.301248 | | 11.45786 | 0.376726 | 0.649725 | |
| NSMF | 8.497665 | 0.189148 | 0.999532 | | 10.36025 | 0.565935 | 0.301248 | | 9.585253 | 0.385518 | 0.625173 | |
| DYSF | 6.897679 | -0.17602 | 0.999532 | | 9.994831 | 0.533086 | 0.301248 | | 8.234654 | 0.727011 | 0.209531 | |
| CYBA | 10.04224 | -0.07463 | 0.999532 | | 13.65057 | 0.490183 | 0.301248 | | 11.46977 | 0.58458 | 0.274496 | |
| RPLP0 | 11.13741 | 0.028755 | 0.999532 | | 15.40552 | 0.639032 | 0.301248 | | 13.27688 | 0.628086 | 0.379978 | |
| GUSB | 3.950127 | 0.343881 | 0.999532 | | 5.094219 | 0.723326 | 0.301248 | | 4.754985 | 0.398219 | 0.690611 | |
| SYPL1 | 6.105965 | 0.368697 | 0.999532 | | 7.243265 | 0.632313 | 0.301248 | | 7.045616 | 0.290312 | 0.739301 | |
| LRCOL1 | 3.303157 | -0.03754 | 0.999532 | | 2.993861 | -0.92075 | 0.301248 | | 2.456631 | -0.8597 | 0.458377 | |
| FAM217A | 2.496047 | -0.18097 | 0.999532 | | 2.134767 | -1.13085 | 0.301248 | | 1.728759 | -0.93952 | 0.536289 | |
| ADAM21 | 2.67094 | -0.09589 | 0.999532 | | 2.321913 | -1.18094 | 0.303037 | | 1.834567 | -1.06861 | 0.445382 | |
| GAK | 12.28402 | 0.336532 | 0.999532 | | 14.24679 | 0.449283 | 0.303037 | | 13.50256 | 0.127061 | 0.88476 | |
| WNK2 | 2.915706 | -0.15433 | 0.999532 | | 2.699844 | -1.0795 | 0.303037 | | 1.99419 | -0.90463 | 0.506666 | |
| PRRC1 | 6.652357 | 0.167657 | 0.999532 | | 8.58299 | 0.565553 | 0.303037 | | 7.863307 | 0.419873 | 0.547092 | |
| ARHGAP | 4.518405 | 0.358896 | 0.999532 | | 5.952607 | 0.690783 | 0.303054 | | 5.75501 | 0.349554 | 0.722703 | |
| PDIA3 | 18.18066 | -0.16235 | 0.999532 | | 25.64288 | 0.509358 | 0.303054 | | 20.9718 | 0.681745 | 0.242703 | |
| SYNGR2 | 10.2544 | -0.34566 | 0.999532 | | 12.42173 | 0.543974 | 0.303054 | | 11.78219 | 0.218693 | 0.776852 | |
| HSP90AA | 12.9573 | -0.02595 | 0.999532 | | 17.82962 | 0.496556 | 0.303339 | | 14.77714 | 0.547288 | 0.389965 | |
| TUBB8 | 4.261718 | 0.29691 | 0.999532 | | 3.289494 | -0.92451 | 0.303339 | | 3.179163 | -1.20341 | 0.189136 | |
| UGP2 | 4.958446 | 0.261527 | 0.999532 | | 6.242375 | 0.690531 | 0.303896 | | 5.979634 | 0.434843 | 0.666566 | |
| TMEM165 | 9.182548 | 0.295328 | 0.999532 | | 11.8534 | 0.531961 | 0.303896 | | 10.89799 | 0.261672 | 0.737539 | |
| PIGH | 5.138469 | 0.026144 | 0.999532 | | 4.61315 | -0.8116 | 0.304163 | | 3.925071 | -0.81661 | 0.403558 | |
| USO1 | 6.45799 | 0.328528 | 0.999532 | | 7.916632 | 0.614385 | 0.304163 | | 7.763312 | 0.297481 | 0.732486 | |
| HTATIP2 | 4.486348 | -0.49037 | 0.999532 | | 4.634025 | -0.76881 | 0.304773 | | 3.253048 | -0.26077 | 0.854533 | |
| FSIP1 | 2.948284 | -0.18875 | 0.999532 | | 2.573809 | -0.9969 | 0.305082 | | 2.205251 | -0.79008 | 0.557909 | |
| BCL2L11 | 6.614645 | 0.067271 | 0.999532 | | 5.658928 | -0.74047 | 0.307126 | | 5.128971 | -0.7922 | 0.292884 | |
| TUSC3 | 4.365784 | 0.648259 | 0.999532 | | 5.21259 | 0.792912 | 0.308039 | | 5.351085 | 0.168917 | 0.895237 | |
| SENP6 | 5.901968 | 0.215903 | 0.999532 | | 7.779457 | 0.598196 | 0.308051 | | 7.137669 | 0.402657 | 0.621895 | |
| NBR1 | 7.635524 | 0.081318 | 0.999532 | | 10.24268 | 0.542275 | 0.308051 | | 9.071165 | 0.479975 | 0.513365 | |
| SNRPD3 | 9.680333 | 0.121215 | 0.999532 | | 12.20988 | 0.516194 | 0.308424 | | 11.03255 | 0.411174 | 0.539774 | |
| SNAPC1 | 6.020912 | -0.01101 | 0.999532 | | 5.683913 | -0.69238 | 0.308628 | | 4.779887 | -0.64604 | 0.493137 | |
| COX6C | 11.46909 | 0.239024 | 0.999532 | | 14.33316 | 0.499662 | 0.30883 | | 12.79692 | 0.283602 | 0.718735 | |
| SLC38A2 | 3.058419 | 0.616409 | 0.999532 | | 4.019179 | 0.950176 | 0.308898 | | 3.834044 | 0.389909 | 0.794473 | |
| PAH | 2.828992 | 0.406352 | 0.999532 | | 4.754666 | 1.31199 | 0.309119 | | 4.417122 | 0.930681 | 0.484303 | |
| UBE4B | 3.346453 | 0.44226 | 0.999532 | | 4.113305 | 0.781052 | 0.309119 | | 4.003699 | 0.350949 | 0.765852 | |
| SND1 | 9.24558 | 0.426392 | 0.999532 | | 10.80715 | 0.520866 | 0.309119 | | 10.64363 | 0.115994 | 0.900848 | |
| CTSF | 5.29858 | -0.02792 | 0.999532 | | 7.729863 | 0.699861 | 0.312059 | | 6.600673 | 0.752866 | 0.254225 | |
| RRS1 | 2.383612 | 0.483119 | 0.999532 | | 3.31719 | 0.930675 | 0.312059 | | 3.201733 | 0.474521 | 0.724386 | |
| ADD3 | 11.0823 | 0.237924 | 0.999532 | | 13.26121 | 0.492183 | 0.312109 | | 12.04155 | 0.274992 | 0.702463 | |
| CAMP | 3.996197 | 0.013825 | 0.999532 | | 3.362081 | -0.89327 | 0.312784 | | 2.948472 | -0.89636 | 0.426561 | |
| CNNM4 | 3.239783 | -0.13007 | 0.999532 | | 2.884279 | -0.95611 | 0.312784 | | 2.398768 | -0.80742 | 0.534852 | |
| TGOLN2 | 11.67866 | 0.278498 | 0.999532 | | 14.28746 | 0.518763 | 0.312784 | | 12.94222 | 0.2606 | 0.759211 | |
| LRP1 | 9.832473 | -0.33417 | 0.999532 | | 14.71289 | 0.506536 | 0.313154 | | 11.22378 | 0.854863 | 0.154583 | |
| PSMD4 | 8.140787 | 0.154557 | 0.999532 | | 10.68777 | 0.544077 | 0.313384 | | 9.6851 | 0.408691 | 0.542124 | |
| OR8G1 | 5.502296 | -0.16327 | 0.999532 | | 5.252041 | -0.73774 | 0.313384 | | 4.206193 | -0.55553 | 0.611214 | |
| SERPINB | 14.78326 | -0.26736 | 0.999532 | | 21.53975 | 0.454496 | 0.313785 | | 17.08776 | 0.746571 | 0.107433 | |
| AFF1 | 8.131868 | -0.3482 | 0.999532 | | 12.52612 | 0.490739 | 0.313785 | | 9.441595 | 0.865896 | 0.150518 | |
| SMARCA | 7.367545 | 0.172181 | 0.999532 | | 9.441947 | 0.571383 | 0.313785 | | 8.35611 | 0.417486 | 0.575547 | |
| SMIM26 | 4.35784 | 0.269197 | 0.999532 | | 5.855753 | 0.67847 | 0.313785 | | 5.282894 | 0.426013 | 0.647218 | |
| BORCS7 | 3.001551 | 0.382782 | 0.999532 | | 3.947315 | 0.795452 | 0.313785 | | 3.67264 | 0.427647 | 0.709825 | |
| PADI1 | 2.592291 | 0.026336 | 0.999532 | | 2.160391 | -1.12484 | 0.313785 | | 1.86842 | -1.13439 | 0.397994 | |
| SRP9 | 12.90691 | 0.222107 | 0.999532 | | 15.37999 | 0.435487 | 0.313785 | | 14.36364 | 0.233155 | 0.720106 | |
| PRAMEF | 3.619641 | -0.02247 | 0.999532 | | 3.027468 | -0.95932 | 0.313785 | | 2.651036 | -0.91599 | 0.416028 | |
| TSG101 | 2.342597 | 0.358594 | 0.999532 | | 3.189992 | 0.881278 | 0.313907 | | 2.866429 | 0.536641 | 0.660958 | |
| RPS8 | 35.38257 | 0.079968 | 0.999532 | | 46.0452 | 0.468305 | 0.313938 | | 39.9587 | 0.413775 | 0.515762 | |
| OAT | 3.258342 | 0.666322 | 0.999532 | | 3.986765 | 0.780457 | 0.313938 | | 4.105749 | 0.130666 | 0.925486 | |
| TMCO2 | 2.965034 | -0.28253 | 0.999532 | | 2.771987 | -0.99107 | 0.313938 | | 2.160409 | -0.68974 | 0.655993 | |
| ZNF346 | 12.11311 | -0.15249 | 0.999532 | | 11.99124 | -0.59776 | 0.313938 | | 9.814156 | -0.42254 | 0.660097 | |
| ELOVL1 | 5.317294 | 0.592571 | 0.999532 | | 6.095664 | 0.652764 | 0.314269 | | 6.276093 | 0.078911 | 0.953948 | |
| BMP7 | 6.437044 | 0.484904 | 0.999532 | | 7.782825 | 0.630931 | 0.314462 | | 7.299026 | 0.16371 | 0.883792 | |
| TMEM190 | 2.477023 | -0.56639 | 0.999532 | | 2.480695 | -1.06588 | 0.314462 | | 1.744068 | -0.48396 | 0.792416 | |
| CALR | 47.08293 | 0.109534 | 0.999532 | | 62.45959 | 0.536218 | 0.314462 | | 54.92721 | 0.447583 | 0.451915 | |
| CFI | 2.378455 | 0.740573 | 0.999532 | | 2.841204 | 0.923357 | 0.314462 | | 2.852721 | 0.202298 | 0.895006 | |
| CRABP1 | 3.13037 | 0.574098 | 0.999532 | | 2.237293 | -1.13793 | 0.314462 | | 2.329974 | -1.69121 | 0.107433 | |
| PIGT | 5.622429 | 0.261443 | 0.999532 | | 7.431267 | 0.612361 | 0.314462 | | 6.587491 | 0.356788 | 0.722703 | |
| CHKA | 3.331865 | -0.12678 | 0.999532 | | 2.892869 | -0.91593 | 0.314783 | | 2.411093 | -0.77302 | 0.532088 | |
| KRT7 | 2.241013 | 0.683632 | 0.999532 | | 2.627123 | 0.960265 | 0.315273 | | 2.783237 | 0.287302 | 0.857394 | |
| RPL23 | 16.71828 | -0.22982 | 0.999532 | | 24.04451 | 0.499957 | 0.316978 | | 19.27895 | 0.743649 | 0.150518 | |
| AIF1 | 10.29225 | 0.432147 | 0.999532 | | 11.88477 | 0.526963 | 0.316978 | | 11.70798 | 0.115946 | 0.904801 | |
| CSNK1A1 | 3.212698 | -0.03104 | 0.999532 | | 2.675812 | -1.02358 | 0.316978 | | 2.235375 | -0.96385 | 0.455544 | |
| ABCC8 | 2.438802 | -0.82426 | 0.999532 | | 2.658163 | -1.0201 | 0.316978 | | 1.680488 | -0.17781 | 0.939549 | |
| MRPL22 | 2.676526 | 0.609281 | 0.999532 | | 3.486434 | 0.837715 | 0.319209 | | 3.49803 | 0.248 | 0.858767 | |
| FADD | 4.049821 | 0.537739 | 0.999532 | | 4.971672 | 0.695388 | 0.319209 | | 5.021611 | 0.175267 | 0.881837 | |
| CROT | 3.871219 | 0.166901 | 0.999532 | | 3.258083 | -0.89885 | 0.319209 | | 2.863851 | -1.04124 | 0.282041 | |
| VCAM1 | 5.421356 | -0.53177 | 0.999532 | | 9.289008 | 0.674956 | 0.319209 | | 6.969818 | 1.226309 | 0.075829 | UP |
| CNOT2 | 3.886289 | 0.486853 | 0.999532 | | 4.757908 | 0.741261 | 0.319372 | | 4.510003 | 0.278146 | 0.803301 | |
| CTCF | 2.943716 | 0.360294 | 0.999532 | | 3.945404 | 0.792798 | 0.319372 | | 3.725934 | 0.448164 | 0.723933 | |
| SPECC1 | 2.537968 | 0.658551 | 0.999532 | | 3.239765 | 0.881015 | 0.319419 | | 3.355387 | 0.241444 | 0.869579 | |
| PDILT | 3.499475 | -0.71208 | 0.999532 | | 3.946182 | -0.83786 | 0.319419 | | 2.382872 | -0.09827 | 0.963492 | |
| ATXN10 | 8.385163 | 0.467653 | 0.999532 | | 9.430106 | 0.556561 | 0.319495 | | 9.354276 | 0.103552 | 0.920527 | |
| SSX4B | 3.857516 | 0.040992 | 0.999532 | | 3.31893 | -0.85548 | 0.320635 | | 2.97513 | -0.87882 | 0.397548 | |
| DPH1 | 6.001276 | 0.243525 | 0.999532 | | 7.653618 | 0.637899 | 0.320635 | | 7.066994 | 0.419831 | 0.630705 | |
| RNF225 | 2.908315 | 0.017003 | 0.999532 | | 2.356591 | -1.06229 | 0.320635 | | 2.214308 | -1.06669 | 0.422058 | |
| ALDH4A1 | 6.037163 | -0.18506 | 0.999532 | | 9.603957 | 0.695628 | 0.320635 | | 7.568633 | 0.912682 | 0.323846 | |
| RAB5C | 18.24827 | -0.00946 | 0.999532 | | 24.17535 | 0.455728 | 0.320635 | | 20.72065 | 0.482519 | 0.333968 | |
| TUBB3 | 2.228196 | -0.99519 | 0.999532 | | 2.532941 | -0.97925 | 0.320635 | | 1.580576 | 0.03637 | 0.992286 | |
| GPR84 | 2.530963 | -0.4199 | 0.999532 | | 2.380293 | -1.06042 | 0.321983 | | 1.791613 | -0.62328 | 0.717138 | |
| MED8 | 3.066649 | 0.457839 | 0.999532 | | 3.917955 | 0.809362 | 0.322176 | | 3.746887 | 0.366378 | 0.759211 | |
| DECR1 | 6.777452 | 0.253075 | 0.999532 | | 8.331015 | 0.567581 | 0.322176 | | 7.587204 | 0.312775 | 0.760572 | |
| SLC2A4R | 5.676479 | 0.205878 | 0.999532 | | 7.17955 | 0.59909 | 0.322665 | | 6.642896 | 0.402288 | 0.652925 | |
| OAZ1 | 36.08001 | -0.09805 | 0.999532 | | 50.28543 | 0.480691 | 0.323274 | | 41.64676 | 0.600266 | 0.188098 | |
| HOXC12 | 2.429496 | -0.27699 | 0.999532 | | 2.148864 | -1.07959 | 0.323724 | | 1.824522 | -0.78907 | 0.630705 | |
| NUTM2G | 9.648274 | 0.129543 | 0.999532 | | 8.640155 | -0.64187 | 0.32417 | | 7.763384 | -0.75614 | 0.264219 | |
| GNPDA2 | 3.782408 | -0.04497 | 0.999532 | | 3.202523 | -0.88677 | 0.324328 | | 2.875395 | -0.81981 | 0.483244 | |
| NEGR1 | 4.294149 | 0.331193 | 0.999532 | | 5.519333 | 0.712542 | 0.324328 | | 5.051491 | 0.403075 | 0.682968 | |
| BTAF1 | 4.800686 | 0.049019 | 0.999532 | | 6.993047 | 0.644039 | 0.324328 | | 5.781401 | 0.62074 | 0.456688 | |
| FKBP8 | 9.400038 | 0.068302 | 0.999532 | | 12.18949 | 0.561056 | 0.324328 | | 10.79619 | 0.483727 | 0.536289 | |
| CRAT | 5.40224 | 0.391709 | 0.999532 | | 6.934758 | 0.666538 | 0.324328 | | 6.548706 | 0.300902 | 0.790999 | |
| RTF2 | 5.43023 | 0.37864 | 0.999532 | | 6.522227 | 0.6232 | 0.324328 | | 6.274227 | 0.253976 | 0.821113 | |
| PPP6R1 | 5.162737 | 0.020041 | 0.999532 | | 7.673989 | 0.61195 | 0.324328 | | 6.426084 | 0.612942 | 0.445382 | |
| PPP1R26 | 5.169046 | 0.03391 | 0.999532 | | 4.759625 | -0.80042 | 0.325612 | | 4.210455 | -0.80919 | 0.418768 | |
| ANO8 | 4.041687 | -0.43264 | 0.999532 | | 4.426599 | -0.7938 | 0.325612 | | 2.817384 | -0.34877 | 0.806394 | |
| METRNL | 5.549878 | -0.26823 | 0.999532 | | 5.775621 | -0.73609 | 0.325612 | | 4.368381 | -0.43102 | 0.718643 | |
| CELA2A | 2.34428 | -0.4742 | 0.999532 | | 2.321184 | -1.04942 | 0.325612 | | 1.669051 | -0.55848 | 0.750371 | |
| SUPT5H | 7.440852 | 0.400624 | 0.999532 | | 8.477625 | 0.552586 | 0.325612 | | 8.173708 | 0.172293 | 0.855083 | |
| TMTC1 | 9.634606 | 0.091378 | 0.999532 | | 12.82834 | 0.561194 | 0.325612 | | 11.49174 | 0.479088 | 0.520178 | |
| ZNF726 | 3.566576 | -0.33054 | 0.999532 | | 3.540171 | -0.95599 | 0.325612 | | 2.506958 | -0.60378 | 0.661666 | |
| TTLL1 | 2.851212 | -0.23987 | 0.999532 | | 2.611522 | -0.94626 | 0.325612 | | 2.000492 | -0.6876 | 0.630705 | |
| RAMP2 | 7.727962 | -0.02449 | 0.999532 | | 11.14541 | 0.638775 | 0.325612 | | 9.441375 | 0.688777 | 0.282041 | |
| TMEM9B | 5.00344 | 0.378579 | 0.999532 | | 6.012922 | 0.649368 | 0.325612 | | 5.898404 | 0.288015 | 0.776852 | |
| MSMP | 2.395736 | -0.90708 | 0.999532 | | 2.601892 | -0.98419 | 0.326363 | | 1.680043 | -0.0626 | 0.98514 | |
| HMGN2 | 39.03289 | 0.003275 | 0.999532 | | 39.89996 | -0.29015 | 0.326363 | | 34.05211 | -0.27262 | 0.486743 | |
| IL1RL1 | 30.80999 | -0.97254 | 0.124973 | | 39.00354 | -0.48487 | 0.326374 | | 23.56923 | 0.49251 | 0.577942 | |
| SLC6A9 | 3.075338 | 0.107678 | 0.999532 | | 2.507156 | -0.99641 | 0.326374 | | 2.201358 | -1.08456 | 0.340167 | |
| RRM1 | 2.640126 | 0.074728 | 0.999532 | | 2.198084 | -1.02948 | 0.326374 | | 1.964949 | -1.08648 | 0.369628 | |
| MRPL33 | 8.230706 | 0.23269 | 0.999532 | | 9.789549 | 0.541783 | 0.326374 | | 9.081053 | 0.30519 | 0.735469 | |
| BAX | 6.586341 | -0.08134 | 0.999532 | | 9.33665 | 0.58698 | 0.326374 | | 7.964208 | 0.680976 | 0.300261 | |
| SPATA46 | 4.217735 | -0.00032 | 0.999532 | | 3.740186 | -0.83916 | 0.326374 | | 3.173515 | -0.81917 | 0.456688 | |
| KCTD12 | 2.584972 | -0.49035 | 0.999532 | | 4.250592 | 0.791415 | 0.326374 | | 3.356536 | 1.278948 | 0.15668 | |
| ST6GALN | 25.01376 | 0.16405 | 0.999532 | | 30.40256 | 0.392622 | 0.326374 | | 27.30955 | 0.254897 | 0.589729 | |
| SPIN2B | 6.576316 | -0.22607 | 0.999532 | | 6.553212 | -0.63898 | 0.326374 | | 5.08956 | -0.39505 | 0.708052 | |
| SLC25A4 | 2.09602 | 0.600554 | 0.999532 | | 2.70039 | 0.928558 | 0.326374 | | 2.712541 | 0.346384 | 0.809832 | |
| PACSIN2 | 6.464947 | 0.006307 | 0.999532 | | 8.797468 | 0.534298 | 0.326374 | | 7.61992 | 0.54685 | 0.436389 | |
| RIDA | 3.899237 | 0.464763 | 0.999532 | | 4.926895 | 0.918946 | 0.326374 | | 4.810624 | 0.446715 | 0.764149 | |
| PLXNB2 | 12.09202 | -0.01595 | 0.999532 | | 16.32253 | 0.487143 | 0.32813 | | 13.60731 | 0.530894 | 0.344798 | |
| HCLS1 | 3.024141 | 0.059864 | 0.999532 | | 4.060187 | 0.782072 | 0.328927 | | 3.749745 | 0.706232 | 0.519358 | |
| RHOD | 3.672689 | -0.12543 | 0.999532 | | 3.353093 | -0.85601 | 0.329255 | | 2.708817 | -0.71185 | 0.542124 | |
| MT1A | 11.91098 | -0.51848 | 0.999532 | | 13.19699 | -0.65044 | 0.329255 | | 8.885494 | -0.118 | 0.907968 | |
| CGN | 2.342576 | 0.52515 | 0.999532 | | 2.992463 | 0.858204 | 0.329255 | | 2.911937 | 0.348987 | 0.793131 | |
| CSKMT | 3.164593 | -0.42459 | 0.999532 | | 3.115535 | -0.91447 | 0.329378 | | 2.378845 | -0.47481 | 0.758124 | |
| DYNLL1 | 23.05283 | 0.006845 | 0.999532 | | 29.70268 | 0.368245 | 0.329378 | | 25.13762 | 0.381144 | 0.476841 | |
| SMYD1 | 2.549327 | -0.56159 | 0.999532 | | 2.594573 | -1.0473 | 0.329378 | | 1.775019 | -0.46571 | 0.799944 | |
| SELENOI | 16.94633 | 0.001786 | 0.999532 | | 22.48688 | 0.454156 | 0.329378 | | 19.22916 | 0.476222 | 0.374565 | |
| HLA-C | 42.40101 | 0.06947 | 0.999532 | | 55.58323 | 0.493274 | 0.329378 | | 48.64449 | 0.442003 | 0.459697 | |
| TNKS2 | 5.64529 | 0.196766 | 0.999532 | | 7.039974 | 0.579994 | 0.329378 | | 6.547741 | 0.405477 | 0.630278 | |
| RPL9 | 25.49606 | 0.021983 | 0.999532 | | 33.64486 | 0.476233 | 0.329378 | | 29.09962 | 0.477806 | 0.263754 | |
| POLR2G | 4.235927 | -0.00777 | 0.999532 | | 5.882292 | 0.638818 | 0.329378 | | 5.200824 | 0.667089 | 0.436389 | |
| TFPI | 10.49906 | -0.03733 | 0.999532 | | 13.98399 | 0.46903 | 0.329378 | | 11.84706 | 0.524003 | 0.362433 | |
| TMEM94 | 4.975342 | 0.596946 | 0.999532 | | 5.971238 | 0.669455 | 0.329378 | | 5.889663 | 0.095611 | 0.946572 | |
| WDTC1 | 4.278999 | 0.244215 | 0.999532 | | 5.770943 | 0.666697 | 0.329378 | | 5.207709 | 0.445116 | 0.647581 | |
| AGAP9 | 10.53185 | 0.127551 | 0.999532 | | 9.266633 | -0.54575 | 0.329378 | | 8.523413 | -0.66536 | 0.289131 | |
| H3Y2 | 3.041076 | -0.71552 | 0.999532 | | 3.269128 | -0.94162 | 0.329378 | | 2.042694 | -0.20514 | 0.915533 | |
| OR10A2 | 2.712258 | -0.05519 | 0.999532 | | 2.439654 | -1.06059 | 0.329378 | | 1.883308 | -0.98232 | 0.465298 | |
| TFG | 10.88357 | 0.104243 | 0.999532 | | 14.2056 | 0.498817 | 0.329378 | | 12.38817 | 0.410363 | 0.546858 | |
| ADH1B | 5.709432 | 0.626536 | 0.999532 | | 6.460884 | 0.727684 | 0.329378 | | 6.865491 | 0.109741 | 0.942695 | |
| CCDC51 | 3.567808 | -0.05266 | 0.999532 | | 3.420244 | -0.87396 | 0.329378 | | 2.779904 | -0.77811 | 0.532278 | |
| NRG4 | 3.819589 | 0.109035 | 0.999532 | | 3.110583 | -0.8993 | 0.329378 | | 2.897714 | -0.99487 | 0.359441 | |
| PLEKHO1 | 6.754125 | 0.250889 | 0.999532 | | 9.053063 | 0.600251 | 0.329378 | | 8.181064 | 0.37444 | 0.668481 | |
| SOS1 | 4.846201 | -0.04559 | 0.999532 | | 6.578775 | 0.648526 | 0.329378 | | 5.887839 | 0.69323 | 0.421517 | |
| RNF175 | 3.043949 | -0.35312 | 0.999532 | | 2.815005 | -0.94195 | 0.329596 | | 2.077434 | -0.57381 | 0.702463 | |
| SUGP2 | 5.10479 | -0.058 | 0.999532 | | 7.161264 | 0.586489 | 0.329634 | | 5.899102 | 0.658759 | 0.366512 | |
| ALOX12B | 2.60971 | -0.0411 | 0.999532 | | 2.240162 | -1.08937 | 0.329634 | | 1.829441 | -1.02889 | 0.452319 | |
| OR2T29 | 3.369879 | -0.33127 | 0.999532 | | 3.413632 | -0.92591 | 0.329918 | | 2.514309 | -0.56834 | 0.698562 | |
| TFF1 | 2.55881 | -0.64395 | 0.999532 | | 2.633353 | -1.00265 | 0.330751 | | 1.683042 | -0.34097 | 0.863979 | |
| BAP1 | 7.006222 | 0.098261 | 0.999532 | | 9.247095 | 0.531958 | 0.330751 | | 8.144789 | 0.452731 | 0.540342 | |
| IGF2 | 3.816418 | 0.269228 | 0.999532 | | 5.526743 | 0.81936 | 0.330751 | | 5.011125 | 0.569909 | 0.60629 | |
| ATG4B | 6.284404 | 0.169777 | 0.999532 | | 8.37257 | 0.564721 | 0.33116 | | 7.450242 | 0.414222 | 0.60606 | |
| MARS2 | 2.904819 | -0.35808 | 0.999532 | | 2.66042 | -0.99297 | 0.33116 | | 2.137623 | -0.61938 | 0.699971 | |
| TRIM64B | 4.216036 | -0.02496 | 0.999532 | | 3.739531 | -0.83962 | 0.331232 | | 3.188851 | -0.79728 | 0.465298 | |
| KRTAP19 | 3.418056 | -0.01479 | 0.999532 | | 2.910407 | -0.92577 | 0.331232 | | 2.579308 | -0.89551 | 0.465298 | |
| DRD5 | 5.861069 | -0.54508 | 0.999532 | | 6.293732 | -0.77924 | 0.331232 | | 4.527165 | -0.22888 | 0.881837 | |
| DOC2B | 3.573952 | 0.463487 | 0.999532 | | 4.463142 | 0.741615 | 0.331232 | | 4.467471 | 0.292532 | 0.808571 | |
| H2AJ | 9.580871 | 0.276661 | 0.999532 | | 12.19821 | 0.523128 | 0.331971 | | 11.169 | 0.26807 | 0.735233 | |
| IFI16 | 8.692243 | -0.21636 | 0.999532 | | 13.46945 | 0.651126 | 0.331971 | | 10.85577 | 0.894484 | 0.186007 | |
| PEBP1 | 15.04833 | 0.278796 | 0.999532 | | 17.01382 | 0.415665 | 0.331971 | | 16.05473 | 0.136948 | 0.872591 | |
| C5orf34 | 2.721913 | -0.32035 | 0.999532 | | 2.628595 | -1.01642 | 0.331971 | | 1.921484 | -0.6752 | 0.661666 | |
| S100A10 | 10.13241 | -0.00805 | 0.999532 | | 14.05996 | 0.52116 | 0.331971 | | 11.69736 | 0.561185 | 0.402522 | |
| SLC35G6 | 4.017886 | -0.38485 | 0.999532 | | 4.033663 | -0.82106 | 0.331971 | | 3.010279 | -0.41548 | 0.776852 | |
| VPS8 | 3.545067 | 0.498831 | 0.999532 | | 4.368249 | 0.74196 | 0.331971 | | 4.158622 | 0.261538 | 0.831749 | |
| ENTR1 | 4.046349 | -0.26881 | 0.999532 | | 3.805923 | -0.80371 | 0.331971 | | 3.145026 | -0.52851 | 0.67937 | |
| EMC2 | 6.354907 | 0.510407 | 0.999532 | | 7.191898 | 0.59057 | 0.332398 | | 7.175446 | 0.097668 | 0.934572 | |
| RPL15 | 46.9096 | -0.03965 | 0.999532 | | 60.54089 | 0.348868 | 0.332398 | | 50.70831 | 0.411548 | 0.3556 | |
| HTRA1 | 74.19793 | 0.546415 | 0.999532 | | 83.9709 | 0.561271 | 0.332398 | | 84.51614 | 0.041523 | 0.97179 | |
| IGF2R | 5.028848 | 0.032087 | 0.999532 | | 7.405809 | 0.657006 | 0.332398 | | 6.244071 | 0.652292 | 0.468942 | |
| SERPINH | 8.422972 | 0.093148 | 0.999532 | | 11.18568 | 0.588094 | 0.332398 | | 9.656929 | 0.524277 | 0.515762 | |
| POMP | 7.201755 | -0.27974 | 0.999532 | | 10.15401 | 0.508342 | 0.332682 | | 8.298632 | 0.797723 | 0.175031 | |
| RBM33 | 2.374228 | 0.094657 | 0.999532 | | 3.507923 | 0.784492 | 0.333207 | | 3.042877 | 0.709227 | 0.516948 | |
| SUMO3 | 5.211229 | 0.008561 | 0.999532 | | 7.695066 | 0.658429 | 0.334394 | | 6.38623 | 0.669494 | 0.397004 | |
| XPO1 | 4.671752 | 0.487918 | 0.999532 | | 5.569459 | 0.647441 | 0.335281 | | 5.604358 | 0.178112 | 0.880577 | |
| FAM149A | 2.77563 | 1.146446 | 0.871349 | | 2.868015 | 0.913132 | 0.335853 | | 3.343268 | -0.2192 | 0.878742 | |
| ATP6V1E | 14.16283 | -0.06886 | 0.999532 | | 19.25464 | 0.437095 | 0.336198 | | 15.95267 | 0.528516 | 0.266933 | |
| EMILIN1 | 4.820605 | 0.161684 | 0.999532 | | 6.450096 | 0.674519 | 0.336198 | | 5.795289 | 0.528171 | 0.565022 | |
| TFB2M | 3.113533 | -0.47859 | 0.999532 | | 2.936533 | -0.91032 | 0.337286 | | 2.192114 | -0.42677 | 0.799627 | |
| CAT | 6.264458 | 0.024774 | 0.999532 | | 8.794888 | 0.576908 | 0.337286 | | 7.545641 | 0.56718 | 0.410504 | |
| DDN | 13.66309 | 0.575486 | 0.999532 | | 16.30841 | 0.728628 | 0.337286 | | 16.52315 | 0.175157 | 0.894612 | |
| CLDN2 | 3.122197 | 0.388398 | 0.999532 | | 4.077761 | 0.783923 | 0.337286 | | 3.76388 | 0.410329 | 0.724411 | |
| SMIM28 | 3.483527 | -0.36863 | 0.999532 | | 3.572199 | -0.85269 | 0.337736 | | 2.578811 | -0.45699 | 0.750371 | |
| FAM86B1 | 6.821646 | -0.06157 | 0.999532 | | 6.551904 | -0.64799 | 0.33784 | | 5.073789 | -0.56406 | 0.53177 | |
| ECE2 | 4.83487 | -0.14389 | 0.999532 | | 4.528997 | -0.75611 | 0.33784 | | 3.848722 | -0.6005 | 0.573903 | |
| WDR6 | 7.076803 | 0.527387 | 0.999532 | | 8.392469 | 0.551021 | 0.338643 | | 7.940886 | 0.047221 | 0.974421 | |
| WNT8A | 12.87846 | 0.066031 | 0.999532 | | 11.85753 | -0.51129 | 0.339219 | | 10.57415 | -0.56012 | 0.387954 | |
| TRIM49D | 3.413899 | 0.078683 | 0.999532 | | 2.888611 | -0.91249 | 0.339219 | | 2.660971 | -0.97597 | 0.394331 | |
| CAPZB | 7.896154 | -0.11725 | 0.999532 | | 11.17465 | 0.545344 | 0.339702 | | 9.286309 | 0.673031 | 0.323635 | |
| MCRIP2 | 11.18307 | 0.228589 | 0.999532 | | 9.666992 | -0.59473 | 0.339702 | | 9.1324 | -0.8055 | 0.222058 | |
| POTEG | 4.695848 | 0.054227 | 0.999532 | | 4.144315 | -0.85601 | 0.339702 | | 3.741932 | -0.89623 | 0.386771 | |
| TTC21A | 3.335202 | -0.08935 | 0.999532 | | 2.886669 | -0.92239 | 0.340005 | | 2.402043 | -0.81697 | 0.524542 | |
| TMEM258 | 11.73516 | -0.06445 | 0.999532 | | 15.35549 | 0.449908 | 0.340005 | | 13.0241 | 0.531405 | 0.342845 | |
| CALCOCO | 8.217316 | -0.06888 | 0.999532 | | 11.44286 | 0.533308 | 0.340248 | | 9.520023 | 0.620426 | 0.350296 | |
| AQR | 3.493605 | -0.85605 | 0.999532 | | 3.942934 | -0.79815 | 0.340248 | | 2.341981 | 0.074001 | 0.974421 | |
| OR5V1 | 4.146756 | -0.59198 | 0.999532 | | 4.308461 | -0.78959 | 0.340248 | | 3.052326 | -0.19223 | 0.91411 | |
| CLCN7 | 4.140043 | 0.331397 | 0.999532 | | 5.180507 | 0.653183 | 0.340248 | | 4.864719 | 0.337474 | 0.737426 | |
| TACC2 | 7.838661 | 0.57425 | 0.999532 | | 8.659047 | 0.574576 | 0.341618 | | 8.712178 | 0.009332 | 0.996891 | |
| KRTAP12 | 6.531284 | 0.218243 | 0.999532 | | 5.609128 | -0.70443 | 0.341746 | | 5.283604 | -0.90146 | 0.263754 | |
| CXCL9 | 8.5524 | -0.36526 | 0.999532 | | 7.909462 | -1.10196 | 0.341746 | | 5.721777 | -0.70932 | 0.540342 | |
| EIF5AL1 | 2.499659 | -0.34614 | 0.999532 | | 2.41475 | -1.01426 | 0.341746 | | 1.732715 | -0.64797 | 0.686214 | |
| ARL2 | 8.023451 | 0.190982 | 0.999532 | | 10.35866 | 0.479384 | 0.341838 | | 9.055987 | 0.302249 | 0.70257 | |
| TNFAIP1 | 7.045206 | -0.05402 | 0.999532 | | 9.669397 | 0.540523 | 0.342319 | | 8.132612 | 0.61298 | 0.35437 | |
| ABCA8 | 1.993613 | 0.165357 | 0.999532 | | 2.816662 | 0.873408 | 0.343043 | | 2.546664 | 0.728295 | 0.540342 | |
| TRIM11 | 3.59302 | -0.00151 | 0.999532 | | 3.012344 | -0.87304 | 0.343043 | | 2.601368 | -0.85662 | 0.456688 | |
| IGFBP5 | 263.6921 | -0.03403 | 0.999532 | | 348.9679 | 0.407494 | 0.343147 | | 294.4015 | 0.469682 | 0.304062 | |
| METTL9 | 4.068482 | 0.220396 | 0.999532 | | 5.180893 | 0.672188 | 0.343147 | | 4.818763 | 0.473104 | 0.632802 | |
| S1PR3 | 6.595151 | -0.24811 | 0.999532 | | 6.234959 | -0.68303 | 0.343147 | | 4.806407 | -0.40217 | 0.686214 | |
| ZNF91 | 4.332636 | 0.508815 | 0.999532 | | 5.154734 | 0.689631 | 0.34406 | | 5.195996 | 0.200416 | 0.885892 | |
| ERLIN2 | 3.326842 | 1.054376 | 0.635439 | | 3.298223 | 0.787979 | 0.344576 | | 3.915325 | -0.24887 | 0.839554 | |
| LAG3 | 3.21541 | -0.54442 | 0.999532 | | 3.280868 | -0.83695 | 0.344591 | | 2.283702 | -0.27638 | 0.872591 | |
| LMO2 | 13.4402 | 0.263619 | 0.999532 | | 16.73248 | 0.543978 | 0.344591 | | 15.67474 | 0.304489 | 0.647218 | |
| ZNF564 | 3.245424 | -0.15468 | 0.999532 | | 2.898182 | -0.86415 | 0.344816 | | 2.414547 | -0.69831 | 0.579378 | |
| PNISR | 9.383816 | 0.283628 | 0.999532 | | 11.66123 | 0.516915 | 0.344816 | | 10.74164 | 0.25504 | 0.755206 | |
| SLCO3A1 | 6.944214 | -0.02399 | 0.999532 | | 9.769247 | 0.53038 | 0.346271 | | 8.210111 | 0.575727 | 0.421517 | |
| BHMT2 | 11.94898 | 0.103653 | 0.999532 | | 17.79975 | 0.784567 | 0.346271 | | 15.34148 | 0.718333 | 0.587187 | |
| OBP2B | 2.272049 | -0.65517 | 0.999532 | | 2.449244 | -0.98314 | 0.346271 | | 1.665275 | -0.3049 | 0.881837 | |
| PPP1R9B | 3.695935 | 0.326947 | 0.999532 | | 4.623443 | 0.749104 | 0.346271 | | 4.543656 | 0.444984 | 0.672584 | |
| BMP8A | 3.844922 | -0.11421 | 0.999532 | | 3.519679 | -0.80362 | 0.346271 | | 2.851589 | -0.67122 | 0.552672 | |
| RPL8 | 26.03332 | -0.06607 | 0.999532 | | 34.83555 | 0.440138 | 0.346271 | | 28.99485 | 0.524926 | 0.359441 | |
| SMAD1 | 5.241337 | 0.706228 | 0.999532 | | 5.979836 | 0.724929 | 0.346638 | | 6.401347 | 0.025383 | 0.991103 | |
| SLC15A1 | 3.763907 | 0.271483 | 0.999532 | | 3.036237 | -0.88308 | 0.346732 | | 2.805737 | -1.13446 | 0.274496 | |
| RNF24 | 3.999494 | -0.35464 | 0.999532 | | 4.017859 | -0.77862 | 0.346732 | | 3.078517 | -0.40832 | 0.760136 | |
| C9orf152 | 2.891457 | -0.35124 | 0.999532 | | 2.974099 | -0.91454 | 0.347442 | | 2.107603 | -0.5331 | 0.724646 | |
| MTCH2 | 5.30294 | -0.01117 | 0.999532 | | 7.125818 | 0.5934 | 0.347442 | | 6.23412 | 0.628838 | 0.415282 | |
| ZNF83 | 4.470851 | 0.231899 | 0.999532 | | 5.950193 | 0.67454 | 0.348299 | | 5.333581 | 0.465 | 0.628695 | |
| KLK7 | 10.7959 | 0.250205 | 0.999532 | | 14.14956 | 0.598605 | 0.348375 | | 12.84069 | 0.380072 | 0.647218 | |
| SRRM2 | 25.94828 | 0.167858 | 0.999532 | | 30.94389 | 0.357333 | 0.3486 | | 27.6234 | 0.197682 | 0.784779 | |
| RPL37A | 47.52969 | -0.08342 | 0.999532 | | 63.96109 | 0.418181 | 0.3486 | | 53.03673 | 0.524713 | 0.274496 | |
| TSPYL1 | 7.283305 | 0.06183 | 0.999532 | | 9.925227 | 0.547165 | 0.3486 | | 8.427459 | 0.504646 | 0.50916 | |
| MDH2 | 7.262471 | 0.062325 | 0.999532 | | 9.292826 | 0.563689 | 0.3486 | | 8.16125 | 0.508533 | 0.536289 | |
| RPS18 | 21.80114 | -0.23486 | 0.999532 | | 31.00029 | 0.431087 | 0.3486 | | 24.64791 | 0.690445 | 0.121156 | |
| POLR2F | 3.797041 | 0.07383 | 0.999532 | | 5.587573 | 0.661475 | 0.349218 | | 4.757198 | 0.609023 | 0.515762 | |
| PRAC1 | 6.690262 | -0.2195 | 0.999532 | | 6.918267 | -0.5988 | 0.349218 | | 5.241251 | -0.35789 | 0.71343 | |
| FGD3 | 2.822555 | 0.485909 | 0.999532 | | 3.536585 | 0.793671 | 0.349218 | | 3.456733 | 0.324061 | 0.801905 | |
| YY1AP1 | 4.198317 | 0.099247 | 0.999532 | | 5.971924 | 0.616549 | 0.349218 | | 5.086946 | 0.532271 | 0.540342 | |
| ATXN2L | 7.418173 | 0.133947 | 0.999532 | | 9.850051 | 0.566049 | 0.349218 | | 8.69141 | 0.451419 | 0.621015 | |
| LILRB3 | 4.343578 | -0.13242 | 0.999532 | | 3.820328 | -0.80107 | 0.349218 | | 3.376839 | -0.64506 | 0.570037 | |
| NR2F2 | 29.42761 | 0.181072 | 0.999532 | | 38.41533 | 0.574269 | 0.349862 | | 34.67109 | 0.411728 | 0.572756 | |
| CRYAB | 19.36413 | 0.262918 | 0.999532 | | 24.46218 | 0.578659 | 0.349862 | | 22.45724 | 0.339738 | 0.66232 | |
| AP2B1 | 12.80935 | -0.17742 | 0.999532 | | 18.94126 | 0.539756 | 0.349862 | | 15.4343 | 0.735027 | 0.154583 | |
| ATP2B1 | 5.040866 | 0.414033 | 0.999532 | | 6.267066 | 0.653354 | 0.351117 | | 6.014072 | 0.255324 | 0.827488 | |
| GLRX | 6.688333 | -0.20933 | 0.999532 | | 9.730611 | 0.506411 | 0.352762 | | 7.568561 | 0.736182 | 0.254511 | |
| AXIN1 | 4.920995 | -0.24781 | 0.999532 | | 4.906238 | -0.73295 | 0.352893 | | 3.814058 | -0.46524 | 0.699971 | |
| ZCCHC24 | 5.633735 | -0.1502 | 0.999532 | | 7.973475 | 0.541151 | 0.352893 | | 6.68574 | 0.707049 | 0.286063 | |
| ADIPOQ | 2.717692 | -0.34942 | 0.999532 | | 2.751166 | -0.94548 | 0.352893 | | 1.998223 | -0.57168 | 0.715922 | |
| SEC61B | 6.489591 | 0.055072 | 0.999532 | | 8.998676 | 0.530584 | 0.353429 | | 7.675101 | 0.492852 | 0.505689 | |
| NRGN | 22.25325 | 0.056523 | 0.999532 | | 29.70862 | 0.535839 | 0.353429 | | 26.12216 | 0.491861 | 0.522078 | |
| SIRT2 | 5.288601 | 0.041224 | 0.999532 | | 7.207426 | 0.563574 | 0.35361 | | 6.143314 | 0.540244 | 0.484303 | |
| TICRR | 2.732236 | 0.06702 | 0.999532 | | 2.16586 | -1.09473 | 0.353891 | | 1.911413 | -1.13774 | 0.397004 | |
| SMAD4 | 7.88052 | 0.106822 | 0.999532 | | 10.45417 | 0.474694 | 0.353891 | | 9.046814 | 0.386701 | 0.578798 | |

| | | | | | | | | | | | | |
|---|---|---|---|---|---|---|---|---|---|---|---|---|
| EMC4 | 6.247616 | -0.11785 | 0.999532 | | 8.95606 | 0.540821 | 0.353891 | | 7.58316 | 0.680732 | 0.282041 | |
| CAVIN1 | 9.704109 | -0.215 | 0.999532 | | 13.81855 | 0.541767 | 0.353891 | | 11.43324 | 0.76725 | 0.222058 | |
| CFAP157 | 3.56003 | -0.27744 | 0.999532 | | 3.305941 | -0.86314 | 0.354035 | | 2.498393 | -0.56376 | 0.672963 | |
| CLGN | 2.386608 | -0.14887 | 0.999532 | | 2.160508 | -1.03985 | 0.354133 | | 1.707268 | -0.87022 | 0.563729 | |
| ARHGEF | 7.289332 | 0.508021 | 0.999532 | | 8.315971 | 0.580574 | 0.354133 | | 8.505475 | 0.084814 | 0.947322 | |
| CD46 | 21.90927 | -0.09153 | 0.999532 | | 29.83373 | 0.384751 | 0.35421 | | 24.26843 | 0.496976 | 0.271669 | |
| TXNL1 | 6.451379 | 0.503869 | 0.999532 | | 7.721961 | 0.602139 | 0.35421 | | 7.613083 | 0.108251 | 0.920259 | |
| INSIG2 | 2.551339 | -0.26979 | 0.999532 | | 2.405224 | -0.97194 | 0.354257 | | 1.888149 | -0.67946 | 0.66232 | |
| CTTNBP2 | 3.318626 | 0.264486 | 0.999532 | | 4.681569 | 0.70882 | 0.354649 | | 4.1124 | 0.46433 | 0.666566 | |
| KITLG | 3.23339 | 0.200257 | 0.999532 | | 4.241579 | 0.78543 | 0.356139 | | 3.952831 | 0.576388 | 0.630705 | |
| RPS17 | 60.62182 | -0.06707 | 0.999532 | | 79.26321 | 0.386156 | 0.357597 | | 66.11125 | 0.453557 | 0.310637 | |
| ATP6AP2 | 9.256798 | 0.03568 | 0.999532 | | 12.48385 | 0.499017 | 0.358959 | | 10.65904 | 0.486564 | 0.485816 | |
| FAM107A | 4.511128 | -0.99273 | 0.831313 | | 5.291351 | -0.79185 | 0.359225 | | 2.957055 | 0.237553 | 0.886495 | |
| ZNF430 | 3.320163 | -0.07595 | 0.999532 | | 2.951243 | -0.92956 | 0.360435 | | 2.32262 | -0.83436 | 0.515762 | |
| EHHADH | 3.074383 | 0.006385 | 0.999532 | | 4.397591 | 0.758745 | 0.36064 | | 3.789081 | 0.758715 | 0.484303 | |
| TRAPPC1 | 3.354374 | -0.21521 | 0.999532 | | 3.117544 | -0.83322 | 0.36064 | | 2.469048 | -0.60256 | 0.647581 | |
| AKT1 | 3.46965 | 0.041778 | 0.999532 | | 5.259988 | 0.641029 | 0.361106 | | 4.276743 | 0.607884 | 0.515762 | |
| VEGFB | 8.705049 | 0.219212 | 0.999532 | | 11.10854 | 0.505622 | 0.361106 | | 10.16617 | 0.304065 | 0.666566 | |
| SCAMP2 | 4.951781 | -0.057 | 0.999532 | | 7.248016 | 0.569344 | 0.361106 | | 5.798475 | 0.647045 | 0.413122 | |
| CLTC | 23.88351 | -0.12967 | 0.999532 | | 32.88298 | 0.420889 | 0.361603 | | 26.81353 | 0.577464 | 0.168532 | |
| OR2S2 | 4.326427 | -0.15132 | 0.999532 | | 4.201362 | -0.86554 | 0.361603 | | 3.262465 | -0.7007 | 0.542124 | |
| PTPRS | 3.325115 | 0.454318 | 0.999532 | | 4.002174 | 0.731096 | 0.363112 | | 3.925493 | 0.294969 | 0.809832 | |
| SLC22A1 | 4.39088 | 0.477233 | 0.999532 | | 3.137174 | -0.86797 | 0.363112 | | 3.350395 | -1.32836 | 0.129363 | |
| MET | 2.311805 | 0.391919 | 0.999532 | | 3.047987 | 0.826939 | 0.363112 | | 2.86832 | 0.452032 | 0.722703 | |
| COX5A | 8.296773 | 0.205331 | 0.999532 | | 10.50872 | 0.487775 | 0.363112 | | 9.294424 | 0.296035 | 0.718633 | |
| PERM1 | 3.804067 | 0.541744 | 0.999532 | | 2.869162 | -0.9018 | 0.363884 | | 2.915993 | -1.42148 | 0.107433 | |
| CD63 | 39.11288 | -0.06221 | 0.999532 | | 54.78732 | 0.533647 | 0.363884 | | 45.92159 | 0.61546 | 0.295094 | |
| CCL24 | 4.569243 | -0.13107 | 0.999532 | | 4.283857 | -0.85958 | 0.363884 | | 3.465887 | -0.723 | 0.524206 | |
| ASAH2B | 2.589055 | -0.28972 | 0.999532 | | 2.498255 | -0.98398 | 0.363884 | | 1.880209 | -0.67374 | 0.665886 | |
| ATP10D | 3.561906 | 0.097443 | 0.999532 | | 5.289196 | 0.734212 | 0.364663 | | 4.726791 | 0.650741 | 0.535342 | |
| ADGRE1 | 2.895326 | -0.49596 | 0.999532 | | 2.831195 | -0.90465 | 0.364663 | | 2.078764 | -0.39577 | 0.815886 | |
| YWHAE | 35.38366 | 0.018228 | 0.999532 | | 46.23121 | 0.423447 | 0.364663 | | 39.15143 | 0.44006 | 0.454116 | |
| LOX | 3.56757 | 0.921815 | 0.999532 | | 3.634177 | 0.776735 | 0.365798 | | 4.184581 | -0.12499 | 0.933644 | |
| SIRPA | 5.654961 | 0.586441 | 0.999532 | | 6.598521 | 0.664115 | 0.366664 | | 6.502589 | 0.10033 | 0.941197 | |
| HK1 | 5.045612 | -0.06856 | 0.999532 | | 7.482602 | 0.563897 | 0.368719 | | 6.158998 | 0.648335 | 0.359638 | |
| DVL2 | 5.099253 | 0.230487 | 0.999532 | | 6.69877 | 0.579265 | 0.368719 | | 6.076827 | 0.365109 | 0.67937 | |
| SLC24A4 | 3.2345 | -0.32895 | 0.999532 | | 3.333332 | -0.84896 | 0.368784 | | 2.560804 | -0.48556 | 0.746666 | |
| SCO2 | 5.000762 | 0.140369 | 0.999532 | | 4.413664 | -0.70492 | 0.368784 | | 3.947381 | -0.82356 | 0.340167 | |
| MME | 50.99926 | 0.362371 | 0.999532 | | 62.00478 | 0.555614 | 0.368784 | | 58.49379 | 0.222986 | 0.791466 | |
| ALKBH1 | 2.410193 | 0.023219 | 0.999532 | | 2.191028 | -1.03052 | 0.368784 | | 1.722461 | -1.02676 | 0.459697 | |
| PPME1 | 3.626812 | 0.413672 | 0.999532 | | 4.539665 | 0.704153 | 0.368828 | | 4.296868 | 0.30904 | 0.799112 | |
| SEC24B | 5.729337 | 0.208748 | 0.999532 | | 7.28644 | 0.540992 | 0.369052 | | 6.726329 | 0.353434 | 0.666906 | |
| DTD2 | 7.410851 | 0.319746 | 0.999532 | | 6.167661 | -0.66787 | 0.370114 | | 6.256374 | -0.95733 | 0.24893 | |
| TMC8 | 3.126805 | -0.30974 | 0.999532 | | 3.002006 | -0.88176 | 0.370114 | | 2.38891 | -0.55766 | 0.705425 | |
| EPB42 | 3.004723 | -0.21843 | 0.999532 | | 2.853694 | -0.91235 | 0.370114 | | 2.316063 | -0.6736 | 0.647218 | |
| ATP5PO | 11.87834 | -0.27684 | 0.999532 | | 16.80605 | 0.409271 | 0.370114 | | 13.31845 | 0.704208 | 0.121156 | |
| SELENOT | 5.491612 | -0.04959 | 0.999532 | | 7.634665 | 0.562432 | 0.370114 | | 6.537547 | 0.624405 | 0.433186 | |
| TCF4 | 5.252862 | -0.56326 | 0.999532 | | 8.612142 | 0.59674 | 0.370114 | | 6.514729 | 1.169587 | 0.068639 | UP |
| ULBP2 | 2.743809 | 0.035823 | 0.999532 | | 2.495205 | -1.00156 | 0.370114 | | 2.158242 | -1.0207 | 0.436389 | |
| RMND5A | 3.518354 | -0.14655 | 0.999532 | | 5.227378 | 0.633935 | 0.370114 | | 4.385253 | 0.800754 | 0.323635 | |
| UQCRC1 | 10.70967 | 0.306567 | 0.999532 | | 12.79835 | 0.497341 | 0.370114 | | 11.55966 | 0.223297 | 0.793131 | |
| AP2M1 | 16.99862 | -0.04782 | 0.999532 | | 22.50354 | 0.410178 | 0.370114 | | 18.59388 | 0.481899 | 0.413122 | |
| PDCL | 3.289584 | 0.174148 | 0.999532 | | 4.614527 | 0.668398 | 0.370114 | | 4.000634 | 0.504451 | 0.632847 | |
| SLC2A4 | 3.219795 | 0.200136 | 0.999532 | | 2.792166 | -0.89485 | 0.370114 | | 2.490989 | -1.066 | 0.323635 | |
| PDHA1 | 2.403107 | 0.489961 | 0.999532 | | 3.235674 | 0.803419 | 0.370847 | | 3.162996 | 0.332649 | 0.803301 | |
| EPB41L5 | 11.76681 | 0.264033 | 0.999532 | | 14.46835 | 0.501811 | 0.370847 | | 12.95347 | 0.267864 | 0.728277 | |
| CDCA3 | 2.496871 | -0.41235 | 0.999532 | | 2.405697 | -0.96898 | 0.370847 | | 1.694843 | -0.53938 | 0.747677 | |
| HLTF | 4.37644 | -0.09646 | 0.999532 | | 4.248663 | -0.71374 | 0.371635 | | 3.207708 | -0.59574 | 0.566812 | |
| DEGS1 | 4.79793 | 0.010588 | 0.999532 | | 6.741272 | 0.596577 | 0.372223 | | 5.615372 | 0.605481 | 0.452319 | |
| CCL8 | 2.633437 | -0.69191 | 0.999532 | | 2.732207 | -0.93675 | 0.372237 | | 1.7306 | -0.22082 | 0.911581 | |
| ALX1 | 2.583518 | -0.16782 | 0.999532 | | 2.544935 | -0.91118 | 0.372237 | | 1.976815 | -0.71726 | 0.619423 | |
| CCDC144 | 5.070829 | 0.133724 | 0.999532 | | 4.147559 | -0.83516 | 0.372247 | | 3.815168 | -0.93609 | 0.300261 | |
| SAMD10 | 6.818733 | -0.05069 | 0.999532 | | 6.431113 | -0.63594 | 0.372247 | | 5.456721 | -0.57096 | 0.552672 | |
| SLC4A4 | 4.422006 | 0.779064 | 0.999532 | | 4.529318 | 0.733517 | 0.372247 | | 5.18754 | -0.05708 | 0.977359 | |
| EGR3 | 5.43396 | -0.81516 | 0.999532 | | 6.11288 | -0.74093 | 0.373313 | | 3.579298 | 0.115563 | 0.946311 | |
| SOCS3 | 4.551572 | -0.30111 | 0.999532 | | 4.499986 | -0.92894 | 0.373313 | | 3.121325 | -0.62732 | 0.572297 | |
| BRD3 | 4.351723 | 0.218211 | 0.999532 | | 5.983121 | 0.593146 | 0.373313 | | 5.117166 | 0.384848 | 0.694961 | |
| MAFA | 3.556854 | -0.31148 | 0.999532 | | 3.571824 | -0.865 | 0.373313 | | 2.68908 | -0.53 | 0.720106 | |
| PLPP3 | 43.40364 | -0.251 | 0.999532 | | 61.48471 | 0.401191 | 0.373408 | | 48.9534 | 0.680099 | 0.105617 | |
| LRRC73 | 3.610807 | -0.00404 | 0.999532 | | 3.17227 | -0.86192 | 0.374817 | | 2.822734 | -0.84878 | 0.496445 | |
| ABRAXA | 4.012434 | 0.527075 | 0.999532 | | 4.658326 | 0.67573 | 0.375864 | | 4.650272 | 0.168092 | 0.895237 | |
| XKR7 | 2.464201 | -0.49881 | 0.999532 | | 2.523966 | -0.97998 | 0.37588 | | 1.783601 | -0.45943 | 0.805825 | |
| DDX53 | 2.407487 | 0.050795 | 0.999532 | | 2.09382 | -1.02022 | 0.37588 | | 1.788876 | -1.05018 | 0.436389 | |
| ARHGAP | 4.992677 | 0.221786 | 0.999532 | | 6.661438 | 0.594007 | 0.37588 | | 5.933758 | 0.395503 | 0.68597 | |
| RNF103 | 3.668866 | 0.806071 | 0.999532 | | 3.828779 | 0.725712 | 0.375996 | | 4.2411 | -0.06689 | 0.970811 | |
| GPR26 | 2.345661 | -0.15648 | 0.999532 | | 2.042891 | -1.07385 | 0.375996 | | 1.664685 | -0.90221 | 0.570037 | |
| RAB7A | 19.87001 | -0.20476 | 0.999532 | | 27.35526 | 0.380736 | 0.375996 | | 21.88732 | 0.605549 | 0.162189 | |
| COQ5 | 1.8177 | 0.56348 | 0.999532 | | 2.339562 | 0.957888 | 0.377486 | | 2.282096 | 0.407342 | 0.79686 | |
| KRT6A | 3.262302 | -0.23993 | 0.999532 | | 3.182869 | -0.93263 | 0.377486 | | 2.479687 | -0.67959 | 0.634624 | |
| RAB18 | 4.758137 | 0.112061 | 0.999532 | | 6.704372 | 0.594884 | 0.37793 | | 5.613111 | 0.50003 | 0.563461 | |
| LAMTOR | 6.579639 | 0.004227 | 0.999532 | | 9.088053 | 0.539102 | 0.378199 | | 7.55431 | 0.560199 | 0.515762 | |
| FAM106A | 5.753692 | 9.57E-05 | 0.99982 | | 5.028635 | -0.70727 | 0.378199 | | 4.626339 | -0.69171 | 0.515762 | |
| RPL6 | 27.44915 | 0.071523 | 0.999532 | | 35.4128 | 0.446495 | 0.378199 | | 30.78052 | 0.396352 | 0.532889 | |
| DENND2 | 3.362497 | -0.27703 | 0.999532 | | 3.458372 | -0.8229 | 0.379037 | | 2.409284 | -0.5164 | 0.706407 | |
| ERVV-1 | 3.13597 | -0.14921 | 0.999532 | | 3.159814 | -0.89317 | 0.379414 | | 2.306406 | -0.71636 | 0.58626 | |
| MYOM1 | 2.343442 | 0.431094 | 0.999532 | | 2.80763 | 0.850907 | 0.379414 | | 2.850806 | 0.429686 | 0.769793 | |
| CRTAP | 9.441967 | 0.188093 | 0.999532 | | 12.22187 | 0.503764 | 0.379416 | | 10.8741 | 0.346185 | 0.672584 | |
| ADAMTS | 4.252431 | 0.195547 | 0.999532 | | 5.664686 | 0.681988 | 0.379416 | | 4.988644 | 0.515173 | 0.623565 | |
| LUM | 5.903623 | 0.132845 | 0.999532 | | 8.321152 | 0.735761 | 0.379416 | | 7.343224 | 0.621205 | 0.566812 | |
| ISCU | 18.31337 | -0.09829 | 0.999532 | | 25.30231 | 0.434636 | 0.379761 | | 20.71843 | 0.552803 | 0.254225 | |
| MED18 | 2.412251 | -0.52125 | 0.999532 | | 2.429452 | -0.92403 | 0.379761 | | 1.743493 | -0.38448 | 0.837256 | |
| GALE | 2.289861 | -0.09603 | 0.999532 | | 3.58819 | 0.792012 | 0.380128 | | 2.93849 | 0.905721 | 0.396754 | |
| C2CD2 | 4.352542 | -0.3673 | 0.999532 | | 4.465505 | -0.68828 | 0.38091 | | 3.224682 | -0.30533 | 0.819366 | |
| GTF2I | 14.11942 | 0.165585 | 0.999532 | | 17.79075 | 0.489318 | 0.38091 | | 16.06965 | 0.336147 | 0.662584 | |
| ZNF723 | 14.6961 | 0.009417 | 0.999532 | | 14.30946 | -0.42791 | 0.382264 | | 12.51124 | -0.41244 | 0.568257 | |
| ZNF519 | 3.842566 | -0.60035 | 0.999532 | | 4.121342 | -0.75084 | 0.383136 | | 2.763059 | -0.12955 | 0.945759 | |
| DYNC2H1 | 2.453651 | -0.21227 | 0.999532 | | 2.192332 | -0.98017 | 0.383785 | | 1.809124 | -0.75316 | 0.646739 | |
| HIPK2 | 12.31037 | 0.328038 | 0.999532 | | 14.37261 | 0.457598 | 0.384095 | | 13.3252 | 0.156902 | 0.844881 | |
| CIRBP | 28.51321 | 0.21133 | 0.999532 | | 33.22418 | 0.359072 | 0.384134 | | 30.78661 | 0.163048 | 0.791466 | |
| UBE2B | 7.507118 | -0.07198 | 0.999532 | | 10.72608 | 0.48722 | 0.384134 | | 8.76166 | 0.571831 | 0.339404 | |
| CSE1L | 3.04036 | 0.15668 | 0.999532 | | 4.320026 | 0.717613 | 0.384134 | | 3.854197 | 0.576066 | 0.574633 | |
| CDC27 | 4.307008 | 0.33123 | 0.999532 | | 5.419484 | 0.610333 | 0.384419 | | 5.121811 | 0.296837 | 0.761699 | |
| TSPAN2 | 18.57428 | 0.112878 | 0.999532 | | 24.2244 | 0.51708 | 0.38524 | | 21.14148 | 0.429891 | 0.565022 | |
| MSI2 | 7.083275 | 0.277166 | 0.999532 | | 9.170995 | 0.612755 | 0.386024 | | 8.041964 | 0.389857 | 0.699086 | |
| POLR2I | 5.423981 | 0.076638 | 0.999532 | | 7.349989 | 0.548655 | 0.386024 | | 6.362791 | 0.490896 | 0.542124 | |
| PSMD8 | 14.19138 | 0.210596 | 0.999532 | | 17.25208 | 0.375551 | 0.386024 | | 15.70931 | 0.187379 | 0.790999 | |
| TSC22D3 | 10.20145 | -1.19303 | 0.299285 | | 12.83195 | -0.74692 | 0.386024 | | 6.670606 | 0.47056 | 0.566812 | |
| ZC3H11A | 10.15896 | 0.199856 | 0.999532 | | 12.30288 | 0.443943 | 0.386024 | | 11.33405 | 0.262097 | 0.720106 | |
| UBTF | 6.924189 | -0.1329 | 0.999532 | | 9.673114 | 0.509218 | 0.386024 | | 7.762105 | 0.657996 | 0.333577 | |
| PTPN14 | 6.297246 | 0.56986 | 0.999532 | | 7.321535 | 0.617841 | 0.386024 | | 7.387227 | 0.071178 | 0.961366 | |
| TNFRSF1 | 3.288161 | 0.365646 | 0.999532 | | 4.253995 | 0.72355 | 0.386024 | | 3.822667 | 0.377938 | 0.735469 | |
| TRIM44 | 4.688728 | 0.415194 | 0.999532 | | 5.503174 | 0.647421 | 0.386024 | | 5.50668 | 0.250307 | 0.823571 | |
| ARHGEF | 2.580646 | 0.416694 | 0.999532 | | 3.485176 | 0.794349 | 0.38682 | | 3.223498 | 0.399496 | 0.75311 | |
| TAS2R43 | 3.829913 | -0.10353 | 0.999532 | | 3.328584 | -0.81032 | 0.38682 | | 3.015794 | -0.69669 | 0.587596 | |
| PYGB | 5.479347 | -0.50383 | 0.999532 | | 5.988425 | -0.63404 | 0.38695 | | 4.003779 | -0.10107 | 0.952656 | |
| PTGES3 | 9.953612 | -0.06996 | 0.999532 | | 13.41005 | 0.449747 | 0.38695 | | 11.13317 | 0.53882 | 0.366244 | |
| DNASE1L | 2.339329 | 0.25097 | 0.999532 | | 3.311914 | 0.785543 | 0.387445 | | 2.986374 | 0.552414 | 0.652925 | |
| DHX36 | 4.759149 | 0.416847 | 0.999532 | | 5.636953 | 0.628466 | 0.38765 | | 5.384868 | 0.23007 | 0.838268 | |
| PSG7 | 3.87026 | -0.55142 | 0.999532 | | 4.200896 | -0.92378 | 0.38765 | | 2.801626 | -0.33278 | 0.855173 | |
| PLA2R1 | 39.29204 | 0.32927 | 0.999532 | | 49.67491 | 0.625359 | 0.38765 | | 46.48119 | 0.318906 | 0.717627 | |
| LILRB4 | 2.834973 | -0.19589 | 0.999532 | | 2.809271 | -0.92213 | 0.38824 | | 2.180043 | -0.7186 | 0.60646 | |
| ZDHHC22 | 3.092063 | -0.3054 | 0.999532 | | 2.993994 | -0.89864 | 0.3888 | | 2.207421 | -0.5722 | 0.703463 | |
| ADGRG7 | 2.431733 | -0.37926 | 0.999532 | | 2.32888 | -0.99368 | 0.388912 | | 1.81823 | -0.59115 | 0.737452 | |
| DLGAP4 | 10.15373 | -0.01318 | 0.999532 | | 13.11522 | 0.427275 | 0.388912 | | 11.27335 | 0.458767 | 0.454116 | |
| C22orf42 | 2.543614 | -0.2297 | 0.999532 | | 2.363238 | -0.96684 | 0.388912 | | 1.894687 | -0.71871 | 0.649136 | |
| EFNB2 | 12.13646 | 0.425589 | 0.999532 | | 14.37915 | 0.566022 | 0.388912 | | 13.90772 | 0.162095 | 0.874653 | |
| TUFM | 6.29528 | 0.562452 | 0.999532 | | 7.246148 | 0.577657 | 0.389092 | | 7.455759 | 0.03126 | 0.985675 | |
| COX14 | 4.735835 | -0.00837 | 0.999532 | | 6.721329 | 0.550658 | 0.389092 | | 5.427507 | 0.578287 | 0.49299 | |
| NDUFS4 | 6.082879 | 0.339191 | 0.999532 | | 7.382946 | 0.5687 | 0.389092 | | 7.150175 | 0.246272 | 0.824012 | |
| TBC1D22 | 9.998262 | -0.17674 | 0.999532 | | 13.73979 | 0.422254 | 0.389318 | | 11.22889 | 0.614003 | 0.266933 | |
| RAC1 | 12.53424 | 0.306911 | 0.999532 | | 15.03933 | 0.507161 | 0.390039 | | 14.18427 | 0.219563 | 0.796937 | |
| CST2 | 3.345363 | -0.36628 | 0.999532 | | 3.550727 | -0.81665 | 0.390039 | | 2.580722 | -0.42899 | 0.765852 | |
| TNNT2 | 14.44432 | 0.555422 | 0.999532 | | 15.60183 | 0.481574 | 0.390531 | | 15.86698 | -0.04863 | 0.965649 | |
| ECI2 | 4.014276 | 0.304449 | 0.999532 | | 4.898125 | 0.664087 | 0.39122 | | 4.580041 | 0.368209 | 0.746876 | |
| UBE2D1 | 2.676668 | 0.408389 | 0.999532 | | 3.197428 | 0.760503 | 0.391269 | | 3.26025 | 0.370532 | 0.776359 | |
| CPHXL | 2.721459 | 0.044409 | 0.999532 | | 2.168065 | -0.9747 | 0.391269 | | 2.049413 | -0.99916 | 0.452179 | |
| XRCC5 | 9.93658 | -0.26165 | 0.999532 | | 14.1467 | 0.447046 | 0.391269 | | 11.31121 | 0.724698 | 0.158141 | |
| RSRC2 | 4.778214 | 0.25839 | 0.999532 | | 6.220638 | 0.593158 | 0.391629 | | 5.656331 | 0.354754 | 0.713778 | |
| CERS2 | 6.658812 | 0.231694 | 0.999532 | | 8.377012 | 0.568912 | 0.391629 | | 7.552394 | 0.360294 | 0.679818 | |
| PRKCG | 3.328427 | -0.13776 | 0.999532 | | 3.251708 | -0.81835 | 0.392764 | | 2.50035 | -0.64923 | 0.630705 | |
| NCR3 | 3.397014 | -0.02922 | 0.999532 | | 3.165796 | -0.89122 | 0.395166 | | 2.7838 | -0.81152 | 0.57215 | |
| CNTN3 | 3.846573 | 0.000433 | 0.999532 | | 5.422184 | 0.623691 | 0.395166 | | 4.882578 | 0.645656 | 0.493644 | |
| RPS6 | 44.17292 | 0.095137 | 0.999532 | | 56.21679 | 0.434932 | 0.396348 | | 49.1197 | 0.36291 | 0.563285 | |
| ZNF160 | 3.274586 | 0.026437 | 0.999532 | | 4.519702 | 0.675947 | 0.396407 | | 3.981214 | 0.675419 | 0.486743 | |
| PHLDB2 | 5.901167 | -0.22024 | 0.999532 | | 9.109407 | 0.549202 | 0.396407 | | 7.298713 | 0.800418 | 0.254225 | |
| PPBP | 4.526496 | -0.04477 | 0.999532 | | 4.336787 | -0.76233 | 0.396407 | | 3.395053 | -0.70224 | 0.496445 | |
| PILRB | 9.489682 | 0.519395 | 0.999532 | | 11.06231 | 0.649744 | 0.396407 | | 11.09698 | 0.146061 | 0.90424 | |
| LMAN2 | 5.283137 | 0.225902 | 0.999532 | | 6.690779 | 0.57922 | 0.396407 | | 6.214851 | 0.372673 | 0.680428 | |
| ITM2A | 4.048557 | 0.529183 | 0.999532 | | 4.454126 | 0.665046 | 0.396407 | | 4.632057 | 0.154576 | 0.903339 | |
| GPX4 | 11.14445 | 0.011028 | 0.999532 | | 14.43 | 0.467746 | 0.396407 | | 12.64232 | 0.463383 | 0.515852 | |
| TRIB2 | 11.74313 | 0.514185 | 0.999532 | | 12.88772 | 0.487891 | 0.396823 | | 12.79613 | 0.004442 | 0.997987 | |
| PMEPA1 | 8.217309 | 0.415715 | 0.999532 | | 9.772961 | 0.518968 | 0.396921 | | 9.396015 | 0.123726 | 0.903054 | |
| TAX1BP3 | 6.249512 | -0.025 | 0.999532 | | 8.79976 | 0.532037 | 0.396921 | | 7.19393 | 0.586734 | 0.512069 | |
| SLC5A12 | 3.652706 | 0.053769 | 0.999532 | | 5.609317 | 0.878858 | 0.396921 | | 4.76183 | 0.867621 | 0.461117 | |
| C9orf50 | 2.653094 | -0.28199 | 0.999532 | | 2.679034 | -0.91405 | 0.396921 | | 1.985545 | -0.60326 | 0.702463 | |
| TMBIM1 | 15.04525 | -0.10117 | 0.999532 | | 19.78183 | 0.382383 | 0.396921 | | 16.3988 | 0.505592 | 0.271669 | |
| EIF4EBP | 3.134796 | -0.26961 | 0.999532 | | 2.843029 | -0.83753 | 0.396921 | | 2.302655 | -0.56043 | 0.699971 | |
| UBD | 4.38881 | 0.450821 | 0.999532 | | 5.019034 | -1.11866 | 0.397519 | | 3.851356 | -0.77893 | 0.539774 | |
| UBE2J1 | 6.525584 | 0.349385 | 0.999532 | | 7.998425 | 0.566173 | 0.397519 | | 7.485544 | 0.238521 | 0.81294 | |
| SLC17A1 | 4.287395 | 0.34132 | 0.999532 | | 5.666734 | 0.749689 | 0.397519 | | 5.184205 | 0.436581 | 0.751138 | |
| AMZ1 | 5.068823 | 0.01254 | 0.999532 | | 4.614242 | -0.71146 | 0.399603 | | 4.054633 | -0.70517 | 0.515762 | |
| GBP3 | 8.679316 | -0.28741 | 0.999532 | | 9.00574 | -0.52572 | 0.400394 | | 7.09095 | -0.21385 | 0.836935 | |
| PTN | 3.604057 | 0.047491 | 0.999532 | | 5.003212 | 0.675559 | 0.400406 | | 4.288764 | 0.64317 | 0.540342 | |
| BRI3 | 14.56002 | 0.048545 | 0.999532 | | 19.20676 | 0.421036 | 0.400406 | | 16.3723 | 0.403094 | 0.541136 | |
| P3H2 | 7.805681 | 0.475138 | 0.999532 | | 9.063832 | 0.601581 | 0.400406 | | 8.744454 | 0.154612 | 0.90016 | |
| NT5E | 2.947721 | -0.5011 | 0.999532 | | 4.753551 | 0.683983 | 0.400406 | | 3.612571 | 1.20603 | 0.154583 | |
| POLR1E | 3.301393 | 0.253605 | 0.999532 | | 2.515193 | -0.88702 | 0.400406 | | 2.510987 | -1.12957 | 0.300261 | |
| OCIAD1 | 9.851826 | 0.174194 | 0.999532 | | 12.00065 | 0.465092 | 0.400558 | | 10.89797 | 0.312708 | 0.660097 | |
| PRKACB | 3.560606 | 0.326705 | 0.999532 | | 4.569378 | 0.668487 | 0.40109 | | 4.397756 | 0.359317 | 0.741003 | |
| NDUFS2 | 4.586654 | -0.18317 | 0.999532 | | 6.834148 | 0.548323 | 0.40109 | | 5.310819 | 0.745111 | 0.323644 | |
| FUT6 | 4.031617 | 0.676996 | 0.999532 | | 4.92248 | 0.816761 | 0.40109 | | 5.031707 | 0.157522 | 0.916585 | |
| TSPAN9 | 16.53116 | -0.19475 | 0.999532 | | 22.51762 | 0.332823 | 0.40109 | | 17.77781 | 0.547649 | 0.200192 | |
| LCN1 | 3.575797 | -0.41364 | 0.999532 | | 3.59003 | -0.85033 | 0.40109 | | 2.480576 | -0.4107 | 0.791466 | |
| MGST3 | 14.15592 | -0.23194 | 0.999532 | | 20.17142 | 0.427472 | 0.40127 | | 15.95525 | 0.684728 | 0.15668 | |
| ACTR2 | 7.953924 | -0.24201 | 0.999532 | | 11.73713 | 0.506312 | 0.40127 | | 9.264328 | 0.766169 | 0.241605 | |
| CCNO | 4.409652 | -0.33621 | 0.999532 | | 4.52262 | -0.69163 | 0.40127 | | 3.38542 | -0.33636 | 0.79686 | |
| DLD | 2.926409 | 0.225484 | 0.999532 | | 3.928936 | 0.701201 | 0.401319 | | 3.442635 | 0.491012 | 0.660097 | |
| RAPGEF1 | 6.460847 | 0.347267 | 0.999532 | | 8.005266 | 0.579157 | 0.401526 | | 7.491075 | 0.253689 | 0.811123 | |
| MTLN | 2.345458 | -0.36951 | 0.999532 | | 2.235698 | -0.9386 | 0.401647 | | 1.733497 | -0.55382 | 0.742129 | |
| BHMT | 5.54502 | 1.011395 | 0.999532 | | 5.41034 | 0.795529 | 0.401732 | | 6.364931 | -0.23231 | 0.907626 | |
| TOM1L1 | 3.722036 | 0.011642 | 0.999532 | | 5.37966 | 0.66282 | 0.401732 | | 4.346138 | 0.670498 | 0.515473 | |
| MARCKS | 8.352474 | 0.140018 | 0.999532 | | 11.10853 | 0.532898 | 0.401732 | | 9.737554 | 0.420441 | 0.626053 | |
| DYNC1LI | 5.466906 | -0.15134 | 0.999532 | | 8.056345 | 0.573454 | 0.401732 | | 6.495239 | 0.73447 | 0.282041 | |
| CLINT1 | 6.658321 | 0.142375 | 0.999532 | | 8.532869 | 0.552752 | 0.402008 | | 7.603309 | 0.428893 | 0.613126 | |
| P4HB | 15.05634 | -0.03787 | 0.999532 | | 20.08749 | 0.425057 | 0.402008 | | 17.01162 | 0.463059 | 0.380222 | |
| PRRC2C | 5.945589 | 0.050836 | 0.999532 | | 7.934023 | 0.517568 | 0.402008 | | 7.102156 | 0.494888 | 0.524206 | |
| RTP2 | 4.434942 | 0.208085 | 0.999532 | | 3.509172 | -0.7861 | 0.40204 | | 3.423471 | -0.98213 | 0.297055 | |
| AZGP1 | 3.97809 | -0.25279 | 0.999532 | | 5.87642 | 0.664457 | 0.402668 | | 4.913864 | 0.940275 | 0.282041 | |
| CCT8 | 5.109644 | 0.299693 | 0.999532 | | 6.162024 | 0.574444 | 0.403205 | | 5.965062 | 0.29127 | 0.757037 | |
| RPL34 | 57.75649 | 0.036514 | 0.999532 | | 71.48875 | 0.294819 | 0.404511 | | 61.75555 | 0.281947 | 0.515762 | |
| SYK | 2.416327 | 0.589432 | 0.999532 | | 2.965897 | 0.812384 | 0.404554 | | 2.892353 | 0.247364 | 0.872493 | |
| MINK1 | 5.037622 | 0.06252 | 0.999532 | | 7.083695 | 0.548217 | 0.405399 | | 6.000077 | 0.505307 | 0.541136 | |
| FCN3 | 11.32446 | 0.27767 | 0.999532 | | 14.73485 | 0.662623 | 0.405406 | | 13.52944 | 0.41214 | 0.677173 | |
| TMEM30A | 29.04073 | -0.11998 | 0.999532 | | 39.24425 | 0.426154 | 0.405406 | | 32.65722 | 0.56637 | 0.24893 | |
| SELE | 4.007091 | 0.066297 | 0.999532 | | 3.452524 | -0.81058 | 0.405406 | | 3.176542 | -0.86223 | 0.443752 | |
| OR10G7 | 2.61405 | -0.181 | 0.999532 | | 2.440478 | -0.93384 | 0.405406 | | 1.931216 | -0.73266 | 0.630705 | |
| NTRK3 | 4.029588 | -0.15446 | 0.999532 | | 3.65065 | -0.7418 | 0.405949 | | 2.988373 | -0.59071 | 0.644433 | |
| MT3 | 3.590733 | -0.1203 | 0.999532 | | 3.245588 | -0.78788 | 0.406886 | | 2.679107 | -0.65157 | 0.621895 | |
| ZDHHC6 | 17.12827 | 0.478711 | 0.999532 | | 19.15291 | 0.506551 | 0.406886 | | 18.81727 | 0.05053 | 0.971732 | |
| RNF146 | 2.779242 | 0.084581 | 0.999532 | | 3.848056 | 0.705375 | 0.406886 | | 3.477866 | 0.640139 | 0.541136 | |
| NTRK2 | 4.291502 | 0.365982 | 0.999532 | | 5.433999 | 0.593766 | 0.406886 | | 4.980216 | 0.238824 | 0.836935 | |
| EDF1 | 15.44299 | -0.04522 | 0.999532 | | 21.10423 | 0.426069 | 0.406886 | | 17.43962 | 0.484395 | 0.421517 | |
| C4B | 5.310529 | 0.639569 | 0.999532 | | 7.212042 | 1.021846 | 0.407045 | | 7.167298 | 0.425505 | 0.824012 | |
| NDUFAB1 | 5.944932 | 0.021106 | 0.999532 | | 7.965594 | 0.529796 | 0.407212 | | 6.861847 | 0.521261 | 0.515762 | |
| YPEL4 | 2.981191 | -0.36763 | 0.999532 | | 2.927643 | -0.87456 | 0.407212 | | 2.148566 | -0.48682 | 0.75311 | |
| PLA1A | 4.689348 | -0.73023 | 0.999532 | | 5.176802 | -0.66534 | 0.40785 | | 3.444923 | 0.079507 | 0.970268 | |
| POLR2K | 5.816194 | -0.44183 | 0.999532 | | 9.095579 | 0.470404 | 0.40785 | | 6.794079 | 0.928944 | 0.11254 | |
| SELENOF | 28.0627 | 0.093526 | 0.999532 | | 37.33836 | 0.535311 | 0.408176 | | 32.41139 | 0.47346 | 0.51741 | |
| HAO2 | 4.971101 | 0.709366 | 0.999532 | | 5.279577 | 0.713084 | 0.408689 | | 5.610766 | 0.019767 | 0.994192 | |
| DDB1 | 6.669666 | 0.196984 | 0.999532 | | 8.71442 | 0.570434 | 0.408689 | | 7.821703 | 0.39421 | 0.644433 | |
| CCDC90B | 3.766773 | 0.126458 | 0.999532 | | 5.183982 | 0.651335 | 0.408689 | | 4.595658 | 0.543818 | 0.598329 | |
| RHOV | 3.089688 | -0.36911 | 0.999532 | | 3.028848 | -0.81533 | 0.408977 | | 2.222479 | -0.43306 | 0.788426 | |
| OR10S1 | 2.366771 | -0.20709 | 0.999532 | | 2.154231 | -0.9668 | 0.408977 | | 1.699863 | -0.74185 | 0.646821 | |
| VN1R2 | 3.33572 | -0.37611 | 0.999532 | | 3.422247 | -0.77754 | 0.409253 | | 2.543214 | -0.38286 | 0.796847 | |
| JAK3 | 5.619295 | -0.59116 | 0.999532 | | 6.250423 | -0.5616 | 0.409253 | | 4.425475 | 0.044711 | 0.979089 | |
| XAGE5 | 3.191357 | -0.15948 | 0.999532 | | 2.960296 | -0.79826 | 0.409253 | | 2.368103 | -0.61956 | 0.645376 | |
| CLIP3 | 9.226939 | 0.123244 | 0.999532 | | 11.70522 | 0.448686 | 0.409298 | | 10.04553 | 0.334087 | 0.669356 | |
| NUP37 | 1.949142 | 0.66965 | 0.999532 | | 2.589736 | 0.895486 | 0.409298 | | 2.579056 | 0.250538 | 0.88476 | |
| MFSD6 | 4.043656 | 0.512709 | 0.999532 | | 5.109748 | 0.706576 | 0.410567 | | 4.787251 | 0.249105 | 0.862823 | |
| CDS2 | 3.688883 | 0.425128 | 0.999532 | | 4.654429 | 0.692139 | 0.410567 | | 4.386109 | 0.287977 | 0.806394 | |
| CLDN1 | 3.366885 | 0.405108 | 0.999532 | | 4.254062 | 0.747408 | 0.411718 | | 4.070496 | 0.363102 | 0.765012 | |
| COX4I1 | 17.79657 | -0.10413 | 0.999532 | | 24.25925 | 0.40579 | 0.412373 | | 19.83511 | 0.537101 | 0.358109 | |
| MYO5B | 4.529385 | -0.28713 | 0.999532 | | 4.634994 | -0.75147 | 0.412691 | | 3.281521 | -0.43988 | 0.720106 | |
| STOML1 | 6.071448 | -0.0577 | 0.999532 | | 5.627666 | -0.68795 | 0.412691 | | 4.691765 | -0.613 | 0.542124 | |
| AGPAT1 | 6.365658 | 0.02342 | 0.999532 | | 8.632295 | 0.539161 | 0.413198 | | 7.368179 | 0.538368 | 0.485816 | |
| ACP5 | 2.590867 | -0.3104 | 0.999532 | | 4.296114 | 0.700736 | 0.413387 | | 3.287378 | 1.02653 | 0.266933 | |
| BLCAP | 4.113798 | 0.028719 | 0.999532 | | 6.160379 | 0.607 | 0.413387 | | 5.170414 | 0.604932 | 0.515762 | |
| OR52J3 | 2.463244 | -0.00615 | 0.999532 | | 2.053963 | -0.98077 | 0.413387 | | 1.883652 | -0.96267 | 0.515762 | |
| FAM205A | 2.476562 | -0.00725 | 0.999532 | | 2.0489 | -0.98423 | 0.413711 | | 1.942385 | -0.96151 | 0.515762 | |
| CLTA | 15.00285 | 0.007448 | 0.999532 | | 19.02411 | 0.380979 | 0.413711 | | 16.4395 | 0.394903 | 0.474973 | |
| FAM107B | 3.549196 | -0.85893 | 0.999532 | | 3.878244 | -0.76263 | 0.413959 | | 2.422893 | 0.13198 | 0.945902 | |
| HMGB3 | 4.67605 | 0.186967 | 0.999532 | | 4.000591 | -0.70847 | 0.414667 | | 3.749479 | -0.87832 | 0.344878 | |
| PARD3 | 4.724999 | 0.075914 | 0.999532 | | 6.695708 | 0.568845 | 0.414667 | | 5.617543 | 0.509365 | 0.590692 | |
| ARHGAP | 15.6712 | 0.298059 | 0.999532 | | 18.35198 | 0.478442 | 0.414667 | | 17.28307 | 0.207549 | 0.792688 | |
| KIF5A | 2.536678 | 0.098459 | 0.999532 | | 2.070948 | -0.95924 | 0.414667 | | 2.007728 | -1.03045 | 0.456688 | |
| PRR5L | 2.401437 | -0.47361 | 0.999532 | | 2.338416 | -0.91688 | 0.414667 | | 1.704484 | -0.43234 | 0.816372 | |
| ARHGEF | 25.99445 | 0.185773 | 0.999532 | | 31.3097 | 0.395594 | 0.414667 | | 28.65044 | 0.235237 | 0.747677 | |
| NELFB | 2.544524 | 0.498675 | 0.999532 | | 3.346409 | 0.768138 | 0.414667 | | 3.238672 | 0.278601 | 0.837256 | |
| IQCN | 4.034843 | 0.074938 | 0.999532 | | 3.469867 | -0.8847 | 0.414667 | | 2.951494 | -0.93114 | 0.399012 | |
| TYSND1 | 4.081775 | 0.180491 | 0.999532 | | 3.437663 | -0.81988 | 0.414667 | | 3.189942 | -0.98223 | 0.323635 | |
| FYN | 7.569685 | 0.207593 | 0.999532 | | 9.730156 | 0.480042 | 0.414667 | | 8.641559 | 0.288204 | 0.725216 | |
| CXXC5 | 8.983269 | 0.230239 | 0.999532 | | 11.02888 | 0.47574 | 0.41518 | | 9.964737 | 0.264412 | 0.750417 | |
| GAS7 | 2.44792 | -0.35952 | 0.999532 | | 2.392346 | -0.89092 | 0.415364 | | 1.80277 | -0.51167 | 0.755301 | |
| TMEM88 | 8.071968 | -0.24527 | 0.999532 | | 8.117772 | -0.66094 | 0.415431 | | 6.287939 | -0.38957 | 0.745908 | |
| C11orf58 | 11.34211 | -0.07635 | 0.999532 | | 15.26401 | 0.416401 | 0.415905 | | 12.55146 | 0.512196 | 0.465298 | |
| MAX | 8.58734 | 0.127126 | 0.999532 | | 10.96827 | 0.468262 | 0.416525 | | 9.726428 | 0.361346 | 0.630705 | |
| PSMD2 | 8.179893 | -0.23282 | 0.999532 | | 11.65217 | 0.45287 | 0.41663 | | 9.206004 | 0.702985 | 0.222058 | |
| MOGAT3 | 3.434897 | -0.04554 | 0.999532 | | 3.182687 | -0.8014 | 0.41663 | | 2.483058 | -0.73053 | 0.542124 | |
| TMEM19 | 3.511256 | 0.358777 | 0.999532 | | 4.414585 | 0.673505 | 0.417373 | | 4.39303 | 0.342372 | 0.774044 | |
| GSTP1 | 18.62246 | 0.097162 | 0.999532 | | 23.79336 | 0.414934 | 0.417373 | | 20.25139 | 0.348057 | 0.603506 | |
| TUBGCP3 | 3.616632 | 0.369131 | 0.999532 | | 4.394151 | 0.641411 | 0.417455 | | 4.190715 | 0.285791 | 0.816589 | |
| FAM25A | 5.590255 | 0.138036 | 0.999532 | | 4.892993 | -0.6714 | 0.417455 | | 4.546992 | -0.78344 | 0.456688 | |
| SNX7 | 2.559334 | 0.9719 | 0.999532 | | 2.788125 | 0.879211 | 0.417455 | | 3.163334 | -0.07479 | 0.972221 | |
| CTSL | 7.672347 | 0.1082 | 0.999532 | | 10.08281 | 0.524348 | 0.417455 | | 8.837721 | 0.435022 | 0.630278 | |
| ARHGAP | 2.405534 | -0.30839 | 0.999532 | | 2.27986 | -0.93602 | 0.417455 | | 1.675248 | -0.60986 | 0.709825 | |
| SNX6 | 5.138897 | 0.111504 | 0.999532 | | 6.713569 | 0.561636 | 0.417455 | | 6.03917 | 0.468745 | 0.598185 | |
| DIO1 | 3.745734 | 0.632711 | 0.999532 | | 4.305377 | 0.722189 | 0.417455 | | 4.377054 | 0.108096 | 0.945456 | |
| CLSTN1 | 5.986785 | 0.01838 | 0.999532 | | 8.161716 | 0.529323 | 0.417455 | | 6.897149 | 0.527728 | 0.49299 | |
| TIMMDC1 | 4.701446 | -0.20726 | 0.999532 | | 6.971568 | 0.57011 | 0.417455 | | 5.743734 | 0.798151 | 0.271669 | |
| TSPAN4 | 7.19555 | 0.3459 | 0.999532 | | 8.429906 | 0.521141 | 0.417455 | | 8.01596 | 0.201269 | 0.823571 | |
| UBQLN1 | 4.713594 | -0.0106 | 0.999532 | | 6.49734 | 0.565556 | 0.417455 | | 5.635163 | 0.591413 | 0.504551 | |
| KPNB1 | 6.138592 | 0.034465 | 0.999532 | | 7.958903 | 0.513344 | 0.417455 | | 6.92076 | 0.495762 | 0.515762 | |
| EMC10 | 23.35376 | 0.057813 | 0.999532 | | 29.37243 | 0.373655 | 0.417455 | | 25.36728 | 0.337237 | 0.515762 | |
| MTA3 | 4.160063 | 0.481854 | 0.999532 | | 4.896655 | 0.640519 | 0.417455 | | 4.851767 | 0.179343 | 0.882936 | |
| ATP6AP1 | 6.443523 | 0.219998 | 0.999532 | | 8.078354 | 0.552047 | 0.417455 | | 7.224471 | 0.362528 | 0.68528 | |
| OPN1LW | 2.439476 | -0.26445 | 0.999532 | | 2.338389 | -0.93344 | 0.417455 | | 1.845853 | -0.65089 | 0.682165 | |
| TAS2R4 | 2.748631 | 0.203436 | 0.999532 | | 2.310721 | -0.90701 | 0.417455 | | 2.047793 | -1.08653 | 0.359638 | |
| COL27A1 | 4.231935 | 0.272972 | 0.999532 | | 5.220177 | 0.675642 | 0.417455 | | 4.791968 | 0.434503 | 0.677254 | |
| DNAJC13 | 4.282497 | 0.288707 | 0.999532 | | 5.734739 | 0.597997 | 0.417622 | | 5.222722 | 0.320913 | 0.741408 | |
| PNRC2 | 22.26924 | -0.00988 | 0.999532 | | 27.47747 | 0.314043 | 0.417622 | | 23.57565 | 0.345884 | 0.434308 | |
| KRCC1 | 3.864724 | 0.200278 | 0.999532 | | 5.105892 | 0.60258 | 0.418545 | | 4.537014 | 0.419619 | 0.671153 | |
| MPP3 | 3.991487 | -0.23066 | 0.999532 | | 3.876165 | -0.80472 | 0.418702 | | 2.943863 | -0.5525 | 0.666566 | |
| AGK | 3.303817 | 0.508283 | 0.999532 | | 4.044239 | 0.693284 | 0.418702 | | 3.979774 | 0.20414 | 0.881837 | |
| HCRT | 3.429208 | -0.05659 | 0.999532 | | 3.261265 | -0.8234 | 0.418702 | | 2.467487 | -0.7384 | 0.55193 | |
| PHAX | 37.64312 | 0.010185 | 0.999532 | | 36.99574 | -0.383 | 0.418702 | | 32.07985 | -0.38246 | 0.507168 | |
| CSDE1 | 17.89332 | -0.14618 | 0.999532 | | 23.98175 | 0.37415 | 0.419574 | | 19.83132 | 0.534955 | 0.266933 | |
| MAFF | 3.651879 | -0.38813 | 0.999532 | | 3.544215 | -0.95145 | 0.420394 | | 2.518599 | -0.52054 | 0.718643 | |
| EMC1 | 3.375874 | 0.249096 | 0.999532 | | 4.190245 | 0.672378 | 0.421639 | | 3.926862 | 0.432894 | 0.706407 | |
| PSMC5 | 10.11629 | -0.04636 | 0.999532 | | 13.28955 | 0.426497 | 0.421639 | | 11.25729 | 0.491344 | 0.407399 | |
| ZNF771 | 4.43289 | 0.227409 | 0.999532 | | 3.495836 | -0.72966 | 0.422694 | | 3.52094 | -0.94252 | 0.311486 | |
| SLC44A1 | 2.047419 | 0.173267 | 0.999532 | | 2.884169 | 0.814101 | 0.423754 | | 2.6111 | 0.65418 | 0.619775 | |
| FAM20C | 12.52572 | 0.204239 | 0.999532 | | 15.49993 | 0.469126 | 0.424266 | | 13.64704 | 0.309005 | 0.694677 | |
| VSIG10L2 | 2.221741 | -0.71824 | 0.999532 | | 2.345724 | -0.9284 | 0.425139 | | 1.625714 | -0.19568 | 0.93672 | |
| PMEL | 4.076588 | 0.285318 | 0.999532 | | 3.266675 | -0.83273 | 0.425258 | | 3.179658 | -1.09679 | 0.266933 | |
| FMNL1 | 2.407779 | -0.30325 | 0.999532 | | 2.352077 | -0.8894 | 0.425258 | | 1.91174 | -0.56199 | 0.726599 | |
| KRT16 | 3.196015 | 0.220502 | 0.999532 | | 2.578785 | -0.9257 | 0.425258 | | 2.514293 | -1.1312 | 0.332351 | |
| TIRAP | 2.445373 | 0.044967 | 0.999532 | | 2.092255 | -1.00043 | 0.425258 | | 1.854766 | -1.02591 | 0.491726 | |
| EEF1G | 34.76078 | 0.013825 | 0.999532 | | 46.17762 | 0.476426 | 0.425258 | | 39.54975 | 0.481887 | 0.504551 | |
| PLEC | 20.13573 | -0.01192 | 0.999532 | | 26.01384 | 0.359699 | 0.425258 | | 22.23554 | 0.394487 | 0.421517 | |
| VTI1A | 10.52954 | 0.68472 | 0.865241 | | 11.0133 | 0.516721 | 0.425258 | | 11.82303 | -0.14753 | 0.896886 | |
| ZNF20 | 3.277185 | -0.07899 | 0.999532 | | 2.990761 | -0.81771 | 0.425258 | | 2.520204 | -0.71969 | 0.566812 | |
| MAMSTR | 3.708121 | -0.16964 | 0.999532 | | 3.680939 | -0.74293 | 0.425258 | | 2.887434 | -0.55146 | 0.66232 | |
| GASK1A | 2.251624 | -0.13088 | 0.999532 | | 2.097857 | -0.96548 | 0.425258 | | 1.640412 | -0.81488 | 0.606619 | |
| SLC40A1 | 6.309133 | -0.13327 | 0.999532 | | 8.595938 | 0.539861 | 0.425258 | | 7.462236 | 0.72069 | 0.282378 | |
| S100A11 | 29.74793 | -0.02895 | 0.999532 | | 41.35672 | 0.540624 | 0.42623 | | 35.13601 | 0.589193 | 0.271669 | |
| PKM | 11.28075 | -0.03388 | 0.999532 | | 15.50024 | 0.441935 | 0.427391 | | 12.58516 | 0.498726 | 0.502623 | |
| SSR3 | 4.245488 | -0.09877 | 0.999532 | | 6.142818 | 0.54699 | 0.427391 | | 4.991187 | 0.660504 | 0.413122 | |
| ANKRD5 | 2.608275 | 0.116568 | 0.999532 | | 2.044113 | -0.97098 | 0.427391 | | 2.048829 | -1.06447 | 0.456688 | |
| GREM1 | 3.160321 | 0.108081 | 0.999532 | | 2.472354 | -0.88901 | 0.427391 | | 2.386877 | -0.98381 | 0.421517 | |
| RPL32 | 51.57235 | 0.020795 | 0.999532 | | 64.60761 | 0.330104 | 0.427391 | | 55.47319 | 0.329538 | 0.540342 | |
| GPR18 | 2.922282 | 0.39573 | 0.999532 | | 2.219753 | -0.91353 | 0.427391 | | 2.246946 | -1.28934 | 0.241605 | |
| MAPK8IP | 2.430602 | -0.2624 | 0.999532 | | 2.378764 | -0.91507 | 0.427391 | | 1.923538 | -0.62368 | 0.707142 | |
| RPL10A | 23.98573 | 0.008245 | 0.999532 | | 31.23277 | 0.426284 | 0.427391 | | 26.82061 | 0.436302 | 0.455768 | |
| SCYL2 | 5.950815 | 0.428405 | 0.999532 | | 6.984882 | 0.546962 | 0.427391 | | 6.922449 | 0.137877 | 0.899732 | |
| H2AB1 | 2.702994 | -0.17905 | 0.999532 | | 2.384981 | -0.88611 | 0.427432 | | 1.970159 | -0.69559 | 0.655316 | |
| MGAT5 | 11.72582 | 0.365829 | 0.999532 | | 13.93555 | 0.468442 | 0.427432 | | 12.89064 | 0.113155 | 0.899852 | |
| TAF3 | 2.667631 | -0.22841 | 0.999532 | | 2.492895 | -0.8985 | 0.427849 | | 1.912184 | -0.65341 | 0.669356 | |
| MAPK9 | 3.044298 | -0.32443 | 0.999532 | | 5.099041 | 0.633033 | 0.427955 | | 3.918558 | 0.984954 | 0.266797 | |
| CCDC6 | 5.291507 | 0.106648 | 0.999532 | | 6.94288 | 0.5466 | 0.428093 | | 6.013196 | 0.454995 | 0.622543 | |
| SLC9A3R | 4.607325 | 0.125781 | 0.999532 | | 6.175463 | 0.605436 | 0.428393 | | 5.440215 | 0.568811 | 0.521228 | |
| CCDC85A | 3.613394 | 0.053435 | 0.999532 | | 2.973023 | -0.80628 | 0.428866 | | 2.768205 | -0.84864 | 0.505689 | |
| TRAPPC2 | 2.050436 | 0.843987 | 0.999532 | | 2.245599 | 0.90705 | 0.428866 | | 2.545249 | 0.081644 | 0.972135 | |
| KAZN | 5.683947 | 0.170037 | 0.999532 | | 7.164124 | 0.525736 | 0.429046 | | 6.465219 | 0.373744 | 0.666566 | |
| PDE5A | 1.960834 | 0.290906 | 0.999532 | | 2.672753 | 0.800477 | 0.429311 | | 2.44819 | 0.527497 | 0.686214 | |
| SYT12 | 2.449294 | -0.3075 | 0.999532 | | 2.434389 | -0.89696 | 0.429311 | | 1.848613 | -0.56703 | 0.730921 | |
| WFDC10B | 2.750157 | -0.24441 | 0.999532 | | 2.55543 | -0.86037 | 0.429615 | | 2.11096 | -0.60053 | 0.687245 | |
| LYSMD4 | 3.215042 | -0.46477 | 0.999532 | | 3.292422 | -0.74148 | 0.429615 | | 2.379758 | -0.25959 | 0.879947 | |
| ERP44 | 9.988927 | -0.15602 | 0.999532 | | 14.18913 | 0.422892 | 0.429615 | | 11.4905 | 0.596061 | 0.274496 | |
| RHOXF2 | 2.546237 | -0.27603 | 0.999532 | | 2.405014 | -0.90482 | 0.429615 | | 1.896621 | -0.61134 | 0.708052 | |
| ZNF837 | 3.28737 | 0.183029 | 0.999532 | | 2.786018 | -0.85265 | 0.429615 | | 2.577814 | -1.01733 | 0.359441 | |
| PDIA6 | 11.53699 | 0.224775 | 0.999532 | | 13.9375 | 0.448868 | 0.429615 | | 12.80229 | 0.245538 | 0.740093 | |
| MYO1B | 8.140094 | -0.3736 | 0.999532 | | 11.56851 | 0.478259 | 0.429615 | | 9.144908 | 0.846192 | 0.165207 | |
| ANAPC5 | 6.536552 | 0.148894 | 0.999532 | | 8.007901 | 0.525601 | 0.429615 | | 7.366912 | 0.375595 | 0.675955 | |
| MTMR9 | 3.267813 | -0.34042 | 0.999532 | | 3.078075 | -0.75655 | 0.429615 | | 2.354651 | -0.40547 | 0.784955 | |
| RDX | 9.219534 | 0.164856 | 0.999532 | | 11.20401 | 0.421556 | 0.429615 | | 10.16337 | 0.274477 | 0.718633 | |
| GGT1 | 6.21657 | -0.337 | 0.999532 | | 6.475488 | -0.57235 | 0.429615 | | 4.716637 | -0.2184 | 0.861119 | |
| CD34 | 11.39971 | -0.00699 | 0.999532 | | 15.15244 | 0.466285 | 0.429615 | | 13.17016 | 0.533492 | 0.287765 | |
| IFNGR2 | 5.62604 | 0.062811 | 0.999532 | | 7.516414 | 0.602834 | 0.429691 | | 6.605262 | 0.544372 | 0.535682 | |
| MAGEA8 | 3.178876 | 0.310366 | 0.999532 | | 2.344883 | -0.90208 | 0.429691 | | 2.458525 | -1.1909 | 0.266933 | |
| NTNG1 | 10.50442 | 0.497715 | 0.999532 | | 11.60138 | 0.468799 | 0.429691 | | 11.63553 | -0.00728 | 0.996891 | |
| CASKIN1 | 2.476891 | -0.17769 | 0.999532 | | 2.258478 | -0.94137 | 0.429691 | | 1.85069 | -0.7463 | 0.645376 | |
| CXorf66 | 3.603831 | -0.28004 | 0.999532 | | 3.47547 | -0.79355 | 0.429691 | | 2.715218 | -0.49623 | 0.736302 | |
| CAPN6 | 2.858517 | -0.05266 | 0.999532 | | 2.64381 | -0.84685 | 0.429691 | | 2.166813 | -0.7687 | 0.566812 | |
| CDAN1 | 3.693978 | 0.53746 | 0.999532 | | 4.263861 | 0.697028 | 0.429691 | | 4.328783 | 0.1857 | 0.892869 | |
| H1-2 | 13.33107 | -0.14575 | 0.999532 | | 18.29642 | 0.369636 | 0.429691 | | 14.5969 | 0.527153 | 0.297055 | |
| IL18RAP | 2.431791 | -0.37577 | 0.999532 | | 2.291679 | -0.87271 | 0.43149 | | 1.777641 | -0.48503 | 0.782253 | |
| YAP1 | 8.603231 | 0.108635 | 0.999532 | | 11.04384 | 0.419877 | 0.431633 | | 9.499536 | 0.329276 | 0.647581 | |
| CCNB2 | 3.025543 | 0.029452 | 0.999532 | | 2.685957 | -0.857 | 0.431633 | | 2.137859 | -0.86484 | 0.509531 | |
| ZMYM2 | 4.141385 | 0.171264 | 0.999532 | | 5.340347 | 0.584927 | 0.432366 | | 4.790056 | 0.435754 | 0.657467 | |
| TMEM62 | 1.633661 | 0.418031 | 0.999532 | | 2.543231 | 0.917903 | 0.432523 | | 2.310597 | 0.530192 | 0.728015 | |
| HMHB1 | 2.423307 | -0.28395 | 0.999532 | | 2.376582 | -0.90911 | 0.432523 | | 1.806416 | -0.6027 | 0.713778 | |
| PPM1N | 2.886731 | -1.10399 | 0.999532 | | 3.43685 | -0.83236 | 0.433066 | | 1.823159 | 0.288933 | 0.881837 | |
| HNRNPA | 7.157145 | -0.17628 | 0.999532 | | 10.21516 | 0.485112 | 0.433589 | | 8.333371 | 0.677437 | 0.306314 | |
| ANKDD1A | 3.787965 | -0.24543 | 0.999532 | | 3.621023 | -0.75242 | 0.433635 | | 2.848896 | -0.48407 | 0.709825 | |
| PNN | 9.675368 | 0.21986 | 0.999532 | | 11.62234 | 0.457672 | 0.433967 | | 10.7311 | 0.261562 | 0.735469 | |
| TLE2 | 5.166737 | -0.73178 | 0.999532 | | 6.075382 | -0.57818 | 0.433967 | | 4.045867 | 0.178583 | 0.895421 | |
| DIAPH1 | 2.530081 | -0.22778 | 0.999532 | | 3.973936 | 0.707747 | 0.433967 | | 3.218537 | 0.951648 | 0.320564 | |
| FAM234A | 5.245144 | 0.32174 | 0.999532 | | 6.440203 | 0.569828 | 0.433967 | | 5.811077 | 0.26648 | 0.800239 | |
| CRIM1 | 37.23687 | 0.19126 | 0.999532 | | 45.91385 | 0.436491 | 0.433967 | | 41.29875 | 0.266747 | 0.709825 | |
| MAP6 | 6.665993 | 0.292634 | 0.999532 | | 8.338419 | 0.664528 | 0.433967 | | 7.946314 | 0.398225 | 0.689943 | |
| KRT37 | 2.168802 | -0.591 | 0.999532 | | 2.265913 | -0.93425 | 0.433967 | | 1.610268 | -0.32226 | 0.881837 | |
| MORF4L2 | 6.198642 | 0.056384 | 0.999532 | | 8.359882 | 0.518404 | 0.434332 | | 7.201935 | 0.482461 | 0.536234 | |
| ZNF782 | 3.171127 | 0.235904 | 0.999532 | | 2.632828 | -0.87319 | 0.434332 | | 2.327687 | -1.09147 | 0.333192 | |
| ECM1 | 7.248461 | -0.38817 | 0.999532 | | 11.9204 | 0.653891 | 0.434332 | | 9.492698 | 1.055967 | 0.083797 | UP |
| DDX60 | 3.85545 | 0.050351 | 0.999532 | | 5.209891 | 0.628958 | 0.434332 | | 4.371124 | 0.592175 | 0.560424 | |
| NXF2 | 4.613675 | 0.027963 | 0.999532 | | 4.187032 | -0.70447 | 0.434332 | | 3.643914 | -0.71523 | 0.512666 | |
| SATB1 | 3.583285 | 0.271625 | 0.999532 | | 4.75284 | 0.624745 | 0.434332 | | 4.273521 | 0.375874 | 0.734836 | |
| PYGO2 | 2.809589 | 0.147707 | 0.999532 | | 3.739357 | 0.686231 | 0.434332 | | 3.43116 | 0.561533 | 0.625947 | |
| C4orf46 | 29.57036 | 0.205972 | 0.999532 | | 26.431 | -0.5148 | 0.434332 | | 24.85761 | -0.69847 | 0.310637 | |
| EID1 | 16.28591 | 0.219359 | 0.999532 | | 19.53186 | 0.429742 | 0.434332 | | 17.98853 | 0.222555 | 0.753597 | |
| PPIA | 42.1576 | -0.1319 | 0.999532 | | 55.82703 | 0.31788 | 0.434461 | | 45.73605 | 0.472932 | 0.222058 | |
| GPX3 | 101.6603 | 0.116889 | 0.999532 | | 142.7468 | 0.681819 | 0.434821 | | 125.2178 | 0.580454 | 0.709825 | |
| LGALS7B | 4.34516 | 0.017851 | 0.999532 | | 4.203096 | -0.71681 | 0.436205 | | 3.393874 | -0.71709 | 0.491726 | |
| GPR21 | 3.898426 | -0.3341 | 0.999532 | | 3.938712 | -0.69923 | 0.437265 | | 2.750662 | -0.34566 | 0.800239 | |
| GLIPR1L1 | 2.573342 | -0.38912 | 0.999532 | | 2.432467 | -0.89278 | 0.43734 | | 1.924907 | -0.48491 | 0.779708 | |
| F8A2 | 4.6373 | -0.56664 | 0.999532 | | 5.16201 | -0.63639 | 0.437414 | | 3.518063 | -0.05268 | 0.978229 | |
| SLC9A3R | 56.16594 | -0.15078 | 0.999532 | | 74.42335 | 0.315355 | 0.437649 | | 60.82953 | 0.493389 | 0.107433 | |
| HNRNPH | 13.81809 | 0.29513 | 0.999532 | | 16.36168 | 0.412879 | 0.438029 | | 15.31058 | 0.142054 | 0.881689 | |
| DYNLT3 | 10.24817 | 0.070945 | 0.999532 | | 12.96152 | 0.413924 | 0.438072 | | 11.59891 | 0.35731 | 0.636291 | |
| PLS3 | 5.428367 | -0.34709 | 0.999532 | | 8.499465 | 0.537369 | 0.438273 | | 6.56732 | 0.89794 | 0.15668 | |
| ADIPOR1 | 3.83851 | 0.328872 | 0.999532 | | 4.866885 | 0.602942 | 0.438389 | | 4.541667 | 0.290678 | 0.799944 | |
| MAP4K1 | 3.135173 | -0.09699 | 0.999532 | | 2.641185 | -0.83593 | 0.438389 | | 2.37382 | -0.7176 | 0.598329 | |
| DCAF5 | 4.20322 | 0.431564 | 0.999532 | | 4.949804 | 0.614116 | 0.43843 | | 5.048494 | 0.242875 | 0.848617 | |
| COPB1 | 4.351 | 0.107237 | 0.999532 | | 6.02753 | 0.566454 | 0.4386 | | 5.241236 | 0.47853 | 0.637292 | |
| ENPEP | 26.38238 | 0.592474 | 0.999532 | | 28.35402 | 0.488742 | 0.438773 | | 28.9018 | -0.0838 | 0.945902 | |
| BOLA2B | 5.886659 | -0.10183 | 0.999532 | | 5.796932 | -0.56725 | 0.438773 | | 4.832648 | -0.44458 | 0.654161 | |
| RPL37 | 34.06906 | 0.015125 | 0.999532 | | 43.00408 | 0.371403 | 0.438773 | | 37.11791 | 0.366846 | 0.512069 | |
| NPR3 | 11.16011 | 0.214454 | 0.999532 | | 13.93654 | 0.456933 | 0.438773 | | 12.73655 | 0.255572 | 0.722703 | |
| PLPP6 | 2.656642 | -0.62803 | 0.999532 | | 2.893366 | -0.79795 | 0.438773 | | 2.01862 | -0.15133 | 0.945129 | |
| SERPINB | 2.417951 | -0.45059 | 0.999532 | | 2.488151 | -0.83578 | 0.438773 | | 1.714548 | -0.36453 | 0.838733 | |
| CYTH3 | 4.631068 | 0.122512 | 0.999532 | | 6.257439 | 0.576172 | 0.438773 | | 5.699017 | 0.476497 | 0.598589 | |
| TOX2 | 2.590748 | -0.18714 | 0.999532 | | 2.539609 | -0.87011 | 0.438773 | | 2.028349 | -0.66111 | 0.660097 | |
| UBALD1 | 16.27324 | 0.241171 | 0.999532 | | 14.1777 | -0.50053 | 0.439391 | | 13.44072 | -0.70267 | 0.139714 | |
| CAPS | 3.531967 | -0.31533 | 0.999532 | | 3.559426 | -0.70192 | 0.439449 | | 2.674325 | -0.36766 | 0.792539 | |
| FCGR1A | 4.304492 | -0.01629 | 0.999532 | | 4.105083 | -0.70033 | 0.439449 | | 3.389362 | -0.65734 | 0.552672 | |
| RPL30 | 59.58503 | -0.09874 | 0.999532 | | 79.2207 | 0.375543 | 0.43985 | | 65.47345 | 0.494825 | 0.348004 | |
| VPS52 | 4.021709 | 0.480517 | 0.999532 | | 4.839901 | 0.636144 | 0.43985 | | 4.807244 | 0.171567 | 0.892869 | |
| SPATA31 | 2.673685 | -0.43656 | 0.999532 | | 2.701228 | -0.88637 | 0.43985 | | 1.938453 | -0.43382 | 0.803594 | |
| ZNF341 | 3.749277 | -0.0119 | 0.999532 | | 3.456339 | -0.79403 | 0.440196 | | 2.843182 | -0.76321 | 0.515762 | |
| TP53RK | 3.416884 | -0.2434 | 0.999532 | | 3.230845 | -0.85115 | 0.440544 | | 2.524546 | -0.58323 | 0.682165 | |
| ALG10B | 6.431914 | -0.45148 | 0.999532 | | 7.079274 | -0.52665 | 0.441551 | | 5.048976 | -0.0558 | 0.972344 | |
| MRPS28 | 2.433479 | 0.323868 | 0.999532 | | 3.431607 | 0.738051 | 0.44181 | | 3.084375 | 0.437418 | 0.734538 | |
| PABPN1 | 14.68793 | 0.496091 | 0.999532 | | 15.71816 | 0.406106 | 0.44181 | | 15.55158 | -0.07816 | 0.941197 | |
| KIF5B | 6.657572 | -0.43016 | 0.999532 | | 9.938196 | 0.471422 | 0.44181 | | 7.650476 | 0.919395 | 0.107433 | |
| ZNF300 | 2.922325 | 0.228573 | 0.999532 | | 2.365587 | -0.86909 | 0.44181 | | 2.229326 | -1.08088 | 0.339543 | |
| NUP214 | 6.08678 | -0.07289 | 0.999532 | | 6.033689 | -0.56065 | 0.44181 | | 4.706981 | -0.46204 | 0.637292 | |
| ILK | 8.637071 | -0.10127 | 0.999532 | | 11.72025 | 0.451332 | 0.44181 | | 9.732614 | 0.561684 | 0.458056 | |
| C5orf15 | 9.54059 | -0.00272 | 0.999532 | | 12.51429 | 0.502868 | 0.441923 | | 10.92769 | 0.520816 | 0.506666 | |
| PNRC1 | 16.5286 | -0.04369 | 0.999532 | | 21.02573 | 0.341493 | 0.442064 | | 17.4949 | 0.403356 | 0.454841 | |
| ALDOA | 34.61388 | -0.09868 | 0.999532 | | 45.99301 | 0.36592 | 0.442064 | | 37.86283 | 0.488325 | 0.269743 | |
| FBP1 | 4.139699 | 0.566503 | 0.999532 | | 5.084443 | 0.774876 | 0.44266 | | 4.937115 | 0.233307 | 0.88885 | |
| PRAP1 | 5.678473 | 0.304806 | 0.999532 | | 6.783836 | 0.523324 | 0.442948 | | 6.406805 | 0.23716 | 0.820072 | |
| CTXN1 | 3.264526 | -0.11817 | 0.999532 | | 3.11836 | -0.77379 | 0.445053 | | 2.56714 | -0.63366 | 0.634624 | |
| ZFAND5 | 13.37372 | -0.93497 | 0.083111 | DOWN | 17.08765 | -0.43886 | 0.445054 | | 10.20882 | 0.517011 | 0.37317 | |
| AGRN | 18.36709 | 0.381748 | 0.999532 | | 21.02026 | 0.411837 | 0.445054 | | 19.87104 | 0.046313 | 0.961366 | |
| SOX4 | 7.964547 | 0.215395 | 0.999532 | | 10.31867 | 0.59377 | 0.445054 | | 9.354839 | 0.400619 | 0.680577 | |
| RAB15 | 2.723627 | -0.20067 | 0.999532 | | 2.443595 | -0.89065 | 0.445054 | | 1.915062 | -0.67293 | 0.662472 | |
| PLA2G1B | 2.241021 | -0.20678 | 0.999532 | | 2.023217 | -0.96689 | 0.445054 | | 1.615242 | -0.7448 | 0.660958 | |
| IDO1 | 4.671165 | -0.60508 | 0.999532 | | 5.43192 | -0.63024 | 0.445054 | | 3.832535 | -0.00487 | 0.99858 | |
| GALNT18 | 6.538783 | 0.265794 | 0.999532 | | 8.054154 | 0.553168 | 0.445054 | | 7.428968 | 0.305166 | 0.765574 | |
| C8orf37 | 2.280667 | -0.30518 | 0.999532 | | 2.174724 | -0.92148 | 0.445054 | | 1.630253 | -0.59754 | 0.718643 | |
| PRND | 2.644955 | -0.10227 | 0.999532 | | 2.5491 | -0.88409 | 0.445054 | | 2.064397 | -0.75278 | 0.621656 | |
| MUC3A | 3.278642 | -0.60728 | 0.999532 | | 3.666065 | -0.75889 | 0.445054 | | 2.684345 | -0.12997 | 0.952785 | |
| PDE10A | 2.83648 | -0.71251 | 0.999532 | | 2.948817 | -0.82285 | 0.445704 | | 1.927046 | -0.09588 | 0.971732 | |
| SEMA3B | 6.465719 | 0.329521 | 0.999532 | | 7.858558 | 0.593328 | 0.446245 | | 7.273004 | 0.299326 | 0.760955 | |
| BDH2 | 11.07779 | 0.066384 | 0.999532 | | 14.87244 | 0.539159 | 0.447662 | | 12.61473 | 0.51633 | 0.515762 | |
| OR4F21 | 5.052232 | -0.0368 | 0.999532 | | 4.818454 | -0.6794 | 0.447662 | | 3.750809 | -0.62814 | 0.524707 | |
| SNRPC | 21.03409 | 0.078937 | 0.999532 | | 20.30527 | -0.36704 | 0.447662 | | 18.13557 | -0.426 | 0.493137 | |
| MLLT10 | 6.875414 | 0.162624 | 0.999532 | | 8.996062 | 0.498861 | 0.448217 | | 8.017685 | 0.358624 | 0.673405 | |
| THNSL2 | 4.25412 | 0.41903 | 0.999532 | | 3.291239 | -0.82552 | 0.448217 | | 3.154406 | -1.20713 | 0.162189 | |
| ACTA2 | 10.21851 | -0.07756 | 0.999532 | | 16.70933 | 0.84483 | 0.448217 | | 14.01907 | 0.946908 | 0.436389 | |
| COIL | 6.667663 | -0.28227 | 0.999532 | | 7.090878 | -0.52522 | 0.448217 | | 5.367795 | -0.21119 | 0.861815 | |
| TUBGCP2 | 3.217503 | 0.620246 | 0.999532 | | 3.581901 | 0.709334 | 0.448217 | | 3.740183 | 0.110093 | 0.946512 | |
| EGLN2 | 3.414259 | -0.10216 | 0.999532 | | 5.167778 | 0.611834 | 0.448217 | | 4.270717 | 0.735636 | 0.438989 | |
| RUBCN | 2.411638 | -0.33533 | 0.999532 | | 2.305769 | -0.89749 | 0.448217 | | 1.788953 | -0.54547 | 0.750371 | |
| ARMH4 | 17.9551 | 0.468907 | 0.999532 | | 20.52033 | 0.492231 | 0.448217 | | 20.02484 | 0.044804 | 0.972305 | |
| PRR20A | 5.993149 | -0.4107 | 0.999532 | | 6.333373 | -0.69082 | 0.448217 | | 4.495723 | -0.25499 | 0.837256 | |
| STX3 | 4.001525 | 0.218965 | 0.999532 | | 5.216274 | 0.674351 | 0.448217 | | 4.651427 | 0.479067 | 0.668481 | |
| MELTF | 2.535634 | 0.001787 | 0.999532 | | 2.261542 | -0.90166 | 0.448217 | | 1.990669 | -0.88005 | 0.536289 | |
| LHFPL6 | 6.478064 | 0.218133 | 0.999532 | | 8.223492 | 0.530018 | 0.448217 | | 7.435835 | 0.329642 | 0.70375 | |
| CGB2 | 9.972758 | -0.09129 | 0.999532 | | 9.825884 | -0.46094 | 0.448217 | | 8.321922 | -0.35906 | 0.666492 | |
| FAM9C | 2.437177 | -0.17621 | 0.999532 | | 2.258466 | -0.93747 | 0.448217 | | 1.756593 | -0.74338 | 0.647218 | |
| TNS2 | 20.88615 | 0.00652 | 0.999532 | | 26.52149 | 0.338405 | 0.448217 | | 22.36038 | 0.351909 | 0.499025 | |
| LAMC1 | 7.956403 | 0.080565 | 0.999532 | | 10.3627 | 0.525483 | 0.448217 | | 9.215072 | 0.452771 | 0.60646 | |
| GCDH | 2.938076 | -0.45645 | 0.999532 | | 3.018383 | -0.78962 | 0.448217 | | 2.158762 | -0.31451 | 0.858705 | |
| TIA1 | 4.748652 | 0.27079 | 0.999532 | | 6.302435 | 0.619068 | 0.448217 | | 5.880627 | 0.354775 | 0.723509 | |
| SPARC | 134.4676 | 0.032539 | 0.999532 | | 176.3099 | 0.450972 | 0.448217 | | 152.0164 | 0.441069 | 0.406514 | |
| SLC36A1 | 12.66903 | -0.01915 | 0.999532 | | 12.42091 | -0.47925 | 0.448217 | | 10.57883 | -0.44087 | 0.5451 | |
| MSN | 14.00831 | -0.41906 | 0.999532 | | 21.20922 | 0.430907 | 0.448217 | | 16.17978 | 0.87098 | 0.046845 | UP |
| MED22 | 5.180607 | 0.435812 | 0.999532 | | 6.131037 | 0.591873 | 0.448217 | | 5.987534 | 0.177192 | 0.878619 | |
| C11orf1 | 2.507745 | 0.683008 | 0.999532 | | 2.792975 | 0.788197 | 0.448217 | | 2.99351 | 0.122507 | 0.945902 | |
| TMCO4 | 3.936066 | -0.1398 | 0.999532 | | 3.955278 | -0.66212 | 0.448217 | | 3.045622 | -0.49916 | 0.667066 | |
| SEPTIN2 | 14.32659 | -0.09828 | 0.999532 | | 19.80987 | 0.408451 | 0.448217 | | 16.07757 | 0.545637 | 0.457856 | |
| SNAP25 | 3.531298 | 0.025814 | 0.999532 | | 3.131065 | -0.77295 | 0.448474 | | 2.759565 | -0.78079 | 0.515762 | |
| CRISPLD | 2.244798 | -0.23334 | 0.999532 | | 2.02415 | -0.93171 | 0.44932 | | 1.730635 | -0.68346 | 0.692333 | |
| PCSK9 | 2.835876 | 0.151319 | 0.999532 | | 2.289959 | -0.90699 | 0.44932 | | 2.114352 | -1.03918 | 0.418768 | |
| UACA | 7.983076 | 0.345277 | 0.999532 | | 9.823632 | 0.484784 | 0.44932 | | 9.020832 | 0.176673 | 0.881837 | |
| MYO1C | 22.17083 | -0.05523 | 0.999532 | | 28.39793 | 0.302378 | 0.449388 | | 23.65025 | 0.375639 | 0.446959 | |
| DCDC2 | 3.927548 | 0.679623 | 0.999532 | | 4.589882 | 0.707133 | 0.449873 | | 4.432795 | 0.080969 | 0.96129 | |
| PEX19 | 3.0694 | -0.47395 | 0.999532 | | 3.282898 | -0.74724 | 0.449873 | | 2.306953 | -0.253 | 0.88476 | |
| UBA3 | 6.140128 | 0.558079 | 0.999532 | | 6.814502 | 0.536806 | 0.449873 | | 6.889461 | 1.61E-05 | 0.999949 | |
| SERPINF | 3.948736 | 0.061152 | 0.999532 | | 3.324067 | -0.74242 | 0.450375 | | 3.081051 | -0.76431 | 0.531935 | |
| COA1 | 4.384841 | 0.423427 | 0.999532 | | 5.076369 | 0.574227 | 0.450375 | | 4.927444 | 0.165619 | 0.892869 | |
| NSD1 | 3.024081 | -0.20717 | 0.999532 | | 4.610773 | 0.628506 | 0.450375 | | 3.653705 | 0.855745 | 0.359441 | |
| GPR75 | 2.555824 | -0.31469 | 0.999532 | | 2.656555 | -0.82904 | 0.450792 | | 1.958476 | -0.4878 | 0.759211 | |
| RPL36A | 16.36763 | -0.00207 | 0.999532 | | 20.59891 | 0.398559 | 0.4516 | | 17.93057 | 0.407185 | 0.515762 | |
| IFI27 | 14.74084 | -0.28938 | 0.999532 | | 21.14925 | 0.392101 | 0.4516 | | 16.75422 | 0.705971 | 0.119877 | |
| CLTRN | 4.398767 | 0.397264 | 0.999532 | | 6.421445 | 0.994569 | 0.452307 | | 5.962135 | 0.646249 | 0.702463 | |
| PHF11 | 3.519281 | 0.326954 | 0.999532 | | 4.690511 | 0.624784 | 0.452424 | | 4.359681 | 0.315694 | 0.783332 | |
| SOX18 | 11.17612 | 0.213723 | 0.999532 | | 13.76721 | 0.449857 | 0.452424 | | 12.86453 | 0.244328 | 0.755206 | |
| MTMR3 | 4.775776 | 0.067755 | 0.999532 | | 6.642275 | 0.543155 | 0.45267 | | 5.451131 | 0.483698 | 0.572297 | |
| FKBP5 | 5.652811 | -1.10394 | 0.651863 | | 6.936115 | -0.72689 | 0.452908 | | 3.756609 | 0.41829 | 0.737109 | |
| KRTAP5- | 7.367252 | 0.054339 | 0.999532 | | 6.805666 | -0.56537 | 0.453661 | | 6.092729 | -0.60826 | 0.522078 | |
| EZR | 8.813383 | 0.172551 | 0.999532 | | 11.46357 | 0.469695 | 0.453928 | | 9.877324 | 0.339733 | 0.666566 | |
| GMPR2 | 7.426255 | 0.274333 | 0.999532 | | 9.134076 | 0.485833 | 0.453928 | | 8.4511 | 0.228617 | 0.800239 | |
| TNC | 19.04845 | -0.00397 | 0.999532 | | 28.06081 | 0.678783 | 0.453928 | | 23.96172 | 0.702038 | 0.438594 | |
| NPHS1 | 20.63773 | 0.848586 | 0.258167 | | 20.23587 | 0.504781 | 0.454182 | | 22.57846 | -0.31682 | 0.731348 | |
| R3HDM2 | 3.278196 | 0.036245 | 0.999532 | | 4.796253 | 0.613232 | 0.454182 | | 4.060106 | 0.596673 | 0.543211 | |
| HEPACA | 2.941223 | -0.10512 | 0.999532 | | 2.663908 | -0.83869 | 0.454629 | | 2.271396 | -0.71566 | 0.632113 | |
| NDUFA4 | 12.88806 | -0.0296 | 0.999532 | | 16.96371 | 0.412154 | 0.454629 | | 14.29343 | 0.458665 | 0.515762 | |

| | | | | | | | | | | | |
|---|---|---|---|---|---|---|---|---|---|---|---|
| GAL3ST2 | 3.639135 | 0.159035 | 0.999532 | | 2.982888 | -0.7738 | 0.454743 | | 2.849998 | -0.91372 | 0.403717 |
| CYP2A7 | 6.592477 | 0.027991 | 0.999532 | | 6.022284 | -0.59104 | 0.454743 | | 5.543049 | -0.6 | 0.558035 |
| CBX5 | 9.047592 | -0.17903 | 0.999532 | | 12.69806 | 0.441142 | 0.454743 | | 9.874405 | 0.618962 | 0.346962 |
| CNNM2 | 2.666263 | 0.638289 | 0.999532 | | 3.074284 | 0.750434 | 0.454831 | | 3.196581 | 0.130831 | 0.939549 |
| ADRA2B | 3.885132 | 0.02963 | 0.999532 | | 3.693218 | -0.72026 | 0.455225 | | 2.938737 | -0.73053 | 0.515762 |
| ASAP2 | 5.545544 | 0.054012 | 0.999532 | | 7.263652 | 0.536786 | 0.455334 | | 6.331054 | 0.492809 | 0.602678 |
| NAGPA | 2.525829 | -0.21343 | 0.999532 | | 2.496447 | -0.8482 | 0.455386 | | 1.777744 | -0.61342 | 0.68597 |
| YME1L1 | 5.278466 | -0.36195 | 0.999532 | | 7.978459 | 0.489248 | 0.455386 | | 6.432367 | 0.881426 | 0.162189 |
| DNAJB13 | 2.670564 | -0.07991 | 0.999532 | | 2.409499 | -0.88763 | 0.455386 | | 1.94513 | -0.78929 | 0.585657 |
| TAPBP | 18.46185 | 0.115267 | 0.999532 | | 23.4996 | 0.413194 | 0.455386 | | 20.63209 | 0.319428 | 0.644433 |
| IFNAR2 | 4.781331 | -0.50815 | 0.999532 | | 5.011702 | -0.60608 | 0.455386 | | 3.700621 | -0.08317 | 0.961567 |
| YWHAQ | 7.207732 | 0.056519 | 0.999532 | | 9.383539 | 0.453942 | 0.455386 | | 8.149004 | 0.411997 | 0.617561 |
| MYCBP2 | 3.854501 | -0.02665 | 0.999532 | | 5.222702 | 0.579553 | 0.455386 | | 4.362946 | 0.622706 | 0.506666 |
| SEM1 | 10.21609 | 0.193163 | 0.999532 | | 12.20226 | 0.461246 | 0.455386 | | 11.25764 | 0.26611 | 0.75713 |
| ZNF653 | 4.579784 | -0.31111 | 0.999532 | | 4.723919 | -0.61701 | 0.455386 | | 3.575769 | -0.28729 | 0.831749 |
| ST3GAL5 | 2.761768 | -0.08888 | 0.999532 | | 2.513153 | -0.84935 | 0.455386 | | 2.041475 | -0.7433 | 0.60629 |
| CR1L | 5.447708 | -0.23281 | 0.999532 | | 5.627211 | -0.61876 | 0.455386 | | 4.323997 | -0.3615 | 0.764366 |
| COL6A3 | 5.393487 | -0.48424 | 0.999532 | | 10.34831 | 0.960846 | 0.455386 | | 7.967573 | 1.4639 | 0.228254 |
| HSPB7 | 3.07687 | -0.50323 | 0.999532 | | 3.276128 | -0.77656 | 0.455386 | | 2.267971 | -0.2491 | 0.892869 |
| KIAA0408 | 2.991104 | -0.08922 | 0.999532 | | 2.73514 | -0.85855 | 0.455386 | | 2.202789 | -0.74813 | 0.60433 |
| CDC42SE | 4.490798 | 0.416463 | 0.999532 | | 5.302232 | 0.57177 | 0.455386 | | 5.340374 | 0.173051 | 0.886495 |
| ACSF2 | 7.686789 | 0.090546 | 0.999532 | | 9.796799 | 0.47959 | 0.455386 | | 8.395338 | 0.414686 | 0.616301 |
| SEPTIN10 | 5.252472 | 0.184177 | 0.999532 | | 6.933354 | 0.525074 | 0.455386 | | 6.126664 | 0.362346 | 0.702463 |
| RSPH9 | 2.948634 | -0.0443 | 0.999532 | | 2.635567 | -0.83744 | 0.455386 | | 2.30983 | -0.77999 | 0.570037 |
| TLX1 | 2.355628 | 0.154126 | 0.999532 | | 1.97932 | -1.02349 | 0.455386 | | 1.802265 | -1.15887 | 0.423641 |
| CAPNS1 | 18.95616 | -0.20455 | 0.999532 | | 25.70131 | 0.363086 | 0.455386 | | 20.73888 | 0.587412 | 0.202199 |
| SPAG9 | 6.489544 | -0.0321 | 0.999532 | | 9.098789 | 0.457508 | 0.455386 | | 7.521165 | 0.514098 | 0.515473 |
| SLC25A37 | 3.897634 | 0.272069 | 0.999532 | | 4.980702 | 0.635079 | 0.45553 | | 4.706281 | 0.384071 | 0.739496 |
| PTPRE | 3.372416 | -0.52764 | 0.999532 | | 3.598735 | -0.69436 | 0.45553 | | 2.556962 | -0.14517 | 0.938429 |
| VRTN | 2.477181 | -0.04932 | 0.999532 | | 2.316507 | -0.90427 | 0.45553 | | 1.799224 | -0.83793 | 0.566812 |
| POTEE | 3.612252 | 0.494638 | 0.999532 | | 2.764908 | -0.81749 | 0.45553 | | 2.981169 | -1.29455 | 0.175765 |
| CYSRT1 | 3.31687 | -0.3793 | 0.999532 | | 3.290297 | -0.81488 | 0.45553 | | 2.480571 | -0.42033 | 0.794815 |
| MIS12 | 2.97917 | -0.60573 | 0.999532 | | 3.171693 | -0.73711 | 0.455642 | | 2.04009 | -0.11526 | 0.958191 |
| PRKCQ | 2.390953 | -0.42348 | 0.999532 | | 2.437736 | -0.88123 | 0.455829 | | 1.857258 | -0.43735 | 0.814247 |
| MSL3 | 6.035678 | 0.282857 | 0.999532 | | 4.748119 | -0.63329 | 0.455874 | | 4.799515 | -0.91055 | 0.254511 |
| OR2V1 | 2.673666 | -0.04395 | 0.999532 | | 2.392928 | -0.86364 | 0.45641 | | 2.029746 | -0.80201 | 0.566812 |
| DARS1 | 7.480349 | 0.36278 | 0.999532 | | 8.788456 | 0.481262 | 0.45641 | | 8.449096 | 0.135926 | 0.894617 |
| NAPA | 3.974026 | 0.046679 | 0.999532 | | 5.252843 | 0.633272 | 0.456493 | | 4.401235 | 0.619256 | 0.542124 |
| LYZL1 | 2.555719 | -0.18641 | 0.999532 | | 2.385634 | -0.85196 | 0.456493 | | 1.88085 | -0.64926 | 0.668481 |
| SVIP | 5.581942 | 0.20307 | 0.999532 | | 7.021267 | 0.504476 | 0.456493 | | 6.417026 | 0.319748 | 0.713076 |
| TRMT2A | 3.052492 | 0.04133 | 0.999532 | | 2.620048 | -0.81861 | 0.456493 | | 2.324389 | -0.84654 | 0.532278 |
| NFATC2 | 3.218837 | 0.039632 | 0.999532 | | 2.597416 | -0.78437 | 0.456493 | | 2.472718 | -0.82579 | 0.515762 |
| TAB2 | 3.637023 | 0.335453 | 0.999532 | | 4.598341 | 0.622844 | 0.456493 | | 4.376263 | 0.30601 | 0.792539 |
| SCUBE1 | 2.813442 | -0.37334 | 0.999532 | | 2.704566 | -0.76079 | 0.456493 | | 2.088859 | -0.37232 | 0.821463 |
| SLC23A3 | 3.085358 | 0.388261 | 0.999532 | | 4.040374 | 0.710551 | 0.456493 | | 3.902234 | 0.342941 | 0.807942 |
| GOLGA8I | 10.36474 | 0.181129 | 0.999532 | | 9.285258 | -0.44288 | 0.456493 | | 8.400786 | -0.60689 | 0.295502 |
| DCBLD1 | 3.256346 | 0.152351 | 0.999532 | | 4.417617 | 0.653452 | 0.456493 | | 4.02642 | 0.53265 | 0.630705 |
| SENP3 | 6.578 | -0.04814 | 0.999532 | | 6.003751 | -0.55339 | 0.456493 | | 5.009958 | -0.51051 | 0.620745 |
| ZNF81 | 2.718396 | -0.31375 | 0.999532 | | 2.476093 | -0.80932 | 0.456493 | | 2.030493 | -0.48452 | 0.757835 |
| LRRC28 | 2.053407 | 0.038047 | 0.999532 | | 3.018085 | 0.750443 | 0.456493 | | 2.688366 | 0.731873 | 0.554866 |
| SOCS1 | 2.429572 | -0.26141 | 0.999532 | | 2.323046 | -0.85402 | 0.456846 | | 1.742597 | -0.577 | 0.723933 |
| PCDH12 | 4.961755 | 0.029606 | 0.999532 | | 6.591132 | 0.519785 | 0.457409 | | 5.765654 | 0.507813 | 0.557909 |
| GPC6 | 2.082574 | 0.217773 | 0.999532 | | 2.922488 | 0.742341 | 0.457409 | | 2.647198 | 0.542644 | 0.672279 |
| CLPX | 2.744956 | 0.333645 | 0.999532 | | 3.336862 | 0.711582 | 0.457409 | | 3.171996 | 0.391454 | 0.765012 |
| GNAI2 | 38.97823 | -0.18946 | 0.999532 | | 54.11897 | 0.402942 | 0.458108 | | 43.86642 | 0.613892 | 0.235029 |
| PAM | 5.995317 | 0.059166 | 0.999532 | | 8.348561 | 0.537117 | 0.45914 | | 6.920046 | 0.498352 | 0.599002 |
| CDK5 | 3.778505 | -0.12126 | 0.999532 | | 3.463397 | -0.69078 | 0.459417 | | 2.932522 | -0.55271 | 0.657538 |
| HECTD1 | 3.785811 | -0.05707 | 0.999532 | | 5.361085 | 0.570909 | 0.45963 | | 4.577134 | 0.648139 | 0.465298 |
| GPAT4 | 7.426841 | 0.58914 | 0.999532 | | 7.57617 | 0.518778 | 0.45963 | | 7.938871 | -0.04482 | 0.974421 |
| RPL36AL | 5.637978 | 0.110741 | 0.999532 | | 7.53267 | 0.496746 | 0.459661 | | 6.467037 | 0.401039 | 0.64142 |
| NDUFA10 | 5.633832 | -0.16783 | 0.999532 | | 8.183025 | 0.519042 | 0.459661 | | 6.342151 | 0.70499 | 0.300852 |
| CAB39 | 4.416707 | 0.178294 | 0.999532 | | 5.566497 | 0.572744 | 0.459661 | | 5.134373 | 0.401472 | 0.709825 |
| TMA16 | 2.346759 | 0.175464 | 0.999532 | | 3.367331 | 0.690619 | 0.459661 | | 2.951492 | 0.535724 | 0.660097 |
| NACA | 22.66198 | -0.01714 | 0.999532 | | 29.69081 | 0.410274 | 0.459661 | | 25.22482 | 0.446103 | 0.455747 |
| SEPTIN11 | 9.356421 | 0.401532 | 0.999532 | | 10.08804 | 0.450956 | 0.459661 | | 10.17773 | 0.06722 | 0.951656 |
| SMARCC | 6.110659 | 0.402647 | 0.999532 | | 7.168809 | 0.516426 | 0.459661 | | 6.855264 | 0.135099 | 0.89901 |
| ZNF547 | 2.406207 | 0.003948 | 0.999532 | | 2.190669 | -0.87583 | 0.459661 | | 1.845849 | -0.85947 | 0.540342 |
| OR2M5 | 3.28615 | -0.25388 | 0.999532 | | 3.283167 | -0.80922 | 0.459661 | | 2.339086 | -0.53437 | 0.709825 |
| ISLR | 3.335387 | -0.02521 | 0.999532 | | 4.690278 | 0.592346 | 0.459661 | | 3.862184 | 0.635362 | 0.515762 |
| GNAZ | 2.673096 | 0.012487 | 0.999532 | | 2.297672 | -0.86483 | 0.459661 | | 1.912169 | -0.86099 | 0.533876 |
| PRICKLE | 4.148369 | 0.329424 | 0.999532 | | 5.331353 | 0.578425 | 0.459661 | | 4.959621 | 0.268863 | 0.811123 |
| ZBTB38 | 6.29933 | 0.302772 | 0.999532 | | 7.338969 | 0.503024 | 0.459661 | | 7.026112 | 0.224681 | 0.809832 |
| ALDH1A2 | 5.469576 | 0.421485 | 0.999532 | | 6.398283 | 0.54367 | 0.459661 | | 6.419611 | 0.141288 | 0.910105 |
| TMEM238 | 3.711687 | -0.13975 | 0.999532 | | 3.395892 | -0.828 | 0.460053 | | 2.749454 | -0.67065 | 0.647581 |
| BCL9L | 18.59005 | 0.287013 | 0.999532 | | 21.06288 | 0.3442 | 0.46127 | | 19.88568 | 0.075818 | 0.920259 |
| CCL2 | 6.562108 | -1.00563 | 0.772121 | | 7.876986 | -0.65403 | 0.46127 | | 4.85638 | 0.393812 | 0.722703 |
| STX7 | 7.510105 | 0.344588 | 0.999532 | | 8.839332 | 0.494624 | 0.46127 | | 8.151065 | 0.169262 | 0.878591 |
| PRR25 | 3.564574 | -0.40458 | 0.999532 | | 3.725596 | -0.71371 | 0.46127 | | 2.576784 | -0.28778 | 0.858705 |
| PCBP1 | 24.55688 | -0.10476 | 0.999532 | | 32.80163 | 0.361433 | 0.46127 | | 26.84589 | 0.491746 | 0.477153 |
| GNAS | 19.16235 | -0.07359 | 0.999532 | | 26.11075 | 0.425005 | 0.46127 | | 21.89612 | 0.513994 | 0.286063 |
| DNAJC50 | 2.658034 | -0.02285 | 0.999532 | | 2.445022 | -0.83406 | 0.46127 | | 2.009667 | -0.79015 | 0.566812 |
| CLDN24 | 3.158418 | -0.30451 | 0.999532 | | 3.262958 | -0.84425 | 0.461788 | | 2.417758 | -0.49553 | 0.776563 |
| HNF1B | 5.085066 | 0.661616 | 0.999532 | | 5.409366 | 0.623792 | 0.461788 | | 5.696932 | -0.00793 | 0.996891 |
| RAF1 | 8.078998 | 0.337779 | 0.999532 | | 9.563786 | 0.478735 | 0.461788 | | 9.060331 | 0.160011 | 0.87831 |
| CISD3 | 6.267583 | 0.484023 | 0.999532 | | 6.874499 | 0.486181 | 0.461788 | | 7.043475 | 0.033123 | 0.984978 |
| BHLHE40 | 4.495709 | -1.07464 | 0.540613 | | 5.799195 | -0.58235 | 0.461788 | | 3.298694 | 0.508348 | 0.666016 |
| PARP4 | 5.151888 | 0.03878 | 0.999532 | | 6.74146 | 0.543073 | 0.461788 | | 5.918223 | 0.527446 | 0.535662 |
| AIF1L | 13.57392 | 0.395677 | 0.999532 | | 15.33917 | 0.413928 | 0.461788 | | 14.66646 | 0.044081 | 0.968173 |
| MPP5 | 9.351401 | 0.4954 | 0.999532 | | 10.79164 | 0.508767 | 0.461788 | | 10.48728 | 0.04496 | 0.974421 |
| DPH3 | 4.211834 | 0.070496 | 0.999532 | | 3.697205 | -0.70236 | 0.461788 | | 3.320083 | -0.75466 | 0.49299 |
| COMMD6 | 6.889902 | 0.127909 | 0.999532 | | 8.855748 | 0.478444 | 0.461788 | | 7.941846 | 0.371343 | 0.65401 |
| FNDC3B | 9.582983 | 0.299199 | 0.999532 | | 11.34733 | 0.487346 | 0.461788 | | 11.01021 | 0.222589 | 0.800031 |
| PER1 | 4.717828 | -1.21103 | 0.999532 | | 5.582008 | -0.96843 | 0.461788 | | 2.847152 | 0.319089 | 0.842637 |
| MBTPS1 | 8.53526 | -0.17969 | 0.999532 | | 11.6755 | 0.397013 | 0.461788 | | 9.399555 | 0.59487 | 0.275253 |
| AEBP1 | 12.70239 | -0.18149 | 0.999532 | | 18.83387 | 0.573544 | 0.461788 | | 15.41049 | 0.776647 | 0.281853 |
| KCNJ11 | 2.516225 | -0.01508 | 0.999532 | | 2.116868 | -0.91596 | 0.461788 | | 1.921679 | -0.88022 | 0.540758 |
| OR1C1 | 3.039658 | 0.044601 | 0.999532 | | 2.580299 | -0.8151 | 0.461788 | | 2.363222 | -0.84854 | 0.518648 |
| ARID3B | 4.552347 | 0.119797 | 0.999532 | | 3.927149 | -0.66756 | 0.461788 | | 3.713038 | -0.77796 | 0.484303 |
| B3GNT8 | 3.381572 | -0.31096 | 0.999532 | | 3.378799 | -0.74101 | 0.461788 | | 2.694304 | -0.42331 | 0.766251 |
| PFKL | 11.95425 | 0.240257 | 0.999532 | | 14.63424 | 0.384017 | 0.461788 | | 13.07061 | 0.165912 | 0.853018 |
| PURA | 10.07854 | 0.10373 | 0.999532 | | 12.57248 | 0.391413 | 0.461788 | | 11.23502 | 0.308659 | 0.658914 |
| ANKZF1 | 5.413533 | 0.150459 | 0.999532 | | 4.96225 | -0.60929 | 0.461788 | | 4.287767 | -0.74262 | 0.378764 |
| ADAMTS4 | 4.418862 | 0.308315 | 0.999532 | | 3.37142 | -0.69547 | 0.461788 | | 3.607513 | -0.98138 | 0.310637 |
| HEBP1 | 7.825346 | 0.37042 | 0.999532 | | 8.942961 | 0.473819 | 0.461788 | | 8.594926 | 0.11833 | 0.914929 |
| TMEM179 | 2.815115 | -0.06746 | 0.999532 | | 2.490871 | -0.86074 | 0.461788 | | 2.099933 | -0.77694 | 0.598329 |
| ZBTB22 | 4.970074 | 0.486145 | 0.999532 | | 5.68452 | 0.579591 | 0.461788 | | 5.520558 | 0.121362 | 0.926513 |
| LAMA2 | 2.521823 | 0.914 | 0.999532 | | 2.419325 | 0.811538 | 0.461788 | | 2.938046 | -0.0961 | 0.963492 |
| ERMARD | 3.355707 | 0.330129 | 0.999532 | | 4.500572 | 0.658859 | 0.461788 | | 4.187789 | 0.346356 | 0.7719 |
| THBS2 | 2.348009 | 0.762865 | 0.999532 | | 2.703062 | 0.808663 | 0.462292 | | 2.728393 | 0.085675 | 0.968434 |
| ARL1 | 3.715407 | 0.508721 | 0.999532 | | 4.602738 | 0.691396 | 0.462634 | | 4.342713 | 0.206582 | 0.877943 |
| RBM3 | 8.140119 | -0.04944 | 0.999532 | | 11.13054 | 0.438377 | 0.46292 | | 9.468664 | 0.510207 | 0.452179 |
| KCNJ4 | 5.474954 | -0.21949 | 0.999532 | | 5.332258 | -0.5906 | 0.46292 | | 4.346768 | -0.35135 | 0.761699 |
| ETNK1 | 5.953833 | -0.59922 | 0.999532 | | 7.107687 | -0.50284 | 0.46292 | | 4.522581 | 0.122766 | 0.928522 |
| MXD3 | 2.846135 | -0.11596 | 0.999532 | | 2.612768 | -0.81653 | 0.46292 | | 2.11058 | -0.68199 | 0.628895 |
| RAVER1 | 5.179965 | 0.177972 | 0.999532 | | 7.032479 | 0.565609 | 0.46292 | | 6.167668 | 0.411766 | 0.668481 |
| DNAJB8 | 2.133579 | -0.35043 | 0.999532 | | 2.067419 | -0.92533 | 0.46292 | | 1.522325 | -0.55548 | 0.757035 |
| TPD52L2 | 6.55735 | 0.380718 | 0.999532 | | 7.834023 | 0.529862 | 0.46292 | | 7.550081 | 0.165345 | 0.876613 |
| DAZAP1 | 7.050716 | 0.03231 | 0.999532 | | 9.210824 | 0.497912 | 0.46292 | | 7.90936 | 0.476584 | 0.619615 |
| SPPL2A | 3.907073 | -0.37764 | 0.999532 | | 6.227085 | 0.527419 | 0.46292 | | 4.630224 | 0.921585 | 0.241605 |
| OVOL1 | 3.214232 | -0.19926 | 0.999532 | | 3.277093 | -0.74231 | 0.46292 | | 2.405896 | -0.5182 | 0.704316 |
| SIX5 | 3.304675 | -0.08686 | 0.999532 | | 3.117999 | -0.7616 | 0.46292 | | 2.612036 | -0.64895 | 0.644433 |
| AURKAIP | 6.83066 | -0.26308 | 0.999532 | | 7.310745 | -0.53227 | 0.46292 | | 5.646837 | -0.25062 | 0.8233 |
| MRPS30 | 3.968214 | 0.100458 | 0.999532 | | 5.110331 | 0.57913 | 0.46292 | | 4.547989 | 0.506773 | 0.606305 |
| RPL38 | 26.95305 | -0.12538 | 0.999532 | | 37.0842 | 0.437979 | 0.46292 | | 30.53138 | 0.582215 | 0.299861 |
| FCHSD1 | 3.295323 | -0.23166 | 0.999532 | | 3.290037 | -0.70679 | 0.46292 | | 2.530605 | -0.45599 | 0.735469 |
| N4BP2 | 2.800096 | -0.65154 | 0.999532 | | 3.0331 | -0.77426 | 0.46292 | | 2.146492 | -0.10706 | 0.965649 |
| IFITM10 | 4.099105 | 0.107155 | 0.999532 | | 3.887842 | -0.66255 | 0.46292 | | 3.282447 | -0.71535 | 0.563461 |
| CHP1 | 8.537142 | -0.19205 | 0.999532 | | 8.986992 | -0.47249 | 0.46292 | | 6.962755 | -0.25832 | 0.791576 |
| PTGER2 | 2.932216 | -0.02096 | 0.999532 | | 2.804908 | -0.78203 | 0.463324 | | 2.321961 | -0.73018 | 0.579064 |
| KLC1 | 10.53874 | 0.022589 | 0.999532 | | 14.21333 | 0.439086 | 0.463324 | | 12.14564 | 0.447303 | 0.542124 |
| PPP1R12 | 5.397464 | 0.015137 | 0.999532 | | 7.203792 | 0.52781 | 0.463324 | | 6.501888 | 0.544702 | 0.540342 |
| ANXA8 | 3.276214 | 0.090343 | 0.999532 | | 3.146244 | -0.76217 | 0.463647 | | 2.566121 | -0.81521 | 0.510412 |
| MAT2A | 6.916963 | -0.21135 | 0.999532 | | 9.807905 | 0.462203 | 0.464083 | | 7.77105 | 0.695515 | 0.263754 |
| CDKN1C | 14.81532 | -0.0341 | 0.999532 | | 20.24712 | 0.50142 | 0.464083 | | 17.15402 | 0.554305 | 0.359441 |
| MAPK12 | 4.582977 | -0.49792 | 0.999532 | | 4.837564 | -0.65752 | 0.464083 | | 3.453353 | -0.12903 | 0.938429 |
| STC2 | 2.285887 | 0.070809 | 0.999532 | | 2.011977 | -0.94798 | 0.464083 | | 1.661314 | -1.00383 | 0.515762 |
| ALAD | 3.679082 | 0.265671 | 0.999532 | | 4.599048 | 0.638134 | 0.464083 | | 4.214727 | 0.392526 | 0.726571 |
| IRAK1BP | 2.681746 | -0.1949 | 0.999532 | | 2.635082 | -0.85617 | 0.464556 | | 2.066753 | -0.64167 | 0.673405 |
| MMP7 | 3.343434 | 0.174275 | 0.999532 | | 5.253946 | 1.020464 | 0.464556 | | 4.581097 | 0.945197 | 0.434766 |
| LHX1 | 2.986136 | -0.3342 | 0.999532 | | 2.920595 | -0.94911 | 0.464863 | | 2.186473 | -0.64378 | 0.660958 |
| CCL4L1 | 3.393955 | 0.007854 | 0.999532 | | 3.053175 | -0.79111 | 0.464863 | | 2.611324 | -0.77956 | 0.528909 |
| TRAM1L1 | 2.629019 | -0.35568 | 0.999532 | | 2.684175 | -0.78175 | 0.464863 | | 1.883643 | -0.40618 | 0.803301 |
| GIPC2 | 2.858543 | -0.42279 | 0.999532 | | 2.88124 | -0.77164 | 0.464863 | | 2.130974 | -0.3304 | 0.849682 |
| TMEM174 | 3.711287 | 0.14872 | 0.999532 | | 4.921864 | 0.641145 | 0.464863 | | 4.542442 | 0.498386 | 0.660097 |
| NIPSNAP | 4.687576 | 0.158216 | 0.999532 | | 6.143901 | 0.554281 | 0.464863 | | 5.485777 | 0.416338 | 0.682165 |
| APPL1 | 8.938886 | 0.15281 | 0.999532 | | 10.751 | 0.419258 | 0.464863 | | 9.58435 | 0.28136 | 0.717627 |
| BRD9 | 3.380807 | 1.217411 | 0.39263 | | 2.946095 | 0.721314 | 0.464863 | | 3.808347 | -0.4778 | 0.663413 |
| C1QL2 | 4.494872 | -0.10777 | 0.999532 | | 4.065932 | -0.66061 | 0.464863 | | 3.477649 | -0.53685 | 0.66232 |
| RCBTB2 | 3.830219 | 0.319786 | 0.999532 | | 4.842726 | 0.571597 | 0.464954 | | 4.497366 | 0.270363 | 0.810054 |
| FKBP1A | 21.72812 | -0.1042 | 0.999532 | | 28.12544 | 0.32372 | 0.464954 | | 23.31342 | 0.441576 | 0.377051 |
| TFB1M | 2.926983 | -0.20681 | 0.999532 | | 2.913205 | -0.75531 | 0.464954 | | 2.265677 | -0.52579 | 0.718643 |
| COX6A2 | 6.828351 | -0.29652 | 0.999532 | | 7.268792 | -0.56202 | 0.464954 | | 5.557285 | -0.24667 | 0.835554 |
| TRH | 2.732923 | 0.374534 | 0.999532 | | 1.999021 | -0.94432 | 0.464954 | | 2.224396 | -1.30232 | 0.275253 |
| CTSA | 8.914689 | 0.035639 | 0.999532 | | 11.81499 | 0.446049 | 0.464954 | | 9.860928 | 0.440375 | 0.566812 |
| LIG3 | 1.897468 | 0.470071 | 0.999532 | | 2.369076 | 0.804211 | 0.464954 | | 2.343318 | 0.350671 | 0.824012 |
| FOXB1 | 3.358551 | -0.08241 | 0.999532 | | 3.161886 | -0.75362 | 0.464954 | | 2.570911 | -0.65053 | 0.622204 |
| SLC39A9 | 3.1396 | 0.008486 | 0.999532 | | 4.308329 | 0.606016 | 0.464954 | | 3.76667 | 0.615897 | 0.542124 |
| SPDYE11 | 12.34784 | 0.158548 | 0.999532 | | 11.16011 | -0.60082 | 0.464954 | | 10.21648 | -0.74244 | 0.359441 |
| FOXO6 | 3.509499 | -0.25336 | 0.999532 | | 3.505364 | -0.74221 | 0.464954 | | 2.719983 | -0.46874 | 0.739496 |
| HDAC9 | 3.487131 | -0.17641 | 0.999532 | | 5.192622 | 0.563853 | 0.464954 | | 4.222039 | 0.758217 | 0.374949 |
| BBX | 3.614936 | 0.150973 | 0.999532 | | 4.893044 | 0.573768 | 0.464954 | | 4.355372 | 0.440391 | 0.66232 |
| SYNPO | 46.28467 | 0.09321 | 0.999532 | | 57.12442 | 0.340316 | 0.464954 | | 49.7728 | 0.267304 | 0.669356 |
| CCDC140 | 3.14727 | -0.04552 | 0.999532 | | 2.911008 | -0.80924 | 0.464954 | | 2.506481 | -0.7486 | 0.57215 |
| RBBP9 | 2.347069 | 0.207969 | 0.999532 | | 3.156966 | 0.703548 | 0.464954 | | 2.951022 | 0.513981 | 0.683901 |
| EIF3B | 4.388156 | -0.22639 | 0.999532 | | 6.19863 | 0.51728 | 0.464954 | | 4.938373 | 0.757292 | 0.326658 |
| ARL6 | 2.522673 | 0.457693 | 0.999532 | | 3.337603 | 0.694745 | 0.464954 | | 3.212074 | 0.260187 | 0.859522 |
| OR2L5 | 3.558184 | -0.29866 | 0.999532 | | 3.578183 | -0.77778 | 0.464954 | | 2.793481 | -0.46656 | 0.75311 |
| POTED | 3.445278 | 0.000396 | 0.999532 | | 3.097189 | -0.71395 | 0.464954 | | 2.82951 | -0.69763 | 0.566812 |
| HPD | 2.897592 | -0.14085 | 0.999532 | | 4.322473 | 0.747596 | 0.464954 | | 3.645617 | 0.876858 | 0.481527 |
| TNFSF13 | 4.009011 | -0.02629 | 0.999532 | | 3.794505 | -0.65807 | 0.464954 | | 3.199239 | -0.61376 | 0.595502 |
| MAP3K1 | 2.233512 | 0.008551 | 0.999532 | | 3.093577 | 0.719905 | 0.464954 | | 2.639208 | 0.728648 | 0.541136 |
| STK32C | 2.519554 | -0.31101 | 0.999532 | | 2.473677 | -0.78661 | 0.464954 | | 1.92515 | -0.45117 | 0.791466 |
| CMA1 | 2.719246 | 0.758287 | 0.999532 | | 3.141576 | 0.721266 | 0.465245 | | 3.344952 | -0.01888 | 0.995112 |
| DCTN3 | 5.190269 | 0.138708 | 0.999532 | | 6.852867 | 0.529987 | 0.465245 | | 6.076246 | 0.410574 | 0.660958 |
| CTSV | 5.439103 | 0.960205 | 0.999532 | | 5.0345 | 0.592809 | 0.465253 | | 5.82272 | -0.37698 | 0.728328 |
| ROCK2 | 8.829351 | -0.0067 | 0.999532 | | 11.70028 | 0.44325 | 0.465253 | | 9.928062 | 0.468992 | 0.515762 |
| ANAPC13 | 6.340077 | -0.25932 | 0.999532 | | 9.159634 | 0.442704 | 0.465253 | | 7.442897 | 0.722233 | 0.254511 |
| ZNF585A | 4.321371 | -0.19145 | 0.999532 | | 4.236804 | -0.63422 | 0.465253 | | 3.437408 | -0.42453 | 0.723933 |
| SUSD4 | 2.213918 | 0.26174 | 0.999532 | | 1.714897 | -0.97797 | 0.465253 | | 1.689553 | -1.22449 | 0.37596 |
| ANKRD16 | 3.037705 | -0.11976 | 0.999532 | | 2.82879 | -0.78648 | 0.465253 | | 2.328682 | -0.64995 | 0.630705 |
| H4C4 | 4.970915 | 0.074993 | 0.999532 | | 6.560975 | 0.514652 | 0.46572 | | 5.567356 | 0.456149 | 0.630705 |
| BAALC | 1.725897 | 1.328393 | 0.999532 | | 1.705955 | 0.974071 | 0.46572 | | 2.082532 | -0.3377 | 0.842754 |
| ABLIM3 | 2.898008 | -0.68596 | 0.999532 | | 3.222663 | -0.73006 | 0.46572 | | 2.18083 | -0.02369 | 0.99505 |
| NADK | 2.618883 | 0.887663 | 0.999532 | | 2.638653 | 0.762802 | 0.46572 | | 3.006497 | -0.10889 | 0.952785 |
| HNRNPM | 12.12409 | -0.23893 | 0.999532 | | 17.10763 | 0.383014 | 0.46572 | | 13.44519 | 0.642568 | 0.209531 |
| RAB3GAI | 4.539278 | -0.09965 | 0.999532 | | 6.168446 | 0.588638 | 0.46572 | | 5.257919 | 0.703914 | 0.433186 |
| RPS10 | 27.51268 | -0.13264 | 0.999532 | | 29.4989 | -0.32147 | 0.46572 | | 23.2602 | -0.15054 | 0.844581 |
| RPS27A | 55.40564 | 0.000463 | 0.999532 | | 69.18503 | 0.301736 | 0.46572 | | 58.92382 | 0.32224 | 0.515762 |
| NANOS3 | 3.327656 | -0.36815 | 0.999532 | | 3.530697 | -0.73239 | 0.465998 | | 2.741336 | -0.34003 | 0.83841 |
| SEMA7A | 2.258442 | 0.41184 | 0.999532 | | 3.018316 | 0.735337 | 0.465998 | | 2.917908 | 0.349154 | 0.807646 |
| C8orf89 | 2.53839 | -0.26229 | 0.999532 | | 2.554576 | -0.7928 | 0.466314 | | 1.942876 | -0.50547 | 0.747457 |
| ARHGEF | 2.915902 | -0.51963 | 0.999532 | | 3.308669 | -0.72746 | 0.466314 | | 2.070614 | -0.16482 | 0.935604 |
| CEBPZ | 2.913115 | -0.21395 | 0.999532 | | 2.748502 | -0.77203 | 0.466314 | | 2.136169 | -0.54047 | 0.717627 |
| INTS5 | 3.45708 | 0.199022 | 0.999532 | | 2.788285 | -0.76383 | 0.466314 | | 2.777111 | -0.937 | 0.411291 |
| PRPF19 | 3.667708 | 0.269011 | 0.999532 | | 4.615424 | 0.619887 | 0.466314 | | 4.240886 | 0.367021 | 0.750371 |
| THG1L | 2.710924 | -0.08806 | 0.999532 | | 2.491084 | -0.82223 | 0.466314 | | 1.992458 | -0.71474 | 0.623462 |
| PRPF39 | 7.581548 | -0.13262 | 0.999532 | | 10.44483 | 0.420672 | 0.466314 | | 8.641534 | 0.575272 | 0.415282 |
| KRTAP10 | 2.718454 | 0.309973 | 0.999532 | | 2.061661 | -0.88884 | 0.466314 | | 2.081349 | -1.18714 | 0.329409 |
| NELL1 | 1.947708 | -0.45066 | 0.999532 | | 3.541502 | 0.848652 | 0.466376 | | 2.820855 | 1.29015 | 0.277462 |
| PTOV1 | 9.588171 | 0.307291 | 0.999532 | | 11.39392 | 0.481236 | 0.466376 | | 10.43981 | 0.211169 | 0.836133 |
| NDUFS6 | 10.93769 | 0.145917 | 0.999532 | | 13.03824 | 0.397149 | 0.466376 | | 12.06672 | 0.265588 | 0.722703 |
| AP1S2 | 8.624702 | 0.103132 | 0.999532 | | 11.00767 | 0.501934 | 0.466376 | | 10.02671 | 0.414778 | 0.653214 |
| GRK3 | 2.560382 | -0.51335 | 0.999532 | | 4.150517 | 0.615852 | 0.466376 | | 3.148214 | 1.14629 | 0.222058 |
| UBE2L3 | 10.07293 | 0.237169 | 0.999532 | | 11.80373 | 0.394105 | 0.466773 | | 11.00096 | 0.175408 | 0.842686 |
| EIF3D | 6.766758 | -0.01203 | 0.999532 | | 8.783044 | 0.467942 | 0.466773 | | 7.42997 | 0.496315 | 0.527774 |
| NANOGP | 3.356775 | -0.07358 | 0.999532 | | 3.186042 | -0.7524 | 0.467188 | | 2.646295 | -0.64795 | 0.64142 |
| PCMTD2 | 5.772487 | 0.325313 | 0.999532 | | 7.05048 | 0.489282 | 0.467188 | | 6.521447 | 0.184965 | 0.864923 |
| NEBL | 14.08304 | 0.703296 | 0.912432 | | 14.95239 | 0.534998 | 0.467188 | | 15.65953 | -0.13071 | 0.920259 |
| ATP9A | 2.342464 | 0.402534 | 0.999532 | | 2.930631 | 0.706394 | 0.467195 | | 2.787437 | 0.319433 | 0.821752 |
| NAT10 | 1.957581 | 0.729058 | 0.999532 | | 2.464475 | 0.83234 | 0.467195 | | 2.396971 | 0.118264 | 0.954597 |
| INTS8 | 3.22215 | 0.314996 | 0.999532 | | 3.815268 | 0.624442 | 0.467195 | | 3.657514 | 0.327864 | 0.789815 |
| KRT18 | 19.59827 | 0.052093 | 0.999532 | | 18.84556 | -0.46622 | 0.467195 | | 16.56589 | -0.4976 | 0.504551 |
| PEX1 | 2.704856 | 0.052483 | 0.999532 | | 2.390916 | -0.80806 | 0.467195 | | 2.038746 | -0.84184 | 0.515762 |
| DCAF12 | 2.651066 | 0.336137 | 0.999532 | | 3.433356 | 0.706323 | 0.467195 | | 3.075029 | 0.381357 | 0.767184 |
| BCR | 3.822238 | 0.382995 | 0.999532 | | 4.913799 | 0.568486 | 0.467195 | | 4.48465 | 0.204152 | 0.874653 |
| SNX4 | 2.675992 | 0.029018 | 0.999532 | | 3.71824 | 0.652421 | 0.467195 | | 3.042971 | 0.642164 | 0.572084 |
| AGO4 | 3.189341 | 0.529597 | 0.999532 | | 3.561698 | 0.645993 | 0.467195 | | 3.568813 | 0.13351 | 0.929549 |
| POLR2J3 | 10.47746 | -0.51579 | 0.999532 | | 11.85542 | -0.59145 | 0.467195 | | 8.557956 | -0.04903 | 0.982124 |
| ZNF750 | 2.430603 | 0.209228 | 0.999532 | | 1.927944 | -0.89543 | 0.467195 | | 1.902624 | -1.08876 | 0.412558 |
| FRAS1 | 1.986247 | 0.903973 | 0.999532 | | 2.22574 | 0.838649 | 0.467195 | | 2.441065 | -0.0477 | 0.985675 |
| LYNX1 | 6.786047 | 0.189845 | 0.999532 | | 6.113223 | -0.569 | 0.467195 | | 5.529075 | -0.73253 | 0.407399 |
| FAM216A | 3.564402 | -0.31913 | 0.999532 | | 3.516254 | -0.67748 | 0.467195 | | 2.684644 | -0.34377 | 0.816618 |
| SELENON | 5.983719 | 0.497312 | 0.999532 | | 6.933599 | 0.581037 | 0.467195 | | 6.740509 | 0.112448 | 0.938929 |
| CAMK2N1 | 8.428028 | 0.038211 | 0.999532 | | 11.23856 | 0.486466 | 0.467195 | | 9.619207 | 0.467524 | 0.506666 |
| ACAA2 | 4.679784 | 0.317778 | 0.999532 | | 5.820863 | 0.594509 | 0.467195 | | 5.407504 | 0.296257 | 0.795612 |
| PRRC2A | 9.123921 | 0.19664 | 0.999532 | | 11.16111 | 0.405505 | 0.467195 | | 9.903801 | 0.22447 | 0.800724 |
| TEK | 5.937205 | -0.31592 | 0.999532 | | 8.786209 | 0.468291 | 0.467195 | | 6.967233 | 0.806942 | 0.234174 |
| DCXR | 4.060107 | 0.687231 | 0.999532 | | 4.525995 | 0.637142 | 0.467195 | | 4.510116 | -0.02815 | 0.989536 |
| PRR32 | 2.33082 | -0.09539 | 0.999532 | | 2.002331 | -0.87784 | 0.467195 | | 1.766837 | -0.77386 | 0.628895 |
| LCT | 2.624558 | -0.0582 | 0.999532 | | 2.462024 | -0.81216 | 0.467195 | | 1.949491 | -0.73397 | 0.600289 |
| MAPK15 | 2.748252 | 0.766895 | 0.999532 | | 3.24706 | 0.750466 | 0.467195 | | 3.423654 | 0.010516 | 0.996891 |
| FAM110C | 3.087898 | 0.292451 | 0.999532 | | 2.652201 | -0.78518 | 0.467195 | | 2.369645 | -1.04678 | 0.328367 |
| CZIB | 3.245883 | 0.394052 | 0.999532 | | 4.356998 | 0.646529 | 0.467195 | | 3.850826 | 0.254351 | 0.851503 |
| NPIPB3 | 15.13669 | 0.260758 | 0.999532 | | 17.42084 | 0.389314 | 0.467195 | | 16.15059 | 0.146059 | 0.862165 |
| GSAP | 3.186293 | -0.18125 | 0.999532 | | 3.045784 | -0.75015 | 0.467195 | | 2.35353 | -0.54973 | 0.700565 |
| P3H1 | 2.876783 | 0.003167 | 0.999532 | | 3.933169 | 0.620587 | 0.467195 | | 3.211272 | 0.631897 | 0.5451 |
| RAPGEFL | 3.15428 | -0.61045 | 0.999532 | | 3.523793 | -0.68665 | 0.467195 | | 2.432535 | -0.05468 | 0.983199 |
| KXD1 | 3.429109 | 0.00151 | 0.999532 | | 4.779714 | 0.586156 | 0.467195 | | 4.207702 | 0.606406 | 0.542124 |
| EIF3L | 9.662665 | 0.206505 | 0.999532 | | 12.32558 | 0.509159 | 0.467195 | | 11.26868 | 0.316578 | 0.704316 |
| C11orf54 | 5.785194 | 0.021267 | 0.999532 | | 8.530736 | 0.648111 | 0.467195 | | 6.985823 | 0.69684 | 0.531575 |
| CLN5 | 4.031706 | 0.562492 | 0.999532 | | 4.432205 | 0.593869 | 0.467195 | | 4.537905 | 0.048612 | 0.977541 |
| NCOA4 | 9.205193 | -0.42168 | 0.999532 | | 10.28931 | -0.42678 | 0.467195 | | 7.260271 | 0.011065 | 0.996681 |
| TPM4 | 50.69903 | 0.011776 | 0.999532 | | 65.55097 | 0.398391 | 0.467707 | | 56.43383 | 0.409277 | 0.462115 |
| SSR4 | 8.147053 | 0.454472 | 0.999532 | | 9.458969 | 0.483114 | 0.467707 | | 9.225024 | 0.055162 | 0.972221 |
| STAG1 | 2.556564 | 0.455507 | 0.999532 | | 3.142242 | 0.730628 | 0.468049 | | 3.147892 | 0.294169 | 0.844216 |
| RPS19BP | 5.033727 | 0.076311 | 0.999532 | | 6.598206 | 0.537107 | 0.468572 | | 5.749429 | 0.473414 | 0.632113 |
| CDKN2AI | 4.497734 | -0.5002 | 0.999532 | | 4.7656 | -0.60778 | 0.468572 | | 3.480758 | -0.09105 | 0.96129 |
| LGALS1 | 44.94896 | -0.05126 | 0.999532 | | 61.56871 | 0.496941 | 0.468572 | | 52.17059 | 0.573091 | 0.281994 |
| OR5H15 | 5.888436 | 0.031995 | 0.999532 | | 5.38438 | -0.59819 | 0.468572 | | 4.598321 | -0.60997 | 0.531045 |
| MICU2 | 4.197801 | 0.184679 | 0.999532 | | 5.558579 | 0.552958 | 0.468572 | | 4.921915 | 0.389179 | 0.699971 |
| ZNF628 | 3.235599 | 0.043926 | 0.999532 | | 2.853359 | -0.7658 | 0.468572 | | 2.434175 | -0.79046 | 0.524206 |
| TAF15 | 9.60117 | 0.089001 | 0.999532 | | 12.2067 | 0.429891 | 0.468572 | | 10.97367 | 0.356972 | 0.680428 |
| CHAT | 2.248383 | -0.52135 | 0.999532 | | 2.253845 | -0.82671 | 0.468572 | | 1.675843 | -0.29613 | 0.886495 |
| RACK1 | 26.24677 | -0.05915 | 0.999532 | | 35.94932 | 0.482704 | 0.469234 | | 30.09893 | 0.56187 | 0.421517 |
| SYNGR3 | 3.673211 | -0.13135 | 0.999532 | | 3.647522 | -0.67337 | 0.469499 | | 2.973226 | -0.51302 | 0.688074 |
| PPM1D | 4.530389 | -0.32807 | 0.999532 | | 4.439239 | -0.64529 | 0.469562 | | 3.389377 | -0.2946 | 0.829716 |
| CXCL14 | 7.878265 | 0.209965 | 0.999532 | | 10.55908 | 0.662328 | 0.470112 | | 9.222703 | 0.483438 | 0.765012 |
| OR4D2 | 2.629897 | 0.039095 | 0.999532 | | 2.187614 | -0.87006 | 0.470276 | | 2.042727 | -0.89665 | 0.552672 |
| SNX14 | 3.985366 | -0.20671 | 0.999532 | | 5.776133 | 0.562116 | 0.47048 | | 4.624497 | 0.791106 | 0.309493 |
| STOM | 12.05896 | -0.43877 | 0.999532 | | 13.68674 | -0.36991 | 0.47048 | | 9.992196 | 0.085435 | 0.93614 |
| ELOVL2 | 3.080513 | 0.052305 | 0.999532 | | 2.955369 | -0.81907 | 0.47048 | | 2.35209 | -0.83857 | 0.515762 |
| MOK | 4.199562 | 0.018995 | 0.999532 | | 3.691097 | -0.69153 | 0.47048 | | 3.202896 | -0.6906 | 0.540342 |
| GRHPR | 8.4811 | -0.11464 | 0.999532 | | 11.62712 | 0.392939 | 0.470912 | | 9.256465 | 0.529808 | 0.424976 |
| TSGA10 | 3.332058 | -0.39098 | 0.999532 | | 3.316172 | -0.69794 | 0.470912 | | 2.395068 | -0.29097 | 0.860036 |
| VAT1L | 3.748019 | -0.00442 | 0.999532 | | 3.305455 | -0.77743 | 0.470912 | | 2.935959 | -0.76256 | 0.52516 |
| TRO | 3.580037 | 0.030547 | 0.999532 | | 3.278715 | -0.70528 | 0.470912 | | 2.716612 | -0.71244 | 0.542124 |
| SELENBP | 8.555678 | 0.269328 | 0.999532 | | 10.40638 | 0.41414 | 0.470912 | | 9.491031 | 0.161593 | 0.870694 |
| CIBAR2 | 3.037844 | -0.04282 | 0.999532 | | 2.935282 | -0.78523 | 0.470912 | | 2.440383 | -0.73397 | 0.566812 |
| LRRC37A | 4.23009 | -0.14318 | 0.999532 | | 4.234759 | -0.77847 | 0.470912 | | 3.200437 | -0.65259 | 0.553283 |
| E2F5 | 2.144286 | 0.946097 | 0.999532 | | 2.437852 | 0.850174 | 0.470912 | | 2.762488 | -0.07733 | 0.973356 |
| PPP5D1 | 2.352969 | -0.3756 | 0.999532 | | 2.190412 | -0.85789 | 0.470996 | | 1.788016 | -0.4632 | 0.803726 |
| ALOX12 | 3.376799 | -0.10653 | 0.999532 | | 3.301747 | -0.73592 | 0.470996 | | 2.639367 | -0.59561 | 0.668481 |
| TEX55 | 2.461849 | -0.1076 | 0.999532 | | 2.412532 | -0.79663 | 0.471568 | | 1.893531 | -0.65798 | 0.660958 |
| PNLIPRP | 3.351268 | -0.3551 | 0.999532 | | 3.568711 | -0.68954 | 0.471702 | | 2.620264 | -0.30672 | 0.85077 |
| API5 | 5.440217 | 0.207916 | 0.999532 | | 7.040405 | 0.524635 | 0.471702 | | 6.259736 | 0.335106 | 0.747457 |
| ZNF555 | 1.934003 | 0.583074 | 0.999532 | | 2.249617 | 0.822319 | 0.472455 | | 2.333412 | 0.253782 | 0.889853 |
| DACT3 | 2.74032 | 0.248654 | 0.999532 | | 2.234104 | -0.83362 | 0.473158 | | 2.137663 | -1.06367 | 0.378764 |
| AP1S1 | 2.450355 | 0.880594 | 0.999532 | | 2.725064 | 0.804555 | 0.473429 | | 2.926155 | -0.05995 | 0.977746 |
| CPT1C | 2.633518 | 0.05864 | 0.999532 | | 2.254432 | -0.85473 | 0.473739 | | 2.075265 | -0.89445 | 0.53177 |
| ASXL2 | 4.230111 | 0.243378 | 0.999532 | | 5.697258 | 0.56097 | 0.473739 | | 4.914601 | 0.362015 | 0.742171 |
| MAML2 | 2.372731 | -0.10361 | 0.999532 | | 3.357422 | 0.660027 | 0.474151 | | 2.778831 | 0.778042 | 0.49299 |
| ANXA1 | 15.48937 | -0.24521 | 0.999532 | | 22.09032 | 0.407584 | 0.475692 | | 17.30771 | 0.679348 | 0.222058 |
| TTPA | 2.945261 | -0.13916 | 0.999532 | | 2.589365 | -0.79681 | 0.475692 | | 2.229374 | -0.64102 | 0.66549 |
| PALLD | 18.95463 | 0.449717 | 0.999532 | | 20.75907 | 0.363731 | 0.475692 | | 20.21092 | -0.05733 | 0.954304 |
| IQGAP2 | 14.19753 | 0.295412 | 0.999532 | | 16.79972 | 0.440439 | 0.475692 | | 15.42251 | 0.170904 | 0.842686 |
| RPS11 | 40.31558 | 0.033828 | 0.999532 | | 51.1012 | 0.378713 | 0.475839 | | 44.18147 | 0.364239 | 0.515762 |
| SLTM | 7.703633 | 0.266533 | 0.999532 | | 9.265416 | 0.428067 | 0.475839 | | 8.402133 | 0.177826 | 0.861119 |
| ALDH7A1 | 7.919694 | -0.08835 | 0.999532 | | 10.92616 | 0.434428 | 0.475839 | | 8.763761 | 0.529902 | 0.465298 |
| CTBP2 | 17.90249 | -0.14175 | 0.999532 | | 18.55425 | -0.34787 | 0.475839 | | 15.56715 | -0.18927 | 0.801409 |
| CRYBA1 | 2.868825 | -0.18718 | 0.999532 | | 2.896437 | -0.74283 | 0.475839 | | 2.374679 | -0.52712 | 0.720526 |
| CDKL3 | 2.363703 | -0.30977 | 0.999532 | | 2.176586 | -0.85073 | 0.475839 | | 1.715582 | -0.53297 | 0.760955 |
| LECT2 | 2.69115 | 0.226107 | 0.999532 | | 2.208454 | -0.8529 | 0.476308 | | 2.018004 | -1.06002 | 0.399012 |
| ZNF429 | 3.032583 | -0.09974 | 0.999532 | | 2.931513 | -0.71119 | 0.476308 | | 2.332984 | -0.59014 | 0.660097 |
| FBXL8 | 5.468152 | 0.002288 | 0.999532 | | 5.186439 | -0.57278 | 0.477033 | | 4.355108 | -0.55621 | 0.563455 |
| STN1 | 4.573697 | -0.1716 | 0.999532 | | 6.746392 | 0.503201 | 0.477392 | | 5.411209 | 0.697557 | 0.37596 |
| SLC10A2 | 1.860988 | 0.763884 | 0.999532 | | 2.075167 | 0.869579 | 0.477392 | | 2.22982 | 0.124244 | 0.955022 |
| KRT81 | 3.152895 | -0.22839 | 0.999532 | | 3.113458 | -0.76373 | 0.47794 | | 2.404475 | -0.5151 | 0.723509 |
| WFDC1 | 5.053912 | -0.73494 | 0.999532 | | 5.894126 | -0.63 | 0.47794 | | 4.063436 | 0.110104 | 0.950616 |
| AFDN | 5.798966 | -0.10837 | 0.999532 | | 8.153729 | 0.465738 | 0.47794 | | 6.637057 | 0.59513 | 0.421517 |
| PECAM1 | 19.27722 | -0.10385 | 0.999532 | | 27.19886 | 0.522578 | 0.478281 | | 22.76721 | 0.639897 | 0.284353 |
| RNF181 | 10.65975 | 0.203116 | 0.999532 | | 12.90663 | 0.382564 | 0.478315 | | 11.59092 | 0.19888 | 0.791576 |
| GID8 | 3.990287 | 0.214003 | 0.999532 | | 5.229955 | 0.554114 | 0.478315 | | 4.618242 | 0.363913 | 0.742171 |
| SENP1 | 3.872228 | -0.3076 | 0.999532 | | 3.947722 | -0.63338 | 0.478315 | | 2.893541 | -0.30638 | 0.829716 |
| TNFSF14 | 2.98311 | -0.41781 | 0.999532 | | 2.891482 | -0.74013 | 0.478315 | | 2.328946 | -0.32179 | 0.864245 |
| PRM2 | 3.51743 | -0.2307 | 0.999532 | | 3.479609 | -0.72805 | 0.478341 | | 2.670201 | -0.47679 | 0.728015 |
| NDUFA2 | 9.433574 | -0.01402 | 0.999532 | | 12.20384 | 0.385181 | 0.478609 | | 10.30135 | 0.417976 | 0.515762 |
| ADCK5 | 4.505468 | -0.21441 | 0.999532 | | 4.38931 | -0.59578 | 0.478784 | | 3.598699 | -0.37215 | 0.767184 |
| SPDYE3 | 7.97407 | -0.1357 | 0.999532 | | 8.160797 | -0.70345 | 0.478784 | | 6.195667 | -0.55385 | 0.605041 |
| NELFE | 5.306454 | 0.028323 | 0.999532 | | 7.264202 | 0.507109 | 0.479545 | | 6.244381 | 0.496507 | 0.578798 |
| ANK2 | 3.102886 | 0.513191 | 0.999532 | | 3.805389 | 0.737885 | 0.480164 | | 3.709677 | 0.251746 | 0.883679 |
| AGXT2 | 3.271821 | 0.727454 | 0.999532 | | 3.428284 | 0.757132 | 0.480164 | | 3.769393 | 0.063113 | 0.97502 |
| ZNF587 | 42.66588 | 0.264727 | 0.999532 | | 38.28629 | -0.4126 | 0.480164 | | 36.64516 | -0.66744 | 0.199116 |
| MARK3 | 10.52299 | 0.230387 | 0.999532 | | 12.25907 | 0.439971 | 0.480183 | | 11.37787 | 0.223362 | 0.804499 |
| PRKACA | 7.963307 | -0.04811 | 0.999532 | | 10.52023 | 0.445506 | 0.480627 | | 9.067195 | 0.521694 | 0.456688 |
| ZFYVE1 | 2.838907 | -0.1624 | 0.999532 | | 2.651903 | -0.80558 | 0.480627 | | 2.173082 | -0.62637 | 0.682485 |
| MED9 | 2.523154 | -0.79589 | 0.999532 | | 2.844039 | -0.76175 | 0.480627 | | 1.836129 | 0.051874 | 0.987274 |
| SERINC3 | 8.027759 | -0.15264 | 0.999532 | | 10.9837 | 0.413914 | 0.480627 | | 9.048844 | 0.583559 | 0.359441 |
| FOXD4L1 | 5.948086 | -0.18974 | 0.999532 | | 6.007092 | -0.56966 | 0.480627 | | 4.764281 | -0.36317 | 0.735469 |
| AGR2 | 2.569348 | 0.11565 | 0.999532 | | 2.313808 | -0.85801 | 0.480627 | | 2.006979 | -0.94602 | 0.504551 |
| KCNG3 | 2.855521 | -0.23835 | 0.999532 | | 2.817907 | -0.80173 | 0.480695 | | 2.219467 | -0.54374 | 0.723933 |
| DCAF6 | 4.400022 | -0.10713 | 0.999532 | | 6.142501 | 0.522006 | 0.480695 | | 5.043725 | 0.645807 | 0.456688 |
| SLC39A13 | 3.629373 | 0.038637 | 0.999532 | | 5.041198 | 0.610146 | 0.480809 | | 4.193648 | 0.603361 | 0.545731 |
| CT45A1 | 4.80311 | -0.08678 | 0.999532 | | 4.520061 | -0.68899 | 0.480809 | | 3.849864 | -0.57977 | 0.626811 |
| TRAPPC1 | 6.63064 | -0.23767 | 0.999532 | | 9.516853 | 0.413686 | 0.480809 | | 7.579327 | 0.671291 | 0.274496 |
| COMT | 10.58306 | 0.060216 | 0.999532 | | 14.08772 | 0.414737 | 0.480809 | | 12.09708 | 0.376256 | 0.589923 |
| B4GALT1 | 9.569209 | -0.20033 | 0.999532 | | 12.76464 | 0.403045 | 0.480809 | | 10.31331 | 0.617145 | 0.300852 |
| BASP1 | 12.98642 | 0.393733 | 0.999532 | | 14.56614 | 0.408991 | 0.480809 | | 14.22722 | 0.04354 | 0.976441 |
| GK | 3.364943 | 0.432963 | 0.999532 | | 4.050683 | 0.637924 | 0.480809 | | 3.787186 | 0.231546 | 0.865363 |
| NDUFB10 | 11.4844 | -0.10482 | 0.999532 | | 14.81555 | 0.349313 | 0.480809 | | 12.21481 | 0.467127 | 0.419745 |
| EGFR | 4.427299 | 0.696775 | 0.999532 | | 4.809397 | 0.574009 | 0.480809 | | 5.007702 | -0.10282 | 0.940744 |
| DDR1 | 5.510254 | 0.587748 | 0.999532 | | 6.086602 | 0.550226 | 0.480809 | | 5.957561 | -0.00786 | 0.996891 |
| PBXIP1 | 6.408091 | 0.129725 | 0.999532 | | 8.284649 | 0.446623 | 0.480809 | | 7.168137 | 0.336158 | 0.699971 |
| HSPB2 | 3.717918 | -0.22138 | 0.999532 | | 3.476666 | -0.65263 | 0.480865 | | 2.811289 | -0.41585 | 0.750371 |
| HMGB1 | 31.0574 | -0.04579 | 0.999532 | | 38.6617 | 0.271295 | 0.481097 | | 32.2941 | 0.340128 | 0.397004 |
| TSKU | 3.92609 | -0.19497 | 0.999532 | | 5.714026 | 0.550792 | 0.48181 | | 4.469129 | 0.769232 | 0.379978 |
| DCLRE1C | 2.851655 | -0.32808 | 0.999532 | | 2.798607 | -0.76373 | 0.48181 | | 2.154876 | -0.41561 | 0.806394 |
| NEK7 | 11.30508 | -0.00558 | 0.999532 | | 14.7225 | 0.397921 | 0.48181 | | 12.72323 | 0.433154 | 0.496445 |
| SUGT1 | 4.910189 | 0.440051 | 0.999532 | | 5.751583 | 0.519185 | 0.48181 | | 5.738264 | 0.101277 | 0.939913 |
| GIT2 | 6.087581 | 0.098361 | 0.999532 | | 7.700678 | 0.451022 | 0.48226 | | 6.694007 | 0.366565 | 0.666906 |
| SKI | 9.057289 | 0.102444 | 0.999532 | | 11.56096 | 0.496075 | 0.482735 | | 10.20789 | 0.412777 | 0.622543 |
| ABAT | 2.21523 | 0.036138 | 0.999532 | | 3.125564 | 0.740494 | 0.483925 | | 2.777234 | 0.712909 | 0.573454 |
| IFIH1 | 2.48395 | 0.146896 | 0.999532 | | 3.663373 | 0.650509 | 0.483925 | | 3.08347 | 0.529149 | 0.66549 |
| PROCR | 2.750908 | -0.34624 | 0.999532 | | 2.649735 | -0.76561 | 0.484764 | | 2.014002 | -0.40415 | 0.808129 |
| OTUB1 | 6.42474 | 0.242961 | 0.999532 | | 7.66936 | 0.467178 | 0.484835 | | 7.304935 | 0.246422 | 0.791466 |
| PGLYRP4 | 2.800321 | -0.2098 | 0.999532 | | 2.667657 | -0.76969 | 0.485534 | | 2.25524 | -0.55267 | 0.722703 |
| DOC2A | 2.781144 | -0.02318 | 0.999532 | | 2.544927 | -0.82195 | 0.485976 | | 2.190407 | -0.77468 | 0.620745 |
| PLEKHG6 | 3.128198 | -0.4197 | 0.999532 | | 3.293052 | -0.69972 | 0.486749 | | 2.509027 | -0.26146 | 0.88476 |
| LNPEP | 4.59262 | 0.15566 | 0.999532 | | 5.7149 | 0.53112 | 0.486749 | | 5.166183 | 0.384788 | 0.717627 |
| PIGL | 5.405248 | -0.38735 | 0.999532 | | 5.813928 | -0.58196 | 0.486749 | | 4.053983 | -0.17667 | 0.894612 |
| TACSTD2 | 2.349797 | 0.289827 | 0.999532 | | 3.085633 | 0.698828 | 0.487044 | | 2.754343 | 0.427449 | 0.750371 |
| GNG3 | 2.768825 | -0.37928 | 0.999532 | | 3.005824 | -0.76823 | 0.487044 | | 2.283891 | -0.36037 | 0.838733 |
| TULP4 | 3.09907 | 0.460552 | 0.999532 | | 3.491702 | 0.638193 | 0.488023 | | 3.54425 | 0.196911 | 0.892869 |
| KCNJ2 | 2.342321 | -0.67806 | 0.999532 | | 2.481235 | -0.77087 | 0.488145 | | 1.715568 | -0.08244 | 0.975753 |
| EEF1D | 34.81724 | -0.03437 | 0.999532 | | 44.80874 | 0.337614 | 0.488756 | | 37.78609 | 0.395849 | 0.414073 |
| FCRLB | 2.169573 | -0.11159 | 0.999532 | | 2.005861 | -0.8585 | 0.488891 | | 1.633226 | -0.72766 | 0.652925 |
| KRTAP2-3 | 3.408619 | 0.083553 | 0.999532 | | 2.886471 | -0.73723 | 0.490092 | | 2.784679 | -0.80071 | 0.515762 |
| SDR16C5 | 2.911618 | -0.01246 | 0.999532 | | 2.494826 | -0.81048 | 0.490092 | | 2.171777 | -0.78137 | 0.566812 |
| SOX17 | 3.307922 | 0.255654 | 0.999532 | | 4.242359 | 0.629728 | 0.490184 | | 4.147306 | 0.40226 | 0.717627 |
| RAB14 | 6.183506 | 0.128437 | 0.999532 | | 8.220277 | 0.441435 | 0.490184 | | 7.097628 | 0.33525 | 0.703463 |
| FAM174C | 3.588087 | -0.92725 | 0.999532 | | 4.791923 | -0.60223 | 0.490184 | | 2.89032 | 0.375815 | 0.799944 |
| DDX5 | 53.99914 | -0.23815 | 0.999532 | | 71.34002 | 0.254676 | 0.490184 | | 56.58174 | 0.509386 | 0.145782 |
| AMPD2 | 6.440927 | -0.2412 | 0.999532 | | 6.641229 | -0.49807 | 0.490184 | | 5.209535 | -0.23813 | 0.837256 |
| ARPC4 | 9.188006 | 0.081112 | 0.999532 | | 12.11239 | 0.494936 | 0.490184 | | 10.54833 | 0.433867 | 0.524206 |
| PCSK1N | 6.693969 | -0.18722 | 0.999532 | | 10.14541 | 0.662786 | 0.490184 | | 7.969369 | 0.924194 | 0.280916 |
| ZNF467 | 2.750678 | 0.068017 | 0.999532 | | 2.380828 | -0.83187 | 0.490184 | | 2.040788 | -0.88705 | 0.531575 |
| MYOZ3 | 3.014697 | -0.08933 | 0.999532 | | 2.761 | -0.78583 | 0.490184 | | 2.314342 | -0.68189 | 0.647581 |
| GAB4 | 2.334238 | -0.11163 | 0.999532 | | 2.130856 | -0.86655 | 0.490301 | | 1.737558 | -0.73555 | 0.647581 |
| C10orf95 | 3.850485 | 0.198767 | 0.999532 | | 3.255469 | -0.73716 | 0.490301 | | 3.180241 | -0.92175 | 0.444687 |
| MARVELD | 3.869439 | -0.0669 | 0.999532 | | 5.529665 | 0.574408 | 0.490301 | | 4.631663 | 0.662137 | 0.515762 |
| IMPDH1 | 7.527913 | -0.34842 | 0.999532 | | 8.17708 | -0.50622 | 0.490769 | | 6.22388 | -0.13542 | 0.923223 |
| HNRNPU | 28.73261 | 0.151139 | 0.999532 | | 35.09563 | 0.3785 | 0.490837 | | 31.06811 | 0.259666 | 0.722703 |
| MUC13 | 3.143452 | 0.330515 | 0.999532 | | 2.488587 | -0.83498 | 0.490837 | | 2.545617 | -1.14493 | 0.359441 |
| CCR1 | 4.183457 | -0.16489 | 0.999532 | | 4.301475 | -0.63057 | 0.490837 | | 3.357068 | -0.45297 | 0.702463 |
| KDM3B | 4.359227 | 0.338027 | 0.999532 | | 5.466258 | 0.532448 | 0.490837 | | 5.074196 | 0.213297 | 0.858705 |
| TMEM158 | 2.93131 | -0.30698 | 0.999532 | | 3.056246 | -0.72393 | 0.490837 | | 2.22943 | -0.39322 | 0.809485 |
| ZBTB1 | 3.8843 | 0.414405 | 0.999532 | | 4.40204 | 0.59501 | 0.491641 | | 4.431052 | 0.198667 | 0.883116 |
| CIART | 3.083551 | 0.08815 | 0.999532 | | 2.731193 | -0.78825 | 0.491954 | | 2.405217 | -0.85378 | 0.515762 |
| NOC4L | 2.241356 | 0.722472 | 0.999532 | | 2.568004 | 0.762709 | 0.491954 | | 2.776108 | 0.060284 | 0.978282 |
| MYO1G | 2.792 | 0.146886 | 0.999532 | | 2.487382 | -0.81569 | 0.492355 | | 2.148277 | -0.95865 | 0.423641 |
| DHRS3 | 22.34066 | 0.232204 | 0.999532 | | 26.29174 | 0.370912 | 0.492355 | | 24.12558 | 0.163084 | 0.835554 |
| HSP90AB | 24.50749 | 0.021846 | 0.999532 | | 30.85092 | 0.323077 | 0.492355 | | 26.21126 | 0.320598 | 0.619775 |
| TMEM147 | 3.650991 | 0.014355 | 0.999532 | | 4.873415 | 0.591633 | 0.492377 | | 4.303699 | 0.596932 | 0.540342 |
| NIT2 | 1.892432 | 0.949305 | 0.999532 | | 2.007049 | 0.856072 | 0.492587 | | 2.189452 | -0.07602 | 0.974421 |
| WASHC3 | 4.610364 | 0.050598 | 0.999532 | | 6.299649 | 0.522605 | 0.492604 | | 5.225353 | 0.485474 | 0.582673 |
| MAP4K4 | 7.105765 | -0.05403 | 0.999532 | | 9.369676 | 0.436177 | 0.492604 | | 8.230669 | 0.511742 | 0.484718 |
| CXCL8 | 3.85807 | -0.63618 | 0.999532 | | 3.809522 | -0.99694 | 0.492604 | | 2.602989 | -0.31368 | 0.855088 |
| SGSH | 2.627234 | 0.772197 | 0.999532 | | 2.778343 | 0.704493 | 0.492604 | | 3.078651 | -0.04782 | 0.982124 |
| MGST2 | 4.312618 | 0.150866 | 0.999532 | | 5.466263 | 0.52502 | 0.492604 | | 4.830739 | 0.388877 | 0.712621 |
| COPS5 | 2.357261 | -0.00492 | 0.999532 | | 3.337314 | 0.656373 | 0.493173 | | 2.901883 | 0.676813 | 0.560424 |
| SLC1A6 | 3.274548 | -0.13877 | 0.999532 | | 3.248261 | -0.75225 | 0.493193 | | 2.403303 | -0.59459 | 0.659727 |
| KIF12 | 11.25255 | 0.563389 | 0.999532 | | 12.29703 | 0.532883 | 0.493193 | | 12.30448 | -0.01217 | 0.996681 |
| CDC42EP | 12.11442 | -0.68676 | 0.955244 | | 14.64271 | -0.41569 | 0.493194 | | 9.687929 | 0.290752 | 0.722703 |
| PITPNB | 7.054417 | 0.279352 | 0.999532 | | 8.612548 | 0.495376 | 0.493194 | | 7.984905 | 0.235969 | 0.815493 |
| DRGX | 3.354924 | 0.024474 | 0.999532 | | 3.352004 | -0.85624 | 0.493194 | | 2.598358 | -0.79303 | 0.598329 |
| CDK2AP1 | 11.32187 | -0.12829 | 0.999532 | | 15.50356 | 0.4231 | 0.493194 | | 12.73839 | 0.576695 | 0.273545 |
| TXN | 11.26142 | -0.00062 | 0.999532 | | 14.62082 | 0.443281 | 0.493194 | | 12.26943 | 0.47738 | 0.499146 |
| H3C1 | 3.787801 | -0.26351 | 0.999532 | | 3.854968 | -0.67272 | 0.493194 | | 2.909424 | -0.38929 | 0.791466 |
| ATXN7L2 | 3.274654 | -0.04304 | 0.999532 | | 2.8797 | -0.78973 | 0.493194 | | 2.38046 | -0.72688 | 0.60629 |
| S100B | 3.263802 | 0.00371 | 0.999532 | | 3.041851 | -0.70617 | 0.493194 | | 2.58189 | -0.68766 | 0.581287 |
| C7orf31 | 4.206328 | -0.02006 | 0.999532 | | 4.121005 | -0.62202 | 0.493194 | | 3.393213 | -0.57094 | 0.631504 |
| OR9G1 | 2.205227 | -0.02028 | 0.999532 | | 2.051147 | -0.87991 | 0.493194 | | 1.727246 | -0.84066 | 0.579378 |
| MANEAL | 2.509841 | -0.41586 | 0.999532 | | 2.555119 | -0.773 | 0.493194 | | 1.908582 | -0.33936 | 0.858705 |
| CAP1 | 14.3198 | 0.001531 | 0.999532 | | 18.55197 | 0.392778 | 0.493194 | | 15.77836 | 0.4096 | 0.582673 |
| HCFC1R1 | 7.769094 | 0.315002 | 0.999532 | | 9.581894 | 0.433669 | 0.493194 | | 8.841536 | 0.138618 | 0.889493 |
| AEN | 6.2994 | 0.031947 | 0.999532 | | 6.067861 | -0.54125 | 0.493269 | | 5.319182 | -0.5434 | 0.588428 |
| CRYZ | 3.963844 | 0.114695 | 0.999532 | | 5.307732 | 0.632584 | 0.493532 | | 4.619695 | 0.533108 | 0.668481 |
| ADAT2 | 2.872986 | -0.53746 | 0.999532 | | 2.939321 | -0.69687 | 0.49357 | | 2.247089 | -0.14953 | 0.943248 |
| SPSB3 | 3.776108 | 0.53826 | 0.999532 | | 4.302222 | 0.579014 | 0.494154 | | 4.380568 | 0.05921 | 0.972758 |
| NUPR2 | 4.878578 | 0.2128 | 0.999532 | | 4.261099 | -0.64227 | 0.49448 | | 3.83299 | -0.83368 | 0.323635 |
| CA14 | 3.063001 | -0.24694 | 0.999532 | | 3.195915 | -0.78762 | 0.494597 | | 2.241029 | -0.53216 | 0.718643 |
| PCF11 | 5.58882 | -0.78355 | 0.999532 | | 6.682989 | -0.50265 | 0.494597 | | 4.244386 | 0.300913 | 0.803301 |
| FGFBP1 | 2.706735 | -0.25181 | 0.999532 | | 2.534351 | -0.80786 | 0.494597 | | 1.976595 | -0.5311 | 0.735469 |
| GOLIM4 | 18.35891 | 0.519903 | 0.999532 | | 19.56496 | 0.406527 | 0.494659 | | 19.45533 | -0.08312 | 0.934597 |
| GLYR1 | 5.432486 | -0.03957 | 0.999532 | | 7.105128 | 0.508787 | 0.494659 | | 6.061732 | 0.565343 | 0.515762 |
| TMEM100 | 2.910119 | -0.09103 | 0.999532 | | 2.797166 | -0.75174 | 0.494781 | | 2.248078 | -0.64078 | 0.647581 |
| PARP15 | 2.74327 | 0.46831 | 0.999532 | | 1.989105 | -0.89799 | 0.494781 | | 2.078035 | -1.35004 | 0.265037 |
| CCDC167 | 3.681665 | -0.52016 | 0.999532 | | 3.876027 | -0.62354 | 0.494781 | | 2.831264 | -0.09557 | 0.963309 |
| CAMK4 | 3.090542 | -0.18915 | 0.999532 | | 2.92382 | -0.71301 | 0.494781 | | 2.456885 | -0.51436 | 0.724363 |
| TBCD | 3.465194 | 0.661225 | 0.999532 | | 4.274556 | 0.660483 | 0.494781 | | 4.193664 | 0.019326 | 0.994537 |
| RBBP8NL | 2.404191 | -0.17175 | 0.999532 | | 2.366093 | -0.81765 | 0.494781 | | 1.875679 | -0.62518 | 0.697497 |
| OSBPL9 | 5.300355 | 0.116615 | 0.999532 | | 6.782137 | 0.482101 | 0.494781 | | 5.850847 | 0.380918 | 0.672739 |
| OR9Q1 | 2.725344 | -0.18871 | 0.999532 | | 2.667008 | -0.79052 | 0.495027 | | 2.052606 | -0.58153 | 0.701966 |
| NAP1L5 | 3.517325 | 0.004072 | 0.999532 | | 5.306827 | 0.594208 | 0.495027 | | 4.598509 | 0.601201 | 0.549065 |
| HSFX1 | 2.696213 | 0.035092 | 0.999532 | | 2.257664 | -0.80396 | 0.495829 | | 2.177568 | -0.82035 | 0.553283 |
| FCRLA | 2.447431 | -0.23015 | 0.999532 | | 2.441121 | -0.84283 | 0.495854 | | 1.844985 | -0.59093 | 0.723933 |
| OR2T35 | 3.085216 | -0.37388 | 0.999532 | | 3.241147 | -0.70007 | 0.496248 | | 2.125512 | -0.30965 | 0.85621 |
| PRRC2B | 14.02008 | 0.056271 | 0.999532 | | 17.46002 | 0.366034 | 0.496293 | | 15.22073 | 0.330265 | 0.617462 |
| LFNG | 5.045142 | 0.352005 | 0.999532 | | 5.841077 | 0.541038 | 0.496655 | | 5.60972 | 0.212279 | 0.859522 |
| NMB | 2.53868 | 0.080546 | 0.999532 | | 2.079009 | -0.8344 | 0.497498 | | 2.058156 | -0.89878 | 0.521228 |
| SEC61A1 | 7.115969 | -0.11125 | 0.999532 | | 9.598981 | 0.433659 | 0.497801 | | 7.773427 | 0.564128 | 0.415282 |
| RRP12 | 3.156348 | 0.049099 | 0.999532 | | 3.072903 | -0.68747 | 0.498657 | | 2.443794 | -0.70484 | 0.562298 |
| RPL41 | 50.44589 | 0.032801 | 0.999532 | | 62.10222 | 0.279837 | 0.498719 | | 53.24807 | 0.270741 | 0.536289 |
| TM9SF3 | 10.27581 | 0.15764 | 0.999532 | | 12.60229 | 0.398912 | 0.498719 | | 11.31517 | 0.261128 | 0.739496 |
| DDC | 3.412221 | -0.29533 | 0.999532 | | 5.093286 | 0.653559 | 0.498719 | | 4.211182 | 0.91789 | 0.394956 |
| RPS27 | 40.50836 | -0.0212 | 0.999532 | | 50.34709 | 0.291647 | 0.498719 | | 42.84804 | 0.328324 | 0.481527 |
| FGF19 | 2.392319 | -0.45949 | 0.999532 | | 2.516768 | -0.77946 | 0.498719 | | 1.715044 | -0.30153 | 0.876613 |
| BACH2 | 3.303426 | -0.11389 | 0.999532 | | 3.125731 | -0.69221 | 0.498816 | | 2.637603 | -0.54799 | 0.707135 |
| SLC17A5 | 5.772453 | 0.604971 | 0.999532 | | 6.010781 | 0.576219 | 0.498925 | | 6.544529 | -0.01962 | 0.993618 |
| ZNF264 | 3.09116 | 0.575332 | 0.999532 | | 3.475118 | 0.629269 | 0.498925 | | 3.562841 | 0.067798 | 0.972305 |
| ZNF222 | 2.345826 | -0.3282 | 0.999532 | | 2.569686 | -0.80353 | 0.498925 | | 1.87598 | -0.44003 | 0.806394 |
| ZFP90 | 5.778829 | 0.108848 | 0.999532 | | 5.169888 | -0.56606 | 0.500159 | | 4.898142 | -0.65934 | 0.515762 |
| RPA3 | 3.180782 | 0.800516 | 0.999532 | | 3.29332 | 0.658509 | 0.500159 | | 3.622415 | -0.12174 | 0.938429 |
| MED27 | 4.389703 | -0.24225 | 0.999532 | | 4.130181 | -0.63121 | 0.500159 | | 3.355966 | -0.38026 | 0.792539 |
| TRAK2 | 6.187669 | 0.265918 | 0.999532 | | 7.702464 | 0.50826 | 0.500159 | | 6.855333 | 0.259677 | 0.791576 |
| PMVK | 2.428594 | -0.10471 | 0.999532 | | 2.261187 | -0.80726 | 0.500159 | | 1.825834 | -0.68099 | 0.661666 |
| BOC | 5.582473 | 0.444331 | 0.999532 | | 6.546545 | 0.475379 | 0.500159 | | 6.309165 | 0.054655 | 0.972221 |
| ACTR3 | 10.14146 | -0.15085 | 0.999532 | | 13.8084 | 0.44375 | 0.500159 | | 11.3188 | 0.603858 | 0.43637 |
| NEURL1 | 2.587573 | 0.074075 | 0.999532 | | 2.217167 | -0.82121 | 0.500159 | | 1.928956 | -0.87837 | 0.53177 |
| SERPING | 11.55442 | 0.547119 | 0.999532 | | 12.39596 | 0.404347 | 0.500159 | | 12.40052 | -0.12655 | 0.898369 |
| ZNF385C | 2.699589 | 0.117718 | 0.999532 | | 2.277941 | -0.83556 | 0.500159 | | 2.187965 | -0.93934 | 0.505783 |
| TRIM17 | 3.256646 | -0.12355 | 0.999532 | | 3.117724 | -0.67676 | 0.500159 | | 2.611156 | -0.53358 | 0.682165 |
| YTHDC1 | 13.12131 | 0.068309 | 0.999532 | | 16.13464 | 0.324597 | 0.500159 | | 14.04956 | 0.275241 | 0.666906 |
| MTCH1 | 12.96475 | -0.02322 | 0.999532 | | 16.59916 | 0.358187 | 0.500159 | | 13.94259 | 0.399497 | 0.541754 |
| LGALS3B | 13.32758 | 0.259767 | 0.999532 | | 15.77881 | 0.418419 | 0.500159 | | 14.65401 | 0.177877 | 0.837256 |
| BEND3 | 2.333665 | -0.88306 | 0.999532 | | 2.675348 | -0.75917 | 0.500159 | | 1.665123 | 0.141505 | 0.954547 |
| ADM | 6.020289 | -0.06397 | 0.999532 | | 8.299342 | 0.440882 | 0.501652 | | 6.814532 | 0.524607 | 0.522078 |
| RASL11A | 4.929573 | -0.35767 | 0.999532 | | 5.270779 | -0.5275 | 0.50241 | | 4.010865 | -0.1493 | 0.913765 |
| FAU | 20.58164 | -0.10082 | 0.999532 | | 27.12121 | 0.319871 | 0.502573 | | 22.11814 | 0.441053 | 0.446459 |
| MTFP1 | 3.363359 | -0.01973 | 0.999532 | | 2.983847 | -0.71816 | 0.502573 | | 2.739328 | -0.68084 | 0.631504 |
| TCEAL8 | 7.317773 | -0.11512 | 0.999532 | | 9.679665 | 0.459098 | 0.502573 | | 8.450465 | 0.593652 | 0.389965 |
| CHRNB1 | 3.547077 | 0.062671 | 0.999532 | | 3.173106 | -0.7114 | 0.502714 | | 2.704884 | -0.75471 | 0.520848 |
| CCL13 | 2.055173 | -0.14564 | 0.999532 | | 1.960933 | -0.86039 | 0.502714 | | 1.67297 | -0.69338 | 0.680496 |
| TCTN1 | 2.505011 | 0.608179 | 0.999532 | | 2.922401 | 0.69416 | 0.502714 | | 2.965648 | 0.104699 | 0.954304 |
| CXCR6 | 3.228496 | -0.25665 | 0.999532 | | 3.269525 | -0.7119 | 0.502714 | | 2.53098 | -0.4358 | 0.764083 |
| GPNMB | 5.335018 | 0.924821 | 0.547204 | | 5.546856 | 0.625415 | 0.502714 | | 6.310424 | -0.28736 | 0.801572 |
| GCNT7 | 2.375957 | -0.07986 | 0.999532 | | 2.139941 | -0.83981 | 0.502714 | | 1.758394 | -0.74303 | 0.646863 |
| SLC25A4 | 2.299077 | 0.606868 | 0.999532 | | 2.781887 | 0.737689 | 0.502714 | | 2.671503 | 0.144184 | 0.938429 |
| MISP3 | 2.460524 | 0.106558 | 0.999532 | | 2.008404 | -0.83494 | 0.502714 | | 1.920813 | -0.92781 | 0.515762 |
| GPR141 | 2.908233 | 0.351783 | 0.999532 | | 2.318588 | -0.80324 | 0.502714 | | 2.292458 | -1.13325 | 0.323635 |
| TIPRL | 2.984219 | 0.218412 | 0.999532 | | 3.571425 | 0.631203 | 0.502714 | | 3.466204 | 0.41923 | 0.728015 |

| | | | | | | | | | | | | |
|---|---|---|---|---|---|---|---|---|---|---|---|---|
| ZNF516 | 5.126617 | 0.327207 | 0.999532 | | 6.13653 | 0.485222 | 0.502714 | | 5.753198 | 0.176294 | 0.877268 | |
| TTC36 | 2.67965 | -0.51101 | 0.999532 | | 2.766854 | -0.76264 | 0.502714 | | 1.911008 | -0.23392 | 0.905952 | |
| OCIAD2 | 6.249842 | 0.15901 | 0.999532 | | 8.583953 | 0.578313 | 0.502714 | | 7.370202 | 0.470434 | 0.662584 | |
| SLC18A3 | 2.140593 | -0.30898 | 0.999532 | | 2.051666 | -0.83611 | 0.502714 | | 1.593802 | -0.51384 | 0.78258 | |
| GRAPL | 6.704945 | -0.23862 | 0.999532 | | 7.13759 | -0.55871 | 0.502714 | | 5.60358 | -0.28593 | 0.824012 | |
| ZNF649 | 2.142489 | -0.10766 | 0.999532 | | 1.951345 | -0.86746 | 0.502714 | | 1.63966 | -0.7409 | 0.652925 | |
| JAGN1 | 3.364619 | -0.11066 | 0.999532 | | 3.169197 | -0.67964 | 0.502714 | | 2.482374 | -0.54981 | 0.664512 | |
| POGZ | 5.312828 | 0.466869 | 0.999532 | | 5.909187 | 0.534432 | 0.502714 | | 5.651405 | 0.098231 | 0.939913 | |
| CAPS2 | 2.42432 | 0.721848 | 0.999532 | | 2.827442 | 0.720182 | 0.502714 | | 2.825448 | 0.012193 | 0.996891 | |
| RPL24 | 26.56967 | -0.16279 | 0.999532 | | 35.84419 | 0.341504 | 0.502714 | | 28.87383 | 0.529558 | 0.280329 | |
| CDK9 | 10.76667 | -0.18551 | 0.999532 | | 14.50585 | 0.358181 | 0.502714 | | 11.47559 | 0.554794 | 0.359441 | |
| PRCD | 2.648318 | 0.324459 | 0.999532 | | 2.078643 | -0.87449 | 0.502714 | | 2.030372 | -1.18031 | 0.339543 | |
| ATOH8 | 4.135338 | -0.43391 | 0.999532 | | 4.380132 | -0.60564 | 0.502714 | | 3.043269 | -0.15407 | 0.925487 | |
| PERP | 5.62652 | 0.310215 | 0.999532 | | 7.157253 | 0.496215 | 0.502715 | | 6.429235 | 0.197159 | 0.861258 | |
| AMFR | 4.195157 | -0.27833 | 0.999532 | | 6.259198 | 0.548195 | 0.50384 | | 4.811809 | 0.858402 | 0.30001 | |
| ACVR2B | 1.98271 | 0.368743 | 0.999532 | | 2.373029 | 0.762097 | 0.50384 | | 2.30971 | 0.40781 | 0.794815 | |
| SLC24A3 | 2.053806 | -0.0063 | 0.999532 | | 3.499367 | 0.989987 | 0.50384 | | 3.00418 | 0.986646 | 0.515762 | |
| TMEM191 | 3.723058 | -0.1308 | 0.999532 | | 3.521313 | -0.66711 | 0.504188 | | 2.862451 | -0.51619 | 0.686214 | |
| PCMTD1 | 9.015009 | 0.665586 | 0.999532 | | 8.991798 | 0.465912 | 0.504188 | | 9.530233 | -0.19036 | 0.861119 | |
| F5 | 2.008584 | 0.815091 | 0.999532 | | 2.18975 | 0.796346 | 0.50463 | | 2.408854 | -0.00096 | 0.999635 | |
| B4GALNT | 3.337565 | 0.358795 | 0.999532 | | 2.722112 | -0.74904 | 0.50463 | | 2.7904 | -1.09201 | 0.283785 | |
| RIPOR1 | 17.99311 | -0.00298 | 0.999532 | | 23.20295 | 0.381506 | 0.50463 | | 19.87794 | 0.409127 | 0.507289 | |
| KIFC3 | 5.517343 | 0.189306 | 0.999532 | | 6.814573 | 0.470336 | 0.505131 | | 6.196459 | 0.299566 | 0.740755 | |
| CYTIP | 3.725038 | -0.2432 | 0.999532 | | 3.676853 | -0.62465 | 0.505277 | | 2.889002 | -0.36611 | 0.791576 | |
| PCBD1 | 4.753945 | 0.342639 | 0.999532 | | 5.745475 | 0.502747 | 0.505533 | | 5.412629 | 0.18003 | 0.881743 | |
| CHST3 | 3.719775 | 0.663088 | 0.999532 | | 4.049693 | 0.601848 | 0.505533 | | 4.303167 | -0.04204 | 0.982949 | |
| MFAP4 | 4.95079 | 0.518259 | 0.999532 | | 5.811947 | 0.622631 | 0.505713 | | 5.802917 | 0.123381 | 0.940744 | |
| ZNF330 | 3.808357 | -0.00472 | 0.999532 | | 5.673524 | 0.62902 | 0.506119 | | 4.640392 | 0.682441 | 0.524206 | |
| ADH4 | 2.152529 | -0.15293 | 0.999532 | | 1.905092 | -0.87222 | 0.506119 | | 1.640937 | -0.71217 | 0.682485 | |
| ECM2 | 3.397082 | -0.03927 | 0.999532 | | 4.75769 | 0.554074 | 0.506286 | | 3.990905 | 0.609808 | 0.552672 | |
| TRAPPC2 | 4.274589 | -0.45214 | 0.999532 | | 4.696038 | -0.56359 | 0.507109 | | 3.280878 | -0.08964 | 0.960773 | |
| B3GNT4 | 3.393665 | 0.399336 | 0.999532 | | 2.709945 | -0.72063 | 0.507109 | | 2.735699 | -1.10045 | 0.266933 | |
| PSMC2 | 5.275919 | 0.232548 | 0.999532 | | 6.556476 | 0.479839 | 0.508026 | | 5.997989 | 0.2665 | 0.79686 | |
| ILF3 | 15.64505 | 0.015402 | 0.999532 | | 20.33079 | 0.390074 | 0.508026 | | 17.43787 | 0.397293 | 0.560424 | |
| CDC42 | 11.51288 | -0.1122 | 0.999532 | | 15.10603 | 0.353998 | 0.508026 | | 12.44869 | 0.484019 | 0.411291 | |
| SOS2 | 4.43758 | -0.08808 | 0.999532 | | 6.53304 | 0.503033 | 0.508046 | | 5.219519 | 0.616207 | 0.515762 | |
| BCAR1 | 10.55063 | -0.28764 | 0.999532 | | 15.17205 | 0.403863 | 0.508344 | | 12.03524 | 0.715171 | 0.162261 | |
| NEK4 | 2.841555 | 0.622172 | 0.999532 | | 3.609001 | 0.685874 | 0.508344 | | 3.437204 | 0.092599 | 0.962597 | |
| TBCEL | 3.544023 | -0.05894 | 0.999532 | | 4.841605 | 0.543871 | 0.508344 | | 4.282233 | 0.625197 | 0.515762 | |
| PTGIS | 2.510562 | 0.064025 | 0.999532 | | 3.459162 | 0.758559 | 0.508344 | | 3.16849 | 0.701318 | 0.617899 | |
| OR14J1 | 2.795266 | 0.393323 | 0.999532 | | 2.099234 | -0.82752 | 0.508344 | | 2.300546 | -1.18851 | 0.323635 | |
| KRTAP10 | 5.557763 | 0.348988 | 0.999532 | | 4.483119 | -0.66336 | 0.509081 | | 4.558121 | -0.99264 | 0.277362 | |
| MYCBP | 2.953563 | 0.605381 | 0.999532 | | 3.528123 | 0.655688 | 0.509081 | | 3.530844 | 0.072973 | 0.971683 | |
| DOCK5 | 4.281869 | 0.106021 | 0.999532 | | 5.691046 | 0.533928 | 0.509081 | | 4.965256 | 0.442362 | 0.647581 | |
| AGO2 | 3.87158 | 0.215709 | 0.999532 | | 4.983684 | 0.57329 | 0.509081 | | 4.430448 | 0.374637 | 0.740093 | |
| IDH2 | 4.078257 | 0.229822 | 0.999532 | | 5.459061 | 0.578235 | 0.509081 | | 4.709977 | 0.371713 | 0.812053 | |
| CCT5 | 6.30883 | 0.118101 | 0.999532 | | 8.040325 | 0.460022 | 0.509081 | | 7.25604 | 0.360541 | 0.660958 | |
| LYG1 | 2.492767 | -0.0523 | 0.999532 | | 2.330522 | -0.79098 | 0.509081 | | 1.914332 | -0.71815 | 0.636913 | |
| DNAH8 | 2.611473 | -0.16305 | 0.999532 | | 2.614951 | -0.75194 | 0.509081 | | 2.100388 | -0.56234 | 0.716171 | |
| TMEM245 | 5.056812 | 0.251424 | 0.999532 | | 6.196434 | 0.503331 | 0.509081 | | 5.630747 | 0.27032 | 0.806797 | |
| KNOP1 | 3.587093 | 0.027409 | 0.999532 | | 3.306525 | -0.67432 | 0.509081 | | 2.681331 | -0.6808 | 0.55841 | |
| RASGRF1 | 2.530977 | -0.62368 | 0.999532 | | 2.893862 | -0.7082 | 0.509081 | | 1.853583 | -0.0548 | 0.985693 | |
| CEACAM | 4.078627 | 0.03847 | 0.999532 | | 3.905093 | -0.67155 | 0.509081 | | 3.387259 | -0.68948 | 0.566812 | |
| LANCL1 | 3.339132 | -0.04526 | 0.999532 | | 4.750147 | 0.556652 | 0.509081 | | 3.961764 | 0.619319 | 0.532137 | |
| ADD2 | 2.825544 | 0.241571 | 0.999532 | | 2.292622 | -0.78946 | 0.509081 | | 2.235956 | -1.01158 | 0.414073 | |
| ELMOD2 | 3.3314 | 0.238372 | 0.999532 | | 4.164873 | 0.575488 | 0.509081 | | 3.916134 | 0.356711 | 0.757835 | |
| EYA1 | 2.250252 | -0.30971 | 0.999532 | | 2.356441 | -0.85247 | 0.509081 | | 1.734573 | -0.51647 | 0.783005 | |
| SPART | 4.591731 | 0.11824 | 0.999532 | | 6.118916 | 0.554982 | 0.509081 | | 5.330483 | 0.468528 | 0.660958 | |
| KLHDC2 | 5.874532 | 0.654326 | 0.999532 | | 5.785602 | 0.490584 | 0.50941 | | 6.320524 | -0.14813 | 0.892869 | |
| EXOSC2 | 2.187992 | 0.228057 | 0.999532 | | 2.793481 | 0.702109 | 0.50988 | | 2.644133 | 0.492808 | 0.723294 | |
| BMERB1 | 42.12505 | 0.303381 | 0.999532 | | 37.92119 | -0.37421 | 0.510063 | | 36.38303 | -0.65688 | 0.107433 | |
| NPR1 | 10.41791 | -0.11854 | 0.999532 | | 14.09812 | 0.350539 | 0.510481 | | 11.42046 | 0.489589 | 0.400624 | |
| KLHL25 | 2.745213 | -0.32797 | 0.999532 | | 2.90952 | -0.75431 | 0.510547 | | 2.19179 | -0.40308 | 0.816112 | |
| SLC25A4 | 6.149047 | 0.398731 | 0.999532 | | 7.1223 | 0.495628 | 0.510674 | | 7.089 | 0.116956 | 0.923864 | |
| PPP1R27 | 2.143776 | -0.02439 | 0.999532 | | 2.001613 | -0.8287 | 0.511238 | | 1.652599 | -0.7827 | 0.620976 | |
| LIX1 | 3.301041 | 0.86763 | 0.999532 | | 3.038756 | 0.651171 | 0.511238 | | 3.687069 | -0.19613 | 0.892869 | |
| SDHC | 22.38703 | 0.071788 | 0.999532 | | 27.43027 | 0.312642 | 0.511344 | | 23.95408 | 0.260909 | 0.647218 | |
| CNN3 | 21.26726 | -0.14761 | 0.999532 | | 28.44749 | 0.355123 | 0.511601 | | 23.13511 | 0.524285 | 0.333577 | |
| NHLRC3 | 3.572012 | 0.206394 | 0.999532 | | 3.14479 | -0.67692 | 0.511601 | | 2.757386 | -0.85917 | 0.445661 | |
| TAOK2 | 2.786227 | 0.260853 | 0.999532 | | 3.753913 | 0.657545 | 0.511601 | | 3.415598 | 0.412427 | 0.739189 | |
| MMP13 | 2.505212 | -0.10637 | 0.999532 | | 2.318563 | -0.79734 | 0.511601 | | 1.880628 | -0.67277 | 0.657538 | |
| HDAC2 | 3.376785 | 0.533696 | 0.999532 | | 3.77792 | 0.621427 | 0.511601 | | 3.715401 | 0.104255 | 0.948896 | |
| NAV1 | 2.278694 | -1.03228 | 0.999532 | | 2.710916 | -0.72013 | 0.511601 | | 1.680555 | 0.330606 | 0.869378 | |
| KCTD13 | 4.160015 | -0.41062 | 0.999532 | | 4.496517 | -0.56571 | 0.511601 | | 3.362215 | -0.13614 | 0.931942 | |
| RNH1 | 8.287593 | -0.1237 | 0.999532 | | 11.14543 | 0.395796 | 0.511846 | | 9.043454 | 0.534669 | 0.469205 | |
| SSR2 | 4.483836 | -0.03099 | 0.999532 | | 6.081189 | 0.499142 | 0.511846 | | 4.979707 | 0.548587 | 0.552672 | |
| RERGL | 2.668417 | 0.15791 | 0.999532 | | 2.490511 | -0.83938 | 0.512384 | | 2.190746 | -0.98948 | 0.451774 | |
| ZDBF2 | 3.78829 | -0.25766 | 0.999532 | | 3.833428 | -0.60326 | 0.512384 | | 2.890792 | -0.32481 | 0.816618 | |
| MTA2 | 6.610992 | -0.13934 | 0.999532 | | 6.66979 | -0.46508 | 0.512646 | | 5.322145 | -0.308 | 0.753585 | |
| ZNF789 | 2.771944 | -0.21485 | 0.999532 | | 2.825244 | -0.71718 | 0.512646 | | 2.100343 | -0.47367 | 0.756957 | |
| BCAS4 | 4.836863 | 0.062375 | 0.999532 | | 4.562201 | -0.60562 | 0.512672 | | 3.730158 | -0.64637 | 0.515762 | |
| ILVBL | 3.641864 | 0.221796 | 0.999532 | | 4.80796 | 0.544237 | 0.512775 | | 4.234158 | 0.337646 | 0.757037 | |
| CCER1 | 2.0943 | -0.64869 | 0.999532 | | 2.206286 | -0.79867 | 0.512775 | | 1.638514 | -0.13149 | 0.960773 | |
| PIGS | 6.961705 | 0.004716 | 0.999532 | | 8.966585 | 0.41114 | 0.512775 | | 7.697388 | 0.424021 | 0.570037 | |
| RGPD2 | 3.330284 | -0.28005 | 0.999532 | | 3.42595 | -0.67738 | 0.513034 | | 2.712248 | -0.3828 | 0.797739 | |
| CPO | 3.604429 | -0.317 | 0.999532 | | 3.658158 | -0.63228 | 0.513676 | | 2.829272 | -0.29518 | 0.850928 | |
| TMPRSS1 | 2.227708 | -0.34836 | 0.999532 | | 2.260973 | -0.82052 | 0.51428 | | 1.621309 | -0.45303 | 0.809485 | |
| MAP2K2 | 8.076673 | -0.07104 | 0.999532 | | 8.009227 | -0.46266 | 0.514723 | | 6.754785 | -0.37125 | 0.668553 | |
| TANC1 | 8.60511 | 0.586974 | 0.999532 | | 8.84672 | 0.432875 | 0.514997 | | 9.298345 | -0.14086 | 0.896374 | |
| ZNF595 | 2.953588 | -0.0499 | 0.999532 | | 2.741471 | -0.69795 | 0.514997 | | 2.309968 | -0.6296 | 0.644433 | |
| RPS24 | 35.1908 | 0.111737 | 0.999532 | | 43.51048 | 0.334011 | 0.516585 | | 37.82255 | 0.257251 | 0.722703 | |
| FAM228A | 2.185377 | -0.32122 | 0.999532 | | 2.090351 | -0.82976 | 0.516585 | | 1.617846 | -0.4915 | 0.792539 | |
| ZNF483 | 3.786904 | -0.33687 | 0.999532 | | 3.729975 | -0.60155 | 0.516585 | | 3.096352 | -0.25623 | 0.873155 | |
| PLPP5 | 3.030447 | 0.150315 | 0.999532 | | 2.608675 | -0.72493 | 0.516585 | | 2.368092 | -0.85707 | 0.475974 | |
| RNF152 | 5.267579 | 0.105994 | 0.999532 | | 7.018047 | 0.521907 | 0.517655 | | 6.163597 | 0.440539 | 0.718633 | |
| ZNF320 | 1.819161 | 0.438993 | 0.999532 | | 2.165898 | 0.829162 | 0.517705 | | 2.21828 | 0.39308 | 0.827429 | |
| NUP62CL | 3.571862 | 0.121683 | 0.999532 | | 3.062972 | -0.70515 | 0.517705 | | 2.749474 | -0.80903 | 0.49299 | |
| POLR2E | 14.80597 | 0.273277 | 0.999532 | | 16.54396 | 0.331956 | 0.517705 | | 15.73984 | 0.087146 | 0.909259 | |
| DND1 | 3.847874 | -0.27033 | 0.999532 | | 4.152395 | -0.65324 | 0.517793 | | 3.022632 | -0.34736 | 0.814402 | |
| FAAP24 | 2.713341 | -0.0616 | 0.999532 | | 2.527862 | -0.77572 | 0.517793 | | 2.187756 | -0.69194 | 0.656224 | |
| SPAG1 | 2.664496 | 0.258659 | 0.999532 | | 2.208168 | -0.80949 | 0.517793 | | 2.182052 | -1.04846 | 0.415282 | |
| COL11A2 | 3.502567 | -0.10531 | 0.999532 | | 3.557784 | -0.67168 | 0.517793 | | 2.845689 | -0.54333 | 0.673659 | |
| SLC2A12 | 2.981963 | 0.36665 | 0.999532 | | 3.929862 | 0.625246 | 0.517793 | | 3.453633 | 0.291262 | 0.837256 | |
| STRAP | 5.635977 | -0.16271 | 0.999532 | | 8.066607 | 0.437784 | 0.517793 | | 6.391832 | 0.620646 | 0.446558 | |
| MAMDC2 | 2.042327 | 0.363822 | 0.999532 | | 2.683306 | 0.728234 | 0.518901 | | 2.589884 | 0.385904 | 0.801905 | |
| IRF2BP1 | 3.092176 | 0.673843 | 0.999532 | | 3.494499 | 0.634 | 0.520526 | | 3.747479 | -0.02107 | 0.993618 | |
| ECH1 | 8.108854 | 0.298371 | 0.999532 | | 9.687463 | 0.413398 | 0.520526 | | 8.657349 | 0.133559 | 0.90424 | |
| PRIMA1 | 4.884775 | -0.17291 | 0.999532 | | 5.062982 | -0.58596 | 0.520526 | | 4.00705 | -0.38972 | 0.752155 | |
| NPAP1L | 33.73714 | 0.308645 | 0.999532 | | 29.93092 | -0.40955 | 0.520526 | | 28.93684 | -0.69875 | 0.15668 | |
| UBE2D3 | 8.516272 | -0.19597 | 0.999532 | | 12.10893 | 0.388489 | 0.520526 | | 9.62578 | 0.607589 | 0.315376 | |
| PROSER2 | 3.305851 | -0.39953 | 0.999532 | | 3.456627 | -0.64442 | 0.520526 | | 2.465958 | -0.22398 | 0.894612 | |
| DIPK1A | 2.592448 | 0.104166 | 0.999532 | | 2.310088 | -0.76902 | 0.520526 | | 2.032356 | -0.85268 | 0.53177 | |
| FECH | 2.865483 | 0.128871 | 0.999532 | | 4.137437 | 0.57132 | 0.520526 | | 3.470045 | 0.458465 | 0.682165 | |
| CBFA2T2 | 5.243958 | 0.4055 | 0.999532 | | 6.325712 | 0.521229 | 0.520526 | | 6.050745 | 0.14511 | 0.910218 | |
| SIAH3 | 2.470991 | 0.248761 | 0.999532 | | 2.145892 | -0.83164 | 0.520526 | | 1.926581 | -1.05973 | 0.413122 | |
| UBL7 | 2.74449 | 0.171409 | 0.999532 | | 3.679353 | 0.617355 | 0.520526 | | 3.237772 | 0.464239 | 0.699971 | |
| TBC1D4 | 3.229744 | -0.00261 | 0.999532 | | 4.634587 | 0.555558 | 0.520526 | | 3.986274 | 0.577401 | 0.566812 | |
| UBAP2L | 5.630313 | -0.25562 | 0.999532 | | 7.969086 | 0.475293 | 0.520526 | | 6.276422 | 0.745198 | 0.37216 | |
| MPLKIP | 3.327939 | 0.023141 | 0.999532 | | 2.928442 | -0.72416 | 0.520526 | | 2.59457 | -0.73232 | 0.53691 | |
| CCDC68 | 2.443651 | -0.28194 | 0.999532 | | 2.492818 | -0.7373 | 0.520526 | | 1.890965 | -0.43544 | 0.796434 | |
| NOP16 | 4.346209 | -0.09604 | 0.999532 | | 3.935468 | -0.59511 | 0.520526 | | 3.545226 | -0.48769 | 0.706407 | |
| BBOX1 | 4.00628 | 0.214412 | 0.999532 | | 6.299497 | 1.023965 | 0.520526 | | 5.701342 | 0.81298 | 0.647581 | |
| KCTD20 | 3.834528 | 0.333255 | 0.999532 | | 4.933229 | 0.612765 | 0.520526 | | 4.618889 | 0.287264 | 0.796512 | |
| SMYD3 | 1.693783 | 0.364276 | 0.999532 | | 2.120479 | 0.801522 | 0.520526 | | 2.06451 | 0.449865 | 0.791576 | |
| HES7 | 10.34472 | -0.10948 | 0.999532 | | 10.13075 | -0.58508 | 0.520937 | | 8.58569 | -0.47372 | 0.702463 | |
| H2BC11 | 4.283234 | -0.531 | 0.999532 | | 7.079655 | 0.507482 | 0.520937 | | 5.16076 | 1.066026 | 0.196145 | |
| OR4A15 | 2.059049 | -0.20141 | 0.999532 | | 1.963073 | -0.8644 | 0.520937 | | 1.568165 | -0.64203 | 0.720106 | |
| TUB | 2.395816 | 0.225066 | 0.999532 | | 3.252772 | 0.673896 | 0.520937 | | 2.930114 | 0.470641 | 0.722703 | |
| GNG11 | 17.30728 | -0.21747 | 0.999532 | | 25.04016 | 0.482271 | 0.520937 | | 20.14291 | 0.721673 | 0.256218 | |
| ADCY3 | 5.036029 | -0.8282 | 0.999532 | | 5.9681 | -0.49496 | 0.520937 | | 3.885059 | 0.339752 | 0.784329 | |
| PAX2 | 4.682764 | 0.67124 | 0.999532 | | 5.060928 | 0.55926 | 0.520937 | | 5.190216 | -0.09374 | 0.948097 | |
| RPL7L1 | 20.80561 | 0.010076 | 0.999532 | | 20.72754 | -0.32843 | 0.520937 | | 17.95884 | -0.31831 | 0.595502 | |
| TMEM40 | 2.515492 | -0.13501 | 0.999532 | | 2.404786 | -0.75968 | 0.520937 | | 1.95292 | -0.60498 | 0.697806 | |
| CCDC200 | 2.466351 | 0.361862 | 0.999532 | | 1.958652 | -0.85546 | 0.520937 | | 1.990909 | -1.19601 | 0.344878 | |
| C19orf54 | 4.10372 | -0.28829 | 0.999532 | | 4.206743 | -0.60801 | 0.520937 | | 3.149221 | -0.30116 | 0.835554 | |
| H2BC5 | 11.43231 | -0.28967 | 0.999532 | | 16.24216 | 0.342667 | 0.520937 | | 12.58993 | 0.654424 | 0.266933 | |
| TIMM17A | 3.510719 | 0.592027 | 0.999532 | | 3.872034 | 0.592092 | 0.520937 | | 3.973794 | 0.018835 | 0.994192 | |
| TMEM204 | 3.265635 | -0.5426 | 0.999532 | | 4.932117 | 0.529674 | 0.521025 | | 3.752491 | 1.085293 | 0.196145 | |
| C1orf159 | 4.007355 | -0.06263 | 0.999532 | | 3.842578 | -0.63139 | 0.521248 | | 3.274932 | -0.55228 | 0.657467 | |
| PRKAG3 | 2.315598 | -0.15515 | 0.999532 | | 2.313158 | -0.84427 | 0.522518 | | 1.723253 | -0.66779 | 0.686214 | |
| COPG1 | 3.948659 | -0.15044 | 0.999532 | | 5.665768 | 0.490912 | 0.522728 | | 4.511296 | 0.658842 | 0.454116 | |
| SMG7 | 5.346944 | 0.060573 | 0.999532 | | 6.763625 | 0.458608 | 0.523629 | | 5.972035 | 0.414987 | 0.645127 | |
| TM9SF2 | 15.63991 | 0.190971 | 0.999532 | | 19.48004 | 0.392626 | 0.52387 | | 17.45058 | 0.228733 | 0.766974 | |
| HSFX3 | 2.321977 | 0.278357 | 0.999532 | | 1.947606 | -0.83224 | 0.52387 | | 1.935963 | -1.08774 | 0.412558 | |
| TCHH | 2.3687 | 0.2288 | 0.999532 | | 1.905464 | -0.85876 | 0.52396 | | 1.888671 | -1.07138 | 0.442315 | |
| PLK1 | 2.627267 | -0.23386 | 0.999532 | | 2.593971 | -0.73301 | 0.524026 | | 1.928359 | -0.48215 | 0.760473 | |
| MYO3A | 2.777812 | 0.281646 | 0.999532 | | 3.266057 | 0.643784 | 0.524026 | | 3.21642 | 0.354251 | 0.808571 | |
| MUC4 | 2.952821 | -0.29182 | 0.999532 | | 3.092011 | -0.71364 | 0.524026 | | 2.426348 | -0.39571 | 0.816618 | |
| CYB561 | 3.026874 | 0.368874 | 0.999532 | | 3.754229 | 0.591528 | 0.524026 | | 3.572308 | 0.241482 | 0.861656 | |
| ALKBH2 | 2.619502 | 0.617179 | 0.999532 | | 2.977027 | 0.665036 | 0.524026 | | 3.028719 | 0.066444 | 0.974421 | |
| NOTCH2 | 4.338884 | 0.299751 | 0.999532 | | 5.245397 | 0.544646 | 0.524026 | | 4.861599 | 0.264926 | 0.825415 | |
| E2F2 | 2.607789 | 0.320335 | 0.999532 | | 2.085431 | -0.8066 | 0.52448 | | 2.153105 | -1.11042 | 0.359441 | |
| DYNLT1 | 5.886248 | 0.091259 | 0.999532 | | 7.44065 | 0.456176 | 0.5246 | | 6.58837 | 0.388518 | 0.647218 | |
| PLEKHD1 | 2.277692 | 0.170553 | 0.999532 | | 1.924794 | -0.85716 | 0.524637 | | 1.824561 | -1.00809 | 0.49299 | |
| CNIH1 | 4.66543 | -0.05862 | 0.999532 | | 6.623799 | 0.504675 | 0.524637 | | 5.44463 | 0.579082 | 0.566812 | |
| AKT2 | 7.106224 | 0.307152 | 0.999532 | | 8.656642 | 0.437039 | 0.524637 | | 7.772369 | 0.145506 | 0.892869 | |
| RPL13A | 10.60684 | -0.1255 | 0.999532 | | 14.22671 | 0.390033 | 0.525542 | | 11.52398 | 0.535469 | 0.451915 | |
| SPTBN1 | 64.09609 | 0.056722 | 0.999532 | | 79.60074 | 0.33563 | 0.525542 | | 69.2658 | 0.302081 | 0.619423 | |
| ARHGAP | 10.057 | -0.02021 | 0.999532 | | 12.83743 | 0.375652 | 0.525971 | | 11.12254 | 0.419442 | 0.541754 | |
| KRTAP23 | 2.379967 | -0.30437 | 0.999532 | | 2.338839 | -0.79116 | 0.525971 | | 1.70595 | -0.46775 | 0.794049 | |
| NPRL3 | 6.462959 | 0.384841 | 0.999532 | | 5.084216 | -0.52569 | 0.525971 | | 5.090286 | -0.88759 | 0.203949 | |
| ELP1 | 2.599688 | -0.30442 | 0.999532 | | 4.129555 | 0.588961 | 0.525971 | | 3.147714 | 0.9114 | 0.352256 | |
| VDR | 3.802011 | -0.24043 | 0.999532 | | 3.723079 | -0.62691 | 0.525971 | | 2.784088 | -0.37583 | 0.794815 | |
| RCOR3 | 3.499294 | 0.434796 | 0.999532 | | 3.968795 | 0.602246 | 0.525971 | | 4.035325 | 0.200257 | 0.886495 | |
| SMC5 | 3.181736 | -0.5413 | 0.999532 | | 3.522624 | -0.62984 | 0.525971 | | 2.419091 | -0.06825 | 0.976441 | |
| H2AC6 | 11.48003 | -0.09598 | 0.999532 | | 15.0543 | 0.361229 | 0.525971 | | 12.35729 | 0.47477 | 0.455196 | |
| ARNT | 4.920916 | -0.09187 | 0.999532 | | 6.965709 | 0.489459 | 0.525971 | | 5.817011 | 0.603194 | 0.49299 | |
| EXOC3 | 4.968724 | 0.431968 | 0.999532 | | 5.83751 | 0.514367 | 0.525971 | | 5.572161 | 0.105305 | 0.937656 | |
| SOCS7 | 3.022731 | 0.217041 | 0.999532 | | 3.837311 | 0.597737 | 0.525971 | | 3.615318 | 0.402159 | 0.736481 | |
| NDUFV1 | 4.821878 | 0.111312 | 0.999532 | | 6.095105 | 0.468863 | 0.525971 | | 5.234126 | 0.372041 | 0.703463 | |
| ZBTB4 | 2.472002 | 0.354284 | 0.999532 | | 3.186294 | 0.677015 | 0.525971 | | 2.998203 | 0.341985 | 0.810121 | |
| LAMB2 | 16.06834 | -0.14907 | 0.999532 | | 21.99379 | 0.345552 | 0.525971 | | 17.65888 | 0.52471 | 0.423641 | |
| UNC50 | 4.758722 | 0.28155 | 0.999532 | | 5.864678 | 0.50706 | 0.525971 | | 5.220251 | 0.244321 | 0.820226 | |
| GBA | 5.047218 | 0.119086 | 0.999532 | | 6.105691 | 0.464364 | 0.525971 | | 5.607595 | 0.359876 | 0.705425 | |
| MED23 | 3.895519 | 0.67372 | 0.999532 | | 4.093834 | 0.591854 | 0.525971 | | 4.2829 | -0.06128 | 0.972221 | |
| HSD17B1 | 10.37123 | -0.02108 | 0.999532 | | 13.73096 | 0.412236 | 0.526033 | | 11.4349 | 0.457329 | 0.49299 | |
| VAMP7 | 1.957927 | -0.27518 | 0.999532 | | 2.943523 | 0.665267 | 0.526033 | | 2.448963 | 0.956024 | 0.428819 | |
| TAGLN2 | 19.68816 | -0.34162 | 0.999532 | | 28.7432 | 0.432918 | 0.527307 | | 22.5065 | 0.786144 | 0.165207 | |
| ENPP4 | 3.28567 | 0.436002 | 0.999532 | | 3.910896 | 0.600923 | 0.527307 | | 3.864079 | 0.188551 | 0.894612 | |
| NCR3LG1 | 2.698394 | -0.35847 | 0.999532 | | 2.728963 | -0.75182 | 0.527307 | | 2.021794 | -0.3725 | 0.836935 | |
| MIDN | 7.135837 | -0.47147 | 0.999532 | | 8.279306 | -0.45102 | 0.527307 | | 5.750359 | 0.032039 | 0.985693 | |
| TMSB4X | 221.7184 | -0.07467 | 0.999532 | | 290.6571 | 0.352067 | 0.527307 | | 242.5915 | 0.451843 | 0.247358 | |
| PPIE | 3.526507 | 0.063597 | 0.999532 | | 4.764453 | 0.522205 | 0.527307 | | 4.115546 | 0.477161 | 0.64694 | |
| ZNF583 | 1.974262 | 0.329377 | 0.999532 | | 2.612079 | 0.716831 | 0.527307 | | 2.451391 | 0.409539 | 0.791466 | |
| TRIM8 | 6.589571 | 0.213252 | 0.999532 | | 8.244025 | 0.473059 | 0.527307 | | 7.28866 | 0.278703 | 0.757035 | |
| UBE3C | 2.363014 | 0.165769 | 0.999532 | | 3.008972 | 0.640678 | 0.527307 | | 2.740597 | 0.490521 | 0.700565 | |
| TDG | 5.333556 | -0.37209 | 0.999532 | | 5.725488 | -0.49235 | 0.527307 | | 4.458082 | -0.10247 | 0.946645 | |
| ARHGEF | 7.01763 | 0.314019 | 0.999532 | | 5.890675 | -0.54612 | 0.527307 | | 5.989249 | -0.84659 | 0.340167 | |
| IGSF5 | 2.060146 | -0.4435 | 0.999532 | | 2.026864 | -0.83255 | 0.527834 | | 1.586502 | -0.37696 | 0.862823 | |
| BORCS8 | 3.250977 | -0.06794 | 0.999532 | | 2.99246 | -0.68523 | 0.528846 | | 2.532912 | -0.60178 | 0.660958 | |
| FANCD2 | 3.699206 | -0.05758 | 0.999532 | | 3.286366 | -0.67365 | 0.528846 | | 2.728698 | -0.59417 | 0.647581 | |
| OR2A1 | 6.06562 | 0.162138 | 0.999532 | | 5.465197 | -0.52408 | 0.528985 | | 5.006648 | -0.66608 | 0.451774 | |
| ILDR2 | 2.881824 | 0.684679 | 0.999532 | | 3.003324 | 0.648878 | 0.529583 | | 3.220482 | -0.02195 | 0.993618 | |
| ATP5MG | 2.738589 | -0.12159 | 0.999532 | | 2.690018 | -0.72555 | 0.529583 | | 2.148909 | -0.58093 | 0.697497 | |
| CHPF | 4.097867 | 0.143503 | 0.999532 | | 5.518194 | 0.494681 | 0.529738 | | 4.640798 | 0.370557 | 0.718633 | |
| USP44 | 2.923536 | -0.09301 | 0.999532 | | 2.723068 | -0.72923 | 0.529913 | | 2.36704 | -0.62531 | 0.68597 | |
| C9orf153 | 2.689439 | -0.03965 | 0.999532 | | 2.4237 | -0.79441 | 0.529913 | | 2.073671 | -0.73746 | 0.625223 | |
| TMEM37 | 9.210369 | 0.422214 | 0.999532 | | 10.01946 | 0.391063 | 0.529913 | | 9.795351 | -0.02601 | 0.988853 | |
| ARL16 | 3.558205 | 0.601179 | 0.999532 | | 3.912812 | 0.611106 | 0.529913 | | 4.064159 | 0.03222 | 0.989148 | |
| HMGA2 | 4.455083 | 0.145488 | 0.999532 | | 4.138395 | -0.62678 | 0.529913 | | 3.661537 | -0.75145 | 0.467412 | |
| TRAPPC1 | 7.096469 | 0.51983 | 0.999532 | | 7.095976 | 0.444013 | 0.529913 | | 7.699078 | -0.04884 | 0.972758 | |
| SLFN11 | 4.672732 | 0.063553 | 0.999532 | | 6.094832 | 0.538804 | 0.529913 | | 5.554236 | 0.478348 | 0.661217 | |
| HSPA12A | 4.669765 | 0.748588 | 0.999532 | | 4.598478 | 0.532893 | 0.529913 | | 4.903285 | -0.20035 | 0.865925 | |
| PLG | 11.52828 | 0.345848 | 0.999532 | | 12.78532 | 0.407126 | 0.529913 | | 12.29087 | 0.061194 | 0.965876 | |
| COA8 | 5.345706 | 0.561639 | 0.999532 | | 6.017616 | 0.510712 | 0.529913 | | 6.036475 | -0.03234 | 0.985873 | |
| COL18A1 | 7.487992 | -0.53633 | 0.999532 | | 11.43167 | 0.438306 | 0.529913 | | 8.570684 | 0.989546 | 0.075829 | UP |
| PPID | 3.059584 | -0.036 | 0.999532 | | 4.186979 | 0.559141 | 0.529913 | | 3.518803 | 0.612416 | 0.552672 | |
| ANPEP | 7.153338 | 0.194897 | 0.999532 | | 8.957719 | 0.578419 | 0.529913 | | 8.196679 | 0.379735 | 0.749588 | |
| LTBP3 | 7.252613 | 0.143207 | 0.999532 | | 9.465858 | 0.499241 | 0.529913 | | 8.199086 | 0.394575 | 0.649304 | |
| PLA2G3 | 4.473228 | -0.01021 | 0.999532 | | 4.167557 | -0.62329 | 0.529913 | | 3.799231 | -0.59963 | 0.630705 | |
| DNAJB4 | 3.18789 | -0.41912 | 0.999532 | | 4.744982 | 0.545311 | 0.529913 | | 3.659312 | 0.978987 | 0.272442 | |
| MANF | 3.763052 | 0.070638 | 0.999532 | | 4.714677 | 0.52883 | 0.529913 | | 4.266388 | 0.482761 | 0.635381 | |
| PPP2R5A | 6.140952 | 0.399884 | 0.999532 | | 6.906315 | 0.490438 | 0.529913 | | 6.653332 | 0.120863 | 0.915888 | |
| SLC25A3 | 2.084945 | 0.046677 | 0.999532 | | 1.878435 | -0.8869 | 0.529913 | | 1.558094 | -0.9142 | 0.560424 | |
| PTMA | 135.9764 | -0.07615 | 0.999532 | | 178.2156 | 0.351488 | 0.529913 | | 148.5032 | 0.452743 | 0.389066 | |
| TMEM171 | 3.146437 | 0.098826 | 0.999532 | | 3.04341 | -0.72468 | 0.529913 | | 2.699896 | -0.79156 | 0.541136 | |
| ARHGAP | 2.703293 | 0.482261 | 0.999532 | | 3.126982 | 0.628896 | 0.529913 | | 3.153064 | 0.16448 | 0.914692 | |
| COL5A1 | 2.231538 | 0.206435 | 0.999532 | | 3.014436 | 0.647775 | 0.529913 | | 2.688627 | 0.45687 | 0.722703 | |
| FBLN1 | 4.116421 | 0.117644 | 0.999532 | | 5.224451 | 0.545604 | 0.530031 | | 4.813564 | 0.454804 | 0.705888 | |
| MYBBP1A | 3.485898 | -0.39365 | 0.999532 | | 3.835159 | -0.63239 | 0.530107 | | 2.69852 | -0.22276 | 0.889493 | |
| RARG | 3.887571 | 0.707007 | 0.999532 | | 3.893409 | 0.584758 | 0.530387 | | 4.288176 | -0.12309 | 0.939107 | |
| SCRN2 | 3.058651 | 0.377345 | 0.999532 | | 3.564023 | 0.606493 | 0.530387 | | 3.471604 | 0.247548 | 0.86209 | |
| MYOCD | 2.81694 | -0.69555 | 0.999532 | | 3.235044 | -0.67978 | 0.530387 | | 2.127222 | 0.041092 | 0.989985 | |
| GTF2H5 | 2.352564 | 0.050784 | 0.999532 | | 2.12178 | -0.81327 | 0.530387 | | 1.903102 | -0.8384 | 0.580033 | |
| CNOT1 | 3.237189 | -0.04131 | 0.999532 | | 4.718456 | 0.543193 | 0.530387 | | 3.886105 | 0.606622 | 0.55841 | |
| CDX2 | 3.191766 | 0.012899 | 0.999532 | | 3.013788 | -0.67444 | 0.530387 | | 2.503862 | -0.66753 | 0.599002 | |
| IFNA14 | 3.031601 | -0.52357 | 0.999532 | | 3.297095 | -0.6582 | 0.530387 | | 2.366915 | -0.1135 | 0.959593 | |
| ANKRD1 | 2.355031 | 0.009012 | 0.999532 | | 2.0297 | -0.80822 | 0.530387 | | 1.888756 | -0.80193 | 0.602786 | |
| OR1K1 | 3.940882 | -0.07762 | 0.999532 | | 3.822301 | -0.60728 | 0.530387 | | 3.059677 | -0.50953 | 0.679818 | |
| BAG5 | 6.339179 | 0.065228 | 0.999532 | | 8.125012 | 0.417019 | 0.530387 | | 7.025101 | 0.372327 | 0.668566 | |
| C5orf38 | 3.0121 | 0.162371 | 0.999532 | | 2.59559 | -0.73129 | 0.530387 | | 2.441256 | -0.87632 | 0.508751 | |
| MRPS18A | 6.026059 | 0.111212 | 0.999532 | | 7.773684 | 0.435158 | 0.530387 | | 6.766275 | 0.345594 | 0.697497 | |
| CXCR3 | 3.801538 | -0.40852 | 0.999532 | | 4.113541 | -0.67506 | 0.530387 | | 3.002688 | -0.25037 | 0.881316 | |
| NR5A1 | 2.759314 | 0.50435 | 0.999532 | | 2.105394 | -0.80766 | 0.530387 | | 2.171604 | -1.29111 | 0.24893 | |
| GPATCH3 | 3.796963 | 0.028812 | 0.999532 | | 3.403742 | -0.63981 | 0.530387 | | 2.94527 | -0.64908 | 0.566812 | |
| C10orf82 | 2.155051 | 1.017824 | 0.999532 | | 2.076445 | 0.806748 | 0.530387 | | 2.529307 | -0.19501 | 0.914687 | |
| TXNDC15 | 5.222964 | 0.016177 | 0.999532 | | 6.88265 | 0.480534 | 0.530387 | | 6.014317 | 0.481498 | 0.570037 | |
| HEXA | 4.911887 | 0.220037 | 0.999532 | | 5.993701 | 0.467894 | 0.530387 | | 5.573267 | 0.26545 | 0.791576 | |
| OST4 | 12.62653 | -0.29928 | 0.999532 | | 17.06158 | 0.340051 | 0.530387 | | 13.28538 | 0.653338 | 0.203949 | |
| SRRD | 4.805714 | -0.10954 | 0.999532 | | 4.640993 | -0.57382 | 0.530387 | | 3.772659 | -0.44326 | 0.688266 | |
| FKTN | 3.005882 | 0.047429 | 0.999532 | | 3.945699 | 0.556612 | 0.53073 | | 3.41177 | 0.524012 | 0.641721 | |
| MCL1 | 12.4944 | -0.6177 | 0.999532 | | 14.7504 | -0.41746 | 0.53073 | | 9.588534 | 0.236233 | 0.779082 | |
| PLA2G4E | 2.327501 | 0.05591 | 0.999532 | | 2.225278 | -0.81633 | 0.530948 | | 1.801299 | -0.85252 | 0.555282 | |
| RNASE1 | 38.70771 | -0.25968 | 0.999532 | | 54.28468 | 0.386239 | 0.530948 | | 43.21803 | 0.67587 | 0.107433 | |
| EIF4H | 14.29045 | 0.025796 | 0.999532 | | 17.65013 | 0.36439 | 0.530948 | | 15.57855 | 0.357365 | 0.563729 | |
| NAXE | 4.488158 | -0.29936 | 0.999532 | | 4.722823 | -0.54411 | 0.530948 | | 3.388104 | -0.22529 | 0.877562 | |
| APOL3 | 6.600502 | 0.246444 | 0.999532 | | 7.860328 | 0.460712 | 0.530948 | | 7.507321 | 0.231517 | 0.825769 | |
| MTHFD1 | 3.131653 | 0.935487 | 0.999532 | | 2.985098 | 0.665713 | 0.531027 | | 3.657811 | -0.23861 | 0.873714 | |
| CTSC | 7.109942 | 0.428838 | 0.999532 | | 9.559737 | 0.841061 | 0.531092 | | 8.984503 | 0.466791 | 0.799944 | |
| MYOG | 2.951751 | -0.26157 | 0.999532 | | 2.938895 | -0.76967 | 0.531882 | | 2.261724 | -0.485 | 0.770105 | |
| PAPOLA | 8.337761 | 0.257205 | 0.999532 | | 10.08896 | 0.412411 | 0.531882 | | 9.299912 | 0.172662 | 0.851503 | |
| ANAPC7 | 4.181645 | 0.110213 | 0.999532 | | 5.530785 | 0.497304 | 0.531962 | | 4.999849 | 0.409769 | 0.678411 | |
| CRNN | 2.641906 | -0.08746 | 0.999532 | | 2.392178 | -0.77516 | 0.531987 | | 2.12894 | -0.67959 | 0.668481 | |
| FGFR4 | 4.078053 | 0.06188 | 0.999532 | | 3.76187 | -0.62967 | 0.531987 | | 3.239762 | -0.67193 | 0.535079 | |
| ASPSCR1 | 2.71878 | 0.310405 | 0.999532 | | 3.578747 | 0.609737 | 0.531987 | | 3.299643 | 0.317194 | 0.810635 | |
| IFITM3 | 70.75453 | -0.10166 | 0.999532 | | 92.451 | 0.320519 | 0.531987 | | 76.60722 | 0.443914 | 0.274496 | |
| CYP1B1 | 4.523581 | -0.05453 | 0.999532 | | 6.126796 | 0.482035 | 0.531987 | | 5.199562 | 0.557453 | 0.515762 | |
| PFDN5 | 13.3134 | -0.14435 | 0.999532 | | 17.93958 | 0.369774 | 0.533662 | | 14.46253 | 0.535846 | 0.456849 | |
| RPL31 | 39.76137 | 0.01268 | 0.999532 | | 49.87497 | 0.321033 | 0.534305 | | 42.5415 | 0.333832 | 0.555282 | |
| RFX3 | 3.621436 | 0.412797 | 0.999532 | | 4.503605 | 0.580877 | 0.534305 | | 4.295417 | 0.186913 | 0.892869 | |
| MYH9 | 115.8259 | 0.209782 | 0.999532 | | 131.1074 | 0.233793 | 0.534305 | | 119.3978 | 0.048048 | 0.946765 | |
| NPIPA1 | 13.07266 | 0.074954 | 0.999532 | | 16.4824 | 0.342064 | 0.534305 | | 14.24294 | 0.288534 | 0.679452 | |
| BSCL2 | 2.855462 | -0.07313 | 0.999532 | | 3.937336 | 0.582198 | 0.534305 | | 3.212573 | 0.670419 | 0.542124 | |
| GPR3 | 2.448701 | -0.44213 | 0.999532 | | 2.601479 | -0.77838 | 0.534635 | | 1.81105 | -0.31659 | 0.875951 | |
| OPA1 | 2.730034 | 0.113643 | 0.999532 | | 3.813265 | 0.607972 | 0.534715 | | 3.331953 | 0.511674 | 0.664512 | |
| FAM76A | 3.482343 | 0.291915 | 0.999532 | | 2.802597 | -0.72096 | 0.534715 | | 2.692823 | -0.98751 | 0.350296 | |
| UHRF1BP | 3.091976 | 0.059974 | 0.999532 | | 2.727719 | -0.69499 | 0.534715 | | 2.419956 | -0.73634 | 0.570828 | |
| SNCG | 6.660076 | -0.21005 | 0.999532 | | 6.652957 | -0.52834 | 0.535128 | | 5.394831 | -0.28597 | 0.801572 | |
| IL4I1 | 13.56887 | -0.10513 | 0.999532 | | 13.4268 | -0.56061 | 0.535675 | | 11.18832 | -0.4365 | 0.723933 | |
| PDCD4 | 15.75334 | 0.274023 | 0.999532 | | 17.53549 | 0.336588 | 0.535675 | | 16.71128 | 0.093233 | 0.903339 | |
| GLG1 | 5.830253 | 0.014353 | 0.999532 | | 7.810819 | 0.432832 | 0.535675 | | 6.49081 | 0.432701 | 0.59668 | |
| CCDC115 | 2.46459 | -0.44387 | 0.999532 | | 3.81961 | 0.611492 | 0.535698 | | 2.876342 | 1.067898 | 0.30043 | |
| DEDD | 4.36326 | 0.232043 | 0.999532 | | 3.90724 | -0.58301 | 0.535701 | | 3.551941 | -0.79082 | 0.413519 | |
| DDTL | 3.029837 | -0.02784 | 0.999532 | | 2.860365 | -0.69366 | 0.535701 | | 2.423022 | -0.64668 | 0.644433 | |
| RESP18 | 3.04765 | -0.19711 | 0.999532 | | 2.833087 | -0.67067 | 0.535701 | | 2.399888 | -0.45796 | 0.758124 | |
| UGT2A3 | 2.653937 | 0.616762 | 0.999532 | | 3.071178 | 0.71207 | 0.535701 | | 3.100729 | 0.117089 | 0.949669 | |
| H2AC14 | 2.742488 | -0.48792 | 0.999532 | | 2.963856 | -0.69333 | 0.535701 | | 2.111092 | -0.18792 | 0.923864 | |
| SHOX2 | 2.768044 | -0.53962 | 0.999532 | | 2.93695 | -0.67183 | 0.535701 | | 2.118378 | -0.12209 | 0.958699 | |
| METRN | 3.765433 | -0.49139 | 0.999532 | | 4.249777 | -0.61243 | 0.535701 | | 3.033761 | -0.09573 | 0.963866 | |
| FRG2C | 6.988619 | 0.416051 | 0.999532 | | 5.743744 | -0.61349 | 0.535701 | | 5.922216 | -1.02474 | 0.2187 | |
| EFCAB2 | 2.306987 | 0.246064 | 0.999532 | | 2.854875 | 0.649878 | 0.535701 | | 2.664619 | 0.420278 | 0.755206 | |
| PSMD1 | 4.013302 | 0.083115 | 0.999532 | | 5.519492 | 0.504243 | 0.535701 | | 4.470705 | 0.446362 | 0.6777 | |
| DCX | 3.409352 | -0.03965 | 0.999532 | | 3.284424 | -0.67538 | 0.535701 | | 2.659375 | -0.61713 | 0.633246 | |
| KANSL3 | 3.756335 | 0.292778 | 0.999532 | | 4.619253 | 0.539384 | 0.535701 | | 4.190505 | 0.267112 | 0.82381 | |
| YIF1A | 4.123114 | 0.474675 | 0.999532 | | 4.665755 | 0.555161 | 0.535701 | | 4.736725 | 0.098836 | 0.949746 | |
| JMY | 6.601167 | 0.364118 | 0.999532 | | 7.531804 | 0.43543 | 0.535701 | | 7.158875 | 0.087436 | 0.945129 | |
| CIT | 2.726485 | -0.19853 | 0.999532 | | 2.742963 | -0.68197 | 0.535701 | | 2.176903 | -0.46386 | 0.757822 | |
| PDK1 | 2.521517 | -0.35993 | 0.999532 | | 2.509234 | -0.7176 | 0.535701 | | 1.921627 | -0.3393 | 0.851503 | |
| WDR45B | 6.501584 | 0.21686 | 0.999532 | | 7.770918 | 0.460869 | 0.535701 | | 7.011427 | 0.26589 | 0.795612 | |
| ACTG2 | 3.203117 | 0.176341 | 0.999532 | | 2.694688 | -0.69456 | 0.535701 | | 2.541671 | -0.85533 | 0.485816 | |
| TMSB10 | 57.6305 | -0.13296 | 0.999532 | | 78.06463 | 0.405753 | 0.535701 | | 64.46491 | 0.558838 | 0.272442 | |
| PCOLCE2 | 23.17609 | 0.29937 | 0.999532 | | 28.61609 | 0.544654 | 0.535701 | | 26.50406 | 0.27362 | 0.795612 | |
| PTH1R | 11.65745 | 0.248627 | 0.999532 | | 13.96314 | 0.466388 | 0.535701 | | 12.79254 | 0.243861 | 0.800239 | |
| PMM2 | 2.658526 | 0.670497 | 0.999532 | | 2.822693 | 0.698338 | 0.535701 | | 3.037093 | 0.046847 | 0.98556 | |
| CEP120 | 3.365779 | -0.36292 | 0.999532 | | 3.39982 | -0.61433 | 0.535701 | | 2.635985 | -0.23417 | 0.886495 | |
| ALG1L2 | 4.558141 | -0.11993 | 0.999532 | | 4.360205 | -0.57697 | 0.535701 | | 3.609889 | -0.44198 | 0.724411 | |
| ADRA1A | 2.148713 | 0.301095 | 0.999532 | | 1.816975 | -0.85988 | 0.535701 | | 1.695027 | -1.14012 | 0.412398 | |
| PIP5K1A | 4.156496 | -0.56283 | 0.999532 | | 4.617008 | -0.58027 | 0.535701 | | 3.359681 | 0.004435 | 0.998655 | |
| TP53AIP1 | 2.083746 | -0.21005 | 0.999532 | | 1.919505 | -0.82415 | 0.535701 | | 1.566598 | -0.59618 | 0.735469 | |
| DCAF10 | 2.936512 | 0.56171 | 0.999532 | | 3.367649 | 0.616534 | 0.535701 | | 3.445078 | 0.073011 | 0.970457 | |
| ERRFI1 | 5.890388 | -1.46834 | 0.912432 | | 7.532488 | -0.90318 | 0.535701 | | 3.583251 | 0.567333 | 0.626555 | |
| CLEC19A | 2.222274 | -0.35515 | 0.999532 | | 2.056922 | -0.78111 | 0.535701 | | 1.731433 | -0.41037 | 0.836935 | |
| TTC16 | 2.767533 | -0.18442 | 0.999532 | | 2.541726 | -0.72818 | 0.535701 | | 2.109318 | -0.52422 | 0.735469 | |
| CFAP161 | 2.085557 | 0.557962 | 0.999532 | | 2.306381 | 0.760553 | 0.535701 | | 2.331907 | 0.22922 | 0.897958 | |
| LY6H | 2.097955 | 0.018085 | 0.999532 | | 1.859314 | -0.84176 | 0.535701 | | 1.572731 | -0.83969 | 0.602029 | |
| TNS1 | 14.38638 | -0.13628 | 0.999532 | | 18.75893 | 0.331029 | 0.535701 | | 15.45562 | 0.487633 | 0.335411 | |
| RNF144A | 7.725922 | -0.20196 | 0.999532 | | 7.950247 | -0.44919 | 0.535701 | | 6.288964 | -0.22714 | 0.823193 | |
| H1-10 | 5.850458 | 0.349257 | 0.999532 | | 6.522258 | 0.46462 | 0.535701 | | 6.353265 | 0.140653 | 0.909074 | |
| KCNQ3 | 2.963775 | 0.021481 | 0.999532 | | 2.679108 | -0.70735 | 0.535701 | | 2.271018 | -0.71097 | 0.579064 | |
| SMAD5 | 5.008312 | -0.09757 | 0.999532 | | 6.953996 | 0.474376 | 0.535701 | | 5.674459 | 0.592152 | 0.515762 | |
| PSEN1 | 7.70581 | -0.50597 | 0.999532 | | 8.932474 | -0.39766 | 0.535701 | | 6.306944 | 0.125966 | 0.914929 | |
| SLC6A6 | 5.280837 | -0.19019 | 0.999532 | | 7.254416 | 0.470977 | 0.535701 | | 5.999255 | 0.664994 | 0.434766 | |
| ANXA5 | 13.14545 | 0.062763 | 0.999532 | | 17.1125 | 0.465469 | 0.535701 | | 14.70655 | 0.430206 | 0.617899 | |
| DDX49 | 2.756572 | 0.009314 | 0.999532 | | 3.768229 | 0.585136 | 0.535701 | | 3.186242 | 0.594324 | 0.600081 | |
| SH2D6 | 3.57594 | -0.22071 | 0.999532 | | 3.665601 | -0.67358 | 0.535701 | | 2.901007 | -0.42369 | 0.784889 | |
| RAB6A | 8.048881 | -0.39024 | 0.999532 | | 11.78409 | 0.389646 | 0.535701 | | 9.018361 | 0.798687 | 0.154583 | |
| TRMT10C | 3.458735 | -0.44112 | 0.999532 | | 3.687177 | -0.60522 | 0.535701 | | 2.760264 | -0.14634 | 0.936689 | |
| CCL5 | 3.79775 | -0.73187 | 0.999532 | | 4.671429 | -0.54113 | 0.535701 | | 3.084525 | 0.209404 | 0.889853 | |
| CD163L1 | 2.636247 | -0.28378 | 0.999532 | | 2.653515 | -0.70485 | 0.535701 | | 2.121603 | -0.40281 | 0.806394 | |
| FBXO45 | 3.140181 | -0.04348 | 0.999532 | | 3.011174 | -0.69042 | 0.535701 | | 2.577846 | -0.62884 | 0.647218 | |
| SLC6A4 | 3.417438 | 0.203462 | 0.999532 | | 2.879363 | -0.67612 | 0.535701 | | 2.777125 | -0.86453 | 0.474973 | |
| HSPA14 | 3.068248 | -0.26216 | 0.999532 | | 2.975141 | -0.67286 | 0.535701 | | 2.322764 | -0.39321 | 0.801572 | |
| SOX12 | 3.22648 | 0.749895 | 0.999532 | | 3.369879 | 0.625248 | 0.535701 | | 3.83829 | -0.10993 | 0.946765 | |
| TTLL11 | 2.625302 | 0.504597 | 0.999532 | | 1.846606 | -0.80553 | 0.535701 | | 2.166722 | -1.29466 | 0.24893 | |
| DCANP1 | 2.655016 | -0.24408 | 0.999532 | | 2.511911 | -0.73042 | 0.535701 | | 2.126663 | -0.47828 | 0.772912 | |
| ITPK1 | 3.471777 | 0.133364 | 0.999532 | | 4.544229 | 0.527782 | 0.535701 | | 3.983358 | 0.41193 | 0.715437 | |
| PCSK1 | 2.330317 | -0.05226 | 0.999532 | | 2.108416 | -0.80781 | 0.535701 | | 1.745851 | -0.73714 | 0.647218 | |
| SASH3 | 2.863172 | -0.2161 | 0.999532 | | 2.674175 | -0.72076 | 0.535701 | | 2.256891 | -0.48674 | 0.742171 | |
| GEMIN7 | 4.665528 | 0.325612 | 0.999532 | | 3.987625 | -0.59709 | 0.535701 | | 3.798513 | -0.90364 | 0.282041 | |
| CRIP2 | 26.77593 | 0.103467 | 0.999532 | | 33.55063 | 0.411746 | 0.536014 | | 29.77709 | 0.330707 | 0.632923 | |
| RASAL1 | 2.461189 | -0.29215 | 0.999532 | | 2.435868 | -0.75398 | 0.536014 | | 1.907981 | -0.44554 | 0.79951 | |
| ASCL5 | 3.230261 | -0.3553 | 0.999532 | | 3.31986 | -0.64622 | 0.536014 | | 2.42335 | -0.27378 | 0.87591 | |
| NAGS | 2.048046 | -0.09917 | 0.999532 | | 1.925906 | -0.83526 | 0.536185 | | 1.552606 | -0.71618 | 0.67176 | |
| HMGA1 | 7.562263 | 0.13051 | 0.999532 | | 6.936893 | -0.46009 | 0.536185 | | 5.972723 | -0.57745 | 0.484303 | |
| CENPQ | 2.221832 | 0.530746 | 0.999532 | | 2.842556 | 0.676922 | 0.536185 | | 2.703001 | 0.164475 | 0.923864 | |
| SLC6A19 | 3.766334 | -0.16799 | 0.999532 | | 5.662183 | 0.590728 | 0.536185 | | 4.34155 | 0.784962 | 0.456849 | |
| PTPRO | 13.525 | 0.363632 | 0.999532 | | 15.67209 | 0.474332 | 0.536185 | | 14.85792 | 0.142344 | 0.897064 | |
| MED28 | 4.404003 | 0.004787 | 0.999532 | | 6.195645 | 0.45399 | 0.536185 | | 5.209626 | 0.470601 | 0.600289 | |
| C2orf92 | 2.43988 | -0.15294 | 0.999532 | | 2.242893 | -0.73918 | 0.536185 | | 1.952534 | -0.56917 | 0.715922 | |
| TMEM25 | 2.386609 | 0.397736 | 0.999532 | | 2.769116 | 0.661981 | 0.536185 | | 2.696655 | 0.278688 | 0.861119 | |
| PDLIM5 | 6.266856 | 0.07401 | 0.999532 | | 8.031683 | 0.428642 | 0.536185 | | 6.912131 | 0.374699 | 0.652925 | |
| PRR22 | 3.929112 | 0.437559 | 0.999532 | | 3.121609 | -0.69709 | 0.536185 | | 3.294209 | -1.12048 | 0.266797 | |
| KRTAP10 | 3.198685 | 0.090003 | 0.999532 | | 2.960655 | -0.70976 | 0.536185 | | 2.801943 | -0.7846 | 0.549022 | |
| ZNF578 | 8.737928 | -0.12318 | 0.999532 | | 8.841687 | -0.43818 | 0.536185 | | 7.505203 | -0.29832 | 0.780676 | |
| RIMKLA | 2.21065 | -0.09093 | 0.999532 | | 2.009275 | -0.82801 | 0.536185 | | 1.601086 | -0.71781 | 0.663104 | |
| RPRD1A | 5.323362 | 0.233477 | 0.999532 | | 6.699288 | 0.475282 | 0.536185 | | 6.12719 | 0.258291 | 0.795612 | |
| DDAH2 | 2.82498 | -0.10505 | 0.999532 | | 3.828144 | 0.56525 | 0.536185 | | 3.195772 | 0.683426 | 0.526728 | |
| DNAI1 | 2.547824 | -0.58953 | 0.999532 | | 2.849855 | -0.68131 | 0.536185 | | 1.985534 | -0.07016 | 0.978282 | |
| POLR2J2 | 11.79425 | -0.12845 | 0.999532 | | 12.58795 | -0.40672 | 0.536185 | | 10.18974 | -0.23526 | 0.816589 | |
| NBPF3 | 2.302431 | 0.841204 | 0.999532 | | 2.377993 | 0.748529 | 0.536185 | | 2.649841 | -0.07666 | 0.972758 | |
| CFHR1 | 2.676234 | -0.13951 | 0.999532 | | 2.652029 | -0.70033 | 0.536185 | | 2.124348 | -0.53868 | 0.717627 | |
| ZFP69 | 3.175485 | -0.07728 | 0.999532 | | 3.082406 | -0.66914 | 0.536185 | | 2.346451 | -0.57102 | 0.663089 | |
| TMEM107 | 5.038396 | -0.4495 | 0.999532 | | 5.506532 | -0.5286 | 0.536185 | | 3.892507 | -0.06237 | 0.974421 | |
| NKPD1 | 2.294126 | 0.010431 | 0.999532 | | 2.100737 | -0.79881 | 0.536185 | | 1.885147 | -0.78831 | 0.62216 | |
| S100A12 | 2.634852 | -0.72157 | 0.999532 | | 3.06553 | -0.66249 | 0.536185 | | 2.014101 | 0.078733 | 0.974421 | |
| LIF | 2.863731 | 0.14595 | 0.999532 | | 2.485268 | -0.74202 | 0.536185 | | 2.15829 | -0.87358 | 0.511836 | |
| TAB3 | 3.5707 | 0.16343 | 0.999532 | | 4.697383 | 0.587285 | 0.536185 | | 4.329635 | 0.440469 | 0.707142 | |
| LY6E | 9.753426 | 0.104107 | 0.999532 | | 12.05456 | 0.381935 | 0.536185 | | 10.68801 | 0.299388 | 0.669726 | |
| H3C11 | 2.297486 | 0.562709 | 0.999532 | | 2.646641 | 0.729982 | 0.536185 | | 2.688567 | 0.182427 | 0.919493 | |
| DLG1 | 2.724184 | 0.186491 | 0.999532 | | 3.539383 | 0.598049 | 0.536185 | | 3.23295 | 0.427118 | 0.728015 | |
| AK4 | 7.666555 | 0.384134 | 0.999532 | | 8.814298 | 0.481835 | 0.536689 | | 8.513442 | 0.107018 | 0.941197 | |
| ZNF540 | 4.102693 | 0.393179 | 0.999532 | | 5.014873 | 0.521049 | 0.536689 | | 4.722289 | 0.144549 | 0.909857 | |
| NEDD8 | 10.50185 | 0.294909 | 0.999532 | | 11.80759 | 0.358842 | 0.53734 | | 11.32931 | 0.087558 | 0.927375 | |
| VCX | 2.839119 | 0.322917 | 0.999532 | | 2.144566 | -0.76785 | 0.53734 | | 2.332499 | -1.07571 | 0.397004 | |
| SPRYD4 | 3.94344 | -0.10682 | 0.999532 | | 3.707858 | -0.59347 | 0.53757 | | 3.071513 | -0.47626 | 0.710808 | |
| NPR2 | 6.685008 | -0.62987 | 0.999532 | | 7.796225 | -0.4293 | 0.537841 | | 5.079974 | 0.210138 | 0.861217 | |
| HMGCS1 | 4.033979 | 0.470059 | 0.999532 | | 4.476285 | 0.522126 | 0.537841 | | 4.52982 | 0.071244 | 0.965649 | |
| SRSF9 | 11.33581 | 0.280415 | 0.999532 | | 12.59076 | 0.345848 | 0.537841 | | 12.07573 | 0.081429 | 0.939628 | |
| MAP4 | 7.986329 | 0.132256 | 0.999532 | | 9.959591 | 0.432409 | 0.537841 | | 8.730337 | 0.318052 | 0.724411 | |
| CCKBR | 2.693543 | -0.20596 | 0.999532 | | 2.582234 | -0.70659 | 0.537841 | | 2.131244 | -0.48244 | 0.757311 | |
| RNPEPL1 | 5.287896 | 0.356985 | 0.999532 | | 6.632731 | 0.47405 | 0.538929 | | 6.073417 | 0.137973 | 0.911296 | |
| NKX2-4 | 9.596768 | 0.3217 | 0.999532 | | 8.49264 | -0.46463 | 0.538929 | | 8.449341 | -0.7682 | 0.273244 | |
| SNRPD2 | 9.44621 | 0.102256 | 0.999532 | | 11.83104 | 0.365174 | 0.538929 | | 10.49118 | 0.283611 | 0.709825 | |
| OXR1 | 5.430061 | 0.23955 | 0.999532 | | 6.492829 | 0.464565 | 0.538929 | | 5.851091 | 0.244806 | 0.809832 | |
| EPOR | 2.926565 | 0.038471 | 0.999532 | | 2.636689 | -0.69588 | 0.539596 | | 2.316226 | -0.71805 | 0.595883 | |
| COX7A2L | 12.0437 | 0.213515 | 0.999532 | | 14.65697 | 0.360006 | 0.539596 | | 13.28063 | 0.173667 | 0.840357 | |
| CAV1 | 3.176512 | 0.095058 | 0.999532 | | 4.076219 | 0.579532 | 0.539596 | | 3.69603 | 0.483928 | 0.694301 | |
| KIAA0895 | 5.353811 | -0.07909 | 0.999532 | | 5.128962 | -0.49287 | 0.539596 | | 4.272298 | -0.40258 | 0.702463 | |
| LAMB1 | 6.773891 | 0.192814 | 0.999532 | | 8.580583 | 0.481688 | 0.539596 | | 7.519735 | 0.315791 | 0.760473 | |
| PRR14 | 3.843127 | -0.10726 | 0.999532 | | 5.469957 | 0.486498 | 0.539596 | | 4.437392 | 0.614271 | 0.515762 | |
| HSPA9 | 13.78308 | -0.002 | 0.999532 | | 17.29017 | 0.350021 | 0.539596 | | 14.46798 | 0.380193 | 0.540342 | |
| M6PR | 3.317095 | 0.479019 | 0.999532 | | 3.712615 | 0.58132 | 0.539596 | | 3.62383 | 0.123225 | 0.937825 | |
| PRR5 | 4.064772 | -0.15775 | 0.999532 | | 5.843241 | 0.493795 | 0.539596 | | 4.658831 | 0.67242 | 0.464413 | |
| TRIM51 | 4.740606 | -0.17441 | 0.999532 | | 4.823398 | -0.53917 | 0.539596 | | 4.026929 | -0.34857 | 0.782299 | |
| AMOTL1 | 6.012591 | 0.104494 | 0.999532 | | 7.957212 | 0.48399 | 0.539596 | | 6.91392 | 0.394153 | 0.652925 | |
| MLXIP | 6.207174 | 0.095835 | 0.999532 | | 8.042924 | 0.417979 | 0.539596 | | 6.866423 | 0.331765 | 0.713778 | |
| SPATA17 | 3.708695 | -0.26633 | 0.999532 | | 3.72248 | -0.63585 | 0.539596 | | 2.731674 | -0.34191 | 0.816526 | |
| PGBD5 | 2.88542 | 0.019366 | 0.999532 | | 2.735692 | -0.72232 | 0.539596 | | 2.320717 | -0.72186 | 0.60646 | |
| COA3 | 3.707253 | 0.492339 | 0.999532 | | 4.195553 | 0.535458 | 0.539596 | | 4.077227 | 0.058317 | 0.973152 | |
| MEF2A | 8.255638 | 0.261305 | 0.999532 | | 9.383034 | 0.419904 | 0.539596 | | 8.9587 | 0.161806 | 0.88476 | |
| SAP30BP | 4.462061 | 0.222018 | 0.999532 | | 5.337949 | 0.502177 | 0.539596 | | 4.917239 | 0.292969 | 0.796512 | |
| ABCF2 | 3.480194 | -0.07078 | 0.999532 | | 4.967706 | 0.524832 | 0.539596 | | 4.248136 | 0.614006 | 0.515762 | |
| DPY30 | 4.649167 | -0.29918 | 0.999532 | | 4.891283 | -0.51586 | 0.541517 | | 3.576797 | -0.1969 | 0.888003 | |
| NHEJ1 | 1.438273 | 0.260201 | 0.999532 | | 1.990387 | 0.776003 | 0.541517 | | 1.874121 | 0.537301 | 0.739496 | |
| S100A2 | 3.135457 | -0.32745 | 0.999532 | | 3.149179 | -0.68581 | 0.541568 | | 2.369163 | -0.3383 | 0.831123 | |
| SEC61G | 7.238861 | 0.164141 | 0.999532 | | 9.080834 | 0.42745 | 0.541961 | | 7.985847 | 0.285508 | 0.736302 | |
| OR4F4 | 5.130591 | -0.0303 | 0.999532 | | 5.079174 | -0.64494 | 0.541961 | | 4.163537 | -0.58115 | 0.645376 | |
| TMEM117 | 2.47966 | -0.35927 | 0.999532 | | 2.55339 | -0.70456 | 0.542725 | | 1.827189 | -0.32664 | 0.859898 | |
| BTN2A2 | 2.350054 | 0.608442 | 0.999532 | | 2.562184 | 0.672187 | 0.543883 | | 2.733623 | 0.081475 | 0.969504 | |
| UBE2V2 | 5.886399 | 0.001486 | 0.999532 | | 7.929793 | 0.413672 | 0.543883 | | 6.681351 | 0.434293 | 0.602786 | |
| A2M | 49.31115 | -0.28061 | 0.999532 | | 73.53366 | 0.497729 | 0.54393 | | 58.19013 | 0.795468 | 0.154583 | |
| PAQR9 | 3.023897 | 0.203247 | 0.999532 | | 2.533899 | -0.72842 | 0.544106 | | 2.507719 | -0.91474 | 0.468942 | |
| SIGLEC6 | 2.338358 | 0.078522 | 0.999532 | | 2.076396 | -0.79496 | 0.54418 | | 1.678586 | -0.85438 | 0.557909 | |
| SCNN1G | 2.128591 | -0.0047 | 0.999532 | | 2.935841 | 0.651899 | 0.54418 | | 2.527289 | 0.676318 | 0.5991 | |
| ATM | 2.924363 | 0.567091 | 0.999532 | | 3.340635 | 0.611812 | 0.54418 | | 3.406987 | 0.060672 | 0.975473 | |
| LRRC36 | 2.565722 | -0.16243 | 0.999532 | | 2.475682 | -0.73881 | 0.54418 | | 1.961654 | -0.55744 | 0.725637 | |
| RPL14 | 30.49176 | -0.04793 | 0.999532 | | 39.03165 | 0.33218 | 0.54418 | | 32.72938 | 0.398083 | 0.515762 | |
| BMP1 | 4.836755 | 0.413634 | 0.999532 | | 5.60569 | 0.518245 | 0.54418 | | 5.243122 | 0.127448 | 0.92697 | |
| STARD7 | 8.154597 | 0.033075 | 0.999532 | | 10.33548 | 0.396173 | 0.544197 | | 8.896537 | 0.379836 | 0.643272 | |
| CIDEB | 3.762542 | -0.10931 | 0.999532 | | 3.623173 | -0.59343 | 0.544233 | | 2.921624 | -0.46644 | 0.709825 | |
| ZFP36L2 | 8.314305 | -0.45409 | 0.999532 | | 9.475768 | -0.38089 | 0.544233 | | 6.83798 | 0.091542 | 0.938624 | |
| SRSF1 | 10.67804 | -0.05698 | 0.999532 | | 13.99957 | 0.349552 | 0.544233 | | 11.75443 | 0.427823 | 0.515762 | |
| BLM | 3.572714 | -0.59516 | 0.999532 | | 3.873213 | -0.59438 | 0.544233 | | 2.797533 | 0.011437 | 0.996891 | |
| CYB5R3 | 14.36201 | -0.07755 | 0.999532 | | 18.62072 | 0.309312 | 0.544233 | | 15.37559 | 0.405396 | 0.499673 | |
| CASTOR2 | 3.174833 | -0.80945 | 0.999532 | | 3.786747 | -0.57244 | 0.544233 | | 2.268041 | 0.254606 | 0.880137 | |
| QPRT | 4.840367 | 0.50788 | 0.999532 | | 5.668834 | 0.562363 | 0.544233 | | 5.493891 | 0.095804 | 0.956011 | |
| OGA | 10.71367 | 0.015542 | 0.999532 | | 14.03829 | 0.38572 | 0.544233 | | 11.84401 | 0.396424 | 0.623565 | |
| SLC7A7 | 4.251196 | 0.399847 | 0.999532 | | 4.656872 | 0.534202 | 0.544233 | | 4.717579 | 0.145751 | 0.913432 | |
| RPL39 | 15.02268 | 0.127116 | 0.999532 | | 18.18413 | 0.383653 | 0.544233 | | 16.2563 | 0.263068 | 0.766618 | |
| TOR3A | 3.110574 | 0.073513 | 0.999532 | | 2.784477 | -0.72931 | 0.544233 | | 2.229279 | -0.77507 | 0.542124 | |
| FAM104B | 3.882785 | -0.34864 | 0.999532 | | 4.199474 | -0.59561 | 0.544233 | | 3.125409 | -0.23293 | 0.880949 | |
| MYSM1 | 2.403738 | 0.115118 | 0.999532 | | 3.211545 | 0.600334 | 0.544565 | | 2.790985 | 0.501683 | 0.686214 | |
| ZNF565 | 2.274963 | -0.20738 | 0.999532 | | 2.088923 | -0.75465 | 0.545317 | | 1.758653 | -0.53078 | 0.760473 | |
| IMMP1L | 4.993173 | -0.18619 | 0.999532 | | 5.161705 | -0.62151 | 0.545447 | | 4.067271 | -0.41379 | 0.753585 | |
| ATP8B4 | 2.425167 | -0.45637 | 0.999532 | | 2.52275 | -0.69315 | 0.545447 | | 1.725626 | -0.21818 | 0.913083 | |
| FAM161B | 3.25637 | 0.360676 | 0.999532 | | 2.467475 | -0.70214 | 0.54552 | | 2.555474 | -1.05289 | 0.363418 | |
| ASCL4 | 2.217678 | 0.087559 | 0.999532 | | 1.880018 | -0.81009 | 0.545528 | | 1.771805 | -0.88092 | 0.552672 | |
| NDUFAF3 | 7.122984 | 0.099878 | 0.999532 | | 8.631828 | 0.412352 | 0.545839 | | 7.800177 | 0.314628 | 0.734912 | |
| DLX1 | 1.946805 | -0.27634 | 0.999532 | | 1.833096 | -0.82637 | 0.545956 | | 1.477951 | -0.53525 | 0.783339 | |
| PPP1R15 | 10.24331 | -0.74357 | 0.999532 | | 12.13699 | -0.47548 | 0.546414 | | 7.704944 | 0.330275 | 0.686214 | |
| CERT1 | 3.627236 | -0.20285 | 0.999532 | | 5.356974 | 0.491153 | 0.546414 | | 4.350445 | 0.714587 | 0.421517 | |
| KDM4B | 8.164228 | 0.106291 | 0.999532 | | 9.852132 | 0.385251 | 0.546414 | | 8.761962 | 0.293071 | 0.757035 | |
| COMMD5 | 3.147996 | -0.50069 | 0.999532 | | 3.347789 | -0.62657 | 0.546414 | | 2.488174 | -0.11925 | 0.956011 | |
| KCNG4 | 3.857997 | 0.771231 | 0.999532 | | 2.650811 | -0.74002 | 0.546586 | | 3.275037 | -1.49582 | 0.121098 | |
| GSC | 3.440778 | 0.019986 | 0.999532 | | 3.146078 | -0.62335 | 0.546709 | | 2.704703 | -0.6262 | 0.60646 | |
| SLC29A4 | 3.586623 | -0.18352 | 0.999532 | | 3.584681 | -0.59316 | 0.546774 | | 2.931643 | -0.39117 | 0.776852 | |
| SYVN1 | 7.512693 | 0.011491 | 0.999532 | | 7.22678 | -0.48828 | 0.546774 | | 6.193002 | -0.47898 | 0.578798 | |
| BNIP3 | 6.630963 | 0.430159 | 0.999532 | | 7.747578 | 0.421566 | 0.546774 | | 7.406216 | 0.018481 | 0.992286 | |
| ATG5 | 3.52286 | 0.025328 | 0.999532 | | 5.053345 | 0.520432 | 0.547094 | | 4.239447 | 0.525134 | 0.632802 | |
| SLC4A11 | 3.913005 | 0.12178 | 0.999532 | | 4.721478 | 0.547309 | 0.547104 | | 4.326473 | 0.42651 | 0.720609 | |
| LHX6 | 2.315863 | -0.45971 | 0.999532 | | 2.552079 | -0.71699 | 0.547174 | | 1.841262 | -0.23516 | 0.907626 | |
| IGF1 | 2.745453 | 0.717956 | 0.999532 | | 3.016527 | 0.639112 | 0.547174 | | 3.2377 | -0.05721 | 0.978229 | |
| AGBL3 | 3.639282 | -0.05356 | 0.999532 | | 3.372792 | -0.62413 | 0.547391 | | 2.779235 | -0.55787 | 0.666566 | |
| SIRT3 | 2.663341 | 0.271409 | 0.999532 | | 3.598939 | 0.595996 | 0.547391 | | 3.255023 | 0.346372 | 0.795612 | |
| WDR13 | 4.538763 | 0.090744 | 0.999532 | | 5.890576 | 0.511066 | 0.547391 | | 4.863065 | 0.444411 | 0.663412 | |
| MYO15B | 7.008159 | 0.287753 | 0.999532 | | 8.265519 | 0.405345 | 0.547391 | | 7.760186 | 0.137693 | 0.893828 | |
| MRM1 | 3.498964 | -0.33444 | 0.999532 | | 3.717755 | -0.61333 | 0.548113 | | 2.79844 | -0.25832 | 0.87591 | |
| NKX6-2 | 5.325331 | -0.18406 | 0.999532 | | 5.441701 | -0.50392 | 0.548113 | | 4.306666 | -0.30117 | 0.802341 | |
| HIF3A | 5.296872 | -0.59034 | 0.999532 | | 6.162059 | -0.54674 | 0.548113 | | 3.923432 | 0.053145 | 0.977473 | |
| GRIN2B | 2.580989 | 0.449255 | 0.999532 | | 2.059138 | -0.76576 | 0.548232 | | 2.122031 | -1.19626 | 0.282041 | |
| USP13 | 3.029537 | -0.14055 | 0.999532 | | 2.882656 | -0.64977 | 0.548232 | | 2.405068 | -0.49497 | 0.737743 | |
| HCFC2 | 3.158268 | 0.300598 | 0.999532 | | 4.002898 | 0.569401 | 0.548232 | | 3.823294 | 0.284452 | 0.827267 | |
| PPP1R14 | 15.92076 | -0.12951 | 0.999532 | | 17.52887 | -0.29581 | 0.548232 | | 13.63683 | -0.15141 | 0.837256 | |
| ZNF800 | 3.206433 | -0.14233 | 0.999532 | | 4.463806 | 0.510459 | 0.548232 | | 3.668246 | 0.666873 | 0.515762 | |
| THRAP3 | 11.55685 | 0.231869 | 0.999532 | | 13.83718 | 0.333126 | 0.548232 | | 12.49864 | 0.127261 | 0.884838 | |
| TNFAIP3 | 4.092459 | -0.90877 | 0.999532 | | 4.725281 | -0.53373 | 0.548232 | | 2.942299 | 0.379236 | 0.787644 | |
| SMAD2 | 8.979365 | 0.084548 | 0.999532 | | 10.81103 | 0.382685 | 0.548232 | | 9.566089 | 0.313685 | 0.707855 | |
| FERMT2 | 14.09752 | 0.12821 | 0.999532 | | 17.15242 | 0.350255 | 0.548232 | | 15.52371 | 0.248471 | 0.701201 | |
| FUT8 | 2.518463 | 0.185506 | 0.999532 | | 3.299457 | 0.610688 | 0.548232 | | 2.830143 | 0.442227 | 0.738702 | |
| SLC33A1 | 4.572377 | -0.55014 | 0.999532 | | 5.205391 | -0.5168 | 0.548232 | | 3.57238 | 0.052285 | 0.978312 | |
| OTOF | 3.14406 | -0.18192 | 0.999532 | | 3.121758 | -0.64861 | 0.548232 | | 2.371083 | -0.44769 | 0.757035 | |
| NAT14 | 2.918314 | 0.505364 | 0.999532 | | 2.392696 | -0.74853 | 0.548232 | | 2.434062 | -1.22968 | 0.254511 | |
| OR7D2 | 2.496747 | 0.083818 | 0.999532 | | 2.129003 | -0.77171 | 0.548232 | | 1.985821 | -0.84114 | 0.560424 | |
| WDHD1 | 3.268831 | -0.19201 | 0.999532 | | 3.583027 | -0.6464 | 0.548232 | | 2.752238 | -0.41469 | 0.783005 | |
| ETNK2 | 4.51046 | -0.25081 | 0.999532 | | 4.423858 | -0.56395 | 0.548232 | | 3.449002 | -0.29071 | 0.829716 | |
| CHMP1B | 5.758182 | 0.21086 | 0.999532 | | 6.855783 | 0.43297 | 0.548232 | | 6.326029 | 0.243102 | 0.79686 | |
| FARP2 | 5.538145 | 0.140489 | 0.999532 | | 6.674157 | 0.430746 | 0.548555 | | 6.006841 | 0.306971 | 0.737109 | |
| NDUFV3 | 4.543192 | 0.310151 | 0.999532 | | 5.398492 | 0.492156 | 0.548555 | | 5.023296 | 0.195008 | 0.882613 | |
| OLR1 | 3.011913 | 0.002603 | 0.999532 | | 2.940036 | -0.64361 | 0.548555 | | 2.426713 | -0.61411 | 0.652925 | |
| MT1F | 14.30012 | -0.09286 | 0.999532 | | 20.10205 | 0.492761 | 0.549073 | | 16.36386 | 0.61233 | 0.442182 | |

| | | | | | | | | | | | |
|---|---|---|---|---|---|---|---|---|---|---|---|
| ADAT1 | 3.528635 | -0.05876 | 0.999532 | | 3.215043 | -0.65011 | 0.549193 | | 2.777119 | -0.57296 | 0.665886 |
| LOXHD1 | 2.478189 | 0.114129 | 0.999532 | | 2.193123 | -0.7469 | 0.549193 | | 1.961723 | -0.84338 | 0.540342 |
| RNF135 | 2.72891 | 0.816834 | 0.999532 | | 2.869271 | 0.674649 | 0.549193 | | 3.074744 | -0.11996 | 0.945902 |
| PTMS | 34.98453 | -0.00442 | 0.999532 | | 43.84628 | 0.311814 | 0.549193 | | 37.62945 | 0.343121 | 0.469608 |
| UTP3 | 4.871801 | 0.257064 | 0.999532 | | 5.81857 | 0.530275 | 0.549193 | | 5.456731 | 0.297215 | 0.795928 |
| ZNF816 | 4.677524 | -0.29961 | 0.999532 | | 5.155747 | -0.52257 | 0.549193 | | 3.977008 | -0.20943 | 0.880375 |
| NUDC | 6.94618 | -0.09721 | 0.999532 | | 9.54126 | 0.408843 | 0.549356 | | 7.589084 | 0.530859 | 0.484303 |
| CNFN | 2.286791 | -0.5365 | 0.999532 | | 2.360157 | -0.71095 | 0.549489 | | 1.721027 | -0.16049 | 0.945129 |
| BEGAIN | 3.544118 | 0.119874 | 0.999532 | | 3.12382 | -0.6298 | 0.549512 | | 2.876593 | -0.73441 | 0.531575 |
| ZNF638 | 4.425009 | -0.24132 | 0.999532 | | 6.141588 | 0.448808 | 0.549512 | | 4.869303 | 0.70432 | 0.386321 |
| PPP1R17 | 2.206941 | -0.09783 | 0.999532 | | 2.111056 | -0.77568 | 0.549512 | | 1.776522 | -0.65488 | 0.693035 |
| TMEM184 | 2.62023 | -0.36233 | 0.999532 | | 2.615385 | -0.71351 | 0.549512 | | 2.064492 | -0.33079 | 0.860694 |
| GTF2A2 | 7.147017 | 0.413207 | 0.999532 | | 7.980282 | 0.437814 | 0.549714 | | 7.770082 | 0.042643 | 0.977473 |
| SLC22A5 | 2.730677 | 0.68369 | 0.999532 | | 2.800031 | 0.672362 | 0.549715 | | 3.003351 | 0.012862 | 0.996891 |
| FUT4 | 3.557291 | -0.18738 | 0.999532 | | 3.695357 | -0.5815 | 0.549907 | | 2.835987 | -0.36866 | 0.792416 |
| CDC14A | 4.652132 | 0.576858 | 0.999532 | | 5.129039 | 0.498866 | 0.549907 | | 5.363754 | -0.06106 | 0.969504 |
| SPC24 | 2.905867 | -0.46447 | 0.999532 | | 3.126889 | -0.63904 | 0.549907 | | 2.221869 | -0.15633 | 0.93826 |
| FAM243A | 2.532864 | 0.59191 | 0.999532 | | 1.864289 | -0.81473 | 0.549907 | | 2.075189 | -1.38802 | 0.235624 |
| MKLN1 | 4.652113 | 0.440561 | 0.999532 | | 5.173139 | 0.494003 | 0.549907 | | 5.347432 | 0.072108 | 0.963868 |
| SERINC4 | 2.308205 | -0.32965 | 0.999532 | | 2.306484 | -0.75831 | 0.549907 | | 1.765664 | -0.41113 | 0.831297 |
| SERGEF | 3.417329 | -0.57935 | 0.999532 | | 3.796142 | -0.56572 | 0.55006 | | 2.768287 | 0.033745 | 0.990864 |
| ARMC3 | 2.502655 | -0.32919 | 0.999532 | | 2.605863 | -0.71904 | 0.55006 | | 1.863226 | -0.3703 | 0.836453 |
| ZNF397 | 6.553122 | -0.36574 | 0.999532 | | 7.158108 | -0.46621 | 0.55006 | | 5.40302 | -0.0852 | 0.955834 |
| TMEM169 | 2.027602 | -0.8317 | 0.999532 | | 2.308855 | -0.73237 | 0.55006 | | 1.507786 | 0.115083 | 0.968235 |
| LRP10 | 7.749939 | -0.37382 | 0.999532 | | 11.03762 | 0.392942 | 0.55006 | | 8.404106 | 0.780656 | 0.225969 |
| TOB1 | 12.90015 | -0.22126 | 0.999532 | | 13.71143 | -0.35413 | 0.55006 | | 10.63933 | -0.11765 | 0.905952 |
| ZNF761 | 9.22545 | -0.25615 | 0.999532 | | 9.902249 | -0.39555 | 0.55006 | | 7.560839 | -0.1228 | 0.914929 |
| CCDC184 | 26.15565 | 0.133566 | 0.999532 | | 24.0076 | -0.55076 | 0.55006 | | 21.33104 | -0.69998 | 0.19389 |
| LCA5L | 3.258761 | -0.27383 | 0.999532 | | 3.115933 | -0.65704 | 0.55006 | | 2.469495 | -0.36064 | 0.8314 |
| KRTAP12 | 4.356665 | 0.04159 | 0.999532 | | 4.062433 | -0.5983 | 0.55006 | | 3.696495 | -0.63112 | 0.572297 |
| ADGRF2 | 2.344469 | -0.14221 | 0.999532 | | 2.252661 | -0.73628 | 0.55026 | | 1.869883 | -0.57479 | 0.722703 |
| H3C10 | 3.617937 | 0.386853 | 0.999532 | | 4.147757 | 0.542494 | 0.551291 | | 4.154974 | 0.165269 | 0.908698 |
| LCK | 3.367238 | -0.24803 | 0.999532 | | 3.330682 | -0.6141 | 0.552214 | | 2.694247 | -0.35443 | 0.816775 |
| NDUFS3 | 6.889476 | 0.324681 | 0.999532 | | 7.849797 | 0.411483 | 0.552511 | | 7.291108 | 0.10383 | 0.929473 |
| TRIM49B | 2.479618 | -0.10274 | 0.999532 | | 2.189 | -0.74178 | 0.55256 | | 1.886273 | -0.62827 | 0.694961 |
| MICALL2 | 6.652352 | -0.05126 | 0.999532 | | 9.37376 | 0.494282 | 0.55273 | | 7.649955 | 0.594391 | 0.515762 |
| BPTF | 2.45146 | -0.07065 | 0.999532 | | 3.744024 | 0.581285 | 0.552884 | | 3.060835 | 0.68045 | 0.554813 |
| HYI | 3.925641 | -0.27784 | 0.999532 | | 4.441584 | -0.55959 | 0.552884 | | 3.242615 | -0.2588 | 0.859522 |
| TBCA | 17.00371 | 0.070157 | 0.999532 | | 20.88096 | 0.338711 | 0.552884 | | 18.4949 | 0.280286 | 0.656473 |
| PAIP1 | 6.363373 | -0.04779 | 0.999532 | | 8.93395 | 0.394317 | 0.552884 | | 7.220457 | 0.473192 | 0.566812 |
| ALDH6A1 | 4.404298 | 0.105391 | 0.999532 | | 5.919026 | 0.488125 | 0.552884 | | 4.972211 | 0.406966 | 0.699971 |
| MTSS1 | 8.168387 | -0.08258 | 0.999532 | | 11.12599 | 0.378925 | 0.552884 | | 8.846672 | 0.472367 | 0.515762 |
| NDN | 3.43319 | 0.459248 | 0.999532 | | 3.876505 | 0.576777 | 0.552884 | | 3.955716 | 0.1274 | 0.938429 |
| ASCL2 | 3.128785 | -0.18755 | 0.999532 | | 3.195704 | -0.62407 | 0.552884 | | 2.415053 | -0.41712 | 0.776158 |
| PLD3 | 15.44767 | 0.098004 | 0.999532 | | 19.35058 | 0.363777 | 0.552884 | | 17.02445 | 0.280545 | 0.647581 |
| PIGN | 3.706288 | -0.44094 | 0.999532 | | 3.807914 | -0.57324 | 0.552884 | | 2.883289 | -0.12316 | 0.947322 |
| GPR4 | 10.45069 | -0.23453 | 0.999532 | | 11.27743 | -0.3463 | 0.552884 | | 9.073022 | -0.09246 | 0.926702 |
| KDM5C | 11.1702 | 0.345967 | 0.999532 | | 12.20867 | 0.34381 | 0.552884 | | 11.89205 | 0.007894 | 0.996891 |
| ALDOB | 41.1416 | 0.401784 | 0.999532 | | 53.77124 | 0.774139 | 0.552884 | | 51.18967 | 0.382413 | 0.865574 |
| UBL3 | 6.678399 | -0.08905 | 0.999532 | | 6.327871 | -0.45498 | 0.552884 | | 5.236794 | -0.35718 | 0.722703 |
| PPFIA1 | 5.329711 | 0.132927 | 0.999532 | | 6.483593 | 0.482921 | 0.552884 | | 5.992375 | 0.369679 | 0.717627 |
| PCBP4 | 6.280091 | 0.233742 | 0.999532 | | 7.821632 | 0.428728 | 0.552884 | | 7.197087 | 0.217817 | 0.831749 |
| GBP1 | 3.984672 | -0.04626 | 0.999532 | | 3.701037 | -0.60857 | 0.552884 | | 3.301377 | -0.55406 | 0.717138 |
| ARL11 | 2.836626 | -0.42364 | 0.999532 | | 2.973813 | -0.65412 | 0.552884 | | 2.126024 | -0.21261 | 0.9094 |
| SLC2A3 | 4.070356 | -0.28923 | 0.999532 | | 4.304573 | -0.64406 | 0.552884 | | 3.069088 | -0.33866 | 0.816618 |
| ITGA10 | 3.057183 | -0.15634 | 0.999532 | | 2.904633 | -0.6435 | 0.552884 | | 2.394717 | -0.47059 | 0.75311 |
| FANCM | 2.350441 | -0.37118 | 0.999532 | | 2.40279 | -0.71735 | 0.552884 | | 1.664696 | -0.32396 | 0.864245 |
| ZFP2 | 2.994644 | -0.15828 | 0.999532 | | 2.888813 | -0.72798 | 0.552884 | | 2.256166 | -0.55344 | 0.706749 |
| CHSY1 | 5.30522 | -0.23632 | 0.999532 | | 5.68618 | -0.51626 | 0.552884 | | 4.221394 | -0.26059 | 0.834593 |
| LMNB2 | 1.778443 | 0.297283 | 0.999532 | | 2.305802 | 0.726543 | 0.552884 | | 2.203489 | 0.448116 | 0.78258 |
| CCT8L2 | 2.221109 | -0.3137 | 0.999532 | | 2.266832 | -0.74015 | 0.552884 | | 1.783268 | -0.40598 | 0.836453 |
| MARCKS | 8.069281 | 0.108736 | 0.999532 | | 9.560588 | 0.414814 | 0.552884 | | 8.616301 | 0.329827 | 0.692762 |
| SRSF10 | 5.899788 | 0.278455 | 0.999532 | | 7.282226 | 0.42457 | 0.552884 | | 6.54805 | 0.163367 | 0.88476 |
| PPP1R7 | 6.009794 | 0.225233 | 0.999532 | | 7.646328 | 0.460185 | 0.552884 | | 6.795353 | 0.257888 | 0.803301 |
| ZFR2 | 2.767058 | 0.163898 | 0.999532 | | 2.178728 | -0.74045 | 0.552884 | | 2.184795 | -0.88839 | 0.515762 |
| NDUFS8 | 5.942847 | 0.319541 | 0.999532 | | 7.029632 | 0.434708 | 0.552884 | | 6.606575 | 0.133176 | 0.903892 |
| INPP5K | 3.242362 | 0.332885 | 0.999532 | | 3.860693 | 0.552484 | 0.552884 | | 3.646274 | 0.235987 | 0.864245 |
| IQGAP1 | 7.165093 | -0.24692 | 0.999532 | | 9.956008 | 0.442096 | 0.552884 | | 8.056223 | 0.699873 | 0.312156 |
| ZNF106 | 3.586756 | -0.03027 | 0.999532 | | 4.83892 | 0.518578 | 0.552884 | | 4.043825 | 0.567241 | 0.566812 |
| SNAI2 | 5.099517 | 0.13316 | 0.999532 | | 6.642459 | 0.493755 | 0.552884 | | 5.922237 | 0.374538 | 0.702463 |
| SF1 | 15.09612 | 0.147326 | 0.999532 | | 18.20082 | 0.318649 | 0.552884 | | 15.8768 | 0.190023 | 0.791466 |
| CHRFAM | 7.204202 | -0.01335 | 0.999532 | | 7.190462 | -0.50775 | 0.552884 | | 6.062134 | -0.48608 | 0.603483 |
| HSP90B1 | 21.54383 | 0.239019 | 0.999532 | | 25.97553 | 0.424816 | 0.552884 | | 23.93485 | 0.203081 | 0.799112 |
| ZNF140 | 20.51113 | 0.071674 | 0.999532 | | 25.01885 | 0.305533 | 0.552884 | | 21.38543 | 0.259269 | 0.709589 |
| AP5M1 | 2.551456 | 0.013113 | 0.999532 | | 2.472813 | -0.70041 | 0.552884 | | 1.913375 | -0.69526 | 0.632923 |
| SDF4 | 12.71841 | -0.04935 | 0.999532 | | 16.28557 | 0.339521 | 0.552884 | | 13.8039 | 0.409118 | 0.503444 |
| BID | 2.56969 | -0.35844 | 0.999532 | | 2.792297 | -0.7044 | 0.552884 | | 1.99995 | -0.32645 | 0.859065 |
| H4C15 | 10.43763 | 0.112662 | 0.999532 | | 12.89387 | 0.36879 | 0.552884 | | 11.18662 | 0.278151 | 0.72567 |
| ANKRD24 | 2.318362 | -0.12596 | 0.999532 | | 2.009378 | -0.7998 | 0.552884 | | 1.756159 | -0.64128 | 0.717138 |
| VPS41 | 5.609899 | 0.212651 | 0.999532 | | 6.937388 | 0.456478 | 0.552884 | | 6.294624 | 0.261728 | 0.794815 |
| C19orf73 | 3.161291 | -0.30677 | 0.999532 | | 3.40536 | -0.65066 | 0.55292 | | 2.463727 | -0.31174 | 0.857916 |
| SLC24A1 | 2.255238 | -0.59507 | 0.999532 | | 2.538353 | -0.72079 | 0.55292 | | 1.64127 | -0.10448 | 0.969678 |
| RAMAC | 4.515742 | -0.27556 | 0.999532 | | 4.714577 | -0.52211 | 0.55292 | | 3.689585 | -0.22645 | 0.877268 |
| BRMS1 | 4.801746 | 0.417112 | 0.999532 | | 5.659557 | 0.476568 | 0.55298 | | 5.483935 | 0.081186 | 0.955834 |
| PAPPA2 | 2.960141 | 0.892982 | 0.999532 | | 2.039346 | -0.75364 | 0.55298 | | 2.415423 | -1.61105 | 0.107433 |
| TMX4 | 4.546128 | 0.588899 | 0.999532 | | 4.744619 | 0.506252 | 0.55298 | | 4.938767 | -0.06655 | 0.967145 |
| RBBP7 | 8.488061 | -0.06006 | 0.999532 | | 11.13533 | 0.356846 | 0.55298 | | 9.313894 | 0.4356 | 0.519883 |
| PPP2CA | 7.365528 | -0.06089 | 0.999532 | | 9.553739 | 0.414631 | 0.55298 | | 8.077687 | 0.491396 | 0.536234 |
| NDUFAF8 | 3.035498 | 0.104122 | 0.999532 | | 4.257201 | 0.522854 | 0.55298 | | 3.589301 | 0.438007 | 0.699971 |
| LRPPRC | 7.080522 | 0.094063 | 0.999532 | | 8.841249 | 0.392441 | 0.55298 | | 7.653115 | 0.319902 | 0.695561 |
| SRM | 2.541425 | -0.20931 | 0.999532 | | 2.408097 | -0.69036 | 0.55298 | | 1.962711 | -0.46465 | 0.772323 |
| UBR4 | 5.631898 | -0.01372 | 0.999532 | | 7.325532 | 0.429663 | 0.55298 | | 6.094439 | 0.458057 | 0.595502 |
| DDI2 | 3.482275 | 0.088522 | 0.999532 | | 4.846911 | 0.51224 | 0.553321 | | 4.162747 | 0.450563 | 0.686214 |
| DUSP3 | 7.126926 | -0.00383 | 0.999532 | | 9.038575 | 0.396053 | 0.553321 | | 7.695173 | 0.423757 | 0.566812 |
| STING1 | 6.517521 | 0.161128 | 0.999532 | | 7.807003 | 0.464463 | 0.553321 | | 7.212138 | 0.306912 | 0.758124 |
| NME2 | 15.66987 | 0.09833 | 0.999532 | | 19.95505 | 0.439091 | 0.553321 | | 17.70442 | 0.351463 | 0.655993 |
| RPS14 | 29.66136 | -0.14577 | 0.999532 | | 38.46275 | 0.295566 | 0.554651 | | 31.31573 | 0.45311 | 0.450852 |
| TRIM22 | 4.823662 | -0.20961 | 0.999532 | | 5.146998 | -0.47806 | 0.554675 | | 3.986694 | -0.24822 | 0.839811 |
| PDLIM2 | 10.34681 | 0.372919 | 0.999532 | | 11.96063 | 0.406044 | 0.554675 | | 11.30183 | 0.054119 | 0.967577 |
| PPP1R21 | 3.253867 | 0.23574 | 0.999532 | | 4.285355 | 0.561056 | 0.554675 | | 3.72676 | 0.341763 | 0.777828 |
| ARMC5 | 3.433169 | -0.74706 | 0.999532 | | 4.087148 | -0.58204 | 0.555726 | | 2.525132 | 0.183918 | 0.914812 |
| MRPS16 | 4.639893 | -0.06359 | 0.999532 | | 6.271145 | 0.446448 | 0.555726 | | 5.211636 | 0.530635 | 0.549065 |
| ZNF865 | 2.567581 | -0.36345 | 0.999532 | | 2.657848 | -0.64804 | 0.555726 | | 1.970769 | -0.26786 | 0.881837 |
| EBNA1BP | 3.954655 | 0.492034 | 0.999532 | | 4.560152 | 0.522606 | 0.555726 | | 4.411134 | 0.049686 | 0.978229 |
| ZNF484 | 3.088579 | -0.04695 | 0.999532 | | 3.037005 | -0.63093 | 0.556321 | | 2.537311 | -0.56885 | 0.663009 |
| TIMM23 | 5.19742 | 0.768298 | 0.999532 | | 5.176753 | 0.524256 | 0.556321 | | 5.733136 | -0.22223 | 0.850451 |
| EXTL3 | 5.235503 | -0.04311 | 0.999532 | | 6.854174 | 0.436134 | 0.556321 | | 5.735505 | 0.494075 | 0.57926 |
| PSMD3 | 7.453669 | -0.12943 | 0.999532 | | 10.05084 | 0.359589 | 0.556321 | | 8.057718 | 0.504691 | 0.454223 |
| KIF14 | 2.280316 | 0.177237 | 0.999532 | | 3.191088 | 0.752965 | 0.556321 | | 2.844515 | 0.588522 | 0.712556 |
| ATF7 | 5.136887 | 0.322335 | 0.999532 | | 5.908027 | 0.474074 | 0.556781 | | 5.682129 | 0.166295 | 0.893853 |
| SAMD14 | 4.389455 | 0.038884 | 0.999532 | | 4.088454 | -0.57586 | 0.556947 | | 3.600404 | -0.59942 | 0.566812 |
| CTNNA1 | 21.72407 | 0.039579 | 0.999532 | | 26.90746 | 0.330099 | 0.557402 | | 23.20492 | 0.308168 | 0.68597 |
| GPX2 | 2.006866 | -0.35779 | 0.999532 | | 2.106679 | -0.75892 | 0.557666 | | 1.633923 | -0.38097 | 0.849682 |
| MTMR6 | 4.181535 | 0.085651 | 0.999532 | | 5.2119 | 0.479298 | 0.557729 | | 4.719098 | 0.419376 | 0.678492 |
| SMIM20 | 2.417086 | 0.925323 | 0.999532 | | 2.577591 | 0.678653 | 0.557782 | | 2.878889 | -0.22648 | 0.886495 |
| ARHGDIA | 14.59607 | 0.04459 | 0.999532 | | 17.68894 | 0.334235 | 0.557782 | | 15.37432 | 0.303202 | 0.66232 |
| JARID2 | 4.72324 | -0.2324 | 0.999532 | | 4.808378 | -0.52098 | 0.557782 | | 3.774091 | -0.27083 | 0.842754 |
| WSB1 | 10.95233 | 0.016266 | 0.999532 | | 13.97515 | 0.336833 | 0.558323 | | 11.96464 | 0.339871 | 0.61086 |
| KIR2DS2 | 2.903333 | -0.09405 | 0.999532 | | 2.833771 | -0.72171 | 0.558323 | | 2.380387 | -0.61684 | 0.672279 |
| CD164 | 7.08372 | -0.01723 | 0.999532 | | 8.905776 | 0.398778 | 0.558432 | | 7.699795 | 0.419872 | 0.632464 |
| FRA10AC | 4.575894 | 0.454682 | 0.999532 | | 5.266546 | 0.483872 | 0.558436 | | 5.174378 | 0.047331 | 0.977473 |
| STEEP1 | 2.047284 | 0.597959 | 0.999532 | | 2.391878 | 0.666303 | 0.558436 | | 2.327867 | 0.085257 | 0.969693 |
| CPT1A | 6.661673 | 0.284923 | 0.999532 | | 8.030236 | 0.445483 | 0.558436 | | 7.435302 | 0.178665 | 0.864923 |
| LEPROTL | 5.234719 | -0.43077 | 0.999532 | | 7.559133 | 0.409645 | 0.558436 | | 5.735415 | 0.857337 | 0.209531 |
| MALSU1 | 3.888532 | 0.650312 | 0.999532 | | 4.348864 | 0.524897 | 0.558436 | | 4.386971 | -0.1087 | 0.942743 |
| EIF2D | 1.803593 | 0.881324 | 0.999532 | | 2.117004 | 0.760975 | 0.558869 | | 2.177555 | -0.09728 | 0.966652 |
| RAB33A | 2.156642 | 0.149469 | 0.999532 | | 1.921043 | -0.83286 | 0.558932 | | 1.674028 | -0.95754 | 0.539079 |
| ZNF512 | 3.753064 | 0.699372 | 0.999532 | | 3.665234 | 0.558479 | 0.558932 | | 4.167359 | -0.13057 | 0.933644 |
| RAPH1 | 9.96027 | -0.0184 | 0.999532 | | 13.1838 | 0.445782 | 0.558932 | | 11.32032 | 0.483164 | 0.486743 |
| H4C9 | 3.294322 | 0.29518 | 0.999532 | | 4.020326 | 0.5564 | 0.558932 | | 3.756768 | 0.282386 | 0.839373 |
| VPS13D | 3.95794 | 0.34298 | 0.999532 | | 4.609147 | 0.507827 | 0.558932 | | 4.561186 | 0.190005 | 0.887703 |
| FDXACB1 | 3.101216 | -0.51756 | 0.999532 | | 3.517514 | -0.6001 | 0.558934 | | 2.382681 | -0.05756 | 0.980456 |
| ID1 | 22.94307 | -0.06751 | 0.999532 | | 29.23817 | 0.299485 | 0.55913 | | 24.48478 | 0.389469 | 0.533876 |
| SLC50A1 | 6.561333 | 0.211922 | 0.999532 | | 5.650707 | -0.49308 | 0.55913 | | 5.585193 | -0.67515 | 0.421517 |
| GIMAP7 | 13.34415 | 0.132506 | 0.999532 | | 16.27252 | 0.371365 | 0.55913 | | 14.68841 | 0.260539 | 0.740755 |
| NXPH4 | 2.770791 | -0.23822 | 0.999532 | | 2.693987 | -0.70769 | 0.55913 | | 2.055353 | -0.44939 | 0.787995 |
| BIRC6 | 4.665451 | -0.50423 | 0.999532 | | 7.055639 | 0.438682 | 0.55913 | | 5.266923 | 0.956576 | 0.200192 |
| C16orf86 | 3.468081 | 0.178952 | 0.999532 | | 3.050716 | -0.6913 | 0.55913 | | 2.801447 | -0.85478 | 0.486743 |
| TSPAN33 | 2.069959 | 0.586102 | 0.999532 | | 2.474786 | 0.67499 | 0.559287 | | 2.443773 | 0.107433 | 0.956322 |
| SORD | 5.130983 | -0.30278 | 0.999532 | | 5.429698 | -0.4814 | 0.559588 | | 4.066266 | -0.16215 | 0.905746 |
| TFAP4 | 2.877193 | -0.02783 | 0.999532 | | 2.906156 | -0.68771 | 0.560333 | | 2.186779 | -0.63515 | 0.651876 |
| TCF7L1 | 7.792979 | -0.20683 | 0.999532 | | 10.81731 | 0.367811 | 0.560333 | | 8.344275 | 0.586488 | 0.415282 |
| JOSD2 | 3.633985 | -0.3395 | 0.999532 | | 5.20204 | 0.462982 | 0.560333 | | 4.157604 | 0.816567 | 0.339543 |
| FGD1 | 2.865761 | -0.83467 | 0.999532 | | 3.311067 | -0.5926 | 0.560333 | | 2.179417 | 0.253299 | 0.88885 |
| FLT3LG | 3.587537 | 0.243973 | 0.999532 | | 2.939761 | -0.60885 | 0.560333 | | 2.937106 | -0.84451 | 0.439339 |
| CCDC192 | 2.064645 | -0.05511 | 0.999532 | | 1.829866 | -0.78047 | 0.560494 | | 1.622 | -0.70956 | 0.666492 |
| NAT8 | 10.57596 | -0.09517 | 0.999532 | | 14.17342 | 0.494582 | 0.560494 | | 11.92882 | 0.573798 | 0.552672 |
| PAIP2B | 2.557391 | -0.30453 | 0.999532 | | 2.466017 | -0.6972 | 0.561601 | | 1.914111 | -0.37801 | 0.836453 |
| IGSF10 | 2.210201 | 0.101766 | 0.999532 | | 1.970644 | -0.78278 | 0.561601 | | 1.734896 | -0.86491 | 0.563455 |
| EIF2S3 | 5.625671 | 0.179534 | 0.999532 | | 7.279308 | 0.464151 | 0.561601 | | 6.222003 | 0.328304 | 0.759304 |
| ZYX | 10.20457 | -0.19159 | 0.999532 | | 14.06531 | 0.405762 | 0.562703 | | 11.32845 | 0.615714 | 0.386321 |
| EML3 | 3.380273 | 0.447922 | 0.999532 | | 3.993092 | 0.566973 | 0.563063 | | 3.916292 | 0.13822 | 0.92697 |
| NRDC | 4.523561 | -0.06782 | 0.999532 | | 5.946286 | 0.483786 | 0.563063 | | 4.916587 | 0.565409 | 0.563885 |
| IGHMBP2 | 3.243762 | 0.008676 | 0.999532 | | 4.421908 | 0.509264 | 0.563139 | | 3.72355 | 0.517717 | 0.628895 |
| ALG14 | 3.822356 | -0.12101 | 0.999532 | | 3.621413 | -0.57062 | 0.563139 | | 2.956756 | -0.43852 | 0.737426 |
| FAM72B | 7.476804 | -0.15956 | 0.999532 | | 7.497725 | -0.59677 | 0.563139 | | 6.088313 | -0.44462 | 0.707142 |
| CC2D2A | 2.145469 | 0.546847 | 0.999532 | | 2.534736 | 0.670476 | 0.563139 | | 2.519712 | 0.141713 | 0.940162 |
| TGFB1 | 6.480762 | 0.119174 | 0.999532 | | 8.1151 | 0.416032 | 0.563139 | | 7.272771 | 0.320043 | 0.712888 |
| AP1M1 | 4.286792 | 0.561469 | 0.999532 | | 4.588574 | 0.522739 | 0.563139 | | 4.740743 | -0.02097 | 0.992459 |
| GHITM | 6.751293 | 0.250012 | 0.999532 | | 7.784663 | 0.396126 | 0.563175 | | 7.204061 | 0.162576 | 0.874653 |
| RPIA | 2.411528 | -0.21874 | 0.999532 | | 2.278251 | -0.73514 | 0.563184 | | 1.806962 | -0.50204 | 0.773215 |
| RNPEP | 8.596471 | -0.01633 | 0.999532 | | 10.98741 | 0.362837 | 0.563187 | | 9.182666 | 0.390773 | 0.630705 |
| EEF1AKM | 2.669563 | 0.125034 | 0.999532 | | 2.290028 | -0.70077 | 0.563187 | | 2.094182 | -0.81149 | 0.543177 |
| SFT2D1 | 5.460401 | -0.02417 | 0.999532 | | 7.023387 | 0.412117 | 0.563232 | | 6.092917 | 0.453367 | 0.598329 |
| KRTAP5-1 | 3.126604 | 0.363809 | 0.999532 | | 2.630122 | -0.698 | 0.563238 | | 2.570662 | -1.04564 | 0.323635 |
| CTR9 | 5.134054 | 0.390677 | 0.999532 | | 5.789626 | 0.467762 | 0.563238 | | 5.599738 | 0.098196 | 0.940945 |
| CMBL | 3.476139 | 0.416429 | 0.999532 | | 4.221906 | 0.583948 | 0.563537 | | 3.813319 | 0.170189 | 0.905746 |
| GNGT2 | 2.026408 | 0.68203 | 0.999532 | | 2.321448 | 0.721397 | 0.563537 | | 2.367713 | 0.05794 | 0.981739 |
| MRPS18C | 6.39627 | -0.21052 | 0.999532 | | 6.599665 | -0.44893 | 0.563537 | | 5.273172 | -0.22198 | 0.858899 |
| CSTF1 | 2.548078 | 0.593842 | 0.999532 | | 2.877829 | 0.635807 | 0.563537 | | 3.12879 | 0.065228 | 0.975284 |
| SMDT1 | 10.42336 | 0.294537 | 0.999532 | | 11.85071 | 0.367749 | 0.563537 | | 10.8519 | 0.096981 | 0.926513 |
| HOXB3 | 15.88958 | 0.081781 | 0.999532 | | 15.4457 | -0.35156 | 0.563537 | | 13.64293 | -0.41122 | 0.53177 |
| SIPA1L3 | 4.679443 | 0.525815 | 0.999532 | | 4.907336 | 0.521724 | 0.563537 | | 5.028072 | 0.014193 | 0.996681 |
| BANF2 | 2.861957 | 0.37611 | 0.999532 | | 2.216002 | -0.73461 | 0.563537 | | 2.349955 | -1.09918 | 0.359441 |
| CYBC1 | 4.788771 | 0.258995 | 0.999532 | | 6.19692 | 0.457832 | 0.563537 | | 5.621941 | 0.214262 | 0.846155 |
| OFD1 | 5.335867 | -0.08515 | 0.999532 | | 5.052397 | -0.48085 | 0.563537 | | 4.480391 | -0.3863 | 0.734276 |
| TINAG | 3.041441 | -0.103 | 0.999532 | | 4.188935 | 0.547113 | 0.563537 | | 3.466613 | 0.669892 | 0.53177 |
| FAM162A | 5.821254 | -0.04701 | 0.999532 | | 7.688896 | 0.435171 | 0.563631 | | 6.378755 | 0.508585 | 0.541754 |
| PCGF1 | 2.921751 | 0.44919 | 0.999532 | | 3.738109 | 0.592275 | 0.563643 | | 3.666757 | 0.158911 | 0.918694 |
| ZDHHC13 | 2.577654 | -0.62596 | 0.999532 | | 2.726614 | -0.667 | 0.563643 | | 1.827389 | -0.02167 | 0.996681 |
| SV2B | 2.607757 | -0.1963 | 0.999532 | | 2.560742 | -0.69794 | 0.563643 | | 2.054135 | -0.48529 | 0.75713 |
| AMZ2 | 4.427204 | 0.278826 | 0.999532 | | 5.598385 | 0.505153 | 0.563643 | | 5.155204 | 0.244953 | 0.839373 |
| PSMB4 | 8.659434 | -0.14227 | 0.999532 | | 11.94448 | 0.35955 | 0.563643 | | 9.644289 | 0.522463 | 0.431254 |
| C20orf202 | 2.654477 | -0.25007 | 0.999532 | | 2.759597 | -0.6628 | 0.563643 | | 2.14709 | -0.39491 | 0.809832 |
| RASGRP | 6.525194 | 0.17797 | 0.999532 | | 7.85616 | 0.40377 | 0.563643 | | 7.25492 | 0.243365 | 0.806662 |
| KANK1 | 5.351355 | 0.108845 | 0.999532 | | 6.687531 | 0.43665 | 0.563643 | | 5.775621 | 0.347264 | 0.718643 |
| SFXN1 | 2.501207 | 0.046005 | 0.999532 | | 3.626942 | 0.62335 | 0.564395 | | 3.193353 | 0.576299 | 0.650381 |
| AARS1 | 4.986753 | 0.170987 | 0.999532 | | 6.268835 | 0.465161 | 0.564395 | | 5.691297 | 0.310004 | 0.764149 |
| XRCC6 | 9.68575 | -0.28689 | 0.999532 | | 13.71135 | 0.32457 | 0.564395 | | 10.4268 | 0.629407 | 0.282041 |
| FZD5 | 2.777631 | -0.40711 | 0.999532 | | 2.893613 | -0.64118 | 0.564395 | | 2.223701 | -0.21876 | 0.908274 |
| NFIA | 3.078101 | -0.00489 | 0.999532 | | 4.004945 | 0.524527 | 0.564395 | | 3.462161 | 0.538345 | 0.644433 |
| NUDT6 | 2.438126 | 0.036968 | 0.999532 | | 2.267941 | -0.72436 | 0.564395 | | 1.890295 | -0.742 | 0.61123 |
| KCNV1 | 2.354242 | 0.167333 | 0.999532 | | 1.993192 | -0.79638 | 0.564395 | | 1.857274 | -0.94372 | 0.515762 |
| ZDHHC16 | 3.159302 | -0.52707 | 0.999532 | | 3.547941 | -0.57238 | 0.564395 | | 2.619087 | -0.02545 | 0.993618 |
| TNFRSF1 | 3.671137 | -0.30191 | 0.999532 | | 3.61616 | -0.59983 | 0.564395 | | 2.860271 | -0.28117 | 0.864245 |
| LCP2 | 6.669068 | -0.32948 | 0.999532 | | 7.070302 | -0.42315 | 0.564395 | | 5.398992 | -0.07934 | 0.955834 |
| NONO | 12.65266 | 0.006145 | 0.999532 | | 16.08851 | 0.321062 | 0.564781 | | 13.54424 | 0.339031 | 0.622543 |
| GNB3 | 2.239668 | -0.18779 | 0.999532 | | 2.175459 | -0.72511 | 0.564781 | | 1.706681 | -0.51719 | 0.758124 |
| PPT2 | 7.16713 | 0.315387 | 0.999532 | | 8.281492 | 0.41143 | 0.564835 | | 7.792906 | 0.117343 | 0.920527 |
| C9orf40 | 5.826833 | 0.151055 | 0.999532 | | 5.460717 | -0.52387 | 0.565668 | | 4.944573 | -0.65883 | 0.484303 |
| SNX1 | 6.18501 | -0.08628 | 0.999532 | | 8.061831 | 0.411001 | 0.56576 | | 6.571329 | 0.522634 | 0.510412 |
| FTCD | 4.565599 | -0.04035 | 0.999532 | | 4.473606 | -0.52937 | 0.56576 | | 3.722228 | -0.46883 | 0.682989 |
| C17orf58 | 3.728282 | -0.32677 | 0.999532 | | 3.751232 | -0.58381 | 0.56576 | | 2.817637 | -0.24064 | 0.881771 |
| SEC14L4 | 3.520972 | 0.353363 | 0.999532 | | 3.037457 | -0.6336 | 0.56576 | | 3.039998 | -0.96975 | 0.333968 |
| MEST | 6.820569 | -0.36063 | 0.999532 | | 7.964833 | -0.43764 | 0.56576 | | 5.726548 | -0.05497 | 0.974421 |
| ECPAS | 5.03988 | 0.219155 | 0.999532 | | 6.564214 | 0.447421 | 0.56605 | | 5.906049 | 0.25256 | 0.813143 |
| AUNIP | 2.501556 | 0.217874 | 0.999532 | | 2.110065 | -0.74299 | 0.56605 | | 2.050167 | -0.94052 | 0.504265 |
| RMDN2 | 4.340372 | -0.04458 | 0.999532 | | 3.808556 | -0.5406 | 0.56605 | | 3.390163 | -0.49953 | 0.682165 |
| BRINP1 | 3.121165 | 0.157391 | 0.999532 | | 2.633756 | -0.66969 | 0.56605 | | 2.468089 | -0.81364 | 0.520407 |
| LARP6 | 2.433058 | -0.69242 | 0.999532 | | 2.701022 | -0.64085 | 0.56605 | | 1.763316 | 0.066258 | 0.979434 |
| FZD9 | 2.880639 | 0.13697 | 0.999532 | | 2.37185 | -0.6744 | 0.56605 | | 2.320304 | -0.79924 | 0.540342 |
| SULT1C4 | 3.422886 | 0.167645 | 0.999532 | | 4.401254 | 0.532114 | 0.56605 | | 3.774717 | 0.374527 | 0.75311 |
| AFAP1L2 | 7.33843 | 0.152659 | 0.999532 | | 8.690318 | 0.387253 | 0.56605 | | 7.729252 | 0.251377 | 0.78258 |
| LPP | 16.77237 | 0.31381 | 0.999532 | | 18.88432 | 0.323504 | 0.56605 | | 18.1247 | 0.025619 | 0.984624 |
| LGALS3 | 6.791613 | -0.08836 | 0.999532 | | 9.110354 | 0.444991 | 0.56605 | | 7.585173 | 0.54615 | 0.544921 |
| MSX1 | 3.473085 | -0.21889 | 0.999532 | | 3.267502 | -0.64514 | 0.56605 | | 2.587137 | -0.40035 | 0.791466 |
| ABCG1 | 4.004142 | -0.06169 | 0.999532 | | 5.339245 | 0.510757 | 0.56605 | | 4.429051 | 0.585527 | 0.574633 |
| TSHZ1 | 4.465238 | 0.511867 | 0.999532 | | 5.132657 | 0.517522 | 0.56605 | | 5.025705 | 0.030197 | 0.989148 |
| OR13C9 | 2.559499 | 0.087859 | 0.999532 | | 2.377383 | -0.6863 | 0.56605 | | 2.073601 | -0.75253 | 0.572297 |
| DMRTC2 | 4.824641 | 0.176804 | 0.999532 | | 5.974646 | 0.464167 | 0.56605 | | 5.294188 | 0.306528 | 0.764702 |
| RPL18A | 24.87782 | 0.150817 | 0.999532 | | 28.98664 | 0.278791 | 0.56697 | | 26.23908 | 0.14332 | 0.826947 |
| CBLN3 | 4.378766 | 0.044902 | 0.999532 | | 4.035079 | -0.55117 | 0.56697 | | 3.679495 | -0.58323 | 0.611214 |
| ACYP2 | 3.347799 | -0.41694 | 0.999532 | | 3.602934 | -0.57268 | 0.56697 | | 2.603539 | -0.13631 | 0.941197 |
| HEXIM1 | 12.76535 | 0.086522 | 0.999532 | | 16.67808 | 0.395721 | 0.56697 | | 14.43625 | 0.341426 | 0.672452 |
| EIF4EBP2 | 5.788839 | -0.16693 | 0.999532 | | 7.876336 | 0.399746 | 0.56697 | | 6.558311 | 0.584547 | 0.445661 |
| LDB1 | 6.963052 | 0.042712 | 0.999532 | | 8.957506 | 0.384324 | 0.56697 | | 7.694149 | 0.361565 | 0.647218 |
| FOXC1 | 6.525666 | 0.202829 | 0.999532 | | 7.894433 | 0.453896 | 0.56697 | | 7.065257 | 0.272132 | 0.791576 |
| TRIP6 | 3.666466 | 0.25601 | 0.999532 | | 4.256103 | 0.515719 | 0.56697 | | 3.925772 | 0.275325 | 0.824012 |
| SOD1 | 14.31949 | -0.05242 | 0.999532 | | 18.97207 | 0.338532 | 0.56697 | | 15.53817 | 0.407858 | 0.533876 |
| CALD1 | 34.08684 | -0.22107 | 0.999532 | | 46.76221 | 0.346732 | 0.56697 | | 37.68045 | 0.590913 | 0.154583 |
| TMEM106 | 2.652576 | 0.535164 | 0.999532 | | 2.858217 | 0.599396 | 0.56697 | | 2.962019 | 0.071857 | 0.973224 |
| RNF213 | 6.771006 | -0.18557 | 0.999532 | | 9.509794 | 0.389615 | 0.567782 | | 7.531442 | 0.590948 | 0.414154 |
| ZNF613 | 2.251562 | -0.13623 | 0.999532 | | 2.182821 | -0.72664 | 0.568699 | | 1.757607 | -0.57121 | 0.722703 |
| FASTKD2 | 2.680747 | 0.066227 | 0.999532 | | 3.506533 | 0.571112 | 0.568755 | | 3.14991 | 0.52321 | 0.666906 |
| WDR55 | 3.591212 | 0.001407 | 0.999532 | | 3.2424 | -0.58059 | 0.568755 | | 2.909359 | -0.56756 | 0.647581 |
| CD99L2 | 3.909241 | 0.126171 | 0.999532 | | 4.92092 | 0.474304 | 0.568755 | | 4.424965 | 0.362653 | 0.735469 |
| MYT1 | 3.07997 | 0.06583 | 0.999532 | | 2.869933 | -0.68624 | 0.568755 | | 2.336438 | -0.72738 | 0.573429 |
| ZGPAT | 3.251779 | 0.166034 | 0.999532 | | 2.705598 | -0.63199 | 0.568755 | | 2.597189 | -0.7896 | 0.523065 |
| NF2 | 4.78911 | -0.31274 | 0.999532 | | 5.24378 | -0.51141 | 0.568755 | | 3.98991 | -0.16979 | 0.909157 |
| ANXA2R | 2.655956 | -0.35442 | 0.999532 | | 2.658309 | -0.66163 | 0.568755 | | 2.042751 | -0.28639 | 0.877703 |
| SF3B3 | 8.194031 | 0.106176 | 0.999532 | | 10.20259 | 0.39653 | 0.568755 | | 8.910953 | 0.313527 | 0.707745 |
| CORO7 | 2.725201 | -0.55212 | 0.999532 | | 3.074819 | -0.62022 | 0.568755 | | 2.197276 | -0.04884 | 0.987164 |
| ACE2 | 2.119783 | 0.861278 | 0.999532 | | 2.274201 | 0.740301 | 0.568755 | | 2.487284 | -0.09485 | 0.968885 |
| FAT1 | 8.829899 | 0.051231 | 0.999532 | | 11.0434 | 0.38682 | 0.568755 | | 9.377402 | 0.351859 | 0.678492 |
| PIK3R4 | 2.61877 | 0.244522 | 0.999532 | | 3.316534 | 0.586477 | 0.568755 | | 2.931414 | 0.360519 | 0.791576 |
| PRAF2 | 2.204065 | 0.203604 | 0.999532 | | 1.929681 | -0.78374 | 0.568755 | | 1.635822 | -0.96491 | 0.508751 |
| MAPK11 | 2.390278 | 0.346829 | 0.999532 | | 2.998628 | 0.631053 | 0.568918 | | 2.955555 | 0.300216 | 0.847417 |
| YIPF1 | 3.313755 | -0.66416 | 0.999532 | | 3.907213 | -0.56411 | 0.569168 | | 2.370544 | 0.121574 | 0.951926 |
| MTRNR2L | 2.405705 | 0.276321 | 0.999532 | | 1.923914 | -0.75732 | 0.569168 | | 1.999922 | -1.01957 | 0.451774 |
| CSHL1 | 3.082039 | -0.59016 | 0.999532 | | 3.586495 | -0.61979 | 0.569168 | | 2.520463 | -0.01144 | 0.996891 |
| AQP6 | 2.16324 | -0.23703 | 0.999532 | | 2.084579 | -0.74917 | 0.569168 | | 1.571653 | -0.49308 | 0.785012 |
| IL18R1 | 3.853165 | -0.60929 | 0.999532 | | 4.452871 | -0.58138 | 0.570295 | | 2.956431 | 0.038179 | 0.988379 |
| NKX1-1 | 3.109172 | -0.28143 | 0.999532 | | 3.307345 | -0.56701 | 0.570295 | | 2.445045 | -0.25887 | 0.87591 |
| TRIP12 | 12.33322 | 0.025432 | 0.999532 | | 15.26714 | 0.319041 | 0.570714 | | 13.1483 | 0.314227 | 0.645127 |
| SHROOM | 3.414231 | -0.26409 | 0.999532 | | 5.012561 | 0.49078 | 0.570714 | | 3.983603 | 0.775131 | 0.413122 |
| CYP20A1 | 3.806773 | 0.194234 | 0.999532 | | 5.136256 | 0.511448 | 0.570965 | | 4.376694 | 0.355829 | 0.760473 |
| RALGDS | 3.713481 | 0.483106 | 0.999532 | | 4.260681 | 0.508589 | 0.570965 | | 4.310404 | 0.045899 | 0.978931 |
| HERPUD1 | 10.72932 | 0.369709 | 0.999532 | | 11.93442 | 0.345806 | 0.570965 | | 11.46494 | -0.00568 | 0.996891 |
| PPP1CB | 16.14728 | -0.22524 | 0.999532 | | 21.72319 | 0.312968 | 0.570965 | | 16.81283 | 0.556156 | 0.363418 |
| MBOAT2 | 3.138425 | 0.198773 | 0.999532 | | 3.787317 | 0.552354 | 0.571172 | | 3.604304 | 0.364319 | 0.791466 |
| RBM11 | 2.293328 | -0.04021 | 0.999532 | | 2.232342 | -0.71807 | 0.571172 | | 1.791827 | -0.64665 | 0.682165 |
| SNX29 | 3.353433 | 0.233964 | 0.999532 | | 4.367297 | 0.533806 | 0.571476 | | 3.623553 | 0.298594 | 0.824012 |
| TEAD1 | 5.697108 | 0.082445 | 0.999532 | | 7.008824 | 0.424481 | 0.571476 | | 6.09826 | 0.359756 | 0.700565 |
| TWIST1 | 2.937362 | -0.40803 | 0.999532 | | 3.029041 | -0.62825 | 0.572149 | | 2.375009 | -0.20721 | 0.914692 |
| UNC45A | 4.979448 | 0.212207 | 0.999532 | | 6.341561 | 0.434363 | 0.572186 | | 5.609512 | 0.243552 | 0.821463 |
| MAEA | 4.779931 | -0.0969 | 0.999532 | | 6.409433 | 0.431114 | 0.572186 | | 5.255154 | 0.545714 | 0.533876 |
| BAIAP2 | 3.485025 | -0.36288 | 0.999532 | | 3.540144 | -0.60515 | 0.572186 | | 2.508166 | -0.2281 | 0.893452 |
| ABR | 5.287534 | -0.24018 | 0.999532 | | 7.672419 | 0.387769 | 0.572186 | | 6.105221 | 0.651224 | 0.37596 |
| RAB29 | 2.414979 | 0.672998 | 0.999532 | | 2.490801 | 0.65805 | 0.572186 | | 2.725082 | 0.001203 | 0.99959 |
| DUSP6 | 10.9058 | -0.63107 | 0.871349 | | 13.36467 | -0.35053 | 0.572186 | | 8.893277 | 0.308732 | 0.717627 |
| SCGB2B2 | 2.791103 | -0.285 | 0.999532 | | 2.976144 | -0.6344 | 0.57229 | | 2.223663 | -0.32889 | 0.844187 |
| C17orf49 | 5.237825 | -0.01094 | 0.999532 | | 6.813191 | 0.419915 | 0.572457 | | 6.012482 | 0.454706 | 0.622543 |
| LIX1L | 5.696139 | -0.0252 | 0.999532 | | 7.434891 | 0.407574 | 0.572838 | | 6.282397 | 0.45348 | 0.570037 |
| ACOT8 | 3.093157 | 0.27615 | 0.999532 | | 3.849711 | 0.523369 | 0.57368 | | 3.548278 | 0.265255 | 0.837256 |
| SPOPL | 2.444751 | 0.410769 | 0.999532 | | 3.182918 | 0.61569 | 0.573765 | | 2.881791 | 0.227895 | 0.88588 |
| TOR1AIP | 4.045374 | -0.20571 | 0.999532 | | 5.831295 | 0.469622 | 0.573765 | | 4.789859 | 0.696339 | 0.445382 |
| UBE2S | 6.023875 | -0.05754 | 0.999532 | | 6.035399 | -0.48619 | 0.573765 | | 4.845826 | -0.40029 | 0.703463 |
| SEMA5A | 12.77398 | 0.278012 | 0.999532 | | 14.8373 | 0.424408 | 0.573765 | | 13.86926 | 0.170441 | 0.869378 |
| PAPLN | 5.914842 | 0.116503 | 0.999532 | | 7.701751 | 0.425241 | 0.573765 | | 6.505443 | 0.338447 | 0.730921 |
| DLX6 | 3.023941 | 0.155555 | 0.999532 | | 2.788356 | -0.65864 | 0.573765 | | 2.541349 | -0.79228 | 0.532055 |
| SYPL2 | 2.87475 | -0.05786 | 0.999532 | | 2.663668 | -0.63317 | 0.573765 | | 2.371156 | -0.56349 | 0.669183 |
| BNIP5 | 2.338185 | -0.02342 | 0.999532 | | 2.110066 | -0.73073 | 0.573765 | | 1.914392 | -0.69548 | 0.665886 |
| SLC2A5 | 3.910619 | 0.03276 | 0.999532 | | 3.476447 | -0.56793 | 0.573765 | | 3.151324 | -0.58352 | 0.63352 |
| HMBS | 2.740919 | -0.04829 | 0.999532 | | 2.734318 | -0.67205 | 0.573765 | | 2.194105 | -0.60032 | 0.677493 |
| TMEM239 | 3.071368 | 0.036743 | 0.999532 | | 2.765573 | -0.63311 | 0.573765 | | 2.478366 | -0.65343 | 0.628895 |
| SLC27A1 | 3.768675 | 0.050419 | 0.999532 | | 3.616807 | -0.55477 | 0.573765 | | 3.204587 | -0.58292 | 0.623462 |
| SMIM12 | 3.333168 | -0.214 | 0.999532 | | 3.297547 | -0.58985 | 0.573834 | | 2.556213 | -0.35826 | 0.809832 |
| S100A9 | 5.286885 | -0.50951 | 0.999532 | | 5.365411 | -0.79507 | 0.573937 | | 3.897446 | -0.21399 | 0.894612 |
| TMEM220 | 2.020452 | 0.305347 | 0.999532 | | 2.522946 | 0.65019 | 0.573937 | | 2.425203 | 0.362209 | 0.816589 |
| LORICRIN | 2.184539 | -0.18911 | 0.999532 | | 2.067651 | -0.76853 | 0.573937 | | 1.663392 | -0.56003 | 0.750371 |
| ACSS2 | 3.641109 | 0.269623 | 0.999532 | | 4.40516 | 0.48668 | 0.573937 | | 4.008222 | 0.231032 | 0.859426 |
| UBXN4 | 7.482981 | 0.083754 | 0.999532 | | 8.88705 | 0.384284 | 0.573937 | | 7.936056 | 0.319123 | 0.706407 |
| LMNA | 18.15193 | -0.64047 | 0.396029 | | 22.5021 | -0.34046 | 0.573937 | | 14.6675 | 0.347444 | 0.67937 |
| RNF111 | 3.311665 | 0.203198 | 0.999532 | | 4.219391 | 0.573022 | 0.573937 | | 3.796137 | 0.398543 | 0.743246 |
| SSH2 | 2.411697 | -0.05993 | 0.999532 | | 2.292355 | -0.70276 | 0.573949 | | 1.865622 | -0.62416 | 0.680577 |
| TUBA8 | 2.686625 | 0.066082 | 0.999532 | | 2.33377 | -0.70981 | 0.574148 | | 2.13124 | -0.7579 | 0.607748 |
| DGKZ | 5.346586 | 0.027026 | 0.999532 | | 7.166372 | 0.413655 | 0.574148 | | 6.334194 | 0.410173 | 0.643959 |
| CFD | 2.720602 | -0.2907 | 0.999532 | | 2.824268 | -0.63854 | 0.574285 | | 2.303596 | -0.33648 | 0.837256 |
| PSMA5 | 4.397135 | -0.00304 | 0.999532 | | 5.951852 | 0.437826 | 0.57433 | | 4.955942 | 0.458135 | 0.643427 |
| VASP | 9.84466 | -0.19158 | 0.999532 | | 13.49844 | 0.401384 | 0.576087 | | 11.28433 | 0.590482 | 0.437893 |
| HNRNPA2 | 45.76428 | -0.10564 | 0.999532 | | 58.05704 | 0.238695 | 0.576087 | | 47.15927 | 0.375991 | 0.413122 |
| DYNLL2 | 9.078831 | 0.113809 | 0.999532 | | 11.14482 | 0.366901 | 0.576148 | | 9.781944 | 0.272076 | 0.747457 |
| RPL35 | 33.32723 | 0.14647 | 0.999532 | | 40.22969 | 0.315176 | 0.577058 | | 35.75495 | 0.194 | 0.791576 |
| HIPK1 | 5.972597 | -0.01236 | 0.999532 | | 7.703302 | 0.421659 | 0.577058 | | 6.531109 | 0.450953 | 0.623286 |
| ZNF853 | 3.356253 | 0.147911 | 0.999532 | | 2.938749 | -0.61338 | 0.577135 | | 2.768157 | -0.74621 | 0.53177 |
| TMEM176 | 3.350676 | 0.430138 | 0.999532 | | 4.109948 | 0.589111 | 0.577202 | | 3.772931 | 0.174356 | 0.910105 |
| FASLG | 2.098217 | 0.066491 | 0.999532 | | 1.936793 | -0.79102 | 0.577246 | | 1.659478 | -0.83736 | 0.602786 |
| FRS3 | 2.721487 | -0.32863 | 0.999532 | | 2.776185 | -0.6292 | 0.577246 | | 2.082986 | -0.28749 | 0.876781 |
| AK2 | 6.292887 | -0.33491 | 0.999532 | | 9.025666 | 0.365155 | 0.577526 | | 6.902484 | 0.717303 | 0.254511 |
| GRAP2 | 4.173635 | 0.256636 | 0.999532 | | 3.340094 | -0.56133 | 0.578029 | | 3.41209 | -0.81549 | 0.445661 |
| MICOS10 | 9.298353 | 0.050095 | 0.999532 | | 11.85829 | 0.351277 | 0.578166 | | 10.37291 | 0.318187 | 0.669356 |
| ARPC1A | 4.053023 | -0.37719 | 0.999532 | | 6.051996 | 0.445571 | 0.578166 | | 4.557274 | 0.841043 | 0.280329 |
| CPEB2 | 4.23271 | 0.371574 | 0.999532 | | 5.035796 | 0.505308 | 0.578286 | | 4.898896 | 0.146289 | 0.915618 |
| IMPDH2 | 3.638417 | 0.386522 | 0.999532 | | 4.34168 | 0.526081 | 0.578286 | | 4.179977 | 0.154845 | 0.912799 |
| SPATA13 | 8.974361 | -0.08747 | 0.999532 | | 11.80628 | 0.345104 | 0.578286 | | 9.812126 | 0.452096 | 0.515762 |
| ZFYVE27 | 3.444262 | 0.184888 | 0.999532 | | 3.204665 | -0.56445 | 0.578464 | | 2.744716 | -0.71997 | 0.536289 |
| LRRC41 | 7.83619 | 0.151115 | 0.999532 | | 9.741493 | 0.360437 | 0.578464 | | 8.55181 | 0.232849 | 0.799944 |
| PTCRA | 2.3583 | -0.01951 | 0.999532 | | 2.055812 | -0.73074 | 0.578464 | | 1.953004 | -0.70251 | 0.66232 |
| ZNF728 | 2.048887 | -0.35469 | 0.999532 | | 2.007255 | -0.72381 | 0.578464 | | 1.532632 | -0.35597 | 0.862655 |
| NFATC3 | 4.712671 | 0.175708 | 0.999532 | | 5.421615 | 0.441961 | 0.578659 | | 5.083459 | 0.285226 | 0.791466 |
| PRKAR1A | 25.01254 | 0.03634 | 0.999532 | | 30.72547 | 0.271065 | 0.578848 | | 26.07672 | 0.255421 | 0.663089 |
| MARCHF | 5.661359 | 0.023129 | 0.999532 | | 7.076315 | 0.409893 | 0.578848 | | 6.19613 | 0.402619 | 0.658332 |
| TMEM44 | 5.137369 | -0.07298 | 0.999532 | | 4.735281 | -0.47837 | 0.578848 | | 4.054392 | -0.39714 | 0.718633 |
| TBKBP1 | 2.418088 | 0.164687 | 0.999532 | | 3.191881 | 0.591539 | 0.578848 | | 2.830242 | 0.449966 | 0.741408 |
| LARP1 | 4.907966 | -0.03292 | 0.999532 | | 4.619505 | -0.49872 | 0.578848 | | 3.909148 | -0.45517 | 0.67937 |
| BRSK2 | 2.659036 | -0.21716 | 0.999532 | | 2.723278 | -0.65847 | 0.578848 | | 2.227646 | -0.42317 | 0.795928 |
| PPP1R1B | 2.333842 | -0.5416 | 0.999532 | | 2.653859 | -0.66365 | 0.578848 | | 1.81677 | -0.0858 | 0.974421 |
| JMJD7 | 4.268706 | -0.19464 | 0.999532 | | 4.355968 | -0.50692 | 0.578848 | | 3.235798 | -0.29382 | 0.820977 |
| MCUR1 | 3.54214 | 0.365384 | 0.999532 | | 4.132343 | 0.5363 | 0.578848 | | 4.004877 | 0.188651 | 0.894404 |
| SAXO1 | 2.305498 | -0.01497 | 0.999532 | | 2.147226 | -0.68558 | 0.578848 | | 1.810011 | -0.65223 | 0.660958 |
| UBL5 | 10.15221 | 0.131867 | 0.999532 | | 12.32645 | 0.37037 | 0.578848 | | 10.69365 | 0.26243 | 0.749206 |
| INAVA | 3.606341 | 0.322101 | 0.999532 | | 3.091903 | -0.70875 | 0.578848 | | 2.97899 | -1.02678 | 0.342845 |
| PLIN4 | 2.960481 | 0.162023 | 0.999532 | | 2.802095 | -0.6525 | 0.579406 | | 2.453635 | -0.79466 | 0.515762 |
| CASZ1 | 3.35511 | 0.336445 | 0.999532 | | 4.445793 | 0.517981 | 0.57944 | | 4.040964 | 0.203137 | 0.881689 |
| RPL26 | 37.8757 | -0.1474 | 0.999532 | | 49.4539 | 0.290775 | 0.579533 | | 40.3048 | 0.458158 | 0.352276 |
| ADAM15 | 136.018 | -0.1865 | 0.999532 | | 180.4978 | 0.286705 | 0.579735 | | 145.4477 | 0.498775 | 0.452179 |
| AFG3L2 | 3.66251 | 0.333031 | 0.999532 | | 4.385919 | 0.543461 | 0.579887 | | 4.19218 | 0.227875 | 0.874653 |
| SPIN2A | 2.515706 | 0.806692 | 0.999532 | | 2.572044 | 0.657948 | 0.579887 | | 2.808705 | -0.13102 | 0.944993 |
| MED19 | 4.304654 | 0.08417 | 0.999532 | | 3.97244 | -0.54871 | 0.579887 | | 3.514328 | -0.61462 | 0.563455 |
| NOP53 | 12.05599 | -0.37254 | 0.999532 | | 16.77731 | 0.319203 | 0.580418 | | 12.91146 | 0.711366 | 0.150518 |
| PITX2 | 3.61753 | 0.281957 | 0.999532 | | 2.9642 | -0.63665 | 0.580418 | | 2.963498 | -0.90662 | 0.455196 |
| EPN1 | 8.747227 | 0.116405 | 0.999532 | | 10.98797 | 0.420237 | 0.580418 | | 9.324928 | 0.335605 | 0.663089 |
| SEC62 | 9.41771 | -0.03186 | 0.999532 | | 12.06301 | 0.342601 | 0.580778 | | 10.09289 | 0.395478 | 0.549815 |
| MAFK | 10.08579 | 0.390369 | 0.999532 | | 10.96492 | 0.356429 | 0.582085 | | 10.54867 | -0.01167 | 0.995112 |
| FNIP1 | 3.102492 | 0.369731 | 0.999532 | | 3.762951 | 0.547308 | 0.582085 | | 3.513257 | 0.195547 | 0.893828 |
| TH | 2.361926 | 0.277701 | 0.999532 | | 1.874595 | -0.78293 | 0.582258 | | 1.862283 | -1.04359 | 0.464674 |
| CHMP3 | 7.100168 | 0.406721 | 0.999532 | | 7.955159 | 0.405434 | 0.5825 | | 7.767624 | 0.025125 | 0.989437 |
| PSG3 | 3.156233 | -0.18935 | 0.999532 | | 3.195101 | -0.60185 | 0.58259 | | 2.550403 | -0.39244 | 0.79686 |
| LIMD1 | 5.473755 | 0.431729 | 0.999532 | | 4.417965 | -0.51778 | 0.58259 | | 4.693301 | -0.94071 | 0.209531 |
| ADCY6 | 7.163724 | 0.102753 | 0.999532 | | 8.666588 | 0.428576 | 0.58259 | | 7.746243 | 0.337052 | 0.724646 |
| DPM2 | 3.640107 | -0.19023 | 0.999532 | | 3.760186 | -0.54678 | 0.583122 | | 2.840529 | -0.33648 | 0.809832 |
| SUN1 | 7.567366 | 0.133558 | 0.999532 | | 9.253477 | 0.379172 | 0.583124 | | 8.128392 | 0.262504 | 0.764702 |
| CLIC5 | 61.96599 | 0.384705 | 0.999532 | | 69.15222 | 0.355026 | 0.583235 | | 66.16429 | -0.00285 | 0.99858 |
| CPNE8 | 5.995749 | 0.060181 | 0.999532 | | 5.444076 | -0.48558 | 0.583235 | | 4.75186 | -0.53264 | 0.600081 |
| OVOL2 | 2.630781 | 0.100857 | 0.999532 | | 2.341863 | -0.70124 | 0.584151 | | 2.092919 | -0.78323 | 0.566812 |
| RABGGT | 3.469276 | 0.568381 | 0.999532 | | 3.845304 | 0.53182 | 0.584151 | | 3.914573 | -0.01933 | 0.994192 |
| ZNF251 | 2.986808 | -0.3115 | 0.999532 | | 4.655803 | 0.513329 | 0.584151 | | 3.516739 | 0.839289 | 0.379978 |
| PYCR1 | 2.972498 | 0.062911 | 0.999532 | | 2.610124 | -0.67682 | 0.584151 | | 2.202158 | -0.71854 | 0.60629 |
| CCND1 | 12.35214 | -0.23183 | 0.999532 | | 17.26752 | 0.404915 | 0.584151 | | 14.06039 | 0.646015 | 0.295502 |
| CYB5B | 6.714561 | 0.630381 | 0.999532 | | 6.577886 | 0.426282 | 0.584151 | | 7.133713 | -0.19237 | 0.864245 |
| DNMT1 | 3.881407 | 0.525496 | 0.999532 | | 4.028231 | 0.532298 | 0.584153 | | 4.353176 | -0.00862 | 0.996893 |
| MRPL49 | 4.68905 | 0.017388 | 0.999532 | | 6.15294 | 0.422645 | 0.584825 | | 5.313454 | 0.425059 | 0.65401 |
| PPP1R15 | 4.475148 | -0.21445 | 0.999532 | | 6.266646 | 0.467456 | 0.584825 | | 4.918055 | 0.707519 | 0.386771 |
| RGN | 4.271914 | -0.17697 | 0.999532 | | 5.946331 | 0.440937 | 0.584989 | | 5.076986 | 0.638864 | 0.485816 |
| TLR2 | 4.723479 | 0.010897 | 0.999532 | | 4.626672 | -0.49504 | 0.584989 | | 3.922382 | -0.48579 | 0.647955 |
| CCR10 | 3.184079 | 0.291444 | 0.999532 | | 2.743565 | -0.65847 | 0.584989 | | 2.516191 | -0.93265 | 0.416169 |
| KDM4E | 2.276436 | 0.00084 | 0.999532 | | 1.909785 | -0.7458 | 0.585194 | | 1.799425 | -0.73391 | 0.654321 |
| AFM | 3.265473 | 0.396199 | 0.999532 | | 3.990262 | 0.665428 | 0.585194 | | 3.786657 | 0.294662 | 0.859522 |
| AJUBA | 3.230568 | -0.53667 | 0.999532 | | 3.63736 | -0.61098 | 0.585194 | | 2.510919 | -0.05938 | 0.979282 |
| IGSF9B | 4.012241 | -0.11245 | 0.999532 | | 3.975773 | -0.67721 | 0.585194 | | 3.084 | -0.54306 | 0.680577 |
| TECRL | 2.322169 | -0.38113 | 0.999532 | | 2.421494 | -0.67552 | 0.585347 | | 1.926271 | -0.28012 | 0.892004 |
| RFPL4AL | 2.754986 | 0.0366 | 0.999532 | | 2.41907 | -0.68209 | 0.585347 | | 2.129631 | -0.70611 | 0.64142 |
| LEP | 3.121116 | -0.31226 | 0.999532 | | 3.215765 | -0.64151 | 0.585347 | | 2.460627 | -0.31031 | 0.864923 |
| CLDN8 | 2.118898 | 0.60477 | 0.999532 | | 2.433243 | 0.64114 | 0.585347 | | 2.451764 | 0.050092 | 0.985306 |
| TRIM32 | 2.026966 | 0.651923 | 0.999532 | | 2.422073 | 0.683852 | 0.585347 | | 2.474991 | 0.057591 | 0.981929 |
| SLC27A2 | 2.066635 | 0.227291 | 0.999532 | | 2.458334 | 0.634473 | 0.585347 | | 2.393681 | 0.425648 | 0.78258 |
| NUSAP1 | 2.467368 | -0.33543 | 0.999532 | | 2.483646 | -0.6727 | 0.585347 | | 1.877823 | -0.3187 | 0.870966 |
| PPP1R1A | 2.942652 | 1.249865 | 0.396029 | | 2.435195 | 0.628986 | 0.585347 | | 3.234274 | -0.60275 | 0.598589 |
| GRIN2C | 5.460849 | 0.234958 | 0.999532 | | 4.897311 | -0.54498 | 0.585347 | | 4.655854 | -0.76438 | 0.442315 |
| SLC16A14 | 2.940863 | -0.14711 | 0.999532 | | 2.852346 | -0.6651 | 0.585347 | | 2.348567 | -0.50321 | 0.737714 |
| FAM81B | 2.478745 | -0.15695 | 0.999532 | | 2.438411 | -0.6764 | 0.585347 | | 1.944238 | -0.50164 | 0.752299 |
| BNIP3L | 15.15287 | -0.02121 | 0.999532 | | 18.8955 | 0.30904 | 0.585347 | | 15.92988 | 0.344336 | 0.622543 |
| APLNR | 5.572895 | -0.52384 | 0.999532 | | 8.737855 | 0.430614 | 0.585347 | | 6.600297 | 0.98665 | 0.107433 |
| GZF1 | 2.921031 | -0.494 | 0.999532 | | 3.274227 | -0.57546 | 0.585347 | | 2.26829 | -0.06205 | 0.97843 |
| LMO4 | 6.321404 | 0.035041 | 0.999532 | | 7.856997 | 0.471137 | 0.585347 | | 6.962421 | 0.417348 | 0.712621 |
| RNF125 | 2.086468 | -0.60623 | 0.999532 | | 2.321245 | -0.67458 | 0.585347 | | 1.675238 | -0.05042 | 0.986853 |
| TRAPPC4 | 4.734465 | 0.126945 | 0.999532 | | 6.000799 | 0.451211 | 0.58593 | | 5.40764 | 0.339757 | 0.738861 |
| CPLX2 | 10.74302 | 0.061345 | 0.999532 | | 10.46092 | -0.39044 | 0.58593 | | 9.393359 | -0.4435 | 0.602786 |
| SNRPN | 11.98354 | -0.11104 | 0.999532 | | 16.00437 | 0.278874 | 0.5866 | | 12.73359 | 0.417122 | 0.510412 |
| OR5A2 | 3.745816 | 0.246529 | 0.999532 | | 3.414508 | -0.64437 | 0.5866 | | 3.041241 | -0.86209 | 0.440178 |
| TEX261 | 4.131629 | -0.17773 | 0.999532 | | 5.593301 | 0.456707 | 0.5866 | | 4.655941 | 0.651895 | 0.465731 |
| NR1D1 | 2.298192 | 0.256805 | 0.999532 | | 2.042918 | -0.72258 | 0.5866 | | 1.918688 | -0.95682 | 0.483244 |
| HMX2 | 2.35308 | -0.03394 | 0.999532 | | 2.143923 | -0.69064 | 0.5866 | | 1.863912 | -0.63943 | 0.67176 |
| CLCA1 | 2.576338 | 0.063622 | 0.999532 | | 2.393812 | -0.70643 | 0.587058 | | 2.067304 | -0.75504 | 0.598671 |
| AKAP11 | 4.495801 | 0.374768 | 0.999532 | | 4.842274 | 0.463555 | 0.587308 | | 4.918262 | 0.108694 | 0.943248 |
| TCEA1 | 4.899922 | -0.01767 | 0.999532 | | 4.502333 | -0.48679 | 0.587308 | | 4.064387 | -0.45772 | 0.675955 |
| GRIK5 | 2.149557 | -0.70371 | 0.999532 | | 2.446572 | -0.65188 | 0.587308 | | 1.70748 | 0.068833 | 0.980022 |
| PTPRJ | 2.636221 | 0.211414 | 0.999532 | | 3.224067 | 0.573636 | 0.587308 | | 3.070654 | 0.376253 | 0.782921 |
| LRRC37B | 2.628439 | 0.08904 | 0.999532 | | 2.311809 | -0.66291 | 0.587308 | | 2.116627 | -0.73255 | 0.598329 |
| SRPRB | 3.823905 | 0.468254 | 0.999532 | | 4.412439 | 0.496806 | 0.587308 | | 4.223036 | 0.045365 | 0.978927 |
| TIE1 | 5.748574 | 0.053411 | 0.999532 | | 7.858565 | 0.466383 | 0.587308 | | 6.765839 | 0.450674 | 0.662584 |
| KCNB1 | 2.826694 | -0.35103 | 0.999532 | | 3.0057 | -0.65096 | 0.58775 | | 2.36907 | -0.28291 | 0.885086 |
| ZNF48 | 4.601814 | 0.04135 | 0.999532 | | 4.346622 | -0.53948 | 0.587881 | | 3.948004 | -0.56227 | 0.665886 |
| CSGALNA | 4.308073 | -0.54979 | 0.999532 | | 4.989275 | -0.50601 | 0.587881 | | 3.378422 | 0.0594 | 0.97502 |
| GLUD1 | 10.99011 | -0.02816 | 0.999532 | | 13.6954 | 0.299859 | 0.587881 | | 11.90662 | 0.352704 | 0.573023 |
| CC2D1B | 2.482959 | 0.540803 | 0.999532 | | 2.688082 | 0.623815 | 0.587941 | | 2.80858 | 0.102069 | 0.960427 |
| EIF1AX | 10.69728 | 0.221084 | 0.999532 | | 12.62528 | 0.362261 | 0.588029 | | 11.51299 | 0.159076 | 0.88885 |
| NAT8L | 3.694234 | -0.16045 | 0.999532 | | 3.637643 | -0.5679 | 0.588756 | | 2.95187 | -0.38857 | 0.784263 |
| POLR2M | 4.555859 | -0.27859 | 0.999532 | | 4.8061 | -0.52681 | 0.588756 | | 3.708362 | -0.23045 | 0.87591 |
| LCE1F | 2.398996 | -0.25199 | 0.999532 | | 2.375891 | -0.67451 | 0.588756 | | 1.840371 | -0.40827 | 0.819366 |
| C1GALT1 | 3.694923 | 0.361772 | 0.999532 | | 4.387358 | 0.509155 | 0.588756 | | 4.134174 | 0.167393 | 0.902567 |
| MAPKAP | 3.627902 | -0.11317 | 0.999532 | | 5.026755 | 0.454398 | 0.588756 | | 4.106934 | 0.583969 | 0.535079 |
| ELAVL3 | 3.194781 | -0.15356 | 0.999532 | | 3.230879 | -0.61675 | 0.588756 | | 2.737938 | -0.45775 | 0.749206 |
| SYT7 | 3.847317 | -0.47483 | 0.999532 | | 4.343533 | -0.576 | 0.588756 | | 3.053487 | -0.0864 | 0.966643 |
| APOA5 | 3.166119 | 0.1449 | 0.999532 | | 2.641806 | -0.66913 | 0.588756 | | 2.659063 | -0.79734 | 0.553283 |
| MCM2 | 3.320004 | -0.1634 | 0.999532 | | 3.237957 | -0.5987 | 0.588756 | | 2.70481 | -0.42291 | 0.774747 |
| DPRX | 2.446992 | -0.56494 | 0.999532 | | 2.654512 | -0.65999 | 0.588888 | | 1.817963 | -0.07634 | 0.977473 |
| HAGHL | 4.243549 | 0.562187 | 0.999532 | | 3.162989 | -0.62839 | 0.589009 | | 3.455563 | -1.17591 | 0.245834 |
| PBX4 | 2.315218 | -0.52965 | 0.999532 | | 2.398119 | -0.6834 | 0.589009 | | 1.734791 | -0.13646 | 0.955834 |
| NAA38 | 4.710225 | -0.31394 | 0.999532 | | 6.773122 | 0.415814 | 0.589009 | | 5.483038 | 0.769042 | 0.280916 |
| IREB2 | 7.362631 | 0.209705 | 0.999532 | | 9.005936 | 0.371953 | 0.589009 | | 8.268267 | 0.180314 | 0.852123 |
| TKFC | 4.122304 | 0.275655 | 0.999532 | | 4.960155 | 0.460843 | 0.589142 | | 4.558175 | 0.203343 | 0.872493 |
| BTLA | 2.109222 | -0.15639 | 0.999532 | | 1.978906 | -0.71969 | 0.589142 | | 1.743275 | -0.546 | 0.750524 |
| GLOD4 | 6.665477 | 0.462792 | 0.999532 | | 7.005531 | 0.393184 | 0.589614 | | 7.010145 | -0.05318 | 0.969614 |
| CELSR3 | 3.066401 | 0.370887 | 0.999532 | | 2.490863 | -0.64318 | 0.590016 | | 2.624617 | -1.00827 | 0.320564 |
| P2RY6 | 4.482926 | -0.02229 | 0.999532 | | 4.116665 | -0.53562 | 0.590183 | | 3.528298 | -0.49656 | 0.678541 |
| CD300LB | 2.824568 | -0.3959 | 0.999532 | | 3.025883 | -0.6517 | 0.590183 | | 2.294819 | -0.23988 | 0.899732 |
| EYS | 2.844521 | 0.048339 | 0.999532 | | 2.551482 | -0.65766 | 0.590805 | | 2.317335 | -0.68708 | 0.630705 |
| ARHGAP | 4.065646 | 0.063507 | 0.999532 | | 5.357312 | 0.441996 | 0.591151 | | 4.59978 | 0.398038 | 0.706749 |
| KRT34 | 5.006407 | 0.275544 | 0.999532 | | 4.122345 | -0.5173 | 0.591151 | | 4.187991 | -0.78783 | 0.397004 |
| LTK | 2.915705 | -0.29326 | 0.999532 | | 3.084202 | -0.59612 | 0.591151 | | 2.298321 | -0.28432 | 0.864245 |
| LMO7 | 6.762155 | 0.003439 | 0.999532 | | 8.614976 | 0.42287 | 0.591151 | | 7.590786 | 0.444789 | 0.578798 |
| EPS8 | 5.814871 | -0.40209 | 0.999532 | | 8.517499 | 0.385258 | 0.591151 | | 6.583036 | 0.808938 | 0.191229 |
| GALNT15 | 3.04806 | -0.02107 | 0.999532 | | 2.868951 | -0.62622 | 0.591151 | | 2.387604 | -0.58285 | 0.668553 |
| MAF1 | 5.491847 | 0.511104 | 0.999532 | | 5.869881 | 0.433436 | 0.591151 | | 6.051764 | -0.05784 | 0.969691 |
| CCDC175 | 2.633571 | -0.30305 | 0.999532 | | 2.654615 | -0.69417 | 0.592582 | | 2.183538 | -0.3792 | 0.836935 |
| GSTT4 | 1.888351 | 0.351067 | 0.999532 | | 2.48752 | 0.644397 | 0.592582 | | 2.3622 | 0.312528 | 0.850013 |
| HBB | 2.280815 | 0.693257 | 0.999532 | | 2.841395 | 0.75839 | 0.592582 | | 2.759252 | 0.155883 | 0.947126 |
| OSMR | 5.637868 | -0.96924 | 0.396029 | | 7.268936 | -0.44606 | 0.592582 | | 4.295205 | 0.552374 | 0.605329 |
| NYNRIN | 1.876961 | 0.432693 | 0.999532 | | 2.283984 | 0.671956 | 0.592582 | | 2.126334 | 0.255537 | 0.88588 |
| CNNM1 | 2.062584 | 0.044644 | 0.999532 | | 1.843692 | -0.74277 | 0.592582 | | 1.607341 | -0.77152 | 0.638796 |
| OLFML2B | 3.229498 | 0.211368 | 0.999532 | | 4.060031 | 0.538805 | 0.592582 | | 3.582084 | 0.349207 | 0.772025 |
| ZIC5 | 2.259729 | -0.1527 | 0.999532 | | 2.216474 | -0.68295 | 0.592582 | | 1.818041 | -0.51197 | 0.753597 |
| CCS | 6.0099 | -0.03122 | 0.999532 | | 8.046512 | 0.397799 | 0.592582 | | 6.647757 | 0.450684 | 0.58295 |
| SLC28A1 | 3.482019 | 0.638158 | 0.999532 | | 3.883686 | 0.540844 | 0.592582 | | 4.052548 | -0.07977 | 0.963866 |
| ZC3H8 | 2.205768 | -0.12673 | 0.999532 | | 2.011312 | -0.73465 | 0.592582 | | 1.802534 | -0.58953 | 0.736481 |
| SNRPB2 | 4.500835 | -0.1557 | 0.999532 | | 6.486243 | 0.430883 | 0.592582 | | 4.896874 | 0.595612 | 0.515762 |
| APOL6 | 4.625422 | -0.17599 | 0.999532 | | 4.591704 | -0.50058 | 0.592582 | | 3.92477 | -0.30133 | 0.806394 |
| CPM | 7.469561 | -0.62963 | 0.999532 | | 8.926573 | -0.44875 | 0.592582 | | 5.962815 | 0.203089 | 0.867976 |
| DDOST | 9.459329 | 0.03168 | 0.999532 | | 12.50202 | 0.438139 | 0.592582 | | 10.36771 | 0.444644 | 0.632847 |
| METTL8 | 3.73903 | -0.0982 | 0.999532 | | 3.454171 | -0.57342 | 0.592582 | | 2.84933 | -0.45412 | 0.725216 |
| EEF1E1 | 4.080351 | 0.321089 | 0.999532 | | 4.744552 | 0.498704 | 0.59303 | | 4.487131 | 0.197429 | 0.881837 |
| LRRTM2 | 2.772242 | 0.166795 | 0.999532 | | 2.399826 | -0.68657 | 0.594018 | | 2.206004 | -0.83449 | 0.541754 |
| SFT2D2 | 7.704305 | -0.16713 | 0.999532 | | 10.67789 | 0.36964 | 0.594018 | | 8.818572 | 0.557597 | 0.411291 |
| CTDNEP1 | 10.6237 | 0.157887 | 0.999532 | | 13.43326 | 0.412487 | 0.594018 | | 11.91142 | 0.275268 | 0.764149 |
| UTRN | 11.71736 | 0.087059 | 0.999532 | | 14.52575 | 0.34741 | 0.594271 | | 12.56816 | 0.28375 | 0.702463 |
| KLHL17 | 3.339203 | 0.100256 | 0.999532 | | 3.183237 | -0.57807 | 0.594271 | | 2.797598 | -0.65426 | 0.587597 |
| SYTL1 | 2.43444 | -0.35774 | 0.999532 | | 2.479009 | -0.65671 | 0.594271 | | 1.952069 | -0.27947 | 0.883792 |
| GSTO1 | 4.927424 | 0.068277 | 0.999532 | | 6.26785 | 0.418744 | 0.594271 | | 5.449059 | 0.368417 | 0.69872 |
| OTUD7B | 5.027308 | 0.394909 | 0.999532 | | 5.613272 | 0.444199 | 0.594415 | | 5.39285 | 0.069371 | 0.96169 |
| SLAMF1 | 2.458987 | -0.66326 | 0.999532 | | 2.902694 | -0.60095 | 0.594415 | | 1.90334 | 0.082251 | 0.974375 |
| TGFBR2 | 28.35943 | -0.03914 | 0.999532 | | 35.73656 | 0.321007 | 0.594415 | | 30.30028 | 0.375842 | 0.578798 |
| GAB2 | 3.937964 | 0.277061 | 0.999532 | | 4.856026 | 0.461293 | 0.594551 | | 4.354425 | 0.201564 | 0.870966 |
| FHAD1 | 2.565195 | -0.35748 | 0.999532 | | 2.597789 | -0.63249 | 0.594551 | | 2.066882 | -0.25712 | 0.892225 |
| SSBP3 | 10.50456 | 0.25633 | 0.999532 | | 12.583 | 0.331178 | 0.594551 | | 11.4021 | 0.100886 | 0.919181 |
| GJA1 | 2.836819 | 0.129649 | 0.999532 | | 3.720567 | 0.546243 | 0.594551 | | 3.222284 | 0.432349 | 0.722703 |
| FAIM2 | 2.764989 | -0.44273 | 0.999532 | | 2.738694 | -0.60457 | 0.594597 | | 2.112855 | -0.15307 | 0.942503 |
| BMI1 | 8.338465 | -0.09169 | 0.999532 | | 10.99397 | 0.338622 | 0.594597 | | 8.946333 | 0.452291 | 0.515762 |
| MTRNR2L | 2.470635 | -0.26259 | 0.999532 | | 2.519082 | -0.64676 | 0.594653 | | 1.907194 | -0.36425 | 0.836935 |
| ZSCAN30 | 2.542816 | 0.462838 | 0.999532 | | 3.143158 | 0.601495 | 0.595227 | | 3.099009 | 0.16148 | 0.927433 |

| | | | | | | | | | | | |
|---|---|---|---|---|---|---|---|---|---|---|---|
| PTDSS2 | 2.830279 | 0.318347 | 0.999532 | | 3.125647 | 0.572983 | 0.595515 | | 2.998976 | 0.246052 | 0.881011 |
| ZNF319 | 2.86422 | 0.629004 | 0.999532 | | 1.987025 | -0.71911 | 0.59552 | | 2.335381 | -1.33852 | 0.234754 |
| CDK5R1 | 3.045356 | -0.31113 | 0.999532 | | 3.15248 | -0.62615 | 0.59552 | | 2.433962 | -0.29483 | 0.86667 |
| OR14A2 | 2.060571 | -0.26761 | 0.999532 | | 2.038959 | -0.70597 | 0.59552 | | 1.603055 | -0.42252 | 0.824012 |
| TUBB6 | 4.125053 | 0.289021 | 0.999532 | | 4.858603 | 0.483729 | 0.59552 | | 4.548126 | 0.209499 | 0.872933 |
| CASD1 | 2.396701 | 0.36133 | 0.999532 | | 2.927118 | 0.606632 | 0.595806 | | 2.83817 | 0.266389 | 0.871957 |
| PSMB5 | 3.709718 | 0.145469 | 0.999532 | | 4.553798 | 0.479504 | 0.595901 | | 4.305599 | 0.355203 | 0.757835 |
| HSBP1 | 4.92413 | 0.17734 | 0.999532 | | 6.193436 | 0.421209 | 0.596819 | | 5.426075 | 0.261202 | 0.803301 |
| OXA1L | 4.0349 | 0.372213 | 0.999532 | | 4.341936 | 0.509344 | 0.597282 | | 4.232283 | 0.144552 | 0.920655 |
| PAFAH1B | 7.959301 | 0.039782 | 0.999532 | | 9.777135 | 0.354076 | 0.597387 | | 8.519629 | 0.328898 | 0.677254 |
| ARMH1 | 2.876122 | -0.00978 | 0.999532 | | 2.753377 | -0.64589 | 0.597387 | | 2.457764 | -0.62118 | 0.663709 |
| PIDD1 | 2.689762 | 0.110742 | 0.999532 | | 2.385478 | -0.67451 | 0.597491 | | 2.205588 | -0.76267 | 0.59686 |
| DFFA | 6.074449 | 0.078423 | 0.999532 | | 7.547446 | 0.415003 | 0.597707 | | 6.779312 | 0.351719 | 0.717627 |
| MRPS17 | 3.636502 | -0.13231 | 0.999532 | | 3.564709 | -0.54871 | 0.597834 | | 2.816637 | -0.39715 | 0.769793 |
| KCNK7 | 2.528225 | -0.10009 | 0.999532 | | 2.262457 | -0.67938 | 0.59786 | | 2.08374 | -0.56829 | 0.725586 |
| CD1B | 2.666538 | -0.07822 | 0.999532 | | 2.548033 | -0.72494 | 0.59786 | | 2.107188 | -0.62705 | 0.692064 |
| ATP5MC3 | 10.08757 | -0.12425 | 0.999532 | | 13.463 | 0.317495 | 0.59786 | | 10.79601 | 0.465021 | 0.497845 |
| DPPA5 | 2.957548 | 0.291919 | 0.999532 | | 2.400057 | -0.68638 | 0.59786 | | 2.326525 | -0.95679 | 0.436389 |
| PLXDC2 | 4.994389 | 0.205931 | 0.999532 | | 6.373149 | 0.450822 | 0.597941 | | 5.378567 | 0.232127 | 0.859426 |
| FBXW10 | 3.399895 | 0.550853 | 0.999532 | | 2.489131 | -0.6533 | 0.598344 | | 2.87005 | -1.19083 | 0.281423 |
| NEK3 | 1.753488 | 0.250144 | 0.999532 | | 2.191181 | 0.661809 | 0.598401 | | 2.111358 | 0.427161 | 0.795612 |
| NTMT1 | 3.063771 | 0.235225 | 0.999532 | | 3.943936 | 0.530867 | 0.598403 | | 3.563951 | 0.319269 | 0.811123 |
| GPATCH1 | 6.07076 | 0.093144 | 0.999532 | | 5.555313 | -0.48674 | 0.598403 | | 5.213295 | -0.57038 | 0.570037 |
| MZF1 | 4.960097 | 0.311794 | 0.999532 | | 6.061198 | 0.447805 | 0.598403 | | 5.612737 | 0.152181 | 0.899247 |
| C9orf43 | 2.998482 | 0.529321 | 0.999532 | | 2.182933 | -0.69052 | 0.598408 | | 2.44423 | -1.20853 | 0.277362 |
| PLSCR1 | 8.216201 | -0.14262 | 0.999532 | | 10.79255 | 0.364678 | 0.598594 | | 8.990732 | 0.520223 | 0.486743 |
| COX16 | 4.890269 | 0.181011 | 0.999532 | | 5.772178 | 0.447916 | 0.598594 | | 5.351884 | 0.282608 | 0.799944 |
| DECR2 | 3.439535 | -0.08482 | 0.999532 | | 3.344388 | -0.58805 | 0.598594 | | 2.57276 | -0.48628 | 0.724411 |
| KSR2 | 1.636265 | 0.551569 | 0.999532 | | 1.928637 | 0.70762 | 0.598594 | | 1.958763 | 0.173841 | 0.932589 |
| ARHGAP | 4.851411 | -0.37042 | 0.999532 | | 5.18391 | -0.44468 | 0.598594 | | 3.912347 | -0.07037 | 0.971732 |
| LASP1 | 8.900912 | -0.07011 | 0.999532 | | 11.84648 | 0.408792 | 0.598594 | | 10.08594 | 0.48483 | 0.532137 |
| GGH | 2.524945 | -0.13119 | 0.999532 | | 3.27833 | 0.56783 | 0.598594 | | 2.762882 | 0.714955 | 0.553283 |
| GPALPP1 | 4.917842 | -0.29542 | 0.999532 | | 4.975166 | -0.55478 | 0.598594 | | 3.940847 | -0.21779 | 0.870663 |
| GSTK1 | 7.990313 | 0.023707 | 0.999532 | | 10.27711 | 0.384785 | 0.598594 | | 8.620645 | 0.385596 | 0.647218 |
| CLK2 | 10.63635 | 0.374179 | 0.999532 | | 11.68422 | 0.346874 | 0.598594 | | 11.26583 | -0.0096 | 0.996681 |
| ADAM22 | 1.439853 | 0.321226 | 0.999532 | | 1.834978 | 0.738502 | 0.598594 | | 1.685974 | 0.432762 | 0.811123 |
| RSKR | 2.629799 | -0.16085 | 0.999532 | | 2.601262 | -0.64454 | 0.598594 | | 2.151707 | -0.46922 | 0.760337 |
| SCLY | 2.7836 | 0.462044 | 0.999532 | | 3.39822 | 0.582682 | 0.598594 | | 3.246369 | 0.139022 | 0.934665 |
| POMK | 3.547662 | 0.025951 | 0.999532 | | 3.343469 | -0.60018 | 0.598594 | | 2.972995 | -0.61223 | 0.647218 |
| SCNN1D | 2.584788 | -0.0677 | 0.999532 | | 2.383245 | -0.65757 | 0.598594 | | 2.016467 | -0.57358 | 0.705993 |
| MIP | 2.769325 | -0.50893 | 0.999532 | | 3.008055 | -0.6199 | 0.598972 | | 2.260578 | -0.09443 | 0.972221 |
| TARDBP | 11.29699 | 0.23604 | 0.999532 | | 12.81031 | 0.340846 | 0.598972 | | 11.92437 | 0.121099 | 0.894617 |
| ARHGDIB | 18.33603 | -0.31053 | 0.999532 | | 26.3257 | 0.391173 | 0.599049 | | 20.82876 | 0.719706 | 0.244096 |
| NDFIP1 | 4.968736 | 0.269016 | 0.999532 | | 6.168721 | 0.472231 | 0.599758 | | 5.642162 | 0.225942 | 0.852074 |
| TIGD5 | 2.930512 | 0.202977 | 0.999532 | | 2.615209 | -0.66564 | 0.600039 | | 2.352032 | -0.84268 | 0.531045 |
| IQCF1 | 2.633676 | 0.575779 | 0.999532 | | 1.909237 | -0.73859 | 0.600039 | | 2.155487 | -1.29995 | 0.266933 |
| DPAGT1 | 3.430736 | 0.056603 | 0.999532 | | 3.307536 | -0.54639 | 0.600163 | | 2.781791 | -0.58241 | 0.628234 |
| RITA1 | 2.360705 | 0.706397 | 0.999532 | | 2.539647 | 0.616917 | 0.60032 | | 2.625557 | -0.07232 | 0.973152 |
| BLVRB | 2.121761 | 0.527506 | 0.999532 | | 2.401055 | 0.618669 | 0.60032 | | 2.398102 | 0.107309 | 0.958191 |
| SKP1 | 33.618 | 0.097962 | 0.999532 | | 39.45328 | 0.218783 | 0.60032 | | 34.33895 | 0.144678 | 0.791576 |
| CYTH1 | 4.687627 | -0.04605 | 0.999532 | | 4.630079 | -0.49888 | 0.60032 | | 3.839816 | -0.43485 | 0.705046 |
| ZAR1L | 2.316541 | -0.32187 | 0.999532 | | 2.333188 | -0.67215 | 0.60032 | | 1.779493 | -0.33557 | 0.864245 |
| LACTB2 | 2.758189 | -0.08162 | 0.999532 | | 3.711374 | 0.535913 | 0.60032 | | 3.111587 | 0.634755 | 0.589729 |
| ZFP3 | 3.422565 | -0.14654 | 0.999532 | | 3.417315 | -0.55466 | 0.60032 | | 2.853242 | -0.38801 | 0.789815 |
| TUBA1C | 43.61572 | -0.27316 | 0.999532 | | 61.329 | 0.350362 | 0.60032 | | 47.99853 | 0.657479 | 0.241605 |
| CD180 | 2.682781 | 0.503434 | 0.999532 | | 2.038675 | -0.70694 | 0.60032 | | 2.230471 | -1.19395 | 0.286063 |
| LOXL2 | 2.568202 | -0.02986 | 0.999532 | | 2.540814 | -0.65759 | 0.60032 | | 2.046644 | -0.59834 | 0.698371 |
| DNER | 3.151405 | -0.2282 | 0.999532 | | 3.143815 | -0.60814 | 0.60032 | | 2.32388 | -0.35705 | 0.81294 |
| DDX24 | 4.223196 | 0.258574 | 0.999532 | | 5.11759 | 0.446937 | 0.60032 | | 4.729722 | 0.208341 | 0.865065 |
| LRRC55 | 2.27083 | -0.14613 | 0.999532 | | 2.143851 | -0.71043 | 0.60032 | | 1.808246 | -0.54552 | 0.741408 |
| OR4N2 | 2.481901 | 0.242943 | 0.999532 | | 2.147873 | -0.68156 | 0.60032 | | 2.003083 | -0.90497 | 0.497845 |
| ADGRE2 | 3.479736 | -0.43686 | 0.999532 | | 3.649873 | -0.56284 | 0.60032 | | 2.661569 | -0.10397 | 0.959907 |
| CTAG2 | 3.146502 | -0.20384 | 0.999532 | | 3.058393 | -0.57918 | 0.600339 | | 2.482753 | -0.36454 | 0.820125 |
| PIEZO1 | 4.934912 | -0.1339 | 0.999532 | | 6.801339 | 0.43954 | 0.600368 | | 5.394482 | 0.597959 | 0.530776 |
| HDHD3 | 2.816314 | 0.332836 | 0.999532 | | 3.322583 | 0.564393 | 0.600446 | | 3.100499 | 0.246895 | 0.872493 |
| C12orf57 | 19.72761 | -0.0835 | 0.999532 | | 25.30882 | 0.278556 | 0.600883 | | 21.32333 | 0.383808 | 0.515762 |
| SLCO1B7 | 3.011546 | -0.02774 | 0.999532 | | 2.750788 | -0.62207 | 0.600883 | | 2.488879 | -0.57435 | 0.69872 |
| PEPD | 4.395158 | -0.49792 | 0.999532 | | 7.020624 | 0.43536 | 0.600883 | | 4.961575 | 0.971601 | 0.241605 |
| PINK1 | 5.940989 | -0.28635 | 0.999532 | | 8.301144 | 0.388037 | 0.601404 | | 6.481101 | 0.6771 | 0.318193 |
| SLC35G2 | 2.360729 | -0.29819 | 0.999532 | | 2.375937 | -0.68385 | 0.601708 | | 1.773654 | -0.3679 | 0.843463 |
| RPS3 | 36.37939 | -0.13375 | 0.999532 | | 47.64911 | 0.305415 | 0.60178 | | 38.74317 | 0.466346 | 0.441823 |
| TAF7 | 5.538992 | 0.025341 | 0.999532 | | 7.144334 | 0.399998 | 0.60178 | | 6.176371 | 0.39251 | 0.660958 |
| YWHAG | 4.792207 | -0.22627 | 0.999532 | | 6.716359 | 0.405015 | 0.60178 | | 5.368062 | 0.648659 | 0.462115 |
| CKM | 2.16611 | 0.436422 | 0.999532 | | 1.654502 | -0.75644 | 0.60178 | | 1.7781 | -1.17002 | 0.363418 |
| ZFC3H1 | 5.248646 | 0.089106 | 0.999532 | | 6.747883 | 0.41072 | 0.601891 | | 5.873988 | 0.341546 | 0.718643 |
| PSMB3 | 8.437169 | 0.118074 | 0.999532 | | 10.43344 | 0.400687 | 0.601891 | | 9.30241 | 0.299102 | 0.724411 |
| TRAF4 | 6.131193 | 0.259505 | 0.999532 | | 5.37677 | -0.46888 | 0.601891 | | 4.936714 | -0.70257 | 0.39437 |
| ZNF747 | 2.501857 | -0.02036 | 0.999532 | | 2.347406 | -0.68308 | 0.601922 | | 2.136947 | -0.65325 | 0.666566 |
| CHCHD1 | 3.603024 | -0.15969 | 0.999532 | | 3.635494 | -0.54804 | 0.601922 | | 2.753526 | -0.37658 | 0.791576 |
| CKS1B | 10.46526 | 0.009365 | 0.999532 | | 10.6272 | -0.36932 | 0.601922 | | 9.127956 | -0.36136 | 0.65401 |
| NXPE3 | 2.159967 | -0.05316 | 0.999532 | | 2.01988 | -0.70433 | 0.602095 | | 1.727623 | -0.63499 | 0.68597 |
| RAET1G | 2.961374 | 0.178461 | 0.999532 | | 2.650799 | -0.67291 | 0.602111 | | 2.475295 | -0.83419 | 0.520848 |
| FBXL2 | 2.384951 | 0.443248 | 0.999532 | | 3.132976 | 0.622312 | 0.602686 | | 3.035004 | 0.195563 | 0.908274 |
| CDC123 | 3.052447 | 0.101758 | 0.999532 | | 3.950375 | 0.532695 | 0.602686 | | 3.476883 | 0.449921 | 0.712171 |
| ACADVL | 15.32225 | 0.097281 | 0.999532 | | 18.82266 | 0.359846 | 0.602686 | | 16.38026 | 0.285299 | 0.738316 |
| NVL | 2.804802 | -0.78847 | 0.999532 | | 3.243747 | -0.5532 | 0.602686 | | 2.18338 | 0.246972 | 0.892004 |
| ITPR2 | 2.949579 | 0.044294 | 0.999532 | | 3.994527 | 0.504211 | 0.602686 | | 3.399466 | 0.473891 | 0.680885 |
| ZNF594 | 4.16807 | 0.043077 | 0.999532 | | 4.096271 | -0.49801 | 0.602714 | | 3.579838 | -0.51882 | 0.644433 |
| MRPL39 | 4.203865 | -0.33053 | 0.999532 | | 4.584927 | -0.4808 | 0.602714 | | 3.340457 | -0.13072 | 0.93603 |
| MTDH | 5.906388 | 0.270658 | 0.999532 | | 6.482813 | 0.399889 | 0.602714 | | 6.2057 | 0.141372 | 0.903329 |
| UBXN2B | 2.761086 | 0.944006 | 0.999532 | | 2.580506 | 0.611571 | 0.602714 | | 3.048097 | -0.315 | 0.819366 |
| TRIB1 | 2.651811 | -0.03398 | 0.999532 | | 2.56897 | -0.63912 | 0.602714 | | 2.034279 | -0.58387 | 0.701406 |
| SLC25A1 | 6.453398 | 0.248069 | 0.999532 | | 7.400296 | 0.395919 | 0.602714 | | 6.904317 | 0.159108 | 0.896886 |
| PTPRD | 3.859466 | 0.312073 | 0.999532 | | 4.53712 | 0.491343 | 0.602714 | | 4.29761 | 0.198829 | 0.881689 |
| COPS8 | 4.596609 | -0.30947 | 0.999532 | | 6.705877 | 0.418604 | 0.602714 | | 5.299332 | 0.747766 | 0.333173 |
| TCF23 | 2.912447 | -0.25291 | 0.999532 | | 3.171379 | -0.58338 | 0.602714 | | 2.305746 | -0.30754 | 0.853737 |
| CSNK1G1 | 2.373225 | 0.030383 | 0.999532 | | 2.165835 | -0.66511 | 0.602714 | | 1.88048 | -0.68028 | 0.647218 |
| RNF166 | 3.90445 | 0.240753 | 0.999532 | | 4.69319 | 0.462188 | 0.602714 | | 4.453865 | 0.242492 | 0.842754 |
| PLAT | 47.24371 | -0.59013 | 0.893099 | | 58.57419 | -0.23985 | 0.602714 | | 41.04076 | 0.37947 | 0.540342 |
| RIOX1 | 2.934448 | -0.34772 | 0.999532 | | 3.033779 | -0.57937 | 0.602714 | | 2.338575 | -0.21593 | 0.903329 |
| ZMYND11 | 5.433793 | -0.06272 | 0.999532 | | 7.241802 | 0.395599 | 0.602714 | | 6.157825 | 0.476945 | 0.574369 |
| C9 | 2.353136 | -0.40051 | 0.999532 | | 2.645761 | -0.64936 | 0.602714 | | 1.844418 | -0.23072 | 0.906811 |
| SEMA6D | 3.18643 | 0.410762 | 0.999532 | | 3.794494 | 0.531553 | 0.602714 | | 3.715559 | 0.139344 | 0.929271 |
| PIGK | 5.585327 | -0.11592 | 0.999532 | | 7.851185 | 0.385628 | 0.602714 | | 6.402718 | 0.521016 | 0.515762 |
| TBX20 | 1.923979 | -0.37 | 0.999532 | | 2.0119 | -0.72004 | 0.602714 | | 1.547787 | -0.33068 | 0.87764 |
| ZNF888 | 6.143594 | -0.05728 | 0.999532 | | 6.17366 | -0.41613 | 0.602714 | | 5.051903 | -0.33982 | 0.730921 |
| FNDC10 | 2.755708 | 0.42584 | 0.999532 | | 2.10076 | -0.68765 | 0.602714 | | 2.221654 | -1.09831 | 0.359441 |
| TRIOBP | 7.822294 | -0.11619 | 0.999532 | | 10.35376 | 0.37059 | 0.602714 | | 8.701424 | 0.509434 | 0.48078 |
| SCARF2 | 2.741222 | 0.002117 | 0.999532 | | 3.600793 | 0.510772 | 0.602714 | | 3.291504 | 0.5389 | 0.65401 |
| BTBD19 | 4.198499 | -0.12535 | 0.999532 | | 5.863249 | 0.433538 | 0.60498 | | 4.588786 | 0.575853 | 0.5258 |
| SPDYE2 | 4.938391 | -0.19231 | 0.999532 | | 5.095678 | -0.47964 | 0.60498 | | 4.060547 | -0.26671 | 0.849682 |
| TAMM41 | 4.53892 | -0.34581 | 0.999532 | | 5.113286 | -0.45851 | 0.605223 | | 3.748212 | -0.08699 | 0.961426 |
| KIDINS22 | 4.068637 | -0.08483 | 0.999532 | | 5.693317 | 0.455442 | 0.60642 | | 4.764129 | 0.557917 | 0.572297 |
| CYSTM1 | 10.64872 | 0.340993 | 0.999532 | | 11.91755 | 0.355624 | 0.606463 | | 11.25388 | 0.032543 | 0.984283 |
| TMEM120 | 3.577811 | 0.27421 | 0.999532 | | 4.284628 | 0.470099 | 0.606504 | | 4.008725 | 0.213891 | 0.877898 |
| PSME1 | 4.815515 | 0.504171 | 0.999532 | | 5.304411 | 0.477527 | 0.606504 | | 5.253664 | -0.00484 | 0.997967 |
| UBA7 | 10.72926 | -0.00229 | 0.999532 | | 13.58706 | 0.318231 | 0.606504 | | 11.42574 | 0.340912 | 0.616301 |
| KRTAP10 | 3.38149 | -0.15912 | 0.999532 | | 3.653611 | -0.60804 | 0.606504 | | 2.796112 | -0.41089 | 0.790337 |
| ING4 | 4.898877 | 0.003612 | 0.999532 | | 6.418798 | 0.413544 | 0.606504 | | 5.505128 | 0.428855 | 0.660958 |
| PSG5 | 6.382799 | 0.120927 | 0.999532 | | 6.105963 | -0.46997 | 0.606504 | | 5.573084 | -0.57737 | 0.542124 |
| GSN | 10.10155 | -0.15181 | 0.999532 | | 13.43624 | 0.314888 | 0.606504 | | 10.6196 | 0.489395 | 0.474523 |
| CLDND2 | 3.60311 | -0.41393 | 0.999532 | | 3.958367 | -0.58856 | 0.606592 | | 2.991031 | -0.15668 | 0.938429 |
| PNPLA6 | 2.301757 | 0.172658 | 0.999532 | | 2.800497 | 0.572795 | 0.607697 | | 2.613896 | 0.415835 | 0.765012 |
| ZNF70 | 2.950581 | 0.414124 | 0.999532 | | 3.618302 | 0.529084 | 0.607874 | | 3.408643 | 0.137574 | 0.934172 |
| FLRT1 | 2.242524 | 0.372992 | 0.999532 | | 2.596268 | 0.594061 | 0.608162 | | 2.545186 | 0.236202 | 0.886495 |
| MRPL21 | 2.650186 | 0.602821 | 0.999532 | | 2.850178 | 0.567233 | 0.608226 | | 2.987639 | -0.01799 | 0.996407 |
| RNF224 | 2.664027 | -0.13056 | 0.999532 | | 2.614964 | -0.62188 | 0.608226 | | 2.244217 | -0.47143 | 0.765012 |
| SEPHS1 | 4.622561 | 0.329588 | 0.999532 | | 5.401484 | 0.448681 | 0.608226 | | 5.020978 | 0.136611 | 0.914687 |
| PLGLB1 | 6.491953 | 0.022419 | 0.999532 | | 6.307191 | -0.42006 | 0.608317 | | 5.442715 | -0.42378 | 0.647218 |
| ITGA8 | 6.506994 | -0.2573 | 0.999532 | | 9.257408 | 0.453317 | 0.608317 | | 7.470049 | 0.755271 | 0.247358 |
| RPS6KB1 | 8.621795 | -0.0606 | 0.999532 | | 8.769092 | -0.37137 | 0.60854 | | 7.277061 | -0.29077 | 0.737109 |
| IRAG1 | 4.482489 | -9.93E-05 | 0.99982 | | 6.202481 | 0.424778 | 0.608889 | | 5.272646 | 0.446093 | 0.647218 |
| DLX2 | 3.111946 | -0.12617 | 0.999532 | | 3.120157 | -0.58144 | 0.60899 | | 2.610215 | -0.43779 | 0.764702 |
| RAB1B | 13.35977 | 0.163772 | 0.999532 | | 15.29591 | 0.301994 | 0.60899 | | 14.01855 | 0.154277 | 0.858647 |
| RNF4 | 4.741604 | 0.474047 | 0.999532 | | 5.038667 | 0.453628 | 0.60899 | | 5.023147 | 0.000366 | 0.999867 |
| SGCB | 6.018263 | -0.22897 | 0.999532 | | 8.144003 | 0.412781 | 0.60899 | | 6.685577 | 0.643202 | 0.456688 |
| M1AP | 3.042344 | 0.553443 | 0.999532 | | 2.401812 | -0.68938 | 0.60899 | | 2.533813 | -1.22065 | 0.274496 |
| GTF2H1 | 3.877599 | 0.044379 | 0.999532 | | 5.052478 | 0.454855 | 0.609014 | | 4.342928 | 0.430813 | 0.693549 |
| AQP2 | 6.587657 | -0.67374 | 0.999532 | | 7.073127 | -0.80874 | 0.609014 | | 4.540244 | -0.01401 | 0.996681 |
| MCCC1 | 2.46691 | 0.829164 | 0.999532 | | 2.524308 | 0.593963 | 0.609014 | | 2.86271 | -0.21493 | 0.88885 |
| MPC1L | 2.776514 | 0.026453 | 0.999532 | | 2.644591 | -0.63297 | 0.609014 | | 2.283429 | -0.64082 | 0.653214 |
| RSL1D1 | 5.294137 | 0.335642 | 0.999532 | | 5.957051 | 0.434077 | 0.609389 | | 5.838185 | 0.113688 | 0.928522 |
| UGT2B7 | 7.125693 | -0.02808 | 0.999532 | | 9.920216 | 0.525592 | 0.609422 | | 8.088596 | 0.585707 | 0.588428 |
| PAMR1 | 2.125419 | -0.44734 | 0.999532 | | 2.255317 | -0.65466 | 0.609422 | | 1.627375 | -0.18826 | 0.932589 |
| FCHSD2 | 5.039468 | -0.13853 | 0.999532 | | 7.17964 | 0.404678 | 0.610292 | | 5.702019 | 0.554068 | 0.506666 |
| SUV39H2 | 2.155414 | -0.19123 | 0.999532 | | 2.086805 | -0.71658 | 0.610914 | | 1.670685 | -0.50749 | 0.784602 |
| MTRNR2L | 6.225091 | 0.826728 | 0.999532 | | 4.191834 | -0.55835 | 0.61097 | | 5.163211 | -1.39607 | 0.154583 |
| ZBTB2 | 3.448014 | -0.12618 | 0.999532 | | 3.461041 | -0.56517 | 0.61097 | | 2.791454 | -0.41959 | 0.759211 |
| ANKLE1 | 2.655213 | -0.1685 | 0.999532 | | 2.562534 | -0.62346 | 0.61097 | | 2.081911 | -0.43921 | 0.788243 |
| UBA52 | 17.39603 | 0.024812 | 0.999532 | | 21.34595 | 0.32126 | 0.61097 | | 18.41255 | 0.31114 | 0.673405 |
| CENPO | 2.101691 | 0.782552 | 0.999532 | | 2.253494 | 0.666752 | 0.61097 | | 2.450927 | -0.09973 | 0.964434 |
| GIMAP1 | 2.079949 | -0.84011 | 0.999532 | | 2.451276 | -0.63577 | 0.61097 | | 1.751962 | 0.223467 | 0.918694 |
| SMIM33 | 2.607552 | -0.04797 | 0.999532 | | 2.576144 | -0.62461 | 0.611355 | | 2.173259 | -0.54638 | 0.720106 |
| TXNIP | 37.3844 | -0.0117 | 0.999532 | | 46.59577 | 0.26457 | 0.611355 | | 39.32339 | 0.301381 | 0.622543 |
| USP6 | 2.674497 | -0.09364 | 0.999532 | | 2.83865 | -0.61284 | 0.611355 | | 2.160778 | -0.4846 | 0.749222 |
| C4orf3 | 14.20567 | 0.02852 | 0.999532 | | 16.9458 | 0.315144 | 0.611355 | | 14.75185 | 0.30091 | 0.663633 |
| MAP3K14 | 3.813133 | 0.031324 | 0.999532 | | 3.410928 | -0.5626 | 0.611355 | | 2.942593 | -0.57715 | 0.649779 |
| MYO1E | 16.27926 | -0.18567 | 0.999532 | | 22.25565 | 0.391527 | 0.611355 | | 18.28438 | 0.596521 | 0.299463 |
| SLC5A4 | 3.443283 | 0.852565 | 0.999532 | | 3.337989 | 0.579242 | 0.612964 | | 3.989675 | -0.24637 | 0.87591 |
| ACOX2 | 4.277591 | 0.510742 | 0.999532 | | 5.100939 | 0.477823 | 0.612964 | | 5.016259 | -0.00334 | 0.998898 |
| SDHD | 12.16485 | 0.141522 | 0.999532 | | 14.55917 | 0.294303 | 0.612964 | | 12.80663 | 0.172045 | 0.835554 |
| CFDP1 | 6.169645 | 0.394023 | 0.999532 | | 7.156491 | 0.413667 | 0.612964 | | 6.811467 | 0.04075 | 0.97843 |
| STEAP1 | 3.170739 | 0.011333 | 0.999532 | | 2.978397 | -0.58408 | 0.612972 | | 2.644581 | -0.57452 | 0.673405 |
| ATRN | 6.371093 | -0.06556 | 0.999532 | | 8.201878 | 0.357541 | 0.614466 | | 6.721806 | 0.439072 | 0.577689 |
| DNAAF3 | 2.68383 | -0.13674 | 0.999532 | | 2.476015 | -0.65191 | 0.614573 | | 2.064732 | -0.49236 | 0.757035 |
| PVR | 6.245004 | 0.161725 | 0.999532 | | 7.481658 | 0.389269 | 0.614573 | | 6.899027 | 0.241283 | 0.810635 |
| TIMM29 | 2.259828 | -0.53056 | 0.999532 | | 2.536118 | -0.65856 | 0.614816 | | 1.745162 | -0.10486 | 0.970268 |
| FAH | 2.287039 | -0.01873 | 0.999532 | | 3.270513 | 0.546569 | 0.614816 | | 2.729266 | 0.581766 | 0.643959 |
| GHR | 2.497054 | 0.274788 | 0.999532 | | 2.063918 | -0.68016 | 0.614816 | | 2.017703 | -0.93692 | 0.477483 |
| RGMA | 5.216347 | 0.053054 | 0.999532 | | 4.943878 | -0.49494 | 0.614816 | | 4.324423 | -0.52761 | 0.630705 |
| TMEM51 | 3.371607 | -0.24145 | 0.999532 | | 4.985702 | 0.459867 | 0.614816 | | 3.818457 | 0.710642 | 0.456688 |
| FUBP3 | 6.820073 | 0.074668 | 0.999532 | | 8.400189 | 0.368477 | 0.614869 | | 7.36371 | 0.307912 | 0.731674 |
| ACER2 | 3.358769 | -0.14949 | 0.999532 | | 3.356941 | -0.54676 | 0.616311 | | 2.78024 | -0.3757 | 0.796702 |
| STAT3 | 12.01405 | -0.24661 | 0.999532 | | 16.55231 | 0.30561 | 0.616345 | | 13.11319 | 0.572136 | 0.30099 |
| ATP1B1 | 30.83557 | 0.193341 | 0.999532 | | 37.86138 | 0.404675 | 0.616345 | | 33.75532 | 0.238569 | 0.840357 |
| ENTPD8 | 4.085845 | -0.00818 | 0.999532 | | 4.175824 | -0.52669 | 0.616345 | | 3.477877 | -0.4812 | 0.715922 |
| MAP4K3 | 3.935097 | 0.029625 | 0.999532 | | 4.69334 | 0.457893 | 0.617119 | | 4.319794 | 0.441016 | 0.68597 |
| SMIM24 | 5.322285 | 0.219479 | 0.999532 | | 6.436708 | 0.519993 | 0.617119 | | 5.866538 | 0.312506 | 0.821463 |
| ZBTB34 | 2.517882 | -0.33853 | 0.999532 | | 2.712103 | -0.59368 | 0.61731 | | 1.976706 | -0.2329 | 0.900167 |
| MINDY1 | 2.59931 | 0.265993 | 0.999532 | | 3.484972 | 0.544404 | 0.617456 | | 2.986454 | 0.297603 | 0.836453 |
| SLC35A4 | 3.672426 | 0.529669 | 0.999532 | | 3.876682 | 0.511336 | 0.617456 | | 4.233264 | -0.00575 | 0.997967 |
| CTH | 3.026432 | 0.475097 | 0.999532 | | 2.448664 | -0.63349 | 0.617456 | | 2.49641 | -1.08848 | 0.30043 |
| ZNF385A | 6.125653 | 0.018978 | 0.999532 | | 7.80987 | 0.380638 | 0.617456 | | 6.757246 | 0.383996 | 0.664512 |
| KLHL5 | 4.520438 | 0.206946 | 0.999532 | | 5.548182 | 0.432239 | 0.617456 | | 5.037611 | 0.246989 | 0.834165 |
| FADS3 | 3.969195 | 0.223367 | 0.999532 | | 4.953459 | 0.450716 | 0.618052 | | 4.428712 | 0.249047 | 0.836453 |
| ERC1 | 4.239602 | 0.34844 | 0.999532 | | 4.824342 | 0.452408 | 0.618222 | | 4.544698 | 0.120328 | 0.932589 |
| BCL11B | 2.393746 | -0.39771 | 0.999532 | | 2.413429 | -0.63071 | 0.618566 | | 1.779986 | -0.21928 | 0.914692 |
| WWC1 | 2.082511 | 0.593349 | 0.999532 | | 2.353854 | 0.613092 | 0.618566 | | 2.396295 | 0.037228 | 0.989437 |
| FN3KRP | 2.543688 | 0.591254 | 0.999532 | | 2.834937 | 0.580848 | 0.618566 | | 2.887549 | 0.009106 | 0.996891 |
| MRPL55 | 5.24308 | 0.375198 | 0.999532 | | 5.806727 | 0.431172 | 0.618566 | | 5.478941 | 0.071733 | 0.960773 |
| ESRRA | 4.242707 | -0.24038 | 0.999532 | | 4.308051 | -0.49339 | 0.618718 | | 3.250738 | -0.23724 | 0.870663 |
| BAG3 | 4.972908 | 0.026838 | 0.999532 | | 6.557157 | 0.438756 | 0.61891 | | 5.413513 | 0.44552 | 0.659727 |
| SMYD2 | 6.019529 | -0.01051 | 0.999532 | | 7.518513 | 0.374865 | 0.619105 | | 6.614868 | 0.400109 | 0.647218 |
| F13A1 | 2.555058 | -0.42798 | 0.999532 | | 2.502046 | -0.60177 | 0.619105 | | 2.01677 | -0.15788 | 0.941197 |
| KLRC2 | 5.899367 | 0.110991 | 0.999532 | | 5.461795 | -0.58384 | 0.619105 | | 4.910359 | -0.67743 | 0.563885 |
| STXBP5 | 2.980046 | 0.025845 | 0.999532 | | 2.788365 | -0.56911 | 0.619105 | | 2.361563 | -0.5752 | 0.660097 |
| COQ4 | 3.458547 | 0.433762 | 0.999532 | | 4.101769 | 0.488053 | 0.619168 | | 3.906971 | 0.072713 | 0.96703 |
| ZNF461 | 6.725611 | -0.34445 | 0.999532 | | 7.670854 | -0.37205 | 0.619168 | | 5.644447 | -0.00737 | 0.996891 |
| MKRN3 | 2.302185 | -0.0013 | 0.999532 | | 2.156126 | -0.67165 | 0.619302 | | 1.874852 | -0.65518 | 0.673405 |
| RAD9A | 6.141464 | 0.194345 | 0.999532 | | 7.50324 | 0.377554 | 0.619302 | | 6.636925 | 0.202815 | 0.845814 |
| TTN | 2.824655 | -0.54674 | 0.999532 | | 3.237934 | -0.56335 | 0.619302 | | 2.333304 | -0.00278 | 0.999385 |
| ALG1L | 4.635374 | 0.203897 | 0.999532 | | 4.217478 | -0.52526 | 0.619302 | | 4.001486 | -0.71208 | 0.504551 |
| NRAS | 3.629902 | -0.75057 | 0.999532 | | 4.287133 | -0.47678 | 0.619302 | | 2.791352 | 0.283842 | 0.853068 |
| RGS20 | 2.667647 | -0.03313 | 0.999532 | | 2.562806 | -0.62263 | 0.619302 | | 2.028011 | -0.57032 | 0.698562 |
| UBE2U | 2.828633 | 0.275788 | 0.999532 | | 2.454077 | -0.6569 | 0.619302 | | 2.345793 | -0.91355 | 0.474081 |
| ANKRD2 | 2.44484 | -0.47453 | 0.999532 | | 2.823955 | -0.63798 | 0.619302 | | 2.006282 | -0.14273 | 0.949669 |
| SWAP70 | 6.612088 | 0.175255 | 0.999532 | | 8.017572 | 0.373947 | 0.619302 | | 7.218504 | 0.221279 | 0.835289 |
| KLHL36 | 3.110825 | 0.595653 | 0.999532 | | 3.234117 | 0.536423 | 0.619302 | | 3.378983 | -0.0394 | 0.986208 |
| MAPRE3 | 4.457133 | -0.25837 | 0.999532 | | 5.052551 | -0.47423 | 0.619302 | | 3.977035 | -0.16069 | 0.920259 |
| PDE4C | 3.205095 | 0.902535 | 0.999532 | | 3.023369 | 0.56534 | 0.619302 | | 3.524144 | -0.31886 | 0.816618 |
| TM4SF4 | 5.036897 | -0.18381 | 0.999532 | | 5.092097 | -0.45754 | 0.619302 | | 4.255143 | -0.24952 | 0.85585 |
| MIDEAS | 6.552392 | -0.03776 | 0.999532 | | 6.449507 | -0.40218 | 0.619302 | | 5.360539 | -0.34816 | 0.723933 |
| SERAC1 | 2.445236 | -0.04752 | 0.999532 | | 2.436323 | -0.66524 | 0.619302 | | 2.034936 | -0.59536 | 0.705046 |
| RNASE8 | 2.47304 | -0.23225 | 0.999532 | | 2.430298 | -0.67113 | 0.619302 | | 1.819398 | -0.41537 | 0.814884 |
| RASGRP | 3.381951 | 0.17467 | 0.999532 | | 2.854309 | -0.56576 | 0.619302 | | 2.726185 | -0.72702 | 0.540342 |
| CCNI2 | 2.291336 | -0.27959 | 0.999532 | | 2.262144 | -0.64803 | 0.619302 | | 1.838604 | -0.35393 | 0.852985 |
| ALDH3A1 | 2.255331 | -0.11598 | 0.999532 | | 2.119444 | -0.6623 | 0.619302 | | 1.807133 | -0.53016 | 0.746581 |
| SBSPON | 6.243692 | 0.090757 | 0.999532 | | 5.928632 | -0.4497 | 0.619302 | | 4.816103 | -0.51622 | 0.59686 |
| NEK9 | 3.038492 | 0.046677 | 0.999532 | | 2.699224 | -0.58994 | 0.619302 | | 2.430886 | -0.62457 | 0.647581 |
| SH2B3 | 4.425967 | -0.14586 | 0.999532 | | 6.3465 | 0.426057 | 0.619302 | | 4.972761 | 0.598346 | 0.524206 |
| CLEC4E | 2.208055 | -0.29965 | 0.999532 | | 2.291082 | -0.65706 | 0.619302 | | 1.713425 | -0.33883 | 0.867976 |
| PQBP1 | 3.812625 | 0.697629 | 0.999532 | | 4.022429 | 0.507458 | 0.619302 | | 4.248887 | -0.17035 | 0.89901 |
| TDRD6 | 2.130468 | -0.20002 | 0.999532 | | 2.136976 | -0.67154 | 0.619302 | | 1.650615 | -0.45063 | 0.804392 |
| VHL | 2.881944 | 0.937694 | 0.999532 | | 2.912409 | 0.572997 | 0.619302 | | 3.369112 | -0.34061 | 0.79686 |
| ZDHHC11 | 6.089053 | -0.02518 | 0.999532 | | 6.1277 | -0.41024 | 0.619302 | | 5.049451 | -0.36549 | 0.707135 |
| ZNF331 | 4.458767 | -0.86204 | 0.999532 | | 5.475943 | -0.47228 | 0.619302 | | 3.365449 | 0.408417 | 0.735469 |
| ANKRD2 | 2.999813 | -0.103 | 0.999532 | | 2.942377 | -0.5791 | 0.619302 | | 2.43914 | -0.459 | 0.749407 |
| CBLB | 8.236398 | 0.576577 | 0.999532 | | 8.442568 | 0.376598 | 0.619302 | | 8.582064 | -0.18494 | 0.864245 |
| H1-3 | 4.060456 | -0.63314 | 0.999532 | | 4.594775 | -0.47318 | 0.619302 | | 3.159134 | 0.162984 | 0.920259 |
| ANK3 | 6.441567 | 0.044304 | 0.999532 | | 7.992786 | 0.367134 | 0.619302 | | 6.853738 | 0.343992 | 0.678492 |
| MBNL1 | 4.256876 | -0.1964 | 0.999532 | | 5.541439 | 0.413536 | 0.619302 | | 4.619651 | 0.619687 | 0.515762 |
| SCAF1 | 7.888489 | -0.34002 | 0.999532 | | 9.162553 | -0.35446 | 0.619302 | | 6.631557 | 0.005139 | 0.997967 |
| SPRR2E | 2.196594 | -0.3405 | 0.999532 | | 2.204802 | -0.65651 | 0.619302 | | 1.731329 | -0.29384 | 0.883679 |
| SP2 | 5.199465 | -0.50042 | 0.999532 | | 5.816394 | -0.42511 | 0.619302 | | 4.136843 | 0.069251 | 0.956322 |
| SP9 | 5.636988 | 0.230292 | 0.999532 | | 4.946074 | -0.48043 | 0.619302 | | 4.823238 | -0.70034 | 0.458377 |
| AMHR2 | 2.314074 | -0.02687 | 0.999532 | | 2.047929 | -0.73288 | 0.619302 | | 1.774139 | -0.68245 | 0.669689 |
| TRAF3IP2 | 3.639359 | -0.28639 | 0.999532 | | 3.919633 | -0.55587 | 0.619302 | | 2.87368 | -0.24878 | 0.876613 |
| KCND1 | 1.998454 | -1.13979 | 0.999532 | | 2.591952 | -0.62529 | 0.619302 | | 1.615185 | 0.538499 | 0.779186 |
| EXOC1 | 2.985455 | 0.688786 | 0.999532 | | 3.034751 | 0.553701 | 0.619302 | | 3.270688 | -0.11766 | 0.945283 |
| GALNT8 | 2.105521 | -0.35869 | 0.999532 | | 2.249285 | -0.6785 | 0.619302 | | 1.640427 | -0.29689 | 0.885892 |
| CLYBL | 3.136012 | 0.184703 | 0.999532 | | 2.556833 | -0.60031 | 0.619302 | | 2.463165 | -0.77301 | 0.531732 |
| MGAT4B | 4.962917 | 0.303219 | 0.999532 | | 5.645244 | 0.436448 | 0.619302 | | 5.247971 | 0.152582 | 0.901033 |
| MCOLN2 | 2.208105 | -0.19473 | 0.999532 | | 2.165459 | -0.67443 | 0.619302 | | 1.753471 | -0.46148 | 0.795612 |
| DNAJC3 | 6.709098 | 0.198718 | 0.999532 | | 8.004288 | 0.374653 | 0.619302 | | 7.1427 | 0.195494 | 0.84579 |
| PHLDB1 | 11.23759 | 0.414433 | 0.999532 | | 12.42493 | 0.382556 | 0.619302 | | 12.11677 | -0.01226 | 0.996471 |
| TGFBR3L | 7.982944 | -0.30307 | 0.999532 | | 8.819996 | -0.40623 | 0.619423 | | 6.670086 | -0.07819 | 0.958889 |
| MEIS1 | 4.421558 | 0.044776 | 0.999532 | | 5.499529 | 0.466119 | 0.619621 | | 4.815389 | 0.430005 | 0.706407 |
| PPHLN1 | 3.911952 | 0.075095 | 0.999532 | | 4.955554 | 0.459372 | 0.619764 | | 4.30631 | 0.403178 | 0.70366 |
| PAGR1 | 5.320051 | 0.267597 | 0.999532 | | 6.379401 | 0.400876 | 0.619764 | | 5.844864 | 0.15242 | 0.894617 |
| ZNF181 | 2.303824 | -0.65033 | 0.999532 | | 2.60222 | -0.61792 | 0.619764 | | 1.910963 | 0.051275 | 0.987491 |
| ABLIM2 | 4.434907 | 0.319696 | 0.999532 | | 5.171539 | 0.437461 | 0.619893 | | 4.865454 | 0.137083 | 0.920259 |
| UFC1 | 4.436135 | 0.364689 | 0.999532 | | 4.998691 | 0.451695 | 0.620407 | | 5.009548 | 0.101524 | 0.943229 |
| MAN2A1 | 3.652146 | -0.01117 | 0.999532 | | 4.906152 | 0.442532 | 0.621815 | | 4.131391 | 0.471906 | 0.646939 |
| ARHGEF7 | 7.050709 | -0.13816 | 0.999532 | | 8.907373 | 0.357252 | 0.622094 | | 7.379869 | 0.507879 | 0.515762 |
| CARM1 | 3.997333 | 0.481423 | 0.999532 | | 4.201944 | 0.47852 | 0.622312 | | 4.342936 | 0.009752 | 0.996891 |
| GSTA1 | 14.92842 | 0.112707 | 0.999532 | | 20.45065 | 0.620274 | 0.622312 | | 17.82976 | 0.532705 | 0.724363 |
| SPEGNB | 2.038392 | 0.837042 | 0.999532 | | 2.155359 | 0.695046 | 0.622312 | | 2.453067 | -0.12576 | 0.955834 |
| RBBP6 | 3.939434 | 0.01445 | 0.999532 | | 5.288525 | 0.430443 | 0.622312 | | 4.330703 | 0.428856 | 0.68528 |
| MEIOB | 2.246884 | 0.161788 | 0.999532 | | 1.827859 | -0.72102 | 0.622312 | | 1.824341 | -0.86322 | 0.572084 |
| MTRNR2L | 4.822492 | 0.475363 | 0.999532 | | 3.96062 | -0.5208 | 0.622312 | | 4.060821 | -0.97479 | 0.271669 |
| RIBC1 | 2.646549 | 0.326628 | 0.999532 | | 2.301922 | -0.66516 | 0.622312 | | 2.102267 | -0.97 | 0.432927 |
| OR4C6 | 2.051061 | -0.02915 | 0.999532 | | 1.986126 | -0.68453 | 0.622312 | | 1.591417 | -0.63444 | 0.696959 |
| NUDT12 | 2.089041 | 0.188043 | 0.999532 | | 2.864661 | 0.586051 | 0.622312 | | 2.532886 | 0.415651 | 0.774747 |
| CALML4 | 2.929181 | 0.450655 | 0.999532 | | 3.267468 | 0.546067 | 0.622312 | | 3.172773 | 0.11318 | 0.948099 |
| NUFIP1 | 2.182267 | 0.541523 | 0.999532 | | 2.606682 | 0.599486 | 0.622312 | | 2.541294 | 0.077037 | 0.972221 |
| CHRNA4 | 2.108028 | -0.42445 | 0.999532 | | 2.254891 | -0.63803 | 0.622572 | | 1.734388 | -0.19446 | 0.92697 |
| TOMM20 | 7.273576 | -0.15305 | 0.999532 | | 9.771182 | 0.38507 | 0.622706 | | 7.911413 | 0.567528 | 0.433783 |
| OSBPL2 | 3.559115 | 0.641686 | 0.999532 | | 3.595684 | 0.490361 | 0.622706 | | 3.95719 | -0.13322 | 0.92403 |
| RWDD4 | 7.57464 | 0.093876 | 0.999532 | | 7.165104 | -0.38881 | 0.622706 | | 6.336044 | -0.46769 | 0.586203 |
| NT5C3B | 3.119681 | 0.594639 | 0.999532 | | 3.582186 | 0.552998 | 0.624152 | | 3.583384 | -0.02091 | 0.994192 |
| WWTR1 | 8.134632 | -0.04466 | 0.999532 | | 10.71345 | 0.335962 | 0.624152 | | 8.850733 | 0.393844 | 0.581238 |
| POLR3D | 3.390685 | -0.18413 | 0.999532 | | 3.658579 | -0.56312 | 0.624152 | | 2.89483 | -0.36545 | 0.803301 |
| AXIN2 | 3.48933 | -0.29321 | 0.999532 | | 3.541899 | -0.57945 | 0.624152 | | 2.573859 | -0.24532 | 0.87892 |
| SCD | 6.133289 | 0.080257 | 0.999532 | | 5.787008 | -0.4267 | 0.624152 | | 5.374491 | -0.48925 | 0.614824 |
| FOXH1 | 3.14904 | 0.014446 | 0.999532 | | 2.907217 | -0.60208 | 0.624152 | | 2.618535 | -0.59748 | 0.707142 |
| TGM5 | 2.826576 | 0.222681 | 0.999532 | | 2.493348 | -0.65241 | 0.624152 | | 2.327968 | -0.85555 | 0.523065 |
| THRA | 5.455858 | -0.37705 | 0.999532 | | 5.975756 | -0.41525 | 0.624152 | | 4.213435 | -0.02636 | 0.991273 |
| SLC38A2 | 13.57873 | -0.3288 | 0.999532 | | 18.87055 | 0.309906 | 0.624152 | | 14.52181 | 0.656264 | 0.244096 |
| MCM7 | 3.095247 | 0.030759 | 0.999532 | | 4.092391 | 0.474855 | 0.624152 | | 3.583301 | 0.463076 | 0.681128 |
| EXOSC3 | 5.30401 | -0.35869 | 0.999532 | | 5.851084 | -0.41376 | 0.62465 | | 4.357435 | -0.03737 | 0.98514 |
| MARK2 | 4.436332 | 0.088422 | 0.999532 | | 5.354732 | 0.424236 | 0.624694 | | 4.933148 | 0.345388 | 0.751138 |
| DUSP15 | 2.742401 | 0.311712 | 0.999532 | | 2.203354 | -0.64875 | 0.624776 | | 2.223818 | -0.94498 | 0.469804 |
| ATP5MF | 18.07662 | 0.061003 | 0.999532 | | 21.88375 | 0.260368 | 0.624928 | | 19.07285 | 0.220186 | 0.710512 |
| CREB3L2 | 5.173067 | 0.164585 | 0.999532 | | 6.413268 | 0.397012 | 0.625363 | | 5.733897 | 0.252107 | 0.806797 |
| MMP19 | 3.449754 | -0.18348 | 0.999532 | | 3.488766 | -0.5494 | 0.625363 | | 2.9387 | -0.34622 | 0.823571 |
| TP53I11 | 5.672088 | 0.097481 | 0.999532 | | 7.067793 | 0.424608 | 0.625363 | | 6.303935 | 0.339417 | 0.722703 |
| P4HTM | 7.293667 | 0.124623 | 0.999532 | | 9.081011 | 0.364513 | 0.625363 | | 7.869493 | 0.26348 | 0.774747 |
| SPP1 | 17.09148 | 0.066096 | 0.999532 | | 24.39563 | 0.652903 | 0.625363 | | 20.8911 | 0.611868 | 0.718643 |
| OR10A6 | 2.329092 | -0.46359 | 0.999532 | | 2.437215 | -0.62785 | 0.625363 | | 1.880155 | -0.14875 | 0.948127 |
| REEP2 | 3.240911 | 0.037318 | 0.999532 | | 2.898718 | -0.55125 | 0.625363 | | 2.680372 | -0.57439 | 0.663089 |
| SDHB | 8.52313 | -0.1085 | 0.999532 | | 11.7291 | 0.321815 | 0.625415 | | 9.267141 | 0.465912 | 0.560424 |
| NAA15 | 3.476186 | 0.179943 | 0.999532 | | 3.076963 | -0.66338 | 0.625415 | | 2.710832 | -0.8306 | 0.486743 |
| CMAS | 3.624606 | 0.813012 | 0.999532 | | 3.418292 | 0.523705 | 0.625415 | | 3.77274 | -0.26762 | 0.833306 |
| NNT | 2.929912 | 0.240583 | 0.999532 | | 3.839363 | 0.510671 | 0.625415 | | 3.299658 | 0.28234 | 0.836453 |
| BTBD9 | 2.706778 | -0.04379 | 0.999532 | | 3.86151 | 0.488468 | 0.625415 | | 3.155836 | 0.548147 | 0.636525 |
| RPL36 | 10.40905 | -0.05089 | 0.999532 | | 13.41479 | 0.406545 | 0.625415 | | 11.31544 | 0.471441 | 0.595502 |
| ZNF85 | 2.684777 | -0.21031 | 0.999532 | | 2.676434 | -0.61307 | 0.625415 | | 2.021993 | -0.38334 | 0.816618 |
| PTPRK | 4.382254 | 0.287201 | 0.999532 | | 5.259207 | 0.482675 | 0.625415 | | 4.785454 | 0.23177 | 0.84413 |
| MFGE8 | 8.679779 | -0.15722 | 0.999532 | | 11.34942 | 0.350269 | 0.625415 | | 9.260772 | 0.516097 | 0.510286 |
| CFB | 2.144167 | 0.325541 | 0.999532 | | 2.701713 | 0.577072 | 0.625415 | | 2.374382 | 0.266887 | 0.867378 |
| BEX4 | 7.81082 | 0.3359 | 0.999532 | | 8.907385 | 0.34784 | 0.625415 | | 8.275899 | 0.033821 | 0.980758 |
| RPL13 | 39.46945 | -0.08849 | 0.999532 | | 51.72358 | 0.336157 | 0.625415 | | 42.6156 | 0.450653 | 0.528307 |
| CYB5R1 | 4.005599 | 0.011436 | 0.999532 | | 5.562042 | 0.43372 | 0.625415 | | 4.649402 | 0.441059 | 0.666906 |
| FAM13A | 2.743284 | -0.17159 | 0.999532 | | 2.601792 | -0.58727 | 0.625655 | | 2.142951 | -0.40657 | 0.799944 |
| INSL6 | 2.379077 | 0.374362 | 0.999532 | | 1.935955 | -0.69906 | 0.625672 | | 1.927298 | -1.05528 | 0.421517 |
| NIBAN1 | 2.447138 | -0.23873 | 0.999532 | | 2.39484 | -0.64395 | 0.625814 | | 1.928375 | -0.38556 | 0.827127 |
| ETHE1 | 3.73514 | -0.00478 | 0.999532 | | 5.149622 | 0.434874 | 0.625814 | | 4.085955 | 0.465886 | 0.663633 |
| CES3 | 2.969534 | -0.04173 | 0.999532 | | 2.722654 | -0.59147 | 0.625814 | | 2.408686 | -0.5326 | 0.713778 |
| HGF | 2.114041 | 0.907215 | 0.999532 | | 2.043232 | 0.687548 | 0.625814 | | 2.473246 | -0.21129 | 0.910105 |
| DMAC1 | 7.920137 | 0.39564 | 0.999532 | | 8.642242 | 0.337515 | 0.625814 | | 8.410901 | -0.03983 | 0.974421 |
| BCKDK | 2.91162 | 0.616576 | 0.999532 | | 3.086836 | 0.532489 | 0.625814 | | 3.182404 | -0.06583 | 0.973356 |
| SLC35C2 | 6.838553 | 0.219222 | 0.999532 | | 8.016378 | 0.382909 | 0.625814 | | 7.562947 | 0.187127 | 0.865065 |
| CDKN2A | 2.695272 | -0.03323 | 0.999532 | | 2.430377 | -0.61919 | 0.625814 | | 2.131986 | -0.5717 | 0.704316 |
| CFAP46 | 3.294334 | 0.23059 | 0.999532 | | 3.077772 | -0.58483 | 0.625814 | | 2.749654 | -0.7938 | 0.507289 |
| ULK1 | 3.515392 | 0.344359 | 0.999532 | | 4.21321 | 0.465825 | 0.625814 | | 3.941217 | 0.138108 | 0.92697 |
| SLC5A2 | 4.006103 | 0.45739 | 0.999532 | | 4.391063 | 0.484984 | 0.625814 | | 4.352755 | 0.036786 | 0.98732 |
| MOSMO | 2.67911 | 0.640076 | 0.999532 | | 2.871131 | 0.578976 | 0.625814 | | 3.059722 | -0.04644 | 0.985299 |
| CASP5 | 3.22147 | -0.19064 | 0.999532 | | 3.44003 | -0.53691 | 0.625814 | | 2.754291 | -0.3139 | 0.846576 |
| BTG3 | 2.628349 | 0.49434 | 0.999532 | | 3.050345 | 0.534741 | 0.625814 | | 2.946257 | 0.058829 | 0.977401 |
| TRIP10 | 8.898469 | -0.05864 | 0.999532 | | 11.22573 | 0.330812 | 0.625814 | | 9.416686 | 0.408548 | 0.570037 |
| HACE1 | 3.459069 | -0.27323 | 0.999532 | | 3.553993 | -0.5347 | 0.625814 | | 2.889688 | -0.25243 | 0.882613 |
| MAG | 2.32858 | 0.011944 | 0.999532 | | 2.059881 | -0.65256 | 0.625814 | | 1.846578 | -0.65055 | 0.669717 |
| PLEKHM1 | 5.714793 | -0.13838 | 0.999532 | | 7.532457 | 0.385158 | 0.625814 | | 6.243188 | 0.543137 | 0.533876 |
| IL13 | 4.74352 | -0.04759 | 0.999532 | | 4.615116 | -0.51011 | 0.625814 | | 3.847099 | -0.44215 | 0.709825 |
| ZNF727 | 2.904789 | 0.051651 | 0.999532 | | 2.589145 | -0.61985 | 0.625814 | | 2.32223 | -0.64852 | 0.644433 |
| KDR | 18.68573 | 0.123689 | 0.999532 | | 23.06527 | 0.364265 | 0.625814 | | 20.69202 | 0.256466 | 0.675955 |
| G0S2 | 6.025853 | 0.391668 | 0.999532 | | 6.743937 | 0.400357 | 0.625814 | | 6.363249 | 0.005319 | 0.997967 |
| PHACTR | 2.645335 | -0.25173 | 0.999532 | | 2.642172 | -0.59931 | 0.625814 | | 2.071455 | -0.32955 | 0.849021 |
| BMPR2 | 9.20495 | 0.017614 | 0.999532 | | 11.26687 | 0.34867 | 0.625814 | | 9.982533 | 0.345397 | 0.662584 |
| SLC25A2 | 2.898271 | -0.73489 | 0.999532 | | 3.609464 | -0.56283 | 0.625814 | | 2.207904 | 0.196923 | 0.917499 |
| LARS1 | 6.394385 | 0.347076 | 0.999532 | | 7.121119 | 0.396555 | 0.625814 | | 6.79386 | 0.072912 | 0.956011 |
| FAAH | 2.064073 | 0.782211 | 0.999532 | | 2.280712 | 0.716426 | 0.625814 | | 2.468344 | -0.05683 | 0.98556 |
| CBX2 | 3.321138 | -0.08204 | 0.999532 | | 3.487895 | -0.58371 | 0.625814 | | 2.952718 | -0.49283 | 0.728015 |
| HOXB5 | 4.365825 | 0.118144 | 0.999532 | | 3.848274 | -0.4982 | 0.625814 | | 3.630993 | -0.59898 | 0.60646 |
| ZG16 | 2.327745 | -0.18614 | 0.999532 | | 2.36513 | -0.65819 | 0.625814 | | 1.851503 | -0.45251 | 0.796937 |
| GPANK1 | 4.798718 | -0.02266 | 0.999532 | | 4.666256 | -0.45123 | 0.625814 | | 3.931891 | -0.4087 | 0.712891 |
| CORO1C | 4.786851 | -0.11415 | 0.999532 | | 6.451104 | 0.419163 | 0.625814 | | 5.486047 | 0.552052 | 0.515762 |
| AZU1 | 2.335837 | -0.30329 | 0.999532 | | 2.443236 | -0.63205 | 0.625814 | | 1.84111 | -0.30971 | 0.871957 |
| PSMD7 | 6.675983 | -0.22848 | 0.999532 | | 9.579631 | 0.337521 | 0.625814 | | 7.2953 | 0.571399 | 0.407399 |
| SHANK1 | 2.27486 | -0.38033 | 0.999532 | | 2.187336 | -0.63472 | 0.625814 | | 1.778091 | -0.25091 | 0.903339 |
| SPATA24 | 3.591744 | -0.04437 | 0.999532 | | 3.628068 | -0.53501 | 0.625814 | | 3.064807 | -0.45439 | 0.745939 |
| MAP6D1 | 3.832197 | 0.162616 | 0.999532 | | 3.309575 | -0.55416 | 0.625814 | | 3.232718 | -0.70067 | 0.548279 |
| SF3A3 | 4.232039 | 0.43611 | 0.999532 | | 4.515342 | 0.493604 | 0.625814 | | 4.53141 | 0.065043 | 0.972305 |
| EGR4 | 2.551469 | -0.20547 | 0.999532 | | 2.689094 | -0.70684 | 0.625814 | | 2.087136 | -0.49504 | 0.774747 |
| RNMT | 3.050347 | 0.264293 | 0.999532 | | 3.689952 | 0.534255 | 0.625814 | | 3.412548 | 0.290555 | 0.835605 |
| ACTR10 | 6.132922 | 0.19322 | 0.999532 | | 7.410811 | 0.362984 | 0.625814 | | 6.666026 | 0.190731 | 0.859255 |
| KRTAP22 | 1.935935 | -0.34314 | 0.999532 | | 2.044388 | -0.68613 | 0.625814 | | 1.588205 | -0.31993 | 0.881837 |
| IDE | 4.251376 | 0.295887 | 0.999532 | | 4.83674 | 0.442471 | 0.625814 | | 4.613204 | 0.159308 | 0.904801 |
| MAP2 | 2.019248 | 0.331353 | 0.999532 | | 2.477958 | 0.602743 | 0.625814 | | 2.323431 | 0.288896 | 0.863536 |
| HSPA5 | 13.62143 | 0.137473 | 0.999532 | | 17.30034 | 0.433867 | 0.625814 | | 15.29405 | 0.318405 | 0.747677 |
| H2BS1 | 11.75628 | -0.17133 | 0.999532 | | 15.55201 | 0.324456 | 0.625814 | | 12.5254 | 0.516239 | 0.441078 |
| SOD3 | 4.91635 | -0.56412 | 0.999532 | | 5.833471 | -0.4914 | 0.625814 | | 4.084246 | 0.077417 | 0.965619 |
| LY6G5C | 2.339429 | -0.48495 | 0.999532 | | 2.484676 | -0.66804 | 0.625814 | | 1.835147 | -0.15837 | 0.946482 |
| TDGF1 | 3.31826 | -0.41452 | 0.999532 | | 3.670256 | -0.51758 | 0.625814 | | 2.745501 | -0.07803 | 0.972305 |
| DNAJC7 | 5.357562 | 0.31172 | 0.999532 | | 6.089965 | 0.394359 | 0.625814 | | 5.623105 | 0.098548 | 0.939549 |
| UVSSA | 4.811923 | -0.2071 | 0.999532 | | 5.062637 | -0.4434 | 0.625814 | | 3.883608 | -0.22192 | 0.864245 |
| ATP6V1D | 3.501276 | 0.795082 | 0.999532 | | 3.49664 | 0.505937 | 0.625814 | | 3.86491 | -0.27112 | 0.824012 |
| ZNF419 | 4.31178 | -0.45359 | 0.999532 | | 4.867727 | -0.44109 | 0.625814 | | 3.663119 | 0.032844 | 0.989437 |
| TLNRD1 | 2.051374 | -0.35157 | 0.999532 | | 2.096941 | -0.64122 | 0.625814 | | 1.632545 | -0.27205 | 0.892869 |
| RASA1 | 7.448144 | 0.232699 | 0.999532 | | 8.237442 | 0.349654 | 0.625814 | | 7.862628 | 0.122091 | 0.912963 |
| TRIM38 | 1.864821 | -0.20774 | 0.999532 | | 2.811547 | 0.565006 | 0.625814 | | 2.351335 | 0.792601 | 0.539565 |
| RAD51AP | 2.091658 | 0.064963 | 0.999532 | | 1.920311 | -0.73863 | 0.627049 | | 1.617745 | -0.78455 | 0.64142 |
| MCUB | 2.679116 | 0.48575 | 0.999532 | | 2.132095 | -0.66926 | 0.628182 | | 2.076848 | -1.13195 | 0.309288 |
| GALK2 | 3.21394 | 0.184984 | 0.999532 | | 3.777028 | 0.485505 | 0.628182 | | 3.549373 | 0.313142 | 0.812437 |
| SEMA6A | 4.487417 | -0.16583 | 0.999532 | | 6.261331 | 0.431073 | 0.628182 | | 5.189847 | 0.614431 | 0.508751 |
| NPFF | 4.54855 | -0.44237 | 0.999532 | | 5.259937 | -0.48396 | 0.628182 | | 3.572468 | -0.02174 | 0.993618 |
| MAD2L1 | 2.747661 | -0.02469 | 0.999532 | | 2.543109 | -0.60092 | 0.628407 | | 2.168299 | -0.55912 | 0.688542 |
| PKHD1 | 4.920307 | 0.631539 | 0.999532 | | 5.362702 | 0.501565 | 0.628407 | | 5.480234 | -0.11285 | 0.940266 |
| WDR18 | 3.907074 | -0.15952 | 0.999532 | | 3.867766 | -0.52844 | 0.628407 | | 3.00383 | -0.34527 | 0.806394 |
| TNS3 | 9.880427 | -0.15257 | 0.999532 | | 13.37327 | 0.302754 | 0.628408 | | 10.55743 | 0.4827 | 0.515762 |
| LAD1 | 4.449713 | 0.059722 | 0.999532 | | 5.486246 | 0.411764 | 0.628498 | | 4.73413 | 0.368222 | 0.717627 |
| RPL36A | 21.94555 | -0.02115 | 0.999532 | | 26.49333 | 0.236499 | 0.629179 | | 22.54394 | 0.274132 | 0.622543 |
| CSRNP3 | 2.945202 | -0.26366 | 0.999532 | | 3.209519 | -0.55044 | 0.629344 | | 2.299622 | -0.26347 | 0.87591 |
| NRBF2 | 2.737656 | -0.27304 | 0.999532 | | 2.882624 | -0.58578 | 0.629344 | | 2.236817 | -0.29752 | 0.864245 |
| WARS2 | 2.234199 | 0.397858 | 0.999532 | | 1.788461 | -0.68994 | 0.629344 | | 1.83771 | -1.06703 | 0.415282 |
| CDK10 | 3.454605 | 0.48164 | 0.999532 | | 4.017646 | 0.487471 | 0.629344 | | 3.850584 | 0.029294 | 0.990309 |
| GPN1 | 4.421185 | 0.067654 | 0.999532 | | 5.883765 | 0.409891 | 0.629453 | | 4.924867 | 0.362296 | 0.718643 |
| CDIP1 | 3.619494 | 0.376401 | 0.999532 | | 4.397405 | 0.531551 | 0.629719 | | 4.231887 | 0.172523 | 0.910105 |
| GFM1 | 3.314255 | 0.181577 | 0.999532 | | 4.354173 | 0.489303 | 0.629719 | | 3.740211 | 0.334933 | 0.803301 |
| YAE1 | 2.910186 | -0.01603 | 0.999532 | | 2.852361 | -0.57166 | 0.630249 | | 2.408712 | -0.53707 | 0.693634 |
| ZMAT1 | 2.887992 | 0.56778 | 0.999532 | | 3.440545 | 0.547359 | 0.630249 | | 3.198181 | -0.00707 | 0.997967 |
| PDPN | 2.876222 | -0.48943 | 0.999532 | | 4.318629 | 0.49342 | 0.630605 | | 3.39759 | 1.017813 | 0.266933 |
| ARL3 | 3.843833 | -0.12813 | 0.999532 | | 3.726358 | -0.50393 | 0.631221 | | 3.104603 | -0.36225 | 0.792539 |
| BLOC1S4 | 3.340577 | -0.3432 | 0.999532 | | 3.887165 | -0.52012 | 0.631221 | | 2.724716 | -0.1329 | 0.945129 |
| AQP1 | 10.4371 | -0.52418 | 0.999532 | | 15.01355 | 0.336094 | 0.631221 | | 11.24139 | 0.84375 | 0.28307 |
| RBMX2 | 4.66904 | -0.25901 | 0.999532 | | 4.779696 | -0.47878 | 0.631221 | | 3.654542 | -0.19657 | 0.892869 |
| ADORA3 | 3.2752 | 0.248602 | 0.999532 | | 3.051613 | -0.61141 | 0.631221 | | 2.756057 | -0.8164 | 0.532137 |
| TPSG1 | 2.225703 | 0.067686 | 0.999532 | | 2.023893 | -0.69186 | 0.631221 | | 1.836724 | -0.74402 | 0.647218 |
| P4HA1 | 3.134529 | -0.2594 | 0.999532 | | 4.483118 | 0.488023 | 0.631221 | | 3.619562 | 0.757922 | 0.499025 |
| TPP2 | 5.113886 | 0.296741 | 0.999532 | | 6.125531 | 0.408139 | 0.631221 | | 5.591327 | 0.124957 | 0.918694 |
| CD276 | 4.262671 | 0.304497 | 0.999532 | | 5.197653 | 0.431212 | 0.631221 | | 4.777272 | 0.143326 | 0.908698 |
| ZNF506 | 11.05 | 0.198533 | 0.999532 | | 12.95798 | 0.325768 | 0.631221 | | 11.70896 | 0.150717 | 0.876613 |
| SIL1 | 3.393246 | 0.157619 | 0.999532 | | 4.656409 | 0.472072 | 0.631221 | | 4.133243 | 0.324902 | 0.784263 |
| TAOK3 | 4.233362 | -0.21977 | 0.999532 | | 4.817767 | -0.47044 | 0.631221 | | 3.436601 | -0.20851 | 0.888554 |
| MGA | 3.143367 | -0.27428 | 0.999532 | | 3.169429 | -0.53756 | 0.631221 | | 2.46025 | -0.2507 | 0.881689 |
| MICAL2 | 4.161006 | -0.43498 | 0.999532 | | 4.455811 | -0.4628 | 0.631221 | | 3.272349 | -0.00966 | 0.996891 |
| CCZ1 | 5.850727 | 0.121594 | 0.999532 | | 7.217965 | 0.409842 | 0.631222 | | 6.367184 | 0.309627 | 0.75322 |
| UNC5A | 6.835819 | -0.14645 | 0.999532 | | 7.212748 | -0.41585 | 0.631745 | | 5.702335 | -0.23189 | 0.85585 |
| LUZP6 | 5.519509 | -0.10932 | 0.999532 | | 7.644864 | 0.380706 | 0.632485 | | 6.487754 | 0.521113 | 0.581238 |
| TTC17 | 5.16295 | 0.693567 | 0.999532 | | 4.941911 | 0.449751 | 0.632485 | | 5.391984 | -0.2269 | 0.848938 |
| TNRC18 | 5.091466 | 0.342163 | 0.999532 | | 5.928582 | 0.459791 | 0.632485 | | 5.478732 | 0.152195 | 0.902699 |
| PON2 | 3.576095 | -0.3046 | 0.999532 | | 4.918552 | 0.434087 | 0.632485 | | 4.01271 | 0.745063 | 0.436389 |
| TTC23L | 2.086368 | -0.53268 | 0.999532 | | 2.176349 | -0.64119 | 0.632485 | | 1.646568 | -0.09056 | 0.974421 |
| ZBED5 | 3.924349 | -0.15143 | 0.999532 | | 5.310471 | 0.430153 | 0.632485 | | 4.353272 | 0.60025 | 0.533679 |
| PCNP | 11.75921 | -0.09685 | 0.999532 | | 14.80745 | 0.294143 | 0.632784 | | 12.1048 | 0.412958 | 0.515762 |
| ATN1 | 10.17323 | -0.29682 | 0.999532 | | 13.90146 | 0.273368 | 0.632895 | | 10.48999 | 0.588279 | 0.269743 |
| PRICKLE | 5.493296 | -0.02915 | 0.999532 | | 5.503243 | -0.44406 | 0.632895 | | 4.648459 | -0.3959 | 0.714657 |
| GLIPR1L | 2.320863 | -0.2237 | 0.999532 | | 2.416633 | -0.68334 | 0.632895 | | 1.766289 | -0.44293 | 0.806394 |
| GUCD1 | 3.178878 | 0.298399 | 0.999532 | | 3.531042 | 0.499329 | 0.632895 | | 3.480707 | 0.218473 | 0.88082 |
| CA5A | 3.242527 | 0.33073 | 0.999532 | | 2.737212 | -0.59696 | 0.632895 | | 2.828626 | -0.90847 | 0.455544 |
| DNAH1 | 3.54466 | 0.05416 | 0.999532 | | 3.269897 | -0.53443 | 0.632895 | | 2.846754 | -0.57056 | 0.647218 |
| FAM114A | 3.333006 | 0.044354 | 0.999532 | | 4.231879 | 0.460408 | 0.632895 | | 3.646339 | 0.430318 | 0.706407 |
| ODF3B | 3.137458 | -0.04434 | 0.999532 | | 3.056952 | -0.58051 | 0.632895 | | 2.626656 | -0.52385 | 0.70375 |
| PPP1R3B | 3.752548 | 0.030951 | 0.999532 | | 3.696487 | -0.50927 | 0.632895 | | 3.095992 | -0.51761 | 0.672763 |
| ERFL | 2.325881 | 0.138371 | 0.999532 | | 2.127381 | -0.67318 | 0.632895 | | 1.858802 | -0.78744 | 0.598185 |
| TBX3 | 6.184572 | -0.23191 | 0.999532 | | 9.247761 | 0.448834 | 0.632895 | | 7.426523 | 0.686616 | 0.403558 |
| PCGF5 | 5.869396 | 0.247351 | 0.999532 | | 6.94056 | 0.393289 | 0.632895 | | 6.538655 | 0.156738 | 0.892869 |
| OSTF1 | 5.528312 | 0.096011 | 0.999532 | | 7.299072 | 0.388085 | 0.632895 | | 6.227335 | 0.31659 | 0.740755 |
| EXOG | 4.292619 | -0.4872 | 0.999532 | | 4.568962 | -0.48343 | 0.632895 | | 3.369304 | 8.30E-05 | 0.999949 |
| CREBL2 | 7.156066 | 0.449735 | 0.999532 | | 7.700627 | 0.363078 | 0.632895 | | 7.607905 | -0.07094 | 0.955834 |
| CTDP1 | 4.020485 | -0.56837 | 0.999532 | | 4.191329 | -0.5091 | 0.632895 | | 3.027483 | 0.086156 | 0.968434 |
| IBSP | 2.328558 | -0.25696 | 0.999532 | | 2.342672 | -0.61602 | 0.632895 | | 1.901394 | -0.34116 | 0.856551 |
| RAI2 | 3.304893 | -0.50758 | 0.999532 | | 5.167399 | 0.425885 | 0.632895 | | 3.720015 | 0.953846 | 0.274496 |
| GLRX2 | 1.923244 | 0.310821 | 0.999532 | | 2.255367 | 0.634071 | 0.632986 | | 2.160398 | 0.338697 | 0.850099 |
| FUCA2 | 2.070274 | -0.28837 | 0.999532 | | 3.022912 | 0.523142 | 0.632986 | | 2.503785 | 0.828512 | 0.49299 |
| RDH8 | 2.548579 | 0.215226 | 0.999532 | | 2.166603 | -0.67347 | 0.632986 | | 2.032878 | -0.87398 | 0.542124 |
| LRRC75A | 2.482301 | -0.60517 | 0.999532 | | 2.877347 | -0.55432 | 0.63331 | | 1.927412 | 0.070686 | 0.977635 |
| PDCD2L | 2.555199 | 0.506945 | 0.999532 | | 1.915829 | -0.68073 | 0.633413 | | 2.173828 | -1.17224 | 0.340167 |
| GALR2 | 2.888822 | 0.046319 | 0.999532 | | 2.598126 | -0.6026 | 0.633752 | | 2.333402 | -0.62991 | 0.667066 |
| VPS37D | 4.111766 | 0.042375 | 0.999532 | | 4.075334 | -0.54349 | 0.633871 | | 3.571062 | -0.56696 | 0.660097 |
| SUFU | 3.67999 | -0.05745 | 0.999532 | | 4.790026 | 0.430706 | 0.63398 | | 4.12094 | 0.505572 | 0.627474 |
| CUL4B | 5.04442 | -0.01006 | 0.999532 | | 4.733535 | -0.44203 | 0.63398 | | 4.101604 | -0.42469 | 0.699971 |
| ARF3 | 7.682425 | 0.029395 | 0.999532 | | 9.655372 | 0.327891 | 0.63398 | | 8.238182 | 0.316741 | 0.709825 |
| PPT1 | 6.733598 | -0.36927 | 0.999532 | | 9.955658 | 0.332267 | 0.63398 | | 7.346271 | 0.727149 | 0.282041 |
| RBM25 | 4.956077 | 0.560106 | 0.999532 | | 5.036527 | 0.442695 | 0.63398 | | 5.395507 | -0.10108 | 0.946765 |
| KYAT3 | 2.825395 | 0.749682 | 0.999532 | | 2.999488 | 0.573881 | 0.63398 | | 3.106339 | -0.14528 | 0.934597 |
| ZNF17 | 2.618774 | 0.044355 | 0.999532 | | 2.292668 | -0.62911 | 0.634585 | | 2.100133 | -0.6662 | 0.66232 |
| UBTD1 | 2.475049 | 0.218014 | 0.999532 | | 2.86252 | 0.556911 | 0.634585 | | 2.741985 | 0.352256 | 0.818918 |
| LYPLAL1 | 2.75426 | 0.232638 | 0.999532 | | 3.615173 | 0.500426 | 0.634585 | | 3.176191 | 0.284301 | 0.836453 |
| COPS4 | 4.062851 | 0.207426 | 0.999532 | | 5.262465 | 0.447278 | 0.634585 | | 4.60134 | 0.255645 | 0.829716 |
| MRPL51 | 5.274444 | -0.09969 | 0.999532 | | 7.160493 | 0.378791 | 0.634585 | | 6.00722 | 0.497651 | 0.563455 |
| GGCT | 3.585442 | 0.221834 | 0.999532 | | 3.089122 | -0.58219 | 0.634585 | | 2.916442 | -0.79027 | 0.52894 |
| PKN1 | 6.821505 | 0.202133 | 0.999532 | | 7.987254 | 0.378036 | 0.634585 | | 7.517432 | 0.184314 | 0.876781 |
| VPS26B | 3.268788 | 0.297758 | 0.999532 | | 3.911209 | 0.504461 | 0.634585 | | 3.639358 | 0.22641 | 0.876107 |
| APOM | 3.563454 | 0.725151 | 0.999532 | | 3.631476 | 0.524743 | 0.634585 | | 4.010631 | -0.19389 | 0.892869 |
| DHX9 | 9.693007 | -0.14368 | 0.999532 | | 12.78384 | 0.333438 | 0.634585 | | 10.32311 | 0.499853 | 0.476728 |
| CYCS | 5.949699 | 0.115896 | 0.999532 | | 7.556259 | 0.359779 | 0.634585 | | 6.519624 | 0.26922 | 0.799627 |
| VCL | 4.449377 | 0.186861 | 0.999532 | | 5.254782 | 0.425972 | 0.634585 | | 4.849831 | 0.238999 | 0.861815 |
| TM7SF3 | 5.49817 | 0.043328 | 0.999532 | | 6.924496 | 0.367704 | 0.634585 | | 6.00318 | 0.343971 | 0.709825 |
| RNF112 | 2.152984 | 0.685848 | 0.999532 | | 2.666231 | 0.632873 | 0.634585 | | 2.693534 | -0.02859 | 0.992459 |
| SLC25A2 | 2.491924 | -0.61684 | 0.999532 | | 2.865416 | -0.55288 | 0.634585 | | 2.030195 | 0.084005 | 0.973788 |
| DUSP5 | 6.86683 | -0.34025 | 0.999532 | | 7.49344 | -0.38278 | 0.634585 | | 5.471068 | -0.01987 | 0.992286 |
| HDLBP | 9.346213 | -0.14418 | 0.999532 | | 13.37492 | 0.334581 | 0.634585 | | 10.46981 | 0.547737 | 0.511836 |
| CMIP | 5.042153 | 0.095067 | 0.999532 | | 6.331085 | 0.413412 | 0.634585 | | 5.76882 | 0.337066 | 0.736481 |
| NUP85 | 2.447757 | -0.15966 | 0.999532 | | 2.458907 | -0.58813 | 0.634585 | | 1.956274 | -0.40983 | 0.797544 |
| GPM6A | 2.899597 | -0.10399 | 0.999532 | | 2.865069 | -0.56962 | 0.634585 | | 2.4905 | -0.45027 | 0.75311 |
| MBNL2 | 9.07781 | 0.041344 | 0.999532 | | 11.44959 | 0.319685 | 0.634585 | | 9.710075 | 0.297905 | 0.690003 |
| RPL17 | 45.13864 | 0.167463 | 0.999532 | | 51.2744 | 0.224876 | 0.634585 | | 46.33101 | 0.071694 | 0.924131 |
| RBM47 | 3.472455 | -0.05121 | 0.999532 | | 4.633425 | 0.493411 | 0.634585 | | 3.813498 | 0.557104 | 0.656993 |
| PFKFB4 | 3.582186 | -0.44446 | 0.999532 | | 4.136047 | -0.49305 | 0.634775 | | 2.833862 | -0.0096 | 0.996891 |
| IPPK | 2.505955 | -0.3115 | 0.999532 | | 2.695049 | -0.58023 | 0.634821 | | 2.058183 | -0.24817 | 0.893828 |
| COG3 | 4.031753 | 0.432525 | 0.999532 | | 4.508872 | 0.456371 | 0.635581 | | 4.314404 | 0.040958 | 0.983211 |
| RBBP5 | 2.495074 | -0.42643 | 0.999532 | | 2.767754 | -0.58514 | 0.636368 | | 1.87139 | -0.1381 | 0.948127 |
| RER1 | 4.109833 | -0.16413 | 0.999532 | | 5.741472 | 0.400707 | 0.636368 | | 4.677701 | 0.584762 | 0.528909 |
| USP11 | 8.200243 | -0.02498 | 0.999532 | | 10.47754 | 0.333082 | 0.636368 | | 8.644823 | 0.377039 | 0.662576 |
| SLC22A2 | 3.87839 | -0.26632 | 0.999532 | | 5.466023 | 0.459176 | 0.636368 | | 4.240427 | 0.751048 | 0.445382 |
| NCL | 14.96387 | -0.00145 | 0.999532 | | 18.70925 | 0.302984 | 0.636368 | | 15.89637 | 0.325233 | 0.658469 |
| MUC20 | 8.076077 | -0.36197 | 0.999532 | | 9.159026 | -0.34542 | 0.636368 | | 6.697124 | 0.040881 | 0.978723 |
| VWC2 | 2.402065 | -0.05492 | 0.999532 | | 2.132314 | -0.63917 | 0.636368 | | 1.925776 | -0.5756 | 0.724646 |
| CTSD | 73.57348 | -0.02133 | 0.999532 | | 89.88487 | 0.209667 | 0.636368 | | 75.58947 | 0.255078 | 0.598934 |

| | | | | | | | | | | | |
|---|---|---|---|---|---|---|---|---|---|---|---|
| HNRNPD | 20.6016 | -0.10987 | 0.999532 | | 26.20011 | 0.259761 | 0.636368 | | 21.60868 | 0.389714 | 0.46286 |
| AMH | 3.596876 | 0.2005 | 0.999532 | | 3.011168 | -0.54512 | 0.636368 | | 2.891677 | -0.73793 | 0.540342 |
| CUL4A | 4.384196 | 0.520388 | 0.999532 | | 4.687564 | 0.456385 | 0.636368 | | 4.747874 | -0.04544 | 0.978805 |
| PROC | 2.460257 | -0.30162 | 0.999532 | | 2.478424 | -0.59581 | 0.636368 | | 1.912454 | -0.27944 | 0.881837 |
| INPP5A | 4.242234 | -0.11424 | 0.999532 | | 5.821983 | 0.412967 | 0.636375 | | 4.75794 | 0.543232 | 0.549022 |
| DHRS7 | 4.526889 | -0.0244 | 0.999532 | | 6.099035 | 0.39398 | 0.636422 | | 5.035443 | 0.434891 | 0.644433 |
| AKAP9 | 3.539634 | -0.22219 | 0.999532 | | 5.040473 | 0.457934 | 0.636422 | | 4.011923 | 0.698842 | 0.486743 |
| ASS1 | 8.653727 | -0.06717 | 0.999532 | | 11.1119 | 0.348153 | 0.636422 | | 9.130137 | 0.418876 | 0.692127 |
| ZNF510 | 2.969275 | -0.28903 | 0.999532 | | 2.893921 | -0.56686 | 0.636634 | | 2.375928 | -0.26368 | 0.88082 |
| NPC1L1 | 2.147749 | -0.00357 | 0.999532 | | 2.105004 | -0.6587 | 0.636634 | | 1.734016 | -0.632 | 0.69872 |
| TNFRSF6 | 2.687594 | -0.06069 | 0.999532 | | 2.639802 | -0.59289 | 0.636634 | | 2.282925 | -0.51089 | 0.739496 |
| TMEM132 | 2.098962 | -0.27136 | 0.999532 | | 2.125835 | -0.63126 | 0.636634 | | 1.753557 | -0.34108 | 0.862823 |
| ZNF485 | 2.216935 | 0.130596 | 0.999532 | | 1.946633 | -0.68117 | 0.636634 | | 1.69157 | -0.79173 | 0.60629 |
| SRSF8 | 6.192221 | 0.377225 | 0.999532 | | 6.952027 | 0.366675 | 0.636634 | | 6.75893 | 0.015118 | 0.994537 |
| LIPT2 | 3.122364 | 0.306201 | 0.999532 | | 2.644316 | -0.57507 | 0.636634 | | 2.626229 | -0.86187 | 0.455544 |
| PTGR1 | 4.891289 | 0.05503 | 0.999532 | | 6.049312 | 0.408642 | 0.636634 | | 5.276076 | 0.364837 | 0.735774 |
| ZNF573 | 3.048856 | 0.045717 | 0.999532 | | 2.754039 | -0.54949 | 0.636634 | | 2.475402 | -0.5799 | 0.658332 |
| ABCC10 | 2.877171 | 0.379711 | 0.999532 | | 3.319377 | 0.512743 | 0.636634 | | 3.141353 | 0.145974 | 0.92697 |
| RNF227 | 3.621361 | 0.398553 | 0.999532 | | 2.864365 | -0.58781 | 0.636634 | | 3.040911 | -0.96783 | 0.359441 |
| DIXDC1 | 2.983057 | -0.14901 | 0.999532 | | 3.065503 | -0.54921 | 0.636644 | | 2.358887 | -0.3785 | 0.803301 |
| LMAN1 | 8.061182 | -0.20634 | 0.999532 | | 10.86341 | 0.326092 | 0.636644 | | 8.555726 | 0.547537 | 0.512428 |
| CCR8 | 3.624878 | 0.659255 | 0.999532 | | 2.757275 | -0.59505 | 0.636644 | | 2.996168 | -1.23398 | 0.2006 |
| TAFA5 | 2.611722 | -0.08556 | 0.999532 | | 2.62129 | -0.62846 | 0.636644 | | 2.144443 | -0.52181 | 0.740093 |
| AGO1 | 4.455016 | 0.129299 | 0.999532 | | 5.504063 | 0.412742 | 0.636644 | | 4.888748 | 0.300588 | 0.785695 |
| LRRC15 | 2.10955 | -0.05163 | 0.999532 | | 2.064186 | -0.71018 | 0.636644 | | 1.630412 | -0.63885 | 0.713778 |
| BHLHE41 | 2.373367 | 0.156625 | 0.999532 | | 2.977688 | 0.563751 | 0.636719 | | 2.81018 | 0.421415 | 0.770077 |
| TMEM242 | 2.589236 | 0.324151 | 0.999532 | | 2.203603 | -0.61917 | 0.636841 | | 2.126645 | -0.9239 | 0.456688 |
| OR2A4 | 2.862139 | -0.19962 | 0.999532 | | 2.874576 | -0.54434 | 0.636841 | | 2.337456 | -0.32674 | 0.843463 |
| SIPA1L1 | 3.341204 | 0.183455 | 0.999532 | | 2.926418 | -0.5463 | 0.636841 | | 2.775189 | -0.71345 | 0.534852 |
| ABCD2 | 2.886952 | -0.43049 | 0.999532 | | 2.988604 | -0.58274 | 0.637308 | | 2.294577 | -0.13601 | 0.951656 |
| CARD10 | 3.916619 | 0.236329 | 0.999532 | | 4.636699 | 0.442224 | 0.637661 | | 4.416058 | 0.219998 | 0.864245 |
| FAM120C | 3.461703 | -0.35704 | 0.999532 | | 3.570615 | -0.51175 | 0.637725 | | 2.635404 | -0.14124 | 0.939913 |
| TNRC6C | 4.363601 | 0.296485 | 0.999532 | | 5.19383 | 0.421313 | 0.637725 | | 4.760222 | 0.141066 | 0.912867 |
| KRTAP10 | 3.961596 | 0.036027 | 0.999532 | | 3.871471 | -0.55036 | 0.637725 | | 3.225821 | -0.55941 | 0.663009 |
| RPS6KB2 | 4.763546 | 0.144636 | 0.999532 | | 6.011785 | 0.388808 | 0.637725 | | 5.261803 | 0.261328 | 0.806394 |
| RET | 3.155371 | -0.11559 | 0.999532 | | 3.224904 | -0.54029 | 0.637725 | | 2.550372 | -0.40547 | 0.769877 |
| CCT4 | 7.06989 | 0.299019 | 0.999532 | | 7.776503 | 0.382872 | 0.637725 | | 7.26123 | 0.09318 | 0.945032 |
| CPNE9 | 3.060083 | 0.28015 | 0.999532 | | 2.625368 | -0.60929 | 0.637725 | | 2.422055 | -0.86975 | 0.485816 |
| ARIH2 | 3.506046 | -0.16452 | 0.999532 | | 5.153366 | 0.423023 | 0.637725 | | 3.991591 | 0.599722 | 0.534789 |
| CCND2 | 4.112713 | 0.068377 | 0.999532 | | 5.470356 | 0.439741 | 0.637725 | | 4.523384 | 0.385582 | 0.725814 |
| CSPP1 | 2.689606 | 0.575444 | 0.999532 | | 2.932142 | 0.54217 | 0.637725 | | 2.884736 | -0.0107 | 0.996891 |
| TRIAP1 | 3.007913 | -0.18649 | 0.999532 | | 2.759713 | -0.58786 | 0.637767 | | 2.394258 | -0.39193 | 0.824012 |
| UCHL3 | 3.099492 | -0.17449 | 0.999532 | | 3.104466 | -0.53837 | 0.637767 | | 2.484187 | -0.34542 | 0.819366 |
| CES4A | 3.806672 | -0.22529 | 0.999532 | | 4.059052 | -0.49429 | 0.637767 | | 3.072483 | -0.24315 | 0.87591 |
| INPP5B | 3.150623 | 0.125808 | 0.999532 | | 3.749829 | 0.472207 | 0.637767 | | 3.521717 | 0.365177 | 0.7719 |
| CDYL2 | 2.922904 | -0.57667 | 0.999532 | | 3.334399 | -0.53554 | 0.638911 | | 2.176367 | 0.060786 | 0.979434 |
| NET1 | 4.442288 | 0.055371 | 0.999532 | | 5.552663 | 0.424197 | 0.6393 | | 4.958927 | 0.380987 | 0.717627 |
| CDHR3 | 2.930237 | -0.61198 | 0.999532 | | 3.428154 | -0.54275 | 0.6393 | | 2.310717 | 0.08689 | 0.970811 |
| ATP6V1B | 4.574041 | 0.906545 | 0.999532 | | 4.421428 | 0.510562 | 0.639729 | | 4.808567 | -0.40927 | 0.780291 |
| DAP | 9.19118 | -0.03486 | 0.999532 | | 11.55978 | 0.303427 | 0.639729 | | 9.771263 | 0.353351 | 0.663009 |
| GRINA | 4.836997 | 0.250292 | 0.999532 | | 5.7387 | 0.408717 | 0.640305 | | 5.171914 | 0.174643 | 0.883792 |
| ZNF415 | 3.456913 | 0.147449 | 0.999532 | | 3.141334 | -0.52979 | 0.640305 | | 2.920116 | -0.66171 | 0.566812 |
| CDH2 | 2.358773 | 0.638981 | 0.999532 | | 2.763679 | 0.567938 | 0.640338 | | 2.626786 | -0.04629 | 0.985693 |
| DDAH1 | 3.346906 | -0.27613 | 0.999532 | | 4.652322 | 0.422651 | 0.64037 | | 3.615413 | 0.710371 | 0.470694 |
| STX5 | 3.324465 | 0.550384 | 0.999532 | | 3.641824 | 0.494207 | 0.6404 | | 3.488765 | -0.0375 | 0.98637 |
| SLC25A4 | 2.352349 | -0.5618 | 0.999532 | | 2.833597 | -0.57988 | 0.640962 | | 1.974317 | 0.008774 | 0.997967 |
| C3orf80 | 1.931759 | -0.37967 | 0.999532 | | 2.016273 | -0.65688 | 0.640962 | | 1.686086 | -0.25581 | 0.906811 |
| GEN1 | 2.554459 | -0.10961 | 0.999532 | | 2.381445 | -0.62483 | 0.641626 | | 1.960395 | -0.5032 | 0.759369 |
| PIGZ | 1.637683 | 0.507779 | 0.999532 | | 1.809162 | 0.677289 | 0.641626 | | 1.887687 | 0.183898 | 0.931488 |
| ANKIB1 | 3.32838 | 0.116628 | 0.999532 | | 4.051713 | 0.473163 | 0.641824 | | 3.570093 | 0.379115 | 0.745908 |
| TMOD4 | 2.35081 | -0.10656 | 0.999532 | | 2.285391 | -0.61049 | 0.641824 | | 1.935728 | -0.48473 | 0.759211 |
| MTRNR2L | 5.945941 | 0.225385 | 0.999532 | | 7.191932 | 0.397523 | 0.642356 | | 6.570703 | 0.193063 | 0.875488 |
| ZFP37 | 3.661651 | 0.34787 | 0.999532 | | 3.300331 | -0.53312 | 0.642356 | | 3.197883 | -0.85627 | 0.413519 |
| COPB2 | 8.787408 | -0.1017 | 0.999532 | | 11.53043 | 0.309012 | 0.642356 | | 9.539049 | 0.430923 | 0.524206 |
| TMIGD1 | 2.68051 | 0.411974 | 0.999532 | | 2.043876 | -0.6883 | 0.642905 | | 2.261762 | -1.07666 | 0.442274 |
| IL6ST | 22.36595 | -0.32918 | 0.999532 | | 30.86946 | 0.268827 | 0.643149 | | 23.91354 | 0.619044 | 0.204889 |
| SETMAR | 3.014017 | -0.21028 | 0.999532 | | 3.059838 | -0.59129 | 0.643149 | | 2.381639 | -0.36008 | 0.82381 |
| GJB7 | 2.986106 | 0.410944 | 0.999532 | | 2.543262 | -0.64926 | 0.643235 | | 2.500295 | -1.04462 | 0.363418 |
| NUP107 | 2.754258 | -0.1243 | 0.999532 | | 3.706772 | 0.476142 | 0.643235 | | 3.086816 | 0.61068 | 0.618451 |
| ATG14 | 3.324927 | -0.2992 | 0.999532 | | 4.795494 | 0.452299 | 0.643294 | | 3.765858 | 0.770637 | 0.435423 |
| CHTOP | 5.781992 | -0.05081 | 0.999532 | | 7.652114 | 0.396164 | 0.643337 | | 6.30977 | 0.471175 | 0.565022 |
| ANO1 | 4.562943 | -0.5652 | 0.999532 | | 6.754728 | 0.439084 | 0.643579 | | 5.222594 | 0.957614 | 0.323586 |
| OXCT1 | 1.837197 | 0.318836 | 0.999532 | | 2.100987 | 0.612822 | 0.643639 | | 2.074115 | 0.309403 | 0.862823 |
| PIANP | 4.629066 | 0.212638 | 0.999532 | | 4.119425 | -0.49422 | 0.644411 | | 3.97186 | -0.7025 | 0.511836 |
| DHX37 | 2.551958 | -0.18377 | 0.999532 | | 2.452941 | -0.60392 | 0.644524 | | 2.018756 | -0.41033 | 0.813438 |
| DAPL1 | 2.033756 | 0.57698 | 0.999532 | | 2.208541 | 0.62049 | 0.644524 | | 2.337845 | 0.060418 | 0.980617 |
| DDT | 4.068936 | 0.472383 | 0.999532 | | 4.95618 | 0.436863 | 0.645179 | | 4.487331 | -0.02034 | 0.993111 |
| PRAG1 | 3.494361 | -0.38047 | 0.999532 | | 3.623161 | -0.51024 | 0.645179 | | 2.780273 | -0.10397 | 0.958191 |
| ZNF197 | 2.746638 | 0.394826 | 0.999532 | | 3.379029 | 0.516779 | 0.645721 | | 3.223042 | 0.143356 | 0.931619 |
| TSSK1B | 2.196404 | -0.17495 | 0.999532 | | 2.161159 | -0.66488 | 0.645721 | | 1.742718 | -0.4716 | 0.79686 |
| AK9 | 3.768482 | 0.118121 | 0.999532 | | 3.655747 | -0.49566 | 0.645721 | | 3.201456 | -0.59519 | 0.590442 |
| RUFY1 | 5.817175 | 0.579969 | 0.999532 | | 6.289233 | 0.401082 | 0.645721 | | 6.40854 | -0.15633 | 0.894612 |
| GLRX3 | 4.631771 | 0.11397 | 0.999532 | | 5.709387 | 0.397506 | 0.645721 | | 4.988132 | 0.303332 | 0.765852 |
| CCL23 | 2.289084 | -0.21421 | 0.999532 | | 2.256626 | -0.62866 | 0.645721 | | 1.898289 | -0.39862 | 0.832794 |
| INSYN2A | 2.20383 | 0.083698 | 0.999532 | | 2.122446 | -0.65837 | 0.645721 | | 1.786983 | -0.71485 | 0.649779 |
| FGFBP3 | 2.522371 | -0.17623 | 0.999532 | | 2.544291 | -0.62835 | 0.645721 | | 1.912305 | -0.42894 | 0.79686 |
| MRM3 | 6.289856 | 0.20897 | 0.999532 | | 5.673088 | -0.42855 | 0.645721 | | 5.385961 | -0.61871 | 0.534495 |
| EPAS1 | 99.65353 | -0.30646 | 0.999532 | | 135.6786 | 0.256045 | 0.645721 | | 105.9941 | 0.585426 | 0.168532 |
| FERD3L | 2.632131 | 0.014397 | 0.999532 | | 2.397249 | -0.62695 | 0.645721 | | 2.07313 | -0.62261 | 0.679818 |
| C1QTNF4 | 2.785436 | 0.597012 | 0.999532 | | 2.011185 | -0.67028 | 0.645721 | | 2.250574 | -1.25086 | 0.271669 |
| KMT2D | 6.121478 | 0.252947 | 0.999532 | | 7.48546 | 0.419138 | 0.645721 | | 6.520089 | 0.206776 | 0.870694 |
| LCE1D | 5.722115 | 0.019679 | 0.999532 | | 5.469979 | -0.45557 | 0.645749 | | 4.844936 | -0.46216 | 0.705046 |
| ZEB1 | 3.737604 | -0.29141 | 0.999532 | | 5.537641 | 0.419102 | 0.645749 | | 4.446632 | 0.732665 | 0.423641 |
| ZSCAN23 | 2.768436 | 0.085461 | 0.999532 | | 2.476314 | -0.59245 | 0.645749 | | 2.145005 | -0.66473 | 0.646821 |
| GNRH2 | 2.835153 | -0.18673 | 0.999532 | | 2.730196 | -0.59392 | 0.645857 | | 2.331591 | -0.39705 | 0.816589 |
| IGHG1 | 7.071817 | 0.324102 | 0.999532 | | 5.577974 | -0.7669 | 0.646852 | | 5.526094 | -1.02093 | 0.566812 |
| HCAR2 | 3.433578 | -0.30334 | 0.999532 | | 3.560494 | -0.51651 | 0.646852 | | 2.732495 | -0.19554 | 0.906985 |
| ANAPC16 | 8.995354 | 0.170531 | 0.999532 | | 10.96087 | 0.379617 | 0.647392 | | 9.695382 | 0.241927 | 0.817974 |
| TGFBR3 | 23.39857 | 0.335608 | 0.999532 | | 26.74959 | 0.35779 | 0.647392 | | 24.99934 | 0.053453 | 0.96913 |
| ARHGAP | 5.252941 | -0.30916 | 0.999532 | | 5.573654 | -0.40754 | 0.647392 | | 4.302827 | -0.08333 | 0.958529 |
| TFIP11 | 3.790155 | 0.611303 | 0.999532 | | 4.239092 | 0.46139 | 0.647678 | | 4.27926 | -0.13062 | 0.926957 |
| C4orf45 | 4.228941 | 0.099218 | 0.999532 | | 4.133201 | -0.51371 | 0.647719 | | 3.599595 | -0.58826 | 0.626709 |
| AP2A1 | 5.572892 | -0.27156 | 0.999532 | | 7.598551 | 0.365629 | 0.647814 | | 6.196516 | 0.657003 | 0.391643 |
| MBD5 | 3.251194 | 0.192619 | 0.999532 | | 2.96075 | -0.53035 | 0.647814 | | 2.814971 | -0.69991 | 0.546846 |
| RAB35 | 3.230827 | 0.139077 | 0.999532 | | 4.355349 | 0.458085 | 0.647825 | | 3.765536 | 0.344616 | 0.785012 |
| ESF1 | 3.566997 | 0.018932 | 0.999532 | | 3.345475 | -0.51118 | 0.647866 | | 2.942384 | -0.51205 | 0.668481 |
| NDUFB1 | 15.57583 | -0.18966 | 0.999532 | | 20.53474 | 0.27888 | 0.647866 | | 16.61301 | 0.487557 | 0.356879 |
| CYP11A1 | 2.38416 | -0.40149 | 0.999532 | | 2.535048 | -0.63184 | 0.6479 | | 1.824301 | -0.20793 | 0.920259 |
| SRF | 4.262941 | -0.03342 | 0.999532 | | 5.324748 | 0.409938 | 0.6479 | | 4.495418 | 0.462818 | 0.647218 |
| ATF6B | 7.239162 | 0.083468 | 0.999532 | | 8.967787 | 0.345489 | 0.6479 | | 7.797738 | 0.281372 | 0.737952 |
| EMX1 | 3.463313 | -0.10335 | 0.999532 | | 4.666142 | 0.442917 | 0.6479 | | 3.850223 | 0.562121 | 0.602151 |
| DOLPP1 | 3.107505 | -0.139 | 0.999532 | | 3.055668 | -0.55958 | 0.6479 | | 2.495664 | -0.40171 | 0.791466 |
| SEMA4A | 2.422001 | -0.40529 | 0.999532 | | 2.485274 | -0.58914 | 0.6479 | | 1.78666 | -0.17012 | 0.939049 |
| TFDP1 | 6.011695 | -0.13645 | 0.999532 | | 6.277014 | -0.38069 | 0.6479 | | 5.02984 | -0.22538 | 0.842754 |
| FIS1 | 5.334115 | -0.02671 | 0.999532 | | 6.856819 | 0.389088 | 0.6479 | | 5.757978 | 0.437812 | 0.632802 |
| ZNF432 | 3.005198 | -0.42136 | 0.999532 | | 3.317373 | -0.49974 | 0.6479 | | 2.397358 | -0.05475 | 0.982124 |
| ORM2 | 6.811861 | 0.445238 | 0.999532 | | 5.561899 | -0.44161 | 0.6479 | | 5.795195 | -0.86853 | 0.254511 |
| SRP68 | 6.874063 | -0.19252 | 0.999532 | | 7.525248 | -0.35412 | 0.6479 | | 5.642911 | -0.14992 | 0.904801 |
| GCAT | 2.652744 | 0.389464 | 0.999532 | | 2.28334 | -0.63115 | 0.6479 | | 2.254484 | -1.00233 | 0.406785 |
| H3C12 | 2.772572 | 0.130331 | 0.999532 | | 2.360408 | -0.62539 | 0.6479 | | 2.208757 | -0.73609 | 0.611084 |
| PARP6 | 5.366647 | 0.150305 | 0.999532 | | 6.384562 | 0.409779 | 0.6479 | | 5.621842 | 0.282676 | 0.791466 |
| UBXN7 | 3.228154 | 0.429198 | 0.999532 | | 3.689341 | 0.478771 | 0.648061 | | 3.595941 | 0.067824 | 0.971732 |
| GP5 | 3.589839 | -0.1081 | 0.999532 | | 3.596837 | -0.49958 | 0.64879 | | 2.996459 | -0.37063 | 0.794473 |
| FAM118B | 2.478378 | 0.916263 | 0.999532 | | 2.603294 | 0.577272 | 0.64879 | | 2.797318 | -0.32027 | 0.823571 |
| USP34 | 5.694783 | 0.269772 | 0.999532 | | 6.56548 | 0.391155 | 0.64879 | | 5.996158 | 0.136203 | 0.911127 |
| ERBIN | 6.013337 | 0.074104 | 0.999532 | | 7.344497 | 0.359986 | 0.64879 | | 6.640493 | 0.315608 | 0.739496 |
| CDC40 | 3.402278 | 0.32314 | 0.999532 | | 2.730656 | -0.56269 | 0.64879 | | 2.689682 | -0.88097 | 0.468942 |
| CXCL1 | 2.897071 | -0.0744 | 0.999532 | | 2.793571 | -0.56189 | 0.64879 | | 2.349214 | -0.47423 | 0.757722 |
| MAGOH | 5.010025 | 0.22845 | 0.999532 | | 6.341839 | 0.371487 | 0.64879 | | 5.718942 | 0.164575 | 0.88476 |
| CCDC148 | 3.039342 | 0.263024 | 0.999532 | | 2.600724 | -0.59889 | 0.64879 | | 2.452914 | -0.84462 | 0.515762 |
| ADAM28 | 2.517347 | 0.03441 | 0.999532 | | 2.35333 | -0.64659 | 0.64879 | | 1.92439 | -0.65934 | 0.662584 |
| ACLY | 4.770933 | -0.33937 | 0.999532 | | 6.787828 | 0.395301 | 0.64879 | | 5.288657 | 0.753643 | 0.348004 |
| KIAA2026 | 2.433206 | 0.395728 | 0.999532 | | 3.000109 | 0.533936 | 0.64879 | | 2.845408 | 0.156589 | 0.92697 |
| VPS35 | 5.106986 | 0.221162 | 0.999532 | | 6.027434 | 0.3829 | 0.64879 | | 5.642084 | 0.180008 | 0.876781 |
| LRP4 | 2.768169 | -0.30044 | 0.999532 | | 2.81952 | -0.5499 | 0.64879 | | 2.270093 | -0.23574 | 0.893853 |
| DAZAP2 | 17.84611 | -0.09624 | 0.999532 | | 22.92094 | 0.283546 | 0.64879 | | 18.97467 | 0.379256 | 0.486743 |
| ANTXR1 | 11.11079 | -0.12971 | 0.999532 | | 14.16946 | 0.295584 | 0.64879 | | 12.03352 | 0.443543 | 0.486743 |
| ZMIZ2 | 7.595305 | -0.01161 | 0.999532 | | 9.756868 | 0.371845 | 0.64879 | | 8.512188 | 0.384664 | 0.682165 |
| CREG1 | 6.819375 | 0.068503 | 0.999532 | | 8.581773 | 0.357272 | 0.64879 | | 7.374298 | 0.308978 | 0.716189 |
| UBE2K | 6.413462 | 0.017677 | 0.999532 | | 8.162172 | 0.357527 | 0.64879 | | 6.954407 | 0.357847 | 0.690003 |
| HPCA | 2.951303 | 0.363575 | 0.999532 | | 2.341368 | -0.62478 | 0.64879 | | 2.579109 | -0.98485 | 0.444687 |
| POR | 6.083168 | -0.21522 | 0.999532 | | 6.20218 | -0.39305 | 0.64879 | | 4.931893 | -0.16702 | 0.897096 |
| CORO2B | 3.343004 | 0.064454 | 0.999532 | | 4.464141 | 0.504988 | 0.64879 | | 3.806879 | 0.460263 | 0.722703 |
| PKNOX2 | 4.930529 | 0.445077 | 0.999532 | | 4.08975 | -0.523 | 0.64879 | | 4.236198 | -0.9482 | 0.310043 |
| ATG16L2 | 2.413029 | -0.11241 | 0.999532 | | 2.387774 | -0.58985 | 0.64879 | | 1.963648 | -0.45514 | 0.782299 |
| G3BP1 | 5.114309 | -0.28441 | 0.999532 | | 7.60016 | 0.399509 | 0.64879 | | 5.871226 | 0.699051 | 0.396754 |
| TTC1 | 4.794983 | 0.047294 | 0.999532 | | 6.01647 | 0.395561 | 0.64879 | | 5.23068 | 0.369851 | 0.70284 |
| NUP88 | 3.850127 | -0.10038 | 0.999532 | | 3.771998 | -0.4674 | 0.64879 | | 3.13607 | -0.35639 | 0.792539 |
| IGSF21 | 2.935155 | -0.27576 | 0.999532 | | 3.008325 | -0.5565 | 0.64879 | | 2.369907 | -0.26386 | 0.883116 |
| PDE4A | 2.075435 | 0.365967 | 0.999532 | | 2.456979 | 0.583245 | 0.648795 | | 2.439969 | 0.236622 | 0.892328 |
| CRYL1 | 3.8081 | 0.198527 | 0.999532 | | 4.676995 | 0.487065 | 0.648795 | | 4.072089 | 0.32198 | 0.792688 |
| ZNF229 | 2.555183 | -0.01982 | 0.999532 | | 2.528248 | -0.5865 | 0.648795 | | 2.042647 | -0.54206 | 0.718643 |
| RAB12 | 7.327467 | 0.251855 | 0.999532 | | 8.337867 | 0.349738 | 0.648795 | | 7.811571 | 0.11864 | 0.907968 |
| BCL7A | 1.960166 | -0.09347 | 0.999532 | | 1.855843 | -0.66307 | 0.648795 | | 1.538087 | -0.55222 | 0.751444 |
| SDC3 | 9.407081 | -0.50304 | 0.999532 | | 11.2186 | -0.31906 | 0.648795 | | 7.804763 | 0.209597 | 0.827267 |
| ARPC2 | 10.94026 | -0.25162 | 0.999532 | | 15.07086 | 0.297549 | 0.648795 | | 11.98939 | 0.574848 | 0.282041 |
| BCL9 | 3.42115 | -0.39262 | 0.999532 | | 3.83544 | -0.48746 | 0.648795 | | 2.91386 | -0.07049 | 0.974421 |
| TTC8 | 2.141882 | -0.09007 | 0.999532 | | 2.831293 | 0.541096 | 0.649261 | | 2.315932 | 0.64537 | 0.634624 |
| RHBDF1 | 3.818819 | -0.4604 | 0.999532 | | 4.094867 | -0.47661 | 0.649972 | | 2.883175 | 0.006097 | 0.997967 |
| TNF | 2.436635 | -0.00674 | 0.999532 | | 2.393915 | -0.60068 | 0.649972 | | 2.089568 | -0.57622 | 0.703463 |
| VXN | 2.081121 | -0.64248 | 0.999532 | | 2.24341 | -0.59694 | 0.650058 | | 1.610255 | 0.057876 | 0.985693 |
| IPO7 | 6.2079 | 0.115129 | 0.999532 | | 5.581477 | -0.39957 | 0.650058 | | 5.194635 | -0.51553 | 0.600983 |
| PTGFRN | 2.508795 | -0.67724 | 0.999532 | | 3.018357 | -0.55099 | 0.650058 | | 1.945231 | 0.147933 | 0.945129 |
| DUSP8 | 6.699482 | 0.101502 | 0.999532 | | 6.352105 | -0.44365 | 0.650058 | | 5.847645 | -0.52412 | 0.646821 |
| SRD5A3 | 2.27113 | 0.499804 | 0.999532 | | 2.721831 | 0.567829 | 0.650058 | | 2.64265 | 0.087512 | 0.967951 |
| ZNF442 | 1.715221 | 0.281249 | 0.999532 | | 2.192525 | 0.613589 | 0.650058 | | 2.059866 | 0.352065 | 0.839811 |
| HSD17B3 | 3.022616 | 0.216662 | 0.999532 | | 2.488097 | -0.56612 | 0.650058 | | 2.518116 | -0.76898 | 0.542124 |
| KAT2B | 3.936022 | 0.288701 | 0.999532 | | 4.569389 | 0.431914 | 0.650058 | | 4.397245 | 0.153196 | 0.911296 |
| SEPTIN7 | 17.13391 | 0.008621 | 0.999532 | | 21.10825 | 0.248185 | 0.650058 | | 17.99839 | 0.263368 | 0.702463 |
| RPS6KA1 | 2.627301 | -0.04659 | 0.999532 | | 3.573073 | 0.502725 | 0.650058 | | 2.974762 | 0.568719 | 0.680097 |
| AHCY | 3.978079 | -0.01653 | 0.999532 | | 5.41683 | 0.42579 | 0.650058 | | 4.484854 | 0.460122 | 0.658045 |
| C16orf72 | 3.09439 | -0.09151 | 0.999532 | | 4.280335 | 0.473639 | 0.650058 | | 3.364864 | 0.566335 | 0.624566 |
| CEP44 | 1.900958 | 0.657227 | 0.999532 | | 2.203758 | 0.622969 | 0.650058 | | 2.226116 | -0.01315 | 0.996891 |
| ARG2 | 2.546924 | -0.57973 | 0.999532 | | 2.872384 | -0.568 | 0.650058 | | 1.925905 | 0.030751 | 0.992586 |
| CISD2 | 6.100719 | 0.426884 | 0.999532 | | 4.986207 | -0.46384 | 0.650058 | | 5.328172 | -0.88418 | 0.225902 |
| COP1 | 6.512294 | -0.22463 | 0.999532 | | 7.220473 | -0.35786 | 0.650058 | | 5.729499 | -0.11182 | 0.92883 |
| ROBO4 | 11.69501 | -0.16853 | 0.999532 | | 15.67444 | 0.261345 | 0.650058 | | 12.60815 | 0.455324 | 0.464674 |
| CD24 | 6.165534 | 0.283293 | 0.999532 | | 7.570019 | 0.48292 | 0.650058 | | 6.617524 | 0.259836 | 0.818918 |
| WDFY2 | 1.974787 | 0.127132 | 0.999532 | | 2.65259 | 0.557977 | 0.650058 | | 2.296679 | 0.447577 | 0.756702 |
| IRX6 | 2.806085 | -0.34228 | 0.999532 | | 2.851632 | -0.55424 | 0.650058 | | 2.242856 | -0.19051 | 0.921 |
| PRADC1 | 2.244106 | 0.275305 | 0.999532 | | 3.102759 | 0.569222 | 0.650058 | | 2.757191 | 0.323059 | 0.840357 |
| DDA1 | 5.364321 | -0.1458 | 0.999532 | | 5.653203 | -0.41099 | 0.650058 | | 4.469322 | -0.24702 | 0.836453 |
| H1-1 | 2.842188 | 0.33722 | 0.999532 | | 2.33197 | -0.58952 | 0.650058 | | 2.373621 | -0.91002 | 0.445382 |
| GALNT4 | 2.401578 | -0.19177 | 0.999532 | | 2.214977 | -0.59563 | 0.650058 | | 1.94294 | -0.39721 | 0.82526 |
| RFESD | 2.305384 | -0.01351 | 0.999532 | | 2.156713 | -0.62832 | 0.650147 | | 1.924381 | -0.59943 | 0.713173 |
| LGALS14 | 3.232547 | 0.641679 | 0.999532 | | 2.276343 | -0.64974 | 0.650147 | | 2.683912 | -1.27268 | 0.241605 |
| CD99 | 7.941653 | -0.29311 | 0.999532 | | 8.566504 | -0.32715 | 0.650147 | | 6.567628 | -0.02168 | 0.991159 |
| LBP | 2.635561 | -0.13951 | 0.999532 | | 2.506649 | -0.62002 | 0.650147 | | 2.080262 | -0.46032 | 0.791466 |
| BCL7C | 8.676433 | -0.13328 | 0.999532 | | 11.56117 | 0.287175 | 0.650147 | | 9.306043 | 0.439886 | 0.521228 |
| DDX54 | 4.65299 | 0.175111 | 0.999532 | | 5.367627 | 0.404069 | 0.650147 | | 5.103244 | 0.241639 | 0.839373 |
| RNF207 | 2.596359 | -0.00884 | 0.999532 | | 2.572144 | -0.56523 | 0.650147 | | 2.087696 | -0.53275 | 0.717627 |
| TYK2 | 5.456351 | 0.483019 | 0.999532 | | 5.947235 | 0.420159 | 0.650147 | | 6.036113 | -0.04705 | 0.977514 |
| ROCK1 | 6.665469 | 0.242983 | 0.999532 | | 7.819802 | 0.351118 | 0.650147 | | 7.158545 | 0.129515 | 0.917499 |
| PADI6 | 2.557392 | 0.274903 | 0.999532 | | 2.209997 | -0.64872 | 0.650147 | | 2.2376 | -0.9098 | 0.515762 |
| CDK17 | 4.452922 | 0.080108 | 0.999532 | | 5.200599 | 0.397281 | 0.650234 | | 4.827854 | 0.335969 | 0.751503 |
| DEFB106 | 2.544482 | 0.607786 | 0.999532 | | 1.901403 | -0.6629 | 0.650328 | | 2.07465 | -1.25194 | 0.26986 |
| NAT8B | 8.589215 | 0.384893 | 0.999532 | | 9.506504 | 0.33315 | 0.650328 | | 9.025077 | -0.02735 | 0.986539 |
| TTBK2 | 3.695275 | -0.32101 | 0.999532 | | 4.061058 | -0.49077 | 0.650328 | | 3.033582 | -0.15675 | 0.923834 |
| SLC13A3 | 3.83138 | 0.292785 | 0.999532 | | 4.628228 | 0.451973 | 0.650328 | | 4.250506 | 0.175475 | 0.90424 |
| MAMLD1 | 2.633302 | 0.153109 | 0.999532 | | 2.315047 | -0.59127 | 0.650333 | | 2.183648 | -0.72936 | 0.579378 |
| MUC8 | 2.492825 | 0.122939 | 0.999532 | | 2.209063 | -0.61572 | 0.650333 | | 1.946652 | -0.71944 | 0.619423 |
| VEGFD | 3.067238 | -0.00839 | 0.999532 | | 2.889802 | -0.55385 | 0.650333 | | 2.514426 | -0.53056 | 0.709825 |
| AK6 | 3.780664 | -0.21918 | 0.999532 | | 4.039479 | -0.45668 | 0.650379 | | 3.173923 | -0.21847 | 0.881837 |
| SH2D2A | 2.366936 | -0.09268 | 0.999532 | | 2.086495 | -0.61725 | 0.650379 | | 1.902455 | -0.52444 | 0.754884 |
| TMEM249 | 2.386752 | 0.131723 | 0.999532 | | 2.160666 | -0.63893 | 0.650379 | | 2.042129 | -0.75226 | 0.622543 |
| RFPL1 | 1.884792 | -0.37103 | 0.999532 | | 1.889562 | -0.65646 | 0.650379 | | 1.481773 | -0.26858 | 0.903339 |
| NECAP1 | 2.396493 | 0.096998 | 0.999532 | | 2.258858 | -0.60901 | 0.650379 | | 1.967108 | -0.68507 | 0.644433 |
| TCAF2 | 3.517472 | 0.583816 | 0.999532 | | 2.655226 | -0.59125 | 0.650379 | | 2.936893 | -1.16149 | 0.282041 |
| NDUFB5 | 6.679098 | 0.277465 | 0.999532 | | 7.942718 | 0.342505 | 0.650812 | | 7.25406 | 0.080357 | 0.945129 |
| PRLH | 3.392804 | 0.415753 | 0.999532 | | 2.738253 | -0.56327 | 0.650953 | | 2.950207 | -0.96305 | 0.359441 |
| C15orf62 | 3.252498 | -0.15087 | 0.999532 | | 3.454272 | -0.50522 | 0.651791 | | 2.745621 | -0.33045 | 0.824012 |
| TENT5B | 2.909923 | 0.014366 | 0.999532 | | 2.760824 | -0.58699 | 0.651791 | | 2.49444 | -0.58459 | 0.686214 |
| FXYD2 | 26.94206 | -0.10407 | 0.999532 | | 37.27367 | 0.446175 | 0.651791 | | 30.45866 | 0.566657 | 0.740755 |
| IL2RG | 2.908964 | 0.561562 | 0.999532 | | 2.087002 | -0.63846 | 0.651791 | | 2.439205 | -1.18114 | 0.282041 |
| TAF9 | 2.366608 | -0.27484 | 0.999532 | | 2.349523 | -0.57463 | 0.651791 | | 1.901788 | -0.28569 | 0.87892 |
| TSC22D1 | 28.19715 | -0.76208 | 0.050433 | DOWN | 36.04746 | -0.30629 | 0.651791 | | 23.46174 | 0.494853 | 0.342845 |
| TOMM70 | 3.254598 | 0.775277 | 0.999532 | | 3.155176 | 0.499703 | 0.651791 | | 3.446897 | -0.25581 | 0.846488 |
| ZNF717 | 2.200995 | 0.652186 | 0.999532 | | 2.53721 | 0.56213 | 0.651791 | | 2.580863 | -0.0721 | 0.973356 |
| UFL1 | 3.280192 | 0.288587 | 0.999532 | | 3.734631 | 0.463719 | 0.651791 | | 3.62039 | 0.190671 | 0.894617 |
| CARS1 | 3.534382 | -0.43431 | 0.999532 | | 3.946016 | -0.45697 | 0.651791 | | 2.989764 | -0.00327 | 0.999385 |
| ATOH1 | 2.157996 | -0.45323 | 0.999532 | | 2.315282 | -0.60684 | 0.651791 | | 1.628457 | -0.13772 | 0.955834 |
| NKAPL | 2.479175 | 0.805431 | 0.999532 | | 1.954409 | -0.67441 | 0.651791 | | 2.107141 | -1.44569 | 0.177152 |
| OR7E24 | 4.797459 | 0.452937 | 0.999532 | | 3.885836 | -0.48793 | 0.651791 | | 4.273387 | -0.92435 | 0.332351 |
| TPD52L3 | 2.417527 | 0.081285 | 0.999532 | | 2.181838 | -0.61898 | 0.651791 | | 1.900642 | -0.68284 | 0.652925 |
| CALM1 | 13.27063 | -0.1369 | 0.999532 | | 17.43956 | 0.334815 | 0.651791 | | 14.55383 | 0.474919 | 0.506334 |
| GPR107 | 5.169238 | 0.311162 | 0.999532 | | 5.898237 | 0.377087 | 0.651791 | | 5.624803 | 0.083022 | 0.950529 |
| CCDC157 | 2.217892 | -0.39327 | 0.999532 | | 2.34014 | -0.60964 | 0.651791 | | 1.682894 | -0.19833 | 0.92697 |
| H2AC17 | 2.741651 | -0.42643 | 0.999532 | | 2.880953 | -0.54635 | 0.651791 | | 2.107086 | -0.09633 | 0.965649 |
| EPO | 2.269488 | -0.2587 | 0.999532 | | 2.198696 | -0.63946 | 0.651791 | | 1.786004 | -0.36591 | 0.857377 |
| PIGF | 3.425894 | 0.058755 | 0.999532 | | 3.258354 | -0.56375 | 0.651791 | | 2.700819 | -0.61806 | 0.635045 |
| SS18L1 | 4.866764 | 0.173863 | 0.999532 | | 4.871289 | -0.42282 | 0.651791 | | 4.143243 | -0.56379 | 0.557909 |
| HNRNPH | 2.529984 | -0.26045 | 0.999532 | | 3.782349 | 0.483717 | 0.651791 | | 3.02148 | 0.764461 | 0.510412 |
| SLK | 9.754548 | -0.15106 | 0.999532 | | 12.75165 | 0.268353 | 0.651791 | | 10.44383 | 0.439435 | 0.456849 |
| NTAQ1 | 2.087044 | 0.670768 | 0.999532 | | 2.497248 | 0.612032 | 0.651791 | | 2.611084 | -0.03715 | 0.990309 |
| AKAP8L | 5.620879 | 0.240942 | 0.999532 | | 6.449051 | 0.37735 | 0.651791 | | 6.154199 | 0.14505 | 0.906302 |
| ZYG11B | 2.984469 | 0.211087 | 0.999532 | | 3.774105 | 0.478884 | 0.651791 | | 3.32879 | 0.289152 | 0.82977 |
| ZNF135 | 2.283858 | 0.410062 | 0.999532 | | 2.547987 | 0.560314 | 0.651791 | | 2.498199 | 0.163394 | 0.928522 |
| ATP1A1 | 13.68073 | 0.202761 | 0.999532 | | 16.89491 | 0.424915 | 0.651791 | | 14.82058 | 0.246933 | 0.849128 |
| PEAK1 | 5.949678 | -0.28926 | 0.999532 | | 8.601787 | 0.383654 | 0.652254 | | 6.74419 | 0.691458 | 0.359638 |
| ABT1 | 3.75257 | -0.02459 | 0.999532 | | 3.854081 | -0.50309 | 0.652576 | | 3.19081 | -0.43452 | 0.757035 |
| NRP1 | 14.9394 | -0.06138 | 0.999532 | | 19.53497 | 0.387038 | 0.652576 | | 16.5647 | 0.48188 | 0.468942 |
| HEXB | 2.968484 | -0.06069 | 0.999532 | | 3.925498 | 0.452722 | 0.652576 | | 3.237096 | 0.529896 | 0.647218 |
| PLA2G4F | 2.120719 | 0.6029 | 0.999532 | | 2.539441 | 0.572337 | 0.652603 | | 2.497506 | -0.01375 | 0.996891 |
| EVA1B | 12.78178 | -0.02868 | 0.999532 | | 16.13914 | 0.300882 | 0.652603 | | 13.54899 | 0.348566 | 0.609581 |
| PI16 | 2.885098 | -0.02154 | 0.999532 | | 2.727283 | -0.55957 | 0.652603 | | 2.438715 | -0.51976 | 0.717627 |
| ZNF267 | 8.455487 | 0.021379 | 0.999532 | | 8.288932 | -0.33927 | 0.652603 | | 7.104531 | -0.3412 | 0.694677 |
| CDC25B | 5.614548 | -0.71058 | 0.999532 | | 6.447453 | -0.42087 | 0.652603 | | 4.257189 | 0.335104 | 0.764366 |
| RUVBL2 | 6.133662 | 0.204096 | 0.999532 | | 6.965989 | 0.368765 | 0.6527 | | 6.439049 | 0.179729 | 0.87591 |
| COL8A1 | 2.589639 | 0.30467 | 0.999532 | | 3.268593 | 0.511776 | 0.652765 | | 2.966922 | 0.227965 | 0.881837 |
| CEP170 | 3.135946 | 0.040065 | 0.999532 | | 3.833304 | 0.459519 | 0.652765 | | 3.450576 | 0.433597 | 0.714657 |
| BSN | 2.590903 | -0.07656 | 0.999532 | | 2.619524 | -0.58191 | 0.653072 | | 2.180457 | -0.48586 | 0.752299 |
| SQLE | 3.226674 | 0.038141 | 0.999532 | | 3.053457 | -0.5242 | 0.653072 | | 2.570089 | -0.54305 | 0.672452 |
| SMIM17 | 6.208527 | 0.201907 | 0.999532 | | 7.444465 | 0.343358 | 0.653072 | | 6.435251 | 0.147781 | 0.899732 |
| NPAS4 | 2.581998 | -0.26679 | 0.999532 | | 2.751617 | -0.56617 | 0.653073 | | 2.025265 | -0.27878 | 0.87764 |
| NACC2 | 6.214996 | 0.20818 | 0.999532 | | 6.99018 | 0.358814 | 0.653073 | | 6.67459 | 0.172653 | 0.87591 |
| SPANXA2 | 3.224695 | 0.417486 | 0.999532 | | 2.590085 | -0.5726 | 0.653073 | | 2.799987 | -0.97677 | 0.396743 |
| KANSL1L | 5.07494 | 0.150255 | 0.999532 | | 5.950758 | 0.381869 | 0.653073 | | 5.276984 | 0.23637 | 0.842754 |
| CHD5 | 2.720555 | 0.227364 | 0.999532 | | 2.38877 | -0.59902 | 0.653073 | | 2.351444 | -0.81077 | 0.548531 |
| WRN | 1.778895 | 1.044306 | 0.999532 | | 1.830994 | 0.674358 | 0.653576 | | 1.958539 | -0.35041 | 0.835289 |
| ZNF358 | 8.759896 | 0.112764 | 0.999532 | | 10.27884 | 0.349649 | 0.654518 | | 9.369476 | 0.262649 | 0.766067 |
| SIRPG | 2.877554 | 0.003785 | 0.999532 | | 2.745943 | -0.57028 | 0.654616 | | 2.352006 | -0.56098 | 0.679818 |
| ZNF548 | 2.936199 | 0.266155 | 0.999532 | | 2.522241 | -0.57831 | 0.654616 | | 2.452821 | -0.82903 | 0.505689 |
| AGXT | 2.558222 | 0.111223 | 0.999532 | | 2.308845 | -0.61093 | 0.654616 | | 2.105079 | -0.70406 | 0.638728 |
| COX6B2 | 2.142625 | -0.23761 | 0.999532 | | 2.251234 | -0.64688 | 0.654616 | | 1.808185 | -0.38485 | 0.849223 |
| SLC15A3 | 3.956533 | -0.34429 | 0.999532 | | 5.445192 | 0.392651 | 0.654616 | | 4.495861 | 0.748778 | 0.413122 |
| DDX25 | 3.683642 | -0.29092 | 0.999532 | | 3.767811 | -0.49485 | 0.654616 | | 2.973705 | -0.19133 | 0.910105 |
| RNASE2 | 3.0855 | -0.02299 | 0.999532 | | 2.855737 | -0.60087 | 0.654616 | | 2.525819 | -0.56328 | 0.706738 |
| EXOC4 | 5.435258 | -0.04309 | 0.999532 | | 6.933325 | 0.36328 | 0.654616 | | 5.641577 | 0.419024 | 0.660958 |
| NOL11 | 3.516531 | 0.985863 | 0.999532 | | 3.141736 | 0.519527 | 0.654736 | | 3.842183 | -0.4493 | 0.702463 |
| DYNC1I2 | 7.963277 | -0.16027 | 0.999532 | | 10.20551 | 0.306156 | 0.654736 | | 8.229131 | 0.485104 | 0.484303 |
| PHF3 | 5.179304 | 0.056976 | 0.999532 | | 6.706017 | 0.354994 | 0.654828 | | 5.765964 | 0.320529 | 0.736481 |
| OTOS | 2.540327 | -0.22876 | 0.999532 | | 2.53191 | -0.57991 | 0.655043 | | 2.035351 | -0.33453 | 0.858767 |
| GTF2H2C | 4.64019 | 0.464937 | 0.999532 | | 5.118916 | 0.429359 | 0.655088 | | 5.128662 | -0.02223 | 0.992286 |
| FBXW12 | 2.570533 | 0.209605 | 0.999532 | | 2.189592 | -0.63531 | 0.655184 | | 2.162865 | -0.83464 | 0.563455 |
| EPHA10 | 2.060608 | 0.05845 | 0.999532 | | 1.831847 | -0.67782 | 0.655184 | | 1.654281 | -0.71792 | 0.665886 |
| HADHB | 3.31962 | 0.094627 | 0.999532 | | 4.552104 | 0.425787 | 0.655184 | | 3.90194 | 0.356504 | 0.756927 |
| AP3S1 | 12.43918 | 0.134551 | 0.999532 | | 14.59507 | 0.282917 | 0.655184 | | 13.20807 | 0.175963 | 0.823677 |
| HDAC5 | 4.748236 | -0.25987 | 0.999532 | | 6.74098 | 0.379296 | 0.655184 | | 5.189094 | 0.658839 | 0.44598 |
| MTMR10 | 5.292148 | 0.148058 | 0.999532 | | 6.441586 | 0.385484 | 0.655184 | | 5.703541 | 0.259495 | 0.816589 |
| CHRNB2 | 2.668528 | 0.168003 | 0.999532 | | 2.537856 | -0.58694 | 0.655404 | | 2.12973 | -0.72663 | 0.5991 |
| SPIRE1 | 2.920224 | 0.333012 | 0.999532 | | 3.633156 | 0.489103 | 0.655404 | | 3.453633 | 0.174601 | 0.908595 |
| ATAD2B | 2.586322 | -0.17563 | 0.999532 | | 2.357266 | -0.58147 | 0.655404 | | 2.054157 | -0.38198 | 0.818918 |
| TMEM143 | 2.911828 | 0.247196 | 0.999532 | | 2.506105 | -0.56556 | 0.655404 | | 2.454938 | -0.79715 | 0.515762 |
| DNASE1L | 3.047335 | 0.28386 | 0.999532 | | 2.581455 | -0.57492 | 0.655404 | | 2.466213 | -0.83964 | 0.49299 |
| CCDC106 | 2.994062 | -0.22558 | 0.999532 | | 3.108311 | -0.49828 | 0.655638 | | 2.361968 | -0.25327 | 0.877268 |
| C11orf96 | 4.870462 | -1.12171 | 0.396029 | | 6.358651 | -0.49476 | 0.655638 | | 3.489762 | 0.698988 | 0.541136 |
| SMC3 | 3.311544 | -0.20557 | 0.999532 | | 4.546883 | 0.433968 | 0.655638 | | 3.6636 | 0.654517 | 0.536289 |
| MAF | 13.23741 | 0.086916 | 0.999532 | | 15.84193 | 0.339876 | 0.655638 | | 13.98446 | 0.283962 | 0.705046 |
| NR2F6 | 4.880242 | 0.580704 | 0.999532 | | 5.395804 | 0.40999 | 0.655955 | | 5.480294 | -0.16058 | 0.88885 |
| CBLC | 4.527816 | 0.015919 | 0.999532 | | 4.480587 | -0.50134 | 0.656087 | | 3.974023 | -0.4971 | 0.707745 |
| C11orf21 | 4.779902 | -0.06922 | 0.999532 | | 4.713185 | -0.43021 | 0.656087 | | 4.183936 | -0.34816 | 0.784185 |
| DLL4 | 3.5721 | 0.094127 | 0.999532 | | 4.44637 | 0.469349 | 0.656087 | | 3.931715 | 0.390104 | 0.746876 |
| EMC3 | 6.213138 | 0.051517 | 0.999532 | | 7.426126 | 0.366262 | 0.656087 | | 6.726269 | 0.331393 | 0.718643 |
| PCGF2 | 8.351314 | -0.18562 | 0.999532 | | 11.23974 | 0.300907 | 0.656087 | | 8.954581 | 0.507818 | 0.445661 |
| LCP1 | 4.075822 | -0.14966 | 0.999532 | | 3.857576 | -0.5335 | 0.656087 | | 3.316314 | -0.36324 | 0.791466 |
| TRPM1 | 2.515723 | -0.11325 | 0.999532 | | 2.46349 | -0.59092 | 0.656087 | | 1.938229 | -0.46141 | 0.78258 |
| SLAIN2 | 4.834071 | -0.22713 | 0.999532 | | 6.8746 | 0.403942 | 0.656087 | | 5.403454 | 0.660452 | 0.465298 |
| PATE1 | 1.783614 | 0.493729 | 0.999532 | | 2.40309 | 0.633885 | 0.656087 | | 2.241518 | 0.169425 | 0.934701 |
| ARRB1 | 11.81364 | -0.02089 | 0.999532 | | 15.35078 | 0.347034 | 0.656087 | | 12.89061 | 0.389493 | 0.617899 |
| C1QB | 8.599303 | -0.67779 | 0.999532 | | 10.26297 | -0.46283 | 0.656087 | | 6.731874 | 0.236489 | 0.864245 |
| PHIP | 7.177571 | 0.564168 | 0.999532 | | 7.229865 | 0.353517 | 0.656087 | | 7.739958 | -0.19014 | 0.843564 |
| ASPRV1 | 3.823095 | 0.018521 | 0.999532 | | 3.645554 | -0.49928 | 0.656087 | | 3.324241 | -0.51222 | 0.712556 |
| ARHGEF | 3.158862 | 0.101848 | 0.999532 | | 3.917846 | 0.454326 | 0.656087 | | 3.346492 | 0.369064 | 0.757037 |
| BZW2 | 2.673883 | -0.61614 | 0.999532 | | 3.141899 | -0.52277 | 0.656381 | | 2.159851 | 0.113421 | 0.95999 |
| TNK1 | 3.193722 | -0.19955 | 0.999532 | | 3.243273 | -0.5092 | 0.657055 | | 2.599282 | -0.29712 | 0.858767 |
| ACADSB | 4.85741 | 0.216196 | 0.999532 | | 5.658915 | 0.383493 | 0.657055 | | 5.328969 | 0.181369 | 0.880949 |
| CTNND2 | 2.943303 | -0.22456 | 0.999532 | | 2.921643 | -0.54779 | 0.657454 | | 2.352467 | -0.31263 | 0.861119 |
| LGALS9C | 4.746566 | 0.276735 | 0.999532 | | 5.437893 | 0.415057 | 0.657454 | | 5.205407 | 0.154396 | 0.904576 |
| RPS19 | 7.725616 | -0.12568 | 0.999532 | | 10.10412 | 0.308584 | 0.657454 | | 8.243292 | 0.451939 | 0.540342 |
| ATXN2 | 5.806231 | -0.27787 | 0.999532 | | 8.040711 | 0.321233 | 0.657454 | | 6.30669 | 0.616669 | 0.387954 |
| RPS27L | 9.202588 | -0.27898 | 0.999532 | | 12.66576 | 0.277877 | 0.657454 | | 9.670323 | 0.573055 | 0.386771 |
| SLC66A1 | 9.476848 | 0.180773 | 0.999532 | | 8.946884 | -0.34718 | 0.657454 | | 8.253302 | -0.50816 | 0.515762 |
| KRTAP29 | 2.192969 | -0.13551 | 0.999532 | | 2.078513 | -0.62682 | 0.657892 | | 1.664256 | -0.47392 | 0.791576 |
| CLCN3 | 4.637021 | 0.460678 | 0.999532 | | 5.28135 | 0.417889 | 0.65814 | | 5.275901 | -0.01859 | 0.994192 |
| RARRES2 | 11.37598 | 0.321763 | 0.999532 | | 12.90338 | 0.352857 | 0.658258 | | 11.87878 | 0.061118 | 0.965649 |
| ETFA | 5.169224 | 0.106436 | 0.999532 | | 5.912889 | 0.376801 | 0.658258 | | 5.300995 | 0.269331 | 0.809832 |
| ING5 | 3.909773 | 0.080101 | 0.999532 | | 4.899742 | 0.409846 | 0.658258 | | 4.218784 | 0.34663 | 0.752562 |
| ZBTB48 | 3.347966 | -0.44229 | 0.999532 | | 3.509954 | -0.51 | 0.658258 | | 2.602915 | -0.05166 | 0.984587 |
| CFAP99 | 2.323577 | 0.066621 | 0.999532 | | 2.062414 | -0.61659 | 0.658258 | | 1.966991 | -0.66915 | 0.666566 |
| SFXN2 | 2.509442 | 0.814172 | 0.999532 | | 2.590695 | 0.554009 | 0.658258 | | 2.72345 | -0.23225 | 0.881837 |
| SNAPC3 | 4.478534 | -0.22999 | 0.999532 | | 6.164871 | 0.369236 | 0.658688 | | 4.910328 | 0.615919 | 0.484303 |
| SRGAP1 | 4.039451 | 0.098231 | 0.999532 | | 5.182093 | 0.420384 | 0.658688 | | 4.475839 | 0.341632 | 0.788433 |
| GDPD3 | 2.573657 | -0.54322 | 0.999532 | | 2.778761 | -0.52828 | 0.658688 | | 2.109317 | 0.031995 | 0.992286 |
| BAK1 | 7.992875 | -0.10366 | 0.999532 | | 8.260801 | -0.36075 | 0.658688 | | 6.745525 | -0.23496 | 0.809832 |
| LAMTOR | 3.216634 | 0.161035 | 0.999532 | | 4.374621 | 0.506183 | 0.658688 | | 3.724062 | 0.320647 | 0.79686 |
| DUOXA1 | 3.555769 | 0.169462 | 0.999532 | | 3.362908 | -0.49788 | 0.659079 | | 2.907272 | -0.65288 | 0.558035 |
| SLURP2 | 2.526528 | -0.04761 | 0.999532 | | 2.58102 | -0.6079 | 0.659079 | | 2.108582 | -0.54052 | 0.728015 |
| DCAF8 | 8.461462 | 0.036684 | 0.999532 | | 10.42021 | 0.300545 | 0.659079 | | 8.79642 | 0.283767 | 0.714657 |
| LBH | 4.617562 | 0.159988 | 0.999532 | | 5.919189 | 0.503738 | 0.659079 | | 5.324688 | 0.360006 | 0.75311 |
| HSD11B2 | 4.621443 | 0.695748 | 0.999532 | | 4.899477 | 0.593066 | 0.659079 | | 4.95486 | -0.04122 | 0.988853 |
| BMP2K | 2.637472 | 0.763042 | 0.999532 | | 2.743292 | 0.554774 | 0.659079 | | 2.856634 | -0.18662 | 0.908595 |
| NSFL1C | 5.277261 | -0.10558 | 0.999532 | | 5.321615 | -0.39803 | 0.659079 | | 4.501016 | -0.27362 | 0.815713 |
| SPACA5B | 2.577629 | 0.006746 | 0.999532 | | 2.354397 | -0.6255 | 0.659079 | | 2.067963 | -0.61458 | 0.686214 |
| DELE1 | 10.36759 | -0.15674 | 0.999532 | | 11.21352 | -0.28051 | 0.659079 | | 8.968277 | -0.10511 | 0.917592 |
| PPP2R3C | 3.148193 | -0.13733 | 0.999532 | | 4.512089 | 0.439677 | 0.659079 | | 3.770958 | 0.604595 | 0.570037 |
| CNRIP1 | 2.682148 | -0.26972 | 0.999532 | | 2.62012 | -0.5365 | 0.659232 | | 2.205108 | -0.25343 | 0.886495 |
| TRABD2B | 2.949011 | -0.57152 | 0.999532 | | 3.250762 | -0.49638 | 0.659232 | | 2.360508 | 0.085506 | 0.972221 |
| CEACAM | 2.424839 | 0.009269 | 0.999532 | | 2.17194 | -0.59426 | 0.659232 | | 1.963185 | -0.59141 | 0.702463 |
| ARHGEF | 3.781652 | 0.172575 | 0.999532 | | 4.619754 | 0.436244 | 0.659232 | | 4.157907 | 0.283343 | 0.816112 |
| NIPBL | 3.551372 | 0.420344 | 0.999532 | | 4.129269 | 0.473223 | 0.659232 | | 3.767917 | 0.081046 | 0.963359 |
| DBNL | 8.926933 | -0.08329 | 0.999532 | | 11.54926 | 0.299369 | 0.659878 | | 9.409403 | 0.401561 | 0.563455 |
| ZNF143 | 3.231364 | -0.26618 | 0.999532 | | 3.563533 | -0.48075 | 0.659878 | | 2.622449 | -0.19001 | 0.907968 |
| OR5H1 | 2.443726 | 0.135278 | 0.999532 | | 2.147256 | -0.62462 | 0.66058 | | 2.018335 | -0.74226 | 0.629744 |
| METTL17 | 5.600572 | -0.17381 | 0.999532 | | 5.930148 | -0.39667 | 0.660756 | | 4.527363 | -0.20504 | 0.866167 |
| NDUFC1 | 9.542915 | 0.101284 | 0.999532 | | 11.68212 | 0.301581 | 0.660756 | | 9.927278 | 0.224185 | 0.791466 |
| TSSK6 | 1.908634 | 0.231436 | 0.999532 | | 2.485739 | 0.61626 | 0.661715 | | 2.217981 | 0.402439 | 0.815293 |
| CABLES1 | 4.453224 | -0.03011 | 0.999532 | | 5.911143 | 0.39667 | 0.661855 | | 4.907212 | 0.443531 | 0.664209 |
| C1orf189 | 2.234391 | 0.078405 | 0.999532 | | 2.085521 | -0.62099 | 0.661855 | | 1.860643 | -0.6787 | 0.662576 |
| H1-5 | 3.033187 | -0.34302 | 0.999532 | | 4.94334 | 0.723044 | 0.661855 | | 4.006551 | 1.019211 | 0.515852 |
| DYNC1LI | 2.873455 | 0.226903 | 0.999532 | | 2.499437 | -0.54974 | 0.661855 | | 2.385376 | -0.75864 | 0.536289 |
| FAM83D | 2.238538 | 0.628845 | 0.999532 | | 1.655844 | -0.69638 | 0.661855 | | 1.918592 | -1.30828 | 0.283338 |
| LY6G5B | 3.815645 | -0.07564 | 0.999532 | | 3.869317 | -0.45635 | 0.662098 | | 3.153968 | -0.36117 | 0.783395 |
| TPGS2 | 4.377466 | 0.090001 | 0.999532 | | 5.649207 | 0.3771 | 0.662098 | | 4.74062 | 0.306323 | 0.771685 |
| RGL1 | 11.9973 | 0.089579 | 0.999532 | | 14.48798 | 0.311027 | 0.662098 | | 12.91994 | 0.240647 | 0.771685 |
| BLNK | 2.067208 | 0.478904 | 0.999532 | | 2.47938 | 0.565063 | 0.662098 | | 2.33433 | 0.104371 | 0.960427 |
| CLDN34 | 2.571789 | 0.431347 | 0.999532 | | 2.072351 | -0.63774 | 0.66212 | | 2.201988 | -1.05547 | 0.398915 |
| PML | 5.194438 | -0.23848 | 0.999532 | | 5.456637 | -0.40052 | 0.66229 | | 4.237383 | -0.1427 | 0.914692 |
| PBX1 | 11.00708 | -0.20922 | 0.999532 | | 14.39879 | 0.273887 | 0.66229 | | 11.61768 | 0.496649 | 0.421517 |
| JCAD | 4.054996 | -0.26742 | 0.999532 | | 5.522656 | 0.387235 | 0.66229 | | 4.376886 | 0.668497 | 0.465298 |
| COL23A1 | 2.915772 | -0.44656 | 0.999532 | | 3.224707 | -0.4962 | 0.66229 | | 2.331626 | -0.03029 | 0.992286 |
| FHDC1 | 2.506695 | 0.103249 | 0.999532 | | 2.257295 | -0.60526 | 0.66229 | | 2.077992 | -0.69117 | 0.650381 |
| BIN3 | 2.601412 | -0.43518 | 0.999532 | | 2.902181 | -0.546 | 0.66229 | | 2.046637 | -0.09257 | 0.970498 |
| RIMBP2 | 2.288736 | -0.24502 | 0.999532 | | 2.415419 | -0.60738 | 0.66229 | | 1.836499 | -0.33786 | 0.862823 |
| FNDC11 | 2.049296 | -0.4069 | 0.999532 | | 2.18604 | -0.60788 | 0.66256 | | 1.620259 | -0.18154 | 0.938429 |
| TYW1 | 7.632132 | 0.138516 | 0.999532 | | 7.252966 | -0.35327 | 0.66267 | | 6.38564 | -0.47089 | 0.551201 |
| PPP4R2 | 16.97266 | -0.03051 | 0.999532 | | 17.69276 | -0.26678 | 0.66267 | | 15.30848 | -0.21554 | 0.803301 |
| CERKL | 2.814404 | 0.380576 | 0.999532 | | 2.113698 | -0.60676 | 0.66267 | | 2.350256 | -0.98156 | 0.456688 |
| GFM2 | 7.4104 | 0.269798 | 0.999532 | | 8.810687 | 0.34126 | 0.66267 | | 8.354189 | 0.092046 | 0.939913 |
| TMPRSS1 | 2.270425 | 0.940857 | 0.999532 | | 2.106162 | 0.600859 | 0.66267 | | 2.545337 | -0.32694 | 0.843043 |
| GPC1 | 4.12418 | -0.32744 | 0.999532 | | 4.422025 | -0.43632 | 0.66267 | | 3.377056 | -0.09358 | 0.960773 |
| MAP3K11 | 3.99968 | -0.23112 | 0.999532 | | 5.423484 | 0.406376 | 0.66267 | | 4.370863 | 0.64058 | 0.520499 |
| FXR2 | 7.629523 | 0.120304 | 0.999532 | | 9.506759 | 0.324627 | 0.66267 | | 8.125374 | 0.221081 | 0.81294 |
| CAMKK2 | 4.173055 | -0.42319 | 0.999532 | | 4.547591 | -0.43013 | 0.66267 | | 3.202311 | 0.010558 | 0.996891 |
| PPP4R3B | 6.520332 | -0.14094 | 0.999532 | | 8.655986 | 0.328028 | 0.66306 | | 6.988208 | 0.489873 | 0.536289 |
| LIME1 | 4.836473 | 0.293642 | 0.999532 | | 5.154708 | 0.403126 | 0.66306 | | 5.090015 | 0.121481 | 0.93452 |
| MIPOL1 | 2.526829 | -0.27277 | 0.999532 | | 2.490803 | -0.58164 | 0.663567 | | 1.995166 | -0.27633 | 0.889853 |
| VWA3A | 2.870749 | 0.369655 | 0.999532 | | 2.37502 | -0.60333 | 0.663567 | | 2.276453 | -0.94985 | 0.442693 |
| ISL2 | 2.357858 | -0.27956 | 0.999532 | | 2.511074 | -0.58261 | 0.663685 | | 1.988246 | -0.28431 | 0.881772 |
| UBXN10 | 2.762447 | -0.34839 | 0.999532 | | 3.051582 | -0.5814 | 0.663755 | | 2.199115 | -0.21832 | 0.906811 |
| SUCO | 4.065809 | 0.291382 | 0.999532 | | 4.969933 | 0.434687 | 0.664022 | | 4.402757 | 0.15325 | 0.907163 |
| PRR11 | 2.029055 | 0.924289 | 0.999532 | | 1.894617 | 0.619484 | 0.664129 | | 2.200502 | -0.28935 | 0.864923 |
| TLCD4 | 2.950645 | 0.275693 | 0.999532 | | 2.513762 | -0.56247 | 0.664129 | | 2.401635 | -0.82633 | 0.515762 |
| RAB8A | 4.693112 | -0.18134 | 0.999532 | | 6.448852 | 0.354746 | 0.664129 | | 5.176337 | 0.555551 | 0.524206 |
| DNAJC8 | 10.19999 | -0.48655 | 0.999532 | | 11.93443 | -0.3126 | 0.664129 | | 8.639721 | 0.200043 | 0.837256 |
| FAM241B | 2.691246 | -0.11654 | 0.999532 | | 2.639035 | -0.5452 | 0.664129 | | 2.102422 | -0.40907 | 0.792538 |
| ITGA1 | 8.344839 | 0.001496 | 0.999532 | | 10.40206 | 0.353515 | 0.664129 | | 8.989204 | 0.369212 | 0.680097 |
| SLA2 | 2.17343 | 0.025625 | 0.999532 | | 1.937002 | -0.64382 | 0.664208 | | 1.769382 | -0.66422 | 0.699971 |
| PSMB2 | 9.089976 | 0.044209 | 0.999532 | | 10.78323 | 0.307458 | 0.664208 | | 9.662146 | 0.298189 | 0.713076 |
| GOLGA8 | 2.803772 | 0.034292 | 0.999532 | | 2.64919 | -0.56199 | 0.664264 | | 2.293778 | -0.5785 | 0.693549 |
| TSEN15 | 3.243701 | 0.377567 | 0.999532 | | 3.96889 | 0.441432 | 0.66427 | | 3.753626 | 0.082014 | 0.96129 |
| SRSF5 | 26.61748 | 0.164042 | 0.999532 | | 30.37788 | 0.240329 | 0.66427 | | 26.9072 | 0.090919 | 0.918694 |
| PDXDC1 | 12.30449 | -0.15667 | 0.999532 | | 15.94193 | 0.273773 | 0.664736 | | 12.9624 | 0.45503 | 0.411291 |
| NAMPT | 6.400196 | -0.79692 | 0.999532 | | 8.047247 | -0.36703 | 0.664736 | | 5.258021 | 0.454541 | 0.644433 |
| CDH5 | 6.67824 | -0.19222 | 0.999532 | | 9.496337 | 0.350406 | 0.664736 | | 7.569029 | 0.568136 | 0.49299 |
| COL6A1 | 14.81193 | 0.008334 | 0.999532 | | 19.59161 | 0.46738 | 0.664736 | | 16.81342 | 0.477897 | 0.663709 |
| TET1 | 2.50272 | -0.4786 | 0.999532 | | 2.649719 | -0.55691 | 0.664736 | | 2.070748 | -0.05803 | 0.984155 |
| NOTCH4 | 13.97099 | -0.04389 | 0.999532 | | 17.41048 | 0.289814 | 0.664736 | | 14.69067 | 0.350153 | 0.65401 |
| TAPT1 | 3.799464 | 0.015909 | 0.999532 | | 3.598115 | -0.4969 | 0.664736 | | 3.140956 | -0.49279 | 0.70375 |
| CDRT15L | 2.3534 | 0.129303 | 0.999532 | | 2.052132 | -0.60516 | 0.664736 | | 2.029727 | -0.72455 | 0.620745 |
| DPH2 | 3.405692 | -0.11438 | 0.999532 | | 3.313195 | -0.4936 | 0.664736 | | 2.743432 | -0.3618 | 0.800239 |
| LHPP | 2.868954 | 0.261625 | 0.999532 | | 3.341675 | 0.481713 | 0.664745 | | 3.209643 | 0.241581 | 0.874653 |
| DEGS2 | 4.648804 | -0.31455 | 0.999532 | | 4.924159 | -0.4398 | 0.664942 | | 3.852424 | -0.10863 | 0.950728 |
| DCST2 | 2.25546 | 0.255786 | 0.999532 | | 1.925968 | -0.64252 | 0.664953 | | 1.806099 | -0.87951 | 0.532889 |
| ERAP2 | 3.036853 | -0.54905 | 0.999532 | | 4.527722 | 0.443306 | 0.664972 | | 3.551694 | 1.028658 | 0.300852 |
| BEX5 | 4.548261 | 0.124778 | 0.999532 | | 5.330246 | 0.450424 | 0.664972 | | 4.838893 | 0.344952 | 0.770048 |
| STK40 | 8.164614 | 0.018498 | 0.999532 | | 9.997852 | 0.301157 | 0.664972 | | 8.832483 | 0.301608 | 0.722703 |
| AFF4 | 7.748848 | -0.22637 | 0.999532 | | 10.84791 | 0.315845 | 0.664972 | | 8.390707 | 0.56088 | 0.397994 |
| PSMG2 | 5.187096 | 0.155955 | 0.999532 | | 6.62584 | 0.377854 | 0.664972 | | 5.701651 | 0.253757 | 0.823571 |
| CEP85L | 3.631848 | -0.3798 | 0.999532 | | 3.781753 | -0.44873 | 0.664972 | | 3.010347 | -0.05707 | 0.97843 |
| TSPAN17 | 4.305963 | 0.337462 | 0.999532 | | 4.994519 | 0.40219 | 0.664972 | | 4.747598 | 0.084356 | 0.955834 |
| NCSTN | 5.43288 | 0.060391 | 0.999532 | | 6.69486 | 0.363899 | 0.664981 | | 5.835407 | 0.323019 | 0.735469 |
| AGAP2 | 2.757104 | -0.01824 | 0.999532 | | 2.852995 | -0.55405 | 0.664981 | | 2.406462 | -0.51068 | 0.728372 |
| RNF113A | 2.962613 | 0.303488 | 0.999532 | | 2.438739 | -0.5516 | 0.664981 | | 2.51262 | -0.83721 | 0.49299 |
| CRB2 | 9.792988 | 0.288682 | 0.999532 | | 11.35793 | 0.393555 | 0.664981 | | 10.51629 | 0.131539 | 0.920181 |
| B2M | 415.1763 | -0.13976 | 0.999532 | | 547.7287 | 0.313255 | 0.664981 | | 447.9176 | 0.477554 | 0.428819 |
| RANGAP | 7.723641 | 0.55987 | 0.999532 | | 7.630513 | 0.350511 | 0.665316 | | 7.95995 | -0.20058 | 0.85309 |
| KCNAB3 | 5.331237 | -0.02146 | 0.999532 | | 5.39387 | -0.40878 | 0.665316 | | 4.524704 | -0.36595 | 0.739496 |
| POLN | 2.323926 | -0.52522 | 0.999532 | | 2.509938 | -0.56949 | 0.665316 | | 1.889347 | -0.03261 | 0.992586 |
| FCGRT | 16.33071 | -0.06611 | 0.999532 | | 20.8388 | 0.28975 | 0.665316 | | 17.1708 | 0.379945 | 0.643601 |
| NFKBIA | 12.51154 | -0.85216 | 0.124973 | | 16.31512 | -0.27855 | 0.666285 | | 10.09343 | 0.595813 | 0.282041 |
| FAM210A | 2.113423 | -0.13702 | 0.999532 | | 2.184878 | -0.61309 | 0.666285 | | 1.856591 | -0.45392 | 0.799944 |
| COQ8A | 2.623351 | -0.11503 | 0.999532 | | 2.712416 | -0.55076 | 0.666285 | | 2.083529 | -0.41398 | 0.79686 |
| RAD51AP | 2.029111 | -0.14528 | 0.999532 | | 1.976393 | -0.64405 | 0.666285 | | 1.698453 | -0.47877 | 0.799157 |
| SLC19A2 | 3.24535 | -0.12259 | 0.999532 | | 3.080224 | -0.5632 | 0.666285 | | 2.514611 | -0.40264 | 0.792539 |
| GBA3 | 2.488207 | 0.095571 | 0.999532 | | 3.231224 | 0.545335 | 0.666285 | | 2.970352 | 0.444251 | 0.762271 |
| GPR83 | 2.422509 | 0.417409 | 0.999532 | | 1.843402 | -0.64306 | 0.666285 | | 2.007588 | -1.05161 | 0.443676 |
| OR2T1 | 3.139541 | 0.328932 | 0.999532 | | 2.684991 | -0.54761 | 0.666285 | | 2.686076 | -0.86035 | 0.484413 |
| GLRX5 | 2.184244 | 0.335702 | 0.999532 | | 2.925575 | 0.579756 | 0.666285 | | 2.666782 | 0.267983 | 0.880375 |
| CLDN6 | 3.159575 | 0.235282 | 0.999532 | | 2.837149 | -0.53178 | 0.666285 | | 2.598683 | -0.74799 | 0.531732 |
| JMJD1C | 6.394188 | -0.21908 | 0.999532 | | 8.744816 | 0.315851 | 0.666285 | | 7.004231 | 0.552526 | 0.461117 |
| PELP1 | 3.390215 | -0.21276 | 0.999532 | | 3.349565 | -0.53797 | 0.666285 | | 2.745404 | -0.30384 | 0.85077 |
| RADX | 2.154016 | -0.44867 | 0.999532 | | 2.310009 | -0.59289 | 0.666395 | | 1.718598 | -0.12516 | 0.96129 |
| ORMDL3 | 6.760396 | 0.360115 | 0.999532 | | 7.40022 | 0.334745 | 0.667015 | | 7.217048 | -0.00658 | 0.996891 |
| PTK7 | 3.446431 | -0.08234 | 0.999532 | | 4.591444 | 0.426324 | 0.66761 | | 3.577342 | 0.516898 | 0.646821 |
| HNRNPCI | 12.42003 | 0.260036 | 0.999532 | | 11.58179 | -0.33533 | 0.66761 | | 10.93333 | -0.5843 | 0.310637 |
| STPG2 | 2.549294 | 0.285696 | 0.999532 | | 2.14613 | -0.60199 | 0.66761 | | 2.109645 | -0.87052 | 0.515762 |
| LONRF2 | 3.630189 | 0.188221 | 0.999532 | | 3.419273 | -0.49421 | 0.66761 | | 3.058977 | -0.66277 | 0.552672 |
| ST13 | 11.33128 | 0.077745 | 0.999532 | | 11.11279 | -0.30569 | 0.667817 | | 9.469231 | -0.36327 | 0.622543 |
| TYRO3 | 21.58471 | 0.21205 | 0.999532 | | 25.25541 | 0.297571 | 0.667903 | | 22.84435 | 0.107285 | 0.900428 |
| PVALEF | 3.069178 | -0.37575 | 0.999532 | | 3.286416 | -0.55356 | 0.66828 | | 2.662927 | -0.17014 | 0.939549 |
| RGS5 | 4.291564 | -0.46307 | 0.999532 | | 6.727737 | 0.526166 | 0.668558 | | 5.223109 | 0.982374 | 0.407399 |
| WBP2NL | 3.204426 | 0.297934 | 0.999532 | | 2.665731 | -0.5775 | 0.668558 | | 2.801169 | -0.84424 | 0.526384 |
| PRAMEF | 2.938466 | -0.1141 | 0.999532 | | 2.920279 | -0.52855 | 0.668558 | | 2.419033 | -0.39668 | 0.79686 |
| LIPT1 | 3.211186 | -0.2209 | 0.999532 | | 3.108506 | -0.49769 | 0.668558 | | 2.482808 | -0.26539 | 0.873268 |
| TASOR | 3.809091 | 0.225836 | 0.999532 | | 4.318541 | 0.42422 | 0.668558 | | 4.246933 | 0.231503 | 0.857633 |
| ANKRD62 | 2.56346 | 0.423173 | 0.999532 | | 2.253296 | -0.60222 | 0.668558 | | 2.149684 | -0.99902 | 0.417552 |
| XPNPEP3 | 2.774468 | 0.304362 | 0.999532 | | 2.261223 | -0.58191 | 0.668558 | | 2.326613 | -0.8691 | 0.515762 |
| TESC | 2.802932 | -0.29046 | 0.999532 | | 2.890357 | -0.53443 | 0.668661 | | 2.268441 | -0.2299 | 0.903339 |
| PPIAL4F | 12.85608 | -0.06272 | 0.999532 | | 13.54809 | -0.28524 | 0.6688 | | 11.53272 | -0.21266 | 0.78258 |
| DNAAF9 | 4.499829 | 0.263722 | 0.999532 | | 5.333552 | 0.432331 | 0.66905 | | 4.85008 | 0.195211 | 0.885712 |
| SLC22A4 | 2.758369 | 0.075313 | 0.999532 | | 2.539604 | -0.55716 | 0.669161 | | 2.191229 | -0.61321 | 0.66232 |
| TBC1D9B | 8.624551 | 0.057792 | 0.999532 | | 10.36827 | 0.296141 | 0.66977 | | 9.085868 | 0.251198 | 0.78518 |
| C12orf29 | 3.155113 | 0.236552 | 0.999532 | | 3.74931 | 0.472071 | 0.66977 | | 3.537897 | 0.248171 | 0.869161 |
| KIAA1328 | 2.642178 | 0.087609 | 0.999532 | | 3.599223 | 0.483551 | 0.66977 | | 3.141738 | 0.417546 | 0.752299 |
| SNX25 | 4.182547 | 0.125809 | 0.999532 | | 5.10349 | 0.391953 | 0.66977 | | 4.494935 | 0.282481 | 0.806394 |
| MUS81 | 5.056561 | 0.32366 | 0.999532 | | 4.219798 | -0.46349 | 0.66977 | | 4.172984 | -0.76404 | 0.400624 |
| ADORA1 | 3.794782 | 0.447358 | 0.999532 | | 4.150123 | 0.453372 | 0.66977 | | 4.196265 | 0.029888 | 0.991273 |
| DNMT3A | 3.622038 | 0.554589 | 0.999532 | | 3.759917 | 0.45683 | 0.66977 | | 3.935856 | -0.07799 | 0.963866 |
| RNASE3 | 3.353713 | 0.00371 | 0.999532 | | 3.281128 | -0.49854 | 0.66977 | | 2.799309 | -0.47903 | 0.718643 |
| RPS4X | 31.54694 | 0.192925 | 0.999532 | | 36.77724 | 0.316413 | 0.66977 | | 33.42588 | 0.140907 | 0.88885 |
| EDIL3 | 6.001193 | -0.46909 | 0.999532 | | 9.202504 | 0.38851 | 0.66977 | | 6.572416 | 0.893287 | 0.240311 |
| ERVV-2 | 2.918807 | 0.107144 | 0.999532 | | 2.612525 | -0.58177 | 0.66977 | | 2.44445 | -0.66953 | 0.652925 |
| TMCO6 | 3.058016 | 0.090018 | 0.999532 | | 2.826299 | -0.53571 | 0.66977 | | 2.536822 | -0.60563 | 0.65401 |
| WIPI2 | 3.452335 | 0.055434 | 0.999532 | | 4.392749 | 0.420331 | 0.66977 | | 3.837228 | 0.382406 | 0.738349 |
| EHD2 | 3.587745 | 0.047596 | 0.999532 | | 4.95 | 0.437056 | 0.670036 | | 4.121774 | 0.430945 | 0.728015 |
| ADNP | 6.683531 | -0.19888 | 0.999532 | | 9.00437 | 0.295199 | 0.670115 | | 7.158008 | 0.510497 | 0.515762 |
| ECHDC3 | 1.967728 | 0.291904 | 0.999532 | | 2.558776 | 0.569389 | 0.670134 | | 2.235877 | 0.297014 | 0.865065 |
| HIPK3 | 6.478309 | 0.249038 | 0.999532 | | 7.590601 | 0.333979 | 0.670134 | | 6.988599 | 0.102906 | 0.928522 |
| XG | 2.5136 | -0.74978 | 0.999532 | | 2.909148 | -0.52923 | 0.670449 | | 2.163683 | 0.234615 | 0.907271 |
| FAM172A | 3.772775 | 0.312579 | 0.999532 | | 3.293943 | -0.48443 | 0.670449 | | 3.097805 | -0.77393 | 0.484303 |

| | | | | | | | | | | | | |
|---|---|---|---|---|---|---|---|---|---|---|---|---|
| PODXL2 | 4.142307 | 0.20128 | 0.999532 | | 3.65254 | -0.49324 | 0.67045 | | 3.426825 | -0.67715 | 0.541754 | |
| GAS2L3 | 3.146198 | -0.20937 | 0.999532 | | 3.199837 | -0.52657 | 0.670573 | | 2.548557 | -0.29269 | 0.861119 | |
| OR5T1 | 2.167222 | -0.13365 | 0.999532 | | 2.080053 | -0.62465 | 0.670573 | | 1.728825 | -0.47282 | 0.797147 | |
| REC8 | 4.011856 | -0.04076 | 0.999532 | | 3.992806 | -0.4506 | 0.670573 | | 3.321038 | -0.39128 | 0.751138 | |
| ISY1 | 3.77836 | 0.204741 | 0.999532 | | 4.778261 | 0.406171 | 0.670573 | | 4.373323 | 0.229318 | 0.861119 | |
| CCNQ | 3.518478 | -0.35738 | 0.999532 | | 3.84838 | -0.45948 | 0.670573 | | 2.997612 | -0.08417 | 0.969678 | |
| CARF | 4.88142 | -0.1406 | 0.999532 | | 4.961245 | -0.41727 | 0.670573 | | 4.188505 | -0.2601 | 0.849682 | |
| ARFGAP2 | 2.860357 | 0.138251 | 0.999532 | | 3.644406 | 0.460572 | 0.670573 | | 3.172901 | 0.342668 | 0.793131 | |
| CEP57 | 3.30233 | 0.246853 | 0.999532 | | 4.264719 | 0.47702 | 0.670573 | | 3.937807 | 0.261804 | 0.862823 | |
| KREMEN2 | 2.120671 | -0.24566 | 0.999532 | | 2.320186 | -0.60778 | 0.670573 | | 1.803088 | -0.34377 | 0.861119 | |
| ZNF616 | 3.880545 | -0.18238 | 0.999532 | | 4.237802 | -0.44843 | 0.670573 | | 3.39422 | -0.2391 | 0.874203 | |
| POU2F2 | 1.915325 | 0.633838 | 0.999532 | | 2.210874 | 0.571273 | 0.670723 | | 2.176687 | -0.04582 | 0.986811 | |
| HNMT | 2.754709 | 0.000901 | 0.999532 | | 3.681622 | 0.448833 | 0.670738 | | 3.242401 | 0.467871 | 0.69872 | |
| DTX3L | 2.677359 | 0.071127 | 0.999532 | | 3.448851 | 0.469312 | 0.670738 | | 3.023688 | 0.413662 | 0.757035 | |
| SYNE1 | 6.638312 | 0.334394 | 0.999532 | | 7.776344 | 0.338593 | 0.670805 | | 7.20398 | 0.012154 | 0.996312 | |
| TVP23B | 8.046223 | -0.08769 | 0.999532 | | 8.2389 | -0.32386 | 0.670805 | | 6.900391 | -0.21636 | 0.826705 | |
| DYNC2LI1 | 3.580381 | 0.393869 | 0.999532 | | 4.266771 | 0.434384 | 0.670817 | | 4.166745 | 0.071205 | 0.970811 | |
| ETS1 | 10.3034 | -0.09578 | 0.999532 | | 13.27428 | 0.277622 | 0.671522 | | 11.01268 | 0.392876 | 0.531935 | |
| IKBKE | 2.646435 | 0.107935 | 0.999532 | | 2.289876 | -0.56589 | 0.671974 | | 2.160599 | -0.65573 | 0.647218 | |
| KLRG1 | 2.04619 | -0.16645 | 0.999532 | | 1.923313 | -0.61968 | 0.672421 | | 1.634534 | -0.43766 | 0.815713 | |
| NME4 | 3.030521 | -0.29935 | 0.999532 | | 4.414595 | 0.417868 | 0.672421 | | 3.489879 | 0.736415 | 0.459697 | |
| ZNF182 | 2.431351 | -0.09102 | 0.999532 | | 2.45524 | -0.57265 | 0.672421 | | 2.061156 | -0.46114 | 0.774747 | |
| TMEM164 | 5.28558 | -0.31607 | 0.999532 | | 5.92806 | -0.37571 | 0.672451 | | 4.290281 | -0.03714 | 0.986869 | |
| CCDC134 | 3.080877 | 0.104839 | 0.999532 | | 2.876681 | -0.522 | 0.672451 | | 2.602011 | -0.60735 | 0.652114 | |
| SLC35F5 | 3.778676 | 0.132994 | 0.999532 | | 4.996441 | 0.442493 | 0.672451 | | 4.0767 | 0.325031 | 0.791576 | |
| KANK3 | 16.94622 | 0.023635 | 0.999532 | | 17.02879 | -0.30926 | 0.672832 | | 14.72988 | -0.31512 | 0.720106 | |
| TUBB4B | 9.551194 | -0.10447 | 0.999532 | | 12.11772 | 0.282555 | 0.672832 | | 10.15946 | 0.406087 | 0.542124 | |
| SAMD4B | 6.455192 | 0.238891 | 0.999532 | | 7.218249 | 0.346151 | 0.673167 | | 6.949809 | 0.128222 | 0.913859 | |
| GANAB | 10.42434 | 0.052116 | 0.999532 | | 13.28669 | 0.317576 | 0.673167 | | 11.06171 | 0.292064 | 0.750371 | |
| RIMBP3B | 4.120307 | -0.32294 | 0.999532 | | 4.690239 | -0.42877 | 0.673167 | | 3.582695 | -0.0811 | 0.965649 | |
| SPSB1 | 4.190517 | 0.34026 | 0.999532 | | 3.603053 | -0.48715 | 0.673167 | | 3.586496 | -0.81449 | 0.398915 | |
| GPR137B | 2.92383 | -0.44317 | 0.999532 | | 3.187763 | -0.47725 | 0.673167 | | 2.341926 | -0.02296 | 0.994722 | |
| SRI | 7.126499 | -0.2987 | 0.999532 | | 9.79988 | 0.349768 | 0.673167 | | 7.93459 | 0.66956 | 0.324867 | |
| GFPT1 | 5.086175 | 0.11225 | 0.999532 | | 6.069602 | 0.355778 | 0.673167 | | 5.488983 | 0.255312 | 0.810276 | |
| TFPI2 | 3.446839 | -0.04255 | 0.999532 | | 4.314766 | 0.487291 | 0.673167 | | 3.768489 | 0.535374 | 0.658914 | |
| GZMA | 2.267804 | -0.02952 | 0.999532 | | 2.144662 | -0.61937 | 0.673167 | | 1.936108 | -0.58132 | 0.721294 | |
| PRSS53 | 3.617726 | 0.234601 | 0.999532 | | 3.219109 | -0.49543 | 0.673167 | | 3.126892 | -0.6968 | 0.595927 | |
| MAJIN | 2.40117 | 0.478653 | 0.999532 | | 1.776553 | -0.6519 | 0.673167 | | 1.98036 | -1.11841 | 0.397004 | |
| SVIL | 6.114833 | -0.03696 | 0.999532 | | 7.863869 | 0.340646 | 0.673167 | | 6.771313 | 0.395238 | 0.653409 | |
| TUBG1 | 2.8494 | -0.29352 | 0.999532 | | 2.972611 | -0.48853 | 0.673167 | | 2.356955 | -0.17844 | 0.919493 | |
| ITGB4 | 2.895346 | -0.12245 | 0.999532 | | 2.797455 | -0.52484 | 0.673167 | | 2.324569 | -0.38853 | 0.801502 | |
| COMMD1 | 4.248775 | 0.737123 | 0.999532 | | 4.100531 | 0.423813 | 0.673167 | | 4.655567 | -0.29289 | 0.793454 | |
| CADM4 | 2.53589 | 0.111864 | 0.999532 | | 3.453027 | 0.472114 | 0.673167 | | 2.881022 | 0.386962 | 0.777682 | |
| ATL1 | 2.13112 | -0.12307 | 0.999532 | | 2.114744 | -0.60497 | 0.673167 | | 1.657215 | -0.46349 | 0.795612 | |
| PPDPFL | 2.444147 | 0.162581 | 0.999532 | | 2.207606 | -0.61185 | 0.673167 | | 2.076207 | -0.75749 | 0.629345 | |
| KRTAP10 | 5.248275 | 0.093097 | 0.999532 | | 4.943943 | -0.45481 | 0.673167 | | 4.453789 | -0.53588 | 0.647581 | |
| IFITM1 | 17.39254 | -0.13592 | 0.999532 | | 22.70806 | 0.317888 | 0.673167 | | 18.92646 | 0.478184 | 0.383088 | |
| NKIRAS2 | 4.36507 | -0.03559 | 0.999532 | | 5.362509 | 0.37168 | 0.673167 | | 4.692477 | 0.422053 | 0.674041 | |
| KCNAB1 | 2.491297 | 0.521655 | 0.999532 | | 2.872491 | 0.523496 | 0.673167 | | 2.805408 | 0.023118 | 0.994192 | |
| MTX3 | 2.528928 | 0.228265 | 0.999532 | | 3.283438 | 0.488767 | 0.673167 | | 2.958144 | 0.280709 | 0.851623 | |
| EHD3 | 49.13207 | 0.151814 | 0.999532 | | 57.02826 | 0.26658 | 0.673167 | | 51.27326 | 0.154806 | 0.810208 | |
| SLC25A3 | 5.455142 | 0.23077 | 0.999532 | | 6.430727 | 0.346023 | 0.673167 | | 5.667923 | 0.128557 | 0.913083 | |
| PWWP3A | 6.107516 | 0.383085 | 0.999532 | | 6.868229 | 0.347068 | 0.673167 | | 6.467996 | -0.01633 | 0.993618 | |
| EN2 | 2.447731 | 0.035612 | 0.999532 | | 2.339934 | -0.60167 | 0.673167 | | 2.028913 | -0.6188 | 0.690003 | |
| UBXN11 | 3.896269 | 0.282836 | 0.999532 | | 4.386611 | 0.41521 | 0.673167 | | 4.119628 | 0.147112 | 0.914692 | |
| SEMA6B | 2.316746 | -0.41627 | 0.999532 | | 2.395424 | -0.5537 | 0.673167 | | 1.890103 | -0.12974 | 0.956011 | |
| EMP1 | 3.766839 | 0.056632 | 0.999532 | | 4.571377 | 0.432588 | 0.673167 | | 3.986453 | 0.390997 | 0.736098 | |
| LGALS2 | 3.260909 | 0.202033 | 0.999532 | | 4.20591 | 0.491517 | 0.673167 | | 3.812838 | 0.301223 | 0.827267 | |
| LIMK1 | 3.299827 | 0.317288 | 0.999532 | | 3.657552 | 0.456661 | 0.673167 | | 3.615158 | 0.145775 | 0.927809 | |
| TIAL1 | 12.21844 | 0.156896 | 0.999532 | | 14.6386 | 0.274819 | 0.673167 | | 12.93249 | 0.141687 | 0.879947 | |
| SNRNP70 | 11.94032 | 0.037129 | 0.999532 | | 14.77475 | 0.267556 | 0.67358 | | 12.84689 | 0.249039 | 0.707142 | |
| USP18 | 7.419705 | 0.137708 | 0.999532 | | 7.278776 | -0.34031 | 0.673623 | | 6.756823 | -0.4507 | 0.606305 | |
| SAYSD1 | 3.20737 | -0.01451 | 0.999532 | | 4.486461 | 0.49998 | 0.673623 | | 4.015873 | 0.520337 | 0.713076 | |
| GALNT1 | 2.975539 | 0.04377 | 0.999532 | | 3.867426 | 0.439223 | 0.673623 | | 3.377895 | 0.413718 | 0.722703 | |
| SLC6A17 | 3.944167 | -0.05342 | 0.999532 | | 3.752405 | -0.45639 | 0.673623 | | 3.221942 | -0.39007 | 0.761337 | |
| EARS2 | 2.909784 | 0.863986 | 0.999532 | | 2.809546 | 0.496906 | 0.673623 | | 3.189115 | -0.3501 | 0.779082 | |
| H2BC4 | 8.45786 | 0.07157 | 0.999532 | | 10.85169 | 0.306786 | 0.673623 | | 9.394165 | 0.259479 | 0.761337 | |
| CHID1 | 4.000667 | 0.16743 | 0.999532 | | 4.914644 | 0.411327 | 0.673623 | | 4.398183 | 0.261603 | 0.835554 | |
| LRRC25 | 2.643025 | -0.1624 | 0.999532 | | 3.661404 | 0.476508 | 0.673623 | | 2.984103 | 0.651724 | 0.609671 | |
| HNRNPD | 11.11636 | 0.019732 | 0.999532 | | 13.94027 | 0.298757 | 0.673623 | | 11.69109 | 0.308819 | 0.722703 | |
| PABIR3 | 2.288722 | 0.143339 | 0.999532 | | 2.101284 | -0.60636 | 0.673623 | | 1.848967 | -0.72992 | 0.628895 | |
| AQP7 | 5.686113 | -0.3713 | 0.999532 | | 6.241381 | -0.37897 | 0.673623 | | 4.681949 | 0.008791 | 0.996891 | |
| RAB37 | 2.403938 | -0.11929 | 0.999532 | | 2.338057 | -0.57493 | 0.673623 | | 1.988699 | -0.43912 | 0.795748 | |
| RPS3A | 48.60996 | -0.11794 | 0.999532 | | 60.22014 | 0.18719 | 0.673623 | | 49.50887 | 0.326152 | 0.456688 | |
| CHGA | 2.418795 | 0.228114 | 0.999532 | | 2.31349 | -0.56797 | 0.673623 | | 2.081717 | -0.7635 | 0.570037 | |
| FAM151A | 3.241365 | 0.689716 | 0.999532 | | 3.346309 | 0.53645 | 0.673623 | | 3.571213 | -0.13881 | 0.942503 | |
| FAF2 | 3.620767 | -0.0419 | 0.999532 | | 4.654189 | 0.416982 | 0.673623 | | 4.003623 | 0.473738 | 0.678492 | |
| RPL29 | 27.13084 | -0.12699 | 0.999532 | | 35.29588 | 0.257368 | 0.673623 | | 28.49545 | 0.42138 | 0.508751 | |
| PGAM2 | 2.836718 | -0.28129 | 0.999532 | | 3.035329 | -0.51222 | 0.673623 | | 2.374508 | -0.21061 | 0.908698 | |
| TNR | 3.263061 | -0.06049 | 0.999532 | | 3.274539 | -0.5376 | 0.673623 | | 2.794698 | -0.46882 | 0.740093 | |
| MDP1 | 3.446651 | 0.347551 | 0.999532 | | 2.714431 | -0.50169 | 0.673623 | | 2.891842 | -0.83662 | 0.446959 | |
| DCDC1 | 2.089111 | 1.016449 | 0.999532 | | 1.981532 | 0.643207 | 0.673623 | | 2.424095 | -0.35898 | 0.839373 | |
| RAB2B | 3.280082 | -0.73536 | 0.999532 | | 3.989491 | -0.432 | 0.673623 | | 2.668882 | 0.321439 | 0.827429 | |
| SLC35F2 | 2.149796 | 0.886289 | 0.999532 | | 2.157726 | 0.608066 | 0.673623 | | 2.391741 | -0.2591 | 0.881689 | |
| PDIA5 | 3.546342 | 0.001654 | 0.999532 | | 4.944928 | 0.425166 | 0.673623 | | 4.119766 | 0.450105 | 0.692333 | |
| IL3 | 2.486972 | 0.240209 | 0.999532 | | 1.952682 | -0.63339 | 0.673623 | | 1.966986 | -0.86899 | 0.566812 | |
| ITPA | 3.21178 | -0.32696 | 0.999532 | | 4.456898 | 0.416858 | 0.673782 | | 3.502553 | 0.758972 | 0.445661 | |
| STAB1 | 9.721701 | -0.59314 | 0.999532 | | 11.31472 | -0.44044 | 0.674195 | | 7.878334 | 0.188627 | 0.877682 | |
| AAMDC | 4.901251 | 0.436799 | 0.999532 | | 5.298052 | 0.376792 | 0.674195 | | 5.126549 | -0.04195 | 0.978805 | |
| DENND5A | 3.965951 | 0.06333 | 0.999532 | | 4.804719 | 0.39381 | 0.674203 | | 4.265132 | 0.340429 | 0.761337 | |
| PLCE1 | 12.07649 | 0.742811 | 0.69276 | | 11.77304 | 0.399785 | 0.674203 | | 12.70948 | -0.30582 | 0.752173 | |
| KLRC4 | 3.302054 | -0.6034 | 0.999532 | | 3.900609 | -0.50263 | 0.674203 | | 2.811031 | 0.12108 | 0.95816 | |
| PRMT6 | 4.619077 | 0.14638 | 0.999532 | | 5.861273 | 0.376042 | 0.674203 | | 5.162697 | 0.250103 | 0.82512 | |
| PABPC1L | 4.459863 | -0.19788 | 0.999532 | | 4.81374 | -0.43661 | 0.674203 | | 3.595355 | -0.22139 | 0.872493 | |
| DRAP1 | 5.879255 | 0.381555 | 0.999532 | | 6.527126 | 0.349855 | 0.674203 | | 6.231993 | -0.01468 | 0.995112 | |
| FEZF2 | 3.277686 | 0.209892 | 0.999532 | | 2.960837 | -0.51444 | 0.674203 | | 2.860734 | -0.70729 | 0.563885 | |
| KCTD15 | 2.427076 | 0.095999 | 0.999532 | | 3.026186 | 0.524851 | 0.674203 | | 2.76782 | 0.440239 | 0.752746 | |
| USP8 | 5.414994 | 0.162698 | 0.999532 | | 6.697536 | 0.338224 | 0.674203 | | 5.90793 | 0.197072 | 0.858705 | |
| NPAS2 | 2.93239 | -0.38717 | 0.999532 | | 3.071129 | -0.49807 | 0.674203 | | 2.287783 | -0.09348 | 0.966643 | |
| ZNF501 | 3.031388 | 0.595124 | 0.999532 | | 2.350488 | -0.58127 | 0.674203 | | 2.572763 | -1.14908 | 0.295502 | |
| TMED2 | 6.360909 | 0.222887 | 0.999532 | | 7.379274 | 0.339209 | 0.674203 | | 6.81419 | 0.136966 | 0.900848 | |
| PDE1C | 2.554312 | 0.240546 | 0.999532 | | 2.184218 | -0.60865 | 0.674203 | | 2.084189 | -0.8324 | 0.557909 | |
| VCY | 2.279206 | -0.42018 | 0.999532 | | 2.363685 | -0.599 | 0.674203 | | 1.856162 | -0.16698 | 0.946572 | |
| MTA1 | 13.32843 | 0.089496 | 0.999532 | | 15.39332 | 0.238534 | 0.674203 | | 13.75766 | 0.163881 | 0.844873 | |
| HINFP | 4.864478 | -0.13291 | 0.999532 | | 5.057165 | -0.41708 | 0.674203 | | 4.128602 | -0.26488 | 0.836935 | |
| UFM1 | 8.975682 | -0.05586 | 0.999532 | | 10.80742 | 0.280418 | 0.674203 | | 9.420979 | 0.35714 | 0.647218 | |
| THRB | 2.803028 | 0.277663 | 0.999532 | | 3.219888 | 0.475312 | 0.674203 | | 3.088451 | 0.217898 | 0.888003 | |
| URB1 | 2.867503 | -0.54073 | 0.999532 | | 3.230989 | -0.52414 | 0.674203 | | 2.16406 | 0.04243 | 0.989148 | |
| HS3ST2 | 2.058318 | -0.14486 | 0.999532 | | 2.122082 | -0.64015 | 0.674203 | | 1.595003 | -0.47243 | 0.801572 | |
| ZNF497 | 2.518669 | -0.17531 | 0.999532 | | 2.583003 | -0.56653 | 0.674203 | | 2.025286 | -0.37003 | 0.837256 | |
| STK11IP | 2.509907 | -0.70736 | 0.999532 | | 2.942968 | -0.50139 | 0.674203 | | 2.007528 | 0.223061 | 0.906811 | |
| NRG2 | 2.559427 | 0.078835 | 0.999532 | | 2.439892 | -0.57008 | 0.674462 | | 2.06046 | -0.62788 | 0.66478 | |
| GSK3A | 7.264127 | 0.199336 | 0.999532 | | 8.446178 | 0.331174 | 0.674511 | | 7.596998 | 0.146277 | 0.895421 | |
| ALDH2 | 10.37517 | 0.171589 | 0.999532 | | 12.30084 | 0.313822 | 0.674511 | | 10.81924 | 0.17407 | 0.859426 | |
| VPS45 | 3.64625 | -0.27061 | 0.999532 | | 3.909939 | -0.43516 | 0.674511 | | 3.003182 | -0.14532 | 0.927544 | |
| SCLT1 | 3.762201 | -0.29492 | 0.999532 | | 3.910367 | -0.44408 | 0.674511 | | 3.07015 | -0.13014 | 0.943463 | |
| WDR27 | 2.348135 | -0.16992 | 0.999532 | | 2.353269 | -0.5562 | 0.674511 | | 1.799446 | -0.36568 | 0.837256 | |
| RAG1 | 2.621877 | 0.009757 | 0.999532 | | 2.605263 | -0.54255 | 0.674511 | | 2.15303 | -0.52747 | 0.720106 | |
| EPB41L4A | 4.382393 | 0.134009 | 0.999532 | | 5.18646 | 0.381076 | 0.674511 | | 4.598655 | 0.262405 | 0.816618 | |
| BUD31 | 7.482216 | -0.06481 | 0.999532 | | 9.620912 | 0.309505 | 0.674511 | | 7.917785 | 0.396485 | 0.619775 | |
| ZNF680 | 4.802917 | 0.085981 | 0.999532 | | 5.710954 | 0.374956 | 0.674511 | | 5.249533 | 0.298654 | 0.787995 | |
| CBX6 | 11.36543 | 0.138097 | 0.999532 | | 13.4258 | 0.305133 | 0.674511 | | 12.00231 | 0.189055 | 0.816611 | |
| NGFR | 4.186634 | -0.87019 | 0.999532 | | 5.047371 | -0.46065 | 0.674511 | | 3.527277 | 0.469693 | 0.717627 | |
| NAA20 | 5.266654 | 0.219225 | 0.999532 | | 5.917606 | 0.366956 | 0.674511 | | 5.635339 | 0.153651 | 0.896807 | |
| NCF1 | 4.457466 | 0.084319 | 0.999532 | | 4.159671 | -0.43187 | 0.674511 | | 3.803897 | -0.50189 | 0.647218 | |
| ZNF18 | 2.259654 | -0.29334 | 0.999532 | | 2.240066 | -0.55046 | 0.674511 | | 1.797189 | -0.24384 | 0.900481 | |
| POMGNT | 2.667518 | 0.155054 | 0.999532 | | 3.429627 | 0.447413 | 0.674511 | | 2.96389 | 0.307817 | 0.816589 | |
| CFL2 | 5.756804 | 0.291997 | 0.999532 | | 6.570405 | 0.358021 | 0.674511 | | 6.178235 | 0.066485 | 0.946958 | |
| PANK3 | 3.869573 | -0.71477 | 0.999532 | | 4.882697 | -0.40155 | 0.674511 | | 3.318525 | 0.339867 | 0.801445 | |
| ZNF608 | 2.262112 | 0.274059 | 0.999532 | | 2.74581 | 0.520215 | 0.674511 | | 2.483951 | 0.258364 | 0.87591 | |
| FBXO11 | 6.541993 | -0.03968 | 0.999532 | | 8.266435 | 0.312322 | 0.674511 | | 6.879068 | 0.368701 | 0.666906 | |
| CDK7 | 2.40438 | -0.19323 | 0.999532 | | 3.448335 | 0.47212 | 0.674511 | | 2.769959 | 0.685292 | 0.578798 | |
| TFAP2D | 2.093897 | 0.080309 | 0.999532 | | 1.866019 | -0.63495 | 0.674511 | | 1.743031 | -0.69506 | 0.666566 | |
| MYO5A | 2.030715 | 0.334152 | 0.999532 | | 2.358778 | 0.538895 | 0.674511 | | 2.351886 | 0.225941 | 0.895609 | |
| IFT52 | 5.014054 | 0.184701 | 0.999532 | | 4.42238 | -0.42994 | 0.674511 | | 4.136705 | -0.59286 | 0.542124 | |
| MIA3 | 5.206569 | 0.011431 | 0.999532 | | 7.003504 | 0.3965 | 0.674511 | | 5.784949 | 0.358785 | 0.722703 | |
| HIGD2A | 12.6258 | -0.05903 | 0.999532 | | 13.10357 | -0.26843 | 0.674511 | | 10.77256 | -0.18783 | 0.815713 | |
| PXK | 3.976514 | 0.05789 | 0.999532 | | 3.710255 | -0.47958 | 0.674511 | | 3.282986 | -0.52232 | 0.668532 | |
| TMC6 | 1.84733 | 0.239621 | 0.999532 | | 2.375119 | 0.558804 | 0.674511 | | 2.182301 | 0.338144 | 0.842686 | |
| SP3 | 8.985901 | 0.214201 | 0.999532 | | 9.953155 | 0.307117 | 0.674511 | | 9.148273 | 0.108043 | 0.918194 | |
| HCN1 | 2.568449 | 0.02765 | 0.999532 | | 2.441228 | -0.5666 | 0.674511 | | 2.064603 | -0.57571 | 0.704227 | |
| TMEM88 | 4.913425 | -0.57779 | 0.999532 | | 5.765017 | -0.37108 | 0.674511 | | 4.10145 | 0.22527 | 0.86306 | |
| XRN2 | 5.449981 | -0.40047 | 0.999532 | | 7.620764 | 0.356141 | 0.674511 | | 6.063979 | 0.771745 | 0.300399 | |
| SNIP1 | 2.191018 | 0.273637 | 0.999532 | | 1.950946 | -0.62516 | 0.674511 | | 1.902686 | -0.87562 | 0.542124 | |
| GIMAP5 | 1.766461 | 0.056087 | 0.999532 | | 2.27064 | 0.552217 | 0.674511 | | 2.074381 | 0.515318 | 0.736481 | |
| OR5D16 | 2.550884 | -0.04224 | 0.999532 | | 2.639852 | -0.54759 | 0.674511 | | 2.184177 | -0.46217 | 0.78258 | |
| KBTBD2 | 4.745382 | 0.502574 | 0.999532 | | 5.0448 | 0.399122 | 0.674511 | | 4.964504 | -0.08292 | 0.955834 | |
| LY96 | 2.698698 | -0.40614 | 0.999532 | | 2.791858 | -0.51478 | 0.674511 | | 2.057148 | -0.09932 | 0.966652 | |
| BEST1 | 2.867463 | -0.41862 | 0.999532 | | 3.173095 | -0.49394 | 0.674511 | | 2.432339 | -0.05687 | 0.982124 | |
| RPL22 | 45.2065 | -0.05312 | 0.999532 | | 56.3544 | 0.230513 | 0.674511 | | 46.95999 | 0.31224 | 0.535079 | |
| C4orf47 | 2.521337 | -0.14471 | 0.999532 | | 2.508132 | -0.56085 | 0.674534 | | 2.099008 | -0.39756 | 0.816775 | |
| TSPYL2 | 8.091576 | -0.47515 | 0.999532 | | 9.342897 | -0.32771 | 0.674649 | | 6.700932 | 0.166267 | 0.881837 | |
| CDC23 | 4.137521 | 0.240816 | 0.999532 | | 5.563623 | 0.395885 | 0.674796 | | 4.814892 | 0.200175 | 0.87892 | |
| HOOK1 | 2.705587 | -0.44935 | 0.999532 | | 2.811639 | -0.5047 | 0.674796 | | 2.121535 | -0.04431 | 0.988853 | |
| NXNL1 | 3.205915 | 0.39529 | 0.999532 | | 2.651924 | -0.56076 | 0.674796 | | 2.720201 | -0.93938 | 0.419745 | |
| PKHD1L1 | 2.55437 | -0.51283 | 0.999532 | | 2.636743 | -0.52828 | 0.674796 | | 2.02187 | -0.0003 | 0.999949 | |
| ETFDH | 4.671524 | 0.323425 | 0.999532 | | 5.479513 | 0.369118 | 0.674796 | | 5.157415 | 0.064785 | 0.965649 | |
| LHFPL2 | 2.666873 | -0.12064 | 0.999532 | | 3.348793 | 0.489187 | 0.674796 | | 2.920256 | 0.59142 | 0.666906 | |
| ADGRG5 | 2.649448 | 0.439276 | 0.999532 | | 2.168198 | -0.58338 | 0.674796 | | 2.273079 | -1.00364 | 0.421517 | |
| PLEKHB1 | 3.844 | -0.13041 | 0.999532 | | 3.644307 | -0.45536 | 0.675367 | | 3.120553 | -0.31086 | 0.827488 | |
| LACTBL1 | 2.654674 | -0.15272 | 0.999532 | | 2.736905 | -0.61277 | 0.675367 | | 2.080041 | -0.44426 | 0.791576 | |
| DNAJB9 | 7.214692 | 0.351564 | 0.999532 | | 8.314834 | 0.354289 | 0.675367 | | 7.863194 | 0.025117 | 0.989437 | |
| IFI27L2 | 3.51948 | 0.122491 | 0.999532 | | 4.457329 | 0.403423 | 0.676063 | | 3.94205 | 0.300835 | 0.806394 | |
| CACYBP | 5.039073 | 0.47688 | 0.999532 | | 5.48209 | 0.372369 | 0.676063 | | 5.347785 | -0.08358 | 0.953949 | |
| FABP7 | 2.325009 | -0.01155 | 0.999532 | | 2.247371 | -0.59119 | 0.676126 | | 1.932572 | -0.55725 | 0.737109 | |
| PHACTR2 | 7.656399 | 0.494407 | 0.999532 | | 8.115366 | 0.342725 | 0.676126 | | 8.254571 | -0.12457 | 0.909074 | |
| ZP1 | 2.340712 | 0.212813 | 0.999532 | | 2.155283 | -0.61447 | 0.676126 | | 1.981055 | -0.80443 | 0.59686 | |
| PLEKHG3 | 5.23962 | 0.415329 | 0.999532 | | 5.679226 | 0.3624 | 0.676126 | | 5.632882 | -0.03219 | 0.985693 | |
| C2orf81 | 4.425318 | 0.427547 | 0.999532 | | 3.711596 | -0.48571 | 0.676126 | | 3.929738 | -0.892 | 0.368714 | |
| ZNF407 | 4.615382 | -0.19577 | 0.999532 | | 4.738913 | -0.40043 | 0.676126 | | 3.869181 | -0.18666 | 0.894612 | |
| SLC13A1 | 2.91577 | 0.67637 | 0.999532 | | 3.09987 | 0.509527 | 0.676126 | | 3.19867 | -0.15165 | 0.927668 | |
| RPP25 | 3.87082 | -0.51269 | 0.999532 | | 4.519656 | -0.46009 | 0.676126 | | 3.281067 | 0.070401 | 0.973356 | |
| OSCAR | 2.584677 | 0.433573 | 0.999532 | | 2.141707 | -0.60125 | 0.676126 | | 2.105145 | -1.01498 | 0.414154 | |
| ATP6V0E | 14.83208 | -0.03474 | 0.999532 | | 18.91391 | 0.261997 | 0.676126 | | 15.49492 | 0.309046 | 0.686214 | |
| LOXL4 | 3.008305 | 0.045066 | 0.999532 | | 2.696638 | -0.57486 | 0.676126 | | 2.306419 | -0.59442 | 0.680564 | |
| COL12A1 | 7.954219 | -0.31459 | 0.999532 | | 11.65009 | 0.426769 | 0.676126 | | 9.003558 | 0.770597 | 0.363418 | |
| TMEM80 | 4.381539 | 0.465748 | 0.999532 | | 4.728073 | 0.400935 | 0.676126 | | 4.905834 | -0.04366 | 0.980581 | |
| TRPV2 | 2.983951 | 0.062118 | 0.999532 | | 3.820395 | 0.433802 | 0.676126 | | 3.337859 | 0.391424 | 0.750371 | |
| HOXD11 | 2.784268 | 0.755184 | 0.999532 | | 2.856016 | 0.501973 | 0.676126 | | 3.026153 | -0.23262 | 0.87764 | |
| HMGN1 | 33.53483 | 0.220976 | 0.999532 | | 37.14984 | 0.186521 | 0.676126 | | 34.0917 | -0.0135 | 0.990309 | |
| P2RY2 | 2.503046 | -0.70504 | 0.999532 | | 3.011211 | -0.49072 | 0.676126 | | 1.969165 | 0.235583 | 0.90066 | |
| CFAP58 | 2.713523 | 0.202491 | 0.999532 | | 2.35726 | -0.56719 | 0.676126 | | 2.313442 | -0.75488 | 0.579378 | |
| SLITRK4 | 2.671682 | -0.10294 | 0.999532 | | 2.57882 | -0.55712 | 0.676126 | | 2.089991 | -0.43267 | 0.786076 | |
| ATP11B | 3.00064 | 0.533398 | 0.999532 | | 2.904605 | 0.483932 | 0.67644 | | 3.214416 | -0.03756 | 0.988379 | |
| KRTAP5- | 2.922408 | 0.157696 | 0.999532 | | 2.609183 | -0.53803 | 0.676502 | | 2.460907 | -0.68228 | 0.628234 | |
| DAXX | 4.214327 | 0.099065 | 0.999532 | | 5.254516 | 0.371843 | 0.676822 | | 4.661451 | 0.291894 | 0.786052 | |
| EPHA6 | 2.112503 | -0.11011 | 0.999532 | | 2.029655 | -0.59745 | 0.676822 | | 1.708453 | -0.47053 | 0.791576 | |
| BRWD1 | 6.389208 | -0.14431 | 0.999532 | | 6.412528 | -0.3809 | 0.67685 | | 5.430898 | -0.20997 | 0.867976 | |
| RANBP3L | 2.666294 | 0.071917 | 0.999532 | | 2.562728 | -0.54089 | 0.67685 | | 2.160524 | -0.59251 | 0.668277 | |
| RPN2 | 13.57685 | 0.107879 | 0.999532 | | 16.2813 | 0.355274 | 0.677304 | | 14.60548 | 0.258069 | 0.790337 | |
| APH1A | 3.829555 | -0.08021 | 0.999532 | | 5.483771 | 0.382963 | 0.677304 | | 4.335801 | 0.486018 | 0.647218 | |
| GALR3 | 9.16109 | 0.164071 | 0.999532 | | 9.060555 | -0.41835 | 0.677304 | | 8.012475 | -0.54926 | 0.557909 | |
| ALDH3A2 | 4.750495 | -0.17497 | 0.999532 | | 6.609239 | 0.359645 | 0.677304 | | 5.16332 | 0.556469 | 0.536706 | |
| STAMBP | 5.484981 | 0.0304 | 0.999532 | | 5.261892 | -0.39833 | 0.678496 | | 4.391603 | -0.40661 | 0.695449 | |
| SLC18B1 | 2.532811 | -0.20415 | 0.999532 | | 2.647474 | -0.53262 | 0.678539 | | 2.047899 | -0.30915 | 0.863808 | |
| B4GALT6 | 3.505673 | -0.19245 | 0.999532 | | 3.509768 | -0.48251 | 0.678539 | | 2.929407 | -0.25845 | 0.867271 | |
| CSNK1G3 | 2.930705 | 0.682325 | 0.999532 | | 3.131187 | 0.510676 | 0.678569 | | 3.188503 | -0.15158 | 0.92697 | |
| BRF1 | 4.99459 | -0.01713 | 0.999532 | | 4.867933 | -0.40439 | 0.678569 | | 4.100429 | -0.36837 | 0.750371 | |
| ATP23 | 2.124125 | -0.40297 | 0.999532 | | 2.275364 | -0.56021 | 0.678792 | | 1.700769 | -0.13993 | 0.95366 | |
| SELENOO | 4.11196 | 0.119966 | 0.999532 | | 5.050716 | 0.393988 | 0.678792 | | 4.535739 | 0.293072 | 0.797138 | |
| ATF4 | 18.74898 | -0.11389 | 0.999532 | | 23.64241 | 0.241986 | 0.678792 | | 19.54656 | 0.369845 | 0.552672 | |
| MECR | 3.349186 | -0.21651 | 0.999532 | | 3.388682 | -0.48921 | 0.678792 | | 2.666175 | -0.25562 | 0.877814 | |
| PRPF18 | 4.841376 | 0.27403 | 0.999532 | | 5.366979 | 0.38619 | 0.6789 | | 5.255838 | 0.140799 | 0.914692 | |
| PROX2 | 2.899751 | 0.104008 | 0.999532 | | 2.694134 | -0.56872 | 0.6789 | | 2.394848 | -0.65383 | 0.661666 | |
| EPHX1 | 6.363419 | 0.087054 | 0.999532 | | 7.539024 | 0.317196 | 0.6789 | | 6.595681 | 0.242344 | 0.803942 | |
| ZNF518A | 4.196291 | 0.421782 | 0.999532 | | 4.901464 | 0.44928 | 0.6789 | | 4.62989 | 0.053148 | 0.978229 | |
| OLIG1 | 3.541717 | -0.29765 | 0.999532 | | 3.774526 | -0.47226 | 0.6789 | | 2.822503 | -0.1566 | 0.929335 | |
| GOLGA4 | 5.309555 | 0.148263 | 0.999532 | | 6.463002 | 0.374605 | 0.6789 | | 5.61472 | 0.252446 | 0.811123 | |
| MEI1 | 2.315944 | 0.080023 | 0.999532 | | 2.20854 | -0.60353 | 0.6789 | | 1.933059 | -0.66551 | 0.666566 | |
| DNM1L | 3.688403 | 0.284641 | 0.999532 | | 4.449071 | 0.423925 | 0.6789 | | 4.020479 | 0.158163 | 0.907016 | |
| PDLIM1 | 15.04617 | -0.40507 | 0.999532 | | 17.4009 | -0.24053 | 0.6789 | | 12.88182 | 0.182598 | 0.806394 | |
| TSPAN7 | 13.14928 | -0.28213 | 0.999532 | | 17.93533 | 0.309666 | 0.6789 | | 14.47448 | 0.610527 | 0.335411 | |
| CARD17 | 2.204523 | 0.154364 | 0.999532 | | 1.89174 | -0.60637 | 0.6789 | | 1.843524 | -0.74387 | 0.638728 | |
| RELL2 | 2.720088 | 0.275225 | 0.999532 | | 2.413127 | -0.54752 | 0.6789 | | 2.343037 | -0.80507 | 0.532055 | |
| MRPS31 | 1.980771 | 0.505012 | 0.999532 | | 2.275151 | 0.546239 | 0.6789 | | 2.287662 | 0.058018 | 0.979432 | |
| STRA6 | 2.987155 | 0.077492 | 0.999532 | | 2.828994 | -0.53169 | 0.6789 | | 2.380407 | -0.59239 | 0.657538 | |
| SF3A2 | 5.95945 | 0.116221 | 0.999532 | | 6.768048 | 0.334595 | 0.679804 | | 6.191358 | 0.225115 | 0.834593 | |
| PELI3 | 2.273896 | 0.768726 | 0.999532 | | 2.17022 | 0.564864 | 0.679804 | | 2.42201 | -0.18868 | 0.915533 | |
| ITPRIPL2 | 3.689897 | -0.36158 | 0.999532 | | 5.430917 | 0.369417 | 0.679804 | | 4.337982 | 0.756164 | 0.374949 | |
| CNP | 4.538988 | -0.19403 | 0.999532 | | 6.127892 | 0.363798 | 0.679891 | | 5.161188 | 0.5748 | 0.536234 | |
| RNASE13 | 2.547332 | 0.220065 | 0.999532 | | 2.287295 | -0.58025 | 0.679891 | | 2.174241 | -0.78588 | 0.563461 | |
| PLXNA1 | 1.961783 | -0.00233 | 0.999532 | | 2.806601 | 0.508873 | 0.679891 | | 2.398502 | 0.531235 | 0.707142 | |
| EIF3M | 7.3997 | 0.084029 | 0.999532 | | 8.805451 | 0.294797 | 0.679891 | | 7.600657 | 0.221112 | 0.810635 | |
| OR5D13 | 2.119911 | -0.51421 | 0.999532 | | 2.314695 | -0.5919 | 0.679891 | | 1.589116 | -0.06274 | 0.985377 | |
| ZNF157 | 1.937664 | -0.00166 | 0.999532 | | 1.834258 | -0.61934 | 0.679891 | | 1.664713 | -0.59795 | 0.723294 | |
| ZBTB49 | 2.649537 | 0.077864 | 0.999532 | | 2.593473 | -0.53727 | 0.679891 | | 2.248238 | -0.58368 | 0.68597 | |
| ACBD6 | 4.32045 | 0.286888 | 0.999532 | | 5.261259 | 0.42693 | 0.679891 | | 4.734984 | 0.161079 | 0.903339 | |
| PRKAA1 | 7.850553 | -0.33387 | 0.999532 | | 10.97688 | 0.283785 | 0.679891 | | 8.658939 | 0.642126 | 0.266933 | |
| SLIT3 | 10.71244 | -0.17261 | 0.999532 | | 15.03245 | 0.378203 | 0.680302 | | 11.99868 | 0.559822 | 0.418768 | |
| SLC6A16 | 2.918565 | 0.021118 | 0.999532 | | 2.955171 | -0.5352 | 0.68037 | | 2.455052 | -0.52082 | 0.722703 | |
| PRMT9 | 2.924112 | 0.124712 | 0.999532 | | 2.458193 | -0.57668 | 0.68037 | | 2.276381 | -0.69105 | 0.647833 | |
| OXT | 2.116744 | 0.054502 | 0.999532 | | 2.02218 | -0.60657 | 0.680393 | | 1.778901 | -0.63986 | 0.694961 | |
| FAM83H | 3.263416 | 0.324316 | 0.999532 | | 4.060429 | 0.428162 | 0.680533 | | 3.681225 | 0.116757 | 0.940162 | |
| CBLIF | 2.402266 | 0.227824 | 0.999532 | | 2.13777 | -0.5919 | 0.680725 | | 2.104949 | -0.79782 | 0.589729 | |
| ACOT11 | 2.855991 | 0.170937 | 0.999532 | | 3.50256 | 0.4783 | 0.680725 | | 3.039065 | 0.330652 | 0.814434 | |
| CLEC10A | 5.387271 | -0.17522 | 0.999532 | | 5.600889 | -0.37148 | 0.680786 | | 4.435325 | -0.17902 | 0.889853 | |
| TEPP | 4.004348 | -0.28768 | 0.999532 | | 4.341621 | -0.46033 | 0.681526 | | 3.305641 | -0.1566 | 0.925421 | |
| CTBS | 3.834845 | 0.429706 | 0.999532 | | 4.320215 | 0.424282 | 0.681526 | | 4.18086 | 0.014573 | 0.996681 | |
| ISX | 2.213033 | -0.18326 | 0.999532 | | 2.220943 | -0.60206 | 0.681816 | | 1.744353 | -0.4 | 0.833306 | |
| ZFP28 | 2.463796 | 0.710442 | 0.999532 | | 2.45403 | 0.529974 | 0.681816 | | 2.756224 | -0.16184 | 0.926219 | |
| MTERF4 | 3.617728 | 0.013227 | 0.999532 | | 4.680585 | 0.400986 | 0.681816 | | 4.083525 | 0.407704 | 0.712664 | |
| RBM5 | 8.846767 | -0.01486 | 0.999532 | | 11.2062 | 0.279818 | 0.681981 | | 9.313611 | 0.312915 | 0.669356 | |
| ABHD15 | 2.805707 | 0.297754 | 0.999532 | | 2.526878 | -0.53761 | 0.681981 | | 2.385443 | -0.81238 | 0.515762 | |
| ARAP2 | 2.950757 | 0.130906 | 0.999532 | | 3.623913 | 0.467977 | 0.681981 | | 3.262947 | 0.352077 | 0.803301 | |
| MBL2 | 2.054043 | -0.08709 | 0.999532 | | 2.129532 | -0.59437 | 0.682188 | | 1.633363 | -0.48546 | 0.783005 | |
| LGALS8 | 7.994336 | 0.055473 | 0.999532 | | 9.60443 | 0.310103 | 0.682188 | | 8.307758 | 0.272285 | 0.753585 | |
| TMCO1 | 7.736442 | 0.193399 | 0.999532 | | 8.967373 | 0.296177 | 0.682188 | | 7.992938 | 0.123124 | 0.904801 | |
| DEFB134 | 3.056461 | 1.187699 | 0.654806 | | 1.745643 | -0.64757 | 0.682188 | | 2.71621 | -1.82023 | 0.062627 | DOWN |
| SH3RF3 | 2.907522 | -0.08322 | 0.999532 | | 2.885491 | -0.51759 | 0.682188 | | 2.486805 | -0.41668 | 0.789885 | |
| POLR3F | 2.227035 | 0.068528 | 0.999532 | | 2.991455 | 0.521867 | 0.682188 | | 2.705465 | 0.473586 | 0.750371 | |
| GARRE1 | 3.132311 | -0.39631 | 0.999532 | | 4.474765 | 0.42086 | 0.682188 | | 3.406032 | 0.839904 | 0.378764 | |
| PSORS1C | 3.364567 | -0.14383 | 0.999532 | | 3.421143 | -0.47641 | 0.682188 | | 2.719565 | -0.31488 | 0.837256 | |
| FKRP | 4.062627 | -0.1345 | 0.999532 | | 3.799884 | -0.45436 | 0.682188 | | 3.261495 | -0.32624 | 0.831293 | |
| TMEM219 | 7.145467 | -0.01901 | 0.999532 | | 8.994866 | 0.314461 | 0.682188 | | 7.740052 | 0.351964 | 0.666906 | |
| BUB3 | 5.715712 | 0.106875 | 0.999532 | | 7.134164 | 0.353316 | 0.682188 | | 6.155379 | 0.266657 | 0.801572 | |
| KIF4A | 2.277021 | -0.22097 | 0.999532 | | 2.144025 | -0.56372 | 0.682188 | | 1.886022 | -0.32888 | 0.868512 | |
| ZNF8 | 2.478201 | -0.03972 | 0.999532 | | 2.178281 | -0.57734 | 0.682188 | | 1.92152 | -0.52754 | 0.749222 | |
| IL17F | 2.585679 | -0.11181 | 0.999532 | | 2.430246 | -0.56144 | 0.682188 | | 1.998772 | -0.43044 | 0.79686 | |
| DYRK1B | 2.96481 | 0.012922 | 0.999532 | | 3.806487 | 0.438585 | 0.682188 | | 3.310165 | 0.441885 | 0.723933 | |
| MPV17L | 3.934063 | -0.10976 | 0.999532 | | 3.986923 | -0.46563 | 0.682188 | | 3.264727 | -0.33707 | 0.806662 | |
| PTGIR | 2.421902 | -0.50766 | 0.999532 | | 2.596001 | -0.52459 | 0.682188 | | 1.97888 | -0.00809 | 0.997987 | |
| STAT1 | 9.33462 | 0.14436 | 0.999532 | | 11.19752 | 0.336719 | 0.682188 | | 10.05379 | 0.212688 | 0.806797 | |
| PDX1 | 2.054666 | 0.577675 | 0.999532 | | 2.240691 | 0.58786 | 0.682188 | | 2.443341 | 0.020539 | 0.996681 | |
| MCMBP | 3.472987 | 0.34885 | 0.999532 | | 3.888968 | 0.417987 | 0.682188 | | 3.818041 | 0.08882 | 0.95099 | |
| EMX2 | 3.444888 | 0.708376 | 0.999532 | | 3.570301 | 0.469889 | 0.682188 | | 3.685664 | -0.21709 | 0.877898 | |
| KCNA7 | 2.352525 | 0.019994 | 0.999532 | | 2.205935 | -0.59728 | 0.682188 | | 1.946085 | -0.5988 | 0.712621 | |
| AKR7A3 | 3.820672 | -0.14531 | 0.999532 | | 4.023997 | -0.46379 | 0.682188 | | 3.327884 | -0.3059 | 0.827267 | |
| PARD6B | 4.372002 | 0.121453 | 0.999532 | | 5.758438 | 0.382546 | 0.682188 | | 4.623315 | 0.272285 | 0.813143 | |
| UNC13B | 2.565795 | 0.23782 | 0.999532 | | 3.235814 | 0.471027 | 0.682188 | | 2.983007 | 0.256581 | 0.868998 | |
| ESM1 | 3.403992 | -0.2672 | 0.999532 | | 3.478521 | -0.47765 | 0.682188 | | 2.81737 | -0.18121 | 0.911296 | |
| PPP2R1A | 5.275914 | 0.303418 | 0.999532 | | 6.222529 | 0.356749 | 0.682298 | | 5.483984 | 0.064158 | 0.972221 | |
| PRRG4 | 3.26905 | 0.147229 | 0.999532 | | 3.106388 | -0.5001 | 0.682298 | | 2.883116 | -0.62735 | 0.630705 | |
| ZNF343 | 2.773339 | 0.461879 | 0.999532 | | 3.124101 | 0.477457 | 0.682298 | | 3.158803 | 0.036676 | 0.988853 | |
| ZC3H15 | 4.062751 | -0.25891 | 0.999532 | | 5.670662 | 0.363416 | 0.682298 | | 4.427226 | 0.640676 | 0.484303 | |
| UBE2A | 3.583197 | 0.340068 | 0.999532 | | 4.234985 | 0.412442 | 0.682298 | | 3.827968 | 0.09204 | 0.954087 | |
| SLC35E2 | 2.698351 | -0.00997 | 0.999532 | | 2.469496 | -0.54286 | 0.682298 | | 2.138636 | -0.51479 | 0.732486 | |
| FBLN5 | 6.670269 | 0.161844 | 0.999532 | | 8.806536 | 0.604324 | 0.682343 | | 7.856795 | 0.462937 | 0.783005 | |
| UBAP1 | 4.06919 | -0.00563 | 0.999532 | | 5.157865 | 0.378136 | 0.682343 | | 4.481944 | 0.401871 | 0.706749 | |
| VCP | 11.6969 | -0.1713 | 0.999532 | | 15.08087 | 0.266332 | 0.682343 | | 12.14959 | 0.457918 | 0.456688 | |
| TAS2R10 | 3.337598 | 0.143295 | 0.999532 | | 3.178858 | -0.47758 | 0.682343 | | 2.936909 | -0.60184 | 0.629744 | |
| GPR183 | 3.209073 | -0.18535 | 0.999532 | | 3.33808 | -0.50327 | 0.682343 | | 2.592403 | -0.29512 | 0.859522 | |
| TTC39A | 2.069957 | -0.22255 | 0.999532 | | 2.014575 | -0.60327 | 0.682363 | | 1.582017 | -0.36519 | 0.858767 | |
| SULF2 | 5.933267 | 0.056922 | 0.999532 | | 7.584933 | 0.334935 | 0.682363 | | 6.384044 | 0.3088 | 0.76158 | |
| DMRTB1 | 2.264405 | 0.16511 | 0.999532 | | 2.13433 | -0.60552 | 0.682363 | | 1.934071 | -0.74991 | 0.630705 | |
| CCNG1 | 4.103177 | 0.229394 | 0.999532 | | 5.005782 | 0.400104 | 0.682363 | | 4.596963 | 0.191523 | 0.88476 | |
| HYAL3 | 2.630236 | -0.4483 | 0.999532 | | 3.01909 | -0.53626 | 0.682363 | | 2.267491 | -0.07067 | 0.978282 | |
| EWSR1 | 14.04923 | -0.11804 | 0.999532 | | 18.30809 | 0.272139 | 0.682363 | | 14.96177 | 0.408346 | 0.49299 | |
| PRR23D2 | 2.704976 | -0.01725 | 0.999532 | | 2.611696 | -0.58844 | 0.682363 | | 2.13931 | -0.5468 | 0.723933 | |
| PRMT3 | 2.047181 | 0.36045 | 0.999532 | | 2.409743 | 0.539329 | 0.682363 | | 2.306309 | 0.19772 | 0.911296 | |
| EIF3J | 5.758821 | 0.09176 | 0.999532 | | 5.259133 | -0.37939 | 0.682377 | | 4.838963 | -0.46642 | 0.660097 | |
| SNAP29 | 5.742158 | -0.04687 | 0.999532 | | 5.924149 | -0.36554 | 0.68251 | | 4.954493 | -0.29762 | 0.790523 | |
| ERVW-1 | 2.161459 | 0.277764 | 0.999532 | | 1.935851 | -0.65289 | 0.682514 | | 1.620584 | -0.90686 | 0.538234 | |
| TMEM198 | 3.192472 | 0.342383 | 0.999532 | | 2.592294 | -0.51542 | 0.682514 | | 2.676553 | -0.84004 | 0.477483 | |
| NPNT | 106.5721 | 0.57085 | 0.912432 | | 107.2501 | 0.284785 | 0.682514 | | 110.7976 | -0.2567 | 0.69872 | |
| PLA2G15 | 3.022693 | 0.325012 | 0.999532 | | 3.573434 | 0.461141 | 0.682514 | | 3.313064 | 0.155063 | 0.920259 | |
| LRRTM1 | 1.957487 | -0.09605 | 0.999532 | | 1.871722 | -0.60998 | 0.682514 | | 1.579395 | -0.4972 | 0.791466 | |
| COG6 | 2.837994 | 0.912858 | 0.999532 | | 2.64354 | 0.512817 | 0.682514 | | 3.071873 | -0.38674 | 0.781295 | |
| ASPHD1 | 2.443634 | 0.097086 | 0.999532 | | 2.241991 | -0.59165 | 0.682514 | | 2.0337 | -0.67179 | 0.657538 | |
| USP1 | 2.387718 | -0.16172 | 0.999532 | | 3.148223 | 0.460477 | 0.682514 | | 2.598324 | 0.634859 | 0.622543 | |
| TECR | 4.407774 | 0.072055 | 0.999532 | | 5.414054 | 0.397968 | 0.682514 | | 4.674287 | 0.348057 | 0.750711 | |
| COX8A | 18.83036 | -0.00934 | 0.999532 | | 22.99755 | 0.253396 | 0.682514 | | 19.2114 | 0.281102 | 0.723453 | |
| COL9A3 | 2.481634 | 0.319507 | 0.999532 | | 1.989811 | -0.6051 | 0.682514 | | 2.002004 | -0.9204 | 0.527774 | |
| QSOX2 | 1.932585 | 0.561555 | 0.999532 | | 2.271639 | 0.549296 | 0.682514 | | 2.132004 | -0.00064 | 0.999758 | |
| FYB2 | 2.416131 | 0.281935 | 0.999532 | | 2.074054 | -0.58626 | 0.682514 | | 2.025829 | -0.85078 | 0.536289 | |
| CASP9 | 2.140205 | -0.00204 | 0.999532 | | 2.086616 | -0.60599 | 0.682514 | | 1.653442 | -0.58309 | 0.723933 | |
| DUSP9 | 2.780039 | 0.355185 | 0.999532 | | 2.322701 | -0.57848 | 0.682514 | | 2.243279 | -0.91245 | 0.48639 | |
| PKP4 | 6.892749 | -0.19149 | 0.999532 | | 9.464819 | 0.32774 | 0.682514 | | 7.246852 | 0.542285 | 0.504771 | |
| LRP5L | 2.384852 | 0.108965 | 0.999532 | | 2.223897 | -0.60003 | 0.682514 | | 1.901676 | -0.6894 | 0.65516 | |
| PERCC1 | 2.881034 | -0.56886 | 0.999532 | | 3.10288 | -0.49659 | 0.682514 | | 2.20622 | 0.091478 | 0.970498 | |
| RGP1 | 3.219715 | 0.02426 | 0.999532 | | 4.332303 | 0.418201 | 0.682514 | | 3.61451 | 0.420263 | 0.720526 | |
| CA7 | 2.381376 | 0.033977 | 0.999532 | | 2.346615 | -0.56811 | 0.682514 | | 1.961816 | -0.58159 | 0.705888 | |
| CDH16 | 3.31842 | 0.270864 | 0.999532 | | 4.167642 | 0.461656 | 0.682514 | | 3.553305 | 0.254971 | 0.886601 | |
| MFNG | 3.222711 | -0.22639 | 0.999532 | | 3.342407 | -0.45584 | 0.682718 | | 2.585672 | -0.20714 | 0.901033 | |
| CPT1B | 2.889769 | -0.24546 | 0.999532 | | 2.872852 | -0.50196 | 0.682718 | | 2.294123 | -0.24431 | 0.892004 | |
| HMGCR | 3.166961 | -0.32756 | 0.999532 | | 3.469189 | -0.47938 | 0.682742 | | 2.54436 | -0.13231 | 0.947941 | |
| HIVEP2 | 3.074825 | -0.02373 | 0.999532 | | 4.189441 | 0.438746 | 0.682742 | | 3.550412 | 0.480806 | 0.679818 | |
| DNAJC27 | 3.611958 | 0.146496 | 0.999532 | | 3.341007 | -0.50319 | 0.682742 | | 3.184196 | -0.63715 | 0.626811 | |
| STBD1 | 7.477716 | -0.42145 | 0.999532 | | 8.937558 | -0.33523 | 0.682742 | | 6.217938 | 0.098298 | 0.940266 | |
| CD72 | 2.847372 | 0.333542 | 0.999532 | | 2.526022 | -0.55153 | 0.682742 | | 2.550712 | -0.87392 | 0.49299 | |
| HDAC7 | 8.455836 | -0.20519 | 0.999532 | | 11.34966 | 0.293743 | 0.682753 | | 9.01899 | 0.518706 | 0.413122 | |
| TRMT112 | 10.76392 | -0.24506 | 0.999532 | | 14.56276 | 0.265095 | 0.682753 | | 11.34385 | 0.534544 | 0.392322 | |
| ODAD3 | 2.605136 | 0.089299 | 0.999532 | | 2.377743 | -0.56622 | 0.68282 | | 2.254007 | -0.63859 | 0.674878 | |
| LAMTOR | 6.49615 | -0.03925 | 0.999532 | | 8.290842 | 0.323992 | 0.683322 | | 7.020576 | 0.384053 | 0.645149 | |
| TFPT | 3.075924 | -0.17449 | 0.999532 | | 3.1726 | -0.48448 | 0.683395 | | 2.547297 | -0.29007 | 0.859426 | |
| APOLD1 | 2.856321 | -0.84406 | 0.999532 | | 3.331832 | -0.73466 | 0.683673 | | 2.089031 | 0.162569 | 0.945902 | |
| CLPS | 2.654939 | 0.131061 | 0.999532 | | 2.390987 | -0.54993 | 0.683719 | | 2.263542 | -0.66755 | 0.651111 | |
| CNGA3 | 2.293439 | 0.159777 | 0.999532 | | 2.003722 | -0.58757 | 0.683719 | | 1.890027 | -0.73517 | 0.643272 | |
| FANCD2 | 2.533376 | 0.069 | 0.999532 | | 2.41696 | -0.57987 | 0.683719 | | 2.195776 | -0.63899 | 0.667704 | |
| ZNF589 | 2.395689 | 0.328722 | 0.999532 | | 2.865477 | 0.483288 | 0.683719 | | 2.810033 | 0.175685 | 0.914929 | |
| CRHBP | 38.55692 | -0.2195 | 0.999532 | | 52.52995 | 0.322765 | 0.683719 | | 41.97303 | 0.568539 | 0.277362 | |
| LRFN5 | 2.848186 | 0.36077 | 0.999532 | | 2.371998 | -0.56083 | 0.683719 | | 2.473887 | -0.90147 | 0.474973 | |
| AUH | 3.649958 | 0.357829 | 0.999532 | | 4.103877 | 0.417307 | 0.683831 | | 4.006793 | 0.077424 | 0.965619 | |
| MMADHC | 6.26242 | -0.04473 | 0.999532 | | 8.215662 | 0.329972 | 0.683831 | | 6.78072 | 0.39863 | 0.659727 | |
| TXNRD1 | 4.797214 | 0.03117 | 0.999532 | | 5.901235 | 0.371069 | 0.683831 | | 5.137451 | 0.355106 | 0.723933 | |
| PIWIL1 | 2.24219 | -0.49088 | 0.999532 | | 2.391376 | -0.56629 | 0.683831 | | 1.857178 | -0.05869 | 0.985873 | |
| PTPA | 3.505792 | 0.342713 | 0.999532 | | 4.199782 | 0.454963 | 0.683831 | | 3.967573 | 0.12914 | 0.938429 | |
| LILRA6 | 3.909078 | -0.08627 | 0.999532 | | 4.039469 | -0.44071 | 0.683831 | | 3.283428 | -0.33525 | 0.800239 | |
| NT5DC2 | 3.719637 | -0.48029 | 0.999532 | | 4.234332 | -0.43959 | 0.683831 | | 3.143734 | 0.066609 | 0.974421 | |
| PRR13 | 9.221583 | -0.40986 | 0.999532 | | 10.96534 | -0.29508 | 0.683831 | | 7.480912 | 0.121413 | 0.907968 | |
| TEDDM1 | 2.858281 | 0.196073 | 0.999532 | | 2.602826 | -0.55434 | 0.683831 | | 2.43128 | -0.72324 | 0.617899 | |
| REM2 | 3.681169 | -0.02084 | 0.999532 | | 3.870448 | -0.4684 | 0.68418 | | 3.038946 | -0.42779 | 0.740093 | |
| SNRNP25 | 2.734655 | -0.12087 | 0.999532 | | 2.740899 | -0.5252 | 0.68418 | | 2.417086 | -0.38484 | 0.815293 | |
| EPPK1 | 2.096355 | 0.485044 | 0.999532 | | 2.255447 | 0.543718 | 0.684181 | | 2.279665 | 0.073599 | 0.974421 | |
| NCOA5 | 4.527304 | 0.49521 | 0.999532 | | 4.690206 | 0.391717 | 0.684322 | | 4.629859 | -0.08437 | 0.955834 | |
| OSBP2 | 2.090991 | -0.28579 | 0.999532 | | 2.109431 | -0.59332 | 0.68477 | | 1.722144 | -0.28761 | 0.892004 | |
| B4GALT2 | 3.742509 | -0.00744 | 0.999532 | | 3.720715 | -0.46938 | 0.68477 | | 3.101201 | -0.44762 | 0.717627 | |
| FAM126A | 2.745344 | -0.07294 | 0.999532 | | 3.632625 | 0.438902 | 0.686149 | | 3.058291 | 0.530421 | 0.654321 | |
| TIMP4 | 3.042046 | 0.18834 | 0.999532 | | 2.813933 | -0.51296 | 0.686231 | | 2.530802 | -0.68015 | 0.59686 | |
| HSF5 | 2.01651 | -0.4249 | 0.999532 | | 2.021671 | -0.60432 | 0.686368 | | 1.582019 | -0.15995 | 0.949669 | |
| MAP2K3 | 5.283482 | -0.24017 | 0.999532 | | 5.57544 | -0.38351 | 0.686502 | | 4.312035 | -0.12561 | 0.934121 | |
| ART4 | 4.028033 | -0.18977 | 0.999532 | | 4.132347 | -0.43254 | 0.686611 | | 3.502063 | -0.22651 | 0.88588 | |
| WASF2 | 11.10734 | -0.07367 | 0.999532 | | 14.02411 | 0.272831 | 0.686761 | | 11.5388 | 0.367451 | 0.610861 | |
| CALHM4 | 4.107883 | 0.724302 | 0.999532 | | 2.99587 | -0.48867 | 0.687245 | | 3.516379 | -1.19528 | 0.154583 | |
| NCAM2 | 2.159105 | -0.07564 | 0.999532 | | 2.25521 | -0.56146 | 0.687325 | | 1.836821 | -0.46179 | 0.788243 | |
| TMEM60 | 4.647454 | -0.31603 | 0.999532 | | 5.068822 | -0.39954 | 0.687325 | | 3.941482 | -0.06175 | 0.974421 | |
| THOP1 | 5.316193 | 0.101747 | 0.999532 | | 5.016229 | -0.37492 | 0.687487 | | 4.46151 | -0.45902 | 0.647218 | |
| JADE1 | 2.97576 | 0.523128 | 0.999532 | | 3.498343 | 0.450363 | 0.687523 | | 3.259164 | -0.05187 | 0.978927 | |
| CER1 | 2.29465 | -0.258 | 0.999532 | | 2.22936 | -0.57113 | 0.687523 | | 1.802956 | -0.29671 | 0.881837 | |
| CEP131 | 3.025896 | 0.205302 | 0.999532 | | 2.439464 | -0.53076 | 0.687523 | | 2.433593 | -0.73152 | 0.595502 | |
| PROKR1 | 2.22029 | 0.020907 | 0.999532 | | 2.228132 | -0.55197 | 0.687523 | | 1.912667 | -0.54797 | 0.722703 | |
| FAN1 | 3.022702 | -0.37769 | 0.999532 | | 3.349135 | -0.47335 | 0.68785 | | 2.339626 | -0.07434 | 0.974421 | |
| SPTLC2 | 7.09734 | 0.079959 | 0.999532 | | 8.509124 | 0.310212 | 0.688112 | | 7.45103 | 0.249181 | 0.791466 | |
| ZNF443 | 3.255638 | 0.29542 | 0.999532 | | 2.931345 | -0.50208 | 0.688112 | | 2.722397 | -0.77659 | 0.511836 | |
| BABAM2 | 5.026539 | -0.42935 | 0.999532 | | 7.32935 | 0.345254 | 0.688112 | | 5.419335 | 0.800012 | 0.299128 | |
| CLEC18C | 6.125325 | 0.15114 | 0.999532 | | 7.434985 | 0.348521 | 0.688112 | | 6.634765 | 0.215608 | 0.836935 | |
| APC2 | 3.022419 | 0.086922 | 0.999532 | | 2.858998 | -0.54211 | 0.688112 | | 2.440855 | -0.60638 | 0.659367 | |
| CDH3 | 2.378222 | -0.12367 | 0.999532 | | 2.368065 | -0.57425 | 0.688112 | | 1.812368 | -0.42895 | 0.800239 | |
| RNF7 | 6.649835 | -0.07153 | 0.999532 | | 8.469161 | 0.322472 | 0.688112 | | 7.192744 | 0.42174 | 0.647218 | |
| C2orf80 | 2.085068 | -0.17836 | 0.999532 | | 2.121777 | -0.59021 | 0.688151 | | 1.741598 | -0.39361 | 0.836453 | |
| IDI1 | 6.440939 | -0.06914 | 0.999532 | | 6.372097 | -0.35408 | 0.688151 | | 5.194667 | -0.25893 | 0.806394 | |
| LAMTOR2 | 5.046996 | -0.37675 | 0.999532 | | 7.281655 | 0.333783 | 0.688151 | | 5.612997 | 0.729843 | 0.332359 | |
| PRY | 2.737218 | 4.83E-05 | 0.999912 | | 2.73421 | -0.5948 | 0.688151 | | 2.132374 | -0.57797 | 0.717138 | |
| ARHGEF1 | 11.19057 | 0.034106 | 0.999532 | | 13.21746 | 0.281768 | 0.688151 | | 11.63624 | 0.257529 | 0.761699 | |
| SOX5 | 3.101967 | -0.16678 | 0.999532 | | 3.05385 | -0.4753 | 0.688151 | | 2.632323 | -0.29943 | 0.852074 | |
| NMNAT2 | 2.201066 | -0.15955 | 0.999532 | | 2.213026 | -0.57489 | 0.688661 | | 1.82049 | -0.39666 | 0.831013 | |
| VOPP1 | 4.052239 | -0.33715 | 0.999532 | | 5.47615 | 0.369256 | 0.688661 | | 4.357203 | 0.725082 | 0.421517 | |
| DEFB128 | 3.000622 | 0.268147 | 0.999532 | | 2.531144 | -0.56777 | 0.688934 | | 2.503741 | -0.81869 | 0.535079 | |
| CYP11B1 | 2.517353 | 0.1669 | 0.999532 | | 2.228819 | -0.56684 | 0.688934 | | 2.093491 | -0.71854 | 0.64142 | |
| HES6 | 2.005428 | 0.039852 | 0.999532 | | 1.860155 | -0.62122 | 0.688934 | | 1.725093 | -0.64593 | 0.708052 | |
| PITRM1 | 3.779025 | 0.640663 | 0.999532 | | 3.840754 | 0.458118 | 0.688934 | | 4.018924 | -0.15352 | 0.915991 | |
| AKTIP | 3.477025 | 0.250253 | 0.999532 | | 4.319315 | 0.426103 | 0.688934 | | 3.903407 | 0.205856 | 0.889853 | |
| DAAM1 | 2.31437 | 0.370859 | 0.999532 | | 2.886372 | 0.502274 | 0.688934 | | 2.60375 | 0.154767 | 0.931619 | |
| AGTRAP | 3.166856 | -0.33642 | 0.999532 | | 3.32613 | -0.45155 | 0.688934 | | 2.426047 | -0.09908 | 0.96129 | |
| SAMD4A | 2.807414 | -0.32053 | 0.999532 | | 3.002208 | -0.51962 | 0.688934 | | 2.423929 | -0.19875 | 0.910105 | |
| ABCB10 | 5.306269 | 0.880091 | 0.547204 | | 4.793424 | 0.388294 | 0.688934 | | 5.720931 | -0.47794 | 0.600081 | |
| POLD2 | 2.950521 | -0.42506 | 0.999532 | | 4.338952 | 0.404122 | 0.688934 | | 3.333544 | 0.84628 | 0.39437 | |
| ATP2C1 | 5.422035 | 0.306247 | 0.999532 | | 5.875739 | 0.352204 | 0.688934 | | 5.736769 | 0.061501 | 0.96832 | |
| RPLP1 | 29.41685 | -0.2187 | 0.999532 | | 38.46102 | 0.223532 | 0.688934 | | 30.2223 | 0.465779 | 0.407399 | |
| TTC30A | 3.767473 | 0.494253 | 0.999532 | | 2.970181 | -0.49377 | 0.688934 | | 3.331583 | -0.97504 | 0.312156 | |
| WDR45 | 3.967464 | 0.093314 | 0.999532 | | 5.224896 | 0.385776 | 0.688934 | | 4.71118 | 0.324651 | 0.789312 | |
| SCML4 | 2.293528 | -0.35904 | 0.999532 | | 2.414222 | -0.58213 | 0.688934 | | 1.884838 | -0.20848 | 0.921708 | |
| MT1M | 15.04666 | -0.27712 | 0.999532 | | 16.88605 | -0.28302 | 0.688934 | | 12.76443 | 0.027708 | 0.983749 | |
| CLK4 | 3.734554 | 0.314556 | 0.999532 | | 4.37866 | 0.420988 | 0.688934 | | 4.142076 | 0.124542 | 0.934354 | |
| PRMT2 | 7.927575 | 0.498266 | 0.999532 | | 7.808738 | 0.326046 | 0.688934 | | 8.119023 | -0.17883 | 0.87591 | |
| NDUFV2 | 7.994077 | 0.153426 | 0.999532 | | 9.139128 | 0.289261 | 0.688934 | | 8.357671 | 0.14376 | 0.89901 | |
| PTPN21 | 7.706316 | 0.418861 | 0.999532 | | 8.584531 | 0.334576 | 0.688934 | | 8.261176 | -0.07364 | 0.948484 | |
| TNNC2 | 3.605681 | -0.23956 | 0.999532 | | 3.806496 | -0.44181 | 0.688934 | | 2.960287 | -0.18174 | 0.914687 | |
| GHSR | 2.138224 | -0.13916 | 0.999532 | | 2.062812 | -0.5747 | 0.689005 | | 1.74152 | -0.41994 | 0.816618 | |
| PUS7L | 1.815061 | 0.885525 | 0.999532 | | 1.792941 | 0.60692 | 0.6891 | | 1.95224 | -0.263 | 0.885783 | |
| PEX11G | 2.135628 | -0.04331 | 0.999532 | | 2.098541 | -0.58221 | 0.6891 | | 1.758627 | -0.52069 | 0.755301 | |
| LIN37 | 2.806857 | 0.627967 | 0.999532 | | 3.080399 | 0.469969 | 0.689194 | | 3.18038 | -0.13794 | 0.933104 | |
| HBZ | 3.693171 | 0.116189 | 0.999532 | | 3.523461 | -0.5005 | 0.689201 | | 3.028449 | -0.59262 | 0.632802 | |
| SIMC1 | 3.738671 | 0.332998 | 0.999532 | | 3.117147 | -0.49984 | 0.689201 | | 3.070055 | -0.81601 | 0.456688 | |
| FUNDC1 | 3.897236 | 0.337963 | 0.999532 | | 4.633746 | 0.41416 | 0.689201 | | 4.460057 | 0.0973 | 0.951656 | |
| THEM6 | 5.25256 | 0.050896 | 0.999532 | | 5.003973 | -0.39866 | 0.689201 | | 4.524372 | -0.43657 | 0.699971 | |
| ELOB | 21.36855 | -0.04013 | 0.999532 | | 26.04066 | 0.193002 | 0.689496 | | 21.6388 | 0.253373 | 0.652925 | |
| CST4 | 2.924002 | 0.095102 | 0.999532 | | 2.904127 | -0.51274 | 0.689496 | | 2.492415 | -0.59138 | 0.660958 | |
| RRM2 | 3.132764 | 0.28389 | 0.999532 | | 2.61907 | -0.51258 | 0.689496 | | 2.634387 | -0.78267 | 0.515762 | |
| BOK | 3.382691 | 0.194673 | 0.999532 | | 3.982679 | 0.418503 | 0.689578 | | 3.661924 | 0.242846 | 0.859898 | |
| TIGAR | 2.469268 | -0.30635 | 0.999532 | | 2.641579 | -0.52423 | 0.689578 | | 1.944257 | -0.19628 | 0.920259 | |
| SMAGP | 2.929653 | -0.00072 | 0.999532 | | 2.90116 | -0.50465 | 0.689684 | | 2.425381 | -0.48362 | 0.736481 | |
| NUMBL | 4.142667 | -0.14877 | 0.999532 | | 4.083172 | -0.42954 | 0.690172 | | 3.5882 | -0.26752 | 0.853549 | |
| UNC93B1 | 2.5714 | 0.174344 | 0.999532 | | 3.140941 | 0.477023 | 0.690449 | | 2.88227 | 0.319074 | 0.827267 | |
| OR4D10 | 2.319958 | 0.081705 | 0.999532 | | 1.939911 | -0.5774 | 0.690449 | | 1.995554 | -0.64297 | 0.686214 | |
| ELP5 | 2.786421 | 0.016617 | 0.999532 | | 3.581836 | 0.449653 | 0.690499 | | 3.203095 | 0.446659 | 0.723933 | |
| ZFYVE19 | 2.77794 | -0.03703 | 0.999532 | | 2.703633 | -0.50113 | 0.690499 | | 2.247016 | -0.44499 | 0.760955 | |
| STX11 | 3.329387 | 0.087687 | 0.999532 | | 3.175153 | -0.48421 | 0.690499 | | 2.749316 | -0.55454 | 0.665188 | |
| SCAF11 | 5.986566 | -0.2245 | 0.999532 | | 7.457572 | 0.329511 | 0.690499 | | 6.160594 | 0.541019 | 0.540342 | |
| TMEM74 | 2.753055 | 0.266808 | 0.999532 | | 2.222955 | -0.58935 | 0.690621 | | 2.152953 | -0.87036 | 0.552672 | |
| PIK3R2 | 4.994648 | 0.046276 | 0.999532 | | 6.312408 | 0.339232 | 0.690713 | | 5.543052 | 0.313224 | 0.755738 | |
| AHSG | 3.177329 | 0.013525 | 0.999532 | | 3.030469 | -0.51248 | 0.690745 | | 2.651843 | -0.51086 | 0.720106 | |
| LHFPL4 | 2.489417 | 0.342603 | 0.999532 | | 2.108593 | -0.5957 | 0.690858 | | 2.140587 | -0.92522 | 0.515762 | |
| MAP1B | 3.818222 | 0.23397 | 0.999532 | | 4.207235 | 0.460741 | 0.691201 | | 4.064401 | 0.21022 | 0.897096 | |
| C1orf167 | 2.154848 | -0.23794 | 0.999532 | | 2.316484 | -0.55358 | 0.691201 | | 1.833087 | -0.2903 | 0.883785 | |
| PHF2 | 5.50795 | -0.26644 | 0.999532 | | 6.177019 | -0.35652 | 0.691201 | | 4.717203 | -0.06556 | 0.969614 | |
| HECTD4 | 3.939761 | 0.076486 | 0.999532 | | 5.161848 | 0.39746 | 0.691411 | | 4.377034 | 0.33167 | 0.765852 | |
| DENND3 | 4.882666 | -0.33993 | 0.999532 | | 5.495986 | -0.38518 | 0.691411 | | 3.897778 | -0.02655 | 0.991159 | |
| TCF24 | 2.798802 | 0.041166 | 0.999532 | | 2.733438 | -0.55048 | 0.691411 | | 2.356602 | -0.57685 | 0.695449 | |
| RAB5B | 8.402367 | 0.040878 | 0.999532 | | 10.42368 | 0.286398 | 0.691411 | | 9.117951 | 0.263185 | 0.736481 | |
| L3MBTL1 | 2.97472 | 0.63199 | 0.999532 | | 2.405262 | -0.54164 | 0.691465 | | 2.570232 | -1.14599 | 0.266933 | |
| SLC34A3 | 3.654165 | 0.372486 | 0.999532 | | 2.855056 | -0.49774 | 0.691604 | | 3.107377 | -0.86625 | 0.47874 | |
| PGC | 2.384243 | -0.14269 | 0.999532 | | 2.297496 | -0.56812 | 0.691604 | | 1.853691 | -0.41055 | 0.82205 | |
| USP38 | 3.365724 | 0.011043 | 0.999532 | | 3.159956 | -0.46996 | 0.691604 | | 2.934531 | -0.46554 | 0.730719 | |
| TPST2 | 4.467861 | 0.220644 | 0.999532 | | 3.90851 | -0.46598 | 0.691687 | | 3.607966 | -0.65334 | 0.535662 | |
| OR1J1 | 2.468922 | -0.01289 | 0.999532 | | 2.26012 | -0.54721 | 0.691688 | | 2.047467 | -0.51975 | 0.740093 | |
| FADS6 | 2.413478 | -0.18623 | 0.999532 | | 2.512478 | -0.53638 | 0.691688 | | 2.080725 | -0.32645 | 0.861815 | |
| SIK3 | 2.21879 | -0.31276 | 0.999532 | | 2.253456 | -0.55117 | 0.691688 | | 1.671697 | -0.2211 | 0.914687 | |
| MAMDC4 | 2.286712 | -0.18917 | 0.999532 | | 2.32686 | -0.53295 | 0.691867 | | 1.852439 | -0.32556 | 0.858705 | |
| VAT1 | 4.3985 | -0.13481 | 0.999532 | | 6.112654 | 0.353415 | 0.691875 | | 4.759765 | 0.517176 | 0.616854 | |
| ASCC1 | 2.379133 | 0.295995 | 0.999532 | | 2.764005 | 0.483551 | 0.691875 | | 2.663075 | 0.206895 | 0.901033 | |
| FGD6 | 3.478144 | -0.1873 | 0.999532 | | 3.383293 | -0.46535 | 0.691875 | | 2.88568 | -0.27156 | 0.868998 | |
| IFNB1 | 2.615526 | 0.134809 | 0.999532 | | 2.520021 | -0.55555 | 0.692819 | | 2.358208 | -0.66569 | 0.658332 | |
| H3C4 | 4.066163 | 0.178094 | 0.999532 | | 5.182888 | 0.443542 | 0.692819 | | 4.561432 | 0.292314 | 0.816589 | |
| UMOD | 16.66931 | 0.82721 | 0.999532 | | 18.13433 | 0.743614 | 0.692819 | | 19.73287 | -0.05129 | 0.991341 | |
| MUC17 | 3.564772 | 0.595025 | 0.999532 | | 2.749893 | -0.51794 | 0.692819 | | 3.093969 | -1.09567 | 0.269743 | |
| KRTAP9- | 4.154634 | 0.55872 | 0.999532 | | 3.45281 | -0.45821 | 0.692819 | | 3.618806 | -0.99392 | 0.266933 | |
| DOCK1 | 6.054218 | -0.27227 | 0.999532 | | 8.470334 | 0.316801 | 0.692819 | | 6.42194 | 0.616269 | 0.421517 | |
| HIC2 | 2.347301 | 0.251362 | 0.999532 | | 2.736108 | 0.485589 | 0.692819 | | 2.497681 | 0.250637 | 0.877451 | |
| LSM4 | 6.375451 | -0.56948 | 0.999532 | | 7.905996 | -0.32944 | 0.692859 | | 5.423016 | 0.254076 | 0.811437 | |
| EML1 | 7.589397 | -0.05466 | 0.999532 | | 9.819192 | 0.295522 | 0.692859 | | 8.248446 | 0.369629 | 0.643272 | |
| EID2B | 2.090641 | -0.40721 | 0.999532 | | 2.157378 | -0.56803 | 0.69305 | | 1.647031 | -0.14921 | 0.952581 | |
| FN3K | 6.322375 | 0.222872 | 0.999532 | | 7.15113 | 0.342042 | 0.69305 | | 6.464884 | 0.126322 | 0.924131 | |
| RPL18 | 33.95628 | -0.01231 | 0.999532 | | 41.64005 | 0.245448 | 0.69305 | | 35.51093 | 0.2767 | 0.645149 | |
| FHOD1 | 2.789993 | -0.0245 | 0.999532 | | 3.734832 | 0.428025 | 0.69305 | | 3.177194 | 0.471312 | 0.69977 | |
| FNDC9 | 2.025029 | -0.21389 | 0.999532 | | 1.972294 | -0.5865 | 0.693141 | | 1.607762 | -0.35792 | 0.86194 | |
| TIMM13 | 8.116886 | 0.244637 | 0.999532 | | 9.170799 | 0.306059 | 0.693141 | | 8.574023 | 0.084236 | 0.94109 | |
| MDM2 | 2.451607 | -0.10079 | 0.999532 | | 2.477872 | -0.51428 | 0.693141 | | 1.902618 | -0.3946 | 0.813172 | |
| POLG | 2.892918 | -0.30298 | 0.999532 | | 4.189329 | 0.399468 | 0.693141 | | 3.309822 | 0.720155 | 0.505689 | |
| XRCC2 | 20.3841 | 0.112593 | 0.999532 | | 20.1271 | -0.31894 | 0.693141 | | 17.86338 | -0.41256 | 0.566812 | |
| PRKDC | 2.809085 | 0.17868 | 0.999532 | | 3.483926 | 0.438536 | 0.693141 | | 3.149597 | 0.279093 | 0.84579 | |
| FKBP4 | 7.949894 | 0.005714 | 0.999532 | | 8.312977 | -0.29996 | 0.693141 | | 6.925881 | -0.28323 | 0.742839 | |
| KLHL24 | 4.127296 | 0.390682 | 0.999532 | | 4.467525 | 0.397306 | 0.693141 | | 4.385285 | 0.022408 | 0.992286 | |
| GRPEL2 | 4.276125 | -0.33795 | 0.999532 | | 6.191452 | 0.407011 | 0.693141 | | 4.795675 | 0.767923 | 0.403558 | |
| CDC45 | 3.358916 | 0.003353 | 0.999532 | | 3.09117 | -0.50446 | 0.693141 | | 2.654109 | -0.48494 | 0.728277 | |
| PPP2R2A | 7.186175 | -0.30651 | 0.999532 | | 7.98439 | -0.30404 | 0.693141 | | 6.264686 | 0.019712 | 0.992286 | |
| EFCC1 | 2.326059 | 0.051545 | 0.999532 | | 2.240571 | -0.57802 | 0.693141 | | 1.997998 | -0.61354 | 0.69872 | |
| OR56B1 | 2.718242 | 0.317917 | 0.999532 | | 2.400154 | -0.58938 | 0.693141 | | 2.193959 | -0.8869 | 0.510095 | |
| SEC23IP | 3.240794 | 0.035183 | 0.999532 | | 4.234108 | 0.397754 | 0.693141 | | 3.556464 | 0.380516 | 0.746581 | |
| TRAPPC6 | 2.937021 | 0.062039 | 0.999532 | | 3.871334 | 0.418044 | 0.693141 | | 3.362755 | 0.375238 | 0.759211 | |
| PPP6R3 | 4.498975 | 0.113904 | 0.999532 | | 5.790453 | 0.363988 | 0.693141 | | 4.945293 | 0.268395 | 0.810121 | |

| | | | | | | | | | | | |
|---|---|---|---|---|---|---|---|---|---|---|---|
| ATP9B | 2.644806 | -0.26805 | 0.999532 | | 3.553659 | 0.43018 | 0.693141 | | 2.938912 | 0.702688 | 0.566812 |
| C1orf112 | 2.432254 | 0.064355 | 0.999532 | | 2.324634 | -0.55214 | 0.693141 | | 2.07046 | -0.5991 | 0.698562 |
| STOML2 | 3.968129 | 0.327237 | 0.999532 | | 4.627594 | 0.378561 | 0.693141 | | 4.410499 | 0.070915 | 0.965649 |
| TMEM63A | 7.319828 | 0.316397 | 0.999532 | | 7.786201 | 0.32998 | 0.693141 | | 7.762155 | 0.030119 | 0.988379 |
| DPY19L1 | 3.816934 | 0.288746 | 0.999532 | | 3.411848 | -0.45202 | 0.693141 | | 3.146783 | -0.72204 | 0.512069 |
| RAB20 | 4.890898 | -0.21286 | 0.999532 | | 5.40652 | -0.37517 | 0.693141 | | 4.207855 | -0.14004 | 0.920486 |
| SCML1 | 2.697045 | 0.19967 | 0.999532 | | 2.429555 | -0.52983 | 0.693141 | | 2.190003 | -0.71077 | 0.600081 |
| ME2 | 3.287582 | -0.17834 | 0.999532 | | 3.43324 | -0.4501 | 0.693141 | | 2.73062 | -0.25186 | 0.876428 |
| PANK1 | 4.135629 | -0.12014 | 0.999532 | | 4.244032 | -0.41957 | 0.693141 | | 3.390442 | -0.27708 | 0.837514 |
| PRSS33 | 1.884211 | 0.487884 | 0.999532 | | 2.330112 | 0.569703 | 0.693141 | | 2.357498 | 0.099147 | 0.967145 |
| RAI1 | 2.590442 | -0.41562 | 0.999532 | | 3.976816 | 0.428155 | 0.693141 | | 2.976561 | 0.862692 | 0.421517 |
| PDE2A | 5.017303 | -0.66901 | 0.999532 | | 6.24066 | -0.34641 | 0.693141 | | 4.08605 | 0.341356 | 0.759398 |
| EXOC2 | 3.395467 | 0.688809 | 0.999532 | | 3.346419 | 0.433141 | 0.693141 | | 3.701002 | -0.23446 | 0.864245 |
| ENTPD2 | 3.288107 | 0.440305 | 0.999532 | | 2.71032 | -0.51618 | 0.693141 | | 3.001636 | -0.94634 | 0.406785 |
| CYP51A1 | 7.177244 | 0.080235 | 0.999532 | | 6.914867 | -0.36961 | 0.693141 | | 6.025965 | -0.42481 | 0.654619 |
| PILRA | 3.293831 | -0.08145 | 0.999532 | | 3.200017 | -0.4732 | 0.693141 | | 2.784368 | -0.38633 | 0.786076 |
| ERAP1 | 5.658778 | -0.1507 | 0.999532 | | 5.744129 | -0.35038 | 0.693141 | | 4.756313 | -0.18706 | 0.881379 |
| PTK2B | 4.692412 | -0.28756 | 0.999532 | | 5.260313 | -0.37314 | 0.693141 | | 3.886857 | -0.05846 | 0.974512 |
| MYO6 | 8.088832 | 0.055161 | 0.999532 | | 10.18227 | 0.28144 | 0.693141 | | 8.477318 | 0.243355 | 0.790999 |
| FAM229B | 2.966124 | 0.402127 | 0.999532 | | 2.392973 | -0.54282 | 0.693141 | | 2.48762 | -0.92438 | 0.446959 |
| TMEM218 | 3.648342 | -0.12289 | 0.999532 | | 5.100613 | 0.379266 | 0.693141 | | 4.278979 | 0.523025 | 0.619939 |
| ARHGAP | 5.199722 | 0.21486 | 0.999532 | | 6.158899 | 0.343589 | 0.693141 | | 5.805608 | 0.171521 | 0.892869 |
| PSMD11 | 4.848893 | -0.0907 | 0.999532 | | 4.8356 | -0.40087 | 0.693141 | | 3.958556 | -0.28574 | 0.808457 |
| CLRN2 | 2.594542 | 0.148028 | 0.999532 | | 2.241469 | -0.54054 | 0.693141 | | 2.218802 | -0.67693 | 0.646821 |
| H2AC13 | 3.328541 | -0.21515 | 0.999532 | | 3.666788 | -0.46914 | 0.693141 | | 2.879186 | -0.23632 | 0.887389 |
| LRP2BP | 2.512379 | -0.07795 | 0.999532 | | 2.420959 | -0.52766 | 0.693141 | | 1.978528 | -0.43138 | 0.791576 |
| FH | 4.055953 | 0.162527 | 0.999532 | | 3.720494 | -0.41432 | 0.693141 | | 3.309095 | -0.55913 | 0.605842 |
| HACD2 | 2.698687 | 0.523872 | 0.999532 | | 2.925528 | 0.465774 | 0.693141 | | 3.041815 | -0.03912 | 0.987491 |
| MANSC4 | 2.279713 | 0.033723 | 0.999532 | | 2.198193 | -0.59215 | 0.693141 | | 1.87131 | -0.60627 | 0.717138 |
| C9orf78 | 3.266202 | 0.276614 | 0.999532 | | 3.746847 | 0.426697 | 0.693141 | | 3.397767 | 0.171113 | 0.905952 |
| NAAA | 2.569122 | 0.035358 | 0.999532 | | 3.179141 | 0.454996 | 0.693141 | | 2.866186 | 0.435482 | 0.738631 |
| LOXL1 | 6.249288 | -0.40876 | 0.999532 | | 8.951287 | 0.334442 | 0.693141 | | 6.765221 | 0.765002 | 0.266933 |
| PITPNM3 | 2.317 | 0.174511 | 0.999532 | | 2.745456 | 0.493661 | 0.693141 | | 2.592682 | 0.331011 | 0.847148 |
| SLC14A1 | 3.158593 | -0.28446 | 0.999532 | | 3.454583 | -0.47695 | 0.693141 | | 2.576509 | -0.17158 | 0.925523 |
| KDM6A | 4.63098 | 0.156978 | 0.999532 | | 4.106019 | -0.40861 | 0.693141 | | 3.84211 | -0.55399 | 0.617899 |
| TMLHE | 3.242803 | 0.021789 | 0.999532 | | 3.177767 | -0.46368 | 0.693141 | | 2.680951 | -0.46466 | 0.722703 |
| SOX11 | 2.456106 | 0.050384 | 0.999532 | | 2.353046 | -0.55321 | 0.693141 | | 2.077151 | -0.58238 | 0.712621 |
| PCBD2 | 1.996171 | 0.22889 | 0.999532 | | 2.487993 | 0.515465 | 0.693141 | | 2.217463 | 0.303586 | 0.858767 |
| CUL2 | 3.602313 | 0.296449 | 0.999532 | | 4.053671 | 0.411507 | 0.693141 | | 3.880013 | 0.123629 | 0.939549 |
| USHBP1 | 2.895935 | -0.31877 | 0.999532 | | 3.247829 | -0.48038 | 0.693141 | | 2.449128 | -0.1435 | 0.941197 |
| HPF1 | 2.896344 | 0.267905 | 0.999532 | | 3.495294 | 0.449224 | 0.693141 | | 3.32065 | 0.200733 | 0.897096 |
| XPC | 4.089511 | -0.09252 | 0.999532 | | 4.168122 | -0.39494 | 0.693141 | | 3.542933 | -0.28346 | 0.824012 |
| EMB | 4.469105 | -0.14551 | 0.999532 | | 4.773676 | -0.38495 | 0.693141 | | 3.694904 | -0.21985 | 0.872493 |
| SMIM3 | 2.256123 | -0.20146 | 0.999532 | | 2.222476 | -0.55475 | 0.693141 | | 1.905728 | -0.34333 | 0.862823 |
| DONSON | 1.795871 | -0.04568 | 0.999532 | | 2.477919 | 0.511927 | 0.693141 | | 2.164673 | 0.58054 | 0.699971 |
| OPRL1 | 2.208881 | 0.006168 | 0.999532 | | 2.102351 | -0.56651 | 0.693141 | | 1.921532 | -0.55857 | 0.724411 |
| TMTC2 | 2.68096 | -0.19529 | 0.999532 | | 2.642042 | -0.48684 | 0.693141 | | 2.203916 | -0.27735 | 0.872493 |
| SUCNR1 | 2.293675 | 0.34213 | 0.999532 | | 2.90086 | 0.511933 | 0.693141 | | 2.701332 | 0.19209 | 0.911296 |
| AZIN1 | 6.605127 | 0.268673 | 0.999532 | | 7.310369 | 0.32246 | 0.693141 | | 6.722655 | 0.064661 | 0.963866 |
| SRD5A1 | 6.139457 | -0.13071 | 0.999532 | | 6.354008 | -0.3993 | 0.693317 | | 5.289485 | -0.25352 | 0.855083 |
| POFUT2 | 4.591647 | 0.087588 | 0.999532 | | 5.19581 | 0.39657 | 0.693317 | | 4.76988 | 0.319482 | 0.800239 |
| SIGLEC1 | 2.213584 | -0.58333 | 0.999532 | | 2.288478 | -0.53251 | 0.693484 | | 1.746432 | 0.070318 | 0.979282 |
| KLHL20 | 3.353857 | 0.09285 | 0.999532 | | 4.412774 | 0.40078 | 0.693484 | | 3.95351 | 0.324264 | 0.791576 |
| AMT | 2.450048 | -0.05028 | 0.999532 | | 2.402432 | -0.53587 | 0.69374 | | 2.183816 | -0.45878 | 0.784779 |
| ACTL8 | 2.128666 | 0.304404 | 0.999532 | | 1.902264 | -0.58883 | 0.693849 | | 1.820605 | -0.87723 | 0.531575 |
| PUSL1 | 3.263831 | -0.66355 | 0.999532 | | 3.675274 | -0.42683 | 0.694414 | | 2.61176 | 0.243412 | 0.886495 |
| MLIP | 2.618462 | -0.33378 | 0.999532 | | 2.939431 | -0.48793 | 0.694414 | | 2.094925 | -0.12953 | 0.950616 |
| SIDT1 | 2.082506 | -0.19477 | 0.999532 | | 2.168546 | -0.60148 | 0.694414 | | 1.761155 | -0.39056 | 0.846786 |
| EFHD1 | 10.67474 | 0.00965 | 0.999532 | | 13.22099 | 0.26295 | 0.694414 | | 10.9081 | 0.263325 | 0.784603 |
| PATJ | 4.660194 | 0.107317 | 0.999532 | | 5.330257 | 0.363732 | 0.694587 | | 4.690302 | 0.231697 | 0.870694 |
| DCTD | 3.738287 | -0.07502 | 0.999532 | | 4.912636 | 0.392528 | 0.694882 | | 3.999178 | 0.487427 | 0.653409 |
| DSG2 | 3.368383 | 0.501008 | 0.999532 | | 3.687492 | 0.432438 | 0.694882 | | 3.501024 | -0.04395 | 0.983211 |
| DNMBP | 2.34659 | 0.878543 | 0.999532 | | 2.471891 | 0.547471 | 0.694882 | | 2.748791 | -0.30796 | 0.849608 |
| RESF1 | 6.768332 | -0.30926 | 0.999532 | | 9.119303 | 0.317726 | 0.694882 | | 7.258359 | 0.647004 | 0.421517 |
| ADPRH | 3.705751 | -0.32733 | 0.999532 | | 3.99384 | -0.41307 | 0.694882 | | 3.073754 | -0.07059 | 0.972221 |
| CBR4 | 4.626153 | 0.148902 | 0.999532 | | 4.379163 | -0.41129 | 0.694882 | | 4.009661 | -0.54481 | 0.626811 |
| C21orf62 | 2.644056 | -0.37535 | 0.999532 | | 2.810001 | -0.49731 | 0.694882 | | 2.146255 | -0.10206 | 0.966316 |
| TIGD2 | 2.219425 | -0.07469 | 0.999532 | | 2.101424 | -0.57125 | 0.694882 | | 1.734331 | -0.47997 | 0.789312 |
| SYMPK | 5.783075 | 0.246218 | 0.999532 | | 6.927695 | 0.314824 | 0.694953 | | 6.212937 | 0.094656 | 0.943229 |
| SLC16A8 | 3.282992 | -0.01103 | 0.999532 | | 3.278345 | -0.45748 | 0.694953 | | 2.875192 | -0.42487 | 0.758977 |
| PLEKHO2 | 3.168883 | -0.19751 | 0.999532 | | 4.36712 | 0.391255 | 0.694953 | | 3.573644 | 0.607019 | 0.566812 |
| ATP6V0D | 6.020205 | -0.07412 | 0.999532 | | 7.930494 | 0.311246 | 0.694953 | | 6.374461 | 0.406985 | 0.641721 |
| SAR1B | 3.251894 | -0.14179 | 0.999532 | | 4.661808 | 0.39949 | 0.694953 | | 3.935955 | 0.557183 | 0.589729 |
| EPHX4 | 2.234544 | 0.223238 | 0.999532 | | 1.98983 | -0.59952 | 0.695208 | | 1.917368 | -0.8053 | 0.612864 |
| GAS2L1 | 5.614376 | -0.31927 | 0.999532 | | 6.198118 | -0.35672 | 0.695208 | | 4.707882 | -0.01943 | 0.993618 |
| OLFML3 | 8.319347 | -0.1652 | 0.999532 | | 8.886536 | -0.30766 | 0.695583 | | 7.235863 | -0.12282 | 0.914929 |
| POSTN | 19.42215 | -0.31341 | 0.999532 | | 28.57565 | 0.441781 | 0.696215 | | 22.4258 | 0.775276 | 0.282041 |
| DEPDC5 | 2.110776 | 0.437777 | 0.999532 | | 2.493514 | 0.53087 | 0.696215 | | 2.507434 | 0.105335 | 0.96129 |
| ALDH3B1 | 4.460758 | -0.16865 | 0.999532 | | 5.844929 | 0.357189 | 0.696215 | | 4.720949 | 0.543658 | 0.563455 |
| SNRPA1 | 6.368296 | 0.050475 | 0.999532 | | 5.898867 | -0.3367 | 0.696446 | | 5.432751 | -0.38041 | 0.703463 |
| ATXN3 | 4.845392 | -0.2579 | 0.999532 | | 6.525837 | 0.353079 | 0.696446 | | 5.087575 | 0.638517 | 0.448851 |
| CDHR2 | 2.109642 | -0.525 | 0.999532 | | 2.290404 | -0.54187 | 0.696512 | | 1.653271 | -0.00024 | 0.999949 |
| CACFD1 | 3.054751 | 0.580897 | 0.999532 | | 3.503257 | 0.473656 | 0.69684 | | 3.499467 | -0.09671 | 0.956322 |
| LMLN2 | 2.321684 | -0.11096 | 0.999532 | | 2.298538 | -0.57888 | 0.697501 | | 1.909409 | -0.45495 | 0.79686 |
| NMT1 | 4.139261 | 0.121724 | 0.999532 | | 5.444183 | 0.38633 | 0.697501 | | 4.708835 | 0.286041 | 0.809485 |
| METTL5 | 4.606152 | 0.051393 | 0.999532 | | 5.708764 | 0.356758 | 0.697542 | | 4.829187 | 0.31704 | 0.764702 |
| PCGF3 | 4.482524 | 0.229332 | 0.999532 | | 5.12782 | 0.369262 | 0.697764 | | 4.561572 | 0.161662 | 0.900917 |
| LEFTY2 | 2.551195 | 0.016956 | 0.999532 | | 2.519185 | -0.53337 | 0.697867 | | 2.172987 | -0.53117 | 0.723933 |
| FAM50A | 5.329162 | 0.045484 | 0.999532 | | 6.758369 | 0.344812 | 0.698023 | | 5.674143 | 0.31799 | 0.757035 |
| TBC1D8 | 4.725389 | -0.64672 | 0.999532 | | 5.464256 | -0.36597 | 0.698271 | | 3.767708 | 0.28204 | 0.835289 |
| TSPAN14 | 4.683075 | -0.1033 | 0.999532 | | 5.876251 | 0.349051 | 0.698345 | | 5.061082 | 0.470736 | 0.632113 |
| C20orf20 | 2.535865 | -0.1095 | 0.999532 | | 2.462939 | -0.52997 | 0.698345 | | 2.059806 | -0.40737 | 0.816589 |
| PTGES | 3.43052 | 0.261461 | 0.999532 | | 2.853935 | -0.483 | 0.698345 | | 2.899915 | -0.73243 | 0.536289 |
| STX12 | 4.943801 | -0.03291 | 0.999532 | | 6.424495 | 0.329983 | 0.698626 | | 5.412166 | 0.382594 | 0.688542 |
| C3orf38 | 4.254499 | 0.10009 | 0.999532 | | 5.250203 | 0.37099 | 0.69863 | | 4.66345 | 0.290161 | 0.801905 |
| LAPTM4B | 5.837109 | -0.2201 | 0.999532 | | 8.211602 | 0.352729 | 0.69863 | | 6.412463 | 0.604567 | 0.540342 |
| BZW1 | 5.32586 | 0.153307 | 0.999532 | | 6.196431 | 0.362999 | 0.69863 | | 5.592908 | 0.22636 | 0.849128 |
| FUBP1 | 11.43949 | 0.306256 | 0.999532 | | 12.57596 | 0.30102 | 0.699117 | | 11.97931 | 0.014053 | 0.993618 |
| ADAM17 | 4.06529 | 0.125478 | 0.999532 | | 4.796137 | 0.3711 | 0.699117 | | 4.217381 | 0.257536 | 0.836935 |
| CKS2 | 5.084475 | -0.16696 | 0.999532 | | 5.144728 | -0.41372 | 0.699154 | | 4.138832 | -0.22594 | 0.877703 |
| CAMK2B | 2.528224 | 0.780839 | 0.999532 | | 1.700694 | -0.62314 | 0.699219 | | 2.131129 | -1.38181 | 0.265037 |
| TJP3 | 2.777828 | -0.29195 | 0.999532 | | 3.217854 | -0.49923 | 0.69943 | | 2.329732 | -0.16045 | 0.939724 |
| ZNF454 | 2.214369 | -0.30695 | 0.999532 | | 2.377262 | -0.5192 | 0.69943 | | 1.941841 | -0.19168 | 0.926702 |
| KCND3 | 3.306863 | -0.15607 | 0.999532 | | 3.368619 | -0.44391 | 0.699434 | | 2.800912 | -0.27157 | 0.863808 |
| BARX2 | 3.350102 | 0.215298 | 0.999532 | | 3.099228 | -0.47175 | 0.699883 | | 2.828432 | -0.66672 | 0.570037 |
| ZNF669 | 3.99143 | -0.44772 | 0.999532 | | 4.501931 | -0.40223 | 0.699883 | | 3.274195 | 0.061037 | 0.976441 |
| MRPL13 | 3.396449 | 0.347357 | 0.999532 | | 3.740172 | 0.416209 | 0.699883 | | 3.634248 | 0.093393 | 0.954616 |
| GATAD1 | 5.829887 | 0.206712 | 0.999532 | | 7.126712 | 0.315832 | 0.699883 | | 6.379359 | 0.131589 | 0.906811 |
| PCNA | 4.957892 | -0.05746 | 0.999532 | | 6.659111 | 0.319489 | 0.699904 | | 5.463065 | 0.396128 | 0.663774 |
| VENTX | 2.711609 | 0.157383 | 0.999532 | | 2.329319 | -0.54725 | 0.700642 | | 2.224115 | -0.68796 | 0.644433 |
| GTF3C6 | 6.642156 | -0.10499 | 0.999532 | | 8.813233 | 0.300276 | 0.700642 | | 7.048359 | 0.428341 | 0.600254 |
| DUOX1 | 4.0022 | 0.047003 | 0.999532 | | 3.826316 | -0.4642 | 0.700663 | | 3.341338 | -0.494 | 0.682485 |
| FBRSL1 | 3.048415 | -0.11547 | 0.999532 | | 3.12344 | -0.46368 | 0.700686 | | 2.506938 | -0.32953 | 0.825415 |
| MRPS24 | 2.926058 | -0.02992 | 0.999532 | | 3.833988 | 0.412665 | 0.700713 | | 3.067989 | 0.45651 | 0.702463 |
| TMEM45A | 2.758909 | 0.101326 | 0.999532 | | 3.314398 | 0.468714 | 0.700713 | | 3.009154 | 0.3812 | 0.791576 |
| ZNF22 | 3.301638 | 0.310457 | 0.999532 | | 3.976399 | 0.416469 | 0.70085 | | 3.724879 | 0.127215 | 0.938429 |
| FAM186B | 2.391466 | 0.280559 | 0.999532 | | 1.995826 | -0.58142 | 0.701022 | | 2.016773 | -0.84653 | 0.558035 |
| NRBP1 | 8.253235 | 0.284894 | 0.999532 | | 8.945247 | 0.310765 | 0.701022 | | 8.476228 | 0.023093 | 0.991159 |
| ATP5F1D | 7.198876 | -0.03923 | 0.999532 | | 9.248277 | 0.288866 | 0.701065 | | 7.305048 | 0.345323 | 0.717627 |
| TRAPPC8 | 2.674493 | -0.37317 | 0.999532 | | 3.821339 | 0.407265 | 0.701842 | | 3.047012 | 0.798673 | 0.456688 |
| PEBP4 | 2.616489 | -0.11067 | 0.999532 | | 2.591777 | -0.50677 | 0.701955 | | 2.197055 | -0.37645 | 0.82512 |
| NAA10 | 5.023727 | 0.06992 | 0.999532 | | 6.049865 | 0.342073 | 0.701955 | | 5.667869 | 0.321381 | 0.773057 |
| HNRNPF | 7.390792 | -0.25945 | 0.999532 | | 9.839593 | 0.282805 | 0.701955 | | 7.906934 | 0.571222 | 0.385491 |
| CXorf51B | 2.31536 | -0.25995 | 0.999532 | | 2.402188 | -0.54017 | 0.701955 | | 1.885103 | -0.26089 | 0.894612 |
| DNAJA1 | 11.12 | -0.11999 | 0.999532 | | 14.28965 | 0.278117 | 0.701955 | | 11.87848 | 0.41131 | 0.552672 |
| ZNF791 | 3.51327 | 0.216701 | 0.999532 | | 3.008071 | -0.50392 | 0.702367 | | 2.814483 | -0.68751 | 0.554813 |
| EIF2B4 | 5.831083 | -0.03217 | 0.999532 | | 7.295921 | 0.31235 | 0.703233 | | 6.195989 | 0.358029 | 0.703463 |
| SEC11A | 7.12321 | 0.169637 | 0.999532 | | 8.051693 | 0.309049 | 0.703716 | | 7.443439 | 0.14096 | 0.892869 |
| ANXA7 | 7.261353 | -0.4164 | 0.999532 | | 10.02986 | 0.290074 | 0.703716 | | 7.687164 | 0.727229 | 0.241605 |
| OSTM1 | 5.524426 | -0.59587 | 0.999532 | | 6.842094 | -0.33481 | 0.703716 | | 4.637243 | 0.288428 | 0.806394 |
| HOXB8 | 3.33073 | 0.057941 | 0.999532 | | 3.251452 | -0.47887 | 0.703716 | | 2.619735 | -0.50615 | 0.699971 |
| EEF1AKM | 4.557715 | 0.194073 | 0.999532 | | 5.097671 | 0.371787 | 0.703716 | | 4.63016 | 0.19973 | 0.874341 |
| SRSF6 | 6.64786 | 0.063103 | 0.999532 | | 7.959749 | 0.31945 | 0.704135 | | 6.989862 | 0.267625 | 0.791466 |
| ZNF787 | 4.629323 | 0.011616 | 0.999532 | | 4.589478 | -0.39676 | 0.704241 | | 3.893467 | -0.38903 | 0.732916 |
| PEX5 | 3.326934 | 0.256306 | 0.999532 | | 3.975594 | 0.421162 | 0.704241 | | 3.609169 | 0.18434 | 0.900848 |
| WBP2 | 5.085457 | -0.31316 | 0.999532 | | 7.223061 | 0.366604 | 0.704241 | | 5.558632 | 0.703866 | 0.436389 |
| DKKL1 | 2.904052 | 0.132864 | 0.999532 | | 2.554346 | -0.51836 | 0.704371 | | 2.462952 | -0.64409 | 0.663413 |
| OR5A1 | 3.012634 | 0.349847 | 0.999532 | | 2.577807 | -0.52048 | 0.704371 | | 2.467018 | -0.84898 | 0.474973 |
| CAMK2A | 3.973167 | 0.269128 | 0.999532 | | 3.515193 | -0.43479 | 0.704371 | | 3.373863 | -0.68462 | 0.536289 |
| AHR | 4.912212 | -0.31966 | 0.999532 | | 7.003024 | 0.354514 | 0.704511 | | 5.48567 | 0.693405 | 0.386771 |
| RPUSD4 | 2.693551 | -0.24404 | 0.999532 | | 2.881556 | -0.4893 | 0.704511 | | 2.268645 | -0.22114 | 0.904476 |
| INSYN1 | 4.709041 | 0.245475 | 0.999532 | | 5.593563 | 0.379418 | 0.704511 | | 4.971902 | 0.165607 | 0.905332 |
| ARRDC1 | 5.968947 | 0.249747 | 0.999532 | | 7.07635 | 0.316589 | 0.704511 | | 6.457242 | 0.085143 | 0.945902 |
| TENM1 | 2.00283 | -0.2327 | 0.999532 | | 2.10105 | -0.5657 | 0.704511 | | 1.710077 | -0.3103 | 0.881837 |
| SDF2L1 | 4.200791 | 0.236048 | 0.999532 | | 3.679935 | -0.4246 | 0.704511 | | 3.665793 | -0.65084 | 0.546858 |
| FSTL1 | 14.51866 | 0.114515 | 0.999532 | | 17.71145 | 0.317322 | 0.704586 | | 15.73012 | 0.2181 | 0.816967 |
| SRGN | 11.57952 | -1.06955 | 0.22007 | | 15.82904 | -0.33881 | 0.705207 | | 9.471668 | 0.751334 | 0.241605 |
| SFRP2 | 2.007599 | 0.306418 | 0.999532 | | 2.406566 | 0.510044 | 0.705373 | | 2.284248 | 0.221831 | 0.900376 |
| SRC | 4.442054 | -0.29482 | 0.999532 | | 4.879058 | -0.37484 | 0.705373 | | 3.72619 | -0.06096 | 0.974421 |
| TDRP | 2.387106 | 0.367727 | 0.999532 | | 2.826089 | 0.468494 | 0.705373 | | 2.616178 | 0.123715 | 0.946808 |
| POLR1A | 3.319583 | 0.355076 | 0.999532 | | 4.032125 | 0.407316 | 0.70544 | | 3.764003 | 0.070891 | 0.969614 |
| AIFM1 | 3.970154 | -0.10508 | 0.999532 | | 5.37104 | 0.358012 | 0.70544 | | 4.243686 | 0.484445 | 0.657538 |
| BICDL2 | 2.402003 | -0.07583 | 0.999532 | | 2.530441 | -0.56753 | 0.705474 | | 1.933786 | -0.47111 | 0.782253 |
| LAIR1 | 2.688073 | 0.181845 | 0.999532 | | 2.657198 | -0.50325 | 0.705474 | | 2.346435 | -0.65808 | 0.632923 |
| THUMPD | 6.55652 | -0.07323 | 0.999532 | | 6.955159 | -0.33494 | 0.706082 | | 5.572553 | -0.23526 | 0.832834 |
| SNRNP4 | 3.736567 | 0.316936 | 0.999532 | | 4.228171 | 0.390958 | 0.706082 | | 4.052525 | 0.087535 | 0.957758 |
| PLAA | 3.630958 | 0.468241 | 0.999532 | | 3.84343 | 0.437184 | 0.706082 | | 4.072968 | -0.02397 | 0.992459 |
| GOLGB1 | 4.018671 | 0.372281 | 0.999532 | | 4.239788 | 0.413132 | 0.706082 | | 4.160232 | 0.057903 | 0.976381 |
| BPIFA3 | 2.257297 | -0.14854 | 0.999532 | | 2.348222 | -0.54833 | 0.706082 | | 1.939247 | -0.37671 | 0.839811 |
| ASIC3 | 3.655369 | -0.32311 | 0.999532 | | 4.129494 | -0.41566 | 0.706082 | | 3.145793 | -0.07084 | 0.973356 |
| C8orf82 | 4.235543 | 0.47624 | 0.999532 | | 4.29645 | 0.383245 | 0.706082 | | 4.498639 | -0.07135 | 0.966549 |
| WDR41 | 4.318422 | 0.420247 | 0.999532 | | 4.56474 | 0.40205 | 0.706082 | | 4.762738 | -0.017 | 0.995304 |
| METTL23 | 3.267248 | -0.45945 | 0.999532 | | 3.404498 | -0.43071 | 0.706082 | | 2.667464 | 0.003879 | 0.999385 |
| GPD2 | 2.449365 | 0.105144 | 0.999532 | | 2.338191 | -0.51194 | 0.706082 | | 2.022212 | -0.59782 | 0.670068 |
| AREG | 2.065597 | 0.112534 | 0.999532 | | 2.945913 | 0.561891 | 0.706082 | | 2.598579 | 0.482531 | 0.777828 |
| TUBAL3 | 2.200296 | -0.17423 | 0.999532 | | 2.159199 | -0.53753 | 0.706082 | | 1.8227 | -0.34733 | 0.858705 |
| SYT2 | 4.605199 | 0.079269 | 0.999532 | | 4.711944 | -0.41633 | 0.706082 | | 4.202824 | -0.44755 | 0.723933 |
| GJD3 | 2.640764 | -0.37656 | 0.999532 | | 2.826058 | -0.46544 | 0.706082 | | 2.058618 | -0.07214 | 0.975633 |
| FER1L6 | 2.452223 | -0.18256 | 0.999532 | | 2.619552 | -0.52403 | 0.706082 | | 2.026516 | -0.3187 | 0.86306 |
| SLC30A9 | 4.920813 | 0.227947 | 0.999532 | | 5.709366 | 0.349672 | 0.706082 | | 5.293244 | 0.140219 | 0.914687 |
| IL17B | 1.929172 | -0.39898 | 0.999532 | | 1.988903 | -0.55452 | 0.706082 | | 1.563017 | -0.13685 | 0.956322 |
| ZNF286A | 2.839421 | -0.17041 | 0.999532 | | 2.811659 | -0.47911 | 0.706082 | | 2.353407 | -0.29408 | 0.864245 |
| GORASP | 5.616209 | 0.188328 | 0.999532 | | 6.616875 | 0.348203 | 0.706082 | | 5.801446 | 0.185267 | 0.877703 |
| OR10A5 | 2.784329 | -0.4193 | 0.999532 | | 3.106922 | -0.52391 | 0.706082 | | 2.355946 | -0.09081 | 0.972758 |
| IL31RA | 2.34045 | 0.050649 | 0.999532 | | 2.132989 | -0.54835 | 0.706082 | | 1.975302 | -0.58211 | 0.714657 |
| CREG2 | 2.998312 | 0.046563 | 0.999532 | | 2.838078 | -0.52734 | 0.706082 | | 2.491067 | -0.55348 | 0.718633 |
| POLK | 3.233416 | -0.20949 | 0.999532 | | 4.443006 | 0.378578 | 0.706082 | | 3.672023 | 0.609383 | 0.563455 |
| CRPPA | 2.977473 | 0.148453 | 0.999532 | | 2.870819 | -0.48078 | 0.706082 | | 2.518252 | -0.61237 | 0.636525 |
| TMEM18 | 2.20256 | -0.09792 | 0.999532 | | 2.274845 | -0.52819 | 0.706082 | | 1.805307 | -0.4076 | 0.813438 |
| HINT2 | 4.952489 | 0.269087 | 0.999532 | | 5.765267 | 0.335815 | 0.706082 | | 5.287987 | 0.084792 | 0.948773 |
| H4C12 | 11.98552 | -0.52269 | 0.999532 | | 17.52515 | 0.344127 | 0.706422 | | 13.21549 | 0.870643 | 0.222058 |
| GNB4 | 3.714472 | 0.231949 | 0.999532 | | 4.419381 | 0.380056 | 0.706422 | | 4.043242 | 0.165854 | 0.901813 |
| SPICE1 | 1.927076 | 0.401649 | 0.999532 | | 2.477246 | 0.518626 | 0.706422 | | 2.256373 | 0.141884 | 0.945129 |
| CASP12 | 3.704854 | -0.16162 | 0.999532 | | 3.905976 | -0.41125 | 0.706422 | | 3.079646 | -0.23147 | 0.877943 |
| RFX2 | 4.133071 | 0.033559 | 0.999532 | | 3.953691 | -0.41917 | 0.706422 | | 3.523239 | -0.43675 | 0.730921 |
| WHAMM | 2.324694 | 0.201294 | 0.999532 | | 2.858784 | 0.46335 | 0.706422 | | 2.64969 | 0.282797 | 0.859426 |
| RNF185 | 4.542767 | -0.22749 | 0.999532 | | 4.768099 | -0.39296 | 0.706422 | | 3.723042 | -0.14546 | 0.928522 |
| IGF1R | 3.664672 | -0.07038 | 0.999532 | | 3.733292 | -0.41505 | 0.706422 | | 2.866985 | -0.32217 | 0.813438 |
| CCDC14 | 2.794884 | -0.23109 | 0.999532 | | 2.787622 | -0.50181 | 0.706422 | | 2.347906 | -0.27694 | 0.88476 |
| THNSL1 | 2.321214 | 0.393367 | 0.999532 | | 1.865415 | -0.59095 | 0.706422 | | 1.867984 | -0.96674 | 0.49299 |
| CHEK2 | 2.764852 | -0.22946 | 0.999532 | | 2.963086 | -0.49013 | 0.706422 | | 2.332433 | -0.24164 | 0.892869 |
| MRPL23 | 3.590961 | -0.59392 | 0.999532 | | 5.31378 | 0.347735 | 0.70644 | | 3.85321 | 0.955159 | 0.254611 |
| HNRNPL | 12.49156 | -0.07645 | 0.999532 | | 15.66546 | 0.252319 | 0.70644 | | 12.95701 | 0.352064 | 0.626811 |
| KBTBD6 | 2.801148 | 0.338727 | 0.999532 | | 3.338944 | 0.449304 | 0.70644 | | 3.331585 | 0.135344 | 0.938429 |
| PYHIN1 | 2.680698 | -0.24397 | 0.999532 | | 2.672603 | -0.49963 | 0.70644 | | 2.319628 | -0.24307 | 0.89901 |
| TIMM21 | 2.546781 | -0.14836 | 0.999532 | | 3.662434 | 0.425963 | 0.70644 | | 3.032324 | 0.598091 | 0.632464 |
| ANGPTL4 | 3.459422 | 0.04395 | 0.999532 | | 3.389686 | -0.45286 | 0.706515 | | 2.994068 | -0.48034 | 0.704363 |
| USP29 | 2.192358 | -0.12207 | 0.999532 | | 2.279713 | -0.5648 | 0.706547 | | 1.907802 | -0.41855 | 0.82268 |
| SLC25A3 | 2.479874 | -0.31537 | 0.999532 | | 2.664019 | -0.48934 | 0.706919 | | 2.028311 | -0.15553 | 0.940114 |
| CD27 | 3.016994 | 0.053107 | 0.999532 | | 2.847682 | -0.47747 | 0.706919 | | 2.603269 | -0.50568 | 0.717627 |
| LGR4 | 2.73135 | 0.172528 | 0.999532 | | 3.341765 | 0.443824 | 0.707613 | | 2.955947 | 0.292223 | 0.835554 |
| ZFPL1 | 2.601198 | 0.357888 | 0.999532 | | 3.031604 | 0.461305 | 0.707822 | | 2.907499 | 0.122037 | 0.946512 |
| MOCS3 | 4.426279 | 0.196612 | 0.999532 | | 3.960782 | -0.44007 | 0.708149 | | 3.721745 | -0.62046 | 0.625947 |
| RLIM | 3.132263 | 0.010463 | 0.999532 | | 3.818065 | 0.422718 | 0.708496 | | 3.228004 | 0.436849 | 0.718633 |
| DGKK | 2.079769 | -0.56929 | 0.999532 | | 2.294066 | -0.52813 | 0.708496 | | 1.647321 | 0.056488 | 0.985873 |
| PGM1 | 2.507803 | 0.434365 | 0.999532 | | 2.857425 | 0.475216 | 0.708496 | | 2.892791 | 0.055855 | 0.978927 |
| TMEM11 | 4.187362 | -0.1523 | 0.999532 | | 5.346002 | 0.364052 | 0.708496 | | 4.334524 | 0.537386 | 0.595157 |
| PJA2 | 8.560249 | -0.15836 | 0.999532 | | 10.6797 | 0.284019 | 0.708496 | | 8.872101 | 0.457091 | 0.536289 |
| SCARB1 | 3.250047 | -0.36815 | 0.999532 | | 3.590179 | -0.44149 | 0.708496 | | 2.727729 | -0.05534 | 0.979967 |
| ARL8A | 15.01963 | 0.15882 | 0.999532 | | 14.716 | -0.30925 | 0.708496 | | 13.25613 | -0.44472 | 0.572297 |
| GNA13 | 7.432837 | 0.300587 | 0.999532 | | 8.001913 | 0.303233 | 0.708496 | | 7.809339 | 0.023299 | 0.990309 |
| POLI | 4.090779 | 0.278707 | 0.999532 | | 4.751656 | 0.372383 | 0.708496 | | 4.354432 | 0.111839 | 0.938429 |
| RRNAD1 | 3.488083 | -0.26827 | 0.999532 | | 3.750387 | -0.40865 | 0.708544 | | 2.786823 | -0.12361 | 0.944635 |
| KDSR | 6.358216 | 0.229799 | 0.999532 | | 7.513803 | 0.320201 | 0.708544 | | 6.62714 | 0.111353 | 0.923864 |
| COL5A3 | 2.208919 | -0.28883 | 0.999532 | | 2.312131 | -0.5096 | 0.708544 | | 1.908062 | -0.20403 | 0.918278 |
| METTL27 | 2.816565 | 0.089477 | 0.999532 | | 3.511225 | 0.419101 | 0.708544 | | 3.202105 | 0.350383 | 0.791576 |
| COQ10B | 2.826839 | 0.191259 | 0.999532 | | 3.175071 | 0.453437 | 0.708544 | | 3.214698 | 0.275271 | 0.871946 |
| TNRC6B | 3.316097 | 0.243559 | 0.999532 | | 4.014178 | 0.386744 | 0.708544 | | 3.663486 | 0.161462 | 0.908829 |
| YWHAH | 7.141944 | -0.4287 | 0.999532 | | 10.3364 | 0.324392 | 0.708673 | | 7.826723 | 0.77661 | 0.332351 |
| NECAB2 | 4.355243 | 0.054049 | 0.999532 | | 4.202094 | -0.40436 | 0.709274 | | 3.824864 | -0.44082 | 0.709825 |
| CCL19 | 2.870168 | 0.317663 | 0.999532 | | 3.262546 | 0.496679 | 0.709394 | | 3.197268 | 0.147109 | 0.947322 |
| NEDD9 | 5.047289 | -0.55652 | 0.999532 | | 5.870407 | -0.35108 | 0.709639 | | 3.874307 | 0.222056 | 0.865384 |
| LRPAP1 | 9.783456 | 0.474382 | 0.999532 | | 10.16997 | 0.279757 | 0.709639 | | 10.17545 | -0.17528 | 0.843564 |
| DOK1 | 3.520661 | -0.56791 | 0.999532 | | 4.190232 | -0.38481 | 0.709639 | | 2.931248 | 0.201633 | 0.895609 |
| CHMP4B | 5.830398 | 0.194883 | 0.999532 | | 6.793347 | 0.374704 | 0.709639 | | 6.176929 | 0.199143 | 0.876781 |
| TMEM13 | 4.311883 | 0.068612 | 0.999532 | | 5.510145 | 0.351783 | 0.709639 | | 4.615629 | 0.291395 | 0.806394 |
| ORC5 | 3.150862 | 0.062882 | 0.999532 | | 3.157688 | -0.47832 | 0.709718 | | 2.659771 | -0.51807 | 0.707745 |
| CENPA | 2.122836 | 0.093558 | 0.999532 | | 1.867895 | -0.58352 | 0.709854 | | 1.8683 | -0.66 | 0.698562 |
| PRKN | 2.255067 | -0.23489 | 0.999532 | | 2.198633 | -0.55703 | 0.709854 | | 1.855961 | -0.29965 | 0.883588 |
| HPRT1 | 2.773485 | -0.36754 | 0.999532 | | 3.079046 | -0.44714 | 0.709854 | | 2.216948 | -0.0621 | 0.97843 |
| KAT8 | 2.99431 | -0.43964 | 0.999532 | | 3.373459 | -0.43746 | 0.709854 | | 2.391535 | 0.018113 | 0.996681 |
| PRIMPOL | 4.069733 | 0.27952 | 0.999532 | | 5.064642 | 0.382685 | 0.709854 | | 4.483094 | 0.121783 | 0.932589 |
| MRPL41 | 6.730342 | 0.550841 | 0.999532 | | 6.806348 | 0.333527 | 0.709854 | | 6.810914 | -0.18983 | 0.856999 |
| GIMAP4 | 6.180002 | -0.34159 | 0.999532 | | 8.844231 | 0.320285 | 0.710059 | | 7.020796 | 0.682919 | 0.336968 |
| DIABLO | 6.408929 | -0.26339 | 0.999532 | | 7.190627 | -0.35672 | 0.710059 | | 5.279057 | -0.07072 | 0.964445 |
| UBA5 | 3.706298 | 0.109328 | 0.999532 | | 4.676932 | 0.388698 | 0.710059 | | 3.950889 | 0.277062 | 0.816618 |
| BRI3BP | 2.01347 | 0.509532 | 0.999532 | | 2.261759 | 0.527487 | 0.710059 | | 2.422507 | 0.040269 | 0.989638 |
| WNK1 | 9.306281 | 0.007271 | 0.999532 | | 11.70154 | 0.276671 | 0.710059 | | 9.539104 | 0.246267 | 0.774044 |
| OR7A10 | 2.988723 | -0.37117 | 0.999532 | | 3.244646 | -0.52591 | 0.710189 | | 2.347758 | -0.12514 | 0.954538 |
| UBE2G1 | 3.86414 | -0.22472 | 0.999532 | | 5.355535 | 0.358114 | 0.710189 | | 4.304121 | 0.600697 | 0.540342 |
| CCN2 | 80.89506 | -0.03843 | 0.999532 | | 103.5932 | 0.310672 | 0.710258 | | 86.4536 | 0.377496 | 0.629006 |
| HPS4 | 4.841646 | 0.460788 | 0.999532 | | 5.404779 | 0.359797 | 0.710366 | | 5.393237 | -0.08071 | 0.956011 |
| MERTK | 3.592943 | 0.013822 | 0.999532 | | 3.677806 | -0.41556 | 0.710393 | | 3.005516 | -0.41132 | 0.737331 |
| OR3A2 | 2.207073 | 0.126735 | 0.999532 | | 1.906382 | -0.56608 | 0.710393 | | 1.855778 | -0.67853 | 0.672584 |
| SELENO | 3.633488 | 0.176915 | 0.999532 | | 3.362032 | -0.42873 | 0.710393 | | 2.980831 | -0.58414 | 0.625476 |
| RPL11 | 16.75322 | -0.16459 | 0.999532 | | 21.72864 | 0.235263 | 0.710393 | | 17.31894 | 0.426388 | 0.515852 |
| ULK2 | 2.164684 | 0.28536 | 0.999532 | | 2.511618 | 0.484468 | 0.710393 | | 2.338649 | 0.213392 | 0.902239 |
| AKIP1 | 4.204935 | 0.452324 | 0.999532 | | 4.19821 | 0.383205 | 0.710393 | | 4.400257 | -0.06706 | 0.972031 |
| MPHOSP | 3.861691 | -0.28499 | 0.999532 | | 4.22027 | -0.38523 | 0.710393 | | 3.187509 | -0.08379 | 0.964621 |
| TJAP1 | 3.501619 | -0.04661 | 0.999532 | | 3.573079 | -0.44252 | 0.710494 | | 2.939577 | -0.37544 | 0.789815 |
| PDLIM4 | 3.151104 | 0.139141 | 0.999532 | | 3.627529 | 0.413663 | 0.710494 | | 3.258001 | 0.283671 | 0.843564 |
| NDUFB9 | 12.2136 | -0.01929 | 0.999532 | | 15.06448 | 0.263557 | 0.710494 | | 12.43201 | 0.299941 | 0.66232 |
| TRA2B | 7.737778 | -0.03608 | 0.999532 | | 9.408386 | 0.300821 | 0.710494 | | 7.922643 | 0.348409 | 0.695903 |
| ESYT2 | 6.744436 | -0.1494 | 0.999532 | | 8.505379 | 0.313237 | 0.710494 | | 7.08157 | 0.48228 | 0.535662 |
| VPS36 | 4.746604 | -0.12476 | 0.999532 | | 6.588495 | 0.319175 | 0.710494 | | 5.286203 | 0.465523 | 0.621895 |
| HMGB4 | 1.907546 | -0.44993 | 0.999532 | | 2.15486 | -0.55641 | 0.710494 | | 1.670762 | -0.08607 | 0.976603 |
| ZNF778 | 2.458985 | -0.143 | 0.999532 | | 2.348711 | -0.52522 | 0.710494 | | 2.006806 | -0.37074 | 0.842686 |
| SRCIN1 | 2.386858 | -0.18329 | 0.999532 | | 2.392282 | -0.5241 | 0.710494 | | 2.010577 | -0.32361 | 0.863134 |
| MAGEC3 | 2.647267 | 0.252871 | 0.999532 | | 2.338943 | -0.51486 | 0.710494 | | 2.281694 | -0.74967 | 0.566812 |
| SCRN3 | 3.132643 | 0.738463 | 0.999532 | | 2.992364 | 0.479533 | 0.710494 | | 3.444183 | -0.24267 | 0.874653 |
| FCER2 | 2.527537 | -0.11864 | 0.999532 | | 2.550002 | -0.50868 | 0.710494 | | 2.090849 | -0.37192 | 0.824012 |
| ICAM1 | 6.003088 | -0.51286 | 0.999532 | | 6.773144 | -0.43264 | 0.710651 | | 4.825993 | 0.113081 | 0.944202 |
| MAPK3 | 4.252548 | -0.35279 | 0.999532 | | 5.856605 | 0.359753 | 0.710729 | | 4.695509 | 0.73122 | 0.414073 |
| KCNMB2 | 2.493254 | 0.15836 | 0.999532 | | 2.26429 | -0.55031 | 0.71081 | | 1.994128 | -0.68965 | 0.652925 |
| CYP1A1 | 1.860919 | 0.645049 | 0.999532 | | 2.061472 | 0.566819 | 0.711075 | | 2.206164 | -0.05768 | 0.984587 |
| ERBB4 | 1.91873 | 0.719725 | 0.999532 | | 1.95953 | 0.575411 | 0.711075 | | 2.093226 | -0.12543 | 0.955834 |
| PTGDR | 2.228147 | -0.27152 | 0.999532 | | 2.430595 | -0.51642 | 0.711075 | | 1.839222 | -0.21704 | 0.914929 |
| COL4A3 | 8.629639 | 0.322741 | 0.999532 | | 9.507994 | 0.317446 | 0.711075 | | 8.873576 | 0.024155 | 0.988853 |
| WDR91 | 5.730542 | -0.07296 | 0.999532 | | 5.957986 | -0.34606 | 0.711075 | | 4.720926 | -0.24785 | 0.827267 |
| CDR1 | 2.250952 | -0.03724 | 0.999532 | | 2.157877 | -0.53567 | 0.71137 | | 1.85807 | -0.47617 | 0.78258 |
| XRCC1 | 2.908202 | -0.10838 | 0.999532 | | 3.858529 | 0.404942 | 0.71137 | | 3.222579 | 0.529362 | 0.659727 |
| OGDH | 3.264155 | 0.09308 | 0.999532 | | 4.023722 | 0.393781 | 0.711376 | | 3.660761 | 0.316878 | 0.806394 |
| UQCR11 | 12.88237 | -0.15167 | 0.999532 | | 16.88623 | 0.298683 | 0.711376 | | 13.64009 | 0.478364 | 0.515762 |
| HEBP2 | 3.603188 | 0.536557 | 0.999532 | | 3.748315 | 0.422274 | 0.711461 | | 3.800242 | -0.09261 | 0.955167 |
| LYRM7 | 2.207592 | 0.155071 | 0.999532 | | 1.965606 | -0.58226 | 0.711461 | | 1.75542 | -0.72542 | 0.660958 |
| LITAFD | 2.083241 | -0.10606 | 0.999532 | | 1.983287 | -0.56431 | 0.711461 | | 1.784812 | -0.44421 | 0.814884 |
| TIPIN | 5.274482 | 0.114707 | 0.999532 | | 4.854951 | -0.37614 | 0.711782 | | 4.44614 | -0.48809 | 0.673405 |
| STUB1 | 2.887189 | -0.06344 | 0.999532 | | 3.829382 | 0.399929 | 0.711923 | | 3.119652 | 0.481158 | 0.682485 |
| MCTS1 | 4.351819 | 0.335575 | 0.999532 | | 4.851359 | 0.369045 | 0.711923 | | 4.617928 | 0.054392 | 0.974421 |
| TM2D1 | 5.691373 | -0.00676 | 0.999532 | | 7.070432 | 0.344431 | 0.711928 | | 6.088269 | 0.37624 | 0.692534 |
| FXR1 | 10.89006 | 0.160136 | 0.999532 | | 12.39767 | 0.25774 | 0.711928 | | 11.35047 | 0.123749 | 0.901033 |
| LIMD2 | 3.783401 | -0.61917 | 0.999532 | | 4.443647 | -0.38013 | 0.712224 | | 3.246926 | 0.257969 | 0.864245 |
| TCF7L2 | 5.181606 | 0.374277 | 0.999532 | | 5.605214 | 0.368277 | 0.712353 | | 5.414639 | 0.019079 | 0.993396 |
| KIF7 | 3.979726 | -0.25998 | 0.999532 | | 4.062613 | -0.42027 | 0.712353 | | 3.279543 | -0.13883 | 0.939549 |
| TP53I13 | 3.033707 | 0.327546 | 0.999532 | | 2.61137 | -0.53821 | 0.712445 | | 2.478968 | -0.84626 | 0.49299 |
| STK24 | 5.362355 | -0.04301 | 0.999532 | | 7.006949 | 0.331029 | 0.712445 | | 5.90742 | 0.393386 | 0.679452 |
| HAS3 | 2.922681 | 0.07264 | 0.999532 | | 2.634476 | -0.52035 | 0.712445 | | 2.577049 | -0.5765 | 0.709825 |
| H4C8 | 9.203742 | -0.23283 | 0.999532 | | 12.06739 | 0.242686 | 0.712445 | | 9.611769 | 0.491833 | 0.451915 |
| IRF7 | 3.550808 | -0.05057 | 0.999532 | | 4.529764 | 0.373837 | 0.712445 | | 3.726876 | 0.436831 | 0.704316 |
| LRCH1 | 7.434191 | -0.01143 | 0.999532 | | 9.308665 | 0.320265 | 0.712685 | | 8.016793 | 0.338669 | 0.717627 |
| PPM1A | 5.239418 | 0.049236 | 0.999532 | | 6.473457 | 0.339341 | 0.712782 | | 5.777085 | 0.314174 | 0.749206 |
| MAGEA9 | 2.823187 | 0.240707 | 0.999532 | | 2.468658 | -0.51246 | 0.713017 | | 2.446044 | -0.7393 | 0.590442 |
| CEACAM | 2.513426 | -0.01251 | 0.999532 | | 2.371523 | -0.53158 | 0.713156 | | 2.032642 | -0.50022 | 0.752582 |
| CBR3 | 2.717159 | 0.296902 | 0.999532 | | 2.303516 | -0.51096 | 0.713411 | | 2.349689 | -0.79104 | 0.542124 |
| ZNRF2 | 4.159374 | 0.236334 | 0.999532 | | 4.984833 | 0.394882 | 0.713579 | | 4.287405 | 0.173716 | 0.906168 |
| ASH2L | 3.785134 | 0.033128 | 0.999532 | | 3.68228 | -0.43239 | 0.713579 | | 3.002906 | -0.44199 | 0.720106 |
| VEZT | 2.990164 | 0.345169 | 0.999532 | | 3.445557 | 0.423049 | 0.713579 | | 3.415794 | 0.101271 | 0.955834 |
| NME1 | 3.138243 | 0.124278 | 0.999532 | | 4.081787 | 0.410035 | 0.713579 | | 3.492652 | 0.29886 | 0.819366 |
| SLX4IP | 3.25331 | -0.63955 | 0.999532 | | 3.757982 | -0.42896 | 0.713579 | | 2.580407 | 0.222954 | 0.899732 |
| SPACA9 | 2.969815 | -0.24608 | 0.999532 | | 3.100882 | -0.45164 | 0.713579 | | 2.514018 | -0.19098 | 0.912963 |
| ARSD | 4.547836 | 0.171102 | 0.999532 | | 5.491723 | 0.345528 | 0.713644 | | 4.840496 | 0.194248 | 0.87591 |
| NKD1 | 4.833256 | 0.678755 | 0.999532 | | 4.901715 | 0.428227 | 0.715103 | | 5.180292 | -0.22897 | 0.864777 |
| TDRD10 | 2.899433 | 0.098341 | 0.999532 | | 2.855308 | -0.4726 | 0.715103 | | 2.464633 | -0.54767 | 0.682485 |
| MORN3 | 2.737965 | -0.18019 | 0.999532 | | 2.801401 | -0.46737 | 0.715361 | | 2.320009 | -0.27094 | 0.87591 |
| ALDH9A1 | 4.975747 | 0.159068 | 0.999532 | | 5.654836 | 0.335526 | 0.715361 | | 5.282627 | 0.189974 | 0.87591 |
| WASHC5 | 3.59095 | 0.156184 | 0.999532 | | 4.052954 | 0.390759 | 0.715361 | | 3.801194 | 0.234695 | 0.869378 |
| BCAN | 2.744006 | 0.159751 | 0.999532 | | 2.518975 | -0.51049 | 0.715361 | | 2.274319 | -0.65215 | 0.64142 |
| GPR132 | 3.396225 | 0.01412 | 0.999532 | | 3.29313 | -0.45501 | 0.715361 | | 2.883345 | -0.45166 | 0.741003 |
| CDH4 | 2.286328 | 0.070479 | 0.999532 | | 2.065029 | -0.54509 | 0.715361 | | 1.837227 | -0.59933 | 0.706407 |
| PSMF1 | 6.681082 | -0.07853 | 0.999532 | | 8.512753 | 0.311834 | 0.715361 | | 7.161351 | 0.410703 | 0.629006 |
| PAQR7 | 9.612632 | 0.267505 | 0.999532 | | 10.49511 | 0.253914 | 0.715361 | | 9.769232 | 0.001266 | 0.999385 |
| ATG2B | 3.47502 | 0.083439 | 0.999532 | | 4.290775 | 0.37595 | 0.715361 | | 3.85169 | 0.308438 | 0.801905 |
| MTFR2 | 2.313083 | -0.22105 | 0.999532 | | 2.276623 | -0.50821 | 0.715454 | | 1.850803 | -0.27089 | 0.88885 |
| TMEM47 | 8.539146 | -0.56264 | 0.999532 | | 12.64658 | 0.265636 | 0.715454 | | 8.973987 | 0.85191 | 0.158141 |
| WDR43 | 3.056312 | 1.049266 | 0.999532 | | 2.545729 | 0.490069 | 0.715454 | | 3.404474 | -0.5331 | 0.652925 |
| LMBR1 | 2.828398 | 0.398943 | 0.999532 | | 3.193108 | 0.43046 | 0.715454 | | 3.228765 | 0.049435 | 0.980581 |
| RGL4 | 1.853818 | 0.068337 | 0.999532 | | 1.731815 | -0.60083 | 0.715454 | | 1.60413 | -0.65069 | 0.717138 |
| BPIFC | 2.184808 | -0.51008 | 0.999532 | | 2.562578 | -0.54061 | 0.715642 | | 1.779211 | -0.00962 | 0.997967 |
| RUNDC1 | 3.947215 | -0.24498 | 0.999532 | | 4.104124 | -0.38935 | 0.71582 | | 3.382146 | -0.12622 | 0.941197 |
| GET4 | 3.405797 | 0.387654 | 0.999532 | | 3.642727 | 0.405822 | 0.71582 | | 3.51088 | 0.032668 | 0.989278 |
| MAPK1 | 12.27273 | -0.04981 | 0.999532 | | 15.03631 | 0.235625 | 0.71582 | | 12.68921 | 0.303528 | 0.647581 |
| ERG | 8.67192 | 0.177412 | 0.999532 | | 9.870815 | 0.292375 | 0.71582 | | 9.254418 | 0.124699 | 0.911581 |
| ZBTB25 | 3.431811 | 0.522222 | 0.999532 | | 3.596145 | 0.421896 | 0.71582 | | 3.604038 | -0.08397 | 0.963492 |
| OR4M2 | 2.404606 | 0.371667 | 0.999532 | | 2.087639 | -0.54765 | 0.71582 | | 1.952874 | -0.90747 | 0.522078 |
| PRR12 | 4.218165 | 0.01631 | 0.999532 | | 5.190755 | 0.351276 | 0.71582 | | 4.503121 | 0.350089 | 0.740093 |
| ECD | 3.468895 | 0.26231 | 0.999532 | | 3.825487 | 0.413977 | 0.71582 | | 3.505062 | 0.158816 | 0.920259 |
| FNBP1L | 7.164021 | 0.410794 | 0.999532 | | 7.618745 | 0.312567 | 0.71582 | | 7.648843 | -0.08511 | 0.945456 |
| EYA3 | 3.315403 | 0.381317 | 0.999532 | | 3.810871 | 0.41977 | 0.71582 | | 3.536398 | 0.063636 | 0.974421 |
| DIP2C-AS | 2.434641 | 0.152266 | 0.999532 | | 2.44408 | -0.5233 | 0.716285 | | 2.091559 | -0.6486 | 0.660958 |
| COL8A2 | 2.424288 | -0.62691 | 0.999532 | | 2.981641 | -0.48093 | 0.71637 | | 2.11114 | 0.162231 | 0.940618 |
| GLTPD2 | 3.516995 | -0.01817 | 0.999532 | | 3.460333 | -0.43972 | 0.716386 | | 3.045682 | -0.40836 | 0.751138 |
| GNAI1 | 6.173175 | 0.174999 | 0.999532 | | 6.962243 | 0.319877 | 0.716386 | | 6.510289 | 0.157139 | 0.892456 |
| GPBAR1 | 4.273751 | 0.532003 | 0.999532 | | 3.499403 | -0.42225 | 0.716521 | | 3.550857 | -0.93346 | 0.281853 |
| RILP | 2.76825 | 0.185886 | 0.999532 | | 2.426001 | -0.50683 | 0.716627 | | 2.317044 | -0.66917 | 0.621015 |
| AAGAB | 3.319005 | 0.61537 | 0.999532 | | 3.476159 | 0.441478 | 0.716627 | | 3.597628 | -0.15496 | 0.920259 |
| TACR3 | 2.538455 | 0.182109 | 0.999532 | | 2.16829 | -0.53075 | 0.716844 | | 2.017872 | -0.69639 | 0.636291 |
| SLC6A8 | 4.394382 | 0.057948 | 0.999532 | | 4.353336 | -0.38981 | 0.717073 | | 3.617736 | -0.42588 | 0.701251 |
| CD1A | 2.176212 | -0.55662 | 0.999532 | | 2.503383 | -0.54674 | 0.717073 | | 1.669665 | 0.032141 | 0.993418 |
| FES | 2.957418 | 0.006389 | 0.999532 | | 3.909515 | 0.392753 | 0.717073 | | 3.221975 | 0.403136 | 0.736481 |
| ARHGEF | 5.053039 | 0.034768 | 0.999532 | | 6.164892 | 0.340797 | 0.717073 | | 5.450426 | 0.323587 | 0.752299 |
| MT1G | 36.39542 | -0.07623 | 0.999532 | | 50.30271 | 0.472095 | 0.717284 | | 41.59648 | 0.574266 | 0.65401 |
| CCDC142 | 3.646997 | -0.25707 | 0.999532 | | 4.01276 | -0.44835 | 0.717373 | | 3.046117 | -0.17203 | 0.920259 |
| NQO1 | 4.629864 | 0.097346 | 0.999532 | | 5.841719 | 0.353054 | 0.717373 | | 5.080822 | 0.27626 | 0.814483 |
| ELAVL1 | 5.287231 | -0.41865 | 0.999532 | | 6.036871 | -0.33458 | 0.717373 | | 4.235486 | 0.105062 | 0.94366 |
| HSPB11 | 4.144956 | 0.141514 | 0.999532 | | 4.903681 | 0.36407 | 0.717373 | | 4.510167 | 0.244966 | 0.83841 |
| PUM1 | 3.573993 | 0.04512 | 0.999532 | | 4.64322 | 0.363195 | 0.717373 | | 3.835332 | 0.330964 | 0.766467 |
| ZER1 | 4.712883 | 0.440129 | 0.999532 | | 5.170009 | 0.352371 | 0.71764 | | 5.08372 | -0.06972 | 0.964728 |
| MLLT6 | 17.23906 | -0.04286 | 0.999532 | | 21.61226 | 0.206728 | 0.71764 | | 17.96669 | 0.278586 | 0.662584 |
| TAX1BP1 | 4.836608 | -0.03585 | 0.999532 | | 6.418781 | 0.320019 | 0.717826 | | 5.332547 | 0.380149 | 0.705425 |
| LAMA5 | 9.881083 | -0.30906 | 0.999532 | | 13.55983 | 0.229519 | 0.717826 | | 10.32451 | 0.55889 | 0.359441 |
| BAMBI | 5.926682 | -0.19885 | 0.999532 | | 7.64654 | 0.314401 | 0.717826 | | 6.230956 | 0.520663 | 0.563455 |
| ARL6IP1 | 4.809531 | 0.126032 | 0.999532 | | 5.872719 | 0.349676 | 0.717826 | | 5.038958 | 0.23948 | 0.842754 |
| LRMDA | 2.469098 | 0.331173 | 0.999532 | | 2.785438 | 0.46901 | 0.717826 | | 2.63999 | 0.15746 | 0.928522 |
| RUSF1 | 4.70442 | 0.145886 | 0.999532 | | 5.668478 | 0.355133 | 0.717826 | | 4.907871 | 0.232729 | 0.846786 |
| AKR1C3 | 4.45984 | 0.117698 | 0.999532 | | 5.553817 | 0.434102 | 0.717826 | | 5.05265 | 0.325035 | 0.816589 |
| PLP2 | 5.006435 | -0.32247 | 0.999532 | | 5.432431 | -0.33907 | 0.717826 | | 4.14685 | -0.01199 | 0.996824 |
| TRMO | 3.147181 | -0.05772 | 0.999532 | | 3.212195 | -0.42819 | 0.717826 | | 2.710799 | -0.34407 | 0.813755 |
| SNED1 | 5.934247 | -0.05756 | 0.999532 | | 7.763338 | 0.320243 | 0.717826 | | 6.534329 | 0.398783 | 0.666566 |
| TMEM95 | 2.19232 | -0.31619 | 0.999532 | | 2.370881 | -0.51702 | 0.717826 | | 1.781283 | -0.18041 | 0.934665 |
| OR13C5 | 2.66741 | 0.460181 | 0.999532 | | 2.067689 | -0.548 | 0.717826 | | 2.274792 | -0.99193 | 0.446959 |
| SLC22A1 | 3.402703 | -0.01775 | 0.999532 | | 3.290812 | -0.43853 | 0.717826 | | 2.885369 | -0.40667 | 0.765574 |
| PCBP3 | 3.761883 | 0.393073 | 0.999532 | | 2.931635 | -0.45326 | 0.717826 | | 3.172858 | -0.84025 | 0.454116 |
| HSPA8 | 20.86089 | -0.35388 | 0.999532 | | 28.8932 | 0.229968 | 0.717826 | | 21.61 | 0.618378 | 0.209531 |
| MEX3D | 5.572878 | 0.271936 | 0.999532 | | 6.099958 | 0.327149 | 0.717826 | | 5.858699 | 0.065728 | 0.966652 |
| TMEM97 | 2.788823 | -0.0543 | 0.999532 | | 2.566183 | -0.49494 | 0.717826 | | 2.287718 | -0.42147 | 0.792416 |
| ZNF446 | 3.486346 | 0.608803 | 0.999532 | | 3.905471 | 0.456381 | 0.717826 | | 4.032495 | -0.12905 | 0.942744 |
| PEF1 | 6.035246 | -0.08483 | 0.999532 | | 7.486831 | 0.321377 | 0.717826 | | 6.59677 | 0.410043 | 0.702463 |
| CHST1 | 4.429319 | 0.177769 | 0.999532 | | 4.257133 | -0.39341 | 0.717826 | | 3.760996 | -0.55021 | 0.603483 |
| YBX2 | 2.177731 | -0.11549 | 0.999532 | | 2.130601 | -0.53813 | 0.717826 | | 1.778155 | -0.40507 | 0.824444 |
| DBNDD2 | 4.014208 | -0.17443 | 0.999532 | | 5.279406 | 0.336348 | 0.717826 | | 4.31287 | 0.522848 | 0.608862 |
| SEMA3G | 5.309759 | -0.40874 | 0.999532 | | 7.373289 | 0.334647 | 0.717826 | | 5.61679 | 0.754097 | 0.386771 |
| SUCLA2 | 4.572796 | -0.53032 | 0.999532 | | 6.72345 | 0.30889 | 0.717826 | | 4.961932 | 0.857206 | 0.26539 |
| SLC2A8 | 2.579001 | 0.252927 | 0.999532 | | 2.327963 | -0.51471 | 0.717826 | | 2.228126 | -0.75059 | 0.566812 |
| ANKRD20 | 6.994855 | 0.036324 | 0.999532 | | 7.070322 | -0.37708 | 0.717826 | | 6.053538 | -0.38453 | 0.740755 |
| TIMM23B | 5.885024 | 0.152217 | 0.999532 | | 6.937134 | 0.3215 | 0.717974 | | 6.223975 | 0.18735 | 0.863401 |
| STX18 | 6.673791 | 0.34712 | 0.999532 | | 7.521632 | 0.292748 | 0.717974 | | 7.075242 | -0.03324 | 0.982518 |
| SMIM14 | 8.212871 | 0.500568 | 0.999532 | | 8.080528 | 0.295206 | 0.718206 | | 8.283314 | -0.20049 | 0.842754 |
| HADH | 3.93289 | 0.25505 | 0.999532 | | 4.615858 | 0.383692 | 0.718206 | | 4.059171 | 0.152189 | 0.914687 |
| GPCPD1 | 3.647837 | -0.37088 | 0.999532 | | 3.809814 | -0.41758 | 0.718206 | | 2.981132 | -0.0479 | 0.985569 |
| VWA5B1 | 3.016767 | 0.304234 | 0.999532 | | 2.485001 | -0.48569 | 0.718206 | | 2.542811 | -0.77654 | 0.533876 |
| IL5 | 2.153351 | -0.14123 | 0.999532 | | 2.157391 | -0.54609 | 0.718206 | | 1.677167 | -0.38197 | 0.839299 |
| UNC5B | 16.04386 | -0.09237 | 0.999532 | | 20.11111 | 0.288005 | 0.718527 | | 17.13233 | 0.396099 | 0.577942 |
| CA4 | 3.340742 | 0.330012 | 0.999532 | | 2.891883 | -0.4721 | 0.718527 | | 2.763574 | -0.78198 | 0.49299 |
| SYDE2 | 2.252592 | -0.49503 | 0.999532 | | 2.52047 | -0.48474 | 0.718676 | | 1.902926 | 0.026816 | 0.994537 |
| RRBP1 | 6.243995 | 0.104623 | 0.999532 | | 7.216322 | 0.301072 | 0.718901 | | 6.656628 | 0.220497 | 0.817974 |
| EXOSC7 | 3.427844 | 0.457773 | 0.999532 | | 3.763446 | 0.407831 | 0.719033 | | 3.745623 | -0.03027 | 0.990309 |
| RPL27 | 13.58902 | -0.46654 | 0.999532 | | 18.48313 | 0.246653 | 0.7191 | | 13.9209 | 0.706337 | 0.213497 |
| DAPP1 | 2.317695 | 0.897884 | 0.999532 | | 2.31523 | 0.517448 | 0.7191 | | 2.661828 | -0.36167 | 0.809832 |
| ZNF566 | 2.372415 | -0.18475 | 0.999532 | | 2.289538 | -0.50382 | 0.7191 | | 1.936829 | -0.31305 | 0.869449 |
| MBLAC1 | 2.602964 | 0.017686 | 0.999532 | | 2.686238 | -0.4951 | 0.719306 | | 2.235624 | -0.49118 | 0.740093 |
| WDR62 | 2.160079 | -0.02417 | 0.999532 | | 2.094759 | -0.53184 | 0.719391 | | 1.846212 | -0.49114 | 0.769793 |
| GIPC3 | 2.469308 | -0.52954 | 0.999532 | | 2.934022 | -0.46564 | 0.719864 | | 2.130263 | 0.083929 | 0.973126 |
| NPDC1 | 17.30606 | -0.11881 | 0.999532 | | 22.00498 | 0.263239 | 0.719864 | | 18.32827 | 0.403067 | 0.524707 |
| DMAP1 | 3.28317 | 0.210506 | 0.999532 | | 4.235879 | 0.416054 | 0.719864 | | 3.854164 | 0.230138 | 0.876781 |
| KMT5B | 3.473335 | 0.006783 | 0.999532 | | 4.487515 | 0.360864 | 0.719864 | | 3.756646 | 0.371678 | 0.737952 |
| CREBZF | 7.863993 | -0.04444 | 0.999532 | | 10.14903 | 0.277333 | 0.720074 | | 8.207542 | 0.344154 | 0.664965 |
| C15orf39 | 4.195784 | -0.00154 | 0.999532 | | 4.042072 | -0.38513 | 0.72009 | | 3.616013 | -0.37082 | 0.765012 |
| RLN2 | 2.420002 | 0.589566 | 0.999532 | | 2.441574 | 0.492818 | 0.720119 | | 2.649758 | -0.08004 | 0.972221 |
| EYA2 | 1.896876 | 0.526662 | 0.999532 | | 2.084451 | 0.54716 | 0.720119 | | 2.052181 | 0.039307 | 0.990309 |
| B3GNTL1 | 3.048897 | 0.308418 | 0.999532 | | 2.786439 | -0.49718 | 0.720119 | | 2.720691 | -0.7958 | 0.515762 |
| F8 | 1.919817 | 0.574001 | 0.999532 | | 2.025285 | 0.519838 | 0.720119 | | 2.072646 | -0.03768 | 0.990309 |
| ARF4 | 8.82809 | 0.293741 | 0.999532 | | 9.611416 | 0.265284 | 0.720119 | | 9.139411 | -0.01008 | 0.996681 |
| CABP4 | 2.410039 | -0.23982 | 0.999532 | | 2.529561 | -0.51324 | 0.720119 | | 1.865896 | -0.25201 | 0.895421 |
| TUBGCP | 2.845287 | 0.726128 | 0.999532 | | 2.04593 | -0.53484 | 0.720119 | | 2.4006 | -1.2458 | 0.263754 |
| VKORC1L | 5.082409 | 0.306943 | 0.999532 | | 5.786146 | 0.331486 | 0.720119 | | 5.415717 | 0.043724 | 0.978229 |
| NPM1 | 33.80976 | -0.15666 | 0.999532 | | 42.60329 | 0.163241 | 0.720119 | | 34.21055 | 0.341887 | 0.456688 |
| LURAP1 | 2.735466 | 0.299396 | 0.999532 | | 2.46197 | -0.49946 | 0.720119 | | 2.340994 | -0.76915 | 0.575703 |
| HLA-A | 49.0844 | -0.33169 | 0.999532 | | 68.64926 | 0.305862 | 0.720119 | | 53.0827 | 0.658749 | 0.283338 |
| RFC5 | 3.126206 | -0.07793 | 0.999532 | | 3.167933 | -0.43422 | 0.720121 | | 2.558186 | -0.33759 | 0.821752 |
| MFSD13A | 2.443437 | -0.31875 | 0.999532 | | 2.6118 | -0.49054 | 0.72025 | | 1.966579 | -0.15475 | 0.944635 |
| MFN2 | 4.992284 | -0.39175 | 0.999532 | | 5.347612 | -0.36254 | 0.72025 | | 3.946604 | 0.046615 | 0.979996 |
| RASA3 | 4.676516 | -0.58148 | 0.999532 | | 5.616984 | -0.38319 | 0.720365 | | 3.987793 | 0.220153 | 0.878232 |
| IRS2 | 5.338535 | -0.37817 | 0.999532 | | 5.917389 | -0.32825 | 0.720501 | | 4.385893 | 0.065647 | 0.969693 |
| DGCR8 | 3.274819 | -0.03756 | 0.999532 | | 4.167036 | 0.373292 | 0.720598 | | 3.603431 | 0.425184 | 0.717627 |
| YBX1 | 26.48346 | -0.00379 | 0.999532 | | 32.18516 | 0.237554 | 0.720598 | | 27.27928 | 0.263153 | 0.717627 |
| KIF1A | 2.351322 | 0.689252 | 0.999532 | | 2.385075 | 0.481697 | 0.720598 | | 2.573012 | -0.19033 | 0.908698 |
| KIT | 2.790882 | -0.05297 | 0.999532 | | 2.855591 | -0.50814 | 0.720598 | | 2.218258 | -0.44445 | 0.765852 |
| ATOX1 | 8.274033 | -0.15219 | 0.999532 | | 10.41241 | 0.258275 | 0.720598 | | 8.566973 | 0.417265 | 0.598158 |
| AIP | 3.997169 | 0.19768 | 0.999532 | | 4.475875 | 0.360328 | 0.720598 | | 4.252099 | 0.192896 | 0.892869 |
| DDX60L | 1.620126 | -0.12346 | 0.999532 | | 2.251414 | 0.503487 | 0.720598 | | 1.96042 | 0.64901 | 0.667066 |
| OR8H2 | 2.165776 | 0.053119 | 0.999532 | | 2.087637 | -0.52614 | 0.720598 | | 1.82248 | -0.55815 | 0.728015 |
| AGGF1 | 4.320997 | 0.636119 | 0.999532 | | 4.53019 | 0.383482 | 0.720598 | | 4.738977 | -0.23701 | 0.846125 |
| COA4 | 3.059488 | -0.28802 | 0.999532 | | 3.231022 | -0.4157 | 0.720598 | | 2.567535 | -0.11306 | 0.954304 |
| IGFBP3 | 2.37663 | 0.495706 | 0.999532 | | 2.449308 | 0.491375 | 0.720598 | | 2.485209 | 0.011941 | 0.996891 |
| NGRN | 4.67863 | -0.12724 | 0.999532 | | 4.855263 | -0.35789 | 0.720598 | | 3.893885 | -0.21255 | 0.877943 |
| CD48 | 2.629296 | 0.106126 | 0.999532 | | 2.331247 | -0.51836 | 0.720598 | | 2.083568 | -0.61303 | 0.686214 |
| TRIM60 | 2.381793 | -0.3146 | 0.999532 | | 2.708219 | -0.49623 | 0.720598 | | 2.064019 | -0.15485 | 0.944635 |
| PHF24 | 2.631523 | 0.441483 | 0.999532 | | 1.992901 | -0.54829 | 0.720598 | | 2.320157 | -0.97452 | 0.505689 |
| TMEM119 | 3.274439 | -0.17362 | 0.999532 | | 3.293036 | -0.44095 | 0.720598 | | 2.928502 | -0.24982 | 0.881316 |
| ADAM10 | 3.55621 | -0.1731 | 0.999532 | | 4.45204 | 0.384939 | 0.720598 | | 3.797941 | 0.561962 | 0.621895 |
| FHL1 | 10.00208 | 0.477371 | 0.999532 | | 10.56927 | 0.298247 | 0.720598 | | 10.46336 | -0.15402 | 0.87892 |
| PLA2G12 | 3.132348 | 0.067214 | 0.999532 | | 2.965443 | -0.44774 | 0.720598 | | 2.651455 | -0.49538 | 0.712849 |
| SLC39A1 | 3.464474 | 0.429232 | 0.999532 | | 3.736432 | 0.399265 | 0.720598 | | 3.722829 | -0.01767 | 0.995943 |
| PHLDB3 | 4.100516 | -0.18724 | 0.999532 | | 4.12733 | -0.40412 | 0.720598 | | 3.468431 | -0.19426 | 0.903329 |
| ARHGAP | 4.157944 | -0.24281 | 0.999532 | | 5.431115 | 0.384126 | 0.720598 | | 4.648174 | 0.669169 | 0.465298 |
| GPR135 | 1.968196 | -0.26812 | 0.999532 | | 1.958738 | -0.57419 | 0.720598 | | 1.565222 | -0.28811 | 0.895865 |
| HAUS5 | 3.386676 | -0.00665 | 0.999532 | | 3.277878 | -0.47405 | 0.720598 | | 2.872631 | -0.45064 | 0.765852 |
| APOC3 | 2.485438 | -0.10436 | 0.999532 | | 2.43116 | -0.5157 | 0.720598 | | 2.050737 | -0.38756 | 0.823193 |
| MTARC2 | 1.875342 | 0.894523 | 0.999532 | | 1.965022 | 0.580727 | 0.720598 | | 2.12291 | -0.29209 | 0.87591 |
| RANBP6 | 4.046314 | 0.577186 | 0.999532 | | 3.961087 | 0.390255 | 0.720598 | | 4.203756 | -0.18273 | 0.900641 |
| CHMP4A | 9.161749 | -0.19491 | 0.999532 | | 9.986557 | -0.27432 | 0.720598 | | 7.591012 | -0.06414 | 0.961567 |
| DCAF1 | 4.629637 | 0.232921 | 0.999532 | | 4.175964 | -0.41432 | 0.720598 | | 3.741178 | -0.62508 | 0.536289 |
| KIAA195 | 2.411487 | -0.01778 | 0.999532 | | 3.205332 | 0.427661 | 0.720598 | | 2.643362 | 0.461186 | 0.723933 |
| ACP3 | 4.05398 | 0.366242 | 0.999532 | | 3.548724 | -0.45398 | 0.720598 | | 3.292245 | -0.82319 | 0.484718 |
| P2RY11 | 3.913018 | -0.34672 | 0.999532 | | 4.175322 | -0.69914 | 0.720598 | | 2.949475 | -0.4497 | 0.750371 |
| ORC3 | 3.64046 | 0.132029 | 0.999532 | | 4.577464 | 0.368149 | 0.720598 | | 4.101167 | 0.257161 | 0.839373 |
| EME2 | 4.985742 | 0.515784 | 0.999532 | | 5.401675 | 0.389655 | 0.720598 | | 5.50422 | -0.12115 | 0.932446 |
| C22orf15 | 3.861154 | -0.33863 | 0.999532 | | 4.039433 | -0.41831 | 0.720598 | | 3.342261 | -0.07491 | 0.975284 |
| MUSTN1 | 2.021833 | 0.074597 | 0.999532 | | 2.759423 | 0.46831 | 0.720598 | | 2.456842 | 0.411181 | 0.786052 |
| AMDHD2 | 7.078758 | 0.007669 | 0.999532 | | 9.128726 | 0.338864 | 0.720598 | | 7.476827 | 0.332171 | 0.705425 |
| FER | 2.300252 | 0.044607 | 0.999532 | | 3.258946 | 0.45016 | 0.720598 | | 2.799488 | 0.430791 | 0.761699 |
| PLGLB2 | 8.243027 | -0.00134 | 0.999532 | | 8.379368 | -0.29316 | 0.720598 | | 7.266434 | -0.26823 | 0.78597 |
| UBB | 23.43251 | -0.00041 | 0.999532 | | 29.12703 | 0.228056 | 0.720598 | | 24.47191 | 0.256341 | 0.70375 |
| TCIM | 22.86484 | -0.11693 | 0.999532 | | 29.31351 | 0.281938 | 0.720598 | | 24.47406 | 0.452988 | 0.359638 |
| PSMD6 | 4.332745 | -0.07272 | 0.999532 | | 5.701683 | 0.330892 | 0.720598 | | 4.582339 | 0.418864 | 0.666566 |
| NUP188 | 3.310175 | 0.32456 | 0.999532 | | 3.626859 | 0.414879 | 0.720598 | | 3.518315 | 0.114015 | 0.944993 |
| ENDOD1 | 3.54772 | -0.08674 | 0.999532 | | 4.715542 | 0.377609 | 0.720598 | | 3.854388 | 0.483321 | 0.663412 |
| ZSWIM8 | 4.837581 | 0.119853 | 0.999532 | | 5.924161 | 0.352903 | 0.720598 | | 5.264282 | 0.253022 | 0.833685 |
| PTCD1 | 3.566492 | -0.06141 | 0.999532 | | 3.562323 | -0.41862 | 0.720598 | | 2.938649 | -0.3384 | 0.810635 |
| NKAIN2 | 2.199468 | -0.19534 | 0.999532 | | 2.271886 | -0.51537 | 0.720598 | | 1.885677 | -0.30269 | 0.87591 |
| MRPS5 | 3.777485 | 0.274922 | 0.999532 | | 4.46516 | 0.374435 | 0.720598 | | 4.002172 | 0.119891 | 0.9402 |
| HNF4G | 3.089051 | 0.12895 | 0.999532 | | 2.799074 | -0.46472 | 0.720598 | | 2.625931 | -0.57303 | 0.671791 |
| TOPORS | 4.224813 | -0.34878 | 0.999532 | | 4.580113 | -0.38773 | 0.720598 | | 3.493491 | -0.01932 | 0.994811 |
| FBXW7 | 4.774792 | -0.27097 | 0.999532 | | 4.936709 | -0.38244 | 0.720598 | | 3.817197 | -0.09919 | 0.956322 |
| SLC7A6 | 2.893918 | 0.022105 | 0.999532 | | 2.647492 | -0.48024 | 0.720598 | | 2.451558 | -0.49075 | 0.737109 |
| TSC22D2 | 3.532242 | -0.40796 | 0.999532 | | 4.25306 | -0.37775 | 0.720598 | | 2.94477 | 0.06878 | 0.974421 |
| SUMO4 | 4.040106 | -0.10085 | 0.999532 | | 5.394099 | 0.352794 | 0.720598 | | 4.492717 | 0.475186 | 0.647581 |
| IGFL4 | 2.215223 | -0.08474 | 0.999532 | | 2.216317 | -0.52434 | 0.720598 | | 1.858627 | -0.41627 | 0.814483 |
| PDZD8 | 8.922663 | -0.08862 | 0.999532 | | 11.589 | 0.246127 | 0.720598 | | 9.429505 | 0.353641 | 0.652925 |
| TRAPPC3 | 4.187746 | -0.10877 | 0.999532 | | 5.556084 | 0.330297 | 0.720598 | | 4.538075 | 0.456917 | 0.647218 |

| | | | | | | | | | | | |
|---|---|---|---|---|---|---|---|---|---|---|---|
| DNAJC5 | 4.797383 | 0.206677 | 0.999532 | | 5.758788 | 0.349554 | 0.720598 | | 5.053141 | 0.169873 | 0.889853 |
| SLC25A2 | 3.892414 | 0.279912 | 0.999532 | | 3.535127 | -0.41653 | 0.720598 | | 3.345197 | -0.6725 | 0.540342 |
| GATAD2A | 5.553429 | -0.35009 | 0.999532 | | 6.007351 | -0.34327 | 0.720598 | | 4.516117 | 0.020546 | 0.993435 |
| MPC2 | 3.809912 | 0.096339 | 0.999532 | | 4.812413 | 0.363187 | 0.720598 | | 4.073305 | 0.280401 | 0.810635 |
| MYO18B | 2.585002 | 0.133083 | 0.999532 | | 2.240273 | -0.50973 | 0.720598 | | 2.074271 | -0.63101 | 0.667066 |
| SBF1 | 2.099646 | 0.604218 | 0.999532 | | 2.181499 | 0.492075 | 0.720598 | | 2.28701 | -0.094 | 0.965619 |
| TOMM6 | 9.604911 | -0.01687 | 0.999532 | | 12.06046 | 0.262102 | 0.720598 | | 10.03526 | 0.299975 | 0.694961 |
| CFAP221 | 2.145498 | 0.038233 | 0.999532 | | 2.11063 | -0.52741 | 0.720598 | | 1.728714 | -0.54206 | 0.739496 |
| HOXC11 | 2.180105 | -0.45665 | 0.999532 | | 2.417825 | -0.5298 | 0.720598 | | 1.803056 | -0.05562 | 0.986898 |
| PRDX6 | 4.620255 | 0.08847 | 0.999532 | | 6.232558 | 0.326404 | 0.720598 | | 5.212774 | 0.267911 | 0.817974 |
| ADI1 | 7.36169 | 0.315705 | 0.999532 | | 8.199042 | 0.299532 | 0.720598 | | 7.679289 | 0.004891 | 0.997967 |
| HS3ST5 | 1.970019 | -0.39482 | 0.999532 | | 2.236564 | -0.51759 | 0.720598 | | 1.690179 | -0.09799 | 0.972221 |
| MAP3K6 | 6.769224 | -0.24623 | 0.999532 | | 7.621202 | -0.3008 | 0.720598 | | 5.938518 | -0.03317 | 0.985693 |
| DDX6 | 5.599217 | -0.14275 | 0.999532 | | 7.386285 | 0.3071 | 0.720598 | | 5.947004 | 0.468809 | 0.566812 |
| WNT2 | 1.967021 | -0.37952 | 0.999532 | | 2.055632 | -0.52881 | 0.720598 | | 1.595011 | -0.13417 | 0.958191 |
| KLK2 | 2.90228 | 0.095395 | 0.999532 | | 2.701043 | -0.5009 | 0.720598 | | 2.428432 | -0.57812 | 0.702463 |
| TIAM1 | 1.924937 | -0.03165 | 0.999532 | | 2.77933 | 0.494718 | 0.720598 | | 2.403356 | 0.543623 | 0.720729 |
| OR5M1 | 3.423218 | 0.406406 | 0.999532 | | 2.874872 | -0.46704 | 0.720598 | | 2.933609 | -0.85631 | 0.432473 |
| FASTKD3 | 2.146156 | 0.759768 | 0.999532 | | 2.094534 | 0.534978 | 0.720598 | | 2.310649 | -0.20956 | 0.907968 |
| GDPD4 | 2.534717 | 0.083214 | 0.999532 | | 2.285671 | -0.51494 | 0.720598 | | 1.984972 | -0.58186 | 0.702463 |
| MMP1 | 2.699534 | 0.140061 | 0.999532 | | 2.372874 | -0.53135 | 0.720598 | | 2.284816 | -0.64417 | 0.663412 |
| PAIP2 | 6.259964 | -0.09059 | 0.999532 | | 7.961769 | 0.302657 | 0.720598 | | 6.539357 | 0.408933 | 0.657538 |
| GGA2 | 4.510822 | 0.336385 | 0.999532 | | 4.875976 | 0.363013 | 0.720598 | | 4.623764 | 0.042538 | 0.98153 |
| MYO9B | 7.914415 | 0.112602 | 0.999532 | | 7.61599 | -0.3014 | 0.720598 | | 6.630446 | -0.39242 | 0.652925 |
| WNT5B | 1.997443 | 0.285754 | 0.999532 | | 1.756739 | -0.55947 | 0.720833 | | 1.663855 | -0.82344 | 0.578798 |
| ULK4 | 3.412222 | 0.069247 | 0.999532 | | 3.331545 | -0.41219 | 0.720833 | | 2.769334 | -0.46527 | 0.723933 |
| ALDH1B1 | 2.870181 | 0.307226 | 0.999532 | | 3.484454 | 0.430748 | 0.720833 | | 3.240071 | 0.135249 | 0.934354 |
| RHNO1 | 2.386321 | -0.14739 | 0.999532 | | 2.30313 | -0.50127 | 0.720996 | | 1.9369 | -0.33843 | 0.855173 |
| VSNL1 | 2.507183 | -0.43992 | 0.999532 | | 2.988165 | -0.49274 | 0.72101 | | 2.130667 | -0.01299 | 0.996891 |
| TMEM192 | 2.524537 | 0.094151 | 0.999532 | | 3.248819 | 0.421301 | 0.721198 | | 2.909239 | 0.35339 | 0.806394 |
| ERF | 9.817262 | -0.26101 | 0.999532 | | 10.82954 | -0.27991 | 0.721664 | | 8.319795 | 0.005623 | 0.997153 |
| RNF168 | 5.995287 | 0.207508 | 0.999532 | | 7.165669 | 0.341574 | 0.72205 | | 6.848242 | 0.136886 | 0.904801 |
| JMJD6 | 3.613661 | -0.10471 | 0.999532 | | 3.872043 | -0.38576 | 0.72205 | | 3.041441 | -0.25643 | 0.859426 |
| SLC25A2 | 4.878212 | 0.457103 | 0.999532 | | 5.208027 | 0.348778 | 0.72205 | | 5.20841 | -0.08988 | 0.949136 |
| DCUN1D2 | 1.864006 | 0.230475 | 0.999532 | | 2.242886 | 0.510748 | 0.722419 | | 2.227388 | 0.29934 | 0.869378 |
| SLC46A1 | 3.107945 | -0.0025 | 0.999532 | | 2.993469 | -0.45062 | 0.722419 | | 2.551584 | -0.42944 | 0.754671 |
| PLEKHB2 | 4.650728 | -0.07166 | 0.999532 | | 5.694118 | 0.3384 | 0.722419 | | 4.755946 | 0.418542 | 0.686214 |
| DPPA3 | 3.543853 | 0.20576 | 0.999532 | | 3.062492 | -0.45647 | 0.722419 | | 3.042473 | -0.65715 | 0.5991 |
| CA13 | 3.278776 | -0.27451 | 0.999532 | | 3.635923 | -0.40581 | 0.722757 | | 2.663145 | -0.10979 | 0.953211 |
| NMRK2 | 4.264452 | 0.365227 | 0.999532 | | 3.696589 | -0.40685 | 0.722757 | | 3.583901 | -0.75447 | 0.465298 |
| CCSAP | 3.05162 | 0.137767 | 0.999532 | | 2.772844 | -0.46416 | 0.722845 | | 2.457586 | -0.58399 | 0.66232 |
| PEA15 | 11.31455 | -0.16248 | 0.999532 | | 14.80977 | 0.255782 | 0.722845 | | 11.7887 | 0.442706 | 0.541754 |
| OR1D2 | 2.098504 | 0.355048 | 0.999532 | | 1.709054 | -0.59401 | 0.722845 | | 1.697302 | -0.93269 | 0.536289 |
| GJA9 | 3.316348 | 0.26377 | 0.999532 | | 3.142219 | -0.46593 | 0.722845 | | 2.648528 | -0.70927 | 0.540342 |
| GPS2 | 7.79284 | -0.02178 | 0.999532 | | 10.17009 | 0.266526 | 0.722845 | | 8.485489 | 0.309543 | 0.709825 |
| PLPPR3 | 2.331697 | -0.12528 | 0.999532 | | 2.406126 | -0.51701 | 0.722845 | | 1.843625 | -0.37248 | 0.835554 |
| CCDC34 | 2.696854 | 0.017764 | 0.999532 | | 2.6219 | -0.49791 | 0.722845 | | 2.252187 | -0.49797 | 0.745939 |
| MTRF1L | 3.612969 | -0.04945 | 0.999532 | | 3.567025 | -0.42342 | 0.722845 | | 3.075358 | -0.36 | 0.791466 |
| CRK | 6.816431 | -0.33105 | 0.999532 | | 9.144377 | 0.272329 | 0.722845 | | 7.050934 | 0.618545 | 0.383113 |
| EIF2B3 | 2.330034 | 0.263915 | 0.999532 | | 2.82524 | 0.466979 | 0.722845 | | 2.609804 | 0.221659 | 0.89433 |
| SH3GL1 | 6.150559 | -0.23401 | 0.999532 | | 6.774425 | -0.31866 | 0.722845 | | 5.131364 | -0.07065 | 0.967145 |
| TMEM65 | 2.354502 | -0.39045 | 0.999532 | | 2.649541 | -0.51428 | 0.722965 | | 1.912276 | -0.10559 | 0.966344 |
| FAM90A1 | 10.25394 | 0.471219 | 0.999532 | | 8.570687 | -0.3301 | 0.722965 | | 9.026665 | -0.78952 | 0.282041 |
| MIER1 | 5.677158 | -0.14632 | 0.999532 | | 5.860101 | -0.32885 | 0.722965 | | 4.733044 | -0.16621 | 0.894612 |
| H2AC4 | 2.861457 | -0.33669 | 0.999532 | | 3.063377 | -0.47078 | 0.722965 | | 2.392189 | -0.1138 | 0.958698 |
| HYAL4 | 2.106038 | -0.09351 | 0.999532 | | 2.22876 | -0.53994 | 0.722965 | | 1.807103 | -0.41843 | 0.823193 |
| ZNF396 | 2.790321 | 0.451607 | 0.999532 | | 2.15697 | -0.51867 | 0.722965 | | 2.315798 | -0.9529 | 0.446959 |
| ZDHHC2 | 3.260612 | 0.079792 | 0.999532 | | 3.254324 | -0.47112 | 0.722965 | | 2.672673 | -0.55981 | 0.666555 |
| RNF212 | 2.821566 | -0.06739 | 0.999532 | | 2.865387 | -0.49572 | 0.722965 | | 2.28641 | -0.40246 | 0.803337 |
| KLF17 | 2.380671 | -0.12462 | 0.999532 | | 2.453359 | -0.52044 | 0.722965 | | 2.015205 | -0.37561 | 0.839373 |
| CWH43 | 2.437302 | -0.53159 | 0.999532 | | 2.849121 | -0.49367 | 0.722965 | | 1.901984 | 0.058842 | 0.984155 |
| BNC1 | 2.221695 | 0.111267 | 0.999532 | | 2.0931 | -0.54231 | 0.722965 | | 1.707876 | -0.63168 | 0.686214 |
| CENPN | 2.968635 | 0.240526 | 0.999532 | | 2.432793 | -0.49953 | 0.722965 | | 2.496464 | -0.71572 | 0.647218 |
| PIGW | 3.376656 | 0.361844 | 0.999532 | | 2.886615 | -0.44211 | 0.722965 | | 2.840754 | -0.78415 | 0.481527 |
| CD164L2 | 2.280074 | -0.20437 | 0.999532 | | 2.257327 | -0.5033 | 0.722965 | | 1.931624 | -0.28407 | 0.88476 |
| OR2T34 | 2.671213 | 0.025544 | 0.999532 | | 2.898486 | -0.46843 | 0.722965 | | 2.280737 | -0.46056 | 0.75311 |
| IRF6 | 2.457669 | -0.23976 | 0.999532 | | 2.589399 | -0.46059 | 0.722965 | | 1.925711 | -0.20084 | 0.914929 |
| IRGC | 3.107762 | 0.072861 | 0.999532 | | 2.866683 | -0.46389 | 0.722965 | | 2.629515 | -0.52875 | 0.713076 |
| OR51E1 | 2.331082 | -0.17076 | 0.999532 | | 2.28203 | -0.50796 | 0.723115 | | 2.046952 | -0.32715 | 0.861119 |
| ACYP1 | 2.204218 | -0.42278 | 0.999532 | | 2.447808 | -0.50149 | 0.723115 | | 1.845697 | -0.0595 | 0.984624 |
| RNF122 | 3.493764 | -0.37281 | 0.999532 | | 3.881114 | -0.40089 | 0.723115 | | 2.815533 | -0.00912 | 0.996891 |
| TJP1 | 22.11464 | 0.230832 | 0.999532 | | 24.57959 | 0.214558 | 0.723275 | | 22.27269 | -0.004 | 0.997967 |
| KLF16 | 5.929839 | 0.035136 | 0.999532 | | 7.529206 | 0.307389 | 0.723275 | | 6.474427 | 0.292612 | 0.766251 |
| HLA-DPA | 15.58768 | -0.17347 | 0.999532 | | 20.49654 | 0.309101 | 0.723596 | | 16.69865 | 0.500717 | 0.526728 |
| SFMBT1 | 2.599133 | 0.65821 | 0.999532 | | 2.703427 | 0.456262 | 0.723608 | | 2.897283 | -0.18322 | 0.906891 |
| SEPTIN9 | 9.263418 | -0.15631 | 0.999532 | | 11.82261 | 0.245945 | 0.723608 | | 9.655523 | 0.417154 | 0.563729 |
| NBDY | 3.516907 | 0.348684 | 0.999532 | | 4.078143 | 0.403852 | 0.723608 | | 3.783549 | 0.077995 | 0.965649 |
| SPANXN2 | 2.330366 | -0.00536 | 0.999532 | | 2.212985 | -0.51933 | 0.723763 | | 1.967774 | -0.4993 | 0.761337 |
| CLIC1 | 17.49574 | -0.16905 | 0.999532 | | 23.13041 | 0.300107 | 0.723763 | | 18.56712 | 0.494439 | 0.540342 |
| FRMD1 | 2.686131 | -0.08467 | 0.999532 | | 2.578042 | -0.48097 | 0.723887 | | 2.179637 | -0.37718 | 0.815713 |
| ADAMTS | 2.83683 | -0.24551 | 0.999532 | | 2.944432 | -0.442 | 0.723936 | | 2.426548 | -0.17941 | 0.920259 |
| RUSC2 | 6.927734 | 0.297499 | 0.999532 | | 7.55265 | 0.297056 | 0.724085 | | 7.132284 | 0.018164 | 0.992286 |
| C19orf47 | 2.31135 | 0.245446 | 0.999532 | | 2.904605 | 0.453624 | 0.724088 | | 2.612375 | 0.227298 | 0.892004 |
| HLA-DQB | 3.145727 | 0.066865 | 0.999532 | | 3.679938 | 0.400881 | 0.72428 | | 3.368381 | 0.353231 | 0.792539 |
| COQ6 | 2.412012 | -0.13079 | 0.999532 | | 2.444194 | -0.49674 | 0.72428 | | 2.096854 | -0.34106 | 0.858705 |
| H2AC16 | 2.115794 | -0.75781 | 0.999532 | | 2.428322 | -0.4972 | 0.724892 | | 1.637106 | 0.279788 | 0.892316 |
| ZNF345 | 2.450745 | -0.25868 | 0.999532 | | 2.498459 | -0.48567 | 0.724892 | | 2.08055 | -0.20908 | 0.918637 |
| EVI2B | 4.567284 | -0.103 | 0.999532 | | 4.593437 | -0.37135 | 0.724892 | | 3.943474 | -0.26177 | 0.864923 |
| CCRL2 | 3.081127 | 0.079877 | 0.999532 | | 2.821934 | -0.45866 | 0.724892 | | 2.812792 | -0.51097 | 0.714749 |
| FMR1NB | 2.283542 | -0.06198 | 0.999532 | | 2.341191 | -0.51313 | 0.724892 | | 2.018718 | -0.43287 | 0.803301 |
| CFAP65 | 2.417023 | 0.068422 | 0.999532 | | 2.376274 | -0.48587 | 0.724892 | | 2.049112 | -0.53047 | 0.722703 |
| FOLH1 | 2.856905 | 0.114719 | 0.999532 | | 2.81877 | -0.45508 | 0.724892 | | 2.429898 | -0.5479 | 0.682485 |
| NSUN3 | 2.304777 | -0.0182 | 0.999532 | | 2.949894 | 0.445308 | 0.724892 | | 2.555384 | 0.47795 | 0.736481 |
| HECA | 4.542326 | -0.10506 | 0.999532 | | 4.626945 | -0.3698 | 0.724892 | | 3.658152 | -0.23871 | 0.859898 |
| TRAPPC1 | 4.270569 | -0.22753 | 0.999532 | | 6.048223 | 0.322004 | 0.724892 | | 4.539044 | 0.551712 | 0.552672 |
| GIPR | 2.432661 | -0.01417 | 0.999532 | | 2.442971 | -0.48725 | 0.724892 | | 2.13528 | -0.45373 | 0.775843 |
| ABI3BP | 2.847697 | 0.288358 | 0.999532 | | 3.434109 | 0.412711 | 0.724892 | | 3.149172 | 0.152249 | 0.934572 |
| GPR32 | 3.127757 | 0.230545 | 0.999532 | | 2.696265 | -0.48802 | 0.724892 | | 2.705956 | -0.69883 | 0.623462 |
| KIF20B | 2.444079 | 0.490446 | 0.999532 | | 2.053472 | -0.53303 | 0.724892 | | 2.135615 | -1.00447 | 0.421517 |
| ELL | 3.459037 | -0.06998 | 0.999532 | | 3.359695 | -0.41088 | 0.724892 | | 2.812729 | -0.32549 | 0.817471 |
| NKX1-2 | 2.945504 | -0.16007 | 0.999532 | | 3.083216 | -0.45836 | 0.724892 | | 2.470174 | -0.27993 | 0.867739 |
| BICC1 | 2.129547 | -0.19379 | 0.999532 | | 2.890688 | 0.446345 | 0.724892 | | 2.234499 | 0.659434 | 0.630705 |
| C11orf49 | 3.419219 | 0.488832 | 0.999532 | | 3.619401 | 0.418793 | 0.724892 | | 3.713071 | -0.05375 | 0.979239 |
| TPST1 | 1.952832 | -0.34365 | 0.999532 | | 2.671836 | 0.448553 | 0.724892 | | 2.266906 | 0.809549 | 0.542124 |
| TSC22D4 | 3.598278 | 0.184783 | 0.999532 | | 4.379837 | 0.361507 | 0.724892 | | 3.887468 | 0.197846 | 0.88395 |
| MAVS | 5.053471 | -0.04698 | 0.999532 | | 6.290584 | 0.308456 | 0.724892 | | 5.246203 | 0.368985 | 0.705425 |
| DHRS7B | 3.927453 | 0.224012 | 0.999532 | | 3.740718 | -0.41229 | 0.724892 | | 3.53246 | -0.61253 | 0.619423 |
| AHCTF1 | 4.333691 | 0.10039 | 0.999532 | | 5.314655 | 0.347781 | 0.724892 | | 4.621792 | 0.266722 | 0.819337 |
| SETD6 | 2.814788 | -0.26713 | 0.999532 | | 2.895583 | -0.44729 | 0.724892 | | 2.291825 | -0.164 | 0.931488 |
| RIPK1 | 4.303097 | -0.51777 | 0.999532 | | 5.165581 | -0.33161 | 0.724892 | | 3.505656 | 0.201959 | 0.883679 |
| GRP | 2.36354 | 0.248412 | 0.999532 | | 1.999435 | -0.55521 | 0.724892 | | 1.916736 | -0.78613 | 0.613126 |
| SLCO2B1 | 3.922547 | 0.43854 | 0.999532 | | 4.309368 | 0.380868 | 0.724892 | | 4.30659 | -0.03796 | 0.985693 |
| TAS2R39 | 2.191288 | 0.647724 | 0.999532 | | 1.667566 | -0.57354 | 0.724892 | | 1.878602 | -1.20503 | 0.314028 |
| CENPB | 5.45239 | 0.031694 | 0.999532 | | 6.773176 | 0.327803 | 0.724892 | | 5.722896 | 0.319281 | 0.752472 |
| SLC25A1 | 3.185663 | -0.66844 | 0.999532 | | 3.666834 | -0.39069 | 0.724892 | | 2.684707 | 0.29262 | 0.852762 |
| KLF5 | 2.929494 | 0.178915 | 0.999532 | | 2.631095 | -0.46701 | 0.724892 | | 2.517201 | -0.62854 | 0.649468 |
| MTFR1 | 3.00664 | -0.20539 | 0.999532 | | 4.468798 | 0.362328 | 0.724892 | | 3.355595 | 0.586144 | 0.603483 |
| OLFML2A | 6.660425 | 0.44698 | 0.999532 | | 6.821628 | 0.308037 | 0.724892 | | 6.84563 | -0.13512 | 0.919002 |
| TCN2 | 3.620407 | 0.075368 | 0.999532 | | 4.50034 | 0.345924 | 0.724892 | | 3.884933 | 0.288555 | 0.810635 |
| AOC3 | 3.53672 | -0.00245 | 0.999532 | | 4.490738 | 0.386886 | 0.724892 | | 3.706151 | 0.416709 | 0.726599 |
| ZFHX3 | 2.298864 | 0.17412 | 0.999532 | | 2.909016 | 0.427095 | 0.724892 | | 2.666966 | 0.273322 | 0.858767 |
| SPRR4 | 2.295564 | -0.18413 | 0.999532 | | 2.301208 | -0.52872 | 0.724892 | | 1.900807 | -0.33338 | 0.865065 |
| ACAP3 | 4.760298 | -0.03394 | 0.999532 | | 5.004215 | -0.37525 | 0.724892 | | 4.248564 | -0.32792 | 0.787104 |
| AANAT | 2.922468 | -0.22787 | 0.999532 | | 2.919828 | -0.4652 | 0.724892 | | 2.462371 | -0.21879 | 0.909074 |
| TXNDC8 | 2.511636 | -0.15392 | 0.999532 | | 2.547172 | -0.48488 | 0.724892 | | 2.177409 | -0.31653 | 0.863134 |
| MCM5 | 3.117759 | 0.760031 | 0.999532 | | 2.884428 | 0.445789 | 0.724892 | | 3.210049 | -0.29527 | 0.834173 |
| IGSF22 | 2.396605 | 0.380016 | 0.999532 | | 2.08972 | -0.53401 | 0.724892 | | 2.078195 | -0.88686 | 0.519358 |
| MRPL52 | 4.22106 | 0.596624 | 0.999532 | | 4.239947 | 0.359605 | 0.724892 | | 4.468165 | -0.22018 | 0.863134 |
| MLLT1 | 5.120087 | -0.20279 | 0.999532 | | 6.780903 | 0.305494 | 0.724892 | | 5.383029 | 0.527149 | 0.542124 |
| RAB3GAI | 3.654436 | -0.06572 | 0.999532 | | 3.548497 | -0.4212 | 0.724892 | | 2.947002 | -0.33181 | 0.811123 |
| CCDC117 | 4.241935 | 0.148063 | 0.999532 | | 5.179681 | 0.350913 | 0.724892 | | 4.774447 | 0.222006 | 0.864923 |
| KIAA1614 | 4.024211 | 0.26806 | 0.999532 | | 3.733573 | -0.40879 | 0.724892 | | 3.47796 | -0.65078 | 0.563627 |
| C1orf210 | 2.421535 | 0.261752 | 0.999532 | | 2.015201 | -0.53611 | 0.724892 | | 2.083201 | -0.77896 | 0.602786 |
| PLAGL2 | 2.884461 | -0.49436 | 0.999532 | | 3.37575 | -0.43695 | 0.724892 | | 2.397592 | 0.076437 | 0.974421 |
| PPP1CC | 8.266188 | 0.036803 | 0.999532 | | 10.29222 | 0.264671 | 0.724892 | | 8.730778 | 0.246333 | 0.783005 |
| C1orf116 | 9.041414 | 0.225878 | 0.999532 | | 8.861616 | -0.32096 | 0.724892 | | 8.139043 | -0.52005 | 0.532278 |
| NAPG | 2.663426 | -0.11922 | 0.999532 | | 3.502096 | 0.401191 | 0.724892 | | 2.872927 | 0.535407 | 0.665886 |
| PPP1R10 | 4.736391 | 0.045936 | 0.999532 | | 6.092252 | 0.366247 | 0.724892 | | 5.118083 | 0.347398 | 0.742839 |
| TAC4 | 2.251366 | -0.09148 | 0.999532 | | 2.174833 | -0.50429 | 0.724892 | | 1.872253 | -0.39499 | 0.826947 |
| SLAMF8 | 2.531437 | -0.11812 | 0.999532 | | 2.568338 | -0.46014 | 0.724892 | | 2.1813 | -0.32044 | 0.854633 |
| MTARC1 | 3.279429 | -0.15116 | 0.999532 | | 3.537706 | -0.44037 | 0.724892 | | 2.880662 | -0.27227 | 0.865363 |
| NUDT4B | 7.72231 | 0.046294 | 0.999532 | | 7.828248 | -0.31204 | 0.724892 | | 6.407464 | -0.34366 | 0.734538 |
| HRNR | 2.400802 | -0.2126 | 0.999532 | | 2.550704 | -0.47517 | 0.724892 | | 1.925013 | -0.24214 | 0.89901 |
| HNRNPR | 9.31808 | -0.08289 | 0.999532 | | 12.18413 | 0.292197 | 0.724892 | | 10.07941 | 0.391385 | 0.628895 |
| KLHL42 | 3.768747 | 0.171619 | 0.999532 | | 3.767456 | -0.39793 | 0.724892 | | 3.326583 | -0.54523 | 0.635045 |
| SMARCE | 12.99053 | -0.32224 | 0.999532 | | 17.33217 | 0.224275 | 0.724892 | | 13.47636 | 0.566212 | 0.274496 |
| ZSCAN22 | 2.618598 | 0.111008 | 0.999532 | | 2.426957 | -0.48228 | 0.724892 | | 2.138171 | -0.57548 | 0.692534 |
| ZNF92 | 1.901403 | -0.10109 | 0.999532 | | 1.804054 | -0.5555 | 0.724892 | | 1.562268 | -0.43674 | 0.821953 |
| SKIDA1 | 2.107907 | 0.135683 | 0.999532 | | 1.890148 | -0.54663 | 0.724892 | | 1.710927 | -0.66388 | 0.672739 |
| PRRT1 | 2.576087 | 0.176051 | 0.999532 | | 3.220289 | 0.438101 | 0.724892 | | 2.892784 | 0.279821 | 0.858426 |
| UBE2Q2 | 3.148259 | -0.12288 | 0.999532 | | 4.112569 | 0.380314 | 0.724892 | | 3.413814 | 0.513187 | 0.678411 |
| RPAIN | 3.416933 | 0.069615 | 0.999532 | | 4.236646 | 0.360796 | 0.724892 | | 3.663448 | 0.309748 | 0.800239 |
| TMEM255 | 4.849441 | -0.45734 | 0.999532 | | 5.824099 | -0.3299 | 0.724892 | | 4.341251 | 0.145386 | 0.918194 |
| PRKCZ | 4.603021 | 0.737377 | 0.999532 | | 4.354415 | 0.374458 | 0.724892 | | 4.628333 | -0.35104 | 0.737109 |
| CMTM3 | 3.322513 | -0.57996 | 0.999532 | | 4.779356 | 0.410283 | 0.724892 | | 3.533755 | 1.007308 | 0.310637 |
| NKAP | 2.013489 | 0.502263 | 0.999532 | | 2.103198 | 0.505569 | 0.724892 | | 2.210764 | 0.026143 | 0.993618 |
| RAB42 | 2.255484 | -0.39257 | 0.999532 | | 2.541834 | -0.52535 | 0.724892 | | 1.904573 | -0.11675 | 0.964769 |
| THEM5 | 5.441132 | 0.562048 | 0.999532 | | 5.924021 | 0.351977 | 0.724892 | | 5.949099 | -0.18568 | 0.881837 |
| CIR1 | 5.074637 | -0.282 | 0.999532 | | 5.480869 | -0.35997 | 0.724892 | | 4.281088 | -0.05305 | 0.976783 |
| KBTBD4 | 2.479157 | -0.13204 | 0.999532 | | 3.290571 | 0.411986 | 0.724892 | | 2.769997 | 0.564803 | 0.660097 |
| RAB11FII | 5.306821 | 0.03942 | 0.999532 | | 5.382838 | -0.35451 | 0.724892 | | 4.490203 | -0.37985 | 0.724411 |
| DNAJA2 | 3.588708 | -0.36589 | 0.999532 | | 5.077665 | 0.355769 | 0.724892 | | 4.012532 | 0.738967 | 0.444687 |
| BBLN | 6.266163 | 0.100242 | 0.999532 | | 7.321575 | 0.280335 | 0.724892 | | 6.567395 | 0.194 | 0.852482 |
| FAM71D | 2.635751 | 0.015945 | 0.999532 | | 2.497838 | -0.4747 | 0.725131 | | 2.14892 | -0.47252 | 0.752299 |
| XYLT1 | 1.836856 | -0.00183 | 0.999532 | | 2.461439 | 0.471257 | 0.725131 | | 2.087982 | 0.490493 | 0.740755 |
| HOXB7 | 3.248264 | 0.134855 | 0.999532 | | 3.07154 | -0.42146 | 0.725131 | | 2.788093 | -0.53793 | 0.662584 |
| RNASEH2 | 7.541171 | 0.018195 | 0.999532 | | 7.933256 | -0.27824 | 0.725468 | | 6.705486 | -0.27237 | 0.764553 |
| TCEA3 | 6.889681 | 0.004295 | 0.999532 | | 8.563271 | 0.286607 | 0.725937 | | 7.021209 | 0.281567 | 0.760473 |
| SLC35E4 | 6.215966 | 0.048779 | 0.999532 | | 6.380226 | -0.37135 | 0.725937 | | 5.432147 | -0.3978 | 0.718643 |
| RAD9B | 2.746808 | 0.102541 | 0.999532 | | 2.667015 | -0.47928 | 0.725937 | | 2.286176 | -0.55883 | 0.695252 |
| AMPD3 | 2.21077 | 0.302099 | 0.999532 | | 1.946302 | -0.5394 | 0.725937 | | 1.837892 | -0.82272 | 0.566578 |
| HAVCR1 | 1.918964 | 0.633015 | 0.999532 | | 1.92686 | 0.547425 | 0.725937 | | 2.062733 | -0.06921 | 0.978472 |
| DOCK8 | 2.442511 | -0.16601 | 0.999532 | | 3.312452 | 0.418417 | 0.725937 | | 2.898989 | 0.611407 | 0.636184 |
| DNMT3B | 1.553127 | 0.187833 | 0.999532 | | 1.905356 | 0.531244 | 0.726341 | | 1.752331 | 0.36067 | 0.849682 |
| SNX17 | 6.613736 | -0.14766 | 0.999532 | | 9.057424 | 0.280994 | 0.726341 | | 7.124362 | 0.458982 | 0.595502 |
| COPRS | 5.222139 | 0.094995 | 0.999532 | | 6.283897 | 0.32759 | 0.726341 | | 5.491809 | 0.251505 | 0.813172 |
| PTPN1 | 5.793775 | 0.242815 | 0.999532 | | 6.816353 | 0.290776 | 0.726341 | | 6.167579 | 0.0644 | 0.961784 |
| TUBB2A | 6.102448 | -0.07895 | 0.999532 | | 7.914638 | 0.353758 | 0.726689 | | 6.337584 | 0.448669 | 0.602151 |
| HARBI1 | 3.101985 | 0.033739 | 0.999532 | | 3.137321 | -0.44758 | 0.727331 | | 2.693399 | -0.45711 | 0.746876 |
| U2AF1L4 | 4.575741 | -0.07749 | 0.999532 | | 4.765587 | -0.36466 | 0.727421 | | 3.895354 | -0.26516 | 0.836935 |
| CCN3 | 3.150468 | 0.154196 | 0.999532 | | 2.847359 | -0.46458 | 0.727439 | | 2.637791 | -0.60609 | 0.652925 |
| IQCA1 | 2.262547 | -0.04533 | 0.999532 | | 2.213403 | -0.52483 | 0.727521 | | 1.869934 | -0.46128 | 0.791576 |
| SPATA6L | 2.819654 | -0.13999 | 0.999532 | | 2.981665 | -0.45935 | 0.727598 | | 2.302072 | -0.30111 | 0.859426 |
| STRADA | 3.511914 | 0.247287 | 0.999532 | | 4.031016 | 0.373581 | 0.72761 | | 3.761315 | 0.143491 | 0.923266 |
| PRAMEF | 2.461749 | 0.505086 | 0.999532 | | 1.932641 | -0.5394 | 0.72761 | | 2.061179 | -1.02838 | 0.421517 |
| HMGN4 | 4.62534 | -0.01657 | 0.999532 | | 5.972422 | 0.330419 | 0.72761 | | 4.933931 | 0.370176 | 0.734538 |
| ZNF512B | 3.034504 | 0.19924 | 0.999532 | | 2.761346 | -0.45982 | 0.72761 | | 2.519609 | -0.63985 | 0.630705 |
| MED25 | 2.451346 | -0.16844 | 0.999532 | | 3.197975 | 0.41223 | 0.727776 | | 2.670343 | 0.594258 | 0.647581 |
| ZBTB21 | 4.010402 | -0.12061 | 0.999532 | | 4.067102 | -0.3928 | 0.727776 | | 3.429685 | -0.2509 | 0.859898 |
| PRKAB1 | 2.72846 | 0.228126 | 0.999532 | | 3.336057 | 0.422867 | 0.727967 | | 2.857941 | 0.220624 | 0.88588 |
| HOXD1 | 2.761911 | 0.067509 | 0.999532 | | 2.694649 | -0.47891 | 0.727967 | | 2.35389 | -0.52054 | 0.725266 |
| ARID4A | 2.092981 | 0.2109 | 0.999532 | | 2.77173 | 0.494037 | 0.728059 | | 2.552939 | 0.293323 | 0.868512 |
| TNRC6A | 5.575678 | -0.17805 | 0.999532 | | 7.496588 | 0.305096 | 0.728059 | | 5.917784 | 0.509984 | 0.563285 |
| TMEM185 | 2.602391 | -0.18812 | 0.999532 | | 2.622644 | -0.46861 | 0.728059 | | 2.104146 | -0.26149 | 0.88885 |
| GPR25 | 2.632193 | -0.09575 | 0.999532 | | 2.482496 | -0.48241 | 0.728555 | | 2.112874 | -0.36865 | 0.829716 |
| RSAD1 | 3.282919 | -0.0488 | 0.999532 | | 3.224399 | -0.43414 | 0.728676 | | 2.913619 | -0.37267 | 0.810635 |
| SLC29A3 | 2.25793 | 0.19313 | 0.999532 | | 2.057147 | -0.53793 | 0.728676 | | 2.046859 | -0.70196 | 0.672452 |
| C5 | 2.280609 | -0.1977 | 0.999532 | | 2.296168 | -0.49892 | 0.728721 | | 1.859086 | -0.28489 | 0.883792 |
| CATSPER | 2.176611 | 0.444422 | 0.999532 | | 2.426933 | 0.477956 | 0.728938 | | 2.41762 | 0.051973 | 0.984587 |
| LUZP1 | 3.517565 | 0.02726 | 0.999532 | | 4.669635 | 0.36753 | 0.728938 | | 3.982956 | 0.36735 | 0.760473 |
| GRSF1 | 3.097474 | 0.288074 | 0.999532 | | 2.655989 | -0.45762 | 0.729146 | | 2.669452 | -0.72871 | 0.540342 |
| PCIF1 | 3.726977 | 0.238664 | 0.999532 | | 3.301186 | -0.41358 | 0.729146 | | 3.097885 | -0.63536 | 0.565022 |
| RTN4R | 2.683004 | 0.076659 | 0.999532 | | 2.584922 | -0.47762 | 0.729146 | | 2.419361 | -0.54169 | 0.714657 |
| DBT | 4.071315 | 0.148902 | 0.999532 | | 5.10596 | 0.33953 | 0.729146 | | 4.475715 | 0.209119 | 0.870663 |
| TCP11L1 | 2.806263 | -0.11607 | 0.999532 | | 3.629778 | 0.396109 | 0.729146 | | 3.010798 | 0.530034 | 0.664325 |
| VMAC | 2.599158 | 0.34241 | 0.999532 | | 2.238061 | -0.51184 | 0.729146 | | 2.153263 | -0.8328 | 0.542124 |
| CCDC28B | 2.927657 | 0.023044 | 0.999532 | | 2.715148 | -0.45113 | 0.729146 | | 2.464159 | -0.46388 | 0.75311 |
| TAS1R2 | 2.530569 | 0.21327 | 0.999532 | | 2.143063 | -0.5245 | 0.729146 | | 2.103792 | -0.71862 | 0.637292 |
| TMTC3 | 2.840818 | -0.20425 | 0.999532 | | 4.101456 | 0.377943 | 0.729146 | | 3.303612 | 0.608147 | 0.628053 |
| MORC3 | 2.762639 | 0.531867 | 0.999532 | | 3.005205 | 0.420802 | 0.729146 | | 3.05929 | -0.09267 | 0.959765 |
| TRIM35 | 2.749878 | -0.23496 | 0.999532 | | 2.906888 | -0.44256 | 0.729146 | | 2.150818 | -0.19273 | 0.916361 |
| ATP7B | 1.910203 | 0.103346 | 0.999532 | | 2.551815 | 0.48258 | 0.729146 | | 2.232771 | 0.401492 | 0.809832 |
| NAP1L1 | 18.97975 | -0.20745 | 0.999532 | | 24.72478 | 0.207547 | 0.729146 | | 19.42178 | 0.440205 | 0.462548 |
| NCKAP1 | 6.387127 | -0.21982 | 0.999532 | | 8.676116 | 0.30147 | 0.729181 | | 6.802898 | 0.547307 | 0.530351 |
| SERPINA | 3.839959 | 0.138315 | 0.999532 | | 3.777657 | -0.40715 | 0.72919 | | 3.144145 | -0.52339 | 0.717724 |
| SHISA6 | 2.319279 | -0.3254 | 0.999532 | | 2.491446 | -0.49061 | 0.72919 | | 1.971969 | -0.1503 | 0.948127 |
| LGI3 | 3.118487 | 0.053784 | 0.999532 | | 2.905682 | -0.45524 | 0.72919 | | 2.746698 | -0.49559 | 0.728015 |
| DNAJA4 | 2.365872 | 0.640732 | 0.999532 | | 2.448523 | 0.478058 | 0.72919 | | 2.600155 | -0.14564 | 0.938624 |
| UQCRQ | 8.737198 | -0.27379 | 0.999532 | | 11.6028 | 0.246019 | 0.730156 | | 8.926203 | 0.534766 | 0.411012 |
| PLAUR | 3.007089 | 0.299781 | 0.999532 | | 2.514174 | -0.46164 | 0.730156 | | 2.602677 | -0.75104 | 0.541136 |
| MNX1 | 2.352548 | 0.571031 | 0.999532 | | 1.821827 | -0.54716 | 0.730156 | | 2.038987 | -1.10117 | 0.386771 |
| PI4KA | 4.013506 | -0.07374 | 0.999532 | | 5.295353 | 0.348209 | 0.730156 | | 4.218838 | 0.441903 | 0.695561 |
| AUP1 | 7.650735 | -0.08317 | 0.999532 | | 9.292778 | 0.282233 | 0.730156 | | 7.851285 | 0.376742 | 0.66232 |
| VAMP5 | 9.422548 | 0.279151 | 0.999532 | | 10.76692 | 0.251087 | 0.730156 | | 10.12475 | -0.01031 | 0.996681 |
| MRPL14 | 4.110111 | -0.14566 | 0.999532 | | 4.077268 | -0.38857 | 0.730156 | | 3.178385 | -0.22322 | 0.880577 |
| DHCR24 | 3.272551 | 0.048052 | 0.999532 | | 4.01585 | 0.400578 | 0.730156 | | 3.581723 | 0.357336 | 0.791576 |
| LANCL2 | 2.405345 | 0.830386 | 0.999532 | | 2.291392 | 0.480651 | 0.730156 | | 2.568946 | -0.33423 | 0.823571 |
| SLC29A1 | 2.0493 | 0.593602 | 0.999532 | | 2.239546 | 0.499536 | 0.730342 | | 2.295385 | -0.07321 | 0.974572 |
| CBR1 | 6.972487 | 0.022074 | 0.999532 | | 8.615776 | 0.294852 | 0.730342 | | 7.483164 | 0.287642 | 0.752562 |
| SCUBE3 | 2.22158 | 0.033921 | 0.999532 | | 2.07797 | -0.53969 | 0.73043 | | 1.85612 | -0.55618 | 0.744727 |
| ZNF721 | 10.00639 | 0.007129 | 0.999532 | | 10.08056 | -0.28594 | 0.73043 | | 8.427822 | -0.25736 | 0.75311 |
| WHRN | 4.056012 | -0.12227 | 0.999532 | | 4.18937 | -0.39029 | 0.73043 | | 3.545014 | -0.24026 | 0.882613 |
| PPAT | 3.817202 | -0.03113 | 0.999532 | | 3.887035 | -0.42282 | 0.73043 | | 3.206604 | -0.3702 | 0.791576 |
| DAPK1 | 4.584704 | 0.249377 | 0.999532 | | 5.176451 | 0.330082 | 0.73043 | | 4.76816 | 0.094686 | 0.948127 |
| ETAA1 | 2.434773 | 0.560875 | 0.999532 | | 2.702044 | 0.457818 | 0.73043 | | 2.808691 | -0.08465 | 0.968434 |
| ERLEC1 | 3.449938 | -0.06436 | 0.999532 | | 4.222712 | 0.38451 | 0.73043 | | 3.710286 | 0.452686 | 0.706749 |
| KLHL18 | 1.942475 | -0.5951 | 0.999532 | | 2.250936 | -0.50855 | 0.73043 | | 1.720012 | 0.104292 | 0.971732 |
| GNB2 | 11.6402 | -0.06245 | 0.999532 | | 14.4155 | 0.23056 | 0.73043 | | 11.9464 | 0.309315 | 0.663009 |
| ATP13A5 | 2.275003 | -0.16617 | 0.999532 | | 2.440802 | -0.47493 | 0.73043 | | 1.886935 | -0.28656 | 0.877703 |
| TBX4 | 2.247747 | -0.26994 | 0.999532 | | 2.257836 | -0.50457 | 0.730489 | | 1.801311 | -0.22189 | 0.917499 |
| ENOSF1 | 7.55071 | 0.446461 | 0.999532 | | 8.107941 | 0.310153 | 0.730489 | | 7.634613 | -0.10949 | 0.926957 |
| TMEM265 | 2.46514 | 0.125229 | 0.999532 | | 2.363187 | -0.49621 | 0.730742 | | 2.088173 | -0.60199 | 0.68528 |
| RPL10L | 7.560938 | 0.369836 | 0.999532 | | 8.413169 | 0.330787 | 0.730811 | | 8.032793 | -0.01502 | 0.995112 |
| IQSEC2 | 2.554006 | -0.17587 | 0.999532 | | 2.650401 | -0.4518 | 0.730835 | | 2.048556 | -0.25574 | 0.886495 |
| SPACA6 | 2.818468 | 0.145859 | 0.999532 | | 2.62681 | -0.49372 | 0.730975 | | 2.373555 | -0.6199 | 0.665886 |
| GALNT5 | 2.264935 | 0.061193 | 0.999532 | | 2.214204 | -0.51851 | 0.730975 | | 1.92871 | -0.55942 | 0.728015 |
| PPCS | 4.471339 | 0.372847 | 0.999532 | | 4.557441 | 0.339294 | 0.731162 | | 4.663007 | -0.0259 | 0.991159 |
| TNFSF10 | 8.512175 | -0.00279 | 0.999532 | | 10.75113 | 0.314299 | 0.731305 | | 9.273036 | 0.336698 | 0.668553 |
| PKIG | 6.494858 | 0.054478 | 0.999532 | | 8.041184 | 0.306798 | 0.731305 | | 7.002101 | 0.272079 | 0.791576 |
| HSPA4 | 4.974148 | -0.05701 | 0.999532 | | 6.536699 | 0.322308 | 0.731469 | | 5.214794 | 0.418527 | 0.686214 |
| RFX1 | 3.675545 | 0.015783 | 0.999532 | | 3.482747 | -0.40098 | 0.731472 | | 3.074701 | -0.4035 | 0.755301 |
| KCNJ3 | 2.711081 | -0.22201 | 0.999532 | | 2.757563 | -0.44756 | 0.731472 | | 2.201395 | -0.22043 | 0.907968 |
| TACC3 | 2.304246 | -0.45484 | 0.999532 | | 2.603082 | -0.45682 | 0.731472 | | 1.898519 | 0.016801 | 0.996681 |
| ATP6V1E | 2.668313 | -0.17184 | 0.999532 | | 2.788073 | -0.45344 | 0.731472 | | 2.194135 | -0.26155 | 0.882936 |
| STAT2 | 5.910055 | 0.076389 | 0.999532 | | 7.193018 | 0.283632 | 0.731472 | | 6.394824 | 0.226568 | 0.816526 |
| UBA2 | 5.388397 | -0.21731 | 0.999532 | | 7.079217 | 0.334387 | 0.731749 | | 5.66308 | 0.581281 | 0.515762 |
| GIP | 3.749599 | 0.215165 | 0.999532 | | 3.579162 | -0.43988 | 0.731849 | | 3.303108 | -0.63308 | 0.623565 |
| TUBE1 | 2.974761 | 0.040879 | 0.999532 | | 3.898037 | 0.406262 | 0.731849 | | 3.238709 | 0.393831 | 0.761337 |
| ODAD4 | 1.784196 | 0.596133 | 0.999532 | | 1.96696 | 0.526892 | 0.731849 | | 1.972151 | -0.0526 | 0.985693 |
| PIAS2 | 8.42621 | 0.158172 | 0.999532 | | 9.785904 | 0.245519 | 0.731849 | | 8.736218 | 0.106424 | 0.914929 |
| CREBBP | 8.532466 | 0.008558 | 0.999532 | | 10.91514 | 0.270466 | 0.731849 | | 9.137393 | 0.298051 | 0.739496 |
| LPCAT3 | 3.516948 | 0.353572 | 0.999532 | | 3.872508 | 0.392508 | 0.732091 | | 3.809085 | 0.05487 | 0.977635 |
| NT5C | 3.627809 | 0.388946 | 0.999532 | | 4.088208 | 0.379603 | 0.732391 | | 4.127917 | 0.008309 | 0.996891 |
| VASH1 | 3.21786 | 0.153709 | 0.999532 | | 3.724308 | 0.397504 | 0.732391 | | 3.665355 | 0.287611 | 0.842696 |
| RTCA | 2.794966 | 0.310577 | 0.999532 | | 3.216496 | 0.412962 | 0.732391 | | 2.918444 | 0.118663 | 0.946512 |
| RILPL2 | 6.041081 | 0.067786 | 0.999532 | | 7.432177 | 0.308148 | 0.732391 | | 6.438651 | 0.260369 | 0.792688 |
| ZNF436 | 3.346963 | 0.090251 | 0.999532 | | 3.119951 | -0.44356 | 0.732391 | | 2.874886 | -0.51718 | 0.686906 |
| GCN1 | 2.857185 | -0.45534 | 0.999532 | | 3.192466 | -0.41732 | 0.732391 | | 2.24917 | 0.055337 | 0.981929 |
| C1orf226 | 2.344566 | -0.19958 | 0.999532 | | 2.486923 | -0.49685 | 0.732391 | | 1.825524 | -0.27242 | 0.886495 |
| NMD3 | 2.970216 | 0.144188 | 0.999532 | | 3.484394 | 0.384878 | 0.732391 | | 3.220916 | 0.258033 | 0.862432 |
| SORBS3 | 14.25827 | 0.006959 | 0.999532 | | 17.01854 | 0.237143 | 0.732391 | | 14.84194 | 0.245976 | 0.747457 |
| ZKSCAN1 | 6.116752 | -0.14494 | 0.999532 | | 7.741221 | 0.301567 | 0.732391 | | 6.483175 | 0.450073 | 0.647218 |
| CYB5R4 | 2.377172 | -0.52311 | 0.999532 | | 2.741656 | -0.4578 | 0.732391 | | 1.841175 | 0.083168 | 0.974421 |
| IQGAP3 | 2.797044 | -0.27665 | 0.999532 | | 3.006471 | -0.4218 | 0.732391 | | 2.25928 | -0.12732 | 0.948127 |
| IL27 | 2.765994 | -0.18231 | 0.999532 | | 2.72619 | -0.44428 | 0.732391 | | 2.428915 | -0.24893 | 0.892646 |
| PCYOX1 | 4.945778 | 0.147118 | 0.999532 | | 5.832653 | 0.331549 | 0.732595 | | 5.042144 | 0.210766 | 0.858767 |
| WDR90 | 3.293483 | -0.05814 | 0.999532 | | 3.34064 | -0.42494 | 0.732595 | | 2.635226 | -0.3452 | 0.815713 |
| RARS2 | 4.311786 | 0.250274 | 0.999532 | | 4.787697 | 0.338989 | 0.732595 | | 4.538736 | 0.099475 | 0.946958 |
| C9orf72 | 1.989581 | 0.666748 | 0.999532 | | 2.207779 | 0.501648 | 0.732687 | | 2.253482 | -0.14759 | 0.940246 |
| STEAP2 | 3.276394 | 0.074503 | 0.999532 | | 3.057091 | -0.44268 | 0.732739 | | 2.715429 | -0.49871 | 0.713076 |
| OR9G4 | 2.690466 | -0.05105 | 0.999532 | | 2.687916 | -0.47647 | 0.732739 | | 2.355514 | -0.40802 | 0.806394 |
| CTDSPL | 6.217964 | 0.26965 | 0.999532 | | 7.033826 | 0.298279 | 0.732739 | | 6.40123 | 0.047604 | 0.974421 |
| NR0B2 | 2.346459 | -0.12594 | 0.999532 | | 2.274341 | -0.52516 | 0.732739 | | 1.881633 | -0.38029 | 0.839908 |
| ZZZ3 | 1.950081 | 0.449286 | 0.999532 | | 2.181717 | 0.49707 | 0.732739 | | 2.134212 | 0.067543 | 0.977746 |
| TRMT2B | 2.106593 | 0.410646 | 0.999532 | | 2.347631 | 0.471249 | 0.732739 | | 2.281294 | 0.077792 | 0.973356 |
| RRAS | 6.024688 | 0.228931 | 0.999532 | | 7.191184 | 0.325698 | 0.732739 | | 6.533665 | 0.111439 | 0.926957 |
| RPUSD2 | 2.729823 | 0.228813 | 0.999532 | | 2.400906 | -0.48557 | 0.732739 | | 2.359854 | -0.69633 | 0.619423 |
| MCPH1 | 2.858674 | 0.185434 | 0.999532 | | 2.578407 | -0.47775 | 0.732739 | | 2.347871 | -0.64392 | 0.652925 |
| PTAR1 | 2.021156 | -0.46534 | 0.999532 | | 2.28891 | -0.48295 | 0.732739 | | 1.729899 | 0.001272 | 0.999635 |
| MCCD1 | 3.712401 | -0.00817 | 0.999532 | | 3.682393 | -0.43201 | 0.733192 | | 3.222197 | -0.41023 | 0.771812 |
| NEK2 | 2.685741 | 0.337642 | 0.999532 | | 2.437669 | -0.47861 | 0.733249 | | 2.388838 | -0.79557 | 0.533876 |
| CYP2D6 | 4.829408 | 0.139546 | 0.999532 | | 4.745991 | -0.38362 | 0.733249 | | 4.188363 | -0.50434 | 0.652925 |
| ARHGAP | 3.464416 | -0.12377 | 0.999532 | | 3.606546 | -0.40485 | 0.733249 | | 2.915846 | -0.26164 | 0.865363 |
| DCK | 3.717043 | 0.130239 | 0.999532 | | 4.454251 | 0.351138 | 0.733249 | | 3.998154 | 0.233739 | 0.862147 |
| UBXN8 | 2.117811 | -0.1843 | 0.999532 | | 2.966339 | 0.422171 | 0.733249 | | 2.402411 | 0.625196 | 0.643959 |
| CCL25 | 2.098327 | -0.06892 | 0.999532 | | 1.97978 | -0.53095 | 0.733249 | | 1.761868 | -0.44762 | 0.811782 |
| LRRC43 | 2.156671 | 0.376878 | 0.999532 | | 2.468918 | 0.483304 | 0.733249 | | 2.296882 | 0.124168 | 0.952656 |
| SLC44A2 | 10.06992 | -0.27841 | 0.999532 | | 13.62815 | 0.264629 | 0.733249 | | 10.51182 | 0.573561 | 0.433186 |
| BPGM | 3.264153 | 0.704635 | 0.999532 | | 3.069237 | 0.426169 | 0.733249 | | 3.415906 | -0.25581 | 0.862433 |
| COPG2 | 2.844785 | 0.08999 | 0.999532 | | 2.610047 | -0.4541 | 0.733249 | | 2.312313 | -0.5258 | 0.705425 |
| OR10G4 | 2.77175 | -0.32631 | 0.999532 | | 3.241559 | -0.44798 | 0.733249 | | 2.465129 | -0.09632 | 0.967145 |
| RAB3A | 2.482039 | -0.69648 | 0.999532 | | 2.887493 | -0.44265 | 0.733636 | | 2.074314 | 0.269989 | 0.88885 |
| TBC1D28 | 3.097307 | 0.436617 | 0.999532 | | 2.569407 | -0.46816 | 0.733699 | | 2.622446 | -0.88562 | 0.451774 |
| FCGBP | 2.214849 | 0.527955 | 0.999532 | | 1.916153 | -0.52856 | 0.734 | | 1.867072 | -1.03008 | 0.421517 |
| TUBGCP | 1.979071 | -0.12941 | 0.999532 | | 2.736036 | 0.450077 | 0.734088 | | 2.392134 | 0.601223 | 0.673405 |
| SYNE3 | 3.105405 | 0.026117 | 0.999532 | | 3.062015 | -0.41945 | 0.734306 | | 2.761011 | -0.42544 | 0.754884 |
| SHISA5 | 9.319195 | -0.1545 | 0.999532 | | 12.057 | 0.245374 | 0.734408 | | 9.803901 | 0.420153 | 0.555299 |
| PRAM1 | 2.017892 | 0.221313 | 0.999532 | | 1.873864 | -0.52947 | 0.734408 | | 1.691858 | -0.72861 | 0.644433 |
| KIF4B | 2.404951 | -0.04088 | 0.999532 | | 2.19945 | -0.49983 | 0.734408 | | 1.935075 | -0.44265 | 0.79686 |
| CAAP1 | 2.191611 | -0.53406 | 0.999532 | | 2.353137 | -0.46666 | 0.734408 | | 1.769597 | 0.075939 | 0.977635 |
| KAT6A | 4.246644 | 0.43658 | 0.999532 | | 4.526485 | 0.355598 | 0.734408 | | 4.545451 | -0.05765 | 0.972344 |
| RC3H2 | 4.3071 | -0.00124 | 0.999532 | | 5.531112 | 0.324899 | 0.734408 | | 4.577984 | 0.348437 | 0.748945 |
| HTRA3 | 1.911581 | -0.03677 | 0.999532 | | 1.83631 | -0.54159 | 0.734408 | | 1.595121 | -0.48837 | 0.792539 |
| U2AF1 | 9.302404 | 0.000741 | 0.999532 | | 11.61326 | 0.275398 | 0.734408 | | 9.896258 | 0.294426 | 0.736104 |
| RPUSD1 | 2.341392 | -0.32355 | 0.999532 | | 2.567693 | -0.45916 | 0.734408 | | 1.845347 | -0.12232 | 0.958798 |
| ELK3 | 3.322837 | -0.0472 | 0.999532 | | 3.948067 | 0.366706 | 0.734408 | | 3.50981 | 0.427508 | 0.735469 |
| PANX1 | 2.51061 | -0.39912 | 0.999532 | | 2.834932 | -0.42512 | 0.734408 | | 2.0717 | -0.00527 | 0.998586 |
| AP1AR | 2.218797 | -0.11878 | 0.999532 | | 2.318968 | -0.48345 | 0.734408 | | 1.859256 | -0.3418 | 0.851503 |
| HMMR | 2.571848 | 0.147512 | 0.999532 | | 2.442386 | -0.48893 | 0.734408 | | 2.083418 | -0.61926 | 0.668481 |
| NIPSNAP | 2.084347 | 0.566463 | 0.999532 | | 2.089819 | 0.502832 | 0.734408 | | 2.287834 | -0.04904 | 0.986679 |
| BSDC1 | 6.875639 | 0.043894 | 0.999532 | | 8.491127 | 0.269844 | 0.734408 | | 7.428538 | 0.255128 | 0.794815 |
| RAB8B | 4.67354 | 0.291633 | 0.999532 | | 5.18089 | 0.338758 | 0.734408 | | 4.848996 | 0.067275 | 0.966652 |
| FBXL12 | 2.36359 | 0.18468 | 0.999532 | | 3.155168 | 0.430635 | 0.734761 | | 2.639352 | 0.262777 | 0.868512 |
| IFT140 | 2.691065 | 0.061998 | 0.999532 | | 3.445599 | 0.395749 | 0.735108 | | 3.018494 | 0.351238 | 0.79686 |
| KLF10 | 6.471694 | -0.45697 | 0.999532 | | 7.414012 | -0.31191 | 0.735221 | | 5.322919 | 0.157891 | 0.905952 |
| PNPLA5 | 3.900691 | 0.050757 | 0.999532 | | 3.93737 | -0.38718 | 0.735221 | | 3.396522 | -0.4142 | 0.749222 |
| PLBD1 | 2.685305 | 0.369108 | 0.999532 | | 3.145962 | 0.40126 | 0.735541 | | 3.035943 | 0.049243 | 0.980581 |
| COPA | 8.849692 | 0.158436 | 0.999532 | | 10.39296 | 0.259985 | 0.736105 | | 8.98403 | 0.126152 | 0.900583 |
| TACR2 | 2.434653 | 0.041548 | 0.999532 | | 2.438641 | -0.48415 | 0.736105 | | 2.124197 | -0.50402 | 0.743576 |
| ZNF570 | 2.607581 | -0.02943 | 0.999532 | | 2.795156 | -0.45206 | 0.736268 | | 2.322785 | -0.40149 | 0.794815 |
| CD209 | 2.118439 | -0.17006 | 0.999532 | | 2.255716 | -0.48996 | 0.736625 | | 1.812573 | -0.30079 | 0.876613 |
| STX2 | 3.791873 | 0.017883 | 0.999532 | | 4.44214 | 0.341999 | 0.736625 | | 4.01006 | 0.334176 | 0.790266 |
| SMIM38 | 2.316594 | 0.105545 | 0.999532 | | 2.181332 | -0.50442 | 0.736625 | | 2.033247 | -0.59291 | 0.705425 |
| SLC11A1 | 2.353868 | -0.06668 | 0.999532 | | 2.420847 | -0.50342 | 0.736691 | | 1.999621 | -0.42081 | 0.805825 |
| RETREG1 | 5.882201 | 0.563779 | 0.999532 | | 6.112619 | 0.407655 | 0.736782 | | 6.206438 | -0.11951 | 0.938429 |
| F8A1 | 8.047792 | -0.07222 | 0.999532 | | 8.499181 | -0.26752 | 0.736789 | | 7.038071 | -0.17726 | 0.865065 |
| ZNF704 | 4.613574 | 0.759121 | 0.999532 | | 4.119171 | 0.369537 | 0.736894 | | 4.759222 | -0.37193 | 0.718633 |
| BTF3 | 14.82967 | 0.208299 | 0.999532 | | 16.18963 | 0.240178 | 0.736983 | | 15.13087 | 0.04656 | 0.968434 |
| VAV2 | 3.499402 | 0.190299 | 0.999532 | | 4.375678 | 0.357334 | 0.737001 | | 3.834471 | 0.18136 | 0.896865 |
| STRN4 | 5.902335 | 0.370184 | 0.999532 | | 6.249215 | 0.296738 | 0.737001 | | 5.971537 | -0.06561 | 0.967145 |
| PHOX2A | 2.310804 | 0.254104 | 0.999532 | | 2.162266 | -0.50605 | 0.737001 | | 1.993291 | -0.74006 | 0.60629 |
| RNF151 | 3.820007 | 0.171148 | 0.999532 | | 3.452118 | -0.41801 | 0.737001 | | 3.140751 | -0.57051 | 0.644932 |
| UBR1 | 3.404791 | -0.01364 | 0.999532 | | 4.333169 | 0.361243 | 0.737001 | | 3.746239 | 0.403154 | 0.727175 |
| CERS5 | 5.590449 | 0.126305 | 0.999532 | | 6.461438 | 0.312185 | 0.737001 | | 5.803422 | 0.20177 | 0.859898 |
| FAM120A | 8.005389 | 0.114271 | 0.999532 | | 9.149005 | 0.313013 | 0.737001 | | 8.244468 | 0.200323 | 0.862433 |
| SLC35B2 | 5.11051 | -0.07269 | 0.999532 | | 6.43097 | 0.298622 | 0.737001 | | 5.392074 | 0.391678 | 0.666492 |
| MLYCD | 3.294507 | 0.308816 | 0.999532 | | 3.599106 | 0.417483 | 0.737001 | | 3.530956 | 0.123184 | 0.945129 |
| UBXN2A | 5.131639 | -0.03112 | 0.999532 | | 5.229816 | -0.34128 | 0.737001 | | 4.22006 | -0.29 | 0.800239 |
| DNAJC15 | 5.047245 | -0.16974 | 0.999532 | | 6.764303 | 0.286917 | 0.737001 | | 5.496656 | 0.475583 | 0.578798 |
| TET2 | 3.654234 | 0.350457 | 0.999532 | | 4.102234 | 0.358758 | 0.737001 | | 3.912989 | 0.026356 | 0.991159 |
| CLPTM1 | 4.748446 | -0.5452 | 0.999532 | | 7.177057 | 0.2951 | 0.737001 | | 4.900846 | 0.852996 | 0.251853 |
| TOP1MT | 2.736349 | 0.332933 | 0.999532 | | 2.227847 | -0.50434 | 0.737001 | | 2.289051 | -0.82162 | 0.546846 |
| TDP2 | 4.323078 | 0.332615 | 0.999532 | | 4.781125 | 0.338807 | 0.737001 | | 4.558264 | 0.022517 | 0.992286 |
| H2BU1 | 3.365559 | 0.020656 | 0.999532 | | 3.445924 | -0.45406 | 0.737001 | | 3.085772 | -0.46733 | 0.747457 |
| SHMT2 | 3.872637 | 0.013329 | 0.999532 | | 4.697833 | 0.345811 | 0.737509 | | 4.182857 | 0.342625 | 0.765312 |
| USP4 | 4.673448 | 0.286371 | 0.999532 | | 5.429231 | 0.339769 | 0.737509 | | 5.007549 | 0.072668 | 0.962255 |
| NR1H3 | 4.637972 | -0.03105 | 0.999532 | | 5.921621 | 0.305705 | 0.737509 | | 5.024197 | 0.357834 | 0.738316 |
| ARFRP1 | 7.034783 | 0.058467 | 0.999532 | | 8.642846 | 0.293869 | 0.737509 | | 7.254457 | 0.265741 | 0.806797 |
| EPB41L1 | 5.596163 | -0.04461 | 0.999532 | | 7.157333 | 0.302053 | 0.737509 | | 5.958151 | 0.366543 | 0.717627 |
| MIB1 | 6.336358 | 0.450872 | 0.999532 | | 6.965936 | 0.287721 | 0.737509 | | 6.713501 | -0.14098 | 0.899161 |
| TFR2 | 2.541141 | -0.19546 | 0.999532 | | 2.710932 | -0.46675 | 0.737509 | | 2.109917 | -0.25043 | 0.893259 |
| C11orf91 | 1.842149 | -0.59991 | 0.999532 | | 2.031858 | -0.51276 | 0.737509 | | 1.40564 | 0.101359 | 0.972568 |
| DLEC1 | 1.894611 | 0.282326 | 0.999532 | | 2.154928 | 0.492401 | 0.737548 | | 2.124517 | 0.226827 | 0.907626 |
| SMIM30 | 5.339266 | 0.206032 | 0.999532 | | 6.273593 | 0.336172 | 0.737615 | | 5.842855 | 0.146839 | 0.909074 |
| TNFRSF8 | 2.709457 | 0.126557 | 0.999532 | | 2.503142 | -0.47245 | 0.737615 | | 2.404469 | -0.58867 | 0.69872 |
| KIF16B | 3.312493 | -0.23202 | 0.999532 | | 4.603467 | 0.346067 | 0.737615 | | 3.790081 | 0.60275 | 0.563885 |
| RTN2 | 1.873877 | 0.022426 | 0.999532 | | 2.466131 | 0.458968 | 0.737615 | | 2.17944 | 0.45504 | 0.769793 |
| KIF3C | 2.188143 | -0.23959 | 0.999532 | | 2.209256 | -0.48394 | 0.737615 | | 1.756731 | -0.23031 | 0.909157 |
| SLC25A2 | 3.895532 | -0.00776 | 0.999532 | | 3.89881 | -0.37181 | 0.737615 | | 3.196119 | -0.34577 | 0.785409 |
| IGFBP1 | 2.351966 | -0.10487 | 0.999532 | | 2.468639 | -0.46315 | 0.737615 | | 2.04386 | -0.33855 | 0.849021 |
| BLZF1 | 4.526123 | 0.485653 | 0.999532 | | 4.895882 | 0.331319 | 0.737615 | | 4.629315 | -0.13828 | 0.913083 |
| TRAPPC5 | 5.388144 | -0.24403 | 0.999532 | | 7.398276 | 0.280931 | 0.737615 | | 5.603801 | 0.545437 | 0.515762 |
| ZNF75A | 2.405653 | 0.708061 | 0.999532 | | 2.610166 | 0.470435 | 0.737615 | | 2.616221 | -0.21276 | 0.898369 |
| CCDC97 | 2.480244 | -0.49965 | 0.999532 | | 2.776679 | -0.43367 | 0.737615 | | 2.059996 | 0.07652 | 0.975305 |
| NT5DC4 | 2.192309 | -0.08204 | 0.999532 | | 2.086782 | -0.50209 | 0.737615 | | 1.901291 | -0.40429 | 0.826947 |
| DIRAS2 | 2.80621 | 0.27951 | 0.999532 | | 2.51917 | -0.45718 | 0.737615 | | 2.345621 | -0.71895 | 0.562298 |
| PXMP4 | 2.668418 | -0.1495 | 0.999532 | | 2.85037 | -0.44127 | 0.737615 | | 2.22316 | -0.26671 | 0.880375 |
| SRRT | 5.880315 | 0.304598 | 0.999532 | | 6.728578 | 0.285556 | 0.737615 | | 6.21264 | 0.001618 | 0.999385 |
| OMD | 2.289068 | -0.61255 | 0.999532 | | 2.841563 | -0.44489 | 0.737615 | | 1.94235 | 0.196897 | 0.923864 |
| FAM220A | 2.477994 | 0.536734 | 0.999532 | | 1.873588 | -0.52491 | 0.737615 | | 2.130106 | -1.04548 | 0.434308 |
| CDH6 | 2.249708 | 0.486024 | 0.999532 | | 2.311829 | 0.471575 | 0.737615 | | 2.424022 | -0.01042 | 0.996891 |
| WDR93 | 2.856172 | -0.12734 | 0.999532 | | 2.80605 | -0.46746 | 0.737615 | | 2.483122 | -0.31755 | 0.864245 |
| PLOD1 | 8.896212 | -0.15981 | 0.999532 | | 11.64765 | 0.24833 | 0.737615 | | 9.548461 | 0.422695 | 0.540342 |
| TMEM230 | 6.13316 | 0.3534 | 0.999532 | | 6.828322 | 0.313593 | 0.737615 | | 6.445916 | -0.01921 | 0.992586 |
| REXO4 | 2.628772 | 0.209124 | 0.999532 | | 3.344957 | 0.409971 | 0.737615 | | 3.006894 | 0.221573 | 0.887811 |
| CD53 | 3.094455 | -0.38537 | 0.999532 | | 3.275964 | -0.40432 | 0.737615 | | 2.431543 | -0.01122 | 0.996891 |
| FIGNL2 | 2.72264 | -0.32268 | 0.999532 | | 2.988353 | -0.44784 | 0.737784 | | 2.196885 | -0.10402 | 0.961807 |
| PANO1 | 2.616707 | -0.2214 | 0.999532 | | 2.658903 | -0.46148 | 0.738389 | | 2.264534 | -0.22614 | 0.905952 |
| METTL25 | 2.039232 | 0.613684 | 0.999532 | | 2.066513 | 0.496126 | 0.738515 | | 2.219909 | -0.10068 | 0.965649 |
| DLC1 | 4.3592 | -0.26413 | 0.999532 | | 5.850324 | 0.333671 | 0.738515 | | 4.577157 | 0.623466 | 0.499025 |
| OR14C36 | 2.196232 | -0.18691 | 0.999532 | | 2.397339 | -0.50957 | 0.738702 | | 1.892504 | -0.30621 | 0.877814 |
| LRTM2 | 2.64299 | 0.014524 | 0.999532 | | 2.539065 | -0.47426 | 0.738702 | | 2.309236 | -0.48512 | 0.746876 |
| TSTD3 | 2.645021 | 0.084465 | 0.999532 | | 2.58568 | -0.48515 | 0.738702 | | 2.392193 | -0.56053 | 0.717138 |
| HDAC6 | 8.238725 | 0.279444 | 0.999532 | | 9.196712 | 0.258478 | 0.738774 | | 8.425767 | -0.0047 | 0.997967 |
| WNK3 | 1.882845 | 0.496235 | 0.999532 | | 2.248881 | 0.511045 | 0.738774 | | 2.08976 | 0.029741 | 0.992586 |
| KLHL9 | 6.56993 | 0.247328 | 0.999532 | | 7.230504 | 0.282424 | 0.738774 | | 6.831001 | 0.045659 | 0.977635 |
| RAD23B | 14.22302 | 0.177035 | 0.999532 | | 15.96643 | 0.219875 | 0.738774 | | 14.57503 | 0.054461 | 0.962716 |
| BACE1 | 6.175229 | 0.032001 | 0.999532 | | 7.690806 | 0.29061 | 0.738774 | | 6.84142 | 0.270355 | 0.784263 |
| RXRA | 7.133641 | -0.1261 | 0.999532 | | 9.36874 | 0.266615 | 0.738774 | | 7.589577 | 0.409718 | 0.598934 |
| WASHC1 | 41.39358 | 0.067327 | 0.999532 | | 48.18799 | 0.174295 | 0.738774 | | 41.73079 | 0.132401 | 0.795612 |
| PEX7 | 2.526586 | -0.16258 | 0.999532 | | 2.540732 | -0.47335 | 0.738774 | | 2.109549 | -0.29183 | 0.87591 |
| INKA2 | 2.135391 | -0.18661 | 0.999532 | | 2.112952 | -0.49459 | 0.738774 | | 1.72699 | -0.29504 | 0.881837 |
| TIGD1 | 3.130351 | 0.036376 | 0.999532 | | 3.036548 | -0.42721 | 0.738774 | | 2.665589 | -0.44621 | 0.743576 |
| PRSS27 | 2.524985 | 0.354234 | 0.999532 | | 2.034889 | -0.51011 | 0.738774 | | 2.2764 | -0.85457 | 0.533871 |
| SLC66A2 | 2.602747 | -0.00643 | 0.999532 | | 3.243498 | 0.408019 | 0.738774 | | 2.921948 | 0.428574 | 0.753597 |
| ATG4D | 4.693992 | 0.227571 | 0.999532 | | 4.248883 | -0.39977 | 0.738774 | | 3.855949 | -0.58747 | 0.558035 |
| APOA2 | 2.030482 | 0.46865 | 0.999532 | | 1.626426 | -0.56324 | 0.738774 | | 1.717051 | -1.01562 | 0.484303 |
| DNALI1 | 3.114861 | 0.125894 | 0.999532 | | 2.976974 | -0.44265 | 0.738774 | | 2.584333 | -0.55088 | 0.682165 |
| ADRM1 | 9.574022 | -0.26528 | 0.999532 | | 10.79053 | -0.24199 | 0.738774 | | 8.510088 | 0.041927 | 0.974421 |
| AFAP1L1 | 7.049935 | -0.10932 | 0.999532 | | 7.034374 | -0.34673 | 0.738774 | | 5.796883 | -0.19025 | 0.87591 |
| AEBP2 | 6.445303 | 0.367421 | 0.999532 | | 6.799387 | 0.310645 | 0.738774 | | 6.542655 | -0.03916 | 0.979967 |
| ANAPC10 | 3.263371 | -0.41147 | 0.999532 | | 3.661062 | -0.41883 | 0.738774 | | 2.629329 | 0.016712 | 0.996681 |
| CC2D1A | 2.775727 | 0.225153 | 0.999532 | | 3.316733 | 0.40339 | 0.738774 | | 2.942602 | 0.192598 | 0.901813 |
| LSM3 | 6.016315 | 0.283863 | 0.999532 | | 6.910186 | 0.285139 | 0.738774 | | 6.186773 | 0.014821 | 0.995112 |
| DHPS | 5.287657 | 0.169945 | 0.999532 | | 6.294652 | 0.300927 | 0.738774 | | 5.544731 | 0.150032 | 0.900167 |
| ZNF821 | 2.892573 | 0.680535 | 0.999532 | | 2.7915 | 0.422401 | 0.738774 | | 3.128999 | -0.23927 | 0.87591 |
| CBY3 | 2.454573 | -0.06924 | 0.999532 | | 2.395837 | -0.48708 | 0.73879 | | 2.163048 | -0.40304 | 0.821463 |
| MCAM | 7.029829 | -0.66619 | 0.999532 | | 10.94671 | 0.362963 | 0.738843 | | 7.900023 | 1.026575 | 0.26986 |
| CDK12 | 4.978619 | 0.102089 | 0.999532 | | 4.987332 | -0.32569 | 0.738892 | | 4.347229 | -0.40796 | 0.686214 |
| QRSL1 | 3.096618 | -0.37997 | 0.999532 | | 3.259977 | -0.40008 | 0.73938 | | 2.665376 | -0.01188 | 0.996891 |
| PIAS3 | 3.681495 | -0.24393 | 0.999532 | | 3.657259 | -0.37801 | 0.739619 | | 3.085242 | -0.1212 | 0.945129 |
| ARVCF | 6.383655 | 0.023576 | 0.999532 | | 7.495478 | 0.277407 | 0.739655 | | 6.837605 | 0.299718 | 0.779082 |
| ALDOC | 3.909511 | -0.13086 | 0.999532 | | 4.169044 | -0.35963 | 0.739655 | | 3.32486 | -0.20783 | 0.885788 |
| P3H4 | 3.728751 | 0.171897 | 0.999532 | | 3.384906 | -0.40503 | 0.740363 | | 3.106068 | -0.56113 | 0.647581 |
| TOMM7 | 13.66199 | -0.28043 | 0.999532 | | 18.4025 | 0.212397 | 0.740363 | | 14.13011 | 0.516626 | 0.323586 |
| RNPC3 | 5.790571 | 0.420742 | 0.999532 | | 6.620819 | 0.31096 | 0.740901 | | 6.252504 | -0.07446 | 0.96129 |
| GIGYF1 | 3.627744 | 0.324193 | 0.999532 | | 4.466758 | 0.358662 | 0.740901 | | 4.108881 | 0.056182 | 0.975399 |
| ZFP57 | 2.154296 | -0.25261 | 0.999532 | | 2.362354 | -0.48772 | 0.740982 | | 1.841005 | -0.20347 | 0.925486 |
| ACSM2A | 3.679619 | 0.541383 | 0.999532 | | 4.025467 | 0.458771 | 0.740982 | | 3.873629 | -0.03121 | 0.991159 |
| CTSE | 1.944966 | -0.0932 | 0.999532 | | 1.947106 | -0.51187 | 0.740982 | | 1.696818 | -0.39927 | 0.836935 |
| RNF31 | 3.215809 | 0.302535 | 0.999532 | | 3.745689 | 0.377846 | 0.740982 | | 3.492125 | 0.094302 | 0.957645 |
| HIBCH | 6.900758 | 0.195655 | 0.999532 | | 8.069667 | 0.304451 | 0.740982 | | 6.993008 | 0.140779 | 0.903339 |
| MCRIP1 | 7.537666 | -0.40674 | 0.999532 | | 8.503392 | -0.29148 | 0.741058 | | 6.149886 | 0.140076 | 0.904576 |
| CLEC3A | 2.440871 | 0.021366 | 0.999532 | | 2.408071 | -0.47616 | 0.741058 | | 1.938127 | -0.47739 | 0.761699 |
| GPIHBP1 | 2.417168 | 0.253881 | 0.999532 | | 2.234879 | -0.48072 | 0.74155 | | 2.028374 | -0.71347 | 0.615566 |
| ZBTB44 | 3.301718 | 0.083749 | 0.999532 | | 3.846504 | 0.368431 | 0.74155 | | 3.331655 | 0.29976 | 0.819366 |
| SETBP1 | 3.977642 | 0.069962 | 0.999532 | | 3.942783 | -0.38001 | 0.74155 | | 3.321165 | -0.42911 | 0.717138 |
| KLHL32 | 2.807992 | 0.460853 | 0.999532 | | 2.535716 | -0.45834 | 0.74155 | | 2.390999 | -0.88724 | 0.446959 |
| RAB13 | 11.69677 | 0.134498 | 0.999532 | | 14.22909 | 0.278629 | 0.74155 | | 12.47903 | 0.178253 | 0.867739 |
| ZNF35 | 2.81073 | -0.50885 | 0.999532 | | 3.344702 | -0.41311 | 0.74155 | | 2.288321 | 0.114163 | 0.955834 |
| MESD | 3.074042 | -0.02271 | 0.999532 | | 2.900276 | -0.43238 | 0.74155 | | 2.433171 | -0.39148 | 0.792539 |
| PRR30 | 2.210491 | -0.27017 | 0.999532 | | 2.377081 | -0.48102 | 0.74155 | | 1.815805 | -0.19039 | 0.929529 |
| CDX1 | 2.252117 | 0.248027 | 0.999532 | | 2.089531 | -0.49562 | 0.74155 | | 1.968772 | -0.71219 | 0.647581 |
| PHYHIPL | 1.967839 | 0.449737 | 0.999532 | | 2.078125 | 0.514004 | 0.74155 | | 2.144151 | 0.078096 | 0.976783 |
| FANCL | 2.726939 | -0.17627 | 0.999532 | | 2.634146 | -0.43404 | 0.74155 | | 2.319672 | -0.24547 | 0.892869 |
| VEGFA | 59.91153 | 0.240287 | 0.999532 | | 67.89823 | 0.251745 | 0.74155 | | 62.17968 | 0.03905 | 0.965619 |
| CRTC2 | 4.916814 | 0.055397 | 0.999532 | | 4.990209 | -0.34505 | 0.74155 | | 4.270568 | -0.37595 | 0.737651 |
| TADA3 | 5.014485 | -0.03416 | 0.999532 | | 6.42262 | 0.29374 | 0.74155 | | 5.196507 | 0.346876 | 0.718633 |
| TWF1 | 3.501863 | -0.03648 | 0.999532 | | 4.438566 | 0.344003 | 0.74155 | | 3.786298 | 0.396957 | 0.725637 |
| LMBRD1 | 7.074332 | 0.041994 | 0.999532 | | 8.59504 | 0.281226 | 0.74155 | | 7.313245 | 0.255701 | 0.801905 |
| COG4 | 4.774671 | 0.395772 | 0.999532 | | 5.619872 | 0.321568 | 0.74155 | | 5.102275 | -0.05244 | 0.974421 |
| C1QBP | 3.929078 | 0.009039 | 0.999532 | | 4.996401 | 0.32859 | 0.74155 | | 4.302189 | 0.338464 | 0.754186 |
| FAM240C | 2.972395 | 0.020063 | 0.999532 | | 2.915394 | -0.47604 | 0.74155 | | 2.484847 | -0.47486 | 0.763873 |
| SLC16A3 | 4.844569 | -0.20346 | 0.999532 | | 5.16885 | -0.32169 | 0.74155 | | 4.205572 | -0.09521 | 0.950529 |
| NEO1 | 5.3181 | -0.27041 | 0.999532 | | 7.440937 | 0.312029 | 0.74155 | | 5.664904 | 0.592335 | 0.486743 |
| CBFB | 6.637186 | 0.370207 | 0.999532 | | 7.169873 | 0.294442 | 0.74155 | | 7.102646 | -0.05632 | 0.971797 |
| SP6 | 2.402195 | -0.13396 | 0.999532 | | 2.521125 | -0.4681 | 0.74155 | | 2.072445 | -0.31679 | 0.862823 |
| UBR5 | 4.632436 | -0.21223 | 0.999532 | | 6.167733 | 0.3053 | 0.74155 | | 4.849915 | 0.533286 | 0.568257 |
| WBP1L | 4.361271 | 0.426153 | 0.999532 | | 4.885236 | 0.330485 | 0.74155 | | 4.522384 | -0.08067 | 0.959377 |
| CDKN2D | 7.633746 | 0.196897 | 0.999532 | | 7.421031 | -0.30188 | 0.74155 | | 6.709094 | -0.4678 | 0.60629 |
| CELSR2 | 3.910621 | -0.01986 | 0.999532 | | 4.909495 | 0.332807 | 0.74155 | | 4.077118 | 0.368399 | 0.749206 |
| ZBTB40 | 3.144465 | -0.26289 | 0.999532 | | 3.651499 | -0.38812 | 0.74155 | | 2.66691 | -0.10503 | 0.955834 |
| IL20 | 2.462063 | 0.063336 | 0.999532 | | 2.569584 | -0.46879 | 0.74155 | | 2.204815 | -0.50822 | 0.735469 |
| PARP12 | 2.569829 | -0.04421 | 0.999532 | | 3.493977 | 0.392113 | 0.74155 | | 2.895654 | 0.453329 | 0.722703 |
| LIN7C | 3.735738 | 0.191515 | 0.999532 | | 4.41713 | 0.348158 | 0.741939 | | 3.911867 | 0.169286 | 0.906811 |
| H3C13 | 7.113177 | -0.13767 | 0.999532 | | 7.632904 | -0.30916 | 0.742186 | | 6.235697 | -0.14738 | 0.914692 |
| CBX7 | 11.20526 | 0.002526 | 0.999532 | | 13.29509 | 0.231182 | 0.743068 | | 11.56549 | 0.248957 | 0.763092 |
| EPHA1 | 1.961049 | 0.625585 | 0.999532 | | 2.445444 | 0.51469 | 0.7431 | | 2.452965 | -0.07876 | 0.975194 |
| NUPR1 | 8.913051 | 0.111186 | 0.999532 | | 10.60343 | 0.27753 | 0.7431 | | 9.233098 | 0.189618 | 0.852152 |
| CDK4 | 6.500809 | -0.05445 | 0.999532 | | 8.095471 | 0.289777 | 0.7431 | | 6.947787 | 0.353376 | 0.718633 |

| | | | | | | | | | | | |
|---|---|---|---|---|---|---|---|---|---|---|---|
| EEA1 | 6.167962 | 0.247416 | 0.999532 | | 7.236716 | 0.273086 | 0.7431 | | 6.565456 | 0.049806 | 0.973356 |
| GAREM2 | 3.903958 | -0.23513 | 0.999532 | | 4.251366 | -0.37781 | 0.7431 | | 3.335486 | -0.12276 | 0.945366 |
| GSE1 | 4.238402 | -0.15412 | 0.999532 | | 5.781186 | 0.304744 | 0.743493 | | 4.52577 | 0.481064 | 0.629006 |
| D2HGDH | 6.085864 | 0.017261 | 0.999532 | | 5.84642 | -0.31552 | 0.743493 | | 5.191934 | -0.30896 | 0.770105 |
| SNCB | 2.128695 | -0.40855 | 0.999532 | | 2.440911 | -0.47727 | 0.743493 | | 1.80141 | -0.04661 | 0.989278 |
| AVP | 2.231323 | 0.389528 | 0.999532 | | 1.822221 | -0.53239 | 0.743493 | | 1.95175 | -0.90981 | 0.532278 |
| KRTAP9- | 6.089784 | -0.01385 | 0.999532 | | 6.261846 | -0.3456 | 0.743493 | | 5.458141 | -0.31603 | 0.816618 |
| SIGIRR | 6.603685 | -0.09601 | 0.999532 | | 8.47334 | 0.275419 | 0.743493 | | 6.994474 | 0.390473 | 0.647218 |
| ZNF124 | 7.076206 | 0.182293 | 0.999532 | | 6.55284 | -0.39096 | 0.743493 | | 6.047868 | -0.56398 | 0.600254 |
| TTC32 | 2.342437 | -0.17587 | 0.999532 | | 2.391395 | -0.47691 | 0.743493 | | 1.887462 | -0.28288 | 0.881837 |
| C10orf12 | 2.253631 | -0.05591 | 0.999532 | | 2.293589 | -0.49174 | 0.743493 | | 1.890857 | -0.41234 | 0.816589 |
| GOT1L1 | 3.01617 | 0.044973 | 0.999532 | | 3.002263 | -0.44976 | 0.743572 | | 2.529304 | -0.47015 | 0.740755 |
| IQSEC1 | 8.958322 | 0.089662 | 0.999532 | | 10.37969 | 0.254825 | 0.74363 | | 9.177386 | 0.183653 | 0.845773 |
| HBA1 | 12.56514 | 0.3387 | 0.999532 | | 14.61286 | 0.411003 | 0.74363 | | 13.76467 | 0.104319 | 0.952785 |
| USP14 | 6.170249 | 0.351053 | 0.999532 | | 6.756985 | 0.303232 | 0.743803 | | 6.417518 | -0.03056 | 0.987274 |
| SULT1A4 | 12.16001 | -0.01804 | 0.999532 | | 12.50106 | -0.22833 | 0.743803 | | 10.87568 | -0.19038 | 0.827267 |
| NOA1 | 2.744238 | -0.09622 | 0.999532 | | 2.792827 | -0.41413 | 0.743803 | | 2.278904 | -0.29941 | 0.852487 |
| PRPF3 | 8.510576 | 0.230788 | 0.999532 | | 9.638422 | 0.26035 | 0.743803 | | 8.738591 | 0.052155 | 0.967999 |
| TBL1XR1 | 3.368973 | 0.265468 | 0.999532 | | 3.782367 | 0.385377 | 0.743803 | | 3.452462 | 0.145792 | 0.923834 |
| MTO1 | 4.128482 | 0.272692 | 0.999532 | | 3.845171 | -0.38215 | 0.743987 | | 3.687361 | -0.63583 | 0.552672 |
| MAN1C1 | 4.814886 | 0.020282 | 0.999532 | | 4.850567 | -0.36532 | 0.74426 | | 3.880317 | -0.37288 | 0.735469 |
| BANP | 2.648811 | -0.18906 | 0.999532 | | 2.759888 | -0.43408 | 0.74426 | | 2.32789 | -0.22189 | 0.900848 |
| MATK | 2.367636 | -0.50352 | 0.999532 | | 2.614886 | -0.43047 | 0.744295 | | 1.9396 | 0.08226 | 0.974421 |
| KCNJ8 | 2.760082 | 0.006296 | 0.999532 | | 2.85953 | -0.42231 | 0.744295 | | 2.319281 | -0.40165 | 0.792416 |
| GGTLC2 | 5.410153 | 0.042039 | 0.999532 | | 5.368058 | -0.38493 | 0.744295 | | 4.685967 | -0.41407 | 0.724797 |
| RDH14 | 3.407132 | 0.052044 | 0.999532 | | 3.182685 | -0.38712 | 0.744295 | | 2.920567 | -0.42513 | 0.735469 |
| CPOX | 2.017892 | -0.21132 | 0.999532 | | 2.003761 | -0.50453 | 0.744295 | | 1.655627 | -0.2769 | 0.894612 |
| NPTN | 12.75183 | -0.0906 | 0.999532 | | 16.37594 | 0.206117 | 0.744295 | | 13.50882 | 0.317315 | 0.62216 |
| TRPC1 | 2.124739 | 0.488183 | 0.999532 | | 2.458345 | 0.458103 | 0.744295 | | 2.364858 | -0.01075 | 0.996891 |
| CALM3 | 17.42502 | 0.038642 | 0.999532 | | 20.74489 | 0.224049 | 0.744295 | | 18.09287 | 0.213933 | 0.749999 |
| ZNF395 | 4.994751 | 0.630615 | 0.999532 | | 4.816923 | 0.340821 | 0.744295 | | 5.314138 | -0.27921 | 0.806797 |
| STK25 | 9.172613 | 0.262618 | 0.999532 | | 10.36436 | 0.243597 | 0.744295 | | 9.51971 | 0.004245 | 0.997967 |
| GRAP | 2.381612 | -0.55268 | 0.999532 | | 2.741632 | -0.45233 | 0.744295 | | 1.801445 | 0.123551 | 0.956011 |
| COPS7A | 4.517978 | 0.0346 | 0.999532 | | 5.428284 | 0.32297 | 0.744295 | | 4.662803 | 0.307912 | 0.783905 |
| IGSF8 | 3.193346 | 0.554741 | 0.999532 | | 3.431319 | 0.454302 | 0.744295 | | 3.459386 | -0.07002 | 0.974421 |
| CETN3 | 3.289007 | 0.055719 | 0.999532 | | 4.113211 | 0.352199 | 0.744295 | | 3.68055 | 0.320141 | 0.806394 |
| CTNNBL1 | 3.499858 | 0.038941 | 0.999532 | | 4.367636 | 0.355433 | 0.744295 | | 3.671312 | 0.332645 | 0.787995 |
| LRP12 | 3.173863 | -0.36134 | 0.999532 | | 3.44325 | -0.42282 | 0.744295 | | 2.568716 | -0.03987 | 0.989148 |
| DARS2 | 3.247717 | -0.17475 | 0.999532 | | 3.491911 | -0.39915 | 0.744295 | | 2.883979 | -0.21238 | 0.894612 |
| AFTPH | 2.913261 | 0.347386 | 0.999532 | | 3.473719 | 0.383479 | 0.744295 | | 3.219587 | 0.055107 | 0.978229 |
| EZH1 | 5.617701 | -0.12379 | 0.999532 | | 7.016434 | 0.30035 | 0.744295 | | 6.057078 | 0.44241 | 0.654619 |
| PCDH10 | 2.863514 | 0.64503 | 0.999532 | | 2.182619 | -0.50061 | 0.744295 | | 2.384133 | -1.12671 | 0.304062 |
| SERPINB | 2.810437 | 0.529704 | 0.999532 | | 2.235144 | -0.49721 | 0.744295 | | 2.536352 | -1.0157 | 0.386809 |
| STMP1 | 8.855848 | 0.225799 | 0.999532 | | 10.34337 | 0.250616 | 0.744295 | | 9.209022 | 0.048123 | 0.970498 |
| SCN2B | 3.197329 | -0.34501 | 0.999532 | | 3.476093 | -0.40889 | 0.744295 | | 2.600791 | -0.04156 | 0.988379 |
| DOK6 | 2.061702 | 0.475389 | 0.999532 | | 2.239457 | 0.472518 | 0.744538 | | 2.264234 | 0.012817 | 0.996891 |
| SEPSEC5 | 2.202568 | 0.47651 | 0.999532 | | 2.310379 | 0.465362 | 0.744538 | | 2.337424 | 0.002305 | 0.999385 |
| LRRC31 | 2.196479 | 0.077059 | 0.999532 | | 2.093289 | -0.48453 | 0.744538 | | 1.897006 | -0.54184 | 0.730921 |
| FKBP9 | 11.56037 | -0.27185 | 0.999532 | | 15.70379 | 0.201081 | 0.744538 | | 12.22277 | 0.506885 | 0.436389 |
| SCAMP4 | 4.892864 | 0.13473 | 0.999532 | | 6.045778 | 0.30762 | 0.744545 | | 5.159894 | 0.197528 | 0.881837 |
| SLC35F4 | 2.556686 | -0.02207 | 0.999532 | | 2.506058 | -0.47927 | 0.744545 | | 2.028493 | -0.43563 | 0.791466 |
| TTC7B | 2.724701 | 0.274515 | 0.999532 | | 3.179098 | 0.403991 | 0.744545 | | 2.856001 | 0.148922 | 0.928552 |
| KIAA1217 | 4.445414 | 0.348022 | 0.999532 | | 5.1725 | 0.332192 | 0.744545 | | 4.658658 | 0.012817 | 0.996681 |
| TMEM178 | 1.841906 | -0.23065 | 0.999532 | | 2.56393 | 0.448036 | 0.744545 | | 2.153648 | 0.698919 | 0.636525 |
| RAD51C | 2.962294 | -0.02746 | 0.999532 | | 3.791879 | 0.40193 | 0.744699 | | 3.185194 | 0.450573 | 0.720526 |
| PRR4 | 2.291737 | 0.461889 | 0.999532 | | 2.545037 | 0.449137 | 0.744785 | | 2.556653 | 0.004295 | 0.998898 |
| FIGN | 2.966349 | 0.24674 | 0.999532 | | 2.808887 | -0.43375 | 0.745106 | | 2.443181 | -0.66161 | 0.595927 |
| ART5 | 2.452661 | -0.02106 | 0.999532 | | 2.36379 | -0.48782 | 0.745183 | | 1.986223 | -0.44441 | 0.784263 |
| CEBPZOS | 3.638701 | -0.52085 | 0.999532 | | 4.418349 | -0.37549 | 0.745862 | | 3.102434 | 0.167167 | 0.920259 |
| SAMD13 | 2.255879 | 0.541195 | 0.999532 | | 2.507288 | 0.446095 | 0.745862 | | 2.560283 | -0.07766 | 0.972221 |
| SYNJ2 | 3.585961 | 0.135362 | 0.999532 | | 4.472862 | 0.361535 | 0.745862 | | 3.939042 | 0.239935 | 0.849885 |
| SLC22A1 | 2.274505 | -0.75042 | 0.999532 | | 2.562068 | -0.44878 | 0.745995 | | 1.686644 | 0.308722 | 0.87813 |
| H4C6 | 3.564729 | -0.53926 | 0.999532 | | 3.917166 | -0.36877 | 0.746182 | | 2.903268 | 0.175753 | 0.918172 |
| EPS8L1 | 3.339675 | 0.247413 | 0.999532 | | 3.936765 | 0.357617 | 0.746182 | | 3.424477 | 0.123171 | 0.9402 |
| ZNF134 | 4.095393 | -0.48186 | 0.999532 | | 4.751012 | -0.35312 | 0.746182 | | 3.485056 | 0.145099 | 0.92697 |
| ZNF394 | 3.920823 | -0.11968 | 0.999532 | | 4.057958 | -0.37149 | 0.746182 | | 3.421647 | -0.23609 | 0.878619 |
| PREPL | 3.397409 | 0.03939 | 0.999532 | | 4.404094 | 0.341074 | 0.746182 | | 3.823024 | 0.320588 | 0.790999 |
| UGT3A1 | 2.211822 | 0.537702 | 0.999532 | | 2.438888 | 0.441023 | 0.746182 | | 2.47227 | -0.07791 | 0.972221 |
| XRRA1 | 2.289336 | 0.226252 | 0.999532 | | 2.17129 | -0.51736 | 0.746182 | | 1.938083 | -0.72803 | 0.630705 |
| BMP5 | 1.978235 | 0.510389 | 0.999532 | | 2.401163 | 0.491346 | 0.746182 | | 2.355031 | 0.005074 | 0.998655 |
| BATF2 | 3.021382 | -0.71999 | 0.999532 | | 3.870821 | -0.38725 | 0.746199 | | 2.704471 | 0.344102 | 0.82268 |
| TNFRSF4 | 4.006354 | -0.05016 | 0.999532 | | 5.021512 | 0.34209 | 0.74651 | | 4.368285 | 0.401199 | 0.723933 |
| SMARCA | 6.162424 | 0.031543 | 0.999532 | | 7.494491 | 0.276661 | 0.746849 | | 6.363806 | 0.262309 | 0.795612 |
| RHPN1 | 2.039664 | 0.890562 | 0.999532 | | 1.944037 | 0.506249 | 0.74709 | | 2.188287 | -0.36555 | 0.816775 |
| RNF39 | 2.285197 | 0.317313 | 0.999532 | | 2.12337 | -0.50482 | 0.747307 | | 1.894208 | -0.80052 | 0.566812 |
| GORASP | 3.015142 | -0.19163 | 0.999532 | | 4.072988 | 0.358739 | 0.747307 | | 3.428337 | 0.568652 | 0.64142 |
| ARSF | 1.977109 | 0.119654 | 0.999532 | | 1.813436 | -0.52603 | 0.747307 | | 1.606224 | -0.62873 | 0.70882 |
| DGKQ | 4.361231 | 0.423016 | 0.999532 | | 3.8758 | -0.38698 | 0.747307 | | 3.88266 | -0.78014 | 0.448851 |
| TBCK | 3.037738 | -0.10762 | 0.999532 | | 3.092779 | -0.40877 | 0.747307 | | 2.588902 | -0.28173 | 0.861119 |
| BBS5 | 2.612502 | 0.286859 | 0.999532 | | 3.103778 | 0.438437 | 0.747307 | | 2.943128 | 0.168093 | 0.92697 |
| EIF2AK1 | 4.202653 | 0.090487 | 0.999532 | | 4.970918 | 0.330524 | 0.747356 | | 4.583548 | 0.274723 | 0.825181 |
| CTSO | 6.227479 | 0.248829 | 0.999532 | | 6.805641 | 0.288343 | 0.747356 | | 6.458172 | 0.054345 | 0.972039 |
| ZNF28 | 13.78777 | -0.11358 | 0.999532 | | 14.91427 | -0.25629 | 0.747356 | | 12.35822 | -0.12667 | 0.90424 |
| NARS1 | 6.95518 | 0.109827 | 0.999532 | | 8.363475 | 0.269482 | 0.747587 | | 7.294432 | 0.1827 | 0.869378 |
| MPRIP | 10.89355 | 0.156801 | 0.999532 | | 12.46286 | 0.266563 | 0.747587 | | 11.36324 | 0.123012 | 0.905746 |
| TIMM44 | 2.419841 | 0.399307 | 0.999532 | | 2.804006 | 0.417022 | 0.747587 | | 2.740029 | 0.037699 | 0.989033 |
| CLDN9 | 2.457464 | 0.321888 | 0.999532 | | 2.146176 | -0.51286 | 0.747587 | | 2.019676 | -0.81532 | 0.569849 |
| TNFAIP8 | 2.970007 | -0.77654 | 0.999532 | | 3.669618 | -0.36546 | 0.747587 | | 2.49469 | 0.427875 | 0.759211 |
| MRPS26 | 3.081283 | 0.45005 | 0.999532 | | 3.486405 | 0.393936 | 0.747587 | | 3.334387 | -0.03085 | 0.991103 |
| ZNF528 | 4.885914 | -0.17285 | 0.999532 | | 5.366894 | -0.34432 | 0.747587 | | 4.281262 | -0.14749 | 0.920259 |
| SYT15 | 2.565573 | 0.104635 | 0.999532 | | 2.430157 | -0.47361 | 0.747587 | | 2.330209 | -0.57378 | 0.704227 |
| SPON1 | 2.461498 | 0.407316 | 0.999532 | | 2.044485 | -0.47965 | 0.747587 | | 2.077323 | -0.86944 | 0.499025 |
| RFC2 | 2.904191 | -0.05902 | 0.999532 | | 2.967308 | -0.44044 | 0.747587 | | 2.527896 | -0.35134 | 0.836935 |
| ACD | 3.562469 | -0.00442 | 0.999532 | | 3.820589 | -0.39576 | 0.747587 | | 3.140665 | -0.34727 | 0.814626 |
| TBC1D31 | 2.335582 | 0.241873 | 0.999532 | | 2.149838 | -0.49129 | 0.747587 | | 2.092319 | -0.71699 | 0.631563 |
| GGN | 2.715686 | 0.118074 | 0.999532 | | 2.670578 | -0.44335 | 0.747587 | | 2.29804 | -0.53699 | 0.705046 |
| TEX9 | 2.800359 | 0.130926 | 0.999532 | | 3.402363 | 0.389006 | 0.747587 | | 3.092286 | 0.277139 | 0.851503 |
| CARD8 | 6.383151 | 0.20651 | 0.999532 | | 7.353139 | 0.284978 | 0.747587 | | 6.998121 | 0.093724 | 0.944262 |
| PTPN7 | 4.225179 | 0.175872 | 0.999532 | | 3.694154 | -0.36466 | 0.747587 | | 3.615954 | -0.5334 | 0.646821 |
| NMI | 3.335019 | 0.074041 | 0.999532 | | 4.032916 | 0.354034 | 0.747587 | | 3.586519 | 0.299496 | 0.810635 |
| RANBP1 | 7.084536 | 0.210735 | 0.999532 | | 8.138253 | 0.25816 | 0.747587 | | 7.185267 | 0.063211 | 0.960773 |
| ACADM | 3.950124 | 0.309498 | 0.999532 | | 4.470426 | 0.34364 | 0.747587 | | 4.048598 | 0.056542 | 0.974421 |
| ICA1L | 4.564382 | 0.313262 | 0.999532 | | 4.027633 | -0.37781 | 0.747587 | | 3.964867 | -0.67604 | 0.515762 |
| LMLN | 2.166084 | 0.057968 | 0.999532 | | 1.978742 | -0.51482 | 0.747587 | | 1.724052 | -0.5567 | 0.742171 |
| TMEM272 | 2.435566 | -0.03104 | 0.999532 | | 2.460703 | -0.46761 | 0.747587 | | 2.00371 | -0.41562 | 0.806366 |
| ERMN | 2.054766 | 0.283345 | 0.999532 | | 1.75399 | -0.52868 | 0.747587 | | 1.760093 | -0.7974 | 0.600081 |
| IL36RN | 3.857683 | 0.326735 | 0.999532 | | 3.436835 | -0.42048 | 0.747587 | | 3.547489 | -0.73736 | 0.540342 |
| PLEKHA5 | 5.456004 | 0.126271 | 0.999532 | | 6.174123 | 0.309543 | 0.747587 | | 5.352292 | 0.212292 | 0.851785 |
| PTCD3 | 5.615443 | 0.08027 | 0.999532 | | 7.169787 | 0.284082 | 0.747587 | | 6.003908 | 0.236303 | 0.835554 |
| MRPL15 | 3.169534 | -0.18148 | 0.999532 | | 3.281034 | -0.3924 | 0.747587 | | 2.55486 | -0.1948 | 0.906811 |
| CPNE7 | 2.291792 | 0.422754 | 0.999532 | | 1.914358 | -0.50055 | 0.747587 | | 1.946536 | -0.89695 | 0.515762 |
| FBXL20 | 3.102628 | -0.16875 | 0.999532 | | 3.261841 | -0.3882 | 0.747587 | | 2.635465 | -0.19947 | 0.902504 |
| RTN4IP1 | 2.066908 | 0.045739 | 0.999532 | | 1.927042 | -0.49521 | 0.747587 | | 1.772599 | -0.52476 | 0.750371 |
| PIWIL2 | 3.212625 | 0.521501 | 0.999532 | | 2.567581 | -0.44342 | 0.747587 | | 2.76508 | -0.94718 | 0.394452 |
| LHX9 | 2.092049 | -0.01332 | 0.999532 | | 2.063228 | -0.50097 | 0.747587 | | 1.778144 | -0.4695 | 0.792416 |
| CLPP | 4.705658 | 0.210907 | 0.999532 | | 5.338885 | 0.311797 | 0.747587 | | 4.94352 | 0.11492 | 0.93275 |
| AKR1B15 | 2.453053 | -0.2498 | 0.999532 | | 2.746671 | -0.43299 | 0.747587 | | 2.128795 | -0.15492 | 0.940333 |
| ADGRG1 | 7.924428 | -0.00409 | 0.999532 | | 10.05728 | 0.292655 | 0.747587 | | 8.068024 | 0.31971 | 0.793355 |
| FDFT1 | 4.69645 | -0.09062 | 0.999532 | | 6.116009 | 0.317359 | 0.747587 | | 4.981163 | 0.427007 | 0.654619 |
| MTMR7 | 2.470837 | -0.11458 | 0.999532 | | 2.523193 | -0.5041 | 0.747587 | | 2.014871 | -0.37107 | 0.836453 |
| HOXC4 | 5.550441 | -0.44295 | 0.999532 | | 6.421149 | -0.31787 | 0.747587 | | 5.087225 | 0.132255 | 0.933547 |
| RRP7A | 3.266926 | -0.40381 | 0.999532 | | 3.727681 | -0.37863 | 0.747587 | | 2.742485 | 0.044521 | 0.985693 |
| TSHB | 2.976762 | 0.02041 | 0.999532 | | 2.915935 | -0.44235 | 0.747587 | | 2.417838 | -0.44304 | 0.758124 |
| ZNF614 | 5.697702 | 0.110575 | 0.999532 | | 5.385615 | -0.31316 | 0.747587 | | 4.846474 | -0.40424 | 0.686214 |
| MCU | 2.809363 | 0.311532 | 0.999532 | | 3.238052 | 0.384224 | 0.747592 | | 3.046016 | 0.089842 | 0.961567 |
| ANKRD33 | 3.80292 | 0.244797 | 0.999532 | | 3.262524 | -0.42604 | 0.747592 | | 3.241419 | -0.64945 | 0.636184 |
| SSB | 9.896043 | 0.140921 | 0.999532 | | 11.71566 | 0.232593 | 0.747592 | | 10.13387 | 0.116048 | 0.917875 |
| MDK | 4.059333 | 0.089385 | 0.999532 | | 3.569045 | -0.37616 | 0.747592 | | 3.36759 | -0.46144 | 0.709825 |
| NPPB | 2.425901 | -0.05826 | 0.999532 | | 2.308658 | -0.46136 | 0.747592 | | 2.037856 | -0.38686 | 0.818118 |
| RPS15 | 26.38598 | -0.03082 | 0.999532 | | 32.13952 | 0.208486 | 0.747592 | | 26.95581 | 0.264047 | 0.693035 |
| ADAMTS | 2.097057 | 0.2355 | 0.999532 | | 2.490057 | 0.471664 | 0.747592 | | 2.095721 | 0.275091 | 0.877155 |
| ABHD5 | 2.922645 | -0.37985 | 0.999532 | | 3.300318 | -0.3928 | 0.747592 | | 2.503182 | 0.006737 | 0.997967 |
| ZMYM4 | 7.363683 | 0.336781 | 0.999532 | | 7.874476 | 0.256132 | 0.747592 | | 7.617993 | -0.06259 | 0.959727 |
| AP1G2 | 2.50452 | 0.30528 | 0.999532 | | 2.897588 | 0.414768 | 0.747592 | | 2.775151 | 0.126079 | 0.945456 |
| MTX2 | 2.46602 | 0.235837 | 0.999532 | | 3.2106 | 0.411392 | 0.747649 | | 2.93215 | 0.198063 | 0.904801 |
| DEF8 | 4.833267 | 0.259905 | 0.999532 | | 4.639905 | -0.33479 | 0.747874 | | 4.326075 | -0.56521 | 0.583461 |
| GYG1 | 6.564961 | -0.05983 | 0.999532 | | 6.866653 | -0.29296 | 0.748724 | | 5.791997 | -0.21562 | 0.859898 |
| SCAND1 | 4.928887 | -0.3299 | 0.999532 | | 5.263817 | -0.31481 | 0.74878 | | 4.023357 | 0.023899 | 0.992286 |
| LIN9 | 1.8747 | -0.19173 | 0.999532 | | 2.018612 | -0.52315 | 0.749051 | | 1.689191 | -0.30747 | 0.88885 |
| FAM221A | 2.197023 | -0.13326 | 0.999532 | | 2.182887 | -0.48874 | 0.749461 | | 1.78262 | -0.33899 | 0.864245 |
| SMIM10L | 4.361494 | -0.60925 | 0.999532 | | 5.320808 | -0.34305 | 0.749461 | | 3.772459 | 0.287089 | 0.835554 |
| NBL1 | 6.564496 | -0.53742 | 0.999532 | | 9.673003 | 0.282027 | 0.749485 | | 7.070408 | 0.832866 | 0.282041 |
| C1D | 5.444135 | 0.113633 | 0.999532 | | 5.475313 | -0.32507 | 0.750039 | | 4.664719 | -0.4207 | 0.68549 |
| IL22 | 2.224975 | 0.132577 | 0.999532 | | 2.179707 | -0.48959 | 0.750039 | | 1.865347 | -0.60053 | 0.701419 |
| H2BC15 | 3.220518 | 0.11233 | 0.999532 | | 2.995352 | -0.41669 | 0.750096 | | 2.688558 | -0.52078 | 0.717627 |
| IRF9 | 6.469573 | 0.115456 | 0.999532 | | 7.550131 | 0.284553 | 0.750096 | | 6.81543 | 0.181953 | 0.87591 |
| POLG2 | 3.990639 | -0.06351 | 0.999532 | | 4.012496 | -0.3859 | 0.750096 | | 3.429429 | -0.30167 | 0.821113 |
| KRT19 | 4.140762 | 0.086335 | 0.999532 | | 4.968674 | 0.344828 | 0.750096 | | 4.401632 | 0.274251 | 0.835972 |
| QRICH1 | 2.414765 | -0.1537 | 0.999532 | | 2.562765 | -0.42039 | 0.750519 | | 2.053807 | -0.24504 | 0.890261 |
| IFNW1 | 2.592293 | 0.029267 | 0.999532 | | 2.668903 | -0.48265 | 0.750648 | | 2.260435 | -0.49226 | 0.758124 |
| KIZ | 2.91004 | 0.171105 | 0.999532 | | 2.563511 | -0.44126 | 0.750648 | | 2.490324 | -0.60061 | 0.673405 |
| VAPA | 13.68141 | -0.08033 | 0.999532 | | 17.08738 | 0.212894 | 0.750911 | | 14.10325 | 0.312947 | 0.660958 |
| C2CD4B | 2.22732 | -0.27054 | 0.999532 | | 2.373184 | -0.48829 | 0.750981 | | 1.886377 | -0.20025 | 0.92697 |
| KDELR1 | 5.091934 | 0.183701 | 0.999532 | | 5.835714 | 0.297861 | 0.751065 | | 5.130568 | 0.134764 | 0.914687 |
| RASSF4 | 4.729718 | 0.328604 | 0.999532 | | 5.236333 | 0.345257 | 0.751065 | | 4.960843 | 0.031101 | 0.988853 |
| KCTD10 | 5.077772 | -0.30348 | 0.999532 | | 5.638412 | -0.31085 | 0.751065 | | 4.556736 | 0.007894 | 0.996891 |
| C7orf25 | 2.850315 | -0.11556 | 0.999532 | | 2.949225 | -0.40522 | 0.751065 | | 2.417293 | -0.27205 | 0.865363 |
| NOMO3 | 7.74566 | -0.09974 | 0.999532 | | 9.473546 | 0.241841 | 0.751065 | | 7.79627 | 0.356444 | 0.657221 |
| FIBP | 3.404694 | 0.076186 | 0.999532 | | 4.082109 | 0.38296 | 0.751065 | | 3.544817 | 0.334636 | 0.79686 |
| CLCN6 | 4.466858 | -0.29844 | 0.999532 | | 4.884137 | -0.33423 | 0.751065 | | 3.884765 | -0.0223 | 0.993618 |
| CCDC89 | 2.637217 | -0.20577 | 0.999532 | | 2.669176 | -0.43159 | 0.751065 | | 2.233591 | -0.20126 | 0.914929 |
| ING1 | 5.502253 | 0.250185 | 0.999532 | | 4.739378 | -0.3218 | 0.751065 | | 4.71895 | -0.56327 | 0.552672 |
| NOTCH2N | 10.32527 | 0.364966 | 0.999532 | | 11.25731 | 0.239899 | 0.751202 | | 10.59954 | -0.10141 | 0.918694 |
| RPF1 | 3.351386 | 0.047463 | 0.999532 | | 4.135675 | 0.349593 | 0.751202 | | 3.396842 | 0.317008 | 0.800239 |
| EPCAM | 2.452096 | 0.688961 | 0.999532 | | 2.381642 | 0.445652 | 0.751202 | | 2.569101 | -0.23048 | 0.890746 |
| ZRANB2 | 7.022913 | -0.21078 | 0.999532 | | 8.820443 | 0.264365 | 0.751832 | | 7.124083 | 0.476729 | 0.597289 |
| SLC3A2 | 11.43402 | -0.13827 | 0.999532 | | 15.25648 | 0.282453 | 0.752119 | | 11.84515 | 0.450251 | 0.600081 |
| MINDY3 | 4.057892 | -0.11581 | 0.999532 | | 4.503906 | -0.37357 | 0.752119 | | 3.516545 | -0.21745 | 0.886495 |
| SPRING1 | 3.813874 | -0.38059 | 0.999532 | | 4.186895 | -0.36518 | 0.752119 | | 3.197021 | 0.030435 | 0.991452 |
| GRIN2D | 2.679844 | -0.22897 | 0.999532 | | 2.763414 | -0.42171 | 0.752119 | | 2.194464 | -0.17742 | 0.926957 |
| IL6R | 2.272596 | -0.03251 | 0.999532 | | 2.275548 | -0.48991 | 0.752119 | | 1.970577 | -0.44372 | 0.79686 |
| LDB2 | 18.05622 | -0.14439 | 0.999532 | | 22.85484 | 0.248636 | 0.752119 | | 18.85144 | 0.41343 | 0.533876 |
| LMAN2L | 4.063404 | -0.21028 | 0.999532 | | 4.487641 | -0.37335 | 0.752119 | | 3.588393 | -0.14901 | 0.92697 |
| GAS6 | 9.20236 | -0.36603 | 0.999532 | | 12.98706 | 0.241236 | 0.752119 | | 9.667851 | 0.625596 | 0.300852 |
| SSTR4 | 9.279466 | -0.09255 | 0.999532 | | 9.721955 | -0.31854 | 0.752119 | | 8.038545 | -0.218 | 0.849682 |
| ORAI1 | 4.245023 | 0.006877 | 0.999532 | | 4.251359 | -0.35248 | 0.752119 | | 3.752479 | -0.34221 | 0.783005 |
| OR4A47 | 2.874837 | 0.473717 | 0.999532 | | 3.13705 | 0.427546 | 0.752119 | | 3.222703 | -0.03316 | 0.991159 |
| MTRNR2L | 2.164714 | -0.61887 | 0.999532 | | 2.502055 | -0.44473 | 0.752119 | | 1.681885 | 0.19188 | 0.928847 |
| ITGB1BP | 5.076682 | -0.12697 | 0.999532 | | 6.298884 | 0.306187 | 0.752119 | | 5.326804 | 0.460327 | 0.637292 |
| CHERP | 5.094932 | 0.076592 | 0.999532 | | 5.83713 | 0.310866 | 0.752119 | | 5.360375 | 0.239331 | 0.858899 |
| PDHB | 3.629591 | 0.392477 | 0.999532 | | 3.973802 | 0.379073 | 0.752306 | | 3.891684 | 0.009286 | 0.996891 |
| MSTO1 | 4.508813 | -0.04013 | 0.999532 | | 4.297759 | -0.34521 | 0.752306 | | 3.72422 | -0.29075 | 0.817974 |
| ASH1L | 5.656576 | 0.107575 | 0.999532 | | 6.727517 | 0.285647 | 0.752306 | | 5.903143 | 0.197013 | 0.865065 |
| CHST11 | 2.072954 | 0.230066 | 0.999532 | | 1.899097 | -0.51817 | 0.752306 | | 1.738763 | -0.72777 | 0.649779 |
| MORN5 | 2.179613 | -0.05214 | 0.999532 | | 2.266241 | -0.49665 | 0.752306 | | 1.910912 | -0.42461 | 0.816589 |
| BRAT1 | 3.435497 | 0.148886 | 0.999532 | | 3.889166 | 0.373258 | 0.752306 | | 3.585408 | 0.246483 | 0.862823 |
| SLC22A6 | 4.697485 | -0.36597 | 0.999532 | | 6.790875 | 0.323567 | 0.752306 | | 5.144287 | 0.702895 | 0.436389 |
| ZNF420 | 2.033759 | 0.447229 | 0.999532 | | 2.369206 | 0.465069 | 0.752306 | | 2.395358 | 0.035176 | 0.991159 |
| RNF10 | 3.43481 | 0.072755 | 0.999532 | | 4.272932 | 0.332878 | 0.752306 | | 3.704092 | 0.277131 | 0.825327 |
| RAB5IF | 4.705466 | -0.00943 | 0.999532 | | 5.750504 | 0.311804 | 0.752306 | | 4.923295 | 0.339666 | 0.749222 |
| WNT10B | 2.79371 | 0.150326 | 0.999532 | | 2.53994 | -0.45994 | 0.752534 | | 2.368472 | -0.60082 | 0.67176 |
| MEF2B | 2.538031 | 0.038804 | 0.999532 | | 2.248404 | -0.47503 | 0.752534 | | 2.06733 | -0.50822 | 0.75311 |
| ELOA2 | 8.485383 | 0.261259 | 0.999532 | | 8.109744 | -0.30934 | 0.752534 | | 7.378243 | -0.54891 | 0.485781 |
| KLHL2 | 2.608549 | -0.20452 | 0.999532 | | 3.81836 | 0.375845 | 0.752534 | | 3.018191 | 0.586927 | 0.626811 |
| FBXO34 | 5.196754 | 0.026371 | 0.999532 | | 6.330712 | 0.301025 | 0.752534 | | 5.401641 | 0.291088 | 0.791576 |
| MYADM | 9.326923 | -0.28588 | 0.999532 | | 12.98513 | 0.289027 | 0.752534 | | 9.674666 | 0.61445 | 0.421517 |
| TLCD3A | 3.229235 | -0.26102 | 0.999532 | | 3.207375 | -0.38872 | 0.752534 | | 2.69924 | -0.12098 | 0.950669 |
| TREM2 | 2.176806 | 0.328874 | 0.999532 | | 1.956407 | -0.50589 | 0.752534 | | 1.943193 | -0.81889 | 0.566812 |
| CCM2 | 1.963932 | 0.367394 | 0.999532 | | 2.045918 | 0.467445 | 0.752534 | | 2.168627 | 0.111957 | 0.96129 |
| PF4 | 3.345578 | -0.33144 | 0.999532 | | 3.647901 | -0.38166 | 0.752534 | | 2.919099 | -0.03658 | 0.989985 |
| BPY2 | 2.837187 | -0.22651 | 0.999532 | | 3.2015 | -0.42221 | 0.752807 | | 2.365997 | -0.15598 | 0.945456 |
| SLC16A7 | 2.313994 | 0.539087 | 0.999532 | | 1.823913 | -0.51694 | 0.752807 | | 1.951326 | -1.03639 | 0.433783 |
| TCEAL2 | 4.23571 | 0.433408 | 0.999532 | | 4.661041 | 0.337625 | 0.752807 | | 4.513462 | -0.07655 | 0.963702 |
| METTL16 | 4.387539 | -0.23872 | 0.999532 | | 4.694914 | -0.33444 | 0.752807 | | 3.692712 | -0.07571 | 0.966664 |
| ESPN | 8.123215 | 0.265852 | 0.999532 | | 7.584274 | -0.34157 | 0.752807 | | 6.844811 | -0.58219 | 0.48296 |
| RRP15 | 2.412383 | 0.389632 | 0.999532 | | 2.135511 | -0.50196 | 0.752807 | | 2.113336 | -0.87272 | 0.520848 |
| RNF169 | 2.215806 | 0.484183 | 0.999532 | | 2.55089 | 0.460584 | 0.752807 | | 2.484127 | -0.00254 | 0.999385 |
| DACT1 | 1.875785 | 0.008354 | 0.999532 | | 2.476572 | 0.453261 | 0.752807 | | 2.178335 | 0.464686 | 0.769793 |
| ITGA2B | 2.253024 | 0.445773 | 0.999532 | | 1.968193 | -0.49052 | 0.752807 | | 1.980837 | -0.91241 | 0.504265 |
| IRAK3 | 3.879751 | -0.46646 | 0.999532 | | 4.689718 | -0.34121 | 0.752807 | | 3.391241 | 0.147581 | 0.926957 |
| NOM1 | 3.383386 | -0.13357 | 0.999532 | | 3.468193 | -0.39315 | 0.752807 | | 2.802717 | -0.24449 | 0.883565 |
| MC1R | 2.837332 | -0.05954 | 0.999532 | | 2.740193 | -0.42878 | 0.752807 | | 2.363258 | -0.35195 | 0.824855 |
| ALG13 | 3.318963 | -0.046 | 0.999532 | | 3.478405 | -0.38724 | 0.752919 | | 2.770186 | -0.32519 | 0.821752 |
| SLC13A5 | 2.190067 | 0.13341 | 0.999532 | | 2.09188 | -0.48146 | 0.752919 | | 1.817057 | -0.58989 | 0.709589 |
| TXNDC16 | 2.499827 | -0.50561 | 0.999532 | | 2.804069 | -0.42093 | 0.752919 | | 1.937561 | 0.101695 | 0.965649 |
| NDEL1 | 4.335905 | -0.43803 | 0.999532 | | 6.461339 | 0.282822 | 0.752919 | | 4.891412 | 0.743168 | 0.348451 |
| PYDC5 | 4.015724 | 0.081389 | 0.999532 | | 3.780614 | -0.40569 | 0.752919 | | 3.42302 | -0.4556 | 0.738702 |
| SMIM32 | 1.967533 | 0.708822 | 0.999532 | | 2.008891 | 0.480723 | 0.752919 | | 2.216603 | -0.21033 | 0.907271 |
| TPR | 5.261887 | -0.6329 | 0.999532 | | 7.607373 | 0.284602 | 0.752919 | | 5.355797 | 0.93668 | 0.203949 |
| RPS6KL1 | 2.562123 | 0.00966 | 0.999532 | | 2.411734 | -0.46232 | 0.752919 | | 2.045733 | -0.45027 | 0.777828 |
| CLPTM1L | 4.769461 | -0.14706 | 0.999532 | | 6.075577 | 0.296103 | 0.752919 | | 4.863032 | 0.456493 | 0.646821 |
| ZNF736 | 3.073563 | 0.191278 | 0.999532 | | 3.830699 | 0.393177 | 0.753123 | | 3.642951 | 0.20886 | 0.893262 |
| C1QTNF1 | 5.848169 | -0.73863 | 0.999532 | | 7.306519 | -0.29087 | 0.753123 | | 4.833233 | 0.469897 | 0.632923 |
| CHN1 | 2.413737 | -0.39351 | 0.999532 | | 2.651715 | -0.45129 | 0.753123 | | 1.935845 | -0.04048 | 0.991103 |
| GNA11 | 6.120918 | -0.27305 | 0.999532 | | 6.772485 | -0.28771 | 0.753168 | | 4.904665 | -0.00162 | 0.999385 |
| KRT79 | 2.1574 | -0.14995 | 0.999532 | | 2.232292 | -0.4778 | 0.753248 | | 1.693504 | -0.30686 | 0.87591 |
| DNAJC9 | 2.994865 | 0.036439 | 0.999532 | | 3.789358 | 0.361479 | 0.753248 | | 3.120488 | 0.340982 | 0.796512 |
| ZNF609 | 7.311408 | 0.060416 | 0.999532 | | 8.747246 | 0.249942 | 0.753248 | | 7.221257 | 0.211335 | 0.820834 |
| SLC12A2 | 3.677386 | -0.12292 | 0.999532 | | 3.93712 | -0.36292 | 0.753248 | | 3.117201 | -0.21866 | 0.888142 |
| CCDC126 | 2.545428 | 0.186296 | 0.999532 | | 2.969116 | 0.400723 | 0.753248 | | 2.678424 | 0.232244 | 0.88476 |
| ARHGAP | 7.993185 | -0.08873 | 0.999532 | | 9.963974 | 0.245095 | 0.753982 | | 8.283602 | 0.348425 | 0.686214 |
| MAT2B | 8.133839 | 0.120897 | 0.999532 | | 9.886094 | 0.298683 | 0.754019 | | 8.735051 | 0.195434 | 0.851372 |
| PLCZ1 | 2.724896 | 0.020535 | 0.999532 | | 2.635678 | -0.42995 | 0.75506 | | 2.356565 | -0.43186 | 0.783332 |
| TARS1 | 3.569607 | 0.591925 | 0.999532 | | 3.655624 | 0.389495 | 0.75506 | | 3.549919 | -0.19258 | 0.892869 |
| MICU3 | 2.102333 | -0.1778 | 0.999532 | | 2.92049 | 0.427616 | 0.755129 | | 2.459423 | 0.618325 | 0.660958 |
| LTBP4 | 6.673702 | -0.22131 | 0.999532 | | 8.703828 | 0.29045 | 0.755143 | | 7.025413 | 0.538057 | 0.49299 |
| SAMD8 | 3.311343 | 0.241224 | 0.999532 | | 3.056272 | -0.39533 | 0.755143 | | 2.728486 | -0.61271 | 0.615428 |
| UXS1 | 6.921957 | -0.24242 | 0.999532 | | 7.656602 | -0.26027 | 0.755143 | | 5.915909 | 0.000593 | 0.999635 |
| SNX30 | 3.834678 | -0.19027 | 0.999532 | | 5.236305 | 0.316188 | 0.755143 | | 4.333158 | 0.525178 | 0.609671 |
| H3C6 | 2.061864 | 0.113554 | 0.999532 | | 1.949073 | -0.49162 | 0.755143 | | 1.79266 | -0.58851 | 0.717627 |
| FLRT3 | 5.399511 | 0.101387 | 0.999532 | | 6.708711 | 0.334761 | 0.755143 | | 5.630934 | 0.250979 | 0.816618 |
| CCDC78 | 2.602646 | 0.299696 | 0.999532 | | 2.320141 | -0.48043 | 0.755143 | | 2.278876 | -0.76031 | 0.598589 |
| N4BP1 | 2.771032 | 0.374654 | 0.999532 | | 3.054409 | 0.399948 | 0.755143 | | 3.004757 | 0.041679 | 0.987274 |
| ZNF426 | 8.270379 | 0.020643 | 0.999532 | | 9.932888 | 0.241694 | 0.755143 | | 8.641426 | 0.239802 | 0.784955 |
| SDCCAG | 4.075203 | 0.293337 | 0.999532 | | 4.942027 | 0.321646 | 0.755143 | | 4.305104 | 0.0353 | 0.98715 |
| AREL1 | 2.657139 | 0.672226 | 0.999532 | | 2.625965 | 0.41455 | 0.755304 | | 2.850923 | -0.24029 | 0.87591 |
| APAF1 | 2.55112 | 0.609473 | 0.999532 | | 2.877832 | 0.434515 | 0.755922 | | 2.862869 | -0.15538 | 0.929529 |
| CEP95 | 2.454705 | -0.22731 | 0.999532 | | 2.471641 | -0.43549 | 0.755922 | | 2.015423 | -0.19695 | 0.920259 |
| NBN | 2.832304 | -0.11705 | 0.999532 | | 3.721749 | 0.344004 | 0.755922 | | 3.134095 | 0.481111 | 0.686214 |
| CCT7 | 6.330446 | -0.1352 | 0.999532 | | 8.285897 | 0.271449 | 0.755922 | | 6.610761 | 0.425795 | 0.632113 |
| DMGDH | 2.160161 | 0.255266 | 0.999532 | | 2.743835 | 0.451308 | 0.756024 | | 2.387396 | 0.225063 | 0.901033 |
| FAM50B | 2.414201 | 0.237933 | 0.999532 | | 2.952368 | 0.424914 | 0.756024 | | 2.58151 | 0.213892 | 0.899732 |
| YTHDF2 | 4.499797 | 0.068973 | 0.999532 | | 5.40847 | 0.306204 | 0.756024 | | 4.794386 | 0.236456 | 0.846488 |
| PRAMEF | 3.676232 | 0.317302 | 0.999532 | | 3.517135 | -0.39527 | 0.756041 | | 3.2308 | -0.668 | 0.568022 |
| GINS4 | 2.635006 | -0.21843 | 0.999532 | | 2.625923 | -0.41869 | 0.756041 | | 2.23628 | -0.18471 | 0.923864 |
| MPND | 2.995625 | 0.302259 | 0.999532 | | 2.746971 | -0.42391 | 0.756304 | | 2.643817 | -0.69291 | 0.598589 |
| EFNB3 | 2.571823 | 0.392114 | 0.999532 | | 2.270887 | -0.46145 | 0.756304 | | 2.105391 | -0.83161 | 0.522078 |
| CXorf49 | 2.081802 | 0.025588 | 0.999532 | | 2.077399 | -0.48135 | 0.756304 | | 1.7599 | -0.48271 | 0.780904 |
| ZNF619 | 2.490036 | -0.41088 | 0.999532 | | 2.970181 | -0.42328 | 0.756304 | | 2.061124 | 0.020293 | 0.996681 |
| IFNL1 | 2.248829 | -0.07497 | 0.999532 | | 2.225215 | -0.47738 | 0.756459 | | 1.866092 | -0.38553 | 0.82977 |
| GPR27 | 2.888697 | -0.46012 | 0.999532 | | 3.316876 | -0.41085 | 0.75691 | | 2.51514 | 0.059616 | 0.979967 |
| BMF | 2.476403 | -0.39145 | 0.999532 | | 2.633415 | -0.42159 | 0.75691 | | 2.009268 | -0.01348 | 0.996891 |
| CCL1 | 1.999806 | 0.925816 | 0.999532 | | 2.027016 | 0.51216 | 0.757002 | | 2.258167 | -0.3925 | 0.815509 |
| TSSK2 | 1.953733 | -0.01159 | 0.999532 | | 1.869022 | -0.51133 | 0.757277 | | 1.618001 | -0.48096 | 0.796937 |
| GPHA2 | 2.502081 | -0.4854 | 0.999532 | | 2.829952 | -0.42395 | 0.757277 | | 2.001308 | 0.080341 | 0.974421 |
| STX1B | 13.00749 | 0.119929 | 0.999532 | | 12.74327 | -0.3115 | 0.757277 | | 11.59654 | -0.43237 | 0.666906 |
| CIDEA | 2.978073 | 0.524273 | 0.999532 | | 2.353194 | -0.46377 | 0.757277 | | 2.594779 | -0.97423 | 0.414073 |
| FBN2 | 2.063909 | 0.111313 | 0.999532 | | 1.946827 | -0.50505 | 0.757277 | | 1.769204 | -0.60045 | 0.71841 |
| POLDIP3 | 4.023252 | 0.054976 | 0.999532 | | 4.991629 | 0.327456 | 0.75739 | | 4.211227 | 0.295982 | 0.806394 |
| POMZP3 | 12.2993 | -0.34124 | 0.999532 | | 13.98793 | -0.21266 | 0.75739 | | 10.61666 | 0.149511 | 0.865065 |
| NIPA2 | 5.241962 | -0.04421 | 0.999532 | | 6.322795 | 0.290113 | 0.75739 | | 5.324067 | 0.351732 | 0.723294 |
| GNPTG | 7.185534 | -0.25175 | 0.999532 | | 9.721649 | 0.239197 | 0.75739 | | 7.524564 | 0.511193 | 0.495181 |
| CNTN1 | 4.707015 | -0.09564 | 0.999532 | | 6.696984 | 0.340357 | 0.75739 | | 5.341986 | 0.487586 | 0.668083 |
| AURKB | 2.81531 | 0.103743 | 0.999532 | | 2.876297 | -0.44077 | 0.75739 | | 2.604023 | -0.51974 | 0.722703 |
| TRA2A | 8.840388 | -0.06344 | 0.999532 | | 11.2407 | 0.22823 | 0.758258 | | 9.190248 | 0.301951 | 0.718643 |
| ZNF280D | 5.444439 | -0.10709 | 0.999532 | | 7.150342 | 0.280574 | 0.758258 | | 5.636269 | 0.406028 | 0.666566 |
| SMCHD1 | 3.295179 | 0.544965 | 0.999532 | | 3.479357 | 0.367686 | 0.758258 | | 3.511315 | -0.15723 | 0.915533 |
| VASN | 12.37772 | 0.353253 | 0.999532 | | 12.9906 | 0.224327 | 0.758258 | | 12.24359 | -0.10766 | 0.907586 |
| SDHAF3 | 3.252611 | 0.11917 | 0.999532 | | 2.973179 | -0.3982 | 0.758386 | | 2.777477 | -0.50395 | 0.704316 |
| CCT6A | 9.840263 | 0.039574 | 0.999532 | | 11.82118 | 0.24112 | 0.758386 | | 10.36185 | 0.214678 | 0.807942 |
| ORAI3 | 4.558839 | 0.181883 | 0.999532 | | 4.373339 | -0.35804 | 0.758386 | | 4.026553 | -0.52456 | 0.634624 |
| SYCP1 | 2.417673 | -0.16112 | 0.999532 | | 2.685849 | -0.44264 | 0.758477 | | 2.153212 | -0.2495 | 0.89689 |
| NMUR2 | 2.7408 | -0.16331 | 0.999532 | | 2.971696 | -0.45057 | 0.758486 | | 2.310664 | -0.27341 | 0.876781 |
| FUT5 | 2.357052 | -0.00922 | 0.999532 | | 2.310345 | -0.4705 | 0.758486 | | 2.015916 | -0.44414 | 0.792538 |
| KNTC1 | 2.744956 | 0.151283 | 0.999532 | | 2.473506 | -0.42997 | 0.758486 | | 2.221541 | -0.56544 | 0.68597 |
| XAGE3 | 2.728428 | 0.220277 | 0.999532 | | 2.305257 | -0.45021 | 0.758486 | | 2.354432 | -0.66112 | 0.650636 |
| NPM2 | 3.850553 | -0.20535 | 0.999532 | | 3.956507 | -0.34313 | 0.758626 | | 3.17868 | -0.12372 | 0.941197 |
| TRPM7 | 2.34184 | 0.178609 | 0.999532 | | 2.700821 | 0.40976 | 0.758812 | | 2.464653 | 0.243296 | 0.88476 |
| ORMDL2 | 3.665284 | -0.09799 | 0.999532 | | 4.618097 | 0.315276 | 0.758812 | | 4.063465 | 0.435728 | 0.682485 |
| MVB12B | 4.268505 | -0.12748 | 0.999532 | | 4.464655 | -0.34125 | 0.758812 | | 3.737209 | -0.19684 | 0.895807 |
| KIAA0930 | 3.706549 | 0.602262 | 0.999532 | | 3.70647 | 0.36901 | 0.758812 | | 3.992397 | -0.21881 | 0.872591 |
| FAT3 | 2.207235 | 0.622971 | 0.999532 | | 2.265536 | 0.462124 | 0.758812 | | 2.33269 | -0.14438 | 0.942744 |
| HIVEP1 | 3.492169 | 0.247195 | 0.999532 | | 3.089026 | -0.41303 | 0.758812 | | 2.758221 | -0.63679 | 0.600081 |
| PSTPIP1 | 2.115811 | -0.194 | 0.999532 | | 2.280875 | -0.46227 | 0.758996 | | 1.825941 | -0.24963 | 0.900583 |
| HOMER3 | 6.103717 | -0.03525 | 0.999532 | | 7.543786 | 0.26742 | 0.758996 | | 6.497625 | 0.319609 | 0.728015 |
| BANF1 | 10.15859 | 0.141603 | 0.999532 | | 12.0436 | 0.218109 | 0.758996 | | 10.29001 | 0.09336 | 0.920259 |
| CCL7 | 2.367232 | 0.128829 | 0.999532 | | 2.365278 | -0.47033 | 0.759121 | | 2.10963 | -0.57801 | 0.709825 |
| PNMA8B | 2.472697 | -0.20111 | 0.999532 | | 2.480306 | -0.43906 | 0.759121 | | 2.006655 | -0.22098 | 0.908595 |
| ZNF33B | 3.508804 | -0.26492 | 0.999532 | | 3.740554 | -0.35683 | 0.759121 | | 2.876097 | -0.07526 | 0.972924 |
| ECEL1 | 2.792623 | 0.423634 | 0.999532 | | 2.328149 | -0.4539 | 0.759121 | | 2.43497 | -0.86251 | 0.493137 |
| MAGI2 | 2.703567 | 0.444116 | 0.999532 | | 2.377044 | -0.4594 | 0.759121 | | 2.221515 | -0.87791 | 0.487828 |
| TGFBRA | 2.985941 | -0.29387 | 0.999532 | | 3.159049 | -0.38814 | 0.759121 | | 2.636327 | -0.07332 | 0.974482 |
| GNPDA1 | 4.370095 | -0.1455 | 0.999532 | | 5.611016 | 0.302477 | 0.759121 | | 4.518301 | 0.463492 | 0.667066 |
| SNRPB | 3.334532 | 0.150177 | 0.999532 | | 4.042008 | 0.351665 | 0.75918 | | 3.638588 | 0.21986 | 0.874653 |
| CCDC96 | 3.137244 | -0.24375 | 0.999532 | | 3.217707 | -0.47749 | 0.75918 | | 2.540659 | -0.21138 | 0.917499 |
| MON1B | 5.960744 | 0.426959 | 0.999532 | | 6.35753 | 0.279749 | 0.75918 | | 6.328735 | -0.12789 | 0.908829 |
| RAPGEF | 2.734516 | 0.418396 | 0.999532 | | 2.860364 | 0.40237 | 0.75918 | | 2.877097 | -0.00674 | 0.997967 |
| ARHGEF | 3.811073 | -0.01449 | 0.999532 | | 4.779356 | 0.321165 | 0.75918 | | 4.101497 | 0.354516 | 0.755977 |
| SNURF | 6.731263 | -0.15813 | 0.999532 | | 9.075884 | 0.248414 | 0.75918 | | 7.139113 | 0.423034 | 0.598329 |
| TXNL4A | 4.108156 | 0.772241 | 0.999532 | | 3.736716 | 0.349218 | 0.75918 | | 4.198995 | -0.40552 | 0.700983 |
| CEP350 | 2.787377 | -0.8102 | 0.999532 | | 3.577335 | -0.37932 | 0.75918 | | 2.280491 | 0.450265 | 0.759369 |
| RNF165 | 3.740175 | 0.282268 | 0.999532 | | 3.466615 | -0.38377 | 0.75918 | | 3.323005 | -0.6392 | 0.598329 |
| FAM98A | 6.962076 | -0.09653 | 0.999532 | | 9.14609 | 0.268878 | 0.75918 | | 7.392215 | 0.394782 | 0.664512 |
| UNC119B | 2.91426 | 0.288726 | 0.999532 | | 3.27904 | 0.378303 | 0.75918 | | 3.070151 | 0.107538 | 0.951656 |
| TARP | 3.00513 | -0.20746 | 0.999532 | | 3.123777 | -0.41078 | 0.75918 | | 2.625131 | -0.1873 | 0.918194 |
| CARD19 | 3.031916 | 0.332862 | 0.999532 | | 3.45203 | 0.365507 | 0.759818 | | 3.380799 | 0.05446 | 0.978282 |
| CCDC61 | 2.850593 | 0.330969 | 0.999532 | | 3.149209 | 0.382077 | 0.759847 | | 3.033711 | 0.06762 | 0.974421 |
| TLCD2 | 2.496938 | 0.351832 | 0.999532 | | 2.08904 | -0.46707 | 0.759847 | | 2.097028 | -0.8007 | 0.557909 |
| BIN2 | 2.589144 | -0.47409 | 0.999532 | | 2.991657 | -0.40399 | 0.759847 | | 2.264024 | 0.08934 | 0.969783 |
| LAMA4 | 2.82224 | 0.175771 | 0.999532 | | 3.261956 | 0.385156 | 0.759847 | | 3.07152 | 0.219845 | 0.892004 |
| NOP14 | 2.566859 | 0.462044 | 0.999532 | | 2.996619 | 0.411254 | 0.759924 | | 2.855995 | -0.02779 | 0.992286 |
| DENND4C | 4.137414 | 0.010855 | 0.999532 | | 5.071524 | 0.320391 | 0.759924 | | 4.385435 | 0.333167 | 0.769793 |
| C14orf28 | 2.090821 | 0.634069 | 0.999532 | | 2.393814 | 0.452437 | 0.759953 | | 2.353978 | -0.15767 | 0.934597 |
| DMXL2 | 6.1825 | -0.27986 | 0.999532 | | 7.826676 | 0.257542 | 0.759953 | | 6.38299 | 0.543947 | 0.515762 |
| HGD | 2.986722 | 0.500194 | 0.999532 | | 3.11629 | 0.386667 | 0.759953 | | 3.141726 | -0.09446 | 0.960254 |
| AQP9 | 2.659462 | -1.00463 | 0.999532 | | 3.399524 | -0.37916 | 0.759953 | | 2.146808 | 0.635413 | 0.666566 |
| SCGB1C2 | 2.173742 | 0.116851 | 0.999532 | | 2.195482 | -0.47906 | 0.759953 | | 1.844369 | -0.57262 | 0.717627 |
| FBXO48 | 2.496481 | 0.569325 | 0.999532 | | 2.168728 | -0.50552 | 0.759953 | | 2.08683 | -1.04612 | 0.421517 |
| PSEN2 | 2.821636 | 0.235865 | 0.999532 | | 3.467365 | 0.374624 | 0.759953 | | 3.225982 | 0.156057 | 0.922679 |
| CD52 | 2.708377 | -0.35195 | 0.999532 | | 2.883585 | -0.39193 | 0.759953 | | 2.259731 | -0.02304 | 0.994617 |
| ATP5PD | 15.80086 | 0.188731 | 0.999532 | | 18.12911 | 0.195922 | 0.759953 | | 15.89209 | 0.026729 | 0.982455 |
| FMOD | 2.76046 | -0.24143 | 0.999532 | | 2.941462 | -0.40345 | 0.759953 | | 2.287158 | -0.14326 | 0.941197 |
| SETD9 | 2.057564 | 0.182342 | 0.999532 | | 2.527972 | 0.434901 | 0.759953 | | 2.301048 | 0.271547 | 0.877268 |
| BNC2 | 1.927512 | 0.046635 | 0.999532 | | 2.256642 | 0.443237 | 0.760336 | | 2.124367 | 0.410197 | 0.806394 |
| PDE4DIP | 5.780306 | 0.365763 | 0.999532 | | 6.550052 | 0.310065 | 0.760336 | | 6.033414 | -0.03608 | 0.982338 |
| GLRA2 | 1.892405 | -0.42963 | 0.999532 | | 2.006811 | -0.49151 | 0.760675 | | 1.463717 | -0.04411 | 0.991134 |
| SLC5A9 | 2.434422 | 0.589927 | 0.999532 | | 2.350566 | 0.448978 | 0.761121 | | 2.606119 | -0.11775 | 0.955834 |
| SCD5 | 4.500558 | -0.11976 | 0.999532 | | 4.82538 | -0.33019 | 0.761121 | | 3.891386 | -0.18886 | 0.893262 |
| ATPAF2 | 2.363585 | 0.011176 | 0.999532 | | 3.01112 | 0.383541 | 0.761121 | | 2.559647 | 0.390977 | 0.789815 |
| ZMYM1 | 2.428136 | 0.726023 | 0.999532 | | 2.272467 | 0.460096 | 0.761142 | | 2.531849 | -0.25931 | 0.885495 |
| ZDHHC19 | 1.911037 | 0.564441 | 0.999532 | | 2.007089 | 0.471008 | 0.761975 | | 2.07113 | -0.0741 | 0.975183 |
| ARAP3 | 3.308975 | 0.299865 | 0.999532 | | 3.923504 | 0.380044 | 0.761975 | | 3.625762 | 0.072693 | 0.970268 |
| ACSF3 | 3.692854 | 0.233413 | 0.999532 | | 3.49203 | -0.36962 | 0.761975 | | 3.197359 | -0.58016 | 0.628985 |
| TMF1 | 5.439616 | -0.15247 | 0.999532 | | 7.15603 | 0.259717 | 0.762006 | | 5.557169 | 0.42331 | 0.644077 |
| NOTCH3 | 9.333758 | -0.40333 | 0.999532 | | 12.93963 | 0.259141 | 0.762124 | | 10.00821 | 0.680696 | 0.281423 |
| GVQW3 | 2.613206 | -0.597 | 0.999532 | | 3.064889 | -0.449 | 0.762181 | | 2.163759 | 0.164772 | 0.941197 |
| MBLAC2 | 2.234593 | 0.463532 | 0.999532 | | 1.983933 | -0.48572 | 0.762344 | | 1.916837 | -0.92501 | 0.488953 |
| GDE1 | 4.688101 | -0.4471 | 0.999532 | | 6.775624 | 0.285942 | 0.762415 | | 5.048216 | 0.746451 | 0.337353 |
| CNNM3 | 4.768962 | 0.221619 | 0.999532 | | 5.595783 | 0.291312 | 0.762415 | | 5.094777 | 0.091358 | 0.948097 |
| AHCYL1 | 12.14291 | -0.18693 | 0.999532 | | 15.4523 | 0.196446 | 0.762415 | | 12.30639 | 0.40144 | 0.515762 |
| IRF5 | 2.693471 | -0.07622 | 0.999532 | | 2.642119 | -0.41474 | 0.762415 | | 2.243283 | -0.32202 | 0.849128 |
| CLRN3 | 2.320914 | 0.115018 | 0.999532 | | 2.205379 | -0.47062 | 0.762415 | | 2.036683 | -0.56588 | 0.722703 |
| FADS2 | 3.638753 | 0.138419 | 0.999532 | | 4.213748 | 0.372638 | 0.762415 | | 3.918613 | 0.27887 | 0.833604 |
| BAZ1A | 3.161898 | 0.432588 | 0.999532 | | 2.518073 | -0.4365 | 0.762415 | | 2.614107 | -0.86143 | 0.481527 |
| PKDCC | 2.999639 | 0.154118 | 0.999532 | | 3.549457 | 0.359294 | 0.762415 | | 3.269248 | 0.226502 | 0.877268 |
| TUT1 | 3.269196 | 0.392538 | 0.999532 | | 3.641722 | 0.358614 | 0.762501 | | 3.432057 | -0.01418 | 0.996681 |
| FAIM | 3.459305 | 0.341975 | 0.999532 | | 2.976803 | -0.40821 | 0.762784 | | 3.055302 | -0.73427 | 0.574633 |
| TMEM98 | 4.841105 | 0.020203 | 0.999532 | | 5.893609 | 0.29176 | 0.762807 | | 5.201075 | 0.292283 | 0.795612 |
| JKAMP | 4.028175 | 0.69374 | 0.999532 | | 3.994047 | 0.345534 | 0.762807 | | 4.275789 | -0.32886 | 0.765012 |
| CHSY3 | 3.112449 | 0.088782 | 0.999532 | | 2.867476 | -0.39916 | 0.763072 | | 2.65724 | -0.48159 | 0.734836 |
| ZNF324 | 4.141588 | -0.34553 | 0.999532 | | 4.51959 | -0.34815 | 0.763072 | | 3.489819 | 0.009631 | 0.996891 |
| DEPDC4 | 2.567373 | -0.3294 | 0.999532 | | 2.86978 | -0.43165 | 0.763072 | | 2.209681 | -0.08495 | 0.974421 |
| IL17C | 2.686425 | -0.24983 | 0.999532 | | 2.694047 | -0.43484 | 0.763072 | | 2.146065 | -0.17443 | 0.934354 |
| KLHL6 | 2.430238 | -0.28884 | 0.999532 | | 2.697752 | -0.43122 | 0.763072 | | 2.15001 | -0.12975 | 0.952656 |
| KLC4 | 3.285998 | 0.588429 | 0.999532 | | 3.297759 | 0.40296 | 0.763072 | | 3.294881 | -0.15666 | 0.920181 |
| BCS1L | 3.754278 | -0.3214 | 0.999532 | | 4.261536 | -0.33924 | 0.763072 | | 3.167644 | -0.00169 | 0.999429 |
| ZPR1 | 3.699516 | -0.4444 | 0.999532 | | 4.265857 | -0.38744 | 0.763072 | | 2.936339 | 0.08134 | 0.969614 |
| SEL1L3 | 2.540721 | -0.11129 | 0.999532 | | 2.614186 | -0.43799 | 0.763534 | | 2.16213 | -0.30642 | 0.867009 |
| HCG22 | 2.117522 | -0.47646 | 0.999532 | | 2.298986 | -0.4447 | 0.763534 | | 1.771371 | 0.040467 | 0.991273 |
| ZNF841 | 5.888697 | 0.063086 | 0.999532 | | 5.680014 | -0.30631 | 0.763534 | | 4.933801 | -0.35101 | 0.736481 |
| ATIC | 3.043219 | -0.15577 | 0.999532 | | 4.17037 | 0.341925 | 0.763534 | | 3.432609 | 0.515591 | 0.658914 |
| CTSH | 4.38519 | -0.31431 | 0.999532 | | 5.672562 | 0.329551 | 0.763534 | | 4.42011 | 0.633582 | 0.568257 |
| CDRT15 | 2.786672 | 0.563924 | 0.999532 | | 2.336604 | -0.46988 | 0.763534 | | 2.466112 | -1.01797 | 0.386771 |
| GUCY2F | 3.331205 | 0.157873 | 0.999532 | | 3.059315 | -0.38447 | 0.763559 | | 2.882812 | -0.52503 | 0.668553 |
| SPOP | 4.512845 | -0.00739 | 0.999532 | | 5.58603 | 0.295664 | 0.763559 | | 4.700442 | 0.319606 | 0.774747 |
| RHOT2 | 5.292446 | 0.211959 | 0.999532 | | 6.352792 | 0.27576 | 0.763559 | | 5.712767 | 0.088212 | 0.948127 |
| OGT | 7.143514 | 0.657116 | 0.999532 | | 6.77526 | 0.287383 | 0.763611 | | 7.259871 | -0.34584 | 0.682165 |
| ABCB9 | 4.51871 | -0.21172 | 0.999532 | | 4.81111 | -0.32419 | 0.763611 | | 3.713032 | -0.09926 | 0.954087 |
| RB1 | 2.632112 | -0.0971 | 0.999532 | | 3.540223 | 0.360134 | 0.763792 | | 2.903841 | 0.473841 | 0.713173 |
| DNAJC25 | 2.192848 | -0.18476 | 0.999532 | | 2.257523 | -0.46438 | 0.763792 | | 1.783032 | -0.26053 | 0.895246 |
| OR5C74 | 2.553685 | 0.017682 | 0.999532 | | 2.607338 | -0.41965 | 0.763792 | | 2.173492 | -0.41569 | 0.788935 |
| CENPT | 5.252115 | 0.082787 | 0.999532 | | 6.454592 | 0.303594 | 0.763792 | | 5.806698 | 0.235994 | 0.840285 |
| ODC1 | 4.190596 | 0.178942 | 0.999532 | | 5.04545 | 0.364095 | 0.763792 | | 4.513685 | 0.210104 | 0.877238 |
| SBK2 | 2.391026 | 0.18388 | 0.999532 | | 2.098212 | -0.48996 | 0.763792 | | 2.069688 | -0.65225 | 0.686214 |
| HBEGF | 4.585677 | -0.77233 | 0.999532 | | 5.948765 | -0.28603 | 0.763792 | | 3.753706 | 0.502304 | 0.647218 |
| TMEM244 | 2.468536 | 0.503197 | 0.999532 | | 2.706408 | 0.42234 | 0.763792 | | 2.714335 | -0.06616 | 0.975194 |
| CSNK1A1 | 5.531172 | -0.28107 | 0.999532 | | 7.587183 | 0.26614 | 0.763792 | | 5.829306 | 0.566373 | 0.489933 |
| LEMD3 | 6.392215 | 0.014703 | 0.999532 | | 7.645093 | 0.267639 | 0.764056 | | 6.80944 | 0.2635 | 0.794256 |
| DTNA | 2.380196 | 0.034388 | 0.999532 | | 2.394745 | -0.46482 | 0.764366 | | 1.950315 | -0.48 | 0.761337 |
| ZNF77 | 3.152394 | 0.076762 | 0.999532 | | 2.926613 | -0.41543 | 0.764366 | | 2.654771 | -0.47532 | 0.737426 |
| KRTAP12 | 3.328325 | -0.10704 | 0.999532 | | 3.259571 | -0.38771 | 0.764517 | | 2.699823 | -0.26435 | 0.870663 |
| SMAD6 | 10.85593 | -0.10208 | 0.999532 | | 13.6586 | 0.258654 | 0.764517 | | 11.25775 | 0.380824 | 0.649779 |
| SEC24D | 2.721739 | 0.087246 | 0.999532 | | 3.13126 | 0.371259 | 0.764833 | | 2.845068 | 0.289973 | 0.854282 |
| SERP1 | 4.355888 | -0.17403 | 0.999532 | | 5.858428 | 0.294392 | 0.764833 | | 4.862087 | 0.498594 | 0.603912 |
| TOMM34 | 3.600398 | 0.201728 | 0.999532 | | 4.218693 | 0.334613 | 0.764833 | | 3.753459 | 0.15047 | 0.917362 |
| CMPK1 | 3.256782 | -0.15611 | 0.999532 | | 3.377165 | -0.39614 | 0.764869 | | 2.754437 | -0.22084 | 0.892674 |
| OR4F17 | 3.424558 | -0.17564 | 0.999532 | | 3.509844 | -0.38534 | 0.765292 | | 2.896628 | -0.20654 | 0.907968 |
| TICAM1 | 4.813234 | -0.49434 | 0.999532 | | 5.669125 | -0.2998 | 0.765312 | | 4.236908 | 0.207974 | 0.885783 |
| AQP3 | 6.596668 | -0.26893 | 0.999532 | | 8.59643 | 0.257435 | 0.765312 | | 6.749512 | 0.545263 | 0.49299 |
| FZD4 | 15.53348 | -0.41635 | 0.999532 | | 18.9633 | -0.20451 | 0.765312 | | 13.82611 | 0.239558 | 0.75038 |
| MFSD8 | 3.621235 | 0.322315 | 0.999532 | | 3.093551 | -0.38423 | 0.765351 | | 3.19139 | -0.68962 | 0.547092 |
| CIDEC | 3.980758 | 0.163199 | 0.999532 | | 3.540157 | -0.36111 | 0.765351 | | 3.289611 | -0.51415 | 0.663412 |
| WRAP73 | 3.965338 | -0.05728 | 0.999532 | | 4.863432 | 0.325048 | 0.765351 | | 4.108233 | 0.399537 | 0.724386 |
| ZBTB46 | 7.322379 | 0.019232 | 0.999532 | | 8.693278 | 0.244917 | 0.765351 | | 7.70794 | 0.268764 | 0.765852 |
| FANCE | 3.144705 | 0.265503 | 0.999532 | | 2.737649 | -0.43307 | 0.765351 | | 2.779699 | -0.67799 | 0.602721 |
| TMPRSS1 | 2.865561 | 0.059621 | 0.999532 | | 2.888109 | -0.42197 | 0.765351 | | 2.544892 | -0.46757 | 0.74314 |
| TSC2 | 4.454554 | 0.11635 | 0.999532 | | 5.580493 | 0.2991 | 0.765351 | | 4.661952 | 0.20893 | 0.872591 |
| RNASEL | 2.698352 | 0.226295 | 0.999532 | | 3.275266 | 0.381031 | 0.765351 | | 2.941664 | 0.176032 | 0.914687 |
| SLC32A1 | 2.548804 | -0.09524 | 0.999532 | | 2.53562 | -0.47138 | 0.765351 | | 2.176349 | -0.35751 | 0.858767 |
| UNKL | 2.379324 | -0.23076 | 0.999532 | | 2.498622 | -0.45304 | 0.765351 | | 2.015099 | -0.2024 | 0.920124 |
| INSL4 | 2.369662 | -0.16436 | 0.999532 | | 2.45853 | -0.444 | 0.765351 | | 2.080841 | -0.25788 | 0.895721 |
| REXO5 | 2.295329 | -0.23605 | 0.999532 | | 2.465498 | -0.44039 | 0.765636 | | 1.897069 | -0.1797 | 0.932589 |
| TCTN2 | 3.009537 | 0.14883 | 0.999532 | | 3.79097 | 0.384695 | 0.765992 | | 3.547846 | 0.252373 | 0.872561 |
| CXCR1 | 2.177986 | -0.31504 | 0.999532 | | 2.289451 | -0.48678 | 0.766022 | | 1.910522 | -0.16366 | 0.947515 |
| TRAPPC1 | 1.844441 | 0.296755 | 0.999532 | | 2.286424 | 0.445858 | 0.76611 | | 2.142633 | 0.169035 | 0.933547 |
| PRODH2 | 7.052226 | 0.310685 | 0.999532 | | 8.057701 | 0.340557 | 0.766531 | | 7.324735 | 0.051586 | 0.976848 |
| SH3GLB1 | 12.40176 | -0.50727 | 0.999532 | | 17.19181 | 0.21175 | 0.766531 | | 12.62733 | 0.742421 | 0.145782 |
| RTL8C | 18.83994 | -0.02916 | 0.999532 | | 23.02723 | 0.215217 | 0.766531 | | 19.36258 | 0.272932 | 0.699971 |
| NDUFA6 | 4.292048 | 0.250981 | 0.999532 | | 4.876322 | 0.319224 | 0.766531 | | 4.606134 | 0.090653 | 0.950751 |
| ZFP82 | 4.189675 | -0.77018 | 0.999532 | | 5.370504 | -0.29845 | 0.766602 | | 3.735254 | 0.492207 | 0.671984 |
| KCNQ4 | 2.698073 | 0.152614 | 0.999532 | | 2.460839 | -0.44368 | 0.766602 | | 2.239382 | -0.58044 | 0.706407 |
| STAR | 2.43249 | 0.653705 | 0.999532 | | 1.910335 | -0.49914 | 0.766602 | | 2.122786 | -1.1336 | 0.359638 |
| MATN3 | 3.09262 | 0.111326 | 0.999532 | | 3.008172 | -0.39857 | 0.766602 | | 2.710024 | -0.49279 | 0.713778 |
| GCM2 | 2.654204 | 0.540059 | 0.999532 | | 2.204105 | -0.47597 | 0.766602 | | 2.294379 | -0.98914 | 0.43621 |
| HK3 | 2.922632 | -0.56678 | 0.999532 | | 3.425677 | -0.36657 | 0.766602 | | 2.493525 | 0.218275 | 0.900848 |
| GDA | 2.785573 | -0.03682 | 0.999532 | | 3.691018 | 0.359185 | 0.766602 | | 3.007325 | 0.409247 | 0.742839 |
| STARD3 | 4.848061 | -0.20289 | 0.999532 | | 6.69882 | 0.274015 | 0.766602 | | 5.339904 | 0.49895 | 0.575703 |
| REPS1 | 3.73713 | 0.180334 | 0.999532 | | 4.381848 | 0.33118 | 0.766602 | | 3.882045 | 0.169004 | 0.901033 |
| CCDC152 | 2.130847 | 0.132682 | 0.999532 | | 1.91705 | -0.49211 | 0.766602 | | 1.892329 | -0.60793 | 0.717627 |
| CLEC7A | 1.804397 | 0.640244 | 0.999532 | | 1.946712 | 0.480625 | 0.766602 | | 1.999116 | -0.14093 | 0.946849 |
| HOXC8 | 3.123681 | 0.05646 | 0.999532 | | 3.081014 | -0.38632 | 0.766602 | | 2.643565 | -0.42298 | 0.760473 |
| FAM13B | 8.123324 | 0.331573 | 0.999532 | | 8.729523 | 0.242845 | 0.766602 | | 8.488116 | -0.0647 | 0.963866 |
| CHFR | 2.59043 | 0.070788 | 0.999532 | | 2.431645 | -0.4276 | 0.766602 | | 2.09407 | -0.48176 | 0.743246 |
| NREP | 8.736993 | -0.31267 | 0.999532 | | 12.44146 | 0.277463 | 0.766602 | | 9.289274 | 0.653512 | 0.456688 |
| BCLAF1 | 6.002002 | 0.270946 | 0.999532 | | 5.412524 | -0.30471 | 0.766602 | | 4.981895 | -0.55432 | 0.535662 |
| H2AC15 | 2.069831 | -0.07247 | 0.999532 | | 2.121842 | -0.45193 | 0.766602 | | 1.767572 | -0.35801 | 0.843564 |
| AVPR1A | 4.48457 | -0.26077 | 0.999532 | | 4.91289 | -0.36646 | 0.766602 | | 3.69413 | -0.07951 | 0.968434 |
| IL16 | 3.392238 | 0.074095 | 0.999532 | | 3.260589 | -0.38423 | 0.766602 | | 2.922702 | -0.43943 | 0.740755 |
| SBNO1 | 3.672759 | 0.25074 | 0.999532 | | 3.974285 | 0.338391 | 0.766602 | | 3.748712 | 0.099058 | 0.954538 |
| CDX4 | 2.329539 | 0.114327 | 0.999532 | | 2.244271 | -0.45731 | 0.766602 | | 2.067217 | -0.55091 | 0.72206 |
| STYX | 3.35597 | -0.06979 | 0.999532 | | 3.478162 | -0.37717 | 0.766602 | | 2.858282 | -0.28608 | 0.855088 |
| FBH1 | 6.164465 | -0.1757 | 0.999532 | | 7.636473 | 0.256387 | 0.766609 | | 6.497227 | 0.441429 | 0.635381 |
| NOLC1 | 3.776912 | -0.34508 | 0.999532 | | 4.096924 | -0.34764 | 0.767151 | | 3.100717 | 0.00555 | 0.99853 |
| MINDY4B | 2.158882 | 0.363837 | 0.999532 | | 1.849311 | -0.49231 | 0.767151 | | 1.831707 | -0.83865 | 0.566812 |
| ZNF385B | 2.281587 | 0.765255 | 0.999532 | | 2.120893 | 0.457052 | 0.767157 | | 2.35342 | -0.29692 | 0.863401 |
| SERPINE | 2.142793 | -0.07772 | 0.999532 | | 2.035555 | -0.47288 | 0.767377 | | 1.790385 | -0.38429 | 0.842754 |
| HID1 | 6.313095 | 0.391656 | 0.999532 | | 5.429087 | -0.3087 | 0.767377 | | 5.546498 | -0.68378 | 0.371531 |
| MPV17L2 | 2.860621 | 0.277817 | 0.999532 | | 2.402486 | -0.42479 | 0.767581 | | 2.417556 | -0.69203 | 0.602029 |
| GSPT1 | 3.345164 | -0.11566 | 0.999532 | | 4.490772 | 0.347647 | 0.767783 | | 3.628936 | 0.488 | 0.684642 |
| C17orf10 | 2.703393 | 0.329829 | 0.999532 | | 2.276493 | -0.45031 | 0.767865 | | 2.275846 | -0.76402 | 0.566812 |
| SPHK2 | 4.221742 | 0.646068 | 0.999532 | | 4.095528 | 0.343544 | 0.767865 | | 4.40729 | -0.28632 | 0.809832 |
| GABPA | 7.426312 | 0.215441 | 0.999532 | | 8.584758 | 0.247259 | 0.768032 | | 7.788824 | 0.051744 | 0.971683 |
| ZNF836 | 3.069524 | -0.14196 | 0.999532 | | 3.321207 | -0.40239 | 0.768106 | | 2.706018 | -0.2333 | 0.895421 |
| NBPF6 | 1.827623 | -0.2762 | 0.999532 | | 1.988911 | -0.47463 | 0.768106 | | 1.620521 | -0.17897 | 0.940744 |
| ZNHIT1 | 5.957822 | -0.02868 | 0.999532 | | 7.473711 | 0.26016 | 0.768183 | | 6.231069 | 0.307344 | 0.746581 |
| RABEP1 | 2.078015 | -0.04246 | 0.999532 | | 2.651738 | 0.402183 | 0.768183 | | 2.307906 | 0.45972 | 0.757395 |
| ZNF254 | 4.638587 | -0.05595 | 0.999532 | | 5.799787 | 0.29043 | 0.768183 | | 4.944996 | 0.36406 | 0.735469 |
| EIF2A | 2.369258 | 0.263025 | 0.999532 | | 2.963541 | 0.405471 | 0.768183 | | 2.716705 | 0.153656 | 0.929087 |
| PRR7 | 4.847498 | 0.309205 | 0.999532 | | 4.500566 | -0.35668 | 0.768183 | | 4.352931 | -0.65174 | 0.533876 |
| CGREF1 | 2.141035 | 0.623995 | 0.999532 | | 2.491066 | 0.457857 | 0.768183 | | 2.554545 | -0.14335 | 0.944262 |
| APOBEC | 3.919239 | 0.204193 | 0.999532 | | 3.686993 | -0.35331 | 0.768183 | | 3.56041 | -0.53867 | 0.644433 |
| ZHX3 | 2.99261 | -0.13445 | 0.999532 | | 3.714195 | 0.357908 | 0.768183 | | 3.153009 | 0.501757 | 0.704227 |
| CYLD | 5.605481 | -0.22323 | 0.999532 | | 6.113493 | -0.27433 | 0.768183 | | 4.92779 | -0.03421 | 0.985693 |
| CGGBP1 | 7.968463 | 0.249834 | 0.999532 | | 8.638122 | 0.247187 | 0.768183 | | 7.926583 | 0.016935 | 0.992286 |
| PPP1R14 | 2.792869 | 0.395097 | 0.999532 | | 2.39315 | -0.47335 | 0.768183 | | 2.621343 | -0.85351 | 0.557467 |
| SREBF1 | 3.104961 | 0.446174 | 0.999532 | | 3.396828 | 0.375663 | 0.768183 | | 3.323599 | -0.05344 | 0.979282 |
| YIPF5 | 4.129014 | 0.072583 | 0.999532 | | 5.175105 | 0.29993 | 0.768183 | | 4.352368 | 0.244164 | 0.836935 |
| GULP1 | 4.321136 | 0.043203 | 0.999532 | | 5.119116 | 0.295021 | 0.768183 | | 4.476547 | 0.266582 | 0.819366 |
| BMP10 | 2.224177 | 0.090976 | 0.999532 | | 2.193613 | -0.47493 | 0.768183 | | 1.922063 | -0.54245 | 0.740755 |
| KRTAP21 | 2.259859 | -0.52014 | 0.999532 | | 2.615145 | -0.43356 | 0.768183 | | 1.990279 | 0.102606 | 0.968885 |

| | | | | | | | | | | | |
|---|---|---|---|---|---|---|---|---|---|---|---|
| CAD | 3.730315 | 0.086152 | 0.999532 | | 4.728465 | 0.311485 | 0.768203 | | 4.058849 | 0.248895 | 0.843463 |
| ATP2A2 | 13.71714 | -0.06923 | 0.999532 | | 17.1669 | 0.248813 | 0.768527 | | 14.29237 | 0.335024 | 0.694961 |
| OCM | 2.455554 | 0.167924 | 0.999532 | | 2.285346 | -0.45084 | 0.768531 | | 2.086503 | -0.59931 | 0.684642 |
| MCOLN3 | 2.171895 | 0.175005 | 0.999532 | | 2.031273 | -0.47049 | 0.768601 | | 1.859828 | -0.62737 | 0.683846 |
| NUDT4 | 3.6692 | 0.011373 | 0.999532 | | 3.709647 | -0.36319 | 0.768601 | | 3.060235 | -0.35501 | 0.799944 |
| TMEM11 | 2.576253 | -0.63374 | 0.999532 | | 3.066911 | -0.39104 | 0.768602 | | 2.117014 | 0.26092 | 0.88588 |
| MOAP1 | 2.402789 | 0.212742 | 0.999532 | | 2.812908 | 0.395885 | 0.768608 | | 2.557402 | 0.20092 | 0.905952 |
| C6orf163 | 1.863725 | 0.97014 | 0.999532 | | 1.764877 | 0.491196 | 0.768608 | | 2.013902 | -0.46101 | 0.765852 |
| MED10 | 4.687171 | 0.339476 | 0.999532 | | 5.365042 | 0.301457 | 0.768735 | | 4.93842 | -0.02108 | 0.992286 |
| NFS1 | 1.952175 | 0.025 | 0.999532 | | 1.86024 | -0.48597 | 0.768812 | | 1.631495 | -0.49651 | 0.78815 |
| NSUN4 | 2.404878 | 0.088584 | 0.999532 | | 3.097289 | 0.381313 | 0.768822 | | 2.565936 | 0.321216 | 0.836453 |
| IMP4 | 4.229739 | 0.117152 | 0.999532 | | 3.954348 | -0.34853 | 0.768822 | | 3.746048 | -0.44725 | 0.703463 |
| KCNH2 | 3.875099 | -0.12591 | 0.999532 | | 3.92937 | -0.35234 | 0.768916 | | 3.244616 | -0.21495 | 0.893828 |
| ISCA1 | 2.604426 | 0.6355 | 0.999532 | | 2.867971 | 0.404301 | 0.768916 | | 2.887047 | -0.21666 | 0.88885 |
| FGGY | 2.546301 | -0.3009 | 0.999532 | | 2.828211 | -0.41315 | 0.768916 | | 2.072437 | -0.09476 | 0.968434 |
| SBK3 | 2.171034 | 0.00967 | 0.999532 | | 2.133974 | -0.47837 | 0.768916 | | 1.806911 | -0.46937 | 0.791576 |
| CALCR | 2.048266 | 0.447883 | 0.999532 | | 2.424946 | 0.449396 | 0.768916 | | 2.3948 | 0.02121 | 0.996109 |
| CHD2 | 8.562649 | -0.15244 | 0.999532 | | 9.532251 | -0.25533 | 0.768916 | | 7.650785 | -0.07411 | 0.95824 |
| HSPB9 | 2.421629 | -0.07167 | 0.999532 | | 2.516993 | -0.45218 | 0.768916 | | 2.133385 | -0.35545 | 0.851503 |
| PLXNA2 | 3.317953 | 0.080772 | 0.999532 | | 3.169694 | -0.38663 | 0.768916 | | 2.727386 | -0.4528 | 0.742183 |
| P2RY1 | 2.884646 | -0.36878 | 0.999532 | | 3.151276 | -0.39944 | 0.768916 | | 2.59063 | -0.01381 | 0.996891 |
| MPDU1 | 4.806709 | 0.205432 | 0.999532 | | 4.241826 | -0.34618 | 0.768916 | | 4.006998 | -0.53694 | 0.632113 |
| KCTD1 | 5.258147 | 0.378555 | 0.999532 | | 4.413035 | -0.32905 | 0.768916 | | 4.426808 | -0.6848 | 0.436389 |
| GNL2 | 2.641461 | 0.411321 | 0.999532 | | 2.944759 | 0.412582 | 0.768916 | | 2.757127 | 0.029011 | 0.991783 |
| DZIP3 | 1.915501 | 0.296706 | 0.999532 | | 2.25271 | 0.432474 | 0.768916 | | 2.205041 | 0.154223 | 0.94013 |
| TRIM56 | 9.93239 | -0.22203 | 0.999532 | | 10.63039 | -0.22727 | 0.768916 | | 8.589412 | -0.01024 | 0.996681 |
| CYP4F8 | 2.308582 | 0.350174 | 0.999532 | | 2.078899 | -0.46827 | 0.768916 | | 1.995528 | -0.79792 | 0.563729 |
| CDS1 | 2.218741 | 0.782647 | 0.999532 | | 2.102115 | 0.461199 | 0.768916 | | 2.267229 | -0.30354 | 0.860368 |
| GADD45G | 6.038259 | 0.332303 | 0.999532 | | 6.220795 | 0.278789 | 0.768916 | | 6.189577 | -0.04921 | 0.974808 |
| GRK1 | 2.330117 | 0.325741 | 0.999532 | | 2.097842 | -0.45681 | 0.768916 | | 1.994807 | -0.75635 | 0.59686 |
| FAM181B | 3.142589 | -0.51307 | 0.999532 | | 3.734816 | -0.36506 | 0.768916 | | 2.798886 | 0.161523 | 0.929529 |
| GSTM4 | 4.211661 | -0.08872 | 0.999532 | | 4.405747 | -0.33123 | 0.768916 | | 3.553057 | -0.22418 | 0.870663 |
| CYP4B1 | 2.059738 | -0.24826 | 0.999532 | | 2.209476 | -0.45364 | 0.768916 | | 1.725334 | -0.18598 | 0.934172 |
| GASK1B | 5.484206 | 0.489472 | 0.999532 | | 5.836458 | 0.327796 | 0.768916 | | 5.897238 | -0.14057 | 0.914929 |
| HPCAL1 | 3.232004 | 0.098665 | 0.999532 | | 4.064858 | 0.33367 | 0.768916 | | 3.60014 | 0.25318 | 0.849848 |
| IL17RE | 2.651341 | 0.309262 | 0.999532 | | 3.101539 | 0.386165 | 0.768916 | | 3.012537 | 0.094705 | 0.962716 |
| DEFB132 | 2.366261 | 0.311627 | 0.999532 | | 2.10532 | -0.46431 | 0.768916 | | 2.042371 | -0.7554 | 0.600291 |
| LRCH4 | 3.852196 | -0.17284 | 0.999532 | | 4.068097 | -0.33467 | 0.768916 | | 3.060605 | -0.14481 | 0.92697 |
| MUTYH | 1.905136 | 0.690636 | 0.999532 | | 2.056028 | 0.475915 | 0.768916 | | 2.22101 | -0.1917 | 0.92403 |
| RTP4 | 1.950611 | 0.405144 | 0.999532 | | 2.24979 | 0.449298 | 0.768916 | | 2.220144 | 0.062484 | 0.979282 |
| FOXO1 | 3.986891 | 0.102047 | 0.999532 | | 4.95729 | 0.313525 | 0.768916 | | 4.095413 | 0.228687 | 0.856551 |
| MAGT1 | 8.696204 | -0.36876 | 0.999532 | | 12.04898 | 0.229009 | 0.768916 | | 8.915265 | 0.628058 | 0.359441 |
| LYRM9 | 3.033784 | 0.525936 | 0.999532 | | 3.236487 | 0.38241 | 0.768916 | | 3.086753 | -0.13084 | 0.941197 |
| BFAR | 3.48147 | 0.584089 | 0.999532 | | 3.687693 | 0.354321 | 0.768916 | | 3.688874 | -0.20649 | 0.881837 |
| AP3S2 | 3.430708 | 0.219521 | 0.999532 | | 4.229751 | 0.342555 | 0.768916 | | 3.822591 | 0.146296 | 0.925023 |
| SH3GL2 | 2.037751 | -0.16535 | 0.999532 | | 1.995036 | -0.45387 | 0.768916 | | 1.695078 | -0.27025 | 0.892869 |
| ARFIP2 | 3.596096 | -0.00788 | 0.999532 | | 3.554736 | -0.35761 | 0.768916 | | 3.006267 | -0.33127 | 0.806394 |
| TCF12 | 8.617059 | 0.156811 | 0.999532 | | 10.1291 | 0.230757 | 0.768916 | | 8.945432 | 0.102657 | 0.936689 |
| TUFT1 | 2.037618 | -0.67448 | 0.999532 | | 2.402173 | -0.4552 | 0.768916 | | 1.653067 | 0.236281 | 0.914761 |
| ABHD12 | 2.934445 | 0.465517 | 0.999532 | | 3.278816 | 0.380288 | 0.768916 | | 3.147807 | -0.06516 | 0.974421 |
| CRYGS | 2.867601 | 0.062791 | 0.999532 | | 2.895771 | -0.40152 | 0.768916 | | 2.502472 | -0.4419 | 0.754671 |
| NHLH2 | 2.32729 | 0.546517 | 0.999532 | | 2.441198 | 0.435707 | 0.768916 | | 2.424551 | -0.0914 | 0.966652 |
| KREMEN | 1.865914 | 0.250526 | 0.999532 | | 2.319018 | 0.44102 | 0.768916 | | 2.160352 | 0.207663 | 0.911289 |
| SPCS3 | 7.527476 | -0.05546 | 0.999532 | | 9.849088 | 0.260013 | 0.768916 | | 8.044285 | 0.344865 | 0.692072 |
| RAB21 | 9.206533 | -0.03794 | 0.999532 | | 9.841686 | -0.23186 | 0.768916 | | 7.906953 | -0.16798 | 0.867101 |
| IFT20 | 3.711634 | 0.101557 | 0.999532 | | 4.431271 | 0.31983 | 0.768916 | | 4.020613 | 0.235594 | 0.864347 |
| FBXL17 | 4.315916 | 0.279374 | 0.999532 | | 4.430175 | 0.334923 | 0.768916 | | 4.410232 | 0.063291 | 0.972758 |
| FAM174B | 2.482149 | -0.24067 | 0.999532 | | 2.795264 | -0.42057 | 0.768916 | | 2.069361 | -0.15294 | 0.941197 |
| HSD11B1 | 2.521535 | -0.00169 | 0.999532 | | 2.314221 | -0.42801 | 0.768916 | | 2.217488 | -0.41807 | 0.796937 |
| ADAM9 | 7.753971 | -0.29622 | 0.999532 | | 10.61056 | 0.249386 | 0.768916 | | 8.204241 | 0.563376 | 0.396754 |
| ABHD18 | 3.324532 | 0.037258 | 0.999532 | | 3.333095 | -0.36918 | 0.768916 | | 2.88041 | -0.38999 | 0.785012 |
| DRG2 | 3.824834 | 0.316866 | 0.999532 | | 4.235291 | 0.375726 | 0.768916 | | 4.122922 | 0.080686 | 0.96382 |
| NDUFA4L | 2.871067 | -0.2406 | 0.999532 | | 3.171461 | -0.37969 | 0.768916 | | 2.522991 | -0.11834 | 0.954087 |
| ACP1 | 5.761121 | 0.216297 | 0.999532 | | 6.438624 | 0.307194 | 0.768916 | | 6.047153 | 0.097196 | 0.945129 |
| OBSL1 | 6.876373 | 0.015674 | 0.999532 | | 8.437459 | 0.259751 | 0.768916 | | 6.971232 | 0.264328 | 0.784603 |
| CD74 | 129.6502 | -0.387 | 0.999532 | | 181.8203 | 0.272129 | 0.768916 | | 138.572 | 0.6839 | 0.165207 |
| CYP2U1 | 2.950583 | 0.070728 | 0.999532 | | 2.878878 | -0.38453 | 0.768916 | | 2.622312 | -0.43692 | 0.751444 |
| BRD1 | 3.504363 | -0.03473 | 0.999532 | | 3.333359 | -0.37209 | 0.768916 | | 2.92602 | -0.32465 | 0.827267 |
| NDRG4 | 1.990391 | -0.19161 | 0.999532 | | 2.043566 | -0.47023 | 0.768916 | | 1.679559 | -0.25851 | 0.901033 |
| PRR14L | 3.128253 | 0.307452 | 0.999532 | | 3.424749 | 0.369141 | 0.768916 | | 3.322149 | 0.083549 | 0.965649 |
| KRT73 | 2.480931 | 0.379043 | 0.999532 | | 2.047684 | -0.48191 | 0.768916 | | 2.02235 | -0.8429 | 0.546636 |
| SEC22C | 8.809921 | 0.255827 | 0.999532 | | 9.868517 | 0.232483 | 0.768916 | | 9.049428 | -0.00323 | 0.99858 |
| FOXL3 | 3.249106 | 0.914007 | 0.999532 | | 2.222766 | -0.47082 | 0.768916 | | 2.910384 | -1.36942 | 0.15668 |
| TCAP | 2.250523 | 0.386202 | 0.999532 | | 1.913919 | -0.48991 | 0.768916 | | 1.903833 | -0.85607 | 0.543177 |
| PARP9 | 4.17014 | -0.02819 | 0.999532 | | 4.879547 | 0.312915 | 0.768916 | | 4.481367 | 0.378662 | 0.738861 |
| YY1 | 6.810835 | -0.05244 | 0.999532 | | 8.724831 | 0.27526 | 0.768916 | | 7.00194 | 0.355544 | 0.694281 |
| GPATCH | 3.093855 | 0.363589 | 0.999532 | | 2.741744 | -0.41967 | 0.768916 | | 2.529276 | -0.75188 | 0.558026 |
| APOBEC | 17.9575 | 0.06603 | 0.999532 | | 21.11735 | 0.216291 | 0.768916 | | 18.51001 | 0.177372 | 0.805466 |
| ALDH1L1 | 4.556289 | -0.44259 | 0.999532 | | 5.283691 | -0.33598 | 0.768916 | | 3.619316 | 0.139718 | 0.92697 |
| FAM200A | 3.139399 | 0.067131 | 0.999532 | | 2.903737 | -0.40234 | 0.768916 | | 2.684922 | -0.45867 | 0.740755 |
| MKRN2O | 2.192112 | -0.33593 | 0.999532 | | 2.322789 | -0.45036 | 0.768916 | | 1.927175 | -0.09737 | 0.972221 |
| CHD4 | 12.21639 | 0.064097 | 0.999532 | | 14.67488 | 0.229981 | 0.768916 | | 12.65454 | 0.190667 | 0.837256 |
| DHX33 | 2.250454 | 0.416249 | 0.999532 | | 2.521806 | 0.421992 | 0.768916 | | 2.55563 | 0.024919 | 0.994192 |
| DOCK11 | 2.401862 | 0.369111 | 0.999532 | | 2.69241 | 0.399869 | 0.768916 | | 2.565284 | 0.047776 | 0.985299 |
| HMGN3 | 9.029487 | 0.341109 | 0.999532 | | 9.75644 | 0.241812 | 0.768916 | | 9.110544 | -0.07723 | 0.945129 |
| OR2B11 | 3.052861 | -0.01665 | 0.999532 | | 3.035676 | -0.38352 | 0.768916 | | 2.57303 | -0.34892 | 0.81294 |
| ZMYM3 | 2.9516 | 0.298147 | 0.999532 | | 3.37757 | 0.368695 | 0.768916 | | 3.243463 | 0.087593 | 0.962014 |
| DOT1L | 3.615547 | -0.17251 | 0.999532 | | 3.802333 | -0.3528 | 0.768916 | | 3.014159 | -0.16175 | 0.915533 |
| SLC25A1 | 2.636344 | 0.063557 | 0.999532 | | 2.601897 | -0.42754 | 0.768916 | | 2.365483 | -0.46924 | 0.764366 |
| HOMER1 | 3.763597 | 0.117225 | 0.999532 | | 3.498745 | -0.38541 | 0.768916 | | 3.1922 | -0.48293 | 0.706749 |
| PAXX | 5.791957 | -0.17223 | 0.999532 | | 6.004857 | -0.27608 | 0.768916 | | 5.147893 | -0.08479 | 0.954304 |
| MUC22 | 2.696675 | 0.05754 | 0.999532 | | 2.740607 | -0.43018 | 0.768916 | | 2.333069 | -0.45132 | 0.788106 |
| AGPAT5 | 3.385817 | -0.3345 | 0.999532 | | 3.4896 | -0.36452 | 0.768916 | | 2.826075 | 0.001507 | 0.999463 |
| FANCG | 2.476032 | 0.14591 | 0.999532 | | 2.342801 | -0.43901 | 0.768916 | | 2.107527 | -0.56627 | 0.704455 |
| DNASE1L | 4.207828 | 0.249715 | 0.999532 | | 4.713715 | 0.308906 | 0.768916 | | 4.338249 | 0.073919 | 0.965649 |
| SGF29 | 3.585428 | 0.45631 | 0.999532 | | 3.681485 | 0.343772 | 0.768916 | | 3.75432 | -0.09469 | 0.954304 |
| PAK6 | 3.941741 | 0.533826 | 0.999532 | | 3.246709 | -0.37969 | 0.768916 | | 3.453361 | -0.8917 | 0.359441 |
| HTR3C | 2.468006 | 0.287978 | 0.999532 | | 2.142919 | -0.45694 | 0.768916 | | 2.065043 | -0.72845 | 0.623462 |
| RASGEF | 4.838988 | 0.162121 | 0.999532 | | 5.825849 | 0.316817 | 0.768916 | | 5.289271 | 0.186513 | 0.88082 |
| NAA25 | 4.251463 | 0.491083 | 0.999532 | | 4.498222 | 0.333275 | 0.768916 | | 4.650491 | -0.143 | 0.920805 |
| CLTB | 10.23566 | -0.05955 | 0.999532 | | 12.50615 | 0.208572 | 0.768916 | | 10.26141 | 0.284112 | 0.707745 |
| PDGFRB | 19.37851 | -0.30662 | 0.999532 | | 26.50087 | 0.21634 | 0.768926 | | 20.44291 | 0.541786 | 0.329469 |
| PSMA6 | 5.182141 | -0.2112 | 0.999532 | | 6.797981 | 0.268568 | 0.768926 | | 5.454398 | 0.497499 | 0.570037 |
| SCRT2 | 2.388556 | 0.258761 | 0.999532 | | 2.16712 | -0.46948 | 0.768926 | | 2.027649 | -0.71038 | 0.636525 |
| OR2A2 | 2.830527 | 0.281327 | 0.999532 | | 2.436825 | -0.43912 | 0.768926 | | 2.514015 | -0.70154 | 0.629744 |
| STK16 | 3.212385 | -0.15758 | 0.999532 | | 3.382102 | -0.367 | 0.768926 | | 2.861752 | -0.19008 | 0.909195 |
| GPR152 | 2.357303 | -0.15663 | 0.999532 | | 2.451768 | -0.45076 | 0.768926 | | 1.887913 | -0.27387 | 0.889853 |
| RAP1GAP | 5.159691 | 0.297647 | 0.999532 | | 5.714182 | 0.383963 | 0.768926 | | 5.30505 | 0.117342 | 0.947322 |
| SMARCA | 2.134533 | -0.33474 | 0.999532 | | 3.10228 | 0.38639 | 0.76896 | | 2.459584 | 0.741647 | 0.579378 |
| FARSA | 5.961651 | 0.146238 | 0.999532 | | 6.777439 | 0.265617 | 0.76896 | | 6.111632 | 0.126267 | 0.915925 |
| PCDH15 | 2.605696 | 0.4973 | 0.999532 | | 2.164034 | -0.46452 | 0.76896 | | 2.303709 | -0.94339 | 0.464413 |
| KRT76 | 2.442372 | -0.48111 | 0.999532 | | 2.76165 | -0.41875 | 0.76896 | | 2.1168 | 0.08125 | 0.97502 |
| TGFB3 | 4.484079 | 0.119823 | 0.999532 | | 4.455093 | -0.3828 | 0.76896 | | 3.891549 | -0.48304 | 0.674647 |
| VWA7 | 2.997975 | -0.34023 | 0.999532 | | 3.538429 | -0.36002 | 0.76896 | | 2.60677 | -0.00689 | 0.997967 |
| IDH1 | 2.303211 | 0.398711 | 0.999532 | | 2.54201 | 0.398188 | 0.76896 | | 2.443994 | 0.016888 | 0.996681 |
| AKR1B1 | 4.43109 | 0.174296 | 0.999532 | | 5.145018 | 0.299893 | 0.76896 | | 4.468006 | 0.141143 | 0.913083 |
| CRLF1 | 2.170005 | 0.424296 | 0.999532 | | 1.874991 | -0.47934 | 0.769013 | | 1.905262 | -0.88611 | 0.533876 |
| PELI1 | 4.175256 | 0.022559 | 0.999532 | | 4.178167 | -0.33167 | 0.769013 | | 3.396602 | -0.33712 | 0.785012 |
| ENPP2 | 4.834245 | -0.45332 | 0.999532 | | 5.831864 | -0.32009 | 0.769334 | | 4.344555 | 0.148804 | 0.920716 |
| TLE5 | 30.60211 | -0.05347 | 0.999532 | | 37.20753 | 0.154562 | 0.769399 | | 31.03068 | 0.228704 | 0.649779 |
| LRRC38 | 2.510743 | 0.39237 | 0.999532 | | 2.186488 | -0.4527 | 0.769399 | | 2.140867 | -0.82382 | 0.534789 |
| SALL4 | 2.118294 | 0.003822 | 0.999532 | | 2.031008 | -0.45967 | 0.769399 | | 1.819607 | -0.44351 | 0.804594 |
| PA2G4 | 7.488382 | 0.382007 | 0.999532 | | 7.017734 | -0.29281 | 0.769399 | | 6.722214 | -0.64344 | 0.426859 |
| BPNT1 | 2.196512 | 0.468977 | 0.999532 | | 2.265945 | 0.419173 | 0.769399 | | 2.294819 | -0.0338 | 0.991159 |
| CMTM2 | 3.491964 | -0.11823 | 0.999532 | | 3.428305 | -0.37737 | 0.769399 | | 3.029135 | -0.26514 | 0.881689 |
| CPLX1 | 4.814769 | -0.33478 | 0.999532 | | 5.708732 | -0.29529 | 0.769399 | | 4.278221 | 0.066324 | 0.972125 |
| NIBAN3 | 2.420145 | -0.36883 | 0.999532 | | 2.770042 | -0.43905 | 0.769399 | | 2.121384 | -0.05394 | 0.986693 |
| ATG10 | 2.208781 | -0.51091 | 0.999532 | | 2.414175 | -0.43184 | 0.769399 | | 1.863558 | 0.096 | 0.972221 |
| TEX52 | 2.31949 | -0.05145 | 0.999532 | | 2.325345 | -0.4483 | 0.769399 | | 2.06291 | -0.37892 | 0.832834 |
| COL16A1 | 3.818892 | -0.15617 | 0.999532 | | 4.027413 | -0.34392 | 0.769399 | | 3.230502 | -0.16774 | 0.906891 |
| GSR | 3.397705 | 0.362419 | 0.999532 | | 3.702859 | 0.348109 | 0.769399 | | 3.7338 | 0.005692 | 0.997967 |
| MFSD3 | 3.381248 | 0.13162 | 0.999532 | | 3.409332 | -0.36574 | 0.769399 | | 2.936339 | -0.46238 | 0.724363 |
| ZNF117 | 5.346454 | -0.153 | 0.999532 | | 5.859472 | -0.28455 | 0.769399 | | 4.84875 | -0.11375 | 0.934354 |
| DDX46 | 4.179619 | 0.13602 | 0.999532 | | 5.004532 | 0.298156 | 0.769399 | | 4.616159 | 0.191888 | 0.883588 |
| CCDC9 | 5.378093 | -0.21532 | 0.999532 | | 6.037157 | -0.2726 | 0.769399 | | 4.639683 | -0.03536 | 0.986811 |
| PLAC9 | 3.68628 | -0.63529 | 0.999532 | | 4.676769 | -0.3073 | 0.769399 | | 3.188984 | 0.350774 | 0.790999 |
| FEZ2 | 2.925606 | 0.102506 | 0.999532 | | 3.587988 | 0.343441 | 0.769399 | | 3.220344 | 0.258765 | 0.853053 |
| TEX15 | 2.839278 | 0.595165 | 0.999532 | | 2.393671 | -0.45444 | 0.769399 | | 2.629066 | -1.02924 | 0.404063 |
| SMIM15 | 3.620395 | 0.568024 | 0.999532 | | 3.446733 | 0.343236 | 0.769399 | | 3.821433 | -0.20996 | 0.877943 |
| DDX58 | 6.416921 | -0.01192 | 0.999532 | | 7.929157 | 0.264164 | 0.769399 | | 6.698412 | 0.294395 | 0.762924 |
| MREG | 2.328608 | -0.10466 | 0.999532 | | 2.327468 | -0.4288 | 0.769399 | | 1.943053 | -0.30297 | 0.873172 |
| MEI4 | 2.067057 | 0.109742 | 0.999532 | | 1.996407 | -0.47055 | 0.769399 | | 1.709365 | -0.5598 | 0.732486 |
| CACNA2 | 3.689467 | 0.329576 | 0.999532 | | 4.229418 | 0.323419 | 0.769399 | | 3.732622 | 0.009928 | 0.996891 |
| PARD3B | 6.028787 | 0.419042 | 0.999532 | | 6.60572 | 0.271686 | 0.769399 | | 6.162374 | -0.13504 | 0.914687 |
| SYT5 | 3.324711 | 0.203265 | 0.999532 | | 3.230218 | -0.39402 | 0.769399 | | 2.878957 | -0.58093 | 0.647581 |
| FGL2 | 11.0459 | -0.04315 | 0.999532 | | 11.42891 | -0.28187 | 0.769399 | | 9.616645 | -0.22039 | 0.809498 |
| HEY2 | 2.604186 | 0.021086 | 0.999532 | | 2.615762 | -0.44143 | 0.769399 | | 2.125114 | -0.43781 | 0.782753 |
| GRB14 | 3.772357 | -0.28137 | 0.999532 | | 4.34323 | -0.36085 | 0.769399 | | 3.147122 | -0.09366 | 0.961567 |
| CEBPA | 2.649018 | 0.126663 | 0.999532 | | 3.362847 | 0.358364 | 0.769399 | | 3.041843 | 0.252654 | 0.867009 |
| HMGB2 | 3.624707 | 0.273262 | 0.999532 | | 3.483742 | -0.37034 | 0.769399 | | 3.051285 | -0.63249 | 0.563455 |
| DNLZ | 2.511485 | 0.775512 | 0.999532 | | 2.212384 | 0.419701 | 0.769399 | | 2.742822 | -0.32906 | 0.829158 |
| SOCS5 | 5.83602 | -0.11218 | 0.999532 | | 6.137539 | -0.28573 | 0.769399 | | 4.911466 | -0.15258 | 0.901033 |
| OTOP2 | 2.415567 | 0.0931 | 0.999532 | | 2.393501 | -0.43937 | 0.769399 | | 2.08563 | -0.5119 | 0.740093 |
| SLC25A3 | 4.509162 | -0.48091 | 0.999532 | | 5.525483 | -0.30111 | 0.769399 | | 3.828395 | 0.211706 | 0.881011 |
| PAK3 | 1.819159 | 0.543712 | 0.999532 | | 1.942756 | 0.493093 | 0.769399 | | 1.949308 | -0.03169 | 0.992586 |
| LDLR | 2.274284 | -0.19264 | 0.999532 | | 2.37559 | -0.42397 | 0.769399 | | 1.879805 | -0.21342 | 0.911845 |
| RTL3 | 2.034412 | 0.133072 | 0.999532 | | 1.865354 | -0.46936 | 0.769399 | | 1.692296 | -0.58379 | 0.720106 |
| DEFB125 | 2.314522 | -0.85623 | 0.999532 | | 2.904695 | -0.43747 | 0.769399 | | 1.965546 | 0.436541 | 0.823571 |
| PODXL | 298.7426 | 0.417349 | 0.999532 | | 309.5152 | 0.202326 | 0.769399 | | 303.6948 | -0.18514 | 0.735469 |
| TOR1B | 3.940529 | -0.0508 | 0.999532 | | 3.992586 | -0.35924 | 0.769399 | | 3.53651 | -0.28184 | 0.851785 |
| SLC35A1 | 2.853395 | -0.28358 | 0.999532 | | 3.70383 | 0.333422 | 0.769399 | | 3.09703 | 0.641138 | 0.563455 |
| SLIRP | 4.002546 | -0.00447 | 0.999532 | | 4.817194 | 0.299287 | 0.769399 | | 4.188231 | 0.319338 | 0.78258 |
| CHST15 | 2.719232 | 0.199613 | 0.999532 | | 3.196448 | 0.379113 | 0.769399 | | 2.928997 | 0.200774 | 0.899852 |
| FANCC | 4.529901 | -0.29641 | 0.999532 | | 5.178999 | -0.30337 | 0.769399 | | 3.977301 | 0.014294 | 0.996681 |
| ANO6 | 11.94121 | -0.16292 | 0.999532 | | 15.13089 | 0.20231 | 0.769399 | | 12.35954 | 0.383879 | 0.530351 |
| FGF18 | 3.65327 | -0.22753 | 0.999532 | | 4.13804 | -0.35902 | 0.769399 | | 3.190747 | -0.10506 | 0.956322 |
| TMEM254 | 2.828151 | 0.244761 | 0.999532 | | 3.58533 | 0.356095 | 0.769399 | | 3.213238 | 0.139911 | 0.933313 |
| PLA2G4C | 2.127189 | -0.13574 | 0.999532 | | 2.975893 | 0.40484 | 0.769399 | | 2.61135 | 0.561795 | 0.702463 |
| MAL | 6.728798 | 0.704411 | 0.999532 | | 6.623743 | 0.33869 | 0.769399 | | 6.837564 | -0.35198 | 0.779082 |
| TTC24 | 2.009654 | 0.245733 | 0.999532 | | 1.814028 | -0.47878 | 0.769576 | | 1.817934 | -0.70398 | 0.657538 |
| NAGK | 6.576455 | 0.070128 | 0.999532 | | 7.828234 | 0.246104 | 0.769691 | | 6.866465 | 0.195853 | 0.849682 |
| PYM1 | 2.120627 | 0.072041 | 0.999532 | | 2.73922 | 0.416282 | 0.769691 | | 2.335266 | 0.373133 | 0.823193 |
| ABHD4 | 2.892935 | 0.274224 | 0.999532 | | 3.373386 | 0.358735 | 0.769691 | | 3.06536 | 0.104143 | 0.953926 |
| PRDM10 | 2.843149 | 0.190156 | 0.999532 | | 2.398233 | -0.40899 | 0.770007 | | 2.455645 | -0.5919 | 0.673405 |
| ARHGEF | 4.539211 | 0.189293 | 0.999532 | | 5.221558 | 0.296179 | 0.770254 | | 4.833113 | 0.12219 | 0.934597 |
| CCDC88 | 7.446132 | -0.01556 | 0.999532 | | 7.855327 | -0.30469 | 0.770411 | | 6.407647 | -0.26975 | 0.800239 |
| ZNF775 | 4.07262 | 0.019365 | 0.999532 | | 4.058351 | -0.33683 | 0.770411 | | 3.420833 | -0.33893 | 0.800239 |
| MTHFD2 | 3.908483 | -0.51854 | 0.999532 | | 4.671054 | -0.34027 | 0.770411 | | 3.258859 | 0.207229 | 0.896374 |
| USP37 | 2.119162 | 0.502527 | 0.999532 | | 2.541693 | 0.410885 | 0.770411 | | 2.336643 | -0.07227 | 0.974421 |
| NME5 | 3.027593 | -0.17461 | 0.999532 | | 2.990946 | -0.39379 | 0.770411 | | 2.464457 | -0.19744 | 0.911286 |
| CCDC169 | 2.066323 | 0.105251 | 0.999532 | | 1.894865 | -0.4769 | 0.770411 | | 1.740392 | -0.56651 | 0.738861 |
| ZFAND2B | 5.521903 | 0.268743 | 0.999532 | | 6.18515 | 0.286779 | 0.770411 | | 5.837511 | 0.034449 | 0.985299 |
| RNF170 | 3.567577 | 0.273338 | 0.999532 | | 3.824684 | 0.333843 | 0.770411 | | 3.637336 | 0.073507 | 0.968235 |
| IKBIP | 3.480732 | 0.264567 | 0.999532 | | 4.295721 | 0.360416 | 0.770411 | | 4.018184 | 0.108326 | 0.94861 |
| KLC3 | 1.968317 | 0.163305 | 0.999532 | | 1.778864 | -0.47989 | 0.770411 | | 1.670429 | -0.62786 | 0.705425 |
| GLYATL3 | 2.077367 | -0.0065 | 0.999532 | | 2.082396 | -0.44562 | 0.770411 | | 1.781524 | -0.41724 | 0.810635 |
| CA9 | 2.642168 | 0.227398 | 0.999532 | | 2.404888 | -0.43475 | 0.770411 | | 2.162473 | -0.64577 | 0.657538 |
| C11orf52 | 2.401443 | 0.052386 | 0.999532 | | 2.404917 | -0.42546 | 0.770652 | | 2.112223 | -0.45833 | 0.766974 |
| FAM210B | 10.61404 | -0.11433 | 0.999532 | | 11.58988 | -0.22055 | 0.770652 | | 9.668044 | -0.0776 | 0.952656 |
| KBTBD3 | 3.023326 | -0.13367 | 0.999532 | | 3.292184 | -0.40798 | 0.770652 | | 2.623485 | -0.28523 | 0.856551 |
| ELFN1 | 1.976268 | -0.49538 | 0.999532 | | 2.09513 | -0.45369 | 0.770652 | | 1.558716 | 0.054412 | 0.987608 |
| PLCL2 | 3.571321 | -0.35634 | 0.999532 | | 3.949838 | -0.35532 | 0.770652 | | 2.83488 | 0.025733 | 0.992459 |
| BCL2A1 | 2.007313 | 0.693763 | 0.999532 | | 2.137522 | 0.46516 | 0.770827 | | 2.258465 | -0.21005 | 0.911845 |
| ID4 | 6.311094 | -0.8466 | 0.950261 | | 8.24464 | -0.26594 | 0.770827 | | 5.249411 | 0.600327 | 0.49299 |
| ZCCHC7 | 2.376857 | -0.13767 | 0.999532 | | 2.356154 | -0.43879 | 0.770827 | | 1.982892 | -0.28583 | 0.883116 |
| PPP1R12 | 10.76581 | -0.07192 | 0.999532 | | 11.09543 | -0.21717 | 0.770827 | | 9.331376 | -0.1324 | 0.892869 |
| ITFG1 | 5.20934 | -0.14388 | 0.999532 | | 6.59935 | 0.298145 | 0.771016 | | 5.504157 | 0.448759 | 0.66549 |
| TRAP1 | 2.929277 | 0.160105 | 0.999532 | | 3.668905 | 0.351344 | 0.771049 | | 3.098111 | 0.218016 | 0.88476 |
| EVI2A | 2.304083 | -0.30343 | 0.999532 | | 2.37729 | -0.42595 | 0.771291 | | 1.84477 | -0.11008 | 0.965619 |
| ZIC3 | 14.101 | -0.12509 | 0.999532 | | 15.14894 | -0.2673 | 0.771291 | | 12.64592 | -0.14279 | 0.905952 |
| SERINC2 | 2.676303 | 0.362626 | 0.999532 | | 2.983535 | 0.380824 | 0.771551 | | 2.752546 | 0.040851 | 0.988359 |
| PDLIM7 | 7.878453 | 0.164583 | 0.999532 | | 9.110488 | 0.239634 | 0.771551 | | 8.223339 | 0.096053 | 0.929529 |
| DNAJC28 | 2.117439 | 0.341568 | 0.999532 | | 2.538578 | 0.414739 | 0.771551 | | 2.328522 | 0.094833 | 0.965649 |
| PCED1B | 3.130385 | 0.868088 | 0.999532 | | 2.829321 | 0.39072 | 0.771911 | | 3.400399 | -0.45989 | 0.717138 |
| MAGI1 | 3.938701 | 0.281631 | 0.999532 | | 4.424374 | 0.308802 | 0.771915 | | 4.254824 | 0.045378 | 0.979717 |
| LCOR | 3.601347 | -0.22673 | 0.999532 | | 4.042488 | -0.32346 | 0.771915 | | 3.189215 | -0.06302 | 0.965649 |
| ABCC1 | 3.362734 | -0.12227 | 0.999532 | | 4.357447 | 0.34817 | 0.771915 | | 3.646951 | 0.495722 | 0.680577 |
| FAM162B | 2.223676 | -0.61453 | 0.999532 | | 2.772129 | -0.42419 | 0.771915 | | 2.033972 | 0.195462 | 0.92697 |
| CPLX4 | 2.155241 | -0.10126 | 0.999532 | | 2.192993 | -0.44777 | 0.771915 | | 1.848611 | -0.32799 | 0.864923 |
| UQCRC2 | 11.7521 | 0.150929 | 0.999532 | | 13.16291 | 0.224419 | 0.771961 | | 11.80661 | 0.09356 | 0.92697 |
| WDR19 | 2.078607 | 0.292062 | 0.999532 | | 2.424505 | 0.431085 | 0.771961 | | 2.21849 | 0.158826 | 0.938429 |
| NSA2 | 7.532142 | 0.380852 | 0.999532 | | 8.051862 | 0.254703 | 0.771961 | | 7.71192 | -0.1067 | 0.922679 |
| SRPRA | 5.519735 | 0.18101 | 0.999532 | | 6.774723 | 0.319512 | 0.771961 | | 5.801058 | 0.172673 | 0.903339 |
| USP47 | 4.003519 | -0.23371 | 0.999532 | | 4.285817 | -0.32159 | 0.772144 | | 3.444656 | -0.07217 | 0.971732 |
| PUS7 | 2.977932 | -0.05483 | 0.999532 | | 2.943392 | -0.38714 | 0.772412 | | 2.514472 | -0.31286 | 0.843043 |
| C2orf66 | 2.058658 | 0.723203 | 0.999532 | | 2.283251 | 0.455681 | 0.772483 | | 2.399038 | -0.23179 | 0.903329 |
| TOMM40L | 2.368223 | 0.136098 | 0.999532 | | 2.191509 | -0.44442 | 0.772483 | | 2.077084 | -0.56323 | 0.720106 |
| KRT76 | 2.131668 | -0.27412 | 0.999532 | | 2.247832 | -0.43275 | 0.772483 | | 1.870009 | -0.13611 | 0.952783 |
| DHFR | 5.109928 | 0.284764 | 0.999532 | | 4.81411 | -0.30106 | 0.772569 | | 4.447509 | -0.56431 | 0.536289 |
| CAST | 9.470089 | 0.570374 | 0.999532 | | 9.302452 | 0.241743 | 0.773105 | | 9.644226 | -0.30735 | 0.699971 |
| PYROXD | 2.74994 | -0.20486 | 0.999532 | | 3.682382 | 0.397614 | 0.773153 | | 3.127132 | 0.607383 | 0.664512 |
| SMC1A | 4.010355 | -0.01611 | 0.999532 | | 5.259165 | 0.323876 | 0.773687 | | 4.482818 | 0.360742 | 0.76158 |
| RSRC1 | 2.420533 | 0.597212 | 0.999532 | | 2.475933 | 0.408141 | 0.773687 | | 2.626415 | -0.17294 | 0.920259 |
| NCBP2 | 4.625285 | 0.371218 | 0.999532 | | 4.840756 | 0.298501 | 0.773687 | | 4.89838 | -0.05093 | 0.977409 |
| TMEM16 | 8.089432 | -0.17475 | 0.999532 | | 10.30507 | 0.235865 | 0.773687 | | 8.272398 | 0.434457 | 0.563627 |
| OR2G3 | 1.833681 | 1.170945 | 0.999532 | | 1.675382 | 0.5131 | 0.773687 | | 2.043544 | -0.63799 | 0.666906 |
| OR4Q3 | 4.706046 | 0.315938 | 0.999532 | | 4.382695 | -0.3493 | 0.773687 | | 4.145751 | -0.65062 | 0.515762 |
| LEPR | 3.241229 | 0.3056 | 0.999532 | | 3.645004 | 0.353004 | 0.773687 | | 3.348943 | 0.066775 | 0.972221 |
| ATP13A3 | 4.930878 | -0.47007 | 0.999532 | | 7.037088 | 0.322458 | 0.773687 | | 5.464954 | 0.810799 | 0.359441 |
| GBE1 | 2.924667 | 0.267369 | 0.999532 | | 3.491189 | 0.33927 | 0.774026 | | 3.178215 | 0.090723 | 0.95999 |
| PHKB | 3.271463 | 0.252839 | 0.999532 | | 3.726147 | 0.352531 | 0.774026 | | 3.342444 | 0.121768 | 0.942744 |
| FERMT1 | 1.922288 | -0.1724 | 0.999532 | | 1.91704 | -0.46775 | 0.774026 | | 1.576581 | -0.27721 | 0.89433 |
| H3-4 | 2.285338 | -0.2836 | 0.999532 | | 2.320073 | -0.4143 | 0.774026 | | 1.936825 | -0.11771 | 0.960773 |
| CHRD | 2.420731 | 0.223414 | 0.999532 | | 2.375497 | -0.44625 | 0.774158 | | 2.054708 | -0.64447 | 0.660958 |
| KLHL26 | 2.670617 | 0.127635 | 0.999532 | | 2.583925 | -0.40124 | 0.774308 | | 2.287961 | -0.51055 | 0.713173 |
| OR52E6 | 2.322358 | 0.278854 | 0.999532 | | 2.001845 | -0.4606 | 0.774308 | | 1.98729 | -0.72486 | 0.637895 |
| RPS9 | 29.49273 | -0.07291 | 0.999532 | | 36.38658 | 0.212093 | 0.774308 | | 30.14409 | 0.303233 | 0.706407 |
| DNAJC19 | 3.059627 | 0.199415 | 0.999532 | | 2.885829 | -0.39046 | 0.774308 | | 2.658562 | -0.5696 | 0.662576 |
| PLN | 3.221597 | 0.199612 | 0.999532 | | 3.04654 | -0.43576 | 0.774308 | | 2.746037 | -0.6107 | 0.670068 |
| KHK | 2.949713 | 0.116116 | 0.999532 | | 3.698376 | 0.369351 | 0.774308 | | 3.193625 | 0.269848 | 0.859522 |
| KRTAP9- | 5.900225 | -0.02161 | 0.999532 | | 6.299116 | -0.30132 | 0.774308 | | 5.295774 | -0.25269 | 0.837256 |
| HOXD10 | 3.963886 | -0.12663 | 0.999532 | | 5.59184 | 0.310278 | 0.774308 | | 4.4094 | 0.454421 | 0.680564 |
| TENT4B | 3.08995 | -0.55031 | 0.999532 | | 3.639576 | -0.34255 | 0.774308 | | 2.495648 | 0.224934 | 0.891408 |
| SIRT5 | 4.276944 | 0.015835 | 0.999532 | | 5.110968 | 0.291529 | 0.774308 | | 4.401115 | 0.291756 | 0.799835 |
| MYBPHL | 3.857046 | 0.04743 | 0.999532 | | 3.738961 | -0.35398 | 0.774308 | | 3.397251 | -0.38557 | 0.771685 |
| SUN2 | 9.04698 | 0.137923 | 0.999532 | | 10.08115 | 0.230001 | 0.774308 | | 8.99509 | 0.107839 | 0.918082 |
| COX7A1 | 8.863437 | -0.21357 | 0.999532 | | 11.40761 | 0.236934 | 0.774308 | | 9.109185 | 0.463667 | 0.566812 |
| EMP3 | 15.29852 | -0.22807 | 0.999532 | | 20.09633 | 0.251539 | 0.774419 | | 16.16729 | 0.50423 | 0.511836 |
| CST1 | 2.174328 | 0.144621 | 0.999532 | | 2.059891 | -0.46352 | 0.774419 | | 1.823256 | -0.58945 | 0.717627 |
| UROS | 4.399641 | 0.167937 | 0.999532 | | 5.193867 | 0.292453 | 0.774946 | | 4.457677 | 0.148112 | 0.906439 |
| ANKMY1 | 3.763389 | -0.003 | 0.999532 | | 3.687117 | -0.34251 | 0.774946 | | 3.246032 | -0.32984 | 0.807738 |
| INTS6L | 3.173046 | -0.58894 | 0.999532 | | 4.994763 | 0.317309 | 0.775006 | | 3.463067 | 0.920805 | 0.304062 |
| VMP1 | 3.501352 | 0.308008 | 0.999532 | | 3.973934 | 0.337288 | 0.775006 | | 3.79024 | 0.046254 | 0.981579 |
| FOCAD | 2.406712 | -0.51555 | 0.999532 | | 2.801919 | -0.42355 | 0.775093 | | 2.14903 | 0.112492 | 0.967 |
| SGTB | 3.141798 | -0.46918 | 0.999532 | | 3.614678 | -0.35299 | 0.775161 | | 2.680095 | 0.138089 | 0.942744 |
| B4GALT5 | 5.840882 | 0.231345 | 0.999532 | | 6.816001 | 0.280961 | 0.775161 | | 6.184337 | 0.068162 | 0.963359 |
| PIPOX | 3.815186 | 0.437545 | 0.999532 | | 3.943798 | 0.366929 | 0.775583 | | 4.104824 | -0.0787 | 0.96734 |
| SMARCD | 3.044089 | 0.237649 | 0.999532 | | 2.779454 | -0.38261 | 0.775583 | | 2.652144 | -0.60084 | 0.641721 |
| ARL15 | 3.662296 | -0.28205 | 0.999532 | | 5.147712 | 0.307294 | 0.776168 | | 4.016355 | 0.612106 | 0.557909 |
| FAM221B | 2.208619 | 0.29879 | 0.999532 | | 2.058561 | -0.45699 | 0.776311 | | 1.884495 | -0.72852 | 0.630705 |
| DESI1 | 3.291933 | -0.4644 | 0.999532 | | 3.842422 | -0.33321 | 0.776475 | | 2.926965 | 0.147292 | 0.934701 |
| RFXANK | 4.356243 | -0.25306 | 0.999532 | | 4.702245 | -0.29806 | 0.776475 | | 3.698868 | -0.0262 | 0.991341 |
| TRIP4 | 3.981486 | 0.485796 | 0.999532 | | 4.120582 | 0.330894 | 0.776475 | | 4.152193 | -0.13439 | 0.924186 |
| OTUD6B | 4.363445 | -0.25996 | 0.999532 | | 4.816401 | -0.30288 | 0.776475 | | 3.839978 | -0.01982 | 0.994347 |
| ICA1 | 2.864697 | -0.32836 | 0.999532 | | 3.313512 | -0.36591 | 0.776475 | | 2.433828 | -0.01853 | 0.996681 |
| LDLRAD4 | 5.547746 | -0.06224 | 0.999532 | | 5.470244 | -0.29612 | 0.776475 | | 4.765914 | -0.22399 | 0.862823 |
| THAP3 | 4.410453 | -0.48796 | 0.999532 | | 5.37342 | -0.28641 | 0.776695 | | 3.67546 | 0.22777 | 0.865363 |
| MPO | 4.215966 | 0.163115 | 0.999532 | | 3.952469 | -0.36339 | 0.776825 | | 3.522358 | -0.50451 | 0.670068 |
| NUS1 | 6.915644 | -0.13143 | 0.999532 | | 7.712206 | -0.24581 | 0.776871 | | 6.147606 | -0.09251 | 0.941197 |
| DIS3L | 2.739196 | 0.908616 | 0.999532 | | 2.454377 | 0.419462 | 0.776898 | | 2.961285 | -0.47401 | 0.720106 |
| FOXI2 | 2.437395 | -0.11331 | 0.999532 | | 2.433019 | -0.41962 | 0.776918 | | 1.997093 | -0.29082 | 0.876781 |
| TPMT | 3.444056 | 0.606067 | 0.999532 | | 3.327177 | 0.352219 | 0.776918 | | 3.521766 | -0.24056 | 0.864245 |
| CYP2R1 | 2.012128 | -0.2731 | 0.999532 | | 2.135476 | -0.43718 | 0.777137 | | 1.757275 | -0.14862 | 0.947322 |
| ZNF440 | 3.787814 | -0.25908 | 0.999532 | | 4.32848 | -0.35693 | 0.777137 | | 3.251637 | -0.073 | 0.972305 |
| ESRP1 | 2.994096 | -0.07064 | 0.999532 | | 3.133854 | -0.37032 | 0.777137 | | 2.581675 | -0.27895 | 0.867739 |
| RAB40C | 2.926195 | 0.111772 | 0.999532 | | 2.721813 | -0.38017 | 0.777246 | | 2.47656 | -0.47535 | 0.720526 |
| NOG | 1.99235 | 0.087827 | 0.999532 | | 1.883818 | -0.48188 | 0.777246 | | 1.716999 | -0.55253 | 0.752523 |
| SERPINB | 3.088162 | -0.11869 | 0.999532 | | 3.74801 | 0.350757 | 0.777246 | | 3.202237 | 0.475117 | 0.717627 |
| MOB4 | 3.509407 | 0.075904 | 0.999532 | | 4.481517 | 0.313538 | 0.777372 | | 3.774291 | 0.256567 | 0.842754 |
| ANKRD3 | 6.664308 | -0.01987 | 0.999532 | | 6.895333 | -0.25488 | 0.777372 | | 5.859744 | -0.2188 | 0.849682 |
| IGFN1 | 2.249486 | 0.103495 | 0.999532 | | 2.225899 | -0.45068 | 0.777372 | | 1.889343 | -0.53569 | 0.735469 |
| MGME1 | 2.307579 | 0.117403 | 0.999532 | | 2.750306 | 0.376364 | 0.777372 | | 2.390637 | 0.271887 | 0.868512 |
| PET100 | 7.423055 | -0.14468 | 0.999532 | | 9.276878 | 0.227462 | 0.777656 | | 7.482565 | 0.390363 | 0.625947 |
| LRAT | 3.200894 | -0.31054 | 0.999532 | | 3.736917 | -0.35105 | 0.778366 | | 2.785864 | -0.01646 | 0.996681 |
| MRPL58 | 3.897637 | 0.336927 | 0.999532 | | 4.198644 | 0.324065 | 0.778366 | | 4.148521 | 0.002463 | 0.999385 |
| MUC16 | 2.199185 | -0.09274 | 0.999532 | | 2.348665 | -0.43312 | 0.778493 | | 1.785086 | -0.3145 | 0.867246 |
| AKT3 | 9.713921 | 0.190462 | 0.999532 | | 10.41775 | 0.213224 | 0.778559 | | 9.889144 | 0.038324 | 0.976783 |
| MAST4 | 3.971496 | 0.425767 | 0.999532 | | 4.375038 | 0.324052 | 0.778559 | | 4.134229 | -0.0848 | 0.962597 |
| DNAJA3 | 1.911995 | 0.083183 | 0.999532 | | 2.396532 | 0.415416 | 0.778559 | | 2.173552 | 0.354562 | 0.836453 |
| SLC4A3 | 2.612981 | -0.39974 | 0.999532 | | 2.983599 | -0.3842 | 0.778559 | | 2.154711 | 0.034682 | 0.991273 |
| RPL7 | 48.7625 | -0.03999 | 0.999532 | | 57.72069 | 0.12106 | 0.778559 | | 48.55925 | 0.181512 | 0.660958 |
| TMEM82 | 2.849079 | 0.358857 | 0.999532 | | 3.33486 | 0.372257 | 0.778635 | | 3.154387 | 0.035893 | 0.989437 |
| ALOX15 | 2.867026 | 0.038864 | 0.999532 | | 2.966158 | -0.41604 | 0.778635 | | 2.367423 | -0.43521 | 0.769793 |
| MUC5B | 5.3877 | 0.272092 | 0.999532 | | 4.797317 | -0.33694 | 0.778635 | | 4.743029 | -0.59683 | 0.60629 |
| RAB5A | 9.433475 | 0.157418 | 0.999532 | | 11.20812 | 0.210306 | 0.779049 | | 9.65937 | 0.080851 | 0.941197 |
| ZNF287 | 3.0332 | 0.060961 | 0.999532 | | 3.113862 | -0.37341 | 0.779049 | | 2.771804 | -0.41368 | 0.767184 |
| KRT86 | 2.594345 | 0.231776 | 0.999532 | | 2.231985 | -0.42625 | 0.779049 | | 2.189112 | -0.64815 | 0.663709 |
| CSTB | 8.769477 | 0.283193 | 0.999532 | | 9.899634 | 0.268088 | 0.779051 | | 9.110013 | 0.03555 | 0.982781 |
| KIAA1841 | 2.175689 | 0.41203 | 0.999532 | | 2.350607 | 0.419048 | 0.779348 | | 2.388821 | 0.026698 | 0.993618 |
| SMU1 | 5.506756 | 0.075295 | 0.999532 | | 6.691501 | 0.265694 | 0.779469 | | 5.770221 | 0.209397 | 0.852179 |
| EIF3C | 19.31614 | 0.254699 | 0.999532 | | 21.5275 | 0.211972 | 0.779469 | | 19.74086 | -0.01349 | 0.994192 |
| HS3ST3A | 5.818626 | -0.66048 | 0.999532 | | 7.274776 | -0.24694 | 0.779469 | | 5.027559 | 0.432158 | 0.668211 |
| RETSAT | 4.676038 | 0.125701 | 0.999532 | | 5.762656 | 0.279802 | 0.779469 | | 5.120466 | 0.180463 | 0.88885 |
| GPAT2 | 7.496916 | -0.11713 | 0.999532 | | 8.372985 | -0.28062 | 0.779585 | | 6.633757 | -0.13508 | 0.916888 |
| TCF20 | 4.629874 | -0.47618 | 0.999532 | | 5.24992 | -0.31263 | 0.779883 | | 3.732834 | 0.184476 | 0.900848 |
| GTF3A | 4.127323 | 0.095345 | 0.999532 | | 5.214998 | 0.298559 | 0.779883 | | 4.519813 | 0.220971 | 0.861656 |
| TCEANC | 2.427268 | -0.02757 | 0.999532 | | 2.418584 | -0.409 | 0.779883 | | 2.080932 | -0.36327 | 0.827267 |
| HOXB4 | 3.514047 | 0.011379 | 0.999532 | | 3.656145 | -0.35062 | 0.779883 | | 3.019148 | -0.34152 | 0.800724 |
| PSMC4 | 4.443539 | 0.123705 | 0.999532 | | 5.301889 | 0.29666 | 0.779925 | | 4.579169 | 0.195513 | 0.881771 |
| APOA4 | 2.31289 | 0.721251 | 0.999532 | | 2.487903 | 0.47313 | 0.779925 | | 2.52078 | -0.21014 | 0.911581 |
| FBXO46 | 2.642705 | 0.091901 | 0.999532 | | 2.661964 | -0.40501 | 0.779925 | | 2.314315 | -0.47496 | 0.749222 |
| CSPG4 | 6.017413 | -1.30978 | 0.042135 | DOWN | 8.788745 | -0.25114 | 0.779925 | | 5.12075 | 1.082554 | 0.139095 |
| FDX1 | 5.511815 | 0.077628 | 0.999532 | | 7.021019 | 0.266844 | 0.779925 | | 5.737851 | 0.185064 | 0.872493 |
| RAPSN | 4.350581 | 0.165977 | 0.999532 | | 4.125571 | -0.33329 | 0.779925 | | 3.972217 | -0.47324 | 0.709825 |
| OTX1 | 2.904439 | 0.101353 | 0.999532 | | 2.809653 | -0.41911 | 0.779925 | | 2.624891 | -0.50465 | 0.742839 |
| PCDH17 | 4.079365 | 0.20274 | 0.999532 | | 4.474636 | 0.316954 | 0.779925 | | 4.251027 | 0.12352 | 0.940162 |
| ZDHHC7 | 2.249884 | 0.540335 | 0.999532 | | 1.796388 | -0.4592 | 0.779966 | | 1.926903 | -0.98168 | 0.442315 |
| NFATC4 | 3.447277 | -0.13491 | 0.999532 | | 3.64981 | -0.35399 | 0.78015 | | 2.776405 | -0.19469 | 0.902504 |
| VSIG2 | 4.071268 | 0.050065 | 0.999532 | | 3.868506 | -0.32977 | 0.780162 | | 3.40545 | -0.3611 | 0.771685 |
| TBC1D19 | 2.958573 | 0.149038 | 0.999532 | | 2.869679 | -0.38619 | 0.780162 | | 2.486301 | -0.52207 | 0.694961 |
| FOXK2 | 4.671546 | -0.08287 | 0.999532 | | 4.820705 | -0.31559 | 0.780162 | | 3.95012 | -0.21004 | 0.88082 |
| REV1 | 3.683026 | -0.02855 | 0.999532 | | 4.596172 | 0.325335 | 0.780162 | | 3.933332 | 0.372558 | 0.75311 |
| CCDC146 | 2.622479 | -0.23145 | 0.999532 | | 2.654229 | -0.40845 | 0.780162 | | 2.216924 | -0.15273 | 0.941197 |
| ANKRD45 | 2.813317 | -0.13999 | 0.999532 | | 2.892817 | -0.38562 | 0.780162 | | 2.417555 | -0.23077 | 0.89901 |
| CHST14 | 3.671828 | -0.06659 | 0.999532 | | 3.760134 | -0.36026 | 0.780162 | | 3.21352 | -0.27906 | 0.859426 |
| C12orf42 | 1.749103 | -0.27649 | 0.999532 | | 1.937423 | -0.46541 | 0.780162 | | 1.517614 | -0.16837 | 0.945902 |
| TMEM115 | 3.418063 | -0.32748 | 0.999532 | | 3.764994 | -0.33779 | 0.780162 | | 2.703023 | -0.00066 | 0.999746 |
| ARHGAP | 2.616857 | 0.099443 | 0.999532 | | 3.208033 | 0.367133 | 0.780162 | | 2.787357 | 0.285706 | 0.850013 |
| SMAP2 | 3.456336 | -0.45466 | 0.999532 | | 3.8934 | -0.32706 | 0.780162 | | 2.964541 | 0.140528 | 0.939107 |
| CNTFR | 2.182586 | -0.05969 | 0.999532 | | 2.284706 | -0.42408 | 0.780162 | | 1.832106 | -0.34188 | 0.851785 |
| MPDZ | 2.854872 | 0.143492 | 0.999532 | | 3.510953 | 0.338935 | 0.780162 | | 3.201074 | 0.224485 | 0.883785 |
| PHTF1 | 2.309749 | 0.299137 | 0.999532 | | 2.666577 | 0.402138 | 0.780162 | | 2.513826 | 0.123052 | 0.950616 |
| SNX19 | 7.133448 | 0.078075 | 0.999532 | | 8.393764 | 0.237533 | 0.780162 | | 7.195986 | 0.179376 | 0.857043 |
| ATP6V1F | 5.761361 | 0.228301 | 0.999532 | | 6.557628 | 0.286609 | 0.780162 | | 5.955664 | 0.082812 | 0.951656 |
| PWP1 | 2.973253 | 0.23774 | 0.999532 | | 3.458827 | 0.399017 | 0.780162 | | 3.224065 | 0.169341 | 0.923223 |
| SEC22A | 3.020221 | 0.131874 | 0.999532 | | 3.563153 | 0.357802 | 0.780162 | | 3.145796 | 0.244952 | 0.872062 |
| TPPP3 | 17.2311 | 0.269745 | 0.999532 | | 19.47459 | 0.257459 | 0.78023 | | 17.87194 | 0.006254 | 0.996891 |
| LZTS2 | 5.976624 | 0.320275 | 0.999532 | | 6.567792 | 0.276859 | 0.780325 | | 6.096614 | -0.01831 | 0.992586 |
| EDDM13 | 2.637822 | 0.178348 | 0.999532 | | 2.491261 | -0.42677 | 0.780418 | | 2.353278 | -0.59209 | 0.682165 |
| ACSL5 | 2.169969 | -0.35569 | 0.999532 | | 2.406076 | -0.41046 | 0.780461 | | 1.851013 | -0.03368 | 0.992286 |
| MYL6B | 3.760679 | -0.15706 | 0.999532 | | 4.95926 | 0.285219 | 0.780461 | | 3.912255 | 0.459719 | 0.664512 |
| MAN1A1 | 3.754465 | -0.00143 | 0.999532 | | 4.738063 | 0.309499 | 0.7806 | | 4.004008 | 0.330774 | 0.782299 |
| CDC20 | 2.574826 | 0.167693 | 0.999532 | | 2.411568 | -0.40415 | 0.781093 | | 2.233121 | -0.55474 | 0.68943 |
| PRSS23 | 6.928126 | -0.06348 | 0.999532 | | 8.901854 | 0.294148 | 0.781093 | | 7.046189 | 0.38108 | 0.762271 |
| TP63 | 2.421374 | 0.383993 | 0.999532 | | 2.014585 | -0.4546 | 0.78137 | | 2.076332 | -0.8228 | 0.562307 |
| CHM | 3.61298 | 0.318155 | 0.999532 | | 3.848285 | 0.331767 | 0.781573 | | 3.836071 | 0.024981 | 0.992286 |
| XPNPEP | 3.472916 | 0.023807 | 0.999532 | | 4.193218 | 0.305176 | 0.781573 | | 3.743638 | 0.301185 | 0.806394 |
| MDM1 | 2.451648 | -0.4057 | 0.999532 | | 2.850986 | -0.38276 | 0.781573 | | 2.215216 | 0.038098 | 0.989985 |
| STX4 | 7.781841 | -0.10175 | 0.999532 | | 10.20719 | 0.232503 | 0.781573 | | 8.412593 | 0.346887 | 0.658234 |
| SDHAF1 | 2.934217 | -0.11374 | 0.999532 | | 3.046318 | -0.40109 | 0.781573 | | 2.528901 | -0.27049 | 0.875488 |
| SAP30 | 2.877222 | -0.38445 | 0.999532 | | 3.03674 | -0.36826 | 0.781573 | | 2.473499 | 0.024881 | 0.994537 |
| SARDH | 7.606286 | 0.40322 | 0.999532 | | 8.023639 | 0.259287 | 0.781573 | | 8.170289 | -0.12697 | 0.915533 |
| SMG9 | 1.944892 | 0.265975 | 0.999532 | | 2.150902 | 0.427209 | 0.781573 | | 2.122468 | 0.18203 | 0.926957 |
| PARN | 3.468798 | -0.47759 | 0.999532 | | 4.141721 | -0.32375 | 0.781573 | | 2.852377 | 0.180013 | 0.911296 |
| GRB10 | 5.042197 | -0.34021 | 0.999532 | | 5.804681 | -0.28852 | 0.781815 | | 4.312902 | 0.065567 | 0.969691 |
| IDS | 5.904114 | 0.248073 | 0.999532 | | 6.412622 | 0.260697 | 0.781891 | | 6.00563 | 0.026685 | 0.989148 |
| MLLT11 | 2.537645 | 0.496457 | 0.999532 | | 2.121627 | -0.44541 | 0.781987 | | 2.226596 | -0.92574 | 0.456688 |
| ZNF696 | 4.810741 | -0.08146 | 0.999532 | | 4.632848 | -0.29496 | 0.782158 | | 4.236104 | -0.20127 | 0.883785 |
| HINT3 | 2.86943 | -0.56786 | 0.999532 | | 3.57258 | -0.344 | 0.782158 | | 2.469903 | 0.244474 | 0.883785 |
| CWC27 | 2.334206 | -0.29053 | 0.999532 | | 3.193552 | 0.368513 | 0.782158 | | 2.575122 | 0.683822 | 0.600081 |
| CDKAL1 | 4.096248 | -0.03084 | 0.999532 | | 3.884861 | -0.32705 | 0.782158 | | 3.558674 | -0.29508 | 0.836935 |
| SEZ6 | 2.210083 | -0.21142 | 0.999532 | | 2.353121 | -0.44304 | 0.782158 | | 1.983359 | -0.21386 | 0.918694 |
| CXCL16 | 3.070498 | 0.025493 | 0.999532 | | 3.79881 | 0.327797 | 0.782158 | | 3.298469 | 0.318219 | 0.806394 |
| TTLL6 | 2.393874 | -0.02816 | 0.999532 | | 2.432352 | -0.43458 | 0.782158 | | 2.048387 | -0.38694 | 0.827488 |
| ICAM2 | 6.162116 | -0.48409 | 0.999532 | | 8.956478 | 0.25053 | 0.782158 | | 6.725659 | 0.754027 | 0.298614 |
| TCTN3 | 3.679756 | -0.27013 | 0.999532 | | 4.877488 | 0.297944 | 0.782158 | | 3.861721 | 0.578342 | 0.59136 |
| ALS2 | 2.666981 | -0.13802 | 0.999532 | | 2.734136 | -0.38443 | 0.782229 | | 2.276863 | -0.23023 | 0.896369 |
| DDX23 | 4.09797 | -0.11827 | 0.999532 | | 5.210092 | 0.290427 | 0.782665 | | 4.199757 | 0.422791 | 0.699971 |
| DIP2A | 3.03328 | -0.35968 | 0.999532 | | 4.378237 | 0.316791 | 0.782665 | | 3.430165 | 0.697962 | 0.516313 |
| STYXL1 | 3.637266 | 0.177878 | 0.999532 | | 3.383396 | -0.34169 | 0.782951 | | 3.188681 | -0.50108 | 0.668289 |
| COL6A2 | 8.284702 | -0.12132 | 0.999532 | | 10.86995 | 0.290094 | 0.782951 | | 8.807846 | 0.431213 | 0.705888 |
| FBXO42 | 4.824706 | 0.25627 | 0.999532 | | 3.966784 | -0.32412 | 0.783026 | | 4.020032 | -0.59725 | 0.57215 |
| OGFOD1 | 2.959205 | -0.42189 | 0.999532 | | 3.330417 | -0.33689 | 0.783038 | | 2.467285 | 0.101294 | 0.95099 |
| ARAP1 | 2.984786 | 0.032212 | 0.999532 | | 3.802574 | 0.356544 | 0.783068 | | 3.202895 | 0.349861 | 0.803301 |
| ORMDL1 | 3.195356 | -0.03102 | 0.999532 | | 3.90078 | 0.319777 | 0.783236 | | 3.572939 | 0.389868 | 0.75311 |
| S100A8 | 3.363105 | -0.04737 | 0.999532 | | 3.285297 | -0.38591 | 0.783236 | | 2.845056 | -0.32622 | 0.824012 |
| SAMD5 | 1.975542 | 0.180759 | 0.999532 | | 2.381751 | 0.405694 | 0.783236 | | 2.131633 | 0.242725 | 0.892869 |
| REG4 | 2.244862 | -0.01691 | 0.999532 | | 2.182315 | -0.44788 | 0.783284 | | 1.879202 | -0.41298 | 0.818918 |
| GABPB2 | 2.296449 | -0.12631 | 0.999532 | | 2.314179 | -0.4107 | 0.783661 | | 1.979472 | -0.26522 | 0.88588 |
| ARIH1 | 4.594438 | 0.133275 | 0.999532 | | 5.482284 | 0.296199 | 0.783661 | | 4.859623 | 0.180681 | 0.88885 |
| MTOR | 3.761055 | 0.044945 | 0.999532 | | 4.536678 | 0.311301 | 0.783661 | | 3.849116 | 0.285978 | 0.816618 |
| KPNA2 | 2.903442 | 0.335741 | 0.999532 | | 3.475793 | 0.349782 | 0.783661 | | 3.269769 | 0.031723 | 0.990309 |
| MYLIP | 11.35917 | 0.066648 | 0.999532 | | 13.5805 | 0.242875 | 0.783661 | | 11.38699 | 0.212378 | 0.816372 |
| ATP11A | 3.203436 | -0.07134 | 0.999532 | | 3.959027 | 0.321349 | 0.783661 | | 3.284173 | 0.406061 | 0.740755 |
| NFYB | 3.937468 | 0.458372 | 0.999532 | | 4.529756 | 0.315651 | 0.783686 | | 4.296136 | -0.11287 | 0.941264 |
| SERTAD2 | 3.210588 | -0.23976 | 0.999532 | | 4.279684 | 0.31242 | 0.78384 | | 3.544151 | 0.566867 | 0.633201 |
| CTTNBP2 | 6.899234 | 0.244054 | 0.999532 | | 7.514607 | 0.258777 | 0.783942 | | 6.970018 | 0.033769 | 0.984624 |
| IQCF2 | 2.253947 | 0.519483 | 0.999532 | | 2.520438 | 0.446836 | 0.784233 | | 2.591247 | -0.05982 | 0.982338 |
| STK19 | 7.573186 | 0.023543 | 0.999532 | | 7.886418 | -0.24196 | 0.784445 | | 6.900564 | -0.24226 | 0.803301 |
| NSUN7 | 3.545445 | 0.270787 | 0.999532 | | 3.982412 | 0.321309 | 0.784445 | | 3.77968 | 0.068628 | 0.971732 |
| SAFB | 7.873332 | 0.069033 | 0.999532 | | 9.578537 | 0.232156 | 0.784682 | | 8.060423 | 0.184655 | 0.844873 |
| GPR89A | 4.797163 | 0.347131 | 0.999532 | | 4.415163 | -0.31022 | 0.785034 | | 4.220267 | -0.63489 | 0.516831 |
| SESN2 | 5.027237 | 0.24836 | 0.999532 | | 4.663494 | -0.30786 | 0.785377 | | 4.515571 | -0.54383 | 0.57215 |
| CNOT6 | 4.407686 | 0.010949 | 0.999532 | | 4.429899 | -0.30425 | 0.785813 | | 3.689419 | -0.29616 | 0.806394 |
| EFNB1 | 9.173404 | 0.304585 | 0.999532 | | 9.94907 | 0.231813 | 0.786293 | | 9.439662 | -0.05658 | 0.965649 |
| NUDCD1 | 3.866 | -0.15389 | 0.999532 | | 3.957305 | -0.34798 | 0.786365 | | 3.353089 | -0.17742 | 0.910105 |
| IFNL4 | 5.095917 | 0.049714 | 0.999532 | | 5.06751 | -0.32824 | 0.786365 | | 4.537691 | -0.36481 | 0.769334 |
| MX1 | 3.943642 | 0.445992 | 0.999532 | | 3.898126 | 0.312108 | 0.786383 | | 4.097346 | -0.11907 | 0.942744 |
| KCP | 2.472953 | 0.269625 | 0.999532 | | 2.223464 | -0.44002 | 0.786426 | | 2.017064 | -0.68906 | 0.630278 |
| SLC39A3 | 2.569623 | 0.555386 | 0.999532 | | 2.756962 | 0.369374 | 0.786748 | | 2.69293 | -0.1661 | 0.918694 |
| ISG15 | 9.951351 | 0.165295 | 0.999532 | | 11.40228 | 0.229325 | 0.786748 | | 10.74656 | 0.08964 | 0.939549 |
| RNF2 | 3.098736 | -0.16961 | 0.999532 | | 4.27044 | 0.327744 | 0.787827 | | 3.338935 | 0.513301 | 0.662584 |
| TEX264 | 5.34191 | 0.24405 | 0.999532 | | 5.627629 | 0.309795 | 0.78793 | | 5.358628 | 0.096717 | 0.947851 |
| REG3G | 2.184199 | 0.213889 | 0.999532 | | 2.011479 | -0.44224 | 0.78793 | | 1.838953 | -0.6394 | 0.679818 |
| RBM23 | 8.419247 | 0.257823 | 0.999532 | | 9.553418 | 0.21312 | 0.788078 | | 8.3959 | -0.03212 | 0.982124 |
| WDR77 | 3.996683 | -0.31567 | 0.999532 | | 4.324454 | -0.3174 | 0.788083 | | 3.275761 | 0.01529 | 0.996681 |
| ZMYM6 | 2.801632 | 0.370194 | 0.999532 | | 2.378401 | -0.42937 | 0.788083 | | 2.327206 | -0.78572 | 0.56156 |
| ARPP19 | 7.296351 | -0.0503 | 0.999532 | | 9.034335 | 0.24377 | 0.788083 | | 7.542314 | 0.312078 | 0.737952 |
| TMEM127 | 6.402446 | 0.290565 | 0.999532 | | 7.142068 | 0.269929 | 0.788127 | | 6.724195 | -0.0017 | 0.999385 |
| SULT1B1 | 2.944811 | 0.114037 | 0.999532 | | 2.874762 | -0.39596 | 0.788129 | | 2.596511 | -0.49512 | 0.734912 |
| GCK | 1.95142 | 0.036561 | 0.999532 | | 1.850578 | -0.46716 | 0.788129 | | 1.571446 | -0.48343 | 0.791576 |
| INTS14 | 2.634015 | 0.017436 | 0.999532 | | 2.680806 | -0.39564 | 0.788129 | | 2.256391 | -0.39099 | 0.809832 |
| ZNF41 | 2.734476 | -0.17362 | 0.999532 | | 2.812636 | -0.38477 | 0.788313 | | 2.302055 | -0.19269 | 0.917499 |
| METAP1 | 3.600502 | -0.31472 | 0.999532 | | 3.793319 | -0.33896 | 0.788414 | | 2.799378 | -0.01427 | 0.996891 |
| PAQR4 | 2.803249 | 0.157123 | 0.999532 | | 2.56784 | -0.42452 | 0.788426 | | 2.479925 | -0.57433 | 0.692762 |
| TMEM108 | 1.983109 | -0.41097 | 0.999532 | | 2.296271 | -0.45 | 0.788452 | | 1.792308 | -0.01967 | 0.996681 |
| IL17RB | 2.443535 | 0.382361 | 0.999532 | | 2.851848 | 0.395945 | 0.788491 | | 2.620546 | 0.035911 | 0.989985 |
| BCAS2 | 5.242868 | 0.369744 | 0.999532 | | 5.536809 | 0.27441 | 0.788491 | | 5.354865 | -0.07671 | 0.958191 |
| SNX8 | 2.755186 | 0.122747 | 0.999532 | | 2.556278 | -0.39084 | 0.788491 | | 2.299643 | -0.49547 | 0.733499 |
| SNHG32 | 7.691439 | -0.20257 | 0.999532 | | 10.31025 | 0.213957 | 0.788491 | | 7.958322 | 0.440811 | 0.563455 |
| DHX15 | 10.01962 | -0.17275 | 0.999532 | | 11.07213 | -0.21407 | 0.788491 | | 9.031066 | -0.02583 | 0.986898 |
| GPR182 | 3.107224 | -0.19418 | 0.999532 | | 3.190195 | -0.38461 | 0.788491 | | 2.691379 | -0.16648 | 0.933644 |
| GUF1 | 2.380461 | 0.133795 | 0.999532 | | 2.861292 | 0.381626 | 0.788491 | | 2.526886 | 0.267931 | 0.874255 |
| ANAPC4 | 2.796626 | -0.15124 | 0.999532 | | 3.46366 | 0.327822 | 0.788491 | | 2.893045 | 0.48844 | 0.702463 |
| PGR | 6.113405 | -0.37455 | 0.999532 | | 7.388782 | -0.24818 | 0.788491 | | 5.559184 | 0.15413 | 0.903339 |
| EXOC1L | 2.346005 | 0.187249 | 0.999532 | | 2.200337 | -0.42679 | 0.788491 | | 2.024983 | -0.5979 | 0.689943 |
| MTMR12 | 5.967424 | -0.36166 | 0.999532 | | 6.648808 | -0.25031 | 0.788491 | | 5.08616 | 0.116651 | 0.932589 |
| MGST1 | 3.053274 | 0.263209 | 0.999532 | | 3.686288 | 0.381607 | 0.788491 | | 3.294529 | 0.142887 | 0.934354 |
| SHOC2 | 4.194969 | 0.189512 | 0.999532 | | 4.669403 | 0.297189 | 0.788491 | | 4.411598 | 0.125206 | 0.93222 |
| PLA2G4B | 2.263787 | 0.633078 | 0.999532 | | 2.314323 | 0.420391 | 0.788491 | | 2.47261 | -0.19689 | 0.915991 |
| SP8 | 2.293726 | 0.763967 | 0.999532 | | 2.185834 | 0.4237 | 0.788491 | | 2.461582 | -0.3213 | 0.851503 |
| KRTAP10 | 5.500763 | 0.166225 | 0.999532 | | 5.365922 | -0.29236 | 0.788491 | | 4.980307 | -0.44174 | 0.662584 |
| FAT2 | 2.428913 | 0.195816 | 0.999532 | | 2.144322 | -0.4307 | 0.788491 | | 2.064332 | -0.6089 | 0.682165 |
| RAN | 9.272193 | 0.052191 | 0.999532 | | 10.87996 | 0.20795 | 0.788491 | | 9.464364 | 0.174179 | 0.847311 |
| MAP9 | 4.051106 | 0.141217 | 0.999532 | | 3.864945 | -0.32165 | 0.788491 | | 3.58928 | -0.44722 | 0.69941 |
| GCNT3 | 2.458937 | 0.166454 | 0.999532 | | 2.405227 | -0.42563 | 0.788491 | | 2.324081 | -0.5717 | 0.718643 |
| MRGPRX | 3.091369 | 0.784443 | 0.999532 | | 2.19809 | -0.43327 | 0.788578 | | 2.648784 | -1.19783 | 0.247358 |
| OVCA2 | 4.420625 | -0.30599 | 0.999532 | | 5.156799 | -0.2823 | 0.788578 | | 3.828359 | 0.042928 | 0.982187 |
| MLPH | 2.176109 | 0.307008 | 0.999532 | | 2.004755 | -0.4623 | 0.788578 | | 1.931384 | -0.75319 | 0.623462 |
| FNBP1 | 9.54495 | 0.03764 | 0.999532 | | 11.52742 | 0.2272 | 0.788578 | | 10.08301 | 0.202857 | 0.814402 |
| PTPRA | 9.229781 | -0.16433 | 0.999532 | | 11.69252 | 0.221766 | 0.788578 | | 9.471329 | 0.409958 | 0.552672 |
| DNAJC1 | 4.733242 | 0.323891 | 0.999532 | | 4.914065 | 0.285148 | 0.788578 | | 4.998533 | -0.02195 | 0.992286 |
| CMTR2 | 1.839659 | 0.134912 | 0.999532 | | 2.163543 | 0.410607 | 0.788578 | | 2.03881 | 0.292634 | 0.872435 |
| AMDHD1 | 2.510323 | 0.54217 | 0.999532 | | 1.984235 | -0.43674 | 0.788595 | | 2.159497 | -0.9618 | 0.436389 |
| H2BC18 | 10.43811 | -0.04479 | 0.999532 | | 11.06612 | -0.22121 | 0.788645 | | 9.12377 | -0.15408 | 0.872435 |
| OR7C2 | 3.024095 | 0.151445 | 0.999532 | | 3.087418 | -0.36092 | 0.788716 | | 2.735562 | -0.47158 | 0.736481 |
| CCDC24 | 5.535002 | -0.36637 | 0.999532 | | 6.301922 | -0.28507 | 0.788947 | | 4.709739 | 0.108575 | 0.938429 |
| TET3 | 10.72865 | -0.04052 | 0.999532 | | 11.23387 | -0.20978 | 0.788947 | | 9.431448 | -0.14951 | 0.869224 |
| CHRNA10 | 2.511383 | 0.125688 | 0.999532 | | 2.419103 | -0.41471 | 0.788947 | | 2.199964 | -0.52351 | 0.728015 |
| RPL27A | 36.44057 | -0.21756 | 0.999532 | | 47.51302 | 0.213462 | 0.789763 | | 37.91964 | 0.453571 | 0.49299 |
| RAB11FI | 2.310082 | 0.22191 | 0.999532 | | 2.021099 | -0.42905 | 0.789774 | | 2.083956 | -0.62532 | 0.686214 |
| GET1 | 7.946569 | 0.065691 | 0.999532 | | 9.924233 | 0.231245 | 0.789873 | | 8.447216 | 0.187199 | 0.844187 |
| ABHD8 | 2.226616 | -0.12549 | 0.999532 | | 2.314065 | -0.40907 | 0.789873 | | 1.845236 | -0.26508 | 0.890062 |
| PPIL1 | 2.356234 | 0.053491 | 0.999532 | | 2.434731 | -0.40412 | 0.789949 | | 2.093184 | -0.43136 | 0.791576 |
| FRMD8 | 3.451845 | 0.03518 | 0.999532 | | 4.312828 | 0.310504 | 0.790156 | | 3.622931 | 0.29473 | 0.816589 |
| TSPYL5 | 2.395481 | 0.046499 | 0.999532 | | 2.398654 | -0.42154 | 0.790156 | | 1.945683 | -0.45496 | 0.769793 |
| UHRF1 | 2.032796 | -0.22159 | 0.999532 | | 2.18427 | -0.41835 | 0.790311 | | 1.743279 | -0.17384 | 0.939107 |
| TMEM250 | 5.626713 | 0.169014 | 0.999532 | | 5.535627 | -0.28352 | 0.790311 | | 5.042513 | -0.43465 | 0.652925 |
| NCK2 | 9.124718 | 0.020637 | 0.999532 | | 11.08179 | 0.21902 | 0.790572 | | 9.538389 | 0.216435 | 0.805576 |
| HEXD | 4.160927 | 0.437015 | 0.999532 | | 4.426215 | 0.315015 | 0.790802 | | 4.36594 | -0.10176 | 0.947322 |
| TNFAIP8L | 2.72742 | 0.207386 | 0.999532 | | 2.419196 | -0.40936 | 0.791148 | | 2.340135 | -0.59916 | 0.671153 |
| TRADD | 2.878184 | 0.208217 | 0.999532 | | 3.204113 | 0.345524 | 0.791247 | | 2.985777 | 0.158985 | 0.920527 |
| ZFAND2A | 3.394867 | 0.058749 | 0.999532 | | 3.499235 | -0.35225 | 0.791247 | | 3.040797 | -0.39046 | 0.773057 |
| PABPC3 | 2.300879 | 0.299785 | 0.999532 | | 2.089299 | -0.43717 | 0.791409 | | 1.973516 | -0.71706 | 0.626895 |
| LYZ | 5.857452 | 0.144061 | 0.999532 | | 6.801499 | 0.355599 | 0.791409 | | 6.340758 | 0.227351 | 0.881011 |
| ZSCAN5A | 2.601542 | -0.06401 | 0.999532 | | 2.765049 | -0.42969 | 0.791409 | | 2.340214 | -0.35615 | 0.837256 |
| DNAJB2 | 7.286359 | 0.440067 | 0.999532 | | 7.401433 | 0.257685 | 0.791409 | | 7.232719 | -0.16037 | 0.883792 |
| SLC16A1 | 4.22563 | 0.376484 | 0.999532 | | 3.510336 | -0.39752 | 0.791409 | | 3.437476 | -0.73887 | 0.524206 |
| WIPI1 | 3.560547 | -0.38569 | 0.999532 | | 4.872634 | 0.284395 | 0.791436 | | 3.774155 | 0.685615 | 0.478728 |
| HCN2 | 2.790343 | -0.17151 | 0.999532 | | 2.939773 | -0.44235 | 0.791561 | | 2.275424 | -0.23816 | 0.901847 |

| | | | | | | | | | | | |
|---|---|---|---|---|---|---|---|---|---|---|---|
| TM2D3 | 7.847108 | -0.354 | 0.999532 | | 10.78916 | 0.218968 | 0.791561 | | 8.008489 | 0.590709 | 0.387955 |
| CYP4Z1 | 6.233302 | 0.248434 | 0.999532 | | 6.021816 | -0.30001 | 0.791561 | | 5.875499 | -0.55727 | 0.552672 |
| ZNF568 | 4.437993 | 0.318496 | 0.999532 | | 5.338085 | 0.294255 | 0.79161 | | 5.021991 | -0.0094 | 0.996891 |
| BRD2 | 12.02517 | -0.22117 | 0.999532 | | 15.54075 | 0.20105 | 0.79161 | | 11.98765 | 0.45223 | 0.515762 |
| ANKRD29 | 2.645056 | 0.431267 | 0.999532 | | 2.193769 | -0.43081 | 0.79161 | | 2.346232 | -0.84457 | 0.543211 |
| MAGEE1 | 2.872743 | 0.528529 | 0.999532 | | 3.208884 | 0.365368 | 0.79161 | | 3.195204 | -0.13632 | 0.939549 |
| LZTR1 | 2.427403 | 0.187207 | 0.999532 | | 2.741548 | 0.376588 | 0.79161 | | 2.536745 | 0.205146 | 0.904801 |
| TBC1D22 | 3.711625 | 0.370158 | 0.999532 | | 4.074517 | 0.32451 | 0.79161 | | 3.931646 | -0.02232 | 0.992459 |
| PLCG1 | 5.824642 | -0.25505 | 0.999532 | | 7.827131 | 0.241419 | 0.79161 | | 5.941268 | 0.507503 | 0.575703 |
| VAV3 | 2.836376 | 0.197061 | 0.999532 | | 3.624512 | 0.327523 | 0.79161 | | 3.122705 | 0.163803 | 0.921563 |
| LILRB2 | 4.971584 | -0.37845 | 0.999532 | | 5.63716 | -0.27519 | 0.792008 | | 4.28995 | 0.11714 | 0.94109 |
| DTX1 | 3.199023 | 0.196791 | 0.999532 | | 2.841269 | -0.37867 | 0.792151 | | 2.637914 | -0.56138 | 0.672452 |
| AADACL4 | 2.143281 | 0.007252 | 0.999532 | | 2.130074 | -0.43444 | 0.792151 | | 1.737314 | -0.42096 | 0.811123 |
| COX19 | 3.728101 | -0.00176 | 0.999532 | | 3.725564 | -0.31846 | 0.792151 | | 3.346608 | -0.30386 | 0.818918 |
| IBTK | 3.34997 | 0.375295 | 0.999532 | | 3.786153 | 0.344153 | 0.792175 | | 3.720395 | -0.01612 | 0.996681 |
| CDHR5 | 3.276358 | 0.495392 | 0.999532 | | 3.246293 | 0.369527 | 0.792526 | | 3.336311 | -0.12113 | 0.948127 |
| ASF1A | 2.91601 | 0.363599 | 0.999532 | | 3.308433 | 0.354762 | 0.792526 | | 2.975841 | 0.016546 | 0.996681 |
| PRDM15 | 2.98024 | 0.135732 | 0.999532 | | 2.813579 | -0.37967 | 0.792526 | | 2.533171 | -0.49828 | 0.723294 |
| EMID1 | 5.608385 | -0.04633 | 0.999532 | | 7.43272 | 0.251547 | 0.792737 | | 6.055228 | 0.321319 | 0.740445 |
| PSMB10 | 6.718867 | 0.186102 | 0.999532 | | 6.030698 | -0.28777 | 0.792737 | | 5.890873 | -0.45608 | 0.652925 |
| SUPT20H | 2.524103 | 0.212185 | 0.999532 | | 2.934441 | 0.363501 | 0.792918 | | 2.61098 | 0.172184 | 0.918194 |
| MCM6 | 4.153108 | 0.342411 | 0.999532 | | 4.561272 | 0.298376 | 0.793117 | | 4.475238 | -0.02342 | 0.992286 |
| SSRP1 | 9.218635 | 0.098552 | 0.999532 | | 10.69084 | 0.21916 | 0.793189 | | 9.318469 | 0.140526 | 0.890746 |
| ARHGAP | 2.555925 | 0.555352 | 0.999532 | | 2.756069 | 0.367846 | 0.79332 | | 2.799506 | -0.1662 | 0.920259 |
| CPA3 | 2.288538 | 0.16753 | 0.999532 | | 2.108806 | -0.43613 | 0.793376 | | 1.968809 | -0.58664 | 0.712891 |
| HOPX | 3.185494 | 0.239877 | 0.999532 | | 3.773013 | 0.325633 | 0.793544 | | 3.406625 | 0.107807 | 0.948865 |
| SKA3 | 2.023186 | -0.03659 | 0.999532 | | 1.824777 | -0.46426 | 0.793905 | | 1.681147 | -0.41718 | 0.837256 |
| COMMD4 | 6.803088 | 0.015887 | 0.999532 | | 8.282774 | 0.246453 | 0.79414 | | 7.267052 | 0.251037 | 0.814402 |
| TAF4 | 3.559729 | 0.049877 | 0.999532 | | 4.419192 | 0.315463 | 0.79414 | | 3.94711 | 0.282635 | 0.829716 |
| NKX2-5 | 2.399569 | 0.332626 | 0.999532 | | 2.168794 | -0.4437 | 0.79414 | | 2.117682 | -0.76073 | 0.598934 |
| DCTN2 | 6.319724 | -0.48106 | 0.999532 | | 9.094986 | 0.237484 | 0.79414 | | 6.687189 | 0.7382 | 0.283338 |
| PYROXD2 | 2.11496 | -0.31638 | 0.999532 | | 2.238254 | -0.42618 | 0.79414 | | 1.758086 | -0.09837 | 0.972221 |
| CBS | 4.035368 | -0.03152 | 0.999532 | | 4.127773 | -0.31956 | 0.79414 | | 3.595635 | -0.27259 | 0.843564 |
| C1orf109 | 3.907799 | -0.09303 | 0.999532 | | 3.981139 | -0.31766 | 0.79414 | | 3.292981 | -0.20751 | 0.88885 |
| SNAPC2 | 4.285543 | -0.07573 | 0.999532 | | 4.306619 | -0.30937 | 0.79414 | | 3.647016 | -0.21478 | 0.881316 |
| CMC2 | 3.592411 | 0.203581 | 0.999532 | | 3.869776 | 0.330743 | 0.79414 | | 3.750685 | 0.119316 | 0.946572 |
| GDF11 | 3.505106 | 0.233579 | 0.999532 | | 3.172278 | -0.34151 | 0.79414 | | 3.037944 | -0.55921 | 0.637967 |
| PRKCA | 7.377498 | -0.01123 | 0.999532 | | 8.750172 | 0.243755 | 0.79414 | | 7.548407 | 0.273038 | 0.7719 |
| KTN1 | 7.40034 | -0.38926 | 0.999532 | | 10.20539 | 0.219819 | 0.79414 | | 7.843421 | 0.629149 | 0.30352 |
| UBP1 | 4.423828 | -0.24638 | 0.999532 | | 5.632634 | 0.272483 | 0.79414 | | 4.562933 | 0.536549 | 0.572297 |
| SLC25A1 | 7.264732 | 0.466728 | 0.999532 | | 6.245081 | -0.35031 | 0.79414 | | 6.051334 | -0.81816 | 0.401964 |
| ZNF45 | 2.522213 | -0.18506 | 0.999532 | | 2.771147 | -0.38016 | 0.79414 | | 2.225149 | -0.16805 | 0.934172 |
| SLC22A1 | 2.582735 | 0.414144 | 0.999532 | | 2.749339 | 0.383706 | 0.79414 | | 2.734782 | -0.01029 | 0.996891 |
| PPP1R14 | 2.837197 | -0.1333 | 0.999532 | | 3.080403 | -0.35077 | 0.79414 | | 2.373809 | -0.19685 | 0.907106 |
| LRTOMT | 3.208686 | -0.26705 | 0.999532 | | 3.362331 | -0.35373 | 0.79414 | | 2.826464 | -0.07054 | 0.974588 |
| PARVB | 3.394063 | 0.410759 | 0.999532 | | 3.928777 | 0.322166 | 0.79414 | | 3.834056 | -0.06545 | 0.972758 |
| TBXAS1 | 3.214249 | 0.46448 | 0.999532 | | 3.62353 | 0.320865 | 0.79414 | | 3.540286 | -0.12971 | 0.93042 |
| WFDC3 | 2.464714 | -0.06818 | 0.999532 | | 2.541386 | -0.40954 | 0.79414 | | 2.156023 | -0.32229 | 0.862045 |
| PHF1 | 4.660254 | 0.01213 | 0.999532 | | 5.624651 | 0.264632 | 0.79414 | | 4.823889 | 0.269268 | 0.809832 |
| HAUS1 | 3.216952 | -0.06474 | 0.999532 | | 3.334597 | -0.35086 | 0.79414 | | 2.933419 | -0.26961 | 0.864923 |
| ADIRF | 8.035524 | -0.22677 | 0.999532 | | 10.58893 | 0.249905 | 0.79414 | | 8.654341 | 0.495516 | 0.505783 |
| AGFG2 | 3.138549 | -0.30842 | 0.999532 | | 3.444265 | -0.35154 | 0.79414 | | 2.460892 | -0.02254 | 0.994537 |
| ZNF362 | 9.231082 | -0.10515 | 0.999532 | | 11.86663 | 0.219521 | 0.79414 | | 9.716726 | 0.33698 | 0.652925 |
| TCAF1 | 9.336012 | -0.26902 | 0.999532 | | 10.53519 | -0.23297 | 0.79414 | | 8.116279 | 0.054075 | 0.972305 |
| XAGE2 | 3.007204 | 0.934231 | 0.999532 | | 2.828982 | 0.390252 | 0.79414 | | 3.243559 | -0.526 | 0.662576 |
| EXOSC1 | 4.735356 | 0.227949 | 0.999532 | | 4.466001 | -0.30748 | 0.79414 | | 4.210541 | -0.51697 | 0.628895 |
| UFD1 | 5.231575 | 0.146042 | 0.999532 | | 5.945236 | 0.258777 | 0.79414 | | 5.528397 | 0.125168 | 0.921185 |
| SLC16A5 | 4.565081 | -0.02755 | 0.999532 | | 5.023326 | -0.30053 | 0.79414 | | 3.860463 | -0.22973 | 0.872493 |
| STK4 | 3.246932 | -0.01812 | 0.999532 | | 3.910345 | 0.336055 | 0.79414 | | 3.525117 | 0.383317 | 0.750371 |
| TNFRSF1 | 5.294254 | -0.36472 | 0.999532 | | 6.447564 | -0.24824 | 0.79414 | | 4.61141 | 0.135456 | 0.919493 |
| SYNCRIP | 8.519847 | -0.04712 | 0.999532 | | 10.44554 | 0.224899 | 0.79414 | | 8.60052 | 0.294788 | 0.720106 |
| DMRTA2 | 3.90433 | 0.772 | 0.999532 | | 2.941336 | -0.37614 | 0.79414 | | 3.513719 | -1.13274 | 0.19389 |
| TMEM243 | 2.811171 | 0.146587 | 0.999532 | | 3.485486 | 0.345962 | 0.79414 | | 3.161012 | 0.218935 | 0.890221 |
| VGLL1 | 2.258926 | 0.306974 | 0.999532 | | 1.940567 | -0.4461 | 0.79414 | | 1.967406 | -0.73357 | 0.638015 |
| OR10G3 | 2.876266 | 0.151099 | 0.999532 | | 2.728388 | -0.41576 | 0.79414 | | 2.462126 | -0.55244 | 0.720106 |
| EIF3F | 14.11892 | -0.02479 | 0.999532 | | 16.9031 | 0.178365 | 0.79414 | | 14.51604 | 0.22084 | 0.756439 |
| FAS | 3.629288 | -0.11018 | 0.999532 | | 3.62888 | -0.33355 | 0.79414 | | 3.074352 | -0.23049 | 0.88885 |
| NFIC | 6.310638 | 0.047028 | 0.999532 | | 7.750657 | 0.247247 | 0.79414 | | 6.36198 | 0.213312 | 0.842696 |
| NLK | 7.372592 | -0.01877 | 0.999532 | | 9.040777 | 0.224962 | 0.79414 | | 7.701166 | 0.263479 | 0.761337 |
| VNN1 | 3.811299 | -0.04335 | 0.999532 | | 3.984344 | -0.3373 | 0.79414 | | 3.281329 | -0.27496 | 0.851503 |
| UBTFL1 | 2.534197 | -0.22503 | 0.999532 | | 2.823158 | -0.38872 | 0.794642 | | 2.249908 | -0.13706 | 0.950895 |
| IL4R | 3.738865 | -0.46094 | 0.999532 | | 4.16805 | -0.30896 | 0.794692 | | 3.201766 | 0.17115 | 0.914687 |
| UBR3 | 5.382008 | -0.26552 | 0.999532 | | 6.843717 | 0.242381 | 0.794802 | | 5.409448 | 0.523507 | 0.536289 |
| DCLK1 | 3.019383 | 0.135263 | 0.999532 | | 2.883423 | -0.3538 | 0.794892 | | 2.621474 | -0.47181 | 0.720106 |
| NUP210 | 2.592312 | 0.406912 | 0.999532 | | 2.176793 | -0.42763 | 0.794928 | | 2.197153 | -0.81672 | 0.540342 |
| GTF2F2 | 3.731632 | -0.35468 | 0.999532 | | 5.52735 | 0.268833 | 0.795073 | | 4.131242 | 0.653653 | 0.502623 |
| KRTAP1- | 1.869024 | 0.493891 | 0.999532 | | 1.948323 | 0.441425 | 0.795568 | | 2.064411 | -0.03603 | 0.991434 |
| RDH5 | 2.718702 | -0.02588 | 0.999532 | | 2.646781 | -0.37781 | 0.795568 | | 2.348521 | -0.3426 | 0.842754 |
| MRPL17 | 3.569689 | 0.072957 | 0.999532 | | 4.456115 | 0.287913 | 0.79567 | | 3.743982 | 0.233475 | 0.861815 |
| KCNA5 | 2.317674 | 0.013783 | 0.999532 | | 2.384975 | -0.43977 | 0.79602 | | 1.995248 | -0.43586 | 0.799944 |
| DENR | 4.591794 | 0.133778 | 0.999532 | | 5.35359 | 0.300895 | 0.79615 | | 5.018091 | 0.166024 | 0.906811 |
| CUL1 | 5.971438 | -0.16311 | 0.999532 | | 6.41839 | -0.25287 | 0.796265 | | 5.08843 | -0.0758 | 0.960773 |
| POU3F1 | 2.400717 | 0.321357 | 0.999532 | | 2.195395 | -0.47717 | 0.796355 | | 2.158279 | -0.81833 | 0.563455 |
| APOC1 | 2.836205 | 0.505871 | 0.999532 | | 2.932789 | 0.38622 | 0.796619 | | 3.08352 | -0.11081 | 0.955834 |
| MED7 | 4.363653 | -0.06877 | 0.999532 | | 4.78535 | -0.292 | 0.796673 | | 3.896776 | -0.19885 | 0.885783 |
| SNPH | 2.399297 | 0.484233 | 0.999532 | | 2.561603 | 0.392441 | 0.796673 | | 2.527891 | -0.0726 | 0.974421 |
| SCPEP1 | 3.13921 | 0.215756 | 0.999532 | | 3.663032 | 0.312827 | 0.796673 | | 3.330456 | 0.11385 | 0.945759 |
| PANK4 | 3.859155 | -0.25083 | 0.999532 | | 3.975496 | -0.30801 | 0.796673 | | 3.265783 | -0.04838 | 0.982124 |
| MITF | 3.964407 | 0.085203 | 0.999532 | | 4.169785 | -0.31877 | 0.796673 | | 3.473636 | -0.37326 | 0.764149 |
| TSPAN18 | 3.048764 | -0.19106 | 0.999532 | | 4.077582 | 0.321037 | 0.796673 | | 3.232264 | 0.530904 | 0.663709 |
| KDF1 | 2.942248 | 0.464537 | 0.999532 | | 2.498847 | -0.39732 | 0.796815 | | 2.603942 | -0.83996 | 0.53177 |
| GOLGA3 | 5.294157 | 0.096106 | 0.999532 | | 6.235865 | 0.25523 | 0.796815 | | 5.430924 | 0.176686 | 0.881837 |
| KAT5 | 4.833741 | 0.055206 | 0.999532 | | 5.47319 | 0.281373 | 0.797073 | | 4.933779 | 0.23485 | 0.849128 |
| MSANTD | 2.14948 | -0.06277 | 0.999532 | | 2.316078 | -0.42871 | 0.797073 | | 1.919507 | -0.33927 | 0.862823 |
| H3-3B | 41.42489 | -0.12496 | 0.999532 | | 50.80469 | 0.143373 | 0.797256 | | 41.39464 | 0.288631 | 0.584522 |
| ZNF709 | 3.584392 | -0.16621 | 0.999532 | | 3.734103 | -0.31986 | 0.797337 | | 3.041103 | -0.13872 | 0.93603 |
| YIPF4 | 4.128799 | 0.015087 | 0.999532 | | 4.86629 | 0.307163 | 0.797337 | | 4.214315 | 0.298673 | 0.82512 |
| PAF1 | 6.401502 | -0.26732 | 0.999532 | | 7.046217 | -0.24321 | 0.797458 | | 5.519457 | 0.03625 | 0.984588 |
| PTBP2 | 2.576221 | -0.04113 | 0.999532 | | 3.206939 | 0.337067 | 0.797458 | | 2.698951 | 0.395602 | 0.769793 |
| BTBD18 | 1.888994 | -0.32559 | 0.999532 | | 2.192594 | -0.43177 | 0.797601 | | 1.763317 | -0.07073 | 0.982124 |
| LRRC4 | 1.984242 | 0.323937 | 0.999532 | | 1.797126 | -0.45719 | 0.797601 | | 1.653437 | -0.7579 | 0.632113 |
| TIMM17B | 3.41181 | -0.36836 | 0.999532 | | 3.798023 | -0.32121 | 0.797601 | | 2.735315 | 0.063089 | 0.976178 |
| CCL20 | 2.419147 | -0.12477 | 0.999532 | | 2.575234 | -0.40835 | 0.797601 | | 2.128077 | -0.26157 | 0.892869 |
| MAGED2 | 9.497547 | -0.4477 | 0.999532 | | 13.37281 | 0.238394 | 0.797601 | | 9.964229 | 0.705195 | 0.290091 |
| KIAA0232 | 2.288411 | -0.06315 | 0.999532 | | 2.847988 | 0.353466 | 0.797601 | | 2.347632 | 0.421445 | 0.779082 |
| TRIM49 | 2.250953 | 0.618643 | 0.999532 | | 1.661842 | -0.46913 | 0.797601 | | 1.987178 | -1.07226 | 0.436389 |
| STARD3N | 2.944115 | 0.111049 | 0.999532 | | 2.772314 | -0.36999 | 0.797601 | | 2.465167 | -0.4636 | 0.737539 |
| DHRS1 | 75.90293 | 0.122652 | 0.999532 | | 75.75125 | -0.24246 | 0.797752 | | 67.69821 | -0.34022 | 0.679818 |
| SEPTIN5 | 6.179467 | 0.138008 | 0.999532 | | 7.241884 | 0.261471 | 0.797817 | | 6.653631 | 0.16812 | 0.877487 |
| CCDC43 | 1.861102 | -0.03995 | 0.999532 | | 2.464121 | 0.396437 | 0.797817 | | 2.029286 | 0.454797 | 0.782299 |
| HSPA12B | 2.474159 | 0.044904 | 0.999532 | | 2.832042 | 0.355576 | 0.797817 | | 2.582052 | 0.322031 | 0.835972 |
| RAE1 | 2.668613 | -0.34271 | 0.999532 | | 3.636871 | 0.321922 | 0.797817 | | 2.861455 | 0.679253 | 0.560424 |
| GNB1 | 9.489114 | 0.019325 | 0.999532 | | 10.87601 | 0.216277 | 0.797817 | | 9.759179 | 0.207975 | 0.821752 |
| FBXO9 | 5.193609 | -0.00136 | 0.999532 | | 6.248941 | 0.25351 | 0.797887 | | 5.334742 | 0.273691 | 0.791576 |
| TMEM235 | 2.904511 | 0.356387 | 0.999532 | | 2.396365 | -0.40706 | 0.79795 | | 2.464906 | -0.75024 | 0.578893 |
| MAPRE2 | 2.95066 | -0.02254 | 0.999532 | | 3.017942 | -0.35324 | 0.79795 | | 2.629164 | -0.31255 | 0.843043 |
| ALOX5 | 2.744365 | 0.460569 | 0.999532 | | 2.397997 | -0.39826 | 0.79795 | | 2.392718 | -0.83771 | 0.49299 |
| TRPM6 | 2.279859 | 0.153865 | 0.999532 | | 2.087791 | -0.46085 | 0.79795 | | 1.815078 | -0.58832 | 0.717627 |
| MFSD14A | 4.761022 | -0.60963 | 0.999532 | | 6.745421 | 0.23712 | 0.798614 | | 4.93559 | 0.860677 | 0.24893 |
| TESK1 | 3.15179 | 0.093803 | 0.999532 | | 3.159634 | -0.34369 | 0.798614 | | 2.723537 | -0.41534 | 0.75311 |
| KCNH4 | 2.353107 | -0.16032 | 0.999532 | | 2.301428 | -0.40157 | 0.798791 | | 2.099103 | -0.23767 | 0.906811 |
| TNNI1 | 4.166307 | 0.28888 | 0.999532 | | 3.96207 | -0.34384 | 0.798791 | | 3.584079 | -0.6006 | 0.603483 |
| GTF2E2 | 2.841117 | -0.41875 | 0.999532 | | 4.033187 | 0.2993 | 0.798926 | | 3.020318 | 0.734944 | 0.515762 |
| GPR1 | 2.802699 | 0.555283 | 0.999532 | | 2.84763 | 0.361093 | 0.798926 | | 2.962078 | -0.17049 | 0.915533 |
| PRF1 | 3.511473 | 0.121465 | 0.999532 | | 3.479601 | -0.35687 | 0.798926 | | 3.126867 | -0.45492 | 0.740755 |
| GTF3C1 | 4.291073 | 0.114779 | 0.999532 | | 4.920265 | 0.279303 | 0.798926 | | 4.336649 | 0.170428 | 0.90424 |
| EVL | 13.09446 | -0.02382 | 0.999532 | | 16.14644 | 0.189815 | 0.799058 | | 13.23969 | 0.230593 | 0.806394 |
| PGBD1 | 1.973321 | -0.15539 | 0.999532 | | 2.132019 | -0.43124 | 0.799349 | | 1.795733 | -0.25401 | 0.90424 |
| BMT2 | 2.784805 | -0.00486 | 0.999532 | | 2.453094 | -0.38363 | 0.799755 | | 2.368628 | -0.35767 | 0.828971 |
| CHRNG | 2.059448 | 0.550042 | 0.999532 | | 2.297968 | 0.424228 | 0.799755 | | 2.403646 | -0.11037 | 0.96169 |
| QSOX1 | 4.369481 | 0.055266 | 0.999532 | | 5.488735 | 0.273063 | 0.799786 | | 4.515567 | 0.243184 | 0.842696 |
| USP42 | 3.193752 | -0.02176 | 0.999532 | | 3.274438 | -0.33361 | 0.799786 | | 2.699838 | -0.29274 | 0.843023 |
| ADAMTS7 | 2.38701 | -1.03347 | 0.999532 | | 2.996276 | -0.35436 | 0.799786 | | 1.820967 | 0.693303 | 0.649779 |
| EXOC3L4 | 2.077595 | 0.510028 | 0.999532 | | 2.256287 | 0.413363 | 0.799786 | | 2.260818 | -0.077 | 0.974421 |
| ITPKC | 2.725955 | 0.528193 | 0.999532 | | 2.06439 | -0.40863 | 0.799786 | | 2.325739 | -0.92936 | 0.453945 |
| VWA1 | 2.107492 | -0.00254 | 0.999532 | | 2.264563 | -0.43044 | 0.799786 | | 1.939439 | -0.39892 | 0.832794 |
| ZRANB1 | 2.271649 | -0.35781 | 0.999532 | | 2.380727 | -0.41145 | 0.799786 | | 1.81005 | -0.03829 | 0.991349 |
| P2RX4 | 3.873904 | 0.075118 | 0.999532 | | 4.905681 | 0.285522 | 0.799786 | | 4.081348 | 0.233184 | 0.86639 |
| SAMD11 | 3.692212 | 0.107085 | 0.999532 | | 3.738472 | -0.32725 | 0.799786 | | 3.153281 | -0.41301 | 0.736481 |
| MMGT1 | 4.748236 | -0.3758 | 0.999532 | | 5.499118 | -0.27401 | 0.799786 | | 4.140836 | 0.119183 | 0.939549 |
| AKNAD1 | 2.770027 | 0.513376 | 0.999532 | | 2.194624 | -0.41682 | 0.799786 | | 2.451467 | -0.90099 | 0.505689 |
| PKIA | 2.252268 | 0.222511 | 0.999532 | | 2.177092 | -0.42767 | 0.799786 | | 1.850578 | -0.62424 | 0.682165 |
| UHRF1BP | 2.169768 | 0.249051 | 0.999532 | | 2.468039 | 0.379029 | 0.799786 | | 2.336292 | 0.14748 | 0.939549 |
| TATDN3 | 2.267494 | 0.256298 | 0.999532 | | 2.502029 | 0.38073 | 0.799786 | | 2.473764 | 0.135601 | 0.945902 |
| RAB17 | 2.827565 | 0.244227 | 0.999532 | | 3.219621 | 0.363272 | 0.799786 | | 2.953168 | 0.139911 | 0.939549 |
| LRIF1 | 2.533014 | 0.359261 | 0.999532 | | 2.227738 | -0.39935 | 0.799786 | | 2.142898 | -0.74047 | 0.560207 |
| MACIR | 2.623222 | -0.22572 | 0.999532 | | 2.838991 | -0.36782 | 0.799786 | | 2.143132 | -0.12454 | 0.950751 |
| LARP7 | 4.821734 | -0.45419 | 0.999532 | | 7.284331 | 0.243348 | 0.799786 | | 5.209904 | 0.717724 | 0.37598 |
| PHF10 | 6.428536 | 0.166868 | 0.999532 | | 7.278965 | 0.246742 | 0.799786 | | 6.514722 | 0.101225 | 0.935145 |
| ZNF703 | 3.867293 | -0.19155 | 0.999532 | | 4.172293 | -0.29746 | 0.799786 | | 3.3397 | -0.08698 | 0.96129 |
| ADAM15 | 2.530997 | 0.499825 | 0.999532 | | 2.027856 | -0.44129 | 0.799786 | | 2.093132 | -0.92174 | 0.493397 |
| ARSG | 2.686289 | -0.35548 | 0.999532 | | 2.995281 | -0.36007 | 0.799786 | | 2.180713 | 0.010604 | 0.996891 |
| PRR29 | 2.685126 | 0.394389 | 0.999532 | | 2.23612 | -0.42028 | 0.799786 | | 2.31594 | -0.79961 | 0.554813 |
| SLC25A1 | 6.682333 | 0.149146 | 0.999532 | | 7.910616 | 0.259038 | 0.799786 | | 7.07672 | 0.132719 | 0.920259 |
| CD28 | 2.736074 | -0.4132 | 0.999532 | | 3.179623 | -0.37599 | 0.799858 | | 2.304364 | 0.062455 | 0.979967 |
| NUDT19 | 2.883199 | 0.449191 | 0.999532 | | 2.982436 | 0.358819 | 0.799921 | | 2.867956 | -0.07137 | 0.972758 |
| POLE4 | 4.65696 | -0.20279 | 0.999532 | | 5.021075 | -0.2858 | 0.799969 | | 4.158629 | -0.07313 | 0.967999 |
| MARK1 | 2.195353 | -0.36123 | 0.999532 | | 2.46514 | -0.41114 | 0.799969 | | 1.856633 | -0.02948 | 0.993618 |
| HTR2C | 2.336324 | 0.210286 | 0.999532 | | 2.1511 | -0.43697 | 0.799969 | | 2.005917 | -0.63345 | 0.672763 |
| FBXL15 | 2.072908 | 0.241403 | 0.999532 | | 1.821273 | -0.44635 | 0.799969 | | 1.728132 | -0.67037 | 0.67176 |
| HYAL2 | 10.57386 | -0.72661 | 0.387503 | | 13.70399 | -0.20425 | 0.799969 | | 9.13238 | 0.548953 | 0.412398 |
| CDK18 | 3.642742 | -0.21502 | 0.999532 | | 4.049183 | -0.31457 | 0.799969 | | 3.263114 | -0.08002 | 0.968434 |
| RASSF6 | 2.314023 | 0.248393 | 0.999532 | | 1.957336 | -0.46082 | 0.799969 | | 1.919482 | -0.67957 | 0.675955 |
| SGSM1 | 3.663591 | -0.15652 | 0.999532 | | 3.898746 | -0.32019 | 0.799969 | | 3.202633 | -0.15142 | 0.929529 |
| MPP6 | 2.466224 | 0.34995 | 0.999532 | | 2.637734 | 0.364503 | 0.799969 | | 2.621221 | 0.034236 | 0.990711 |
| SLC25A2 | 4.963243 | 0.39999 | 0.999532 | | 4.41353 | -0.31058 | 0.799969 | | 4.285482 | -0.68892 | 0.450657 |
| ETF1 | 6.65561 | -0.10982 | 0.999532 | | 7.199361 | -0.24757 | 0.799969 | | 5.751784 | -0.10858 | 0.939107 |
| RUNX3 | 3.534165 | -0.10672 | 0.999532 | | 3.693697 | -0.32598 | 0.799969 | | 2.940036 | -0.20058 | 0.901033 |
| MS4A7 | 2.119817 | -0.20659 | 0.999532 | | 2.806994 | 0.374997 | 0.800768 | | 2.39915 | 0.583078 | 0.702463 |
| LCMT1 | 3.11644 | -0.32375 | 0.999532 | | 3.685246 | -0.31763 | 0.800768 | | 2.768628 | 0.031488 | 0.991273 |
| PI4KB | 5.342546 | 0.116741 | 0.999532 | | 6.271877 | 0.249939 | 0.800871 | | 5.713416 | 0.15986 | 0.893262 |
| TGM4 | 2.271298 | -0.38503 | 0.999532 | | 2.777904 | -0.37988 | 0.800871 | | 2.049774 | 0.03043 | 0.992586 |
| NDUFA5 | 9.709047 | 0.018818 | 0.999532 | | 11.43567 | 0.211886 | 0.800871 | | 9.641635 | 0.217753 | 0.799841 |
| PSMD10 | 3.123155 | 0.482289 | 0.999532 | | 3.343825 | 0.330207 | 0.800871 | | 3.276019 | -0.13666 | 0.928522 |
| WDR59 | 2.234826 | -0.21709 | 0.999532 | | 2.136685 | -0.40832 | 0.800984 | | 1.823837 | -0.18074 | 0.936292 |
| P2RY4 | 2.103598 | 0.111168 | 0.999532 | | 2.479399 | 0.386466 | 0.801011 | | 2.417461 | 0.295411 | 0.869224 |
| DGCR2 | 5.32309 | 0.332956 | 0.999532 | | 5.7905 | 0.271461 | 0.801019 | | 5.353179 | -0.03874 | 0.984587 |
| CADM1 | 2.406117 | -0.1099 | 0.999532 | | 2.438021 | -0.3958 | 0.8011 | | 1.960306 | -0.26796 | 0.885892 |
| FBXO8 | 5.192529 | 0.672824 | 0.999532 | | 4.718733 | 0.286346 | 0.8011 | | 5.138833 | -0.36847 | 0.709825 |
| COMP | 2.157944 | 0.124623 | 0.999532 | | 2.039521 | -0.41816 | 0.8011 | | 1.948891 | -0.52355 | 0.749206 |
| RINL | 2.659571 | -0.24312 | 0.999532 | | 3.022826 | -0.36475 | 0.8011 | | 2.350599 | -0.09703 | 0.966817 |
| UBR7 | 3.296441 | 0.409337 | 0.999532 | | 3.412491 | 0.33647 | 0.801643 | | 3.450708 | -0.05602 | 0.978282 |
| SLC22A1 | 5.027122 | 0.121119 | 0.999532 | | 5.870838 | 0.270723 | 0.801644 | | 5.026801 | 0.173778 | 0.889853 |
| TRPM4 | 4.230044 | 0.184811 | 0.999532 | | 4.874161 | 0.2784 | 0.802046 | | 4.474941 | 0.110885 | 0.939549 |
| RSBN1L | 2.978765 | -0.40821 | 0.999532 | | 3.448756 | -0.33829 | 0.802162 | | 2.552592 | 0.091191 | 0.966643 |
| DEFB1 | 15.43534 | -0.19652 | 0.999532 | | 16.11672 | -0.50206 | 0.802162 | | 12.35345 | -0.28402 | 0.902328 |
| MON1A | 2.054383 | -0.24986 | 0.999532 | | 2.191555 | -0.40021 | 0.802162 | | 1.688253 | -0.13025 | 0.956011 |
| CRYBB3 | 3.893443 | 0.423369 | 0.999532 | | 3.480456 | -0.33757 | 0.802162 | | 3.558099 | -0.74074 | 0.49299 |
| H4-16 | 4.540625 | 0.072489 | 0.999532 | | 5.17336 | 0.263868 | 0.802162 | | 4.588889 | 0.207862 | 0.863134 |
| BTNL2 | 2.003359 | 0.112969 | 0.999532 | | 1.960994 | -0.456 | 0.802162 | | 1.744549 | -0.5497 | 0.753585 |
| GNA15 | 2.730983 | 0.082125 | 0.999532 | | 2.618579 | -0.39619 | 0.802162 | | 2.402452 | -0.47419 | 0.742171 |
| AMY1A | 11.63962 | -0.0173 | 0.999532 | | 12.34928 | -0.25293 | 0.802162 | | 10.10465 | -0.19751 | 0.84579 |
| SNX27 | 3.236154 | 0.109041 | 0.999532 | | 4.002388 | 0.301181 | 0.802162 | | 3.535231 | 0.212291 | 0.880137 |
| MRPL44 | 3.190231 | -0.10833 | 0.999532 | | 3.512395 | -0.3492 | 0.802162 | | 2.763456 | -0.22984 | 0.883785 |
| ARF6 | 13.85095 | 0.178474 | 0.999532 | | 15.56318 | 0.187073 | 0.802162 | | 14.11472 | 0.031389 | 0.979282 |
| DBP | 4.157554 | -0.01666 | 0.999532 | | 5.197371 | 0.276658 | 0.802313 | | 4.32206 | 0.310076 | 0.791576 |
| TMCC2 | 2.984116 | -0.14641 | 0.999532 | | 3.202077 | -0.34053 | 0.802313 | | 2.526515 | -0.17464 | 0.920181 |
| ME3 | 2.717539 | 0.307992 | 0.999532 | | 3.192686 | 0.341611 | 0.802365 | | 3.042853 | 0.053664 | 0.979282 |
| MYO19 | 3.132542 | -0.06335 | 0.999532 | | 3.336548 | -0.34947 | 0.802386 | | 2.81388 | -0.27192 | 0.862433 |
| FBXL19 | 4.470504 | 0.037133 | 0.999532 | | 4.587376 | -0.2938 | 0.802386 | | 3.894768 | -0.31069 | 0.797147 |
| CCAR2 | 5.629136 | -0.07786 | 0.999532 | | 7.134756 | 0.240223 | 0.802386 | | 5.638963 | 0.340156 | 0.718633 |
| MAGIX | 2.35127 | 0.238135 | 0.999532 | | 2.179447 | -0.41248 | 0.802386 | | 2.044962 | -0.63485 | 0.663633 |
| PNPLA8 | 2.519533 | 0.238182 | 0.999532 | | 2.844323 | 0.352646 | 0.802386 | | 2.721067 | 0.13122 | 0.945129 |
| SCP2 | 4.673226 | 0.348876 | 0.999532 | | 5.161129 | 0.281805 | 0.802386 | | 4.791935 | -0.04506 | 0.978487 |
| WDFY1 | 3.253417 | -0.04169 | 0.999532 | | 4.104233 | 0.303854 | 0.802386 | | 3.473092 | 0.363044 | 0.7719 |
| NDUFA13 | 6.712723 | -0.12307 | 0.999532 | | 8.785554 | 0.23219 | 0.802386 | | 7.097219 | 0.373427 | 0.666566 |
| SPDEF | 3.252504 | -0.03445 | 0.999532 | | 3.353427 | -0.40592 | 0.802386 | | 2.710372 | -0.34463 | 0.829716 |
| RANGRF | 2.114614 | 0.071716 | 0.999532 | | 2.121987 | -0.42769 | 0.802394 | | 1.862083 | -0.47701 | 0.784602 |
| TSR2 | 2.389246 | 0.730163 | 0.999532 | | 2.298702 | 0.389724 | 0.802557 | | 2.469644 | -0.32088 | 0.831749 |
| RIMKLB | 8.934701 | 0.198294 | 0.999532 | | 9.937558 | 0.21427 | 0.802561 | | 8.958761 | 0.03715 | 0.977501 |
| PMS1 | 2.515271 | 0.345959 | 0.999532 | | 2.360776 | -0.39201 | 0.80259 | | 2.247055 | -0.71624 | 0.584608 |
| BAG2 | 3.221492 | 0.21688 | 0.999532 | | 3.064 | -0.34355 | 0.802695 | | 2.813342 | -0.54698 | 0.652925 |
| RBMS1 | 10.39106 | 0.187468 | 0.999532 | | 11.71103 | 0.215759 | 0.802768 | | 10.35211 | 0.056336 | 0.966672 |
| MAP3K3 | 7.969214 | 0.210354 | 0.999532 | | 8.535344 | 0.22345 | 0.802768 | | 8.022475 | 0.014148 | 0.995112 |
| SNX10 | 2.148407 | 0.825171 | 0.999532 | | 2.211282 | 0.424208 | 0.802768 | | 2.367585 | -0.37785 | 0.810208 |
| JPH4 | 4.755256 | 0.10931 | 0.999532 | | 5.796585 | 0.266401 | 0.802768 | | 5.083505 | 0.188232 | 0.887555 |
| LCN12 | 3.701774 | 0.204378 | 0.999532 | | 3.469414 | -0.32751 | 0.802768 | | 3.040401 | -0.51166 | 0.683089 |
| NISCH | 4.304769 | -0.09494 | 0.999532 | | 4.463079 | -0.29351 | 0.802768 | | 3.645149 | -0.18213 | 0.897958 |
| MAST3 | 3.473047 | 0.683773 | 0.999532 | | 3.438434 | 0.341848 | 0.803011 | | 3.624616 | -0.3241 | 0.806394 |
| OGFOD2 | 1.958737 | -0.03007 | 0.999532 | | 2.044491 | -0.42189 | 0.803011 | | 1.7105 | -0.3711 | 0.843564 |
| SRSF11 | 13.55058 | 0.103718 | 0.999532 | | 15.87698 | 0.190827 | 0.803011 | | 13.8922 | 0.10704 | 0.906962 |
| TSSC4 | 3.483136 | 0.198489 | 0.999532 | | 3.329462 | -0.36124 | 0.803011 | | 3.033344 | -0.54172 | 0.665886 |
| LPL | 2.290831 | 0.599585 | 0.999532 | | 2.384011 | 0.403097 | 0.803011 | | 2.507881 | -0.18074 | 0.920124 |
| OPN1MW | 2.127196 | 0.394229 | 0.999532 | | 1.90501 | -0.43845 | 0.803051 | | 1.81998 | -0.81227 | 0.570375 |
| RALY | 6.84592 | -0.06273 | 0.999532 | | 8.393963 | 0.228086 | 0.803051 | | 7.035525 | 0.308407 | 0.738702 |
| CIITA | 10.11439 | -0.18072 | 0.999532 | | 13.27113 | 0.231837 | 0.803051 | | 10.70899 | 0.439378 | 0.632113 |
| C8orf86 | 1.652489 | -0.14074 | 0.999532 | | 2.133307 | 0.414716 | 0.803051 | | 1.853944 | 0.574848 | 0.735469 |
| SALL3 | 3.043927 | 0.056369 | 0.999532 | | 3.171199 | -0.3498 | 0.803051 | | 2.680807 | -0.38168 | 0.792688 |
| CDC16 | 5.818787 | 0.04924 | 0.999532 | | 7.419893 | 0.250474 | 0.803051 | | 6.158004 | 0.226093 | 0.836935 |
| KLHL29 | 3.015459 | 0.075273 | 0.999532 | | 2.777679 | -0.36229 | 0.803051 | | 2.585106 | -0.42311 | 0.791466 |
| SMARCD | 6.031271 | 0.372582 | 0.999532 | | 5.437628 | -0.26468 | 0.803125 | | 5.259756 | -0.61887 | 0.423641 |
| CMTM5 | 2.367271 | -0.10132 | 0.999532 | | 2.361032 | -0.39688 | 0.803125 | | 2.010352 | -0.28268 | 0.881837 |
| NCOA7 | 4.447262 | -0.14939 | 0.999532 | | 5.768682 | 0.270181 | 0.803125 | | 4.810774 | 0.436296 | 0.678411 |
| TIMM9 | 3.088309 | 0.325531 | 0.999532 | | 3.499158 | 0.331419 | 0.803125 | | 3.298064 | 0.026908 | 0.992286 |
| QTRT2 | 2.69982 | 0.358435 | 0.999532 | | 2.359242 | -0.39023 | 0.803125 | | 2.298331 | -0.72989 | 0.563455 |
| RFLNB | 4.828281 | -0.72913 | 0.999532 | | 6.214642 | -0.27277 | 0.803125 | | 4.176249 | 0.473962 | 0.659727 |
| H2AC11 | 2.552155 | -0.70569 | 0.999532 | | 3.014813 | -0.35106 | 0.803125 | | 2.16698 | 0.370799 | 0.82512 |
| IER2 | 10.83668 | -0.71675 | 0.950261 | | 13.9763 | -0.24976 | 0.803125 | | 8.884231 | 0.518042 | 0.515762 |
| GIMAP8 | 8.636643 | -0.22529 | 0.999532 | | 11.01291 | 0.198725 | 0.803125 | | 9.131911 | 0.446611 | 0.515762 |
| TMEM150 | 2.049728 | -0.73635 | 0.999532 | | 2.568926 | -0.3867 | 0.803125 | | 1.825241 | 0.369535 | 0.854533 |
| N6AMT1 | 4.270531 | 0.066954 | 0.999532 | | 4.306243 | -0.30059 | 0.803125 | | 3.784387 | -0.34776 | 0.774747 |
| INHA | 2.246945 | 0.698196 | 0.999532 | | 2.415251 | 0.380341 | 0.803125 | | 2.368222 | -0.30276 | 0.849128 |
| HSD17B1 | 4.457155 | -0.09229 | 0.999532 | | 6.094062 | 0.250587 | 0.803125 | | 4.759818 | 0.363769 | 0.718643 |
| CCDC177 | 2.436498 | 0.399132 | 0.999532 | | 2.075431 | -0.42138 | 0.803125 | | 2.18636 | -0.80641 | 0.560424 |
| PEX12 | 2.321164 | 0.315889 | 0.999532 | | 2.341155 | 0.372893 | 0.803125 | | 2.439933 | 0.068064 | 0.976783 |
| ARPC5 | 8.636304 | -0.22898 | 0.999532 | | 11.23998 | 0.213609 | 0.803125 | | 8.998298 | 0.46016 | 0.515762 |
| TALDO1 | 4.864887 | -0.07179 | 0.999532 | | 5.864291 | 0.253368 | 0.803125 | | 5.026789 | 0.339654 | 0.737013 |
| TMEM9 | 4.533295 | 0.067595 | 0.999532 | | 5.508908 | 0.285035 | 0.803125 | | 4.934807 | 0.230591 | 0.849179 |
| SF3A1 | 4.507138 | 0.213898 | 0.999532 | | 4.955952 | 0.282816 | 0.803125 | | 4.637057 | 0.093851 | 0.947515 |
| ACY1 | 4.172559 | 0.162402 | 0.999532 | | 4.815526 | 0.291618 | 0.803125 | | 4.328407 | 0.147277 | 0.918172 |
| TEKT2 | 2.351743 | 0.130597 | 0.999532 | | 2.318978 | -0.41406 | 0.803125 | | 2.13794 | -0.52863 | 0.735469 |
| LRRC59 | 3.795521 | -0.27542 | 0.999532 | | 4.375961 | -0.30175 | 0.803125 | | 3.374668 | -0.00224 | 0.999385 |
| KLHL12 | 3.026809 | 0.165053 | 0.999532 | | 3.482577 | 0.322263 | 0.803125 | | 3.286982 | 0.174812 | 0.912867 |
| TMEM27 | 2.457154 | -0.38415 | 0.999532 | | 2.786972 | -0.41016 | 0.803125 | | 2.038773 | -0.00614 | 0.998585 |
| USE1 | 3.730565 | 0.053998 | 0.999532 | | 3.82565 | -0.32248 | 0.803125 | | 3.132468 | -0.35812 | 0.78258 |
| F12 | 4.311295 | -0.17405 | 0.999532 | | 4.557225 | -0.32495 | 0.803207 | | 3.71393 | -0.12633 | 0.94196 |
| KRT13 | 2.338657 | 0.036768 | 0.999532 | | 2.378896 | -0.41145 | 0.803207 | | 2.127643 | -0.43057 | 0.803493 |
| IGFBP6 | 2.736902 | 0.221306 | 0.999532 | | 2.421291 | -0.38468 | 0.803207 | | 2.320534 | -0.5933 | 0.670068 |
| ADA2 | 5.028919 | 0.147525 | 0.999532 | | 5.752296 | 0.308979 | 0.803395 | | 5.250312 | 0.176327 | 0.904721 |
| KCNT1 | 2.114521 | -0.0784 | 0.999532 | | 2.137974 | -0.4365 | 0.803395 | | 1.858702 | -0.3367 | 0.865363 |
| COG8 | 3.316163 | -0.05268 | 0.999532 | | 4.101559 | 0.296436 | 0.803395 | | 3.405988 | 0.368052 | 0.75311 |
| TTF1 | 3.732027 | -0.1496 | 0.999532 | | 4.256457 | -0.3041 | 0.803455 | | 3.22964 | -0.12756 | 0.939549 |
| HLA-DRA | 55.26556 | -0.2063 | 0.999532 | | 73.15115 | 0.27543 | 0.803455 | | 58.86034 | 0.500836 | 0.533876 |
| NSDHL | 2.268414 | -0.12644 | 0.999532 | | 2.97221 | 0.345537 | 0.803455 | | 2.507954 | 0.487455 | 0.723933 |
| FAM169B | 2.202174 | -0.34764 | 0.999532 | | 2.376984 | -0.39271 | 0.803507 | | 1.876099 | -0.02895 | 0.993618 |
| TNK2 | 4.533596 | 0.441497 | 0.999532 | | 4.721741 | 0.301695 | 0.803833 | | 4.924052 | -0.12944 | 0.923864 |
| GPN2 | 2.643753 | 0.1089 | 0.999532 | | 3.033869 | 0.331315 | 0.803833 | | 2.736033 | 0.22715 | 0.88885 |
| UAP1L1 | 2.580096 | -0.05192 | 0.999532 | | 3.365596 | 0.357784 | 0.80391 | | 2.800846 | 0.430648 | 0.759369 |
| PACS1 | 5.110699 | 0.010346 | 0.999532 | | 6.431431 | 0.255787 | 0.80391 | | 5.550731 | 0.260297 | 0.800239 |
| KDM7A | 3.600001 | -0.20081 | 0.999532 | | 4.944741 | 0.288586 | 0.80391 | | 3.923402 | 0.511972 | 0.660958 |
| SPTLC3 | 2.285768 | 0.302268 | 0.999532 | | 2.506863 | 0.379977 | 0.803941 | | 2.31819 | 0.096061 | 0.965649 |
| EAPP | 5.169112 | 0.146139 | 0.999532 | | 6.094029 | 0.278715 | 0.803941 | | 5.448976 | 0.151809 | 0.901033 |
| VPS39 | 4.239153 | 0.032322 | 0.999532 | | 4.988471 | 0.270135 | 0.803941 | | 4.430967 | 0.243123 | 0.849885 |
| BLOC1S3 | 2.110948 | -0.60305 | 0.999532 | | 2.635815 | -0.38066 | 0.803941 | | 1.86687 | 0.238343 | 0.903339 |
| TBK1 | 3.424966 | -0.56242 | 0.999532 | | 4.30557 | -0.3032 | 0.803956 | | 3.04831 | 0.273883 | 0.849223 |
| PPP1R37 | 4.011885 | 0.498205 | 0.999532 | | 4.210576 | 0.309663 | 0.803956 | | 4.164717 | -0.16437 | 0.904801 |
| LST1 | 3.921221 | -0.39919 | 0.999532 | | 4.171531 | -0.30797 | 0.803956 | | 3.337281 | 0.088497 | 0.965649 |
| SPINT1 | 2.333931 | 0.065003 | 0.999532 | | 2.922243 | 0.330784 | 0.803956 | | 2.515058 | 0.283315 | 0.853034 |
| MINPP1 | 2.636636 | -0.00367 | 0.999532 | | 3.312914 | 0.344898 | 0.803956 | | 2.78931 | 0.366747 | 0.798849 |
| SHE | 5.441183 | 0.235979 | 0.999532 | | 6.064162 | 0.257347 | 0.803956 | | 5.619196 | 0.041463 | 0.978931 |
| SH3D21 | 3.66485 | 0.598956 | 0.999532 | | 3.645549 | 0.334474 | 0.803956 | | 3.781473 | -0.24494 | 0.864245 |
| S1PR5 | 1.989533 | 0.333601 | 0.999532 | | 1.695018 | -0.44835 | 0.803974 | | 1.721899 | -0.76405 | 0.636913 |
| GAS1 | 11.93031 | 0.188346 | 0.999532 | | 12.033 | -0.20511 | 0.80424 | | 10.49649 | -0.40042 | 0.600291 |
| BCL7B | 4.732552 | -0.67969 | 0.999532 | | 5.978539 | -0.264 | 0.804385 | | 3.912997 | 0.437649 | 0.696082 |
| PRDM9 | 2.087217 | -0.22979 | 0.999532 | | 2.116598 | -0.414 | 0.804385 | | 1.7614 | -0.17008 | 0.942695 |
| ENHO | 4.484038 | -0.15107 | 0.999532 | | 4.965496 | -0.30263 | 0.804385 | | 4.066393 | -0.12727 | 0.939913 |
| ZNF536 | 1.87595 | 0.865451 | 0.999532 | | 1.914369 | 0.434724 | 0.804385 | | 2.011521 | -0.41321 | 0.796512 |
| ZNF202 | 2.533153 | 0.326927 | 0.999532 | | 2.369195 | -0.39691 | 0.804385 | | 2.308087 | -0.70409 | 0.614824 |
| RPS21 | 14.24858 | -0.09268 | 0.999532 | | 17.27209 | 0.181617 | 0.804385 | | 14.36538 | 0.288993 | 0.673465 |
| TFEB | 2.068024 | -0.03153 | 0.999532 | | 2.080297 | -0.41904 | 0.80442 | | 1.752342 | -0.37078 | 0.839261 |
| TEX22 | 3.81942 | 0.46694 | 0.999532 | | 3.18183 | -0.3624 | 0.804427 | | 3.423331 | -0.81572 | 0.511836 |
| MRPL1 | 2.299591 | 0.614683 | 0.999532 | | 2.196942 | 0.387251 | 0.804447 | | 2.337796 | -0.2131 | 0.902619 |
| RASSF1 | 5.866906 | -0.37099 | 0.999532 | | 6.963306 | -0.23838 | 0.804447 | | 5.051387 | 0.153221 | 0.901033 |
| SNX22 | 2.010167 | -0.07001 | 0.999532 | | 2.855794 | 0.368734 | 0.804447 | | 2.35232 | 0.466856 | 0.762924 |
| DHRS11 | 2.803896 | 0.060247 | 0.999532 | | 2.866657 | -0.38549 | 0.804447 | | 2.511399 | -0.42363 | 0.792484 |
| ZNF780B | 2.925009 | 0.315536 | 0.999532 | | 3.17321 | 0.339169 | 0.804447 | | 3.117669 | 0.03759 | 0.988853 |
| WDR86 | 4.39261 | 0.087319 | 0.999532 | | 4.096946 | -0.33895 | 0.804447 | | 3.896071 | -0.41507 | 0.740755 |
| PTGES2 | 2.414239 | 0.437681 | 0.999532 | | 2.666057 | 0.370086 | 0.804447 | | 2.577693 | -0.04845 | 0.98514 |
| NPL | 1.724573 | 0.504217 | 0.999532 | | 1.78949 | 0.435095 | 0.804447 | | 1.868533 | -0.04933 | 0.987608 |
| ERMAP | 2.324561 | 0.403716 | 0.999532 | | 2.675942 | 0.360494 | 0.804735 | | 2.633947 | -0.02061 | 0.995256 |
| CPQ | 5.465667 | 0.028655 | 0.999532 | | 6.480636 | 0.274977 | 0.805218 | | 5.44951 | 0.282449 | 0.792688 |
| NKAIN3 | 1.865728 | -0.00211 | 0.999532 | | 2.605611 | 0.389566 | 0.805237 | | 2.205962 | 0.41981 | 0.804594 |
| ITGA6 | 2.533799 | 0.48384 | 0.999532 | | 2.78523 | 0.371861 | 0.805374 | | 2.664407 | -0.08219 | 0.971732 |
| RNF223 | 2.322102 | 0.020915 | 0.999532 | | 2.191838 | -0.40456 | 0.805435 | | 1.979671 | -0.40842 | 0.814402 |
| KHNYN | 4.281227 | -0.04423 | 0.999532 | | 5.31654 | 0.267911 | 0.805435 | | 4.444762 | 0.331214 | 0.755206 |
| OR4C13 | 2.730745 | -0.01479 | 0.999532 | | 3.041237 | -0.36592 | 0.805435 | | 2.446952 | -0.31736 | 0.851503 |
| RLF | 3.571018 | -0.54662 | 0.999532 | | 4.278101 | -0.30916 | 0.805435 | | 3.076814 | 0.258747 | 0.864245 |
| RASSF5 | 4.544131 | 0.177984 | 0.999532 | | 4.284703 | -0.29775 | 0.805435 | | 4.005939 | -0.46819 | 0.692072 |
| HERC6 | 2.413709 | 0.594036 | 0.999532 | | 2.615838 | 0.373875 | 0.805435 | | 2.694319 | -0.20113 | 0.903935 |
| PPP1R13 | 6.365576 | -0.15646 | 0.999532 | | 8.434509 | 0.225516 | 0.805435 | | 6.93922 | 0.407441 | 0.647218 |
| JTB | 7.161736 | 0.151356 | 0.999532 | | 8.44034 | 0.215513 | 0.805435 | | 7.279913 | 0.078768 | 0.946808 |
| TOMM5 | 4.120454 | -0.03364 | 0.999532 | | 4.331324 | -0.30804 | 0.805435 | | 3.416089 | -0.25525 | 0.855088 |
| ARHGDIG | 2.799744 | 0.1843 | 0.999532 | | 2.476417 | -0.37484 | 0.805435 | | 2.318862 | -0.54838 | 0.702463 |
| FOXN2 | 4.402073 | 0.111704 | 0.999532 | | 5.288917 | 0.266044 | 0.805533 | | 4.589265 | 0.172436 | 0.892182 |
| FGF2 | 3.703416 | -0.4267 | 0.999532 | | 4.278108 | -0.30126 | 0.805535 | | 3.22906 | 0.144022 | 0.929529 |
| GJB4 | 2.117192 | -0.34948 | 0.999532 | | 2.273008 | -0.39393 | 0.805535 | | 1.806033 | -0.02944 | 0.993618 |
| PACS2 | 4.590252 | 0.09902 | 0.999532 | | 4.747299 | -0.27792 | 0.805535 | | 4.169863 | -0.35311 | 0.75038 |
| HAUS6 | 3.107442 | 0.395954 | 0.999532 | | 3.453808 | 0.324974 | 0.805552 | | 3.221412 | -0.06017 | 0.975445 |
| ZNF658 | 2.839444 | 0.02584 | 0.999532 | | 2.671153 | -0.36499 | 0.805552 | | 2.383922 | -0.3746 | 0.813143 |
| MPST | 5.426489 | 0.254617 | 0.999532 | | 5.980683 | 0.272563 | 0.805552 | | 5.850664 | 0.048853 | 0.979248 |
| CCDC159 | 4.701465 | -0.57574 | 0.999532 | | 5.842352 | -0.26652 | 0.805715 | | 4.211268 | 0.32685 | 0.78258 |
| LGR6 | 1.880233 | -0.33581 | 0.999532 | | 2.03827 | -0.40697 | 0.805849 | | 1.629496 | -0.05221 | 0.988079 |
| MPP7 | 2.241758 | 0.706468 | 0.999532 | | 2.33449 | 0.39465 | 0.806023 | | 2.343964 | -0.29568 | 0.858705 |
| GLT8D2 | 3.093088 | -0.0551 | 0.999532 | | 3.388438 | -0.32885 | 0.806043 | | 2.767325 | -0.23881 | 0.882936 |
| SHARPIN | 4.840692 | 0.085341 | 0.999532 | | 4.755977 | -0.29761 | 0.806043 | | 4.142009 | -0.36057 | 0.769663 |
| VANGL2 | 5.098476 | -0.0717 | 0.999532 | | 5.17032 | -0.27355 | 0.806043 | | 4.322917 | -0.18575 | 0.88956 |
| TRAK1 | 5.458217 | 0.015413 | 0.999532 | | 6.762385 | 0.287515 | 0.806227 | | 5.601728 | 0.308533 | 0.791466 |
| NEK1 | 2.757951 | 0.269546 | 0.999532 | | 2.383255 | -0.38177 | 0.806227 | | 2.353807 | -0.63645 | 0.647581 |
| ASF1B | 2.658652 | -0.20578 | 0.999532 | | 2.705866 | -0.36535 | 0.806227 | | 2.266964 | -0.14268 | 0.945902 |
| ANKRD42 | 2.451765 | -0.024 | 0.999532 | | 2.358125 | -0.37278 | 0.806227 | | 2.045942 | -0.33174 | 0.843564 |
| SLC37A4 | 3.937378 | 0.031539 | 0.999532 | | 4.571126 | 0.283945 | 0.806227 | | 4.032608 | 0.26719 | 0.833941 |
| SUMF1 | 3.873524 | 0.206312 | 0.999532 | | 4.394548 | 0.289286 | 0.806227 | | 3.973809 | 0.102046 | 0.946765 |
| INO80D | 3.963165 | -0.30084 | 0.999532 | | 5.519785 | 0.283298 | 0.806227 | | 4.462729 | 0.601911 | 0.560424 |
| ITCH | 12.6868 | 0.187459 | 0.999532 | | 14.00118 | 0.187358 | 0.806227 | | 12.7274 | 0.021059 | 0.988853 |
| AP2S1 | 4.172126 | -0.189 | 0.999532 | | 5.676378 | 0.26155 | 0.806227 | | 4.568663 | 0.473872 | 0.649779 |
| PPP1R35 | 4.665945 | 0.023617 | 0.999532 | | 5.787857 | 0.266253 | 0.806227 | | 4.947766 | 0.265626 | 0.816618 |
| UQCR10 | 3.946371 | 0.198983 | 0.999532 | | 4.508681 | 0.31849 | 0.806227 | | 4.045091 | 0.136984 | 0.93603 |
| MAPK4 | 2.647038 | 0.031521 | 0.999532 | | 2.484571 | -0.38398 | 0.806227 | | 2.363137 | -0.39647 | 0.806394 |
| SHC2 | 1.921911 | 0.876434 | 0.999532 | | 1.807459 | 0.429143 | 0.806227 | | 2.049339 | -0.42936 | 0.795612 |
| UGT8 | 2.480758 | 0.578018 | 0.999532 | | 1.970456 | -0.42824 | 0.806227 | | 2.036216 | -0.9837 | 0.445661 |
| OVOL3 | 2.340243 | -0.00304 | 0.999532 | | 2.458721 | -0.41111 | 0.806227 | | 2.057233 | -0.39254 | 0.823571 |
| KCNE1 | 5.442397 | -0.00834 | 0.999532 | | 5.77334 | -0.26376 | 0.806227 | | 4.933125 | -0.23332 | 0.84694 |
| GPM6B | 2.750382 | -0.70693 | 0.999532 | | 3.402534 | -0.34414 | 0.806276 | | 2.490951 | 0.369855 | 0.827119 |
| TMA7 | 83.23514 | -0.07977 | 0.999532 | | 103.6191 | 0.19763 | 0.806276 | | 86.22772 | 0.304196 | 0.647218 |
| TAOK1 | 9.899449 | -0.15837 | 0.999532 | | 12.1889 | 0.195573 | 0.8063 | | 10.27918 | 0.367914 | 0.623565 |
| FTMT | 2.738901 | -0.02547 | 0.999532 | | 2.698765 | -0.36795 | 0.8063 | | 2.32568 | -0.33089 | 0.849682 |
| GPD1L | 3.006898 | 0.824972 | 0.999532 | | 2.884327 | 0.359086 | 0.8063 | | 3.112134 | -0.43595 | 0.728277 |
| SPX | 2.550597 | 0.534512 | 0.999532 | | 1.938819 | -0.41143 | 0.8063 | | 2.224623 | -0.92907 | 0.465298 |
| TPP1 | 12.53709 | 0.004847 | 0.999532 | | 14.93093 | 0.171317 | 0.8063 | | 12.40064 | 0.181678 | 0.8233 |
| GNRH1 | 2.803751 | -0.06109 | 0.999532 | | 2.84553 | -0.35108 | 0.8063 | | 2.438674 | -0.27403 | 0.870247 |
| MAPKAP | 3.340036 | 0.009685 | 0.999532 | | 3.21879 | -0.33027 | 0.8063 | | 2.801632 | -0.32013 | 0.817974 |
| C4orf17 | 2.347712 | -0.02446 | 0.999532 | | 2.240873 | -0.39798 | 0.8063 | | 2.062711 | -0.35738 | 0.844187 |
| NFKB1 | 5.806687 | -0.20383 | 0.999532 | | 6.076896 | -0.25909 | 0.8063 | | 5.155806 | -0.02552 | 0.991783 |
| NPTX2 | 1.854613 | 0.049838 | 0.999532 | | 1.846953 | -0.4232 | 0.8063 | | 1.668477 | -0.45381 | 0.800239 |
| POMGNT | 2.957847 | -0.24498 | 0.999532 | | 2.965566 | -0.34355 | 0.8063 | | 2.607799 | -0.08445 | 0.971732 |
| PIAS1 | 4.212473 | -0.26941 | 0.999532 | | 5.935248 | 0.255288 | 0.8063 | | 4.486394 | 0.54079 | 0.55841 |
| ARPC3 | 14.66261 | -0.12501 | 0.999532 | | 18.72122 | 0.17639 | 0.8063 | | 15.02765 | 0.315529 | 0.632923 |
| NEDD4L | 4.097619 | 0.624474 | 0.999532 | | 3.927528 | 0.293265 | 0.8063 | | 4.194025 | -0.31383 | 0.785012 |
| ZNF100 | 2.63904 | 0.392988 | 0.999532 | | 2.374347 | -0.39238 | 0.8063 | | 2.368275 | -0.76682 | 0.551754 |
| TMEM181 | 3.258768 | 0.319121 | 0.999532 | | 3.668053 | 0.317642 | 0.8063 | | 3.2724 | 0.01513 | 0.996681 |
| PLIN2 | 4.280029 | -0.41147 | 0.999532 | | 5.862354 | 0.259612 | 0.8063 | | 4.202649 | 0.68655 | 0.486743 |
| PGAP4 | 4.046403 | 0.5619 | 0.999532 | | 3.811742 | 0.30326 | 0.8063 | | 4.100274 | -0.23882 | 0.86257 |
| TAS2R20 | 2.301777 | 0.032742 | 0.999532 | | 2.772443 | 0.346876 | 0.8063 | | 2.510995 | 0.329399 | 0.836935 |
| TIMM50 | 3.222242 | 0.108509 | 0.999532 | | 3.989717 | 0.296897 | 0.8063 | | 3.520234 | 0.209184 | 0.883565 |
| PRR16 | 2.890175 | -0.31585 | 0.999532 | | 3.094805 | -0.33507 | 0.8063 | | 2.356496 | -0.01208 | 0.996891 |
| ARMC2 | 3.006945 | 0.100883 | 0.999532 | | 2.959654 | -0.35442 | 0.8063 | | 2.618612 | -0.43239 | 0.755977 |
| SBF2 | 4.559038 | 0.480068 | 0.999532 | | 4.723582 | 0.286749 | 0.8063 | | 4.667864 | -0.1727 | 0.893259 |
| TTC38 | 2.629349 | 0.024922 | 0.999532 | | 3.194207 | 0.342305 | 0.8063 | | 2.705194 | 0.337446 | 0.816526 |
| FDPS | 3.856208 | -0.57649 | 0.999532 | | 4.66855 | -0.27984 | 0.806759 | | 3.322203 | 0.311471 | 0.818726 |
| CLPB | 3.874741 | -0.58043 | 0.999532 | | 4.71699 | -0.29096 | 0.806759 | | 3.234648 | 0.307943 | 0.816618 |
| TWF2 | 2.453842 | -0.04206 | 0.999532 | | 2.471235 | -0.37637 | 0.807352 | | 2.065807 | -0.31477 | 0.863401 |
| HDDC2 | 3.461452 | -0.05987 | 0.999532 | | 3.68511 | -0.32277 | 0.80739 | | 2.903632 | -0.2399 | 0.876613 |
| ARMC8 | 3.727728 | 0.062227 | 0.999532 | | 3.719247 | -0.3113 | 0.807451 | | 3.273832 | -0.35426 | 0.791576 |
| SPACA4 | 2.473374 | -0.59237 | 0.999532 | | 2.9659 | -0.3658 | 0.807501 | | 2.148265 | 0.237424 | 0.903329 |
| BAZ1B | 4.596561 | 0.372009 | 0.999532 | | 4.779137 | 0.281937 | 0.807662 | | 4.759031 | -0.07702 | 0.962725 |
| VPS54 | 4.384273 | -0.021 | 0.999532 | | 4.324489 | -0.29108 | 0.807682 | | 3.616182 | -0.25386 | 0.850724 |
| GPR146 | 4.620217 | -0.03331 | 0.999532 | | 4.870294 | -0.27888 | 0.807808 | | 3.83258 | -0.24973 | 0.850928 |
| NSMCE1 | 5.79476 | -0.1576 | 0.999532 | | 7.362609 | 0.230154 | 0.807808 | | 5.909809 | 0.405106 | 0.663089 |
| DNASE2 | 2.818635 | -0.05955 | 0.999532 | | 3.817029 | 0.309794 | 0.807808 | | 3.084353 | 0.391456 | 0.76172 |
| CHDH | 3.599222 | 0.148374 | 0.999532 | | 4.358988 | 0.289476 | 0.807808 | | 3.644994 | 0.157225 | 0.918675 |
| OR2L8 | 2.388179 | -0.26384 | 0.999532 | | 2.684457 | -0.39279 | 0.807808 | | 2.284161 | -0.1058 | 0.968434 |
| ATP6V1A | 6.824307 | 0.155712 | 0.999532 | | 7.803767 | 0.227491 | 0.807808 | | 6.967549 | 0.091448 | 0.942503 |
| SYNPO2 | 2.700163 | -0.44831 | 0.999532 | | 3.077695 | -0.34986 | 0.807808 | | 2.230883 | 0.117961 | 0.956011 |
| PPP2R2B | 3.795476 | 0.570178 | 0.999532 | | 3.05737 | -0.37776 | 0.807808 | | 3.1496 | -0.91156 | 0.387954 |
| TRIM7 | 2.760072 | 0.470286 | 0.999532 | | 2.159179 | -0.39686 | 0.807808 | | 2.435644 | -0.83961 | 0.515762 |
| VPS4B | 5.709877 | -0.15905 | 0.999532 | | 7.117146 | 0.228944 | 0.807808 | | 5.783605 | 0.403595 | 0.658715 |
| OR11A1 | 2.101854 | 0.398115 | 0.999532 | | 1.77928 | -0.45758 | 0.807808 | | 1.740212 | -0.83682 | 0.587597 |
| RIN2 | 9.496788 | 0.073719 | 0.999532 | | 10.98462 | 0.217403 | 0.807808 | | 9.644164 | 0.170045 | 0.859426 |
| ADARB1 | 3.37761 | 0.05961 | 0.999532 | | 4.237304 | 0.295897 | 0.807808 | | 3.658061 | 0.258036 | 0.857377 |
| ERCC2 | 2.790848 | 0.27719 | 0.999532 | | 3.026814 | 0.342131 | 0.807808 | | 3.072457 | 0.084778 | 0.970457 |
| CARTPT | 2.64858 | 0.072972 | 0.999532 | | 2.634968 | -0.3869 | 0.807841 | | 2.379633 | -0.43968 | 0.788938 |
| KCNA4 | 2.856361 | 0.093933 | 0.999532 | | 2.659567 | -0.3608 | 0.807951 | | 2.493605 | -0.43615 | 0.783005 |
| ECE1 | 6.141922 | 0.004187 | 0.999532 | | 7.792459 | 0.303519 | 0.807951 | | 6.70225 | 0.308189 | 0.771812 |
| CKMT1A | 4.005581 | -0.50016 | 0.999532 | | 4.914314 | -0.35194 | 0.808174 | | 3.574118 | 0.163831 | 0.93222 |
| C11orf42 | 2.039765 | 0.508976 | 0.999532 | | 1.774208 | -0.4422 | 0.808174 | | 1.806668 | -0.93186 | 0.515762 |
| PIGV | 3.517825 | -0.01484 | 0.999532 | | 3.708461 | -0.33253 | 0.808174 | | 3.113623 | -0.29721 | 0.843564 |
| ATAD3A | 10.79473 | 0.106315 | 0.999532 | | 12.11688 | 0.190811 | 0.808174 | | 10.70643 | 0.100953 | 0.914929 |
| KRT10 | 2.220957 | -0.34618 | 0.999532 | | 2.52881 | -0.37339 | 0.808174 | | 1.985865 | -0.00733 | 0.998094 |
| MED6 | 3.450754 | -0.00701 | 0.999532 | | 4.120206 | 0.299911 | 0.808174 | | 3.603384 | 0.314781 | 0.81294 |
| POLRMT | 7.565224 | 0.303093 | 0.999532 | | 7.089621 | -0.25269 | 0.808203 | | 6.669468 | -0.53529 | 0.52568 |
| PGLYRP | 2.406924 | 0.154762 | 0.999532 | | 2.442764 | -0.38741 | 0.808203 | | 2.181741 | -0.51708 | 0.735469 |
| CCDC154 | 2.034532 | 0.395663 | 0.999532 | | 2.34721 | 0.384671 | 0.808203 | | 2.267614 | 0.011658 | 0.996891 |
| NUDT9 | 2.571741 | 0.034699 | 0.999532 | | 3.120547 | 0.325384 | 0.808252 | | 2.708549 | 0.310658 | 0.843564 |
| PTCH2 | 2.249674 | -0.35478 | 0.999532 | | 2.591234 | -0.37865 | 0.808252 | | 2.001692 | -0.0118 | 0.996891 |
| CEBPG | 3.583369 | -0.17488 | 0.999532 | | 4.846165 | 0.283275 | 0.808252 | | 3.777507 | 0.481161 | 0.668083 |
| HAGH | 4.222785 | 0.048067 | 0.999532 | | 4.950703 | 0.273873 | 0.808562 | | 4.317062 | 0.242989 | 0.846786 |
| ABCA7 | 2.225124 | -0.31494 | 0.999532 | | 2.336818 | -0.38272 | 0.808562 | | 1.93081 | -0.05631 | 0.985693 |
| ASPN | 1.867098 | 0.64414 | 0.999532 | | 1.897795 | 0.425346 | 0.808562 | | 1.965901 | -0.20096 | 0.918694 |
| RABAC1 | 13.88441 | -0.16772 | 0.999532 | | 17.08184 | 0.161204 | 0.809119 | | 13.89884 | 0.338426 | 0.595502 |
| GPR37L1 | 2.087676 | 0.23623 | 0.999532 | | 1.957198 | -0.43143 | 0.809119 | | 1.783784 | -0.64824 | 0.68528 |
| FBXO10 | 3.662405 | 0.308755 | 0.999532 | | 3.389645 | -0.34018 | 0.809119 | | 3.061473 | -0.63908 | 0.560424 |
| USP3 | 2.882627 | -0.46359 | 0.999532 | | 4.031458 | 0.312744 | 0.809119 | | 3.076502 | 0.795926 | 0.49299 |
| DYNC2I2 | 3.255841 | 0.01068 | 0.999532 | | 3.324817 | -0.33344 | 0.809294 | | 2.695841 | -0.32408 | 0.818918 |
| SLC6A7 | 2.415418 | 0.096947 | 0.999532 | | 2.343045 | -0.37773 | 0.809493 | | 2.177093 | -0.46347 | 0.761337 |
| HCN3 | 2.815022 | 0.24424 | 0.999532 | | 2.419724 | -0.37915 | 0.809493 | | 2.438968 | -0.61008 | 0.666566 |
| FRRS1 | 2.898146 | 0.052279 | 0.999532 | | 2.719432 | -0.36306 | 0.809493 | | 2.373239 | -0.39859 | 0.794815 |
| PRXL2B | 6.308009 | -0.33575 | 0.999532 | | 8.492178 | 0.210684 | 0.809493 | | 6.72712 | 0.56575 | 0.465298 |
| DAPK3 | 7.124511 | -0.07198 | 0.999532 | | 8.516301 | 0.219498 | 0.809493 | | 7.247017 | 0.304075 | 0.740127 |
| PNKD | 4.671648 | 0.164436 | 0.999532 | | 4.691451 | -0.2928 | 0.809882 | | 4.27055 | -0.43531 | 0.697497 |
| NEFL | 2.234936 | -0.27248 | 0.999532 | | 2.270978 | -0.39452 | 0.809882 | | 1.813119 | -0.10569 | 0.967145 |
| ZNF729 | 2.499251 | -0.21326 | 0.999532 | | 2.605918 | -0.35758 | 0.809882 | | 2.07312 | -0.12957 | 0.950895 |
| ITPRID2 | 13.06975 | -0.13461 | 0.999532 | | 16.4659 | 0.173824 | 0.809882 | | 13.60826 | 0.330954 | 0.596335 |
| CAPN14 | 2.674114 | 0.263236 | 0.999532 | | 2.319961 | -0.39407 | 0.809882 | | 2.32658 | -0.64168 | 0.663412 |
| MASP2 | 1.875701 | 0.072645 | 0.999532 | | 1.755772 | -0.42875 | 0.810176 | | 1.593369 | -0.4849 | 0.791466 |
| APOB | 2.172747 | 0.127915 | 0.999532 | | 2.19752 | -0.41282 | 0.810176 | | 1.832163 | -0.51695 | 0.752562 |
| LCE2D | 2.075381 | 0.783566 | 0.999532 | | 2.079638 | 0.410958 | 0.810176 | | 2.273385 | -0.35494 | 0.825411 |
| HLA-B | 164.8293 | -0.1209 | 0.999532 | | 206.7711 | 0.198161 | 0.810203 | | 169.4958 | 0.341351 | 0.6215 |
| LRR1 | 2.391503 | -0.33094 | 0.999532 | | 2.667178 | -0.36824 | 0.810242 | | 2.072836 | -0.02116 | 0.996471 |
| SUB1 | 8.278396 | 0.009111 | 0.999532 | | 9.736381 | 0.202177 | 0.810305 | | 8.465114 | 0.211588 | 0.816775 |
| DDR2 | 2.017634 | -0.17964 | 0.999532 | | 2.54544 | 0.353081 | 0.810305 | | 2.138353 | 0.544835 | 0.717138 |
| PFKFB3 | 4.19392 | -0.39847 | 0.999532 | | 5.128145 | -0.29016 | 0.810305 | | 3.71072 | 0.116933 | 0.941197 |
| MAB21L3 | 2.208292 | 0.766677 | 0.999532 | | 2.367047 | 0.407117 | 0.810305 | | 2.432325 | -0.33384 | 0.837514 |
| STX16 | 3.432329 | 0.476055 | 0.999532 | | 3.492646 | 0.313093 | 0.810305 | | 3.66328 | -0.14472 | 0.921 |
| C1orf122 | 6.329414 | 0.306817 | 0.999532 | | 7.017118 | 0.24049 | 0.810305 | | 6.663919 | -0.04445 | 0.977473 |
| OR5AL1 | 1.877347 | -0.0502 | 0.999532 | | 1.79741 | -0.43159 | 0.81066 | | 1.595061 | -0.36551 | 0.859426 |
| FAM217B | 2.331808 | 0.45121 | 0.999532 | | 2.533047 | 0.377725 | 0.81066 | | 2.326562 | -0.0494 | 0.984978 |
| PTK6 | 10.7325 | 0.372409 | 0.999532 | | 9.840823 | -0.23291 | 0.81066 | | 9.735034 | -0.57825 | 0.473732 |
| KNDC1 | 2.385796 | 0.457298 | 0.999532 | | 2.155828 | -0.40752 | 0.81066 | | 2.067438 | -0.83911 | 0.533876 |
| LDLRAD2 | 1.766089 | 0.339966 | 0.999532 | | 2.120756 | 0.410062 | 0.81066 | | 1.959917 | 0.089734 | 0.972758 |
| NOTUM | 4.961297 | 0.24293 | 0.999532 | | 4.692264 | -0.31843 | 0.81066 | | 4.438286 | -0.5494 | 0.644433 |
| FAM83B | 2.060677 | 0.009342 | 0.999532 | | 2.015952 | -0.39964 | 0.810691 | | 1.737078 | -0.39165 | 0.823571 |
| KCTD21 | 2.222636 | 0.405013 | 0.999532 | | 2.419592 | 0.375691 | 0.810691 | | 2.367617 | -0.01174 | 0.996891 |
| BCL6 | 2.174209 | -0.30374 | 0.999532 | | 2.25131 | -0.39225 | 0.810691 | | 1.835454 | -0.07288 | 0.978805 |
| CAPN12 | 2.510694 | 0.302022 | 0.999532 | | 2.337012 | -0.39358 | 0.810691 | | 2.323123 | -0.66777 | 0.658332 |
| NRG1 | 3.326106 | -0.0284 | 0.999532 | | 3.318732 | -0.33125 | 0.810691 | | 2.782793 | -0.28936 | 0.859583 |
| TOX | 2.01644 | -0.43488 | 0.999532 | | 2.214697 | -0.40019 | 0.810691 | | 1.685225 | 0.050051 | 0.988853 |
| MKNK2 | 8.151342 | -0.17961 | 0.999532 | | 10.15858 | 0.202573 | 0.810782 | | 7.934473 | 0.403774 | 0.599249 |
| HHAT | 1.953431 | 0.430319 | 0.999532 | | 1.994037 | 0.411801 | 0.810948 | | 2.075654 | -0.01096 | 0.996891 |
| SNRNP35 | 3.052306 | 0.314048 | 0.999532 | | 3.383212 | 0.335231 | 0.81138 | | 3.275969 | 0.036734 | 0.988969 |
| CHD1 | 4.431517 | -0.03221 | 0.999532 | | 5.661402 | 0.263704 | 0.811459 | | 4.75144 | 0.314866 | 0.803493 |
| PAX5 | 2.171524 | 0.059513 | 0.999532 | | 2.150706 | -0.38799 | 0.811512 | | 1.828116 | -0.42834 | 0.798028 |
| ESS2 | 4.300627 | 0.332623 | 0.999532 | | 4.621666 | 0.278511 | 0.811512 | | 4.398996 | -0.03805 | 0.985675 |
| MFN1 | 3.202204 | 0.245117 | 0.999532 | | 3.64684 | 0.311064 | 0.811547 | | 3.453924 | 0.082504 | 0.96556 |
| FAHD2B | 8.230769 | 0.023176 | 0.999532 | | 8.618003 | -0.24156 | 0.811547 | | 7.363087 | -0.2566 | 0.772912 |
| EIF4B | 25.33619 | 0.081256 | 0.999532 | | 29.07328 | 0.152828 | 0.811774 | | 25.67981 | 0.092276 | 0.903659 |
| NUDT11 | 2.69665 | -0.06511 | 0.999532 | | 2.68363 | -0.38144 | 0.811884 | | 2.257585 | -0.29031 | 0.867976 |
| GSTM1 | 2.905856 | 1.091176 | 0.955244 | | 2.542873 | 0.41122 | 0.811939 | | 3.082021 | -0.6542 | 0.60629 |
| PRELID2 | 5.019031 | -0.13206 | 0.999532 | | 5.589612 | -0.26311 | 0.811939 | | 4.485694 | -0.10445 | 0.946512 |
| SLC35E2 | 4.901737 | -0.25898 | 0.999532 | | 5.528387 | -0.285 | 0.811939 | | 4.107579 | 0.00023 | 0.999949 |
| ARMC7 | 7.306167 | -0.15145 | 0.999532 | | 8.005721 | -0.24121 | 0.812183 | | 6.504408 | -0.07121 | 0.959593 |
| DDX31 | 1.761094 | 0.144581 | 0.999532 | | 2.076486 | 0.385712 | 0.812209 | | 1.965106 | 0.257179 | 0.892004 |
| PRLHR | 2.599983 | 0.024073 | 0.999532 | | 2.87128 | -0.38742 | 0.812556 | | 2.325678 | -0.3647 | 0.837256 |
| BBS7 | 2.164109 | 0.110002 | 0.999532 | | 2.705855 | 0.353573 | 0.812556 | | 2.425814 | 0.262912 | 0.87764 |
| TXNDC17 | 3.876039 | -0.08527 | 0.999532 | | 5.170506 | 0.291827 | 0.812556 | | 4.073131 | 0.39167 | 0.728015 |
| RBM4B | 3.370966 | 0.895755 | 0.999532 | | 2.928213 | 0.332316 | 0.812556 | | 3.442456 | -0.54705 | 0.631504 |
| NPIPB2 | 2.743467 | 0.027174 | 0.999532 | | 2.714462 | -0.35447 | 0.812556 | | 2.433321 | -0.36571 | 0.816775 |
| HUWE1 | 3.847138 | 0.217075 | 0.999532 | | 4.345544 | 0.291187 | 0.812556 | | 4.075426 | 0.094581 | 0.953572 |
| S100P | 2.283228 | -0.42551 | 0.999532 | | 2.699416 | -0.38534 | 0.812556 | | 1.970469 | 0.054729 | 0.985693 |
| EPHB3 | 2.99857 | 0.283989 | 0.999532 | | 2.96662 | -0.3584 | 0.812556 | | 2.780497 | -0.60384 | 0.66232 |
| RNASEH1 | 6.78122 | -0.28364 | 0.999532 | | 7.502932 | -0.24322 | 0.812556 | | 5.601737 | 0.064168 | 0.965649 |

| | | | | | | | | | | | |
|---|---|---|---|---|---|---|---|---|---|---|---|
| DLGAP3 | 5.051264 | 0.160276 | 0.999532 | | 4.926476 | -0.28761 | 0.812556 | | 4.551118 | -0.42907 | 0.709825 |
| NAXD | 3.115885 | -0.76198 | 0.999532 | | 4.003392 | -0.29543 | 0.812556 | | 2.633286 | 0.484817 | 0.709825 |
| MED29 | 5.970057 | 0.229946 | 0.999532 | | 6.577551 | 0.230896 | 0.812556 | | 5.933826 | 0.015451 | 0.994537 |
| SF3B4 | 2.572255 | -0.11654 | 0.999532 | | 3.501079 | 0.33232 | 0.812556 | | 2.741141 | 0.47013 | 0.735469 |
| TRIM6 | 2.37285 | 0.06778 | 0.999532 | | 2.954821 | 0.353143 | 0.812556 | | 2.671769 | 0.296645 | 0.863401 |
| SBDS | 9.602173 | -0.29514 | 0.999532 | | 12.92277 | 0.178889 | 0.812556 | | 10.14688 | 0.49821 | 0.421517 |
| MRPL11 | 5.343494 | 0.129512 | 0.999532 | | 4.94836 | -0.26885 | 0.812556 | | 4.723429 | -0.37613 | 0.718643 |
| SUMO2 | 7.398795 | -0.34298 | 0.999532 | | 8.654425 | -0.20848 | 0.812556 | | 6.641832 | 0.158785 | 0.893262 |
| SLC45A2 | 2.189282 | -0.28113 | 0.999532 | | 2.441954 | -0.3979 | 0.812556 | | 1.887614 | -0.09591 | 0.972221 |
| BRIX1 | 4.761698 | 0.40483 | 0.999532 | | 5.03319 | 0.260008 | 0.812556 | | 4.74161 | -0.12898 | 0.920259 |
| PNPT1 | 2.540123 | 0.193125 | 0.999532 | | 2.903656 | 0.337541 | 0.812556 | | 2.753948 | 0.150527 | 0.935073 |
| PSENEN | 5.267022 | 0.238416 | 0.999532 | | 5.904484 | 0.246362 | 0.812556 | | 5.427441 | 0.026071 | 0.989638 |
| FGF17 | 3.854687 | 0.29848 | 0.999532 | | 4.602725 | 0.31263 | 0.812556 | | 4.30036 | 0.035803 | 0.988853 |
| CCDC124 | 6.632346 | 0.062486 | 0.999532 | | 6.880201 | -0.23692 | 0.812556 | | 5.90483 | -0.27875 | 0.78258 |
| CLCA2 | 1.741038 | 0.081456 | 0.999532 | | 2.113215 | 0.400309 | 0.812556 | | 1.867239 | 0.337022 | 0.859898 |
| TRANK1 | 3.821821 | -0.22737 | 0.999532 | | 4.085499 | -0.2984 | 0.812556 | | 3.404522 | -0.05711 | 0.977473 |
| TMEM63C | 2.160037 | -0.08907 | 0.999532 | | 2.075269 | -0.4073 | 0.812556 | | 1.799494 | -0.30629 | 0.881689 |
| MORN4 | 2.424706 | 0.362841 | 0.999532 | | 2.181299 | -0.39719 | 0.812556 | | 2.183626 | -0.7325 | 0.622543 |
| CCR4 | 2.19234 | -0.27183 | 0.999532 | | 2.310184 | -0.39613 | 0.812877 | | 1.938777 | -0.10144 | 0.970935 |
| ATP6V1G | 15.60271 | 0.172091 | 0.999532 | | 17.51371 | 0.164215 | 0.812877 | | 15.632 | 0.01292 | 0.992286 |
| DCTN6 | 4.814935 | 0.334833 | 0.999532 | | 5.368143 | 0.263257 | 0.81291 | | 5.101383 | -0.05189 | 0.974421 |
| MAP2K6 | 3.182861 | 0.270694 | 0.999532 | | 3.059649 | -0.3429 | 0.81291 | | 2.785053 | -0.58554 | 0.647218 |
| KIAA1586 | 2.649017 | 0.266594 | 0.999532 | | 3.14377 | 0.352474 | 0.81291 | | 2.816065 | 0.108365 | 0.958284 |
| P2RX3 | 2.234239 | -0.44717 | 0.999532 | | 2.515285 | -0.37601 | 0.812954 | | 1.953553 | 0.092146 | 0.972221 |
| RAB25 | 2.793449 | 0.10049 | 0.999532 | | 2.539339 | -0.37066 | 0.812954 | | 2.39592 | -0.45099 | 0.764718 |
| C6orf223 | 1.939421 | 0.068883 | 0.999532 | | 1.894149 | -0.42448 | 0.812954 | | 1.661738 | -0.4746 | 0.794815 |
| MCM9 | 2.459772 | -0.26393 | 0.999532 | | 2.668532 | -0.34857 | 0.812954 | | 2.17341 | -0.06714 | 0.978229 |
| C2CD3 | 2.51557 | 0.140366 | 0.999532 | | 2.265581 | -0.3704 | 0.812954 | | 2.202174 | -0.49426 | 0.739496 |
| CDK5RAP | 2.728109 | -0.36905 | 0.999532 | | 3.154975 | -0.3276 | 0.812954 | | 2.259205 | 0.061905 | 0.97843 |
| CSF3R | 2.673425 | -0.15131 | 0.999532 | | 2.835185 | -0.34966 | 0.812954 | | 2.166299 | -0.17862 | 0.923834 |
| CKLF | 5.876381 | -0.02244 | 0.999532 | | 6.198775 | -0.27048 | 0.812954 | | 5.100261 | -0.22004 | 0.872933 |
| HELQ | 3.795526 | 0.156719 | 0.999532 | | 4.738118 | 0.271281 | 0.812954 | | 4.065114 | 0.134389 | 0.924297 |
| RBM12B | 3.031385 | -0.06548 | 0.999532 | | 3.063371 | -0.33195 | 0.812954 | | 2.493264 | -0.24864 | 0.880375 |
| CALB1 | 4.697463 | -0.58885 | 0.999532 | | 5.332621 | -0.51661 | 0.812954 | | 3.557361 | 0.19472 | 0.900848 |
| OGFRL1 | 2.933644 | 0.342331 | 0.999532 | | 3.267803 | 0.324452 | 0.812954 | | 3.099536 | 0.001222 | 0.999573 |
| TBC1D23 | 3.450779 | -0.31652 | 0.999532 | | 3.726373 | -0.32602 | 0.812954 | | 2.809555 | 0.01184 | 0.996891 |
| TENT5A | 2.622703 | 0.392117 | 0.999532 | | 2.222885 | -0.39075 | 0.812954 | | 2.347461 | -0.76837 | 0.563455 |
| C18orf21 | 2.984293 | 0.681756 | 0.999532 | | 2.981834 | 0.349619 | 0.812954 | | 3.303458 | -0.31638 | 0.827625 |
| EMD | 4.260589 | -0.21198 | 0.999532 | | 5.530457 | 0.245438 | 0.812954 | | 4.379784 | 0.472613 | 0.637179 |
| ONECUT2 | 3.122032 | -0.18062 | 0.999532 | | 3.254429 | -0.38133 | 0.812954 | | 2.674803 | -0.18744 | 0.924131 |
| EDDM3A | 2.024109 | -0.22757 | 0.999532 | | 2.135893 | -0.4023 | 0.812954 | | 1.766701 | -0.15294 | 0.948127 |
| SFTPC | 2.878601 | 0.154808 | 0.999532 | | 2.781743 | -0.37029 | 0.812954 | | 2.550541 | -0.50789 | 0.728015 |
| TTC39B | 3.456967 | -0.04106 | 0.999532 | | 3.492193 | -0.31342 | 0.812954 | | 2.967774 | -0.25685 | 0.868465 |
| GBX2 | 3.209471 | -0.11849 | 0.999532 | | 3.1855 | -0.33254 | 0.812954 | | 2.706266 | -0.20438 | 0.907568 |
| DVL3 | 7.679459 | 0.375619 | 0.999532 | | 7.892158 | 0.223229 | 0.812954 | | 7.809589 | -0.13365 | 0.900848 |
| PAX6 | 2.346527 | 0.298713 | 0.999532 | | 2.144304 | -0.41165 | 0.812954 | | 2.236776 | -0.70241 | 0.639801 |
| MANBAL | 4.384351 | -0.0272 | 0.999532 | | 4.177968 | -0.28824 | 0.813198 | | 3.765995 | -0.25757 | 0.85907 |
| PPM1G | 4.997087 | 0.298268 | 0.999532 | | 5.146095 | 0.264196 | 0.813198 | | 5.082479 | -0.01396 | 0.996681 |
| TRIM45 | 2.208649 | 0.604186 | 0.999532 | | 2.299921 | 0.385929 | 0.813406 | | 2.364014 | -0.19898 | 0.910762 |
| RAB11B | 19.17591 | -0.04109 | 0.999532 | | 23.11689 | 0.166169 | 0.813588 | | 19.33135 | 0.229005 | 0.740755 |
| KANSL1 | 4.168239 | -0.58303 | 0.999532 | | 5.151517 | -0.26102 | 0.813588 | | 3.661599 | 0.341415 | 0.776541 |
| KIRREL1 | 7.97023 | 0.189291 | 0.999532 | | 9.006851 | 0.224725 | 0.813588 | | 7.916695 | 0.061964 | 0.967 |
| PRKD3 | 2.368309 | 0.01582 | 0.999532 | | 2.936323 | 0.331051 | 0.813588 | | 2.57035 | 0.333237 | 0.824012 |
| FLAD1 | 4.198091 | -0.23854 | 0.999532 | | 5.597536 | 0.257187 | 0.813588 | | 4.336972 | 0.508962 | 0.61958 |
| ASPDH | 3.418429 | -0.22363 | 0.999532 | | 3.628334 | -0.31384 | 0.813717 | | 2.992624 | -0.07514 | 0.972758 |
| KLHL34 | 2.041664 | 0.127918 | 0.999532 | | 2.609384 | 0.374054 | 0.813717 | | 2.293958 | 0.265829 | 0.883116 |
| DTX4 | 3.958182 | -0.41597 | 0.999532 | | 4.738104 | -0.30982 | 0.813717 | | 3.443952 | 0.130186 | 0.941197 |
| INSRR | 2.250289 | 0.619657 | 0.999532 | | 2.212809 | 0.388723 | 0.813717 | | 2.46411 | -0.20993 | 0.908891 |
| NUDT1 | 2.484287 | 0.320329 | 0.999532 | | 2.200803 | -0.39535 | 0.813717 | | 2.200605 | -0.69624 | 0.649779 |
| KLF13 | 8.246982 | 0.001773 | 0.999532 | | 9.915999 | 0.207044 | 0.813717 | | 8.179514 | 0.219092 | 0.813067 |
| ZNF674 | 1.904469 | 0.100459 | 0.999532 | | 2.459819 | 0.375182 | 0.813717 | | 2.077261 | 0.295938 | 0.872493 |
| ZNF107 | 2.026419 | 0.351746 | 0.999532 | | 2.103367 | 0.384097 | 0.813717 | | 2.211354 | 0.047916 | 0.987274 |
| GALT | 3.73173 | -0.161 | 0.999532 | | 4.999784 | 0.272767 | 0.813717 | | 4.074836 | 0.448953 | 0.67176 |
| SOX3 | 2.673001 | 0.685212 | 0.999532 | | 1.831973 | -0.40883 | 0.813717 | | 2.400873 | -1.06646 | 0.37598 |
| CRYM | 3.228845 | 0.447917 | 0.999532 | | 3.532296 | 0.31766 | 0.813717 | | 3.453785 | -0.09961 | 0.958191 |
| CYGB | 4.657549 | -0.31869 | 0.999532 | | 6.820514 | 0.252968 | 0.813717 | | 5.18901 | 0.584515 | 0.515762 |
| GBF1 | 3.183995 | -0.17314 | 0.999532 | | 4.263276 | 0.284016 | 0.813717 | | 3.404345 | 0.475542 | 0.672584 |
| GCNT1 | 2.085345 | -0.08807 | 0.999532 | | 2.202291 | -0.40237 | 0.813717 | | 1.83935 | -0.29225 | 0.883116 |
| GEMIN2 | 4.162355 | 0.050839 | 0.999532 | | 4.241484 | -0.3031 | 0.813717 | | 3.678902 | -0.33248 | 0.796512 |
| CCT2 | 6.963236 | -0.14045 | 0.999532 | | 8.911991 | 0.206107 | 0.813733 | | 7.244425 | 0.367665 | 0.650439 |
| GPR137 | 4.40271 | -0.14771 | 0.999532 | | 4.568136 | -0.29158 | 0.813733 | | 3.505116 | -0.11269 | 0.945129 |
| EGF | 3.110864 | 0.491186 | 0.999532 | | 3.36483 | 0.32839 | 0.813878 | | 3.092478 | -0.18824 | 0.913156 |
| GOT1 | 3.631028 | 0.105819 | 0.999532 | | 4.410135 | 0.288431 | 0.813878 | | 3.613827 | 0.187243 | 0.893259 |
| USP2 | 2.01071 | -0.37357 | 0.999532 | | 2.200147 | -0.38886 | 0.813878 | | 1.726414 | -0.00378 | 0.999385 |
| RASGRF2 | 2.377131 | 0.251945 | 0.999532 | | 2.740337 | 0.336558 | 0.814048 | | 2.499235 | 0.100564 | 0.960427 |
| MSR1 | 2.750455 | -0.0688 | 0.999532 | | 2.645548 | -0.35111 | 0.814048 | | 2.322841 | -0.26885 | 0.874653 |
| PYDC1 | 2.429772 | 0.142488 | 0.999532 | | 2.29033 | -0.37436 | 0.814137 | | 2.182231 | -0.50225 | 0.735469 |
| ZNF845 | 8.353228 | 0.232605 | 0.999532 | | 8.193641 | -0.25779 | 0.814137 | | 7.83043 | -0.47993 | 0.628053 |
| FUT10 | 1.765558 | 0.04997 | 0.999532 | | 2.157625 | 0.387936 | 0.814137 | | 1.950365 | 0.354391 | 0.849682 |
| C2CD4A | 2.732362 | 0.414684 | 0.999532 | | 2.355826 | -0.38008 | 0.814137 | | 2.399544 | -0.7753 | 0.565022 |
| UBE2E3 | 4.268046 | 0.384514 | 0.999532 | | 4.271808 | 0.287632 | 0.814263 | | 4.469213 | -0.07616 | 0.963866 |
| NBPF11 | 9.313343 | -0.10735 | 0.999532 | | 10.42146 | -0.21981 | 0.814263 | | 8.40819 | -0.08924 | 0.94366 |
| IGBP1 | 3.162661 | -0.04329 | 0.999532 | | 3.962314 | 0.294743 | 0.81434 | | 3.459426 | 0.355791 | 0.790952 |
| PFN1 | 43.85418 | -0.05625 | 0.999532 | | 53.84177 | 0.207047 | 0.814578 | | 45.1408 | 0.283734 | 0.717627 |
| WWP2 | 3.215418 | 0.172874 | 0.999532 | | 3.73145 | 0.291064 | 0.814648 | | 3.403805 | 0.134999 | 0.929087 |
| ECHDC2 | 9.106003 | 0.262796 | 0.999532 | | 9.835223 | 0.213947 | 0.814801 | | 8.953353 | -0.02489 | 0.988853 |
| DBN1 | 7.14523 | 0.224642 | 0.999532 | | 7.889256 | 0.213805 | 0.814801 | | 7.181478 | 0.010174 | 0.996681 |
| HAMP | 2.125798 | 0.493317 | 0.999532 | | 1.813101 | -0.43256 | 0.815089 | | 1.781753 | -0.90673 | 0.526942 |
| TSGA10IP | 3.521227 | 0.408541 | 0.999532 | | 3.157024 | -0.33916 | 0.81513 | | 3.10272 | -0.72019 | 0.536289 |
| ZC3H10 | 2.268228 | 0.574581 | 0.999532 | | 2.319766 | 0.393073 | 0.81515 | | 2.477583 | -0.17077 | 0.929604 |
| ADAMTS | 2.711364 | 0.464107 | 0.999532 | | 2.246379 | -0.38494 | 0.81515 | | 2.335651 | -0.83554 | 0.522078 |
| HAX1 | 6.628205 | 0.04345 | 0.999532 | | 7.259932 | -0.22821 | 0.81515 | | 6.043392 | -0.23402 | 0.827643 |
| TDP1 | 2.387273 | 0.434117 | 0.999532 | | 2.746515 | 0.354945 | 0.81515 | | 2.550413 | -0.05494 | 0.982124 |
| VMA21 | 5.906248 | -0.11133 | 0.999532 | | 7.438078 | 0.224512 | 0.81515 | | 6.234132 | 0.351721 | 0.709293 |
| SURF6 | 2.858881 | 1.044117 | 0.999532 | | 2.371679 | 0.371877 | 0.81515 | | 3.073684 | -0.66409 | 0.58911 |
| CLOCK | 4.298401 | 0.082424 | 0.999532 | | 5.250964 | 0.265946 | 0.81515 | | 4.662553 | 0.202874 | 0.87764 |
| METTL22 | 2.232907 | 0.204361 | 0.999532 | | 2.727065 | 0.351838 | 0.815293 | | 2.480056 | 0.167823 | 0.927544 |
| MAN2B2 | 3.459487 | -0.61514 | 0.999532 | | 4.160137 | -0.31002 | 0.815296 | | 2.842593 | 0.327512 | 0.819366 |
| TRIM31 | 2.06664 | 0.07876 | 0.999532 | | 1.983135 | -0.3922 | 0.815384 | | 1.837451 | -0.45262 | 0.792416 |
| PPP5C | 5.216525 | 0.201056 | 0.999532 | | 6.306405 | 0.252904 | 0.815384 | | 5.595055 | 0.071527 | 0.961544 |
| ATP4A | 2.471 | 0.187233 | 0.999532 | | 2.327676 | -0.38915 | 0.815384 | | 2.138871 | -0.55919 | 0.720106 |
| THOC5 | 3.765716 | 0.503161 | 0.999532 | | 3.713441 | 0.289121 | 0.815384 | | 3.775617 | -0.20363 | 0.882613 |
| MIS18A | 2.583138 | -0.22089 | 0.999532 | | 2.812194 | -0.341 | 0.815384 | | 2.21562 | -0.09652 | 0.96588 |
| JRKL | 2.509306 | 0.254588 | 0.999532 | | 3.064185 | 0.346313 | 0.815384 | | 2.793927 | 0.122711 | 0.950669 |
| CALML6 | 3.662185 | 0.4732 | 0.999532 | | 2.913701 | -0.35232 | 0.815384 | | 3.263215 | -0.80694 | 0.510412 |
| PYCR3 | 2.175649 | 0.001688 | 0.999532 | | 2.19341 | -0.3854 | 0.815384 | | 1.902649 | -0.36872 | 0.837256 |
| NPY4R | 2.607028 | 0.233877 | 0.999532 | | 2.443446 | -0.38293 | 0.815384 | | 2.251818 | -0.59636 | 0.679818 |
| WDR37 | 2.955328 | -0.11068 | 0.999532 | | 3.167858 | -0.32567 | 0.815438 | | 2.593792 | -0.1971 | 0.904801 |
| ACVR2A | 2.378683 | 0.284116 | 0.999532 | | 2.828154 | 0.346095 | 0.815621 | | 2.635693 | 0.080549 | 0.972031 |
| DHRSX | 2.502795 | 0.322599 | 0.999532 | | 2.741138 | 0.341837 | 0.815621 | | 2.638078 | 0.035858 | 0.989646 |
| TBRG1 | 4.045242 | -0.25449 | 0.999532 | | 5.541081 | 0.24898 | 0.815621 | | 4.371881 | 0.52488 | 0.600081 |
| UBE2F | 2.077675 | 0.647247 | 0.999532 | | 2.039491 | 0.388086 | 0.815621 | | 2.226546 | -0.24501 | 0.891051 |
| TMED1 | 2.769385 | 0.582363 | 0.999532 | | 2.757892 | 0.340785 | 0.815621 | | 2.84499 | -0.22543 | 0.883116 |
| PIK3AP1 | 2.88229 | 0.072603 | 0.999532 | | 3.603736 | 0.307836 | 0.815621 | | 3.218051 | 0.254159 | 0.863401 |
| TMEM209 | 4.230764 | 0.282915 | 0.999532 | | 4.676261 | 0.280746 | 0.815621 | | 4.421001 | 0.016072 | 0.995943 |
| UCHL1 | 4.254929 | -0.42405 | 0.999532 | | 6.498089 | 0.252207 | 0.815621 | | 4.813662 | 0.685334 | 0.436389 |
| USP24 | 3.914787 | -0.31966 | 0.999532 | | 4.580146 | -0.27555 | 0.815621 | | 3.284419 | 0.07205 | 0.971732 |
| STK35 | 1.984611 | -0.01106 | 0.999532 | | 2.032393 | -0.41163 | 0.815625 | | 1.699994 | -0.38021 | 0.843043 |
| MYCN | 2.266374 | 0.2747 | 0.999532 | | 2.531828 | 0.361836 | 0.816006 | | 2.480731 | 0.105678 | 0.961964 |
| NT5C2 | 4.935564 | 0.442523 | 0.999532 | | 5.311135 | 0.250884 | 0.816093 | | 5.258233 | -0.16315 | 0.895864 |
| NKAPD1 | 2.836833 | -0.52016 | 0.999532 | | 3.457724 | -0.31543 | 0.816093 | | 2.614121 | 0.230001 | 0.896228 |
| SLC39A2 | 2.571679 | 0.200447 | 0.999532 | | 2.399995 | -0.3819 | 0.816093 | | 2.272269 | -0.56463 | 0.702463 |
| NIF3L1 | 2.836957 | -0.42607 | 0.999532 | | 3.067805 | -0.36263 | 0.816093 | | 2.227209 | 0.093357 | 0.969693 |
| ZNF23 | 2.84911 | 0.690629 | 0.999532 | | 2.486579 | 0.351838 | 0.816093 | | 2.931785 | -0.34574 | 0.827856 |
| C1orf54 | 3.87022 | -0.22404 | 0.999532 | | 4.70772 | 0.268116 | 0.816093 | | 3.842756 | 0.496395 | 0.660097 |
| TDRKH | 2.054334 | -0.23399 | 0.999532 | | 2.233634 | -0.38869 | 0.816093 | | 1.893624 | -0.13493 | 0.956011 |
| CILK1 | 2.538604 | 0.492061 | 0.999532 | | 2.761931 | 0.344855 | 0.816093 | | 2.716404 | -0.12887 | 0.94366 |
| MYRIP | 2.248774 | 0.01681 | 0.999532 | | 2.197483 | -0.3861 | 0.816101 | | 1.940911 | -0.38601 | 0.827267 |
| C6orf62 | 5.883698 | -0.01151 | 0.999532 | | 7.065989 | 0.235065 | 0.816182 | | 6.070921 | 0.264205 | 0.797544 |
| ZNF737 | 2.384723 | 0.198456 | 0.999532 | | 2.905735 | 0.33378 | 0.81628 | | 2.688669 | 0.154696 | 0.930262 |
| DDX55 | 3.806713 | 0.155642 | 0.999532 | | 3.504488 | -0.31669 | 0.816499 | | 3.294192 | -0.47103 | 0.725408 |
| BACH1 | 7.325521 | -0.03062 | 0.999532 | | 8.591903 | 0.204869 | 0.816573 | | 7.366448 | 0.247386 | 0.791576 |
| PIP4K2B | 4.165722 | 0.367516 | 0.999532 | | 4.45613 | 0.270626 | 0.816922 | | 4.373724 | -0.07509 | 0.963359 |
| C11orf80 | 2.228285 | -0.14595 | 0.999532 | | 2.322528 | -0.37856 | 0.816922 | | 1.924207 | -0.21556 | 0.914761 |
| PPM1B | 7.311568 | -0.31652 | 0.999532 | | 8.439492 | -0.21209 | 0.816922 | | 6.668091 | 0.120569 | 0.920124 |
| PPIG | 7.835204 | 0.109644 | 0.999532 | | 9.56333 | 0.212561 | 0.816924 | | 8.384947 | 0.125028 | 0.906811 |
| WDR33 | 6.469688 | 0.18873 | 0.999532 | | 7.013678 | 0.217002 | 0.816924 | | 6.508865 | 0.047296 | 0.97386 |
| FEV | 2.410623 | -0.37647 | 0.999532 | | 2.702449 | -0.35729 | 0.816924 | | 2.105586 | 0.039283 | 0.991103 |
| OR10H3 | 2.421813 | -0.21604 | 0.999532 | | 2.701915 | -0.36225 | 0.818017 | | 2.151113 | -0.12616 | 0.955834 |
| GGA3 | 3.560338 | 0.22722 | 0.999532 | | 3.983912 | 0.288345 | 0.818103 | | 3.602647 | 0.080262 | 0.963359 |
| BET1L | 5.09311 | -0.02276 | 0.999532 | | 6.097248 | 0.236196 | 0.818103 | | 5.290374 | 0.277386 | 0.792539 |
| ADSS2 | 5.126758 | -0.33117 | 0.999532 | | 6.784259 | 0.242969 | 0.818103 | | 5.389879 | 0.585605 | 0.560424 |
| TYMP | 5.67343 | -0.09372 | 0.999532 | | 5.907234 | -0.29154 | 0.818103 | | 5.068673 | -0.15299 | 0.903339 |
| PRKCD | 2.907304 | 0.633627 | 0.999532 | | 3.009931 | 0.328149 | 0.818103 | | 3.030955 | -0.28349 | 0.842754 |
| PNPLA4 | 3.312835 | 0.403638 | 0.999532 | | 3.051112 | -0.3384 | 0.818103 | | 2.863733 | -0.71968 | 0.532055 |
| RUNX1T1 | 5.020356 | 0.334393 | 0.999532 | | 5.403624 | 0.255119 | 0.818103 | | 5.238177 | -0.05823 | 0.972221 |
| RPP30 | 3.736475 | 0.044303 | 0.999532 | | 4.013711 | -0.28989 | 0.818103 | | 3.281363 | -0.31704 | 0.806797 |
| SAMM50 | 2.426859 | 0.136161 | 0.999532 | | 2.771354 | 0.337946 | 0.818103 | | 2.48834 | 0.215208 | 0.90016 |
| NEFH | 2.082011 | 0.319606 | 0.999532 | | 2.006739 | -0.40953 | 0.818103 | | 1.795696 | -0.70732 | 0.644433 |
| ZNF552 | 3.787945 | 0.725261 | 0.999532 | | 2.78887 | -0.34242 | 0.818103 | | 3.390579 | -1.05308 | 0.266933 |
| CDK2 | 2.730669 | 0.714279 | 0.999532 | | 2.612247 | 0.343566 | 0.818119 | | 2.759158 | -0.35366 | 0.79686 |
| HPGD | 12.69545 | 0.30568 | 0.999532 | | 14.26818 | 0.292431 | 0.818119 | | 13.57889 | 0.023431 | 0.991273 |
| GGPS1 | 2.696952 | 0.326589 | 0.999532 | | 2.906859 | 0.341254 | 0.818147 | | 2.73012 | 0.031486 | 0.991273 |
| GTPBP6 | 5.755774 | 0.100078 | 0.999532 | | 6.879071 | 0.238006 | 0.818147 | | 6.007814 | 0.157722 | 0.895421 |
| RPEL1 | 2.462994 | 0.023787 | 0.999532 | | 2.384937 | -0.37579 | 0.818147 | | 2.07796 | -0.38063 | 0.821752 |
| USP43 | 2.935849 | 0.007299 | 0.999532 | | 2.983836 | -0.36277 | 0.818147 | | 2.554466 | -0.35078 | 0.829716 |
| ZNF439 | 2.022731 | 0.096477 | 0.999532 | | 1.921685 | -0.40026 | 0.818147 | | 1.712367 | -0.48199 | 0.783395 |
| BIRC7 | 1.753058 | -0.04347 | 0.999532 | | 2.154591 | 0.380809 | 0.818147 | | 1.917001 | 0.441017 | 0.800239 |
| ASB13 | 2.080927 | 0.033194 | 0.999532 | | 2.337453 | 0.367431 | 0.818147 | | 2.224725 | 0.342888 | 0.852482 |
| SLC6A20 | 3.031668 | -0.32709 | 0.999532 | | 3.464718 | -0.33708 | 0.818147 | | 2.622623 | 0.011041 | 0.996891 |
| OR5C1 | 2.631933 | 0.210076 | 0.999532 | | 2.59095 | -0.35424 | 0.818147 | | 2.430551 | -0.532 | 0.715437 |
| GJC2 | 4.861793 | -0.20163 | 0.999532 | | 5.462155 | -0.2646 | 0.818147 | | 4.395177 | -0.0331 | 0.989437 |
| CLDN7 | 4.96728 | 0.179106 | 0.999532 | | 4.767854 | -0.29094 | 0.818147 | | 4.222824 | -0.45093 | 0.660097 |
| AP3M1 | 3.76938 | 0.641133 | 0.999532 | | 3.726493 | 0.309468 | 0.818147 | | 4.087982 | -0.32361 | 0.792691 |
| GPR45 | 2.035929 | -0.43403 | 0.999532 | | 2.325934 | -0.38627 | 0.818499 | | 1.708998 | 0.066171 | 0.982124 |
| OR5AN1 | 2.153285 | -0.10832 | 0.999532 | | 2.289654 | -0.36809 | 0.818536 | | 1.871385 | -0.23782 | 0.901813 |
| CYP4F3 | 2.932205 | 0.179598 | 0.999532 | | 3.614443 | 0.328127 | 0.818709 | | 2.938685 | 0.183329 | 0.906829 |
| ZNF652 | 3.563588 | 0.202011 | 0.999532 | | 4.091043 | 0.27522 | 0.818871 | | 3.744116 | 0.091046 | 0.955834 |
| MSH5 | 3.880088 | 0.024702 | 0.999532 | | 5.111125 | 0.283842 | 0.818912 | | 4.233645 | 0.280571 | 0.82512 |
| ZNF334 | 2.538258 | -0.06884 | 0.999532 | | 2.701208 | -0.34409 | 0.819061 | | 2.201851 | -0.24976 | 0.88885 |
| RNLS | 2.374075 | 0.128293 | 0.999532 | | 2.357941 | -0.35802 | 0.819061 | | 2.039611 | -0.45997 | 0.760473 |
| LUC7L3 | 9.719401 | -0.05111 | 0.999532 | | 11.70249 | 0.192976 | 0.819061 | | 9.77288 | 0.267405 | 0.722703 |
| GP1BA | 2.285484 | 0.095532 | 0.999532 | | 2.137234 | -0.39393 | 0.819061 | | 1.884089 | -0.47213 | 0.78258 |
| SRGAP2B | 34.35418 | 0.149215 | 0.999532 | | 39.05335 | 0.168889 | 0.819061 | | 34.65688 | 0.041063 | 0.965649 |
| SPRED1 | 9.515349 | 0.172539 | 0.999532 | | 10.38897 | 0.195144 | 0.819061 | | 9.745365 | 0.040716 | 0.974588 |
| ZNF347 | 2.646102 | -0.16269 | 0.999532 | | 3.154009 | 0.319294 | 0.819061 | | 2.62269 | 0.496126 | 0.718643 |
| SLC12A9 | 3.638664 | -0.08159 | 0.999532 | | 3.662178 | -0.30271 | 0.819061 | | 3.153262 | -0.20872 | 0.892328 |
| MTSS2 | 2.762228 | -0.12149 | 0.999532 | | 3.568761 | 0.301891 | 0.819061 | | 3.089805 | 0.442107 | 0.735469 |
| C3orf62 | 2.222962 | 0.365945 | 0.999532 | | 1.899845 | -0.40744 | 0.819061 | | 1.902621 | -0.75727 | 0.619775 |
| FIBCD1 | 3.08303 | 0.383319 | 0.999532 | | 2.769417 | -0.35655 | 0.819061 | | 2.611644 | -0.71796 | 0.542124 |
| CEP19 | 1.880145 | 0.138711 | 0.999532 | | 1.814855 | -0.41365 | 0.819061 | | 1.700183 | -0.53534 | 0.75311 |
| ABHD11 | 2.430706 | 0.280288 | 0.999532 | | 2.86427 | 0.365492 | 0.819061 | | 2.547729 | 0.10013 | 0.960773 |
| LMF2 | 3.881631 | 0.100654 | 0.999532 | | 4.577645 | 0.259299 | 0.819061 | | 3.844764 | 0.170093 | 0.90267 |
| ERG28 | 4.032625 | -0.03124 | 0.999532 | | 5.27287 | 0.258342 | 0.819061 | | 4.274463 | 0.318565 | 0.790208 |
| TRIM62 | 3.265446 | -0.16914 | 0.999532 | | 3.42073 | -0.31615 | 0.819061 | | 2.73298 | -0.13038 | 0.945129 |
| PRKRIP1 | 5.657421 | 0.519501 | 0.999532 | | 6.131339 | 0.268543 | 0.819061 | | 6.087482 | -0.21391 | 0.865065 |
| DIP2C | 2.194143 | 0.004474 | 0.999532 | | 2.270741 | -0.37026 | 0.819061 | | 1.815136 | -0.35324 | 0.83987 |
| MXD4 | 12.98171 | -0.17246 | 0.999532 | | 16.29694 | 0.19133 | 0.819061 | | 13.19015 | 0.381137 | 0.573454 |
| C19orf12 | 2.66846 | 0.244812 | 0.999532 | | 2.51577 | -0.36972 | 0.819061 | | 2.256035 | -0.59275 | 0.668481 |
| ZNF133 | 3.975413 | 0.708948 | 0.999532 | | 3.768765 | 0.326816 | 0.819061 | | 4.341955 | -0.34878 | 0.783127 |
| GRIP2 | 4.792831 | -0.26693 | 0.999532 | | 5.400669 | -0.27002 | 0.819061 | | 4.277557 | 0.01521 | 0.996681 |
| SLC7A2 | 3.144459 | 0.567376 | 0.999532 | | 2.315595 | -0.37047 | 0.819354 | | 2.728108 | -0.93526 | 0.420356 |
| STK3 | 3.31453 | -0.08937 | 0.999532 | | 3.354148 | -0.32428 | 0.819354 | | 2.799734 | -0.21498 | 0.894612 |
| SLC49A4 | 1.916325 | -0.17678 | 0.999532 | | 2.629206 | 0.340708 | 0.819354 | | 2.167091 | 0.537375 | 0.716722 |
| BRINP2 | 2.365089 | 0.836249 | 0.999532 | | 2.50327 | 0.374874 | 0.819354 | | 2.689459 | -0.4295 | 0.76302 |
| PIM1 | 3.407287 | -0.14705 | 0.999532 | | 3.810104 | -0.30274 | 0.819437 | | 3.053526 | -0.13794 | 0.93603 |
| CERK | 5.965998 | 0.201232 | 0.999532 | | 6.651595 | 0.235164 | 0.819437 | | 6.043496 | 0.05266 | 0.972924 |
| ETV7 | 2.554821 | -0.32317 | 0.999532 | | 2.953238 | -0.34093 | 0.819683 | | 2.184958 | 0.001743 | 0.999429 |
| RECQL4 | 2.141903 | -0.234 | 0.999532 | | 2.850298 | 0.349639 | 0.819683 | | 2.27074 | 0.602936 | 0.685955 |
| NEPRO | 2.863825 | -0.51137 | 0.999532 | | 3.257003 | -0.30568 | 0.819683 | | 2.475945 | 0.226229 | 0.89416 |
| RALB | 5.718283 | 0.256537 | 0.999532 | | 6.308915 | 0.232211 | 0.819683 | | 5.925526 | -0.00681 | 0.996891 |
| F2RL1 | 1.836326 | -0.22754 | 0.999532 | | 2.397981 | 0.351915 | 0.819732 | | 2.030994 | 0.59516 | 0.69941 |
| PLEKHF2 | 2.785685 | -0.05412 | 0.999532 | | 2.816828 | -0.34186 | 0.819777 | | 2.436608 | -0.26963 | 0.872493 |
| TOR4A | 2.225482 | -0.04119 | 0.999532 | | 2.331481 | -0.36405 | 0.819777 | | 2.008496 | -0.30782 | 0.861545 |
| NECAB1 | 2.867763 | 0.345159 | 0.999532 | | 2.827173 | -0.38816 | 0.819948 | | 2.58939 | -0.71504 | 0.599857 |
| EXOSC8 | 3.616358 | -0.08155 | 0.999532 | | 3.789482 | -0.29665 | 0.819948 | | 3.138535 | -0.19696 | 0.896374 |
| NPTX1 | 1.917459 | 0.072131 | 0.999532 | | 2.484699 | 0.367149 | 0.819993 | | 2.149186 | 0.314209 | 0.861815 |
| NTRK1 | 2.011232 | -0.023 | 0.999532 | | 2.178732 | -0.3962 | 0.819993 | | 1.882731 | -0.34223 | 0.863907 |
| SP100 | 8.423436 | -0.20282 | 0.999532 | | 9.497608 | -0.19855 | 0.819993 | | 7.39575 | 0.026469 | 0.988379 |
| CEMP1 | 2.358617 | 0.223704 | 0.999532 | | 2.277577 | -0.37583 | 0.819993 | | 2.002157 | -0.57303 | 0.703463 |
| TGFB2 | 3.0012 | -0.33431 | 0.999532 | | 3.347419 | -0.32787 | 0.820203 | | 2.60264 | 0.022152 | 0.99497 |
| METTL1 | 4.000967 | 0.376626 | 0.999532 | | 3.372339 | -0.30681 | 0.82037 | | 3.368132 | -0.665 | 0.524206 |
| CD4 | 5.879854 | -0.56756 | 0.999532 | | 7.419045 | -0.23734 | 0.820424 | | 5.100565 | 0.350216 | 0.737952 |
| FAM117B | 3.538513 | 0.598394 | 0.999532 | | 3.606873 | 0.288626 | 0.820424 | | 3.782203 | -0.29197 | 0.803301 |
| RPS2 | 26.84124 | -0.10107 | 0.999532 | | 33.29131 | 0.176883 | 0.820496 | | 27.15731 | 0.308483 | 0.703463 |
| IP6K1 | 3.039822 | -0.21928 | 0.999532 | | 4.27204 | 0.271302 | 0.820533 | | 3.421798 | 0.512108 | 0.652925 |
| GALC | 2.913695 | 0.598536 | 0.999532 | | 3.004784 | 0.32951 | 0.820893 | | 3.112737 | -0.24924 | 0.864245 |
| BLVRA | 3.003907 | -0.01296 | 0.999532 | | 3.797713 | 0.289023 | 0.820893 | | 3.157674 | 0.318245 | 0.810635 |
| DIDO1 | 5.035379 | -0.40626 | 0.999532 | | 5.733519 | -0.26668 | 0.820893 | | 4.103186 | 0.155269 | 0.915533 |
| ANTXRL | 2.508499 | 0.553921 | 0.999532 | | 2.168458 | -0.40277 | 0.820893 | | 2.14416 | -0.93795 | 0.456688 |
| MLF2 | 7.98503 | -0.31617 | 0.999532 | | 9.267575 | -0.20386 | 0.820893 | | 6.960397 | 0.134308 | 0.900848 |
| SHROOM | 4.370129 | -0.17649 | 0.999532 | | 4.826207 | -0.26738 | 0.820907 | | 3.601187 | -0.07081 | 0.969693 |
| PHACTR | 2.846437 | 0.121697 | 0.999532 | | 2.69844 | -0.343 | 0.820907 | | 2.5105 | -0.45342 | 0.755206 |
| ZNF408 | 2.291961 | -0.0068 | 0.999532 | | 2.43602 | -0.37338 | 0.820907 | | 1.892394 | -0.34246 | 0.847986 |
| ISG20L2 | 3.911484 | 0.235576 | 0.999532 | | 3.58418 | -0.30632 | 0.820907 | | 3.540652 | -0.51657 | 0.663633 |
| LMF1 | 2.208847 | 0.116729 | 0.999532 | | 2.804 | 0.343699 | 0.820955 | | 2.47492 | 0.247085 | 0.885086 |
| SH2D4A | 2.217433 | 0.69905 | 0.999532 | | 2.005065 | 0.376713 | 0.820955 | | 2.294926 | -0.31008 | 0.857043 |
| GTF2H4 | 2.738863 | -0.15955 | 0.999532 | | 2.82242 | -0.3254 | 0.820955 | | 2.312429 | -0.15056 | 0.938429 |
| GCC1 | 2.339358 | 0.399166 | 0.999532 | | 2.132467 | -0.39587 | 0.820955 | | 2.198317 | -0.7886 | 0.553283 |
| OSTC | 5.970678 | -0.01225 | 0.999532 | | 7.240089 | 0.219762 | 0.820955 | | 6.060095 | 0.248055 | 0.809485 |
| ZNF146 | 5.017354 | 0.046282 | 0.999532 | | 6.362964 | 0.237194 | 0.820955 | | 5.294265 | 0.217426 | 0.85585 |
| ALKBH5 | 6.560798 | -0.2159 | 0.999532 | | 8.766211 | 0.205039 | 0.820955 | | 6.74157 | 0.435259 | 0.59686 |
| FAM53C | 2.978282 | -0.13826 | 0.999532 | | 3.998943 | 0.307404 | 0.820955 | | 3.298477 | 0.461203 | 0.70719 |
| CD8A | 2.806433 | -0.00252 | 0.999532 | | 2.923457 | -0.33864 | 0.820955 | | 2.556095 | -0.31119 | 0.849682 |
| SPATA19 | 2.548411 | 0.216222 | 0.999532 | | 2.317859 | -0.35704 | 0.820955 | | 2.210935 | -0.55602 | 0.694961 |
| GALM | 5.356779 | 0.046295 | 0.999532 | | 6.475602 | 0.246616 | 0.820955 | | 5.579901 | 0.220896 | 0.849128 |
| PRTG | 3.224074 | 0.262765 | 0.999532 | | 3.114997 | -0.33641 | 0.820955 | | 2.94326 | -0.57195 | 0.662584 |
| PHPT1 | 4.7416 | 0.588494 | 0.999532 | | 4.604987 | 0.282587 | 0.820955 | | 4.888209 | -0.2887 | 0.809832 |
| IKBKB | 4.474559 | 0.235846 | 0.999532 | | 4.864816 | 0.263526 | 0.820955 | | 4.64525 | 0.051504 | 0.975473 |
| GDI1 | 7.802984 | -0.03812 | 0.999532 | | 9.75759 | 0.228974 | 0.820955 | | 8.284086 | 0.286557 | 0.767184 |
| PRDX3 | 5.674384 | 0.006938 | 0.999532 | | 6.752155 | 0.224884 | 0.820955 | | 5.926954 | 0.23634 | 0.825181 |
| NSUN6 | 1.800937 | -0.23585 | 0.999532 | | 1.79495 | -0.41092 | 0.820955 | | 1.586199 | -0.15647 | 0.951656 |
| CLEC2B | 3.330248 | -0.2602 | 0.999532 | | 3.788539 | -0.28749 | 0.820955 | | 2.826267 | -0.00591 | 0.998094 |
| TLE1 | 4.415465 | -0.12081 | 0.999532 | | 4.516116 | -0.27633 | 0.820955 | | 3.69442 | -0.14071 | 0.92697 |
| C9orf64 | 2.256535 | 0.383765 | 0.999532 | | 2.671845 | 0.347461 | 0.820955 | | 2.457401 | -0.00937 | 0.99716 |
| PDCD7 | 2.691934 | -0.24125 | 0.999532 | | 3.482187 | 0.292514 | 0.820955 | | 2.808417 | 0.548967 | 0.660097 |
| ZNF558 | 2.869665 | -0.05519 | 0.999532 | | 3.057806 | -0.33019 | 0.820955 | | 2.527908 | -0.2541 | 0.881743 |
| KAT2A | 4.437107 | -0.0986 | 0.999532 | | 5.75835 | 0.236885 | 0.820955 | | 4.498713 | 0.351877 | 0.737109 |
| MYLK3 | 2.299584 | -0.41111 | 0.999532 | | 3.279793 | 0.345193 | 0.820955 | | 2.491971 | 0.781151 | 0.577942 |
| SMG1 | 4.733954 | 0.015127 | 0.999532 | | 4.761064 | -0.26073 | 0.820955 | | 3.954328 | -0.25753 | 0.83086 |
| NAA80 | 3.036166 | 0.443529 | 0.999532 | | 2.549169 | -0.34374 | 0.820955 | | 2.619003 | -0.77035 | 0.512428 |
| SMOX | 2.36251 | -0.11714 | 0.999532 | | 2.497009 | -0.346 | 0.82103 | | 1.993253 | -0.21049 | 0.910105 |
| SIAE | 4.89316 | 0.011693 | 0.999532 | | 6.120118 | 0.247693 | 0.82103 | | 5.189867 | 0.257631 | 0.819366 |
| RBCK1 | 8.442265 | -0.13268 | 0.999532 | | 10.69736 | 0.208879 | 0.82103 | | 8.552191 | 0.373634 | 0.660097 |
| DHX16 | 2.923549 | 0.164653 | 0.999532 | | 2.646751 | -0.33897 | 0.82103 | | 2.51261 | -0.49689 | 0.722703 |
| RGS16 | 6.091627 | -0.11102 | 0.999532 | | 6.606182 | -0.23252 | 0.82103 | | 5.38857 | -0.1011 | 0.941973 |
| PNO1 | 2.446497 | -0.61941 | 0.999532 | | 2.930045 | -0.3449 | 0.82103 | | 2.200546 | 0.283601 | 0.883785 |
| ANGPTL7 | 1.949005 | -0.25713 | 0.999532 | | 2.189831 | -0.38243 | 0.82103 | | 1.789323 | -0.10976 | 0.965649 |
| UTP25 | 2.14909 | 0.16023 | 0.999532 | | 2.113776 | -0.3722 | 0.82103 | | 1.853245 | -0.51372 | 0.735469 |
| FNTA | 5.082381 | -0.0753 | 0.999532 | | 5.393205 | -0.24528 | 0.82103 | | 4.367778 | -0.15385 | 0.906811 |
| RTN4RL2 | 4.074891 | 0.078988 | 0.999532 | | 4.183125 | -0.28292 | 0.82103 | | 3.556086 | -0.34188 | 0.791466 |
| SRPK2 | 4.624416 | -0.1213 | 0.999532 | | 4.986787 | -0.25795 | 0.821095 | | 4.131825 | -0.11706 | 0.937326 |
| OSBPL10 | 3.348104 | 0.199001 | 0.999532 | | 3.67782 | 0.293112 | 0.821095 | | 3.446171 | 0.106591 | 0.951656 |
| CDK11A | 6.194968 | -0.02806 | 0.999532 | | 7.341237 | 0.218156 | 0.821249 | | 6.322768 | 0.26462 | 0.801905 |
| POC1B | 2.317601 | 0.410186 | 0.999532 | | 2.401169 | 0.363916 | 0.821364 | | 2.330555 | -0.0292 | 0.992286 |
| C1QTNF2 | 2.28037 | -0.1969 | 0.999532 | | 2.327425 | -0.37033 | 0.821465 | | 1.905897 | -0.15691 | 0.943845 |
| TMEM109 | 6.021207 | -0.09783 | 0.999532 | | 7.502399 | 0.218594 | 0.821496 | | 6.192986 | 0.335121 | 0.717138 |
| MED4 | 7.297893 | 0.437989 | 0.999532 | | 7.460036 | 0.212232 | 0.821496 | | 7.609335 | -0.20413 | 0.835289 |
| S100A5 | 2.153352 | 0.214797 | 0.999532 | | 1.93193 | -0.39497 | 0.821496 | | 1.836359 | -0.59316 | 0.708378 |
| ACSL1 | 5.749432 | -0.09216 | 0.999532 | | 7.27529 | 0.281492 | 0.821496 | | 5.913145 | 0.407454 | 0.705425 |
| GMEB2 | 4.287671 | -0.09051 | 0.999532 | | 4.674076 | -0.27675 | 0.821504 | | 3.564845 | -0.1687 | 0.908698 |
| CYP2A13 | 2.0002 | 0.021691 | 0.999532 | | 1.948003 | -0.39757 | 0.821504 | | 1.706001 | -0.40221 | 0.826947 |
| COL7A1 | 3.475692 | -0.14895 | 0.999532 | | 3.772344 | -0.29791 | 0.822049 | | 2.789487 | -0.14023 | 0.935799 |
| NAT16 | 3.235978 | 0.619295 | 0.999532 | | 2.692439 | -0.34374 | 0.822262 | | 2.968085 | -0.94018 | 0.385915 |
| GALNT7 | 1.786995 | 0.28397 | 0.999532 | | 2.047414 | 0.372956 | 0.822393 | | 1.9128 | 0.106326 | 0.963492 |
| GTSE1 | 2.167667 | 0.138026 | 0.999532 | | 2.0818 | -0.39928 | 0.822393 | | 1.815189 | -0.51842 | 0.757035 |
| MUC1 | 3.843581 | 0.155097 | 0.999532 | | 3.79933 | -0.32584 | 0.822393 | | 3.113863 | -0.44871 | 0.738349 |
| PPP2R5D | 2.894289 | 0.15298 | 0.999532 | | 3.373624 | 0.304681 | 0.822393 | | 2.975736 | 0.17061 | 0.913859 |
| WDR26 | 6.407005 | -0.21163 | 0.999532 | | 8.080434 | 0.21883 | 0.822393 | | 6.349769 | 0.449354 | 0.610861 |
| DOCK3 | 2.297243 | 0.059986 | 0.999532 | | 2.136035 | -0.38227 | 0.82251 | | 2.010963 | -0.42212 | 0.806394 |
| PCNX2 | 2.860103 | 0.091744 | 0.999532 | | 2.902378 | -0.34769 | 0.82251 | | 2.365752 | -0.42929 | 0.766067 |
| MECP2 | 4.331774 | -0.17572 | 0.999532 | | 5.783831 | 0.247525 | 0.82251 | | 4.609051 | 0.437415 | 0.660097 |
| PIP4K2C | 3.836263 | -0.13886 | 0.999532 | | 4.217639 | -0.27997 | 0.82251 | | 3.357318 | -0.12143 | 0.941452 |
| GPATCH2 | 4.393313 | 0.109941 | 0.999532 | | 4.992937 | 0.253146 | 0.82251 | | 4.502457 | 0.156406 | 0.907163 |
| USP54 | 2.777865 | 0.050758 | 0.999532 | | 2.67578 | -0.3509 | 0.82251 | | 2.351128 | -0.38958 | 0.809832 |
| MS4A1 | 1.628031 | 0.496613 | 0.999532 | | 1.813215 | 0.409908 | 0.82251 | | 1.886704 | -0.07034 | 0.978463 |
| DNAL1 | 3.166551 | -0.46619 | 0.999532 | | 3.528407 | -0.29493 | 0.82251 | | 2.599216 | 0.182136 | 0.914929 |
| ZNF784 | 2.818992 | 0.150411 | 0.999532 | | 2.666878 | -0.34398 | 0.822533 | | 2.557254 | -0.46539 | 0.75311 |
| AVL9 | 1.658349 | -0.02071 | 0.999532 | | 2.307696 | 0.365819 | 0.822533 | | 1.991122 | 0.409928 | 0.810208 |
| RHCE | 3.5212 | 0.149846 | 0.999532 | | 3.362403 | -0.32234 | 0.822533 | | 3.082952 | -0.45601 | 0.746876 |
| PRMT1 | 6.712257 | 0.010379 | 0.999532 | | 8.003919 | 0.211624 | 0.822533 | | 6.857607 | 0.214088 | 0.835972 |
| HTR2A | 2.484655 | -0.21004 | 0.999532 | | 2.512043 | -0.35802 | 0.822621 | | 2.109011 | -0.13585 | 0.950669 |
| TAF5L | 2.35898 | 0.087999 | 0.999532 | | 2.481252 | -0.35713 | 0.822621 | | 2.168472 | -0.41976 | 0.792416 |
| KRTAP3-3 | 2.349511 | 0.09462 | 0.999532 | | 2.260042 | -0.37812 | 0.823061 | | 2.084001 | -0.45956 | 0.791576 |
| ZNF792 | 2.60318 | 0.014227 | 0.999532 | | 3.281648 | 0.306916 | 0.823129 | | 3.037581 | 0.313826 | 0.831013 |
| PHKG1 | 2.853985 | 0.022637 | 0.999532 | | 2.755939 | -0.33527 | 0.823129 | | 2.471289 | -0.34379 | 0.828869 |
| NDUFA8 | 4.237865 | 0.351313 | 0.999532 | | 4.605951 | 0.264599 | 0.823129 | | 4.385217 | -0.06845 | 0.968235 |
| RFTN1 | 7.910431 | -0.60228 | 0.999532 | | 9.944182 | -0.19451 | 0.823129 | | 7.182895 | 0.442147 | 0.568812 |
| H3C8 | 2.500329 | -0.18122 | 0.999532 | | 2.884027 | -0.34473 | 0.823307 | | 2.257074 | -0.13804 | 0.946765 |
| FAM171A | 3.110011 | -0.14844 | 0.999532 | | 3.33919 | -0.30715 | 0.823307 | | 2.677826 | -0.14082 | 0.938839 |
| CARNS1 | 2.124408 | 0.373869 | 0.999532 | | 1.874492 | -0.39936 | 0.823307 | | 1.862887 | -0.75891 | 0.616063 |
| OR4F6 | 2.758041 | -0.05303 | 0.999532 | | 2.830336 | -0.34778 | 0.823307 | | 2.362702 | -0.27786 | 0.876781 |
| FBXO47 | 2.425408 | 0.315215 | 0.999532 | | 2.263285 | -0.38072 | 0.823307 | | 2.04777 | -0.67204 | 0.645376 |
| DOCK4 | 3.518206 | -0.0324 | 0.999532 | | 4.202319 | 0.268365 | 0.823365 | | 3.672443 | 0.318835 | 0.795612 |
| SLC27A5 | 2.431095 | -0.05313 | 0.999532 | | 2.336551 | -0.3697 | 0.823365 | | 2.124308 | -0.30699 | 0.872493 |
| PARP16 | 3.507185 | -0.00352 | 0.999532 | | 3.768259 | -0.31072 | 0.823365 | | 3.175504 | -0.28307 | 0.852152 |
| OR51S1 | 2.053653 | -0.21383 | 0.999532 | | 2.244032 | -0.39581 | 0.823951 | | 1.77352 | -0.16435 | 0.944229 |
| ZNF184 | 2.186146 | 0.297647 | 0.999532 | | 2.0947 | -0.38731 | 0.824159 | | 1.938889 | -0.66822 | 0.649779 |
| ANTKMT | 6.039791 | 0.180731 | 0.999532 | | 5.811527 | -0.24832 | 0.824159 | | 5.400107 | -0.41267 | 0.666566 |
| FCER1G | 4.459194 | -0.45043 | 0.999532 | | 4.875628 | -0.27074 | 0.824535 | | 3.834358 | 0.194476 | 0.894612 |
| CLUAP1 | 2.588172 | -0.0095 | 0.999532 | | 3.178468 | 0.3092 | 0.824535 | | 2.711216 | 0.333254 | 0.824012 |
| PLEKHG2 | 5.712145 | -0.20026 | 0.999532 | | 7.523681 | 0.215582 | 0.824535 | | 5.966543 | 0.434236 | 0.629345 |
| FBXL7 | 2.593158 | 0.298491 | 0.999532 | | 2.723967 | 0.330757 | 0.824535 | | 2.71368 | 0.056858 | 0.979996 |
| JPT2 | 3.057523 | 0.221066 | 0.999532 | | 3.380498 | 0.304953 | 0.824535 | | 3.272988 | 0.101802 | 0.956322 |
| BTBD1 | 4.934631 | -0.08916 | 0.999532 | | 6.369892 | 0.247147 | 0.824535 | | 5.404614 | 0.349944 | 0.739496 |
| XRCC4 | 2.669048 | 0.481236 | 0.999532 | | 3.009704 | 0.345904 | 0.824535 | | 2.87463 | -0.11309 | 0.954224 |
| SOST | 10.61425 | -0.14044 | 0.999532 | | 13.00798 | 0.197233 | 0.824535 | | 10.8529 | 0.372429 | 0.660958 |
| PEAK3 | 3.006036 | 0.246285 | 0.999532 | | 2.640588 | -0.35076 | 0.824535 | | 2.650814 | -0.56903 | 0.666566 |
| AMBP | 1.940481 | 0.082941 | 0.999532 | | 1.9113 | -0.39851 | 0.824535 | | 1.722448 | -0.46027 | 0.795928 |
| DEXI | 6.804892 | 0.029831 | 0.999532 | | 6.945857 | -0.22625 | 0.824535 | | 5.999756 | -0.23675 | 0.816775 |
| ZNF689 | 2.057073 | -0.12178 | 0.999532 | | 2.005125 | -0.38722 | 0.824535 | | 1.689517 | -0.25696 | 0.901813 |
| PRR18 | 2.356609 | -0.13591 | 0.999532 | | 2.449976 | -0.36098 | 0.824535 | | 2.050691 | -0.20776 | 0.920181 |
| E2F6 | 7.08272 | 0.30691 | 0.999532 | | 6.470817 | -0.28248 | 0.824535 | | 6.390146 | -0.59305 | 0.552672 |
| RNF220 | 11.90022 | 0.391241 | 0.999532 | | 12.63811 | 0.171575 | 0.824535 | | 11.94663 | -0.19991 | 0.803301 |
| GEMIN5 | 2.651946 | 0.338097 | 0.999532 | | 2.347645 | -0.35717 | 0.824535 | | 2.362605 | -0.67672 | 0.62216 |
| GABPB1 | 2.555548 | -0.09531 | 0.999532 | | 2.779414 | -0.34496 | 0.824535 | | 2.338603 | -0.23097 | 0.899161 |
| ASIC1 | 2.115035 | 0.243513 | 0.999532 | | 1.798076 | -0.40839 | 0.824535 | | 1.878831 | -0.64459 | 0.699971 |
| ACBD5 | 3.366974 | -0.16845 | 0.999532 | | 3.814988 | -0.27627 | 0.824535 | | 2.981018 | -0.08359 | 0.965619 |
| ZSWIM4 | 2.127274 | 0.066356 | 0.999532 | | 2.666818 | 0.353892 | 0.824535 | | 2.454228 | 0.301789 | 0.866245 |
| TMBIM4 | 9.471153 | 0.123905 | 0.999532 | | 10.45144 | 0.199415 | 0.824535 | | 9.516506 | 0.083684 | 0.944202 |
| DHX8 | 4.466808 | -0.05388 | 0.999532 | | 5.497417 | 0.247291 | 0.824535 | | 4.591803 | 0.318752 | 0.774747 |
| ZBTB47 | 3.501463 | 0.715105 | 0.999532 | | 3.175583 | 0.303915 | 0.824535 | | 3.697434 | -0.39527 | 0.737426 |
| SESN1 | 2.307346 | 0.396799 | 0.999532 | | 2.004244 | -0.38716 | 0.824535 | | 1.973394 | -0.76447 | 0.598177 |
| CNTROB | 2.61395 | -0.255 | 0.999532 | | 2.926141 | -0.32776 | 0.824535 | | 2.286445 | -0.05296 | 0.984581 |
| RTKN | 1.906746 | 0.159965 | 0.999532 | | 2.370375 | 0.351329 | 0.824535 | | 2.162709 | 0.207961 | 0.908891 |
| FAAP20 | 5.9341 | -0.14451 | 0.999532 | | 7.348453 | 0.214941 | 0.824535 | | 5.939916 | 0.377248 | 0.668158 |
| CWC15 | 4.208117 | -0.01963 | 0.999532 | | 4.452582 | -0.27362 | 0.824535 | | 3.578484 | -0.23402 | 0.864245 |
| ANKAR | 2.796868 | -0.24452 | 0.999532 | | 3.008829 | -0.32045 | 0.824535 | | 2.350314 | -0.06719 | 0.978004 |
| POU3F4 | 2.725201 | 0.519423 | 0.999532 | | 2.171 | -0.37496 | 0.824535 | | 2.469029 | -0.87986 | 0.515762 |
| ENTPD4 | 6.330819 | -0.07085 | 0.999532 | | 7.639879 | 0.219248 | 0.824535 | | 6.293757 | 0.293919 | 0.794473 |
| TSR1 | 2.599354 | -0.03881 | 0.999532 | | 2.759839 | -0.33423 | 0.824535 | | 2.256616 | -0.26513 | 0.881771 |
| UTS2R | 1.963978 | -0.09436 | 0.999532 | | 1.99342 | -0.38452 | 0.824535 | | 1.683301 | -0.27369 | 0.89433 |
| ATPAF1 | 2.62228 | -0.12237 | 0.999532 | | 3.347682 | 0.298726 | 0.824535 | | 2.716008 | 0.436438 | 0.740093 |
| CRLS1 | 2.913112 | 0.406511 | 0.999532 | | 3.118467 | 0.313231 | 0.824535 | | 3.08401 | -0.07647 | 0.971387 |
| RABEP2 | 4.795188 | 0.000406 | 0.999532 | | 5.962432 | 0.237204 | 0.824535 | | 5.031388 | 0.251889 | 0.823571 |
| MXRA7 | 12.19093 | 0.140079 | 0.999532 | | 12.59357 | -0.17263 | 0.824535 | | 11.18013 | -0.29149 | 0.680564 |
| PDCL3 | 4.926917 | 0.34599 | 0.999532 | | 5.403483 | 0.272687 | 0.824535 | | 5.346239 | -0.07214 | 0.965721 |
| PREB | 4.577786 | 0.205892 | 0.999532 | | 4.536385 | -0.272 | 0.824535 | | 3.887211 | -0.46568 | 0.661154 |
| IFNA8 | 2.204039 | -0.02367 | 0.999532 | | 2.198746 | -0.38465 | 0.824535 | | 1.999676 | -0.3448 | 0.861837 |
| SIAH2 | 5.037018 | 0.076454 | 0.999532 | | 5.849396 | 0.23383 | 0.824535 | | 5.20573 | 0.179398 | 0.897958 |
| SH3PXD2 | 4.366587 | 0.079074 | 0.999532 | | 5.149261 | 0.269037 | 0.824535 | | 4.56557 | 0.210229 | 0.869224 |
| SLC25A4 | 6.020538 | -0.35177 | 0.999532 | | 8.495915 | 0.205832 | 0.824535 | | 6.327221 | 0.585298 | 0.456849 |
| ABHD6 | 2.275646 | 0.327274 | 0.999532 | | 2.480129 | 0.341675 | 0.824535 | | 2.43308 | 0.027956 | 0.992459 |
| DHCR7 | 3.687145 | -0.00175 | 0.999532 | | 3.901776 | -0.30624 | 0.824535 | | 3.211207 | -0.27479 | 0.857886 |
| NOL10 | 3.459159 | -0.15541 | 0.999532 | | 4.115775 | 0.282676 | 0.824535 | | 3.403119 | 0.45466 | 0.710512 |
| PHOSPHO | 3.507099 | 0.075209 | 0.999532 | | 3.51368 | -0.31678 | 0.824535 | | 3.064819 | -0.3723 | 0.792538 |
| RBPJ | 5.905215 | 0.142634 | 0.999532 | | 7.24533 | 0.236989 | 0.824535 | | 6.245836 | 0.125004 | 0.922679 |
| VPS72 | 3.285512 | -0.12123 | 0.999532 | | 3.490087 | -0.28922 | 0.824535 | | 2.821723 | -0.14852 | 0.927668 |
| PBX2 | 11.86814 | 0.033383 | 0.999532 | | 14.237 | 0.190387 | 0.824535 | | 12.35592 | 0.178028 | 0.829716 |
| ADCYAP1 | 2.041065 | 0.065871 | 0.999532 | | 2.084185 | -0.40012 | 0.824535 | | 1.82708 | -0.44623 | 0.806394 |
| C7orf61 | 3.17015 | 0.37753 | 0.999532 | | 2.730261 | -0.3586 | 0.824535 | | 2.826288 | -0.71818 | 0.574633 |
| SFRP1 | 3.755489 | -0.25824 | 0.999532 | | 4.311493 | -0.30136 | 0.824535 | | 3.389311 | -0.0299 | 0.991273 |
| A4GALT | 5.386045 | 0.149271 | 0.999532 | | 6.058816 | 0.249068 | 0.824535 | | 5.473486 | 0.118469 | 0.931691 |
| DKK2 | 1.804565 | 1.039283 | 0.999532 | | 1.72262 | 0.433152 | 0.824535 | | 2.006481 | -0.58262 | 0.713076 |
| ABHD14B | 6.310062 | -0.07016 | 0.999532 | | 7.86159 | 0.223589 | 0.824535 | | 6.531558 | 0.315591 | 0.765852 |
| HLA-DQB | 9.579066 | -0.51964 | 0.999532 | | 14.54576 | 0.389329 | 0.824535 | | 10.81159 | 0.988306 | 0.11254 |
| ZFAND6 | 6.20527 | -0.05344 | 0.999532 | | 7.820024 | 0.21017 | 0.824535 | | 6.352601 | 0.285443 | 0.765336 |
| MRPL4 | 5.037709 | 0.22689 | 0.999532 | | 4.848934 | -0.26749 | 0.824535 | | 4.458076 | -0.47462 | 0.649468 |
| BCL2L15 | 2.474931 | 0.244382 | 0.999532 | | 2.287324 | -0.36145 | 0.824535 | | 2.131068 | -0.58783 | 0.686214 |
| UBQLN3 | 2.621228 | -0.13664 | 0.999532 | | 2.731495 | -0.3568 | 0.824535 | | 2.202296 | -0.19999 | 0.918234 |
| RHBDL2 | 3.236273 | 0.115378 | 0.999532 | | 3.234027 | -0.3352 | 0.824535 | | 2.813696 | -0.43051 | 0.760955 |
| IRF3 | 12.96269 | -0.18245 | 0.999532 | | 14.52472 | -0.16853 | 0.824535 | | 11.47286 | 0.03417 | 0.977541 |
| ZFP41 | 4.037827 | -0.21035 | 0.999532 | | 4.490854 | -0.25955 | 0.824535 | | 3.489695 | -0.03177 | 0.989646 |
| RAC2 | 3.8785 | 0.150137 | 0.999532 | | 3.655628 | -0.28806 | 0.824535 | | 3.498492 | -0.42054 | 0.715437 |
| CAPN10 | 2.673622 | 0.398662 | 0.999532 | | 3.034233 | 0.30558 | 0.824535 | | 2.885578 | -0.07332 | 0.971732 |
| TNFRSF1 | 3.696387 | -0.34792 | 0.999532 | | 5.057005 | 0.243545 | 0.824535 | | 3.844303 | 0.609456 | 0.533876 |
| ESCO1 | 2.121894 | 0.095895 | 0.999532 | | 2.608233 | 0.338776 | 0.824535 | | 2.272839 | 0.260642 | 0.881837 |
| MID1IP1 | 4.876652 | -0.10644 | 0.999532 | | 6.473735 | 0.224565 | 0.824535 | | 5.312177 | 0.352832 | 0.718643 |
| BECN2 | 2.154115 | -0.08609 | 0.999532 | | 2.214806 | -0.36483 | 0.824535 | | 1.850164 | -0.26093 | 0.892869 |
| ENGASE | 3.799959 | 0.617334 | 0.999532 | | 3.593597 | 0.296412 | 0.824535 | | 3.838326 | -0.29472 | 0.806394 |
| RNF38 | 3.6887 | 0.010361 | 0.999532 | | 4.405101 | 0.264191 | 0.824535 | | 3.832677 | 0.271607 | 0.834627 |
| EVC | 9.044295 | 0.615641 | 0.999532 | | 8.861536 | 0.242236 | 0.824535 | | 8.987759 | -0.33173 | 0.722703 |
| SV2A | 2.231998 | 0.425656 | 0.999532 | | 1.942703 | -0.40004 | 0.824535 | | 2.01733 | -0.80989 | 0.568812 |
| HCRTR2 | 2.127199 | -0.21044 | 0.999532 | | 2.273791 | -0.37657 | 0.824535 | | 1.80419 | -0.14655 | 0.948099 |
| SRSF2 | 11.45119 | -0.03426 | 0.999532 | | 13.13654 | 0.172005 | 0.824535 | | 11.34736 | 0.209902 | 0.79686 |
| TRIM72 | 4.836169 | 0.405402 | 0.999532 | | 4.564873 | -0.29833 | 0.824535 | | 4.351558 | -0.67185 | 0.498401 |
| ZNF213 | 3.322851 | -0.17146 | 0.999532 | | 3.73001 | -0.28544 | 0.824535 | | 2.885196 | -0.09765 | 0.956322 |
| AZIN2 | 3.270275 | 0.820676 | 0.999532 | | 3.120672 | 0.332448 | 0.824535 | | 3.499644 | -0.46721 | 0.704316 |
| LAS1L | 3.545092 | -0.27843 | 0.999532 | | 4.842463 | 0.266974 | 0.824535 | | 3.806634 | 0.565569 | 0.619423 |
| DDIT4L | 2.42412 | -0.1925 | 0.999532 | | 2.544732 | -0.36075 | 0.824535 | | 1.977352 | -0.15324 | 0.941197 |
| DIRAS1 | 2.875786 | 0.49777 | 0.999532 | | 3.203209 | 0.338943 | 0.824535 | | 3.11356 | -0.1402 | 0.938429 |
| OSGEPL1 | 2.214432 | 0.041263 | 0.999532 | | 2.074583 | -0.36647 | 0.824535 | | 1.922107 | -0.39276 | 0.816775 |
| ZFP1 | 4.987139 | 0.112563 | 0.999532 | | 4.996639 | -0.27586 | 0.824535 | | 4.643538 | -0.38588 | 0.736481 |
| KRTAP10 | 3.700074 | 0.304163 | 0.999532 | | 3.382376 | -0.31481 | 0.824535 | | 3.28345 | -0.59878 | 0.632113 |
| KRTCAP2 | 13.25709 | 0.035816 | 0.999532 | | 15.85997 | 0.168766 | 0.824535 | | 13.50071 | 0.152976 | 0.843463 |
| ZNF592 | 3.301539 | -0.05541 | 0.999532 | | 3.906816 | 0.271493 | 0.824535 | | 3.4417 | 0.339919 | 0.796702 |
| OR2Z1 | 2.232749 | 0.078931 | 0.999532 | | 1.97212 | -0.38826 | 0.824535 | | 1.895575 | -0.45682 | 0.79686 |
| RAD17 | 4.858464 | -0.27574 | 0.999532 | | 5.56898 | -0.24931 | 0.824535 | | 4.178988 | 0.045032 | 0.979967 |
| ATXN1L | 5.45782 | 0.109996 | 0.999532 | | 6.304065 | 0.240896 | 0.824535 | | 5.593484 | 0.151332 | 0.903329 |
| LRP5 | 3.288832 | 0.253975 | 0.999532 | | 3.709576 | 0.314762 | 0.824535 | | 3.472109 | 0.074281 | 0.971732 |
| CEL | 5.414734 | 0.417844 | 0.999532 | | 4.940135 | -0.27644 | 0.824535 | | 4.904716 | -0.677 | 0.454223 |
| SUSD2 | 2.316031 | 0.730171 | 0.999532 | | 2.177063 | 0.361378 | 0.824535 | | 2.411411 | -0.35089 | 0.816618 |
| SP4 | 3.642055 | -0.1071 | 0.999532 | | 3.92141 | -0.27561 | 0.824535 | | 3.065849 | -0.14571 | 0.92697 |
| RAD21 | 7.336924 | -0.17427 | 0.999532 | | 9.184435 | 0.206874 | 0.824535 | | 7.345206 | 0.39882 | 0.652925 |
| BBS2 | 4.722743 | -0.04215 | 0.999532 | | 6.135965 | 0.225857 | 0.824535 | | 4.854768 | 0.292092 | 0.790999 |
| SHQ1 | 3.583103 | 0.294852 | 0.999532 | | 4.13037 | 0.275432 | 0.824535 | | 3.844124 | 8.86E-05 | 0.999949 |
| SLC38A6 | 2.694059 | 0.301849 | 0.999532 | | 2.971681 | 0.318336 | 0.824535 | | 2.787142 | 0.027769 | 0.992286 |
| MTRES1 | 2.886289 | 0.604604 | 0.999532 | | 3.046212 | 0.314878 | 0.824535 | | 3.047256 | -0.27873 | 0.839402 |
| FOXP3 | 2.169261 | 0.117439 | 0.999532 | | 2.140671 | -0.37579 | 0.824535 | | 1.895246 | -0.47435 | 0.769793 |
| SPEG | 1.974502 | 0.293598 | 0.999532 | | 1.742994 | -0.41433 | 0.824535 | | 1.72526 | -0.69106 | 0.667066 |
| ADH5 | 6.732593 | 0.183048 | 0.999532 | | 7.735117 | 0.241995 | 0.824535 | | 6.797112 | 0.092511 | 0.946572 |
| DTX3 | 7.681997 | 0.141219 | 0.999532 | | 9.105272 | 0.197171 | 0.824535 | | 7.901009 | 0.089145 | 0.952195 |
| VCAN | 3.684653 | 0.004301 | 0.999532 | | 3.755206 | -0.28738 | 0.824535 | | 3.026422 | -0.2767 | 0.855083 |
| SAPCD2 | 2.997405 | 0.041555 | 0.999532 | | 2.980777 | -0.34231 | 0.824535 | | 2.74341 | -0.37139 | 0.815713 |
| SEC16B | 2.525026 | 0.116646 | 0.999532 | | 2.342178 | -0.36204 | 0.824535 | | 2.189067 | -0.46835 | 0.767179 |
| NHSL2 | 3.150266 | 0.237674 | 0.999532 | | 2.842495 | -0.33068 | 0.824587 | | 2.738668 | -0.54877 | 0.663104 |
| MSRB3 | 9.182963 | 0.392989 | 0.999532 | | 9.827872 | 0.212142 | 0.824832 | | 9.181725 | -0.15689 | 0.881837 |
| MRPL32 | 4.699981 | -0.18565 | 0.999532 | | 6.250422 | 0.249178 | 0.824905 | | 4.981589 | 0.452087 | 0.660958 |
| HSPA6 | 4.395868 | -0.15203 | 0.999532 | | 5.438605 | 0.253016 | 0.82494 | | 4.314291 | 0.421265 | 0.70375 |
| MAP2K1 | 6.98814 | -0.23573 | 0.999532 | | 7.780956 | -0.2109 | 0.82494 | | 6.358899 | 0.039855 | 0.981069 |
| EPN2 | 6.400959 | -0.18078 | 0.999532 | | 7.390747 | -0.21411 | 0.82494 | | 5.934566 | -0.00504 | 0.997967 |
| KIFBP | 2.638529 | 0.01117 | 0.999532 | | 3.208306 | 0.301748 | 0.824998 | | 2.877262 | 0.307791 | 0.836453 |
| TP53I3 | 2.349972 | 0.119892 | 0.999532 | | 2.706413 | 0.326249 | 0.825098 | | 2.368215 | 0.215279 | 0.90234 |
| GMNN | 2.29716 | 0.271814 | 0.999532 | | 2.428851 | 0.342338 | 0.825107 | | 2.42173 | 0.085489 | 0.972135 |
| AMER2 | 2.266816 | 0.062269 | 0.999532 | | 2.384014 | -0.36152 | 0.825157 | | 1.993077 | -0.40251 | 0.806797 |
| OR6N2 | 1.991732 | -0.09131 | 0.999532 | | 1.973716 | -0.38613 | 0.825157 | | 1.742727 | -0.27825 | 0.892869 |
| GSDMA | 3.34058 | 0.944171 | 0.999532 | | 3.048237 | 0.334983 | 0.825599 | | 3.588626 | -0.60103 | 0.584522 |
| IL33 | 1.896178 | 0.675818 | 0.999532 | | 1.951047 | 0.394105 | 0.825611 | | 2.130849 | -0.26111 | 0.88885 |
| ZNF71 | 2.46819 | 1.03837 | 0.999532 | | 2.193543 | 0.378315 | 0.825611 | | 2.699901 | -0.64377 | 0.630278 |
| PPM1J | 2.829914 | 0.147751 | 0.999532 | | 2.664838 | -0.34839 | 0.825611 | | 2.401124 | -0.47762 | 0.745881 |
| SMIM40 | 4.716607 | 0.243807 | 0.999532 | | 4.42554 | -0.31423 | 0.825702 | | 4.288461 | -0.56854 | 0.644433 |
| PRKAR2B | 6.193533 | 0.086239 | 0.999532 | | 6.133417 | -0.25441 | 0.826116 | | 5.333806 | -0.31987 | 0.765852 |
| ZNF423 | 3.132891 | 0.139731 | 0.999532 | | 3.738914 | 0.282977 | 0.826171 | | 3.332887 | 0.161291 | 0.915533 |
| OR5AR1 | 1.92182 | -0.01058 | 0.999532 | | 1.831418 | -0.40705 | 0.826171 | | 1.636782 | -0.37827 | 0.851785 |
| WDR35 | 2.597269 | 0.199153 | 0.999532 | | 2.983296 | 0.338053 | 0.826171 | | 2.706488 | 0.15742 | 0.931619 |
| LMTK2 | 3.60995 | 0.366269 | 0.999532 | | 3.210253 | -0.31354 | 0.826171 | | 3.094329 | -0.65896 | 0.552842 |
| NGB | 1.722693 | 0.269636 | 0.999532 | | 2.129032 | 0.398028 | 0.826171 | | 1.961006 | 0.149053 | 0.948127 |
| CDO1 | 2.314556 | -0.50372 | 0.999532 | | 2.708303 | -0.32984 | 0.826171 | | 1.908202 | 0.192768 | 0.923834 |
| PEMT | 2.903264 | 0.338653 | 0.999532 | | 3.200644 | 0.306624 | 0.826171 | | 2.981082 | -0.00665 | 0.997967 |
| GTF2F1 | 7.111423 | 0.049613 | 0.999532 | | 8.095044 | 0.211105 | 0.826298 | | 6.968828 | 0.179826 | 0.869224 |
| IRS1 | 2.949761 | 0.285353 | 0.999532 | | 2.464935 | -0.35101 | 0.826298 | | 2.469282 | -0.63124 | 0.659727 |
| NPAS3 | 2.702007 | 0.618208 | 0.999532 | | 2.383847 | -0.37919 | 0.826298 | | 2.318303 | -0.97647 | 0.399012 |
| MRGPRD | 3.856124 | -0.23709 | 0.999532 | | 4.405461 | -0.29916 | 0.826298 | | 3.504461 | -0.05019 | 0.982187 |
| HOXD9 | 2.567282 | 0.05727 | 0.999532 | | 3.189996 | 0.314538 | 0.826298 | | 2.569071 | 0.275726 | 0.859898 |
| ATP2B4 | 6.158755 | -0.17392 | 0.999532 | | 8.218386 | 0.203745 | 0.826298 | | 6.514909 | 0.408015 | 0.660097 |
| SH3GLB2 | 5.43611 | 0.123967 | 0.999532 | | 6.444173 | 0.225805 | 0.826298 | | 5.400146 | 0.12581 | 0.9195 |
| ABI2 | 5.825562 | 0.328394 | 0.999532 | | 6.097878 | 0.22413 | 0.826298 | | 6.008258 | -0.08355 | 0.947557 |
| RTL5 | 2.194889 | 0.142115 | 0.999532 | | 2.770745 | 0.329106 | 0.826298 | | 2.381351 | 0.213724 | 0.90424 |
| TYW1B | 2.186583 | 0.175131 | 0.999532 | | 2.028835 | -0.37731 | 0.826298 | | 1.888329 | -0.53472 | 0.734322 |
| PDGFRA | 6.387876 | -0.13213 | 0.999532 | | 8.065494 | 0.231426 | 0.8263 | | 6.649322 | 0.385749 | 0.668481 |
| SLC25A1 | 2.788291 | 0.382563 | 0.999532 | | 2.831103 | 0.324678 | 0.8263 | | 2.853567 | -0.04223 | 0.987274 |
| INSIG1 | 2.75831 | 0.135715 | 0.999532 | | 3.530462 | 0.29485 | 0.8263 | | 3.205101 | 0.184108 | 0.907271 |
| DPPA2 | 3.440914 | 0.237236 | 0.999532 | | 3.261527 | -0.3136 | 0.8263 | | 3.092875 | -0.53878 | 0.664512 |
| CSK | 8.507873 | 0.072367 | 0.999532 | | 9.505464 | 0.194555 | 0.826567 | | 8.667494 | 0.136493 | 0.895865 |
| FHL5 | 2.245616 | 0.352635 | 0.999532 | | 2.567226 | 0.341247 | 0.826567 | | 2.463468 | 0.007053 | 0.997967 |
| KPNA1 | 2.983666 | -0.41945 | 0.999532 | | 3.366069 | -0.30634 | 0.826567 | | 2.439778 | 0.127625 | 0.948127 |
| TTC9 | 2.757433 | -0.25747 | 0.999532 | | 2.780374 | -0.33857 | 0.826567 | | 2.278208 | -0.05813 | 0.982435 |
| PAFAH1B | 2.321764 | 0.816866 | 0.999532 | | 2.02065 | 0.381457 | 0.826567 | | 2.35229 | -0.42474 | 0.791466 |
| DCAF16 | 5.9509 | 0.032057 | 0.999532 | | 7.315405 | 0.216747 | 0.826567 | | 6.27658 | 0.206543 | 0.856551 |
| CLEC11A | 9.388999 | 0.078862 | 0.999532 | | 11.12046 | 0.197286 | 0.826567 | | 9.868688 | 0.14704 | 0.889126 |
| SP5 | 2.751865 | -0.4212 | 0.999532 | | 3.324538 | -0.32699 | 0.826567 | | 2.430212 | 0.122359 | 0.955167 |
| TMEM170 | 2.628496 | 0.01784 | 0.999532 | | 2.661927 | -0.34909 | 0.826567 | | 2.314154 | -0.34385 | 0.84246 |
| TPPP2 | 2.826867 | 0.451788 | 0.999532 | | 2.463332 | -0.36425 | 0.826567 | | 2.488103 | -0.79748 | 0.532278 |
| NUP50 | 5.434392 | -0.34165 | 0.999532 | | 6.377172 | -0.22849 | 0.826567 | | 4.75941 | 0.131278 | 0.922679 |
| TAF8 | 4.109591 | -0.15766 | 0.999532 | | 5.158547 | 0.254276 | 0.826584 | | 4.411641 | 0.43014 | 0.701251 |
| PKP2 | 2.182002 | 0.025366 | 0.999532 | | 2.236013 | -0.3531 | 0.826584 | | 1.881524 | -0.35817 | 0.836935 |
| DNAAF4 | 2.227432 | 0.171221 | 0.999532 | | 2.077817 | -0.37857 | 0.826584 | | 1.940288 | -0.53437 | 0.738861 |
| SEMA3A | 2.325558 | 0.246337 | 0.999532 | | 2.112738 | -0.36508 | 0.826584 | | 2.022015 | -0.59451 | 0.685366 |
| GABRR1 | 2.231779 | 0.31945 | 0.999532 | | 1.990794 | -0.38532 | 0.826584 | | 1.934851 | -0.68735 | 0.652925 |
| ACKR4 | 2.26333 | 0.359953 | 0.999532 | | 2.090639 | -0.39273 | 0.826584 | | 2.029193 | -0.73355 | 0.630705 |

| | | | | | | | | | | | | |
|---|---|---|---|---|---|---|---|---|---|---|---|---|
| PRPSAP1 | 4.422073 | -0.50381 | 0.999532 | | 5.371159 | -0.28554 | 0.826584 | | 3.949261 | 0.229721 | 0.872435 | |
| GATA3 | 11.66712 | 0.271493 | 0.999532 | | 12.87746 | 0.206197 | 0.826584 | | 12.32367 | -0.04637 | 0.972221 | |
| CKAP2 | 2.203518 | -0.0199 | 0.999532 | | 2.418225 | -0.37835 | 0.826584 | | 2.03351 | -0.33103 | 0.862823 | |
| SLC66A3 | 3.605564 | 0.078749 | 0.999532 | | 4.200283 | 0.278366 | 0.826624 | | 3.873678 | 0.21631 | 0.876781 | |
| ACSL4 | 8.338844 | -0.1221 | 0.999532 | | 10.69403 | 0.231542 | 0.827019 | | 8.814221 | 0.370578 | 0.675955 | |
| IZUMO1 | 2.726437 | 0.326666 | 0.999532 | | 2.362971 | -0.38105 | 0.827019 | | 2.399032 | -0.69755 | 0.646939 | |
| FAM47A | 2.069511 | 0.114035 | 0.999532 | | 2.114681 | -0.38601 | 0.827019 | | 1.771961 | -0.47714 | 0.78258 | |
| LPGAT1 | 2.738878 | -0.24049 | 0.999532 | | 3.554659 | 0.302337 | 0.827019 | | 3.041325 | 0.542865 | 0.698562 | |
| SMIM27 | 3.032265 | -0.0792 | 0.999532 | | 3.087707 | -0.30384 | 0.827053 | | 2.685108 | -0.20748 | 0.898369 | |
| RALGAPB | 2.495371 | -0.06641 | 0.999532 | | 3.320694 | 0.297384 | 0.827053 | | 2.760044 | 0.383329 | 0.784263 | |
| VAMP1 | 4.056444 | 0.301561 | 0.999532 | | 4.496417 | 0.257206 | 0.827703 | | 4.34487 | -0.02374 | 0.99176 | |
| OR52N1 | 1.995685 | -0.26559 | 0.999532 | | 2.099877 | -0.36793 | 0.827844 | | 1.679103 | -0.08686 | 0.974421 | |
| DIRC1 | 1.955935 | 0.403657 | 0.999532 | | 1.722894 | -0.4172 | 0.827844 | | 1.725708 | -0.80077 | 0.617899 | |
| TRIM16L | 2.901998 | 0.178241 | 0.999532 | | 2.685205 | -0.33563 | 0.827844 | | 2.570193 | -0.49461 | 0.737952 | |
| ACTA1 | 2.166402 | 0.942964 | 0.999532 | | 2.048386 | 0.388218 | 0.827844 | | 2.326335 | -0.53523 | 0.713173 | |
| EFNA2 | 2.775272 | 0.267192 | 0.999532 | | 2.434851 | -0.35709 | 0.827844 | | 2.395316 | -0.61073 | 0.662584 | |
| PDSS1 | 2.99306 | 0.389089 | 0.999532 | | 2.791032 | -0.3338 | 0.827844 | | 2.618084 | -0.70153 | 0.563627 | |
| CTAGE9 | 4.758077 | 0.063027 | 0.999532 | | 4.790398 | -0.28752 | 0.827844 | | 4.174943 | -0.33505 | 0.797186 | |
| HLA-DQA | 3.604325 | 0.466604 | 0.999532 | | 4.245088 | 0.370181 | 0.827844 | | 4.162523 | -0.10337 | 0.961451 | |
| NUP35 | 3.044659 | 0.032923 | 0.999532 | | 2.955485 | -0.32 | 0.827844 | | 2.726308 | -0.3362 | 0.836012 | |
| LTA4H | 3.337505 | -0.17378 | 0.999532 | | 4.140857 | 0.263582 | 0.827844 | | 3.487113 | 0.451059 | 0.699971 | |
| OR2A7 | 3.204103 | 0.236132 | 0.999532 | | 2.959827 | -0.33917 | 0.827844 | | 2.789304 | -0.55726 | 0.672739 | |
| CYRIB | 2.763562 | -0.47509 | 0.999532 | | 3.126074 | -0.32673 | 0.827844 | | 2.223991 | 0.166101 | 0.936689 | |
| SCAMP1 | 7.105209 | 0.304911 | 0.999532 | | 7.412846 | 0.205605 | 0.827844 | | 7.091736 | -0.08387 | 0.944635 | |
| SNRNP20 | 4.412824 | -0.29357 | 0.999532 | | 5.678048 | 0.235278 | 0.827844 | | 4.395491 | 0.532035 | 0.616301 | |
| RENBP | 4.404245 | -0.26297 | 0.999532 | | 4.769214 | -0.26367 | 0.827844 | | 3.766795 | 0.007269 | 0.997967 | |
| PTGDR2 | 2.708111 | -0.15884 | 0.999532 | | 2.90784 | -0.32596 | 0.828086 | | 2.380067 | -0.14847 | 0.940162 | |
| H4C2 | 4.452094 | 0.112595 | 0.999532 | | 5.172882 | 0.239085 | 0.828095 | | 4.477358 | 0.145362 | 0.910105 | |
| POLR3E | 2.585989 | 0.025964 | 0.999532 | | 3.495343 | 0.288858 | 0.828095 | | 2.964313 | 0.286423 | 0.843066 | |
| FAM53B | 5.12077 | -0.06778 | 0.999532 | | 6.243205 | 0.230679 | 0.828095 | | 5.218881 | 0.320299 | 0.749206 | |
| TPM3 | 16.77804 | -0.14575 | 0.999532 | | 21.27165 | 0.163926 | 0.828095 | | 17.2697 | 0.334786 | 0.618451 | |
| FAM131A | 2.552256 | 0.374385 | 0.999532 | | 2.287356 | -0.36052 | 0.828095 | | 2.308249 | -0.71801 | 0.598329 | |
| FSBP | 2.044165 | 0.452867 | 0.999532 | | 2.019607 | 0.373996 | 0.828095 | | 2.115419 | -0.06833 | 0.97843 | |
| ZC3H7A | 7.536647 | 0.148463 | 0.999532 | | 8.746121 | 0.209608 | 0.828287 | | 7.886266 | 0.079714 | 0.947941 | |
| GTF2IRD1 | 3.231382 | 0.252667 | 0.999532 | | 3.552865 | 0.295136 | 0.828287 | | 3.360416 | 0.063484 | 0.974421 | |
| GIN1 | 2.834852 | -0.13734 | 0.999532 | | 2.781222 | -0.3395 | 0.828287 | | 2.263357 | -0.18303 | 0.924292 | |
| ACR | 3.687135 | 0.408531 | 0.999532 | | 3.185769 | -0.31302 | 0.828287 | | 3.284252 | -0.70484 | 0.545883 | |
| NFE2L2 | 4.537068 | 0.107063 | 0.999532 | | 5.288402 | 0.267057 | 0.828287 | | 4.530638 | 0.174139 | 0.908891 | |
| MRPL46 | 2.57201 | 0.688222 | 0.999532 | | 2.471031 | 0.330266 | 0.828287 | | 2.644432 | -0.33989 | 0.809832 | |
| SRRM5 | 1.999125 | 0.694773 | 0.999532 | | 2.076519 | 0.378152 | 0.828361 | | 2.229052 | -0.29568 | 0.867769 | |
| RAX2 | 2.896214 | 0.664124 | 0.999532 | | 2.413995 | -0.37116 | 0.828361 | | 2.562187 | -1.01823 | 0.359441 | |
| ZNF580 | 5.238526 | -0.16236 | 0.999532 | | 5.58901 | -0.22764 | 0.828361 | | 4.446478 | -0.05318 | 0.975194 | |
| NSMCE4A | 2.460784 | 0.04391 | 0.999532 | | 3.059348 | 0.299762 | 0.828361 | | 2.652459 | 0.274237 | 0.858705 | |
| TMEM102 | 3.307487 | 0.220042 | 0.999532 | | 3.046984 | -0.32431 | 0.828361 | | 2.944896 | -0.5417 | 0.668481 | |
| CD300A | 2.409619 | 0.123801 | 0.999532 | | 2.844687 | 0.329181 | 0.828361 | | 2.597331 | 0.232155 | 0.892869 | |
| KIF2A | 1.958 | -0.07189 | 0.999532 | | 2.446843 | 0.338435 | 0.828361 | | 2.152091 | 0.427118 | 0.791576 | |
| TMEM42 | 3.862203 | 0.278835 | 0.999532 | | 3.504691 | -0.28749 | 0.828361 | | 3.523111 | -0.54497 | 0.636913 | |
| INSR | 4.227732 | 0.209511 | 0.999532 | | 4.982778 | 0.254974 | 0.828361 | | 4.379096 | 0.068629 | 0.969783 | |
| TRIM74 | 2.863009 | 0.262385 | 0.999532 | | 2.906089 | -0.3552 | 0.828361 | | 2.481776 | -0.58889 | 0.66478 | |
| ABHD13 | 2.354468 | 0.040175 | 0.999532 | | 3.032197 | 0.314174 | 0.828361 | | 2.557356 | 0.288602 | 0.853577 | |
| TM2D2 | 2.501814 | 0.243768 | 0.999532 | | 2.852956 | 0.307587 | 0.828361 | | 2.630573 | 0.080697 | 0.969693 | |
| CDC42EP | 8.683818 | 0.222337 | 0.999532 | | 9.675018 | 0.190841 | 0.828361 | | 8.822596 | -0.01225 | 0.995304 | |
| TMED5 | 4.989307 | -0.08754 | 0.999532 | | 6.177556 | 0.229076 | 0.828446 | | 5.13524 | 0.335293 | 0.737952 | |
| RTP1 | 2.64997 | -0.08997 | 0.999532 | | 2.943729 | -0.32024 | 0.828574 | | 2.204687 | -0.20822 | 0.905952 | |
| SCN2A | 2.124339 | 0.05398 | 0.999532 | | 2.143854 | -0.38155 | 0.82867 | | 1.809994 | -0.41809 | 0.811123 | |
| ENKUR | 2.534408 | -0.00996 | 0.999532 | | 2.755829 | -0.359 | 0.828972 | | 2.269713 | -0.32218 | 0.861119 | |
| VPS25 | 3.550365 | 0.422669 | 0.999532 | | 3.911604 | 0.294003 | 0.828972 | | 3.931196 | -0.10694 | 0.953949 | |
| ZCWPW1 | 2.691401 | -0.04911 | 0.999532 | | 2.893956 | -0.33506 | 0.828972 | | 2.495995 | -0.26847 | 0.87813 | |
| HGS | 5.269082 | 0.074127 | 0.999532 | | 6.444658 | 0.226345 | 0.828972 | | 5.565072 | 0.171555 | 0.883819 | |
| GPR176 | 2.754357 | 0.445324 | 0.999532 | | 2.87382 | 0.331618 | 0.828972 | | 2.997242 | -0.11147 | 0.955834 | |
| DPY19L2 | 5.087308 | -0.14092 | 0.999532 | | 5.704257 | -0.23164 | 0.829001 | | 4.514657 | -0.07063 | 0.965619 | |
| CCDC22 | 3.140802 | 0.319501 | 0.999532 | | 3.243216 | 0.299446 | 0.829001 | | 3.340087 | 0.010751 | 0.996891 | |
| CABYR | 2.899616 | -0.17483 | 0.999532 | | 3.117685 | -0.31766 | 0.829001 | | 2.636388 | -0.12445 | 0.951792 | |
| CYS1 | 6.224036 | 0.583959 | 0.999532 | | 6.080921 | 0.240959 | 0.829001 | | 6.500453 | -0.3275 | 0.746876 | |
| SMIM10L | 3.06847 | 0.348356 | 0.999532 | | 2.762575 | -0.32543 | 0.829001 | | 2.817332 | -0.65447 | 0.600081 | |
| PADI4 | 2.817732 | 0.360508 | 0.999532 | | 2.395907 | -0.36 | 0.829001 | | 2.485048 | -0.70411 | 0.606305 | |
| DENND4A | 3.223486 | 0.424097 | 0.999532 | | 3.515025 | 0.301231 | 0.829114 | | 3.365949 | -0.10281 | 0.955834 | |
| MRGPRE | 4.202741 | -0.09722 | 0.999532 | | 4.328114 | -0.26811 | 0.829114 | | 3.680982 | -0.15122 | 0.921 | |
| HAS1 | 2.452443 | -0.11863 | 0.999532 | | 2.568022 | -0.36074 | 0.829114 | | 2.040822 | -0.22348 | 0.909084 | |
| IL24 | 7.55192 | -0.24605 | 0.999532 | | 10.00478 | 0.2413 | 0.829274 | | 7.827516 | 0.518685 | 0.531575 | |
| IL5RA | 2.443683 | -0.09708 | 0.999532 | | 2.469024 | -0.34143 | 0.829274 | | 2.011097 | -0.2264 | 0.903205 | |
| EXOC5 | 3.095809 | 0.32501 | 0.999532 | | 3.267839 | 0.303664 | 0.829274 | | 3.186297 | -0.00976 | 0.996891 | |
| FASTK | 3.35507 | -0.55409 | 0.999532 | | 4.744954 | 0.267602 | 0.829274 | | 3.336564 | 0.845625 | 0.397004 | |
| STXBP2 | 4.053477 | 0.461653 | 0.999532 | | 4.410411 | 0.255663 | 0.8293 | | 4.273779 | -0.17955 | 0.89268 | |
| ZNF567 | 3.926662 | 0.514088 | 0.999532 | | 3.550576 | -0.30924 | 0.8293 | | 3.501895 | -0.80098 | 0.421517 | |
| NMNAT3 | 2.737637 | 0.271238 | 0.999532 | | 2.591822 | -0.32069 | 0.8293 | | 2.383303 | -0.56661 | 0.67176 | |
| MACROD | 4.809762 | 0.587516 | 0.999532 | | 4.667557 | 0.278631 | 0.8293 | | 5.021919 | -0.31216 | 0.805825 | |
| FKBP1B | 2.846985 | 0.020685 | 0.999532 | | 2.931124 | -0.326 | 0.8293 | | 2.660717 | -0.33298 | 0.837256 | |
| WDR46 | 5.993354 | 0.277215 | 0.999532 | | 6.559697 | 0.22834 | 0.8293 | | 6.311554 | -0.01689 | 0.994617 | |
| UTP20 | 1.890491 | 0.267931 | 0.999532 | | 1.760807 | -0.40862 | 0.8293 | | 1.710547 | -0.66592 | 0.686214 | |
| TRMT1 | 4.242554 | -0.02661 | 0.999532 | | 4.408686 | -0.26236 | 0.8293 | | 3.615035 | -0.21534 | 0.87591 | |
| ZIC1 | 2.530509 | 0.346812 | 0.999532 | | 2.263857 | -0.3714 | 0.8293 | | 2.15369 | -0.69739 | 0.628895 | |
| WEE1 | 3.466814 | -0.10746 | 0.999532 | | 3.641261 | -0.29354 | 0.8293 | | 2.913223 | -0.17072 | 0.917817 | |
| ARID2 | 4.347753 | 0.031132 | 0.999532 | | 4.561607 | -0.27461 | 0.8293 | | 3.800165 | -0.28983 | 0.815713 | |
| TMEM199 | 5.552165 | -0.10661 | 0.999532 | | 5.894111 | -0.23276 | 0.8293 | | 4.870518 | -0.10726 | 0.940744 | |
| ZNF785 | 5.19152 | 0.095027 | 0.999532 | | 5.350373 | -0.24177 | 0.8293 | | 4.641632 | -0.31588 | 0.768974 | |
| KMT2A | 4.228517 | -0.15385 | 0.999532 | | 5.447762 | 0.232984 | 0.8293 | | 4.294969 | 0.402624 | 0.703463 | |
| ZFAND1 | 2.943128 | 0.236691 | 0.999532 | | 3.198598 | 0.307762 | 0.8293 | | 2.904505 | 0.091635 | 0.963492 | |
| TAF6L | 3.292774 | -0.08475 | 0.999532 | | 4.329989 | 0.281844 | 0.8293 | | 3.570579 | 0.387712 | 0.75937 | |
| WWOX | 4.491178 | 0.277124 | 0.999532 | | 5.108461 | 0.266524 | 0.8293 | | 4.829428 | 0.00227 | 0.999385 | |
| PDE4D | 2.388732 | -0.17202 | 0.999532 | | 3.239735 | 0.301861 | 0.8293 | | 2.689184 | 0.490389 | 0.717627 | |
| BRF2 | 2.569457 | 0.375268 | 0.999532 | | 2.929572 | 0.318292 | 0.8293 | | 2.872342 | -0.03767 | 0.989148 | |
| C7orf50 | 3.646565 | 0.32482 | 0.999532 | | 4.247766 | 0.259578 | 0.8293 | | 3.792543 | -0.05 | 0.97843 | |
| CYBRD1 | 5.366069 | -0.08218 | 0.999532 | | 6.723819 | 0.225434 | 0.8293 | | 5.637317 | 0.327093 | 0.753746 | |
| SMAD7 | 6.420687 | -0.20956 | 0.999532 | | 6.908328 | -0.22471 | 0.8293 | | 5.548134 | 0.008065 | 0.996891 | |
| RAVER2 | 3.230693 | -0.15318 | 0.999532 | | 3.439802 | -0.28822 | 0.8293 | | 2.723461 | -0.11855 | 0.94861 | |
| ZC3HAV1 | 5.735489 | -0.09026 | 0.999532 | | 6.234449 | -0.23672 | 0.8293 | | 4.865903 | -0.14005 | 0.909259 | |
| CYB561A | 3.265352 | -0.36497 | 0.999532 | | 4.576353 | 0.259872 | 0.8293 | | 3.357247 | 0.641495 | 0.5451 | |
| LAP3 | 7.496232 | -0.18837 | 0.999532 | | 9.599487 | 0.206923 | 0.8293 | | 7.836612 | 0.422906 | 0.628895 | |
| CDC42BP | 6.446489 | -0.0827 | 0.999532 | | 7.812476 | 0.203138 | 0.829353 | | 6.587108 | 0.300243 | 0.741003 | |
| MARCHF | 1.939855 | -0.45694 | 0.999532 | | 2.233136 | -0.37407 | 0.829353 | | 1.637782 | 0.099811 | 0.972516 | |
| TRIM34 | 2.007492 | 0.655429 | 0.999532 | | 2.088557 | 0.386299 | 0.829353 | | 2.061214 | -0.25055 | 0.892869 | |
| CEP135 | 2.434389 | 0.813412 | 0.999532 | | 2.16918 | 0.364428 | 0.829353 | | 2.491105 | -0.43255 | 0.765852 | |
| MYRF | 2.156485 | 0.049353 | 0.999532 | | 2.825773 | 0.331206 | 0.829353 | | 2.202034 | 0.286284 | 0.865065 | |
| FUNDC2 | 4.915809 | -0.07027 | 0.999532 | | 5.347488 | -0.23516 | 0.829386 | | 4.263482 | -0.1407 | 0.915888 | |
| FSCN1 | 10.55629 | -0.15045 | 0.999532 | | 12.75217 | 0.181619 | 0.829386 | | 10.7306 | 0.343966 | 0.66232 | |
| ZNF621 | 3.669976 | 0.073569 | 0.999532 | | 3.70096 | -0.27989 | 0.829399 | | 3.29417 | -0.33323 | 0.809832 | |
| STXBP1 | 9.947577 | -0.15163 | 0.999532 | | 12.37506 | 0.178263 | 0.829399 | | 10.29393 | 0.343837 | 0.659846 | |
| ERCC6L2 | 2.694906 | 0.051773 | 0.999532 | | 3.234259 | 0.301593 | 0.829399 | | 2.720285 | 0.268044 | 0.863134 | |
| HIGD1A | 14.20115 | 0.015386 | 0.999532 | | 16.90501 | 0.157205 | 0.829489 | | 14.27597 | 0.157501 | 0.836935 | |
| TRAM2 | 7.15365 | 0.354842 | 0.999532 | | 7.591711 | 0.203134 | 0.829559 | | 7.344528 | -0.12591 | 0.91707 | |
| SRRM1 | 5.782058 | 0.02576 | 0.999532 | | 7.104281 | 0.210039 | 0.829607 | | 6.163384 | 0.212424 | 0.851503 | |
| SNX3 | 14.55277 | -0.00387 | 0.999532 | | 17.27737 | 0.167085 | 0.829607 | | 14.77069 | 0.207421 | 0.814884 | |
| ODF3L2 | 2.235088 | 0.367001 | 0.999532 | | 2.607765 | 0.333059 | 0.829607 | | 2.521468 | -0.00985 | 0.996891 | |
| PDP1 | 4.007819 | 0.219203 | 0.999532 | | 4.248767 | 0.270958 | 0.829684 | | 4.123076 | 0.062725 | 0.974421 | |
| SLC17A9 | 2.775818 | 0.558916 | 0.999532 | | 2.401279 | -0.36318 | 0.829769 | | 2.518073 | -0.91105 | 0.438594 | |
| GGCX | 4.489345 | -0.03399 | 0.999532 | | 4.719646 | -0.25508 | 0.829769 | | 3.951456 | -0.2028 | 0.883116 | |
| ZNF714 | 5.807481 | -0.0662 | 0.999532 | | 6.327276 | -0.21934 | 0.829769 | | 5.310846 | -0.13109 | 0.918194 | |
| ACKR2 | 3.680221 | 0.463051 | 0.999532 | | 3.051648 | -0.32964 | 0.829769 | | 3.252334 | -0.76792 | 0.515762 | |
| SC5D | 4.356063 | -0.54086 | 0.999532 | | 5.100936 | -0.25063 | 0.829769 | | 3.696745 | 0.319822 | 0.799944 | |
| ZCCHC14 | 3.813327 | 0.172913 | 0.999532 | | 4.111572 | 0.261381 | 0.829769 | | 3.875325 | 0.099438 | 0.949685 | |
| CYP46A1 | 2.165711 | 0.200245 | 0.999532 | | 2.227519 | -0.37166 | 0.83035 | | 2.014841 | -0.54035 | 0.736481 | |
| FLT4 | 6.778979 | -0.15669 | 0.999532 | | 8.540249 | 0.192548 | 0.830659 | | 6.895361 | 0.367749 | 0.676565 | |
| SLC25A1 | 2.860654 | -0.15533 | 0.999532 | | 3.029993 | -0.30796 | 0.830894 | | 2.517627 | -0.13304 | 0.945129 | |
| SMC4 | 3.880758 | -0.01529 | 0.999532 | | 4.07129 | -0.27377 | 0.830894 | | 3.398911 | -0.24022 | 0.865065 | |
| RUFY3 | 4.928862 | -0.21032 | 0.999532 | | 6.012395 | 0.223142 | 0.830894 | | 4.892831 | 0.448369 | 0.647218 | |
| SH3BP1 | 2.239626 | -0.29405 | 0.999532 | | 2.39186 | -0.35175 | 0.830894 | | 1.99387 | -0.04905 | 0.988328 | |
| CEPT1 | 2.889962 | 0.666553 | 0.999532 | | 2.16205 | -0.35503 | 0.830894 | | 2.574781 | -1.00657 | 0.350296 | |
| RPF2 | 4.224637 | -0.06801 | 0.999532 | | 4.36758 | -0.25854 | 0.830894 | | 3.679473 | -0.17544 | 0.901813 | |
| HACD3 | 5.963526 | 0.280686 | 0.999532 | | 6.331272 | 0.228128 | 0.830894 | | 5.893285 | -0.04887 | 0.978229 | |
| CCNH | 3.663264 | -0.57129 | 0.999532 | | 4.490082 | -0.25893 | 0.830894 | | 3.072018 | 0.331189 | 0.806394 | |
| CD300E | 2.605082 | 0.035947 | 0.999532 | | 2.723138 | -0.32185 | 0.830894 | | 2.29861 | -0.33465 | 0.836935 | |
| GTF2B | 3.792384 | 0.216464 | 0.999532 | | 4.399709 | 0.254221 | 0.830894 | | 3.847505 | 0.069417 | 0.974421 | |
| DAGLB | 3.31874 | 0.239897 | 0.999532 | | 2.893037 | -0.30496 | 0.830894 | | 2.857425 | -0.52998 | 0.678541 | |
| DLGAP1 | 2.61326 | 0.171152 | 0.999532 | | 2.308067 | -0.34892 | 0.830894 | | 2.260979 | -0.51447 | 0.741408 | |
| FLVCR1 | 3.268079 | 0.48097 | 0.999532 | | 3.425029 | 0.313597 | 0.830894 | | 3.47163 | -0.15099 | 0.929529 | |
| KDM2B | 3.329948 | -0.20352 | 0.999532 | | 3.556261 | -0.29264 | 0.830894 | | 3.066107 | -0.07139 | 0.974421 | |
| C1QC | 5.266672 | -0.85948 | 0.999532 | | 6.649737 | -0.41993 | 0.830894 | | 4.211941 | 0.464762 | 0.707142 | |
| PSMC6 | 5.097596 | -0.29232 | 0.999532 | | 5.76593 | -0.2228 | 0.830894 | | 4.515164 | 0.08573 | 0.955167 | |
| NAA50 | 2.766122 | 0.350233 | 0.999532 | | 2.955222 | 0.344699 | 0.831031 | | 2.835236 | 0.033918 | 0.991134 | |
| TMPO | 3.494554 | 0.127101 | 0.999532 | | 3.278996 | -0.28961 | 0.831031 | | 2.948823 | -0.40669 | 0.759211 | |
| ANGPTL2 | 2.699737 | -0.09919 | 0.999532 | | 3.293363 | 0.299661 | 0.831031 | | 2.843545 | 0.415156 | 0.762271 | |
| LDHAL6A | 2.713371 | 0.191247 | 0.999532 | | 2.529768 | -0.32191 | 0.831031 | | 2.348199 | -0.49737 | 0.722703 | |
| GLB1 | 2.086455 | 0.083832 | 0.999532 | | 2.379258 | 0.328794 | 0.831171 | | 2.190262 | 0.259722 | 0.881837 | |
| IFI44 | 3.863266 | 0.103918 | 0.999532 | | 3.781468 | -0.27958 | 0.831207 | | 3.456766 | -0.3658 | 0.774044 | |
| BUB1B | 2.306078 | 0.09597 | 0.999532 | | 2.250274 | -0.3645 | 0.831207 | | 1.929342 | -0.44047 | 0.793454 | |
| HS3ST3B | 5.354767 | -0.35392 | 0.999532 | | 6.425216 | -0.22426 | 0.831207 | | 4.747119 | 0.149874 | 0.907568 | |
| SLC23A2 | 3.614559 | 0.138876 | 0.999532 | | 3.33425 | -0.29523 | 0.831238 | | 3.142154 | -0.4226 | 0.739496 | |
| UBE2D4 | 2.684613 | -0.46007 | 0.999532 | | 3.286746 | -0.30477 | 0.831253 | | 2.367365 | 0.177232 | 0.924131 | |
| NCOA2 | 4.286038 | 0.564159 | 0.999532 | | 4.340603 | 0.252713 | 0.831388 | | 4.39326 | -0.29261 | 0.791576 | |
| ZYG11A | 2.323463 | 0.543253 | 0.999532 | | 1.852055 | -0.38261 | 0.831388 | | 2.099943 | -0.91166 | 0.505689 | |
| NR2E3 | 1.97138 | -0.40024 | 0.999532 | | 2.236077 | -0.38194 | 0.831388 | | 1.681285 | 0.035963 | 0.992286 | |
| ZMYM5 | 3.072884 | 0.081814 | 0.999532 | | 4.005363 | 0.288015 | 0.831388 | | 3.415408 | 0.21935 | 0.881837 | |
| TEP1 | 3.042566 | -0.26949 | 0.999532 | | 3.236118 | -0.29371 | 0.83141 | | 2.522443 | -0.0101 | 0.996891 | |
| DKK3 | 3.337075 | -0.46453 | 0.999532 | | 4.679573 | 0.262649 | 0.83141 | | 3.585052 | 0.73234 | 0.515762 | |
| ZBTB45 | 3.287376 | 0.221097 | 0.999532 | | 3.818894 | 0.304816 | 0.83141 | | 3.426147 | 0.110194 | 0.951149 | |
| S100A14 | 2.582654 | 0.320199 | 0.999532 | | 2.27527 | -0.36299 | 0.83141 | | 2.233732 | -0.66483 | 0.650461 | |
| NUP160 | 2.811605 | 0.276183 | 0.999532 | | 3.114782 | 0.29826 | 0.83141 | | 2.911477 | 0.040424 | 0.987274 | |
| ZNF154 | 3.499592 | 0.303131 | 0.999532 | | 3.754986 | 0.274247 | 0.831477 | | 3.556908 | -0.01388 | 0.996681 | |
| PPP1R8 | 5.79481 | 0.062242 | 0.999532 | | 6.028044 | -0.22945 | 0.831477 | | 4.932856 | -0.26648 | 0.813143 | |
| GAST | 3.163485 | 0.337248 | 0.999532 | | 3.054596 | -0.31127 | 0.831477 | | 2.92143 | -0.632 | 0.594446 | |
| PDCD5 | 5.239737 | 0.091231 | 0.999532 | | 6.370121 | 0.231262 | 0.831477 | | 5.563247 | 0.163157 | 0.897096 | |
| NPRL2 | 1.604488 | -0.19234 | 0.999532 | | 2.203498 | 0.356402 | 0.831479 | | 1.828251 | 0.568904 | 0.730921 | |
| HNRNPH1 | 15.80873 | -0.28621 | 0.999532 | | 20.69801 | 0.148271 | 0.831479 | | 15.60855 | 0.458364 | 0.506666 | |
| NUFIP2 | 3.025893 | 0.231489 | 0.999532 | | 2.791047 | -0.31693 | 0.83156 | | 2.583134 | -0.53097 | 0.682165 | |
| NMU | 2.469386 | 0.248031 | 0.999532 | | 2.249571 | -0.37649 | 0.831712 | | 2.163091 | -0.60485 | 0.709825 | |
| UGGT1 | 3.99643 | -0.2533 | 0.999532 | | 4.45817 | -0.25718 | 0.831724 | | 3.366354 | 0.012107 | 0.996891 | |
| PHYHIP | 7.288558 | -0.02428 | 0.999532 | | 7.435484 | -0.24303 | 0.831724 | | 6.300569 | -0.20678 | 0.881837 | |
| TAS2R14 | 2.250129 | -0.39604 | 0.999532 | | 2.528582 | -0.35985 | 0.831724 | | 1.967158 | 0.054311 | 0.986811 | |
| CREM | 4.118695 | -0.60324 | 0.999532 | | 5.239589 | -0.23382 | 0.831724 | | 3.694026 | 0.392246 | 0.736481 | |
| ZW10 | 4.155246 | 0.13728 | 0.999532 | | 4.806364 | 0.257098 | 0.831754 | | 4.397477 | 0.142199 | 0.915533 | |
| GPRC5C | 6.174389 | 0.174507 | 0.999532 | | 6.986035 | 0.213468 | 0.831754 | | 6.319653 | 0.058875 | 0.968434 | |
| LSP1 | 4.557169 | -0.2013 | 0.999532 | | 5.802938 | 0.27955 | 0.831754 | | 4.959255 | 0.474747 | 0.717627 | |
| EMSY | 4.123913 | -0.41108 | 0.999532 | | 5.010683 | -0.24891 | 0.831754 | | 3.575906 | 0.185289 | 0.896374 | |
| CEP295N | 2.516921 | -0.26621 | 0.999532 | | 3.265216 | 0.300468 | 0.83181 | | 2.589592 | 0.582679 | 0.663633 | |
| ASXL1 | 3.610142 | -0.32429 | 0.999532 | | 4.242194 | -0.26249 | 0.83181 | | 3.170816 | 0.081331 | 0.965649 | |
| VPREB3 | 2.201653 | -0.91059 | 0.999532 | | 2.804446 | -0.32449 | 0.832139 | | 1.821055 | 0.60314 | 0.722703 | |
| SNAP23 | 6.530476 | -0.19997 | 0.999532 | | 7.170072 | -0.21399 | 0.832139 | | 5.770625 | 0.003331 | 0.998586 | |
| HDGF | 7.904094 | -0.08042 | 0.999532 | | 9.754469 | 0.190141 | 0.832139 | | 8.101481 | 0.291908 | 0.722703 | |
| OR52E5 | 1.884494 | 0.108292 | 0.999532 | | 1.81736 | -0.38026 | 0.832457 | | 1.723422 | -0.47088 | 0.791576 | |
| MAGEB6 | 2.390467 | 0.047297 | 0.999532 | | 2.4239 | -0.3335 | 0.832858 | | 2.028226 | -0.3624 | 0.826668 | |
| PSMD14 | 4.063421 | 0.222743 | 0.999532 | | 4.876389 | 0.259334 | 0.832858 | | 4.312055 | 0.051015 | 0.977473 | |
| ITK | 2.421125 | 0.699372 | 0.999532 | | 1.846615 | -0.39678 | 0.832858 | | 2.169985 | -1.0831 | 0.405327 | |
| PHRF1 | 4.337185 | -0.40232 | 0.999532 | | 5.069645 | -0.2381 | 0.832858 | | 3.746158 | 0.182003 | 0.897096 | |
| TUBA4A | 5.677997 | -0.4231 | 0.999532 | | 7.058335 | -0.22206 | 0.832858 | | 5.151057 | 0.223256 | 0.852739 | |
| KLF7 | 5.295365 | -0.1842 | 0.999532 | | 6.98214 | 0.216272 | 0.832858 | | 5.475873 | 0.418695 | 0.658914 | |
| VIPR2 | 2.9462 | 0.783781 | 0.999532 | | 2.393159 | -0.36521 | 0.832858 | | 2.733263 | -1.12608 | 0.286175 | |
| PCNT | 2.83888 | -0.16484 | 0.999532 | | 3.021783 | -0.3125 | 0.832858 | | 2.343681 | -0.12762 | 0.94749 | |
| THAP6 | 2.605102 | 0.674389 | 0.999532 | | 2.529915 | 0.338058 | 0.832858 | | 2.586872 | -0.31659 | 0.834627 | |
| ATG9A | 4.203642 | 0.192057 | 0.999532 | | 4.789341 | 0.247495 | 0.832858 | | 4.365707 | 0.07337 | 0.964675 | |
| RANBP17 | 2.390577 | -0.06793 | 0.999532 | | 2.373431 | -0.3379 | 0.832858 | | 2.005762 | -0.25172 | 0.892869 | |
| DMXL1 | 4.771088 | -0.17667 | 0.999532 | | 6.046608 | 0.229897 | 0.832858 | | 4.882889 | 0.427733 | 0.660958 | |
| FURIN | 6.994137 | -0.3394 | 0.999532 | | 9.208523 | 0.191097 | 0.832858 | | 7.171317 | 0.551312 | 0.45512 | |
| RHOG | 2.851281 | 0.148083 | 0.999532 | | 3.177832 | 0.294322 | 0.832881 | | 2.977734 | 0.161305 | 0.920805 | |
| MFSD10 | 3.417913 | 0.214157 | 0.999532 | | 3.667586 | 0.276522 | 0.832881 | | 3.466795 | 0.070459 | 0.972221 | |
| RND3 | 5.507898 | -0.20852 | 0.999532 | | 6.322464 | -0.23453 | 0.832881 | | 4.833336 | 0.000714 | 0.999635 | |
| ELF2 | 5.650031 | -0.2356 | 0.999532 | | 6.459475 | -0.23593 | 0.832881 | | 4.873617 | 0.023832 | 0.991434 | |
| IFNA6 | 2.416741 | 0.465141 | 0.999532 | | 1.997211 | -0.38229 | 0.832881 | | 2.129729 | -0.83019 | 0.546319 | |
| H1-4 | 18.74571 | -0.17472 | 0.999532 | | 23.25468 | 0.163058 | 0.832926 | | 18.74227 | 0.369575 | 0.504771 | |
| C16orf91 | 2.916054 | -0.23247 | 0.999532 | | 3.116546 | -0.30638 | 0.832926 | | 2.398328 | -0.05611 | 0.98162 | |
| PRR35 | 2.440981 | 0.450639 | 0.999532 | | 2.14188 | -0.36392 | 0.832926 | | 2.134696 | -0.79577 | 0.549065 | |
| TRIM59 | 2.207735 | -0.10795 | 0.999532 | | 2.3552 | -0.33625 | 0.833216 | | 1.943347 | -0.2085 | 0.914692 | |
| LNX2 | 3.004436 | -0.36921 | 0.999532 | | 3.625248 | -0.31113 | 0.833224 | | 2.757969 | 0.083184 | 0.972758 | |
| JAM2 | 3.339177 | -0.31821 | 0.999532 | | 3.878826 | -0.27373 | 0.8333 | | 3.014998 | 0.06267 | 0.975857 | |
| POGLUT2 | 1.771194 | 0.183716 | 0.999532 | | 2.068852 | 0.356646 | 0.8333 | | 1.97518 | 0.190458 | 0.926586 | |
| APOBEC3 | 3.447007 | -0.17767 | 0.999532 | | 3.599798 | -0.27571 | 0.8333 | | 2.885808 | -0.0901 | 0.963866 | |
| CLK1 | 15.98173 | -0.35198 | 0.999532 | | 18.92975 | -0.16295 | 0.83352 | | 13.98194 | 0.217192 | 0.761337 | |
| ADD1 | 10.05066 | -0.03863 | 0.999532 | | 12.27423 | 0.177731 | 0.83352 | | 10.24153 | 0.235909 | 0.764366 | |
| RBBP8 | 3.536627 | -0.31236 | 0.999532 | | 4.154676 | -0.27416 | 0.833683 | | 2.880242 | 0.062822 | 0.974806 | |
| PPP4C | 5.169612 | -0.00059 | 0.999532 | | 6.214796 | 0.225953 | 0.833823 | | 5.212952 | 0.236752 | 0.859898 | |
| ALKBH4 | 2.602728 | -0.51309 | 0.999532 | | 3.160386 | -0.31105 | 0.833881 | | 2.352808 | 0.222939 | 0.906811 | |
| OR4F15 | 2.201537 | 0.204957 | 0.999532 | | 2.148548 | -0.38316 | 0.834058 | | 1.982261 | -0.5682 | 0.732916 | |
| ANKRD53 | 2.395212 | -0.44825 | 0.999532 | | 2.985064 | -0.33044 | 0.834058 | | 2.093263 | 0.17229 | 0.938429 | |
| FLYWCH | 4.509154 | -0.44804 | 0.999532 | | 5.183691 | -0.24612 | 0.834479 | | 4.02359 | 0.206434 | 0.890746 | |
| SIGLECL | 2.781527 | 0.562106 | 0.999532 | | 2.168359 | -0.39341 | 0.834609 | | 2.38869 | -0.92554 | 0.488953 | |
| UPB1 | 4.145928 | 0.719272 | 0.999532 | | 3.855903 | 0.273354 | 0.834609 | | 4.182601 | -0.42727 | 0.689782 | |
| SEMA4C | 6.147549 | -0.32467 | 0.999532 | | 8.177252 | 0.197555 | 0.834743 | | 6.430253 | 0.540935 | 0.504121 | |
| SLC20A1 | 3.791691 | -0.48524 | 0.999532 | | 4.218237 | -0.27756 | 0.834799 | | 3.187914 | 0.199495 | 0.914929 | |
| GPER1 | 3.320457 | 0.455825 | 0.999532 | | 2.811055 | -0.31898 | 0.834799 | | 3.013268 | -0.75704 | 0.540342 | |
| CCL11 | 1.785079 | -0.17489 | 0.999532 | | 2.225394 | 0.346604 | 0.834799 | | 1.894181 | 0.527226 | 0.750371 | |
| SMIM34A | 2.345502 | -0.09 | 0.999532 | | 2.28715 | -0.34591 | 0.834799 | | 1.968535 | -0.24128 | 0.900848 | |
| TMEM223 | 4.708655 | -0.00817 | 0.999532 | | 4.655369 | -0.25536 | 0.834799 | | 4.048141 | -0.23716 | 0.862823 | |
| PHF5A | 2.518939 | -0.40877 | 0.999532 | | 2.907957 | -0.31705 | 0.834878 | | 2.046365 | 0.109786 | 0.96129 | |
| ZNF84 | 3.025544 | -0.17976 | 0.999532 | | 3.251289 | -0.30035 | 0.834915 | | 2.717265 | -0.10084 | 0.96129 | |
| RSL24D1 | 4.234123 | -0.23415 | 0.999532 | | 5.518895 | 0.22641 | 0.834915 | | 4.467518 | 0.47987 | 0.652925 | |
| TXNDC9 | 5.684168 | 0.274627 | 0.999532 | | 5.115818 | -0.23658 | 0.834935 | | 4.97729 | -0.50372 | 0.5991 | |
| DENND5B | 2.930373 | 0.220645 | 0.999532 | | 2.582525 | -0.33278 | 0.834935 | | 2.394402 | -0.55201 | 0.705425 | |
| EIF3A | 13.07835 | -0.05695 | 0.999532 | | 15.7325 | 0.161548 | 0.834935 | | 13.36767 | 0.239016 | 0.750371 | |
| MLC1 | 1.991742 | -0.05327 | 0.999532 | | 2.147113 | -0.36784 | 0.835222 | | 1.798579 | -0.2871 | 0.88867 | |
| EIF2B1 | 5.67682 | 0.19295 | 0.999532 | | 6.526212 | 0.21894 | 0.835222 | | 6.029501 | 0.046349 | 0.976441 | |
| ZBED3 | 3.073623 | 0.397641 | 0.999532 | | 3.096347 | 0.290718 | 0.835306 | | 3.007211 | -0.09233 | 0.960773 | |
| OLA1 | 5.350205 | 0.294604 | 0.999532 | | 5.761879 | 0.232599 | 0.835449 | | 5.638887 | -0.04861 | 0.97652 | |
| ZNF263 | 2.981724 | 0.240168 | 0.999532 | | 3.187286 | 0.285689 | 0.835744 | | 3.096952 | 0.063861 | 0.974421 | |
| DCAF7 | 2.222065 | 0.229829 | 0.999532 | | 2.750605 | 0.320591 | 0.835793 | | 2.518132 | 0.109197 | 0.958191 | |
| NAPRT | 3.830416 | -0.09167 | 0.999532 | | 4.000947 | -0.25914 | 0.835793 | | 3.341942 | -0.15128 | 0.920259 | |
| ASB10 | 1.782578 | -0.24937 | 0.999532 | | 2.653094 | 0.38395 | 0.835793 | | 2.191494 | 0.648054 | 0.717627 | |
| LHFPL1 | 1.955146 | -0.33869 | 0.999532 | | 2.192436 | -0.34791 | 0.835793 | | 1.748342 | 0.011171 | 0.996916 | |
| GPD1 | 4.152115 | -0.2022 | 0.999532 | | 5.295566 | 0.262371 | 0.835793 | | 4.278137 | 0.480159 | 0.664512 | |
| ARHGAP | 2.176461 | -0.06826 | 0.999532 | | 2.273178 | -0.34787 | 0.835793 | | 1.945267 | -0.26288 | 0.889271 | |
| SETD1B | 4.284845 | -0.16112 | 0.999532 | | 4.708739 | -0.256 | 0.835793 | | 3.844346 | -0.08 | 0.965649 | |
| GNG5 | 7.402611 | -0.49755 | 0.999532 | | 9.152896 | -0.19923 | 0.836026 | | 6.567967 | 0.31409 | 0.745908 | |
| UCN | 2.409646 | 0.63993 | 0.999532 | | 2.408767 | 0.344067 | 0.836026 | | 2.451108 | -0.27979 | 0.871957 | |
| AURKA | 3.678003 | 0.167544 | 0.999532 | | 4.317549 | 0.266316 | 0.836026 | | 3.783407 | 0.118497 | 0.9402 | |
| TCHP | 2.897526 | 0.436217 | 0.999532 | | 2.415231 | -0.33386 | 0.836026 | | 2.583459 | -0.74896 | 0.542124 | |
| GPC4 | 3.69376 | 0.127784 | 0.999532 | | 4.093986 | 0.286339 | 0.836354 | | 3.583318 | 0.187311 | 0.901813 | |
| SNX21 | 2.950924 | 0.192755 | 0.999532 | | 2.873828 | -0.3121 | 0.836522 | | 2.601997 | -0.48665 | 0.717627 | |
| CDH15 | 2.062244 | 0.176407 | 0.999532 | | 1.946327 | -0.37272 | 0.836522 | | 1.850013 | -0.52636 | 0.751138 | |
| KCNIP3 | 2.164596 | 0.576077 | 0.999532 | | 1.873817 | -0.40027 | 0.836522 | | 1.911545 | -0.95814 | 0.49299 | |
| SLC46A3 | 2.284275 | -0.23742 | 0.999532 | | 2.538493 | -0.33512 | 0.836718 | | 1.9798 | -0.07847 | 0.975473 | |
| SGK1 | 18.36235 | -0.7182 | 0.484521 | | 26.47664 | 0.158307 | 0.836968 | | 18.21746 | 0.907627 | 0.017097 | UP |
| TBC1D2B | 4.938065 | -0.02463 | 0.999532 | | 6.04745 | 0.230543 | 0.836968 | | 5.17871 | 0.276158 | 0.792538 | |
| SSH1 | 3.026055 | -0.43353 | 0.999532 | | 3.388386 | -0.29212 | 0.837071 | | 2.657779 | 0.16473 | 0.927982 | |
| COL20A1 | 1.839328 | 0.899218 | 0.999532 | | 1.609234 | 0.405237 | 0.837071 | | 1.887458 | -0.4771 | 0.776852 | |
| IRX5 | 3.205397 | 0.359657 | 0.999532 | | 2.809403 | -0.34385 | 0.837071 | | 2.899847 | -0.68092 | 0.614824 | |
| EMILIN2 | 2.709515 | -0.01545 | 0.999532 | | 2.673889 | -0.32428 | 0.837071 | | 2.195 | -0.28802 | 0.865363 | |
| APOL1 | 12.80722 | -0.13102 | 0.999532 | | 16.38122 | 0.201647 | 0.837071 | | 13.35283 | 0.347673 | 0.668532 | |
| MAP4K5 | 4.243733 | -0.45061 | 0.999532 | | 6.110328 | 0.240013 | 0.837071 | | 4.547415 | 0.709474 | 0.451915 | |
| BHLHA15 | 2.953161 | 0.550346 | 0.999532 | | 2.49518 | -0.34065 | 0.837071 | | 2.66791 | -0.87134 | 0.458377 | |
| TRIM23 | 2.360115 | -0.09666 | 0.999532 | | 2.453121 | -0.32686 | 0.837071 | | 2.105714 | -0.20899 | 0.911296 | |
| PLOD2 | 4.133276 | 0.611715 | 0.999532 | | 4.134059 | 0.262909 | 0.837296 | | 4.390435 | -0.3171 | 0.800922 | |
| CCDC110 | 2.619172 | -0.2189 | 0.999532 | | 3.143144 | -0.32143 | 0.837299 | | 2.405707 | -0.05111 | 0.985825 | |
| G6PD | 2.994857 | 0.162697 | 0.999532 | | 2.64932 | -0.32217 | 0.837299 | | 2.483749 | -0.46717 | 0.745979 | |
| C1orf216 | 3.473376 | -0.51986 | 0.999532 | | 4.192275 | -0.26032 | 0.83733 | | 3.209879 | 0.284767 | 0.842696 | |
| TMEM131 | 4.540096 | -0.32554 | 0.999532 | | 5.577577 | -0.22547 | 0.83733 | | 3.997695 | 0.12624 | 0.934701 | |
| IL17D | 2.118632 | -0.03649 | 0.999532 | | 2.200098 | -0.35649 | 0.837406 | | 1.926657 | -0.30184 | 0.877268 | |
| ICE1 | 3.013217 | 0.257784 | 0.999532 | | 3.459057 | 0.291353 | 0.837406 | | 3.278122 | 0.047062 | 0.983281 | |
| DUT | 6.607001 | 0.254842 | 0.999532 | | 6.992251 | 0.220815 | 0.837406 | | 6.649945 | -0.0114 | 0.996681 | |
| ZNHIT2 | 3.655284 | 0.329111 | 0.999532 | | 3.499238 | -0.29737 | 0.837406 | | 3.361871 | -0.59346 | 0.637193 | |
| FAM168A | 7.915598 | -0.45239 | 0.999532 | | 9.754392 | -0.18355 | 0.837406 | | 7.121588 | 0.288081 | 0.739496 | |
| EFR3B | 2.945728 | 0.081226 | 0.999532 | | 2.976481 | -0.30508 | 0.837406 | | 2.506052 | -0.36748 | 0.801572 | |
| PLA2G10 | 2.448683 | 0.349109 | 0.999532 | | 2.262517 | -0.35195 | 0.837406 | | 2.314558 | -0.68078 | 0.644433 | |
| C16orf87 | 1.986742 | 0.104236 | 0.999532 | | 2.423634 | 0.341313 | 0.837406 | | 2.248251 | 0.253989 | 0.890932 | |
| BTBD6 | 7.217001 | -0.22242 | 0.999532 | | 8.16704 | -0.20835 | 0.837406 | | 6.619941 | 0.033707 | 0.98514 | |
| IL1F10 | 3.255593 | -0.02523 | 0.999532 | | 3.618237 | -0.29575 | 0.837406 | | 2.976605 | -0.24826 | 0.876781 | |
| ABCB6 | 2.680762 | 0.229512 | 0.999532 | | 2.58512 | -0.33321 | 0.837406 | | 2.516617 | -0.54495 | 0.705046 | |
| TSFM | 4.110789 | 0.254446 | 0.999532 | | 4.415291 | 0.259801 | 0.837406 | | 4.176788 | 0.011881 | 0.996891 | |
| CFLAR | 10.8235 | -0.2528 | 0.999532 | | 13.86176 | 0.168391 | 0.837406 | | 10.74895 | 0.438479 | 0.515762 | |
| RNF115 | 4.620334 | -0.63686 | 0.999532 | | 5.670424 | -0.22427 | 0.837406 | | 4.055049 | 0.425865 | 0.705425 | |
| NFKBIB | 2.570512 | 0.038342 | 0.999532 | | 2.671007 | -0.31653 | 0.837406 | | 2.245899 | -0.33128 | 0.839811 | |
| PI4K2B | 2.420626 | 0.588886 | 0.999532 | | 2.531319 | 0.329309 | 0.837406 | | 2.659805 | -0.24404 | 0.881837 | |
| SAP18 | 10.69759 | -0.02739 | 0.999532 | | 12.5543 | 0.204004 | 0.837406 | | 10.84676 | 0.243631 | 0.805576 | |
| ZNF575 | 2.662396 | -0.23526 | 0.999532 | | 2.987992 | -0.31165 | 0.837788 | | 2.228178 | -0.05985 | 0.978931 | |
| OMA1 | 3.03533 | -0.01278 | 0.999532 | | 3.884973 | 0.263556 | 0.837788 | | 3.100541 | 0.296421 | 0.827643 | |
| DCAF13 | 3.264172 | -0.23616 | 0.999532 | | 3.589604 | -0.27266 | 0.837788 | | 2.875996 | -0.02093 | 0.995112 | |
| ZMAT5 | 3.847563 | -0.04445 | 0.999532 | | 4.080336 | -0.27951 | 0.837788 | | 3.461444 | -0.21125 | 0.893262 | |
| RGR | 2.621326 | -0.13303 | 0.999532 | | 2.855992 | -0.34449 | 0.837788 | | 2.332923 | -0.1943 | 0.920259 | |
| MYBL2 | 2.211354 | 0.628466 | 0.999532 | | 2.201411 | 0.343513 | 0.837788 | | 2.204579 | -0.26534 | 0.876781 | |
| SETD7 | 7.644246 | 0.686527 | 0.999532 | | 6.759834 | 0.215216 | 0.837788 | | 7.703521 | -0.43623 | 0.566812 | |
| VDAC1 | 13.56969 | 0.006554 | 0.999532 | | 16.08088 | 0.157324 | 0.837788 | | 13.57494 | 0.171071 | 0.82512 | |
| OR5P3 | 2.309734 | 0.391952 | 0.999532 | | 2.254968 | -0.35167 | 0.837788 | | 2.031351 | -0.7149 | 0.614525 | |
| RABGAP | 5.864812 | 0.062982 | 0.999532 | | 6.906261 | 0.206006 | 0.837788 | | 5.999966 | 0.162659 | 0.881837 | |
| MAS1L | 1.892144 | -0.08098 | 0.999532 | | 1.844559 | -0.38041 | 0.837788 | | 1.622122 | -0.28396 | 0.894617 | |
| ADGRF4 | 2.179951 | 0.520514 | 0.999532 | | 2.120355 | 0.346352 | 0.837788 | | 2.335529 | -0.15931 | 0.939549 | |
| ZNF74 | 2.050379 | 0.139931 | 0.999532 | | 1.874689 | -0.36605 | 0.837788 | | 1.849315 | -0.49165 | 0.776158 | |
| FOXS1 | 3.369508 | 0.190957 | 0.999532 | | 3.123922 | -0.29315 | 0.837788 | | 3.043672 | -0.4665 | 0.718643 | |
| PRH2 | 2.642912 | 0.01928 | 0.999532 | | 2.838885 | -0.32669 | 0.837788 | | 2.340426 | -0.32098 | 0.853068 | |
| MPI | 2.204953 | 0.51347 | 0.999532 | | 2.272314 | 0.33539 | 0.837877 | | 2.341003 | -0.15857 | 0.934172 | |
| GDF1 | 9.250798 | 0.255097 | 0.999532 | | 8.905945 | -0.23218 | 0.83803 | | 8.600604 | -0.46618 | 0.65401 | |
| PLAAT2 | 2.298971 | 0.327206 | 0.999532 | | 1.924022 | -0.37601 | 0.838457 | | 1.967257 | -0.68652 | 0.660958 | |
| PIEZO2 | 4.411928 | -0.03586 | 0.999532 | | 5.49127 | 0.238991 | 0.838457 | | 4.605433 | 0.293008 | 0.79686 | |
| IL10RB | 3.303991 | 0.887538 | 0.999532 | | 2.847319 | 0.309893 | 0.838673 | | 3.391568 | -0.56241 | 0.636021 | |
| OSBP | 3.587341 | -0.10213 | 0.999532 | | 3.728094 | -0.27797 | 0.838673 | | 3.126141 | -0.15751 | 0.922922 | |
| PLEKHA1 | 2.277996 | -0.13898 | 0.999532 | | 2.786772 | 0.308179 | 0.838673 | | 2.339692 | 0.470516 | 0.739496 | |
| YWHAZ | 20.05487 | -0.09836 | 0.999532 | | 24.39425 | 0.141596 | 0.838673 | | 19.78624 | 0.27062 | 0.647218 | |
| VAMP3 | 5.655343 | 0.262433 | 0.999532 | | 6.095527 | 0.219209 | 0.838673 | | 5.674125 | -0.03033 | 0.988853 | |
| C3orf14 | 2.790817 | -0.19572 | 0.999532 | | 2.985564 | -0.31282 | 0.838673 | | 2.565324 | -0.11102 | 0.95999 | |
| PLCXD1 | 3.615118 | -0.08288 | 0.999532 | | 3.548911 | -0.27606 | 0.838673 | | 3.009746 | -0.18345 | 0.907968 | |
| UNC13D | 2.887325 | 0.407845 | 0.999532 | | 2.399513 | -0.33656 | 0.838673 | | 2.595166 | -0.71902 | 0.582673 | |
| ZNF282 | 3.326654 | -0.06161 | 0.999532 | | 3.599353 | -0.2745 | 0.838673 | | 2.859794 | -0.19429 | 0.90066 | |
| ACVRL1 | 6.661354 | -0.28967 | 0.999532 | | 9.233281 | 0.204764 | 0.838695 | | 7.204664 | 0.511675 | 0.527585 | |
| GFAP | 1.916015 | 0.403624 | 0.999532 | | 2.119332 | 0.369436 | 0.838695 | | 2.073357 | -0.01576 | 0.996891 | |
| GPLD1 | 3.021608 | -0.32711 | 0.999532 | | 3.462738 | -0.31379 | 0.83897 | | 2.712899 | 0.013579 | 0.996891 | |
| CEND1 | 2.213182 | 0.210284 | 0.999532 | | 2.622655 | 0.338623 | 0.839037 | | 2.587526 | 0.139939 | 0.948484 | |
| SNN | 4.531119 | -0.32896 | 0.999532 | | 5.320117 | -0.23242 | 0.839443 | | 4.039519 | 0.115313 | 0.9402 | |
| GJA4 | 4.433682 | -0.55612 | 0.999532 | | 5.394421 | -0.22736 | 0.839443 | | 3.750227 | 0.345387 | 0.763873 | |
| ACTL10 | 3.023233 | -0.24838 | 0.999532 | | 3.534644 | -0.29476 | 0.840464 | | 2.734382 | -0.02379 | 0.994192 | |
| METTL7A | 9.520936 | 0.152562 | 0.999532 | | 11.11638 | 0.204603 | 0.840486 | | 9.395131 | 0.109755 | 0.934572 | |
| IK | 6.992407 | 0.381878 | 0.999532 | | 7.691535 | 0.198792 | 0.840523 | | 7.243624 | -0.16385 | 0.877268 | |
| CHD8 | 4.615189 | 0.358606 | 0.999532 | | 4.649699 | 0.245337 | 0.840601 | | 4.600368 | -0.0952 | 0.950616 | |
| SAT1 | 15.13678 | -0.3973 | 0.999532 | | 20.69824 | 0.192378 | 0.840601 | | 15.37425 | 0.621775 | 0.387954 | |
| PDGFD | 5.961148 | -0.60343 | 0.999532 | | 8.666814 | 0.213084 | 0.840601 | | 6.218468 | 0.837813 | 0.268933 | |
| HCFC1 | 2.829514 | 0.081075 | 0.999532 | | 3.177618 | 0.289001 | 0.840601 | | 2.923615 | 0.209683 | 0.900848 | |
| FDXR | 2.096169 | -0.12983 | 0.999532 | | 2.234223 | -0.34784 | 0.840601 | | 1.722756 | -0.19645 | 0.923834 | |
| CATSPER | 2.791881 | 0.174414 | 0.999532 | | 2.769055 | -0.33373 | 0.840601 | | 2.689983 | -0.49891 | 0.740755 | |
| MAN2C1 | 6.380611 | 0.005053 | 0.999532 | | 6.871748 | -0.20918 | 0.840601 | | 5.694192 | -0.20139 | 0.852179 | |
| CDC7 | 3.992037 | 0.031941 | 0.999532 | | 4.066748 | -0.2695 | 0.840601 | | 3.48072 | -0.28267 | 0.839373 | |
| SYNGAP1 | 3.115229 | 0.294488 | 0.999532 | | 3.635404 | 0.274763 | 0.840601 | | 3.278111 | 0.001823 | 0.999385 | |
| LPAR1 | 2.792205 | -0.15954 | 0.999532 | | 2.980631 | -0.30092 | 0.840601 | | 2.322894 | -0.12275 | 0.949669 | |
| KIF18B | 1.896019 | -0.73659 | 0.999532 | | 2.422195 | -0.34094 | 0.840601 | | 1.638175 | 0.422142 | 0.831749 | |
| FARS2 | 2.921958 | 0.271756 | 0.999532 | | 2.564151 | -0.31079 | 0.840601 | | 2.59792 | -0.56352 | 0.668481 | |
| ITGB3BP | 2.131509 | -0.1771 | 0.999532 | | 2.191196 | -0.34871 | 0.840601 | | 1.824293 | -0.15418 | 0.945902 | |
| YARS1 | 4.748946 | -0.13425 | 0.999532 | | 5.934788 | 0.217474 | 0.840601 | | 5.003277 | 0.369449 | 0.723933 | |
| KCNK6 | 6.669891 | 0.029386 | 0.999532 | | 7.064841 | -0.22863 | 0.840601 | | 6.196833 | -0.24612 | 0.825997 | |
| MAGEA3 | 6.460698 | -0.0215 | 0.999532 | | 6.998882 | -0.19918 | 0.840601 | | 5.99764 | -0.15491 | 0.890746 | |
| GCA | 4.242693 | 0.411621 | 0.999532 | | 3.520945 | -0.27577 | 0.840601 | | 3.834735 | -0.68378 | 0.542124 | |
| C1QTNF9 | 3.283644 | 0.186067 | 0.999532 | | 3.248617 | -0.3072 | 0.840601 | | 3.067256 | -0.47909 | 0.720106 | |
| ST6GAL1 | 12.80166 | 0.032782 | 0.999532 | | 14.73849 | 0.175939 | 0.840601 | | 12.77753 | 0.141476 | 0.901033 | |
| TBCE | 3.014707 | -0.1045 | 0.999532 | | 4.157891 | 0.260121 | 0.840601 | | 3.28956 | 0.377855 | 0.755977 | |
| GLCCI1 | 2.358697 | -0.20987 | 0.999532 | | 2.580291 | -0.32551 | 0.840601 | | 2.05744 | -0.09689 | 0.968235 | |
| ATG7 | 3.577092 | -0.09964 | 0.999532 | | 3.78207 | -0.27191 | 0.840601 | | 3.221372 | -0.15707 | 0.924131 | |
| POMT1 | 2.86833 | 0.058833 | 0.999532 | | 3.371734 | 0.286809 | 0.840601 | | 3.0138 | 0.249537 | 0.870663 | |
| ZMAT2 | 6.196193 | -0.18769 | 0.999532 | | 8.293453 | 0.193274 | 0.840601 | | 6.38646 | 0.416018 | 0.647581 | |
| KIAA1143 | 4.375786 | -0.23623 | 0.999532 | | 5.066869 | -0.2467 | 0.840601 | | 3.908031 | 0.004896 | 0.998346 | |
| SERTAD1 | 2.886929 | 0.72522 | 0.999532 | | 2.775248 | 0.309015 | 0.840826 | | 2.914473 | -0.39736 | 0.755206 | |
| C1QTNF9 | 2.469836 | 0.089609 | 0.999532 | | 2.505021 | -0.36605 | 0.840863 | | 2.359078 | -0.45496 | 0.794473 | |
| SLITRK2 | 2.288497 | 1.090801 | 0.999532 | | 1.795409 | 0.378314 | 0.840863 | | 2.341574 | -0.69485 | 0.620745 | |
| TRPM3 | 4.83332 | 0.474931 | 0.999532 | | 4.759772 | 0.259045 | 0.840863 | | 4.934696 | -0.18101 | 0.892869 | |
| NASP | 5.478489 | -0.31425 | 0.999532 | | 6.492792 | -0.21127 | 0.840955 | | 4.772119 | 0.12214 | 0.926586 | |
| RNF126 | 2.334882 | -0.49707 | 0.999532 | | 3.28274 | 0.29327 | 0.840955 | | 2.426294 | 0.805395 | 0.532713 | |
| NIN | 3.522801 | -0.11098 | 0.999532 | | 3.798239 | -0.26934 | 0.84124 | | 3.077327 | -0.14199 | 0.934354 | |
| MTFMT | 2.360286 | -0.61543 | 0.999532 | | 2.782326 | -0.3013 | 0.841322 | | 2.030481 | 0.331208 | 0.857071 | |
| ZNF260 | 2.965512 | -0.19375 | 0.999532 | | 3.455527 | -0.29412 | 0.841322 | | 2.666811 | -0.07698 | 0.972305 | |
| SPIC | 2.18495 | 0.087637 | 0.999532 | | 2.148711 | -0.35956 | 0.841571 | | 2.01502 | -0.43057 | 0.811123 | |
| HNF4A | 4.010532 | -0.19454 | 0.999532 | | 5.440356 | 0.277974 | 0.841571 | | 4.216528 | 0.508418 | 0.673405 | |
| CCDC112 | 2.87632 | -0.16025 | 0.999532 | | 3.215755 | -0.28817 | 0.841606 | | 2.618565 | -0.10547 | 0.958191 | |
| DUSP7 | 5.265611 | -0.12793 | 0.999532 | | 6.010787 | -0.21741 | 0.841606 | | 4.452866 | -0.07044 | 0.965649 | |
| RPS6KA3 | 7.034508 | 0.071319 | 0.999532 | | 8.110915 | 0.225645 | 0.841606 | | 7.173791 | 0.155554 | 0.905952 | |
| KRR1 | 2.040578 | -0.35408 | 0.999532 | | 2.915027 | 0.308536 | 0.841606 | | 2.325699 | 0.683161 | 0.638015 | |
| ZNF514 | 2.916146 | 0.291902 | 0.999532 | | 2.41638 | -0.32194 | 0.841764 | | 2.540286 | -0.60782 | 0.660958 | |
| PGBD4 | 3.04615 | 0.154085 | 0.999532 | | 3.063836 | -0.3242 | 0.84198 | | 2.756758 | -0.45584 | 0.760473 | |
| IGDCC4 | 1.926347 | 0.327902 | 0.999532 | | 2.213406 | 0.340692 | 0.84198 | | 1.949194 | 0.020843 | 0.996471 | |
| MEGF9 | 3.550128 | 0.035184 | 0.999532 | | 4.339488 | 0.275372 | 0.84198 | | 3.549744 | 0.259311 | 0.849021 | |
| GGNBP2 | 7.257395 | 0.169355 | 0.999532 | | 8.537336 | 0.215606 | 0.84198 | | 7.425207 | 0.061908 | 0.965619 | |
| SIGLEC9 | 2.810515 | -0.00802 | 0.999532 | | 2.732004 | -0.3003 | 0.84198 | | 2.40377 | -0.2746 | 0.865065 | |
| ERN1 | 3.442994 | 0.246827 | 0.999532 | | 4.002887 | 0.26024 | 0.84198 | | 3.569726 | 0.031592 | 0.989437 | |
| LCE3E | 2.378588 | 0.405434 | 0.999532 | | 2.160926 | -0.34466 | 0.84198 | | 2.273047 | -0.73818 | 0.575108 | |
| HPS5 | 1.859957 | -0.07965 | 0.999532 | | 2.512024 | 0.329062 | 0.84198 | | 2.018405 | 0.430907 | 0.795612 | |
| MBOAT1 | 2.309431 | 0.113347 | 0.999532 | | 2.224001 | -0.34607 | 0.84198 | | 1.949029 | -0.4377 | 0.791466 | |
| SMTN | 5.504684 | -0.43171 | 0.999532 | | 6.54165 | -0.20676 | 0.84198 | | 4.802916 | 0.238384 | 0.837256 | |
| ASMTL | 7.748828 | 0.410102 | 0.999532 | | 7.788549 | 0.199388 | 0.84198 | | 7.886245 | -0.19714 | 0.848095 | |
| LCN10 | 2.006641 | -0.18905 | 0.999532 | | 2.158699 | -0.34432 | 0.84198 | | 1.688426 | -0.13824 | 0.952656 | |
| SPRY3 | 2.290681 | -0.27252 | 0.999532 | | 2.464949 | -0.32426 | 0.84198 | | 1.96642 | -0.03613 | 0.991434 | |
| TNFRSF1 | 2.424505 | 0.391754 | 0.999532 | | 2.154427 | -0.35542 | 0.84198 | | 2.118677 | -0.73036 | 0.605142 | |
| NXF1 | 5.017402 | -0.23228 | 0.999532 | | 6.399055 | 0.208426 | 0.84198 | | 5.209505 | 0.461089 | 0.622543 | |
| OR2T12 | 2.883763 | 0.322907 | 0.999532 | | 2.594504 | -0.33178 | 0.84198 | | 2.528749 | -0.63575 | 0.647581 | |
| TRIM27 | 3.440984 | 0.256589 | 0.999532 | | 3.970206 | 0.286041 | 0.84198 | | 3.766625 | 0.033969 | 0.988853 | |
| CMTR1 | 2.937839 | 0.377607 | 0.999532 | | 2.474763 | -0.32429 | 0.84198 | | 2.569258 | -0.68444 | 0.589729 | |
| B4GAT1 | 5.146106 | 0.203972 | 0.999532 | | 5.929849 | 0.221244 | 0.84198 | | 5.100757 | 0.029163 | 0.989138 | |
| MSH3 | 3.267102 | 0.446147 | 0.999532 | | 2.938286 | -0.29079 | 0.84198 | | 2.862171 | -0.71392 | 0.515762 | |
| AMBRA1 | 3.77651 | 0.690377 | 0.999532 | | 3.491072 | 0.264954 | 0.84198 | | 3.888094 | -0.40814 | 0.708052 | |
| CEACAM | 3.084913 | -0.23404 | 0.999532 | | 3.522125 | -0.28746 | 0.84198 | | 2.854693 | -0.03461 | 0.990467 | |
| HDAC4 | 1.844923 | 0.412599 | 0.999532 | | 2.015782 | 0.375561 | 0.84198 | | 1.965903 | -0.01818 | 0.996681 | |
| CYSLTR1 | 2.939828 | -0.05002 | 0.999532 | | 3.629431 | 0.28943 | 0.84198 | | 3.236099 | 0.344641 | 0.809994 | |
| PIN1 | 2.84999 | -0.32793 | 0.999532 | | 3.248015 | -0.27649 | 0.84198 | | 2.519206 | 0.071445 | 0.974421 | |
| RNF139 | 5.27672 | -0.27405 | 0.999532 | | 6.668205 | 0.204449 | 0.84198 | | 5.282451 | 0.492606 | 0.578798 | |
| RTTN | 1.867559 | 0.422143 | 0.999532 | | 2.011807 | 0.363988 | 0.84198 | | 2.014848 | -0.04217 | 0.989985 | |
| PLA2G6 | 5.408777 | 0.025409 | 0.999532 | | 5.639107 | -0.23829 | 0.84198 | | 4.584154 | -0.23164 | 0.848938 | |
| MRPL3 | 6.020272 | -0.49567 | 0.999532 | | 8.486165 | 0.200689 | 0.84198 | | 6.219338 | 0.717317 | 0.304062 | |
| TBCCD1 | 3.669231 | 0.469538 | 0.999532 | | 3.149891 | -0.289 | 0.84198 | | 3.190024 | -0.74098 | 0.487828 | |
| PTBP1 | 464.2737 | 0.078693 | 0.999532 | | 549.4137 | 0.217918 | 0.84198 | | 479.5453 | 0.164012 | 0.915991 | |
| HSBP1L1 | 2.19095 | 0.503549 | 0.999532 | | 2.392975 | 0.329835 | 0.842105 | | 2.424841 | -0.15252 | 0.938429 | |
| SUPT3H | 1.774763 | 0.231437 | 0.999532 | | 1.959008 | 0.349629 | 0.842127 | | 1.895567 | 0.13744 | 0.950751 | |
| SEMA5B | 4.678858 | -0.15639 | 0.999532 | | 5.182628 | -0.28752 | 0.842127 | | 3.933043 | -0.11184 | 0.946512 | |
| RGL3 | 7.891061 | -0.25625 | 0.999532 | | 9.358777 | -0.19642 | 0.842207 | | 7.056585 | 0.065404 | 0.963866 | |
| TAGLN | 13.11986 | -0.29187 | 0.999532 | | 15.46684 | -0.1735 | 0.842207 | | 11.92863 | 0.143564 | 0.892869 | |
| RNF8 | 2.189429 | 0.423167 | 0.999532 | | 2.372359 | 0.331212 | 0.842207 | | 2.231041 | -0.07364 | 0.974421 | |
| PPP3R2 | 2.131995 | -0.05561 | 0.999532 | | 2.145655 | -0.34775 | 0.842271 | | 1.855616 | -0.27482 | 0.889493 | |
| USP25 | 6.291175 | 0.075743 | 0.999532 | | 7.521859 | 0.206507 | 0.842271 | | 6.543505 | 0.152333 | 0.894612 | |
| SLC52A2 | 3.331218 | -0.10982 | 0.999532 | | 3.330812 | -0.27139 | 0.842295 | | 2.794976 | -0.15352 | 0.928249 | |
| PPP2R5C | 9.504597 | -0.11984 | 0.999532 | | 12.26868 | 0.181148 | 0.842543 | | 10.18653 | 0.317379 | 0.681871 | |
| CCNS | 2.330112 | 0.081014 | 0.999532 | | 2.341633 | -0.34996 | 0.842543 | | 2.047615 | -0.41143 | 0.821463 | |
| ZFP62 | 2.684963 | 0.301927 | 0.999532 | | 2.835539 | 0.292773 | 0.842543 | | 2.774802 | 0.008616 | 0.997084 | |
| PPP1R2 | 9.467129 | 0.054416 | 0.999532 | | 9.933521 | -0.18141 | 0.842543 | | 8.506755 | -0.21687 | 0.810635 | |
| P3H3 | 2.682861 | -0.40273 | 0.999532 | | 2.963032 | -0.28526 | 0.842543 | | 2.202501 | 0.132124 | 0.946572 | |
| ANKS3 | 3.924892 | 0.04043 | 0.999532 | | 3.956835 | -0.26862 | 0.842543 | | 3.338238 | -0.28086 | 0.843043 | |
| KRT15 | 2.425409 | 0.283805 | 0.999532 | | 2.244197 | -0.34793 | 0.842543 | | 2.163709 | -0.613 | 0.676565 | |
| ACAD10 | 3.047025 | -0.35484 | 0.999532 | | 3.551183 | -0.26889 | 0.842543 | | 2.613728 | 0.106389 | 0.955834 | |
| CDC34 | 3.716766 | 0.170532 | 0.999532 | | 3.976253 | 0.247273 | 0.842543 | | 3.742628 | 0.088178 | 0.958191 | |
| TNIP1 | 8.736258 | -0.11333 | 0.999532 | | 9.905908 | -0.17852 | 0.842543 | | 7.866011 | -0.03991 | 0.977473 | |
| GPSM3 | 3.015458 | 0.164492 | 0.999532 | | 2.767937 | -0.29664 | 0.842543 | | 2.773749 | -0.44041 | 0.750371 | |
| ANKRD52 | 4.50339 | 0.42827 | 0.999532 | | 4.798036 | 0.23417 | 0.842543 | | 4.577636 | -0.1759 | 0.892869 | |
| RNF20 | 2.954785 | -0.0215 | 0.999532 | | 3.626352 | 0.271952 | 0.842543 | | 3.077598 | 0.30993 | 0.821724 | |
| C1orf21 | 7.307698 | 0.560598 | 0.999532 | | 7.036476 | 0.208535 | 0.842543 | | 7.430324 | -0.33311 | 0.691465 | |
| H2AC1 | 2.539362 | 0.04749 | 0.999532 | | 2.39682 | -0.31266 | 0.842543 | | 2.288634 | -0.34522 | 0.833604 | |
| ZNF384 | 4.272985 | -0.12177 | 0.999532 | | 4.659919 | -0.23278 | 0.842543 | | 3.723723 | -0.09207 | 0.954304 | |
| SPEN | 5.156365 | -0.13366 | 0.999532 | | 6.262883 | 0.205155 | 0.842543 | | 5.199772 | 0.352057 | 0.718735 | |
| POLQ | 2.381734 | -0.3295 | 0.999532 | | 2.633492 | -0.30733 | 0.842543 | | 2.154143 | 0.038621 | 0.990309 | |
| RNF217 | 2.992697 | -0.08293 | 0.999532 | | 3.22544 | -0.28165 | 0.842543 | | 2.71793 | -0.17558 | 0.916969 | |
| TSEN34 | 3.807031 | -0.46563 | 0.999532 | | 4.475209 | -0.24316 | 0.842543 | | 3.323325 | 0.240736 | 0.865363 | |
| IFNA13 | 2.026492 | -0.0314 | 0.999532 | | 1.91877 | -0.35832 | 0.842543 | | 1.836217 | -0.31081 | 0.879947 | |
| HERC2 | 3.976798 | 0.081478 | 0.999532 | | 4.026776 | -0.2553 | 0.842543 | | 3.39496 | -0.31733 | 0.800239 | |
| SOWAHA | 2.894035 | 0.00525 | 0.999532 | | 3.085053 | -0.31659 | 0.842543 | | 2.676981 | -0.30581 | 0.859255 | |
| CNOT8 | 5.729958 | -0.03593 | 0.999532 | | 6.697124 | 0.202764 | 0.842543 | | 5.785911 | 0.252825 | 0.809832 | |
| HS6ST3 | 2.481114 | -0.29348 | 0.999532 | | 2.925712 | -0.31669 | 0.842543 | | 2.235189 | 0.002465 | 0.999385 | |
| LDAH | 2.494045 | 0.738647 | 0.999532 | | 2.304767 | 0.334174 | 0.842543 | | 2.656757 | -0.38964 | 0.800239 | |
| MRPL18 | 5.593473 | -0.06148 | 0.999532 | | 7.351599 | 0.197837 | 0.842543 | | 5.766537 | 0.292496 | 0.780291 | |
| CD300LF | 2.719218 | 0.304573 | 0.999532 | | 2.510473 | -0.33151 | 0.842543 | | 2.494131 | -0.61915 | 0.660097 | |
| CLUH | 3.925188 | 0.501014 | 0.999532 | | 4.022419 | 0.253025 | 0.842543 | | 3.912953 | -0.22752 | 0.862433 | |
| HSPB3 | 2.163011 | -0.30418 | 0.999532 | | 2.445397 | -0.33882 | 0.842543 | | 1.826883 | -0.0192 | 0.996681 | |
| SF3B1 | 9.129084 | -0.26157 | 0.999532 | | 11.46322 | 0.171034 | 0.842543 | | 9.099043 | 0.445587 | 0.540342 | |
| RAB11FI | 4.084127 | -0.05263 | 0.999532 | | 5.141073 | 0.226133 | 0.842543 | | 4.142335 | 0.29653 | 0.793131 | |
| GUCY1A1 | 3.127012 | 0.108143 | 0.999532 | | 3.644003 | 0.297699 | 0.842543 | | 3.345909 | 0.221229 | 0.88082 | |
| FMC1 | 2.837697 | 0.056208 | 0.999532 | | 3.367756 | 0.270576 | 0.842666 | | 2.902543 | 0.230827 | 0.880375 | |
| TOMM40 | 5.528429 | 0.393275 | 0.999532 | | 5.089569 | -0.24055 | 0.842915 | | 5.136229 | -0.61359 | 0.499673 | |
| GNG12 | 5.505268 | -0.0863 | 0.999532 | | 6.843746 | 0.222932 | 0.842915 | | 5.61402 | 0.334013 | 0.728015 | |
| EIF2B5 | 4.367692 | 0.373585 | 0.999532 | | 3.830401 | -0.26108 | 0.842915 | | 3.706459 | -0.62162 | 0.542124 | |
| PHF20L1 | 3.326508 | -0.15322 | 0.999532 | | 4.510549 | 0.25345 | 0.843061 | | 3.596486 | 0.426038 | 0.717627 | |
| WLS | 3.703404 | -0.2486 | 0.999532 | | 5.231983 | 0.233604 | 0.843092 | | 3.997862 | 0.511564 | 0.64142 | |
| B4GALT7 | 4.417779 | 0.054822 | 0.999532 | | 4.661816 | -0.2457 | 0.843092 | | 3.995777 | -0.2757 | 0.829716 | |
| FXYD4 | 3.592677 | -0.10221 | 0.999532 | | 3.82293 | -0.26559 | 0.843092 | | 3.079573 | -0.14631 | 0.928522 | |
| RAB39B | 2.521616 | -0.4011 | 0.999532 | | 2.832967 | -0.31016 | 0.843092 | | 2.201994 | 0.111568 | 0.961426 | |
| IL3RA | 2.265504 | -0.09455 | 0.999532 | | 2.267241 | -0.32375 | 0.843092 | | 1.973107 | -0.21613 | 0.911127 | |
| COMMD7 | 3.605718 | 0.268239 | 0.999532 | | 4.060856 | 0.26149 | 0.843092 | | 3.799971 | 0.011917 | 0.996891 | |
| EDARADD | 1.989779 | 0.764443 | 0.999532 | | 1.871012 | 0.364158 | 0.843585 | | 2.127699 | -0.38449 | 0.816589 | |
| MOSPD3 | 2.641697 | -0.48495 | 0.999532 | | 2.960401 | -0.29716 | 0.843585 | | 2.287495 | 0.200088 | 0.916888 | |
| EMC7 | 6.639112 | 0.031713 | 0.999532 | | 7.787683 | 0.195195 | 0.843585 | | 6.679644 | 0.180244 | 0.877703 | |
| FGFR1 | 5.541405 | 0.008151 | 0.999532 | | 6.219501 | 0.212538 | 0.843585 | | 5.415781 | 0.220289 | 0.846786 | |
| PER3 | 2.483034 | 0.37367 | 0.999532 | | 2.695008 | 0.317538 | 0.843585 | | 2.499218 | -0.03774 | 0.989437 | |
| SERPINB | 2.646447 | -0.03472 | 0.999532 | | 3.438842 | 0.297716 | 0.843585 | | 2.869041 | 0.352444 | 0.814483 | |
| SULT1C2 | 2.883537 | 0.465847 | 0.999532 | | 2.890453 | 0.318707 | 0.843585 | | 2.986021 | -0.12823 | 0.946512 | |
| RYK | 5.132718 | -0.2158 | 0.999532 | | 7.069863 | 0.205073 | 0.843585 | | 5.609449 | 0.436893 | 0.630705 | |
| MGLL | 5.603408 | -0.206 | 0.999532 | | 7.568308 | 0.214074 | 0.843893 | | 6.024858 | 0.443005 | 0.643272 | |
| AMBN | 2.324683 | -0.00438 | 0.999532 | | 2.365187 | -0.33854 | 0.843893 | | 1.932029 | -0.31098 | 0.865065 | |
| IZUMO4 | 2.679178 | -0.3262 | 0.999532 | | 2.954491 | -0.30824 | 0.843893 | | 2.379603 | 0.03388 | 0.992286 | |
| PTS | 3.048652 | -0.13302 | 0.999532 | | 4.082732 | 0.251544 | 0.843893 | | 3.130156 | 0.3968 | 0.742183 | |
| ZNFX1 | 8.238061 | -0.08345 | 0.999532 | | 10.42997 | 0.191847 | 0.843893 | | 8.735257 | 0.289095 | 0.736481 | |
| SLC16A1 | 2.398269 | 0.251732 | 0.999532 | | 2.728095 | 0.302306 | 0.843954 | | 2.4562 | 0.065419 | 0.97733 | |
| B3GNT7 | 2.735842 | 0.027818 | 0.999532 | | 2.506061 | -0.30446 | 0.844054 | | 2.390852 | -0.31448 | 0.846373 | |
| ARMT1 | 2.007309 | -0.63976 | 0.999532 | | 2.473248 | -0.32763 | 0.844054 | | 1.718235 | 0.330402 | 0.865363 | |
| C1QA | 4.220672 | -0.59861 | 0.999532 | | 5.097539 | -0.30176 | 0.844054 | | 3.589054 | 0.329015 | 0.801502 | |
| SCGB1D4 | 2.186418 | 0.467768 | 0.999532 | | 1.871395 | -0.36695 | 0.844054 | | 2.003029 | -0.81474 | 0.565022 | |
| RAB3IP | 2.617189 | 0.230486 | 0.999532 | | 2.465937 | -0.32967 | 0.844054 | | 2.407749 | -0.5431 | 0.702463 | |
| ALDH3B2 | 3.045273 | -0.40064 | 0.999532 | | 3.475314 | -0.29579 | 0.844357 | | 2.636638 | 0.114682 | 0.958991 | |
| DDX21 | 5.651624 | -0.16338 | 0.999532 | | 6.224046 | -0.23398 | 0.844898 | | 4.912631 | -0.04096 | 0.983281 | |
| ACAD9 | 4.398957 | 0.078311 | 0.999532 | | 5.010169 | 0.237956 | 0.844898 | | 4.481169 | 0.176441 | 0.904592 | |
| MRPL30 | 6.440281 | 0.045525 | 0.999532 | | 7.710312 | 0.199904 | 0.845097 | | 6.655054 | 0.173124 | 0.87591 | |
| CCNT1 | 5.482487 | 0.017504 | 0.999532 | | 5.63298 | -0.22175 | 0.845097 | | 4.883238 | -0.22084 | 0.861119 | |
| RELT | 2.878385 | -0.00651 | 0.999532 | | 2.906676 | -0.30862 | 0.845097 | | 2.429396 | -0.28182 | 0.864923 | |
| ZC3HAV1 | 3.447067 | -0.15859 | 0.999532 | | 4.538148 | 0.242621 | 0.845097 | | 3.665437 | 0.420572 | 0.717627 | |
| KHDC1 | 2.047799 | 0.372547 | 0.999532 | | 2.288976 | 0.33474 | 0.845097 | | 2.205449 | -0.01935 | 0.996681 | |

| | | | | | | | | | | | |
|---|---|---|---|---|---|---|---|---|---|---|---|
| MANSC1 | 4.160176 | 0.114294 | 0.999532 | | 4.933926 | 0.246781 | 0.845099 | | 4.36397 | 0.148864 | 0.913083 |
| CHIC2 | 4.272445 | -0.24939 | 0.999532 | | 4.611494 | -0.25835 | 0.845168 | | 3.602264 | 0.015985 | 0.996681 |
| EPRS1 | 3.275614 | -0.23919 | 0.999532 | | 3.49829 | -0.28031 | 0.845314 | | 2.777303 | -0.0257 | 0.993435 |
| NUP155 | 3.920486 | -0.37522 | 0.999532 | | 4.632065 | -0.24945 | 0.845314 | | 3.351075 | 0.146552 | 0.925618 |
| ZFYVE28 | 2.800736 | -0.03709 | 0.999532 | | 2.977407 | -0.29261 | 0.845314 | | 2.590153 | -0.23344 | 0.889853 |
| CDKN2AI | 3.255531 | 0.448237 | 0.999532 | | 2.878063 | -0.3041 | 0.845314 | | 2.886753 | -0.72867 | 0.542124 |
| DCP1A | 3.753827 | -0.196 | 0.999532 | | 4.271061 | -0.25528 | 0.845558 | | 3.39073 | -0.04023 | 0.986208 |
| KRTAP2- | 2.195488 | -0.32505 | 0.999532 | | 2.419015 | -0.32261 | 0.845629 | | 2.018841 | 0.024106 | 0.995275 |
| ATP6V0D | 3.707402 | 0.01824 | 0.999532 | | 3.675694 | -0.31631 | 0.845669 | | 3.070599 | -0.30707 | 0.852482 |
| ZNF746 | 3.159405 | -0.34236 | 0.999532 | | 3.651431 | -0.2666 | 0.845747 | | 2.683683 | 0.095767 | 0.96129 |
| CALHM6 | 2.787033 | 0.019678 | 0.999532 | | 2.932573 | -0.29344 | 0.845817 | | 2.453619 | -0.2943 | 0.853018 |
| CFAP20 | 3.825976 | 0.013364 | 0.999532 | | 4.845894 | 0.239436 | 0.845993 | | 4.099686 | 0.24838 | 0.849532 |
| EPB41 | 3.375614 | 0.261647 | 0.999532 | | 3.031221 | -0.30003 | 0.845993 | | 2.974724 | -0.54997 | 0.686214 |
| AMER1 | 2.848829 | 0.528579 | 0.999532 | | 2.841743 | 0.306047 | 0.845993 | | 2.991237 | -0.20317 | 0.899732 |
| TRIT1 | 3.311366 | 0.579205 | 0.999532 | | 3.100336 | 0.283179 | 0.845993 | | 3.491838 | -0.29168 | 0.844697 |
| UQCRFS | 2.404445 | 0.117622 | 0.999532 | | 3.005538 | 0.286338 | 0.845993 | | 2.727936 | 0.187485 | 0.907882 |
| APLN | 5.087604 | -0.36103 | 0.999532 | | 6.794032 | 0.21485 | 0.845993 | | 5.360585 | 0.584738 | 0.543177 |
| MAN1B1 | 4.16268 | -0.08553 | 0.999532 | | 5.211344 | 0.238181 | 0.845993 | | 4.35866 | 0.340138 | 0.794815 |
| DNAJC4 | 5.398533 | 0.374384 | 0.999532 | | 5.638634 | 0.229435 | 0.846253 | | 5.432035 | -0.12236 | 0.923864 |
| LPCAT2 | 5.1336 | -0.04808 | 0.999532 | | 6.685837 | 0.217265 | 0.846253 | | 5.324149 | 0.288744 | 0.806797 |
| BMP6 | 2.295871 | 0.442525 | 0.999532 | | 1.952671 | -0.34809 | 0.846393 | | 2.095616 | -0.7726 | 0.566812 |
| MAB21L | 1.901884 | -0.08174 | 0.999532 | | 1.961133 | -0.36414 | 0.846393 | | 1.636758 | -0.26368 | 0.901033 |
| CAMLG | 4.064061 | 0.119817 | 0.999532 | | 4.675701 | 0.230858 | 0.846424 | | 4.040302 | 0.12959 | 0.92697 |
| OSCP1 | 2.829441 | 0.06574 | 0.999532 | | 2.517886 | -0.3136 | 0.846424 | | 2.425876 | -0.37568 | 0.834165 |
| VCX3A | 2.770583 | 0.29437 | 0.999532 | | 2.533687 | -0.31226 | 0.846424 | | 2.457204 | -0.58946 | 0.663104 |
| TIPARP | 2.376551 | -0.27863 | 0.999532 | | 2.561279 | -0.31992 | 0.846651 | | 2.164892 | -0.02281 | 0.995943 |
| TNFSF12 | 3.751378 | -0.33293 | 0.999532 | | 4.880928 | 0.23244 | 0.846651 | | 3.905597 | 0.57685 | 0.57215 |
| OTULINL | 2.08422 | 0.076727 | 0.999532 | | 2.369044 | 0.313433 | 0.846651 | | 2.067306 | 0.251176 | 0.88885 |
| SLC1A2 | 2.672674 | 0.375275 | 0.999532 | | 2.460146 | -0.34162 | 0.846651 | | 2.434626 | -0.69473 | 0.642446 |
| ENTPD6 | 4.117026 | -0.20843 | 0.999532 | | 4.456159 | -0.25003 | 0.846651 | | 3.43717 | -0.02458 | 0.992586 |
| PROSER | 4.326847 | -0.0091 | 0.999532 | | 4.478685 | -0.24369 | 0.846651 | | 3.775128 | -0.21673 | 0.874251 |
| ATG12 | 5.530829 | -0.00143 | 0.999532 | | 6.919532 | 0.220177 | 0.846651 | | 5.961886 | 0.236044 | 0.835289 |
| CLASP1 | 5.692116 | -0.27035 | 0.999532 | | 7.377607 | 0.201116 | 0.846651 | | 5.901382 | 0.489926 | 0.57215 |
| DZANK1 | 2.324145 | 0.719384 | 0.999532 | | 2.390798 | 0.344697 | 0.846651 | | 2.377231 | -0.34627 | 0.829449 |
| BCOR | 2.890501 | -0.2214 | 0.999532 | | 3.95877 | 0.264511 | 0.846686 | | 3.122094 | 0.507395 | 0.678541 |
| DCUN1D | 3.327876 | 0.353425 | 0.999532 | | 3.742576 | 0.266105 | 0.846696 | | 3.342481 | -0.05592 | 0.977514 |
| ZNF12 | 2.647746 | -0.09323 | 0.999532 | | 2.781796 | -0.29941 | 0.84691 | | 2.256215 | -0.18914 | 0.91707 |
| CXADR | 7.728185 | 0.623142 | 0.999532 | | 7.328011 | 0.211546 | 0.847185 | | 7.824595 | -0.39631 | 0.655993 |
| CCDC57 | 2.507646 | 1.051402 | 0.999532 | | 2.102997 | 0.34382 | 0.847185 | | 2.586638 | -0.69228 | 0.563285 |
| SIK2 | 3.160373 | -0.42423 | 0.999532 | | 4.382693 | 0.246043 | 0.847185 | | 3.163116 | 0.690332 | 0.524433 |
| EIF2AK3 | 4.640755 | 0.158962 | 0.999532 | | 4.694749 | -0.26039 | 0.847185 | | 4.123107 | -0.39772 | 0.723933 |
| PDGFB | 3.96465 | -0.55159 | 0.999532 | | 4.99131 | -0.23916 | 0.847185 | | 3.442327 | 0.330406 | 0.806394 |
| RGS3 | 5.067191 | -0.34232 | 0.999532 | | 5.589754 | -0.21973 | 0.847185 | | 4.423046 | 0.132529 | 0.926702 |
| ZNF562 | 3.537304 | 0.272112 | 0.999532 | | 3.292492 | -0.27836 | 0.847239 | | 3.31549 | -0.53242 | 0.659727 |
| EVC2 | 2.378422 | 0.4006 | 0.999532 | | 2.701454 | 0.301481 | 0.847239 | | 2.538572 | -0.07839 | 0.972221 |
| CXCL17 | 2.81073 | -0.16909 | 0.999532 | | 3.211207 | -0.30478 | 0.847239 | | 2.429956 | -0.11205 | 0.958191 |
| C1orf174 | 4.698763 | -0.52064 | 0.999532 | | 6.17203 | -0.22422 | 0.847239 | | 4.207633 | 0.317899 | 0.791576 |
| PDGFC | 3.578219 | 0.146839 | 0.999532 | | 3.527669 | -0.27093 | 0.847239 | | 3.21337 | -0.39907 | 0.750371 |
| SPATA2L | 8.835806 | 0.525841 | 0.999532 | | 9.077514 | 0.300979 | 0.847239 | | 9.382892 | -0.20689 | 0.920259 |
| SERPINE | 3.088197 | 0.468528 | 0.999532 | | 3.503223 | 0.284037 | 0.847239 | | 3.199226 | -0.16325 | 0.915451 |
| RPS25 | 14.15981 | -0.25611 | 0.999532 | | 17.94493 | 0.135699 | 0.847239 | | 13.93643 | 0.400267 | 0.535229 |
| RAB4A | 5.315317 | 0.164633 | 0.999532 | | 6.148761 | 0.206245 | 0.847239 | | 5.42701 | 0.066822 | 0.968235 |
| CHCHD3 | 5.637811 | -0.21596 | 0.999532 | | 6.463559 | -0.21371 | 0.847239 | | 4.860969 | 0.020542 | 0.992286 |
| FAM234B | 2.886712 | -0.06157 | 0.999532 | | 3.086155 | -0.28417 | 0.847298 | | 2.490722 | -0.19556 | 0.914929 |
| IKZF2 | 2.634053 | 0.015201 | 0.999532 | | 2.723811 | -0.31948 | 0.847298 | | 2.315895 | -0.3106 | 0.859898 |
| PDK2 | 3.831169 | 0.106463 | 0.999532 | | 4.610008 | 0.257159 | 0.847449 | | 3.889939 | 0.175768 | 0.900583 |
| KBTBD1 | 4.276995 | 0.609168 | 0.999532 | | 4.276816 | 0.267733 | 0.847498 | | 4.391011 | -0.32038 | 0.791576 |
| GZMB | 3.050696 | 0.401233 | 0.999532 | | 3.225358 | 0.286454 | 0.847628 | | 3.284406 | -0.09308 | 0.961948 |
| ATP6V0A | 2.284059 | 0.758142 | 0.999532 | | 1.96341 | 0.336766 | 0.847727 | | 2.363703 | -0.40304 | 0.809498 |
| ADAM11 | 2.421398 | 0.885215 | 0.999532 | | 2.209342 | 0.339679 | 0.847809 | | 2.538053 | -0.52715 | 0.717627 |
| G2E3 | 3.492887 | -0.08914 | 0.999532 | | 3.901947 | -0.2543 | 0.847828 | | 3.031418 | -0.14298 | 0.93222 |
| RERG | 9.181449 | -0.00604 | 0.999532 | | 11.33946 | 0.208271 | 0.84793 | | 9.396586 | 0.235234 | 0.806797 |
| GNAO1 | 2.530559 | 0.146818 | 0.999532 | | 2.462224 | -0.34612 | 0.84793 | | 2.171836 | -0.46981 | 0.779082 |
| SYF2 | 4.829269 | 0.239185 | 0.999532 | | 5.055049 | 0.229301 | 0.84793 | | 4.828529 | -0.01069 | 0.996891 |
| CIC | 6.575826 | 0.131397 | 0.999532 | | 7.413895 | 0.204682 | 0.84793 | | 6.688945 | 0.092816 | 0.944635 |
| PRRT3 | 3.643501 | 0.132855 | 0.999532 | | 3.455111 | -0.2902 | 0.84793 | | 3.173574 | -0.39267 | 0.776359 |
| PRSS36 | 2.304677 | 0.382714 | 0.999532 | | 2.022035 | -0.34242 | 0.84793 | | 2.137279 | -0.70623 | 0.62216 |
| GZMM | 3.503305 | 0.597873 | 0.999532 | | 2.782025 | -0.30889 | 0.84793 | | 3.131314 | -0.89519 | 0.426859 |
| SOX1 | 2.262816 | 0.083272 | 0.999532 | | 2.105363 | -0.35038 | 0.84793 | | 1.901092 | -0.41688 | 0.816372 |
| BDKRB1 | 2.269766 | -0.18555 | 0.999532 | | 2.268312 | -0.32837 | 0.84793 | | 1.941081 | -0.13469 | 0.955167 |
| TGDS | 2.945429 | 0.529279 | 0.999532 | | 3.078563 | 0.28683 | 0.84793 | | 3.064302 | -0.22258 | 0.887811 |
| CITED4 | 2.973725 | -0.14531 | 0.999532 | | 3.348783 | -0.27445 | 0.84793 | | 2.689354 | -0.10386 | 0.958798 |
| VPS51 | 4.225587 | 0.166893 | 0.999532 | | 4.618871 | 0.243155 | 0.84793 | | 4.384108 | 0.081903 | 0.96556 |
| CRYGB | 1.980461 | 0.569274 | 0.999532 | | 2.040598 | 0.35188 | 0.84793 | | 2.046716 | -0.19801 | 0.918675 |
| TMEM22 | 3.843575 | 0.574892 | 0.999532 | | 3.204246 | -0.2992 | 0.84793 | | 3.439819 | -0.85638 | 0.423641 |
| KIAA167 | 3.832438 | 0.417964 | 0.999532 | | 3.709686 | 0.25733 | 0.84793 | | 4.028676 | -0.10056 | 0.952656 |
| TRIM14 | 3.172604 | 0.156207 | 0.999532 | | 3.05081 | -0.29107 | 0.84793 | | 2.85984 | -0.43207 | 0.752155 |
| HAUS4 | 3.760899 | -0.26076 | 0.999532 | | 4.312529 | -0.24802 | 0.84793 | | 3.419527 | 0.030424 | 0.990309 |
| HNRNPC | 22.54207 | 0.208173 | 0.999532 | | 24.97308 | 0.150937 | 0.84793 | | 22.51201 | -0.02922 | 0.982124 |
| CPVL | 2.687133 | -0.26625 | 0.999532 | | 2.939662 | -0.29612 | 0.84793 | | 2.355457 | -0.01365 | 0.996891 |
| RNASEH | 2.811068 | 0.33235 | 0.999532 | | 3.236706 | 0.271228 | 0.84793 | | 3.053478 | -0.04218 | 0.985675 |
| YTHDF1 | 3.55898 | -0.12317 | 0.999532 | | 4.687767 | 0.251504 | 0.84793 | | 3.82451 | 0.392699 | 0.738631 |
| FIBIN | 3.212911 | -0.14837 | 0.999532 | | 4.191656 | 0.251308 | 0.84793 | | 3.386042 | 0.417735 | 0.746876 |
| POMC | 2.256002 | 0.349514 | 0.999532 | | 2.613142 | 0.310436 | 0.84793 | | 2.467677 | -0.02127 | 0.995112 |
| ADAM12 | 2.583906 | -0.14135 | 0.999532 | | 3.374603 | 0.271243 | 0.84793 | | 2.730926 | 0.430101 | 0.744708 |
| GPR31 | 2.28266 | 0.191072 | 0.999532 | | 2.079423 | -0.35652 | 0.84793 | | 1.911769 | -0.52799 | 0.747457 |
| ABI1 | 4.276048 | 0.010511 | 0.999532 | | 5.303071 | 0.232844 | 0.84793 | | 4.447606 | 0.241387 | 0.84579 |
| ARID1B | 3.977934 | 0.403703 | 0.999532 | | 4.297349 | 0.252474 | 0.84793 | | 4.080991 | -0.14246 | 0.914929 |
| EVI5L | 3.422496 | 0.436347 | 0.999532 | | 3.692202 | 0.259494 | 0.84793 | | 3.664658 | -0.15779 | 0.913136 |
| PHF8 | 3.638847 | 0.511184 | 0.999532 | | 3.712109 | 0.251083 | 0.84793 | | 3.814418 | -0.23948 | 0.857043 |
| OR8B8 | 2.966101 | 0.285451 | 0.999532 | | 2.445909 | -0.31495 | 0.84793 | | 2.651069 | -0.58833 | 0.702463 |
| C3AR1 | 3.640827 | 0.257397 | 0.999532 | | 3.904358 | 0.258111 | 0.84793 | | 3.827958 | 0.014987 | 0.996681 |
| OR1M1 | 2.451944 | 0.16367 | 0.999532 | | 2.282876 | -0.32987 | 0.847943 | | 2.138362 | -0.47829 | 0.75311 |
| GOSR2 | 2.953098 | 0.196206 | 0.999532 | | 3.416954 | 0.268945 | 0.847971 | | 3.095793 | 0.091027 | 0.96129 |
| ARMCX3 | 6.553803 | -0.13344 | 0.999532 | | 8.359644 | 0.189798 | 0.847971 | | 6.687425 | 0.34561 | 0.701419 |
| ACOT4 | 2.886531 | 0.020173 | 0.999532 | | 3.018575 | -0.28832 | 0.847971 | | 2.683675 | -0.2866 | 0.862147 |
| CACNA1( | 3.829029 | 0.317182 | 0.999532 | | 4.160435 | 0.261853 | 0.847971 | | 3.778625 | -0.02478 | 0.992286 |
| BRCC3 | 3.259634 | 0.043157 | 0.999532 | | 3.557709 | -0.26848 | 0.847971 | | 3.02772 | -0.29006 | 0.839373 |
| ZSWIM1 | 2.364699 | -0.08143 | 0.999532 | | 2.447171 | -0.31511 | 0.848134 | | 2.13426 | -0.2171 | 0.906811 |
| GTF2A1L | 2.21215 | -0.18603 | 0.999532 | | 2.25255 | -0.3255 | 0.848257 | | 1.872674 | -0.12557 | 0.958191 |
| TELO2 | 3.871476 | -0.10345 | 0.999532 | | 3.916797 | -0.25821 | 0.848257 | | 3.375808 | -0.14258 | 0.929549 |
| RIMS2 | 2.111828 | 0.136971 | 0.999532 | | 2.025687 | -0.36053 | 0.848257 | | 1.812042 | -0.48077 | 0.791576 |
| GJB5 | 2.046825 | -0.03506 | 0.999532 | | 1.950632 | -0.35176 | 0.848617 | | 1.719254 | -0.30266 | 0.881837 |
| TMOD2 | 2.674236 | 0.584518 | 0.999532 | | 2.283905 | -0.34467 | 0.848617 | | 2.462698 | -0.90387 | 0.511836 |
| GSTM2 | 4.223916 | 0.489622 | 0.999532 | | 4.360311 | 0.23863 | 0.848617 | | 4.411251 | -0.23426 | 0.846786 |
| ITSN2 | 2.607161 | -0.26667 | 0.999532 | | 3.570673 | 0.269257 | 0.848617 | | 2.812912 | 0.553287 | 0.659564 |
| GLI1 | 2.103127 | -0.41731 | 0.999532 | | 2.274017 | -0.32657 | 0.848617 | | 1.771044 | 0.105803 | 0.968241 |
| CEP43 | 2.566915 | -0.08043 | 0.999532 | | 2.758139 | -0.30659 | 0.848617 | | 2.258176 | -0.20568 | 0.911296 |
| UBE2N | 3.762136 | 0.026828 | 0.999532 | | 4.70436 | 0.242588 | 0.848691 | | 3.828133 | 0.220494 | 0.866065 |
| NRROS | 7.079508 | 0.025808 | 0.999532 | | 7.213297 | -0.23269 | 0.8489 | | 6.388926 | -0.23818 | 0.823193 |
| SECISBP | 5.966569 | -0.09712 | 0.999532 | | 7.424624 | 0.191949 | 0.848948 | | 5.963068 | 0.307447 | 0.750371 |
| UTP18 | 4.078981 | -0.31993 | 0.999532 | | 4.639016 | -0.23591 | 0.848948 | | 3.815577 | 0.106096 | 0.948127 |
| ZBTB7B | 4.899791 | -0.0782 | 0.999532 | | 5.993341 | 0.218792 | 0.848948 | | 5.049221 | 0.319289 | 0.755301 |
| HOXA4 | 3.351525 | 0.106619 | 0.999532 | | 3.414886 | -0.27795 | 0.848948 | | 2.95413 | -0.36663 | 0.790208 |
| GTSF1L | 2.354777 | 0.407271 | 0.999532 | | 2.105969 | -0.35468 | 0.848948 | | 2.111067 | -0.73572 | 0.637292 |
| LAX1 | 1.99106 | -0.17401 | 0.999532 | | 2.07922 | -0.33517 | 0.848948 | | 1.791235 | -0.14453 | 0.950616 |
| IL13RA1 | 5.42253 | 0.166449 | 0.999532 | | 6.332559 | 0.205908 | 0.848948 | | 5.560667 | 0.061738 | 0.970811 |
| NAB1 | 4.177248 | 0.31174 | 0.999532 | | 4.388024 | 0.261283 | 0.848948 | | 4.363125 | -0.05336 | 0.97843 |
| MRPL9 | 3.262117 | 0.62976 | 0.999532 | | 3.12842 | 0.272962 | 0.848948 | | 3.445023 | -0.33701 | 0.791576 |
| DDX11 | 2.815435 | -0.00233 | 0.999532 | | 2.897595 | -0.3038 | 0.848948 | | 2.526703 | -0.28035 | 0.874341 |
| PRELID1 | 5.660996 | -0.07152 | 0.999532 | | 5.962275 | -0.22012 | 0.848948 | | 4.936553 | -0.13208 | 0.923223 |
| PMM1 | 2.689075 | 0.530845 | 0.999532 | | 2.795536 | 0.310929 | 0.848948 | | 2.738886 | -0.19804 | 0.906811 |
| LIPF | 1.94294 | -0.37395 | 0.999532 | | 2.255705 | -0.32878 | 0.848948 | | 1.732657 | 0.063356 | 0.983281 |
| ZBED6 | 4.743326 | -0.38978 | 0.999532 | | 5.485062 | -0.22812 | 0.848948 | | 3.855178 | 0.180889 | 0.897958 |
| CHST10 | 4.342194 | 0.055291 | 0.999532 | | 4.663609 | -0.24369 | 0.848948 | | 3.938342 | -0.27758 | 0.824012 |
| PSMB1 | 5.819838 | -0.16095 | 0.999532 | | 6.343282 | -0.21576 | 0.848948 | | 5.35975 | -0.01801 | 0.994537 |
| ADHFE1 | 3.835795 | 0.427582 | 0.999532 | | 4.02015 | 0.256514 | 0.848948 | | 4.004961 | -0.15204 | 0.916511 |
| VSIG10 | 5.682571 | 0.430581 | 0.999532 | | 4.954602 | -0.23182 | 0.848948 | | 5.203259 | -0.64785 | 0.465298 |
| LIG4 | 2.684681 | 0.108002 | 0.999532 | | 2.739035 | -0.29847 | 0.848948 | | 2.358775 | -0.38663 | 0.801409 |
| PDE7B | 3.441187 | -0.18281 | 0.999532 | | 4.354813 | 0.237089 | 0.848948 | | 3.57987 | 0.444149 | 0.70363 |
| FGF12 | 1.95413 | -0.04126 | 0.999532 | | 2.146952 | -0.3364 | 0.848948 | | 1.863076 | -0.27141 | 0.890225 |
| HOXA5 | 2.67184 | 0.129812 | 0.999532 | | 2.992398 | 0.276275 | 0.848948 | | 2.695934 | 0.161871 | 0.923864 |
| NUBPL | 5.103778 | -0.00924 | 0.999532 | | 5.296619 | -0.22938 | 0.848948 | | 4.406284 | -0.19922 | 0.87591 |
| ANTXR2 | 4.892488 | -0.14001 | 0.999532 | | 6.019039 | 0.215613 | 0.848948 | | 5.056011 | 0.373292 | 0.720106 |
| OR6C70 | 2.000603 | -0.03644 | 0.999532 | | 2.14063 | -0.33962 | 0.848948 | | 1.849369 | -0.28125 | 0.886495 |
| ACSM6 | 2.470306 | 0.155653 | 0.999532 | | 2.235834 | -0.33363 | 0.848948 | | 2.157691 | -0.4618 | 0.766832 |
| KIAA010( | 4.229209 | -0.14708 | 0.999532 | | 5.016837 | 0.228551 | 0.848948 | | 4.236862 | 0.392212 | 0.718643 |
| TRIM5 | 3.477779 | 0.124277 | 0.999532 | | 3.962716 | 0.255342 | 0.848948 | | 3.682724 | 0.143131 | 0.927809 |
| MEAF6 | 6.248223 | -0.15062 | 0.999532 | | 7.833986 | 0.187195 | 0.848948 | | 6.250384 | 0.358117 | 0.673405 |
| TIAF1 | 2.769132 | -0.3201 | 0.999532 | | 3.063906 | -0.27938 | 0.849531 | | 2.420864 | 0.0545 | 0.982124 |
| ZFP30 | 3.290382 | -0.38326 | 0.999532 | | 3.977234 | -0.26373 | 0.849689 | | 2.851764 | 0.139431 | 0.938624 |
| RBM17 | 4.478538 | -0.28762 | 0.999532 | | 6.027403 | 0.206125 | 0.849689 | | 4.632242 | 0.512621 | 0.579472 |
| TMEM14 | 3.400928 | -0.08762 | 0.999532 | | 4.302271 | 0.237229 | 0.849794 | | 3.493292 | 0.345028 | 0.790266 |
| BRWD3 | 2.813356 | 0.101709 | 0.999532 | | 3.402707 | 0.272727 | 0.850143 | | 3.018991 | 0.192704 | 0.905952 |
| CACNA2 | 1.871614 | -0.21946 | 0.999532 | | 2.023746 | -0.34743 | 0.850143 | | 1.59479 | -0.10902 | 0.968434 |
| ZNF587B | 3.44179 | 0.244305 | 0.999532 | | 3.207064 | -0.2685 | 0.850143 | | 2.964706 | -0.4998 | 0.682403 |
| TMEM14E | 5.127709 | 0.038579 | 0.999532 | | 6.002941 | 0.212698 | 0.850143 | | 5.173432 | 0.196083 | 0.868182 |
| PPP2R2C | 2.527451 | -0.15962 | 0.999532 | | 2.828119 | -0.33936 | 0.850143 | | 2.316185 | -0.15448 | 0.946512 |
| BRD3OS | 2.762107 | -0.04653 | 0.999532 | | 2.808519 | -0.28984 | 0.850143 | | 2.39779 | -0.22687 | 0.893262 |
| YJU2 | 4.246532 | 0.465161 | 0.999532 | | 4.279696 | 0.254008 | 0.850143 | | 4.241915 | -0.18426 | 0.889853 |
| MAP1LC3 | 5.477759 | 0.066258 | 0.999532 | | 6.349124 | 0.205856 | 0.850143 | | 5.55487 | 0.153103 | 0.90424 |
| C19orf25 | 3.832165 | 0.475027 | 0.999532 | | 3.97565 | 0.271236 | 0.850143 | | 4.153843 | -0.19031 | 0.899161 |
| FBXL4 | 2.771841 | -0.32823 | 0.999532 | | 3.735826 | 0.278525 | 0.850143 | | 2.892218 | 0.627127 | 0.621895 |
| GUCY1B1 | 3.303481 | 0.518261 | 0.999532 | | 3.326288 | 0.260737 | 0.850143 | | 3.505575 | -0.24011 | 0.861737 |
| NGDN | 3.407273 | -0.48413 | 0.999532 | | 3.933117 | -0.25534 | 0.850143 | | 2.844214 | 0.225589 | 0.894612 |
| GTF3C2 | 3.391322 | 0.056707 | 0.999532 | | 3.767257 | 0.249634 | 0.850143 | | 3.576962 | 0.213076 | 0.88885 |
| C2orf15 | 3.153581 | 0.621593 | 0.999532 | | 2.735819 | -0.3164 | 0.850143 | | 2.855395 | -0.91434 | 0.412398 |
| MTERF3 | 2.276062 | -0.02334 | 0.999532 | | 2.867619 | 0.289118 | 0.850143 | | 2.396996 | 0.328049 | 0.842754 |
| GPR158 | 3.078896 | -0.22566 | 0.999532 | | 3.295761 | -0.28506 | 0.850143 | | 2.697085 | -0.04864 | 0.985693 |
| IL32 | 11.20764 | -0.37601 | 0.999532 | | 15.50055 | 0.241229 | 0.850143 | | 11.96829 | 0.640369 | 0.439373 |
| MRPL48 | 2.997713 | 0.516005 | 0.999532 | | 3.036666 | 0.285707 | 0.850143 | | 3.205033 | -0.20952 | 0.893262 |
| FAM193B | 5.78944 | 0.242556 | 0.999532 | | 6.349637 | 0.202521 | 0.850168 | | 5.807848 | -0.02124 | 0.99176 |
| QSER1 | 4.551684 | -0.13372 | 0.999532 | | 4.844285 | -0.22456 | 0.850168 | | 4.011659 | -0.07472 | 0.96556 |
| MZT1 | 3.389684 | 0.180582 | 0.999532 | | 3.289329 | -0.27657 | 0.850199 | | 3.090308 | -0.44132 | 0.723933 |
| ELOC | 14.45834 | -0.45339 | 0.999532 | | 17.90771 | -0.16201 | 0.850199 | | 12.96652 | 0.313961 | 0.652925 |
| SMAP1 | 2.305468 | 0.25885 | 0.999532 | | 2.704405 | 0.300058 | 0.850199 | | 2.591699 | 0.058308 | 0.979282 |
| ADAM23 | 1.847319 | 0.002678 | 0.999532 | | 1.809992 | -0.35696 | 0.850199 | | 1.606936 | -0.34318 | 0.864923 |
| APOF | 2.341574 | 0.375614 | 0.999532 | | 1.944636 | -0.34706 | 0.850199 | | 2.023432 | -0.70661 | 0.644433 |
| INPP5D | 1.776137 | 0.207633 | 0.999532 | | 2.099619 | 0.334304 | 0.850199 | | 1.911609 | 0.145378 | 0.946572 |
| TBC1D15 | 3.296087 | 0.068235 | 0.999532 | | 4.218095 | 0.274766 | 0.850199 | | 3.605084 | 0.233472 | 0.881689 |
| APEH | 2.263818 | 0.066643 | 0.999532 | | 2.720534 | 0.289903 | 0.850199 | | 2.32324 | 0.238685 | 0.88885 |
| ADGRL2 | 7.881487 | -0.47283 | 0.999532 | | 10.85485 | 0.181435 | 0.850199 | | 8.047098 | 0.678309 | 0.283338 |
| KRTDAP | 2.201322 | 0.156369 | 0.999532 | | 2.216162 | -0.3338 | 0.850199 | | 2.0319 | -0.46674 | 0.779182 |
| TSBP1 | 1.977306 | 0.239699 | 0.999532 | | 1.925968 | -0.35427 | 0.850199 | | 1.743142 | -0.56835 | 0.740093 |
| WDR97 | 2.080137 | 0.149928 | 0.999532 | | 1.94977 | -0.35093 | 0.850199 | | 1.769367 | -0.48488 | 0.78258 |
| FAM102A | 3.582516 | -0.27075 | 0.999532 | | 3.826934 | -0.27435 | 0.850199 | | 2.959781 | 0.019485 | 0.99573 |
| PHF21A | 3.160344 | -0.05874 | 0.999532 | | 4.008515 | 0.256869 | 0.850199 | | 3.259809 | 0.33727 | 0.801572 |
| PRUNE1 | 2.426691 | -0.01504 | 0.999532 | | 2.953491 | 0.276423 | 0.850199 | | 2.591031 | 0.310612 | 0.83987 |
| HSD3B1 | 3.099225 | 0.755436 | 0.999532 | | 2.30928 | -0.31807 | 0.850199 | | 2.695965 | -1.06204 | 0.29867 |
| ATP6V1H | 3.538601 | 0.001694 | 0.999532 | | 3.83636 | -0.25707 | 0.850199 | | 3.277696 | -0.2398 | 0.8691 |
| THYN1 | 5.671812 | 0.242176 | 0.999532 | | 6.167017 | 0.205994 | 0.850199 | | 6.005935 | -0.00759 | 0.996891 |
| GHRL | 2.332094 | 0.489207 | 0.999532 | | 1.814484 | -0.3541 | 0.850199 | | 2.073605 | -0.8325 | 0.572084 |
| EFR3A | 4.218512 | -0.06759 | 0.999532 | | 4.799658 | 0.224926 | 0.850199 | | 4.284959 | 0.305219 | 0.794815 |
| TCEAL7 | 2.003145 | -0.40623 | 0.999532 | | 2.186867 | -0.32181 | 0.850199 | | 1.692049 | 0.098454 | 0.971732 |
| PPP1R3E | 3.688803 | 0.207711 | 0.999532 | | 3.250089 | -0.26589 | 0.850199 | | 3.076976 | -0.46577 | 0.707142 |
| YBX3 | 14.29626 | -0.21326 | 0.999532 | | 16.42011 | -0.14459 | 0.850199 | | 12.69152 | 0.087194 | 0.920259 |
| LSM1 | 3.198083 | -0.08899 | 0.999532 | | 3.743099 | 0.258914 | 0.850199 | | 3.324311 | 0.35514 | 0.803301 |
| LEF1 | 2.351587 | -0.07695 | 0.999532 | | 2.412924 | -0.31688 | 0.850199 | | 2.004175 | -0.22353 | 0.906811 |
| OSBPL5 | 4.363487 | -0.00052 | 0.999532 | | 4.687736 | -0.22894 | 0.850199 | | 3.92389 | -0.20925 | 0.87591 |
| CYP8B1 | 4.118219 | 0.674476 | 0.999532 | | 3.277274 | -0.27567 | 0.850199 | | 3.696084 | -0.93278 | 0.30001 |
| ALDH16A | 2.116646 | 0.359723 | 0.999532 | | 2.327 | 0.311767 | 0.850199 | | 2.238144 | -0.03071 | 0.992286 |
| KRTAP10 | 2.205664 | 0.135596 | 0.999532 | | 2.120368 | -0.34198 | 0.850199 | | 1.851342 | -0.46015 | 0.790999 |
| RHOU | 3.043762 | 0.194636 | 0.999532 | | 3.456413 | 0.275412 | 0.850199 | | 3.17872 | 0.097888 | 0.95999 |
| ATXN1 | 7.791931 | 0.0725 | 0.999532 | | 9.314401 | 0.179379 | 0.850199 | | 7.99726 | 0.128533 | 0.901813 |
| LRBA | 2.443067 | 0.360801 | 0.999532 | | 2.809195 | 0.300226 | 0.850406 | | 2.490769 | -0.05505 | 0.979996 |
| NOCT | 2.726537 | 0.485634 | 0.999532 | | 2.33995 | -0.33845 | 0.850519 | | 2.455211 | -0.81079 | 0.524206 |
| ADAMTS | 2.932964 | 0.313776 | 0.999532 | | 3.18361 | 0.288721 | 0.850611 | | 2.8614 | 0.003875 | 0.998898 |
| SHKBP1 | 3.265978 | -0.18901 | 0.999532 | | 4.325927 | 0.238147 | 0.850611 | | 3.407563 | 0.438471 | 0.706749 |
| FKBP10 | 8.789649 | -0.52615 | 0.999532 | | 12.52146 | 0.207761 | 0.850668 | | 9.137221 | 0.758 | 0.271669 |
| DTX2 | 2.248138 | -0.14104 | 0.999532 | | 2.884286 | 0.29099 | 0.850752 | | 2.228212 | 0.445978 | 0.754304 |
| SIRT6 | 9.590522 | -0.04937 | 0.999532 | | 11.84259 | 0.201871 | 0.851258 | | 9.754178 | 0.279482 | 0.785012 |
| ACO1 | 3.148559 | 0.012334 | 0.999532 | | 3.629191 | 0.267011 | 0.851261 | | 3.110722 | 0.272399 | 0.857043 |
| DUSP10 | 2.561172 | 0.789334 | 0.999532 | | 2.569043 | 0.342865 | 0.851261 | | 2.725163 | -0.42375 | 0.776852 |
| CHAF1A | 1.972604 | -0.26219 | 0.999532 | | 2.724357 | 0.315585 | 0.851317 | | 2.119752 | 0.600399 | 0.701826 |
| MLXIPL | 3.828314 | 0.400258 | 0.999532 | | 4.050994 | 0.260139 | 0.851317 | | 3.89254 | -0.12071 | 0.944635 |
| CSDC2 | 6.18146 | 0.284671 | 0.999532 | | 7.000533 | 0.210153 | 0.851317 | | 6.339359 | -0.05917 | 0.967145 |
| KLRK1 | 2.028337 | -0.15271 | 0.999532 | | 2.090308 | -0.33648 | 0.851317 | | 1.805515 | -0.16373 | 0.943307 |
| FAM71F2 | 2.190975 | 0.271398 | 0.999532 | | 2.658756 | 0.313577 | 0.851317 | | 2.339353 | 0.061593 | 0.978931 |
| LSM10 | 2.328479 | 0.394087 | 0.999532 | | 2.411072 | 0.301385 | 0.85156 | | 2.299541 | -0.08072 | 0.972221 |
| NCBP3 | 2.646774 | -0.09382 | 0.999532 | | 3.311134 | 0.267601 | 0.85184 | | 2.756726 | 0.375033 | 0.794049 |
| KIF3B | 5.641082 | 0.057593 | 0.999532 | | 6.768713 | 0.20191 | 0.85184 | | 5.810082 | 0.165033 | 0.88885 |
| CHMP2A | 5.903444 | 0.076749 | 0.999532 | | 7.025677 | 0.212333 | 0.852044 | | 6.171909 | 0.148395 | 0.896374 |
| PGAP6 | 6.210839 | -0.0776 | 0.999532 | | 7.499756 | 0.194213 | 0.852044 | | 6.393636 | 0.283274 | 0.782255 |
| ABHD12B | 2.909497 | 0.36041 | 0.999532 | | 2.689139 | -0.31867 | 0.85224 | | 2.525843 | -0.65812 | 0.628895 |
| NSMAF | 2.416626 | 0.338136 | 0.999532 | | 2.379396 | 0.304891 | 0.852564 | | 2.54214 | -0.03663 | 0.990864 |
| ATP8B1 | 4.724005 | -0.25951 | 0.999532 | | 6.447015 | 0.214125 | 0.852564 | | 5.210216 | 0.499108 | 0.632923 |
| HLA-DRB | 38.96965 | -0.23835 | 0.999532 | | 51.11053 | 0.220956 | 0.852564 | | 40.65989 | 0.482229 | 0.414073 |
| SEH1L | 2.781483 | -0.1401 | 0.999532 | | 3.651197 | 0.26669 | 0.852564 | | 2.81096 | 0.432739 | 0.75311 |
| GJD4 | 3.96154 | 0.441963 | 0.999532 | | 4.270388 | 0.263779 | 0.852566 | | 4.316045 | -0.17382 | 0.904644 |
| IDH3G | 6.034849 | -0.15736 | 0.999532 | | 7.4432 | 0.193474 | 0.852566 | | 6.271492 | 0.363889 | 0.702463 |
| SCIN | 2.068208 | 0.346766 | 0.999532 | | 2.476412 | 0.31063 | 0.852566 | | 2.194562 | -0.01443 | 0.996891 |
| PBDC1 | 2.713787 | -0.17601 | 0.999532 | | 3.073945 | -0.27355 | 0.852566 | | 2.335807 | -0.07745 | 0.972221 |
| DHX30 | 2.939811 | -0.2569 | 0.999532 | | 4.177253 | 0.291625 | 0.852865 | | 3.497262 | 0.543224 | 0.694864 |
| PLIN5 | 4.454221 | 0.305052 | 0.999532 | | 4.784828 | 0.231878 | 0.853242 | | 4.574371 | -0.05483 | 0.976783 |
| TACR1 | 2.558633 | 0.281152 | 0.999532 | | 2.368713 | -0.32413 | 0.853242 | | 2.323622 | -0.58604 | 0.686214 |
| SLC39A4 | 2.132793 | 0.439264 | 0.999532 | | 2.399426 | 0.320381 | 0.853242 | | 2.172244 | -0.10258 | 0.961567 |
| ZNF32 | 4.496964 | 0.293398 | 0.999532 | | 5.350074 | 0.222707 | 0.853242 | | 4.873938 | -0.03101 | 0.989093 |
| MORC1 | 2.419955 | 0.302744 | 0.999532 | | 2.715817 | 0.310848 | 0.853242 | | 2.661124 | 0.023098 | 0.995112 |
| POU3F2 | 2.106749 | 0.017872 | 0.999532 | | 2.101784 | -0.33811 | 0.853242 | | 1.807536 | -0.33794 | 0.860077 |
| OSBPL1 | 2.21711 | -0.32821 | 0.999532 | | 2.52909 | -0.30523 | 0.853242 | | 2.024414 | 0.042423 | 0.989638 |
| DIPK2A | 3.149992 | -0.14289 | 0.999532 | | 3.896072 | 0.24762 | 0.853335 | | 3.290978 | 0.411723 | 0.735469 |
| COX15 | 4.778391 | 0.300979 | 0.999532 | | 4.981704 | 0.23788 | 0.853416 | | 4.880585 | -0.06396 | 0.97358 |
| NSRP1 | 3.569423 | -0.34762 | 0.999532 | | 4.943709 | 0.228981 | 0.853416 | | 3.781328 | 0.597315 | 0.566812 |
| ZDHHC12 | 4.781462 | -0.00526 | 0.999532 | | 5.138081 | -0.22786 | 0.85374 | | 4.254091 | -0.20179 | 0.877943 |
| SLC49A3 | 3.127562 | 0.749299 | 0.999532 | | 2.815503 | 0.286058 | 0.85384 | | 3.064316 | -0.44022 | 0.718468 |
| FAM133B | 2.508797 | 0.619499 | 0.999532 | | 1.884065 | -0.34611 | 0.854122 | | 2.312362 | -0.95134 | 0.474973 |
| TRIM24 | 4.402311 | 0.301448 | 0.999532 | | 4.114807 | -0.23632 | 0.854263 | | 3.931658 | -0.51577 | 0.655993 |
| HLA-E | 96.36099 | -0.18388 | 0.999532 | | 122.0754 | 0.162792 | 0.854263 | | 98.24178 | 0.369264 | 0.546858 |
| TMEM59L | 2.402174 | -0.1413 | 0.999532 | | 2.593105 | -0.30687 | 0.854263 | | 2.194389 | -0.14526 | 0.946311 |
| CDK8 | 2.727247 | -0.44305 | 0.999532 | | 3.163053 | -0.2626 | 0.854263 | | 2.453603 | 0.198013 | 0.907882 |
| OR10G9 | 2.177545 | 0.187891 | 0.999532 | | 2.044349 | -0.35749 | 0.854263 | | 1.868089 | -0.52755 | 0.754906 |
| ACTC1 | 1.963107 | -0.41617 | 0.999532 | | 2.38148 | -0.33576 | 0.854263 | | 1.840631 | 0.088158 | 0.974421 |
| PIP5K1C | 5.000339 | -0.10163 | 0.999532 | | 6.627149 | 0.202488 | 0.854263 | | 5.234615 | 0.317336 | 0.750995 |
| ESCO2 | 3.137245 | 0.736629 | 0.999532 | | 3.185224 | 0.313371 | 0.854263 | | 3.340841 | -0.40518 | 0.761699 |
| GAA | 4.689205 | -0.14628 | 0.999532 | | 5.134562 | -0.21737 | 0.854263 | | 4.16643 | -0.0511 | 0.977635 |
| NDUFS7 | 2.160088 | 0.401875 | 0.999532 | | 2.264821 | 0.310188 | 0.854727 | | 2.229325 | -0.07436 | 0.974421 |
| ENC1 | 4.131502 | -0.23194 | 0.999532 | | 4.479232 | -0.26559 | 0.854814 | | 3.677542 | -0.00931 | 0.996891 |
| LENG1 | 14.52948 | 0.05148 | 0.999532 | | 16.70841 | 0.133794 | 0.854814 | | 14.49082 | 0.102152 | 0.897958 |
| CTSG | 2.558973 | 0.66336 | 0.999532 | | 2.390628 | 0.310372 | 0.854814 | | 2.646977 | -0.33542 | 0.827119 |
| AGTPBP1 | 2.758101 | -0.14068 | 0.999532 | | 3.657092 | 0.2553 | 0.854913 | | 2.984438 | 0.41602 | 0.749206 |
| GUCY1A2 | 3.900756 | 0.353745 | 0.999532 | | 3.969588 | 0.242929 | 0.854913 | | 4.021465 | -0.0974 | 0.952656 |
| PNMA6A | 2.659693 | 0.007113 | 0.999532 | | 2.596712 | -0.31182 | 0.854913 | | 2.294409 | -0.30335 | 0.865065 |
| PKN3 | 2.78382 | 0.10316 | 0.999532 | | 3.391923 | 0.282268 | 0.854913 | | 2.952382 | 0.197961 | 0.906811 |
| ARRDC5 | 2.24317 | 0.153339 | 0.999532 | | 2.23645 | -0.32337 | 0.854913 | | 1.972229 | -0.45524 | 0.779082 |
| SGMS1 | 4.617384 | 0.134865 | 0.999532 | | 5.044845 | 0.212944 | 0.854985 | | 4.655715 | 0.088906 | 0.955834 |
| SOGA1 | 3.501307 | 0.495701 | 0.999532 | | 3.332048 | 0.256031 | 0.855 | | 3.60799 | -0.23164 | 0.87591 |
| UBQLN4 | 3.831924 | -0.38516 | 0.999532 | | 4.744085 | -0.27828 | 0.855 | | 3.544269 | 0.128536 | 0.946512 |
| DSG1 | 2.334584 | 0.052828 | 0.999532 | | 2.173556 | -0.32025 | 0.855 | | 2.022168 | -0.366 | 0.836935 |
| C11orf24 | 2.03499 | 0.093671 | 0.999532 | | 2.456727 | 0.293871 | 0.855052 | | 2.156747 | 0.218826 | 0.901033 |
| PARP10 | 4.887308 | 0.479374 | 0.999532 | | 4.919941 | 0.219819 | 0.855052 | | 5.010538 | -0.24174 | 0.821463 |
| LRRK1 | 2.091288 | -0.22602 | 0.999532 | | 2.33102 | -0.32281 | 0.855052 | | 1.779113 | -0.07716 | 0.977473 |
| KPNA6 | 3.414438 | -0.60059 | 0.999532 | | 4.70079 | 0.223464 | 0.855069 | | 3.468887 | 0.833738 | 0.394452 |
| C13orf46 | 3.237768 | 0.367533 | 0.999532 | | 2.761143 | -0.30381 | 0.855069 | | 2.895594 | -0.66554 | 0.632923 |
| VPS37B | 3.941987 | -0.26998 | 0.999532 | | 4.461715 | -0.236 | 0.855069 | | 3.532592 | 0.054687 | 0.978229 |
| GGTLC1 | 6.531909 | 0.097714 | 0.999532 | | 6.655544 | -0.22894 | 0.855069 | | 5.885133 | -0.30536 | 0.784955 |
| OXSM | 1.720323 | -0.05864 | 0.999532 | | 2.393923 | 0.333553 | 0.855069 | | 2.006091 | 0.424995 | 0.816618 |
| ASCL1 | 2.662825 | -0.28706 | 0.999532 | | 3.117764 | -0.29173 | 0.855069 | | 2.472758 | 0.013986 | 0.996891 |
| CYB5D2 | 2.716445 | -0.18277 | 0.999532 | | 3.503079 | 0.263064 | 0.855268 | | 2.79462 | 0.462177 | 0.722703 |
| ITGAX | 4.588119 | -0.55767 | 0.999532 | | 5.552246 | -0.21285 | 0.855268 | | 4.014944 | 0.352055 | 0.765852 |
| ASNSD1 | 2.708601 | -0.29831 | 0.999532 | | 3.05597 | -0.27188 | 0.855569 | | 2.387396 | 0.045062 | 0.986552 |
| RAPGEF | 4.886145 | 0.008525 | 0.999532 | | 6.117132 | 0.218258 | 0.855569 | | 5.060398 | 0.222523 | 0.849021 |
| GTDC1 | 2.466742 | 0.030255 | 0.999532 | | 2.876785 | 0.277165 | 0.855569 | | 2.550174 | 0.26656 | 0.869224 |
| GRK2 | 5.834986 | -0.30179 | 0.999532 | | 7.930022 | 0.193956 | 0.855569 | | 6.087804 | 0.527613 | 0.566812 |
| OR2T8 | 2.65165 | 0.400922 | 0.999532 | | 2.358876 | -0.3179 | 0.8556 | | 2.454653 | -0.70361 | 0.600779 |
| FABP4 | 5.790073 | -0.77366 | 0.999532 | | 8.61745 | 0.182968 | 0.8556 | | 6.124721 | 0.970465 | 0.178337 |
| SLC45A4 | 2.211997 | 0.138728 | 0.999532 | | 2.759236 | 0.312113 | 0.8556 | | 2.412451 | 0.198109 | 0.915991 |
| FRMD4A | 3.162829 | -0.45548 | 0.999532 | | 4.393323 | 0.253846 | 0.8556 | | 3.169533 | 0.735115 | 0.515762 |
| CNTRL | 3.192923 | 0.0923 | 0.999532 | | 3.14829 | -0.27161 | 0.8556 | | 2.699244 | -0.34928 | 0.815713 |
| CARD11 | 2.493773 | -0.39298 | 0.999532 | | 3.342603 | 0.274473 | 0.8556 | | 2.703626 | 0.682827 | 0.614824 |
| KLK4 | 2.258761 | 0.027886 | 0.999532 | | 2.284107 | -0.31487 | 0.8556 | | 2.025384 | -0.32528 | 0.858705 |
| STK17B | 9.884754 | -0.31671 | 0.999532 | | 13.30575 | 0.21698 | 0.8556 | | 10.49368 | 0.566689 | 0.465298 |
| PRC1 | 2.576849 | -0.25265 | 0.999532 | | 2.909128 | -0.28787 | 0.855636 | | 2.277172 | -0.01754 | 0.996681 |
| NDUFA9 | 5.275793 | -0.08988 | 0.999532 | | 5.673412 | -0.20698 | 0.855675 | | 4.634826 | -0.09802 | 0.946512 |
| NSMCE2 | 4.847696 | -0.22394 | 0.999532 | | 6.084922 | 0.19827 | 0.855675 | | 5.01062 | 0.438647 | 0.66232 |
| NID2 | 5.151992 | -0.20825 | 0.999532 | | 6.726259 | 0.268587 | 0.855903 | | 5.392613 | 0.497137 | 0.661666 |
| CSF1 | 5.236232 | -0.40651 | 0.999532 | | 7.349594 | 0.220253 | 0.855903 | | 5.683033 | 0.636478 | 0.426998 |
| PALM | 2.359811 | -0.01707 | 0.999532 | | 2.896199 | 0.27121 | 0.855942 | | 2.355301 | 0.304967 | 0.844187 |
| ZNF880 | 3.413294 | 0.493512 | 0.999532 | | 3.708681 | 0.259255 | 0.855942 | | 3.61836 | -0.22215 | 0.872493 |
| HMGCLL | 9.781807 | -0.00381 | 0.999532 | | 10.12343 | -0.2426 | 0.855942 | | 8.857653 | -0.2204 | 0.879947 |
| EI24 | 5.520444 | -0.51346 | 0.999532 | | 6.843938 | -0.19271 | 0.855942 | | 4.858664 | 0.33913 | 0.741408 |
| UBLCP1 | 5.261061 | 0.144555 | 0.999532 | | 6.220083 | 0.225225 | 0.855942 | | 5.601498 | 0.092451 | 0.947877 |
| COPZ1 | 4.619336 | 0.098005 | 0.999532 | | 5.180707 | 0.240993 | 0.855942 | | 4.52475 | 0.160963 | 0.911296 |
| ADAMTS | 3.015748 | 0.533776 | 0.999532 | | 2.704844 | -0.31194 | 0.855942 | | 2.775185 | -0.79785 | 0.546319 |
| ZNF654 | 2.677286 | -0.27765 | 0.999532 | | 2.961648 | -0.28412 | 0.855942 | | 2.299872 | 0.012037 | 0.996891 |
| PRELP | 3.417925 | 0.328308 | 0.999532 | | 2.957517 | -0.28675 | 0.855942 | | 2.991689 | -0.59837 | 0.634624 |
| LRTM1 | 2.172061 | 0.628637 | 0.999532 | | 1.769201 | -0.35345 | 0.856232 | | 2.00262 | -0.96677 | 0.465298 |
| LCE3B | 2.675263 | -0.39509 | 0.999532 | | 3.057426 | -0.29969 | 0.85646 | | 2.391275 | 0.094588 | 0.969501 |
| NMRAL1 | 4.628189 | -0.18741 | 0.999532 | | 4.981462 | -0.22246 | 0.856496 | | 4.05498 | -0.02316 | 0.992286 |
| DUSP11 | 5.385248 | -0.1094 | 0.999532 | | 6.444644 | 0.196696 | 0.856721 | | 5.435411 | 0.325073 | 0.743745 |
| BOP1 | 2.120838 | -0.27147 | 0.999532 | | 2.285679 | -0.31487 | 0.856721 | | 1.865344 | -0.02907 | 0.993618 |
| JPT1 | 6.119409 | -0.60778 | 0.999532 | | 7.262435 | -0.20414 | 0.857157 | | 5.234639 | 0.397554 | 0.718643 |
| PLPP7 | 3.055433 | 0.660139 | 0.999532 | | 2.887547 | 0.298953 | 0.857157 | | 3.240824 | -0.35081 | 0.801572 |
| WSB2 | 3.394054 | 0.359553 | 0.999532 | | 3.721713 | 0.247461 | 0.857157 | | 3.542335 | -0.09381 | 0.955834 |
| SUCLG2 | 6.817454 | -0.11073 | 0.999532 | | 8.032272 | 0.180531 | 0.857157 | | 6.778497 | 0.301081 | 0.750371 |
| TMUB1 | 2.364239 | 0.243925 | 0.999532 | | 2.366617 | -0.33872 | 0.857157 | | 2.095237 | -0.54923 | 0.733499 |
| ECSCR | 4.15958 | -0.21435 | 0.999532 | | 5.201191 | 0.221755 | 0.857157 | | 4.204198 | 0.44242 | 0.702463 |
| PPP4R3A | 5.704347 | -0.02285 | 0.999532 | | 7.23642 | 0.191553 | 0.857157 | | 5.902923 | 0.23871 | 0.823066 |
| TNFAIP2 | 8.034333 | -0.30191 | 0.999532 | | 8.953988 | -0.18218 | 0.857157 | | 7.016106 | 0.133943 | 0.905746 |
| AOX1 | 3.379038 | -0.18716 | 0.999532 | | 3.773126 | -0.26958 | 0.857157 | | 2.985162 | -0.06501 | 0.976258 |
| KIFC2 | 5.210265 | 0.251541 | 0.999532 | | 5.605763 | 0.227812 | 0.857157 | | 5.128933 | -0.00713 | 0.99716 |
| HSD17B8 | 4.346216 | 0.038874 | 0.999532 | | 4.586721 | -0.23742 | 0.857157 | | 3.731421 | -0.25631 | 0.846786 |
| ZNF292 | 3.314605 | -0.27217 | 0.999532 | | 4.380121 | 0.236427 | 0.857157 | | 3.328098 | 0.529881 | 0.647218 |
| MRPL35 | 6.899381 | 0.236062 | 0.999532 | | 7.180665 | -0.19336 | 0.857157 | | 6.392646 | -0.38317 | 0.694961 |
| PPARA | 3.307652 | -0.19951 | 0.999532 | | 3.621992 | -0.25107 | 0.857236 | | 2.927377 | -0.03464 | 0.989646 |
| FAM156A | 4.429275 | 0.040815 | 0.999532 | | 5.341273 | 0.241636 | 0.857236 | | 4.555847 | 0.227903 | 0.865065 |
| SLC22A1 | 2.151247 | 0.398572 | 0.999532 | | 2.267549 | 0.313804 | 0.857236 | | 2.257165 | -0.06687 | 0.978282 |
| PYCR2 | 6.726535 | -0.03015 | 0.999532 | | 7.271764 | -0.19049 | 0.857236 | | 5.966107 | -0.14151 | 0.900271 |
| SNRNP27 | 3.306861 | 0.17577 | 0.999532 | | 3.790887 | 0.249399 | 0.857236 | | 3.535376 | 0.091325 | 0.96129 |
| CLRN1 | 2.720079 | 0.098604 | 0.999532 | | 2.640036 | -0.29137 | 0.857236 | | 2.481579 | -0.36608 | 0.816775 |
| PARL | 3.701194 | -0.04823 | 0.999532 | | 3.769478 | -0.24445 | 0.857302 | | 3.241513 | -0.17988 | 0.904871 |
| MECOM | 7.382766 | -0.19464 | 0.999532 | | 9.084722 | 0.196267 | 0.857307 | | 7.509887 | 0.406198 | 0.650636 |
| FAM136A | 2.595057 | 0.673726 | 0.999532 | | 2.509646 | 0.313231 | 0.857307 | | 2.765266 | -0.34241 | 0.832386 |
| BAHD1 | 3.614815 | -0.13199 | 0.999532 | | 3.974793 | -0.23923 | 0.857307 | | 3.01096 | -0.09103 | 0.959377 |
| PLA2G2E | 2.548088 | 0.16007 | 0.999532 | | 2.437312 | -0.32474 | 0.857307 | | 2.255936 | -0.4681 | 0.769793 |
| SNX5 | 8.50159 | -0.14014 | 0.999532 | | 10.53743 | 0.188466 | 0.857307 | | 8.411848 | 0.364487 | 0.667066 |
| TPRA1 | 3.758587 | -0.30859 | 0.999532 | | 4.311054 | -0.23127 | 0.857307 | | 3.400546 | 0.09791 | 0.955834 |
| CRBN | 3.310691 | 0.202894 | 0.999532 | | 3.812004 | 0.248257 | 0.857307 | | 3.505155 | 0.063142 | 0.97386 |
| TBC1D1 | 11.00713 | -0.30227 | 0.999532 | | 14.40307 | 0.157724 | 0.857307 | | 11.13025 | 0.481289 | 0.457896 |
| HRC | 2.744566 | -0.15016 | 0.999532 | | 3.003325 | -0.27511 | 0.857307 | | 2.549083 | -0.10627 | 0.960773 |
| CCDC40 | 2.607994 | 0.161668 | 0.999532 | | 2.590653 | -0.2979 | 0.857319 | | 2.3444 | -0.43881 | 0.771685 |
| TSEN2 | 2.918634 | 0.235469 | 0.999532 | | 2.705002 | -0.29168 | 0.857319 | | 2.66722 | -0.50616 | 0.718643 |
| NRIP1 | 2.854874 | 0.192957 | 0.999532 | | 3.327457 | 0.255109 | 0.857454 | | 3.004423 | 0.080337 | 0.966632 |
| RAD52 | 3.946385 | 0.266838 | 0.999532 | | 4.499839 | 0.232555 | 0.857454 | | 4.224687 | -0.00768 | 0.997084 |
| HELLS | 2.100798 | 0.400353 | 0.999532 | | 2.155714 | 0.317313 | 0.857533 | | 2.213575 | -0.06641 | 0.978282 |
| G6PC1 | 2.242771 | 0.761762 | 0.999532 | | 2.062644 | 0.318939 | 0.857815 | | 2.306183 | -0.44614 | 0.758124 |
| ERP29 | 7.697333 | 0.066671 | 0.999532 | | 8.873368 | 0.186761 | 0.857815 | | 7.905739 | 0.132851 | 0.898682 |
| PLK2 | 2.760472 | -0.39654 | 0.999532 | | 3.793228 | 0.247331 | 0.857897 | | 2.980302 | 0.665691 | 0.560424 |
| NIFK | 4.128085 | 0.021387 | 0.999532 | | 4.305257 | -0.24374 | 0.858 | | 3.620882 | -0.24179 | 0.865065 |
| PFDN6 | 3.767579 | 0.011536 | 0.999532 | | 4.53912 | 0.251851 | 0.858088 | | 3.997119 | 0.247674 | 0.866719 |
| REPIN1 | 5.977418 | 0.186984 | 0.999532 | | 6.783411 | 0.198928 | 0.858088 | | 6.18537 | 0.029225 | 0.987274 |
| CPNE3 | 3.266187 | 0.395349 | 0.999532 | | 3.506421 | 0.248734 | 0.858088 | | 3.305166 | -0.13187 | 0.940114 |
| MRPL57 | 5.469575 | 0.381096 | 0.999532 | | 5.757961 | 0.238806 | 0.858088 | | 5.606082 | -0.12502 | 0.927668 |
| CCBE1 | 4.262441 | 0.259343 | 0.999532 | | 4.801541 | 0.275117 | 0.858088 | | 4.27187 | 0.060616 | 0.974421 |
| PRPF8 | 4.874508 | -0.04921 | 0.999532 | | 5.915147 | 0.210405 | 0.858088 | | 5.057146 | 0.278157 | 0.796702 |
| ZNF318 | 3.926387 | -0.15935 | 0.999532 | | 4.968481 | 0.235782 | 0.858088 | | 4.218186 | 0.419194 | 0.720106 |
| SMS | 3.826994 | 0.289367 | 0.999532 | | 4.436624 | 0.228954 | 0.858088 | | 3.882349 | -0.03997 | 0.98514 |
| DBF4B | 2.287816 | 0.538464 | 0.999532 | | 2.185957 | 0.314607 | 0.858088 | | 2.23388 | -0.20712 | 0.907163 |
| RTL10 | 2.565129 | -0.0391 | 0.999532 | | 2.405384 | -0.31286 | 0.858088 | | 2.251387 | -0.25415 | 0.892869 |
| FAM167B | 2.563873 | 0.163263 | 0.999532 | | 3.005189 | 0.286441 | 0.858088 | | 2.617268 | 0.147238 | 0.938429 |
| PLEKHG | 2.879373 | -0.67891 | 0.999532 | | 3.606268 | -0.24419 | 0.858463 | | 2.420852 | 0.449541 | 0.74497 |
| PAK4 | 2.617202 | 0.248631 | 0.999532 | | 2.944009 | 0.271356 | 0.858497 | | 2.611725 | 0.041171 | 0.987491 |
| ELF1 | 4.186408 | 0.113728 | 0.999532 | | 4.716954 | 0.219772 | 0.858497 | | 4.107664 | 0.124924 | 0.929529 |
| MT1H | 8.410626 | -0.10541 | 0.999532 | | 11.32452 | 0.319033 | 0.858689 | | 9.013165 | 0.49208 | 0.764304 |
| SHLD1 | 3.084961 | -0.13843 | 0.999532 | | 3.854677 | 0.267037 | 0.858689 | | 3.101706 | 0.426844 | 0.746581 |
| IKZF1 | 2.000597 | -0.29726 | 0.999532 | | 2.172996 | -0.32679 | 0.858689 | | 1.64567 | -0.01149 | 0.996893 |
| MAP3K5 | 2.784424 | -0.15565 | 0.999532 | | 3.518847 | 0.261879 | 0.858689 | | 2.789223 | 0.433386 | 0.753597 |
| APEX2 | 2.529376 | 0.714086 | 0.999532 | | 1.862902 | -0.33736 | 0.858689 | | 2.243839 | -1.03254 | 0.389965 |
| PLIN1 | 2.38696 | -0.45021 | 0.999532 | | 2.966903 | -0.2803 | 0.858689 | | 2.146075 | 0.196018 | 0.918421 |
| NFAT5 | 6.153137 | -0.05408 | 0.999532 | | 7.185602 | 0.184666 | 0.858689 | | 6.23576 | 0.262207 | 0.791466 |
| CACNG5 | 2.361063 | 0.499609 | 0.999532 | | 1.823482 | -0.34517 | 0.858689 | | 2.094618 | -0.82759 | 0.563461 |
| TOM1L2 | 3.482142 | 0.200671 | 0.999532 | | 3.242411 | -0.26675 | 0.858689 | | 3.058432 | -0.44583 | 0.718468 |
| COX5B | 9.975309 | -0.08974 | 0.999532 | | 12.46783 | 0.149519 | 0.858689 | | 9.928564 | 0.26024 | 0.751444 |
| MAGEB1 | 1.947219 | -0.20171 | 0.999532 | | 2.109841 | -0.33436 | 0.858689 | | 1.621122 | -0.11436 | 0.965649 |
| GPR34 | 2.412968 | -0.53235 | 0.999532 | | 2.936547 | -0.28317 | 0.858689 | | 2.19184 | 0.262444 | 0.889911 |
| GART | 3.602689 | 0.300237 | 0.999532 | | 3.910217 | 0.244463 | 0.858689 | | 3.750471 | -0.0357 | 0.987274 |
| SORL1 | 7.388353 | 0.304968 | 0.999532 | | 7.678073 | 0.182467 | 0.858689 | | 7.382617 | -0.11329 | 0.923864 |
| ZNF684 | 5.565071 | -0.15272 | 0.999532 | | 6.705633 | 0.195174 | 0.858689 | | 5.635066 | 0.355462 | 0.720106 |
| ARHGEF | 7.512889 | -0.16217 | 0.999532 | | 9.530685 | 0.166922 | 0.858689 | | 7.691578 | 0.349723 | 0.702463 |
| SLC30A5 | 2.480172 | -0.04842 | 0.999532 | | 2.863536 | 0.272176 | 0.858689 | | 2.513813 | 0.346658 | 0.816618 |
| MAFG | 6.301844 | -0.20024 | 0.999532 | | 6.978735 | -0.19624 | 0.858689 | | 5.624895 | 0.021352 | 0.992286 |
| E2F4 | 4.212355 | -0.22304 | 0.999532 | | 5.620475 | 0.211848 | 0.858689 | | 4.380081 | 0.452438 | 0.661666 |
| PAQR5 | 3.622391 | -0.0399 | 0.999532 | | 3.929782 | -0.25486 | 0.858689 | | 3.106055 | -0.20441 | 0.896374 |
| [illegible] | [illegible] | [illegible] | [illegible] | | [illegible] | [illegible] | [illegible] | | [illegible] | [illegible] | [illegible] |
| RFT1 | 2.269872 | 0.098201 | 0.999532 | | 2.133062 | -0.30788 | 0.858689 | | 2.001067 | -0.39183 | 0.814884 |
| ZSCAN20 | 3.059768 | 0.803235 | 0.999532 | | 2.628964 | 0.287087 | 0.858689 | | 3.173037 | -0.50393 | 0.692534 |
| CR1 | 3.700007 | 0.134588 | 0.999532 | | 3.662749 | -0.25776 | 0.858689 | | 3.136367 | -0.36855 | 0.783395 |
| C1orf131 | 2.276709 | 0.516052 | 0.999532 | | 2.216002 | 0.314079 | 0.858689 | | 2.291669 | -0.1861 | 0.920472 |
| ZMIZ1 | 10.94435 | -0.09701 | 0.999532 | | 13.2365 | 0.179905 | 0.858689 | | 10.85925 | 0.304908 | 0.738861 |
| TLR8 | 2.363485 | 0.106272 | 0.999532 | | 2.284051 | -0.3144 | 0.858689 | | 2.104109 | -0.40924 | 0.809498 |
| OR52A5 | 1.914141 | -0.08396 | 0.999532 | | 2.109982 | -0.3226 | 0.858689 | | 1.748772 | -0.21444 | 0.917499 |
| NDUFA12 | 5.949124 | -0.28591 | 0.999532 | | 7.714136 | 0.188568 | 0.858689 | | 6.059862 | 0.492855 | 0.565022 |
| CCDC18 | 3.032833 | -0.57486 | 0.999532 | | 3.746891 | -0.25808 | 0.858689 | | 2.643172 | 0.336763 | 0.827429 |
| DAP3 | 6.596473 | -0.02673 | 0.999532 | | 8.256662 | 0.174318 | 0.858689 | | 6.855996 | 0.22164 | 0.816618 |
| RAPGEF | 5.886361 | -0.22942 | 0.999532 | | 7.311007 | 0.191333 | 0.858689 | | 6.193814 | 0.46609 | 0.595562 |
| UMAD1 | 2.692669 | 0.045155 | 0.999532 | | 3.287163 | 0.263508 | 0.858689 | | 2.816177 | 0.236364 | 0.88082 |
| GLIPR1 | 2.811141 | 0.018064 | 0.999532 | | 3.002965 | -0.2719 | 0.858689 | | 2.566431 | -0.27099 | 0.863808 |
| ROR2 | 2.332081 | -0.32044 | 0.999532 | | 2.548387 | -0.29948 | 0.858689 | | 2.045748 | 0.037328 | 0.991159 |
| RGCC | 2.150131 | -0.48525 | 0.999532 | | 2.296114 | -0.31475 | 0.858689 | | 1.731913 | 0.186942 | 0.934572 |
| H6PD | 5.093522 | 0.095959 | 0.999532 | | 5.759452 | 0.213757 | 0.858689 | | 5.161391 | 0.145666 | 0.906811 |
| MKRN2 | 2.761502 | -0.03867 | 0.999532 | | 2.907206 | -0.27411 | 0.858689 | | 2.390806 | -0.21657 | 0.898369 |
| PTPN12 | 12.39865 | 0.000792 | 0.999532 | | 14.40022 | 0.141965 | 0.858689 | | 12.38717 | 0.158087 | 0.859898 |
| ENY2 | 4.22385 | 0.338041 | 0.999532 | | 4.805344 | 0.223622 | 0.858689 | | 4.505715 | -0.08352 | 0.960773 |
| WAS | 4.617981 | 0.138319 | 0.999532 | | 4.382894 | -0.22463 | 0.858689 | | 4.076165 | -0.34809 | 0.753585 |
| OPRD1 | 2.293335 | 0.481511 | 0.999532 | | 1.897565 | -0.34331 | 0.858689 | | 2.057017 | -0.80716 | 0.573023 |
| TADA2B | 3.600082 | -0.10097 | 0.999532 | | 4.789542 | 0.234625 | 0.858689 | | 3.784654 | 0.355715 | 0.765852 |
| RBM19 | 3.525252 | 0.122247 | 0.999532 | | 3.633046 | -0.25236 | 0.858689 | | 3.188519 | -0.35365 | 0.791576 |
| NUP98 | 4.340218 | -0.33741 | 0.999532 | | 5.387236 | 0.207629 | 0.858689 | | 4.208006 | 0.5444 | 0.600081 |
| EN1 | 2.358204 | -0.16558 | 0.999532 | | 2.558107 | -0.31937 | 0.858689 | | 2.161021 | -0.13764 | 0.954304 |
| CDK19 | 3.051497 | -0.24891 | 0.999532 | | 3.491525 | -0.2509 | 0.858689 | | 2.616677 | 0.0136 | 0.996891 |
| DIP2B | 2.451064 | -0.01574 | 0.999532 | | 3.002833 | 0.26462 | 0.858694 | | 2.672704 | 0.303232 | 0.839961 |
| FTH1 | 214.674 | -0.21258 | 0.999532 | | 272.535 | 0.14974 | 0.858694 | | 217.2323 | 0.383484 | 0.499025 |
| NBPF12 | 40.01009 | 0.152858 | 0.999532 | | 43.74692 | 0.092941 | 0.858694 | | 38.77975 | -0.04351 | 0.946808 |
| MAML3 | 4.792257 | 0.009587 | 0.999532 | | 5.621398 | 0.228842 | 0.858814 | | 4.97472 | 0.246771 | 0.857457 |
| OR5F1 | 2.088323 | 0.364541 | 0.999532 | | 1.895978 | -0.34903 | 0.858814 | | 1.929971 | -0.69511 | 0.660097 |
| PUDP | 3.318288 | -0.17614 | 0.999532 | | 3.602908 | -0.24767 | 0.858814 | | 2.879671 | -0.05476 | 0.978931 |
| MIGA1 | 3.599237 | -0.09385 | 0.999532 | | 4.686912 | 0.229511 | 0.858842 | | 3.961181 | 0.337302 | 0.78258 |
| HEXIM2 | 3.193635 | 0.21572 | 0.999532 | | 3.895471 | 0.267957 | 0.858842 | | 3.488594 | 0.076409 | 0.971732 |
| WNT3 | 2.080399 | -0.16736 | 0.999532 | | 2.651346 | 0.293986 | 0.858851 | | 2.239679 | 0.478783 | 0.752562 |
| TMEM13 | 2.249964 | 0.161628 | 0.999532 | | 2.192621 | -0.31074 | 0.858935 | | 2.050808 | -0.45691 | 0.765012 |
| ATP6AP1 | 2.94679 | 0.539403 | 0.999532 | | 2.507264 | -0.29207 | 0.858944 | | 2.665127 | -0.81223 | 0.484303 |
| SLC9A9 | 3.444727 | -0.93347 | 0.999532 | | 4.765484 | -0.23035 | 0.858944 | | 3.116021 | 0.722952 | 0.515762 |
| LINGO4 | 2.263009 | -0.71522 | 0.999532 | | 2.876926 | -0.28474 | 0.858944 | | 1.975395 | 0.44974 | 0.791576 |
| NBPF19 | 31.54085 | 0.095908 | 0.999532 | | 32.82793 | -0.10041 | 0.858944 | | 29.06371 | -0.17857 | 0.732486 |
| GRPEL1 | 2.399171 | 0.434188 | 0.999532 | | 2.056582 | -0.33923 | 0.858944 | | 2.074782 | -0.75287 | 0.60629 |
| DPH7 | 3.609557 | 0.314436 | 0.999532 | | 3.848668 | 0.243695 | 0.858944 | | 3.833546 | -0.05354 | 0.977635 |
| DNAH14 | 2.306683 | 0.794613 | 0.999532 | | 2.19713 | 0.306847 | 0.859056 | | 2.376161 | -0.47036 | 0.732864 |
| DNAI4 | 2.606295 | -0.01886 | 0.999532 | | 2.641845 | -0.28644 | 0.859155 | | 2.264929 | -0.25039 | 0.881689 |
| NLE1 | 2.096047 | 0.404716 | 0.999532 | | 1.979854 | -0.34236 | 0.859155 | | 1.947447 | -0.72756 | 0.632847 |
| PTPN11 | 8.888238 | -0.0651 | 0.999532 | | 10.58116 | 0.162702 | 0.859155 | | 9.189872 | 0.239014 | 0.791576 |
| RGS18 | 4.509229 | -0.41398 | 0.999532 | | 5.260477 | -0.22102 | 0.859155 | | 3.758588 | 0.211325 | 0.880273 |
| SELENOI | 3.352724 | 0.317954 | 0.999532 | | 3.453548 | 0.25725 | 0.859155 | | 3.332489 | -0.04959 | 0.982124 |
| HILPDA | 3.009895 | 0.211197 | 0.999532 | | 2.826889 | -0.2809 | 0.859227 | | 2.700986 | -0.47913 | 0.722703 |
| CX3CL1 | 6.926718 | -0.26958 | 0.999532 | | 7.582973 | -0.20417 | 0.859227 | | 6.07568 | 0.078597 | 0.957645 |
| ARFGAP | 3.636338 | 0.294029 | 0.999532 | | 4.387123 | 0.237362 | 0.859227 | | 3.792184 | -0.03497 | 0.98811 |
| DUSP23 | 1.878574 | -0.32598 | 0.999532 | | 2.073189 | -0.32032 | 0.859227 | | 1.54404 | 0.02093 | 0.996681 |
| IL26 | 2.286133 | -0.16828 | 0.999532 | | 2.484845 | -0.28914 | 0.859227 | | 2.04445 | -0.09942 | 0.966632 |
| DERA | 3.40992 | -0.43852 | 0.999532 | | 3.825462 | -0.24371 | 0.859289 | | 2.864493 | 0.200943 | 0.901033 |
| TTC7A | 3.299387 | 0.586936 | 0.999532 | | 2.882213 | -0.2843 | 0.859289 | | 2.898032 | -0.84632 | 0.444687 |
| NNAT | 2.25155 | -0.19756 | 0.999532 | | 2.431568 | -0.30682 | 0.859511 | | 1.967448 | -0.09001 | 0.972221 |
| LYN | 3.673789 | -0.13327 | 0.999532 | | 4.730596 | 0.22636 | 0.859654 | | 3.815728 | 0.384155 | 0.753585 |
| MVD | 3.97584 | -0.02243 | 0.999532 | | 4.778602 | 0.225777 | 0.859654 | | 4.029872 | 0.267415 | 0.836453 |
| CACNA1 | 2.861381 | -0.28534 | 0.999532 | | 3.219564 | -0.27361 | 0.859654 | | 2.606185 | 0.028199 | 0.992586 |
| HOXC10 | 4.730672 | -0.33082 | 0.999532 | | 6.500393 | 0.192701 | 0.859654 | | 5.161208 | 0.543874 | 0.52568 |
| SDR39U1 | 4.650255 | 0.05118 | 0.999532 | | 4.851965 | -0.21887 | 0.859806 | | 4.155856 | -0.25216 | 0.839373 |
| ITGA5 | 5.353912 | 0.408318 | 0.999532 | | 5.549041 | 0.210768 | 0.859806 | | 5.524388 | -0.17779 | 0.880375 |
| OSBPL7 | 2.077119 | 0.546161 | 0.999532 | | 2.138109 | 0.311315 | 0.859806 | | 2.179446 | -0.21735 | 0.900848 |
| CDPF1 | 2.84416 | 0.114441 | 0.999532 | | 2.905186 | -0.27047 | 0.859806 | | 2.658281 | -0.367 | 0.79686 |
| POGLUT | 6.365772 | -0.51834 | 0.999532 | | 8.195013 | -0.17725 | 0.859806 | | 5.671823 | 0.358537 | 0.702463 |
| ZKSCAN | 2.4408 | -0.03933 | 0.999532 | | 2.386067 | -0.29819 | 0.859806 | | 2.06444 | -0.24432 | 0.894963 |
| ZNF529 | 6.60042 | 0.317483 | 0.999532 | | 6.79126 | 0.188003 | 0.859806 | | 6.533525 | -0.11785 | 0.920716 |
| MED11 | 2.439677 | -0.06535 | 0.999532 | | 2.919855 | 0.26948 | 0.859806 | | 2.607726 | 0.351089 | 0.818002 |
| TRIL | 2.477052 | -0.51795 | 0.999532 | | 3.150973 | -0.26888 | 0.859806 | | 2.237107 | 0.273925 | 0.877614 |
| CILP | 2.424954 | 0.040049 | 0.999532 | | 2.580327 | -0.31555 | 0.859806 | | 2.168784 | -0.33594 | 0.849532 |
| MPHOSPI | 2.794473 | 0.037176 | 0.999532 | | 3.475064 | 0.270057 | 0.859867 | | 2.994926 | 0.256105 | 0.87591 |
| IGSF3 | 2.938253 | 0.481216 | 0.999532 | | 3.142619 | 0.263436 | 0.860064 | | 3.151987 | -0.19997 | 0.893259 |
| POLR1D | 5.387777 | 0.578649 | 0.999532 | | 5.026974 | 0.224842 | 0.860064 | | 5.46479 | -0.32634 | 0.747677 |
| NDRG3 | 2.71797 | -0.95184 | 0.999532 | | 4.232708 | 0.237913 | 0.860064 | | 2.968848 | 1.214616 | 0.203949 |
| POLR2C | 3.994348 | -0.12075 | 0.999532 | | 4.33334 | -0.243 | 0.860098 | | 3.465666 | -0.09991 | 0.956011 |
| MICB | 2.56378 | 0.118834 | 0.999532 | | 2.584382 | -0.3236 | 0.860098 | | 2.43112 | -0.43577 | 0.794914 |
| WTIP | 2.757263 | -0.32271 | 0.999532 | | 3.559112 | 0.253804 | 0.860098 | | 2.80069 | 0.600989 | 0.623462 |
| FOXE3 | 3.425649 | 0.101439 | 0.999532 | | 3.439858 | -0.26047 | 0.860098 | | 3.131805 | -0.34116 | 0.818373 |
| DICER1 | 5.155517 | 0.021905 | 0.999532 | | 5.773672 | 0.201092 | 0.860098 | | 5.088816 | 0.194639 | 0.87591 |
| PIGQ | 4.689826 | 0.008369 | 0.999532 | | 5.768522 | 0.20898 | 0.860098 | | 4.756423 | 0.211658 | 0.861119 |
| WFDC9 | 2.574646 | 0.246775 | 0.999532 | | 2.395453 | -0.30156 | 0.860098 | | 2.297011 | -0.53122 | 0.709825 |
| ZNF831 | 2.373202 | 0.497015 | 0.999532 | | 2.506775 | 0.311047 | 0.860098 | | 2.596692 | -0.17082 | 0.931488 |
| TOGARA | 3.240092 | 0.399146 | 0.999532 | | 2.933645 | -0.26781 | 0.860098 | | 2.907855 | -0.64783 | 0.566812 |
| C2orf78 | 2.226905 | 0.615914 | 0.999532 | | 1.815161 | -0.35496 | 0.860098 | | 1.963832 | -0.95233 | 0.49299 |
| ZP2 | 2.3224 | 0.458988 | 0.999532 | | 1.969971 | -0.33656 | 0.860098 | | 2.153931 | -0.78333 | 0.598934 |
| CENPV | 2.326057 | -0.40027 | 0.999532 | | 2.834219 | -0.2853 | 0.860246 | | 2.074317 | 0.137828 | 0.949067 |
| NSUN5 | 4.873871 | -0.12872 | 0.999532 | | 6.014627 | 0.201167 | 0.860246 | | 4.783381 | 0.346246 | 0.736481 |
| INPP4B | 5.064233 | 0.046375 | 0.999532 | | 6.068465 | 0.214763 | 0.860246 | | 5.245853 | 0.188457 | 0.892869 |
| ETFB | 3.286777 | -0.04074 | 0.999532 | | 4.057438 | 0.248325 | 0.860294 | | 3.363789 | 0.311677 | 0.806394 |
| CHST12 | 3.62276 | -0.07598 | 0.999532 | | 3.949983 | -0.24131 | 0.860348 | | 3.390059 | -0.1446 | 0.92697 |
| ZNF600 | 1.790548 | 0.302136 | 0.999532 | | 2.04247 | 0.323483 | 0.860348 | | 1.975977 | 0.040068 | 0.990309 |
| AP3M2 | 3.819406 | -0.38735 | 0.999532 | | 4.381049 | -0.24469 | 0.860348 | | 3.313343 | 0.14544 | 0.927375 |
| [illegible] | [illegible] | [illegible] | [illegible] | | [illegible] | [illegible] | [illegible] | | [illegible] | [illegible] | [illegible] |

| | | | | | | | | | | | |
|---|---|---|---|---|---|---|---|---|---|---|---|
| GTF3C5 | 3.097908 | 0.188868 | 0.999532 | | 2.989744 | -0.27516 | 0.860348 | | 2.698698 | -0.44603 | 0.739496 |
| CRYGC | 2.110549 | 0.081162 | 0.999532 | | 2.666341 | 0.303061 | 0.860348 | | 2.335215 | 0.242571 | 0.895647 |
| MSL1 | 8.97219 | 0.071927 | 0.999532 | | 10.62474 | 0.163227 | 0.860348 | | 8.951586 | 0.123962 | 0.904801 |
| PLCB1 | 2.541335 | 0.070003 | 0.999532 | | 2.522454 | -0.29515 | 0.860348 | | 2.208955 | -0.34784 | 0.824012 |
| IMPA2 | 2.63866 | 0.281107 | 0.999532 | | 3.015018 | 0.260052 | 0.860348 | | 2.764635 | -0.00263 | 0.999385 |
| MRPS7 | 3.934075 | 0.053321 | 0.999532 | | 4.53852 | 0.224369 | 0.860348 | | 3.87844 | 0.190942 | 0.888554 |
| RFX7 | 2.785613 | 0.204617 | 0.999532 | | 2.696937 | -0.27794 | 0.860348 | | 2.437058 | -0.46316 | 0.735233 |
| LMOD1 | 2.38043 | 0.183928 | 0.999532 | | 2.340327 | -0.29477 | 0.860348 | | 2.053876 | -0.46035 | 0.764083 |
| ASB7 | 1.92456 | -0.05701 | 0.999532 | | 2.373608 | 0.298673 | 0.860348 | | 2.039406 | 0.375052 | 0.823571 |
| SLC7A10 | 2.369023 | 0.269961 | 0.999532 | | 2.113189 | -0.32104 | 0.860348 | | 2.151651 | -0.5826 | 0.717138 |
| GATA4 | 2.341677 | 0.314819 | 0.999532 | | 2.141434 | -0.31988 | 0.860348 | | 2.147725 | -0.61347 | 0.679818 |
| B3GNT9 | 2.500305 | -0.11763 | 0.999532 | | 2.611746 | -0.28233 | 0.860348 | | 2.257848 | -0.15011 | 0.941197 |
| CLMP | 2.388571 | -0.52541 | 0.999532 | | 2.887063 | -0.28005 | 0.860348 | | 1.988565 | 0.263663 | 0.885078 |
| CPED1 | 2.908291 | 0.614206 | 0.999532 | | 3.034544 | 0.275051 | 0.860431 | | 3.128273 | -0.31892 | 0.816775 |
| NIM1K | 2.262006 | 0.269658 | 0.999532 | | 2.509923 | 0.295505 | 0.860431 | | 2.406928 | 0.042044 | 0.988853 |
| KDM6B | 5.335987 | -0.41796 | 0.999532 | | 6.278698 | -0.19948 | 0.860479 | | 4.415141 | 0.2417 | 0.837514 |
| SOX2 | 1.799352 | 0.355796 | 0.999532 | | 2.120755 | 0.32467 | 0.860634 | | 2.011759 | -0.0102 | 0.997153 |
| FOXC2 | 5.190631 | 0.018825 | 0.999532 | | 5.546748 | -0.19977 | 0.860901 | | 4.688288 | -0.19715 | 0.874653 |
| TTC5 | 1.840234 | 0.355178 | 0.999532 | | 1.942087 | 0.324552 | 0.861063 | | 1.90116 | -0.01458 | 0.996891 |
| C8orf48 | 2.534232 | -0.05366 | 0.999532 | | 2.672548 | -0.28012 | 0.861103 | | 2.251621 | -0.20698 | 0.908829 |
| PGAM5 | 3.438635 | 0.342882 | 0.999532 | | 3.120764 | -0.26927 | 0.861103 | | 2.987883 | -0.59496 | 0.632847 |
| SKAP1 | 2.602774 | 0.846752 | 0.999532 | | 1.851567 | -0.33011 | 0.861103 | | 2.363499 | -1.15982 | 0.282041 |
| TUBA3C | 2.32723 | -0.44858 | 0.999532 | | 2.935975 | -0.29705 | 0.861593 | | 2.168211 | 0.184345 | 0.934354 |
| CNPY2 | 6.841934 | 0.169682 | 0.999532 | | 6.524443 | -0.19456 | 0.861599 | | 6.120219 | -0.34541 | 0.720106 |
| MCM4 | 2.782616 | -0.07616 | 0.999532 | | 2.834918 | -0.2818 | 0.861599 | | 2.590846 | -0.19327 | 0.915533 |
| USP7 | 3.738107 | -0.23129 | 0.999532 | | 4.104624 | -0.23613 | 0.861599 | | 3.170648 | 0.01326 | 0.996681 |
| H3C2 | 3.559773 | -0.26351 | 0.999532 | | 3.761215 | -0.27111 | 0.861599 | | 2.919905 | -0.01181 | 0.996891 |
| REC114 | 1.967488 | -0.10422 | 0.999532 | | 2.085907 | -0.32278 | 0.861702 | | 1.847527 | -0.20121 | 0.92697 |
| LRP6 | 5.962547 | -0.2171 | 0.999532 | | 7.594249 | 0.178796 | 0.861721 | | 5.960908 | 0.405667 | 0.661666 |
| ZNF655 | 3.340397 | 0.268498 | 0.999532 | | 3.605151 | 0.238983 | 0.861721 | | 3.349821 | -0.01318 | 0.996681 |
| CNPY4 | 4.031454 | 0.003545 | 0.999532 | | 4.976476 | 0.217075 | 0.861726 | | 4.182972 | 0.231428 | 0.858767 |
| DNAJC17 | 6.241224 | 0.076018 | 0.999532 | | 6.128093 | -0.21294 | 0.861726 | | 5.578959 | -0.27775 | 0.811123 |
| PCDH1 | 4.548626 | -0.36742 | 0.999532 | | 6.46996 | 0.205491 | 0.86173 | | 4.638682 | 0.607009 | 0.532889 |
| AP5B1 | 2.413958 | 0.610524 | 0.999532 | | 2.395162 | 0.295265 | 0.861792 | | 2.45017 | -0.29636 | 0.851372 |
| SAMD1 | 3.940271 | -0.08126 | 0.999532 | | 4.072419 | -0.25505 | 0.861792 | | 3.46543 | -0.14846 | 0.927433 |
| DCAF4L2 | 2.214314 | -0.21083 | 0.999532 | | 2.355794 | -0.30748 | 0.861792 | | 1.956756 | -0.07804 | 0.976783 |
| BCKDHA | 7.14966 | -0.25171 | 0.999532 | | 8.253731 | -0.18147 | 0.861792 | | 6.531274 | 0.08746 | 0.947941 |
| AMY2B | 3.006368 | 0.129894 | 0.999532 | | 3.639838 | 0.252996 | 0.861792 | | 2.958344 | 0.120072 | 0.948382 |
| CXCL6 | 2.66747 | 0.352945 | 0.999532 | | 2.602504 | -0.31174 | 0.861792 | | 2.339133 | -0.65933 | 0.630705 |
| IBA57 | 2.811974 | 0.314818 | 0.999532 | | 2.853915 | 0.266082 | 0.861926 | | 2.865121 | -0.03316 | 0.991103 |
| CHST13 | 3.290276 | 0.361701 | 0.999532 | | 2.768961 | -0.27854 | 0.861926 | | 2.865664 | -0.6378 | 0.634624 |
| BAD | 6.092019 | -0.49585 | 0.999532 | | 7.341771 | -0.18659 | 0.861926 | | 5.409119 | 0.329718 | 0.746666 |
| KIF1B | 3.855184 | -0.23944 | 0.999532 | | 4.859395 | 0.215885 | 0.861926 | | 4.013463 | 0.471503 | 0.662576 |
| LHX5 | 2.223806 | -0.46834 | 0.999532 | | 2.669241 | -0.30048 | 0.861926 | | 1.987505 | 0.187331 | 0.932589 |
| CATSPER | 2.083037 | 0.245565 | 0.999532 | | 1.97261 | -0.33743 | 0.861926 | | 1.844652 | -0.56125 | 0.735469 |
| ITPKB | 4.927337 | -0.26576 | 0.999532 | | 6.91572 | 0.198183 | 0.861926 | | 5.264659 | 0.502684 | 0.630705 |
| SLC22A2 | 3.261886 | 0.081976 | 0.999532 | | 3.746796 | 0.255473 | 0.861926 | | 3.318572 | 0.190128 | 0.901033 |
| CRY2 | 4.566659 | 0.055305 | 0.999532 | | 5.241991 | 0.225978 | 0.861926 | | 4.567707 | 0.194106 | 0.88476 |
| ELK1 | 3.610759 | -0.33374 | 0.999532 | | 4.335668 | -0.24885 | 0.861926 | | 3.27637 | 0.105467 | 0.955834 |
| IRX2 | 1.874661 | -0.20982 | 0.999532 | | 2.081594 | -0.32621 | 0.861926 | | 1.713072 | -0.09985 | 0.97183 |
| TMEM74B | 3.244079 | 0.342323 | 0.999532 | | 2.790459 | -0.29414 | 0.861926 | | 2.88179 | -0.62619 | 0.659727 |
| SLC20A2 | 4.912659 | -0.05569 | 0.999532 | | 5.297541 | -0.21979 | 0.861926 | | 4.28608 | -0.14218 | 0.923834 |
| MMP24OS | 10.6149 | -0.06963 | 0.999532 | | 11.46475 | -0.15687 | 0.861926 | | 9.360735 | -0.06703 | 0.952656 |
| VAMP2 | 9.400553 | -0.25701 | 0.999532 | | 11.23029 | -0.1643 | 0.861926 | | 8.434376 | 0.102959 | 0.922679 |
| PIK3CG | 2.018498 | 0.011651 | 0.999532 | | 2.071516 | -0.31683 | 0.861926 | | 1.75382 | -0.30937 | 0.870663 |
| DHX32 | 2.319007 | -0.35466 | 0.999532 | | 2.467654 | -0.29236 | 0.861926 | | 1.951747 | 0.065089 | 0.979996 |
| ZNF236 | 2.585424 | 0.136563 | 0.999532 | | 2.650966 | -0.28546 | 0.861926 | | 2.1954 | -0.39576 | 0.799627 |
| CST6 | 2.90099 | -0.01004 | 0.999532 | | 2.838131 | -0.26874 | 0.861926 | | 2.509456 | -0.24409 | 0.883785 |
| RELCH | 4.455408 | -0.23915 | 0.999532 | | 5.297611 | -0.21389 | 0.861926 | | 4.015789 | 0.052516 | 0.977635 |
| SIRT7 | 2.855283 | -0.10283 | 0.999532 | | 3.75962 | 0.252731 | 0.861926 | | 3.124515 | 0.374066 | 0.783825 |
| AKAP6 | 2.877088 | 0.214816 | 0.999532 | | 2.788178 | -0.28127 | 0.861926 | | 2.638399 | -0.47197 | 0.750371 |
| CD36 | 2.388646 | -0.4812 | 0.999532 | | 2.862934 | -0.26599 | 0.861926 | | 2.135101 | 0.232381 | 0.898965 |
| SPATA32 | 1.943511 | -0.03797 | 0.999532 | | 1.963011 | -0.32722 | 0.861926 | | 1.734694 | -0.2722 | 0.894884 |
| ZNF214 | 3.567041 | 0.109118 | 0.999532 | | 3.378258 | -0.26418 | 0.861926 | | 3.006334 | -0.35567 | 0.801905 |
| EPHA4 | 5.06802 | 0.254045 | 0.999532 | | 5.318747 | 0.207095 | 0.861926 | | 5.244829 | -0.03306 | 0.987274 |
| APEX1 | 2.717568 | -0.24398 | 0.999532 | | 3.489716 | 0.240887 | 0.861926 | | 2.851103 | 0.498213 | 0.69872 |
| NAV2 | 2.341443 | -0.10661 | 0.999532 | | 2.33029 | -0.28806 | 0.861926 | | 1.982516 | -0.18269 | 0.927433 |
| MGAT1 | 6.832389 | -0.3145 | 0.999532 | | 8.858078 | 0.177767 | 0.861926 | | 6.705908 | 0.512797 | 0.5451 |
| ELL2 | 6.59328 | 0.112435 | 0.999532 | | 6.827537 | -0.19659 | 0.861926 | | 5.942267 | -0.28671 | 0.780904 |
| CCDC28A | 3.710927 | 0.166634 | 0.999532 | | 3.457762 | -0.25491 | 0.861926 | | 3.377463 | -0.40984 | 0.749206 |
| RRAGC | 2.194761 | 0.031772 | 0.999532 | | 2.599405 | 0.285012 | 0.86194 | | 2.320283 | 0.27126 | 0.879538 |
| ZNF493 | 3.502631 | -0.02314 | 0.999532 | | 3.76993 | -0.24997 | 0.86194 | | 3.223174 | -0.20734 | 0.892869 |
| GDPGP1 | 1.970168 | 0.877622 | 0.999532 | | 1.861686 | 0.349446 | 0.86194 | | 2.139305 | -0.51441 | 0.740445 |
| SLC25A3 | 2.948363 | 0.091767 | 0.999532 | | 2.941611 | -0.26626 | 0.86194 | | 2.66557 | -0.3385 | 0.818666 |
| DISC1 | 3.847754 | 0.279145 | 0.999532 | | 4.193985 | 0.244164 | 0.86194 | | 4.008077 | -0.013 | 0.996681 |
| ZC3H14 | 4.886407 | -0.17442 | 0.999532 | | 5.588489 | -0.21116 | 0.861965 | | 4.226237 | -0.02355 | 0.99186 |
| FREM1 | 2.552991 | 0.707158 | 0.999532 | | 2.511473 | 0.289865 | 0.862011 | | 2.77289 | -0.38729 | 0.796702 |
| TRIM61 | 3.014329 | 0.216737 | 0.999532 | | 2.906866 | -0.29453 | 0.862011 | | 2.754094 | -0.50249 | 0.739496 |
| MGRN1 | 4.893121 | -0.25436 | 0.999532 | | 6.686618 | 0.190947 | 0.862011 | | 5.089044 | 0.47146 | 0.639848 |
| NDST1 | 4.129549 | 0.150782 | 0.999532 | | 4.520394 | 0.215383 | 0.862011 | | 4.062916 | 0.077681 | 0.963492 |
| RASSF7 | 2.849053 | 0.120384 | 0.999532 | | 3.206598 | 0.256248 | 0.862011 | | 2.765325 | 0.152775 | 0.92697 |
| SPINK4 | 2.339037 | 0.128063 | 0.999532 | | 2.269331 | -0.31479 | 0.862011 | | 2.075277 | -0.42681 | 0.807942 |
| SLC2A1 | 2.05865 | 0.153611 | 0.999532 | | 1.929691 | -0.31872 | 0.862011 | | 1.799634 | -0.45497 | 0.791466 |
| IER5L | 2.670632 | 0.307281 | 0.999532 | | 2.91234 | 0.27786 | 0.862011 | | 2.874531 | -0.0173 | 0.996681 |
| NFIB | 3.773285 | -0.23917 | 0.999532 | | 5.011388 | 0.216666 | 0.862011 | | 3.917911 | 0.476337 | 0.664512 |
| SPANXC | 2.278514 | 0.123993 | 0.999532 | | 2.091452 | -0.3274 | 0.862011 | | 1.977385 | -0.43429 | 0.809485 |
| C17orf10 | 2.581436 | -0.23339 | 0.999532 | | 2.744057 | -0.28066 | 0.862011 | | 2.211349 | -0.03722 | 0.991134 |
| LTA | 2.667538 | 0.541135 | 0.999532 | | 2.341984 | -0.31238 | 0.862011 | | 2.450226 | -0.83441 | 0.515762 |
| SLC25A1 | 3.117353 | -0.17878 | 0.999532 | | 3.225603 | -0.25259 | 0.862011 | | 2.921452 | -0.04158 | 0.987859 |
| ANXA6 | 8.453678 | -0.10282 | 0.999532 | | 10.66125 | 0.172718 | 0.862011 | | 8.698375 | 0.290859 | 0.749206 |
| GJB3 | 2.226614 | -0.18654 | 0.999532 | | 2.42862 | -0.28818 | 0.862011 | | 2.006489 | -0.08318 | 0.973356 |
| FAM120B | 3.865416 | -0.21629 | 0.999532 | | 4.77301 | 0.222773 | 0.862011 | | 3.857835 | 0.447952 | 0.706407 |
| SLC1A1 | 3.327983 | 0.04241 | 0.999532 | | 3.389461 | -0.27885 | 0.862011 | | 3.052375 | -0.30995 | 0.849532 |
| C5orf51 | 3.189464 | 0.518065 | 0.999532 | | 3.023931 | 0.269099 | 0.862011 | | 3.185546 | -0.22385 | 0.883679 |
| EHBP1L1 | 3.826223 | -0.48637 | 0.999532 | | 5.287716 | 0.207067 | 0.862011 | | 4.183674 | 0.70309 | 0.484718 |
| RNF6 | 3.369003 | -0.10117 | 0.999532 | | 3.61042 | -0.2479 | 0.862202 | | 3.126571 | -0.13193 | 0.940744 |
| CALHM3 | 3.372177 | 0.338168 | 0.999532 | | 2.897451 | -0.26661 | 0.862202 | | 2.960095 | -0.60155 | 0.647218 |
| SLC9B2 | 2.343417 | 0.34809 | 0.999532 | | 2.28529 | -0.3261 | 0.862253 | | 2.096345 | -0.64924 | 0.659069 |
| PTPRB | 13.663 | -0.47861 | 0.999532 | | 18.85098 | 0.173161 | 0.862452 | | 14.0389 | 0.6744 | 0.216108 |
| PSME3IP | 6.869607 | 0.29207 | 0.999532 | | 6.310313 | -0.19905 | 0.862465 | | 6.164506 | -0.48371 | 0.619423 |
| OR5K1 | 2.186707 | 0.341054 | 0.999532 | | 1.918433 | -0.32542 | 0.862465 | | 1.986633 | -0.64645 | 0.669526 |
| PAQR3 | 2.341785 | 0.259642 | 0.999532 | | 2.066338 | -0.33454 | 0.862524 | | 1.963316 | -0.57919 | 0.723453 |
| CASP6 | 3.187942 | 0.000144 | 0.99982 | | 3.24199 | -0.25532 | 0.862656 | | 2.774746 | -0.23673 | 0.881837 |
| VPS37A | 4.747293 | -0.47398 | 0.999532 | | 5.558823 | -0.20195 | 0.862656 | | 3.998741 | 0.2898 | 0.808547 |
| SIRT1 | 4.609482 | 0.004834 | 0.999532 | | 4.635121 | -0.22832 | 0.862656 | | 4.033373 | -0.22235 | 0.884653 |
| EEF1B2 | 12.72024 | -0.19204 | 0.999532 | | 16.13806 | 0.138672 | 0.862656 | | 12.8398 | 0.352361 | 0.635596 |
| GNG7 | 3.310212 | 0.346982 | 0.999532 | | 3.499671 | 0.253293 | 0.862656 | | 3.404245 | -0.07313 | 0.970532 |
| C1orf43 | 8.54183 | 0.12656 | 0.999532 | | 9.499981 | 0.163033 | 0.862656 | | 8.863667 | 0.05907 | 0.96129 |
| CASC3 | 5.198761 | -0.48186 | 0.999532 | | 6.070584 | -0.19244 | 0.862656 | | 4.538812 | 0.299904 | 0.803493 |
| CDYL | 2.963018 | 0.382938 | 0.999532 | | 2.718802 | -0.27894 | 0.862656 | | 2.679449 | -0.63777 | 0.622543 |
| C1orf74 | 2.920367 | 0.057654 | 0.999532 | | 2.880034 | -0.27781 | 0.862656 | | 2.555809 | -0.31759 | 0.84357 |
| GRIN1 | 2.082066 | 0.49272 | 0.999532 | | 2.233766 | 0.326005 | 0.862656 | | 2.281917 | -0.14986 | 0.945129 |
| ZNF526 | 3.262418 | -0.01396 | 0.999532 | | 3.38777 | -0.2565 | 0.862656 | | 2.965018 | -0.22791 | 0.881837 |
| TIGD6 | 2.120078 | -0.06518 | 0.999532 | | 2.288996 | -0.30509 | 0.862656 | | 1.842538 | -0.2154 | 0.914929 |
| TEX2 | 2.66871 | 0.253638 | 0.999532 | | 2.912695 | 0.275221 | 0.862656 | | 2.620147 | 0.045626 | 0.985825 |
| IGHA1 | 3.555821 | 0.731921 | 0.999532 | | 3.027434 | 0.30338 | 0.862656 | | 3.580756 | -0.47086 | 0.765297 |
| KLHL11 | 2.967321 | 0.117668 | 0.999532 | | 3.412036 | 0.250615 | 0.862656 | | 3.059021 | 0.148889 | 0.930403 |
| RASA4 | 7.632199 | -0.30419 | 0.999532 | | 9.310523 | -0.17534 | 0.862656 | | 7.187185 | 0.170437 | 0.887166 |
| TLK1 | 3.79313 | 0.196832 | 0.999532 | | 4.388684 | 0.223701 | 0.862656 | | 3.81477 | 0.048802 | 0.978931 |
| ANK1 | 2.214211 | -0.24443 | 0.999532 | | 3.242799 | 0.309597 | 0.862817 | | 2.695974 | 0.569858 | 0.734904 |
| MBTD1 | 3.144508 | 0.302603 | 0.999532 | | 3.206541 | 0.25536 | 0.862988 | | 3.050688 | -0.04083 | 0.987274 |
| VASH2 | 3.000716 | 0.395222 | 0.999532 | | 2.748792 | -0.29978 | 0.862988 | | 2.608891 | -0.66825 | 0.628053 |
| C6orf47 | 3.577889 | -0.18895 | 0.999532 | | 5.254033 | 0.228388 | 0.862988 | | 3.989218 | 0.473899 | 0.69977 |
| TRPC6 | 4.985557 | 0.619945 | 0.999532 | | 4.696833 | 0.215482 | 0.862988 | | 5.042081 | -0.38807 | 0.700381 |
| COL25A1 | 3.569798 | -0.27689 | 0.999532 | | 3.900377 | -0.24281 | 0.863336 | | 3.096774 | 0.039246 | 0.989093 |
| AASS | 6.915377 | -0.10875 | 0.999532 | | 7.350121 | -0.1937 | 0.863478 | | 5.915609 | -0.05796 | 0.971092 |
| ARIH2OS | 2.200627 | 0.477682 | 0.999532 | | 1.894436 | -0.32584 | 0.863588 | | 1.976126 | -0.78577 | 0.572084 |
| NAP1L4 | 10.43788 | -0.23719 | 0.999532 | | 13.20953 | 0.175986 | 0.863588 | | 10.45776 | 0.425031 | 0.622543 |
| MT1X | 13.98085 | -0.22981 | 0.999532 | | 18.69058 | 0.249523 | 0.863866 | | 14.4621 | 0.534138 | 0.637179 |
| SCN7A | 1.954756 | 0.0185 | 0.999532 | | 2.013487 | -0.31844 | 0.863866 | | 1.741668 | -0.31827 | 0.868512 |
| DNAJB11 | 5.498649 | 0.217317 | 0.999532 | | 5.912932 | 0.206275 | 0.863866 | | 5.577638 | -0.00163 | 0.999385 |
| OR1Q1 | 1.690312 | 0.431696 | 0.999532 | | 1.867117 | 0.338013 | 0.863866 | | 1.880362 | -0.07556 | 0.977746 |
| FOXJ2 | 2.726258 | -0.23668 | 0.999532 | | 2.999108 | -0.25964 | 0.863866 | | 2.347934 | -0.00524 | 0.998585 |
| GRB7 | 2.739976 | 0.482978 | 0.999532 | | 2.243629 | -0.30806 | 0.863866 | | 2.470208 | -0.78173 | 0.53778 |
| MRPL24 | 5.64009 | 0.006535 | 0.999532 | | 7.062479 | 0.179835 | 0.863866 | | 5.744337 | 0.185048 | 0.864245 |
| ZNF266 | 4.103482 | -0.10564 | 0.999532 | | 4.446245 | -0.21606 | 0.863866 | | 3.691583 | -0.09256 | 0.955834 |
| ZNF136 | 3.91747 | -0.12461 | 0.999532 | | 4.143309 | -0.22496 | 0.863866 | | 3.642695 | -0.08944 | 0.960773 |
| POLR1C | 2.820492 | -0.14672 | 0.999532 | | 3.873156 | 0.239911 | 0.864082 | | 3.100435 | 0.406253 | 0.75038 |
| GXYLT2 | 2.389805 | 0.408675 | 0.999532 | | 2.019807 | -0.30855 | 0.864359 | | 2.055473 | -0.7022 | 0.622543 |
| CARS2 | 3.350587 | 0.294638 | 0.999532 | | 3.869325 | 0.23375 | 0.864359 | | 3.542643 | -0.03815 | 0.986471 |
| SEC31B | 3.463221 | -0.35129 | 0.999532 | | 4.048406 | -0.29507 | 0.864359 | | 2.985504 | 0.068401 | 0.976389 |
| SENP8 | 2.969384 | 0.421454 | 0.999532 | | 2.601505 | -0.29752 | 0.864359 | | 2.479049 | -0.69899 | 0.583371 |
| ANKRD9 | 2.361221 | -0.11946 | 0.999532 | | 2.536376 | -0.29518 | 0.864359 | | 2.054524 | -0.15723 | 0.941197 |
| SLC7A5 | 2.107258 | 0.110367 | 0.999532 | | 1.973595 | -0.33651 | 0.864359 | | 1.735895 | -0.42658 | 0.816618 |
| DDX27 | 2.826967 | 0.034677 | 0.999532 | | 3.667776 | 0.244483 | 0.864359 | | 3.156722 | 0.229861 | 0.879947 |
| TSPO2 | 2.214506 | 0.286648 | 0.999532 | | 1.949001 | -0.31366 | 0.864359 | | 2.008945 | -0.58568 | 0.70375 |
| EBI3 | 2.568747 | -0.61747 | 0.999532 | | 2.979425 | -0.26668 | 0.864807 | | 2.155431 | 0.371318 | 0.820834 |
| SCAMP3 | 6.756401 | 0.10282 | 0.999532 | | 8.022572 | 0.195433 | 0.864807 | | 6.648524 | 0.099869 | 0.939913 |
| GGT6 | 2.121499 | 0.413849 | 0.999532 | | 2.383196 | 0.301639 | 0.864807 | | 2.163014 | -0.09345 | 0.96734 |
| CTDSP1 | 6.647254 | -0.00499 | 0.999532 | | 7.478568 | 0.178711 | 0.864807 | | 6.471875 | 0.186933 | 0.870694 |
| RHEB | 7.968837 | 0.025548 | 0.999532 | | 9.295545 | 0.175846 | 0.864807 | | 8.108388 | 0.171725 | 0.864923 |
| KIF21A | 3.670414 | 0.168793 | 0.999532 | | 3.496494 | -0.25482 | 0.864807 | | 3.343588 | -0.40103 | 0.762039 |
| SPRYD3 | 4.439484 | -0.14611 | 0.999532 | | 5.695963 | 0.210456 | 0.864807 | | 4.370751 | 0.370586 | 0.723453 |
| C10orf62 | 2.410067 | 0.668166 | 0.999532 | | 2.55957 | 0.322155 | 0.864807 | | 2.687763 | -0.3395 | 0.837256 |
| HERC4 | 4.600125 | -0.13682 | 0.999532 | | 5.683554 | 0.205047 | 0.864807 | | 4.522471 | 0.36259 | 0.739496 |
| ZIM2 | 1.89692 | -0.29532 | 0.999532 | | 2.490856 | 0.28969 | 0.864807 | | 1.922794 | 0.600727 | 0.702908 |
| NDUFA11 | 6.435673 | -0.00862 | 0.999532 | | 7.681322 | 0.180959 | 0.864807 | | 6.681752 | 0.205811 | 0.84246 |
| DDX10 | 2.60965 | -0.47189 | 0.999532 | | 3.932533 | 0.244876 | 0.864807 | | 2.92402 | 0.741403 | 0.539079 |
| ZSCAN4 | 2.500901 | -0.37815 | 0.999532 | | 2.954133 | -0.27375 | 0.864807 | | 2.320374 | 0.121216 | 0.955834 |
| PIGA | 2.841925 | 0.201575 | 0.999532 | | 2.752955 | -0.27118 | 0.864807 | | 2.462914 | -0.45346 | 0.740093 |
| ARHGEF | 3.417967 | -0.08797 | 0.999532 | | 3.600743 | -0.26103 | 0.864807 | | 3.023114 | -0.15443 | 0.92697 |
| AAK1 | 3.054255 | -0.11438 | 0.999532 | | 3.660649 | 0.243845 | 0.864807 | | 3.112037 | 0.375905 | 0.774747 |
| ZNF37A | 3.443132 | 0.301953 | 0.999532 | | 3.56193 | 0.258718 | 0.864807 | | 3.505113 | -0.03705 | 0.988853 |
| MOG | 2.163675 | 0.642686 | 0.999532 | | 2.145573 | 0.30861 | 0.864807 | | 2.171618 | -0.32001 | 0.849608 |
| ILF2 | 5.234947 | -0.16091 | 0.999532 | | 6.645653 | 0.202423 | 0.864807 | | 5.136849 | 0.391944 | 0.705046 |
| COL13A1 | 2.276603 | 0.381226 | 0.999532 | | 2.088118 | -0.31376 | 0.864807 | | 2.0942 | -0.67565 | 0.646782 |
| AVPI1 | 3.787677 | 0.327094 | 0.999532 | | 3.575666 | -0.27268 | 0.864807 | | 3.431649 | -0.58773 | 0.632113 |
| SUMO1 | 9.223751 | 0.303596 | 0.999532 | | 10.10955 | 0.170065 | 0.864807 | | 9.355531 | -0.11051 | 0.916969 |
| RECQL | 3.259741 | -0.4306 | 0.999532 | | 4.485617 | 0.219375 | 0.864807 | | 3.474977 | 0.665927 | 0.536289 |
| PLPPR1 | 2.731882 | 0.399716 | 0.999532 | | 2.944145 | 0.287159 | 0.864807 | | 2.816611 | -0.09248 | 0.966817 |
| ARRDC3 | 9.311669 | -0.14327 | 0.999532 | | 11.55469 | 0.158649 | 0.864807 | | 9.150905 | 0.318696 | 0.710338 |
| C9orf135 | 2.627155 | 0.237759 | 0.999532 | | 2.627759 | -0.30314 | 0.864807 | | 2.433757 | -0.51988 | 0.728015 |
| VPS50 | 2.418046 | 0.042194 | 0.999532 | | 2.428948 | -0.29361 | 0.864807 | | 2.086436 | -0.31829 | 0.858705 |
| ADGRG6 | 2.427658 | 0.485086 | 0.999532 | | 2.089637 | -0.31384 | 0.864807 | | 2.088312 | -0.77909 | 0.558035 |
| SCAF4 | 2.519199 | -0.23428 | 0.999532 | | 3.313565 | 0.245485 | 0.864807 | | 2.593268 | 0.495873 | 0.709825 |
| MINAR1 | 2.116374 | -0.09799 | 0.999532 | | 2.221796 | -0.31543 | 0.864807 | | 1.814728 | -0.19864 | 0.92697 |
| SHANK3 | 11.26735 | -0.78875 | 0.256559 | | 15.16482 | -0.13748 | 0.864807 | | 9.933761 | 0.668128 | 0.241605 |
| EPHA8 | 2.256002 | -0.36709 | 0.999532 | | 2.777506 | -0.28042 | 0.864807 | | 2.108856 | 0.10841 | 0.963309 |
| PTTG2 | 2.144236 | 0.39589 | 0.999532 | | 2.468204 | 0.290685 | 0.864807 | | 2.39045 | -0.08616 | 0.970811 |
| CRYZL1 | 5.368844 | 0.061066 | 0.999532 | | 6.697849 | 0.190788 | 0.865057 | | 5.510735 | 0.154946 | 0.900848 |
| FMN2 | 4.675484 | 0.675386 | 0.999532 | | 4.52425 | 0.252917 | 0.865079 | | 4.756293 | -0.395 | 0.717627 |
| CYHR1 | 2.89535 | 0.015168 | 0.999532 | | 3.125601 | -0.26075 | 0.865106 | | 2.566547 | -0.25221 | 0.876781 |
| SEC13 | 2.03328 | 0.425217 | 0.999532 | | 2.177203 | 0.291669 | 0.865106 | | 2.028275 | -0.12062 | 0.955167 |
| MAP4K2 | 4.589252 | 0.191002 | 0.999532 | | 4.696299 | -0.2119 | 0.865215 | | 4.140614 | -0.38102 | 0.720526 |
| FAM135A | 2.768495 | -0.41854 | 0.999532 | | 3.2763 | -0.24987 | 0.865215 | | 2.541156 | 0.18749 | 0.914929 |
| ARNTL | 1.951008 | -0.14821 | 0.999532 | | 2.041333 | -0.30221 | 0.865215 | | 1.784014 | -0.13622 | 0.95365 |
| H4C13 | 4.169844 | -0.00928 | 0.999532 | | 5.070734 | 0.216915 | 0.865215 | | 4.230363 | 0.24687 | 0.842754 |
| FAM193A | 5.11133 | -0.01731 | 0.999532 | | 5.410884 | -0.21721 | 0.865215 | | 4.401047 | -0.17637 | 0.88885 |
| CARMIL1 | 2.700479 | 0.19844 | 0.999532 | | 2.545465 | -0.28176 | 0.865215 | | 2.380018 | -0.45928 | 0.75038 |
| ARHGAP | 3.33797 | -0.52836 | 0.999532 | | 4.118909 | -0.23309 | 0.865259 | | 2.706675 | 0.314121 | 0.836453 |
| PDIA4 | 6.447804 | 0.100227 | 0.999532 | | 7.016112 | 0.198775 | 0.865738 | | 6.239823 | 0.093748 | 0.950616 |
| SULT1A2 | 4.840636 | 0.15801 | 0.999532 | | 4.885638 | -0.21309 | 0.865905 | | 4.367779 | -0.3551 | 0.735774 |
| NCEH1 | 2.947147 | -0.25178 | 0.999532 | | 3.278479 | -0.25263 | 0.865941 | | 2.546709 | 0.016667 | 0.996681 |
| ZBTB7A | 8.322034 | -0.24101 | 0.999532 | | 9.630019 | -0.16105 | 0.865941 | | 7.5406 | 0.097971 | 0.93603 |
| LTO1 | 3.747674 | -0.08881 | 0.999532 | | 4.103001 | -0.22614 | 0.865941 | | 3.454508 | -0.11972 | 0.941197 |
| CBX4 | 3.94951 | -0.60788 | 0.999532 | | 5.08383 | -0.20837 | 0.865941 | | 3.601819 | 0.415785 | 0.722703 |
| AMACR | 1.974522 | 0.270302 | 0.999532 | | 2.261832 | 0.294775 | 0.865941 | | 2.180734 | 0.047206 | 0.987274 |
| SCFD1 | 3.926735 | -0.14706 | 0.999532 | | 5.212054 | 0.203527 | 0.865941 | | 4.351624 | 0.369821 | 0.730921 |
| ZNF511 | 2.437964 | 0.10549 | 0.999532 | | 2.363815 | -0.29703 | 0.865941 | | 2.227219 | -0.37966 | 0.815713 |
| POLA1 | 2.094346 | 0.191392 | 0.999532 | | 2.355117 | 0.286086 | 0.865941 | | 2.134182 | 0.110441 | 0.95999 |
| RAB11FIP | 4.918272 | -0.01637 | 0.999532 | | 5.090316 | -0.20998 | 0.865941 | | 4.311657 | -0.1779 | 0.894617 |
| NRM | 3.484527 | 0.03438 | 0.999532 | | 3.485914 | -0.24786 | 0.865941 | | 2.869856 | -0.26156 | 0.86667 |
| BIVM | 2.861152 | -0.12104 | 0.999532 | | 3.689422 | 0.23753 | 0.865941 | | 3.093637 | 0.3757 | 0.774747 |
| ZDHHC4 | 3.555201 | 0.218873 | 0.999532 | | 3.334378 | -0.25687 | 0.865941 | | 3.101105 | -0.46268 | 0.709825 |
| PRPF4B | 11.12223 | 0.066523 | 0.999532 | | 12.47358 | 0.140321 | 0.865941 | | 11.10605 | 0.091143 | 0.92403 |
| AOC1 | 5.443776 | -0.36739 | 0.999532 | | 7.381428 | 0.354133 | 0.865941 | | 5.789819 | 0.677282 | 0.718643 |
| CR2 | 2.808984 | -0.29773 | 0.999532 | | 3.306893 | -0.25688 | 0.865941 | | 2.552925 | 0.058972 | 0.978931 |
| TEAD2 | 9.698281 | -0.13711 | 0.999532 | | 11.80562 | 0.155858 | 0.865941 | | 9.740381 | 0.321444 | 0.658914 |
| ZFP69B | 2.101108 | 0.118257 | 0.999532 | | 2.184779 | -0.32177 | 0.865941 | | 1.979334 | -0.42622 | 0.809832 |
| MYOM3 | 3.715929 | 0.290439 | 0.999532 | | 3.3053 | -0.24608 | 0.865941 | | 3.421274 | -0.52752 | 0.663353 |
| RASA2 | 3.774101 | -0.10586 | 0.999532 | | 4.883128 | 0.220321 | 0.865941 | | 3.941478 | 0.348531 | 0.774749 |
| ENO2 | 4.558099 | -0.32664 | 0.999532 | | 5.326209 | -0.22604 | 0.865941 | | 3.975341 | 0.127168 | 0.93603 |
| RBMS3 | 7.76018 | -0.16718 | 0.999532 | | 8.777401 | -0.19075 | 0.866074 | | 7.142202 | 0.00222 | 0.999385 |
| ACAD11 | 3.242214 | 0.147892 | 0.999532 | | 3.555317 | -0.2605 | 0.866142 | | 2.87818 | -0.36461 | 0.800239 |
| GAL3ST1 | 2.184759 | 0.198944 | 0.999532 | | 2.503142 | 0.287408 | 0.866403 | | 2.414598 | 0.113848 | 0.957645 |
| ANKHD1 | 6.545741 | -0.19952 | 0.999532 | | 8.162205 | 0.162012 | 0.866412 | | 6.616952 | 0.374402 | 0.66815 |
| C17orf11 | 2.388086 | 0.645204 | 0.999532 | | 2.282393 | 0.298194 | 0.866417 | | 2.472516 | -0.33014 | 0.837256 |
| FLII | 5.485802 | -0.22806 | 0.999532 | | 6.58766 | 0.180709 | 0.866417 | | 5.556851 | 0.412687 | 0.682485 |
| OPTN | 10.93131 | 0.111764 | 0.999532 | | 12.5752 | 0.150978 | 0.866499 | | 10.99812 | 0.056707 | 0.958422 |
| EDC4 | 5.424464 | 0.100431 | 0.999532 | | 5.369232 | -0.20435 | 0.866499 | | 4.742128 | -0.28285 | 0.795777 |
| KMT2B | 3.431313 | 0.315725 | 0.999532 | | 3.777956 | 0.236635 | 0.866499 | | 3.547349 | -0.06007 | 0.974421 |
| ATP13A1 | 4.117146 | 0.009195 | 0.999532 | | 5.047399 | 0.200433 | 0.866499 | | 4.253786 | 0.20966 | 0.867009 |
| RARS1 | 3.677244 | 0.217611 | 0.999532 | | 4.3478 | 0.219853 | 0.866775 | | 3.915079 | 0.023375 | 0.992286 |
| MRNIP | 3.945897 | 0.088084 | 0.999532 | | 3.926346 | -0.22903 | 0.866775 | | 3.348575 | -0.30356 | 0.816618 |
| KLB | 2.396911 | 0.500136 | 0.999532 | | 2.118649 | -0.31804 | 0.866775 | | 2.266195 | -0.80675 | 0.542124 |
| TNKS1BP | 5.124267 | 0.465651 | 0.999532 | | 5.205607 | 0.19768 | 0.866775 | | 5.260206 | -0.24952 | 0.812032 |
| HOXA10 | 3.902615 | 0.09754 | 0.999532 | | 4.317856 | 0.212349 | 0.866775 | | 3.932814 | 0.123565 | 0.936689 |
| DPP6 | 3.575028 | 0.989608 | 0.950261 | | 3.178798 | 0.277762 | 0.866775 | | 3.638275 | -0.69292 | 0.496445 |
| TOM1 | 4.865013 | 0.004341 | 0.999532 | | 4.961387 | -0.21921 | 0.866775 | | 4.174636 | -0.20195 | 0.886495 |
| SAMD9 | 2.909661 | 0.198757 | 0.999532 | | 2.757493 | -0.27565 | 0.866775 | | 2.644894 | -0.46326 | 0.746876 |
| DHX40 | 3.371347 | -0.42132 | 0.999532 | | 4.749986 | 0.208132 | 0.866775 | | 3.670171 | 0.649316 | 0.515762 |
| TMEM123 | 5.517666 | -0.33834 | 0.999532 | | 6.387709 | -0.19098 | 0.866775 | | 4.596612 | 0.171377 | 0.892225 |
| PPP1R16 | 2.914159 | 0.675426 | 0.999532 | | 2.687728 | 0.270462 | 0.866949 | | 2.891683 | -0.38436 | 0.78258 |
| PRTN3 | 2.95858 | -0.06953 | 0.999532 | | 3.160816 | -0.27995 | 0.867008 | | 2.77038 | -0.20541 | 0.90928 |
| CEP76 | 1.743137 | 0.304668 | 0.999532 | | 1.999392 | 0.321626 | 0.867008 | | 1.939558 | 0.037956 | 0.991341 |
| CNST | 3.210466 | 0.00668 | 0.999532 | | 3.528166 | -0.25181 | 0.867008 | | 2.968053 | -0.21987 | 0.89383 |
| PDS5A | 4.565113 | 0.143648 | 0.999532 | | 5.152206 | 0.202147 | 0.867008 | | 4.531652 | 0.072432 | 0.964713 |
| HTATSF1 | 4.969931 | -0.02954 | 0.999532 | | 6.061811 | 0.195149 | 0.867008 | | 4.997615 | 0.23346 | 0.836453 |
| FBXO43 | 2.27441 | 0.276702 | 0.999532 | | 2.046177 | -0.31154 | 0.867008 | | 2.032743 | -0.57048 | 0.720106 |
| MAU2 | 2.973939 | -0.18588 | 0.999532 | | 3.259089 | -0.24714 | 0.867008 | | 2.619482 | -0.04631 | 0.985693 |
| PCK2 | 3.450536 | 0.319798 | 0.999532 | | 3.483707 | 0.244748 | 0.867129 | | 3.430777 | -0.07629 | 0.971732 |
| MRPS27 | 5.891654 | -0.44332 | 0.999532 | | 6.943965 | -0.19338 | 0.867129 | | 4.917877 | 0.272526 | 0.815713 |
| VSIG8 | 5.343653 | 0.103955 | 0.999532 | | 6.286264 | 0.224726 | 0.867235 | | 5.536914 | 0.136513 | 0.925023 |
| NAPB | 2.059374 | 0.330314 | 0.999532 | | 2.056407 | 0.299735 | 0.867235 | | 2.045628 | -0.01909 | 0.996681 |
| GSTA4 | 3.30178 | 0.41184 | 0.999532 | | 3.295172 | 0.242954 | 0.867619 | | 3.205602 | -0.15922 | 0.918694 |
| BTN3A3 | 6.733806 | 0.024036 | 0.999532 | | 8.266631 | 0.176281 | 0.867619 | | 7.168618 | 0.173496 | 0.876781 |
| LYPLA2 | 3.148117 | -0.07178 | 0.999532 | | 4.12593 | 0.232393 | 0.867619 | | 3.443676 | 0.328175 | 0.809832 |
| KDM1B | 2.10288 | 0.011253 | 0.999532 | | 2.217374 | -0.30342 | 0.867619 | | 1.954919 | -0.29276 | 0.879538 |
| STX10 | 3.943149 | 0.085326 | 0.999532 | | 4.828943 | 0.213226 | 0.867619 | | 4.20731 | 0.147092 | 0.915533 |
| INTU | 3.66484 | 0.24584 | 0.999532 | | 3.957622 | 0.237619 | 0.867619 | | 3.798375 | 0.015237 | 0.996681 |
| SUPT16H | 5.125558 | -0.233 | 0.999532 | | 6.334885 | 0.187081 | 0.867619 | | 5.126112 | 0.432348 | 0.660958 |
| CETP | 2.059454 | 0.06911 | 0.999532 | | 2.069195 | -0.31274 | 0.867645 | | 1.868587 | -0.38513 | 0.842152 |
| OAF | 4.27483 | 0.153119 | 0.999532 | | 4.072841 | -0.2241 | 0.867845 | | 3.726682 | -0.36113 | 0.758124 |
| FCGR2C | 9.780198 | -0.00753 | 0.999532 | | 11.70559 | 0.186027 | 0.867928 | | 10.00908 | 0.210415 | 0.870694 |
| DPM1 | 5.407536 | 0.020554 | 0.999532 | | 6.487945 | 0.186986 | 0.867928 | | 5.532468 | 0.186667 | 0.87892 |
| PTPN9 | 4.055153 | 0.018706 | 0.999532 | | 4.161776 | -0.22491 | 0.867928 | | 3.694045 | -0.2252 | 0.87591 |
| BPHL | 4.460024 | -0.17919 | 0.999532 | | 4.742566 | -0.20706 | 0.867928 | | 3.903155 | -0.0167 | 0.995943 |
| IFT22 | 2.871646 | 0.462232 | 0.999532 | | 2.954507 | 0.25971 | 0.867928 | | 2.915937 | -0.18178 | 0.908595 |
| POP5 | 2.467253 | 0.035018 | 0.999532 | | 3.085089 | 0.255145 | 0.867928 | | 2.710677 | 0.241001 | 0.881837 |
| NR3C2 | 7.218576 | 0.161248 | 0.999532 | | 7.231533 | -0.18357 | 0.867928 | | 6.532824 | -0.32539 | 0.722703 |
| SLC25A1 | 2.692411 | -0.01413 | 0.999532 | | 2.712498 | -0.28839 | 0.868216 | | 2.371313 | -0.25579 | 0.88476 |
| PTPN6 | 4.203941 | 0.00835 | 0.999532 | | 4.35118 | -0.21659 | 0.868216 | | 3.678307 | -0.21125 | 0.88082 |
| THADA | 2.185172 | -0.65209 | 0.999532 | | 2.554764 | -0.27753 | 0.868368 | | 1.832577 | 0.38661 | 0.835554 |
| WBP11 | 6.469386 | -0.31554 | 0.999532 | | 7.48355 | -0.17893 | 0.86854 | | 5.646276 | 0.155675 | 0.893853 |
| TAPBPL | 4.684731 | -0.09202 | 0.999532 | | 5.04406 | -0.20965 | 0.86854 | | 4.16551 | -0.09405 | 0.955167 |
| ECT2 | 2.717089 | 0.38143 | 0.999532 | | 2.380875 | -0.2909 | 0.86854 | | 2.568791 | -0.6546 | 0.64142 |
| MSANTD | 2.211222 | -0.00964 | 0.999532 | | 2.197929 | -0.3134 | 0.86854 | | 1.779831 | -0.28165 | 0.88476 |
| USP27X | 2.448547 | 0.51733 | 0.999532 | | 2.553159 | 0.292117 | 0.86854 | | 2.646819 | -0.19685 | 0.914687 |
| ZNF805 | 3.600676 | -0.10042 | 0.999532 | | 3.798414 | -0.22984 | 0.86854 | | 3.138749 | -0.11094 | 0.947557 |
| RALGAP | 3.312572 | 0.040112 | 0.999532 | | 3.391946 | -0.25032 | 0.868625 | | 2.99357 | -0.26805 | 0.86114 |
| TEX30 | 3.130571 | -0.42134 | 0.999532 | | 3.757023 | -0.24349 | 0.868746 | | 2.706192 | 0.19672 | 0.907882 |
| GPT2 | 3.939569 | -0.1364 | 0.999532 | | 5.057448 | 0.219741 | 0.868746 | | 4.151857 | 0.37557 | 0.753585 |
| FAM199X | 2.731184 | -0.08836 | 0.999532 | | 3.184224 | -0.2519 | 0.868746 | | 2.522275 | -0.14553 | 0.936303 |
| GINS2 | 2.959367 | 0.150422 | 0.999532 | | 3.142168 | -0.28759 | 0.868746 | | 2.874017 | -0.42171 | 0.791576 |
| CCDC174 | 2.7726 | -0.30121 | 0.999532 | | 3.216586 | -0.24391 | 0.868746 | | 2.488871 | 0.075592 | 0.972758 |
| PIK3CB | 3.206933 | 0.25744 | 0.999532 | | 3.571993 | 0.252735 | 0.868746 | | 3.202118 | 0.020364 | 0.994537 |
| TINAGL1 | 24.5747 | -0.22689 | 0.999532 | | 31.45017 | 0.138743 | 0.868746 | | 24.92824 | 0.385237 | 0.515762 |
| DNAJB14 | 3.850439 | -0.29079 | 0.999532 | | 5.106096 | 0.213081 | 0.868746 | | 3.924739 | 0.524599 | 0.632113 |
| DRAM1 | 4.087533 | -0.35259 | 0.999532 | | 5.204283 | 0.20577 | 0.868746 | | 4.173366 | 0.557412 | 0.628895 |
| SPATA4 | 2.394665 | 0.357428 | 0.999532 | | 2.214599 | -0.32255 | 0.868746 | | 2.17102 | -0.66063 | 0.666566 |
| SLC30A4 | 4.58356 | -0.12574 | 0.999532 | | 4.844237 | -0.2067 | 0.868746 | | 4.019829 | -0.06714 | 0.972135 |
| UBE2J2 | 2.099448 | 0.034975 | 0.999532 | | 2.50208 | 0.278143 | 0.868746 | | 2.164054 | 0.261723 | 0.881837 |
| C9orf47 | 2.851911 | -0.00553 | 0.999532 | | 3.047521 | -0.26179 | 0.868746 | | 2.651707 | -0.24278 | 0.881837 |
| GNPAT | 1.922949 | -0.17679 | 0.999532 | | 2.474413 | 0.269294 | 0.868746 | | 2.107955 | 0.468072 | 0.757037 |
| SCAMP5 | 2.760585 | 0.205211 | 0.999532 | | 3.089983 | 0.251196 | 0.868746 | | 2.991613 | 0.057935 | 0.978408 |
| CYP4F2 | 3.243532 | -0.2403 | 0.999532 | | 3.675656 | -0.26203 | 0.868746 | | 2.794311 | 0.001693 | 0.999429 |
| PLCL1 | 2.728175 | 0.079096 | 0.999532 | | 3.140016 | 0.240951 | 0.868746 | | 2.751966 | 0.178958 | 0.909074 |
| CXorf65 | 2.156866 | 0.679986 | 0.999532 | | 1.721018 | -0.33562 | 0.868746 | | 1.965357 | -0.99804 | 0.457856 |
| DGAT1 | 4.359357 | -0.14051 | 0.999532 | | 4.602142 | -0.23113 | 0.868746 | | 3.687804 | -0.06618 | 0.972305 |
| PDE1A | 4.740421 | -0.02549 | 0.999532 | | 5.644991 | 0.193233 | 0.86892 | | 4.784473 | 0.234355 | 0.846488 |
| MRPS33 | 3.129087 | -0.04461 | 0.999532 | | 3.804484 | 0.227601 | 0.86892 | | 3.392391 | 0.295853 | 0.837256 |
| KDM5B | 2.474584 | 0.094513 | 0.999532 | | 2.765321 | 0.26691 | 0.86892 | | 2.567295 | 0.178795 | 0.920259 |
| BATF3 | 2.357064 | 0.208153 | 0.999532 | | 2.642707 | 0.274973 | 0.868955 | | 2.359488 | 0.08075 | 0.972396 |
| ANKRD3 | 15.89925 | 0.048419 | 0.999532 | | 16.48521 | -0.15714 | 0.868974 | | 14.30402 | -0.17759 | 0.827267 |
| OR52B2 | 1.923199 | 0.163542 | 0.999532 | | 1.85672 | -0.32131 | 0.868974 | | 1.640425 | -0.46747 | 0.795612 |
| SPNS3 | 2.14307 | -0.12449 | 0.999532 | | 2.402007 | -0.29878 | 0.869311 | | 1.99 | -0.15237 | 0.946512 |
| STAU2 | 5.20442 | -0.19797 | 0.999532 | | 5.697896 | -0.20598 | 0.869311 | | 4.423243 | 0.0126 | 0.996681 |
| KLHDC9 | 2.201421 | 0.129377 | 0.999532 | | 2.292104 | -0.2989 | 0.869311 | | 1.891219 | -0.41174 | 0.806394 |
| TRAPPC | 2.870627 | -0.16997 | 0.999532 | | 3.305869 | -0.25291 | 0.869354 | | 2.578208 | -0.06499 | 0.97733 |
| NFE4 | 2.078443 | 0.361006 | 0.999532 | | 1.960811 | -0.31639 | 0.869354 | | 1.778984 | -0.6574 | 0.662818 |
| ALG6 | 2.906122 | 0.358491 | 0.999532 | | 2.670198 | -0.27023 | 0.869354 | | 2.647796 | -0.60678 | 0.647581 |
| ERI3 | 3.559829 | -0.10447 | 0.999532 | | 4.496292 | 0.230147 | 0.869354 | | 3.735924 | 0.355214 | 0.76172 |
| ZNF69 | 5.439247 | 0.465773 | 0.999532 | | 4.775784 | -0.21823 | 0.869354 | | 4.994156 | -0.66852 | 0.496762 |
| PIKFYVE | 3.093163 | -0.36726 | 0.999532 | | 3.960779 | 0.21948 | 0.869354 | | 3.039723 | 0.598405 | 0.60773 |
| ADAMTS | 2.449844 | 0.252065 | 0.999532 | | 2.798799 | 0.277677 | 0.869354 | | 2.563787 | 0.036875 | 0.989437 |
| DIAPH3 | 2.697992 | 0.464948 | 0.999532 | | 2.383757 | -0.28704 | 0.869354 | | 2.325919 | -0.73827 | 0.568812 |
| GOSR1 | 6.534734 | 0.235548 | 0.999532 | | 7.166029 | 0.174352 | 0.869354 | | 6.311513 | -0.05129 | 0.972758 |
| TEDC1 | 2.30498 | 0.265749 | 0.999532 | | 2.067415 | -0.31457 | 0.869354 | | 2.112869 | -0.57023 | 0.723933 |
| EMP2 | 5.177044 | -0.26529 | 0.999532 | | 6.389722 | 0.186189 | 0.869354 | | 5.24381 | 0.441036 | 0.669689 |
| CCDC25 | 4.766056 | 0.381598 | 0.999532 | | 4.364791 | -0.21699 | 0.869354 | | 4.233786 | -0.57953 | 0.540342 |
| SPRTN | 2.851456 | 0.758364 | 0.999532 | | 2.273443 | -0.28648 | 0.869354 | | 2.567891 | -1.02503 | 0.30352 |
| RPL7A | 53.54938 | -0.02812 | 0.999532 | | 62.4236 | 0.103163 | 0.869354 | | 53.16409 | 0.143285 | 0.808547 |
| CCDC18 | 3.954571 | 0.214055 | 0.999532 | | 3.767067 | -0.23168 | 0.869354 | | 3.468816 | -0.42848 | 0.71963 |
| ACAA1 | 5.436187 | 0.38896 | 0.999532 | | 5.641569 | 0.213309 | 0.869488 | | 5.538358 | -0.16224 | 0.897958 |
| NANOS1 | 2.061971 | 0.426272 | 0.999532 | | 1.864726 | -0.32361 | 0.869548 | | 1.77323 | -0.72773 | 0.631563 |
| KCNE3 | 2.622096 | 0.228745 | 0.999532 | | 2.493596 | -0.27799 | 0.869577 | | 2.279323 | -0.48649 | 0.735469 |
| CNPY3 | 6.186033 | -0.20021 | 0.999532 | | 7.979694 | 0.177461 | 0.869577 | | 6.340028 | 0.397201 | 0.666566 |
| USP31 | 2.076727 | -0.05465 | 0.999532 | | 2.101903 | -0.29122 | 0.869577 | | 1.77287 | -0.22236 | 0.9094 |
| SLFN12 | 3.949644 | 0.150073 | 0.999532 | | 3.96303 | -0.22671 | 0.869577 | | 3.644808 | -0.35517 | 0.779843 |
| KLHDC1 | 4.168602 | -0.11072 | 0.999532 | | 5.213836 | 0.199324 | 0.869647 | | 4.096625 | 0.329181 | 0.765852 |
| [illegible] | [illegible] | [illegible] | [illegible] | | [illegible] | [illegible] | [illegible] | | [illegible] | [illegible] | [illegible] |
| TLK2 | 13.89426 | -0.03372 | 0.999532 | | 14.46976 | -0.12529 | 0.869647 | | 12.77394 | -0.07095 | 0.947877 |
| PAFAH2 | 2.236547 | 0.124937 | 0.999532 | | 2.081782 | -0.30814 | 0.869647 | | 1.935464 | -0.42311 | 0.809832 |
| PABPC1L | 2.522661 | -0.08792 | 0.999532 | | 2.758427 | -0.27789 | 0.869647 | | 2.448994 | -0.17018 | 0.934354 |
| MRGPRX | 1.826792 | -0.17784 | 0.999532 | | 2.321141 | 0.310439 | 0.869768 | | 1.89926 | 0.503631 | 0.779948 |
| RELB | 2.961637 | -0.31898 | 0.999532 | | 3.756013 | 0.234761 | 0.870136 | | 2.813007 | 0.575662 | 0.645376 |
| LDHA | 13.73841 | -0.30233 | 0.999532 | | 18.23498 | 0.12957 | 0.870145 | | 14.06125 | 0.453034 | 0.418768 |
| TRARG1 | 2.639918 | 0.484013 | 0.999532 | | 2.255876 | -0.30849 | 0.870396 | | 2.353005 | -0.78319 | 0.560424 |
| ANKRD3 | 4.21837 | -0.29251 | 0.999532 | | 5.388268 | 0.197689 | 0.870555 | | 3.995388 | 0.510886 | 0.624892 |
| FAM76B | 2.290148 | 0.399031 | 0.999532 | | 1.942437 | -0.30855 | 0.87086 | | 2.06503 | -0.69296 | 0.64142 |
| ILKAP | 3.044609 | -0.29844 | 0.999532 | | 3.511337 | -0.23524 | 0.870888 | | 2.80366 | 0.080149 | 0.971732 |
| RASL10A | 2.759514 | 0.266853 | 0.999532 | | 3.282127 | 0.265061 | 0.871032 | | 3.118386 | 0.007444 | 0.997967 |
| DERL2 | 5.33588 | -0.11663 | 0.999532 | | 5.869802 | -0.19846 | 0.871032 | | 4.791998 | -0.06043 | 0.972924 |
| UPK3BL2 | 20.68044 | -0.15585 | 0.999532 | | 25.963 | 0.14325 | 0.871032 | | 21.30688 | 0.312426 | 0.622543 |
| CD109 | 5.324493 | 0.039513 | 0.999532 | | 6.35747 | 0.187782 | 0.871102 | | 5.406242 | 0.166667 | 0.88885 |
| PEX11A | 2.573821 | -0.04391 | 0.999532 | | 2.78284 | -0.26323 | 0.871102 | | 2.399992 | -0.20115 | 0.909259 |
| HOXB9 | 3.133215 | -0.01656 | 0.999532 | | 3.128338 | -0.25664 | 0.871102 | | 2.830167 | -0.23067 | 0.897096 |
| ING3 | 4.416872 | 0.183318 | 0.999532 | | 4.34119 | -0.21251 | 0.871306 | | 3.871481 | -0.37658 | 0.727913 |
| SERF1A | 4.907481 | -0.63718 | 0.999532 | | 6.398632 | -0.19507 | 0.871306 | | 4.43972 | 0.462053 | 0.666566 |
| CETN1 | 2.602603 | 0.107953 | 0.999532 | | 2.57599 | -0.27609 | 0.871369 | | 2.279814 | -0.365 | 0.824587 |
| WDR47 | 2.346672 | -0.35527 | 0.999532 | | 2.701784 | -0.28546 | 0.871462 | | 2.107244 | 0.06882 | 0.972758 |
| CCNG2 | 7.533503 | 0.055674 | 0.999532 | | 8.980321 | 0.161544 | 0.871462 | | 7.574525 | 0.12332 | 0.909074 |
| SSR1 | 9.256673 | -0.07045 | 0.999532 | | 11.25355 | 0.182322 | 0.871462 | | 9.333529 | 0.276859 | 0.78258 |
| GDF7 | 2.374226 | -0.03368 | 0.999532 | | 2.883342 | 0.271334 | 0.871462 | | 2.495473 | 0.321411 | 0.851785 |
| CASKIN2 | 4.932692 | 0.101139 | 0.999532 | | 5.800908 | 0.190264 | 0.871462 | | 5.07992 | 0.111518 | 0.938429 |
| PSMC1 | 9.205154 | 0.349801 | 0.999532 | | 9.669605 | 0.15706 | 0.871765 | | 9.357816 | -0.17469 | 0.846155 |
| ZNF605 | 2.05714 | 0.23615 | 0.999532 | | 2.507457 | 0.283896 | 0.871817 | | 2.079672 | 0.067989 | 0.977635 |
| CCNL2 | 8.031706 | -0.0474 | 0.999532 | | 9.699234 | 0.161667 | 0.871817 | | 8.110192 | 0.228321 | 0.806339 |
| ONECUT | 2.26008 | 0.295245 | 0.999532 | | 2.198267 | -0.30833 | 0.871974 | | 2.056351 | -0.57524 | 0.720106 |
| KCNMB4 | 4.431527 | -0.11397 | 0.999532 | | 4.924412 | -0.22883 | 0.871974 | | 3.876108 | -0.09295 | 0.958889 |
| FOXJ1 | 2.396288 | 0.430883 | 0.999532 | | 2.129657 | -0.29088 | 0.871974 | | 2.061515 | -0.7027 | 0.5991 |
| EDN1 | 2.309851 | 0.386684 | 0.999532 | | 1.87171 | -0.30538 | 0.871974 | | 2.046673 | -0.67099 | 0.652095 |
| YKT6 | 4.566412 | -0.23369 | 0.999532 | | 5.654626 | 0.188472 | 0.871974 | | 4.440384 | 0.436139 | 0.668481 |
| FOXN1 | 2.404946 | 0.337835 | 0.999532 | | 2.401439 | -0.30458 | 0.872054 | | 2.165572 | -0.6275 | 0.66232 |
| FAM131C | 6.463506 | 0.382889 | 0.999532 | | 5.740044 | -0.2028 | 0.872054 | | 5.855727 | -0.57862 | 0.563455 |
| PTPRQ | 8.041518 | 0.228609 | 0.999532 | | 9.028889 | 0.182956 | 0.872129 | | 8.221687 | -0.03372 | 0.982124 |
| RAB31 | 3.451758 | -0.08906 | 0.999532 | | 3.751006 | -0.24061 | 0.872129 | | 2.921949 | -0.12536 | 0.946512 |
| VPS37C | 4.053706 | -0.38414 | 0.999532 | | 4.80174 | -0.2135 | 0.872495 | | 3.616655 | 0.187409 | 0.900797 |
| ANXA9 | 2.463039 | 0.420128 | 0.999532 | | 2.603465 | 0.272734 | 0.872508 | | 2.436917 | -0.13211 | 0.944757 |
| FER1L5 | 2.370404 | -0.02703 | 0.999532 | | 2.44942 | -0.29305 | 0.872508 | | 2.137243 | -0.24822 | 0.898369 |
| DIAPH2 | 5.054229 | 0.320775 | 0.999532 | | 5.563642 | 0.198084 | 0.872508 | | 5.471241 | -0.11318 | 0.932589 |
| YAF2 | 3.143601 | -0.3054 | 0.999532 | | 3.322294 | -0.24001 | 0.872508 | | 2.801537 | 0.070645 | 0.97502 |
| RNF183 | 2.112793 | -0.14814 | 0.999532 | | 2.394231 | -0.2834 | 0.872508 | | 1.911533 | -0.11185 | 0.962716 |
| PRMT5 | 2.431659 | 0.044781 | 0.999532 | | 3.129641 | 0.256729 | 0.872508 | | 2.618682 | 0.242533 | 0.88885 |
| C10orf99 | 2.688166 | 0.622812 | 0.999532 | | 2.683894 | 0.302622 | 0.872508 | | 2.851101 | -0.3082 | 0.859426 |
| WDR73 | 3.087478 | -0.31768 | 0.999532 | | 4.08073 | 0.219857 | 0.872654 | | 3.009398 | 0.542385 | 0.652925 |
| FGF6 | 2.955635 | 0.208713 | 0.999532 | | 2.757594 | -0.25608 | 0.872735 | | 2.634844 | -0.44889 | 0.739496 |
| DAGLA | 2.172634 | 0.14236 | 0.999532 | | 1.968491 | -0.30477 | 0.872753 | | 1.975905 | -0.43116 | 0.809832 |
| DTD1 | 3.024992 | -0.06346 | 0.999532 | | 3.729698 | 0.245499 | 0.873503 | | 3.117776 | 0.331705 | 0.813143 |
| P2RY8 | 4.097292 | 0.30953 | 0.999532 | | 3.985315 | -0.24266 | 0.873503 | | 3.902424 | -0.53847 | 0.650461 |
| STRC | 2.464542 | -0.02754 | 0.999532 | | 2.583777 | -0.27038 | 0.873503 | | 2.158604 | -0.2267 | 0.901033 |
| HPSE2 | 1.857638 | 0.051301 | 0.999532 | | 1.779916 | -0.31143 | 0.873503 | | 1.679209 | -0.34378 | 0.861737 |
| ORC1 | 2.650116 | 0.224545 | 0.999532 | | 2.619947 | -0.2649 | 0.873503 | | 2.434393 | -0.47167 | 0.732486 |
| ADAP1 | 1.881939 | 0.512197 | 0.999532 | | 1.972628 | 0.304869 | 0.873503 | | 2.029218 | -0.18781 | 0.92403 |
| HEYL | 2.466365 | -0.18941 | 0.999532 | | 3.582782 | 0.253565 | 0.873503 | | 2.802665 | 0.48169 | 0.740093 |
| CASP8AP | 3.380711 | 0.395315 | 0.999532 | | 3.471768 | 0.233633 | 0.873503 | | 3.445539 | -0.14405 | 0.926586 |
| TAF6 | 4.733037 | -0.07736 | 0.999532 | | 5.975266 | 0.199779 | 0.873503 | | 5.199992 | 0.293708 | 0.78258 |
| FAM166A | 2.280435 | 0.953606 | 0.999532 | | 1.917117 | 0.313455 | 0.873503 | | 2.37002 | -0.62382 | 0.653214 |
| GBGT1 | 2.426898 | 0.596666 | 0.999532 | | 2.582136 | 0.276322 | 0.873503 | | 2.624782 | -0.29168 | 0.861656 |
| NLN | 2.445061 | 0.21024 | 0.999532 | | 2.738307 | 0.268446 | 0.873503 | | 2.606173 | 0.073112 | 0.974421 |
| GIMAP2 | 4.375161 | 0.176889 | 0.999532 | | 5.062319 | 0.207248 | 0.873503 | | 4.80856 | 0.066267 | 0.973754 |
| NLRC5 | 2.091325 | 0.038997 | 0.999532 | | 2.582688 | 0.275136 | 0.873503 | | 2.244231 | 0.256487 | 0.889117 |
| GM2A | 2.843706 | 0.077542 | 0.999532 | | 3.359384 | 0.238131 | 0.873503 | | 2.927581 | 0.177925 | 0.909431 |
| SPRR2B | 2.347612 | 0.08122 | 0.999532 | | 3.102572 | 0.287232 | 0.873503 | | 2.58804 | 0.227724 | 0.900848 |
| DNTTIP1 | 3.031104 | -0.03774 | 0.999532 | | 3.09243 | -0.24894 | 0.873503 | | 2.645561 | -0.19519 | 0.907626 |
| MIB2 | 2.953344 | 0.460064 | 0.999532 | | 2.924868 | 0.249951 | 0.873503 | | 2.964753 | -0.19353 | 0.900848 |
| BMP4 | 4.755436 | -0.08583 | 0.999532 | | 5.739318 | 0.183907 | 0.873503 | | 4.999156 | 0.292225 | 0.787995 |
| CLEC3B | 25.39308 | -0.04056 | 0.999532 | | 30.09455 | 0.140067 | 0.873503 | | 25.41648 | 0.205315 | 0.73978 |
| MPG | 2.980645 | -0.49835 | 0.999532 | | 3.497375 | -0.24134 | 0.873503 | | 2.497422 | 0.276655 | 0.864245 |
| ST3GAL3 | 3.861548 | -0.38999 | 0.999532 | | 4.553277 | -0.20946 | 0.873503 | | 3.350593 | 0.198124 | 0.892869 |
| HSPB6 | 2.816436 | -0.03648 | 0.999532 | | 2.76743 | -0.25715 | 0.873769 | | 2.412666 | -0.2108 | 0.905497 |
| IRF1 | 13.26138 | -0.24726 | 0.999532 | | 15.34692 | -0.17691 | 0.873769 | | 11.95228 | 0.088258 | 0.93603 |
| C11orf16 | 2.595477 | -0.02964 | 0.999532 | | 2.643124 | -0.27659 | 0.873769 | | 2.245444 | -0.23305 | 0.90405 |
| NADK2 | 3.302902 | 0.301125 | 0.999532 | | 3.135556 | -0.24944 | 0.87381 | | 2.960513 | -0.53093 | 0.666555 |
| STAM2 | 2.662411 | -0.01909 | 0.999532 | | 2.801686 | -0.25712 | 0.87381 | | 2.409524 | -0.2222 | 0.893262 |
| FNDC4 | 2.920027 | 0.428817 | 0.999532 | | 2.569229 | -0.27042 | 0.87381 | | 2.712113 | -0.68624 | 0.570037 |
| NEMF | 6.343598 | -0.23016 | 0.999532 | | 8.122138 | 0.163228 | 0.87381 | | 6.125445 | 0.396681 | 0.691311 |
| AGMAT | 6.179246 | -0.05718 | 0.999532 | | 7.607552 | 0.234674 | 0.873938 | | 6.278587 | 0.323296 | 0.79686 |
| TMEM35B | 5.403451 | -0.09079 | 0.999532 | | 6.696063 | 0.178694 | 0.873938 | | 5.435528 | 0.288643 | 0.785012 |
| TRUB2 | 3.682969 | 0.367913 | 0.999532 | | 3.314887 | -0.23969 | 0.873938 | | 3.312999 | -0.59101 | 0.614525 |
| MS4A4A | 2.401619 | 0.003019 | 0.999532 | | 2.301912 | -0.29627 | 0.873938 | | 1.944265 | -0.2754 | 0.88476 |
| DPF3 | 2.182649 | 0.852253 | 0.999532 | | 2.029457 | 0.311914 | 0.873938 | | 2.303807 | -0.52448 | 0.718643 |
| METTL7B | 2.923933 | -0.22138 | 0.999532 | | 3.1829 | -0.24248 | 0.873938 | | 2.682433 | -0.00218 | 0.999385 |
| SCX | 4.145791 | 0.415923 | 0.999532 | | 3.572359 | -0.23036 | 0.874012 | | 3.677107 | -0.64026 | 0.566812 |
| ST7 | 2.344473 | 0.194892 | 0.999532 | | 2.562575 | 0.256865 | 0.874012 | | 2.39942 | 0.078199 | 0.972221 |
| TUBB2B | 3.26311 | 0.299659 | 0.999532 | | 3.755581 | 0.224874 | 0.874012 | | 3.343463 | -0.07116 | 0.971092 |
| MOB1A | 8.210951 | -0.13771 | 0.999532 | | 9.944775 | 0.157732 | 0.874012 | | 8.346962 | 0.318641 | 0.717627 |
| TBC1D24 | 2.354155 | 0.208837 | 0.999532 | | 2.256957 | -0.28228 | 0.874012 | | 2.19983 | -0.46927 | 0.760955 |
| FGFRL1 | 3.688444 | 0.552536 | 0.999532 | | 3.467516 | 0.226075 | 0.874012 | | 3.635221 | -0.31975 | 0.803301 |
| PDCD10 | 6.559492 | -0.02214 | 0.999532 | | 7.413373 | -0.16553 | 0.874012 | | 6.035091 | -0.11732 | 0.920716 |
| CNOT6L | 4.592626 | -0.33041 | 0.999532 | | 5.27456 | -0.2048 | 0.874012 | | 3.993208 | 0.143152 | 0.923864 |
| TMCO3 | 4.440071 | -0.02206 | 0.999532 | | 5.488997 | 0.205447 | 0.874012 | | 4.418403 | 0.248771 | 0.842754 |
| TMPPE | 3.28503 | 0.411531 | 0.999532 | | 3.530396 | 0.233549 | 0.874012 | | 3.56214 | -0.15788 | 0.916694 |
| TLR3 | 6.04804 | -0.3341 | 0.999532 | | 8.110208 | 0.173121 | 0.874012 | | 6.395938 | 0.533372 | 0.542134 |
| TREX1 | 2.559173 | 0.709537 | 0.999532 | | 2.076796 | -0.30305 | 0.874012 | | 2.336829 | -0.99481 | 0.397004 |
| EDN2 | 2.146562 | 0.557529 | 0.999532 | | 2.051644 | -0.31841 | 0.874012 | | 2.032497 | -0.84899 | 0.540342 |
| LONP1 | 7.222681 | -0.14374 | 0.999532 | | 9.224451 | 0.172362 | 0.874012 | | 7.217599 | 0.322522 | 0.720106 |
| LYVE1 | 3.783879 | -0.30045 | 0.999532 | | 4.345118 | -0.22935 | 0.874012 | | 3.449682 | 0.076013 | 0.969693 |
| SAMHD1 | 6.596315 | -0.38559 | 0.999532 | | 7.467687 | -0.21459 | 0.874012 | | 5.619613 | 0.189144 | 0.886755 |
| VAC14 | 4.288028 | 0.056511 | 0.999532 | | 4.355992 | -0.2061 | 0.87405 | | 3.934296 | -0.24962 | 0.842754 |
| COL15A1 | 2.372336 | -0.45943 | 0.999532 | | 3.309742 | 0.253915 | 0.874362 | | 2.463445 | 0.733137 | 0.563729 |
| SERP2 | 2.913153 | 0.781608 | 0.999532 | | 2.827646 | 0.267701 | 0.874793 | | 3.069486 | -0.49238 | 0.702463 |
| MMP17 | 2.163219 | -0.01454 | 0.999532 | | 2.251647 | -0.29133 | 0.874793 | | 1.846497 | -0.25791 | 0.893262 |
| BICD1 | 4.345875 | -0.0441 | 0.999532 | | 4.656862 | -0.20117 | 0.874793 | | 3.766542 | -0.13827 | 0.921791 |
| CIAPIN1 | 4.889353 | -0.27282 | 0.999532 | | 5.37808 | -0.20045 | 0.874793 | | 4.346464 | 0.096961 | 0.951381 |
| LYSMD3 | 3.800399 | 0.167273 | 0.999532 | | 4.344492 | 0.231739 | 0.874793 | | 3.84482 | 0.083994 | 0.964153 |
| MSL2 | 3.863234 | -0.45908 | 0.999532 | | 5.105418 | 0.195414 | 0.874793 | | 3.828713 | 0.670376 | 0.499146 |
| RAB33B | 3.485359 | -0.26548 | 0.999532 | | 3.983991 | -0.22505 | 0.874793 | | 3.173995 | 0.057065 | 0.978229 |
| IL13RA2 | 4.071523 | 0.373085 | 0.999532 | | 4.174543 | 0.223129 | 0.874793 | | 4.181045 | -0.121 | 0.938429 |
| SMCO4 | 3.496196 | 0.53664 | 0.999532 | | 2.884928 | -0.25415 | 0.874793 | | 3.092584 | -0.77303 | 0.486743 |
| TMEM212 | 1.620658 | 0.440346 | 0.999532 | | 1.674897 | 0.33078 | 0.874793 | | 1.710668 | -0.09102 | 0.974421 |
| ZNF418 | 3.880112 | -0.18274 | 0.999532 | | 4.459442 | -0.22929 | 0.874793 | | 3.450545 | -0.04116 | 0.98556 |
| AQP4 | 1.974334 | -0.16503 | 0.999532 | | 2.093014 | -0.30612 | 0.874793 | | 1.731121 | -0.12507 | 0.961873 |
| MT2A | 33.27283 | -0.53358 | 0.999532 | | 42.35549 | -0.1499 | 0.874793 | | 29.44939 | 0.410302 | 0.515762 |
| TSSK4 | 2.467934 | -0.01894 | 0.999532 | | 2.439674 | -0.27197 | 0.874793 | | 2.204883 | -0.23793 | 0.896638 |
| FRY | 6.947818 | 0.595497 | 0.999532 | | 6.454858 | 0.189799 | 0.874793 | | 6.755431 | -0.37659 | 0.657538 |
| HMGCS2 | 2.475712 | 0.028101 | 0.999532 | | 3.0867 | 0.246603 | 0.875124 | | 2.526778 | 0.240227 | 0.885078 |
| PLVAP | 3.444868 | -0.63951 | 0.999532 | | 4.720061 | 0.231583 | 0.875443 | | 3.334426 | 0.885865 | 0.413519 |
| ATAD2 | 2.116239 | -0.164 | 0.999532 | | 2.69331 | 0.253001 | 0.875443 | | 2.218966 | 0.434973 | 0.766974 |
| SLC5A10 | 7.376329 | 0.071559 | 0.999532 | | 7.749807 | -0.19094 | 0.875443 | | 6.778926 | -0.24964 | 0.816618 |
| CKB | 7.885619 | -0.16647 | 0.999532 | | 8.850736 | -0.17483 | 0.875443 | | 6.785332 | 0.024007 | 0.989536 |
| SGPL1 | 3.520901 | -0.14297 | 0.999532 | | 3.785809 | -0.23398 | 0.875443 | | 2.957117 | -0.06429 | 0.97502 |
| ARGLU1 | 23.82573 | -0.09015 | 0.999532 | | 28.64048 | 0.145963 | 0.875443 | | 23.75633 | 0.270674 | 0.657617 |
| ACVR1 | 4.427516 | 0.219864 | 0.999532 | | 5.099561 | 0.210531 | 0.875443 | | 4.537434 | 0.010312 | 0.996891 |
| LNP1 | 2.109443 | 0.211379 | 0.999532 | | 2.518115 | 0.277232 | 0.875443 | | 2.271182 | 0.083815 | 0.972221 |
| RGS2 | 6.885179 | -1.25354 | 0.064292 | DOWN | 10.21558 | -0.19758 | 0.875443 | | 6.050173 | 1.081499 | 0.15668 |
| GCNT4 | 3.113931 | -0.42153 | 0.999532 | | 3.711492 | -0.23495 | 0.875501 | | 2.789926 | 0.205346 | 0.901033 |
| SCAF8 | 5.106944 | 0.221826 | 0.999532 | | 5.893948 | 0.187175 | 0.875545 | | 5.399781 | -0.00381 | 0.998586 |
| PRR23B | 1.797664 | 0.314737 | 0.999532 | | 1.633736 | -0.32686 | 0.875545 | | 1.6569 | -0.62385 | 0.712891 |
| ZNF224 | 2.26361 | 0.065283 | 0.999532 | | 2.35889 | -0.27781 | 0.875571 | | 2.11239 | -0.31822 | 0.858767 |
| ZKSCAN | 2.484551 | 0.476765 | 0.999532 | | 2.54832 | 0.264473 | 0.875592 | | 2.581906 | -0.19333 | 0.907882 |
| FILIP1L | 4.062559 | -1.03119 | 0.69276 | | 6.068687 | 0.187523 | 0.875592 | | 3.805381 | 1.235257 | 0.116641 |
| CCDC9B | 2.807999 | -0.17636 | 0.999532 | | 3.188607 | -0.24527 | 0.875592 | | 2.556266 | -0.04872 | 0.985306 |
| CPTP | 8.388436 | 0.193534 | 0.999532 | | 8.148618 | -0.17115 | 0.875592 | | 7.464883 | -0.34547 | 0.692054 |
| SMTNL2 | 3.690145 | 0.361929 | 0.999532 | | 3.789599 | 0.225335 | 0.875592 | | 3.739963 | -0.1167 | 0.94366 |
| OTUD4 | 5.962017 | -0.15785 | 0.999532 | | 6.583167 | -0.1798 | 0.875592 | | 5.341595 | -0.00222 | 0.999385 |
| NRBP2 | 5.257975 | 0.451839 | 0.999532 | | 4.688452 | -0.20071 | 0.875592 | | 4.775143 | -0.63389 | 0.446959 |
| R3HDM4 | 6.254012 | -0.0903 | 0.999532 | | 7.896168 | 0.161065 | 0.875592 | | 6.363614 | 0.271201 | 0.773057 |
| KCTD18 | 2.827977 | 0.464458 | 0.999532 | | 2.460052 | -0.29372 | 0.875592 | | 2.451755 | -0.73449 | 0.557909 |
| EIF5A2 | 2.487036 | 0.308779 | 0.999532 | | 2.631277 | 0.25938 | 0.875644 | | 2.411067 | -0.03867 | 0.989192 |
| CIAO2A | 4.119291 | -0.08044 | 0.999532 | | 4.52096 | -0.20817 | 0.875716 | | 3.754616 | -0.12856 | 0.932589 |
| HCCS | 3.181708 | -0.14931 | 0.999532 | | 4.295062 | 0.22102 | 0.876079 | | 3.36717 | 0.398539 | 0.752562 |
| ACY3 | 2.878243 | -0.10612 | 0.999532 | | 2.928499 | -0.25098 | 0.87644 | | 2.492486 | -0.13721 | 0.945902 |

| | | | | | | | | | | | |
|---|---|---|---|---|---|---|---|---|---|---|---|
| MTUS1 | 4.176006 | -0.48966 | 0.999532 | | 5.0153 | -0.20867 | 0.876661 | | 3.557829 | 0.301406 | 0.814434 |
| SPEF2 | 2.271345 | 0.347377 | 0.999532 | | 2.543356 | 0.268024 | 0.876661 | | 2.428772 | -0.05754 | 0.980581 |
| TENT4A | 4.0913 | -0.08685 | 0.999532 | | 4.352309 | -0.20546 | 0.876664 | | 3.640051 | -0.10115 | 0.951792 |
| TAF1C | 3.719825 | 0.155765 | 0.999532 | | 3.590259 | -0.22663 | 0.876664 | | 3.421974 | -0.3679 | 0.776133 |
| ZNF804A | 1.916635 | -0.35867 | 0.999532 | | 2.222432 | -0.29295 | 0.876664 | | 1.802135 | 0.076368 | 0.977746 |
| EIF1 | 17.36862 | -0.68976 | 0.22007 | | 23.13855 | -0.13534 | 0.876664 | | 15.2509 | 0.595069 | 0.272442 |
| PCTP | 2.745991 | 0.179356 | 0.999532 | | 3.358122 | 0.238338 | 0.876664 | | 2.93425 | 0.088903 | 0.96556 |
| HIRIP3 | 2.650137 | 0.055742 | 0.999532 | | 2.777879 | -0.25845 | 0.876664 | | 2.265313 | -0.29324 | 0.859522 |
| HEATR5B | 2.32967 | -0.28076 | 0.999532 | | 3.120572 | 0.243794 | 0.876664 | | 2.412031 | 0.541362 | 0.695561 |
| KYNU | 2.854258 | -0.03106 | 0.999532 | | 3.070673 | -0.24623 | 0.876833 | | 2.557919 | -0.19552 | 0.907171 |
| GNMT | 2.249723 | 0.580171 | 0.999532 | | 1.831095 | -0.3128 | 0.877259 | | 1.985088 | -0.87571 | 0.536289 |
| AIDA | 8.093469 | -0.41663 | 0.999532 | | 9.862579 | -0.15402 | 0.877332 | | 7.232439 | 0.280695 | 0.754186 |
| CARHSP1 | 6.128298 | -0.11633 | 0.999532 | | 7.593413 | 0.178792 | 0.877515 | | 6.163439 | 0.313359 | 0.759369 |
| ZNF354C | 2.741615 | 0.439735 | 0.999532 | | 2.968547 | 0.24801 | 0.877515 | | 2.734919 | -0.17044 | 0.918694 |
| AK3 | 5.357772 | -0.28007 | 0.999532 | | 5.789235 | -0.19156 | 0.877515 | | 4.471062 | 0.092728 | 0.955167 |
| LIN52 | 2.245555 | -0.00586 | 0.999532 | | 2.763111 | 0.260099 | 0.877583 | | 2.212155 | 0.275285 | 0.874251 |
| CACNB3 | 3.433708 | 0.659692 | 0.999532 | | 2.824804 | -0.25423 | 0.877583 | | 3.112626 | -0.89394 | 0.359441 |
| PPWD1 | 4.967523 | 0.052211 | 0.999532 | | 5.744347 | 0.184456 | 0.8777 | | 4.984227 | 0.151077 | 0.90066 |
| DISP3 | 2.066313 | -0.28376 | 0.999532 | | 2.640534 | 0.265093 | 0.8777 | | 2.060473 | 0.566125 | 0.706407 |
| RSRP1 | 9.409673 | -0.20416 | 0.999532 | | 10.71669 | -0.14156 | 0.8777 | | 8.550643 | 0.079905 | 0.944635 |
| USP6NL | 2.881138 | 0.043473 | 0.999532 | | 3.402711 | 0.232777 | 0.8777 | | 2.99158 | 0.213893 | 0.892646 |
| DOCK2 | 3.87217 | -0.00293 | 0.999532 | | 3.813794 | -0.27411 | 0.8777 | | 3.284677 | -0.24015 | 0.87764 |
| RBM41 | 3.294396 | 0.006036 | 0.999532 | | 3.297196 | -0.23472 | 0.877849 | | 2.918994 | -0.2341 | 0.881837 |
| OR6B1 | 2.003799 | 0.072822 | 0.999532 | | 2.157259 | -0.30601 | 0.877849 | | 1.870992 | -0.35533 | 0.856868 |
| FDCSP | 2.095538 | 0.657343 | 0.999532 | | 1.989465 | 0.306774 | 0.877849 | | 2.179282 | -0.334 | 0.850928 |
| NUBP2 | 5.535201 | 0.175562 | 0.999532 | | 5.341393 | -0.19028 | 0.877849 | | 5.090326 | -0.34843 | 0.725637 |
| PRSS58 | 3.483312 | 0.64642 | 0.999532 | | 2.645243 | -0.26123 | 0.877849 | | 3.015664 | -0.89197 | 0.406014 |
| CIB2 | 2.72174 | 0.494436 | 0.999532 | | 2.885144 | 0.254051 | 0.877849 | | 2.876603 | -0.22168 | 0.889126 |
| KCNA2 | 2.404095 | 0.15103 | 0.999532 | | 2.683363 | -0.27179 | 0.877849 | | 2.236976 | -0.37982 | 0.816112 |
| SMC2 | 2.417165 | 0.06552 | 0.999532 | | 2.416146 | -0.27472 | 0.878235 | | 2.204414 | -0.31825 | 0.85585 |
| LRRN2 | 2.166902 | 0.150199 | 0.999532 | | 2.130782 | -0.28605 | 0.878287 | | 1.92253 | -0.4156 | 0.805466 |
| CCPG1 | 2.687419 | 0.561083 | 0.999532 | | 2.565477 | 0.270457 | 0.878287 | | 2.668989 | -0.28651 | 0.863134 |
| PSMD12 | 7.103981 | 0.222683 | 0.999532 | | 7.836019 | 0.161736 | 0.878287 | | 7.363973 | -0.04561 | 0.974331 |
| AHNAK | 74.88627 | 0.016925 | 0.999532 | | 86.68174 | 0.107653 | 0.878287 | | 74.20124 | 0.114609 | 0.863792 |
| DGCR6L | 3.652627 | 0.028816 | 0.999532 | | 3.714123 | -0.22312 | 0.878287 | | 3.199786 | -0.23371 | 0.87591 |
| TADA2A | 2.875875 | 0.6622 | 0.999532 | | 2.130615 | -0.27979 | 0.878287 | | 2.541212 | -0.92884 | 0.421517 |
| SLC6A1 | 2.252483 | 0.045899 | 0.999532 | | 2.459889 | -0.27765 | 0.878287 | | 2.084178 | -0.30895 | 0.861119 |
| KCTD4 | 2.785342 | 0.351881 | 0.999532 | | 2.596989 | -0.26522 | 0.878287 | | 2.593899 | -0.6015 | 0.652925 |
| PLEKHF1 | 2.090767 | 0.409532 | 0.999532 | | 1.698508 | -0.3168 | 0.878287 | | 1.866502 | -0.72009 | 0.657538 |
| ATXN7L3 | 5.532874 | 0.033136 | 0.999532 | | 6.764799 | 0.170538 | 0.878287 | | 5.455995 | 0.157521 | 0.900583 |
| HBA2 | 3.950104 | -0.02225 | 0.999532 | | 5.495533 | 0.388066 | 0.878287 | | 4.442119 | 0.523086 | 0.816112 |
| NOD2 | 2.09298 | 0.253686 | 0.999532 | | 2.069795 | -0.30449 | 0.878287 | | 1.947545 | -0.54185 | 0.737109 |
| GKN1 | 1.959537 | 0.053632 | 0.999532 | | 1.956263 | -0.29588 | 0.878287 | | 1.650196 | -0.33236 | 0.862823 |
| TSACC | 2.210181 | 0.137937 | 0.999532 | | 2.280218 | -0.29587 | 0.878287 | | 2.127062 | -0.41438 | 0.816618 |
| GPI | 4.494321 | 0.366993 | 0.999532 | | 4.877393 | 0.201419 | 0.878287 | | 4.275031 | -0.15384 | 0.908891 |
| VSIR | 4.883482 | -0.12117 | 0.999532 | | 5.859441 | 0.205515 | 0.878287 | | 4.909712 | 0.349401 | 0.75311 |
| TAS2R3 | 2.659049 | 0.440002 | 0.999532 | | 2.726076 | 0.25703 | 0.878287 | | 2.691991 | -0.16434 | 0.922922 |
| SLC47A2 | 3.226958 | 0.030318 | 0.999532 | | 3.672392 | 0.227274 | 0.878287 | | 3.177762 | 0.212027 | 0.890225 |
| CD300LG | 2.376065 | 0.589453 | 0.999532 | | 2.369211 | 0.28364 | 0.878287 | | 2.519833 | -0.2924 | 0.872435 |
| GPR161 | 2.857551 | 0.186148 | 0.999532 | | 2.852756 | -0.26481 | 0.878287 | | 2.474614 | -0.42841 | 0.764702 |
| RGS9 | 3.818828 | 0.479835 | 0.999532 | | 3.550429 | 0.224956 | 0.878287 | | 3.826697 | -0.23377 | 0.869378 |
| SSPOP | 2.982587 | 0.476904 | 0.999532 | | 2.590482 | -0.2591 | 0.878287 | | 2.643711 | -0.71851 | 0.540342 |
| DHRS12 | 3.427528 | 0.349834 | 0.999532 | | 3.072255 | -0.25365 | 0.878287 | | 2.881504 | -0.58027 | 0.644433 |
| UFSP2 | 7.555156 | -0.13338 | 0.999532 | | 8.64743 | -0.16475 | 0.878287 | | 6.826181 | -0.01798 | 0.992286 |
| RBPMS2 | 2.543372 | -0.01756 | 0.999532 | | 2.96377 | 0.24288 | 0.878287 | | 2.762256 | 0.267062 | 0.876495 |
| NTSR2 | 2.03737 | -0.07516 | 0.999532 | | 2.179514 | -0.28542 | 0.878287 | | 1.824564 | -0.19067 | 0.92697 |
| RABGAP1 | 3.271569 | -0.38163 | 0.999532 | | 3.872061 | -0.21904 | 0.878287 | | 2.89885 | 0.182148 | 0.909084 |
| PLGRKT | 2.760874 | 0.223087 | 0.999532 | | 3.162773 | 0.242012 | 0.878287 | | 2.923208 | 0.035745 | 0.989278 |
| RFC4 | 2.158556 | 0.242412 | 0.999532 | | 2.099467 | -0.30252 | 0.878287 | | 1.904409 | -0.5246 | 0.742782 |
| WDR48 | 2.787152 | -0.07691 | 0.999532 | | 2.955375 | -0.24332 | 0.878287 | | 2.533438 | -0.14552 | 0.939942 |
| SMIM19 | 4.235786 | -0.60932 | 0.999532 | | 5.58054 | 0.200112 | 0.878287 | | 3.969439 | 0.807433 | 0.420623 |
| TREM1 | 2.1113 | -0.40144 | 0.999532 | | 2.322306 | -0.27507 | 0.878287 | | 1.814522 | 0.136708 | 0.955167 |
| ATP5MC2 | 57.62574 | -0.07776 | 0.999532 | | 69.77538 | 0.146567 | 0.878287 | | 57.80019 | 0.239851 | 0.816589 |
| CAVIN3 | 5.197752 | -0.29667 | 0.999532 | | 7.217225 | 0.175236 | 0.878287 | | 5.44709 | 0.472566 | 0.598934 |
| CEP83 | 2.421419 | 0.385172 | 0.999532 | | 2.509846 | 0.260823 | 0.878287 | | 2.614575 | -0.07461 | 0.974421 |
| CFAP45 | 2.075674 | -0.02739 | 0.999532 | | 1.95502 | -0.30306 | 0.878287 | | 1.781802 | -0.26515 | 0.90066 |
| ITGA2 | 3.618985 | 0.067567 | 0.999532 | | 4.261575 | 0.209756 | 0.878287 | | 3.532987 | 0.158035 | 0.912439 |
| P3R3URF | 3.271487 | -0.26947 | 0.999532 | | 3.815651 | -0.24895 | 0.878287 | | 3.081659 | 0.024279 | 0.994458 |
| ZNF490 | 2.911635 | 0.001072 | 0.999532 | | 3.199864 | -0.2375 | 0.878287 | | 2.632157 | -0.21341 | 0.894612 |
| TMEM214 | 5.368041 | -0.15889 | 0.999532 | | 6.360523 | 0.178457 | 0.878287 | | 5.377712 | 0.344134 | 0.746876 |
| RBBP4 | 7.269148 | -0.17139 | 0.999532 | | 8.424954 | -0.15397 | 0.878287 | | 6.458068 | 0.036339 | 0.980581 |
| PPP1R26 | 2.932276 | -0.18239 | 0.999532 | | 3.282727 | -0.24063 | 0.878287 | | 2.604177 | -0.03777 | 0.989536 |
| NTN4 | 10.98489 | -0.47061 | 0.999532 | | 15.33775 | 0.141208 | 0.878287 | | 11.08237 | 0.629662 | 0.266933 |
| LTB4R2 | 3.188069 | -0.13604 | 0.999532 | | 3.54113 | -0.23751 | 0.878287 | | 3.002793 | -0.0864 | 0.965649 |
| TK2 | 4.871065 | -0.27237 | 0.999532 | | 5.476397 | -0.18998 | 0.878287 | | 4.413366 | 0.102255 | 0.946765 |
| OR51H1 | 2.07334 | 0.422405 | 0.999532 | | 1.855064 | -0.31904 | 0.878287 | | 1.878933 | -0.7242 | 0.647218 |
| SLC16A1 | 2.746616 | -0.18615 | 0.999532 | | 3.066776 | -0.24613 | 0.878287 | | 2.338047 | -0.04322 | 0.987405 |
| AGBL2 | 3.173976 | 0.477179 | 0.999532 | | 2.759286 | -0.25622 | 0.878287 | | 2.820727 | -0.71791 | 0.549172 |
| BROX | 3.943824 | -0.55728 | 0.999532 | | 5.472617 | 0.188927 | 0.878287 | | 4.017178 | 0.763578 | 0.413122 |
| CAPSL | 1.977083 | 0.542988 | 0.999532 | | 1.959843 | 0.303608 | 0.878287 | | 2.058867 | -0.22053 | 0.908829 |
| ZNF189 | 3.874328 | -0.05212 | 0.999532 | | 4.60334 | 0.208427 | 0.878287 | | 3.788723 | 0.275675 | 0.836453 |
| TMC4 | 3.343486 | 0.143215 | 0.999532 | | 3.363223 | -0.23387 | 0.878287 | | 3.008269 | -0.35268 | 0.816589 |
| BTBD2 | 4.621724 | -0.02088 | 0.999532 | | 5.609572 | 0.188461 | 0.878287 | | 4.55655 | 0.228887 | 0.849848 |
| BCL6B | 2.797794 | -0.36248 | 0.999532 | | 3.694146 | 0.230809 | 0.878287 | | 2.793803 | 0.615486 | 0.628053 |
| CCDC77 | 2.416683 | 0.417311 | 0.999532 | | 2.060449 | -0.30282 | 0.878287 | | 2.070284 | -0.70131 | 0.636525 |
| ATF1 | 6.505286 | 0.003959 | 0.999532 | | 7.589515 | 0.176209 | 0.878287 | | 6.643793 | 0.181374 | 0.880949 |
| CEP68 | 4.441433 | -0.04338 | 0.999532 | | 5.5436 | 0.194977 | 0.878287 | | 4.579155 | 0.2572 | 0.835289 |
| NCF4 | 2.731627 | 0.293579 | 0.999532 | | 2.423272 | -0.27135 | 0.878287 | | 2.509244 | -0.55239 | 0.706407 |
| LLGL2 | 3.21923 | 0.134282 | 0.999532 | | 3.372801 | -0.23566 | 0.878287 | | 2.906884 | -0.34639 | 0.800724 |
| IL25 | 2.309351 | 0.553482 | 0.999532 | | 1.754997 | -0.31686 | 0.878287 | | 2.062161 | -0.85014 | 0.563885 |
| OR1D5 | 2.308867 | 0.172728 | 0.999532 | | 2.710145 | 0.276471 | 0.878287 | | 2.29889 | 0.123904 | 0.952713 |
| ZNF462 | 2.380988 | 0.213013 | 0.999532 | | 2.664347 | 0.258373 | 0.878287 | | 2.367941 | 0.061807 | 0.978312 |
| CT47A10 | 3.603535 | -0.48399 | 0.999532 | | 4.44234 | -0.22618 | 0.878287 | | 3.326807 | 0.279406 | 0.859255 |
| BCAS3 | 4.48025 | 0.070009 | 0.999532 | | 4.205509 | -0.22051 | 0.878312 | | 3.923449 | -0.28217 | 0.836453 |
| PPAN | 3.14723 | 0.246215 | 0.999532 | | 2.964202 | -0.26148 | 0.878312 | | 2.848531 | -0.4915 | 0.729001 |
| RNF114 | 2.455878 | -0.17372 | 0.999532 | | 3.311748 | 0.242489 | 0.878312 | | 2.548329 | 0.444397 | 0.75311 |
| METTL21 | 2.474025 | 0.801288 | 0.999532 | | 2.279893 | 0.280896 | 0.878355 | | 2.570723 | -0.50255 | 0.717138 |
| ALG10 | 3.387815 | -0.34012 | 0.999532 | | 4.002726 | -0.23823 | 0.878599 | | 3.188813 | 0.125349 | 0.952656 |
| MUL1 | 3.286084 | 0.178602 | 0.999532 | | 2.904573 | -0.2367 | 0.878648 | | 2.871892 | -0.40332 | 0.76302 |
| RAC3 | 8.359117 | 0.060281 | 0.999532 | | 8.827834 | -0.18068 | 0.878835 | | 7.64202 | -0.21834 | 0.847148 |
| ZNF165 | 2.284566 | 0.525004 | 0.999532 | | 2.031078 | -0.30132 | 0.878882 | | 2.040577 | -0.80644 | 0.563729 |
| DCTN1 | 6.210967 | -0.05775 | 0.999532 | | 7.46285 | 0.164027 | 0.878882 | | 6.070435 | 0.239253 | 0.824012 |
| UBFD1 | 3.718119 | -0.42316 | 0.999532 | | 4.42867 | -0.20583 | 0.87889 | | 3.321149 | 0.234461 | 0.871072 |
| KRT20 | 2.561475 | 0.060403 | 0.999532 | | 2.693882 | -0.26162 | 0.87898 | | 2.395603 | -0.29745 | 0.867009 |
| TMED10 | 6.448989 | -0.09209 | 0.999532 | | 7.872503 | 0.164673 | 0.87898 | | 6.497587 | 0.276819 | 0.790999 |
| OXER1 | 3.669813 | 0.189544 | 0.999532 | | 3.524267 | -0.22722 | 0.87898 | | 3.34358 | -0.38863 | 0.806394 |
| MED31 | 2.181545 | 0.04203 | 0.999532 | | 2.219558 | -0.28101 | 0.87898 | | 2.009184 | -0.30076 | 0.872435 |
| TMEM225 | 2.267532 | 0.623129 | 0.999532 | | 1.858439 | -0.31531 | 0.879108 | | 2.036811 | -0.92375 | 0.510412 |
| EIF1B | 5.064432 | -0.32686 | 0.999532 | | 6.784931 | 0.177611 | 0.879116 | | 5.061134 | 0.533417 | 0.579993 |
| RBP7 | 2.192617 | -0.02225 | 0.999532 | | 2.309717 | -0.27391 | 0.879116 | | 1.928377 | -0.23156 | 0.90275 |
| BLK | 2.447719 | -0.30841 | 0.999532 | | 2.613294 | -0.26342 | 0.879116 | | 2.222711 | 0.066136 | 0.978931 |
| CETN2 | 2.812192 | -0.07733 | 0.999532 | | 3.491757 | 0.235412 | 0.879116 | | 2.795085 | 0.333448 | 0.832386 |
| SHF | 1.757047 | 0.295746 | 0.999532 | | 1.929827 | 0.291522 | 0.879116 | | 1.864583 | 0.010991 | 0.996891 |
| RPTN | 1.903019 | -0.23568 | 0.999532 | | 2.152571 | -0.29424 | 0.879116 | | 1.777891 | -0.03762 | 0.992286 |
| CASP10 | 4.875568 | 0.080234 | 0.999532 | | 5.345421 | 0.197763 | 0.879116 | | 4.91691 | 0.167027 | 0.905952 |
| CCDC116 | 2.134381 | 0.04982 | 0.999532 | | 2.10795 | -0.28403 | 0.879116 | | 2.002808 | -0.32051 | 0.864245 |
| TCP1 | 4.727435 | 0.281179 | 0.999532 | | 5.11258 | 0.19567 | 0.879116 | | 4.753577 | -0.06612 | 0.967 |
| RUNDC3A | 2.317372 | 0.103316 | 0.999532 | | 2.144707 | -0.29028 | 0.879116 | | 2.114261 | -0.37867 | 0.839908 |
| CHD9 | 2.432608 | -0.00934 | 0.999532 | | 2.873424 | 0.239163 | 0.879116 | | 2.434904 | 0.26378 | 0.871713 |
| SPINK1 | 2.460583 | 0.331679 | 0.999532 | | 2.164715 | -0.27761 | 0.879116 | | 2.120238 | -0.59565 | 0.678411 |
| ATG101 | 2.925568 | -0.31735 | 0.999532 | | 3.402162 | -0.237 | 0.879116 | | 2.572436 | 0.102864 | 0.96129 |
| RBPMS | 14.82402 | -0.01052 | 0.999532 | | 17.61942 | 0.125559 | 0.879116 | | 14.73141 | 0.159478 | 0.839622 |
| NBPF26 | 26.64113 | 0.281195 | 0.999532 | | 28.29389 | 0.11545 | 0.879116 | | 26.37139 | -0.13945 | 0.858705 |
| RASAL2 | 5.929419 | 0.114047 | 0.999532 | | 6.500752 | 0.187711 | 0.879116 | | 5.767708 | 0.103749 | 0.939549 |
| ROM1 | 2.040113 | 0.321025 | 0.999532 | | 2.008255 | -0.30066 | 0.879116 | | 1.802412 | -0.59967 | 0.702463 |
| APLP1 | 6.084443 | -0.05039 | 0.999532 | | 7.343829 | 0.166307 | 0.879116 | | 6.289294 | 0.24018 | 0.808547 |
| APBB1 | 3.019051 | -0.09227 | 0.999532 | | 3.74035 | 0.220685 | 0.879116 | | 3.084528 | 0.330563 | 0.809485 |
| NR2C1 | 5.160123 | -0.35997 | 0.999532 | | 6.273522 | -0.17386 | 0.879116 | | 4.634598 | 0.20602 | 0.864347 |
| GSTT2 | 6.062708 | -0.19528 | 0.999532 | | 8.102867 | 0.179257 | 0.879116 | | 6.385278 | 0.378442 | 0.668532 |
| WDPCP | 2.375529 | 0.228266 | 0.999532 | | 2.661447 | 0.25526 | 0.879116 | | 2.490023 | 0.045802 | 0.986366 |
| SYNC | 1.972934 | -0.39824 | 0.999532 | | 2.154954 | -0.28366 | 0.879116 | | 1.754537 | 0.13289 | 0.958191 |
| SMPD4 | 3.9138 | -0.02416 | 0.999532 | | 4.295358 | -0.20336 | 0.879116 | | 3.540775 | -0.16144 | 0.908953 |
| FRMD3 | 8.25448 | 0.030636 | 0.999532 | | 9.680222 | 0.157081 | 0.879116 | | 8.454485 | 0.148258 | 0.887555 |
| UGT3A2 | 2.475289 | 0.467906 | 0.999532 | | 2.310005 | 0.264126 | 0.879116 | | 2.540449 | -0.18952 | 0.915095 |
| DCBLD2 | 4.446339 | -0.19221 | 0.999532 | | 4.882385 | -0.19392 | 0.879116 | | 4.041864 | 0.016344 | 0.995943 |
| B4GALT4 | 2.859911 | 0.030041 | 0.999532 | | 3.364505 | 0.219965 | 0.879116 | | 2.979288 | 0.208602 | 0.889853 |
| CAND1 | 7.021377 | 0.132952 | 0.999532 | | 7.638853 | 0.161018 | 0.879116 | | 6.843727 | 0.034063 | 0.984624 |
| COASY | 4.934303 | 0.027146 | 0.999532 | | 5.228356 | -0.1977 | 0.879116 | | 4.292858 | -0.19928 | 0.87831 |
| EFCAB7 | 2.064711 | 1.051349 | 0.999532 | | 1.68827 | 0.315769 | 0.879116 | | 2.137398 | -0.71985 | 0.620745 |
| YIF1B | 4.23179 | 0.343904 | 0.999532 | | 4.725429 | 0.199352 | 0.879116 | | 4.337033 | -0.12331 | 0.929529 |
| TPCN2 | 4.369035 | 0.101313 | 0.999532 | | 4.561385 | -0.19984 | 0.879116 | | 4.048334 | -0.28272 | 0.811123 |
| LGALSL | 3.167769 | -0.12814 | 0.999532 | | 3.631502 | -0.23616 | 0.879116 | | 3.024785 | -0.08049 | 0.971732 |
| SAPCD1 | 2.473455 | 0.296369 | 0.999532 | | 2.609966 | 0.248852 | 0.879116 | | 2.601837 | -0.03107 | 0.991349 |
| SH2B1 | 4.129338 | 0.101661 | 0.999532 | | 4.819274 | 0.205345 | 0.879116 | | 4.266673 | 0.125617 | 0.938429 |
| FOXO4 | 3.112148 | -0.07322 | 0.999532 | | 4.018858 | 0.210971 | 0.879116 | | 3.251949 | 0.303803 | 0.818726 |
| CCDC27 | 2.440883 | 0.45619 | 0.999532 | | 2.057532 | -0.28912 | 0.879116 | | 2.221412 | -0.72486 | 0.606305 |
| THEMIS2 | 3.085638 | -0.17335 | 0.999532 | | 3.489343 | -0.23392 | 0.879116 | | 2.886372 | -0.05096 | 0.982124 |
| ALKBH3 | 2.244797 | 0.032814 | 0.999532 | | 2.683271 | 0.261991 | 0.879116 | | 2.297756 | 0.248674 | 0.886495 |
| SYTL2 | 2.813508 | -0.29818 | 0.999532 | | 3.407912 | 0.236882 | 0.879116 | | 2.796105 | 0.548327 | 0.679452 |
| NCBP2AS | 2.822688 | 0.426842 | 0.999532 | | 3.180129 | 0.234243 | 0.879116 | | 2.906504 | -0.16857 | 0.916969 |
| KALRN | 2.78875 | 0.398098 | 0.999532 | | 2.358771 | -0.28071 | 0.879116 | | 2.398232 | -0.66303 | 0.639848 |
| ENPP7 | 2.165696 | -0.42471 | 0.999532 | | 2.417592 | -0.27297 | 0.879116 | | 1.850443 | 0.16541 | 0.941973 |
| CRIPT | 3.750863 | -0.37867 | 0.999532 | | 4.905782 | 0.194642 | 0.879116 | | 3.838088 | 0.586803 | 0.569849 |
| FKBP11 | 3.792061 | 0.203571 | 0.999532 | | 3.807306 | -0.22612 | 0.879116 | | 3.33478 | -0.41076 | 0.730921 |
| SAE1 | 6.024712 | -0.21795 | 0.999532 | | 6.682451 | -0.18662 | 0.879116 | | 5.472126 | 0.054353 | 0.973931 |
| WIZ | 4.727751 | -0.24351 | 0.999532 | | 6.034683 | 0.17942 | 0.879116 | | 4.714757 | 0.439509 | 0.659545 |
| SLURP1 | 4.094059 | 0.223516 | 0.999532 | | 4.092138 | -0.23449 | 0.879116 | | 3.808619 | -0.44295 | 0.717627 |
| GH1 | 2.180542 | -0.23957 | 0.999532 | | 2.556131 | -0.27107 | 0.879116 | | 1.983697 | -0.00772 | 0.997967 |
| PPIAL4A | 9.968417 | -0.39836 | 0.999532 | | 12.27379 | -0.13637 | 0.879116 | | 8.851507 | 0.281227 | 0.735469 |
| SHB | 4.893976 | -0.28654 | 0.999532 | | 6.230474 | 0.191302 | 0.879116 | | 4.725773 | 0.508884 | 0.590692 |
| CMTM4 | 3.166166 | 0.248401 | 0.999532 | | 3.30527 | 0.237496 | 0.879116 | | 2.926142 | 0.010384 | 0.996891 |
| ZBED6 | 2.406308 | 0.046622 | 0.999532 | | 2.327586 | -0.26971 | 0.879116 | | 2.142527 | -0.30314 | 0.863401 |
| LATS2 | 6.776629 | -0.10485 | 0.999532 | | 8.165168 | 0.162087 | 0.879376 | | 6.800792 | 0.285152 | 0.764702 |
| MSLN | 2.899082 | 0.488549 | 0.999532 | | 3.015239 | 0.268782 | 0.879376 | | 3.023365 | -0.2007 | 0.906698 |
| AP4B1 | 2.265336 | 0.161802 | 0.999532 | | 2.561881 | 0.253559 | 0.879376 | | 2.400081 | 0.109777 | 0.958191 |
| TREH | 2.151809 | 0.248069 | 0.999532 | | 2.447823 | 0.26429 | 0.879376 | | 2.287428 | 0.036521 | 0.991103 |
| TCIRG1 | 4.432003 | 0.220546 | 0.999532 | | 4.789021 | 0.191575 | 0.879644 | | 4.419855 | -0.01304 | 0.996681 |
| USP46 | 3.924897 | 0.100786 | 0.999532 | | 3.871187 | -0.21796 | 0.879644 | | 3.442986 | -0.30218 | 0.816618 |
| SAA2 | 3.600728 | 0.007644 | 0.999532 | | 3.891087 | -0.2179 | 0.879644 | | 3.3519 | -0.2061 | 0.88992 |
| CNOT4 | 5.763894 | -0.06952 | 0.999532 | | 6.788881 | 0.162413 | 0.879644 | | 5.883266 | 0.244866 | 0.811552 |
| ZNF813 | 3.342836 | 0.384079 | 0.999532 | | 2.894577 | -0.26578 | 0.879644 | | 2.828021 | -0.6291 | 0.632847 |
| SPATA3 | 2.493748 | 0.456725 | 0.999532 | | 2.145785 | -0.29507 | 0.879644 | | 2.28139 | -0.74381 | 0.619423 |
| TMEM269 | 2.109958 | 0.373106 | 0.999532 | | 1.97115 | -0.29704 | 0.879644 | | 2.013567 | -0.65338 | 0.664512 |
| METTL6 | 3.213049 | -0.08344 | 0.999532 | | 4.159509 | 0.210686 | 0.879644 | | 3.398971 | 0.314102 | 0.809832 |
| GANC | 3.441922 | -0.18611 | 0.999532 | | 4.616376 | 0.199942 | 0.879683 | | 3.676499 | 0.413868 | 0.734538 |
| PHLDA1 | 4.057603 | -0.88204 | 0.999532 | | 5.526549 | -0.18528 | 0.879811 | | 3.54945 | 0.712474 | 0.49299 |
| SOHLH1 | 2.434634 | 0.03384 | 0.999532 | | 2.463969 | -0.28214 | 0.879985 | | 2.34077 | -0.31281 | 0.866245 |
| LTB4R | 2.893956 | 0.015838 | 0.999532 | | 2.727399 | -0.24575 | 0.879985 | | 2.544789 | -0.25026 | 0.881837 |
| AIG1 | 4.118028 | -0.28088 | 0.999532 | | 4.628271 | -0.20987 | 0.879985 | | 3.411098 | 0.091959 | 0.958991 |
| SLC10A3 | 3.72773 | -0.31101 | 0.999532 | | 4.901176 | 0.203187 | 0.879985 | | 4.045342 | 0.523109 | 0.652925 |
| EED | 4.555391 | -0.22004 | 0.999532 | | 5.364234 | -0.19717 | 0.879985 | | 4.134452 | 0.052295 | 0.978282 |
| BLOC1S1 | 6.902386 | -0.23453 | 0.999532 | | 8.897924 | 0.168781 | 0.88005 | | 6.922414 | 0.427741 | 0.647218 |
| UCHL5 | 4.087527 | 0.266301 | 0.999532 | | 3.852118 | -0.20861 | 0.88005 | | 3.660847 | -0.45604 | 0.670353 |
| SERPINA | 2.605267 | 0.464119 | 0.999532 | | 2.195553 | -0.28031 | 0.88005 | | 2.396261 | -0.72587 | 0.599988 |
| NCAPH | 3.204513 | 0.901003 | 0.999532 | | 2.22548 | -0.28107 | 0.88005 | | 2.93064 | -1.16697 | 0.332846 |
| GRIK3 | 1.784223 | 0.415656 | 0.999532 | | 1.819138 | 0.29642 | 0.88005 | | 1.839507 | -0.10148 | 0.967145 |
| NPHP1 | 2.347153 | -0.41625 | 0.999532 | | 2.824949 | -0.25559 | 0.88005 | | 2.24674 | 0.171846 | 0.931619 |
| MLH3 | 3.26632 | -0.0532 | 0.999532 | | 3.296997 | -0.22692 | 0.88005 | | 2.908323 | -0.16029 | 0.925229 |
| YLPM1 | 4.24254 | -0.13119 | 0.999532 | | 4.54474 | -0.20395 | 0.88005 | | 3.710802 | -0.04984 | 0.978229 |
| FAM184B | 2.900393 | -0.0751 | 0.999532 | | 3.232686 | -0.2392 | 0.88005 | | 2.556632 | -0.14037 | 0.940744 |
| MAGOHB | 3.438805 | 0.187288 | 0.999532 | | 3.267306 | -0.22753 | 0.880118 | | 3.03784 | -0.39697 | 0.758124 |
| CAMTA1 | 5.121455 | -0.67603 | 0.999532 | | 7.547994 | 0.158103 | 0.880118 | | 5.130279 | 0.851584 | 0.246926 |
| TMEM71 | 2.375941 | -0.39794 | 0.999532 | | 2.689056 | -0.25791 | 0.880118 | | 2.165631 | 0.157998 | 0.941197 |
| FBXO3 | 5.094281 | -0.23563 | 0.999532 | | 6.622596 | 0.170804 | 0.880118 | | 5.026268 | 0.422188 | 0.678541 |
| TBC1D14 | 3.103081 | -0.16223 | 0.999532 | | 4.152492 | 0.219048 | 0.880118 | | 3.312227 | 0.397386 | 0.750371 |
| RCOR1 | 3.474887 | 0.414734 | 0.999532 | | 3.710838 | 0.222973 | 0.880118 | | 3.557279 | -0.17209 | 0.905746 |
| INSL3 | 2.729021 | -0.1267 | 0.999532 | | 3.005429 | -0.25496 | 0.880144 | | 2.649176 | -0.08956 | 0.972221 |
| CPEB3 | 7.610352 | 0.140575 | 0.999532 | | 7.768353 | -0.16375 | 0.880144 | | 7.144589 | -0.29066 | 0.757722 |
| ZNF471 | 2.70361 | 0.000444 | 0.999532 | | 3.474776 | 0.237958 | 0.880144 | | 2.749822 | 0.246433 | 0.876679 |
| CYTH2 | 7.601458 | -0.36999 | 0.999532 | | 8.895276 | -0.15008 | 0.880144 | | 6.786376 | 0.241155 | 0.805436 |
| PRKAB2 | 3.779703 | 0.062503 | 0.999532 | | 4.570195 | 0.195896 | 0.880144 | | 4.087708 | 0.154614 | 0.911296 |
| UPF2 | 4.741753 | 0.452694 | 0.999532 | | 4.572759 | 0.196564 | 0.880144 | | 4.792603 | -0.25453 | 0.843043 |
| PEX14 | 2.2982 | -0.07177 | 0.999532 | | 2.772809 | 0.248416 | 0.880144 | | 2.454381 | 0.33733 | 0.834173 |
| CPAMD8 | 2.578487 | 0.404222 | 0.999532 | | 2.280291 | -0.26846 | 0.880144 | | 2.417847 | -0.65222 | 0.641721 |
| NOS2 | 2.143305 | -0.09507 | 0.999532 | | 2.188249 | -0.27776 | 0.880144 | | 1.976104 | -0.1588 | 0.941973 |
| CRYAA | 2.609684 | 0.16778 | 0.999532 | | 2.744003 | 0.249022 | 0.880144 | | 2.670198 | 0.083273 | 0.972221 |
| CMC1 | 5.253787 | 0.154328 | 0.999532 | | 5.157699 | -0.18561 | 0.880144 | | 4.734191 | -0.32489 | 0.759211 |
| FGF9 | 2.214403 | 0.498727 | 0.999532 | | 2.385514 | 0.278124 | 0.880144 | | 2.33655 | -0.20184 | 0.910105 |
| DYNLRB2 | 2.613052 | 0.183005 | 0.999532 | | 2.459036 | -0.26574 | 0.880144 | | 2.329963 | -0.43802 | 0.784955 |
| PTCD2 | 1.953749 | -0.35355 | 0.999532 | | 2.351992 | -0.26442 | 0.880144 | | 1.83693 | 0.109686 | 0.964434 |
| PHTF2 | 4.116006 | -0.44042 | 0.999532 | | 5.756932 | 0.185966 | 0.880144 | | 4.124022 | 0.646266 | 0.495026 |
| MORC2 | 3.425694 | -0.07207 | 0.999532 | | 4.075164 | 0.206154 | 0.880324 | | 3.535321 | 0.291635 | 0.825181 |
| PNP | 3.624447 | -0.30686 | 0.999532 | | 4.667839 | 0.19667 | 0.880324 | | 3.66019 | 0.514911 | 0.649779 |
| BCAP29 | 3.460199 | 0.060107 | 0.999532 | | 3.633447 | 0.213401 | 0.880324 | | 3.454405 | 0.14582 | 0.92968 |
| GINS1 | 2.100148 | 0.674849 | 0.999532 | | 1.993897 | 0.284259 | 0.880324 | | 2.148592 | -0.37407 | 0.814483 |
| HOXB6 | 4.541678 | 0.165977 | 0.999532 | | 4.578689 | -0.19588 | 0.880448 | | 4.104066 | -0.34261 | 0.757722 |
| NPEPL1 | 2.346892 | -0.19248 | 0.999532 | | 2.958189 | 0.243767 | 0.880448 | | 2.521945 | 0.435045 | 0.78258 |
| ZNF620 | 1.960759 | 0.026989 | 0.999532 | | 2.105428 | -0.29852 | 0.880581 | | 1.690527 | -0.3024 | 0.881837 |
| PRELID3 | 11.20918 | 0.071319 | 0.999532 | | 12.83908 | 0.139099 | 0.880581 | | 11.24737 | 0.086973 | 0.927433 |
| PHB2 | 4.844793 | -0.18915 | 0.999532 | | 6.166553 | 0.184849 | 0.880619 | | 4.928147 | 0.384015 | 0.734904 |
| SEC23A | 3.476598 | -0.16536 | 0.999532 | | 4.553241 | 0.200745 | 0.880716 | | 3.729887 | 0.386314 | 0.746882 |
| PRKAR2A | 4.300059 | -0.04415 | 0.999532 | | 5.189249 | 0.195525 | 0.880716 | | 4.427049 | 0.262921 | 0.824012 |
| MTX1 | 5.59279 | 0.189908 | 0.999532 | | 5.529701 | -0.18834 | 0.880768 | | 5.282882 | -0.36218 | 0.742782 |
| ZNF277 | 2.825529 | -0.33403 | 0.999532 | | 2.991664 | -0.23321 | 0.88084 | | 2.445968 | 0.111163 | 0.959377 |
| RMC1 | 2.376563 | -0.29479 | 0.999532 | | 2.673711 | -0.25159 | 0.881152 | | 2.042846 | 0.063903 | 0.979239 |
| GABRE | 2.517083 | 0.026342 | 0.999532 | | 2.935514 | 0.2567 | 0.881186 | | 2.474049 | 0.251019 | 0.88849 |
| ZNF43 | 2.86113 | -0.20199 | 0.999532 | | 2.994397 | -0.23471 | 0.881186 | | 2.530639 | -0.01323 | 0.996891 |
| KLRB1 | 1.881978 | -0.28434 | 0.999532 | | 2.053924 | -0.30147 | 0.881191 | | 1.641586 | 0.00419 | 0.999385 |
| DGLUCY | 3.470052 | 0.035979 | 0.999532 | | 4.244547 | 0.209354 | 0.881571 | | 3.593298 | 0.194532 | 0.892869 |
| TST | 2.436087 | -0.13012 | 0.999532 | | 3.376276 | 0.231116 | 0.881571 | | 2.72544 | 0.382948 | 0.791576 |
| PVRIG | 3.540408 | 0.200562 | 0.999532 | | 3.579503 | -0.23184 | 0.881571 | | 3.059813 | -0.41229 | 0.742839 |
| FXYD5 | 6.346202 | -0.16034 | 0.999532 | | 8.129275 | 0.238864 | 0.881862 | | 6.734311 | 0.39676 | 0.749588 |
| SRFBP1 | 4.652651 | 0.611828 | 0.999532 | | 4.482185 | 0.205704 | 0.881862 | | 4.645679 | -0.38962 | 0.686214 |
| ABCD3 | 2.838754 | 0.419393 | 0.999532 | | 3.0953 | 0.230422 | 0.881862 | | 2.93026 | -0.1701 | 0.914409 |
| TLX2 | 2.439651 | -0.06216 | 0.999532 | | 2.631664 | -0.2775 | 0.881862 | | 2.286684 | -0.1986 | 0.920655 |
| SLC16A4 | 2.840353 | -0.06727 | 0.999532 | | 3.528758 | 0.217186 | 0.881862 | | 2.835098 | 0.302362 | 0.829716 |
| INPP5E | 2.948968 | 0.175955 | 0.999532 | | 2.883464 | -0.26148 | 0.881862 | | 2.593795 | -0.41498 | 0.791576 |
| MRLN | 1.986018 | 0.393198 | 0.999532 | | 2.13994 | 0.28572 | 0.881862 | | 2.053514 | -0.08686 | 0.972758 |
| RIPOR2 | 3.495492 | -0.33676 | 0.999532 | | 4.214971 | -0.21579 | 0.881862 | | 3.005336 | 0.139317 | 0.934354 |
| DPY19L3 | 4.120523 | -0.26768 | 0.999532 | | 4.671221 | -0.22289 | 0.881862 | | 3.64672 | 0.066879 | 0.972758 |
| EVPL | 2.566109 | 0.380633 | 0.999532 | | 2.269808 | -0.27471 | 0.881862 | | 2.297453 | -0.6384 | 0.652925 |
| RHOBTB3 | 3.44204 | -0.53718 | 0.999532 | | 4.387504 | -0.21012 | 0.881862 | | 2.973991 | 0.337842 | 0.801905 |
| CHMP4C | 2.446347 | 0.134606 | 0.999532 | | 2.390669 | -0.26597 | 0.881862 | | 2.138038 | -0.3829 | 0.811123 |
| YIPF3 | 4.521698 | -0.15637 | 0.999532 | | 5.513611 | 0.179071 | 0.881862 | | 4.500319 | 0.354967 | 0.735469 |
| CHTF8 | 4.352215 | -0.34793 | 0.999532 | | 5.787774 | 0.182651 | 0.881862 | | 4.287039 | 0.551661 | 0.563461 |
| CACNG7 | 2.720285 | 0.377422 | 0.999532 | | 2.492033 | -0.26896 | 0.881862 | | 2.592129 | -0.63596 | 0.647218 |
| TEX50 | 1.886446 | -0.13915 | 0.999532 | | 1.936957 | -0.2959 | 0.881862 | | 1.647524 | -0.14154 | 0.955834 |
| HRK | 2.42641 | -0.20307 | 0.999532 | | 2.999835 | 0.255718 | 0.881862 | | 2.482625 | 0.458833 | 0.784263 |
| GDF2 | 2.402527 | 0.455308 | 0.999532 | | 2.205681 | -0.28064 | 0.881862 | | 2.247828 | -0.72151 | 0.598589 |
| CCNK | 3.56646 | 0.029257 | 0.999532 | | 4.592139 | 0.214629 | 0.881862 | | 3.821082 | 0.206019 | 0.886495 |
| CUL3 | 4.661316 | -0.11451 | 0.999532 | | 5.746593 | 0.182213 | 0.881862 | | 4.816254 | 0.31563 | 0.780611 |
| GALNT3 | 2.220166 | 0.25944 | 0.999532 | | 2.407384 | 0.262964 | 0.881862 | | 2.279797 | 0.019078 | 0.996681 |
| MEF2C | 3.624811 | -0.36379 | 0.999532 | | 4.879686 | 0.196714 | 0.881862 | | 3.898214 | 0.575499 | 0.60433 |
| XXYLT1 | 2.506879 | -0.40396 | 0.999532 | | 3.456394 | 0.232099 | 0.881862 | | 2.806506 | 0.651836 | 0.626811 |
| RAET1L | 5.989287 | 0.431986 | 0.999532 | | 5.263593 | -0.21911 | 0.881862 | | 5.680647 | -0.67685 | 0.523476 |
| LUC7L | 14.23111 | 0.314287 | 0.999532 | | 14.8329 | 0.135038 | 0.881862 | | 14.13156 | -0.15801 | 0.842754 |
| GP1BB | 4.939955 | -0.02123 | 0.999532 | | 5.38879 | -0.20759 | 0.881862 | | 4.548876 | -0.17397 | 0.899882 |
| PEDS1 | 2.674666 | 0.315364 | 0.999532 | | 2.428191 | -0.26445 | 0.881862 | | 2.291457 | -0.56684 | 0.68528 |
| UBE3A | 2.61802 | 0.121928 | 0.999532 | | 2.622904 | -0.25654 | 0.881862 | | 2.303135 | -0.35847 | 0.816618 |
| FHIT | 2.680086 | 0.280468 | 0.999532 | | 2.508175 | -0.26265 | 0.881862 | | 2.423167 | -0.52737 | 0.70375 |
| SH2D7 | 1.474681 | -0.24984 | 0.999532 | | 1.951643 | 0.296585 | 0.881862 | | 1.57945 | 0.563717 | 0.757835 |
| SSBP2 | 3.288136 | -0.14463 | 0.999532 | | 3.402142 | -0.2245 | 0.881862 | | 2.883658 | -0.06091 | 0.977473 |
| VPS13A | 1.982017 | 0.08359 | 0.999532 | | 2.284197 | 0.270244 | 0.881862 | | 2.057415 | 0.200135 | 0.919181 |
| TEDC2 | 2.824212 | -0.00124 | 0.999532 | | 2.896395 | -0.25028 | 0.881862 | | 2.571599 | -0.23385 | 0.900167 |
| ITGA4 | 3.947281 | 0.154706 | 0.999532 | | 3.985612 | -0.20836 | 0.881862 | | 3.655148 | -0.33943 | 0.791466 |
| NFRKB | 2.289002 | 0.261726 | 0.999532 | | 2.308071 | -0.25948 | 0.881862 | | 2.059951 | -0.4995 | 0.724411 |
| SULT2B1 | 2.343105 | 0.612146 | 0.999532 | | 1.921472 | -0.30281 | 0.881862 | | 2.114262 | -0.89747 | 0.515762 |
| APOA1 | 2.266515 | 0.210506 | 0.999532 | | 2.248228 | -0.28545 | 0.881862 | | 2.091317 | -0.4756 | 0.7719 |
| STMN4 | 1.925459 | -0.57907 | 0.999532 | | 2.716453 | 0.266244 | 0.881862 | | 1.932777 | 0.872156 | 0.552672 |
| C2orf76 | 2.458445 | 0.496038 | 0.999532 | | 2.696495 | 0.256445 | 0.881862 | | 2.556264 | -0.21329 | 0.900583 |
| HSDL1 | 2.471794 | -0.19655 | 0.999532 | | 3.120634 | 0.241385 | 0.881862 | | 2.593591 | 0.458739 | 0.75311 |
| NUCB1 | 21.47559 | 0.25029 | 0.999532 | | 22.69235 | 0.126011 | 0.881862 | | 21.05495 | -0.10344 | 0.905952 |
| TOP3B | 2.427953 | -0.11675 | 0.999532 | | 2.753161 | -0.26415 | 0.881862 | | 2.152484 | -0.12718 | 0.952822 |
| CLN3 | 3.045425 | -0.44375 | 0.999532 | | 3.753587 | -0.22429 | 0.881862 | | 2.736454 | 0.23462 | 0.88476 |
| CTNNBIP1 | 5.863176 | 0.138638 | 0.999532 | | 6.864086 | 0.163157 | 0.881862 | | 5.847659 | 0.033357 | 0.985528 |
| CTSZ | 21.18073 | -0.26907 | 0.999532 | | 27.62177 | 0.153118 | 0.881862 | | 21.61082 | 0.438646 | 0.515762 |
| SHANK2 | 2.893334 | 0.454445 | 0.999532 | | 2.813941 | 0.234074 | 0.881862 | | 2.853826 | -0.2098 | 0.892869 |
| AP4E1 | 2.395584 | -0.73584 | 0.999532 | | 3.038508 | -0.23653 | 0.881862 | | 2.128803 | 0.518865 | 0.728015 |
| ZNF549 | 3.369191 | 0.285222 | 0.999532 | | 3.066655 | -0.24889 | 0.881862 | | 3.058717 | -0.51511 | 0.695561 |
| ENKD1 | 1.958466 | 0.236029 | 0.999532 | | 2.270043 | 0.265687 | 0.881862 | | 2.053889 | 0.048362 | 0.986446 |
| C2orf88 | 3.003139 | 0.220723 | 0.999532 | | 2.638841 | -0.25319 | 0.881862 | | 2.673998 | -0.44859 | 0.764702 |
| TTC37 | 3.572373 | -0.3602 | 0.999532 | | 4.596562 | 0.194414 | 0.881862 | | 3.722685 | 0.578302 | 0.60646 |
| HCST | 4.217018 | -0.06845 | 0.999532 | | 4.605634 | -0.19936 | 0.881862 | | 3.938189 | -0.10704 | 0.948127 |
| PSMA2 | 7.165594 | -0.23657 | 0.999532 | | 9.039393 | 0.14573 | 0.881862 | | 7.033698 | 0.393324 | 0.652925 |
| CSRP2 | 6.059369 | 0.062518 | 0.999532 | | 7.020105 | 0.185603 | 0.881862 | | 6.209855 | 0.141065 | 0.904576 |
| ADRB1 | 2.304979 | -0.21342 | 0.999532 | | 2.557205 | -0.26445 | 0.881862 | | 2.022822 | -0.03476 | 0.991452 |
| SKAP2 | 3.826151 | 0.485374 | 0.999532 | | 3.899236 | 0.204513 | 0.881862 | | 3.971326 | -0.26275 | 0.8278 |
| WRNIP1 | 5.413953 | -0.04308 | 0.999532 | | 6.30075 | 0.175705 | 0.881862 | | 5.290611 | 0.23385 | 0.836935 |
| KNL1 | 2.727383 | 0.326737 | 0.999532 | | 2.460856 | -0.26235 | 0.881862 | | 2.535197 | -0.5739 | 0.684909 |
| WNK4 | 2.851677 | 0.838104 | 0.999532 | | 2.224008 | -0.26895 | 0.881862 | | 2.492384 | -1.09697 | 0.283338 |
| P2RX2 | 2.645215 | 0.496542 | 0.999532 | | 2.447139 | 0.258755 | 0.881862 | | 2.688747 | -0.22938 | 0.892612 |
| C1orf35 | 1.741433 | -0.28174 | 0.999532 | | 1.949275 | -0.28733 | 0.881862 | | 1.616193 | 0.01755 | 0.996891 |
| WDR83 | 2.221465 | -0.15528 | 0.999532 | | 2.520846 | -0.26041 | 0.881862 | | 1.988547 | -0.08138 | 0.974421 |
| PRKACG | 2.030155 | 0.406134 | 0.999532 | | 2.187973 | 0.27118 | 0.881862 | | 2.09592 | -0.11758 | 0.958011 |
| IRF8 | 2.118119 | -0.10603 | 0.999532 | | 2.607569 | 0.255861 | 0.882125 | | 2.239806 | 0.367024 | 0.829716 |
| OR4S1 | 2.632175 | 0.383834 | 0.999532 | | 2.18728 | -0.28281 | 0.882125 | | 2.289665 | -0.66991 | 0.665431 |
| URM1 | 4.601527 | -0.02476 | 0.999532 | | 5.307278 | 0.179233 | 0.882125 | | 4.543037 | 0.21792 | 0.859522 |
| MARCHF | 4.65389 | -0.11207 | 0.999532 | | 5.895945 | 0.197136 | 0.882125 | | 4.69408 | 0.334202 | 0.771685 |
| PRPF40A | 3.42717 | -0.08917 | 0.999532 | | 3.955347 | -0.22967 | 0.882262 | | 3.170505 | -0.12352 | 0.945129 |
| SEC16A | 6.552669 | 0.000386 | 0.999532 | | 6.667839 | -0.17933 | 0.882262 | | 5.635772 | -0.16622 | 0.897096 |
| RBM26 | 4.863621 | -0.1667 | 0.999532 | | 5.075515 | -0.20117 | 0.882262 | | 4.178237 | -0.03769 | 0.987608 |
| METTL18 | 2.521033 | 0.009592 | 0.999532 | | 2.587324 | -0.26393 | 0.882397 | | 2.248979 | -0.25699 | 0.88885 |
| HGH1 | 3.459089 | -0.16842 | 0.999532 | | 3.864169 | -0.2079 | 0.882397 | | 3.022834 | -0.02235 | 0.993618 |
| SPATA5 | 3.09751 | 0.166546 | 0.999532 | | 3.199187 | -0.23205 | 0.882397 | | 2.770368 | -0.36673 | 0.809845 |
| OSBPL8 | 5.137665 | 0.00087 | 0.999532 | | 5.433494 | -0.19463 | 0.882397 | | 4.800625 | -0.17681 | 0.897958 |
| ADPGK | 3.425183 | -0.31907 | 0.999532 | | 4.611795 | 0.200559 | 0.882397 | | 3.390923 | 0.54653 | 0.647581 |
| TNNI2 | 2.949288 | -0.17713 | 0.999532 | | 3.052139 | -0.23099 | 0.882397 | | 2.662597 | -0.04488 | 0.987274 |
| UCK2 | 2.222642 | 0.053048 | 0.999532 | | 2.337503 | -0.26585 | 0.882397 | | 1.97084 | -0.29754 | 0.869378 |
| LKAAEAR | 2.382682 | 0.401195 | 0.999532 | | 2.136228 | -0.29149 | 0.882397 | | 2.048108 | -0.6712 | 0.65516 |
| TMEM140 | 9.63875 | -0.05782 | 0.999532 | | 11.80403 | 0.130958 | 0.882397 | | 9.575921 | 0.210421 | 0.805576 |
| RAB28 | 2.462006 | 0.024788 | 0.999532 | | 2.934639 | 0.242552 | 0.882397 | | 2.481766 | 0.237099 | 0.88885 |
| COX10 | 4.234014 | 0.343891 | 0.999532 | | 4.369258 | 0.20857 | 0.882397 | | 4.501978 | -0.12694 | 0.939107 |
| PDE8A | 5.073295 | 0.220108 | 0.999532 | | 5.537533 | 0.175093 | 0.882397 | | 5.08259 | -0.02815 | 0.989148 |
| CDKN2C | 2.261775 | -0.59078 | 0.999532 | | 2.686182 | -0.24064 | 0.882397 | | 1.92612 | 0.364604 | 0.835478 |
| KIAA0513 | 3.370079 | 0.132835 | 0.999532 | | 3.866954 | 0.203948 | 0.882397 | | 3.548539 | 0.09248 | 0.957598 |
| WNT9B | 1.95533 | 0.05305 | 0.999532 | | 2.373216 | 0.26071 | 0.882397 | | 2.029593 | 0.225559 | 0.903339 |
| SMPD2 | 2.141548 | -0.02088 | 0.999532 | | 2.636583 | 0.249535 | 0.882397 | | 2.38394 | 0.292124 | 0.863134 |
| CEP89 | 2.04671 | 0.301448 | 0.999532 | | 1.843287 | -0.2915 | 0.882397 | | 1.839982 | -0.57534 | 0.718468 |
| NCKAP5L | 2.517899 | 0.080665 | 0.999532 | | 2.569043 | -0.24815 | 0.882397 | | 2.19858 | -0.30822 | 0.859255 |
| B9D2 | 2.602776 | -0.41444 | 0.999532 | | 2.97491 | -0.24746 | 0.882397 | | 2.183469 | 0.18714 | 0.922437 |
| SREK1IP1 | 4.584502 | -0.29976 | 0.999532 | | 5.621443 | -0.18509 | 0.882473 | | 4.210441 | 0.143874 | 0.920025 |
| MIS18BP1 | 2.280724 | 0.373224 | 0.999532 | | 2.092348 | -0.27939 | 0.882487 | | 2.119335 | -0.6361 | 0.66232 |
| CHCHD7 | 3.487662 | 0.233603 | 0.999532 | | 3.182632 | -0.22344 | 0.882487 | | 3.173428 | -0.43904 | 0.728328 |
| CHML | 2.23162 | -0.3297 | 0.999532 | | 2.467427 | -0.25452 | 0.882487 | | 1.939576 | 0.091827 | 0.971732 |
| ICOSLG | 3.598081 | 0.270991 | 0.999532 | | 4.244881 | 0.203664 | 0.882556 | | 3.584443 | -0.05461 | 0.97733 |
| HOOK3 | 8.335502 | -0.21579 | 0.999532 | | 10.55478 | 0.140202 | 0.882556 | | 8.303166 | 0.375987 | 0.652925 |
| TARS3 | 3.442153 | -0.20756 | 0.999532 | | 4.169071 | -0.23141 | 0.882556 | | 3.107799 | 0.014665 | 0.996681 |
| ZC3H18 | 3.720739 | 0.453369 | 0.999532 | | 3.766311 | 0.206799 | 0.882556 | | 3.798575 | -0.22842 | 0.862433 |
| PIK3CA | 3.504482 | 0.651333 | 0.999532 | | 3.252992 | 0.224993 | 0.882556 | | 3.509733 | -0.40842 | 0.723933 |
| SYT17 | 2.072036 | -0.17337 | 0.999532 | | 2.305458 | -0.26715 | 0.882556 | | 1.846989 | -0.07501 | 0.977473 |
| WTAP | 8.212796 | -0.41913 | 0.999532 | | 9.958475 | -0.13848 | 0.882556 | | 7.540219 | 0.29972 | 0.725814 |
| IFIT5 | 2.841868 | -0.42826 | 0.999532 | | 3.645171 | 0.219146 | 0.882556 | | 2.765127 | 0.660453 | 0.606343 |
| NUDT7 | 4.85576 | -0.18494 | 0.999532 | | 5.578885 | -0.18199 | 0.882556 | | 4.476473 | 0.022023 | 0.992286 |
| PHB | 5.788169 | 0.366893 | 0.999532 | | 5.921585 | 0.17708 | 0.882556 | | 5.835436 | -0.17234 | 0.879386 |
| NR1I2 | 1.863529 | -0.21278 | 0.999532 | | 2.151567 | -0.27869 | 0.882635 | | 1.747996 | -0.04579 | 0.989437 |
| ZSCAN32 | 4.019368 | -0.01801 | 0.999532 | | 4.315717 | -0.20939 | 0.882671 | | 3.769494 | -0.17261 | 0.908698 |
| CACNB1 | 1.966883 | 0.194436 | 0.999532 | | 1.843184 | -0.30367 | 0.882671 | | 1.772365 | -0.48404 | 0.795612 |
| FBXO41 | 2.539459 | 0.791148 | 0.999532 | | 1.893494 | -0.29081 | 0.882671 | | 2.262688 | -1.06573 | 0.38606 |
| POLR1F | 1.788823 | 0.201614 | 0.999532 | | 2.08869 | 0.271067 | 0.883187 | | 1.814155 | 0.086622 | 0.972305 |
| IFNLR1 | 2.771948 | 0.234623 | 0.999532 | | 2.700767 | -0.26727 | 0.883227 | | 2.49366 | -0.48047 | 0.749222 |
| OSGIN1 | 3.306422 | -0.10785 | 0.999532 | | 3.537352 | -0.24213 | 0.883249 | | 3.060721 | -0.12454 | 0.948477 |
| SRPX2 | 3.22675 | -0.14135 | 0.999532 | | 3.606575 | -0.22376 | 0.883249 | | 2.957068 | -0.05323 | 0.982338 |
| COQ9 | 3.759647 | 0.150811 | 0.999532 | | 4.457294 | 0.194969 | 0.883249 | | 3.71645 | 0.05968 | 0.974421 |
| MICOS13 | 7.056042 | -0.10558 | 0.999532 | | 8.909943 | 0.147931 | 0.883303 | | 7.305453 | 0.275492 | 0.753585 |
| SLC25A2 | 3.50883 | 0.223682 | 0.999532 | | 3.522836 | -0.22848 | 0.883303 | | 3.243751 | -0.43181 | 0.740755 |
| POLR2D | 4.393143 | 0.182168 | 0.999532 | | 4.954147 | 0.188652 | 0.883303 | | 4.485369 | 0.024627 | 0.991341 |
| MMACHC | 2.559765 | 0.205127 | 0.999532 | | 2.943381 | 0.237401 | 0.883303 | | 2.705855 | 0.051177 | 0.982864 |
| VANGL1 | 3.277463 | 0.266776 | 0.999532 | | 3.104288 | -0.24068 | 0.883303 | | 2.924904 | -0.48846 | 0.706407 |
| CD320 | 3.308099 | 0.574314 | 0.999532 | | 3.200096 | 0.223733 | 0.883303 | | 3.362086 | -0.33175 | 0.795612 |
| CFAP36 | 2.906514 | -0.25671 | 0.999532 | | 3.319465 | -0.22438 | 0.883303 | | 2.668337 | 0.047711 | 0.985299 |
| SDS | 2.113319 | 0.75371 | 0.999532 | | 2.019351 | 0.288919 | 0.883385 | | 2.316754 | -0.45023 | 0.776852 |
| GCFC2 | 2.658069 | 0.195553 | 0.999532 | | 2.875324 | 0.235213 | 0.883385 | | 2.700222 | 0.047221 | 0.985693 |
| REXO1 | 3.675826 | -0.23053 | 0.999532 | | 4.625243 | 0.197078 | 0.883385 | | 3.531081 | 0.45113 | 0.68597 |
| DCUN1D | 2.056199 | -0.124 | 0.999532 | | 2.187563 | -0.28342 | 0.883718 | | 1.796995 | -0.14184 | 0.952656 |
| GLB1L | 3.623524 | 0.151537 | 0.999532 | | 4.107749 | 0.199645 | 0.883718 | | 3.65782 | 0.065839 | 0.972221 |
| MAPKAP | 3.985655 | -0.12849 | 0.999532 | | 4.215351 | -0.19999 | 0.883718 | | 3.400889 | -0.05899 | 0.976603 |
| C22orf39 | 4.665605 | -0.08914 | 0.999532 | | 4.825934 | -0.19228 | 0.883718 | | 4.076369 | -0.08382 | 0.960427 |
| AGAP1 | 5.331794 | 0.003285 | 0.999532 | | 6.366244 | 0.165099 | 0.883784 | | 5.563079 | 0.177621 | 0.877943 |
| UBA6 | 2.985972 | -0.11171 | 0.999532 | | 3.268368 | -0.22092 | 0.883859 | | 2.667632 | -0.09017 | 0.964445 |
| ESRRB | 2.239935 | 0.194015 | 0.999532 | | 2.095693 | -0.27674 | 0.883859 | | 1.941941 | -0.45431 | 0.785695 |
| PDE8B | 2.021602 | -0.1631 | 0.999532 | | 2.286748 | -0.27059 | 0.883859 | | 1.760314 | -0.07947 | 0.976603 |
| NSG1 | 3.80644 | 0.259965 | 0.999532 | | 3.718099 | -0.22312 | 0.883859 | | 3.277433 | -0.44652 | 0.720106 |
| PRR9 | 1.774791 | -0.38821 | 0.999532 | | 2.141599 | -0.28234 | 0.883859 | | 1.711163 | 0.127854 | 0.961643 |
| CMTM1 | 2.273231 | 0.418233 | 0.999532 | | 2.641497 | 0.268667 | 0.883867 | | 2.612819 | -0.13998 | 0.945129 |
| ZNF584 | 2.599457 | 0.325458 | 0.999532 | | 2.778519 | 0.247314 | 0.883934 | | 2.662664 | -0.06179 | 0.978487 |
| SLC16A2 | 2.551122 | 0.675471 | 0.999532 | | 2.596726 | 0.248344 | 0.883975 | | 2.593119 | -0.40734 | 0.759211 |
| SYNJ1 | 2.886987 | -0.0039 | 0.999532 | | 3.450087 | 0.216106 | 0.884001 | | 2.944639 | 0.238227 | 0.876107 |
| TMEM45B | 3.718583 | 0.220468 | 0.999532 | | 3.663016 | -0.2524 | 0.884001 | | 3.319837 | -0.45374 | 0.720526 |
| LCE3A | 5.143974 | 0.334206 | 0.999532 | | 4.89591 | -0.20981 | 0.884001 | | 4.622983 | -0.52293 | 0.646782 |
| DLST | 3.53382 | -0.08386 | 0.999532 | | 4.576071 | 0.203514 | 0.884001 | | 3.752874 | 0.302378 | 0.81294 |
| INAFM2 | 4.036764 | -0.49743 | 0.999532 | | 5.052255 | -0.19665 | 0.884001 | | 3.804864 | 0.319977 | 0.810105 |
| LNPK | 3.284285 | 0.715933 | 0.999532 | | 2.45478 | -0.25799 | 0.884001 | | 2.929215 | -0.95268 | 0.359441 |
| SWSAP1 | 4.498857 | 0.401329 | 0.999532 | | 3.832085 | -0.20897 | 0.884126 | | 3.939429 | -0.59668 | 0.552672 |
| SLC31A1 | 4.456607 | 0.096451 | 0.999532 | | 4.481298 | -0.1947 | 0.884126 | | 4.070169 | -0.27596 | 0.827429 |
| ATP1B3 | 6.948884 | -0.5774 | 0.999532 | | 9.241682 | -0.14255 | 0.884137 | | 6.469056 | 0.458949 | 0.579064 |
| PROKR2 | 2.277149 | -0.00471 | 0.999532 | | 2.398601 | -0.27147 | 0.884137 | | 2.058068 | -0.24431 | 0.900848 |
| USH1C | 2.617643 | -0.39515 | 0.999532 | | 3.545016 | 0.225367 | 0.884738 | | 2.730319 | 0.639448 | 0.630705 |
| ELOVL3 | 2.606177 | 0.697759 | 0.999532 | | 2.047728 | -0.2958 | 0.884887 | | 2.416917 | -0.97297 | 0.484718 |
| CXXC1 | 5.52587 | 0.141168 | 0.999532 | | 6.135223 | 0.164437 | 0.884887 | | 5.531944 | 0.041021 | 0.978927 |
| LYL1 | 4.305676 | 0.169976 | 0.999532 | | 4.664567 | 0.189388 | 0.884887 | | 4.479705 | 0.045205 | 0.979967 |
| THAP1 | 3.709828 | 0.25287 | 0.999532 | | 4.087512 | 0.206998 | 0.884887 | | 3.821667 | -0.0276 | 0.991159 |
| CNOT9 | 3.663333 | 0.179863 | 0.999532 | | 4.121445 | 0.21312 | 0.884887 | | 3.534591 | 0.050247 | 0.978487 |
| SLC6A12 | 3.099594 | 0.573558 | 0.999532 | | 2.958944 | 0.234431 | 0.884887 | | 3.160757 | -0.32201 | 0.819902 |
| NCOR2 | 9.330409 | -0.08439 | 0.999532 | | 10.96791 | 0.130671 | 0.884894 | | 9.10742 | 0.223638 | 0.806394 |
| TRAF7 | 2.029749 | -0.11446 | 0.999532 | | 2.543773 | 0.245817 | 0.885123 | | 2.079133 | 0.37837 | 0.814247 |
| TMX2 | 4.252705 | 0.06014 | 0.999532 | | 4.509271 | -0.19848 | 0.885123 | | 3.685735 | -0.24068 | 0.855173 |
| GLYCTK | 3.429521 | 0.485723 | 0.999532 | | 3.066314 | 0.229843 | 0.885238 | | 3.333526 | -0.22734 | 0.887487 |
| VEZF1 | 6.842433 | -0.01281 | 0.999532 | | 7.834537 | 0.155769 | 0.885238 | | 6.801633 | 0.174906 | 0.884993 |
| CSAG3 | 2.618641 | 0.416479 | 0.999532 | | 2.4875 | -0.27294 | 0.885238 | | 2.423766 | -0.64385 | 0.666555 |
| ZNF691 | 2.295789 | 0.378982 | 0.999532 | | 2.413545 | 0.260744 | 0.885238 | | 2.351282 | -0.0962 | 0.965087 |
| HDAC11 | 2.290404 | -0.50833 | 0.999532 | | 3.22003 | 0.23611 | 0.885409 | | 2.47584 | 0.754392 | 0.593895 |
| ALKBH6 | 1.9362 | 0.164754 | 0.999532 | | 2.504375 | 0.265396 | 0.885409 | | 2.212916 | 0.141233 | 0.948127 |
| HELB | 2.314483 | 0.328735 | 0.999532 | | 2.332436 | 0.251927 | 0.885409 | | 2.358826 | -0.06527 | 0.978229 |
| ACSM5 | 4.708046 | 0.452371 | 0.999532 | | 4.785397 | 0.198705 | 0.885409 | | 4.969645 | -0.23884 | 0.842754 |
| TP53BP1 | 5.935484 | -0.08616 | 0.999532 | | 6.472394 | -0.18541 | 0.886007 | | 5.423798 | -0.0779 | 0.96556 |
| SMKR1 | 3.212036 | 0.409231 | 0.999532 | | 2.785995 | -0.25046 | 0.886636 | | 2.944866 | -0.6436 | 0.628895 |
| REEP3 | 2.262995 | -0.19407 | 0.999532 | | 2.512272 | -0.24914 | 0.886636 | | 2.031152 | -0.03616 | 0.991273 |
| RAB4B | 2.78913 | 0.279769 | 0.999532 | | 2.485248 | -0.24157 | 0.886636 | | 2.454702 | -0.50469 | 0.705268 |
| NOC3L | 2.352681 | 0.062812 | 0.999532 | | 2.414291 | -0.27485 | 0.886636 | | 2.048146 | -0.31694 | 0.862823 |
| LARP4B | 3.891825 | 0.452755 | 0.999532 | | 3.945939 | 0.219936 | 0.886636 | | 3.785507 | -0.21166 | 0.892869 |
| ZNF14 | 3.063627 | -0.08292 | 0.999532 | | 3.233206 | -0.21733 | 0.886636 | | 2.737333 | -0.11965 | 0.946765 |
| ZFPM1 | 4.545649 | 0.288175 | 0.999532 | | 4.300343 | -0.19455 | 0.886636 | | 4.215045 | -0.46594 | 0.660097 |
| RNF5 | 3.670625 | -0.11853 | 0.999532 | | 4.354711 | 0.201353 | 0.886636 | | 3.685902 | 0.327449 | 0.805825 |
| LRIG3 | 2.450152 | 0.054465 | 0.999532 | | 2.876725 | 0.252996 | 0.886636 | | 2.621172 | 0.208796 | 0.909255 |
| SDC4 | 8.661337 | 0.031136 | 0.999532 | | 10.29388 | 0.155259 | 0.886636 | | 8.429798 | 0.136537 | 0.901916 |
| PPP1R18 | 3.061716 | 0.040789 | 0.999532 | | 3.078605 | -0.22491 | 0.886636 | | 2.720515 | -0.24952 | 0.871072 |
| LTN1 | 3.777396 | 0.516603 | 0.999532 | | 3.706397 | 0.21101 | 0.886636 | | 3.737371 | -0.28827 | 0.82125 |
| UQCC1 | 2.90312 | -0.13483 | 0.999532 | | 3.360781 | -0.22538 | 0.886636 | | 2.591517 | -0.07081 | 0.974421 |
| KCNC4 | 5.952597 | 0.080834 | 0.999532 | | 6.071576 | -0.16442 | 0.886636 | | 5.535892 | -0.22184 | 0.836935 |
| CCDC138 | 2.707053 | 0.312003 | 0.999532 | | 2.274922 | -0.26384 | 0.886636 | | 2.314622 | -0.56406 | 0.706407 |
| SFXN3 | 3.777433 | 0.165825 | 0.999532 | | 4.396783 | 0.197945 | 0.886636 | | 3.847954 | 0.044036 | 0.984725 |
| PPFIBP1 | 7.911126 | -0.1076 | 0.999532 | | 9.63253 | 0.143681 | 0.886636 | | 7.897897 | 0.269742 | 0.754884 |
| C20orf20 | 1.958294 | -0.13271 | 0.999532 | | 2.172316 | -0.2722 | 0.886636 | | 1.765017 | -0.12076 | 0.960773 |
| CIPC | 2.026056 | 0.137335 | 0.999532 | | 2.277304 | 0.271658 | 0.886636 | | 2.067973 | 0.152383 | 0.945129 |
| SAMD15 | 2.682813 | 0.836386 | 0.999532 | | 2.334558 | 0.258771 | 0.886636 | | 2.704329 | -0.56217 | 0.666566 |
| COPS2 | 5.924081 | -0.2852 | 0.999532 | | 7.572298 | 0.160664 | 0.886636 | | 6.045848 | 0.45874 | 0.630705 |
| NR3C1 | 4.397274 | -0.44001 | 0.999532 | | 5.905259 | 0.177645 | 0.886636 | | 4.509431 | 0.654728 | 0.456688 |
| ZBTB8B | 2.344838 | 0.272711 | 0.999532 | | 2.25298 | -0.2797 | 0.886636 | | 2.166549 | -0.53348 | 0.739496 |
| HSPB8 | 6.185097 | -0.63865 | 0.999532 | | 8.945262 | 0.169203 | 0.886636 | | 5.984235 | 0.85085 | 0.280916 |
| NUP93 | 2.86037 | -0.11695 | 0.999532 | | 3.472119 | 0.206553 | 0.886636 | | 2.918352 | 0.340147 | 0.803301 |
| BRAF | 6.943864 | -0.29692 | 0.999532 | | 8.228129 | -0.14842 | 0.886636 | | 6.063138 | 0.167427 | 0.881837 |
| ZNRF1 | 4.83664 | 0.403416 | 0.999532 | | 4.499017 | -0.19692 | 0.886636 | | 4.657751 | -0.58798 | 0.540342 |
| PRDX4 | 2.703974 | -0.12949 | 0.999532 | | 3.379889 | 0.211881 | 0.886636 | | 2.81859 | 0.359463 | 0.795928 |
| ZDHHC18 | 3.159598 | -0.24386 | 0.999532 | | 3.471114 | -0.22044 | 0.886636 | | 2.837177 | 0.036317 | 0.989985 |
| HOXA1 | 2.226859 | -0.25925 | 0.999532 | | 2.611537 | -0.25058 | 0.886636 | | 2.065128 | 0.034906 | 0.99186 |
| HDGFL3 | 5.431289 | -0.14866 | 0.999532 | | 6.495965 | 0.165737 | 0.886636 | | 5.398758 | 0.31941 | 0.7719 |
| SLC35B4 | 2.399854 | 0.581437 | 0.999532 | | 1.914479 | -0.28567 | 0.886636 | | 2.208722 | -0.85827 | 0.515762 |
| HHIP | 2.000121 | 0.26062 | 0.999532 | | 2.071697 | 0.27677 | 0.886636 | | 2.048033 | 0.018653 | 0.996681 |
| PRDM2 | 5.075422 | -0.1907 | 0.999532 | | 6.282001 | 0.167352 | 0.886636 | | 5.060427 | 0.376066 | 0.709825 |
| HMOX2 | 5.389265 | 0.205268 | 0.999532 | | 5.83506 | 0.166313 | 0.886636 | | 5.257174 | -0.02556 | 0.990868 |
| PDIK1L | 2.909603 | 0.503817 | 0.999532 | | 2.562918 | -0.25132 | 0.886636 | | 2.668928 | -0.73799 | 0.535079 |
| RREB1 | 4.497382 | -0.19897 | 0.999532 | | 4.859712 | -0.18572 | 0.886636 | | 3.932351 | 0.017249 | 0.995943 |
| GALNTL5 | 5.788382 | 0.386567 | 0.999532 | | 5.456085 | -0.23428 | 0.886636 | | 5.41966 | -0.61122 | 0.614525 |
| GFI1B | 2.134877 | -0.03648 | 0.999532 | | 2.262495 | -0.26372 | 0.886636 | | 1.964247 | -0.21075 | 0.915533 |
| ZNF586 | 2.752947 | -0.17302 | 0.999532 | | 2.900047 | -0.24284 | 0.886636 | | 2.274247 | -0.05344 | 0.984587 |
| OXNAD1 | 3.294742 | -0.24326 | 0.999532 | | 4.358916 | 0.207844 | 0.886636 | | 3.573505 | 0.463695 | 0.715437 |
| MUC12 | 2.384479 | 0.418281 | 0.999532 | | 2.143917 | -0.28365 | 0.886636 | | 2.177185 | -0.68597 | 0.647581 |
| ADIPOR2 | 2.900443 | 0.253746 | 0.999532 | | 2.759986 | -0.24493 | 0.886636 | | 2.568354 | -0.47889 | 0.735469 |
| MAP1LC3 | 2.833272 | 0.270894 | 0.999532 | | 3.064241 | 0.22922 | 0.886636 | | 2.81846 | -0.02616 | 0.992286 |
| CDK14 | 2.625212 | 0.794024 | 0.999532 | | 2.448697 | 0.251043 | 0.886636 | | 2.723422 | -0.52467 | 0.670068 |
| GCLC | 3.100771 | 0.080517 | 0.999532 | | 3.574937 | 0.206843 | 0.886636 | | 3.198101 | 0.143815 | 0.92697 |
| OR52N2 | 2.310455 | 0.091103 | 0.999532 | | 2.265943 | -0.26509 | 0.886636 | | 2.115794 | -0.33738 | 0.851604 |
| PPP2R3B | 2.553832 | -0.35634 | 0.999532 | | 3.065389 | -0.2207 | 0.886636 | | 2.280096 | 0.156619 | 0.932496 |
| DCST1 | 2.139875 | 0.019634 | 0.999532 | | 2.140594 | -0.27976 | 0.886636 | | 2.009664 | -0.28017 | 0.892869 |
| GYPC | 8.918075 | 0.032402 | 0.999532 | | 9.648812 | 0.153108 | 0.886636 | | 8.755568 | 0.109547 | 0.925618 |
| ZNF217 | 4.736889 | -0.04007 | 0.999532 | | 5.533956 | 0.174436 | 0.886636 | | 5.056274 | 0.237253 | 0.842696 |
| SMIM41 | 1.615384 | -0.19562 | 0.999532 | | 2.099726 | 0.280311 | 0.886636 | | 1.83367 | 0.483363 | 0.799627 |
| RIOK1 | 2.3636 | 0.037782 | 0.999532 | | 2.365778 | -0.25188 | 0.886636 | | 1.99606 | -0.27408 | 0.878619 |
| MISP | 2.555513 | -0.1176 | 0.999532 | | 2.921498 | -0.24524 | 0.886636 | | 2.336882 | -0.09209 | 0.969783 |
| TSPAN6 | 5.247829 | 0.395389 | 0.999532 | | 5.421194 | 0.176623 | 0.886636 | | 5.16569 | -0.19506 | 0.877268 |
| OR4C16 | 1.83365 | 0.161195 | 0.999532 | | 1.911556 | -0.2976 | 0.886636 | | 1.694487 | -0.43562 | 0.816618 |
| LLGL1 | 3.383626 | 0.388638 | 0.999532 | | 3.309823 | 0.217657 | 0.886636 | | 3.450701 | -0.1448 | 0.927433 |
| PHF7 | 2.537345 | 0.215087 | 0.999532 | | 2.512657 | -0.29307 | 0.886636 | | 2.350401 | -0.51709 | 0.735469 |
| RBSN | 2.984513 | 0.236477 | 0.999532 | | 2.931951 | -0.23542 | 0.886636 | | 2.780163 | -0.45247 | 0.736481 |
| CD82 | 3.574971 | -0.1375 | 0.999532 | | 4.036269 | -0.2162 | 0.886636 | | 3.118082 | -0.07117 | 0.972305 |
| RBM12 | 2.177053 | 0.107394 | 0.999532 | | 2.087193 | -0.26737 | 0.886854 | | 1.972408 | -0.36966 | 0.836935 |
| CUTA | 4.744939 | 0.320228 | 0.999532 | | 4.755912 | 0.181311 | 0.886902 | | 4.639353 | -0.11997 | 0.927375 |
| TMEM106 | 100.1615 | 0.018411 | 0.999532 | | 116.8421 | 0.122175 | 0.886902 | | 100.2418 | 0.13205 | 0.886495 |
| TRABD | 3.550767 | 0.058236 | 0.999532 | | 3.466017 | -0.21932 | 0.886902 | | 2.947282 | -0.263 | 0.864245 |
| TMEM54 | 2.314137 | 0.162649 | 0.999532 | | 2.278358 | -0.25579 | 0.886902 | | 2.023702 | -0.39774 | 0.805466 |
| OSR1 | 2.492258 | 0.305494 | 0.999532 | | 2.653266 | 0.240879 | 0.886902 | | 2.677745 | -0.04625 | 0.98637 |
| INTS7 | 2.894235 | 0.302799 | 0.999532 | | 2.603112 | -0.24182 | 0.886902 | | 2.623229 | -0.52811 | 0.702463 |
| ARL4A | 3.287502 | 0.235662 | 0.999532 | | 3.141668 | -0.22547 | 0.886902 | | 2.905428 | -0.44233 | 0.732486 |
| KIF22 | 2.658663 | -0.15978 | 0.999532 | | 2.9895 | -0.2367 | 0.887026 | | 2.472503 | -0.07169 | 0.974421 |
| OPLAH | 2.226039 | 0.354772 | 0.999532 | | 2.141153 | -0.28061 | 0.887026 | | 2.014542 | -0.60799 | 0.702061 |
| NCAM1 | 2.742694 | -0.16494 | 0.999532 | | 2.897777 | -0.2348 | 0.887026 | | 2.416458 | -0.05953 | 0.979996 |
| RAB22A | 3.165551 | -0.2776 | 0.999532 | | 4.163509 | 0.206096 | 0.887781 | | 3.37453 | 0.502143 | 0.686214 |
| LSM6 | 2.433282 | -0.14108 | 0.999532 | | 3.120685 | 0.223058 | 0.887783 | | 2.65918 | 0.383906 | 0.791576 |
| THBS3 | 5.950623 | 0.140728 | 0.999532 | | 6.01744 | -0.17218 | 0.887783 | | 5.211437 | -0.29355 | 0.77448 |
| GTPBP10 | 3.090027 | 0.316803 | 0.999532 | | 3.374633 | 0.216033 | 0.887783 | | 3.140124 | -0.08042 | 0.968434 |

| | | | | | | | | | | | |
|---|---|---|---|---|---|---|---|---|---|---|---|
| TNFRSF1 | 3.879864 | -0.11478 | 0.999532 | | 4.205721 | -0.19792 | 0.887811 | | 3.481503 | -0.06403 | 0.974331 |
| LTBP1 | 2.839502 | -0.65373 | 0.999532 | | 3.551048 | -0.23089 | 0.887811 | | 2.46357 | 0.44286 | 0.764702 |
| SNORC | 3.975065 | 0.344041 | 0.999532 | | 3.533773 | -0.22319 | 0.887825 | | 3.697551 | -0.57579 | 0.629345 |
| HEATR3 | 1.923944 | 0.00136 | 0.999532 | | 2.511182 | 0.247387 | 0.887825 | | 2.166907 | 0.270119 | 0.881837 |
| AGT | 2.076064 | 0.191089 | 0.999532 | | 2.295741 | 0.264991 | 0.887825 | | 2.194244 | 0.088867 | 0.972344 |
| FGF16 | 1.830563 | 0.277337 | 0.999532 | | 2.063461 | 0.276617 | 0.887825 | | 1.877575 | 0.016233 | 0.996891 |
| WWC3 | 6.49626 | 0.01527 | 0.999532 | | 7.326846 | 0.151529 | 0.887825 | | 6.584204 | 0.15853 | 0.885712 |
| C9orf24 | 1.909435 | 0.235275 | 0.999532 | | 2.124598 | 0.266543 | 0.887825 | | 1.957135 | 0.046254 | 0.988768 |
| SLC10A1 | 1.944689 | 0.119261 | 0.999532 | | 1.948765 | -0.28189 | 0.887825 | | 1.790532 | -0.38213 | 0.836935 |
| MNDA | 2.472868 | 0.553154 | 0.999532 | | 2.334812 | 0.2657 | 0.888131 | | 2.442208 | -0.25897 | 0.878742 |
| IYD | 2.096416 | -0.00231 | 0.999532 | | 2.239191 | -0.26337 | 0.888451 | | 1.870129 | -0.24072 | 0.901033 |
| FHL2 | 11.81631 | -0.48892 | 0.999532 | | 15.9398 | 0.180275 | 0.888451 | | 11.96388 | 0.685201 | 0.397994 |
| AGAP3 | 4.261489 | -0.42852 | 0.999532 | | 5.882044 | 0.166458 | 0.888539 | | 4.263099 | 0.611137 | 0.520848 |
| SERPINI1 | 2.797837 | 0.406195 | 0.999532 | | 3.030357 | 0.233002 | 0.888782 | | 2.95758 | -0.15702 | 0.926513 |
| MFSD1 | 3.47606 | -0.19882 | 0.999532 | | 3.930116 | -0.21776 | 0.888782 | | 2.950104 | 0.007242 | 0.997967 |
| BSPRY | 2.668991 | 0.195128 | 0.999532 | | 2.570799 | -0.25061 | 0.888782 | | 2.431484 | -0.42467 | 0.784263 |
| TOR1AIP | 3.970046 | -0.233 | 0.999532 | | 5.175122 | 0.175371 | 0.888782 | | 4.104871 | 0.429167 | 0.699971 |
| CDC26 | 3.076964 | -0.26812 | 0.999532 | | 3.494645 | -0.20984 | 0.888999 | | 2.762111 | 0.078571 | 0.971732 |
| C6orf89 | 5.583647 | 0.170394 | 0.999532 | | 6.247484 | 0.171083 | 0.88903 | | 5.633727 | 0.023733 | 0.991159 |
| TM4SF1 | 16.44247 | -0.41382 | 0.999532 | | 22.15498 | 0.150694 | 0.889036 | | 16.70843 | 0.586875 | 0.359441 |
| C5AR1 | 2.571418 | -0.2546 | 0.999532 | | 3.133237 | 0.240866 | 0.889036 | | 2.602216 | 0.499557 | 0.733499 |
| RBM15 | 2.545627 | 0.462338 | 0.999532 | | 2.428854 | 0.248674 | 0.889036 | | 2.529213 | -0.19851 | 0.911296 |
| SEC24A | 2.579161 | -0.25426 | 0.999532 | | 3.129119 | 0.217871 | 0.889036 | | 2.594628 | 0.485489 | 0.720106 |
| EIF4ENIF | 2.865798 | -0.0372 | 0.999532 | | 2.991855 | -0.22989 | 0.889036 | | 2.483369 | -0.17606 | 0.920399 |
| GCGR | 2.385535 | 0.274255 | 0.999532 | | 2.722422 | 0.23225 | 0.889036 | | 2.412966 | -0.02196 | 0.99505 |
| MAST2 | 2.752286 | -0.23505 | 0.999532 | | 3.191728 | -0.22341 | 0.889036 | | 2.443308 | 0.030516 | 0.991783 |
| TAS2R45 | 3.4441 | 0.215421 | 0.999532 | | 3.856595 | 0.205845 | 0.889036 | | 3.733164 | 0.002725 | 0.999385 |
| GMEB1 | 3.798288 | -0.04259 | 0.999532 | | 4.624125 | 0.185221 | 0.889036 | | 3.955201 | 0.247757 | 0.849021 |
| NDUFAF4 | 3.85346 | -0.1668 | 0.999532 | | 4.395023 | -0.19403 | 0.889036 | | 3.5868 | 0.004714 | 0.998585 |
| PTPN5 | 3.537551 | -0.09402 | 0.999532 | | 3.741159 | -0.21428 | 0.889141 | | 3.303367 | -0.1048 | 0.955834 |
| ARHGEF | 1.931335 | 0.289195 | 0.999532 | | 2.048948 | 0.263715 | 0.889162 | | 1.920687 | -0.0084 | 0.997967 |
| MIEF2 | 2.199848 | -0.00173 | 0.999532 | | 2.310759 | -0.27526 | 0.889162 | | 2.027174 | -0.25339 | 0.900848 |
| RAP1GA | 2.933429 | 0.083873 | 0.999532 | | 2.750472 | -0.24378 | 0.889398 | | 2.590338 | -0.30267 | 0.85585 |
| ERMP1 | 2.881427 | -0.19951 | 0.999532 | | 3.930532 | 0.201769 | 0.889607 | | 3.103864 | 0.434094 | 0.737109 |
| PIP5K1B | 3.199655 | -0.32265 | 0.999532 | | 3.59361 | -0.22412 | 0.889607 | | 2.894688 | 0.131831 | 0.944202 |
| SNF8 | 6.564984 | 0.202793 | 0.999532 | | 7.212931 | 0.164507 | 0.889607 | | 6.507049 | -0.02031 | 0.992286 |
| COL4A4 | 4.979695 | 0.583195 | 0.999532 | | 5.14753 | 0.191242 | 0.889607 | | 5.156404 | -0.36403 | 0.735469 |
| ZSWIM9 | 2.767811 | 0.152032 | 0.999532 | | 3.08003 | 0.229933 | 0.889607 | | 2.876075 | 0.096606 | 0.963866 |
| CCAR1 | 6.162034 | -0.13181 | 0.999532 | | 6.71357 | -0.18503 | 0.889607 | | 5.312221 | -0.01133 | 0.996891 |
| SENP5 | 5.235348 | -0.01575 | 0.999532 | | 6.236604 | 0.164391 | 0.889607 | | 5.487273 | 0.197073 | 0.870965 |
| PHKA2 | 2.629158 | 0.282674 | 0.999532 | | 2.802739 | 0.230081 | 0.889607 | | 2.682061 | -0.04888 | 0.985693 |
| MST1 | 7.364732 | 0.320938 | 0.999532 | | 7.379485 | -0.17506 | 0.889607 | | 6.964455 | -0.46546 | 0.622543 |
| SOCS4 | 2.848251 | 0.28735 | 0.999532 | | 2.752759 | -0.24102 | 0.889607 | | 2.549689 | -0.5084 | 0.706749 |
| KIAA1522 | 4.72578 | 0.026015 | 0.999532 | | 5.520202 | 0.172074 | 0.889607 | | 4.648102 | 0.162036 | 0.898369 |
| CDC5L | 2.867678 | -0.25293 | 0.999532 | | 3.544322 | 0.213225 | 0.889607 | | 2.901688 | 0.479003 | 0.712621 |
| THAP4 | 4.096995 | 0.243177 | 0.999532 | | 4.409749 | 0.198899 | 0.889724 | | 4.039032 | -0.02407 | 0.992286 |
| OR4D1 | 2.272609 | -0.05558 | 0.999532 | | 2.468061 | -0.25198 | 0.889784 | | 2.138206 | -0.17198 | 0.934572 |
| LIPA | 3.481519 | 0.366306 | 0.999532 | | 3.584407 | 0.206534 | 0.889784 | | 3.456091 | -0.14575 | 0.929156 |
| AATF | 4.171693 | 0.041833 | 0.999532 | | 4.510161 | -0.19704 | 0.889784 | | 3.788581 | -0.21745 | 0.876428 |
| DAB2IP | 2.958133 | 0.446281 | 0.999532 | | 2.908446 | 0.220112 | 0.889784 | | 2.951468 | -0.21137 | 0.889271 |
| MLH1 | 2.559807 | -0.29271 | 0.999532 | | 2.926724 | -0.22164 | 0.889784 | | 2.214948 | 0.087541 | 0.969501 |
| NKIRAS1 | 2.832263 | 0.987466 | 0.999532 | | 2.635323 | 0.243482 | 0.889784 | | 3.006126 | -0.72522 | 0.515762 |
| ZNF611 | 4.821984 | 0.187964 | 0.999532 | | 5.058796 | -0.18448 | 0.889784 | | 4.52326 | -0.34557 | 0.753585 |
| GPR139 | 2.139683 | 0.168485 | 0.999532 | | 2.176448 | -0.27613 | 0.889784 | | 1.912472 | -0.42482 | 0.806394 |
| DENND2 | 3.583446 | 0.530866 | 0.999532 | | 3.252722 | -0.22584 | 0.889784 | | 3.344561 | -0.74076 | 0.484128 |
| FABP6 | 2.2477 | 0.143367 | 0.999532 | | 2.218264 | -0.2661 | 0.889784 | | 2.085202 | -0.39654 | 0.821463 |
| TCEAL1 | 3.675419 | 0.679282 | 0.999532 | | 3.372797 | 0.215378 | 0.889784 | | 3.624903 | -0.4461 | 0.688214 |
| ZNF532 | 7.983501 | -0.27545 | 0.999532 | | 10.38768 | 0.145061 | 0.890116 | | 7.88938 | 0.449077 | 0.568811 |
| VWF | 2.86882 | -0.24006 | 0.999532 | | 3.265714 | -0.21807 | 0.890116 | | 2.619654 | 0.040801 | 0.988768 |
| ZBTB17 | 3.656238 | -0.04924 | 0.999532 | | 3.61104 | -0.24331 | 0.890776 | | 3.201587 | -0.12728 | 0.945129 |
| EXD3 | 3.961661 | 0.257218 | 0.999532 | | 3.717823 | -0.21008 | 0.890891 | | 3.493233 | -0.44342 | 0.704316 |
| IFT122 | 2.499469 | 0.322248 | 0.999532 | | 2.654544 | 0.234146 | 0.890891 | | 2.559378 | -0.07139 | 0.974421 |
| OSER1 | 3.360889 | 0.259819 | 0.999532 | | 3.821713 | 0.231205 | 0.890891 | | 3.354921 | -0.00549 | 0.998123 |
| MAP3K20 | 3.07986 | 0.301124 | 0.999532 | | 2.860253 | -0.22826 | 0.890891 | | 2.672343 | -0.51302 | 0.695541 |
| RADIL | 1.971961 | -0.12535 | 0.999532 | | 2.554853 | 0.25685 | 0.890891 | | 2.142065 | 0.397227 | 0.816618 |
| WDR7 | 4.251445 | -0.19988 | 0.999532 | | 5.11531 | -0.18633 | 0.890891 | | 3.928761 | 0.04047 | 0.985299 |
| ACTR1B | 3.684212 | 0.052548 | 0.999532 | | 4.55241 | 0.18603 | 0.890891 | | 3.813061 | 0.154003 | 0.912963 |
| CAMSAP | 3.510912 | 0.089497 | 0.999532 | | 3.983794 | 0.194836 | 0.890891 | | 3.433451 | 0.12363 | 0.938429 |
| OR10H2 | 2.822223 | 0.659599 | 0.999532 | | 2.6264 | 0.252342 | 0.89092 | | 2.943337 | -0.39898 | 0.791466 |
| C1R | 11.96481 | 0.287739 | 0.999532 | | 12.91905 | 0.160357 | 0.89092 | | 12.1656 | -0.10719 | 0.928552 |
| ZNF850 | 1.953397 | 0.236751 | 0.999532 | | 1.906316 | -0.27798 | 0.89092 | | 1.777693 | -0.49413 | 0.766974 |
| IWS1 | 3.381412 | -0.02852 | 0.999532 | | 3.740475 | -0.19643 | 0.89092 | | 3.092571 | -0.14835 | 0.925487 |
| HLA-DOA | 5.700317 | 0.176329 | 0.999532 | | 6.257038 | 0.187437 | 0.89092 | | 5.614385 | 0.028686 | 0.991085 |
| FCSK | 2.122699 | -0.4508 | 0.999532 | | 2.748058 | -0.23798 | 0.89092 | | 2.043955 | 0.242564 | 0.90066 |
| ELF3 | 2.377534 | 0.344719 | 0.999532 | | 2.195323 | -0.2625 | 0.89092 | | 2.23025 | -0.59441 | 0.679818 |
| CCDC120 | 1.893414 | -0.19637 | 0.999532 | | 2.102527 | -0.26443 | 0.89092 | | 1.724105 | -0.0496 | 0.988379 |
| ARC | 2.425991 | 0.085963 | 0.999532 | | 2.504918 | -0.24984 | 0.89092 | | 2.143385 | -0.30996 | 0.864245 |
| RP2 | 2.763453 | 0.713409 | 0.999532 | | 2.372518 | 0.246766 | 0.89092 | | 2.773863 | -0.44951 | 0.740755 |
| KRTAP5- | 2.297289 | 0.254148 | 0.999532 | | 2.306717 | -0.25942 | 0.89092 | | 2.143222 | -0.49356 | 0.741003 |
| NFYA | 40.47422 | 0.00137 | 0.999532 | | 47.06303 | 0.086591 | 0.89092 | | 39.79245 | 0.110663 | 0.859092 |
| FLNB | 4.712279 | -0.181 | 0.999532 | | 5.853701 | 0.164782 | 0.89092 | | 4.754704 | 0.362225 | 0.72206 |
| ABO | 3.242608 | 0.073717 | 0.999532 | | 3.126887 | -0.21329 | 0.89092 | | 2.891533 | -0.27274 | 0.857377 |
| CPSF6 | 3.123065 | 0.293948 | 0.999532 | | 3.433144 | 0.210959 | 0.89092 | | 3.181638 | -0.06372 | 0.974421 |
| CHORDC | 3.70384 | 0.487873 | 0.999532 | | 3.722793 | 0.210965 | 0.89092 | | 3.658307 | -0.25388 | 0.848938 |
| CRELD1 | 4.514485 | -0.18024 | 0.999532 | | 5.068373 | -0.18043 | 0.89092 | | 4.105847 | 0.01782 | 0.994617 |
| GPR68 | 2.320353 | 0.067235 | 0.999532 | | 2.671772 | 0.243153 | 0.89092 | | 2.382731 | 0.185889 | 0.924131 |
| S100A7 | 2.831596 | 0.060765 | 0.999532 | | 3.134487 | -0.23735 | 0.89092 | | 2.769425 | -0.27453 | 0.872561 |
| ZC2HC1C | 2.027026 | 0.267235 | 0.999532 | | 2.240135 | 0.266399 | 0.89092 | | 2.162947 | 0.017245 | 0.996681 |
| TIGIT | 2.148606 | -0.14117 | 0.999532 | | 2.293563 | -0.25724 | 0.89092 | | 1.922959 | -0.09737 | 0.967577 |
| ELAC1 | 2.349614 | 0.008678 | 0.999532 | | 2.586224 | 0.239186 | 0.89092 | | 2.402741 | 0.250232 | 0.892004 |
| XYLT2 | 2.259437 | -0.15803 | 0.999532 | | 2.495012 | -0.23701 | 0.89092 | | 2.00614 | -0.06105 | 0.979432 |
| TOR1A | 2.23719 | -0.22531 | 0.999532 | | 2.451284 | -0.24628 | 0.89092 | | 1.916499 | -0.00258 | 0.999385 |
| FKBP15 | 5.291845 | -0.10296 | 0.999532 | | 5.631685 | -0.16749 | 0.89092 | | 4.925571 | -0.03765 | 0.98556 |
| PLPPR2 | 4.352185 | -0.16101 | 0.999532 | | 5.068225 | -0.19228 | 0.89092 | | 3.955323 | -0.00415 | 0.998585 |
| DHTKD1 | 4.399603 | -0.05454 | 0.999532 | | 4.546358 | -0.18504 | 0.89092 | | 3.802922 | -0.11493 | 0.941197 |
| RPS15A | 24.79314 | -0.0965 | 0.999532 | | 29.59126 | 0.095107 | 0.89092 | | 24.53111 | 0.210958 | 0.719612 |
| NTPCR | 3.550743 | -0.18603 | 0.999532 | | 4.758106 | 0.181697 | 0.89092 | | 3.723948 | 0.391931 | 0.734912 |
| MICALCL | 2.781552 | 0.542982 | 0.999532 | | 2.271229 | -0.2576 | 0.89092 | | 2.487025 | -0.78488 | 0.532278 |
| SOD2 | 7.949135 | -0.34683 | 0.999532 | | 10.51901 | 0.180479 | 0.89092 | | 8.15769 | 0.580538 | 0.515762 |
| ZNF449 | 3.573931 | -0.2023 | 0.999532 | | 3.8755 | -0.19636 | 0.89092 | | 3.184206 | 0.021686 | 0.994192 |
| PLAAT3 | 5.166529 | -0.16763 | 0.999532 | | 6.82889 | 0.156522 | 0.89092 | | 5.424824 | 0.350204 | 0.722703 |
| BCL2 | 4.754896 | 0.590978 | 0.999532 | | 4.672942 | 0.192991 | 0.89092 | | 4.701196 | -0.38152 | 0.720526 |
| METAP1D | 2.234819 | 0.444123 | 0.999532 | | 2.12067 | -0.27928 | 0.89092 | | 2.044283 | -0.70037 | 0.644433 |
| ODF2 | 3.698057 | 0.221132 | 0.999532 | | 3.529834 | -0.20402 | 0.89092 | | 3.441287 | -0.4104 | 0.723021 |
| TMEM16 | 8.419103 | 0.102134 | 0.999532 | | 8.692394 | -0.14772 | 0.89092 | | 7.678365 | -0.23129 | 0.796702 |
| SLC27A4 | 4.867883 | -0.40515 | 0.999532 | | 6.515076 | 0.156026 | 0.890923 | | 5.157853 | 0.584496 | 0.522695 |
| KCTD11 | 5.381846 | -0.3717 | 0.999532 | | 6.284291 | -0.16743 | 0.890923 | | 4.797832 | 0.211612 | 0.876781 |
| BIN1 | 4.475609 | 0.042422 | 0.999532 | | 4.579669 | -0.18496 | 0.890923 | | 3.93553 | -0.20853 | 0.876781 |
| NYAP1 | 4.086664 | 0.214682 | 0.999532 | | 4.805009 | 0.198779 | 0.890968 | | 4.202938 | -0.00435 | 0.99858 |
| TMEM275 | 1.921701 | 0.217497 | 0.999532 | | 2.089895 | 0.2604 | 0.890968 | | 1.931864 | 0.059005 | 0.982338 |
| MMP20 | 2.529033 | 0.291849 | 0.999532 | | 2.535007 | -0.26634 | 0.890968 | | 2.317858 | -0.53506 | 0.722703 |
| APOBR | 1.849603 | 0.148638 | 0.999532 | | 2.063577 | 0.271748 | 0.891272 | | 1.849932 | 0.143046 | 0.949067 |
| TM6SF2 | 3.645197 | -0.13136 | 0.999532 | | 4.167768 | -0.21214 | 0.891504 | | 3.435966 | -0.04955 | 0.984988 |
| SEZ6L2 | 3.042569 | 0.04731 | 0.999532 | | 3.674601 | 0.205306 | 0.891504 | | 3.075314 | 0.179993 | 0.906811 |
| HOXA13 | 2.059746 | -0.36908 | 0.999532 | | 2.821593 | 0.239934 | 0.891504 | | 2.269741 | 0.629345 | 0.676565 |
| FAM110A | 4.484306 | 0.048049 | 0.999532 | | 4.995465 | 0.186701 | 0.891504 | | 4.647271 | 0.175324 | 0.900583 |
| DET1 | 3.687274 | 0.232416 | 0.999532 | | 3.75339 | -0.20349 | 0.891504 | | 3.303136 | -0.41413 | 0.739496 |
| FAM3D | 3.288336 | 0.298693 | 0.999532 | | 3.108707 | -0.22129 | 0.891504 | | 3.041642 | -0.50227 | 0.692333 |
| ABCB1 | 2.820996 | -0.09546 | 0.999532 | | 3.343717 | 0.21527 | 0.891525 | | 2.792942 | 0.321902 | 0.835036 |
| SOSTDC1 | 2.523511 | 0.155468 | 0.999532 | | 2.681587 | 0.22706 | 0.8916 | | 2.517775 | 0.080689 | 0.972221 |
| PCNX3 | 3.261548 | -0.22939 | 0.999532 | | 3.719549 | -0.19754 | 0.891624 | | 3.087782 | 0.053793 | 0.978927 |
| PLA2G2A | 2.085528 | 0.075727 | 0.999532 | | 2.103813 | -0.28012 | 0.891637 | | 1.894402 | -0.33803 | 0.865384 |
| VPS16 | 3.51339 | 0.274205 | 0.999532 | | 3.739031 | 0.207321 | 0.891802 | | 3.536164 | -0.05052 | 0.979996 |
| BEX3 | 7.132643 | 0.222414 | 0.999532 | | 7.971865 | 0.141672 | 0.8919 | | 6.892062 | -0.06614 | 0.961567 |
| ESRRG | 3.370635 | 0.089428 | 0.999532 | | 3.440614 | -0.2242 | 0.891975 | | 2.908142 | -0.29382 | 0.84579 |
| RIMS3 | 2.228503 | 0.450206 | 0.999532 | | 2.302151 | 0.249683 | 0.892207 | | 2.369829 | -0.17865 | 0.923834 |
| NRG3 | 2.358378 | -0.21667 | 0.999532 | | 2.550473 | -0.23991 | 0.892213 | | 2.028317 | -0.00812 | 0.997967 |
| ANXA3 | 3.123776 | 0.298329 | 0.999532 | | 3.524728 | 0.229399 | 0.892213 | | 3.249984 | -0.03909 | 0.988768 |
| ECI1 | 3.805272 | 0.042912 | 0.999532 | | 4.474492 | 0.187413 | 0.892213 | | 3.555737 | 0.157394 | 0.915533 |
| MAP7D3 | 2.158223 | 0.193476 | 0.999532 | | 1.952454 | -0.26992 | 0.892213 | | 1.906259 | -0.44738 | 0.798099 |
| C16orf70 | 4.2789 | 0.008043 | 0.999532 | | 5.127774 | 0.182887 | 0.892213 | | 4.436896 | 0.191969 | 0.88476 |
| RFXAP | 1.934507 | 0.227513 | 0.999532 | | 2.145727 | 0.262345 | 0.892213 | | 2.181842 | 0.050058 | 0.987274 |
| DYRK1A | 5.253227 | -0.2637 | 0.999532 | | 5.883869 | -0.17068 | 0.892349 | | 4.682388 | 0.113618 | 0.938429 |
| TMEM87B | 2.494089 | 0.609593 | 0.999532 | | 2.516641 | 0.237622 | 0.892382 | | 2.542006 | -0.34943 | 0.809498 |
| CCL22 | 3.06108 | 0.372488 | 0.999532 | | 2.825622 | -0.26596 | 0.892382 | | 2.772857 | -0.62294 | 0.654772 |
| TRAF3 | 2.701097 | 0.07046 | 0.999532 | | 3.230788 | 0.221927 | 0.892382 | | 2.7815 | 0.171106 | 0.920124 |
| ENOPH1 | 4.414222 | 0.011149 | 0.999532 | | 5.358529 | 0.173935 | 0.892382 | | 4.553024 | 0.181685 | 0.887683 |
| DNPH1 | 2.924804 | -0.17803 | 0.999532 | | 3.633666 | 0.197648 | 0.892382 | | 2.958929 | 0.390772 | 0.764702 |
| CDC14C | 1.941391 | 0.10329 | 0.999532 | | 2.051109 | -0.27539 | 0.892382 | | 1.805124 | -0.35452 | 0.85585 |
| ZNF768 | 4.561532 | -0.2494 | 0.999532 | | 5.688167 | 0.171595 | 0.892382 | | 4.329365 | 0.434418 | 0.679818 |
| SLC7A9 | 3.118612 | 0.25108 | 0.999532 | | 3.415207 | 0.21602 | 0.892415 | | 3.030147 | -0.01351 | 0.996681 |
| MPPED2 | 3.667065 | 0.255642 | 0.999532 | | 3.856505 | 0.205362 | 0.892415 | | 3.880707 | -0.02968 | 0.991134 |
| ABCA2 | 2.994911 | 0.230656 | 0.999532 | | 3.234344 | 0.213801 | 0.892415 | | 2.915455 | -0.00229 | 0.999385 |
| NEB | 2.257161 | -0.05354 | 0.999532 | | 2.295074 | -0.24933 | 0.892488 | | 2.066124 | -0.17897 | 0.931619 |
| NARF | 3.58489 | 0.556997 | 0.999532 | | 2.826572 | -0.22264 | 0.892488 | | 3.251315 | -0.76348 | 0.479746 |
| TMEM175 | 4.387775 | 0.027347 | 0.999532 | | 5.092277 | 0.176606 | 0.892616 | | 4.490503 | 0.171513 | 0.893262 |
| CH25H | 2.045383 | 0.177897 | 0.999532 | | 2.068292 | -0.27784 | 0.892711 | | 1.839784 | -0.43567 | 0.806797 |
| COG7 | 4.597332 | 0.27637 | 0.999532 | | 5.121862 | 0.17688 | 0.892806 | | 4.737641 | -0.08183 | 0.956322 |
| SOCS6 | 3.352327 | 0.027323 | 0.999532 | | 3.219094 | -0.21535 | 0.892806 | | 3.018574 | -0.23271 | 0.892869 |
| IL21 | 2.097653 | 0.160585 | 0.999532 | | 1.948668 | -0.26835 | 0.892934 | | 1.879492 | -0.41643 | 0.814884 |
| ADGRF3 | 2.398971 | 0.54222 | 0.999532 | | 2.075202 | -0.2711 | 0.892934 | | 2.122742 | -0.79466 | 0.552672 |
| KIAA1549 | 3.092325 | -0.0366 | 0.999532 | | 3.587797 | 0.202087 | 0.892934 | | 3.228951 | 0.258145 | 0.862823 |
| EPM2AIP | 4.646013 | -0.30691 | 0.999532 | | 5.41391 | -0.1738 | 0.893009 | | 3.983313 | 0.151224 | 0.914687 |
| ABCB7 | 3.160933 | 0.087063 | 0.999532 | | 3.385663 | 0.214353 | 0.89302 | | 3.236463 | 0.138337 | 0.939913 |
| DHDDS | 2.320507 | 0.161032 | 0.999532 | | 2.604048 | 0.23304 | 0.893112 | | 2.433762 | 0.108595 | 0.960427 |
| CNTNAP | 2.998466 | 0.131898 | 0.999532 | | 3.480363 | 0.220183 | 0.893112 | | 3.08888 | 0.108206 | 0.955167 |
| ENPP1 | 2.654266 | 0.439794 | 0.999532 | | 2.514556 | 0.244769 | 0.893112 | | 2.66886 | -0.20217 | 0.91707 |
| BRK1 | 8.831803 | -0.33489 | 0.999532 | | 11.58445 | 0.119993 | 0.89343 | | 8.936916 | 0.475493 | 0.49299 |
| MPP2 | 2.986099 | -0.00732 | 0.999532 | | 2.983246 | -0.24536 | 0.893691 | | 2.514914 | -0.21776 | 0.905261 |
| ABCD4 | 4.373985 | 0.306762 | 0.999532 | | 4.536246 | 0.185982 | 0.893691 | | 4.470064 | -0.09695 | 0.950895 |
| PDYN | 2.387265 | 0.017962 | 0.999532 | | 2.366525 | -0.25845 | 0.893973 | | 2.324526 | -0.25911 | 0.89901 |
| RGS12 | 3.238719 | 0.030543 | 0.999532 | | 3.795526 | 0.204523 | 0.893983 | | 3.32956 | 0.19086 | 0.90113 |
| FNDC5 | 2.591292 | 0.365912 | 0.999532 | | 2.353186 | -0.25349 | 0.893983 | | 2.42549 | -0.6054 | 0.667066 |
| NSG2 | 2.030738 | 0.230831 | 0.999532 | | 2.242014 | 0.267096 | 0.893983 | | 2.209497 | 0.039468 | 0.991159 |
| TYR | 2.109576 | -0.33764 | 0.999532 | | 2.944717 | 0.248553 | 0.893983 | | 2.353823 | 0.605829 | 0.703463 |
| RHOBTB | 4.650586 | -0.36134 | 0.999532 | | 6.418967 | 0.166552 | 0.894207 | | 4.849858 | 0.553841 | 0.573023 |
| SPG21 | 2.000316 | 0.172953 | 0.999532 | | 2.371554 | 0.238226 | 0.894371 | | 2.04625 | 0.082882 | 0.972221 |
| CLNS1A | 5.127381 | -0.04347 | 0.999532 | | 6.067688 | 0.166355 | 0.894378 | | 5.173805 | 0.227153 | 0.849682 |
| PRXL2C | 4.83586 | 0.145304 | 0.999532 | | 5.570224 | 0.168167 | 0.894378 | | 4.874022 | 0.046039 | 0.978312 |
| ERGIC2 | 3.747223 | -0.24364 | 0.999532 | | 5.11314 | 0.169249 | 0.894378 | | 4.01942 | 0.432197 | 0.676169 |
| ESR1 | 2.888384 | -0.09794 | 0.999532 | | 3.153462 | -0.23861 | 0.894378 | | 2.591547 | -0.12067 | 0.955834 |
| RASL11B | 3.965664 | 0.148078 | 0.999532 | | 4.00036 | -0.22291 | 0.894378 | | 3.657483 | -0.36412 | 0.791576 |
| RBP1 | 3.892455 | -0.17644 | 0.999532 | | 4.404238 | -0.19548 | 0.894378 | | 3.715012 | -0.00279 | 0.999385 |
| PRKAG1 | 5.885344 | -0.11984 | 0.999532 | | 7.321242 | 0.152016 | 0.894378 | | 5.924899 | 0.288001 | 0.792416 |
| SCG3 | 2.241804 | 0.675854 | 0.999532 | | 2.215118 | 0.266736 | 0.894708 | | 2.379352 | -0.39426 | 0.8089 |
| GDI2 | 9.64798 | -0.32921 | 0.999532 | | 12.19461 | 0.131406 | 0.894708 | | 9.670928 | 0.475638 | 0.499456 |
| AMD1 | 6.249018 | 0.336315 | 0.999532 | | 5.528601 | -0.16563 | 0.894708 | | 5.619683 | -0.487 | 0.600081 |
| ZNF622 | 2.56778 | -0.26638 | 0.999532 | | 3.069036 | -0.23125 | 0.894708 | | 2.292769 | 0.057595 | 0.981849 |
| RNF25 | 2.069863 | -0.44519 | 0.999532 | | 2.277803 | -0.2493 | 0.894708 | | 1.74157 | 0.199994 | 0.927623 |
| LINGO2 | 2.20511 | 0.256279 | 0.999532 | | 2.133506 | -0.27364 | 0.894708 | | 2.011434 | -0.51212 | 0.757822 |
| PIP4P1 | 4.537328 | -0.02389 | 0.999532 | | 5.229905 | 0.173549 | 0.894708 | | 4.418747 | 0.220327 | 0.858705 |
| HSPA4L | 2.669745 | -0.01337 | 0.999532 | | 2.699083 | -0.24895 | 0.894708 | | 2.131566 | -0.18681 | 0.922922 |
| C1orf52 | 5.296117 | 0.069823 | 0.999532 | | 6.059931 | 0.159133 | 0.894708 | | 5.471809 | 0.108918 | 0.93603 |
| CAPN7 | 4.650644 | 0.028396 | 0.999532 | | 4.880295 | -0.17649 | 0.894708 | | 4.079365 | -0.18675 | 0.886495 |
| ARMC9 | 3.08406 | 0.039164 | 0.999532 | | 3.105342 | -0.24542 | 0.894708 | | 2.615908 | -0.25176 | 0.880577 |
| DNAJC14 | 3.673214 | 0.160517 | 0.999532 | | 4.381084 | 0.187114 | 0.894708 | | 3.958725 | 0.051833 | 0.978282 |
| CRACD | 2.690801 | 0.057257 | 0.999532 | | 2.831289 | -0.23623 | 0.894708 | | 2.254997 | -0.26892 | 0.875951 |
| SHPK | 4.285351 | -0.49887 | 0.999532 | | 5.379294 | -0.16533 | 0.894708 | | 3.781292 | 0.352577 | 0.763181 |
| CHRAC1 | 4.520977 | 0.173469 | 0.999532 | | 5.099817 | 0.172392 | 0.894708 | | 4.585303 | 0.020074 | 0.993334 |
| DUSP13 | 2.376872 | 0.102074 | 0.999532 | | 2.29461 | -0.2501 | 0.894708 | | 2.269322 | -0.34457 | 0.848095 |
| N4BP2L2 | 8.331013 | 0.073311 | 0.999532 | | 10.04061 | 0.150515 | 0.894708 | | 8.555079 | 0.105728 | 0.928552 |
| VEGFC | 3.050958 | -0.27512 | 0.999532 | | 4.039664 | 0.195113 | 0.894708 | | 3.207128 | 0.486722 | 0.686214 |
| IPO5 | 4.065365 | 0.271115 | 0.999532 | | 4.383892 | 0.203609 | 0.894708 | | 4.039309 | -0.04128 | 0.985299 |
| ANXA11 | 16.01033 | -0.08012 | 0.999532 | | 19.417 | 0.114001 | 0.894708 | | 16.06861 | 0.19081 | 0.806797 |
| TSPAN13 | 2.804295 | 0.360789 | 0.999532 | | 2.980103 | 0.222136 | 0.894708 | | 2.887592 | -0.11924 | 0.946512 |
| CCDC191 | 3.071722 | 0.843324 | 0.999532 | | 2.49176 | 0.233121 | 0.894708 | | 3.029427 | -0.59185 | 0.625223 |
| PPP3CC | 3.422582 | -0.40392 | 0.999532 | | 4.046397 | -0.19829 | 0.894708 | | 2.971415 | 0.22617 | 0.881837 |
| TRHDE | 2.830328 | 0.016148 | 0.999532 | | 2.804171 | -0.25279 | 0.894708 | | 2.368355 | -0.23855 | 0.892869 |
| SACM1L | 5.511478 | 0.05982 | 0.999532 | | 6.065472 | 0.158497 | 0.894708 | | 5.461015 | 0.11722 | 0.927433 |
| SLC8A1 | 5.726221 | 0.233069 | 0.999532 | | 6.373462 | 0.166915 | 0.894708 | | 5.524753 | -0.10688 | 0.947322 |
| HOXA7 | 2.87787 | -0.1795 | 0.999532 | | 3.257802 | -0.21224 | 0.894708 | | 2.634781 | -0.01429 | 0.996681 |
| ERC2 | 2.184086 | 0.676467 | 0.999532 | | 1.666186 | -0.283 | 0.894708 | | 1.934284 | -0.94324 | 0.49299 |
| CAPN5 | 4.168413 | -0.21445 | 0.999532 | | 4.886365 | -0.18114 | 0.894708 | | 3.801891 | 0.05089 | 0.978282 |
| ANAPC1 | 2.410938 | 0.016243 | 0.999532 | | 2.933404 | 0.225197 | 0.894708 | | 2.431744 | 0.223952 | 0.89433 |
| NAIF1 | 2.624698 | 0.665117 | 0.999532 | | 2.221115 | -0.25494 | 0.894708 | | 2.412074 | -0.90121 | 0.445661 |
| PRUNE2 | 2.073167 | 0.469794 | 0.999532 | | 2.166045 | 0.251707 | 0.894708 | | 2.078884 | -0.20497 | 0.909074 |
| FOXL2NB | 2.207121 | 0.384502 | 0.999532 | | 2.294133 | 0.255884 | 0.894708 | | 2.205081 | -0.11027 | 0.962597 |
| DNAJC16 | 3.241258 | -0.09139 | 0.999532 | | 3.460859 | -0.20935 | 0.894708 | | 2.927692 | -0.09907 | 0.958191 |
| RASGRP | 2.249119 | -0.09581 | 0.999532 | | 2.784902 | 0.231126 | 0.894708 | | 2.42307 | 0.346117 | 0.836453 |
| ADO | 2.189262 | 0.052463 | 0.999532 | | 2.126192 | -0.25671 | 0.894708 | | 1.889901 | -0.29377 | 0.878591 |
| LY75 | 2.144652 | -0.21563 | 0.999532 | | 2.49781 | -0.24375 | 0.894708 | | 1.971255 | -0.01266 | 0.996891 |
| BBC3 | 2.599141 | -0.13629 | 0.999532 | | 2.927419 | -0.23284 | 0.894708 | | 2.492546 | -0.08203 | 0.97358 |
| GAPVD1 | 4.109103 | 0.224714 | 0.999532 | | 4.777236 | 0.183433 | 0.894708 | | 4.223558 | -0.02356 | 0.992286 |
| UBXN6 | 5.980126 | 0.032835 | 0.999532 | | 7.250097 | 0.153006 | 0.894708 | | 5.992018 | 0.146219 | 0.908891 |
| DCHS1 | 4.767877 | -0.03278 | 0.999532 | | 5.582439 | 0.174493 | 0.894708 | | 4.757719 | 0.235061 | 0.839101 |
| FLOT2 | 10.51092 | 0.152257 | 0.999532 | | 11.37379 | 0.134161 | 0.894708 | | 10.59965 | -0.00596 | 0.996891 |
| PAGE2B | 2.745753 | 0.379125 | 0.999532 | | 2.493281 | -0.25741 | 0.894708 | | 2.577603 | -0.61443 | 0.679818 |
| GIT1 | 5.683138 | 0.066674 | 0.999532 | | 6.453117 | 0.168672 | 0.894708 | | 5.684616 | 0.128692 | 0.914692 |
| AQP12B | 3.112879 | 0.27015 | 0.999532 | | 2.809273 | -0.22589 | 0.894708 | | 2.827548 | -0.48443 | 0.723453 |
| RAB24 | 3.41946 | 0.319947 | 0.999532 | | 3.77888 | 0.198033 | 0.894708 | | 3.545819 | -0.1085 | 0.946512 |
| TM4SF20 | 2.808619 | 0.484318 | 0.999532 | | 2.797668 | -0.24701 | 0.894708 | | 2.682165 | -0.67496 | 0.625223 |
| DAB1 | 2.524998 | 0.425919 | 0.999532 | | 2.24462 | -0.26963 | 0.894708 | | 2.27541 | -0.67843 | 0.635381 |
| IKBKG | 5.174405 | -0.1148 | 0.999532 | | 5.620794 | -0.17573 | 0.894708 | | 4.62928 | -0.04245 | 0.984283 |
| URGCP | 2.520383 | 0.098863 | 0.999532 | | 2.501414 | -0.23837 | 0.894708 | | 2.313451 | -0.32236 | 0.849885 |
| RAD51D | 3.331591 | -0.43722 | 0.999532 | | 4.256435 | -0.19949 | 0.894708 | | 2.977185 | 0.274667 | 0.862006 |
| COBLL1 | 5.776063 | 0.077465 | 0.999532 | | 6.873962 | 0.161208 | 0.894708 | | 5.734505 | 0.108602 | 0.939759 |
| NHP2 | 6.476547 | 0.342746 | 0.999532 | | 6.961309 | 0.15852 | 0.894708 | | 6.567069 | -0.16277 | 0.881837 |
| GPRIN3 | 3.492931 | 0.46422 | 0.999532 | | 3.53269 | 0.226364 | 0.894708 | | 3.762434 | -0.24225 | 0.879538 |
| GTF2A1 | 5.05776 | 0.308737 | 0.999532 | | 4.677208 | -0.18507 | 0.894708 | | 4.533466 | -0.47547 | 0.644433 |
| LRRC8C | 2.853646 | 0.048 | 0.999532 | | 3.541034 | 0.205749 | 0.894708 | | 3.03624 | 0.177909 | 0.909087 |
| TNFRSF1 | 2.021107 | 0.170127 | 0.999532 | | 1.997841 | -0.26583 | 0.894708 | | 1.81515 | -0.42171 | 0.800473 |
| CDK6 | 4.321403 | -0.06497 | 0.999532 | | 4.623642 | -0.18973 | 0.894708 | | 3.935432 | -0.10609 | 0.947515 |
| KIF11 | 1.987187 | 0.300166 | 0.999532 | | 2.266188 | 0.259466 | 0.894708 | | 2.217254 | -0.02043 | 0.996681 |
| UBQLNL | 2.543182 | 0.308165 | 0.999532 | | 2.261343 | -0.2573 | 0.894708 | | 2.218227 | -0.54558 | 0.717138 |
| FAM124A | 2.594385 | -0.14798 | 0.999532 | | 2.970054 | 0.219563 | 0.894708 | | 2.586913 | 0.375775 | 0.809832 |
| MED15 | 8.773149 | 0.195628 | 0.999532 | | 9.672035 | 0.131349 | 0.894708 | | 8.766466 | -0.04556 | 0.972221 |
| SRD5A2 | 2.408891 | 0.171315 | 0.999532 | | 2.16952 | -0.25541 | 0.894748 | | 2.17326 | -0.40301 | 0.816589 |
| NOP56 | 6.507036 | 0.210168 | 0.999532 | | 7.30158 | 0.158865 | 0.894857 | | 6.456255 | -0.03412 | 0.98503 |
| NKX6-1 | 2.766467 | 0.502602 | 0.999532 | | 2.373272 | -0.26018 | 0.895098 | | 2.556913 | -0.74807 | 0.563455 |
| TBC1D12 | 4.537689 | -0.11946 | 0.999532 | | 4.971158 | -0.18273 | 0.895407 | | 4.015023 | -0.04333 | 0.98353 |
| VN1R4 | 2.884021 | 0.351421 | 0.999532 | | 3.136662 | 0.213041 | 0.895407 | | 3.044108 | -0.1196 | 0.945129 |
| NEMP1 | 2.494287 | 0.33486 | 0.999532 | | 2.431515 | -0.26077 | 0.895407 | | 2.277749 | -0.58563 | 0.673405 |
| CPSF7 | 5.94686 | 0.231705 | 0.999532 | | 6.227786 | 0.16991 | 0.895407 | | 5.891754 | -0.07073 | 0.968434 |
| RCN1 | 11.48968 | 0.159616 | 0.999532 | | 12.50561 | 0.124222 | 0.895495 | | 11.33243 | -0.01706 | 0.991341 |
| SPPL2C | 2.49995 | 0.184389 | 0.999532 | | 2.418634 | -0.25014 | 0.895495 | | 2.316011 | -0.41726 | 0.795777 |
| BCO2 | 2.016236 | -0.38497 | 0.999532 | | 2.522167 | -0.25516 | 0.895495 | | 1.978875 | 0.144843 | 0.950616 |
| EXT2 | 4.077734 | -0.06099 | 0.999532 | | 4.882035 | 0.179235 | 0.895495 | | 3.988597 | 0.261819 | 0.836935 |
| ARHGAP | 2.451812 | -0.0192 | 0.999532 | | 2.991685 | 0.219983 | 0.895495 | | 2.482489 | 0.258956 | 0.876781 |
| MLX | 6.247531 | -0.28867 | 0.999532 | | 7.424184 | -0.19703 | 0.895495 | | 5.900783 | 0.07939 | 0.963309 |
| C2orf49 | 4.582819 | 0.027522 | 0.999532 | | 5.223344 | 0.16822 | 0.895495 | | 4.629528 | 0.1591 | 0.904804 |
| ABCC9 | 2.588051 | 0.35847 | 0.999532 | | 2.493823 | -0.24265 | 0.895495 | | 2.257268 | -0.57194 | 0.67937 |
| CS | 8.281894 | 0.047576 | 0.999532 | | 9.464901 | 0.13715 | 0.895495 | | 8.20802 | 0.109857 | 0.916388 |
| BST1 | 5.515027 | -0.10783 | 0.999532 | | 6.052666 | -0.19379 | 0.895495 | | 4.695772 | -0.04359 | 0.984362 |
| FBXO33 | 2.734145 | 0.128784 | 0.999532 | | 2.808433 | -0.21631 | 0.895495 | | 2.53862 | -0.31937 | 0.83651 |
| RPS6KA4 | 2.85052 | 0.343743 | 0.999532 | | 2.864261 | 0.220126 | 0.895495 | | 2.931747 | -0.09985 | 0.96129 |
| RBM45 | 2.419603 | -0.18793 | 0.999532 | | 2.514753 | -0.23439 | 0.895495 | | 2.228443 | -0.03256 | 0.992286 |
| PCSK6 | 3.848891 | -0.20257 | 0.999532 | | 4.7892 | 0.177694 | 0.895495 | | 3.928098 | 0.404767 | 0.715437 |
| STX6 | 3.538588 | -0.18077 | 0.999532 | | 4.193956 | 0.184545 | 0.895495 | | 3.531124 | 0.395814 | 0.746876 |
| LMCD1 | 4.736782 | -0.4807 | 0.999532 | | 6.677052 | 0.159856 | 0.895564 | | 4.993585 | 0.662002 | 0.459697 |
| PUM2 | 6.365826 | 0.016317 | 0.999532 | | 7.216495 | 0.146937 | 0.895584 | | 6.241138 | 0.134135 | 0.914687 |
| CHD1L | 3.346995 | -0.21448 | 0.999532 | | 3.524941 | -0.20734 | 0.895584 | | 2.927186 | 0.031011 | 0.991452 |
| ERCC1 | 4.820377 | 0.080896 | 0.999532 | | 5.343007 | 0.174007 | 0.895784 | | 4.735007 | 0.112774 | 0.938429 |
| R3HCC1L | 3.743771 | -0.02604 | 0.999532 | | 4.073943 | -0.19128 | 0.895784 | | 3.286853 | -0.14393 | 0.92697 |
| FOXJ3 | 4.165112 | -0.09385 | 0.999532 | | 4.664167 | -0.1912 | 0.895784 | | 3.791465 | -0.08372 | 0.96129 |
| ZNF175 | 3.024821 | -0.37543 | 0.999532 | | 3.593306 | -0.20095 | 0.895851 | | 2.775923 | 0.188754 | 0.910105 |
| IGSF23 | 2.401118 | -0.2076 | 0.999532 | | 2.859858 | -0.22029 | 0.895851 | | 2.319062 | 0.013518 | 0.996891 |
| KLK13 | 2.16803 | 0.238758 | 0.999532 | | 2.127666 | -0.26638 | 0.895851 | | 1.957513 | -0.48345 | 0.774044 |
| NFAM1 | 4.717856 | 0.183405 | 0.999532 | | 4.739493 | -0.18085 | 0.895851 | | 4.138407 | -0.34224 | 0.754304 |
| PCGF6 | 2.918268 | 0.097718 | 0.999532 | | 2.958948 | -0.21957 | 0.895851 | | 2.571043 | -0.29839 | 0.849682 |
| RBMS2 | 10.72813 | -0.05626 | 0.999532 | | 12.26881 | 0.116052 | 0.896069 | | 10.48695 | 0.181165 | 0.842754 |
| C7orf77 | 2.123405 | 0.64724 | 0.999532 | | 1.803877 | -0.2809 | 0.896119 | | 1.884559 | -0.90828 | 0.515762 |
| BMX | 2.320025 | 0.527558 | 0.999532 | | 1.978777 | -0.26942 | 0.896208 | | 2.150816 | -0.78351 | 0.570037 |
| NANP | 2.599429 | 0.772224 | 0.999532 | | 2.11321 | -0.26458 | 0.896208 | | 2.341983 | -1.01668 | 0.387954 |
| SNAPC4 | 3.795109 | 0.045266 | 0.999532 | | 4.725269 | 0.183027 | 0.896208 | | 3.965714 | 0.142122 | 0.920259 |
| NINJ2 | 2.348937 | -0.51253 | 0.999532 | | 2.915823 | -0.23118 | 0.896208 | | 2.222134 | 0.286822 | 0.877451 |
| C6orf136 | 2.846984 | -0.24347 | 0.999532 | | 3.746299 | 0.192973 | 0.896208 | | 2.932435 | 0.454232 | 0.715437 |
| CRYBG3 | 4.381581 | 0.119025 | 0.999532 | | 4.955343 | 0.176294 | 0.896208 | | 4.541543 | 0.084249 | 0.958191 |
| TPX2 | 1.895614 | -0.51546 | 0.999532 | | 2.232311 | -0.24959 | 0.896208 | | 1.624917 | 0.282754 | 0.892004 |
| POLDIP2 | 6.036495 | 0.050303 | 0.999532 | | 7.010166 | 0.152429 | 0.89639 | | 6.072085 | 0.122167 | 0.920472 |
| RBM10 | 2.380083 | -0.1604 | 0.999532 | | 2.748385 | 0.216006 | 0.896445 | | 2.367649 | 0.395022 | 0.792538 |
| ODF3L1 | 2.201362 | 0.424818 | 0.999532 | | 1.993155 | -0.27065 | 0.896541 | | 2.079266 | -0.67656 | 0.655993 |
| STK11 | 3.731471 | -0.21901 | 0.999532 | | 4.289557 | -0.18418 | 0.896597 | | 3.311283 | 0.055452 | 0.977621 |
| SEMG1 | 2.305159 | -0.53017 | 0.999532 | | 2.737734 | -0.22647 | 0.896597 | | 2.266117 | 0.327256 | 0.859426 |
| CCN4 | 6.20017 | 0.111283 | 0.999532 | | 6.137442 | -0.16566 | 0.896597 | | 5.645506 | -0.28261 | 0.813686 |
| IAH1 | 4.463845 | -0.00546 | 0.999532 | | 4.801808 | -0.17421 | 0.896597 | | 3.891857 | -0.14943 | 0.914687 |
| QPCTL | 3.263716 | 0.338872 | 0.999532 | | 3.348679 | 0.211989 | 0.896597 | | 3.256452 | -0.10756 | 0.952785 |
| SZT2 | 4.994216 | 0.435275 | 0.999532 | | 4.589576 | -0.17918 | 0.896597 | | 4.474425 | -0.59444 | 0.540342 |
| NELFA | 4.72426 | -0.1027 | 0.999532 | | 5.128279 | -0.17248 | 0.896597 | | 4.162112 | -0.04925 | 0.978229 |
| CLDND1 | 4.753819 | 0.065516 | 0.999532 | | 5.261219 | 0.171249 | 0.896597 | | 4.600006 | 0.116685 | 0.938429 |
| TAF1L | 2.175017 | 0.30527 | 0.999532 | | 1.933354 | -0.27885 | 0.896604 | | 1.883864 | -0.5645 | 0.735469 |
| EXTL1 | 2.145443 | -0.59571 | 0.999532 | | 3.243977 | 0.221009 | 0.896675 | | 2.342797 | 0.841189 | 0.540342 |
| TAFA2 | 2.176158 | 0.879287 | 0.999532 | | 2.021945 | 0.270279 | 0.896902 | | 2.305091 | -0.59077 | 0.679452 |
| NEURL1B | 3.470118 | -0.2384 | 0.999532 | | 4.708725 | 0.195622 | 0.896902 | | 3.649769 | 0.444988 | 0.703463 |
| ZDHHC21 | 2.44668 | -0.34514 | 0.999532 | | 3.274286 | 0.217456 | 0.896939 | | 2.506302 | 0.580941 | 0.677493 |
| TNFRSF1 | 2.772568 | 0.013396 | 0.999532 | | 2.933187 | -0.22481 | 0.897035 | | 2.636118 | -0.22076 | 0.90016 |
| SLC9A2 | 2.474997 | 0.414546 | 0.999532 | | 2.149585 | -0.25086 | 0.897092 | | 2.173948 | -0.65039 | 0.652925 |
| SYDE1 | 5.15496 | -0.13931 | 0.999532 | | 5.653412 | -0.17725 | 0.897092 | | 4.703986 | -0.01662 | 0.99572 |
| ZNF846 | 2.083475 | -0.18119 | 0.999532 | | 2.118192 | -0.24969 | 0.897092 | | 1.761827 | -0.05962 | 0.984624 |
| THOC2 | 2.034666 | 0.484527 | 0.999532 | | 1.659889 | -0.27972 | 0.897092 | | 1.822562 | -0.74812 | 0.630705 |
| CTSK | 2.617162 | 0.359586 | 0.999532 | | 2.309893 | -0.24222 | 0.897092 | | 2.344695 | -0.58802 | 0.668481 |
| ITGAE | 2.267791 | 0.632691 | 0.999532 | | 1.928926 | -0.27383 | 0.897092 | | 2.016838 | -0.88753 | 0.515762 |
| TAS2R5 | 2.793514 | 0.628833 | 0.999532 | | 2.217469 | -0.24798 | 0.897092 | | 2.458747 | -0.85994 | 0.481527 |
| GRAMD2 | 5.160443 | 0.255262 | 0.999532 | | 5.486703 | 0.167567 | 0.897092 | | 5.119906 | -0.06801 | 0.965649 |
| QDPR | 2.021043 | 0.710176 | 0.999532 | | 1.663736 | -0.28674 | 0.897092 | | 1.81791 | -0.9764 | 0.484303 |
| TFRC | 4.44316 | -0.5845 | 0.999532 | | 5.639638 | -0.17612 | 0.897092 | | 4.111044 | 0.433185 | 0.733499 |
| GTPBP2 | 4.597832 | 0.07008 | 0.999532 | | 4.828061 | -0.17561 | 0.897092 | | 4.313584 | -0.22456 | 0.862823 |
| MRPL20 | 5.980178 | 0.313829 | 0.999532 | | 6.234601 | 0.168976 | 0.897092 | | 5.865142 | -0.13175 | 0.925747 |
| MGAT4A | 2.721145 | 0.115147 | 0.999532 | | 2.678718 | -0.23283 | 0.897092 | | 2.49107 | -0.32728 | 0.842754 |
| CD8B | 4.063567 | 0.174801 | 0.999532 | | 4.129284 | -0.22299 | 0.897092 | | 3.807788 | -0.38681 | 0.774747 |
| FGF11 | 4.03632 | -0.03423 | 0.999532 | | 4.387219 | -0.18537 | 0.897092 | | 3.562074 | -0.13165 | 0.929613 |
| EMC8 | 2.299204 | -0.0377 | 0.999532 | | 2.646244 | 0.227666 | 0.897092 | | 2.45068 | 0.288082 | 0.87251 |
| PAPOLG | 2.018146 | -0.07249 | 0.999532 | | 2.089697 | -0.24643 | 0.897092 | | 1.736142 | -0.1586 | 0.941976 |
| C19orf38 | 1.85092 | 0.326451 | 0.999532 | | 1.721896 | -0.27803 | 0.897204 | | 1.629681 | -0.58565 | 0.724363 |
| ZNF543 | 2.266542 | 0.053924 | 0.999532 | | 2.508317 | -0.2373 | 0.897225 | | 2.086874 | -0.25816 | 0.890261 |
| CARD16 | 4.344567 | -0.19278 | 0.999532 | | 4.938106 | -0.17006 | 0.897382 | | 3.996599 | 0.042835 | 0.98153 |
| DDX56 | 3.101544 | -0.11993 | 0.999532 | | 3.236144 | -0.19916 | 0.897499 | | 2.720523 | -0.06206 | 0.977473 |
| C2orf42 | 2.796824 | 0.411999 | 0.999532 | | 2.963454 | 0.222689 | 0.897511 | | 3.030861 | -0.17345 | 0.918644 |
| CHCHD6 | 1.83748 | -0.72014 | 0.999532 | | 2.448606 | -0.24921 | 0.897511 | | 1.731966 | 0.492662 | 0.795612 |
| GPX5 | 2.468212 | 0.331576 | 0.999532 | | 2.170234 | -0.25251 | 0.897511 | | 2.195451 | -0.56796 | 0.705425 |
| HEPH | 3.296419 | 0.885761 | 0.999532 | | 3.01624 | 0.221139 | 0.897527 | | 3.570313 | -0.65098 | 0.541136 |
| DBNDD1 | 2.313095 | 0.440347 | 0.999532 | | 2.034814 | -0.2548 | 0.897527 | | 2.080685 | -0.67503 | 0.644433 |
| CALHM2 | 5.576667 | -0.38791 | 0.999532 | | 7.064017 | -0.17433 | 0.897527 | | 5.132826 | 0.233056 | 0.859522 |
| CD2BP2 | 2.632998 | -0.04787 | 0.999532 | | 2.703421 | -0.22399 | 0.8977 | | 2.290402 | -0.15975 | 0.931749 |
| GJB2 | 2.263776 | -0.04435 | 0.999532 | | 2.769725 | 0.223284 | 0.897706 | | 2.254998 | 0.283628 | 0.86549 |
| DDX41 | 2.842717 | -0.12786 | 0.999532 | | 3.549504 | 0.195769 | 0.89778 | | 3.03782 | 0.340749 | 0.808547 |
| EPGN | 2.261387 | 0.120164 | 0.999532 | | 2.289326 | -0.25227 | 0.89778 | | 2.043231 | -0.35679 | 0.837256 |
| PHETA1 | 2.463709 | -0.28772 | 0.999532 | | 2.848686 | -0.22175 | 0.898025 | | 2.273026 | 0.082085 | 0.972758 |
| MYOT | 2.08776 | -0.31767 | 0.999532 | | 2.299838 | -0.24231 | 0.898335 | | 1.916167 | 0.092615 | 0.972305 |
| BEST4 | 2.917715 | -0.0118 | 0.999532 | | 3.02064 | -0.21928 | 0.898455 | | 2.623238 | -0.18854 | 0.916989 |
| KATNA1 | 2.903535 | 0.226252 | 0.999532 | | 2.780165 | -0.21772 | 0.898496 | | 2.449176 | -0.42749 | 0.752299 |
| FBXO30 | 3.129005 | 0.353819 | 0.999532 | | 3.300871 | 0.209766 | 0.898531 | | 3.115382 | -0.12429 | 0.942744 |
| RAPGEF2 | 6.221305 | -0.27432 | 0.999532 | | 7.163424 | -0.16083 | 0.898531 | | 5.762218 | 0.117003 | 0.9402 |
| CTSW | 2.724013 | 0.072763 | 0.999532 | | 2.855147 | -0.22391 | 0.898531 | | 2.567575 | -0.27433 | 0.872435 |
| NTNG2 | 2.420353 | 0.498823 | 0.999532 | | 1.988919 | -0.25988 | 0.898531 | | 2.24301 | -0.74364 | 0.603483 |
| TMEM16 | 3.025806 | -0.23592 | 0.999532 | | 3.625807 | -0.19721 | 0.898531 | | 2.791967 | 0.059768 | 0.977746 |
| EDEM2 | 3.903893 | 0.097157 | 0.999532 | | 4.27758 | 0.176322 | 0.898531 | | 3.801588 | 0.093525 | 0.955167 |
| ZNF256 | 2.427539 | -0.33215 | 0.999532 | | 2.782887 | -0.23057 | 0.898771 | | 2.141132 | 0.120345 | 0.958191 |
| ZFYVE16 | 5.309253 | -0.15425 | 0.999532 | | 6.475302 | 0.157361 | 0.898952 | | 5.390597 | 0.318321 | 0.77448 |
| SNTA1 | 4.234372 | -0.06929 | 0.999532 | | 5.380491 | 0.165741 | 0.898952 | | 4.377462 | 0.258309 | 0.833604 |
| ATP11C | 3.691198 | -0.31808 | 0.999532 | | 4.758307 | 0.169796 | 0.898952 | | 3.983613 | 0.521435 | 0.633741 |
| SLC35F3 | 2.909853 | 0.223385 | 0.999532 | | 2.572073 | -0.23347 | 0.898952 | | 2.552854 | -0.43706 | 0.771685 |
| TLR4 | 4.970932 | -0.29759 | 0.999532 | | 6.359678 | 0.158542 | 0.898952 | | 4.923585 | 0.474976 | 0.619615 |
| COLEC10 | 2.017594 | 0.780635 | 0.999532 | | 1.772142 | 0.278529 | 0.898952 | | 2.12733 | -0.48873 | 0.766067 |
| GTPBP1 | 6.765931 | 0.028698 | 0.999532 | | 7.486754 | 0.1399 | 0.898952 | | 6.600759 | 0.110912 | 0.929473 |
| G6PC3 | 4.474885 | 0.106371 | 0.999532 | | 4.413795 | -0.17558 | 0.898952 | | 3.934501 | -0.26886 | 0.828869 |
| PREX2 | 4.486992 | -0.32131 | 0.999532 | | 5.194119 | -0.18102 | 0.898952 | | 4.220575 | 0.15262 | 0.926957 |
| TENT5C | 2.885528 | 0.314848 | 0.999532 | | 2.79016 | -0.21993 | 0.898952 | | 2.639782 | -0.50824 | 0.714722 |
| MOCS1 | 2.841235 | -0.17234 | 0.999532 | | 3.183127 | -0.20545 | 0.899056 | | 2.554926 | -0.01475 | 0.996681 |
| H2BC8 | 3.462269 | -0.56618 | 0.999532 | | 4.277226 | -0.18102 | 0.899056 | | 2.963637 | 0.398455 | 0.765852 |
| TM6SF1 | 2.807404 | 0.178302 | 0.999532 | | 3.026596 | 0.221301 | 0.899056 | | 2.853618 | 0.055593 | 0.981739 |
| CINP | 2.170997 | 0.188734 | 0.999532 | | 2.434736 | 0.23688 | 0.899056 | | 2.129418 | 0.065868 | 0.978282 |
| SYS1 | 2.646902 | 0.019479 | 0.999532 | | 3.038766 | 0.219404 | 0.899056 | | 2.70685 | 0.204381 | 0.912355 |
| SNRPE | 8.457439 | 0.481568 | 0.999532 | | 7.85356 | -0.14543 | 0.899056 | | 7.975598 | -0.60272 | 0.387493 |
| CA5B | 2.984963 | -0.14993 | 0.999532 | | 3.88819 | 0.200748 | 0.899162 | | 3.18982 | 0.378816 | 0.798849 |
| ARPIN | 2.922909 | -0.03308 | 0.999532 | | 3.815289 | 0.204442 | 0.899162 | | 3.314696 | 0.24548 | 0.872808 |
| ANKRD12 | 3.246904 | -0.12929 | 0.999532 | | 4.098404 | 0.191214 | 0.899162 | | 3.418564 | 0.345404 | 0.791576 |
| RUVBL1 | 4.364994 | 0.367357 | 0.999532 | | 4.504657 | 0.17227 | 0.899162 | | 4.386344 | -0.17567 | 0.895421 |
| DZIP1L | 3.594884 | -0.25508 | 0.999532 | | 4.090511 | -0.18902 | 0.899162 | | 3.079745 | 0.082008 | 0.969041 |
| ACOX3 | 4.009888 | 0.183343 | 0.999532 | | 3.67881 | -0.1888 | 0.899162 | | 3.504032 | -0.35926 | 0.7719 |
| STAG2 | 4.492602 | -0.12375 | 0.999532 | | 5.25782 | 0.177358 | 0.899162 | | 4.516062 | 0.302973 | 0.806797 |
| SPAG16 | 3.077335 | 0.638888 | 0.999532 | | 3.069435 | 0.209147 | 0.899162 | | 3.156178 | -0.41328 | 0.730921 |
| NUB1 | 5.769197 | -0.01175 | 0.999532 | | 6.259008 | -0.15477 | 0.899162 | | 5.296237 | -0.12165 | 0.927047 |
| MYDGF | 5.222147 | -0.06371 | 0.999532 | | 6.342754 | 0.163123 | 0.899162 | | 5.300961 | 0.244988 | 0.824012 |
| IP6K2 | 14.71532 | 0.144976 | 0.999532 | | 16.36257 | 0.104962 | 0.899162 | | 14.21005 | -0.0226 | 0.985675 |
| SYN3 | 2.481954 | 0.464798 | 0.999532 | | 2.186456 | -0.25461 | 0.899162 | | 2.271065 | -0.70304 | 0.628053 |
| SOWAHB | 1.958536 | 0.312189 | 0.999532 | | 1.77685 | -0.27478 | 0.899162 | | 1.915754 | -0.57404 | 0.736481 |
| CTXND1 | 2.597219 | 0.087127 | 0.999532 | | 2.585478 | -0.23221 | 0.899162 | | 2.402705 | -0.30706 | 0.862564 |
| ARFGAP | 5.174355 | -0.46978 | 0.999532 | | 6.383972 | -0.15181 | 0.899162 | | 4.614188 | 0.337517 | 0.754277 |
| CPE | 2.212234 | 0.796544 | 0.999532 | | 2.118851 | 0.247743 | 0.899162 | | 2.319659 | -0.53061 | 0.702908 |
| MCM3 | 3.401805 | 0.266356 | 0.999532 | | 3.636617 | 0.192215 | 0.899162 | | 3.363137 | -0.05737 | 0.976783 |
| PTGDS | 13.79461 | 0.563681 | 0.999532 | | 13.80692 | 0.168417 | 0.899162 | | 13.7762 | -0.33584 | 0.705326 |
| PSMB9 | 9.761789 | -0.18689 | 0.999532 | | 12.22358 | 0.151838 | 0.899162 | | 9.918421 | 0.366204 | 0.68232 |
| NOP10 | 6.299993 | -0.25694 | 0.999532 | | 8.100807 | 0.141846 | 0.899162 | | 6.479645 | 0.418129 | 0.636291 |
| ZNF365 | 1.934815 | 0.538949 | 0.999532 | | 2.116053 | 0.26252 | 0.899162 | | 2.081907 | -0.25592 | 0.892328 |
| IGFBP4 | 67.17692 | -0.24895 | 0.999532 | | 79.94966 | -0.07173 | 0.899162 | | 62.39007 | 0.199066 | 0.65401 |
| EIF4G1 | 6.951372 | -0.32933 | 0.999532 | | 9.078966 | 0.14075 | 0.899162 | | 7.0135 | 0.488479 | 0.566812 |
| DNAJC24 | 3.177112 | 0.231785 | 0.999532 | | 2.92937 | -0.21063 | 0.899162 | | 2.782975 | -0.42504 | 0.747457 |
| DHRS2 | 3.43611 | 0.487773 | 0.999532 | | 3.381615 | 0.200349 | 0.899162 | | 3.686911 | -0.2645 | 0.849682 |
| EDEM3 | 3.933333 | -0.08922 | 0.999532 | | 4.441506 | -0.17856 | 0.899162 | | 3.491012 | -0.06887 | 0.972031 |
| C14orf119 | 6.204441 | 0.127318 | 0.999532 | | 6.07798 | -0.15026 | 0.899162 | | 5.494645 | -0.26857 | 0.801905 |
| PIK3C2B | 5.44493 | -0.22102 | 0.999532 | | 6.291282 | -0.15549 | 0.899162 | | 5.066892 | 0.080663 | 0.956011 |
| KRTAP4- | 2.078726 | -0.22157 | 0.999532 | | 2.322059 | -0.24353 | 0.899162 | | 1.946168 | -0.00365 | 0.999385 |
| ORAI2 | 2.856207 | -0.53965 | 0.999532 | | 3.370502 | -0.19809 | 0.899162 | | 2.525362 | 0.362004 | 0.809832 |
| FAM102B | 3.021625 | -0.33138 | 0.999532 | | 3.314983 | -0.20966 | 0.899619 | | 2.718128 | 0.128005 | 0.948099 |
| RBFOX3 | 2.370788 | -0.07945 | 0.999532 | | 2.693664 | -0.25146 | 0.899619 | | 2.15033 | -0.15148 | 0.944262 |
| TMEM150 | 3.078531 | -0.17642 | 0.999532 | | 3.740577 | 0.196657 | 0.899916 | | 3.064878 | 0.387214 | 0.780676 |
| SNX9 | 5.102819 | 0.012205 | 0.999532 | | 6.228334 | 0.153112 | 0.899916 | | 5.306771 | 0.162317 | 0.892869 |
| KL | 5.595544 | 0.421741 | 0.999532 | | 5.372836 | -0.16655 | 0.899916 | | 5.081343 | -0.56154 | 0.57215 |
| ACADS | 3.357491 | 0.434925 | 0.999532 | | 3.218058 | 0.204577 | 0.899916 | | 3.230066 | -0.22093 | 0.88476 |
| LCMT2 | 2.918513 | 0.093886 | 0.999532 | | 3.059881 | -0.20973 | 0.899916 | | 2.690043 | -0.28264 | 0.858705 |
| DACH1 | 24.98911 | 0.446829 | 0.999532 | | 25.21245 | 0.142425 | 0.899916 | | 24.56393 | -0.28541 | 0.67937 |
| LYRM4 | 3.629226 | 0.518813 | 0.999532 | | 3.44128 | 0.203344 | 0.899916 | | 3.64032 | -0.30449 | 0.825942 |
| GNL3L | 4.471027 | 0.070985 | 0.999532 | | 4.727092 | -0.16826 | 0.899916 | | 4.15592 | -0.22022 | 0.861737 |
| ADCK2 | 2.672388 | 0.208073 | 0.999532 | | 2.934639 | 0.234462 | 0.899916 | | 2.591699 | 0.062608 | 0.977473 |
| HSCB | 2.762123 | -0.06327 | 0.999532 | | 3.021463 | -0.20904 | 0.899916 | | 2.39376 | -0.12676 | 0.946512 |
| LSM11 | 2.681828 | -0.34101 | 0.999532 | | 3.078879 | -0.20986 | 0.899916 | | 2.550884 | 0.148837 | 0.940162 |
| TMEM170 | 3.514066 | 0.118718 | 0.999532 | | 3.568399 | -0.19059 | 0.899916 | | 3.209122 | -0.29103 | 0.827267 |
| AOPEP | 2.434316 | 0.325115 | 0.999532 | | 2.469138 | 0.231309 | 0.899978 | | 2.550858 | -0.07363 | 0.974421 |
| KIAA0753 | 3.343656 | 0.108254 | 0.999532 | | 3.763036 | 0.193701 | 0.899978 | | 3.287059 | 0.105999 | 0.950711 |
| GCHFR | 2.260904 | 0.292364 | 0.999532 | | 2.027546 | -0.25259 | 0.899978 | | 2.044476 | -0.52796 | 0.735469 |
| PPP2R2D | 6.92535 | -0.02027 | 0.999532 | | 7.989088 | 0.134634 | 0.899978 | | 6.804487 | 0.17313 | 0.868512 |
| IL1B | 3.202212 | 0.22527 | 0.999532 | | 2.969378 | -0.24004 | 0.899978 | | 2.996012 | -0.41882 | 0.772025 |
| ZNF662 | 1.941146 | 0.60552 | 0.999532 | | 1.885428 | 0.257985 | 0.900177 | | 2.037973 | -0.32791 | 0.853549 |
| SVOPL | 3.224854 | 0.458677 | 0.999532 | | 3.068504 | -0.21941 | 0.900177 | | 3.134426 | -0.66196 | 0.572297 |
| POLR1B | 3.364704 | -0.5373 | 0.999532 | | 4.154796 | -0.19891 | 0.900237 | | 3.032636 | 0.35418 | 0.816372 |
| C16orf54 | 2.101909 | 0.200639 | 0.999532 | | 2.100756 | -0.25655 | 0.900237 | | 1.954355 | -0.43916 | 0.79686 |
| NFYC | 4.422428 | 0.16409 | 0.999532 | | 4.851273 | 0.172326 | 0.900237 | | 4.428588 | 0.022194 | 0.992286 |
| NCOR1 | 3.734162 | -0.14523 | 0.999532 | | 4.916552 | 0.177717 | 0.900237 | | 3.892111 | 0.343055 | 0.7719 |
| SSC5D | 3.466056 | 0.071546 | 0.999532 | | 3.964562 | 0.181349 | 0.900237 | | 3.457631 | 0.127906 | 0.93452 |
| ZCCHC4 | 2.949208 | 0.230136 | 0.999532 | | 2.799184 | -0.21449 | 0.900237 | | 2.635912 | -0.42731 | 0.752562 |
| ZNF660 | 2.386732 | 0.186675 | 0.999532 | | 2.17505 | -0.24333 | 0.900398 | | 2.077112 | -0.41853 | 0.803217 |
| ARHGEF | 2.168512 | 0.541546 | 0.999532 | | 2.182397 | 0.251742 | 0.90057 | | 2.293626 | -0.27215 | 0.881837 |
| S1PR4 | 2.177468 | 0.523343 | 0.999532 | | 1.900977 | -0.26388 | 0.900717 | | 1.959593 | -0.77129 | 0.572297 |
| FNDC3A | 5.288941 | 0.305118 | 0.999532 | | 5.475122 | 0.173426 | 0.900717 | | 5.330587 | -0.12879 | 0.923864 |
| MASTL | 3.127418 | -0.14138 | 0.999532 | | 3.419324 | -0.20096 | 0.900717 | | 2.758646 | -0.04188 | 0.987274 |
| CYB561D | 3.401329 | -0.20479 | 0.999532 | | 3.803252 | -0.1865 | 0.900717 | | 3.106844 | 0.032228 | 0.990309 |
| PEX3 | 5.387796 | -0.12374 | 0.999532 | | 6.272935 | -0.15412 | 0.900717 | | 5.088193 | -0.00962 | 0.996891 |
| BRD4 | 15.16338 | 0.002658 | 0.999532 | | 17.6674 | 0.110038 | 0.900717 | | 15.11854 | 0.129321 | 0.881771 |
| TCF3 | 2.97319 | 0.270605 | 0.999532 | | 3.093373 | 0.201753 | 0.900717 | | 3.094737 | -0.05653 | 0.978312 |
| NATD1 | 2.70966 | -0.17064 | 0.999532 | | 2.984216 | -0.21441 | 0.900717 | | 2.508077 | -0.02628 | 0.993618 |
| WIPF1 | 4.337801 | -0.21267 | 0.999532 | | 5.573562 | 0.167876 | 0.900717 | | 4.409798 | 0.399189 | 0.713076 |
| DSTYK | 4.374974 | 0.453122 | 0.999532 | | 3.943093 | -0.19328 | 0.900717 | | 3.985479 | -0.6261 | 0.53691 |
| MRTO4 | 3.507203 | -0.5936 | 0.999532 | | 4.75513 | -0.1878 | 0.900717 | | 3.231932 | 0.424232 | 0.740163 |
| NME7 | 2.925518 | -0.17277 | 0.999532 | | 3.225255 | -0.19939 | 0.900717 | | 2.549929 | -0.01 | 0.996891 |
| DNAJC18 | 2.390236 | -0.09848 | 0.999532 | | 2.673476 | -0.21673 | 0.900794 | | 2.21433 | -0.09886 | 0.96382 |
| PRDM4 | 3.794191 | 0.151275 | 0.999532 | | 4.46997 | 0.172797 | 0.900801 | | 3.879731 | 0.043106 | 0.982124 |
| PDZD3 | 2.559199 | 0.261513 | 0.999532 | | 2.833461 | 0.236018 | 0.900801 | | 2.57482 | -0.00152 | 0.999463 |
| CTNS | 2.620265 | -0.30819 | 0.999532 | | 3.026656 | -0.20944 | 0.900801 | | 2.355109 | 0.117951 | 0.955834 |
| L3HYPDH | 2.567393 | 0.254733 | 0.999532 | | 2.715846 | 0.21671 | 0.900801 | | 2.535967 | -0.02077 | 0.995112 |
| NLRP6 | 2.458766 | 0.195265 | 0.999532 | | 2.424578 | -0.25017 | 0.900801 | | 2.243242 | -0.42787 | 0.800239 |
| GH2 | 2.980629 | 0.347017 | 0.999532 | | 3.334406 | 0.206921 | 0.900873 | | 3.156068 | -0.11338 | 0.950895 |
| EEF1AKM | 3.392324 | 0.358551 | 0.999532 | | 3.270515 | -0.22843 | 0.900873 | | 3.076755 | -0.56677 | 0.644433 |
| MIIP | 1.937476 | 0.295996 | 0.999532 | | 1.78991 | -0.26876 | 0.900873 | | 1.721084 | -0.54714 | 0.740755 |
| RDH12 | 1.583796 | -0.87765 | 0.999532 | | 1.998764 | -0.24811 | 0.900873 | | 1.434245 | 0.645587 | 0.735286 |
| SGIP1 | 3.387382 | 0.17958 | 0.999532 | | 3.433544 | -0.21602 | 0.900873 | | 2.953401 | -0.38469 | 0.780676 |
| NAB2 | 4.210597 | 0.245095 | 0.999532 | | 4.07916 | -0.18305 | 0.900873 | | 3.771442 | -0.40559 | 0.723933 |
| CRCP | 4.396342 | -0.19343 | 0.999532 | | 5.90058 | 0.158245 | 0.900873 | | 4.557785 | 0.371106 | 0.720106 |
| CYFIP1 | 8.380536 | -0.26721 | 0.999532 | | 11.15551 | 0.125129 | 0.900873 | | 8.663584 | 0.411445 | 0.578798 |
| SLC35F6 | 4.33239 | -0.00834 | 0.999532 | | 5.105006 | 0.161098 | 0.900873 | | 4.268384 | 0.184859 | 0.88476 |
| SURF1 | 5.828254 | -0.061 | 0.999532 | | 6.726692 | 0.14199 | 0.90094 | | 5.881759 | 0.219302 | 0.836935 |
| ZNF700 | 2.122664 | -0.23293 | 0.999532 | | 2.367466 | -0.2329 | 0.90094 | | 1.907318 | 0.017817 | 0.996681 |
| TRPC3 | 2.373873 | 0.149599 | 0.999532 | | 2.32739 | -0.24193 | 0.90094 | | 2.257569 | -0.37893 | 0.827499 |
| ATP6V0A | 3.558085 | 0.472961 | 0.999532 | | 3.573344 | 0.223314 | 0.90094 | | 3.391355 | -0.2687 | 0.907247 |
| CCDC183 | 2.729514 | -0.19727 | 0.999532 | | 2.993486 | -0.21573 | 0.90094 | | 2.5349 | 0.0032 | 0.999385 |
| UBASH3B | 2.280009 | 0.183687 | 0.999532 | | 2.057778 | -0.25039 | 0.90094 | | 2.026057 | -0.41828 | 0.810276 |
| FAM71C | 1.837203 | 0.083535 | 0.999532 | | 1.828459 | -0.27007 | 0.90094 | | 1.622107 | -0.33551 | 0.872435 |
| CCN1 | 6.14763 | -0.59861 | 0.999532 | | 7.783813 | -0.20642 | 0.90094 | | 5.284859 | 0.42645 | 0.709825 |
| ZEB2 | 9.608146 | -0.20447 | 0.999532 | | 11.98361 | 0.151871 | 0.90094 | | 9.771271 | 0.387881 | 0.644433 |
| FASN | 4.076955 | 0.233443 | 0.999532 | | 4.532905 | 0.178334 | 0.90094 | | 4.191006 | -0.03647 | 0.987062 |
| NEUROG | 3.356699 | 0.15786 | 0.999532 | | 3.545367 | -0.217 | 0.90094 | | 3.211897 | -0.34607 | 0.829716 |
| ZNF559 | 3.099739 | -0.02487 | 0.999532 | | 3.801071 | 0.188488 | 0.90094 | | 3.106162 | 0.233283 | 0.880375 |
| KIF13B | 3.42832 | 0.491321 | 0.999532 | | 3.291427 | 0.196243 | 0.90094 | | 3.354656 | -0.27912 | 0.837256 |
| AKAP5 | 2.853605 | 0.232916 | 0.999532 | | 3.316325 | 0.209838 | 0.90094 | | 3.149216 | 0.00098 | 0.999635 |
| SYT11 | 4.070091 | -0.13471 | 0.999532 | | 5.227197 | 0.174423 | 0.90094 | | 4.296734 | 0.326821 | 0.791466 |
| POU2F1 | 4.100924 | -0.14228 | 0.999532 | | 4.339289 | -0.17494 | 0.90094 | | 3.646727 | -0.01707 | 0.996149 |
| MARCHF | 10.50564 | -0.35475 | 0.999532 | | 13.87889 | 0.122329 | 0.90094 | | 10.55936 | 0.498964 | 0.455544 |
| ZFX | 3.449808 | 0.355759 | 0.999532 | | 3.289808 | 0.195153 | 0.90094 | | 3.348497 | -0.15349 | 0.924131 |
| DSC2 | 2.105687 | 0.089457 | 0.999532 | | 2.284135 | 0.239534 | 0.90094 | | 2.127239 | 0.173801 | 0.931282 |
| TTC13 | 2.417764 | -0.57028 | 0.999532 | | 2.992661 | -0.20209 | 0.90094 | | 2.053304 | 0.383883 | 0.806797 |
| CCDC113 | 1.822332 | 0.18768 | 0.999532 | | 1.954442 | 0.25439 | 0.90094 | | 1.943927 | 0.08638 | 0.974421 |
| IKZF5 | 3.776309 | -0.19508 | 0.999532 | | 4.240637 | -0.18746 | 0.90094 | | 3.120483 | 0.018381 | 0.995112 |
| PITPNC1 | 4.304872 | -0.09002 | 0.999532 | | 5.236499 | 0.180943 | 0.90094 | | 4.342146 | 0.294921 | 0.806853 |
| ARL5A | 3.282912 | -0.08247 | 0.999532 | | 3.48999 | -0.2071 | 0.90094 | | 3.042236 | -0.11789 | 0.949669 |
| RNF44 | 6.861219 | 0.38579 | 0.999532 | | 6.836836 | 0.143653 | 0.90094 | | 6.799939 | -0.22784 | 0.836453 |
| DPYD | 4.042148 | -0.20095 | 0.999532 | | 4.632027 | -0.16637 | 0.90094 | | 3.586038 | 0.052776 | 0.977473 |
| HS3ST1 | 2.114986 | -0.27386 | 0.999532 | | 2.437802 | -0.22445 | 0.90094 | | 1.920644 | 0.067883 | 0.97843 |
| WARS1 | 3.37411 | 0.331681 | 0.999532 | | 3.391333 | 0.193602 | 0.90094 | | 3.353172 | -0.12119 | 0.942743 |
| ZFYVE26 | 3.857968 | 0.085409 | 0.999532 | | 3.919464 | -0.19316 | 0.90094 | | 3.578509 | -0.25843 | 0.861119 |
| OPN1MW | 2.17353 | 0.421151 | 0.999532 | | 2.292751 | 0.245954 | 0.90094 | | 2.28544 | -0.15526 | 0.941197 |
| CYP2E1 | 2.571016 | -0.50875 | 0.999532 | | 3.739007 | 0.216205 | 0.90094 | | 2.95315 | 0.719661 | 0.613126 |

| | | | | | | | | | | | |
|---|---|---|---|---|---|---|---|---|---|---|---|
| CXCR2 | 2.92129 | 0.442807 | 0.999532 | | 2.561218 | -0.23316 | 0.90094 | | 2.696444 | -0.66959 | 0.617899 |
| ZNF615 | 3.11938 | 0.093476 | 0.999532 | | 3.364346 | 0.193673 | 0.90094 | | 3.202221 | 0.117756 | 0.946958 |
| METTL14 | 3.89607 | 0.219706 | 0.999532 | | 3.808896 | -0.19468 | 0.90094 | | 3.385373 | -0.3977 | 0.745628 |
| SREBF2 | 10.74951 | -0.01172 | 0.999532 | | 12.56375 | 0.145345 | 0.90094 | | 10.56727 | 0.179475 | 0.87591 |
| SCN8A | 1.906961 | -0.25805 | 0.999532 | | 2.391398 | 0.226255 | 0.90094 | | 2.00798 | 0.503398 | 0.75311 |
| PIP4K2A | 11.57305 | 0.044036 | 0.999532 | | 13.36617 | 0.123069 | 0.90094 | | 11.67922 | 0.09867 | 0.921808 |
| EFHC1 | 2.822465 | 0.220432 | 0.999532 | | 3.18829 | 0.201067 | 0.90094 | | 2.925308 | -0.00167 | 0.999429 |
| HAND2 | 1.836506 | 0.487554 | 0.999532 | | 1.92785 | 0.261357 | 0.90094 | | 1.974434 | -0.20578 | 0.920181 |
| NHLRC2 | 5.697331 | -0.41299 | 0.999532 | | 6.65606 | -0.15607 | 0.90094 | | 5.019327 | 0.275723 | 0.806394 |
| LCORL | 2.851455 | -0.24036 | 0.999532 | | 3.71691 | 0.188479 | 0.90094 | | 2.882368 | 0.445744 | 0.722703 |
| SEMA4D | 3.139349 | -0.71758 | 0.999532 | | 4.129505 | -0.18159 | 0.90094 | | 2.575148 | 0.562615 | 0.663633 |
| YIPF2 | 2.689381 | -0.05651 | 0.999532 | | 3.010012 | 0.230115 | 0.90094 | | 2.607914 | 0.293882 | 0.8691 |
| B3GALT4 | 3.227581 | -0.20873 | 0.999532 | | 3.788171 | -0.18942 | 0.90094 | | 3.077863 | 0.047521 | 0.984588 |
| SEMA6C | 5.060857 | 0.495171 | 0.999532 | | 4.978002 | 0.178589 | 0.90094 | | 5.103545 | -0.29852 | 0.79622 |
| SVEP1 | 2.661758 | -0.03108 | 0.999532 | | 3.25535 | 0.199121 | 0.90094 | | 2.735263 | 0.249615 | 0.873714 |
| SLC22A1 | 2.608367 | 0.24344 | 0.999532 | | 2.318303 | -0.23931 | 0.90094 | | 2.256726 | -0.47386 | 0.760473 |
| ELP2 | 4.152193 | 0.602319 | 0.999532 | | 3.822209 | 0.184394 | 0.90094 | | 4.048048 | -0.39804 | 0.715437 |
| SPRR2A | 2.473226 | -0.12383 | 0.999532 | | 2.786467 | -0.25851 | 0.90094 | | 2.345692 | -0.13568 | 0.954538 |
| S1PR1 | 3.329317 | -0.42518 | 0.999532 | | 4.562269 | 0.174022 | 0.90094 | | 3.556742 | 0.61814 | 0.557909 |
| LRRC23 | 2.426302 | 0.386555 | 0.999532 | | 2.520652 | 0.23179 | 0.90094 | | 2.392529 | -0.13636 | 0.945456 |
| ORC4 | 3.110772 | 0.381243 | 0.999532 | | 3.270296 | 0.203046 | 0.90094 | | 3.200665 | -0.16041 | 0.918492 |
| CRH | 1.744223 | 0.503845 | 0.999532 | | 1.808968 | 0.272789 | 0.90094 | | 1.864192 | -0.21131 | 0.921 |
| CLMN | 3.208612 | 0.311834 | 0.999532 | | 3.069484 | -0.20657 | 0.90094 | | 2.809305 | -0.4941 | 0.699971 |
| TCFL5 | 2.076021 | 0.512381 | 0.999532 | | 2.209469 | 0.24119 | 0.90094 | | 2.220965 | -0.25044 | 0.88885 |
| KLHDC8A | 2.485383 | 0.227609 | 0.999532 | | 2.323525 | -0.24162 | 0.90094 | | 2.123521 | -0.45088 | 0.776133 |
| CDCA2 | 2.246177 | 0.422777 | 0.999532 | | 2.347354 | 0.241074 | 0.90094 | | 2.304399 | -0.16491 | 0.932589 |
| HPS3 | 3.995688 | 0.530031 | 0.999532 | | 3.976786 | 0.185239 | 0.90094 | | 3.996678 | -0.32866 | 0.774747 |
| TMED8 | 2.287733 | -0.07809 | 0.999532 | | 2.327491 | -0.23317 | 0.90094 | | 1.954578 | -0.14544 | 0.946572 |
| MOXD1 | 2.569768 | -0.2439 | 0.999532 | | 2.819951 | -0.21557 | 0.90094 | | 2.305611 | 0.045069 | 0.988328 |
| SLC8B1 | 3.656987 | -0.19134 | 0.999532 | | 3.928224 | -0.18765 | 0.90094 | | 3.338831 | 0.014761 | 0.996681 |
| KAT6B | 3.522076 | -0.05029 | 0.999532 | | 4.378672 | 0.17707 | 0.901056 | | 3.735229 | 0.247744 | 0.859898 |
| MFSD14B | 4.334037 | 0.000679 | 0.999532 | | 5.228206 | 0.167334 | 0.901056 | | 4.491949 | 0.182963 | 0.88885 |
| MICALL1 | 3.731599 | 0.243324 | 0.999532 | | 4.408205 | 0.187021 | 0.901056 | | 3.958303 | -0.04127 | 0.985675 |
| GOLGA7B | 2.922542 | -0.45159 | 0.999532 | | 3.446293 | -0.19284 | 0.901056 | | 2.551007 | 0.27725 | 0.862823 |
| STRIP2 | 1.984103 | 0.419926 | 0.999532 | | 2.025283 | 0.243333 | 0.901056 | | 2.062524 | -0.15766 | 0.9402 |
| GSTA2 | 3.713467 | 0.074145 | 0.999532 | | 3.881204 | -0.20151 | 0.901289 | | 3.404893 | -0.26712 | 0.849848 |
| C16orf74 | 2.132033 | -0.57126 | 0.999532 | | 2.593433 | -0.22983 | 0.901391 | | 1.863822 | 0.359929 | 0.849021 |
| RUNX1 | 2.600496 | -0.36408 | 0.999532 | | 3.559576 | 0.190355 | 0.901504 | | 2.71211 | 0.573569 | 0.655324 |
| PPP4R1 | 2.684515 | -0.08579 | 0.999532 | | 3.433126 | 0.202892 | 0.901781 | | 2.867574 | 0.304246 | 0.836012 |
| SS18 | 5.546834 | -0.02899 | 0.999532 | | 6.618119 | 0.158118 | 0.90186 | | 5.707066 | 0.20598 | 0.859898 |
| MLST8 | 3.781633 | 0.297334 | 0.999532 | | 3.42587 | -0.19273 | 0.90186 | | 3.473268 | -0.47758 | 0.697497 |
| PSMB8 | 9.474632 | -0.30512 | 0.999532 | | 12.0639 | 0.116039 | 0.90186 | | 9.565255 | 0.434128 | 0.549172 |
| CYB5D1 | 3.71067 | 0.026954 | 0.999532 | | 4.162919 | -0.18464 | 0.90186 | | 3.435162 | -0.18981 | 0.897958 |
| SNX12 | 4.574527 | 0.074605 | 0.999532 | | 4.873939 | -0.17979 | 0.90186 | | 4.016165 | -0.22592 | 0.870791 |
| LCA5 | 2.844196 | -0.18681 | 0.999532 | | 3.514759 | 0.197762 | 0.90186 | | 2.841679 | 0.403577 | 0.765852 |
| DAOA | 4.144543 | 0.447268 | 0.999532 | | 3.666623 | -0.19609 | 0.90186 | | 3.717896 | -0.62498 | 0.557909 |
| TASOR2 | 44.48175 | -0.01601 | 0.999532 | | 48.71538 | -0.0846 | 0.90186 | | 41.66314 | -0.04758 | 0.951851 |
| WDR20 | 3.192765 | -0.15739 | 0.999532 | | 3.360162 | -0.20207 | 0.90186 | | 2.800515 | -0.02977 | 0.99186 |
| NOC2L | 3.881337 | -0.28483 | 0.999532 | | 5.271727 | 0.16304 | 0.90186 | | 3.967378 | 0.467752 | 0.663104 |
| MFSD14C | 2.99818 | 0.140112 | 0.999532 | | 3.559236 | 0.197043 | 0.90186 | | 3.086937 | 0.080485 | 0.969504 |
| SPATA20 | 6.694213 | -0.43975 | 0.999532 | | 8.255069 | -0.12846 | 0.902088 | | 6.13508 | 0.328983 | 0.720576 |
| CLEC14A | 5.224184 | -0.22686 | 0.999532 | | 6.535585 | 0.168349 | 0.902139 | | 5.449544 | 0.395116 | 0.735469 |
| TMEM161 | 3.512597 | 0.064013 | 0.999532 | | 3.346318 | -0.19504 | 0.902162 | | 3.110925 | -0.2489 | 0.869224 |
| KCNMA1 | 2.691672 | 0.990458 | 0.999532 | | 2.405234 | 0.2499 | 0.902162 | | 2.89243 | -0.72802 | 0.566812 |
| TCERG1 | 3.096754 | 0.342043 | 0.999532 | | 3.148043 | 0.196443 | 0.9022 | | 3.181772 | -0.13104 | 0.939913 |
| MEN1 | 6.129875 | -0.14327 | 0.999532 | | 7.793738 | 0.139805 | 0.902233 | | 6.308738 | 0.300148 | 0.740755 |
| PRXL2A | 4.768282 | -0.3094 | 0.999532 | | 5.783795 | -0.17314 | 0.902233 | | 4.242484 | 0.177413 | 0.90539 |
| IMMP2L | 3.370024 | 0.666849 | 0.999532 | | 2.627922 | -0.22543 | 0.902233 | | 3.128025 | -0.88178 | 0.414154 |
| IL1RAP | 2.631902 | 0.092278 | 0.999532 | | 3.130927 | 0.200278 | 0.902421 | | 2.737469 | 0.127842 | 0.945129 |
| PDGFA | 2.610514 | -0.13488 | 0.999532 | | 3.120106 | 0.205041 | 0.902763 | | 2.635278 | 0.362808 | 0.807903 |
| PPIL4 | 3.916849 | -0.41147 | 0.999532 | | 4.752902 | -0.16605 | 0.902763 | | 3.437085 | 0.263532 | 0.836935 |
| ADGRA3 | 3.667122 | 0.091911 | 0.999532 | | 4.191699 | 0.174133 | 0.90283 | | 3.706047 | 0.098293 | 0.952785 |
| STIP1 | 5.945048 | -0.37743 | 0.999532 | | 7.726331 | 0.13873 | 0.902906 | | 5.953787 | 0.533056 | 0.520848 |
| DLL3 | 1.961671 | 0.344437 | 0.999532 | | 1.97576 | -0.25691 | 0.902973 | | 1.874382 | -0.58024 | 0.717138 |
| CYP3A4 | 2.096517 | 0.585642 | 0.999532 | | 1.868704 | -0.26039 | 0.902993 | | 1.873561 | -0.82451 | 0.552672 |
| GRIK2 | 2.273375 | 0.183196 | 0.999532 | | 2.525977 | 0.223413 | 0.903005 | | 2.252075 | 0.05837 | 0.980051 |
| ZNF366 | 3.802698 | 0.107482 | 0.999532 | | 3.929467 | -0.18739 | 0.903005 | | 3.496064 | -0.2758 | 0.838791 |
| CPNE1 | 5.719994 | 0.011413 | 0.999532 | | 6.601093 | 0.157137 | 0.903059 | | 5.772373 | 0.160103 | 0.903339 |
| STRIP1 | 4.039867 | 0.263319 | 0.999532 | | 4.591162 | 0.174829 | 0.903059 | | 4.098874 | -0.07624 | 0.964027 |
| FEM1B | 5.000227 | -0.23572 | 0.999532 | | 5.706258 | -0.15419 | 0.903144 | | 4.262273 | 0.101748 | 0.945129 |
| NANOGNB | 1.911999 | 0.411235 | 0.999532 | | 1.896069 | 0.262339 | 0.903221 | | 2.039233 | -0.13694 | 0.955834 |
| TULP3 | 2.329143 | 0.092341 | 0.999532 | | 2.839634 | 0.206073 | 0.903221 | | 2.43365 | 0.133566 | 0.944202 |
| ZSCAN12 | 1.952688 | 0.210216 | 0.999532 | | 2.501393 | 0.269927 | 0.903255 | | 2.302016 | 0.062693 | 0.984587 |
| SLC25A3 | 3.164271 | 0.11974 | 0.999532 | | 3.618173 | 0.197488 | 0.903353 | | 3.082345 | 0.102642 | 0.955834 |
| HSPA13 | 3.392068 | -0.8609 | 0.999532 | | 5.382266 | 0.151706 | 0.903353 | | 3.599744 | 1.031626 | 0.212897 |
| C2orf69 | 4.389149 | 0.308113 | 0.999532 | | 4.517106 | 0.163951 | 0.903409 | | 4.332679 | -0.12827 | 0.92697 |
| AHCYL2 | 3.030592 | -0.16743 | 0.999532 | | 3.324634 | -0.20165 | 0.903658 | | 2.856445 | -0.01923 | 0.996471 |
| AVPR1B | 2.134326 | -0.07924 | 0.999532 | | 2.382979 | -0.23056 | 0.903658 | | 1.978757 | -0.13144 | 0.952713 |
| TAF1D | 6.188076 | -0.28991 | 0.999532 | | 7.497806 | -0.14623 | 0.903658 | | 5.721468 | 0.16479 | 0.892869 |
| BRICD5 | 2.84756 | -0.32943 | 0.999532 | | 3.630034 | 0.19241 | 0.903658 | | 2.84654 | 0.540274 | 0.674041 |
| EIF4E | 5.296162 | -0.20637 | 0.999532 | | 5.847647 | -0.14975 | 0.903658 | | 4.674712 | 0.073912 | 0.96129 |
| ZFY | 2.066929 | 0.155833 | 0.999532 | | 2.301526 | 0.245328 | 0.903731 | | 2.149099 | 0.095193 | 0.97179 |
| PPA2 | 2.57569 | -0.04335 | 0.999532 | | 2.920227 | 0.203278 | 0.90383 | | 2.557309 | 0.258076 | 0.876613 |
| ERO1A | 5.090362 | -0.31292 | 0.999532 | | 6.285589 | 0.150789 | 0.90383 | | 5.130441 | 0.478814 | 0.640728 |
| MRPL27 | 3.894879 | -0.16843 | 0.999532 | | 4.696309 | 0.167473 | 0.90383 | | 3.980264 | 0.345401 | 0.78258 |
| GPRASP1 | 2.191683 | -0.16851 | 0.999532 | | 2.335905 | -0.23961 | 0.903885 | | 1.868409 | -0.04838 | 0.988328 |
| DNAAF11 | 1.812348 | -0.24601 | 0.999532 | | 2.250638 | 0.231271 | 0.90402 | | 1.887283 | 0.494797 | 0.765852 |
| ARFGEF2 | 5.084178 | 0.08317 | 0.999532 | | 6.100317 | 0.149787 | 0.90402 | | 5.07567 | 0.093402 | 0.946572 |
| LDB3 | 2.225151 | -0.0281 | 0.999532 | | 2.463104 | -0.22644 | 0.90402 | | 2.01253 | -0.17923 | 0.926957 |
| NT5DC1 | 6.423585 | 0.409694 | 0.999532 | | 6.559204 | 0.148974 | 0.90402 | | 6.406228 | -0.23839 | 0.8233 |
| HSD17B1 | 7.031859 | 0.175595 | 0.999532 | | 7.440272 | 0.149391 | 0.90402 | | 6.84734 | 0.003681 | 0.99858 |
| NUDCD2 | 3.832958 | 0.606085 | 0.999532 | | 3.619817 | 0.191408 | 0.90402 | | 3.930107 | -0.38735 | 0.723933 |
| SCARA3 | 4.123504 | 0.080268 | 0.999532 | | 4.265005 | -0.1837 | 0.90402 | | 3.741704 | -0.24557 | 0.859426 |
| HSPBAP1 | 2.730744 | -0.26296 | 0.999532 | | 3.773366 | 0.208842 | 0.904032 | | 2.930256 | 0.497185 | 0.722703 |
| ARID4B | 3.562998 | -0.05485 | 0.999532 | | 4.445885 | 0.171002 | 0.904032 | | 3.597682 | 0.243024 | 0.856868 |
| ZNF593 | 2.922634 | 0.033616 | 0.999532 | | 2.986519 | -0.20005 | 0.904032 | | 2.589092 | -0.21528 | 0.893853 |
| C2orf68 | 2.762358 | -0.042 | 0.999532 | | 2.847301 | -0.21118 | 0.904032 | | 2.556912 | -0.16412 | 0.93222 |
| TOB2 | 5.489268 | -0.07585 | 0.999532 | | 6.691019 | 0.14228 | 0.904032 | | 5.541642 | 0.238081 | 0.823571 |
| RBL1 | 2.754602 | 0.602514 | 0.999532 | | 2.620252 | -0.23547 | 0.904236 | | 2.613153 | -0.7964 | 0.53177 |
| DCTN5 | 5.214534 | -0.33159 | 0.999532 | | 6.283277 | -0.15749 | 0.904559 | | 4.915843 | 0.192908 | 0.881771 |
| ZNHIT6 | 3.190654 | -0.03868 | 0.999532 | | 3.943069 | 0.181762 | 0.904559 | | 3.344989 | 0.24011 | 0.868998 |
| NLGN2 | 3.773979 | 0.082665 | 0.999532 | | 4.495739 | 0.181193 | 0.904559 | | 3.829155 | 0.124306 | 0.938429 |
| TRPC4AP | 4.261246 | -0.1793 | 0.999532 | | 5.364673 | 0.157044 | 0.904559 | | 4.101768 | 0.353927 | 0.750371 |
| CWC22 | 2.829785 | -0.31594 | 0.999532 | | 3.332441 | -0.20394 | 0.904559 | | 2.581053 | 0.124439 | 0.947941 |
| TTC30B | 3.505215 | 0.320194 | 0.999532 | | 3.173134 | -0.20446 | 0.904559 | | 3.153664 | -0.51155 | 0.686214 |
| TCEA2 | 7.913061 | 0.265493 | 0.999532 | | 8.303083 | 0.14141 | 0.904559 | | 7.791907 | -0.11286 | 0.92697 |
| TAS2R41 | 2.226667 | 0.56907 | 0.999532 | | 1.753322 | -0.26446 | 0.904559 | | 2.003829 | -0.822 | 0.573023 |
| IFITM2 | 16.3358 | -0.46358 | 0.999532 | | 20.52947 | -0.09315 | 0.904559 | | 15.20837 | 0.392051 | 0.454223 |
| MEF2D | 6.855758 | -0.10942 | 0.999532 | | 8.317519 | 0.138737 | 0.904559 | | 6.810741 | 0.268932 | 0.78258 |
| ZUP1 | 2.604341 | -0.1635 | 0.999532 | | 3.057942 | -0.21737 | 0.904559 | | 2.521271 | -0.03435 | 0.991349 |
| SART3 | 3.08954 | 0.168302 | 0.999532 | | 3.483637 | 0.187816 | 0.904614 | | 3.020232 | 0.0436 | 0.985108 |
| LIN54 | 2.841041 | 0.521574 | 0.999532 | | 2.317655 | -0.21901 | 0.904661 | | 2.570077 | -0.72945 | 0.540342 |
| ITFG2 | 4.939956 | 0.280623 | 0.999532 | | 5.362804 | 0.156404 | 0.904802 | | 5.019208 | -0.10731 | 0.94109 |
| SCRIB | 4.127796 | -0.01023 | 0.999532 | | 4.730631 | 0.16395 | 0.904802 | | 4.164592 | 0.184689 | 0.895345 |
| RECK | 3.514219 | 0.1699 | 0.999532 | | 3.792696 | 0.180979 | 0.90481 | | 3.395623 | 0.027907 | 0.991289 |
| SLC10A6 | 2.463456 | 0.044098 | 0.999532 | | 2.532786 | -0.23258 | 0.90481 | | 2.229858 | -0.25971 | 0.88885 |
| NRXN3 | 2.228297 | 0.23804 | 0.999532 | | 2.472837 | 0.240564 | 0.90481 | | 2.262542 | 0.024472 | 0.99497 |
| MRGPRX | 2.625549 | -0.60719 | 0.999532 | | 3.330866 | -0.20005 | 0.90481 | | 2.422463 | 0.425206 | 0.786701 |
| PPFIA2 | 2.493405 | 0.480301 | 0.999532 | | 2.546757 | 0.242095 | 0.904834 | | 2.559936 | -0.22283 | 0.90148 |
| SSX5 | 2.319236 | 0.09948 | 0.999532 | | 2.324073 | -0.23134 | 0.904871 | | 2.147797 | -0.31431 | 0.862823 |
| CCDC59 | 8.923091 | 0.123841 | 0.999532 | | 9.965076 | 0.121269 | 0.904881 | | 8.972059 | 0.017383 | 0.992286 |
| APCS | 1.753543 | 0.277426 | 0.999532 | | 1.680755 | -0.27031 | 0.904881 | | 1.597842 | -0.52832 | 0.765852 |
| GNL1 | 3.075603 | -0.04991 | 0.999532 | | 3.191731 | -0.20046 | 0.904881 | | 2.762398 | -0.13621 | 0.9422 |
| RAG2 | 2.315945 | -0.06985 | 0.999532 | | 2.536834 | -0.23205 | 0.904881 | | 2.228052 | -0.13977 | 0.948865 |
| TFCP2 | 2.88321 | 0.117518 | 0.999532 | | 3.40933 | 0.198834 | 0.904881 | | 2.923992 | 0.103945 | 0.955834 |
| CEP250 | 3.921142 | 0.404489 | 0.999532 | | 3.527515 | -0.18647 | 0.905441 | | 3.508049 | -0.57299 | 0.595927 |
| F2RL2 | 3.83996 | -0.12281 | 0.999532 | | 3.925746 | -0.18933 | 0.905441 | | 3.388466 | -0.06958 | 0.974421 |
| ELMO1 | 3.493796 | -0.28061 | 0.999532 | | 4.175683 | -0.18738 | 0.905441 | | 3.142578 | 0.115922 | 0.947941 |
| MAP7 | 4.120892 | 0.359915 | 0.999532 | | 4.257917 | 0.173241 | 0.905441 | | 4.050644 | -0.17081 | 0.900583 |
| CDC42EP | 9.109851 | 0.035164 | 0.999532 | | 10.45211 | 0.126001 | 0.90554 | | 9.195365 | 0.103634 | 0.922437 |
| UROD | 6.274173 | 0.059902 | 0.999532 | | 7.453142 | 0.139041 | 0.905546 | | 6.389586 | 0.100125 | 0.938429 |
| SLC25A5 | 2.636869 | -0.1303 | 0.999532 | | 2.813663 | -0.20679 | 0.905546 | | 2.387365 | -0.06853 | 0.977359 |
| GRID2IP | 2.210018 | -0.15949 | 0.999532 | | 2.52575 | -0.23925 | 0.905596 | | 1.9509 | -0.06035 | 0.98353 |
| TERF2 | 3.034717 | -0.34759 | 0.999532 | | 4.118764 | 0.178266 | 0.905596 | | 3.10972 | 0.552925 | 0.659727 |
| ATG3 | 4.843747 | 0.291205 | 0.999532 | | 5.249329 | 0.172562 | 0.905596 | | 4.977114 | -0.09493 | 0.959377 |
| DESI2 | 2.246813 | 0.056692 | 0.999532 | | 2.30414 | -0.23266 | 0.905596 | | 1.965941 | -0.26924 | 0.881837 |
| MIER2 | 3.321702 | 0.136212 | 0.999532 | | 3.512137 | 0.18687 | 0.905596 | | 3.263314 | 0.06708 | 0.974421 |
| CEP57L1 | 3.356146 | 0.251492 | 0.999532 | | 3.275857 | -0.19506 | 0.905596 | | 3.181439 | -0.43384 | 0.728015 |
| FNIP2 | 4.702896 | -0.03152 | 0.999532 | | 4.817567 | -0.17071 | 0.905596 | | 4.04658 | -0.11158 | 0.941197 |
| STH | 3.391685 | 0.628991 | 0.999532 | | 3.089829 | 0.199711 | 0.905596 | | 3.405827 | -0.4093 | 0.739496 |
| OSR2 | 1.948451 | -0.2538 | 0.999532 | | 2.307314 | -0.24541 | 0.905596 | | 1.783794 | 0.028709 | 0.994347 |
| ISOC2 | 4.300131 | 0.116778 | 0.999532 | | 4.675999 | 0.171383 | 0.905596 | | 4.182933 | 0.078596 | 0.963359 |
| KCNK9 | 4.345004 | 0.764513 | 0.999532 | | 3.313369 | -0.19624 | 0.905661 | | 3.921305 | -0.94387 | 0.254511 |
| NR1I3 | 4.171249 | 0.585127 | 0.999532 | | 3.454803 | -0.19695 | 0.906135 | | 3.688541 | -0.76399 | 0.454116 |
| ZNF121 | 3.240354 | 0.023586 | 0.999532 | | 3.331829 | -0.19112 | 0.906135 | | 2.910949 | -0.19998 | 0.90016 |
| SLC10A7 | 2.974378 | -0.36898 | 0.999532 | | 3.447015 | -0.1886 | 0.906276 | | 2.792311 | 0.195464 | 0.907734 |
| FSCN3 | 1.744373 | -0.2734 | 0.999532 | | 2.091103 | -0.24456 | 0.906276 | | 1.685156 | 0.049568 | 0.988969 |
| EXTL2 | 2.166596 | 0.114593 | 0.999532 | | 2.116437 | -0.23224 | 0.906276 | | 1.945856 | -0.32874 | 0.854633 |
| PHF23 | 2.524449 | -0.43053 | 0.999532 | | 2.954763 | -0.20027 | 0.906276 | | 2.237262 | 0.245328 | 0.889271 |
| EPHB2 | 2.085639 | 0.682205 | 0.999532 | | 1.93499 | 0.249666 | 0.906276 | | 2.18185 | -0.41619 | 0.798572 |
| ZNF786 | 3.684708 | 0.342347 | 0.999532 | | 3.412861 | -0.18913 | 0.906276 | | 3.374873 | -0.51208 | 0.680097 |
| PDCD2 | 3.281451 | 0.249734 | 0.999532 | | 3.306882 | -0.1941 | 0.906276 | | 3.043271 | -0.41805 | 0.753585 |
| MAP10 | 2.172448 | 0.085425 | 0.999532 | | 2.09207 | -0.23656 | 0.906276 | | 1.929348 | -0.31226 | 0.872435 |
| P2RY12 | 1.876739 | 0.009012 | 0.999532 | | 1.861201 | -0.25062 | 0.906276 | | 1.776778 | -0.23521 | 0.912549 |
| TP53INP1 | 2.313519 | -0.68617 | 0.999532 | | 3.249994 | 0.19027 | 0.906276 | | 2.365195 | 0.90073 | 0.458377 |
| CXCL3 | 2.130031 | 0.224362 | 0.999532 | | 1.986024 | -0.2434 | 0.906276 | | 1.922604 | -0.44654 | 0.794473 |
| TTPAL | 2.169298 | -0.62437 | 0.999532 | | 2.811735 | -0.2177 | 0.906276 | | 1.901215 | 0.429144 | 0.806394 |
| TRIM68 | 3.009845 | 0.064347 | 0.999532 | | 3.394067 | 0.194537 | 0.906276 | | 2.96602 | 0.151957 | 0.92697 |
| IFT88 | 3.07702 | 0.148275 | 0.999532 | | 3.554397 | 0.183106 | 0.906276 | | 3.289409 | 0.055491 | 0.978229 |
| KLRG2 | 4.521391 | -0.1118 | 0.999532 | | 4.947862 | -0.16391 | 0.906276 | | 4.064836 | -0.03409 | 0.98811 |
| LRIT1 | 2.283041 | 0.008474 | 0.999532 | | 2.524083 | -0.23146 | 0.906276 | | 2.103325 | -0.2139 | 0.913156 |
| VPS11 | 3.789099 | -0.00314 | 0.999532 | | 4.446888 | 0.172551 | 0.906276 | | 3.938187 | 0.189061 | 0.895865 |
| XKR6 | 2.34166 | -0.52178 | 0.999532 | | 2.942525 | -0.20744 | 0.906276 | | 2.157378 | 0.338984 | 0.851503 |
| UGGT2 | 2.204992 | 0.260297 | 0.999532 | | 2.136044 | -0.2489 | 0.906276 | | 1.912826 | -0.4904 | 0.760473 |
| PKP1 | 2.053148 | 0.102207 | 0.999532 | | 1.981315 | -0.25045 | 0.906276 | | 1.890805 | -0.34108 | 0.86209 |
| EDNRA | 5.210232 | -0.60401 | 0.999532 | | 7.482837 | 0.137711 | 0.906276 | | 5.280563 | 0.761389 | 0.292884 |
| RBM27 | 3.258929 | -0.45766 | 0.999532 | | 3.841336 | -0.18228 | 0.906276 | | 2.80386 | 0.293541 | 0.842754 |
| SQOR | 5.696314 | 0.138113 | 0.999532 | | 6.493159 | 0.155346 | 0.906276 | | 5.7192 | 0.036237 | 0.985306 |
| WDR75 | 3.17651 | 0.353342 | 0.999532 | | 3.233978 | 0.191037 | 0.906276 | | 3.176569 | -0.14359 | 0.927433 |
| PTPN20 | 2.126541 | 0.86418 | 0.999532 | | 1.720449 | 0.259585 | 0.906276 | | 2.178931 | -0.58877 | 0.70375 |
| ZBTB37 | 2.980313 | -0.12406 | 0.999532 | | 3.642098 | 0.186054 | 0.906276 | | 2.914457 | 0.324356 | 0.823571 |
| GAPDHS | 2.380262 | 0.226035 | 0.999532 | | 2.341049 | -0.23055 | 0.906276 | | 2.1782 | -0.43547 | 0.789778 |
| RAB6B | 2.57127 | 0.695006 | 0.999532 | | 2.404953 | 0.214288 | 0.906276 | | 2.604282 | -0.46357 | 0.722703 |
| CSF2 | 2.167717 | 0.245723 | 0.999532 | | 2.152702 | -0.23814 | 0.906276 | | 2.108463 | -0.47151 | 0.765336 |
| RXFP2 | 2.003188 | -0.00717 | 0.999532 | | 2.242483 | -0.23584 | 0.906276 | | 1.849722 | -0.20619 | 0.918694 |
| ANGPTL1 | 1.96968 | 0.702637 | 0.999532 | | 1.868098 | 0.248722 | 0.906276 | | 2.074192 | -0.438 | 0.791466 |
| MSRB1 | 3.168985 | -0.06812 | 0.999532 | | 3.397923 | -0.19829 | 0.906276 | | 2.723145 | -0.11052 | 0.951656 |
| MON2 | 4.889889 | -0.03979 | 0.999532 | | 5.852218 | 0.152192 | 0.906276 | | 4.857481 | 0.21139 | 0.862823 |
| EIF5 | 7.147536 | -0.20989 | 0.999532 | | 8.712991 | 0.127908 | 0.906276 | | 6.92862 | 0.348669 | 0.702463 |
| MCCC2 | 2.706768 | -0.13815 | 0.999532 | | 3.414834 | 0.18894 | 0.906276 | | 2.735529 | 0.347563 | 0.809832 |
| CCL26 | 2.118861 | -0.05733 | 0.999532 | | 2.368563 | -0.23491 | 0.906276 | | 2.000106 | -0.14613 | 0.948127 |
| NOL4L | 2.845101 | 0.118295 | 0.999532 | | 3.232408 | 0.198407 | 0.906276 | | 2.928091 | 0.092093 | 0.964728 |
| POLE2 | 1.875762 | 0.536626 | 0.999532 | | 1.892189 | 0.254067 | 0.906276 | | 1.940651 | -0.2643 | 0.892674 |
| CBX1 | 2.526915 | -0.42877 | 0.999532 | | 3.047664 | -0.19617 | 0.906333 | | 2.305566 | 0.249913 | 0.881689 |
| MAGED4 | 1.957446 | 0.418458 | 0.999532 | | 1.734511 | -0.25975 | 0.906333 | | 1.807902 | -0.66102 | 0.678492 |
| CALR3 | 2.04581 | 0.638437 | 0.999532 | | 1.780678 | -0.26589 | 0.906522 | | 1.934669 | -0.88531 | 0.540342 |
| SEC11C | 2.548953 | 0.60497 | 0.999532 | | 2.457743 | 0.214802 | 0.906522 | | 2.588984 | -0.37203 | 0.794815 |
| DOHH | 1.905951 | 0.057733 | 0.999532 | | 2.276476 | 0.232233 | 0.906522 | | 2.052353 | 0.188072 | 0.926586 |
| ZBTB8A | 2.297593 | -0.11298 | 0.999532 | | 2.741677 | -0.22385 | 0.906795 | | 2.239144 | -0.06885 | 0.978927 |
| CACNA1H | 2.165288 | 0.119833 | 0.999532 | | 2.565461 | 0.210074 | 0.906828 | | 2.239639 | 0.108417 | 0.958889 |
| ABHD17C | 2.525264 | -0.09495 | 0.999532 | | 2.904894 | -0.22827 | 0.906965 | | 2.345837 | -0.11019 | 0.962716 |
| MTHFR | 2.947893 | -0.01629 | 0.999532 | | 3.178349 | -0.20235 | 0.906965 | | 2.646965 | -0.16714 | 0.925487 |
| SPG7 | 6.388527 | 0.218286 | 0.999532 | | 7.002858 | 0.136001 | 0.906965 | | 6.300921 | -0.06324 | 0.963492 |
| GSK3B | 4.833955 | -0.20371 | 0.999532 | | 6.041795 | 0.148686 | 0.906965 | | 4.907809 | 0.369909 | 0.718633 |
| IPO11 | 2.626771 | 0.161842 | 0.999532 | | 2.569444 | -0.21084 | 0.906965 | | 2.3029 | -0.35467 | 0.816618 |
| HDHD5 | 2.558047 | 0.097051 | 0.999532 | | 2.691956 | -0.20702 | 0.907074 | | 2.244438 | -0.28278 | 0.867009 |
| HUS1 | 5.213659 | -0.64698 | 0.999532 | | 7.158692 | 0.135392 | 0.907172 | | 5.20515 | 0.800308 | 0.282041 |
| CEP192 | 1.900256 | 0.254389 | 0.999532 | | 2.075273 | 0.233915 | 0.907612 | | 1.966287 | -0.00186 | 0.999429 |
| GNPNAT1 | 1.951803 | 0.424746 | 0.999532 | | 1.667332 | -0.25404 | 0.907612 | | 1.752975 | -0.66312 | 0.666906 |
| NUDT5 | 4.439656 | -0.22 | 0.999532 | | 5.737761 | 0.149642 | 0.90777 | | 4.474087 | 0.39048 | 0.717627 |
| CSF1R | 2.55142 | -0.30926 | 0.999532 | | 3.346652 | 0.206338 | 0.90777 | | 2.724811 | 0.522513 | 0.718735 |
| OSBPL1A | 5.154874 | 0.009399 | 0.999532 | | 5.502861 | -0.15184 | 0.90777 | | 4.560456 | -0.14866 | 0.910105 |
| TACO1 | 2.238194 | 0.090816 | 0.999532 | | 2.521444 | 0.209783 | 0.908042 | | 2.291884 | 0.133193 | 0.946572 |
| ECRG4 | 4.267203 | 0.155154 | 0.999532 | | 4.602488 | 0.1746 | 0.908153 | | 4.266224 | 0.032393 | 0.990134 |
| RAB40AL | 2.484954 | 0.767262 | 0.999532 | | 1.958474 | -0.23748 | 0.90816 | | 2.299727 | -0.98107 | 0.413122 |
| PRKG1 | 2.231648 | 0.17659 | 0.999532 | | 2.241828 | -0.22857 | 0.908209 | | 2.033951 | -0.38909 | 0.809832 |
| TDO2 | 2.310647 | 0.052451 | 0.999532 | | 2.586386 | 0.221854 | 0.908271 | | 2.403544 | 0.164044 | 0.941446 |
| PDZD4 | 2.187667 | 0.217235 | 0.999532 | | 2.073027 | -0.23881 | 0.908367 | | 2.109483 | -0.43681 | 0.792539 |
| ADH6 | 2.188227 | 0.214219 | 0.999532 | | 2.380489 | 0.218164 | 0.908384 | | 2.228248 | 0.022088 | 0.995304 |
| PAPSS1 | 5.239008 | -0.05791 | 0.999532 | | 6.248315 | 0.151668 | 0.90875 | | 5.058823 | 0.225159 | 0.864245 |
| GOLGA5 | 5.673547 | 0.279467 | 0.999532 | | 5.967974 | 0.145283 | 0.90875 | | 5.698638 | -0.11575 | 0.92697 |
| RMND1 | 1.940236 | 0.116252 | 0.999532 | | 2.152959 | 0.224519 | 0.90875 | | 1.993662 | 0.127404 | 0.954459 |
| TRIO | 6.407775 | -0.25441 | 0.999532 | | 8.155359 | 0.125241 | 0.90875 | | 6.479435 | 0.396514 | 0.646821 |
| MINDY4 | 2.915101 | -0.45054 | 0.999532 | | 4.044062 | -0.19689 | 0.90875 | | 2.849827 | 0.351733 | 0.837256 |
| KCNN1 | 1.933764 | -0.0133 | 0.999532 | | 1.940934 | -0.23704 | 0.90875 | | 1.799277 | -0.20776 | 0.920259 |
| TSR3 | 3.868494 | -0.01155 | 0.999532 | | 4.412688 | 0.172218 | 0.90875 | | 3.878025 | 0.209916 | 0.877943 |
| H2BC9 | 3.233233 | -0.4754 | 0.999532 | | 3.968564 | -0.1977 | 0.90875 | | 2.916651 | 0.295888 | 0.861737 |
| KRI1 | 2.062361 | 0.089627 | 0.999532 | | 2.45118 | 0.214442 | 0.90875 | | 2.138602 | 0.143017 | 0.944229 |
| CRACR2B | 3.458757 | 0.347449 | 0.999532 | | 2.923645 | -0.1992 | 0.90875 | | 3.105261 | -0.51783 | 0.672452 |
| UBTD2 | 3.604403 | -0.29364 | 0.999532 | | 4.601278 | 0.182599 | 0.90875 | | 3.747031 | 0.498281 | 0.668481 |
| KIF13A | 11.38662 | -0.23301 | 0.999532 | | 13.40276 | -0.10865 | 0.90875 | | 10.42206 | 0.145712 | 0.870663 |
| TRMT11 | 3.491567 | 0.076142 | 0.999532 | | 3.833598 | 0.187678 | 0.90875 | | 3.377863 | 0.131616 | 0.939913 |
| PYGM | 2.126664 | 0.218327 | 0.999532 | | 2.026416 | -0.24292 | 0.90875 | | 1.837291 | -0.44602 | 0.792688 |
| SAGE1 | 2.097571 | 0.760645 | 0.999532 | | 1.920302 | 0.253079 | 0.908872 | | 2.146157 | -0.48934 | 0.755301 |
| PSPN | 2.510295 | 0.207587 | 0.999532 | | 2.467095 | -0.21589 | 0.908872 | | 2.308167 | -0.40269 | 0.798849 |
| UGDH | 4.172373 | -0.19027 | 0.999532 | | 5.38624 | 0.160076 | 0.908872 | | 4.12147 | 0.36291 | 0.745628 |
| SPRY4 | 7.612331 | -0.70606 | 0.950261 | | 10.37344 | -0.12961 | 0.908872 | | 6.820956 | 0.612426 | 0.434766 |
| RPRD2 | 4.340225 | -0.51666 | 0.999532 | | 5.955199 | 0.158601 | 0.908892 | | 4.539118 | 0.681053 | 0.484303 |
| GSX2 | 4.700191 | 0.731411 | 0.999532 | | 4.322484 | 0.202121 | 0.909166 | | 4.745282 | -0.5172 | 0.639848 |
| SPDYA | 2.33025 | 0.529111 | 0.999532 | | 1.757125 | -0.25509 | 0.909327 | | 2.085571 | -0.74761 | 0.632847 |
| PCP4 | 2.314424 | 0.332105 | 0.999532 | | 2.170615 | -0.22635 | 0.909415 | | 2.055281 | -0.53715 | 0.719855 |
| LETMD1 | 4.619313 | -0.30565 | 0.999532 | | 5.752875 | 0.149023 | 0.909415 | | 4.519106 | 0.474996 | 0.638728 |
| DENND10 | 4.142721 | 0.098921 | 0.999532 | | 4.148067 | -0.17612 | 0.909415 | | 3.748065 | -0.25825 | 0.858647 |
| CDKN3 | 2.16681 | 0.846143 | 0.999532 | | 1.99254 | 0.249186 | 0.909415 | | 2.206227 | -0.57163 | 0.701196 |
| SMG8 | 2.918226 | 0.294978 | 0.999532 | | 2.670312 | -0.21994 | 0.909415 | | 2.714819 | -0.50498 | 0.722703 |
| DHX58 | 2.485096 | -0.1909 | 0.999532 | | 2.735694 | -0.20342 | 0.909415 | | 2.316585 | 0.001144 | 0.999635 |
| NRN1L | 3.514135 | 0.119883 | 0.999532 | | 3.774209 | -0.1811 | 0.909452 | | 3.314241 | -0.27525 | 0.844321 |
| EIF4G3 | 5.597482 | -0.19804 | 0.999532 | | 7.01705 | 0.132554 | 0.909572 | | 5.574724 | 0.348714 | 0.717627 |
| NUDT17 | 2.574059 | 0.399919 | 0.999532 | | 2.56734 | 0.216667 | 0.909684 | | 2.605151 | -0.16932 | 0.927668 |
| ALPK2 | 2.197091 | 0.185769 | 0.999532 | | 2.278616 | 0.221447 | 0.909852 | | 2.218741 | 0.051434 | 0.985873 |
| CKAP4 | 6.814913 | -0.01126 | 0.999532 | | 8.221593 | 0.13133 | 0.909852 | | 6.72041 | 0.159704 | 0.886495 |
| HARS1 | 6.369799 | -0.07494 | 0.999532 | | 7.098937 | -0.14338 | 0.909852 | | 5.951027 | -0.04801 | 0.977635 |
| TBX6 | 2.969094 | 0.301201 | 0.999532 | | 3.34371 | 0.199835 | 0.909852 | | 3.064782 | -0.08152 | 0.968434 |
| RNF14 | 5.572668 | -0.20149 | 0.999532 | | 7.147175 | 0.133818 | 0.909852 | | 5.487298 | 0.347983 | 0.709825 |
| CKAP5 | 1.924946 | -0.4713 | 0.999532 | | 2.564258 | 0.211425 | 0.909852 | | 2.004086 | 0.694192 | 0.649725 |
| NUDT15 | 4.310178 | 0.527362 | 0.999532 | | 3.990891 | -0.18407 | 0.909868 | | 4.046392 | -0.6886 | 0.49299 |
| SLC38A7 | 2.465023 | 0.878803 | 0.999532 | | 2.052625 | 0.231225 | 0.909958 | | 2.507321 | -0.63259 | 0.643959 |
| SDHAF2 | 2.946136 | -0.08387 | 0.999532 | | 3.27744 | -0.20199 | 0.909958 | | 2.780672 | -0.09902 | 0.963866 |
| PRRG3 | 2.454207 | 0.190117 | 0.999532 | | 2.702434 | 0.20688 | 0.909958 | | 2.472635 | 0.035015 | 0.991103 |
| CAPG | 3.369696 | 0.177591 | 0.999532 | | 3.763306 | 0.177145 | 0.909958 | | 3.365133 | 0.017091 | 0.996109 |
| GHDC | 2.32536 | 0.05847 | 0.999532 | | 2.687098 | 0.200867 | 0.909958 | | 2.348767 | 0.159228 | 0.931282 |
| CTNND1 | 9.954851 | -0.04158 | 0.999532 | | 11.81148 | 0.119662 | 0.909958 | | 9.866341 | 0.180538 | 0.843564 |
| FAM200B | 4.577191 | 0.277528 | 0.999532 | | 5.12949 | 0.161079 | 0.909958 | | 4.518212 | -0.09847 | 0.946958 |
| SF3B5 | 3.495489 | -0.18988 | 0.999532 | | 4.782324 | 0.15619 | 0.909958 | | 3.567092 | 0.355426 | 0.769793 |
| NGF | 2.102873 | 0.233892 | 0.999532 | | 2.227358 | -0.23555 | 0.909958 | | 1.961696 | -0.44434 | 0.791576 |
| GSTA3 | 2.780212 | 0.332118 | 0.999532 | | 2.437633 | -0.21741 | 0.909958 | | 2.5882 | -0.53432 | 0.724411 |
| DLL1 | 3.727742 | -0.4312 | 0.999532 | | 4.38984 | -0.16699 | 0.909958 | | 3.321243 | 0.279277 | 0.849021 |
| KDM4A | 4.242108 | -0.16707 | 0.999532 | | 4.706747 | -0.15998 | 0.909958 | | 3.69926 | 0.023915 | 0.992286 |
| FBN3 | 2.16709 | 0.32228 | 0.999532 | | 2.062628 | -0.2401 | 0.909958 | | 2.083749 | -0.54574 | 0.724411 |
| ZNF576 | 3.180171 | -0.19966 | 0.999532 | | 3.544919 | -0.18507 | 0.910112 | | 2.875016 | 0.031078 | 0.991289 |
| CARD6 | 6.250122 | 0.081305 | 0.999532 | | 7.089902 | 0.139291 | 0.910112 | | 6.120282 | 0.081119 | 0.950616 |
| RGS7 | 1.733208 | 0.520365 | 0.999532 | | 1.820519 | 0.258175 | 0.910135 | | 1.808415 | -0.24287 | 0.903339 |
| COG2 | 3.262366 | -0.13322 | 0.999532 | | 3.450994 | -0.18136 | 0.910268 | | 3.011927 | -0.03828 | 0.988853 |
| METTL4 | 2.540839 | 0.173112 | 0.999532 | | 2.414034 | -0.21663 | 0.910426 | | 2.271924 | -0.37276 | 0.81294 |
| PREX1 | 4.024781 | -0.34981 | 0.999532 | | 4.606637 | -0.16926 | 0.910494 | | 3.541012 | 0.193439 | 0.901033 |
| RCVRN | 1.900329 | 0.418511 | 0.999532 | | 1.679768 | -0.25346 | 0.910494 | | 1.753536 | -0.65409 | 0.679452 |
| SYNRG | 5.365055 | 0.226999 | 0.999532 | | 5.514176 | 0.144635 | 0.910494 | | 5.277321 | -0.0693 | 0.964153 |
| PDHX | 4.242981 | 0.127158 | 0.999532 | | 4.895137 | 0.153715 | 0.910494 | | 4.27268 | 0.044419 | 0.979434 |
| RNF19B | 4.423549 | 0.161013 | 0.999532 | | 4.583523 | 0.158922 | 0.910647 | | 4.352314 | 9.46E-05 | 0.999949 |
| DOP1B | 4.137689 | 0.338997 | 0.999532 | | 3.743587 | -0.18373 | 0.910647 | | 3.822194 | -0.50446 | 0.660958 |
| SAMD3 | 2.878801 | 0.239807 | 0.999532 | | 2.782269 | -0.20034 | 0.910647 | | 2.637341 | -0.42232 | 0.759133 |
| PABIR1 | 6.159566 | -0.6207 | 0.999532 | | 7.887111 | -0.13137 | 0.910647 | | 5.390901 | 0.502456 | 0.616301 |
| ACTL6A | 4.735776 | -0.04331 | 0.999532 | | 5.443227 | 0.144106 | 0.910647 | | 4.759987 | 0.206616 | 0.863134 |
| ATF7IP2 | 2.499511 | 0.256333 | 0.999532 | | 2.633712 | 0.210017 | 0.910865 | | 2.554877 | -0.02786 | 0.992286 |
| DCAF12L | 2.341272 | 1.092596 | 0.999532 | | 1.851921 | 0.245152 | 0.910865 | | 2.371028 | -0.82948 | 0.515762 |
| PGAP2 | 2.213757 | 0.040878 | 0.999532 | | 2.687145 | 0.213292 | 0.910916 | | 2.314148 | 0.198874 | 0.917753 |
| ZC3H3 | 2.493014 | -0.28896 | 0.999532 | | 2.79181 | -0.2007 | 0.911089 | | 2.12406 | 0.103709 | 0.963309 |
| DEFA5 | 2.303868 | 0.732322 | 0.999532 | | 1.778556 | -0.2498 | 0.911089 | | 2.114599 | -0.9649 | 0.456688 |
| CYC1 | 5.687886 | -0.22801 | 0.999532 | | 6.375461 | -0.1566 | 0.911089 | | 4.932172 | 0.096645 | 0.951656 |
| C2orf74 | 3.955752 | -0.0135 | 0.999532 | | 4.411884 | 0.168901 | 0.911089 | | 3.913056 | 0.176208 | 0.905952 |
| CCDC137 | 4.407313 | -0.37828 | 0.999532 | | 5.739705 | 0.154009 | 0.911089 | | 4.49561 | 0.54211 | 0.614525 |
| EDEM1 | 3.750749 | 0.061245 | 0.999532 | | 4.072215 | 0.178608 | 0.911096 | | 3.832349 | 0.131953 | 0.939549 |
| ARHGAP | 2.484787 | 0.246494 | 0.999532 | | 2.424991 | -0.22966 | 0.911096 | | 2.281305 | -0.45686 | 0.769793 |
| C1QTNF6 | 2.178527 | 0.220169 | 0.999532 | | 2.440452 | 0.221044 | 0.911148 | | 2.148716 | 0.020339 | 0.996681 |
| CACNA2D | 2.309908 | -0.11995 | 0.999532 | | 2.484869 | -0.2202 | 0.911328 | | 1.994373 | -0.08151 | 0.974421 |
| SOX9 | 3.969067 | 0.139426 | 0.999532 | | 4.036251 | -0.17496 | 0.911561 | | 3.555436 | -0.29164 | 0.825455 |
| GJD2 | 2.298439 | 0.728046 | 0.999532 | | 1.749015 | -0.25327 | 0.911699 | | 2.120001 | -0.96833 | 0.484303 |
| SF3B2 | 3.562764 | -0.1277 | 0.999532 | | 4.518961 | 0.165003 | 0.911699 | | 3.659714 | 0.311896 | 0.803301 |
| CEP63 | 3.427717 | 0.180774 | 0.999532 | | 3.202962 | -0.18305 | 0.911755 | | 3.113619 | -0.35217 | 0.798849 |
| SRCAP | 5.148212 | -0.13216 | 0.999532 | | 6.197912 | 0.139947 | 0.911755 | | 4.966156 | 0.289743 | 0.784185 |
| PALD1 | 3.754539 | -0.41716 | 0.999532 | | 5.191221 | 0.15588 | 0.911755 | | 3.860896 | 0.59067 | 0.563455 |
| POLR2L | 21.04203 | 0.041362 | 0.999532 | | 24.1671 | 0.114222 | 0.911755 | | 21.00704 | 0.090017 | 0.92697 |
| COQ7 | 4.245841 | 0.652914 | 0.999532 | | 3.893015 | 0.180819 | 0.911755 | | 4.279348 | -0.45529 | 0.686214 |
| RAD23A | 12.56154 | -0.02869 | 0.999532 | | 14.57533 | 0.09917 | 0.911755 | | 12.40661 | 0.145379 | 0.858647 |
| SIGMAR1 | 4.445463 | -0.13887 | 0.999532 | | 5.356928 | 0.15298 | 0.911755 | | 4.606686 | 0.316825 | 0.789312 |
| C1QTNF7 | 1.947795 | 0.252974 | 0.999532 | | 2.042951 | 0.224293 | 0.911755 | | 1.980371 | -0.01423 | 0.996891 |
| CAB39L | 2.582734 | -0.259 | 0.999532 | | 2.833909 | -0.20168 | 0.911953 | | 2.396761 | 0.086489 | 0.972135 |
| CLCN2 | 2.785888 | 0.343849 | 0.999532 | | 3.121827 | 0.198485 | 0.912072 | | 2.856266 | -0.11981 | 0.947515 |
| TMEM266 | 2.520295 | 0.405353 | 0.999532 | | 2.723512 | 0.206325 | 0.912077 | | 2.613732 | -0.16713 | 0.908891 |
| STRADB | 3.692865 | 0.316867 | 0.999532 | | 4.078365 | 0.173408 | 0.912162 | | 3.67137 | -0.12072 | 0.939759 |
| TG | 3.282971 | 0.57676 | 0.999532 | | 2.88474 | -0.20521 | 0.912304 | | 3.0699 | -0.76413 | 0.502578 |
| MAPK10 | 4.657058 | -0.26229 | 0.999532 | | 5.268817 | -0.15709 | 0.912319 | | 4.092552 | 0.122542 | 0.93603 |
| C1orf185 | 1.867721 | -0.01891 | 0.999532 | | 1.92921 | -0.24192 | 0.912319 | | 1.67338 | -0.20592 | 0.925487 |
| FBRS | 12.69611 | -0.19393 | 0.999532 | | 14.54119 | -0.12135 | 0.912479 | | 11.75368 | 0.092667 | 0.934701 |
| KCTD7 | 3.642844 | 0.670259 | 0.999532 | | 3.041707 | 0.200682 | 0.912507 | | 3.636447 | -0.65012 | 0.583283 |
| S1PR2 | 3.490269 | -0.00383 | 0.999532 | | 3.639995 | -0.18516 | 0.912507 | | 3.197303 | -0.1615 | 0.923834 |
| ARL10 | 3.837823 | -0.01628 | 0.999532 | | 4.087279 | -0.17903 | 0.912507 | | 3.338611 | -0.14049 | 0.930403 |
| SOX14 | 2.172733 | 0.145085 | 0.999532 | | 2.64974 | 0.22807 | 0.912507 | | 2.405936 | 0.104125 | 0.965649 |
| BMPER | 3.005124 | -0.16371 | 0.999532 | | 3.257759 | -0.19454 | 0.912507 | | 2.754113 | -0.01849 | 0.996681 |
| GPR155 | 2.833513 | 0.339936 | 0.999532 | | 2.820697 | 0.199675 | 0.912507 | | 2.764831 | -0.12307 | 0.946876 |
| KRT26 | 2.468282 | 0.154918 | 0.999532 | | 2.31134 | -0.2276 | 0.912507 | | 2.230254 | -0.37382 | 0.836012 |
| TMEM68 | 4.14387 | -0.07407 | 0.999532 | | 5.115362 | 0.162051 | 0.912615 | | 4.31001 | 0.25269 | 0.844097 |
| PHF19 | 2.252169 | 0.222261 | 0.999532 | | 2.179504 | -0.22814 | 0.912635 | | 2.025327 | -0.43172 | 0.791576 |
| CLIC3 | 3.370093 | 0.173065 | 0.999532 | | 3.377197 | -0.20448 | 0.912635 | | 3.184546 | -0.37039 | 0.799944 |
| GPAA1 | 8.538404 | -0.10455 | 0.999532 | | 9.809136 | 0.117401 | 0.912635 | | 8.24352 | 0.23166 | 0.806394 |
| CCDC85C | 8.430554 | 0.240785 | 0.999532 | | 8.946112 | 0.117991 | 0.912635 | | 8.418105 | -0.1032 | 0.918194 |
| INCENP | 3.526968 | 0.277262 | 0.999532 | | 3.253696 | -0.19233 | 0.912635 | | 3.201938 | -0.45155 | 0.720106 |
| GOLT1B | 3.341694 | 0.037916 | 0.999532 | | 3.511287 | -0.17776 | 0.912635 | | 3.189884 | -0.19524 | 0.898151 |
| MSX2 | 1.92728 | 0.286931 | 0.999532 | | 2.310394 | 0.245029 | 0.912635 | | 2.138515 | -0.02626 | 0.994617 |
| AASDHPF | 2.768317 | -0.30241 | 0.999532 | | 3.368464 | 0.190159 | 0.912635 | | 2.790224 | 0.495978 | 0.720106 |
| IER5 | 4.502112 | -0.3108 | 0.999532 | | 5.998418 | 0.142592 | 0.912635 | | 4.694265 | 0.474052 | 0.646863 |
| LMBRD2 | 2.81009 | 0.302809 | 0.999532 | | 2.594617 | -0.20714 | 0.912635 | | 2.71192 | -0.49425 | 0.720106 |
| ADAM32 | 2.029545 | 0.534259 | 0.999532 | | 1.660962 | -0.25752 | 0.912635 | | 1.918036 | -0.77348 | 0.626985 |
| TOPBP1 | 3.059797 | 0.216225 | 0.999532 | | 2.987854 | -0.19705 | 0.912635 | | 2.863308 | -0.39314 | 0.784779 |
| RFK | 4.994407 | -0.10084 | 0.999532 | | 6.033354 | 0.147625 | 0.912635 | | 5.074408 | 0.260726 | 0.827267 |
| MAP3K7C | 2.7148 | 0.074836 | 0.999532 | | 2.784764 | -0.20011 | 0.912635 | | 2.403855 | -0.25528 | 0.878619 |
| NT5C3A | 3.615771 | 0.156703 | 0.999532 | | 3.733995 | -0.17683 | 0.912635 | | 3.399038 | -0.31594 | 0.815293 |
| TRBC1 | 2.936326 | 0.246591 | 0.999532 | | 2.889844 | -0.19899 | 0.912635 | | 2.776635 | -0.42967 | 0.75311 |
| SAP30L | 3.320074 | -0.13202 | 0.999532 | | 3.951846 | 0.166684 | 0.912635 | | 3.298453 | 0.310548 | 0.816775 |
| CIAO2B | 3.69183 | 0.118032 | 0.999532 | | 3.584333 | -0.18156 | 0.912635 | | 3.372122 | -0.28304 | 0.840357 |
| GNA14 | 3.419841 | -0.22092 | 0.999532 | | 3.871534 | -0.16849 | 0.912713 | | 3.110858 | 0.070933 | 0.972221 |
| MRPS10 | 3.570913 | 0.227076 | 0.999532 | | 3.895264 | 0.190681 | 0.912828 | | 3.684844 | -0.01956 | 0.995112 |
| ARL4D | 3.263894 | -0.03205 | 0.999532 | | 3.393392 | -0.19246 | 0.912828 | | 2.888308 | -0.1419 | 0.941197 |
| TPH1 | 2.22063 | 0.333988 | 0.999532 | | 2.217042 | 0.223446 | 0.912828 | | 2.258672 | -0.09618 | 0.967 |
| GRIN3A | 3.036727 | 0.225524 | 0.999532 | | 3.107867 | 0.193231 | 0.912828 | | 3.07144 | -0.01545 | 0.996681 |
| AMMECR | 4.190662 | -0.05371 | 0.999532 | | 4.541095 | -0.16317 | 0.913122 | | 3.616852 | -0.09219 | 0.955834 |
| ACP6 | 3.698389 | 0.464247 | 0.999532 | | 3.423841 | 0.187148 | 0.913137 | | 3.5958 | -0.27702 | 0.85585 |
| RABGEF1 | 3.900224 | -0.16974 | 0.999532 | | 4.684175 | 0.158866 | 0.913137 | | 3.882981 | 0.3389 | 0.791466 |
| ITGBL1 | 2.006455 | -0.21839 | 0.999532 | | 2.275279 | -0.22494 | 0.913137 | | 1.88803 | 0.009309 | 0.997967 |
| CUL9 | 2.769289 | 0.473398 | 0.999532 | | 3.005271 | 0.201995 | 0.913137 | | 2.968898 | -0.25621 | 0.868182 |
| CAMK1D | 3.752998 | 0.110345 | 0.999532 | | 3.666133 | -0.17598 | 0.913137 | | 3.416445 | -0.26991 | 0.842696 |
| ARMCX6 | 4.491919 | -0.02884 | 0.999532 | | 4.797544 | -0.15707 | 0.913137 | | 4.066969 | -0.10576 | 0.946512 |
| ZCCHC2 | 3.19587 | -0.62665 | 0.999532 | | 3.968505 | -0.17453 | 0.913137 | | 2.888193 | 0.469697 | 0.728015 |
| CFAP20D | 2.08971 | 0.440684 | 0.999532 | | 1.914306 | -0.23588 | 0.913137 | | 2.000763 | -0.65713 | 0.660958 |
| SCRT1 | 2.298035 | 0.183295 | 0.999532 | | 2.404743 | -0.24082 | 0.913137 | | 2.150818 | -0.397 | 0.826947 |
| ZNF469 | 3.802714 | 0.0603 | 0.999532 | | 4.205469 | -0.17687 | 0.913137 | | 3.668077 | -0.21357 | 0.886495 |
| LY86 | 2.561376 | -0.00483 | 0.999532 | | 3.044666 | 0.188342 | 0.913137 | | 2.66671 | 0.21339 | 0.900848 |
| FREM2 | 1.934064 | 0.277576 | 0.999532 | | 1.751513 | -0.25309 | 0.913137 | | 1.754119 | -0.51265 | 0.773546 |
| FEM1A | 3.596761 | -0.42071 | 0.999532 | | 4.446989 | -0.16804 | 0.913247 | | 3.310925 | 0.272218 | 0.84977 |
| FAM126B | 6.544673 | 0.004785 | 0.999532 | | 7.157184 | -0.13925 | 0.913247 | | 6.169687 | -0.11264 | 0.939107 |
| FRMD4B | 2.470977 | -0.05943 | 0.999532 | | 2.891143 | 0.191345 | 0.913247 | | 2.410878 | 0.262852 | 0.87591 |
| SPRY1 | 4.130948 | -0.34796 | 0.999532 | | 5.387544 | 0.147983 | 0.913247 | | 4.205134 | 0.51439 | 0.60646 |
| TCP10L2 | 2.58222 | 0.293184 | 0.999532 | | 2.678972 | -0.22895 | 0.913247 | | 2.356161 | -0.50993 | 0.722703 |
| SNAPC5 | 2.157388 | 0.221031 | 0.999532 | | 2.410774 | 0.212517 | 0.913247 | | 2.223129 | 0.00782 | 0.997967 |
| SDCBP | 7.481076 | -0.56634 | 0.999532 | | 9.801861 | -0.12174 | 0.913247 | | 6.628336 | 0.460773 | 0.575108 |
| FGD4 | 2.837481 | -0.12942 | 0.999532 | | 3.077475 | -0.18973 | 0.913247 | | 2.464997 | -0.04741 | 0.985693 |
| HSF4 | 3.905485 | 0.620424 | 0.999532 | | 3.293322 | -0.20007 | 0.913247 | | 3.679061 | -0.81278 | 0.446609 |
| KLHDC1 | 4.350344 | 0.52715 | 0.999532 | | 4.188207 | 0.171833 | 0.913247 | | 4.314138 | -0.33915 | 0.768383 |
| COMMD9 | 3.217165 | 0.274472 | 0.999532 | | 3.631031 | 0.173534 | 0.913247 | | 3.349186 | -0.07976 | 0.965649 |
| SP1 | 7.181992 | -0.07835 | 0.999532 | | 8.542106 | 0.129287 | 0.913247 | | 7.508659 | 0.258967 | 0.797739 |
| HTT | 3.059583 | -0.0315 | 0.999532 | | 3.88305 | 0.19355 | 0.913247 | | 3.145792 | 0.243584 | 0.876107 |
| ACO2 | 6.620452 | 0.101669 | 0.999532 | | 7.210771 | 0.141721 | 0.913247 | | 6.475653 | 0.037497 | 0.985693 |
| ZNF708 | 2.339398 | -0.02592 | 0.999532 | | 2.959992 | 0.203249 | 0.913247 | | 2.626983 | 0.235905 | 0.891051 |
| B3GNT2 | 3.271811 | 0.373625 | 0.999532 | | 3.312007 | 0.187348 | 0.913247 | | 3.319175 | -0.17259 | 0.914929 |
| ZNF648 | 1.981274 | -0.02609 | 0.999532 | | 2.183473 | -0.22945 | 0.913247 | | 1.778992 | -0.17329 | 0.941197 |
| UGCG | 5.997486 | -0.30479 | 0.999532 | | 7.703625 | 0.136484 | 0.913247 | | 5.838036 | 0.460056 | 0.628556 |
| ZSCAN1 | 2.377216 | 0.412982 | 0.999532 | | 2.411225 | -0.22382 | 0.913247 | | 2.227666 | -0.61018 | 0.666566 |
| APTX | 3.160647 | -0.03921 | 0.999532 | | 3.47981 | -0.19283 | 0.913247 | | 2.82814 | -0.13093 | 0.943229 |
| ABCB8 | 3.457442 | 0.492025 | 0.999532 | | 3.575364 | 0.176184 | 0.913247 | | 3.416009 | -0.29435 | 0.814884 |
| HS6ST1 | 7.835709 | -0.03172 | 0.999532 | | 9.010918 | 0.124515 | 0.913247 | | 7.64747 | 0.165192 | 0.881837 |
| HOXA6 | 2.031614 | -0.51475 | 0.999532 | | 2.67091 | 0.199054 | 0.913247 | | 2.08126 | 0.729055 | 0.623565 |
| GAS8 | 3.779682 | 0.004453 | 0.999532 | | 4.508571 | 0.163308 | 0.913247 | | 3.743915 | 0.177391 | 0.899852 |
| RHD | 3.472568 | 0.088385 | 0.999532 | | 3.522651 | -0.20055 | 0.913247 | | 3.228177 | -0.26265 | 0.864923 |
| MN51 | 2.376881 | 0.464295 | 0.999532 | | 2.089909 | -0.23764 | 0.913247 | | 2.173885 | -0.68189 | 0.649779 |
| DNPEP | 8.025361 | 0.124708 | 0.999532 | | 9.016336 | 0.126666 | 0.913247 | | 8.033244 | 0.022992 | 0.989148 |
| ZNF687 | 4.222184 | 0.038966 | 0.999532 | | 4.921774 | 0.154222 | 0.913247 | | 4.232344 | 0.133231 | 0.924186 |
| C10orf67 | 2.164504 | 0.342613 | 0.999532 | | 2.335467 | 0.224036 | 0.913247 | | 2.245584 | -0.10034 | 0.965649 |
| TP53TG3 | 2.583387 | -0.4485 | 0.999532 | | 3.215822 | -0.19109 | 0.913247 | | 2.404437 | 0.277747 | 0.874251 |
| KCNJ1 | 4.191485 | 0.115596 | 0.999532 | | 4.343504 | -0.18292 | 0.913247 | | 3.530949 | -0.27373 | 0.864245 |
| CNPPD1 | 5.048854 | -0.03179 | 0.999532 | | 6.11078 | 0.154957 | 0.913247 | | 5.084953 | 0.203508 | 0.865065 |
| BCCIP | 3.779915 | 0.100981 | 0.999532 | | 4.076109 | 0.174781 | 0.913247 | | 3.753043 | 0.073091 | 0.972221 |
| KCNJ14 | 1.856388 | 0.326253 | 0.999532 | | 1.666884 | -0.25016 | 0.913247 | | 1.722279 | -0.55997 | 0.739496 |
| ZNF79 | 2.6027 | 0.380854 | 0.999532 | | 2.346455 | -0.21888 | 0.913463 | | 2.37888 | -0.58222 | 0.676565 |
| EXOC6 | 1.870298 | -0.71075 | 0.999532 | | 2.327227 | -0.21322 | 0.913506 | | 1.736258 | 0.519077 | 0.765852 |
| UPK1B | 2.282156 | 0.533547 | 0.999532 | | 1.988487 | -0.24726 | 0.913506 | | 2.159598 | -0.77102 | 0.581238 |
| MAP7D1 | 4.619678 | 0.194282 | 0.999532 | | 4.795846 | 0.154527 | 0.913506 | | 4.4674 | -0.02495 | 0.991273 |
| FBXO21 | 4.98676 | 0.076624 | 0.999532 | | 5.674079 | 0.16026 | 0.913506 | | 4.942449 | 0.107406 | 0.941512 |
| TTC12 | 5.036012 | 0.029349 | 0.999532 | | 5.121504 | -0.1524 | 0.913506 | | 4.598257 | -0.16294 | 0.902619 |
| ASCC2 | 2.437247 | -0.07161 | 0.999532 | | 2.776538 | 0.192423 | 0.913506 | | 2.485594 | 0.283411 | 0.864245 |
| CHST2 | 3.483563 | 0.295051 | 0.999532 | | 3.615486 | 0.180994 | 0.913527 | | 3.580737 | -0.10532 | 0.947557 |
| ARHGAP | 1.818216 | 0.217604 | 0.999532 | | 1.998216 | 0.232673 | 0.913527 | | 1.872783 | 0.032366 | 0.992286 |
| GBP4 | 6.316238 | -0.47239 | 0.999532 | | 7.550794 | -0.16671 | 0.913527 | | 5.730994 | 0.340944 | 0.757644 |
| ZBTB20 | 6.581468 | 0.097824 | 0.999532 | | 7.6929 | 0.127288 | 0.913527 | | 6.436473 | 0.045291 | 0.974572 |
| YARS2 | 2.325556 | -0.30468 | 0.999532 | | 2.532891 | -0.20447 | 0.913527 | | 2.044499 | 0.112918 | 0.960773 |
| SOX6 | 2.047024 | 0.053199 | 0.999532 | | 2.104813 | -0.2253 | 0.913527 | | 1.872031 | -0.25925 | 0.894612 |
| DNAJB1 | 4.416782 | -0.27306 | 0.999532 | | 5.171065 | -0.15776 | 0.913527 | | 3.780946 | 0.113808 | 0.943248 |
| GATD1 | 7.552853 | -0.02459 | 0.999532 | | 8.998454 | 0.125753 | 0.913527 | | 7.626344 | 0.17076 | 0.863907 |
| TBP | 3.126482 | -0.02249 | 0.999532 | | 3.073246 | -0.18353 | 0.913527 | | 2.733485 | -0.14705 | 0.934572 |
| COPS3 | 5.220433 | 0.256388 | 0.999532 | | 5.597713 | 0.143114 | 0.913527 | | 5.102094 | -0.0996 | 0.943533 |
| SWI5 | 3.167996 | 0.082579 | 0.999532 | | 3.86302 | 0.174122 | 0.913527 | | 3.221182 | 0.110478 | 0.949893 |
| GPATCH | 3.701279 | -0.01115 | 0.999532 | | 4.091175 | -0.16743 | 0.913704 | | 3.385073 | -0.13518 | 0.931827 |
| MRC1 | 2.837059 | -0.3547 | 0.999532 | | 3.811169 | 0.180383 | 0.913722 | | 3.002503 | 0.5538 | 0.671153 |
| CDC25A | 1.787513 | 0.447875 | 0.999532 | | 1.942809 | 0.24222 | 0.913722 | | 1.835428 | -0.18749 | 0.929529 |
| CARNMT | 2.933291 | 0.117887 | 0.999532 | | 3.263579 | 0.182977 | 0.913722 | | 2.961525 | 0.083597 | 0.967002 |
| GRK6 | 3.353865 | -0.01684 | 0.999532 | | 3.406102 | -0.19269 | 0.913722 | | 3.087071 | -0.15709 | 0.923864 |
| PIAS4 | 1.97341 | 0.008367 | 0.999532 | | 2.124529 | -0.23067 | 0.913722 | | 1.797621 | -0.22069 | 0.913005 |
| ARL2BP | 6.853592 | -0.17042 | 0.999532 | | 7.831274 | -0.14163 | 0.913722 | | 6.025042 | 0.054031 | 0.977473 |
| LDLRAP1 | 4.531265 | 0.02387 | 0.999532 | | 5.373321 | 0.150246 | 0.913722 | | 4.49943 | 0.144634 | 0.911581 |
| MALRD1 | 2.373063 | 0.012858 | 0.999532 | | 2.452687 | -0.21478 | 0.913722 | | 2.144008 | -0.21027 | 0.909084 |
| EPM2A | 2.05468 | 0.238055 | 0.999532 | | 2.288954 | 0.215423 | 0.913722 | | 2.089351 | -0.00527 | 0.998586 |
| UHMK1 | 3.757451 | 0.167193 | 0.999532 | | 4.453753 | 0.163273 | 0.913722 | | 3.790508 | 0.022034 | 0.992963 |
| PPIF | 3.573609 | -0.14803 | 0.999532 | | 4.011368 | -0.16684 | 0.913722 | | 3.256399 | -2.54E-05 | 0.999949 |
| NDUFA3 | 11.01398 | -0.1113 | 0.999532 | | 13.74152 | 0.114532 | 0.913722 | | 11.11199 | 0.260012 | 0.781295 |
| SGSM3 | 4.580715 | -0.28278 | 0.999532 | | 5.371041 | -0.15984 | 0.913722 | | 4.149094 | 0.13987 | 0.920527 |
| TF | 2.107634 | 0.196738 | 0.999532 | | 2.085897 | -0.23748 | 0.913722 | | 1.945273 | -0.41655 | 0.815713 |
| NUAK2 | 3.077108 | 0.409332 | 0.999532 | | 2.815794 | -0.20661 | 0.913722 | | 2.79751 | -0.59935 | 0.629006 |
| OR4E1 | 2.587604 | 0.151814 | 0.999532 | | 2.90392 | -0.21918 | 0.913722 | | 2.529578 | -0.34658 | 0.837256 |
| PDE7A | 7.262389 | -0.14988 | 0.999532 | | 8.882292 | 0.12115 | 0.913722 | | 7.158932 | 0.291159 | 0.735469 |
| GPR52 | 2.891646 | 0.09201 | 0.999532 | | 3.419972 | 0.189013 | 0.913722 | | 2.938949 | 0.116953 | 0.94861 |
| AVEN | 2.623723 | 0.280438 | 0.999532 | | 2.774675 | 0.196877 | 0.913722 | | 2.612602 | -0.06579 | 0.976603 |
| ZKSCAN | 5.111744 | 0.019949 | 0.999532 | | 5.833286 | 0.140926 | 0.913722 | | 5.308262 | 0.142621 | 0.920486 |
| BAG4 | 4.845518 | 0.028679 | 0.999532 | | 5.033289 | -0.15345 | 0.913722 | | 4.351875 | -0.16428 | 0.902619 |
| PAOX | 3.318574 | 0.315497 | 0.999532 | | 3.758748 | 0.169676 | 0.913722 | | 3.474379 | -0.12071 | 0.940719 |
| CLEC17A | 1.988233 | -0.18627 | 0.999532 | | 2.185651 | -0.22113 | 0.913722 | | 1.805666 | -0.01776 | 0.996681 |
| KRBA2 | 1.768807 | 0.469697 | 0.999532 | | 1.801666 | 0.239939 | 0.913722 | | 1.771404 | -0.21219 | 0.915991 |
| ACTR1A | 6.51593 | -0.56332 | 0.999532 | | 8.280133 | -0.13018 | 0.913722 | | 5.856016 | 0.451429 | 0.619423 |
| SPCS2 | 3.248491 | -0.06816 | 0.999532 | | 3.779368 | 0.169037 | 0.913722 | | 3.137691 | 0.257108 | 0.854633 |
| GDF5 | 2.054411 | 0.415103 | 0.999532 | | 2.297274 | 0.244275 | 0.913722 | | 2.241267 | -0.15344 | 0.946512 |
| FTSJ1 | 4.445767 | -0.33067 | 0.999532 | | 5.33849 | -0.16156 | 0.913722 | | 3.777131 | 0.183137 | 0.894612 |
| ITGAM | 2.227563 | -0.52443 | 0.999532 | | 2.800737 | 0.191945 | 0.914349 | | 2.244173 | 0.730336 | 0.619423 |
| CD1E | 2.053012 | 0.185268 | 0.999532 | | 2.265687 | 0.220912 | 0.91436 | | 2.061203 | 0.052149 | 0.985709 |
| TLE6 | 2.761328 | 0.344487 | 0.999532 | | 2.842752 | 0.197518 | 0.914755 | | 2.950412 | -0.132 | 0.945456 |
| ESRP2 | 2.768879 | -0.23003 | 0.999532 | | 3.283231 | -0.18869 | 0.914755 | | 2.501192 | 0.062523 | 0.978282 |
| OR52L1 | 2.367987 | 0.684523 | 0.999532 | | 2.278636 | 0.222505 | 0.914755 | | 2.40552 | -0.44242 | 0.760572 |
| SPINK6 | 1.847018 | 0.113924 | 0.999532 | | 1.888294 | -0.24015 | 0.914755 | | 1.715225 | -0.33612 | 0.867378 |
| TCF25 | 8.798096 | 0.186666 | 0.999532 | | 8.7161 | -0.13088 | 0.914755 | | 8.15337 | -0.30391 | 0.735469 |
| RAD1 | 2.360044 | 0.556282 | 0.999532 | | 2.417764 | 0.211436 | 0.914755 | | 2.501668 | -0.32912 | 0.827267 |
| ENTHD1 | 2.206007 | -0.68574 | 0.999532 | | 2.757001 | -0.20488 | 0.914755 | | 1.85001 | 0.500386 | 0.760955 |
| RABIF | 2.082135 | 0.403314 | 0.999532 | | 2.181905 | 0.219081 | 0.914755 | | 2.12173 | -0.16774 | 0.934354 |
| PURB | 3.095659 | 0.137993 | 0.999532 | | 3.724201 | 0.175951 | 0.914755 | | 3.102934 | 0.052287 | 0.979239 |
| ATP2A1 | 4.280137 | -0.44848 | 0.999532 | | 5.457222 | -0.15539 | 0.914755 | | 3.936665 | 0.317327 | 0.809832 |
| DCP1B | 2.407332 | 0.248593 | 0.999532 | | 2.661397 | 0.204937 | 0.914755 | | 2.358737 | -0.02532 | 0.993618 |
| MAPK8IP | 5.909536 | 0.382901 | 0.999532 | | 5.335548 | -0.1559 | 0.914755 | | 5.171104 | -0.51212 | 0.598177 |
| DDX3X | 13.52605 | -0.5318 | 0.999532 | | 17.22459 | -0.10968 | 0.914755 | | 11.90018 | 0.456905 | 0.560424 |
| SHROOM | 2.857083 | -0.38075 | 0.999532 | | 3.312164 | -0.18618 | 0.914755 | | 2.586766 | 0.209668 | 0.905952 |
| AIMP1 | 6.03824 | -0.13407 | 0.999532 | | 7.325977 | 0.135434 | 0.914755 | | 6.208268 | 0.28316 | 0.792416 |
| PPP1R16 | 1.716776 | 0.350331 | 0.999532 | | 1.804129 | 0.238153 | 0.914755 | | 1.761847 | -0.09525 | 0.972039 |
| CCDC80 | 4.467524 | -0.44075 | 0.999532 | | 5.743248 | 0.161391 | 0.914755 | | 4.402033 | 0.612304 | 0.542124 |
| ZNRD2 | 4.393647 | -0.2365 | 0.999532 | | 4.733089 | -0.15292 | 0.914755 | | 3.75371 | 0.090513 | 0.958284 |
| GOLPH3 | 5.043266 | -0.1271 | 0.999532 | | 6.103681 | 0.138875 | 0.914755 | | 4.963121 | 0.283739 | 0.795612 |
| MFSD12 | 6.353957 | 0.059217 | 0.999532 | | 6.449614 | -0.13617 | 0.914755 | | 5.81064 | -0.18649 | 0.881689 |
| MTRF1 | 3.081179 | 0.298073 | 0.999532 | | 3.345588 | 0.181266 | 0.914755 | | 3.276172 | -0.0973 | 0.958191 |
| LAMTOR | 5.136721 | -0.02107 | 0.999532 | | 6.296772 | 0.131261 | 0.914755 | | 5.154569 | 0.170431 | 0.884653 |
| GATB | 3.715989 | 0.397147 | 0.999532 | | 3.836869 | 0.183685 | 0.914755 | | 3.755877 | -0.19265 | 0.900848 |
| MRGBP | 2.607354 | -0.27757 | 0.999532 | | 3.310096 | 0.178875 | 0.914755 | | 2.678532 | 0.472225 | 0.731348 |
| TRIM4 | 3.930776 | 0.348795 | 0.999532 | | 4.107545 | 0.164481 | 0.914755 | | 3.980404 | -0.16611 | 0.903339 |
| YOD1 | 2.656967 | 0.236753 | 0.999532 | | 2.98522 | 0.192161 | 0.914755 | | 2.708115 | -0.02584 | 0.992916 |
| TRIM39 | 3.506748 | 0.008062 | 0.999532 | | 3.825364 | -0.1832 | 0.914755 | | 3.142762 | -0.16989 | 0.914929 |
| NODAL | 2.552565 | 0.581946 | 0.999532 | | 2.213327 | -0.22261 | 0.914755 | | 2.3699 | -0.78684 | 0.538285 |
| SLC35D2 | 2.397784 | 0.147418 | 0.999532 | | 2.620035 | 0.201578 | 0.914755 | | 2.422677 | 0.073046 | 0.974421 |
| OAS2 | 3.677529 | -0.14184 | 0.999532 | | 4.068247 | -0.16532 | 0.914755 | | 3.198818 | -0.00338 | 0.999159 |
| NLRX1 | 2.948465 | 0.164392 | 0.999532 | | 2.861728 | -0.19261 | 0.914755 | | 2.625171 | -0.3436 | 0.818373 |
| PDPK1 | 5.271145 | 0.483671 | 0.999532 | | 4.525325 | -0.15742 | 0.914755 | | 4.689797 | -0.61974 | 0.481527 |
| UBE2H | 5.850753 | -0.35087 | 0.999532 | | 8.011249 | 0.137342 | 0.914755 | | 6.046477 | 0.493029 | 0.566812 |
| CNOT3 | 4.266161 | -0.00862 | 0.999532 | | 5.067743 | 0.1449 | 0.914755 | | 4.269308 | 0.172518 | 0.894612 |
| MBD2 | 8.938656 | -0.15709 | 0.999532 | | 11.21837 | 0.111819 | 0.914755 | | 9.100285 | 0.287324 | 0.706749 |
| BBS1 | 2.249073 | 0.226437 | 0.999532 | | 2.663946 | 0.201831 | 0.914755 | | 2.296632 | -0.00625 | 0.998316 |
| CCDC85B | 2.711336 | -0.3662 | 0.999532 | | 3.384953 | 0.179316 | 0.914755 | | 2.560818 | 0.561921 | 0.664921 |
| EIF3E | 10.18779 | 0.03592 | 0.999532 | | 11.8189 | 0.125172 | 0.914755 | | 9.922956 | 0.118282 | 0.92403 |
| DDX50 | 4.525012 | 0.036438 | 0.999532 | | 4.965638 | -0.16197 | 0.914807 | | 4.269808 | -0.18371 | 0.892869 |

| | | | | | | | | | | | |
|---|---|---|---|---|---|---|---|---|---|---|---|
| STK17A | 3.479061 | 0.141457 | 0.999532 | | 3.554356 | -0.1728 | 0.914807 | | 3.154144 | -0.29559 | 0.824012 |
| PTRH2 | 2.468677 | 0.178355 | 0.999532 | | 2.635728 | -0.20413 | 0.914807 | | 2.313015 | -0.36264 | 0.816526 |
| WDR1 | 9.043464 | -0.22854 | 0.999532 | | 11.16285 | 0.124532 | 0.914807 | | 8.927687 | 0.365167 | 0.673405 |
| KDELR3 | 2.898057 | 0.662046 | 0.999532 | | 2.645177 | 0.193897 | 0.914807 | | 2.897237 | -0.45167 | 0.723453 |
| PTRH1 | 3.446254 | 0.258099 | 0.999532 | | 3.270132 | -0.18943 | 0.914807 | | 3.119395 | -0.43108 | 0.739496 |
| HSF1 | 3.213072 | -0.11463 | 0.999532 | | 3.663332 | -0.17209 | 0.914807 | | 2.877506 | -0.04057 | 0.986898 |
| PFDN1 | 5.773088 | 0.080225 | 0.999532 | | 6.528587 | 0.130776 | 0.914807 | | 5.852539 | 0.068403 | 0.961643 |
| AP4S1 | 2.503529 | 0.205995 | 0.999532 | | 2.389034 | -0.2111 | 0.91481 | | 2.237185 | -0.40058 | 0.806394 |
| HLA-F | 13.28897 | 0.030595 | 0.999532 | | 13.86084 | -0.12526 | 0.91481 | | 12.31022 | -0.11379 | 0.900848 |
| UIMC1 | 6.49916 | 0.002957 | 0.999532 | | 7.641699 | 0.136183 | 0.91481 | | 6.603413 | 0.151039 | 0.894617 |
| CLDN11 | 7.221746 | -0.42221 | 0.999532 | | 9.973173 | 0.188288 | 0.91481 | | 7.397337 | 0.642632 | 0.542124 |
| CDK3 | 3.246507 | 0.602981 | 0.999532 | | 3.216359 | 0.183628 | 0.91481 | | 3.239122 | -0.39565 | 0.753597 |
| C19orf53 | 4.66282 | 0.061412 | 0.999532 | | 5.099327 | 0.147977 | 0.91481 | | 4.643688 | 0.084943 | 0.95999 |
| NUCB2 | 5.350351 | 0.492867 | 0.999532 | | 5.425354 | 0.141275 | 0.91481 | | 5.306836 | -0.3284 | 0.751138 |
| FAM166C | 1.956915 | 0.24625 | 0.999532 | | 2.082694 | 0.224704 | 0.91481 | | 1.964271 | -0.0097 | 0.997694 |
| FOXG1 | 3.595845 | 0.021485 | 0.999532 | | 3.508948 | -0.18763 | 0.91481 | | 3.281451 | -0.22831 | 0.901033 |
| EFHD2 | 3.451035 | -0.55336 | 0.999532 | | 4.392111 | -0.16688 | 0.91481 | | 3.211533 | 0.403579 | 0.769793 |
| MPC1 | 4.76704 | 0.177476 | 0.999532 | | 5.20638 | 0.156351 | 0.91481 | | 4.450028 | 0.018514 | 0.994064 |
| RHOBTB1 | 3.465947 | 0.475895 | 0.999532 | | 3.473545 | 0.175921 | 0.91481 | | 3.511211 | -0.27905 | 0.837256 |
| USP32 | 2.436037 | -0.06562 | 0.999532 | | 2.904983 | 0.183142 | 0.91481 | | 2.430386 | 0.263714 | 0.871713 |
| PRKX | 9.892797 | 0.083016 | 0.999532 | | 10.69012 | -0.13232 | 0.91481 | | 9.233156 | -0.19296 | 0.854633 |
| DDRGK1 | 5.20168 | -0.01706 | 0.999532 | | 6.158679 | 0.136322 | 0.91481 | | 5.177043 | 0.169921 | 0.890261 |
| CRABP2 | 3.935314 | 0.070234 | 0.999532 | | 4.263653 | -0.1673 | 0.914877 | | 3.757531 | -0.21125 | 0.890932 |
| PINX1 | 3.754072 | -0.11343 | 0.999532 | | 3.97491 | -0.16861 | 0.914899 | | 3.339148 | -0.03309 | 0.989536 |
| SDE2 | 4.128854 | -0.31008 | 0.999532 | | 5.249068 | 0.146213 | 0.914899 | | 4.170082 | 0.469946 | 0.663089 |
| FAM204A | 7.395987 | -0.10251 | 0.999532 | | 8.873362 | 0.118689 | 0.914924 | | 7.579894 | 0.244787 | 0.800239 |
| ABL1 | 5.91469 | -0.01402 | 0.999532 | | 6.978394 | 0.131587 | 0.914924 | | 5.600804 | 0.151229 | 0.901813 |
| TNPO3 | 5.044539 | 0.446119 | 0.999532 | | 5.096854 | 0.146591 | 0.914924 | | 5.087474 | -0.28088 | 0.792539 |
| PPOX | 2.596842 | 0.513303 | 0.999532 | | 2.554532 | 0.210231 | 0.914946 | | 2.745236 | -0.28124 | 0.871595 |
| CDC42BPA | 2.609594 | 0.831046 | 0.999532 | | 1.754711 | -0.23191 | 0.914961 | | 2.392812 | -1.05477 | 0.394907 |
| OR1S2 | 3.195662 | 0.508469 | 0.999532 | | 2.595636 | -0.2057 | 0.914961 | | 2.912634 | -0.69705 | 0.565022 |
| SYCP2 | 2.459782 | 0.146443 | 0.999532 | | 2.409883 | -0.22234 | 0.915021 | | 2.248902 | -0.34768 | 0.850928 |
| TXNDC2 | 1.994337 | -0.27763 | 0.999532 | | 2.317336 | -0.22003 | 0.915021 | | 1.86041 | 0.077339 | 0.978004 |
| ROBO1 | 3.02459 | 0.391235 | 0.999532 | | 3.154678 | 0.177052 | 0.915021 | | 2.993484 | -0.19402 | 0.896374 |
| GOLT1A | 3.87918 | 0.401049 | 0.999532 | | 3.520916 | -0.1816 | 0.915021 | | 3.606339 | -0.56726 | 0.632847 |
| ATP5MJ | 7.534771 | -0.07085 | 0.999532 | | 8.825864 | 0.119029 | 0.915021 | | 7.317532 | 0.200356 | 0.851785 |
| PABIR2 | 2.71528 | -0.03111 | 0.999532 | | 3.387175 | 0.170351 | 0.915021 | | 2.751953 | 0.222896 | 0.885892 |
| MANBA | 2.08709 | -0.11341 | 0.999532 | | 2.334554 | -0.20733 | 0.915021 | | 1.850194 | -0.0736 | 0.977359 |
| GNAT3 | 2.076973 | 0.571608 | 0.999532 | | 1.769579 | -0.24459 | 0.915021 | | 1.898236 | -0.80023 | 0.57215 |
| SNX24 | 2.443748 | 0.287793 | 0.999532 | | 2.780505 | 0.191144 | 0.915021 | | 2.60442 | -0.07001 | 0.974772 |
| XYLB | 3.564 | -0.00817 | 0.999532 | | 3.727914 | -0.20442 | 0.915021 | | 3.273901 | -0.19064 | 0.91062 |
| AJM1 | 3.281661 | 0.136801 | 0.999532 | | 3.199655 | -0.18431 | 0.915021 | | 3.133491 | -0.27708 | 0.869666 |
| RAB3B | 1.976387 | 0.158681 | 0.999532 | | 2.16371 | -0.22039 | 0.915021 | | 1.854504 | -0.35392 | 0.844097 |
| PHC3 | 9.439885 | -0.05724 | 0.999532 | | 11.31828 | 0.104725 | 0.915021 | | 9.39114 | 0.181347 | 0.833604 |
| MAP1LC3 | 2.342912 | 0.3078 | 0.999532 | | 2.160626 | -0.21758 | 0.915021 | | 2.23634 | -0.51048 | 0.740755 |
| SLC45A1 | 5.406456 | 0.076137 | 0.999532 | | 5.260595 | -0.16186 | 0.915021 | | 4.796691 | -0.23876 | 0.880375 |
| ZBTB18 | 2.433115 | 0.143601 | 0.999532 | | 2.697659 | 0.188039 | 0.915021 | | 2.457591 | 0.061295 | 0.978229 |
| ZNF232 | 2.145489 | -0.05107 | 0.999532 | | 2.553722 | 0.216396 | 0.915021 | | 2.145173 | 0.294869 | 0.871957 |
| GLT8D1 | 3.694277 | 0.436815 | 0.999532 | | 3.595551 | 0.168351 | 0.915021 | | 3.627545 | -0.25949 | 0.858767 |
| DDIT3 | 6.468781 | -0.26238 | 0.999532 | | 7.401793 | -0.13228 | 0.915021 | | 5.980839 | 0.155354 | 0.90465 |
| RHOA | 6.668467 | -0.57231 | 0.999532 | | 9.234255 | 0.120981 | 0.915021 | | 6.795613 | 0.711295 | 0.286175 |
| RUFY2 | 4.689084 | 0.321513 | 0.999532 | | 4.604459 | 0.148542 | 0.915021 | | 4.719057 | -0.15246 | 0.908985 |
| FBXO15 | 2.311984 | 0.45908 | 0.999532 | | 2.423309 | 0.211462 | 0.915021 | | 2.478918 | -0.23081 | 0.893853 |
| MAEL | 2.480227 | 0.2344 | 0.999532 | | 2.459674 | -0.22182 | 0.915021 | | 2.268178 | -0.43743 | 0.792416 |
| CDK1 | 1.81086 | 0.177871 | 0.999532 | | 1.862951 | 0.225617 | 0.915021 | | 1.796111 | 0.060033 | 0.984587 |
| TMEM25 | 3.407416 | -0.42722 | 0.999532 | | 4.089405 | -0.163 | 0.915021 | | 2.997621 | 0.278813 | 0.851503 |
| DPP3 | 1.912357 | 0.424507 | 0.999532 | | 1.831536 | 0.224054 | 0.915021 | | 1.912036 | -0.18514 | 0.92697 |
| PTP4A3 | 6.082481 | 0.165051 | 0.999532 | | 6.74958 | 0.134722 | 0.915021 | | 6.180315 | -0.014 | 0.995943 |
| TNKS | 3.875886 | -0.28733 | 0.999532 | | 5.040732 | 0.147261 | 0.915021 | | 3.883383 | 0.452745 | 0.679818 |
| BLMH | 3.088129 | 0.354999 | 0.999532 | | 3.25807 | 0.181449 | 0.915021 | | 3.039579 | -0.15527 | 0.920487 |
| MBD1 | 4.122799 | -0.13827 | 0.999532 | | 5.106325 | 0.15524 | 0.915021 | | 4.100788 | 0.30862 | 0.806394 |
| MICU1 | 3.690465 | -0.21174 | 0.999532 | | 4.234535 | -0.16855 | 0.915021 | | 3.230453 | 0.063107 | 0.975473 |
| KRTAP4- | 1.968628 | 0.166492 | 0.999532 | | 2.217483 | 0.226759 | 0.915021 | | 1.981241 | 0.079491 | 0.975753 |
| DCAF4 | 3.476038 | 0.163045 | 0.999532 | | 3.769547 | -0.17513 | 0.915021 | | 3.283253 | -0.3113 | 0.823193 |
| LIN7B | 2.786877 | 0.495289 | 0.999532 | | 2.796224 | -0.21506 | 0.915021 | | 2.648127 | -0.68625 | 0.600254 |
| PRCC | 3.596676 | 0.162481 | 0.999532 | | 3.59717 | -0.17506 | 0.915021 | | 3.272093 | -0.31864 | 0.816589 |
| NME8 | 2.28439 | 0.108397 | 0.999532 | | 2.354276 | -0.21741 | 0.915021 | | 2.127563 | -0.30849 | 0.865065 |
| RAP1B | 15.0224 | -0.22785 | 0.999532 | | 18.8068 | 0.092826 | 0.915021 | | 15.06653 | 0.3387 | 0.57215 |
| GAMT | 4.659142 | 0.005702 | 0.999532 | | 5.362268 | 0.151921 | 0.915021 | | 4.578958 | 0.171491 | 0.899161 |
| PCID2 | 5.88913 | 0.336361 | 0.999532 | | 6.133101 | 0.136873 | 0.915021 | | 5.837994 | -0.18037 | 0.874653 |
| TEPSIN | 4.023059 | 0.105463 | 0.999532 | | 4.657356 | 0.16403 | 0.915021 | | 4.221393 | 0.069055 | 0.971683 |
| CMPK2 | 3.128088 | -0.20461 | 0.999532 | | 3.362026 | -0.17072 | 0.915021 | | 2.811206 | 0.049748 | 0.983211 |
| MRPS22 | 3.749703 | 0.168002 | 0.999532 | | 3.864303 | 0.167823 | 0.915021 | | 3.682541 | 0.008484 | 0.996891 |
| FCHO2 | 4.667592 | 0.2654 | 0.999532 | | 4.893907 | 0.148364 | 0.915021 | | 4.687384 | -0.1029 | 0.944668 |
| BTN2A1 | 6.230776 | 0.188204 | 0.999532 | | 6.883354 | 0.146891 | 0.915021 | | 6.368903 | -0.01652 | 0.995331 |
| KCNIP1 | 2.197887 | -0.68091 | 0.999532 | | 2.791617 | -0.21055 | 0.915021 | | 1.969843 | 0.483548 | 0.794256 |
| TRAC | 2.540658 | -0.08308 | 0.999532 | | 3.212496 | 0.183345 | 0.915021 | | 2.700555 | 0.281496 | 0.863134 |
| TBX2 | 8.976341 | -0.33361 | 0.999532 | | 10.78472 | -0.11059 | 0.915021 | | 8.17542 | 0.243723 | 0.789444 |
| GLA | 2.365454 | 0.357831 | 0.999532 | | 2.670386 | 0.201194 | 0.915021 | | 2.518923 | -0.1395 | 0.941197 |
| LRRC75B | 8.013623 | 0.317259 | 0.999532 | | 7.603845 | -0.13314 | 0.915021 | | 7.516197 | -0.43582 | 0.660958 |
| PGM5 | 2.929684 | -0.02691 | 0.999532 | | 3.025588 | -0.1963 | 0.915021 | | 2.576659 | -0.14896 | 0.939549 |
| ACTL7A | 2.46801 | 0.686695 | 0.999532 | | 2.001554 | -0.22846 | 0.915021 | | 2.231635 | -0.89763 | 0.485816 |
| GALNT10 | 2.301519 | 0.06984 | 0.999532 | | 2.737904 | 0.190451 | 0.915021 | | 2.368058 | 0.143372 | 0.941197 |
| SLC37A1 | 3.557125 | 0.262626 | 0.999532 | | 3.412909 | -0.17929 | 0.915021 | | 3.403048 | -0.43053 | 0.732486 |
| CYP17A1 | 2.825416 | -0.38793 | 0.999532 | | 3.419272 | -0.17979 | 0.915021 | | 2.555249 | 0.227984 | 0.892869 |
| VCPKMT | 3.009855 | 0.11694 | 0.999532 | | 2.922232 | -0.19001 | 0.915021 | | 2.66285 | -0.28915 | 0.85585 |
| RPA2 | 4.297125 | 0.510155 | 0.999532 | | 4.201971 | 0.172949 | 0.915021 | | 4.116579 | -0.30394 | 0.792538 |
| ATP13A4 | 2.151067 | 0.443541 | 0.999532 | | 1.935623 | -0.23689 | 0.915021 | | 1.999508 | -0.66513 | 0.67072 |
| ISM1 | 2.723709 | 0.220363 | 0.999532 | | 3.116947 | 0.187037 | 0.915021 | | 2.730173 | -0.01817 | 0.996471 |
| MB | 1.835558 | 0.076241 | 0.999532 | | 2.032312 | -0.22711 | 0.915021 | | 1.830215 | -0.27261 | 0.893103 |
| PPM1H | 2.357457 | 0.164296 | 0.999532 | | 2.405526 | -0.20729 | 0.915021 | | 2.145328 | -0.35285 | 0.830822 |
| PRR19 | 2.086501 | 0.484684 | 0.999532 | | 1.780195 | -0.2397 | 0.915021 | | 1.882374 | -0.70977 | 0.647218 |
| ABCC4 | 2.542562 | 0.354548 | 0.999532 | | 2.86395 | 0.192664 | 0.915021 | | 2.717191 | -0.13696 | 0.941197 |
| C19orf48 | 2.457765 | 0.426161 | 0.999532 | | 2.276905 | -0.21176 | 0.915021 | | 2.248926 | -0.61964 | 0.652925 |
| TTLL4 | 2.206047 | 0.031667 | 0.999532 | | 2.139217 | -0.2194 | 0.915021 | | 1.907818 | -0.23812 | 0.90424 |
| SEMA4B | 4.335732 | 0.007283 | 0.999532 | | 4.632552 | -0.15392 | 0.915021 | | 3.920685 | -0.14384 | 0.923834 |
| PHC2 | 11.87455 | -0.13439 | 0.999532 | | 13.52552 | -0.09981 | 0.915021 | | 10.79287 | 0.058517 | 0.960773 |
| ZNF799 | 4.38864 | 0.36608 | 0.999532 | | 3.838197 | -0.16363 | 0.915021 | | 4.032658 | -0.50949 | 0.628053 |
| CPD | 6.119011 | 0.248438 | 0.999532 | | 6.528282 | 0.135279 | 0.915021 | | 5.872623 | -0.09938 | 0.938429 |
| CLCN4 | 3.030218 | 0.04766 | 0.999532 | | 3.219428 | -0.19292 | 0.915021 | | 2.673261 | -0.23354 | 0.883116 |
| ZNF738 | 3.017886 | 0.452447 | 0.999532 | | 2.62805 | -0.20038 | 0.915021 | | 2.661686 | -0.63509 | 0.62216 |
| SLC44A4 | 2.109412 | 0.07616 | 0.999532 | | 2.483021 | 0.206241 | 0.915021 | | 2.259618 | 0.147434 | 0.945302 |
| TTC28 | 2.331862 | -0.55134 | 0.999532 | | 2.89232 | -0.18723 | 0.915021 | | 2.086853 | 0.381901 | 0.813755 |
| ERICH4 | 1.910542 | 0.003762 | 0.999532 | | 2.127413 | -0.22959 | 0.915036 | | 1.83311 | -0.21013 | 0.922922 |
| PHLDA3 | 3.690613 | -0.13222 | 0.999532 | | 4.437066 | 0.154928 | 0.915036 | | 3.727958 | 0.309829 | 0.800239 |
| PASD1 | 2.312699 | 0.303567 | 0.999532 | | 2.145557 | -0.22014 | 0.915189 | | 2.0418 | -0.50768 | 0.747677 |
| IPO8 | 3.729242 | 0.090434 | 0.999532 | | 4.08214 | 0.159213 | 0.915189 | | 3.903202 | 0.096054 | 0.958699 |
| RBM6 | 8.469701 | 0.087601 | 0.999532 | | 9.64616 | 0.110268 | 0.915189 | | 8.460757 | 0.043199 | 0.974421 |
| NDUFS1 | 5.580336 | 0.066577 | 0.999532 | | 6.231948 | 0.137843 | 0.915189 | | 5.570494 | 0.087033 | 0.94934 |
| ARL8B | 3.902168 | -0.22852 | 0.999532 | | 5.119149 | 0.150298 | 0.915189 | | 3.918951 | 0.407207 | 0.738316 |
| AP5S1 | 3.342497 | -0.56877 | 0.999532 | | 4.184389 | -0.1577 | 0.915189 | | 2.993612 | 0.429185 | 0.732916 |
| APOH | 2.024232 | -0.1102 | 0.999532 | | 2.078724 | -0.21851 | 0.91535 | | 1.75078 | -0.09525 | 0.972221 |
| EP400 | 2.934554 | 0.378059 | 0.999532 | | 2.894356 | -0.1942 | 0.915364 | | 2.635964 | -0.55825 | 0.660097 |
| DNM3 | 3.74469 | 0.27421 | 0.999532 | | 3.80412 | 0.163432 | 0.915364 | | 3.683213 | -0.09614 | 0.955022 |
| GFER | 2.962242 | -0.0929 | 0.999532 | | 3.044139 | -0.17889 | 0.915364 | | 2.73175 | -0.07024 | 0.974421 |
| TM9SF1 | 3.452707 | 0.35719 | 0.999532 | | 3.6388 | 0.17608 | 0.915609 | | 3.559691 | -0.15897 | 0.918194 |
| GSDMB | 3.163137 | 0.598658 | 0.999532 | | 3.006393 | 0.184149 | 0.915635 | | 3.09595 | -0.39996 | 0.757037 |
| DOCK7 | 3.959975 | -0.36746 | 0.999532 | | 5.252246 | 0.146632 | 0.91607 | | 4.107995 | 0.534261 | 0.630705 |
| TCEAL6 | 2.023896 | 0.177812 | 0.999532 | | 2.013316 | -0.22345 | 0.916122 | | 1.810421 | -0.38221 | 0.834627 |
| KIAA1191 | 4.354005 | 0.32399 | 0.999532 | | 4.590236 | 0.163974 | 0.916162 | | 4.323374 | -0.14161 | 0.920259 |
| POLE3 | 2.270569 | 0.117737 | 0.999532 | | 2.599125 | 0.196109 | 0.916162 | | 2.292967 | 0.095604 | 0.965649 |
| ALG2 | 2.870164 | -0.51044 | 0.999532 | | 3.869916 | 0.159804 | 0.916162 | | 2.803168 | 0.681596 | 0.560424 |
| APCDD1 | 2.239489 | 0.426696 | 0.999532 | | 2.15401 | 0.208735 | 0.916162 | | 2.20835 | -0.2014 | 0.910105 |
| NUP153 | 5.885937 | 0.071193 | 0.999532 | | 5.680717 | -0.14248 | 0.916162 | | 5.150161 | -0.20288 | 0.881837 |
| GLTP | 6.375289 | 0.653747 | 0.999532 | | 5.504639 | 0.149784 | 0.916162 | | 6.124617 | -0.47352 | 0.617899 |
| QARS1 | 8.632684 | 0.085951 | 0.999532 | | 9.72532 | 0.129017 | 0.916162 | | 8.411885 | 0.066748 | 0.9618 |
| REEP4 | 3.038344 | 0.039192 | 0.999532 | | 3.660343 | 0.166353 | 0.916162 | | 3.143249 | 0.146125 | 0.92697 |
| INO80E | 10.46616 | -0.0484 | 0.999532 | | 12.16964 | -0.09917 | 0.916162 | | 9.826465 | -0.02341 | 0.988379 |
| RPS7 | 24.81349 | -0.354 | 0.999532 | | 31.69492 | 0.083285 | 0.916162 | | 23.9868 | 0.452164 | 0.385915 |
| MED17 | 2.433369 | -0.1547 | 0.999532 | | 2.729287 | -0.1929 | 0.916162 | | 2.220737 | -0.01846 | 0.996681 |
| ZC3H13 | 8.113201 | -0.14379 | 0.999532 | | 10.244 | 0.113993 | 0.916162 | | 8.330438 | 0.278068 | 0.744727 |
| ATRX | 45.6476 | 0.181253 | 0.999532 | | 49.72111 | 0.109096 | 0.916162 | | 45.24499 | -0.04546 | 0.974421 |
| ADAMTS9 | 13.517 | 0.22942 | 0.999532 | | 13.39535 | -0.10738 | 0.916162 | | 12.23015 | -0.31813 | 0.649779 |
| ARID5B | 4.409954 | -0.38129 | 0.999532 | | 5.251253 | -0.14438 | 0.916162 | | 3.899905 | 0.255965 | 0.842696 |
| MRRF | 3.651917 | -0.06448 | 0.999532 | | 3.852276 | -0.17377 | 0.916233 | | 3.289296 | -0.09682 | 0.960427 |
| NOL6 | 2.456403 | 0.299142 | 0.999532 | | 2.267256 | -0.21371 | 0.916359 | | 2.180948 | -0.4974 | 0.741408 |
| SLC35A3 | 3.807139 | 0.328372 | 0.999532 | | 4.042284 | 0.160883 | 0.916466 | | 3.9501 | -0.14824 | 0.91993 |
| TPPP | 2.144448 | 0.1455 | 0.999532 | | 2.326908 | -0.21176 | 0.916466 | | 2.025864 | -0.32709 | 0.858705 |
| CLU | 8.899411 | -0.18259 | 0.999532 | | 11.54086 | 0.181086 | 0.916466 | | 8.970122 | 0.413949 | 0.737109 |
| FAM178B | 1.965105 | 0.461368 | 0.999532 | | 2.139061 | 0.219163 | 0.916466 | | 2.13757 | -0.22094 | 0.906811 |
| ZXDB | 3.042795 | -0.08039 | 0.999532 | | 3.22681 | -0.18902 | 0.916466 | | 2.623475 | -0.08639 | 0.969501 |
| MED1 | 3.421619 | -0.18215 | 0.999532 | | 3.773347 | -0.16719 | 0.916466 | | 3.080986 | 0.031264 | 0.991159 |
| ELAC2 | 3.342348 | -0.10896 | 0.999532 | | 4.066895 | 0.165097 | 0.916471 | | 3.518007 | 0.293305 | 0.824718 |
| FGD5 | 11.38331 | -0.04691 | 0.999532 | | 12.76761 | 0.10839 | 0.916715 | | 11.23648 | 0.162975 | 0.87591 |
| DYNC1I1 | 1.98171 | -0.36239 | 0.999532 | | 2.196177 | -0.21375 | 0.916715 | | 1.778412 | 0.165554 | 0.943248 |
| SPSB2 | 3.190104 | 0.129404 | 0.999532 | | 3.184937 | -0.1849 | 0.916715 | | 2.902965 | -0.29528 | 0.851503 |
| OR1N2 | 2.404089 | -0.0977 | 0.999532 | | 2.470592 | -0.1974 | 0.916801 | | 2.277777 | -0.07815 | 0.974421 |
| YBEY | 2.97484 | -0.37536 | 0.999532 | | 3.358266 | -0.18063 | 0.916801 | | 2.566605 | 0.210128 | 0.903339 |
| TMEM237 | 2.298772 | -0.19105 | 0.999532 | | 2.832637 | 0.185331 | 0.916889 | | 2.419153 | 0.397745 | 0.792176 |
| TAAR5 | 2.826264 | 0.27491 | 0.999532 | | 2.63758 | -0.19787 | 0.916889 | | 2.595156 | -0.45439 | 0.752299 |
| HIKESHI | 3.824942 | -0.46267 | 0.999532 | | 4.330361 | -0.1576 | 0.916928 | | 3.355463 | 0.317912 | 0.815293 |
| MZT2A | 3.233322 | 0.450563 | 0.999532 | | 3.307733 | 0.174608 | 0.917176 | | 3.146465 | -0.25937 | 0.857377 |
| TXK | 2.051984 | -0.18388 | 0.999532 | | 2.209335 | -0.21439 | 0.917176 | | 1.890254 | -0.01312 | 0.996891 |
| SOX7 | 3.334969 | -0.13304 | 0.999532 | | 4.110413 | 0.161276 | 0.917207 | | 3.591727 | 0.307896 | 0.827267 |
| CALML5 | 2.706936 | 0.35871 | 0.999532 | | 2.387503 | -0.21545 | 0.917241 | | 2.560509 | -0.56455 | 0.731946 |
| CD83 | 3.566758 | -0.15661 | 0.999532 | | 3.939112 | -0.1658 | 0.917241 | | 3.148683 | 0.01132 | 0.996891 |
| ZNF425 | 2.120606 | -0.13034 | 0.999532 | | 2.21574 | -0.20856 | 0.917241 | | 1.97771 | -0.06481 | 0.980581 |
| B4GALNT | 3.049439 | 0.322306 | 0.999532 | | 2.934992 | -0.1916 | 0.917241 | | 2.695255 | -0.49409 | 0.709825 |
| NOP9 | 5.885186 | 0.075552 | 0.999532 | | 6.775067 | 0.133876 | 0.917241 | | 5.847855 | 0.076547 | 0.95824 |
| FAM229A | 2.372718 | 0.145938 | 0.999532 | | 2.777387 | 0.1831 | 0.917241 | | 2.455851 | 0.058483 | 0.978927 |
| PRDM1 | 4.251413 | -0.30428 | 0.999532 | | 4.81863 | -0.14284 | 0.917241 | | 3.898552 | 0.17302 | 0.903339 |
| HOXA2 | 2.950956 | 0.185546 | 0.999532 | | 3.491966 | 0.169595 | 0.917241 | | 3.186707 | 0.012943 | 0.996891 |
| KMT2C | 4.249434 | 0.29774 | 0.999532 | | 4.643578 | 0.161784 | 0.91732 | | 4.221091 | -0.11854 | 0.938714 |
| FUOM | 5.392877 | -0.22266 | 0.999532 | | 6.141991 | -0.14131 | 0.91732 | | 4.663908 | 0.102021 | 0.943248 |
| LAYN | 2.911409 | -0.58129 | 0.999532 | | 4.159743 | 0.165984 | 0.91732 | | 3.063759 | 0.76967 | 0.515762 |
| GATA2 | 5.078614 | -0.55131 | 0.999532 | | 6.64956 | -0.13097 | 0.91732 | | 4.82691 | 0.441159 | 0.66232 |
| EPHB1 | 4.866833 | 0.15778 | 0.999532 | | 4.938704 | -0.15276 | 0.91732 | | 4.77411 | -0.30114 | 0.801905 |
| KHDC3L | 2.024924 | 0.374505 | 0.999532 | | 1.887692 | -0.22205 | 0.91732 | | 1.832016 | -0.57913 | 0.724363 |
| PRSS22 | 2.266927 | -0.00764 | 0.999532 | | 2.377989 | -0.20392 | 0.91732 | | 2.082466 | -0.18173 | 0.931282 |
| TIMM10 | 2.420839 | 0.164204 | 0.999532 | | 2.65918 | 0.188265 | 0.91732 | | 2.344872 | 0.040299 | 0.988853 |
| TMEM200 | 1.741419 | -0.07438 | 0.999532 | | 1.818019 | -0.226 | 0.91732 | | 1.571574 | -0.13624 | 0.957645 |
| DIPK1B | 2.796821 | -0.6114 | 0.999532 | | 3.642313 | 0.15821 | 0.91732 | | 2.869524 | 0.783095 | 0.49299 |
| NABP1 | 3.087574 | 0.190066 | 0.999532 | | 3.53075 | 0.167508 | 0.91732 | | 3.123925 | -0.00063 | 0.999746 |
| COTL1 | 11.32367 | -0.16749 | 0.999532 | | 13.78648 | 0.13298 | 0.91732 | | 11.34752 | 0.329987 | 0.697271 |
| GLDC | 2.305481 | -0.25711 | 0.999532 | | 2.67632 | -0.19806 | 0.91732 | | 2.048257 | 0.07947 | 0.974421 |
| TRAPPC1 | 4.794096 | 0.082524 | 0.999532 | | 4.49559 | -0.15326 | 0.91732 | | 4.219754 | -0.24904 | 0.860184 |
| PRKRA | 3.949516 | 0.288767 | 0.999532 | | 4.156962 | 0.151862 | 0.91732 | | 3.823997 | -0.12021 | 0.935073 |
| CABP5 | 1.988043 | 0.172007 | 0.999532 | | 2.362588 | 0.207732 | 0.91732 | | 2.13303 | 0.066757 | 0.979591 |
| SRSF3 | 9.867367 | -0.12357 | 0.999532 | | 12.30459 | 0.113162 | 0.91732 | | 9.806859 | 0.271131 | 0.755977 |
| ZNF766 | 2.851789 | -0.39932 | 0.999532 | | 3.317059 | -0.16963 | 0.91732 | | 2.697984 | 0.258374 | 0.87591 |
| PRSS38 | 2.292929 | -0.25951 | 0.999532 | | 2.640391 | -0.20782 | 0.91732 | | 2.051002 | 0.068942 | 0.978931 |
| MRE11 | 2.425744 | 0.240505 | 0.999532 | | 2.49585 | 0.194022 | 0.91732 | | 2.374016 | -0.03185 | 0.99176 |
| OCEL1 | 2.601415 | -0.42818 | 0.999532 | | 3.557028 | 0.168612 | 0.91732 | | 2.678204 | 0.617464 | 0.64142 |
| TMEM53 | 2.70903 | 0.563531 | 0.999532 | | 2.366502 | -0.19986 | 0.91732 | | 2.54111 | -0.74855 | 0.523387 |
| OR8B2 | 2.681489 | 0.223586 | 0.999532 | | 2.744545 | -0.19201 | 0.91732 | | 2.452414 | -0.39697 | 0.784263 |
| ZNF697 | 8.719099 | 0.149788 | 0.999532 | | 9.529313 | 0.120149 | 0.91732 | | 8.878908 | -0.00537 | 0.997153 |
| DEFA1B | 3.544695 | -0.21962 | 0.999532 | | 3.979028 | -0.16261 | 0.91732 | | 3.035757 | 0.076199 | 0.969693 |
| DHX35 | 2.811063 | -0.006 | 0.999532 | | 3.366848 | 0.172835 | 0.91732 | | 2.823613 | 0.195476 | 0.900848 |
| CEP162 | 2.832586 | 0.4224 | 0.999532 | | 2.578109 | -0.20162 | 0.91732 | | 2.72499 | -0.60411 | 0.66232 |
| RDH11 | 4.06964 | -0.00033 | 0.999532 | | 4.897256 | 0.141007 | 0.91732 | | 3.953894 | 0.152895 | 0.910105 |
| TGIF2LY | 2.080519 | 0.522707 | 0.999532 | | 1.72906 | -0.23664 | 0.91732 | | 2.003627 | -0.74345 | 0.64142 |
| CNKSR3 | 5.055683 | -0.04272 | 0.999532 | | 5.554578 | -0.14335 | 0.91732 | | 4.810062 | -0.08076 | 0.961567 |
| DGAT2L6 | 2.122536 | 0.33633 | 0.999532 | | 2.245602 | 0.211529 | 0.91732 | | 2.159653 | -0.10538 | 0.953492 |
| H3C7 | 2.161381 | -0.25356 | 0.999532 | | 2.436827 | -0.20894 | 0.91732 | | 1.979982 | 0.058628 | 0.983749 |
| ASIC5 | 1.8577 | -0.01021 | 0.999532 | | 1.86511 | -0.22638 | 0.91732 | | 1.687163 | -0.20168 | 0.928522 |
| SNAI3 | 2.412957 | -0.09776 | 0.999532 | | 2.505199 | -0.19683 | 0.91732 | | 2.206602 | -0.08263 | 0.972758 |
| ATP6V1G | 2.958508 | 0.423174 | 0.999532 | | 2.41111 | -0.2081 | 0.91732 | | 2.617689 | -0.64544 | 0.660097 |
| C20orf96 | 2.45591 | 0.155169 | 0.999532 | | 2.484734 | -0.19594 | 0.91732 | | 2.283532 | -0.33258 | 0.836935 |
| SHPRH | 3.030718 | 0.19618 | 0.999532 | | 3.556284 | 0.18862 | 0.91732 | | 3.240016 | 0.01267 | 0.996891 |
| AMN1 | 2.161816 | 0.418816 | 0.999532 | | 2.151202 | 0.207604 | 0.91732 | | 2.191355 | -0.1953 | 0.917014 |
| NINJ1 | 9.547443 | -0.14942 | 0.999532 | | 11.74378 | 0.107433 | 0.91732 | | 9.543068 | 0.27463 | 0.722703 |
| HLA-DMA | 6.900072 | 0.041851 | 0.999532 | | 7.221598 | -0.14104 | 0.91732 | | 6.253526 | -0.16223 | 0.901033 |
| REL | 3.960975 | -0.16777 | 0.999532 | | 4.55445 | -0.15913 | 0.91732 | | 3.541383 | 0.027728 | 0.99186 |
| PIWIL4 | 2.444651 | 0.495351 | 0.999532 | | 2.139266 | -0.20962 | 0.91732 | | 2.229816 | -0.68729 | 0.628053 |
| PIGC | 4.188503 | 0.284171 | 0.999532 | | 4.028916 | -0.1592 | 0.91732 | | 3.957255 | -0.42998 | 0.699971 |
| IFI44L | 3.546099 | 0.158207 | 0.999532 | | 3.702142 | 0.16471 | 0.91732 | | 3.555879 | 0.010175 | 0.996891 |
| SLC52A3 | 2.160437 | 0.206773 | 0.999532 | | 2.050274 | -0.21437 | 0.917612 | | 1.903505 | -0.40791 | 0.814402 |
| MTMR14 | 4.061236 | -0.1227 | 0.999532 | | 4.252484 | -0.1601 | 0.917612 | | 3.6037 | -0.02443 | 0.992286 |
| EFCAB14 | 4.075083 | -0.19497 | 0.999532 | | 4.597801 | -0.1445 | 0.917612 | | 3.534724 | 0.063149 | 0.973209 |
| EXOSC10 | 4.018372 | 0.085752 | 0.999532 | | 3.865649 | -0.16392 | 0.917945 | | 3.612934 | -0.23283 | 0.870663 |
| SLC35A2 | 3.333668 | 0.546601 | 0.999532 | | 2.845409 | 0.177786 | 0.917945 | | 3.25529 | -0.34898 | 0.79686 |
| ACVR1C | 2.217698 | 0.70643 | 0.999532 | | 1.80938 | -0.22964 | 0.917945 | | 2.077809 | -0.9186 | 0.49299 |
| RWDD2B | 3.702975 | 0.03236 | 0.999532 | | 3.85419 | -0.16674 | 0.917974 | | 3.311215 | -0.1822 | 0.907106 |
| LILRA5 | 3.947244 | 0.063689 | 0.999532 | | 4.726537 | 0.157384 | 0.917974 | | 4.050925 | 0.110659 | 0.945129 |
| CCDC69 | 2.265799 | 0.045988 | 0.999532 | | 2.729658 | 0.184188 | 0.917974 | | 2.411848 | 0.160669 | 0.931619 |
| C18orf25 | 3.362413 | 0.023101 | 0.999532 | | 3.563528 | -0.16717 | 0.917974 | | 3.137812 | -0.17474 | 0.910793 |
| TUBG2 | 6.159639 | -0.0024 | 0.999532 | | 6.623763 | -0.12959 | 0.917974 | | 5.783066 | -0.11425 | 0.926957 |
| SOCS2 | 5.020392 | -0.18349 | 0.999532 | | 6.581275 | 0.137671 | 0.917974 | | 5.067119 | 0.338975 | 0.754671 |
| OR9A2 | 2.087392 | 0.762451 | 0.999532 | | 1.637224 | -0.24497 | 0.917974 | | 1.928711 | -0.99023 | 0.489034 |
| CA3 | 2.31798 | -0.04526 | 0.999532 | | 2.426728 | -0.19506 | 0.917974 | | 2.06566 | -0.13615 | 0.948097 |
| PPM1M | 3.931453 | 0.40853 | 0.999532 | | 3.728635 | 0.157642 | 0.917974 | | 3.963066 | -0.23047 | 0.865988 |
| MINDY2 | 7.459588 | 0.175193 | 0.999532 | | 8.390047 | 0.124628 | 0.917974 | | 7.870089 | -0.04151 | 0.977473 |
| LZTS1 | 4.472597 | 0.1975 | 0.999532 | | 4.304843 | -0.1562 | 0.917974 | | 4.173425 | -0.3455 | 0.774747 |
| ABHD14A | 3.571136 | 0.29211 | 0.999532 | | 3.43416 | -0.1738 | 0.917974 | | 3.518964 | -0.45932 | 0.712621 |
| NDUFC2 | 2.50109 | 0.450008 | 0.999532 | | 2.548156 | -0.20844 | 0.917974 | | 2.44578 | -0.6359 | 0.647218 |
| CHCHD4 | 2.598828 | 0.381701 | 0.999532 | | 2.479672 | -0.2006 | 0.917974 | | 2.386105 | -0.56037 | 0.682165 |
| RRAGD | 1.912652 | 0.475733 | 0.999532 | | 2.01111 | 0.216309 | 0.917974 | | 1.947182 | -0.23823 | 0.900376 |
| TMEM160 | 5.53195 | -0.21825 | 0.999532 | | 6.516391 | -0.1394 | 0.917974 | | 5.418921 | 0.118575 | 0.945129 |
| GRM4 | 2.126397 | -0.01106 | 0.999532 | | 2.125503 | -0.20914 | 0.917974 | | 1.961052 | -0.17837 | 0.934354 |
| C6orf52 | 2.247674 | 0.47737 | 0.999532 | | 1.970368 | -0.22509 | 0.917974 | | 2.072961 | -0.68489 | 0.652925 |
| C1QL1 | 2.754843 | -0.101 | 0.999532 | | 2.959154 | -0.19349 | 0.917974 | | 2.471098 | -0.07627 | 0.974588 |
| UBE2E1 | 5.634465 | -0.17713 | 0.999532 | | 6.744901 | 0.130113 | 0.917974 | | 5.644374 | 0.324667 | 0.750371 |
| CLN6 | 3.181415 | 0.358759 | 0.999532 | | 3.131356 | 0.169595 | 0.917974 | | 3.147003 | -0.18677 | 0.907586 |
| CERS3 | 2.308399 | -0.33663 | 0.999532 | | 2.668206 | -0.19053 | 0.917974 | | 1.999274 | 0.162348 | 0.939107 |
| SYBU | 2.208008 | -0.16941 | 0.999532 | | 2.457619 | -0.20962 | 0.917974 | | 1.896253 | -0.02337 | 0.995782 |
| ARHGEF5 | 3.020714 | -0.14731 | 0.999532 | | 3.185343 | -0.17194 | 0.917974 | | 2.691095 | -0.00754 | 0.997967 |
| BDP1 | 3.187952 | -0.88862 | 0.999532 | | 4.392871 | -0.15269 | 0.917974 | | 3.046773 | 0.753035 | 0.515762 |
| PXMP2 | 2.549236 | -0.00447 | 0.999532 | | 2.639761 | -0.18894 | 0.917974 | | 2.262992 | -0.16617 | 0.92697 |
| EXD2 | 3.332526 | -0.0156 | 0.999532 | | 3.861532 | 0.160778 | 0.917974 | | 3.2906 | 0.191956 | 0.900797 |
| RING1 | 7.933574 | -0.27387 | 0.999532 | | 10.17851 | 0.11212 | 0.917974 | | 7.855473 | 0.404994 | 0.603483 |
| LGMN | 7.630127 | -0.24416 | 0.999532 | | 9.681176 | 0.116792 | 0.917974 | | 7.471917 | 0.36761 | 0.740755 |
| RASSF10 | 2.860941 | 0.775553 | 0.999532 | | 2.227282 | -0.22336 | 0.917988 | | 2.619514 | -0.97733 | 0.419745 |
| NBPF15 | 36.23113 | -0.02352 | 0.999532 | | 39.8339 | -0.0755 | 0.917988 | | 34.32319 | -0.03131 | 0.972221 |
| WAC | 6.20693 | 0.123355 | 0.999532 | | 6.723243 | 0.121531 | 0.918143 | | 6.028218 | 0.013916 | 0.995112 |
| DCAF17 | 2.344999 | 0.394943 | 0.999532 | | 2.26675 | -0.20773 | 0.918143 | | 2.228984 | -0.58161 | 0.682485 |
| MKS1 | 2.963756 | 0.381993 | 0.999532 | | 2.735395 | -0.19072 | 0.918143 | | 2.737244 | -0.5549 | 0.673405 |
| ST14 | 2.635458 | 0.131481 | 0.999532 | | 2.772741 | 0.184969 | 0.918166 | | 2.483311 | 0.069085 | 0.975183 |
| MDM4 | 4.406021 | 0.099003 | 0.999532 | | 5.046707 | 0.145163 | 0.918166 | | 4.37915 | 0.064202 | 0.971732 |
| CCDC163 | 2.645974 | 0.350337 | 0.999532 | | 2.608837 | -0.19882 | 0.918166 | | 2.434549 | -0.52745 | 0.705888 |
| FAM78A | 2.793764 | 0.393516 | 0.999532 | | 2.489674 | -0.18889 | 0.918166 | | 2.614134 | -0.56813 | 0.660097 |
| SYCP2L | 3.820361 | 0.075399 | 0.999532 | | 3.842788 | -0.1589 | 0.918192 | | 3.577946 | -0.22393 | 0.879969 |
| TTBK1 | 2.06946 | 0.255549 | 0.999532 | | 1.937292 | -0.21922 | 0.918222 | | 1.878524 | -0.4595 | 0.791466 |
| HIVEP3 | 2.561144 | 0.263572 | 0.999532 | | 2.340499 | -0.19836 | 0.918222 | | 2.370701 | -0.44649 | 0.769793 |
| MRPL43 | 6.075947 | 0.127727 | 0.999532 | | 6.753606 | 0.12835 | 0.918279 | | 5.891335 | 0.017892 | 0.993618 |
| ZADH2 | 3.492387 | 0.164256 | 0.999532 | | 3.904415 | 0.160561 | 0.918279 | | 3.56255 | 0.014454 | 0.996681 |
| DCLRE1A | 2.128789 | 0.825916 | 0.999532 | | 1.944209 | 0.220569 | 0.918329 | | 2.084423 | -0.58328 | 0.68528 |
| ARMC10 | 3.344265 | 0.160442 | 0.999532 | | 3.325147 | -0.17156 | 0.918659 | | 3.004006 | -0.3138 | 0.827267 |
| MXD1 | 3.010115 | 0.482597 | 0.999532 | | 3.020819 | 0.184941 | 0.918659 | | 3.137646 | -0.28088 | 0.860036 |
| AGL | 2.092151 | 0.117113 | 0.999532 | | 2.128156 | -0.20635 | 0.918659 | | 1.942792 | -0.3038 | 0.868512 |
| SKA2 | 4.298975 | 0.228845 | 0.999532 | | 4.560366 | 0.146058 | 0.918659 | | 4.305368 | -0.06851 | 0.96913 |
| KHSRP | 12.19832 | 0.206317 | 0.999532 | | 13.13034 | 0.097238 | 0.918659 | | 12.08734 | -0.06875 | 0.921708 |
| EDA2R | 1.952016 | 0.190123 | 0.999532 | | 1.928698 | -0.22394 | 0.918659 | | 1.838667 | -0.39671 | 0.829716 |
| CCL18 | 2.299046 | 0.154944 | 0.999532 | | 2.319889 | -0.20661 | 0.918659 | | 2.181846 | -0.34272 | 0.846786 |
| RPP25L | 2.923411 | 0.260089 | 0.999532 | | 3.158734 | 0.177578 | 0.918659 | | 2.950601 | -0.067 | 0.974512 |
| FRMPD2 | 1.834996 | -0.00228 | 0.999532 | | 1.904957 | -0.22335 | 0.918659 | | 1.674672 | -0.20579 | 0.92697 |
| ADAR | 8.873032 | -0.37838 | 0.999532 | | 11.38903 | 0.109517 | 0.918659 | | 8.744051 | 0.504562 | 0.499673 |
| MRPS12 | 3.073418 | -0.43209 | 0.999532 | | 3.866637 | -0.15861 | 0.918659 | | 2.827658 | 0.294408 | 0.842911 |
| DENND4B | 5.242927 | 0.337004 | 0.999532 | | 5.129975 | -0.14651 | 0.918659 | | 5.058 | -0.46423 | 0.647218 |
| SHC1 | 8.554386 | -0.40986 | 0.999532 | | 10.72334 | -0.11572 | 0.918669 | | 7.881442 | 0.311014 | 0.717627 |
| SLC11A2 | 3.974904 | 0.301919 | 0.999532 | | 3.92865 | 0.151758 | 0.918759 | | 3.936411 | -0.12831 | 0.931827 |
| KCTD2 | 3.79116 | 0.092762 | 0.999532 | | 3.720498 | -0.15983 | 0.918759 | | 3.472523 | -0.23037 | 0.876428 |
| DDX18 | 7.841979 | -0.43198 | 0.999532 | | 10.11366 | 0.111109 | 0.918759 | | 7.452098 | 0.560554 | 0.434766 |
| PDF | 2.924445 | 0.112217 | 0.999532 | | 3.137647 | -0.17772 | 0.918759 | | 2.736729 | -0.26354 | 0.868512 |
| ITGB2 | 2.915724 | 0.278238 | 0.999532 | | 2.98016 | 0.168955 | 0.918759 | | 2.855122 | -0.09672 | 0.958863 |
| SUDS3 | 3.705039 | 0.183063 | 0.999532 | | 4.146845 | 0.165835 | 0.918759 | | 3.778142 | -0.00267 | 0.999385 |
| DDX47 | 5.137283 | 0.177901 | 0.999532 | | 5.84767 | 0.135735 | 0.91882 | | 5.291347 | -0.02333 | 0.991341 |
| SYCE1L | 1.987261 | 0.073935 | 0.999532 | | 2.04122 | -0.22213 | 0.91882 | | 1.844827 | -0.27813 | 0.892869 |
| RFX8 | 1.896746 | -0.26844 | 0.999532 | | 2.108248 | -0.21272 | 0.91882 | | 1.686192 | 0.073563 | 0.976487 |
| GATA6 | 2.322517 | -0.47369 | 0.999532 | | 2.845272 | -0.18926 | 0.91882 | | 2.012531 | 0.302499 | 0.869449 |
| RCN3 | 5.585551 | -0.30279 | 0.999532 | | 7.39812 | 0.137223 | 0.918867 | | 5.642652 | 0.463366 | 0.644433 |
| MORN1 | 2.688036 | 0.090436 | 0.999532 | | 2.752065 | -0.1831 | 0.918924 | | 2.493451 | -0.255 | 0.87764 |
| HSPA2 | 3.839888 | -0.07542 | 0.999532 | | 4.037152 | -0.15958 | 0.919069 | | 3.338812 | -0.06252 | 0.974421 |
| WDR11 | 5.193155 | -0.1645 | 0.999532 | | 6.258779 | 0.143619 | 0.919132 | | 5.235589 | 0.343602 | 0.730921 |
| BCL2L13 | 6.244823 | 0.202056 | 0.999532 | | 6.67583 | 0.12173 | 0.919132 | | 6.142989 | -0.06322 | 0.964357 |
| TRMT10B | 2.903258 | 0.325273 | 0.999532 | | 2.730891 | -0.18261 | 0.919132 | | 2.773563 | -0.48574 | 0.723933 |
| IZUMO2 | 2.14022 | 0.457784 | 0.999532 | | 2.062425 | 0.20589 | 0.919132 | | 2.113674 | -0.23998 | 0.895865 |
| ALG5 | 4.204834 | -0.07924 | 0.999532 | | 4.843926 | 0.139621 | 0.919132 | | 4.169745 | 0.233096 | 0.860036 |
| PPP1R32 | 2.264487 | -0.02364 | 0.999532 | | 2.408531 | -0.19617 | 0.919132 | | 2.044357 | -0.15355 | 0.941197 |
| UBE2NL | 3.509712 | 0.241004 | 0.999532 | | 3.322317 | -0.17261 | 0.919137 | | 3.264135 | -0.39274 | 0.760992 |
| ATP1A2 | 4.171314 | -0.26694 | 0.999532 | | 5.450834 | 0.147974 | 0.919289 | | 4.324631 | 0.42794 | 0.689408 |
| PABPC1 | 31.45642 | -0.34686 | 0.999532 | | 40.92642 | 0.097364 | 0.919289 | | 30.90179 | 0.468011 | 0.475974 |
| TTC33 | 8.890988 | -0.18635 | 0.999532 | | 10.98324 | 0.145735 | 0.919289 | | 9.043078 | 0.339686 | 0.759304 |
| WNT5A | 1.895957 | 0.041408 | 0.999532 | | 2.170903 | 0.207323 | 0.919354 | | 1.87917 | 0.182374 | 0.93042 |
| ZNF75D | 2.769107 | 0.546849 | 0.999532 | | 2.193076 | -0.1991 | 0.919591 | | 2.400298 | -0.73758 | 0.560424 |
| RRAGB | 2.168941 | -0.5952 | 0.999532 | | 2.961193 | 0.172946 | 0.919604 | | 2.164888 | 0.782221 | 0.549065 |
| UCMA | 2.449109 | 0.243089 | 0.999532 | | 2.300794 | -0.21329 | 0.919604 | | 2.121822 | -0.43626 | 0.794815 |
| SNRPD1 | 4.004875 | -0.3038 | 0.999532 | | 5.329238 | 0.156527 | 0.919712 | | 4.085535 | 0.475539 | 0.664512 |
| PSKH1 | 2.788047 | 0.069874 | 0.999532 | | 2.990244 | -0.18119 | 0.919712 | | 2.520492 | -0.22528 | 0.893262 |
| PIGB | 2.209562 | 0.134776 | 0.999532 | | 2.171649 | -0.20526 | 0.919779 | | 2.069387 | -0.32029 | 0.864245 |
| SON | 12.02617 | -0.32279 | 0.999532 | | 15.28853 | 0.100546 | 0.919779 | | 11.96457 | 0.429484 | 0.552672 |
| JSRP1 | 2.307984 | 0.341293 | 0.999532 | | 2.248565 | -0.22024 | 0.919779 | | 2.134483 | -0.53959 | 0.734912 |
| CCDC71L | 3.116696 | 0.047543 | 0.999532 | | 3.377199 | -0.16558 | 0.919786 | | 2.852338 | -0.18879 | 0.904804 |
| RCAN3 | 2.574594 | -0.1645 | 0.999532 | | 2.699141 | -0.19116 | 0.919786 | | 2.246113 | -0.00378 | 0.999385 |
| C8G | 1.888992 | 0.001957 | 0.999532 | | 1.94809 | -0.21707 | 0.919786 | | 1.717442 | -0.20137 | 0.926586 |
| SLC9A4 | 2.300184 | 0.072608 | 0.999532 | | 2.149375 | -0.2114 | 0.9198 | | 2.026325 | -0.26939 | 0.891051 |
| FRYL | 4.047886 | -0.22673 | 0.999532 | | 5.308742 | 0.13609 | 0.9198 | | 4.17341 | 0.381548 | 0.722703 |
| MRFAP1L | 4.103625 | -0.1042 | 0.999532 | | 4.563566 | -0.14283 | 0.9198 | | 3.745198 | -0.0198 | 0.993618 |
| CCDC194 | 2.702453 | -0.24963 | 0.999532 | | 3.135955 | -0.17329 | 0.919852 | | 2.567788 | 0.094303 | 0.96556 |
| GYS1 | 3.692367 | 0.15307 | 0.999532 | | 3.698221 | -0.15774 | 0.919852 | | 3.332265 | -0.2917 | 0.825181 |
| NOSTRIN | 7.955963 | -0.06902 | 0.999532 | | 8.351867 | -0.12673 | 0.919852 | | 7.040368 | -0.03769 | 0.982124 |
| GPC2 | 2.339597 | 0.363623 | 0.999532 | | 2.229841 | -0.21353 | 0.919852 | | 2.076194 | -0.55176 | 0.720106 |
| FAM214A | 2.615007 | 0.072066 | 0.999532 | | 3.304697 | 0.181445 | 0.919857 | | 2.888102 | 0.128286 | 0.946512 |
| WDR74 | 2.682209 | 0.400814 | 0.999532 | | 2.439478 | -0.1903 | 0.919857 | | 2.529409 | -0.57209 | 0.66549 |
| ROGDI | 5.973268 | 0.108148 | 0.999532 | | 6.152992 | -0.14589 | 0.919913 | | 5.639668 | -0.23973 | 0.849608 |
| PPA1 | 10.27521 | -0.28427 | 0.999532 | | 12.73 | 0.099538 | 0.920084 | | 9.866142 | 0.40624 | 0.543177 |
| KDM1A | 4.560974 | 0.229855 | 0.999532 | | 5.17339 | 0.135527 | 0.920218 | | 4.783091 | -0.07302 | 0.962597 |
| AP1S3 | 3.863955 | 0.636382 | 0.999532 | | 3.425862 | 0.169497 | 0.920218 | | 3.769377 | -0.46378 | 0.718643 |
| DEFB121 | 2.356582 | 0.22895 | 0.999532 | | 2.440378 | -0.20117 | 0.920218 | | 2.285767 | -0.40701 | 0.801905 |
| NOSIP | 4.723903 | -0.22644 | 0.999532 | | 5.838998 | -0.14186 | 0.920218 | | 4.484227 | 0.106321 | 0.945129 |
| TSPAN32 | 3.53528 | -0.11904 | 0.999532 | | 3.918966 | -0.1559 | 0.920218 | | 3.294695 | -0.01762 | 0.996149 |
| TCF15 | 2.09487 | 0.180162 | 0.999532 | | 2.621568 | 0.199695 | 0.920218 | | 2.246376 | 0.033455 | 0.991434 |
| SLC39A1- | 2.542586 | -0.00111 | 0.999532 | | 2.669839 | -0.18112 | 0.920218 | | 2.171838 | -0.16653 | 0.928522 |
| ERVFRD- | 1.925312 | 0.422972 | 0.999532 | | 2.095923 | 0.218399 | 0.920218 | | 2.101459 | -0.18562 | 0.929027 |
| UBE2G2 | 7.946134 | 0.227188 | 0.999532 | | 8.554873 | 0.116707 | 0.920218 | | 7.798271 | -0.0902 | 0.942744 |
| KCNQ2 | 3.148937 | 0.122804 | 0.999532 | | 3.290536 | -0.18439 | 0.920218 | | 2.986003 | -0.28519 | 0.862823 |
| BORCS5 | 2.15782 | -0.04654 | 0.999532 | | 2.344999 | -0.19842 | 0.920218 | | 1.952684 | -0.13063 | 0.953949 |
| LIG1 | 2.205361 | -0.24858 | 0.999532 | | 2.843947 | 0.18659 | 0.920218 | | 2.102037 | 0.457864 | 0.763092 |
| ACOT9 | 2.266398 | -0.29187 | 0.999532 | | 2.466508 | -0.19784 | 0.920218 | | 1.905331 | 0.119132 | 0.958798 |
| TMEM259 | 3.420091 | 0.066207 | 0.999532 | | 3.823222 | 0.157799 | 0.920218 | | 3.370701 | 0.099954 | 0.957482 |
| MPHOSPH | 3.269769 | 0.298012 | 0.999532 | | 3.573178 | 0.174041 | 0.920218 | | 3.436082 | -0.11285 | 0.947851 |
| SGK3 | 3.824638 | 0.259207 | 0.999532 | | 3.880993 | 0.153597 | 0.920218 | | 3.718814 | -0.08812 | 0.959437 |
| HIF1AN | 6.464822 | -0.26414 | 0.999532 | | 7.236813 | -0.12405 | 0.920218 | | 5.62864 | 0.151512 | 0.901813 |
| TMED7 | 5.498479 | -0.21043 | 0.999532 | | 6.771241 | 0.13533 | 0.920218 | | 5.517325 | 0.368216 | 0.725814 |
| EPHA3 | 2.690931 | 0.427102 | 0.999532 | | 2.420283 | -0.19726 | 0.92026 | | 2.52937 | -0.60424 | 0.662584 |
| SKA1 | 1.907673 | -0.17739 | 0.999532 | | 2.396989 | 0.200729 | 0.92026 | | 2.01477 | 0.39862 | 0.826048 |
| GIGYF2 | 2.574821 | 0.003333 | 0.999532 | | 3.191747 | 0.167802 | 0.9203 | | 2.670192 | 0.18483 | 0.909157 |
| KLHL28 | 2.685422 | 0.232926 | 0.999532 | | 3.01339 | 0.175872 | 0.9203 | | 2.875074 | -0.0389 | 0.988853 |
| MACROH | 2.880425 | 0.038069 | 0.999532 | | 3.533421 | 0.16169 | 0.9203 | | 2.925096 | 0.14286 | 0.930403 |
| KCNH6 | 2.09681 | 0.024188 | 0.999532 | | 2.142525 | -0.21569 | 0.9203 | | 1.914437 | -0.22493 | 0.914687 |
| SPNS1 | 4.94985 | 0.445757 | 0.999532 | | 4.553607 | -0.14843 | 0.920884 | | 4.680372 | -0.57828 | 0.532625 |
| NEURL3 | 2.879337 | 0.357013 | 0.999532 | | 2.664064 | -0.18783 | 0.92093 | | 2.563307 | -0.52548 | 0.702463 |
| ROMO1 | 3.456286 | 0.292064 | 0.999532 | | 3.715157 | 0.161687 | 0.92093 | | 3.403504 | -0.11118 | 0.946572 |
| TNFSF18 | 2.380163 | -0.08225 | 0.999532 | | 2.40005 | -0.19715 | 0.92093 | | 2.288875 | -0.1039 | 0.968235 |
| DNTTIP2 | 4.506352 | 0.122671 | 0.999532 | | 4.360395 | -0.15189 | 0.92093 | | 4.108777 | -0.25023 | 0.859522 |
| ARHGEF3 | 3.019287 | -0.10335 | 0.999532 | | 3.769731 | 0.17408 | 0.92093 | | 3.087588 | 0.297263 | 0.843564 |
| IPMK | 2.605191 | 0.11121 | 0.999532 | | 2.666433 | -0.18094 | 0.92093 | | 2.465841 | -0.27187 | 0.871595 |
| IARS2 | 4.401916 | 0.336101 | 0.999532 | | 4.641497 | 0.142852 | 0.92093 | | 4.285898 | -0.17212 | 0.893262 |
| PPCDC | 3.676978 | -0.31407 | 0.999532 | | 4.276836 | -0.15243 | 0.92093 | | 3.344801 | 0.178679 | 0.907626 |
| SLC43A2 | 4.551382 | -0.00141 | 0.999532 | | 5.42069 | 0.146017 | 0.92093 | | 4.4563 | 0.163517 | 0.901033 |
| NPEPPS | 3.777529 | 0.033369 | 0.999532 | | 4.086992 | -0.16759 | 0.92093 | | 3.51802 | -0.1884 | 0.898369 |
| C1orf146 | 2.215626 | 0.43767 | 0.999532 | | 2.057658 | -0.22246 | 0.92093 | | 2.032007 | -0.63628 | 0.681871 |
| GOLGA2 | 9.361965 | -0.132 | 0.999532 | | 10.30661 | -0.10533 | 0.920946 | | 8.660709 | 0.050819 | 0.970811 |
| KRIT1 | 3.214395 | -0.32711 | 0.999532 | | 4.256123 | 0.157819 | 0.920946 | | 3.258743 | 0.505889 | 0.699971 |
| CCDC13 | 8.28816 | 0.541327 | 0.999532 | | 7.949807 | 0.147219 | 0.921006 | | 8.334514 | -0.37143 | 0.784602 |
| RHBDL1 | 2.127799 | 0.000974 | 0.999532 | | 2.035101 | -0.21103 | 0.921115 | | 1.907093 | -0.19757 | 0.92697 |
| GALK1 | 2.763792 | 0.16345 | 0.999532 | | 2.938841 | 0.18205 | 0.921115 | | 2.63982 | 0.042434 | 0.987274 |
| ERGIC3 | 7.674363 | 0.198344 | 0.999532 | | 8.272412 | 0.121645 | 0.921115 | | 7.463385 | -0.05229 | 0.973946 |
| VPS18 | 2.744438 | 0.11821 | 0.999532 | | 3.290319 | 0.168713 | 0.921115 | | 2.76863 | 0.069899 | 0.97386 |
| MARCHF | 2.576052 | -0.50643 | 0.999532 | | 3.330694 | -0.17775 | 0.921115 | | 2.365415 | 0.3529 | 0.835554 |
| SLC6A11 | 2.282765 | 0.314011 | 0.999532 | | 2.404211 | 0.200539 | 0.921115 | | 2.418515 | -0.08829 | 0.972305 |
| BTBD10 | 3.836296 | -0.39213 | 0.999532 | | 4.5878 | -0.14102 | 0.921115 | | 3.429598 | 0.266026 | 0.843564 |
| RPS29 | 26.91415 | -0.13543 | 0.999532 | | 31.71137 | 0.069621 | 0.92113 | | 25.60163 | 0.217036 | 0.703689 |
| ICOS | 1.99415 | 0.098792 | 0.999532 | | 1.916757 | -0.21399 | 0.92132 | | 1.835667 | -0.29449 | 0.883565 |
| IFT57 | 4.995736 | 0.148229 | 0.999532 | | 5.189197 | 0.141467 | 0.92132 | | 4.686467 | 0.011351 | 0.996891 |
| NOL9 | 4.616503 | -0.04205 | 0.999532 | | 5.110796 | -0.14471 | 0.921409 | | 4.515425 | -0.09025 | 0.956011 |
| ERP27 | 4.850536 | -0.19406 | 0.999532 | | 5.331662 | -0.146 | 0.921409 | | 4.184522 | 0.061801 | 0.974421 |
| ADCY9 | 5.406451 | -0.38721 | 0.999532 | | 6.583008 | -0.1386 | 0.921418 | | 4.838845 | 0.277998 | 0.824012 |
| PARP2 | 4.343147 | -0.2831 | 0.999532 | | 4.784749 | -0.14859 | 0.921418 | | 3.708583 | 0.1501 | 0.922679 |
| KIF5C | 2.304669 | -0.07903 | 0.999532 | | 2.629534 | -0.19833 | 0.921418 | | 2.24631 | -0.11699 | 0.957663 |
| TFAM | 3.970397 | 0.223539 | 0.999532 | | 4.04678 | -0.14716 | 0.921418 | | 3.555547 | -0.34921 | 0.760473 |
| PMPCA | 3.438071 | 0.294479 | 0.999532 | | 3.62996 | 0.162302 | 0.921418 | | 3.355017 | -0.11176 | 0.945857 |
| NINL | 2.272124 | 0.375817 | 0.999532 | | 2.548974 | 0.186928 | 0.921418 | | 2.40964 | -0.16885 | 0.924131 |
| TNFRSF1 | 4.761078 | -0.2808 | 0.999532 | | 5.445451 | -0.15135 | 0.921418 | | 4.077048 | 0.15175 | 0.920259 |
| COL26A1 | 2.555192 | 0.117314 | 0.999532 | | 2.500254 | -0.19139 | 0.921418 | | 2.341284 | -0.29624 | 0.869666 |
| ARL6IP6 | 4.291439 | 0.487942 | 0.999532 | | 4.132402 | 0.157595 | 0.921418 | | 4.325615 | -0.30574 | 0.794815 |
| CYP2S1 | 2.356335 | 0.292599 | 0.999532 | | 2.230493 | 0.205309 | 0.921418 | | 2.199688 | -0.06587 | 0.979239 |
| ZCCHC9 | 2.616897 | 0.493415 | 0.999532 | | 2.641103 | 0.197802 | 0.921418 | | 2.650839 | -0.27413 | 0.87591 |
| KATNB1 | 3.067034 | 0.339597 | 0.999532 | | 2.944435 | -0.17052 | 0.921418 | | 2.827132 | -0.49058 | 0.691311 |
| ZNF781 | 4.031986 | 0.58395 | 0.999532 | | 3.482124 | -0.16344 | 0.921418 | | 3.81941 | -0.72836 | 0.458377 |
| MBD4 | 3.477612 | -0.30076 | 0.999532 | | 4.17763 | -0.15367 | 0.921418 | | 3.110833 | 0.163774 | 0.915533 |
| UBIAD1 | 3.22424 | 0.233893 | 0.999532 | | 3.168197 | -0.17557 | 0.921418 | | 2.964673 | -0.39074 | 0.776321 |
| PDE6D | 3.643569 | -0.15183 | 0.999532 | | 4.73095 | 0.143679 | 0.921418 | | 3.791905 | 0.313024 | 0.797276 |
| ANXA4 | 3.075625 | -0.05876 | 0.999532 | | 3.811169 | 0.162922 | 0.921418 | | 3.166867 | 0.245939 | 0.872493 |
| SHISA7 | 2.698031 | -0.14521 | 0.999532 | | 2.801522 | -0.19471 | 0.921418 | | 2.33318 | -0.02384 | 0.995112 |
| PSTK | 3.918584 | 0.350481 | 0.999532 | | 3.684453 | -0.16947 | 0.921418 | | 3.611442 | -0.50399 | 0.662952 |
| RECQL5 | 4.65758 | 0.106742 | 0.999532 | | 5.235626 | 0.139118 | 0.921418 | | 4.582182 | 0.052388 | 0.975194 |
| ZFR | 6.282376 | -0.1571 | 0.999532 | | 7.836279 | 0.12236 | 0.921418 | | 6.115372 | 0.308282 | 0.746666 |
| MED13 | 11.67785 | -0.18217 | 0.999532 | | 13.56699 | -0.0988 | 0.92145 | | 10.66885 | 0.098296 | 0.923266 |
| XKR8 | 3.263284 | 0.183032 | 0.999532 | | 4.069617 | 0.166027 | 0.92145 | | 3.519368 | 0.002048 | 0.999385 |
| OR51J1 | 2.302108 | 0.519353 | 0.999532 | | 2.297043 | 0.205577 | 0.92145 | | 2.222457 | -0.28913 | 0.869666 |
| TXNRD2 | 2.546032 | -0.02979 | 0.999532 | | 2.740078 | -0.18763 | 0.92145 | | 2.279287 | -0.13668 | 0.946572 |
| IL23A | 2.591884 | 0.238457 | 0.999532 | | 2.38328 | -0.20151 | 0.92145 | | 2.43696 | -0.42715 | 0.806394 |
| MRPL38 | 5.169625 | 0.264701 | 0.999532 | | 5.63045 | 0.130653 | 0.92145 | | 4.978101 | -0.1126 | 0.931942 |
| METTL2B | 4.204703 | 0.31141 | 0.999532 | | 4.597516 | 0.146681 | 0.92145 | | 4.183351 | -0.14742 | 0.912963 |
| HP | 3.293857 | 0.116017 | 0.999532 | | 3.215509 | -0.17067 | 0.92145 | | 3.161432 | -0.26535 | 0.876291 |
| ZNF445 | 3.380191 | -0.07299 | 0.999532 | | 3.844296 | 0.154439 | 0.92145 | | 3.3948 | 0.235676 | 0.879947 |
| DPP7 | 6.44393 | 0.255593 | 0.999532 | | 6.696766 | 0.13978 | 0.92145 | | 6.217723 | -0.09325 | 0.948127 |
| MEDAG | 2.416855 | -0.02708 | 0.999532 | | 2.388489 | -0.20117 | 0.92145 | | 2.221575 | -0.15669 | 0.944635 |
| MAZ | 55.72606 | 0.052607 | 0.999532 | | 59.34172 | -0.11746 | 0.92145 | | 51.78328 | -0.15132 | 0.87591 |
| BICRA | 2.911908 | -0.28166 | 0.999532 | | 3.5598 | -0.16558 | 0.92145 | | 2.63787 | 0.140455 | 0.939913 |
| PARP11 | 2.713988 | 0.126773 | 0.999532 | | 2.705797 | -0.18087 | 0.92145 | | 2.530806 | -0.2913 | 0.859522 |
| N4BP3 | 2.233333 | 0.143329 | 0.999532 | | 2.310402 | -0.19881 | 0.921761 | | 1.982231 | -0.31712 | 0.860036 |
| CYP2W1 | 5.280343 | -0.07558 | 0.999532 | | 5.674945 | -0.13589 | 0.921761 | | 4.947789 | -0.0352 | 0.98811 |
| DKC1 | 2.809732 | 0.438646 | 0.999532 | | 2.848938 | 0.175602 | 0.921772 | | 2.975252 | -0.24757 | 0.873155 |
| RABEPK | 4.13243 | 0.121983 | 0.999532 | | 4.604086 | 0.141492 | 0.921809 | | 4.110849 | 0.038724 | 0.98506 |
| AKIRIN1 | 5.416716 | -0.10144 | 0.999532 | | 6.165903 | -0.12529 | 0.921809 | | 4.922531 | -0.01084 | 0.996681 |
| STYK1 | 4.148649 | -0.23004 | 0.999532 | | 5.12057 | 0.142505 | 0.922037 | | 4.133703 | 0.369064 | 0.791576 |
| CFAP410 | 3.144287 | -0.22184 | 0.999532 | | 3.470564 | -0.16285 | 0.922037 | | 2.63298 | 0.080091 | 0.969693 |
| GLIS3 | 4.799709 | -0.28203 | 0.999532 | | 5.90731 | 0.127736 | 0.922191 | | 4.855994 | 0.427708 | 0.683901 |
| TCOF1 | 4.21578 | 0.230826 | 0.999532 | | 4.560811 | 0.143659 | 0.922377 | | 4.335999 | -0.05874 | 0.974421 |
| MAOA | 3.498034 | 0.021267 | 0.999532 | | 3.500377 | -0.16027 | 0.922377 | | 3.086455 | -0.17369 | 0.915533 |
| ACRBP | 2.24481 | 0.176104 | 0.999532 | | 2.095533 | -0.19717 | 0.922377 | | 2.142676 | -0.35686 | 0.840699 |
| MBIP | 2.765759 | 0.157002 | 0.999532 | | 2.907229 | -0.17546 | 0.922377 | | 2.577707 | -0.31264 | 0.837256 |
| PGM2L1 | 2.117649 | 0.018803 | 0.999532 | | 2.383509 | -0.19771 | 0.922377 | | 2.02397 | -0.20551 | 0.914687 |
| MPZL1 | 5.069965 | -0.30815 | 0.999532 | | 6.006124 | -0.12465 | 0.922377 | | 4.674348 | 0.199387 | 0.87591 |
| GCNA | 2.564067 | 0.113369 | 0.999532 | | 2.600367 | -0.18972 | 0.922377 | | 2.327762 | -0.28911 | 0.872493 |
| ZSWIM7 | 2.180169 | -0.29596 | 0.999532 | | 2.565823 | -0.18976 | 0.922377 | | 2.03627 | 0.123433 | 0.958191 |
| ZNF664 | 3.484947 | 0.210863 | 0.999532 | | 3.715131 | 0.151469 | 0.922377 | | 3.426571 | -0.04556 | 0.982809 |
| STOX2 | 2.373469 | 0.373178 | 0.999532 | | 2.200513 | -0.19606 | 0.922377 | | 2.053429 | -0.55237 | 0.702463 |
| C20orf27 | 2.237855 | 0.130452 | 0.999532 | | 2.56913 | 0.18701 | 0.922377 | | 2.234311 | 0.075174 | 0.974421 |
| TES | 2.65624 | -0.03426 | 0.999532 | | 3.115187 | 0.163967 | 0.922377 | | 2.666051 | 0.214157 | 0.896374 |
| PUF60 | 7.409515 | -0.26135 | 0.999532 | | 9.527752 | 0.103424 | 0.922377 | | 7.220463 | 0.38349 | 0.647218 |
| SLC48A1 | 8.651036 | 0.022667 | 0.999532 | | 9.540592 | 0.105407 | 0.922377 | | 8.436913 | 0.090165 | 0.940719 |
| SCNN1B | 2.638793 | 0.602487 | 0.999532 | | 2.567354 | 0.180822 | 0.922377 | | 2.572907 | -0.40041 | 0.767781 |
| CPPED1 | 2.432277 | 0.277538 | 0.999532 | | 2.179538 | -0.19349 | 0.922377 | | 2.257703 | -0.45234 | 0.765312 |
| LSS | 3.192959 | -0.051 | 0.999532 | | 3.490764 | -0.15716 | 0.922377 | | 3.056505 | -0.06467 | 0.966652 |
| NDUFA7 | 6.510395 | 0.581704 | 0.999532 | | 6.482671 | 0.132079 | 0.922377 | | 6.684035 | -0.43132 | 0.629006 |
| ZNF367 | 2.521416 | -0.11019 | 0.999532 | | 3.115117 | 0.174851 | 0.922377 | | 2.489705 | 0.305766 | 0.850438 |
| ZSWIM5 | 2.760982 | 0.043121 | 0.999532 | | 2.858983 | -0.18194 | 0.922377 | | 2.572797 | -0.20605 | 0.907626 |
| FERMT3 | 3.032218 | -0.39214 | 0.999532 | | 3.592858 | -0.15818 | 0.922377 | | 2.780457 | 0.250273 | 0.876781 |
| CUTC | 2.300384 | -0.23938 | 0.999532 | | 2.743709 | -0.18665 | 0.922377 | | 2.098762 | 0.0711 | 0.977473 |
| PABPC4 | 5.035683 | 0.317151 | 0.999532 | | 5.015246 | 0.131491 | 0.922377 | | 4.881838 | -0.17516 | 0.890746 |
| ZBED1 | 2.414044 | -0.19499 | 0.999532 | | 2.619883 | -0.18371 | 0.922377 | | 2.2858 | 0.031794 | 0.992286 |
| NUP54 | 3.965157 | 0.279328 | 0.999532 | | 4.287216 | 0.143572 | 0.922377 | | 4.077653 | -0.11394 | 0.941197 |
| TBC1D9 | 4.644047 | -0.14367 | 0.999532 | | 5.455262 | -0.13195 | 0.922377 | | 4.232913 | 0.037197 | 0.985825 |
| C5orf52 | 2.192812 | 0.687244 | 0.999532 | | 2.022769 | 0.213707 | 0.922377 | | 2.326973 | -0.45114 | 0.790999 |
| DUSP16 | 2.555186 | 0.193178 | 0.999532 | | 2.508614 | -0.18311 | 0.922377 | | 2.422558 | -0.35973 | 0.816112 |
| FAM177A | 4.400193 | 0.11944 | 0.999532 | | 4.959898 | 0.138146 | 0.922377 | | 4.383172 | 0.03645 | 0.985693 |
| GET3 | 8.796341 | -0.09208 | 0.999532 | | 10.31613 | 0.101575 | 0.922377 | | 8.63401 | 0.21076 | 0.816618 |
| PNMA1 | 4.018187 | 0.590894 | 0.999532 | | 3.769415 | 0.153132 | 0.922377 | | 3.916507 | -0.42148 | 0.715437 |
| DCTN4 | 4.284229 | -0.02489 | 0.999532 | | 4.574241 | -0.14318 | 0.922377 | | 3.810717 | -0.1022 | 0.948141 |
| TMEM132 | 2.313317 | 0.082367 | 0.999532 | | 2.232025 | -0.19993 | 0.922377 | | 2.110412 | -0.25024 | 0.89433 |
| JAG2 | 3.356252 | -0.20568 | 0.999532 | | 4.100153 | 0.147308 | 0.922377 | | 3.453842 | 0.370296 | 0.765852 |
| BTRC | 2.837298 | 0.400594 | 0.999532 | | 2.907486 | 0.176683 | 0.922377 | | 2.723519 | -0.20273 | 0.895865 |
| CFAP298 | 3.809659 | -0.10738 | 0.999532 | | 4.85125 | 0.147825 | 0.922377 | | 4.155638 | 0.263083 | 0.842754 |
| EBP | 3.38026 | 0.218056 | 0.999532 | | 3.287681 | -0.15919 | 0.922377 | | 3.020315 | -0.35957 | 0.780456 |
| PPP1R11 | 3.416814 | 0.520304 | 0.999532 | | 3.231203 | 0.164357 | 0.922377 | | 3.422812 | -0.3381 | 0.791576 |
| YJEFN3 | 4.396782 | 0.015714 | 0.999532 | | 4.911792 | -0.13909 | 0.922377 | | 4.069174 | -0.12048 | 0.939549 |
| SULT2A1 | 2.242542 | -0.01052 | 0.999532 | | 2.378214 | -0.18841 | 0.922377 | | 2.060132 | -0.16078 | 0.938429 |
| CDC42EP | 2.66495 | -0.45304 | 0.999532 | | 3.252773 | -0.16325 | 0.922377 | | 2.394283 | 0.307588 | 0.845923 |
| DRD3 | 1.727387 | 0.267568 | 0.999532 | | 2.007111 | 0.2129 | 0.922377 | | 1.850956 | -0.02959 | 0.993618 |
| CYP7B1 | 3.375366 | -0.12474 | 0.999532 | | 4.098761 | 0.154315 | 0.922377 | | 3.258223 | 0.297811 | 0.827267 |
| ZNF624 | 3.121851 | -0.1516 | 0.999532 | | 3.430875 | -0.16663 | 0.922377 | | 2.684553 | 0.001937 | 0.999429 |
| SLC4A1AP | 5.095357 | -0.33713 | 0.999532 | | 5.892753 | -0.12551 | 0.922377 | | 4.609022 | 0.226433 | 0.858705 |
| ATF7IP | 8.469479 | 0.140908 | 0.999532 | | 9.339818 | 0.117754 | 0.922377 | | 8.303145 | -0.00138 | 0.999385 |
| CELSR1 | 3.351465 | 0.259197 | 0.999532 | | 3.725194 | 0.152756 | 0.922377 | | 3.340348 | -0.08422 | 0.963492 |
| MESP2 | 2.507012 | 0.026016 | 0.999532 | | 2.472626 | -0.19344 | 0.922377 | | 2.30197 | -0.20673 | 0.915533 |
| PSPH | 3.920458 | 0.070041 | 0.999532 | | 4.236345 | -0.14835 | 0.922377 | | 3.634973 | -0.19544 | 0.890109 |
| SH3TC1 | 3.873687 | -0.25674 | 0.999532 | | 4.686926 | 0.143084 | 0.922377 | | 3.767212 | 0.419463 | 0.717627 |
| CLCC1 | 4.075697 | -0.02074 | 0.999532 | | 4.839344 | 0.137637 | 0.922377 | | 4.046549 | 0.177013 | 0.893828 |
| PER2 | 2.251892 | -0.37008 | 0.999532 | | 2.846753 | -0.2163 | 0.922377 | | 2.054807 | 0.158508 | 0.943463 |
| RXYLT1 | 3.068288 | 0.285381 | 0.999532 | | 3.052532 | -0.17593 | 0.922377 | | 2.953267 | -0.44559 | 0.735469 |
| BEST2 | 2.567207 | 0.367965 | 0.999532 | | 2.200075 | -0.21043 | 0.922377 | | 2.279343 | -0.57854 | 0.722703 |
| MYLPF | 2.128844 | 0.046668 | 0.999532 | | 2.074985 | -0.20055 | 0.922377 | | 1.9227 | -0.23427 | 0.905952 |
| RSAD2 | 2.500403 | 0.237582 | 0.999532 | | 2.80544 | 0.185565 | 0.922377 | | 2.493903 | -0.03009 | 0.992286 |
| AK1 | 5.981073 | 0.139172 | 0.999532 | | 6.670863 | 0.120983 | 0.922377 | | 5.91718 | -0.00017 | 0.999949 |
| ERLIN1 | 3.146853 | -0.4294 | 0.999532 | | 3.725267 | -0.15384 | 0.922377 | | 2.803067 | 0.292666 | 0.843564 |
| FBXO17 | 2.981641 | 0.985748 | 0.999532 | | 2.445335 | 0.195754 | 0.922377 | | 2.833747 | -0.75809 | 0.49299 |
| ANKRD49 | 2.27082 | 0.11236 | 0.999532 | | 2.571962 | 0.180689 | 0.922377 | | 2.285516 | 0.08617 | 0.969691 |
| SPATA45 | 2.017589 | 0.483966 | 0.999532 | | 1.989141 | -0.21503 | 0.922377 | | 1.874742 | -0.6741 | 0.657538 |

| | | | | | | | | | | | |
|---|---|---|---|---|---|---|---|---|---|---|---|
| CD2AP | 4.885514 | -0.18234 | 0.999532 | | 6.051569 | 0.127998 | 0.922377 | | 4.713179 | 0.328908 | 0.748204 |
| CFAP69 | 2.306628 | 0.005606 | 0.999532 | | 2.378954 | -0.18447 | 0.922377 | | 2.067011 | -0.1725 | 0.927668 |
| E2F3 | 3.653524 | 0.187355 | 0.999532 | | 3.516645 | -0.16409 | 0.922377 | | 3.477684 | -0.34829 | 0.799944 |
| CNOT7 | 6.240269 | 0.079282 | 0.999532 | | 6.616136 | -0.12681 | 0.922377 | | 5.788232 | -0.19243 | 0.863134 |
| OR4C15 | 2.077454 | 0.274122 | 0.999532 | | 1.941287 | -0.21254 | 0.922377 | | 1.90334 | -0.47129 | 0.783332 |
| RBM18 | 1.923647 | 0.25491 | 0.999532 | | 1.829026 | -0.21617 | 0.922377 | | 1.784084 | -0.45516 | 0.795612 |
| GYPB | 2.336436 | 0.559798 | 0.999532 | | 2.043743 | -0.22157 | 0.922414 | | 2.343571 | -0.77271 | 0.619615 |
| PFDN4 | 4.006138 | -0.05932 | 0.999532 | | 4.50545 | -0.14219 | 0.92264 | | 3.819397 | -0.06002 | 0.974421 |
| PACSIN1 | 2.101524 | -0.28096 | 0.999532 | | 2.340917 | -0.20024 | 0.92264 | | 1.822691 | 0.099194 | 0.971092 |
| GMPPA | 3.694053 | -0.20185 | 0.999532 | | 4.009971 | -0.14301 | 0.92277 | | 3.264689 | 0.072437 | 0.971732 |
| PALB2 | 2.130742 | -0.1071 | 0.999532 | | 2.695288 | 0.198611 | 0.92277 | | 2.239584 | 0.318655 | 0.864245 |
| NCBP1 | 3.483132 | -0.07796 | 0.999532 | | 4.278523 | 0.146475 | 0.92277 | | 3.648568 | 0.244931 | 0.862823 |
| GTF2IRD | 7.270494 | -0.06532 | 0.999532 | | 7.936009 | -0.11603 | 0.92277 | | 6.619777 | -0.0316 | 0.985693 |
| F2R | 5.597759 | -0.24198 | 0.999532 | | 6.946993 | 0.136327 | 0.92277 | | 5.363353 | 0.411593 | 0.698716 |
| FSTL3 | 2.529777 | 0.298739 | 0.999532 | | 2.379528 | -0.19974 | 0.92277 | | 2.175768 | -0.46868 | 0.755206 |
| AP3B1 | 4.034476 | -0.03472 | 0.999532 | | 4.159274 | -0.1441 | 0.92277 | | 3.660132 | -0.09245 | 0.957624 |
| PPP1R12 | 4.683595 | -0.24251 | 0.999532 | | 5.400575 | -0.13618 | 0.92277 | | 4.295032 | 0.120374 | 0.939549 |
| GAREM1 | 1.877968 | -0.40936 | 0.999532 | | 2.39276 | 0.191361 | 0.922782 | | 1.860554 | 0.613465 | 0.707142 |
| MFAP1 | 2.524589 | -0.12935 | 0.999532 | | 2.747845 | -0.18083 | 0.92286 | | 2.211355 | -0.03306 | 0.991452 |
| PFKFB2 | 3.08753 | 0.050699 | 0.999532 | | 3.476902 | -0.16595 | 0.92286 | | 2.74003 | -0.18488 | 0.911296 |
| TTYH3 | 9.940506 | -0.13196 | 0.999532 | | 12.14882 | 0.091856 | 0.92286 | | 9.815179 | 0.242795 | 0.760463 |
| IL12B | 2.400382 | -0.51452 | 0.999532 | | 3.151021 | -0.17806 | 0.922909 | | 2.33598 | 0.356894 | 0.842696 |
| PARP3 | 3.319927 | 0.179229 | 0.999532 | | 3.402061 | -0.17245 | 0.922909 | | 3.263662 | -0.33293 | 0.823193 |
| DNMT3L | 2.003593 | 0.636977 | 0.999532 | | 1.825943 | 0.209523 | 0.922909 | | 2.068588 | -0.40323 | 0.816775 |
| GAP43 | 1.72746 | 0.126157 | 0.999532 | | 1.795255 | -0.21207 | 0.922909 | | 1.689085 | -0.31778 | 0.877238 |
| PUS10 | 2.219344 | 0.090084 | 0.999532 | | 2.224028 | -0.19427 | 0.922909 | | 2.132539 | -0.26825 | 0.88885 |
| TMOD3 | 6.00345 | -0.37067 | 0.999532 | | 7.347026 | -0.11569 | 0.922909 | | 5.471664 | 0.274311 | 0.797942 |
| MRPL42 | 3.907433 | -0.48723 | 0.999532 | | 5.295813 | 0.136152 | 0.922909 | | 3.888482 | 0.641828 | 0.515762 |
| WDFY4 | 2.77574 | 0.319781 | 0.999532 | | 2.810991 | 0.179316 | 0.922909 | | 2.670776 | -0.12062 | 0.949206 |
| CSNK1G | 9.36951 | -0.16855 | 0.999532 | | 11.94154 | 0.097017 | 0.922909 | | 9.453385 | 0.292966 | 0.717627 |
| SHISA3 | 6.483407 | 0.404244 | 0.999532 | | 6.55734 | 0.144722 | 0.922909 | | 6.377322 | -0.23328 | 0.865065 |
| NOX4 | 2.089639 | -0.16389 | 0.999532 | | 2.500996 | 0.184226 | 0.922909 | | 2.068067 | 0.366136 | 0.830822 |
| GARNL3 | 2.807367 | 0.1442 | 0.999532 | | 2.918198 | -0.17581 | 0.922909 | | 2.490587 | -0.30756 | 0.85621 |
| FHOD3 | 2.502179 | 0.05465 | 0.999532 | | 2.537977 | -0.18346 | 0.922909 | | 2.121428 | -0.22041 | 0.901813 |
| HESX1 | 1.92794 | 0.405105 | 0.999532 | | 1.808661 | -0.20961 | 0.922909 | | 1.829686 | -0.59531 | 0.706407 |
| POLR3C | 3.948823 | -0.06793 | 0.999532 | | 4.501468 | 0.137043 | 0.922909 | | 3.910798 | 0.217055 | 0.876781 |
| ENDOU | 1.742473 | -0.01904 | 0.999532 | | 1.947706 | -0.20985 | 0.922909 | | 1.657412 | -0.16991 | 0.9422 |
| ANKRD3 | 3.168866 | 0.096141 | 0.999532 | | 3.103849 | -0.16315 | 0.922909 | | 2.674873 | -0.24391 | 0.877204 |
| WDR24 | 2.631983 | 0.011352 | 0.999532 | | 2.674144 | -0.18787 | 0.922909 | | 2.306497 | -0.18558 | 0.92697 |
| ZMAT3 | 3.317864 | -0.41416 | 0.999532 | | 3.990812 | -0.16403 | 0.922909 | | 3.0141 | 0.268682 | 0.865363 |
| DNAJB12 | 3.701317 | 0.266975 | 0.999532 | | 4.009384 | 0.148605 | 0.922909 | | 3.725646 | -0.10264 | 0.950372 |
| TRDMT1 | 2.241799 | 0.018489 | 0.999532 | | 2.444081 | -0.18609 | 0.922909 | | 2.166477 | -0.18958 | 0.921 |
| FCRL5 | 2.441066 | 0.423233 | 0.999532 | | 2.329396 | -0.20004 | 0.922909 | | 2.274805 | -0.61097 | 0.660958 |
| EGFLAM | 3.9085 | -0.3236 | 0.999532 | | 4.540379 | -0.15282 | 0.922909 | | 3.556142 | 0.192653 | 0.906811 |
| LYSMD2 | 4.062203 | -0.3448 | 0.999532 | | 4.834493 | -0.15201 | 0.922909 | | 3.471693 | 0.218249 | 0.87764 |
| UBE2W | 3.042012 | -0.0747 | 0.999532 | | 3.496227 | 0.153491 | 0.922909 | | 3.044935 | 0.239892 | 0.87591 |
| WASHC4 | 5.186808 | -0.06235 | 0.999532 | | 6.074864 | 0.133615 | 0.922909 | | 5.179361 | 0.225149 | 0.849885 |
| SPPL3 | 7.479968 | -0.21991 | 0.999532 | | 8.776995 | -0.10513 | 0.922909 | | 6.68641 | 0.133315 | 0.906168 |
| EIF4A1 | 11.71382 | -0.51846 | 0.999532 | | 15.648 | 0.117948 | 0.922909 | | 11.56643 | 0.635396 | 0.37596 |
| HACD1 | 2.340535 | 0.357243 | 0.999532 | | 2.542822 | 0.186009 | 0.922909 | | 2.434267 | -0.15045 | 0.939913 |
| SEBOX | 2.413331 | 0.408782 | 0.999532 | | 2.201012 | -0.19599 | 0.922909 | | 2.162253 | -0.58827 | 0.686214 |
| TSPAN31 | 7.50178 | -0.06888 | 0.999532 | | 9.220835 | 0.113994 | 0.922909 | | 7.495291 | 0.20604 | 0.843564 |
| ACTL9 | 3.063533 | 0.113274 | 0.999532 | | 3.079129 | -0.17047 | 0.922909 | | 2.882706 | -0.27114 | 0.864923 |
| PC | 4.121735 | -0.29941 | 0.999532 | | 4.683966 | -0.1453 | 0.922909 | | 3.70131 | 0.17328 | 0.906811 |
| GNG10 | 6.397725 | 0.107011 | 0.999532 | | 6.911401 | 0.11823 | 0.922909 | | 6.267223 | 0.029505 | 0.987608 |
| TRMT61B | 3.310574 | 0.091897 | 0.999532 | | 3.150458 | -0.15584 | 0.922909 | | 3.023442 | -0.23371 | 0.880949 |
| RIOX2 | 3.65128 | 0.363265 | 0.999532 | | 3.832151 | 0.149038 | 0.922909 | | 3.730624 | -0.19411 | 0.88885 |
| H2AC7 | 2.93064 | 0.169973 | 0.999532 | | 3.092177 | 0.165606 | 0.922909 | | 2.933024 | 0.00591 | 0.99833 |
| APOO | 2.871104 | 0.323002 | 0.999532 | | 2.898855 | 0.171509 | 0.922909 | | 2.967321 | -0.13339 | 0.941197 |
| PARD6G | 3.482081 | -0.1543 | 0.999532 | | 4.259628 | 0.148246 | 0.922909 | | 3.243365 | 0.324063 | 0.802068 |
| NIPAL3 | 3.034618 | -0.1856 | 0.999532 | | 3.418452 | -0.15907 | 0.922909 | | 2.855026 | 0.041541 | 0.987491 |
| PAK2 | 3.562792 | -0.42215 | 0.999532 | | 4.043502 | -0.14293 | 0.922909 | | 3.133327 | 0.290825 | 0.837256 |
| RHEX | 2.193277 | -0.12722 | 0.999532 | | 2.474873 | -0.18612 | 0.922909 | | 2.011748 | -0.04013 | 0.989985 |
| ABHD2 | 3.628891 | 0.115081 | 0.999532 | | 4.156007 | 0.152854 | 0.922909 | | 3.526059 | 0.059482 | 0.975389 |
| SLC25A1 | 2.623033 | 0.351078 | 0.999532 | | 2.531942 | -0.18802 | 0.922909 | | 2.451892 | -0.51819 | 0.717627 |
| MAP1A | 2.065433 | -0.09354 | 0.999532 | | 2.540824 | 0.1841 | 0.922909 | | 2.157217 | 0.294449 | 0.869579 |
| APOOL | 4.157229 | 0.297628 | 0.999532 | | 4.300256 | 0.141478 | 0.922909 | | 4.102017 | -0.13752 | 0.920527 |
| EPB41L2 | 6.491809 | -0.36367 | 0.999532 | | 8.262603 | 0.116031 | 0.922909 | | 6.534825 | 0.523029 | 0.505783 |
| TXNDC5 | 6.836701 | -0.25454 | 0.999532 | | 8.063543 | -0.10848 | 0.922909 | | 6.309031 | 0.165319 | 0.880375 |
| RPL21 | 57.20348 | -0.01426 | 0.999532 | | 65.85334 | 0.050213 | 0.922909 | | 55.27242 | 0.087391 | 0.843043 |
| SMAD9 | 2.675808 | 0.249039 | 0.999532 | | 2.938432 | 0.169409 | 0.922909 | | 2.74821 | -0.06202 | 0.977473 |
| UNC80 | 2.484478 | 0.023944 | 0.999532 | | 2.662946 | -0.18276 | 0.922909 | | 2.390325 | -0.19659 | 0.914367 |
| SLC41A3 | 3.37128 | -0.02836 | 0.999532 | | 4.101841 | 0.144399 | 0.922909 | | 3.513318 | 0.191884 | 0.893103 |
| SCGB3A | 2.244538 | -0.05226 | 0.999532 | | 2.375482 | -0.20143 | 0.922909 | | 2.091151 | -0.13524 | 0.953814 |
| ERICH1 | 2.018286 | 0.421387 | 0.999532 | | 2.042457 | 0.197926 | 0.922909 | | 2.111385 | -0.20212 | 0.915533 |
| PCOLCE | 3.3735 | 0.309103 | 0.999532 | | 3.562344 | 0.166441 | 0.922909 | | 3.33746 | -0.11823 | 0.942931 |
| STK33 | 3.016 | 0.761056 | 0.999532 | | 2.611958 | 0.189434 | 0.922909 | | 2.939972 | -0.55106 | 0.673659 |
| ANKMY2 | 3.381086 | 0.27367 | 0.999532 | | 3.064126 | -0.16507 | 0.922909 | | 2.937188 | -0.4198 | 0.740619 |
| RGS14 | 3.437255 | -0.26326 | 0.999532 | | 4.164121 | -0.15517 | 0.922909 | | 3.236857 | 0.128923 | 0.941197 |
| HBS1L | 3.45296 | -0.51876 | 0.999532 | | 4.253952 | -0.14121 | 0.922909 | | 2.896128 | 0.392353 | 0.755206 |
| MCOLN1 | 3.19922 | -0.403 | 0.999532 | | 3.760181 | -0.15405 | 0.922909 | | 2.789059 | 0.260778 | 0.871072 |
| PPM1K | 4.744454 | 0.054423 | 0.999532 | | 5.593429 | 0.128666 | 0.922909 | | 4.563357 | 0.085856 | 0.955834 |
| FLT1 | 12.86078 | -0.33975 | 0.999532 | | 16.91101 | 0.092474 | 0.922909 | | 12.87391 | 0.452475 | 0.456688 |
| GJA3 | 2.393547 | 0.120778 | 0.999532 | | 2.699563 | 0.191667 | 0.922909 | | 2.496521 | 0.090516 | 0.967577 |
| TMCC1 | 3.412971 | 0.251412 | 0.999532 | | 3.47003 | -0.16544 | 0.922909 | | 2.953967 | -0.39559 | 0.757822 |
| ZFPM2 | 2.625838 | -0.34663 | 0.999532 | | 3.320116 | 0.168303 | 0.922909 | | 2.583187 | 0.537975 | 0.700524 |
| ACSM3 | 2.466854 | 0.315599 | 0.999532 | | 2.61621 | 0.176147 | 0.922909 | | 2.464272 | -0.12119 | 0.948865 |
| PGGT1B | 4.760244 | -0.3225 | 0.999532 | | 5.693056 | -0.12643 | 0.922909 | | 4.172765 | 0.212861 | 0.87591 |
| TMEM26 | 3.900971 | 0.019758 | 0.999532 | | 4.333427 | 0.149945 | 0.922909 | | 3.998853 | 0.112425 | 0.955834 |
| ART1 | 3.107506 | 0.186093 | 0.999532 | | 3.504057 | 0.167029 | 0.922909 | | 3.314244 | 0.004635 | 0.998586 |
| KRT17 | 2.287857 | 0.708533 | 0.999532 | | 1.843163 | -0.20888 | 0.922909 | | 2.151233 | -0.89522 | 0.515762 |
| RFX5 | 2.32946 | -0.08072 | 0.999532 | | 3.032407 | 0.176007 | 0.922909 | | 2.451729 | 0.282287 | 0.869378 |
| PSMC3 | 8.387628 | 0.035934 | 0.999532 | | 9.234296 | 0.111354 | 0.922909 | | 8.074367 | 0.080919 | 0.947851 |
| FAM47B | 3.796916 | -0.14225 | 0.999532 | | 4.169143 | -0.14891 | 0.922909 | | 3.474092 | 0.014976 | 0.996681 |
| MITD1 | 3.724825 | 0.474719 | 0.999532 | | 3.684668 | 0.153456 | 0.922909 | | 3.66809 | -0.30177 | 0.814884 |
| DEPTOR | 3.047836 | -0.26141 | 0.999532 | | 3.700705 | -0.15107 | 0.922909 | | 2.739175 | 0.129062 | 0.941197 |
| IFNGR1 | 4.877496 | -0.0679 | 0.999532 | | 5.637327 | 0.128434 | 0.922909 | | 4.807717 | 0.215254 | 0.857816 |
| ODR4 | 3.548701 | 0.116019 | 0.999532 | | 3.928508 | 0.15136 | 0.922909 | | 3.44696 | 0.05465 | 0.977746 |
| PDCD6IP | 6.106184 | -0.02839 | 0.999532 | | 6.967683 | 0.131006 | 0.922909 | | 6.128874 | 0.182174 | 0.878232 |
| DDX42 | 5.663496 | 0.006167 | 0.999532 | | 5.973379 | -0.12927 | 0.922909 | | 5.301378 | -0.11937 | 0.92924 |
| INAFM1 | 2.718872 | -0.55046 | 0.999532 | | 3.590346 | -0.15565 | 0.922909 | | 2.483923 | 0.415576 | 0.774747 |
| PGPEP1 | 4.260596 | 0.569167 | 0.999532 | | 3.880719 | 0.151216 | 0.922909 | | 4.322206 | -0.38601 | 0.755206 |
| CALU | 6.559891 | -0.39476 | 0.999532 | | 7.700215 | -0.11334 | 0.922909 | | 5.88895 | 0.28633 | 0.791576 |
| ZBTB39 | 2.267502 | -0.04143 | 0.999532 | | 2.679438 | 0.181423 | 0.922909 | | 2.2576 | 0.240852 | 0.893262 |
| DENND1 | 5.705537 | 0.022252 | 0.999532 | | 5.998327 | -0.13476 | 0.922909 | | 5.370942 | -0.13101 | 0.924131 |
| XPO4 | 2.487517 | -0.02046 | 0.999532 | | 2.504879 | -0.18524 | 0.922909 | | 2.284123 | -0.1501 | 0.94386 |
| PHF12 | 3.041363 | 0.428814 | 0.999532 | | 3.195736 | 0.16282 | 0.922909 | | 3.059938 | -0.24736 | 0.864245 |
| CDK16 | 3.991515 | -0.04048 | 0.999532 | | 4.652142 | 0.139948 | 0.922909 | | 3.927205 | 0.200942 | 0.880375 |
| LMTK3 | 2.369051 | 0.390021 | 0.999532 | | 2.084131 | -0.21339 | 0.922909 | | 2.167821 | -0.59236 | 0.717138 |
| RBM48 | 3.026395 | -0.16889 | 0.999532 | | 3.393056 | -0.16072 | 0.922909 | | 2.631189 | 0.020463 | 0.995304 |
| SHH | 2.044311 | -0.37479 | 0.999532 | | 2.568453 | -0.20597 | 0.922909 | | 1.982378 | 0.176647 | 0.939913 |
| ZNF521 | 2.313189 | 0.104585 | 0.999532 | | 2.300688 | -0.18488 | 0.922909 | | 2.061778 | -0.27278 | 0.877943 |
| SNX33 | 6.121778 | 0.168863 | 0.999532 | | 6.926817 | 0.125633 | 0.922909 | | 6.056358 | -0.01357 | 0.996471 |
| FAHD1 | 2.961109 | 0.183723 | 0.999532 | | 2.862903 | -0.17864 | 0.922909 | | 2.711932 | -0.3619 | 0.816618 |
| ADM2 | 2.614867 | 0.19596 | 0.999532 | | 2.900236 | 0.183936 | 0.922909 | | 2.710245 | 0.000362 | 0.999949 |
| KAT7 | 7.148641 | 0.079362 | 0.999532 | | 7.702341 | -0.12791 | 0.922909 | | 6.495994 | -0.18657 | 0.864245 |
| FOXB2 | 2.96297 | 0.496155 | 0.999532 | | 2.529637 | -0.183 | 0.922909 | | 2.800628 | -0.65788 | 0.623462 |
| FRMD6 | 3.492415 | -0.00455 | 0.999532 | | 3.822533 | -0.15368 | 0.922909 | | 3.324621 | -0.1311 | 0.936689 |
| BCO1 | 2.158567 | 0.159846 | 0.999532 | | 2.269504 | -0.20566 | 0.922909 | | 2.120948 | -0.34219 | 0.862823 |
| ARFIP1 | 2.938984 | -0.3674 | 0.999532 | | 4.0463 | 0.154718 | 0.922909 | | 3.210576 | 0.545744 | 0.673405 |
| AAAS | 4.122458 | -0.21967 | 0.999532 | | 4.716412 | -0.13633 | 0.922909 | | 3.796299 | 0.100395 | 0.950529 |
| CA11 | 3.559714 | -0.09578 | 0.999532 | | 3.712557 | -0.16034 | 0.922909 | | 3.244289 | -0.03788 | 0.988379 |
| PJA1 | 2.576026 | -0.08319 | 0.999532 | | 3.146995 | 0.157233 | 0.923356 | | 2.595609 | 0.26076 | 0.872264 |
| TAF11 | 3.90005 | -0.10754 | 0.999532 | | 4.210374 | -0.15148 | 0.923356 | | 3.514162 | -0.02004 | 0.994617 |
| ABCA4 | 3.626584 | -0.02943 | 0.999532 | | 3.912463 | -0.15832 | 0.923356 | | 3.322772 | -0.11106 | 0.953948 |
| SPN | 4.131076 | 0.078464 | 0.999532 | | 4.163682 | -0.15401 | 0.923356 | | 3.887032 | -0.22723 | 0.877703 |
| C8orf33 | 2.910429 | -0.12478 | 0.999532 | | 3.086727 | -0.17685 | 0.923356 | | 2.513741 | -0.02146 | 0.995112 |
| TMEM26 | 2.274721 | -0.13098 | 0.999532 | | 2.593958 | -0.18344 | 0.923356 | | 2.016282 | -0.03165 | 0.992286 |
| SPAG8 | 3.087441 | 0.058536 | 0.999532 | | 3.789442 | 0.174141 | 0.923356 | | 3.414926 | 0.12594 | 0.951149 |
| OR51L1 | 1.824342 | -0.1863 | 0.999532 | | 2.106723 | -0.19683 | 0.923356 | | 1.776148 | 0.005924 | 0.998586 |
| SSTR3 | 2.872569 | 0.03521 | 0.999532 | | 3.145949 | -0.17614 | 0.923356 | | 2.526156 | -0.18486 | 0.915533 |
| GLS2 | 1.93196 | 0.438045 | 0.999532 | | 2.008582 | 0.199959 | 0.923356 | | 2.067084 | -0.22165 | 0.908891 |
| ADAL | 2.617995 | 0.587608 | 0.999532 | | 2.381277 | -0.1954 | 0.923356 | | 2.394671 | -0.78279 | 0.541556 |
| ADAMTS | 2.896429 | -0.08862 | 0.999532 | | 3.137795 | -0.16864 | 0.923356 | | 2.602612 | -0.06184 | 0.978156 |
| APOBEC | 1.918306 | 0.389745 | 0.999532 | | 1.650561 | -0.22162 | 0.923356 | | 1.760613 | -0.59789 | 0.725338 |
| XIAP | 3.820181 | -0.03593 | 0.999532 | | 4.273638 | -0.1407 | 0.923356 | | 3.691546 | -0.07979 | 0.965649 |
| ZNF281 | 2.937386 | -0.04356 | 0.999532 | | 2.939663 | -0.16411 | 0.923356 | | 2.544014 | -0.11281 | 0.955834 |
| INTS4 | 1.957577 | -0.18699 | 0.999532 | | 2.363327 | 0.183264 | 0.923356 | | 1.937116 | 0.385067 | 0.821752 |
| ZKSCAN | 2.858371 | -0.2593 | 0.999532 | | 3.396993 | -0.16565 | 0.923356 | | 2.714062 | 0.114842 | 0.956011 |
| H3C3 | 3.065359 | -0.0413 | 0.999532 | | 3.234468 | -0.16247 | 0.923356 | | 2.824385 | -0.10574 | 0.958191 |
| IFIT2 | 2.488355 | -0.19814 | 0.999532 | | 2.807867 | -0.17346 | 0.923356 | | 2.234037 | 0.044158 | 0.988379 |
| TEN1 | 3.336762 | 0.147494 | 0.999532 | | 3.444035 | -0.16738 | 0.923356 | | 2.982969 | -0.30082 | 0.836453 |
| SAXO2 | 2.278111 | 0.070984 | 0.999532 | | 2.295391 | -0.19195 | 0.923356 | | 2.01531 | -0.24761 | 0.896886 |
| CHRM2 | 1.892504 | 0.149462 | 0.999532 | | 1.874057 | -0.21332 | 0.923356 | | 1.68253 | -0.3451 | 0.863979 |
| HABP4 | 8.399556 | 0.388596 | 0.999532 | | 8.667446 | 0.101259 | 0.923356 | | 8.247718 | -0.27049 | 0.742864 |
| ZNF641 | 2.714567 | 0.197877 | 0.999532 | | 3.032942 | 0.167119 | 0.923356 | | 2.75616 | -0.00841 | 0.997739 |
| CASTOR | 2.43233 | -0.25522 | 0.999532 | | 3.371607 | 0.163578 | 0.923356 | | 2.649077 | 0.442147 | 0.754186 |
| MOCS2 | 2.334821 | 0.07563 | 0.999532 | | 2.523084 | -0.18445 | 0.923356 | | 2.14222 | -0.23897 | 0.895865 |
| TCTA | 2.651484 | 0.276253 | 0.999532 | | 2.835448 | 0.167608 | 0.923356 | | 2.576908 | -0.08953 | 0.964357 |
| GPR153 | 2.305308 | 0.086033 | 0.999532 | | 2.343891 | -0.18573 | 0.923356 | | 2.099572 | -0.25334 | 0.892328 |
| WDR54 | 3.209811 | 0.094915 | 0.999532 | | 3.230678 | -0.1624 | 0.923356 | | 2.91437 | -0.23688 | 0.880997 |
| DYRK2 | 2.948504 | -0.05475 | 0.999532 | | 3.40095 | 0.154714 | 0.923356 | | 2.964893 | 0.223846 | 0.88885 |
| TCEAL4 | 6.080718 | 0.019535 | 0.999532 | | 7.341878 | 0.108714 | 0.923356 | | 6.099922 | 0.108076 | 0.926957 |
| H1-7 | 3.498784 | 0.346238 | 0.999532 | | 3.766853 | 0.160733 | 0.923356 | | 3.666786 | -0.16919 | 0.920259 |
| DUS1L | 4.85975 | -0.07983 | 0.999532 | | 6.237502 | 0.124853 | 0.923356 | | 5.033542 | 0.257928 | 0.842754 |
| ZNF577 | 3.877592 | 0.304317 | 0.999532 | | 4.083326 | 0.139884 | 0.923356 | | 3.94547 | -0.14541 | 0.91707 |
| RHPN2 | 6.279014 | -0.12675 | 0.999532 | | 7.233218 | -0.15111 | 0.923356 | | 6.014421 | -0.02429 | 0.992286 |
| GLMP | 4.177332 | -0.37975 | 0.999532 | | 5.474908 | 0.130968 | 0.923356 | | 4.124715 | 0.52885 | 0.630705 |
| USP19 | 3.909788 | 0.082035 | 0.999532 | | 3.707616 | -0.15153 | 0.923356 | | 3.378334 | -0.22025 | 0.884285 |
| ELF5 | 2.754062 | 0.220409 | 0.999532 | | 2.634143 | -0.18023 | 0.923356 | | 2.520598 | -0.38753 | 0.806208 |
| ISCA2 | 5.613902 | 0.0316 | 0.999532 | | 6.871931 | 0.140214 | 0.923356 | | 6.086399 | 0.108067 | 0.94366 |
| RACGAP | 2.599673 | 0.151004 | 0.999532 | | 3.219046 | 0.180495 | 0.923356 | | 2.670928 | 0.057468 | 0.980581 |
| TRAF3IP | 3.272855 | 0.141414 | 0.999532 | | 3.417259 | -0.15996 | 0.923356 | | 2.927176 | -0.28173 | 0.849682 |
| UPF3A | 7.003938 | -0.28703 | 0.999532 | | 8.604359 | -0.10947 | 0.923356 | | 6.683521 | 0.209236 | 0.854282 |
| EMC9 | 2.475619 | 0.401284 | 0.999532 | | 2.59784 | 0.179373 | 0.923356 | | 2.514658 | -0.20341 | 0.904941 |
| CYP4A22 | 7.30546 | -0.18003 | 0.999532 | | 9.185892 | 0.158038 | 0.923356 | | 7.211935 | 0.377383 | 0.75311 |
| DNAAF5 | 3.356821 | -0.19279 | 0.999532 | | 4.32853 | 0.151231 | 0.923356 | | 3.604616 | 0.352067 | 0.791576 |
| RELA | 4.285009 | -0.17516 | 0.999532 | | 5.293728 | -0.1422 | 0.923356 | | 4.050316 | 0.086269 | 0.962725 |
| APBB2 | 10.50497 | 0.107667 | 0.999532 | | 11.67069 | 0.109491 | 0.923356 | | 10.29968 | 0.022935 | 0.990309 |
| BMP8B | 3.276023 | 0.631441 | 0.999532 | | 3.107278 | 0.168541 | 0.923356 | | 3.276455 | -0.44721 | 0.704363 |
| LSM8 | 3.196116 | 0.44169 | 0.999532 | | 3.284257 | 0.170269 | 0.923356 | | 3.135121 | -0.25442 | 0.865065 |
| NPB | 2.985928 | -0.07706 | 0.999532 | | 3.337197 | -0.17697 | 0.923356 | | 2.866178 | -0.07935 | 0.974421 |
| MCM3AP | 3.696511 | 0.212231 | 0.999532 | | 4.117891 | 0.146004 | 0.923356 | | 3.548745 | -0.04086 | 0.986869 |
| C1QL4 | 2.823919 | 0.446093 | 0.999532 | | 2.591507 | -0.1864 | 0.923356 | | 2.511634 | -0.6134 | 0.647218 |
| SLC5A6 | 2.764149 | 0.402916 | 0.999532 | | 2.763597 | 0.17267 | 0.923356 | | 2.865951 | -0.22194 | 0.900641 |
| C11orf68 | 3.106343 | -0.03656 | 0.999532 | | 3.202739 | -0.15651 | 0.923678 | | 2.757231 | -0.10609 | 0.955834 |
| SLC6A2 | 2.111197 | -0.43353 | 0.999532 | | 2.40218 | -0.18335 | 0.923678 | | 1.769308 | 0.264449 | 0.893828 |
| TSNARE | 2.874907 | 0.095704 | 0.999532 | | 3.031628 | -0.17568 | 0.923678 | | 2.554507 | -0.24908 | 0.880375 |
| SIN3A | 3.591898 | -0.24301 | 0.999532 | | 4.524149 | 0.136512 | 0.923747 | | 3.588654 | 0.397785 | 0.736481 |
| FAM161A | 2.242675 | 0.647851 | 0.999532 | | 1.794666 | -0.20694 | 0.923747 | | 1.99651 | -0.83599 | 0.536289 |
| FBXO31 | 2.263704 | 0.867166 | 0.999532 | | 2.037528 | 0.196545 | 0.923748 | | 2.335397 | -0.65437 | 0.622543 |
| EFL1 | 2.145802 | 0.557773 | 0.999532 | | 1.803189 | -0.21104 | 0.92379 | | 1.908087 | -0.75077 | 0.622543 |
| TUBA3E | 3.669463 | -0.32059 | 0.999532 | | 4.514092 | -0.14282 | 0.92379 | | 3.335774 | 0.199387 | 0.892869 |
| SEPTIN6 | 2.599855 | -0.01284 | 0.999532 | | 3.12979 | 0.160674 | 0.92379 | | 2.665515 | 0.196923 | 0.907586 |
| CCHCR1 | 2.832068 | 0.287185 | 0.999532 | | 2.825323 | -0.17241 | 0.92379 | | 2.783804 | -0.44261 | 0.742129 |
| SH3BP2 | 8.088964 | -0.30592 | 0.999532 | | 9.12816 | -0.11505 | 0.92379 | | 7.111036 | 0.217668 | 0.831123 |
| SEPTIN8 | 6.367709 | 0.063798 | 0.999532 | | 7.489755 | 0.112544 | 0.92379 | | 6.19228 | 0.069665 | 0.960773 |
| TFF3 | 2.50678 | -0.32241 | 0.999532 | | 3.035989 | 0.169478 | 0.92379 | | 2.425221 | 0.481154 | 0.749206 |
| SREK1 | 4.944466 | -0.16706 | 0.999532 | | 6.399961 | 0.122801 | 0.92379 | | 5.10861 | 0.308981 | 0.769793 |
| SIN3B | 3.213164 | -0.03573 | 0.999532 | | 3.694839 | 0.149603 | 0.92379 | | 3.23221 | 0.203461 | 0.892869 |
| CCNL1 | 7.135524 | 0.279985 | 0.999532 | | 7.484024 | 0.120803 | 0.92379 | | 6.821743 | -0.13268 | 0.905746 |
| IDNK | 2.090283 | -0.23819 | 0.999532 | | 2.298383 | -0.18497 | 0.92379 | | 1.827306 | 0.066024 | 0.979996 |
| OR8U1 | 2.120746 | 0.510474 | 0.999532 | | 1.865757 | -0.21263 | 0.92379 | | 1.917558 | -0.70337 | 0.647581 |
| C1orf141 | 2.494276 | 0.320217 | 0.999532 | | 2.411727 | -0.18208 | 0.92379 | | 2.353562 | -0.48371 | 0.735469 |
| TCEAL3 | 6.989208 | 0.009924 | 0.999532 | | 8.166133 | 0.120814 | 0.92379 | | 6.929716 | 0.135147 | 0.907968 |
| CEP72 | 2.444006 | 0.094505 | 0.999532 | | 2.877713 | 0.171811 | 0.92379 | | 2.534356 | 0.096114 | 0.964357 |
| DHDH | 2.408999 | 0.275798 | 0.999532 | | 2.271316 | -0.18837 | 0.92379 | | 2.160825 | -0.44562 | 0.775223 |
| UBXN1 | 9.096706 | 0.050748 | 0.999532 | | 9.950571 | -0.09451 | 0.92379 | | 8.377385 | -0.12179 | 0.908829 |
| TMEM213 | 4.471001 | -0.06329 | 0.999532 | | 5.153125 | -0.13208 | 0.92379 | | 4.139059 | -0.0355 | 0.987608 |
| KY | 2.067959 | 0.560809 | 0.999532 | | 1.957359 | -0.21008 | 0.92379 | | 2.054918 | -0.74969 | 0.6201 |
| TBC1D17 | 7.607033 | -0.25 | 0.999532 | | 8.882847 | -0.10172 | 0.92379 | | 6.915256 | 0.167571 | 0.870791 |
| APOBEC | 2.232237 | 0.879733 | 0.999532 | | 1.92213 | 0.212342 | 0.92379 | | 2.288127 | -0.65271 | 0.653409 |
| CTF1 | 2.650344 | -0.03235 | 0.999532 | | 2.713525 | -0.17502 | 0.92379 | | 2.361225 | -0.12974 | 0.948865 |
| USP12 | 3.124282 | 0.182339 | 0.999532 | | 2.869059 | -0.16626 | 0.92379 | | 2.79463 | -0.33077 | 0.827267 |
| PPIC | 2.555551 | 0.07965 | 0.999532 | | 2.78857 | 0.163835 | 0.92379 | | 2.493488 | 0.095734 | 0.96375 |
| ZC2HC1A | 3.277763 | -0.0268 | 0.999532 | | 3.86956 | 0.146678 | 0.92379 | | 3.305917 | 0.187995 | 0.901916 |
| STAM | 2.321264 | -0.27379 | 0.999532 | | 3.017046 | 0.160803 | 0.92379 | | 2.34108 | 0.452944 | 0.750417 |
| CD93 | 6.336568 | -0.63898 | 0.999532 | | 9.256529 | 0.141636 | 0.92379 | | 6.596649 | 0.792633 | 0.302277 |
| ZNF148 | 4.360472 | 0.052894 | 0.999532 | | 4.715853 | -0.15132 | 0.92379 | | 4.215909 | -0.19004 | 0.901033 |
| UTP6 | 4.651306 | -0.19727 | 0.999532 | | 5.285006 | -0.12818 | 0.923894 | | 4.124936 | 0.085257 | 0.958422 |
| TGM1 | 1.830657 | 0.164782 | 0.999532 | | 1.928701 | -0.20503 | 0.923905 | | 1.765618 | -0.34996 | 0.858705 |
| CNIH3 | 2.498737 | 0.169541 | 0.999532 | | 2.549154 | -0.18613 | 0.923905 | | 2.417171 | -0.34093 | 0.837927 |
| SGTA | 10.07958 | -0.12095 | 0.999532 | | 12.10192 | 0.091538 | 0.923905 | | 9.59065 | 0.230011 | 0.774747 |
| FUS | 17.82677 | -0.16745 | 0.999532 | | 21.79456 | 0.090848 | 0.923915 | | 17.45894 | 0.278964 | 0.71532 |
| MLKL | 2.403662 | -0.1885 | 0.999532 | | 2.749532 | -0.1762 | 0.923915 | | 2.222309 | 0.027842 | 0.993618 |
| ETFRF1 | 3.702721 | 0.716878 | 0.999532 | | 2.965985 | -0.16427 | 0.923915 | | 3.382035 | -0.86293 | 0.350296 |
| OGN | 3.140896 | 0.179736 | 0.999532 | | 3.110341 | -0.17308 | 0.923915 | | 2.862685 | -0.34283 | 0.835289 |
| CCNA1 | 2.031757 | 0.157064 | 0.999532 | | 2.036393 | -0.19209 | 0.923915 | | 1.838146 | -0.3318 | 0.859898 |
| TMEM234 | 6.709313 | -0.21321 | 0.999532 | | 7.98518 | 0.10681 | 0.923915 | | 6.435596 | 0.318081 | 0.749206 |
| PTP4A2 | 8.786798 | -0.37928 | 0.999532 | | 10.37233 | -0.09591 | 0.923915 | | 7.864657 | 0.291379 | 0.733256 |
| HARS2 | 3.020125 | 0.71195 | 0.999532 | | 2.824074 | 0.170516 | 0.924242 | | 3.161382 | -0.52217 | 0.674647 |
| SETDB1 | 3.222114 | 0.784397 | 0.999532 | | 2.985251 | 0.167557 | 0.924242 | | 3.28986 | -0.59056 | 0.626053 |
| GDF6 | 2.267073 | 0.03237 | 0.999532 | | 2.385921 | -0.18652 | 0.924242 | | 1.983473 | -0.19804 | 0.9195 |
| ZCRB1 | 3.04873 | 0.552052 | 0.999532 | | 2.507043 | -0.17374 | 0.924288 | | 2.814601 | -0.71167 | 0.552672 |
| SMIM10 | 2.280458 | 0.462904 | 0.999532 | | 1.980931 | -0.19332 | 0.924288 | | 2.097524 | -0.63998 | 0.660958 |
| PCMT1 | 4.893243 | -0.07195 | 0.999532 | | 5.862701 | 0.125693 | 0.924288 | | 4.849797 | 0.220226 | 0.859898 |
| SLIT1 | 2.203438 | -0.04795 | 0.999532 | | 2.250863 | -0.19562 | 0.924426 | | 1.978519 | -0.13381 | 0.955834 |
| TARBP2 | 3.506764 | 0.061235 | 0.999532 | | 3.858964 | -0.14499 | 0.924426 | | 3.283589 | -0.18216 | 0.90424 |
| WNT10A | 3.950548 | -0.04412 | 0.999532 | | 4.035899 | -0.14239 | 0.924441 | | 3.604642 | -0.08424 | 0.965619 |
| GOLGA6 | 5.712939 | -0.23457 | 0.999532 | | 6.850485 | -0.11372 | 0.924653 | | 5.333251 | 0.143083 | 0.906902 |
| HP1BP3 | 9.114003 | -0.00501 | 0.999532 | | 10.47293 | 0.112886 | 0.924665 | | 8.810703 | 0.146115 | 0.886268 |
| ZBED6CL | 2.19463 | -0.34779 | 0.999532 | | 2.755586 | -0.1834 | 0.92473 | | 2.108498 | 0.187355 | 0.929335 |
| TNIK | 3.091941 | 0.297807 | 0.999532 | | 3.228955 | 0.159768 | 0.92473 | | 3.170722 | -0.12618 | 0.94861 |
| YTHDF3 | 4.629038 | 0.553908 | 0.999532 | | 4.32732 | 0.139136 | 0.92473 | | 4.667656 | -0.39335 | 0.712891 |
| NDST2 | 4.053069 | 0.07806 | 0.999532 | | 4.443379 | 0.132558 | 0.92473 | | 3.926709 | 0.070891 | 0.967002 |
| KPNA3 | 4.097914 | -0.118 | 0.999532 | | 4.482632 | -0.1398 | 0.92473 | | 3.693233 | -0.00926 | 0.996891 |
| ZNF141 | 3.633328 | 0.538105 | 0.999532 | | 3.697124 | 0.149 | 0.92473 | | 3.655647 | -0.36712 | 0.758206 |
| TMIE | 1.971364 | 0.061908 | 0.999532 | | 2.388828 | 0.184965 | 0.92473 | | 2.044734 | 0.143427 | 0.946572 |
| CXCL12 | 4.789179 | 0.146485 | 0.999532 | | 4.747278 | -0.15098 | 0.92473 | | 4.17786 | -0.26893 | 0.837927 |
| PAWR | 7.825965 | 0.131161 | 0.999532 | | 8.824287 | 0.116361 | 0.92473 | | 7.625858 | 0.002954 | 0.998655 |
| KCNMB3 | 2.449199 | 0.264463 | 0.999532 | | 2.606929 | -0.18405 | 0.92473 | | 2.374367 | -0.42489 | 0.786076 |
| MKKS | 4.239106 | -0.43543 | 0.999532 | | 5.698335 | 0.130258 | 0.92473 | | 4.189645 | 0.586113 | 0.546858 |
| GOLGA8 | 11.19982 | -0.05037 | 0.999532 | | 12.22833 | -0.09054 | 0.92473 | | 10.21088 | -0.02177 | 0.989437 |
| ASPM | 1.907313 | 0.084561 | 0.999532 | | 1.938385 | -0.20329 | 0.92473 | | 1.843645 | -0.27227 | 0.895721 |
| PLAGL1 | 3.050976 | 0.100945 | 0.999532 | | 3.449016 | 0.151397 | 0.924918 | | 3.116013 | 0.067377 | 0.97436 |
| MRPL10 | 3.643101 | -0.08375 | 0.999532 | | 4.078545 | -0.13741 | 0.92502 | | 3.364148 | -0.03437 | 0.988853 |
| DKK1 | 5.314301 | 0.443191 | 0.999532 | | 5.044002 | -0.14943 | 0.925456 | | 5.18366 | -0.56763 | 0.623462 |
| WIPF2 | 5.566801 | -0.09728 | 0.999532 | | 6.281383 | -0.11997 | 0.925589 | | 5.211985 | 0.002621 | 0.999385 |
| C1QTNF | 3.673683 | 0.016338 | 0.999532 | | 4.435289 | 0.138924 | 0.925589 | | 3.63492 | 0.146588 | 0.921808 |
| HK2 | 2.391955 | 0.094115 | 0.999532 | | 2.444935 | -0.18171 | 0.925589 | | 2.100295 | -0.25861 | 0.887278 |
| LRRC61 | 2.433649 | 0.734725 | 0.999532 | | 2.146853 | 0.188458 | 0.925589 | | 2.510077 | -0.53323 | 0.705932 |
| UNC5CL | 3.858313 | 0.663317 | 0.999532 | | 3.052612 | -0.15661 | 0.925589 | | 3.480212 | -0.80588 | 0.413122 |
| SCAPER | 4.173657 | 0.282426 | 0.999532 | | 3.943485 | -0.15243 | 0.925589 | | 3.687098 | -0.40066 | 0.720106 |
| EXOC6B | 5.322842 | 0.030046 | 0.999532 | | 5.514291 | -0.13786 | 0.925736 | | 4.983161 | -0.15898 | 0.909157 |
| DGKE | 3.197934 | 0.183359 | 0.999532 | | 3.498698 | 0.151001 | 0.925736 | | 3.064526 | -0.00647 | 0.996916 |
| LSR | 3.456054 | 0.099786 | 0.999532 | | 3.779224 | 0.144307 | 0.925852 | | 3.331178 | 0.06522 | 0.972758 |
| PLSCR3 | 6.398405 | -0.14798 | 0.999532 | | 7.709947 | 0.112212 | 0.925852 | | 6.331953 | 0.278476 | 0.776852 |
| L3MBTL4 | 4.314891 | 0.010941 | 0.999532 | | 5.134383 | 0.129015 | 0.925852 | | 4.331974 | 0.141075 | 0.920486 |
| ABCF3 | 3.101274 | -0.07779 | 0.999532 | | 3.381596 | -0.15208 | 0.925852 | | 2.694727 | -0.05551 | 0.97843 |
| A2ML1 | 1.835385 | 0.106019 | 0.999532 | | 2.010097 | 0.199033 | 0.925855 | | 1.902431 | 0.105717 | 0.969501 |
| FCAR | 2.374122 | 0.003158 | 0.999532 | | 2.752253 | 0.164844 | 0.925928 | | 2.407634 | 0.175863 | 0.924165 |
| TMEM128 | 5.320682 | -0.07614 | 0.999532 | | 6.330604 | 0.115718 | 0.925928 | | 5.342448 | 0.211438 | 0.861119 |
| MDN1 | 2.29161 | 0.274559 | 0.999532 | | 2.582358 | 0.173032 | 0.925928 | | 2.373447 | -0.08158 | 0.972221 |
| IPO4 | 2.557744 | 0.061387 | 0.999532 | | 2.745587 | 0.165094 | 0.925928 | | 2.593955 | 0.120072 | 0.952656 |
| LMX1B | 5.725657 | 0.06703 | 0.999532 | | 7.082342 | 0.140933 | 0.925976 | | 6.019242 | 0.095852 | 0.950669 |
| ZNF827 | 3.491166 | 0.261335 | 0.999532 | | 3.673858 | 0.145382 | 0.925976 | | 3.356724 | -0.09609 | 0.955834 |
| OR6S1 | 2.398347 | 0.262281 | 0.999532 | | 2.474112 | -0.18241 | 0.925976 | | 2.286445 | -0.41609 | 0.796702 |
| RNF141 | 3.533123 | -0.00126 | 0.999532 | | 3.74747 | -0.14215 | 0.925976 | | 3.255286 | -0.12205 | 0.941074 |
| HACD4 | 2.18114 | 0.18223 | 0.999532 | | 2.232258 | -0.18437 | 0.925996 | | 1.950726 | -0.3472 | 0.842696 |
| GFRA3 | 2.014383 | -0.10398 | 0.999532 | | 2.202473 | -0.19543 | 0.926164 | | 1.921735 | -0.07691 | 0.978229 |
| NOL3 | 3.41883 | 0.380901 | 0.999532 | | 3.307521 | -0.14967 | 0.926164 | | 3.171784 | -0.51277 | 0.652095 |
| PI4K2A | 1.997148 | -0.36791 | 0.999532 | | 2.452116 | 0.171288 | 0.926164 | | 2.040935 | 0.563911 | 0.717138 |
| ARPC1B | 6.530865 | -0.34149 | 0.999532 | | 8.158306 | 0.105603 | 0.926164 | | 6.29483 | 0.436708 | 0.65492 |
| VSIG10L | 1.886709 | 0.114087 | 0.999532 | | 2.217907 | 0.191367 | 0.926164 | | 1.962134 | 0.095002 | 0.971732 |
| ILRUN | 4.466766 | -0.28559 | 0.999532 | | 5.755272 | 0.126555 | 0.926164 | | 4.300282 | 0.437497 | 0.680885 |
| TANK | 6.010545 | -0.19122 | 0.999532 | | 7.800696 | 0.105115 | 0.926164 | | 6.097887 | 0.315233 | 0.724363 |
| FAM3C | 7.266527 | -0.15213 | 0.999532 | | 8.724442 | 0.100038 | 0.926164 | | 7.191326 | 0.266139 | 0.776133 |
| ZNF24 | 2.263634 | -0.28461 | 0.999532 | | 2.964322 | 0.168432 | 0.926164 | | 2.425373 | 0.473016 | 0.750371 |
| ARSB | 2.469231 | 0.121443 | 0.999532 | | 2.674659 | -0.16753 | 0.926164 | | 2.241826 | -0.26313 | 0.877943 |
| VILL | 3.122721 | 0.408768 | 0.999532 | | 2.982738 | -0.16999 | 0.92624 | | 2.925454 | -0.56695 | 0.652925 |
| RTL6 | 3.540651 | 0.167645 | 0.999532 | | 3.577373 | 0.157762 | 0.92624 | | 3.429969 | -0.00814 | 0.997967 |
| CLIP1 | 9.123478 | 0.104959 | 0.999532 | | 10.50543 | 0.094471 | 0.92637 | | 8.907274 | 0.017403 | 0.992286 |
| SLC38A1 | 3.758503 | -0.27508 | 0.999532 | | 5.01068 | 0.137496 | 0.92637 | | 3.82193 | 0.4317 | 0.705888 |
| CCNB1IP | 2.273975 | 0.652161 | 0.999532 | | 1.894009 | -0.19802 | 0.926469 | | 1.988772 | -0.83127 | 0.542124 |
| CRAMP1 | 2.990657 | -0.06607 | 0.999532 | | 3.175254 | -0.16588 | 0.926484 | | 2.70787 | -0.07744 | 0.972924 |
| CTNNA3 | 1.813023 | 0.005357 | 0.999532 | | 1.944507 | -0.20217 | 0.926484 | | 1.613435 | -0.18857 | 0.934597 |
| ZNF211 | 2.742184 | 0.049435 | 0.999532 | | 3.142018 | 0.154228 | 0.926484 | | 2.734778 | 0.122001 | 0.946512 |
| CPZ | 2.963154 | 0.364694 | 0.999532 | | 3.065795 | -0.16285 | 0.926484 | | 2.953213 | -0.50149 | 0.69872 |
| CTNNAL | 3.751191 | 0.040886 | 0.999532 | | 4.73164 | 0.130259 | 0.926484 | | 3.792502 | 0.116451 | 0.941197 |
| RAP1A | 8.229887 | -0.10294 | 0.999532 | | 9.728067 | 0.100459 | 0.926484 | | 8.136474 | 0.212719 | 0.827267 |
| LGALS12 | 2.31902 | 0.558807 | 0.999532 | | 2.273047 | 0.180677 | 0.926484 | | 2.402658 | -0.36063 | 0.815293 |
| ZNF296 | 2.333044 | 0.227068 | 0.999532 | | 2.384424 | 0.178839 | 0.926484 | | 2.376905 | -0.03663 | 0.991273 |
| TBRG4 | 5.046902 | -0.24877 | 0.999532 | | 6.234242 | 0.120074 | 0.926484 | | 4.956649 | 0.395288 | 0.686214 |
| ERGIC1 | 7.229891 | -0.03684 | 0.999532 | | 8.150067 | 0.112768 | 0.926484 | | 7.011251 | 0.158097 | 0.891485 |
| GON7 | 2.32607 | 0.005663 | 0.999532 | | 2.759552 | 0.163409 | 0.926484 | | 2.386338 | 0.175936 | 0.923864 |
| C3orf52 | 3.032694 | 0.155709 | 0.999532 | | 3.103286 | -0.17326 | 0.926484 | | 2.872857 | -0.31014 | 0.857043 |
| DLAT | 4.294938 | 0.09977 | 0.999532 | | 4.834004 | 0.129279 | 0.926484 | | 4.446032 | 0.053739 | 0.975857 |
| STT3A | 5.596305 | -0.39005 | 0.999532 | | 7.114274 | 0.110611 | 0.926484 | | 5.323632 | 0.511979 | 0.564783 |
| B3GAT1 | 2.168089 | 0.754824 | 0.999532 | | 2.008571 | 0.204753 | 0.926484 | | 2.246514 | -0.53532 | 0.729933 |
| BRAP | 2.521159 | 0.35734 | 0.999532 | | 2.640206 | 0.164714 | 0.926484 | | 2.560148 | -0.17422 | 0.918024 |
| RPS6KC | 2.903038 | 0.085502 | 0.999532 | | 3.389243 | 0.149549 | 0.926484 | | 2.925137 | 0.08159 | 0.968235 |
| MMS22L | 1.977632 | 0.789243 | 0.999532 | | 1.902011 | 0.197781 | 0.926484 | | 2.022775 | -0.5676 | 0.701251 |
| DAND5 | 4.066674 | 0.383677 | 0.999532 | | 3.724315 | -0.14436 | 0.926504 | | 3.671166 | -0.51141 | 0.659545 |
| UQCC2 | 4.01991 | -0.21221 | 0.999532 | | 4.688401 | 0.128936 | 0.926506 | | 3.908626 | 0.354462 | 0.764149 |
| SSX2IP | 2.398036 | 0.37357 | 0.999532 | | 2.148593 | -0.18634 | 0.926506 | | 2.197332 | -0.54581 | 0.718643 |
| CHPF2 | 4.402029 | 0.182874 | 0.999532 | | 4.998368 | 0.126199 | 0.926506 | | 4.431153 | -0.03766 | 0.98556 |
| PPARGC | 2.536948 | 0.348844 | 0.999532 | | 2.555018 | -0.18202 | 0.926506 | | 2.414485 | -0.50941 | 0.722703 |
| ACTR8 | 2.718394 | -0.09705 | 0.999532 | | 3.354149 | 0.14723 | 0.926506 | | 2.734392 | 0.26214 | 0.862165 |
| SMIM18 | 2.139342 | 0.514526 | 0.999532 | | 2.228558 | 0.189548 | 0.926551 | | 2.211165 | -0.30566 | 0.862823 |
| TSEN54 | 2.25514 | -0.11954 | 0.999532 | | 2.742125 | 0.169346 | 0.926789 | | 2.335202 | 0.301872 | 0.863792 |
| ARMCX4 | 2.679139 | 0.345379 | 0.999532 | | 2.645125 | -0.17114 | 0.926789 | | 2.470696 | -0.4922 | 0.722703 |
| CCDC32 | 3.282743 | 0.246317 | 0.999532 | | 3.487898 | 0.14486 | 0.926789 | | 3.36311 | -0.08082 | 0.965619 |
| LYST | 3.140529 | 0.113894 | 0.999532 | | 3.239069 | -0.15396 | 0.926789 | | 2.832442 | -0.24824 | 0.87591 |
| MZT2B | 658.1899 | -0.28311 | 0.999532 | | 850.8312 | 0.127432 | 0.926851 | | 661.8154 | 0.434806 | 0.646863 |
| CNN1 | 2.332316 | 0.402734 | 0.999532 | | 2.546787 | 0.17449 | 0.926851 | | 2.331057 | -0.19974 | 0.912151 |
| IQSEC3 | 1.994395 | -0.2646 | 0.999532 | | 2.555783 | 0.174359 | 0.926851 | | 2.114117 | 0.463426 | 0.765012 |
| LYPD8 | 1.920522 | 0.079554 | 0.999532 | | 2.039876 | -0.19534 | 0.926851 | | 1.80301 | -0.25417 | 0.901033 |
| INPP5F | 3.080206 | 0.024917 | 0.999532 | | 3.121116 | -0.16093 | 0.926851 | | 2.788278 | -0.16842 | 0.92637 |
| OLFML1 | 4.691737 | 0.047958 | 0.999532 | | 5.464073 | 0.120065 | 0.926851 | | 4.691147 | 0.089959 | 0.951792 |
| SPAAR | 3.535552 | -0.06468 | 0.999532 | | 3.918697 | -0.14436 | 0.927098 | | 3.319756 | -0.06039 | 0.976441 |
| KLHL8 | 2.517132 | 0.398544 | 0.999532 | | 2.551825 | 0.175995 | 0.927098 | | 2.347392 | -0.19961 | 0.906595 |
| FAM3A | 8.388484 | 0.284794 | 0.999532 | | 8.075778 | -0.10232 | 0.927098 | | 7.676057 | -0.37145 | 0.65516 |
| MRPL12 | 6.8905 | 0.088528 | 0.999532 | | 7.827478 | 0.105465 | 0.927119 | | 6.724038 | 0.038025 | 0.978927 |
| PNMA2 | 2.213817 | -0.05267 | 0.999532 | | 2.599442 | 0.174629 | 0.927198 | | 2.259722 | 0.243834 | 0.895237 |
| PLPBP | 4.814106 | -0.3112 | 0.999532 | | 6.047915 | 0.11911 | 0.927383 | | 4.688474 | 0.435978 | 0.682165 |
| TTC21B | 2.076007 | 0.526213 | 0.999532 | | 2.0448 | 0.196987 | 0.927447 | | 2.144831 | -0.30937 | 0.865635 |
| PXN | 8.030553 | -0.32452 | 0.999532 | | 10.28156 | 0.096898 | 0.927447 | | 7.912832 | 0.435155 | 0.626811 |
| KRBOX4 | 3.696789 | 0.139156 | 0.999532 | | 4.02957 | -0.14425 | 0.927447 | | 3.634506 | -0.25502 | 0.859898 |
| OR5H14 | 3.127784 | 0.641968 | 0.999532 | | 2.879537 | 0.167618 | 0.927447 | | 3.161346 | -0.46537 | 0.735469 |
| CCP110 | 2.465078 | -0.05233 | 0.999532 | | 2.90368 | 0.159709 | 0.927447 | | 2.322909 | 0.226282 | 0.894555 |
| GRIA1 | 2.2434 | 0.683676 | 0.999532 | | 1.797654 | -0.20295 | 0.927447 | | 2.07543 | -0.86951 | 0.524206 |
| HOXC5 | 3.432863 | 0.173433 | 0.999532 | | 3.714138 | 0.141325 | 0.927447 | | 3.492699 | -0.01199 | 0.996891 |
| LUC7L2 | 11.49975 | -0.2273 | 0.999532 | | 13.34587 | -0.08686 | 0.927447 | | 10.48042 | 0.158377 | 0.864245 |
| TM4SF18 | 6.379336 | -0.00848 | 0.999532 | | 7.100187 | 0.118493 | 0.927447 | | 6.296392 | 0.153759 | 0.897958 |
| UCKL1 | 4.04773 | 0.102758 | 0.999532 | | 4.804026 | 0.128168 | 0.927447 | | 4.142807 | 0.047523 | 0.978927 |
| CREB1 | 6.052785 | -0.02488 | 0.999532 | | 6.604552 | -0.11864 | 0.927447 | | 5.597645 | -0.07438 | 0.960773 |
| TAS2R60 | 3.759415 | 0.427448 | 0.999532 | | 3.373342 | -0.15653 | 0.927447 | | 3.467744 | -0.57223 | 0.633741 |
| PLCB3 | 4.044382 | -0.17833 | 0.999532 | | 4.857966 | 0.12907 | 0.927447 | | 4.127719 | 0.334805 | 0.783005 |
| FCAMR | 2.364946 | 0.356727 | 0.999532 | | 2.40267 | 0.201397 | 0.927447 | | 2.456685 | -0.18503 | 0.934451 |
| CASP1 | 4.804782 | 0.082427 | 0.999532 | | 5.064197 | -0.12143 | 0.927447 | | 4.534828 | -0.18453 | 0.883116 |
| COG1 | 3.902718 | 0.370427 | 0.999532 | | 3.615086 | -0.1469 | 0.927447 | | 3.567484 | -0.50162 | 0.649779 |
| ADAMTS | 3.198598 | -0.04468 | 0.999532 | | 3.953992 | 0.141726 | 0.927447 | | 3.267456 | 0.206322 | 0.889853 |
| FAM32A | 4.741683 | -0.37899 | 0.999532 | | 5.948234 | -0.1175 | 0.927811 | | 4.163294 | 0.274627 | 0.814922 |
| ST7L | 3.027098 | -0.03411 | 0.999532 | | 3.619588 | 0.14718 | 0.927811 | | 3.005152 | 0.198802 | 0.903339 |
| MAP3K2 | 3.042248 | 0.164158 | 0.999532 | | 3.574688 | 0.146732 | 0.927811 | | 3.105767 | -0.00071 | 0.999741 |
| ING2 | 2.41926 | 0.240625 | 0.999532 | | 2.40397 | -0.1735 | 0.927811 | | 2.345203 | -0.39456 | 0.800239 |
| PPFIBP2 | 2.4808 | -0.30171 | 0.999532 | | 2.955704 | -0.15752 | 0.927811 | | 2.262139 | 0.163911 | 0.929529 |
| L3MBTL2 | 2.000065 | -0.09405 | 0.999532 | | 2.666154 | 0.171296 | 0.927811 | | 2.183286 | 0.282076 | 0.874514 |
| LRRK2 | 4.592268 | 0.018161 | 0.999532 | | 5.472778 | 0.11746 | 0.927811 | | 4.647693 | 0.121267 | 0.927668 |
| RPS4Y1 | 5.248816 | -0.17086 | 0.999532 | | 6.742943 | 0.236919 | 0.927811 | | 5.479081 | 0.433141 | 0.853549 |
| EFNA5 | 1.901748 | 0.00052 | 0.999532 | | 2.318919 | 0.183668 | 0.927811 | | 1.847705 | 0.200808 | 0.920124 |
| COX18 | 4.208524 | -0.34952 | 0.999532 | | 5.274788 | -0.1272 | 0.927811 | | 3.870994 | 0.245733 | 0.85621 |
| PTPN23 | 5.283825 | -0.05052 | 0.999532 | | 6.091364 | 0.117418 | 0.927811 | | 5.021208 | 0.186528 | 0.880137 |
| TUT4 | 3.618336 | 0.381086 | 0.999532 | | 3.492439 | 0.146903 | 0.927965 | | 3.474625 | -0.21515 | 0.881689 |
| LONRF1 | 2.730061 | -0.11857 | 0.999532 | | 3.276138 | 0.167965 | 0.927965 | | 2.780177 | 0.312398 | 0.853699 |
| IRAK1 | 4.611955 | 0.023201 | 0.999532 | | 4.754955 | -0.1312 | 0.927965 | | 4.03456 | -0.14255 | 0.926957 |
| INPPL1 | 6.779852 | 0.157601 | 0.999532 | | 7.34711 | 0.105709 | 0.927983 | | 6.615885 | -0.03408 | 0.982338 |
| CST7 | 2.99068 | -0.16297 | 0.999532 | | 3.159286 | -0.15151 | 0.927983 | | 2.651226 | 0.015022 | 0.996681 |
| ALCAM | 4.406661 | -0.15687 | 0.999532 | | 5.208693 | -0.13763 | 0.928067 | | 3.90517 | 0.058746 | 0.974588 |
| DUSP22 | 5.187517 | 0.00827 | 0.999532 | | 5.418365 | -0.12188 | 0.928067 | | 4.755856 | -0.11437 | 0.940162 |
| ORC6 | 2.327564 | 0.557723 | 0.999532 | | 2.010769 | -0.19087 | 0.928067 | | 2.183111 | -0.7293 | 0.616235 |
| CNR1 | 2.044726 | 0.000502 | 0.999532 | | 2.050277 | -0.18502 | 0.928067 | | 1.847085 | -0.16853 | 0.939628 |
| GCLM | 2.414881 | -0.46868 | 0.999532 | | 2.917748 | -0.15667 | 0.928067 | | 2.262533 | 0.335109 | 0.840407 |
| LARP4 | 3.340053 | -0.13369 | 0.999532 | | 3.737379 | -0.13838 | 0.928067 | | 3.037853 | 0.014818 | 0.996681 |
| SYNE4 | 2.46396 | 0.748812 | 0.999532 | | 1.956119 | -0.1946 | 0.928067 | | 2.186418 | -0.92298 | 0.461117 |
| NNMT | 2.890637 | -0.46978 | 0.999532 | | 3.384884 | -0.14993 | 0.928067 | | 2.428827 | 0.330331 | 0.836453 |
| CD44 | 5.138983 | 0.148317 | 0.999532 | | 5.628024 | 0.118617 | 0.928067 | | 5.169432 | -0.01028 | 0.996891 |
| ZNF234 | 2.276441 | 0.332122 | 0.999532 | | 2.362768 | 0.177451 | 0.928067 | | 2.265339 | -0.14234 | 0.945456 |
| FSD1L | 3.723165 | 0.518362 | 0.999532 | | 3.47969 | 0.14981 | 0.928067 | | 3.621943 | -0.35116 | 0.780607 |
| SPAG11A | 3.688259 | 0.247267 | 0.999532 | | 4.107739 | 0.155302 | 0.928067 | | 3.880689 | -0.085 | 0.965649 |
| SMG5 | 6.178555 | 0.086466 | 0.999532 | | 6.055543 | -0.1129 | 0.928067 | | 5.471881 | -0.20065 | 0.874251 |
| TFE3 | 2.285466 | -0.151 | 0.999532 | | 2.34729 | -0.17054 | 0.928108 | | 1.956617 | -0.01195 | 0.996891 |
| TMEM67 | 2.640331 | 0.398075 | 0.999532 | | 2.595188 | 0.167629 | 0.928108 | | 2.486319 | -0.21451 | 0.89901 |
| JAG1 | 4.359101 | -0.4123 | 0.999532 | | 6.148817 | 0.122674 | 0.928108 | | 4.703435 | 0.552964 | 0.557909 |
| TBL2 | 4.628744 | 0.174918 | 0.999532 | | 4.486207 | -0.12887 | 0.928108 | | 4.205032 | -0.29439 | 0.809832 |
| ZSWIM6 | 4.631603 | 0.090652 | 0.999532 | | 4.893857 | -0.12594 | 0.928108 | | 4.240259 | -0.19835 | 0.883104 |
| ITGB1BP | 2.064829 | 0.484957 | 0.999532 | | 2.066661 | 0.189146 | 0.928108 | | 2.111629 | -0.27817 | 0.882594 |
| UBAP2 | 3.989521 | -0.16528 | 0.999532 | | 4.383928 | -0.13919 | 0.928108 | | 3.401331 | 0.048318 | 0.979996 |
| BEND5 | 2.369282 | 0.264212 | 0.999532 | | 2.413862 | 0.177074 | 0.928108 | | 2.422854 | -0.08549 | 0.972516 |
| ARHGEF | 5.193456 | 0.016964 | 0.999532 | | 6.244413 | 0.109297 | 0.928108 | | 4.973898 | 0.105959 | 0.9462 |
| FAM90A2 | 2.170291 | 0.179408 | 0.999532 | | 2.469399 | 0.175212 | 0.928152 | | 2.21149 | 0.01612 | 0.996681 |
| MPV17 | 3.312386 | 0.242036 | 0.999532 | | 3.5956 | 0.146406 | 0.928152 | | 3.486415 | -0.06973 | 0.973152 |
| CPXM1 | 4.845545 | 1.197359 | 0.396029 | | 3.572976 | 0.165931 | 0.928152 | | 4.482521 | -1.01227 | 0.319712 |
| REM1 | 3.089614 | 0.192891 | 0.999532 | | 3.604825 | 0.157007 | 0.928152 | | 3.268578 | -0.02442 | 0.993076 |
| IFT81 | 2.128457 | 0.294501 | 0.999532 | | 2.383497 | 0.17652 | 0.928152 | | 2.187148 | -0.10072 | 0.963866 |
| MRPS34 | 3.990772 | 0.08196 | 0.999532 | | 4.577245 | 0.13533 | 0.928152 | | 3.963515 | 0.068299 | 0.970878 |
| CUL5 | 4.306483 | 0.014423 | 0.999532 | | 4.648706 | -0.12864 | 0.928152 | | 4.025829 | -0.12426 | 0.93626 |
| KCNH3 | 4.146345 | -0.02901 | 0.999532 | | 4.762195 | -0.13692 | 0.928152 | | 3.949051 | -0.07228 | 0.971732 |
| SYCP3 | 1.843517 | 0.076571 | 0.999532 | | 1.980176 | -0.197 | 0.928152 | | 1.682548 | -0.25451 | 0.903054 |
| LMNB1 | 1.845407 | -0.34598 | 0.999532 | | 2.3143 | 0.171108 | 0.928152 | | 1.908868 | 0.534931 | 0.739496 |
| TSNAXIP | 2.839429 | 0.931426 | 0.999532 | | 2.082564 | -0.18392 | 0.928152 | | 2.685746 | -1.09495 | 0.298628 |
| COL9A2 | 2.766695 | -0.56898 | 0.999532 | | 3.509635 | -0.15475 | 0.928152 | | 2.584775 | 0.44407 | 0.759211 |
| SPATA2 | 2.068995 | 0.211164 | 0.999532 | | 2.075072 | -0.19116 | 0.928288 | | 1.765153 | -0.38318 | 0.832317 |
| SCN4B | 2.644013 | 0.096477 | 0.999532 | | 2.845336 | -0.15863 | 0.928393 | | 2.480195 | -0.23153 | 0.891051 |
| POT1 | 2.678096 | 0.03837 | 0.999532 | | 2.598406 | -0.16219 | 0.928463 | | 2.420006 | -0.18781 | 0.91707 |
| RGS22 | 2.098756 | 0.754171 | 0.999532 | | 1.953251 | 0.191041 | 0.928658 | | 2.165873 | -0.54114 | 0.717627 |
| PPP1R3 | 4.822417 | -0.11494 | 0.999532 | | 5.918447 | 0.133907 | 0.928691 | | 4.757388 | 0.27439 | 0.821967 |
| TTLL5 | 3.115199 | 0.044406 | 0.999532 | | 3.541847 | 0.162267 | 0.928691 | | 3.034663 | 0.145342 | 0.937326 |
| AS3MT | 2.90762 | 0.248033 | 0.999532 | | 2.825405 | 0.155487 | 0.928691 | | 2.879203 | -0.07502 | 0.972758 |
| FMO2 | 3.437056 | -0.20242 | 0.999532 | | 4.089052 | -0.14299 | 0.928691 | | 3.045552 | 0.103833 | 0.960773 |
| ADRA1B | 2.260567 | 0.024839 | 0.999532 | | 2.40896 | -0.18194 | 0.928691 | | 2.091714 | -0.18891 | 0.926702 |
| GOLGA7 | 11.52939 | -0.2056 | 0.999532 | | 13.16381 | -0.07862 | 0.928851 | | 10.74984 | 0.147053 | 0.869224 |
| SPDYC | 2.285404 | 0.648108 | 0.999532 | | 2.119073 | 0.193023 | 0.928852 | | 2.483059 | -0.44322 | 0.792539 |
| SLC5A8 | 1.828099 | 0.211696 | 0.999532 | | 1.805416 | -0.20315 | 0.928855 | | 1.705279 | -0.39637 | 0.839373 |
| ELMO2 | 4.203976 | 0.227669 | 0.999532 | | 4.562945 | 0.129875 | 0.928855 | | 4.146573 | -0.07891 | 0.963309 |
| AARSD1 | 3.644961 | 0.16686 | 0.999532 | | 3.814633 | 0.138084 | 0.928855 | | 3.583667 | -0.01889 | 0.995256 |
| CNGA1 | 3.47978 | 0.239442 | 0.999532 | | 3.581978 | -0.1528 | 0.928855 | | 3.093593 | -0.3577 | 0.808547 |
| CRADD | 3.281067 | -0.3273 | 0.999532 | | 4.054054 | 0.130519 | 0.928855 | | 3.186602 | 0.462617 | 0.720106 |
| IPO9 | 5.253672 | -0.03771 | 0.999532 | | 5.490084 | -0.12272 | 0.928855 | | 4.759029 | -0.06006 | 0.972305 |
| PGD | 6.810198 | 0.11734 | 0.999532 | | 7.267418 | 0.105056 | 0.928855 | | 6.673541 | 0.003619 | 0.99858 |
| H3C15 | 4.995901 | -0.37664 | 0.999532 | | 5.900679 | -0.16011 | 0.928855 | | 4.55935 | 0.225001 | 0.895345 |
| RBM28 | 3.323142 | -0.11147 | 0.999532 | | 3.536893 | -0.13956 | 0.928855 | | 2.983035 | -0.01452 | 0.996681 |
| CAND2 | 2.873936 | 0.024382 | 0.999532 | | 3.157261 | -0.15253 | 0.928855 | | 2.632348 | -0.1538 | 0.931749 |
| MAP3K12 | 2.252688 | -0.29817 | 0.999532 | | 2.860487 | 0.157274 | 0.928855 | | 2.325366 | 0.47529 | 0.743576 |
| LPAR2 | 4.041633 | 0.056542 | 0.999532 | | 4.091702 | -0.13832 | 0.928855 | | 3.839 | -0.17058 | 0.906743 |
| ZNF718 | 2.761724 | 0.431013 | 0.999532 | | 2.925517 | 0.162555 | 0.928855 | | 2.837384 | -0.24628 | 0.881379 |
| DDX59 | 2.970063 | -0.32021 | 0.999532 | | 3.704877 | -0.14741 | 0.928855 | | 2.84072 | 0.194904 | 0.908891 |
| LAMC2 | 2.233811 | -0.02379 | 0.999532 | | 2.253024 | -0.1764 | 0.928855 | | 1.978622 | -0.14109 | 0.948343 |
| TPRN | 3.341854 | 0.328891 | 0.999532 | | 3.154121 | -0.16102 | 0.928855 | | 3.089352 | -0.47438 | 0.720106 |
| IRAK2 | 2.469132 | -0.10398 | 0.999532 | | 2.581997 | -0.17229 | 0.928855 | | 2.167809 | -0.05193 | 0.985693 |
| ASB4 | 2.010811 | 0.798484 | 0.999532 | | 1.802565 | 0.197363 | 0.928855 | | 2.040667 | -0.58257 | 0.69977 |
| OR2T5 | 2.339163 | 0.248796 | 0.999532 | | 2.429301 | -0.18153 | 0.928855 | | 2.185434 | -0.41015 | 0.799627 |
| EFNA1 | 8.636973 | -0.06371 | 0.999532 | | 9.70874 | -0.09854 | 0.928855 | | 8.138272 | -0.01452 | 0.994537 |
| BORCS6 | 2.050662 | 0.303917 | 0.999532 | | 2.169419 | 0.184861 | 0.928964 | | 2.100801 | -0.10323 | 0.965649 |
| FADS1 | 4.126226 | -0.55176 | 0.999532 | | 5.443714 | 0.122517 | 0.928964 | | 4.253925 | 0.685426 | 0.512688 |
| CRNKL1 | 2.836904 | -0.27507 | 0.999532 | | 3.798064 | 0.134601 | 0.92928 | | 2.975678 | 0.429366 | 0.730921 |
| SALL2 | 2.536213 | 0.563689 | 0.999532 | | 2.074771 | -0.18641 | 0.92928 | | 2.407775 | -0.73281 | 0.614824 |
| PDCD6 | 5.538758 | -0.12913 | 0.999532 | | 6.519878 | -0.11433 | 0.92928 | | 5.061443 | 0.044909 | 0.979432 |
| ACSS1 | 3.72606 | 0.359983 | 0.999532 | | 3.854526 | 0.133935 | 0.92928 | | 3.632837 | -0.21317 | 0.88082 |
| TBL3 | 3.726087 | 0.271805 | 0.999532 | | 3.755097 | 0.13625 | 0.92928 | | 3.643261 | -0.11756 | 0.941197 |
| EHMT2 | 2.517689 | 0.298712 | 0.999532 | | 2.789662 | 0.164643 | 0.92928 | | 2.733698 | -0.11701 | 0.954304 |
| PGBD2 | 2.317421 | -0.36627 | 0.999532 | | 2.667639 | -0.16501 | 0.92928 | | 2.136497 | 0.217087 | 0.909259 |
| KIF3A | 2.32067 | -0.2441 | 0.999532 | | 2.609377 | -0.16332 | 0.92928 | | 2.091538 | 0.099019 | 0.9657 |
| CTRL | 2.457868 | 0.477462 | 0.999532 | | 2.234703 | -0.18351 | 0.92928 | | 2.238175 | -0.64252 | 0.647218 |
| TOLLIP | 5.360196 | -0.22263 | 0.999532 | | 6.763861 | 0.107758 | 0.92928 | | 5.373045 | 0.349751 | 0.720526 |
| HACL1 | 3.176965 | 0.08222 | 0.999532 | | 3.834292 | 0.146629 | 0.92928 | | 3.285635 | 0.078214 | 0.968434 |
| CCL28 | 2.278076 | 0.359031 | 0.999532 | | 2.119119 | 0.176413 | 0.92928 | | 2.287663 | -0.18618 | 0.925804 |
| SERPINB | 1.926184 | 0.850635 | 0.999532 | | 1.8155 | 0.199858 | 0.92928 | | 2.024921 | -0.62793 | 0.678864 |
| PPARGC | 3.912581 | 0.247925 | 0.999532 | | 3.684218 | -0.14647 | 0.92928 | | 3.365056 | -0.3751 | 0.763181 |
| PPARG | 4.918385 | -0.16581 | 0.999532 | | 5.8861 | 0.113017 | 0.92928 | | 5.096546 | 0.304632 | 0.784955 |
| ATP6V0E | 3.415519 | -0.05291 | 0.999532 | | 4.146929 | 0.131958 | 0.92928 | | 3.419052 | 0.20594 | 0.88885 |
| WNT1 | 2.052622 | -0.04015 | 0.999532 | | 2.31691 | -0.17949 | 0.92928 | | 1.910275 | -0.11855 | 0.960773 |
| FEZ1 | 4.241412 | -0.25153 | 0.999532 | | 5.145401 | -0.13231 | 0.92928 | | 4.022295 | 0.13711 | 0.928522 |
| NFU1 | 3.271148 | -0.14611 | 0.999532 | | 3.826869 | 0.134383 | 0.92928 | | 3.236465 | 0.299104 | 0.824012 |
| TMEM24 | 2.38621 | -0.08219 | 0.999532 | | 2.532579 | -0.16498 | 0.92928 | | 2.24308 | -0.06593 | 0.978312 |
| TNNC1 | 4.576968 | 0.308124 | 0.999532 | | 4.815233 | 0.125191 | 0.92928 | | 4.490513 | -0.16441 | 0.908829 |
| TRIM9 | 3.268853 | 0.637352 | 0.999532 | | 2.92357 | -0.15808 | 0.92928 | | 3.094567 | -0.76451 | 0.512069 |
| TMEM13 | 3.224977 | 0.626637 | 0.999532 | | 2.844038 | 0.158763 | 0.92928 | | 3.185829 | -0.45225 | 0.722703 |
| RGMB | 2.977149 | 0.173715 | 0.999532 | | 2.942261 | -0.15636 | 0.92928 | | 2.794181 | -0.30588 | 0.849682 |
| SPNS2 | 3.766091 | 0.101665 | 0.999532 | | 4.026542 | 0.134493 | 0.92928 | | 3.766943 | 0.04871 | 0.979434 |
| TYW3 | 1.658632 | -0.0126 | 0.999532 | | 1.889323 | 0.183701 | 0.92928 | | 1.724322 | 0.215352 | 0.916969 |
| KIF26A | 3.218521 | 0.178651 | 0.999532 | | 3.140499 | -0.15325 | 0.92928 | | 2.950411 | -0.32108 | 0.819337 |
| COMMD1 | 6.172243 | 0.020861 | 0.999532 | | 6.623052 | -0.11188 | 0.92928 | | 5.509456 | -0.1112 | 0.932589 |
| TAAR9 | 1.936567 | 0.4025 | 0.999532 | | 2.049062 | 0.187565 | 0.92928 | | 1.989988 | -0.19652 | 0.920487 |
| DCDC2B | 2.056573 | 0.020745 | 0.999532 | | 2.287788 | -0.18239 | 0.92928 | | 1.954103 | -0.18078 | 0.933313 |
| SIGLEC1 | 4.012044 | 0.147363 | 0.999532 | | 4.101563 | -0.14031 | 0.92928 | | 3.980637 | -0.27489 | 0.842754 |
| ST8SIA5 | 2.28495 | 0.363456 | 0.999532 | | 2.145786 | -0.1784 | 0.92928 | | 2.119223 | -0.52273 | 0.72472 |
| PRAMEF | 2.95425 | 0.009837 | 0.999532 | | 3.112505 | -0.15697 | 0.92928 | | 2.681528 | -0.1513 | 0.934572 |
| ZNF774 | 2.055719 | -0.01346 | 0.999532 | | 2.171847 | -0.17594 | 0.92928 | | 1.887695 | -0.14372 | 0.947126 |
| LRRC18 | 2.025441 | 0.25265 | 0.999532 | | 2.354211 | 0.179827 | 0.92928 | | 2.080514 | -0.04741 | 0.987608 |
| GDAP1L1 | 2.481063 | -0.00188 | 0.999532 | | 2.743706 | -0.16981 | 0.929315 | | 2.465704 | -0.14703 | 0.945129 |
| SNAPIN | 5.123821 | -0.26227 | 0.999532 | | 6.081307 | -0.11782 | 0.929479 | | 4.700995 | 0.165049 | 0.898369 |
| SMCR8 | 2.674581 | 0.12638 | 0.999532 | | 2.762887 | -0.16895 | 0.929492 | | 2.526875 | -0.27494 | 0.87591 |
| JDP2 | 3.528523 | -0.14877 | 0.999532 | | 3.919449 | -0.13796 | 0.929492 | | 2.966147 | 0.033211 | 0.989841 |
| ST3GAL2 | 5.580787 | 0.23334 | 0.999532 | | 5.810844 | 0.128285 | 0.929551 | | 5.684896 | -0.08189 | 0.96129 |
| TUSC2 | 9.135413 | 0.198363 | 0.999532 | | 9.070214 | -0.10123 | 0.929551 | | 8.387608 | -0.2774 | 0.740127 |
| PREP | 3.820407 | -0.2461 | 0.999532 | | 4.548432 | -0.13021 | 0.929551 | | 3.306041 | 0.129424 | 0.93626 |
| INTS10 | 3.778936 | -0.31495 | 0.999532 | | 4.704488 | 0.127478 | 0.929612 | | 3.50271 | 0.454866 | 0.699971 |
| HMOX1 | 3.747302 | -0.3099 | 0.999532 | | 4.730186 | 0.121653 | 0.929706 | | 3.77106 | 0.450598 | 0.677493 |
| OSM | 2.751686 | 0.301065 | 0.999532 | | 2.794489 | -0.16742 | 0.929706 | | 2.710132 | -0.4544 | 0.75038 |

| | | | | | | | | | | | |
|---|---|---|---|---|---|---|---|---|---|---|---|
| MIGA2 | 2.28395 | -0.3222 | 0.999532 | | 2.727201 | -0.15681 | 0.929706 | | 2.061155 | 0.184239 | 0.924131 |
| IKZF3 | 3.097254 | 0.032033 | 0.999532 | | 3.131607 | -0.15109 | 0.929706 | | 2.808211 | -0.17381 | 0.920259 |
| OXLD1 | 3.172724 | -0.44623 | 0.999532 | | 3.886459 | -0.1376 | 0.929706 | | 2.860736 | 0.32346 | 0.819366 |
| BIK | 1.824224 | 0.225284 | 0.999532 | | 1.785314 | -0.19666 | 0.929706 | | 1.674388 | -0.40523 | 0.831749 |
| ARMCX2 | 3.307674 | 0.190569 | 0.999532 | | 3.419362 | 0.139671 | 0.929706 | | 3.186528 | -0.03955 | 0.987274 |
| NEXN | 2.589064 | 0.517686 | 0.999532 | | 2.050972 | -0.18213 | 0.929706 | | 2.371012 | -0.68211 | 0.630705 |
| CIZ1 | 4.91335 | -0.14501 | 0.999532 | | 5.972734 | 0.11321 | 0.929809 | | 4.944683 | 0.280534 | 0.798849 |
| ZNF280C | 2.405242 | 0.433796 | 0.999532 | | 2.148815 | -0.17422 | 0.929824 | | 2.218374 | -0.59468 | 0.673405 |
| ELF4 | 1.999814 | -0.09904 | 0.999532 | | 2.433866 | 0.171828 | 0.92983 | | 2.204633 | 0.292842 | 0.874661 |
| POLR3K | 4.113341 | 0.571399 | 0.999532 | | 3.534089 | -0.15382 | 0.92983 | | 3.788384 | -0.70526 | 0.527276 |
| SLC4A1 | 2.994372 | 0.177345 | 0.999532 | | 3.102572 | -0.15239 | 0.929833 | | 2.649145 | -0.31773 | 0.832101 |
| PEX13 | 2.20976 | 0.576018 | 0.999532 | | 2.084494 | 0.178381 | 0.929833 | | 2.189581 | -0.38034 | 0.810635 |
| RBP4 | 2.490028 | -0.04696 | 0.999532 | | 2.99077 | 0.161377 | 0.929833 | | 2.555592 | 0.22267 | 0.901033 |
| CYP4V2 | 3.984999 | -0.18774 | 0.999532 | | 4.922307 | 0.131067 | 0.929998 | | 4.053454 | 0.33309 | 0.796702 |
| DPCD | 2.509556 | 0.52305 | 0.999532 | | 2.519808 | 0.167958 | 0.929998 | | 2.512051 | -0.33658 | 0.824012 |
| LTF | 3.082112 | 0.028744 | 0.999532 | | 3.496747 | 0.154795 | 0.93002 | | 3.116788 | 0.135483 | 0.939549 |
| ZBTB26 | 2.488292 | -0.09908 | 0.999532 | | 2.82937 | -0.1612 | 0.930247 | | 2.239395 | -0.03791 | 0.99013 |
| S100PBP | 3.680946 | 0.489206 | 0.999532 | | 3.654271 | 0.134686 | 0.930247 | | 3.864195 | -0.3243 | 0.792176 |
| ZNF503 | 4.717334 | 0.039976 | 0.999532 | | 5.714571 | 0.135886 | 0.930247 | | 4.806753 | 0.125837 | 0.938429 |
| JAK1 | 4.272325 | -0.30425 | 0.999532 | | 5.081227 | -0.11998 | 0.930247 | | 3.901167 | 0.201891 | 0.881837 |
| IFFO2 | 2.797328 | 0.01259 | 0.999532 | | 3.355277 | 0.144515 | 0.930247 | | 2.785662 | 0.150657 | 0.92883 |
| RNF208 | 2.810983 | 0.154192 | 0.999532 | | 3.372926 | 0.160305 | 0.930247 | | 2.954914 | 0.024262 | 0.993618 |
| SARS1 | 8.59534 | -0.07764 | 0.999532 | | 10.16683 | 0.092303 | 0.930382 | | 8.417001 | 0.18733 | 0.84579 |
| FASTKD1 | 4.442849 | 0.609958 | 0.999532 | | 3.70385 | -0.1391 | 0.930382 | | 3.939374 | -0.7335 | 0.423641 |
| INTS1 | 6.858069 | 0.489424 | 0.999532 | | 6.601228 | 0.107038 | 0.930382 | | 6.871596 | -0.36271 | 0.709825 |
| NAA60 | 4.245475 | -0.22944 | 0.999532 | | 4.880334 | -0.1255 | 0.930382 | | 3.887287 | 0.123938 | 0.935174 |
| EGFL6 | 1.895912 | -0.09531 | 0.999532 | | 2.041452 | -0.1808 | 0.930382 | | 1.748559 | -0.06857 | 0.979432 |
| TM9SF4 | 4.492191 | 0.031733 | 0.999532 | | 4.741479 | -0.12241 | 0.930382 | | 3.988686 | -0.13581 | 0.922437 |
| ABCC5 | 2.389639 | -0.09309 | 0.999532 | | 2.474419 | -0.16562 | 0.930382 | | 2.069593 | -0.05928 | 0.982124 |
| ENDOV | 3.516696 | 0.691986 | 0.999532 | | 2.897962 | -0.156 | 0.930382 | | 3.366948 | -0.83347 | 0.412558 |
| AADAT | 2.531426 | 0.639227 | 0.999532 | | 2.410596 | 0.176846 | 0.930382 | | 2.539435 | -0.44485 | 0.76047 |
| TMEM70 | 2.668135 | -0.22392 | 0.999532 | | 3.415725 | 0.142439 | 0.930382 | | 2.672404 | 0.38494 | 0.785012 |
| ASGR1 | 2.466433 | 0.376467 | 0.999532 | | 2.248196 | -0.17875 | 0.930382 | | 2.262403 | -0.53587 | 0.723933 |
| ZNF219 | 3.31055 | 0.240965 | 0.999532 | | 3.600676 | 0.149181 | 0.930382 | | 3.323395 | -0.074 | 0.972135 |
| BTD | 2.644664 | 0.221437 | 0.999532 | | 2.976867 | 0.151246 | 0.930382 | | 2.653047 | -0.0525 | 0.982187 |
| LXN | 3.524433 | -0.80984 | 0.999532 | | 5.111744 | 0.124147 | 0.930382 | | 3.456778 | 0.951694 | 0.30043 |
| CARD9 | 2.959395 | 0.194492 | 0.999532 | | 2.985714 | -0.16012 | 0.930382 | | 2.921866 | -0.33505 | 0.829716 |
| TP53 | 5.095289 | 0.485545 | 0.999532 | | 4.72926 | 0.138719 | 0.930382 | | 5.10607 | -0.33709 | 0.758124 |
| ETV1 | 2.340478 | 0.343336 | 0.999532 | | 2.282276 | -0.17982 | 0.930382 | | 2.278372 | -0.50883 | 0.740093 |
| SURF4 | 5.8143 | 0.050662 | 0.999532 | | 6.792007 | 0.107355 | 0.930382 | | 5.721099 | 0.078994 | 0.956322 |
| POLR2B | 4.623474 | -0.0854 | 0.999532 | | 5.511315 | 0.113864 | 0.930617 | | 4.678304 | 0.218225 | 0.861737 |
| DEDD2 | 3.988997 | -0.23885 | 0.999532 | | 5.213656 | 0.125731 | 0.930617 | | 3.991401 | 0.37805 | 0.738861 |
| RTL8A | 2.802842 | -0.0083 | 0.999532 | | 2.884406 | -0.15237 | 0.930617 | | 2.536932 | -0.12789 | 0.946572 |
| MTCL1 | 2.198989 | 0.016334 | 0.999532 | | 2.229226 | -0.17618 | 0.930617 | | 1.996283 | -0.18041 | 0.932688 |
| FAR1 | 3.904363 | -0.0673 | 0.999532 | | 4.406901 | 0.126027 | 0.930631 | | 3.980371 | 0.213693 | 0.877682 |
| NUP42 | 3.350525 | -0.16492 | 0.999532 | | 3.586478 | -0.13929 | 0.930825 | | 3.100422 | 0.045391 | 0.985675 |
| PEAR1 | 6.035642 | -0.29215 | 0.999532 | | 7.451796 | -0.1059 | 0.930825 | | 5.750362 | 0.220493 | 0.874251 |
| RIPPLY2 | 2.468766 | 0.702993 | 0.999532 | | 1.890336 | -0.1856 | 0.930825 | | 2.204533 | -0.87229 | 0.511836 |
| GTPBP3 | 2.646621 | 0.02484 | 0.999532 | | 2.555634 | -0.16538 | 0.930825 | | 2.374228 | -0.18067 | 0.92697 |
| SDR42E1 | 2.339693 | 0.628042 | 0.999532 | | 2.213948 | 0.174984 | 0.930918 | | 2.350061 | -0.4358 | 0.774747 |
| MMUT | 1.898379 | -0.24988 | 0.999532 | | 2.385849 | 0.170254 | 0.930978 | | 1.941216 | 0.43737 | 0.801572 |
| PIK3C2A | 5.793127 | -0.0974 | 0.999532 | | 6.244876 | -0.10836 | 0.931134 | | 5.362883 | 0.01637 | 0.995112 |
| ZNF326 | 4.022006 | 0.274192 | 0.999532 | | 4.061639 | -0.13218 | 0.93117 | | 3.687928 | -0.38538 | 0.735469 |
| ANKRD4 | 5.207067 | 0.353724 | 0.999532 | | 5.039214 | -0.12013 | 0.931365 | | 4.884212 | -0.45016 | 0.655324 |
| ZBTB24 | 2.733636 | 0.344956 | 0.999532 | | 2.782544 | 0.174222 | 0.931365 | | 2.773633 | -0.16362 | 0.934701 |
| LSM14A | 4.352599 | -0.09261 | 0.999532 | | 4.520534 | -0.12615 | 0.931365 | | 3.919934 | -0.02103 | 0.993435 |
| AKR1E2 | 3.25264 | -0.24313 | 0.999532 | | 4.287807 | 0.134976 | 0.931365 | | 3.290649 | 0.399986 | 0.75311 |
| GLP2R | 2.107567 | 0.6071 | 0.999532 | | 1.777564 | -0.19686 | 0.931753 | | 1.99275 | -0.78826 | 0.599002 |
| MARCHF | 2.54949 | 0.246218 | 0.999532 | | 2.539699 | -0.17539 | 0.931764 | | 2.425243 | -0.42174 | 0.785012 |
| LRRC42 | 2.690466 | 0.120654 | 0.999532 | | 2.654903 | -0.16482 | 0.931764 | | 2.600419 | -0.26305 | 0.88588 |
| PGRMC1 | 4.326172 | 0.085937 | 0.999532 | | 4.9388 | 0.119995 | 0.931764 | | 4.236267 | 0.051405 | 0.97652 |
| SDHA | 25.05085 | -0.36693 | 0.999532 | | 31.21259 | -0.06218 | 0.931764 | | 22.91113 | 0.32894 | 0.515762 |
| AKT1S1 | 3.128618 | 0.61381 | 0.999532 | | 3.174104 | 0.145817 | 0.931764 | | 3.246508 | -0.44812 | 0.705046 |
| GABRG2 | 1.874084 | 0.2302 | 0.999532 | | 1.777461 | -0.19318 | 0.931764 | | 1.721594 | -0.40685 | 0.830269 |
| INTS2 | 2.479099 | 0.599897 | 0.999532 | | 2.414187 | 0.164292 | 0.931764 | | 2.458514 | -0.41797 | 0.766974 |
| ERCC5 | 3.723129 | 0.256213 | 0.999532 | | 3.89701 | 0.134097 | 0.931764 | | 3.720072 | -0.1069 | 0.948127 |
| MIF4GD | 3.108343 | 0.077365 | 0.999532 | | 3.144502 | -0.14582 | 0.931764 | | 2.740978 | -0.2058 | 0.897096 |
| AAMP | 6.485621 | 0.275515 | 0.999532 | | 6.975694 | 0.103233 | 0.931826 | | 6.329265 | -0.14981 | 0.893262 |
| MRPS6 | 3.865436 | -0.05123 | 0.999532 | | 4.606687 | 0.120798 | 0.931826 | | 3.729104 | 0.18665 | 0.892869 |
| CYP26B1 | 7.075225 | 0.245495 | 0.999532 | | 6.856335 | -0.12418 | 0.932011 | | 6.555341 | -0.34229 | 0.75311 |
| FZR1 | 3.312491 | -0.31471 | 0.999532 | | 4.19881 | 0.129944 | 0.932026 | | 3.266168 | 0.458653 | 0.715799 |
| APBA1 | 2.136209 | 0.180707 | 0.999532 | | 2.13975 | -0.17531 | 0.932026 | | 1.982344 | -0.3358 | 0.859898 |
| SGCD | 2.689478 | -0.0528 | 0.999532 | | 3.02607 | -0.1551 | 0.932112 | | 2.516615 | -0.08094 | 0.972305 |
| HIBADH | 4.802627 | 0.497239 | 0.999532 | | 4.60266 | 0.125571 | 0.932118 | | 4.704927 | -0.3529 | 0.741477 |
| CD47 | 6.322158 | -0.32076 | 0.999532 | | 7.427714 | -0.09845 | 0.932118 | | 5.929884 | 0.236146 | 0.824012 |
| TERF2IP | 8.056529 | 0.094683 | 0.999532 | | 8.765904 | 0.097678 | 0.932118 | | 7.632748 | 0.019211 | 0.992286 |
| CEP41 | 3.119524 | 0.651088 | 0.999532 | | 2.641733 | -0.17063 | 0.932118 | | 2.945413 | -0.80478 | 0.515762 |
| C1orf115 | 4.187047 | -0.15032 | 0.999532 | | 5.00017 | 0.137044 | 0.932118 | | 3.903196 | 0.328499 | 0.792539 |
| TBX19 | 3.574832 | -0.21269 | 0.999532 | | 4.238748 | -0.12781 | 0.932118 | | 3.239475 | 0.108822 | 0.948343 |
| SYNE2 | 6.136076 | -0.11285 | 0.999532 | | 6.892371 | -0.10538 | 0.932118 | | 5.430395 | 0.031615 | 0.986811 |
| HEATR5 | 3.444547 | 0.057321 | 0.999532 | | 3.670275 | 0.14033 | 0.932118 | | 3.337222 | 0.093656 | 0.96129 |
| FAM47C | 2.071103 | 0.397602 | 0.999532 | | 1.988709 | -0.18614 | 0.932118 | | 1.877854 | -0.5659 | 0.720106 |
| CAPZA1 | 4.658948 | -0.26674 | 0.999532 | | 5.950095 | 0.111169 | 0.932118 | | 4.776184 | 0.404072 | 0.686264 |
| TRMT1L | 3.102215 | 0.188689 | 0.999532 | | 3.203619 | -0.14967 | 0.932118 | | 2.805472 | -0.31828 | 0.827119 |
| CHCHD1 | 7.607509 | 0.02485 | 0.999532 | | 8.903405 | 0.091513 | 0.932118 | | 7.405264 | 0.08429 | 0.944993 |
| STARD4 | 1.90623 | 0.231127 | 0.999532 | | 2.114264 | 0.1737 | 0.932118 | | 1.914544 | -0.03763 | 0.991159 |
| HERC1 | 5.336779 | -0.28436 | 0.999532 | | 7.099219 | 0.105807 | 0.932118 | | 5.327413 | 0.41422 | 0.679304 |
| SLC45A3 | 1.967774 | -0.21063 | 0.999532 | | 2.299041 | -0.1738 | 0.932118 | | 1.839531 | 0.056007 | 0.985675 |
| TMEM231 | 3.685115 | 0.480524 | 0.999532 | | 3.35556 | -0.14657 | 0.932118 | | 3.355943 | -0.60275 | 0.590247 |
| CRTC1 | 3.956234 | 0.196712 | 0.999532 | | 4.564902 | 0.13265 | 0.932118 | | 3.968507 | -0.03524 | 0.988853 |
| PITX1 | 1.956517 | -0.06413 | 0.999532 | | 2.565343 | 0.176317 | 0.932118 | | 2.23631 | 0.254876 | 0.89901 |
| BRINP3 | 1.936669 | 0.235526 | 0.999532 | | 2.196836 | 0.178971 | 0.932118 | | 1.99723 | -0.03789 | 0.991159 |
| CDCA4 | 2.560952 | -0.01697 | 0.999532 | | 2.773624 | -0.16801 | 0.932118 | | 2.21707 | -0.11984 | 0.955834 |
| TMEM12 | 2.79929 | -0.10628 | 0.999532 | | 3.027101 | -0.15088 | 0.932118 | | 2.452102 | -0.02559 | 0.993618 |
| ZNF398 | 3.91849 | 0.529181 | 0.999532 | | 3.317292 | -0.14188 | 0.932118 | | 3.609223 | -0.64638 | 0.53564 |
| FAAH2 | 2.065531 | 0.744056 | 0.999532 | | 1.873807 | 0.189269 | 0.932118 | | 2.090696 | -0.53789 | 0.722703 |
| CRIP1 | 9.36756 | -0.51279 | 0.999532 | | 13.17217 | 0.131333 | 0.932118 | | 9.548524 | 0.677535 | 0.515762 |
| GPR20 | 2.127375 | -0.25063 | 0.999532 | | 2.395801 | -0.17066 | 0.932118 | | 1.924226 | 0.093993 | 0.972221 |
| HECTD3 | 3.109501 | 0.069394 | 0.999532 | | 3.240468 | -0.14217 | 0.932118 | | 2.889588 | -0.19384 | 0.902504 |
| STAT5A | 3.600696 | -0.16256 | 0.999532 | | 3.862459 | -0.13513 | 0.932157 | | 3.274031 | 0.03138 | 0.991224 |
| DSE | 3.054801 | 0.021064 | 0.999532 | | 3.369351 | 0.134753 | 0.932157 | | 3.076943 | 0.129019 | 0.939549 |
| PDZD2 | 5.908015 | -0.13231 | 0.999532 | | 6.695579 | -0.11494 | 0.932213 | | 5.46426 | 0.035242 | 0.987405 |
| NAPEPLD | 3.723978 | -0.06614 | 0.999532 | | 4.003915 | -0.13624 | 0.932213 | | 3.536941 | -0.05263 | 0.979996 |
| TGIF1 | 3.912393 | -0.27234 | 0.999532 | | 4.809119 | -0.123 | 0.932213 | | 3.377514 | 0.150108 | 0.923912 |
| GPR156 | 1.829659 | 0.344394 | 0.999532 | | 2.004938 | 0.183381 | 0.932258 | | 1.873212 | -0.14194 | 0.949669 |
| AZI2 | 4.099298 | -0.00817 | 0.999532 | | 4.326901 | -0.12261 | 0.932294 | | 3.587503 | -0.09882 | 0.951656 |
| MYO5C | 2.380249 | 0.656528 | 0.999532 | | 2.329092 | 0.1742 | 0.932294 | | 2.380217 | -0.46456 | 0.751138 |
| ADK | 3.752365 | -0.10031 | 0.999532 | | 4.431876 | 0.121875 | 0.932294 | | 3.615431 | 0.235863 | 0.863401 |
| FAM163A | 1.970719 | -0.44027 | 0.999532 | | 2.414921 | -0.16595 | 0.932294 | | 1.847805 | 0.2918 | 0.883116 |
| ERH | 12.79645 | -0.28032 | 0.999532 | | 15.72645 | 0.077669 | 0.932294 | | 12.22942 | 0.367513 | 0.583626 |
| RIT1 | 3.239311 | -0.00717 | 0.999532 | | 3.509016 | 0.139796 | 0.932294 | | 3.187966 | 0.170167 | 0.920259 |
| ELOA3P | 6.936354 | 0.030913 | 0.999532 | | 7.738259 | -0.1351 | 0.932294 | | 6.491524 | -0.13707 | 0.922922 |
| FYB1 | 2.117989 | 0.237507 | 0.999532 | | 2.064562 | -0.17986 | 0.932294 | | 2.039579 | -0.39781 | 0.823193 |
| PTPRN2 | 2.229387 | 0.211173 | 0.999532 | | 2.076036 | -0.18028 | 0.932294 | | 2.142267 | -0.37594 | 0.837256 |
| PEX10 | 3.824665 | 0.346801 | 0.999532 | | 3.715103 | -0.13834 | 0.932294 | | 3.509564 | -0.46711 | 0.676169 |
| ARMC1 | 4.138439 | 0.223661 | 0.999532 | | 4.484268 | 0.131577 | 0.932294 | | 4.306315 | -0.08213 | 0.96129 |
| MDC1 | 3.992591 | 0.060299 | 0.999532 | | 4.447985 | 0.1282 | 0.932294 | | 3.784184 | 0.09082 | 0.955834 |
| FAM53A | 2.351115 | 0.714345 | 0.999532 | | 2.055702 | -0.18393 | 0.932294 | | 2.263339 | -0.87937 | 0.497845 |
| UTP4 | 2.281411 | -0.31664 | 0.999532 | | 2.64116 | -0.16606 | 0.932294 | | 2.010371 | 0.171088 | 0.934701 |
| DEFB4B | 2.016933 | 0.99015 | 0.999532 | | 1.706391 | 0.202214 | 0.932294 | | 2.056217 | -0.76892 | 0.611506 |
| OARD1 | 3.926508 | -0.04848 | 0.999532 | | 4.568399 | 0.126601 | 0.932391 | | 4.041662 | 0.196477 | 0.88588 |
| PTRHD1 | 4.00254 | 0.262131 | 0.999532 | | 4.04029 | -0.13302 | 0.932391 | | 3.657295 | -0.37657 | 0.752299 |
| TAL1 | 3.882403 | -0.25875 | 0.999532 | | 4.432141 | -0.13256 | 0.932391 | | 3.444497 | 0.145864 | 0.929 |
| ACER3 | 2.704617 | 0.316014 | 0.999532 | | 2.375158 | -0.16441 | 0.932447 | | 2.433492 | -0.47276 | 0.749206 |
| RC3H1 | 4.035143 | 0.093795 | 0.999532 | | 4.298115 | 0.127325 | 0.932447 | | 3.880319 | 0.04367 | 0.983199 |
| TPRG1L | 5.573755 | -0.27115 | 0.999532 | | 7.116684 | 0.098378 | 0.932702 | | 5.56529 | 0.391641 | 0.684642 |
| HLA-DMB | 4.326303 | -0.05298 | 0.999532 | | 5.439834 | 0.116818 | 0.932702 | | 4.399098 | 0.196465 | 0.881837 |
| HIF1A | 6.881797 | -0.28087 | 0.999532 | | 8.337982 | -0.09777 | 0.932702 | | 6.272185 | 0.200829 | 0.871467 |
| BTNL9 | 9.986466 | -0.10131 | 0.999532 | | 10.87141 | -0.10081 | 0.932729 | | 9.273846 | -0.00302 | 0.998655 |
| LSG1 | 2.400482 | -0.1208 | 0.999532 | | 2.671609 | -0.15674 | 0.932729 | | 2.193092 | -0.01803 | 0.996681 |
| CCL17 | 2.628624 | 0.290874 | 0.999532 | | 2.539936 | -0.16776 | 0.932729 | | 2.451702 | -0.44041 | 0.783332 |
| SPIB | 2.493462 | 0.376431 | 0.999532 | | 2.195885 | -0.1757 | 0.932729 | | 2.172276 | -0.53689 | 0.723933 |
| MATN2 | 5.819131 | -0.13789 | 0.999532 | | 6.553888 | -0.10834 | 0.932729 | | 5.222875 | 0.046129 | 0.979996 |
| TTYH1 | 1.620723 | 0.480021 | 0.999532 | | 1.624256 | 0.2009 | 0.932729 | | 1.647434 | -0.26083 | 0.903054 |
| TMEM92 | 2.119524 | 0.424412 | 0.999532 | | 1.916877 | -0.1971 | 0.932729 | | 2.012039 | -0.59727 | 0.730921 |
| DYNLT2 | 2.285585 | 0.500234 | 0.999532 | | 2.099875 | -0.17588 | 0.932729 | | 2.20472 | -0.65472 | 0.652095 |
| SH3RF1 | 3.865488 | -0.13792 | 0.999532 | | 5.007162 | 0.130054 | 0.932729 | | 4.102651 | 0.282422 | 0.829716 |
| XBP1 | 10.48062 | 0.176621 | 0.999532 | | 11.39622 | 0.086741 | 0.932729 | | 10.19122 | -0.07004 | 0.948127 |
| PLCXD3 | 2.40065 | 0.226247 | 0.999532 | | 2.548447 | 0.161941 | 0.932729 | | 2.249768 | -0.04674 | 0.986811 |
| NLRP5 | 1.881158 | -0.13371 | 0.999532 | | 2.222748 | 0.172718 | 0.932729 | | 1.887446 | 0.327003 | 0.864245 |
| KPNA5 | 3.561294 | 0.282519 | 0.999532 | | 3.726149 | 0.138048 | 0.932729 | | 3.58662 | -0.12955 | 0.939107 |
| ZNF221 | 2.513597 | 0.459851 | 0.999532 | | 2.205868 | -0.17185 | 0.932729 | | 2.295603 | -0.61498 | 0.660097 |
| GGTA1 | 4.618253 | 0.021788 | 0.999532 | | 4.854247 | -0.12269 | 0.932729 | | 4.320254 | -0.1246 | 0.927668 |
| PPP2R1B | 4.18746 | 0.025535 | 0.999532 | | 4.653354 | 0.128394 | 0.932729 | | 3.911085 | 0.131405 | 0.929335 |
| SET | 8.091383 | -0.10983 | 0.999532 | | 9.879601 | 0.099102 | 0.932729 | | 7.958364 | 0.236356 | 0.801572 |
| PHF6 | 2.34047 | -0.09474 | 0.999532 | | 2.604802 | -0.16293 | 0.932729 | | 2.031771 | -0.04969 | 0.985873 |
| KIF21B | 1.957425 | 0.249867 | 0.999532 | | 2.058151 | -0.18588 | 0.932827 | | 1.798542 | -0.41351 | 0.815886 |
| OPCML | 2.444575 | -0.11818 | 0.999532 | | 2.805003 | -0.15444 | 0.932891 | | 2.373678 | -0.0099 | 0.997084 |
| SCNM1 | 7.646265 | 0.18223 | 0.999532 | | 8.011357 | 0.094633 | 0.932891 | | 7.769238 | -0.06618 | 0.963309 |
| TAS2R30 | 2.197191 | 0.136676 | 0.999532 | | 2.121941 | -0.1687 | 0.932891 | | 2.03509 | -0.28832 | 0.87591 |
| KDELR2 | 4.46246 | -0.2413 | 0.999532 | | 5.75202 | 0.108139 | 0.93291 | | 4.4392 | 0.367902 | 0.735469 |
| RBM15B | 5.06156 | 0.162637 | 0.999532 | | 5.755565 | 0.122655 | 0.933152 | | 5.022267 | -0.02288 | 0.991434 |
| ZNF473 | 1.901645 | -0.0153 | 0.999532 | | 1.933055 | -0.17672 | 0.933152 | | 1.67811 | -0.14572 | 0.949252 |
| IPO13 | 2.561486 | 0.318909 | 0.999532 | | 2.585559 | 0.151414 | 0.933152 | | 2.526364 | -0.14963 | 0.932757 |
| ABHD16A | 7.105476 | -0.07803 | 0.999532 | | 7.627602 | -0.09682 | 0.933152 | | 6.415667 | -0.00885 | 0.996891 |
| FAM89B | 3.52747 | -0.01413 | 0.999532 | | 4.144312 | 0.125645 | 0.933152 | | 3.522997 | 0.15926 | 0.915533 |
| ZNF93 | 4.362558 | 0.386385 | 0.999532 | | 4.46254 | -0.13583 | 0.933152 | | 4.205012 | -0.45753 | 0.717627 |
| MED24 | 5.803856 | 0.153769 | 0.999532 | | 5.843737 | -0.11111 | 0.933152 | | 5.207221 | -0.24439 | 0.827267 |
| SEMA3D | 2.11926 | 0.112595 | 0.999532 | | 2.14535 | -0.17413 | 0.933152 | | 2.019902 | -0.26837 | 0.890308 |
| RBMXL1 | 3.874313 | 0.217685 | 0.999532 | | 4.407738 | 0.130756 | 0.933152 | | 4.018016 | -0.06839 | 0.971732 |
| TBC1D10 | 2.846635 | 0.035408 | 0.999532 | | 2.930122 | -0.14606 | 0.933152 | | 2.525653 | -0.16426 | 0.923864 |
| SUSD6 | 5.673126 | 0.207478 | 0.999532 | | 5.867224 | 0.110109 | 0.933336 | | 5.446646 | -0.08133 | 0.955834 |
| ARNT2 | 4.49514 | 0.006896 | 0.999532 | | 5.321048 | 0.13306 | 0.933336 | | 4.383139 | 0.133964 | 0.92697 |
| RTF1 | 3.828913 | 0.178785 | 0.999532 | | 3.587918 | -0.14776 | 0.933336 | | 3.25303 | -0.31751 | 0.835289 |
| H2BC7 | 2.287819 | 0.412761 | 0.999532 | | 2.081447 | -0.168 | 0.933336 | | 2.126159 | -0.56498 | 0.691465 |
| VAV1 | 2.0687 | -0.21437 | 0.999532 | | 2.208185 | -0.16591 | 0.933336 | | 1.85874 | 0.060416 | 0.983211 |
| ATP1A3 | 2.581132 | -0.47347 | 0.999532 | | 3.364273 | -0.14832 | 0.933336 | | 2.466506 | 0.350123 | 0.835925 |
| DENND1 | 4.15015 | 0.119976 | 0.999532 | | 4.508619 | 0.120271 | 0.933336 | | 4.01801 | 0.019902 | 0.993618 |
| NCK1 | 3.210658 | -0.33087 | 0.999532 | | 3.761219 | -0.13311 | 0.933336 | | 2.8646 | 0.213755 | 0.892869 |
| SHFL | 8.657386 | 0.124774 | 0.999532 | | 9.542683 | 0.097102 | 0.933411 | | 8.737999 | -0.019 | 0.992286 |
| B9D1 | 4.058399 | -0.11905 | 0.999532 | | 4.755396 | -0.11903 | 0.933411 | | 3.623838 | 0.019986 | 0.994192 |
| FMNL2 | 2.452444 | 0.09657 | 0.999532 | | 2.537352 | -0.15933 | 0.933411 | | 2.200943 | -0.24079 | 0.893259 |
| ZNF625 | 8.404489 | 0.699952 | 0.950261 | | 6.856792 | -0.1175 | 0.933427 | | 7.984861 | -0.81484 | 0.241605 |
| TIMM10B | 4.762378 | 0.01992 | 0.999532 | | 5.024756 | -0.11581 | 0.93346 | | 4.435307 | -0.10719 | 0.946958 |
| COPS7B | 4.071482 | 0.191002 | 0.999532 | | 3.851312 | -0.12801 | 0.933674 | | 3.64006 | -0.30985 | 0.805922 |
| VN1R5 | 2.163225 | 0.423695 | 0.999532 | | 2.255592 | 0.171622 | 0.933674 | | 2.153839 | -0.23103 | 0.89901 |
| NCAPG | 2.489843 | 0.500938 | 0.999532 | | 2.086266 | -0.16917 | 0.933674 | | 2.337895 | -0.65442 | 0.647218 |
| ELK4 | 7.409001 | 0.211116 | 0.999532 | | 7.618205 | -0.10532 | 0.933674 | | 6.894961 | -0.29329 | 0.761699 |
| IMPA1 | 3.661109 | 0.082589 | 0.999532 | | 3.487268 | -0.14111 | 0.933674 | | 3.325729 | -0.20503 | 0.901813 |
| DDO | 2.048848 | 0.456752 | 0.999532 | | 1.846565 | -0.1841 | 0.933762 | | 1.886094 | -0.62277 | 0.686214 |
| NRL | 1.965079 | 0.001846 | 0.999532 | | 2.283217 | -0.17382 | 0.933762 | | 1.898252 | -0.14683 | 0.948097 |
| LIN28A | 2.975278 | 0.231688 | 0.999532 | | 3.219981 | -0.16223 | 0.934167 | | 2.953347 | -0.39473 | 0.791576 |
| FAM118A | 4.443657 | 0.625472 | 0.999532 | | 3.952336 | 0.128059 | 0.934167 | | 4.300837 | -0.48384 | 0.647581 |
| FCRL6 | 2.009227 | -0.01863 | 0.999532 | | 2.211203 | -0.17601 | 0.934366 | | 1.763441 | -0.1377 | 0.953949 |
| KRBA1 | 3.206968 | 0.354774 | 0.999532 | | 3.167459 | -0.14169 | 0.934366 | | 3.029166 | -0.47337 | 0.70363 |
| ZNF585B | 2.910261 | 0.01266 | 0.999532 | | 3.18477 | -0.13935 | 0.934366 | | 2.652784 | -0.13301 | 0.940744 |
| BUD13 | 2.570163 | -0.01805 | 0.999532 | | 3.193817 | 0.142498 | 0.934366 | | 2.679922 | 0.181607 | 0.914687 |
| EIF3K | 12.1523 | 0.085712 | 0.999532 | | 13.73252 | 0.078405 | 0.934366 | | 11.72973 | 0.01523 | 0.992586 |
| CDH1 | 7.120841 | -0.05631 | 0.999532 | | 8.236842 | 0.110215 | 0.934366 | | 6.621089 | 0.191921 | 0.874653 |
| LCE1A | 2.051522 | 0.720635 | 0.999532 | | 1.743365 | -0.1915 | 0.934366 | | 1.965327 | -0.895 | 0.515762 |
| TGM3 | 2.231664 | 1.020623 | 0.999532 | | 1.927482 | 0.177953 | 0.934381 | | 2.223349 | -0.82323 | 0.505689 |
| CCDC30 | 2.068466 | 0.558609 | 0.999532 | | 2.126209 | 0.177038 | 0.934544 | | 2.096112 | -0.36162 | 0.836453 |
| MMAB | 2.016617 | 0.313874 | 0.999532 | | 2.065426 | 0.172459 | 0.934544 | | 2.021137 | -0.12543 | 0.955834 |
| MEX3A | 2.089885 | -0.14157 | 0.999532 | | 2.690883 | 0.159378 | 0.934544 | | 2.122514 | 0.324371 | 0.857522 |
| MTM1 | 2.033323 | -0.49392 | 0.999532 | | 2.493512 | -0.1524 | 0.934544 | | 1.965022 | 0.359548 | 0.839373 |
| PRORP | 3.112301 | 0.12018 | 0.999532 | | 3.73207 | 0.135177 | 0.934544 | | 3.2508 | 0.035804 | 0.988853 |
| FBXW5 | 7.077664 | 0.113398 | 0.999532 | | 8.027046 | 0.103029 | 0.934544 | | 7.19855 | 0.001834 | 0.999385 |
| ZNF142 | 3.078877 | 0.271962 | 0.999532 | | 3.179193 | -0.14237 | 0.934544 | | 2.876674 | -0.38652 | 0.780291 |
| SYP | 4.232981 | 0.166814 | 0.999532 | | 4.377699 | -0.13127 | 0.934544 | | 3.849318 | -0.27389 | 0.836935 |
| KCTD5 | 4.219131 | -0.11525 | 0.999532 | | 4.7977 | -0.12907 | 0.934544 | | 3.869019 | 0.009671 | 0.996891 |
| CCDC18 | 2.342334 | 0.326973 | 0.999532 | | 2.327824 | 0.164134 | 0.934544 | | 2.326874 | -0.14197 | 0.94366 |
| PLEKHA | 4.850795 | 0.257141 | 0.999532 | | 4.66233 | -0.12528 | 0.934544 | | 4.493588 | -0.37224 | 0.767184 |
| DOCK6 | 6.417856 | -0.46567 | 0.999532 | | 8.351096 | 0.101392 | 0.934544 | | 6.230112 | 0.582913 | 0.484303 |
| NRIP2 | 6.873893 | -0.11224 | 0.999532 | | 8.033586 | 0.115766 | 0.934658 | | 6.820567 | 0.254909 | 0.803301 |
| TYROBP | 3.730833 | -0.34885 | 0.999532 | | 4.360926 | -0.13201 | 0.934732 | | 3.417151 | 0.225107 | 0.881837 |
| DHX29 | 3.354954 | 0.220195 | 0.999532 | | 3.796357 | 0.139402 | 0.934732 | | 3.312506 | -0.07037 | 0.972758 |
| POLR3A | 2.80367 | -0.37253 | 0.999532 | | 3.732555 | 0.142672 | 0.934732 | | 3.082241 | 0.540965 | 0.686495 |
| ZNF180 | 2.974509 | 0.237107 | 0.999532 | | 3.465788 | 0.141772 | 0.934732 | | 3.145398 | -0.07683 | 0.971092 |
| ARMC6 | 2.585762 | -0.18509 | 0.999532 | | 3.324023 | 0.139334 | 0.934732 | | 2.752631 | 0.34232 | 0.814884 |
| ZNF438 | 2.1273 | 0.1325 | 0.999532 | | 2.019315 | -0.17843 | 0.934732 | | 1.879235 | -0.30038 | 0.881689 |
| PCYOX1L | 4.866462 | -0.37743 | 0.999532 | | 6.078002 | -0.10839 | 0.934732 | | 4.553278 | 0.288338 | 0.801572 |
| UMODL1 | 2.087316 | 0.036728 | 0.999532 | | 2.064578 | -0.17167 | 0.934732 | | 1.973804 | -0.1907 | 0.93042 |
| SPARCL1 | 3.805832 | 0.107786 | 0.999532 | | 4.32267 | -0.13279 | 0.93478 | | 3.680105 | -0.22533 | 0.87562 |
| SPTBN2 | 2.583117 | 0.066404 | 0.999532 | | 2.76586 | -0.15053 | 0.934885 | | 2.186174 | -0.19879 | 0.908829 |
| TFAP2B | 2.255163 | 0.334964 | 0.999532 | | 2.225891 | -0.16642 | 0.934885 | | 2.082012 | -0.47736 | 0.752299 |
| TECPR1 | 2.245033 | -0.44428 | 0.999532 | | 2.8237 | -0.14741 | 0.934885 | | 2.033064 | 0.317586 | 0.859426 |
| PLK5 | 1.945759 | 0.167962 | 0.999532 | | 1.942821 | -0.17965 | 0.934885 | | 1.886847 | -0.32849 | 0.865065 |
| TFAP2A | 2.741384 | 0.685968 | 0.999532 | | 2.375103 | -0.16258 | 0.934933 | | 2.522629 | -0.81619 | 0.532889 |
| DCUN1D3 | 4.369206 | -0.12806 | 0.999532 | | 5.382688 | 0.110963 | 0.934933 | | 4.359677 | 0.257747 | 0.82512 |
| TPTE | 2.181347 | 0.476449 | 0.999532 | | 1.972868 | -0.17637 | 0.934933 | | 1.925061 | -0.63407 | 0.666566 |
| EID2 | 3.880823 | 0.171692 | 0.999532 | | 4.014145 | -0.13357 | 0.934933 | | 3.458027 | -0.29019 | 0.834593 |
| ZDHHC5 | 4.554704 | -0.0397 | 0.999532 | | 5.331419 | 0.116293 | 0.934933 | | 4.37095 | 0.177274 | 0.892225 |
| GEMIN8 | 2.111174 | 0.618306 | 0.999532 | | 1.96165 | 0.177658 | 0.934933 | | 2.140899 | -0.42244 | 0.791576 |
| SLC35A5 | 3.232372 | 0.076687 | 0.999532 | | 3.751891 | -0.13339 | 0.935002 | | 3.223438 | 0.072692 | 0.971732 |
| ANKRD4 | 5.296501 | -0.14098 | 0.999532 | | 6.325258 | 0.109407 | 0.935002 | | 5.008708 | 0.276098 | 0.79686 |
| CLEC16A | 4.952358 | 0.011004 | 0.999532 | | 5.2689 | -0.11399 | 0.93515 | | 4.299157 | -0.09984 | 0.946876 |
| NCOA3 | 4.396642 | -0.16055 | 0.999532 | | 4.867655 | -0.11594 | 0.93515 | | 3.886014 | 0.058049 | 0.974772 |
| OR7D4 | 2.87976 | 0.311939 | 0.999532 | | 2.65235 | -0.15527 | 0.93518 | | 2.643322 | -0.45073 | 0.758124 |
| PATE2 | 2.168975 | 0.259603 | 0.999532 | | 1.989683 | -0.18343 | 0.93518 | | 1.824678 | -0.41855 | 0.813438 |
| NDE1 | 4.172444 | -0.17021 | 0.999532 | | 5.294139 | 0.110179 | 0.93518 | | 4.26635 | 0.3024 | 0.792539 |
| OPN1MW | 2.781815 | 0.768208 | 0.999532 | | 2.185817 | -0.17053 | 0.935185 | | 2.649151 | -0.92086 | 0.446775 |
| PRIM1 | 2.087299 | 0.474313 | 0.999532 | | 1.923158 | -0.17664 | 0.93526 | | 1.863062 | -0.63108 | 0.668481 |
| PDCD11 | 3.163958 | 0.227194 | 0.999532 | | 2.911862 | -0.141 | 0.935274 | | 2.808785 | -0.35616 | 0.800922 |
| FZD8 | 2.636271 | 0.052344 | 0.999532 | | 3.192066 | 0.13777 | 0.935274 | | 2.696721 | 0.103877 | 0.955834 |
| APOL4 | 2.460777 | 0.199135 | 0.999532 | | 2.517089 | 0.152303 | 0.935348 | | 2.455881 | -0.01784 | 0.996681 |
| IFT80 | 2.398677 | 0.28838 | 0.999532 | | 2.627672 | 0.152966 | 0.935348 | | 2.511149 | -0.11954 | 0.951656 |
| COX4I2 | 2.130647 | -0.50963 | 0.999532 | | 2.694701 | -0.15185 | 0.935348 | | 2.010984 | 0.378631 | 0.828209 |
| XAB2 | 7.121029 | -0.22217 | 0.999532 | | 8.826809 | 0.098444 | 0.935348 | | 6.745114 | 0.344198 | 0.705425 |
| WFS1 | 5.542358 | 0.095612 | 0.999532 | | 6.12213 | 0.106802 | 0.935348 | | 5.334905 | 0.032217 | 0.986208 |
| NRP2 | 3.16262 | 0.156978 | 0.999532 | | 3.240828 | 0.136905 | 0.935348 | | 3.054961 | -0.0162 | 0.996681 |
| ARID5A | 3.681832 | 0.297102 | 0.999532 | | 3.925853 | -0.14382 | 0.935348 | | 3.569419 | -0.37611 | 0.805466 |
| MRTFA | 2.833176 | 0.120189 | 0.999532 | | 3.029471 | -0.15488 | 0.935348 | | 2.584887 | -0.26917 | 0.863134 |
| TMEM18 | 3.394423 | -0.04312 | 0.999532 | | 4.255227 | 0.126245 | 0.935348 | | 3.485719 | 0.187944 | 0.89689 |
| KLHL10 | 1.822988 | 0.485895 | 0.999532 | | 1.719884 | 0.182956 | 0.935348 | | 1.799258 | -0.28708 | 0.883785 |
| SMCO3 | 3.367322 | 0.299139 | 0.999532 | | 3.455449 | 0.133404 | 0.935551 | | 3.230428 | -0.14811 | 0.924131 |
| MUCL1 | 2.615184 | 0.565072 | 0.999532 | | 2.213901 | -0.16724 | 0.935551 | | 2.440204 | -0.71497 | 0.600081 |
| RMI2 | 2.101705 | 0.096633 | 0.999532 | | 2.041956 | -0.17146 | 0.935614 | | 1.818197 | -0.25029 | 0.897958 |
| MCAT | 3.43646 | -0.2055 | 0.999532 | | 4.011411 | -0.12951 | 0.935683 | | 3.075884 | 0.097333 | 0.958239 |
| DYNLT2B | 3.536727 | 0.235797 | 0.999532 | | 4.012121 | 0.123987 | 0.935683 | | 3.658257 | -0.08258 | 0.963492 |
| PPM1L | 2.545369 | 0.130751 | 0.999532 | | 2.677752 | -0.15304 | 0.935683 | | 2.440816 | -0.25491 | 0.887703 |
| ZNF223 | 2.601345 | 0.177875 | 0.999532 | | 2.574549 | -0.15637 | 0.935683 | | 2.279758 | -0.3141 | 0.846284 |
| ZCCHC17 | 2.910409 | 0.194511 | 0.999532 | | 3.186632 | 0.1421 | 0.935683 | | 2.987541 | -0.03809 | 0.988853 |
| TBC1D8B | 2.390635 | 0.565348 | 0.999532 | | 2.298418 | 0.165317 | 0.935683 | | 2.480451 | -0.38645 | 0.79668 |
| TLE4 | 4.234914 | -0.18367 | 0.999532 | | 4.928344 | -0.11792 | 0.935907 | | 4.10317 | 0.08547 | 0.958889 |
| SNTN | 1.965722 | 0.105501 | 0.999532 | | 2.03698 | -0.17431 | 0.935907 | | 1.850371 | -0.26635 | 0.893828 |
| MSANTD | 2.597258 | 0.586581 | 0.999532 | | 2.407789 | 0.160887 | 0.935907 | | 2.560425 | -0.40854 | 0.792538 |
| AHSA1 | 3.846929 | 0.126173 | 0.999532 | | 4.470101 | 0.124795 | 0.935991 | | 3.650695 | 0.028391 | 0.991273 |
| LSM7 | 4.955469 | -0.07949 | 0.999532 | | 5.761234 | 0.10789 | 0.935991 | | 4.775979 | 0.198233 | 0.87813 |
| SGSM2 | 4.672594 | 0.305715 | 0.999532 | | 5.009396 | 0.115981 | 0.936217 | | 4.509521 | -0.18301 | 0.886495 |
| ABCF1 | 2.757041 | 0.637613 | 0.999532 | | 2.634892 | 0.151455 | 0.936217 | | 2.737058 | -0.46938 | 0.712621 |
| ALS2CL | 58.3496 | 0.449685 | 0.999532 | | 53.68329 | -0.10229 | 0.936217 | | 53.82806 | -0.52406 | 0.398915 |
| ZNF808 | 3.038601 | 0.24966 | 0.999532 | | 3.507959 | 0.141972 | 0.936217 | | 3.14825 | -0.08523 | 0.965876 |
| MTPAP | 5.175536 | 0.192086 | 0.999532 | | 5.743315 | 0.116632 | 0.936217 | | 5.380464 | -0.06077 | 0.972305 |
| RSC1A1 | 2.092535 | 0.701396 | 0.999532 | | 1.96795 | 0.172823 | 0.936217 | | 2.082422 | -0.5089 | 0.730921 |
| GSTZ1 | 3.073229 | 0.091958 | 0.999532 | | 3.236848 | -0.14592 | 0.936217 | | 2.779288 | -0.21654 | 0.897096 |
| SIVA1 | 4.067727 | -0.10446 | 0.999532 | | 5.165256 | 0.113736 | 0.936217 | | 4.145505 | 0.233749 | 0.852074 |
| CCDC7 | 2.27703 | 0.031515 | 0.999532 | | 2.312896 | -0.16249 | 0.936217 | | 2.003864 | -0.17885 | 0.929087 |
| ALKBH8 | 2.703001 | 0.020278 | 0.999532 | | 2.90102 | -0.14904 | 0.936266 | | 2.531596 | -0.15342 | 0.939549 |
| PLEKHG | 5.420962 | -0.31316 | 0.999532 | | 7.248459 | 0.105313 | 0.936266 | | 5.539987 | 0.434506 | 0.647581 |
| DCAF11 | 3.945438 | 0.289894 | 0.999532 | | 4.180175 | 0.121658 | 0.936266 | | 4.086582 | -0.15162 | 0.914692 |
| IL37 | 1.916648 | -0.11819 | 0.999532 | | 2.124554 | -0.17471 | 0.936266 | | 1.805079 | -0.04003 | 0.991273 |
| CHD7 | 4.740631 | 0.253792 | 0.999532 | | 4.840538 | 0.112253 | 0.936432 | | 4.597855 | -0.12943 | 0.926957 |
| PHF21B | 4.233133 | 0.489797 | 0.999532 | | 3.516374 | -0.13103 | 0.936432 | | 3.982183 | -0.59755 | 0.598934 |
| SSU72 | 7.604673 | -0.09229 | 0.999532 | | 8.616925 | -0.09035 | 0.936432 | | 6.893588 | 0.023403 | 0.989766 |
| TCAIM | 2.204017 | -0.13876 | 0.999532 | | 2.641204 | 0.147676 | 0.936432 | | 2.17709 | 0.301461 | 0.861815 |
| CLK3 | 4.500926 | -0.13721 | 0.999532 | | 5.518961 | 0.115029 | 0.936432 | | 4.422797 | 0.275356 | 0.821384 |
| HMX1 | 3.920857 | 0.279904 | 0.999532 | | 4.320096 | 0.122723 | 0.936432 | | 3.924183 | -0.14465 | 0.918694 |
| ZNF480 | 2.941286 | 0.161004 | 0.999532 | | 2.996051 | -0.14119 | 0.936432 | | 2.652978 | -0.27863 | 0.861545 |
| UBALD2 | 4.686703 | -0.10022 | 0.999532 | | 5.664552 | 0.111296 | 0.936432 | | 4.772578 | 0.229193 | 0.853729 |
| GLT1D1 | 2.519359 | -0.0317 | 0.999532 | | 2.782801 | -0.15096 | 0.936432 | | 2.406999 | -0.09896 | 0.965619 |
| SLC35C1 | 2.283049 | 0.242004 | 0.999532 | | 2.342234 | -0.16268 | 0.936432 | | 2.199995 | -0.37854 | 0.824012 |
| TAS2R19 | 2.401051 | 0.431826 | 0.999532 | | 2.119294 | -0.16783 | 0.936432 | | 2.269146 | -0.58202 | 0.686214 |
| TMEM201 | 2.808084 | 0.498046 | 0.999532 | | 2.679777 | 0.154134 | 0.936432 | | 2.784235 | -0.32274 | 0.829716 |
| SARS2 | 3.410757 | 0.438324 | 0.999532 | | 3.159693 | 0.137146 | 0.936432 | | 3.320449 | -0.28025 | 0.849682 |
| SYTL5 | 2.965857 | 0.295527 | 0.999532 | | 2.794342 | -0.151 | 0.936432 | | 2.696297 | -0.42858 | 0.763873 |
| ADGRD2 | 3.391081 | 0.07012 | 0.999532 | | 3.724973 | -0.13014 | 0.936432 | | 3.105837 | -0.17889 | 0.905952 |
| FAM83C | 5.990437 | 0.565863 | 0.999532 | | 5.133781 | -0.11775 | 0.936432 | | 5.749287 | -0.66273 | 0.414073 |
| TXLNA | 4.827153 | 0.156215 | 0.999532 | | 5.496406 | 0.11086 | 0.936432 | | 4.834897 | -0.02456 | 0.991273 |
| CASP3 | 2.9527 | 0.500912 | 0.999532 | | 2.944905 | 0.146183 | 0.936432 | | 3.018982 | -0.33521 | 0.814402 |
| XPO7 | 4.753976 | -0.76422 | 0.999532 | | 6.479929 | -0.09946 | 0.936432 | | 4.328817 | 0.679506 | 0.460198 |
| RASGEF1 | 2.385134 | 0.393358 | 0.999532 | | 2.209938 | -0.17178 | 0.936432 | | 2.219446 | -0.5512 | 0.717627 |
| KCNMB1 | 2.701921 | 0.447189 | 0.999532 | | 2.265629 | -0.15432 | 0.936432 | | 2.539787 | -0.58371 | 0.663009 |
| H2BC21 | 4.247683 | 0.414078 | 0.999532 | | 3.7841 | -0.13276 | 0.936432 | | 4.075675 | -0.53931 | 0.661666 |
| RBL2 | 5.12039 | -0.20357 | 0.999532 | | 6.589112 | 0.098134 | 0.936432 | | 5.258012 | 0.320307 | 0.739496 |
| UBE2T | 1.830708 | 0.289677 | 0.999532 | | 1.843197 | 0.180568 | 0.936432 | | 1.860918 | -0.09185 | 0.973356 |
| BCL3 | 6.147049 | -0.34373 | 0.999532 | | 7.549255 | -0.10597 | 0.936432 | | 5.792485 | 0.253397 | 0.824012 |
| HOXB13 | 2.119162 | 0.091566 | 0.999532 | | 2.189139 | -0.17372 | 0.936432 | | 2.070853 | -0.25549 | 0.901033 |
| NUP62 | 3.734494 | -0.24806 | 0.999532 | | 4.668223 | 0.117227 | 0.936432 | | 3.83236 | 0.386492 | 0.75038 |
| MAP3K8 | 3.982521 | 0.15143 | 0.999532 | | 4.180452 | -0.12379 | 0.936432 | | 3.736541 | -0.25589 | 0.846284 |
| OR2C1 | 2.062686 | 0.065765 | 0.999532 | | 2.093114 | -0.17321 | 0.936432 | | 1.858458 | -0.22081 | 0.914929 |
| MUC21 | 3.307402 | 0.737172 | 0.999532 | | 2.653832 | -0.14886 | 0.93648 | | 3.067146 | -0.8689 | 0.397004 |
| KRTAP20 | 2.382775 | -0.47241 | 0.999532 | | 2.86572 | -0.16266 | 0.93648 | | 2.2698 | 0.302577 | 0.88476 |
| RHOF | 2.937542 | 0.064549 | 0.999532 | | 3.018451 | -0.14351 | 0.93648 | | 2.686823 | -0.19113 | 0.910105 |
| NAP1L3 | 2.397185 | -0.49218 | 0.999532 | | 3.173073 | 0.137058 | 0.93648 | | 2.336128 | 0.643823 | 0.644433 |
| TBL1X | 5.386562 | -0.04428 | 0.999532 | | 5.948209 | -0.10301 | 0.93648 | | 4.965805 | -0.04098 | 0.979967 |
| ADA | 2.204196 | -0.53301 | 0.999532 | | 2.78592 | -0.15509 | 0.936731 | | 2.017874 | 0.399304 | 0.830822 |
| SUPT6H | 7.302078 | -0.21993 | 0.999532 | | 8.251425 | -0.09245 | 0.936784 | | 6.416656 | 0.140541 | 0.90424 |
| RAB26 | 3.961066 | 0.506682 | 0.999532 | | 3.958308 | 0.150164 | 0.936784 | | 4.083035 | -0.34036 | 0.837256 |
| COL5A2 | 5.723078 | -0.12429 | 0.999532 | | 6.865632 | 0.103875 | 0.936784 | | 5.763438 | 0.247184 | 0.827267 |
| HLA-DQA | 6.146907 | -0.79045 | 0.999532 | | 9.539245 | 0.264108 | 0.93702 | | 6.508992 | 1.088915 | 0.632923 |
| NFATC1 | 4.067509 | -0.16346 | 0.999532 | | 4.887511 | 0.123959 | 0.937054 | | 3.941032 | 0.317288 | 0.796434 |
| GREB1 | 2.27867 | 0.284352 | 0.999532 | | 2.502355 | 0.169762 | 0.937054 | | 2.267607 | -0.08934 | 0.972221 |
| KIF27 | 2.013885 | 0.412277 | 0.999532 | | 2.000628 | 0.166305 | 0.937054 | | 2.008455 | -0.22971 | 0.902036 |
| MANEA | 4.449553 | 0.390021 | 0.999532 | | 4.44434 | 0.115107 | 0.937054 | | 4.31696 | -0.25711 | 0.8256 |
| CUEDC1 | 4.954726 | -0.36066 | 0.999532 | | 5.896346 | -0.10354 | 0.937054 | | 4.678449 | 0.271566 | 0.833604 |
| GOLGA6 | 2.374894 | 0.168404 | 0.999532 | | 2.388024 | -0.15646 | 0.937054 | | 2.152922 | -0.30641 | 0.859898 |
| PIM3 | 8.280619 | -0.33181 | 0.999532 | | 9.490077 | -0.08244 | 0.937054 | | 7.411897 | 0.257754 | 0.779082 |
| TCEANC2 | 2.816386 | 0.064021 | 0.999532 | | 3.18431 | 0.142406 | 0.937054 | | 2.91722 | 0.101604 | 0.963359 |
| SEC14L5 | 1.798999 | 0.704428 | 0.999532 | | 1.552595 | 0.190734 | 0.937054 | | 1.796526 | -0.49661 | 0.774747 |
| MSANTD | 2.271889 | 0.238427 | 0.999532 | | 2.459582 | 0.152089 | 0.937054 | | 2.211113 | -0.06809 | 0.977473 |
| FHIP2A | 3.960587 | 0.212833 | 0.999532 | | 4.553215 | 0.113776 | 0.937054 | | 4.05578 | -0.07723 | 0.963359 |
| PHYKPL | 3.251059 | -0.08993 | 0.999532 | | 3.553234 | -0.13292 | 0.937054 | | 3.050372 | -0.02816 | 0.992286 |
| CBL | 6.408015 | 0.005867 | 0.999532 | | 7.412265 | 0.096823 | 0.937054 | | 6.331957 | 0.113006 | 0.920259 |
| CDK5RAP | 2.661108 | -0.08927 | 0.999532 | | 2.755038 | -0.14089 | 0.937054 | | 2.367973 | -0.03903 | 0.989148 |
| COA6 | 3.252625 | 0.52031 | 0.999532 | | 2.834087 | -0.14234 | 0.937054 | | 3.020438 | -0.64127 | 0.598934 |
| CRYGN | 2.601657 | 0.411142 | 0.999532 | | 2.545738 | -0.1603 | 0.937054 | | 2.558153 | -0.54439 | 0.713778 |
| WDR25 | 2.30159 | 0.197428 | 0.999532 | | 2.158661 | -0.16463 | 0.937054 | | 1.995015 | -0.34626 | 0.845164 |
| WASF3 | 2.790862 | 0.332503 | 0.999532 | | 2.676613 | -0.15276 | 0.937054 | | 2.5282 | -0.46864 | 0.744727 |
| ASPG | 4.437244 | 0.117697 | 0.999532 | | 4.531114 | -0.11528 | 0.937054 | | 4.106225 | -0.21199 | 0.869579 |
| NRDE2 | 3.435684 | 0.569719 | 0.999532 | | 3.101173 | 0.13398 | 0.937054 | | 3.422976 | -0.41906 | 0.729463 |
| IFIT1B | 2.151476 | 0.510987 | 0.999532 | | 2.230569 | 0.165263 | 0.937054 | | 2.309686 | -0.32708 | 0.849686 |
| RPRD1B | 8.417104 | -0.16878 | 0.999532 | | 10.13915 | 0.083721 | 0.937054 | | 8.042247 | 0.26192 | 0.774747 |
| SNU13 | 16.82977 | -0.10187 | 0.999532 | | 18.80211 | -0.07084 | 0.937054 | | 15.35682 | 0.046632 | 0.968434 |
| CLDN19 | 2.021766 | -0.02755 | 0.999532 | | 2.008682 | -0.16442 | 0.937054 | | 1.7846 | -0.12326 | 0.958889 |
| SRA1 | 3.925948 | -0.01981 | 0.999532 | | 4.713641 | 0.11294 | 0.937054 | | 4.011772 | 0.151825 | 0.912095 |
| FAM8A1 | 2.834642 | -0.44179 | 0.999532 | | 3.470553 | 0.134268 | 0.937054 | | 2.744359 | 0.574158 | 0.676565 |
| RXFP1 | 3.702429 | 0.412308 | 0.999532 | | 3.497815 | 0.128013 | 0.937054 | | 3.726927 | -0.2553 | 0.850915 |
| CPNE4 | 2.121356 | 0.150429 | 0.999532 | | 2.234917 | -0.16283 | 0.937054 | | 2.015382 | -0.29117 | 0.876781 |
| GPR173 | 2.264559 | 0.440131 | 0.999532 | | 2.006805 | -0.16897 | 0.937054 | | 2.112251 | -0.59091 | 0.678411 |
| MARS1 | 3.977252 | 0.238814 | 0.999532 | | 3.818689 | -0.13753 | 0.937054 | | 3.637843 | -0.35174 | 0.791576 |
| GPR160 | 3.160284 | 0.08402 | 0.999532 | | 3.645253 | 0.13739 | 0.937054 | | 3.091961 | 0.082294 | 0.969614 |
| RNF214 | 3.300446 | 0.464613 | 0.999532 | | 3.145528 | 0.146338 | 0.937054 | | 3.129886 | -0.28544 | 0.837927 |
| CRYBB1 | 1.937126 | 0.34976 | 0.999532 | | 2.243748 | 0.163812 | 0.937054 | | 2.122529 | -0.15939 | 0.939913 |
| ARL17B | 9.501197 | 0.403561 | 0.999532 | | 9.037754 | -0.10198 | 0.937084 | | 8.583397 | -0.46784 | 0.515762 |
| SELPLG | 2.485509 | 0.173777 | 0.999532 | | 2.852706 | 0.14213 | 0.937159 | | 2.593604 | -0.0117 | 0.996891 |
| AP5Z1 | 3.296073 | 0.298263 | 0.999532 | | 3.366125 | -0.13973 | 0.937159 | | 3.069804 | -0.43053 | 0.724409 |
| LINGO1 | 4.090628 | -0.04975 | 0.999532 | | 4.425704 | -0.1194 | 0.937352 | | 3.700351 | -0.05366 | 0.978229 |
| CAPN3 | 3.171001 | 0.062557 | 0.999532 | | 3.476731 | -0.12877 | 0.937352 | | 2.902315 | -0.17023 | 0.914687 |
| CAPRIN2 | 3.692253 | 0.171114 | 0.999532 | | 3.638971 | -0.1264 | 0.937352 | | 3.381557 | -0.27934 | 0.836012 |
| UXT | 2.466385 | -0.50971 | 0.999532 | | 3.038367 | -0.13609 | 0.937352 | | 2.150381 | 0.387052 | 0.806394 |
| MRPS11 | 3.536439 | -0.14508 | 0.999532 | | 4.551332 | 0.126188 | 0.937488 | | 3.738648 | 0.283029 | 0.836453 |
| BRCA1 | 2.534026 | 0.345834 | 0.999532 | | 2.286375 | -0.16129 | 0.937543 | | 2.381605 | -0.49359 | 0.749222 |
| PDE1B | 2.383277 | 0.236407 | 0.999532 | | 2.539851 | 0.152453 | 0.937571 | | 2.406087 | -0.06862 | 0.9774 |
| NXF3 | 2.271725 | -0.22701 | 0.999532 | | 2.474878 | -0.1507 | 0.937571 | | 2.077193 | 0.092578 | 0.971458 |
| MS4A18 | 1.927478 | 0.186297 | 0.999532 | | 1.880612 | -0.16908 | 0.937571 | | 1.856083 | -0.33591 | 0.859426 |
| YPEL2 | 2.833493 | -0.30919 | 0.999532 | | 3.765294 | 0.127053 | 0.937571 | | 2.906027 | 0.452547 | 0.722703 |
| SPRY2 | 2.355286 | 0.077108 | 0.999532 | | 2.578611 | 0.153074 | 0.937571 | | 2.323794 | 0.094758 | 0.967145 |
| C9orf131 | 1.93705 | 0.281002 | 0.999532 | | 2.032213 | -0.17905 | 0.937601 | | 1.96489 | -0.43571 | 0.810208 |
| GTF2E1 | 3.018245 | 0.88912 | 0.999532 | | 2.447793 | 0.159042 | 0.937603 | | 2.981705 | -0.70934 | 0.570037 |
| RBMX | 11.7846 | 0.065324 | 0.999532 | | 12.06721 | -0.09233 | 0.937603 | | 10.70044 | -0.14033 | 0.886495 |
| TSPAN12 | 3.086789 | -0.0781 | 0.999532 | | 3.236076 | -0.13206 | 0.937603 | | 2.777115 | -0.04825 | 0.98556 |
| DSCAM | 2.640005 | 0.141718 | 0.999532 | | 2.707261 | -0.16365 | 0.937603 | | 2.253893 | -0.27178 | 0.881316 |
| KIAA075 | 6.312534 | 0.325142 | 0.999532 | | 6.528225 | 0.09659 | 0.937603 | | 6.171918 | -0.21011 | 0.842686 |
| HRH2 | 2.449897 | 0.296184 | 0.999532 | | 2.467232 | -0.16059 | 0.937603 | | 2.327675 | -0.4339 | 0.786555 |
| BCAT1 | 2.230792 | -0.22162 | 0.999532 | | 2.612727 | -0.16314 | 0.937603 | | 2.141838 | 0.074343 | 0.978229 |
| MIPEP | 1.809759 | -0.26751 | 0.999532 | | 2.001968 | -0.16343 | 0.937603 | | 1.773482 | 0.121293 | 0.962716 |
| SPIN1 | 3.727045 | -0.24041 | 0.999532 | | 4.811856 | 0.115986 | 0.937603 | | 3.760622 | 0.379226 | 0.760473 |
| TINF2 | 4.845857 | -0.12144 | 0.999532 | | 5.82824 | 0.103852 | 0.937603 | | 4.962768 | 0.241716 | 0.837256 |
| TMEM87A | 5.202338 | 0.1606 | 0.999532 | | 5.089606 | -0.10737 | 0.937603 | | 4.71338 | -0.2487 | 0.827429 |
| DMTN | 3.702953 | -0.12126 | 0.999532 | | 4.408521 | 0.126514 | 0.937603 | | 3.652902 | 0.269287 | 0.851503 |
| SETD2 | 3.654237 | 0.471545 | 0.999532 | | 3.639723 | 0.126719 | 0.937603 | | 3.65819 | -0.32676 | 0.791576 |
| NR1H2 | 6.45267 | -0.16418 | 0.999532 | | 7.110023 | -0.09885 | 0.937603 | | 5.811041 | 0.084898 | 0.952656 |
| PGGHG | 5.988953 | 0.328344 | 0.999532 | | 5.933955 | -0.10108 | 0.937603 | | 5.719283 | -0.40504 | 0.666555 |
| LENEP | 1.819838 | -0.07421 | 0.999532 | | 2.108648 | -0.17088 | 0.937603 | | 1.811675 | -0.07407 | 0.978927 |
| PIK3R6 | 1.937705 | -0.08994 | 0.999532 | | 2.182952 | -0.1684 | 0.937603 | | 1.828746 | -0.06085 | 0.985528 |
| FNTB | 4.369724 | 0.409957 | 0.999532 | | 4.161597 | 0.129031 | 0.937603 | | 4.213818 | -0.26263 | 0.856173 |
| CDADC1 | 2.507567 | 0.424161 | 0.999532 | | 2.595601 | 0.149929 | 0.937603 | | 2.463992 | -0.25384 | 0.881379 |
| LYSMD1 | 2.209503 | -0.49004 | 0.999532 | | 2.790301 | 0.14307 | 0.937603 | | 2.177989 | 0.651023 | 0.652925 |
| UNC119 | 3.800088 | 0.093189 | 0.999532 | | 4.023337 | -0.12971 | 0.937603 | | 3.55684 | -0.20549 | 0.88885 |
| PLBD2 | 3.175633 | -0.31721 | 0.999532 | | 4.221036 | 0.116371 | 0.937603 | | 3.230786 | 0.452608 | 0.703463 |
| ZNF507 | 3.025217 | 0.352337 | 0.999532 | | 2.946809 | -0.14256 | 0.937603 | | 2.783367 | -0.47403 | 0.720106 |
| C7orf33 | 1.985572 | 0.398622 | 0.999532 | | 1.974601 | 0.165341 | 0.937603 | | 1.93036 | -0.216 | 0.910105 |
| DNAJC10 | 3.858389 | 0.06336 | 0.999532 | | 4.430731 | 0.114472 | 0.937603 | | 3.791706 | 0.066024 | 0.972221 |
| PTPRC | 3.099124 | -0.46119 | 0.999532 | | 3.839181 | -0.14777 | 0.937603 | | 2.935531 | 0.316154 | 0.83994 |
| OR4K15 | 2.339624 | -0.00373 | 0.999532 | | 2.498874 | -0.15922 | 0.937603 | | 2.257707 | -0.14275 | 0.948099 |
| OXCT2 | 2.470377 | 0.21135 | 0.999532 | | 2.832034 | 0.160801 | 0.937603 | | 2.600733 | -0.02788 | 0.993618 |
| SMURF1 | 4.245685 | 0.121114 | 0.999532 | | 4.026441 | -0.12016 | 0.937603 | | 3.854255 | -0.25831 | 0.858767 |
| WDR12 | 3.821897 | -0.05639 | 0.999532 | | 4.306717 | -0.12393 | 0.937603 | | 3.51636 | -0.04422 | 0.984624 |
| RRP1B | 3.635251 | -0.0608 | 0.999532 | | 3.84003 | -0.12506 | 0.937603 | | 3.310926 | -0.05463 | 0.978927 |
| OR2J3 | 2.671002 | 0.798597 | 0.999532 | | 2.399908 | 0.163802 | 0.937603 | | 2.781941 | -0.61988 | 0.647581 |
| PKD1 | 12.53421 | 0.191012 | 0.999532 | | 13.38312 | 0.076676 | 0.937603 | | 12.35055 | -0.09218 | 0.92403 |
| PPM1F | 4.4904 | -0.40245 | 0.999532 | | 5.630772 | 0.106231 | 0.937603 | | 4.330955 | 0.519776 | 0.614824 |
| PROB1 | 2.34851 | 0.512469 | 0.999532 | | 2.002127 | -0.17022 | 0.937603 | | 2.220377 | -0.66488 | 0.660958 |
| DNM1 | 4.517234 | 0.102518 | 0.999532 | | 4.666692 | 0.117807 | 0.937603 | | 4.259226 | 0.02997 | 0.990309 |
| PTGS1 | 2.661093 | -0.00087 | 0.999532 | | 3.208744 | 0.133654 | 0.937603 | | 2.685087 | 0.153856 | 0.928522 |
| NRN1 | 3.122635 | 0.032586 | 0.999532 | | 3.712582 | 0.135926 | 0.937603 | | 3.234617 | 0.128402 | 0.946512 |
| ZNF770 | 3.145417 | 0.477553 | 0.999532 | | 3.290696 | 0.135854 | 0.937603 | | 3.241894 | -0.3095 | 0.835554 |
| PHF14 | 3.208745 | -0.28749 | 0.999532 | | 4.17493 | 0.116331 | 0.937603 | | 3.219198 | 0.421972 | 0.724386 |
| TUT7 | 3.30786 | -0.07972 | 0.999532 | | 3.75799 | -0.12745 | 0.937603 | | 3.081674 | -0.03008 | 0.991159 |
| LATS1 | 5.852159 | -0.01679 | 0.999532 | | 6.758334 | 0.103129 | 0.937603 | | 5.565331 | 0.145217 | 0.908815 |
| ALAS1 | 3.62858 | -0.03391 | 0.999532 | | 4.066273 | 0.11828 | 0.937603 | | 3.596295 | 0.17084 | 0.908274 |
| CXorf38 | 3.846822 | -0.2266 | 0.999532 | | 4.335047 | -0.11731 | 0.937603 | | 3.561886 | 0.125702 | 0.939913 |
| BRME1 | 1.745985 | 0.267129 | 0.999532 | | 1.87082 | 0.177716 | 0.937603 | | 1.734282 | -0.07065 | 0.979432 |
| CERCAM | 2.766191 | -0.15166 | 0.999532 | | 3.397298 | 0.134902 | 0.937603 | | 2.751329 | 0.306543 | 0.849648 |
| YIPF6 | 5.105008 | -0.3629 | 0.999532 | | 6.932036 | 0.097948 | 0.937603 | | 5.207145 | 0.478886 | 0.599857 |
| MAS1 | 2.420851 | 0.203657 | 0.999532 | | 2.339554 | -0.16372 | 0.937603 | | 2.124026 | -0.34323 | 0.843043 |
| USP17L7 | 3.34087 | 0.326674 | 0.999532 | | 3.449291 | -0.14234 | 0.937603 | | 3.370702 | -0.44604 | 0.745908 |
| EPN3 | 2.290912 | 0.185643 | 0.999532 | | 2.208355 | -0.15994 | 0.937603 | | 2.19581 | -0.33067 | 0.854633 |
| B4GALT3 | 4.213059 | 0.004198 | 0.999532 | | 4.332263 | -0.11473 | 0.937603 | | 3.646303 | -0.10701 | 0.946765 |
| FLOT1 | 8.704135 | -0.3719 | 0.999532 | | 10.79072 | -0.10044 | 0.937603 | | 7.874958 | 0.299264 | 0.783005 |
| FBXO44 | 3.418662 | 0.180329 | 0.999532 | | 3.679778 | -0.12757 | 0.937603 | | 3.145829 | -0.28011 | 0.843463 |
| C1QL3 | 2.333212 | 0.253337 | 0.999532 | | 2.515978 | -0.16445 | 0.937603 | | 2.15086 | -0.38546 | 0.824012 |
| CLPSL1 | 2.520791 | 0.626767 | 0.999532 | | 2.088675 | -0.17192 | 0.937603 | | 2.487369 | -0.77553 | 0.626811 |
| POU6F1 | 4.218119 | 0.34006 | 0.999532 | | 4.190242 | 0.118912 | 0.937603 | | 4.144416 | -0.20206 | 0.878742 |
| CHAC2 | 2.65862 | 0.050549 | 0.999532 | | 2.755075 | -0.15005 | 0.937603 | | 2.567819 | -0.18639 | 0.918942 |
| PIP4P2 | 5.055148 | 0.023629 | 0.999532 | | 6.144451 | 0.102291 | 0.937603 | | 5.212716 | 0.094603 | 0.945129 |
| UBE2M | 3.81226 | -0.30506 | 0.999532 | | 4.346468 | -0.11873 | 0.937603 | | 3.303728 | 0.199628 | 0.893853 |
| BTF3L4 | 3.245986 | 0.287764 | 0.999532 | | 3.583434 | 0.134483 | 0.937603 | | 3.303927 | -0.13516 | 0.937656 |
| BAIAP3 | 3.367335 | 0.11215 | 0.999532 | | 3.610569 | -0.13181 | 0.937603 | | 2.967219 | -0.22468 | 0.883792 |
| HMG20B | 6.335973 | -0.00754 | 0.999532 | | 6.859339 | -0.09922 | 0.937603 | | 5.585497 | -0.06815 | 0.965619 |
| PAQR8 | 3.060179 | 0.14622 | 0.999532 | | 3.530968 | 0.128567 | 0.937603 | | 2.968585 | 0.002679 | 0.999385 |
| WDR36 | 2.145078 | -0.03751 | 0.999532 | | 2.523631 | 0.147723 | 0.937603 | | 2.101894 | 0.203068 | 0.910851 |
| TAZ | 3.278842 | 0.249134 | 0.999532 | | 3.301239 | -0.1332 | 0.937603 | | 2.990084 | -0.36379 | 0.784053 |
| CD33 | 2.156061 | -0.1236 | 0.999532 | | 2.723857 | 0.145108 | 0.937603 | | 2.278057 | 0.28779 | 0.868465 |
| CLEC2D | 1.960951 | 0.385119 | 0.999532 | | 1.869075 | 0.165709 | 0.937603 | | 1.960458 | -0.2053 | 0.918194 |
| POLA2 | 2.823263 | 0.603676 | 0.999532 | | 2.648865 | 0.155543 | 0.937603 | | 2.919071 | -0.43635 | 0.741408 |
| AGTR1 | 5.03063 | -0.01906 | 0.999532 | | 5.747174 | 0.118979 | 0.937603 | | 5.055028 | 0.184067 | 0.878619 |
| ITGA11 | 4.580528 | -0.45155 | 0.999532 | | 5.755375 | -0.10667 | 0.937603 | | 4.04724 | 0.361874 | 0.765574 |
| KCNJ15 | 3.395598 | 0.181892 | 0.999532 | | 3.199634 | -0.14068 | 0.937603 | | 2.843617 | -0.31832 | 0.836343 |
| CEP78 | 2.116446 | -0.30168 | 0.999532 | | 2.692816 | -0.15799 | 0.937603 | | 2.12098 | 0.169811 | 0.939913 |
| DRC3 | 2.28044 | 0.55893 | 0.999532 | | 2.195342 | 0.160573 | 0.937603 | | 2.408742 | -0.37799 | 0.818918 |
| HSD3B2 | 2.812324 | 0.465663 | 0.999532 | | 2.84675 | -0.15028 | 0.93781 | | 2.807724 | -0.5944 | 0.652925 |
| RORC | 3.90347 | 0.430455 | 0.999532 | | 4.05933 | 0.11904 | 0.93781 | | 3.771181 | -0.28968 | 0.815293 |
| ZNF675 | 1.941465 | 0.467809 | 0.999532 | | 1.710679 | -0.17171 | 0.93781 | | 1.8141 | -0.62179 | 0.686214 |
| EFTUD2 | 3.345325 | -0.09306 | 0.999532 | | 3.621516 | -0.12265 | 0.937869 | | 3.121877 | -0.01447 | 0.996681 |
| MESP1 | 2.00842 | -0.25619 | 0.999532 | | 2.414321 | 0.158583 | 0.937869 | | 2.012079 | 0.42963 | 0.811123 |
| MAP3K9 | 2.055643 | 0.350338 | 0.999532 | | 2.235528 | 0.160652 | 0.937869 | | 2.167772 | -0.17079 | 0.93222 |
| PHLPP2 | 1.868552 | 0.352626 | 0.999532 | | 1.760651 | -0.1737 | 0.937869 | | 1.730325 | -0.50832 | 0.760506 |
| IFI35 | 4.604643 | 0.230525 | 0.999532 | | 4.660462 | 0.114121 | 0.937869 | | 4.494133 | -0.09829 | 0.950616 |
| TSC1 | 2.547337 | 0.44593 | 0.999532 | | 2.314927 | 0.150843 | 0.937869 | | 2.492347 | -0.2861 | 0.865363 |
| PFN2 | 4.889445 | -0.16242 | 0.999532 | | 5.510347 | -0.10479 | 0.937869 | | 4.517934 | 0.077996 | 0.960798 |
| OR2C3 | 2.163341 | 0.368489 | 0.999532 | | 2.122775 | -0.17433 | 0.937869 | | 2.144859 | -0.53115 | 0.740445 |
| POLL | 3.833867 | -0.0197 | 0.999532 | | 4.077961 | -0.1209 | 0.937869 | | 3.540081 | -0.07684 | 0.966652 |
| GATA5 | 2.869332 | 0.375036 | 0.999532 | | 2.717572 | 0.146727 | 0.937869 | | 2.752821 | -0.2219 | 0.901393 |
| FAM20B | 5.009059 | -0.1679 | 0.999532 | | 5.741394 | -0.10246 | 0.937869 | | 4.538047 | 0.083372 | 0.955834 |
| MMP25 | 1.880273 | -0.2497 | 0.999532 | | 2.157455 | -0.15691 | 0.937869 | | 1.781282 | 0.106746 | 0.96703 |
| FLCN | 3.112676 | 0.193827 | 0.999532 | | 3.278566 | 0.133647 | 0.938015 | | 2.935573 | -0.0395 | 0.987274 |
| KLF11 | 4.349091 | -0.18677 | 0.999532 | | 4.839357 | -0.10978 | 0.938015 | | 3.88524 | 0.096905 | 0.950616 |
| NLRC3 | 1.99721 | 0.304083 | 0.999532 | | 1.877306 | -0.16683 | 0.938039 | | 1.807193 | -0.45516 | 0.790999 |
| RIC8A | 4.618174 | -0.11596 | 0.999532 | | 4.821078 | -0.11205 | 0.93815 | | 4.168143 | 0.012382 | 0.996681 |
| SLC9A3 | 2.222269 | 0.491838 | 0.999532 | | 2.435446 | 0.168147 | 0.938364 | | 2.33584 | -0.27255 | 0.887283 |
| NOB1 | 5.41954 | 0.197938 | 0.999532 | | 5.307156 | -0.11899 | 0.938462 | | 5.104563 | -0.31709 | 0.759304 |
| HIP1 | 5.296696 | -0.27274 | 0.999532 | | 6.179784 | -0.10033 | 0.938464 | | 4.636876 | 0.191273 | 0.88082 |
| ZNF502 | 1.654936 | -0.04381 | 0.999532 | | 2.084187 | 0.167052 | 0.938464 | | 1.834094 | 0.23144 | 0.913156 |
| MYOZ2 | 2.227284 | -0.22491 | 0.999532 | | 2.809427 | 0.141239 | 0.938464 | | 2.295335 | 0.383518 | 0.814434 |
| HSPH1 | 3.504667 | -0.07733 | 0.999532 | | 4.171621 | 0.114312 | 0.938464 | | 3.463846 | 0.209709 | 0.881837 |
| STIL | 2.169494 | 0.694984 | 0.999532 | | 2.000529 | 0.163424 | 0.938464 | | 2.118245 | -0.51455 | 0.723509 |
| OTOR | 2.878247 | 0.631483 | 0.999532 | | 2.500406 | -0.15867 | 0.938464 | | 2.602364 | -0.77357 | 0.520848 |
| ULK3 | 3.480636 | -0.14299 | 0.999532 | | 3.877024 | -0.11975 | 0.93847 | | 3.154478 | 0.041785 | 0.98556 |
| IZUMO1R | 2.743553 | 0.278202 | 0.999532 | | 2.709263 | -0.14593 | 0.938478 | | 2.487477 | -0.40526 | 0.783005 |
| HNRNPA | 33.92517 | -0.01195 | 0.999532 | | 39.32456 | 0.06706 | 0.938478 | | 33.13193 | 0.110015 | 0.896263 |
| ATP5F1E | 2.278068 | 0.076551 | 0.999532 | | 2.445739 | -0.15189 | 0.938478 | | 2.211629 | -0.20397 | 0.915991 |
| ZNF335 | 2.616913 | -0.06238 | 0.999532 | | 3.314718 | 0.138042 | 0.938648 | | 2.692402 | 0.213472 | 0.897655 |
| IL17RA | 3.614473 | 0.302573 | 0.999532 | | 3.75767 | 0.127629 | 0.938648 | | 3.662443 | -0.16108 | 0.914692 |
| PDE3A | 3.312178 | -0.21856 | 0.999532 | | 4.327381 | 0.113825 | 0.938648 | | 3.415771 | 0.352194 | 0.778209 |
| NLRP9 | 2.501647 | -0.31047 | 0.999532 | | 3.014886 | -0.14907 | 0.938648 | | 2.182251 | 0.184235 | 0.926586 |
| PMAIP1 | 2.976668 | -0.503 | 0.999532 | | 3.847205 | -0.13484 | 0.938648 | | 2.733977 | 0.414339 | 0.798028 |
| PLPP4 | 3.523213 | -0.32144 | 0.999532 | | 4.324057 | -0.12659 | 0.938648 | | 3.327068 | 0.213529 | 0.892869 |
| APLF | 2.373801 | 0.133189 | 0.999532 | | 2.442758 | -0.15648 | 0.938661 | | 2.219369 | -0.2723 | 0.885078 |
| DCP2 | 3.691165 | 0.159614 | 0.999532 | | 4.042721 | 0.116271 | 0.938661 | | 3.648964 | -0.02593 | 0.991521 |

| | | | | | | | | | | | |
|---|---|---|---|---|---|---|---|---|---|---|---|
| TAB1 | 3.18749 | 0.201282 | 0.999532 | | 3.72072 | 0.130949 | 0.93885 | | 3.038023 | -0.06708 | 0.974179 |
| EHMT1 | 4.530861 | 0.26242 | 0.999532 | | 4.816771 | 0.108 | 0.939048 | | 4.386935 | -0.13941 | 0.920259 |
| ACTR3B | 2.798799 | -0.24408 | 0.999532 | | 3.153609 | -0.12996 | 0.939048 | | 2.579111 | 0.131081 | 0.945129 |
| SMG6 | 4.471894 | 0.220684 | 0.999532 | | 4.408651 | -0.11609 | 0.939048 | | 4.116669 | -0.32185 | 0.791466 |
| PPP6C | 11.09416 | -0.09268 | 0.999532 | | 12.94386 | 0.082237 | 0.939048 | | 10.65765 | 0.199505 | 0.814402 |
| GLIS1 | 2.296404 | 0.632604 | 0.999532 | | 2.263338 | 0.150319 | 0.939048 | | 2.439972 | -0.46308 | 0.737651 |
| PDPR | 4.726441 | 0.128214 | 0.999532 | | 4.748582 | -0.11278 | 0.939048 | | 4.309164 | -0.21827 | 0.86209 |
| ZHX2 | 3.293128 | -0.24766 | 0.999532 | | 3.876542 | -0.1186 | 0.939048 | | 3.031208 | 0.147062 | 0.926957 |
| ATL3 | 4.022403 | -0.49184 | 0.999532 | | 5.43368 | 0.106773 | 0.939048 | | 4.075695 | 0.608537 | 0.575547 |
| DBR1 | 3.24327 | 0.33035 | 0.999532 | | 3.051251 | -0.13394 | 0.939048 | | 2.93244 | -0.44576 | 0.719913 |
| SMOC2 | 1.965663 | -0.2224 | 0.999532 | | 2.181449 | -0.15606 | 0.939048 | | 1.824165 | 0.087487 | 0.974421 |
| SLMAP | 4.845169 | 0.198778 | 0.999532 | | 4.802413 | -0.11366 | 0.939048 | | 4.454972 | -0.29825 | 0.815713 |
| DEFB103 | 1.839376 | -0.09281 | 0.999532 | | 2.070072 | -0.16699 | 0.939048 | | 1.671641 | -0.05692 | 0.985693 |
| DDHD2 | 5.816529 | -0.22342 | 0.999532 | | 6.876147 | -0.10156 | 0.939048 | | 5.458333 | 0.137897 | 0.910231 |
| RGS1 | 1.710767 | -0.11552 | 0.999532 | | 1.951664 | -0.16601 | 0.939048 | | 1.590464 | -0.02917 | 0.994192 |
| SLCO4A1 | 2.473948 | 0.165939 | 0.999532 | | 2.583689 | -0.15311 | 0.939048 | | 2.273505 | -0.29843 | 0.864245 |
| HNRNPUL | 7.08477 | 0.28985 | 0.999532 | | 7.45014 | 0.097747 | 0.939048 | | 6.925266 | -0.17097 | 0.876781 |
| F3 | 10.66214 | 0.426379 | 0.999532 | | 10.65064 | 0.114354 | 0.939048 | | 10.36448 | -0.27046 | 0.794815 |
| ALG9 | 2.814072 | 0.29685 | 0.999532 | | 2.575498 | -0.14225 | 0.939048 | | 2.499039 | -0.42365 | 0.761699 |
| NABP2 | 3.124489 | 0.589048 | 0.999532 | | 3.13833 | 0.138681 | 0.939048 | | 3.271294 | -0.42792 | 0.739496 |
| IFI27L1 | 2.301723 | -0.17032 | 0.999532 | | 2.544393 | -0.14795 | 0.939048 | | 2.171838 | 0.0404 | 0.990103 |
| PLXDC1 | 2.162758 | 0.231043 | 0.999532 | | 2.177334 | -0.15361 | 0.939094 | | 2.069099 | -0.36288 | 0.832317 |
| GAN | 2.781074 | 0.295459 | 0.999532 | | 2.756239 | -0.14371 | 0.939249 | | 2.611563 | -0.42322 | 0.760473 |
| ARHGEF | 6.713657 | -0.10183 | 0.999532 | | 7.687074 | 0.094227 | 0.939249 | | 6.648386 | 0.205964 | 0.857043 |
| CIB1 | 5.915599 | 0.029869 | 0.999532 | | 6.966263 | 0.098205 | 0.939345 | | 5.974082 | 0.086694 | 0.947941 |
| NIPSNAP | 4.243731 | -0.1565 | 0.999532 | | 5.312841 | 0.111974 | 0.939415 | | 4.344726 | 0.288217 | 0.811123 |
| PHYHD1 | 3.878254 | 0.374286 | 0.999532 | | 3.942641 | 0.123186 | 0.939521 | | 3.879437 | -0.23298 | 0.870247 |
| HCN4 | 2.611854 | 0.374504 | 0.999532 | | 2.65429 | 0.151226 | 0.939521 | | 2.791425 | -0.20436 | 0.915533 |
| NUDT16 | 2.976661 | -0.17225 | 0.999532 | | 3.464131 | -0.12731 | 0.939521 | | 2.859001 | 0.06264 | 0.977473 |
| FAM83F | 3.385825 | 0.28257 | 0.999532 | | 3.248589 | -0.13341 | 0.939521 | | 3.034193 | -0.39952 | 0.763186 |
| NUDT13 | 2.860893 | 0.203373 | 0.999532 | | 2.940603 | -0.13724 | 0.939521 | | 2.673645 | -0.31896 | 0.834867 |
| PELI2 | 3.307955 | 0.090221 | 0.999532 | | 3.464642 | -0.12454 | 0.939521 | | 3.115733 | -0.19236 | 0.900848 |
| TMIGD3 | 2.198752 | 0.04773 | 0.999532 | | 2.253022 | -0.15536 | 0.939521 | | 2.094556 | -0.19132 | 0.92697 |
| KLF14 | 2.162073 | 0.257393 | 0.999532 | | 2.095217 | -0.16689 | 0.939521 | | 2.011084 | -0.40814 | 0.819366 |
| DCAF15 | 2.930528 | 0.419643 | 0.999532 | | 2.821126 | -0.13947 | 0.939521 | | 2.834402 | -0.53845 | 0.674145 |
| CATSPER | 1.773374 | -0.32168 | 0.999532 | | 2.294073 | 0.153515 | 0.939521 | | 1.860062 | 0.497488 | 0.776283 |
| BCL10 | 3.344799 | 0.067115 | 0.999532 | | 3.548106 | 0.119579 | 0.939521 | | 3.36094 | 0.05677 | 0.978229 |
| USP35 | 2.939666 | 0.012129 | 0.999532 | | 3.064079 | -0.1318 | 0.939521 | | 2.694935 | -0.13371 | 0.945032 |
| LZTFL1 | 2.410146 | 0.169498 | 0.999532 | | 2.787627 | 0.143717 | 0.939521 | | 2.457478 | -0.00623 | 0.99833 |
| MX2 | 3.666627 | 0.558719 | 0.999532 | | 3.23688 | 0.126572 | 0.939543 | | 3.50202 | -0.41285 | 0.720106 |
| FBXW11 | 4.554644 | -0.14385 | 0.999532 | | 5.206997 | -0.10704 | 0.939546 | | 4.174904 | 0.056392 | 0.974421 |
| SLC44A5 | 2.726668 | 0.456122 | 0.999532 | | 2.522885 | -0.14743 | 0.939569 | | 2.515732 | -0.58506 | 0.661217 |
| ATG2A | 2.719971 | 0.377891 | 0.999532 | | 2.799051 | 0.135568 | 0.939569 | | 2.750657 | -0.22474 | 0.88588 |
| BAIAP2L | 2.687961 | 0.219985 | 0.999532 | | 3.146365 | 0.143667 | 0.939569 | | 2.737148 | -0.03915 | 0.989437 |
| CD2 | 2.861021 | 0.317085 | 0.999532 | | 2.605117 | -0.14358 | 0.939569 | | 2.649896 | -0.43963 | 0.757035 |
| SSBP1 | 5.879143 | 0.315956 | 0.999532 | | 6.156026 | 0.097162 | 0.939569 | | 5.826288 | -0.19765 | 0.865363 |
| GTPBP4 | 6.113781 | -0.08765 | 0.999532 | | 7.548977 | 0.09082 | 0.939765 | | 6.149765 | 0.198926 | 0.854336 |
| FRG2 | 3.29604 | 0.91638 | 0.999532 | | 2.671997 | 0.136077 | 0.939787 | | 3.271575 | -0.76358 | 0.453945 |
| CFAP44 | 2.056513 | 0.224276 | 0.999532 | | 2.067598 | -0.1545 | 0.939896 | | 1.898823 | -0.35964 | 0.836453 |
| LRCH2 | 1.597377 | -0.2053 | 0.999532 | | 1.794618 | -0.17131 | 0.940051 | | 1.510996 | 0.053248 | 0.988853 |
| RCC2 | 5.175107 | -0.07199 | 0.999532 | | 5.415972 | -0.10226 | 0.940054 | | 4.822699 | -0.00672 | 0.997739 |
| MED14 | 5.536248 | 0.114158 | 0.999532 | | 5.943227 | 0.104731 | 0.940054 | | 5.432622 | 0.00217 | 0.999385 |
| SLC16A1 | 2.301705 | -0.19211 | 0.999532 | | 2.731388 | 0.141322 | 0.940082 | | 2.227885 | 0.352383 | 0.834627 |
| ZBTB8OS | 5.132224 | 0.226404 | 0.999532 | | 5.017686 | -0.10869 | 0.940082 | | 4.656601 | -0.31524 | 0.780676 |
| SLC15A2 | 1.801335 | 0.22261 | 0.999532 | | 1.926008 | 0.162962 | 0.940186 | | 1.783668 | -0.04786 | 0.988853 |
| ZBTB6 | 2.280562 | 0.144532 | 0.999532 | | 2.304417 | -0.15015 | 0.940273 | | 2.142285 | -0.27885 | 0.877703 |
| RAD51B | 6.030898 | 0.245931 | 0.999532 | | 6.41644 | 0.092685 | 0.940278 | | 5.906695 | -0.13724 | 0.906811 |
| KIFAP3 | 3.588285 | 0.349565 | 0.999532 | | 3.533476 | -0.1206 | 0.940363 | | 3.299172 | -0.45108 | 0.699971 |
| CACNG6 | 2.512611 | 0.083748 | 0.999532 | | 2.455355 | -0.14275 | 0.940363 | | 2.398656 | -0.211 | 0.909431 |
| NACAD | 2.034106 | 0.43158 | 0.999532 | | 2.120653 | 0.155443 | 0.940363 | | 1.984634 | -0.25931 | 0.886495 |
| UHRF2 | 5.167405 | 0.128775 | 0.999532 | | 5.092346 | -0.10428 | 0.940363 | | 4.723752 | -0.22898 | 0.862823 |
| USP39 | 2.257832 | -0.14099 | 0.999532 | | 2.644476 | 0.134169 | 0.940481 | | 2.171693 | 0.28754 | 0.865065 |
| MIER3 | 2.809754 | -0.08361 | 0.999532 | | 3.231008 | 0.132714 | 0.940481 | | 2.726907 | 0.238757 | 0.883792 |
| SAC3D1 | 2.645263 | 0.818905 | 0.999532 | | 2.518111 | 0.146851 | 0.940481 | | 2.697014 | -0.64811 | 0.602029 |
| MTRFR | 3.677907 | 0.328137 | 0.999532 | | 3.4121 | -0.12113 | 0.940481 | | 3.386368 | -0.43378 | 0.714479 |
| DUX5 | 6.80614 | 0.084524 | 0.999532 | | 7.612782 | -0.09139 | 0.940481 | | 6.474888 | -0.14817 | 0.897958 |
| TRIM47 | 4.211977 | -0.0327 | 0.999532 | | 4.882273 | 0.109088 | 0.940481 | | 4.210449 | 0.150424 | 0.920259 |
| CCDC125 | 2.576798 | 0.191115 | 0.999532 | | 3.021844 | 0.130697 | 0.940481 | | 2.646888 | -0.04114 | 0.987274 |
| TMCC3 | 3.262934 | 0.115391 | 0.999532 | | 3.492613 | 0.122513 | 0.940481 | | 3.150218 | 0.021496 | 0.994192 |
| TGFA | 2.82728 | 0.359126 | 0.999532 | | 3.046137 | 0.134295 | 0.940481 | | 2.764197 | -0.204 | 0.897958 |
| ALG3 | 3.305089 | -0.24621 | 0.999532 | | 3.947502 | -0.12502 | 0.940481 | | 3.183653 | 0.135196 | 0.940162 |
| NMS | 2.218395 | -0.15756 | 0.999532 | | 2.492886 | -0.14518 | 0.940481 | | 2.101131 | 0.028127 | 0.993618 |
| CCDC87 | 2.182153 | 0.549 | 0.999532 | | 1.939161 | -0.16455 | 0.940481 | | 2.00727 | -0.69357 | 0.646821 |
| NAA30 | 4.033904 | -0.05093 | 0.999532 | | 4.678377 | -0.10862 | 0.940481 | | 3.758118 | -0.03802 | 0.985693 |
| CACNB2 | 2.475164 | 0.443055 | 0.999532 | | 2.351574 | 0.156273 | 0.940481 | | 2.371158 | -0.25881 | 0.883116 |
| MALT1 | 2.736314 | 0.611353 | 0.999532 | | 2.22783 | -0.14853 | 0.940481 | | 2.426166 | -0.7467 | 0.554813 |
| WRAP53 | 2.530073 | 0.165202 | 0.999532 | | 2.80625 | 0.144734 | 0.940481 | | 2.511009 | 0.000185 | 0.999949 |
| EP300 | 5.739544 | 0.109165 | 0.999532 | | 6.46022 | 0.097341 | 0.940481 | | 5.552896 | 0.011124 | 0.996681 |
| CNOT11 | 5.162085 | 0.31262 | 0.999532 | | 5.427796 | 0.099704 | 0.940481 | | 5.058238 | -0.19283 | 0.868512 |
| HEY1 | 2.678408 | -0.06652 | 0.999532 | | 3.095268 | -0.14788 | 0.940481 | | 2.575227 | -0.06843 | 0.977635 |
| COQ2 | 2.906175 | -0.63915 | 0.999532 | | 4.354549 | 0.112574 | 0.940481 | | 2.897634 | 0.768034 | 0.49299 |
| SH2D3C | 5.613926 | -0.28506 | 0.999532 | | 7.310963 | 0.10009 | 0.940481 | | 5.621789 | 0.410358 | 0.666847 |
| OXSR1 | 6.241584 | 0.204583 | 0.999532 | | 6.806381 | 0.098002 | 0.940481 | | 6.082735 | -0.08279 | 0.949136 |
| GRIK4 | 1.938689 | 0.288001 | 0.999532 | | 1.958188 | -0.16199 | 0.940481 | | 1.815108 | -0.42942 | 0.805576 |
| CCDC8 | 3.283464 | 0.049582 | 0.999532 | | 3.475537 | -0.11949 | 0.940481 | | 2.945682 | -0.14845 | 0.92697 |
| RETREG2 | 3.895352 | -0.19372 | 0.999532 | | 4.648256 | 0.111816 | 0.940481 | | 3.858456 | 0.319229 | 0.799944 |
| ANKLE2 | 6.348489 | -0.06191 | 0.999532 | | 7.718224 | 0.085714 | 0.940481 | | 6.350949 | 0.169804 | 0.87591 |
| LBR | 3.386449 | 0.020683 | 0.999532 | | 3.380323 | -0.12781 | 0.940481 | | 3.129671 | -0.13979 | 0.93603 |
| CASP4 | 4.411822 | -0.26815 | 0.999532 | | 5.223996 | -0.10626 | 0.940481 | | 4.206465 | 0.185147 | 0.892869 |
| RNF13 | 2.589621 | 0.194298 | 0.999532 | | 2.645208 | 0.138795 | 0.940481 | | 2.499236 | -0.04131 | 0.988853 |
| IARS1 | 8.674398 | -0.38005 | 0.999532 | | 10.65891 | 0.074884 | 0.940481 | | 8.306691 | 0.451436 | 0.553283 |
| SFR1 | 3.431433 | 0.089255 | 0.999532 | | 3.464833 | -0.12502 | 0.940481 | | 3.070282 | -0.19483 | 0.900376 |
| CYP3A5 | 4.333138 | 0.268724 | 0.999532 | | 4.487946 | 0.113687 | 0.940481 | | 4.00075 | -0.14518 | 0.918675 |
| ACOT1 | 3.459912 | 0.071262 | 0.999532 | | 3.824233 | 0.11674 | 0.940481 | | 3.308343 | 0.06152 | 0.974421 |
| SLA | 5.385337 | -0.197 | 0.999532 | | 6.784958 | 0.098263 | 0.940481 | | 5.491815 | 0.315 | 0.753711 |
| TMEM39A | 6.304006 | 0.238502 | 0.999532 | | 6.211828 | -0.09379 | 0.940481 | | 5.930224 | -0.31572 | 0.735469 |
| NOD1 | 1.98023 | -0.3495 | 0.999532 | | 2.326296 | -0.14628 | 0.940481 | | 1.82935 | 0.222754 | 0.910105 |
| HIC1 | 3.525064 | -1.04413 | 0.831313 | | 5.033193 | -0.10771 | 0.940481 | | 3.043908 | 0.953814 | 0.333968 |
| DSCAML | 2.388076 | 0.254845 | 0.999532 | | 2.465117 | 0.145305 | 0.940481 | | 2.386774 | -0.09478 | 0.965649 |
| TOP2B | 3.687835 | -0.05738 | 0.999532 | | 4.406881 | 0.120654 | 0.940481 | | 3.72334 | 0.197263 | 0.892646 |
| KARS1 | 5.337994 | 0.253372 | 0.999532 | | 5.53862 | 0.099493 | 0.940481 | | 5.075096 | -0.13178 | 0.914692 |
| MAPK13 | 2.826447 | 0.305763 | 0.999532 | | 2.545755 | -0.14942 | 0.940481 | | 2.460114 | -0.43486 | 0.773546 |
| NUDT14 | 4.692275 | 0.119976 | 0.999532 | | 4.703763 | -0.10644 | 0.940481 | | 4.443033 | -0.20934 | 0.871957 |
| MRPS23 | 4.150091 | 0.032009 | 0.999532 | | 4.45122 | -0.10732 | 0.940481 | | 3.729336 | -0.12285 | 0.939549 |
| IL21R | 2.43691 | -0.56889 | 0.999532 | | 3.360071 | 0.136585 | 0.940481 | | 2.542002 | 0.727196 | 0.628053 |
| LPCAT4 | 5.247554 | -0.64027 | 0.999532 | | 7.324495 | 0.095306 | 0.940481 | | 5.376088 | 0.764384 | 0.358109 |
| DHX38 | 4.187788 | -0.23806 | 0.999532 | | 4.940393 | -0.10232 | 0.940481 | | 3.83763 | 0.154788 | 0.913432 |
| NCALD | 2.383016 | 0.50753 | 0.999532 | | 2.215427 | 0.148487 | 0.940481 | | 2.467603 | -0.34074 | 0.838554 |
| RASSF8 | 9.057268 | 0.072427 | 0.999532 | | 10.1399 | 0.099007 | 0.940481 | | 8.863675 | 0.048065 | 0.97502 |
| SMPD1 | 2.615985 | 0.293603 | 0.999532 | | 2.780209 | 0.130633 | 0.940481 | | 2.586166 | -0.14484 | 0.934572 |
| FYCO1 | 2.340578 | 0.100341 | 0.999532 | | 2.818649 | 0.140947 | 0.940481 | | 2.459099 | 0.061435 | 0.978931 |
| GPX7 | 2.038809 | -0.16017 | 0.999532 | | 2.297559 | -0.14771 | 0.940481 | | 1.925207 | 0.031867 | 0.992286 |
| KCNG2 | 2.641735 | -0.27695 | 0.999532 | | 3.443364 | 0.123344 | 0.940481 | | 2.676911 | 0.419295 | 0.760955 |
| C1orf50 | 2.241135 | 0.057029 | 0.999532 | | 2.313191 | -0.14515 | 0.940481 | | 2.069318 | -0.1865 | 0.923864 |
| BICD2 | 3.929057 | 0.097966 | 0.999532 | | 3.849073 | -0.11568 | 0.940481 | | 3.659571 | -0.19586 | 0.892225 |
| C17orf80 | 4.12437 | 0.000419 | 0.999532 | | 4.337156 | -0.11408 | 0.940481 | | 3.845132 | -0.09802 | 0.955167 |
| GPBP1L1 | 5.998432 | 0.043552 | 0.999532 | | 7.040859 | 0.095192 | 0.940481 | | 6.057931 | 0.084232 | 0.957779 |
| CD69 | 2.11421 | -0.06234 | 0.999532 | | 2.184706 | -0.16372 | 0.940481 | | 1.90843 | -0.07994 | 0.977473 |
| HOXC6 | 5.745713 | -0.0837 | 0.999532 | | 6.973335 | 0.094791 | 0.940481 | | 5.995911 | 0.213555 | 0.858705 |
| H2AX | 3.130721 | 0.093333 | 0.999532 | | 3.182857 | -0.12493 | 0.940481 | | 2.862922 | -0.20154 | 0.896228 |
| FBXO38 | 3.162961 | 0.124344 | 0.999532 | | 3.426804 | -0.12709 | 0.940481 | | 3.043283 | -0.22382 | 0.889853 |
| RNF187 | 9.083466 | 0.150744 | 0.999532 | | 9.774565 | 0.07734 | 0.940481 | | 8.745247 | -0.05803 | 0.963918 |
| DPY19L4 | 2.719586 | 0.500492 | 0.999532 | | 2.335975 | -0.1465 | 0.940481 | | 2.465265 | -0.62933 | 0.637292 |
| STAT4 | 2.334131 | 0.144073 | 0.999532 | | 2.674934 | 0.138654 | 0.940481 | | 2.454255 | 0.006241 | 0.998243 |
| PSORS1C | 2.510282 | 0.999965 | 0.999532 | | 1.891012 | -0.17794 | 0.940481 | | 2.278965 | -1.15454 | 0.323586 |
| FAM169A | 2.38248 | -0.06732 | 0.999532 | | 2.914537 | 0.141325 | 0.940481 | | 2.492547 | 0.22308 | 0.904721 |
| LPCAT1 | 3.423596 | -0.03497 | 0.999532 | | 3.848224 | 0.115814 | 0.940481 | | 3.376244 | 0.172942 | 0.906811 |
| TMEM132 | 2.057651 | 0.340335 | 0.999532 | | 1.967868 | -0.15949 | 0.940481 | | 1.874812 | -0.48081 | 0.766974 |
| ILDR1 | 2.370829 | 0.714409 | 0.999532 | | 2.213105 | 0.157025 | 0.940481 | | 2.426112 | -0.53938 | 0.713783 |
| TMEM31 | 2.374027 | 0.183734 | 0.999532 | | 2.324267 | -0.14678 | 0.940481 | | 2.211326 | -0.31463 | 0.858705 |
| NUDT3 | 5.000928 | 0.121666 | 0.999532 | | 5.593985 | 0.103942 | 0.940481 | | 4.928377 | 0.001278 | 0.999429 |
| ZNF470 | 3.064154 | -0.24084 | 0.999532 | | 3.498172 | -0.12953 | 0.940481 | | 2.877884 | 0.130044 | 0.94388 |
| KRT28 | 9.460351 | 0.006721 | 0.999532 | | 10.89486 | 0.087016 | 0.940637 | | 9.289887 | 0.101666 | 0.922679 |
| CEMIP | 2.675397 | 0.63707 | 0.999532 | | 2.221498 | -0.14791 | 0.940637 | | 2.560866 | -0.76218 | 0.552672 |
| MYBPC2 | 2.403757 | 0.121945 | 0.999532 | | 2.701699 | -0.14094 | 0.940637 | | 2.385348 | -0.23549 | 0.894254 |
| FOXRED1 | 2.136925 | 0.047429 | 0.999532 | | 2.359103 | -0.14614 | 0.940637 | | 1.979699 | -0.17483 | 0.928847 |
| SLC34A2 | 2.136159 | 0.694349 | 0.999532 | | 1.667706 | -0.17213 | 0.940637 | | 1.914521 | -0.84859 | 0.551754 |
| NSD2 | 4.209181 | 0.156633 | 0.999532 | | 4.645631 | 0.108845 | 0.940637 | | 4.291835 | -0.04013 | 0.98556 |
| POP4 | 2.130131 | 0.017995 | 0.999532 | | 2.330541 | -0.14313 | 0.940637 | | 1.92283 | -0.14125 | 0.945902 |
| HERPUD2 | 3.74222 | -0.29685 | 0.999532 | | 4.602914 | 0.103383 | 0.940637 | | 3.672895 | 0.417704 | 0.715437 |
| HRCT1 | 2.534172 | 0.812195 | 0.999532 | | 2.379269 | 0.144381 | 0.940637 | | 2.61857 | -0.64347 | 0.609581 |
| IL7R | 2.045858 | 0.658295 | 0.999532 | | 1.885009 | 0.165667 | 0.940637 | | 2.119871 | -0.47842 | 0.760473 |
| ADAMTS | 5.349856 | 0.413136 | 0.999532 | | 4.952094 | 0.110503 | 0.940637 | | 4.971831 | -0.28872 | 0.814402 |
| HS1BP3 | 2.992029 | 0.560208 | 0.999532 | | 2.953702 | 0.133149 | 0.940637 | | 3.028182 | -0.40776 | 0.754884 |
| ENPP6 | 2.135571 | 0.03988 | 0.999532 | | 2.575555 | 0.149662 | 0.940637 | | 2.217729 | 0.131186 | 0.952785 |
| ZNF706 | 7.076697 | -0.01825 | 0.999532 | | 8.231073 | 0.08913 | 0.940637 | | 7.096134 | 0.129688 | 0.928847 |
| SH3BGRL | 7.090876 | -0.30762 | 0.999532 | | 9.216475 | 0.08323 | 0.940637 | | 7.009943 | 0.40783 | 0.621895 |
| DFFB | 2.137681 | -0.00548 | 0.999532 | | 2.176122 | -0.14547 | 0.940637 | | 1.984301 | -0.1298 | 0.954304 |
| BBIP1 | 4.196189 | 0.282305 | 0.999532 | | 4.272922 | 0.114974 | 0.940637 | | 4.256196 | -0.12821 | 0.934701 |
| TPD52 | 6.956805 | 0.391589 | 0.999532 | | 7.195402 | 0.093306 | 0.940637 | | 6.53646 | -0.27678 | 0.784263 |
| CMYA5 | 3.228781 | 0.312682 | 0.999532 | | 3.115992 | -0.13852 | 0.940637 | | 3.001002 | -0.43535 | 0.750988 |
| EPDR1 | 5.344409 | 0.095447 | 0.999532 | | 5.831597 | 0.106959 | 0.940637 | | 5.197706 | 0.026886 | 0.991159 |
| USP48 | 3.603541 | -0.2524 | 0.999532 | | 4.048593 | -0.11547 | 0.940637 | | 3.307672 | 0.158814 | 0.915991 |
| SSH3 | 2.786154 | 0.266649 | 0.999532 | | 2.772645 | 0.130707 | 0.940637 | | 2.652093 | -0.13058 | 0.945129 |
| METTL21 | 4.398797 | 0.318629 | 0.999532 | | 4.168938 | -0.11035 | 0.940637 | | 4.021835 | -0.41477 | 0.714657 |
| PXDC1 | 4.96446 | -0.45377 | 0.999532 | | 6.272429 | -0.09712 | 0.940637 | | 4.531077 | 0.374454 | 0.728015 |
| INHBE | 2.500866 | 0.204316 | 0.999532 | | 2.528407 | -0.15334 | 0.940637 | | 2.352343 | -0.34502 | 0.849021 |
| ZBTB14 | 1.949261 | 0.318319 | 0.999532 | | 2.062656 | 0.157513 | 0.940637 | | 1.889404 | -0.14139 | 0.948127 |
| BOLA3 | 4.114847 | 0.014015 | 0.999532 | | 4.231714 | -0.12187 | 0.940724 | | 3.784908 | -0.1395 | 0.938429 |
| LRATD2 | 4.919613 | 0.242439 | 0.999532 | | 4.915238 | -0.10459 | 0.940724 | | 4.588969 | -0.32808 | 0.760337 |
| RAB23 | 2.625745 | -0.00383 | 0.999532 | | 3.08191 | 0.1252 | 0.940724 | | 2.604515 | 0.148469 | 0.933313 |
| MN1 | 2.739003 | 0.066203 | 0.999532 | | 2.798645 | -0.13388 | 0.940724 | | 2.425546 | -0.18348 | 0.918194 |
| SLC30A3 | 2.001931 | 0.213647 | 0.999532 | | 2.274425 | 0.159145 | 0.940724 | | 2.119703 | -0.04198 | 0.990711 |
| SLC44A3 | 2.615248 | 0.161884 | 0.999532 | | 2.803531 | 0.135687 | 0.940724 | | 2.525941 | -0.00825 | 0.997967 |
| SLC2A9 | 4.55095 | 0.429166 | 0.999532 | | 4.291535 | 0.106742 | 0.940724 | | 4.430212 | -0.3078 | 0.78518 |
| NLRP1 | 3.717338 | -0.1747 | 0.999532 | | 4.338958 | -0.11588 | 0.940724 | | 3.453399 | 0.073949 | 0.969501 |
| NADSYN1 | 4.583932 | 0.006482 | 0.999532 | | 5.313122 | 0.101453 | 0.940724 | | 4.831602 | 0.144656 | 0.920486 |
| KCNJ9 | 6.411487 | 0.090684 | 0.999532 | | 6.827664 | -0.10405 | 0.940724 | | 5.936817 | -0.17698 | 0.892646 |
| EIF2AK4 | 1.850319 | -0.09626 | 0.999532 | | 2.119236 | 0.154399 | 0.940724 | | 1.773407 | 0.270038 | 0.892869 |
| FBXL5 | 7.011502 | -0.28857 | 0.999532 | | 8.730401 | 0.08775 | 0.940724 | | 6.562979 | 0.407363 | 0.644433 |
| CHAF1B | 2.135132 | 0.332609 | 0.999532 | | 1.971062 | -0.16324 | 0.940724 | | 2.030568 | -0.47488 | 0.791466 |
| CLECL1 | 1.998983 | -0.02556 | 0.999532 | | 2.06477 | -0.15101 | 0.940724 | | 1.788116 | -0.11279 | 0.963771 |
| RPL12 | 23.85618 | -0.22188 | 0.999532 | | 28.0388 | -0.06002 | 0.940724 | | 21.88887 | 0.182427 | 0.79686 |
| SRP54 | 4.578283 | 0.339106 | 0.999532 | | 4.502735 | 0.107803 | 0.940724 | | 4.317105 | -0.21878 | 0.86549 |
| ANKS1A | 2.884313 | -0.39831 | 0.999532 | | 3.787548 | 0.126701 | 0.940724 | | 2.710952 | 0.539201 | 0.668481 |
| LRRC2 | 4.489176 | 0.238442 | 0.999532 | | 4.526204 | -0.12063 | 0.940724 | | 4.153044 | -0.3409 | 0.783891 |
| BTN3A2 | 3.802596 | -0.50988 | 0.999532 | | 4.656712 | -0.11554 | 0.940724 | | 3.445134 | 0.412814 | 0.740755 |
| FXYD1 | 4.182318 | 0.326557 | 0.999532 | | 4.217046 | 0.117676 | 0.940724 | | 4.041798 | -0.20066 | 0.892869 |
| DVL1 | 8.124938 | 0.22537 | 0.999532 | | 8.748659 | 0.085628 | 0.940761 | | 8.112877 | -0.1216 | 0.90928 |
| RAP2A | 5.524242 | 0.096008 | 0.999532 | | 6.230081 | 0.091519 | 0.94077 | | 5.479294 | 0.015981 | 0.99505 |
| SMR3A | 2.197063 | 0.190674 | 0.999532 | | 2.323332 | -0.14721 | 0.941045 | | 2.051307 | -0.31434 | 0.859426 |
| ZNF132 | 2.790686 | 0.129679 | 0.999532 | | 3.204588 | 0.136332 | 0.941128 | | 2.851904 | 0.022146 | 0.995112 |
| H2BC3 | 2.944607 | -0.14113 | 0.999532 | | 3.344872 | -0.12574 | 0.941528 | | 2.715247 | 0.03319 | 0.991159 |
| SEMA4F | 2.910311 | 0.527899 | 0.999532 | | 2.671803 | -0.13845 | 0.941638 | | 2.699627 | -0.64702 | 0.600081 |
| GOLGA6 | 2.433479 | 0.165975 | 0.999532 | | 2.573383 | -0.14145 | 0.941638 | | 2.251447 | -0.28365 | 0.872933 |
| PLXNB3 | 2.476112 | -0.1455 | 0.999532 | | 2.762898 | -0.13195 | 0.941665 | | 2.322325 | 0.033298 | 0.991434 |
| ZNF554 | 2.372542 | 0.310403 | 0.999532 | | 2.351058 | -0.14139 | 0.941665 | | 2.195251 | -0.43218 | 0.771011 |
| PDE12 | 4.33488 | -0.17043 | 0.999532 | | 5.639789 | 0.099696 | 0.941665 | | 4.221773 | 0.285199 | 0.806797 |
| LZIC | 3.467794 | 0.108424 | 0.999532 | | 3.654315 | 0.115582 | 0.941665 | | 3.536442 | 0.021123 | 0.994537 |
| P2RX1 | 2.249048 | 0.460845 | 0.999532 | | 2.07655 | -0.15532 | 0.941665 | | 2.11826 | -0.59887 | 0.686214 |
| KYAT1 | 3.498967 | 0.503741 | 0.999532 | | 3.240801 | -0.12934 | 0.941665 | | 3.132466 | -0.61861 | 0.585657 |
| CACNG1 | 2.244681 | 0.344207 | 0.999532 | | 2.398148 | 0.14297 | 0.941665 | | 2.280912 | -0.17907 | 0.924186 |
| POGLUT | 2.515916 | -0.39265 | 0.999532 | | 3.181431 | -0.13078 | 0.941665 | | 2.557756 | 0.279818 | 0.87591 |
| XCR1 | 2.213024 | -0.04994 | 0.999532 | | 2.625703 | -0.15137 | 0.941665 | | 2.184204 | -0.08286 | 0.974421 |
| IHO1 | 2.122258 | 0.486733 | 0.999532 | | 2.065828 | -0.16654 | 0.941665 | | 2.037507 | -0.63089 | 0.686214 |
| DUSP26 | 3.095941 | 0.378547 | 0.999532 | | 2.980136 | -0.14022 | 0.941665 | | 3.020706 | -0.49808 | 0.724411 |
| COG5 | 6.188193 | 0.130782 | 0.999532 | | 6.672035 | 0.091523 | 0.941665 | | 5.766424 | -0.02202 | 0.991341 |
| GORAB | 2.519552 | -0.1565 | 0.999532 | | 3.075878 | 0.127365 | 0.941665 | | 2.523997 | 0.301641 | 0.858647 |
| C16orf89 | 2.827576 | 0.409205 | 0.999532 | | 2.648415 | -0.13597 | 0.941665 | | 2.522656 | -0.52609 | 0.70258 |
| LY6K | 2.187181 | 0.194227 | 0.999532 | | 2.302557 | -0.15734 | 0.941665 | | 1.967936 | -0.33441 | 0.853737 |
| PIWIL3 | 1.837526 | -0.03295 | 0.999532 | | 2.261035 | 0.143764 | 0.941665 | | 1.944729 | 0.195685 | 0.920805 |
| SMARCA | 7.706476 | 0.05042 | 0.999532 | | 8.085315 | -0.09062 | 0.941665 | | 7.145999 | -0.11985 | 0.918172 |
| KLK14 | 2.179912 | 0.008464 | 0.999532 | | 2.712987 | 0.137277 | 0.941665 | | 2.288821 | 0.160895 | 0.938429 |
| PARS2 | 2.27879 | 0.048829 | 0.999532 | | 2.54155 | -0.14555 | 0.941665 | | 2.275075 | -0.17566 | 0.931827 |
| THAP11 | 3.813654 | 0.462863 | 0.999532 | | 3.16196 | -0.12523 | 0.941665 | | 3.476685 | -0.58253 | 0.643272 |
| TKT | 3.566352 | 0.114355 | 0.999532 | | 3.967528 | 0.122209 | 0.941665 | | 3.311208 | 0.023143 | 0.993618 |
| SEC14L1 | 32.85354 | -0.3573 | 0.999532 | | 41.14077 | -0.05801 | 0.941665 | | 30.92006 | 0.317653 | 0.566812 |
| IFT74 | 2.431665 | 0.094906 | 0.999532 | | 2.631983 | -0.13677 | 0.941665 | | 2.291795 | -0.21057 | 0.904721 |
| DDX4 | 2.678212 | 0.156482 | 0.999532 | | 3.091258 | 0.132551 | 0.941751 | | 2.886599 | 0.007264 | 0.997967 |
| SLC35B3 | 3.037642 | 0.014666 | 0.999532 | | 3.639417 | 0.116344 | 0.941751 | | 2.971548 | 0.117422 | 0.945456 |
| CPSF3 | 2.711429 | 0.022645 | 0.999532 | | 3.041488 | 0.127354 | 0.941751 | | 2.7614 | 0.114409 | 0.952656 |
| OR2K2 | 1.681419 | 0.240725 | 0.999532 | | 1.884029 | 0.162307 | 0.941751 | | 1.837729 | -0.05865 | 0.98556 |
| ICAM3 | 2.205296 | -0.41825 | 0.999532 | | 2.612037 | -0.14055 | 0.941751 | | 2.020145 | 0.29372 | 0.880375 |
| RHBG | 3.276708 | 0.267963 | 0.999532 | | 2.969895 | -0.13448 | 0.941751 | | 2.763527 | -0.38205 | 0.794049 |
| ISG20 | 2.545676 | 0.123506 | 0.999532 | | 2.531618 | -0.13744 | 0.941751 | | 2.321761 | -0.24008 | 0.88885 |
| SPATA7 | 2.874189 | 0.617042 | 0.999532 | | 2.382049 | 0.137358 | 0.941782 | | 2.751364 | -0.47673 | 0.737109 |
| PTH2R | 2.275296 | 0.464491 | 0.999532 | | 2.000738 | -0.15155 | 0.941785 | | 2.100436 | -0.59756 | 0.679452 |
| SMYD4 | 2.123926 | -0.09542 | 0.999532 | | 2.684825 | 0.136403 | 0.941785 | | 2.284972 | 0.247569 | 0.889041 |
| PLEKHA4 | 9.155516 | 0.150596 | 0.999532 | | 10.32954 | 0.07941 | 0.941785 | | 9.365222 | -0.04354 | 0.974421 |
| ZNF579 | 5.330753 | 0.352255 | 0.999532 | | 5.528367 | -0.11359 | 0.941785 | | 4.872294 | -0.43593 | 0.702463 |
| SMUG1 | 2.481869 | -0.59033 | 0.999532 | | 3.278277 | -0.1208 | 0.941785 | | 2.34418 | 0.491797 | 0.732916 |
| RASL10B | 2.308038 | 0.220458 | 0.999532 | | 2.213502 | -0.14921 | 0.941785 | | 2.146269 | -0.34658 | 0.843564 |
| POGK | 4.63819 | 0.284859 | 0.999532 | | 4.666311 | 0.103033 | 0.941785 | | 4.574192 | -0.16498 | 0.900146 |
| FIGLA | 1.822774 | 0.507531 | 0.999532 | | 1.786888 | -0.16352 | 0.941785 | | 1.79564 | -0.65062 | 0.672279 |
| CSTA | 1.879931 | -0.17259 | 0.999532 | | 2.344601 | 0.142134 | 0.941785 | | 1.965209 | 0.33549 | 0.858705 |
| IL1R1 | 5.477588 | 0.352132 | 0.999532 | | 5.142911 | -0.10353 | 0.941785 | | 5.309001 | -0.44545 | 0.647218 |
| PHF20 | 3.446376 | 0.216548 | 0.999532 | | 3.168491 | -0.13139 | 0.941785 | | 3.065341 | -0.29812 | 0.837927 |
| UBE2Z | 8.33996 | -0.15775 | 0.999532 | | 9.202422 | -0.0833 | 0.941785 | | 7.728927 | 0.100798 | 0.932589 |
| HDAC10 | 2.997106 | 0.711368 | 0.999532 | | 2.779482 | 0.132878 | 0.941785 | | 3.052493 | -0.55962 | 0.646782 |
| ZMYND19 | 1.805155 | 0.324068 | 0.999532 | | 1.89456 | 0.155809 | 0.941785 | | 1.746921 | -0.14835 | 0.946512 |
| ARCN1 | 3.086914 | 0.410971 | 0.999532 | | 2.77197 | -0.1337 | 0.941785 | | 2.902014 | -0.52792 | 0.668532 |
| USF3 | 3.093365 | 0.04642 | 0.999532 | | 3.269264 | -0.13265 | 0.941785 | | 2.967127 | -0.1705 | 0.92697 |
| SLC12A3 | 4.98744 | -0.21286 | 0.999532 | | 5.569189 | -0.19049 | 0.941785 | | 4.330817 | 0.122734 | 0.949893 |
| PTDSS1 | 3.848604 | 0.104604 | 0.999532 | | 4.185567 | 0.111183 | 0.941857 | | 3.830472 | 0.026921 | 0.991434 |
| NUP205 | 2.796738 | 0.521933 | 0.999532 | | 2.851464 | 0.132125 | 0.941857 | | 2.937945 | -0.37155 | 0.792538 |
| HTRA2 | 3.981201 | 0.036453 | 0.999532 | | 4.59827 | 0.11139 | 0.941857 | | 4.08676 | 0.085075 | 0.961846 |
| ZNF98 | 2.146511 | 0.711337 | 0.999532 | | 2.216914 | 0.161126 | 0.942235 | | 2.316694 | -0.52631 | 0.734904 |
| REX1BD | 4.980544 | 0.1239 | 0.999532 | | 5.702769 | 0.09209 | 0.942235 | | 4.882634 | -0.00561 | 0.997967 |
| PGF | 5.243622 | -0.11152 | 0.999532 | | 5.934003 | -0.10455 | 0.942235 | | 4.903399 | 0.030064 | 0.989138 |
| MAGI3 | 3.454515 | 0.36851 | 0.999532 | | 3.43493 | -0.12769 | 0.942235 | | 3.461466 | -0.49437 | 0.686214 |
| VWA5B2 | 2.60428 | -0.1464 | 0.999532 | | 2.906747 | -0.13514 | 0.942235 | | 2.287378 | 0.030827 | 0.992286 |
| STARD10 | 3.03651 | 0.505543 | 0.999532 | | 2.707495 | -0.13824 | 0.942235 | | 2.778123 | -0.62319 | 0.631504 |
| FBXW8 | 2.551109 | 0.58339 | 0.999532 | | 2.182745 | -0.1475 | 0.942331 | | 2.474547 | -0.7141 | 0.595023 |
| TCP10L | 1.946457 | 0.139605 | 0.999532 | | 2.015264 | -0.15616 | 0.942429 | | 1.89412 | -0.27748 | 0.892646 |
| BIRC5 | 12.60409 | 0.07059 | 0.999532 | | 14.25661 | 0.070412 | 0.942557 | | 12.1715 | 0.023774 | 0.987164 |
| GATAD2B | 4.279697 | -0.09693 | 0.999532 | | 4.961002 | -0.11677 | 0.942557 | | 3.945216 | -0.00179 | 0.999385 |
| RYBP | 3.731004 | -0.0118 | 0.999532 | | 4.573673 | 0.11405 | 0.942557 | | 3.867888 | 0.160892 | 0.927668 |
| NANS | 3.149056 | -0.2144 | 0.999532 | | 4.019239 | 0.11375 | 0.942557 | | 3.016111 | 0.350294 | 0.795796 |
| SENP7 | 6.683756 | 0.322644 | 0.999532 | | 6.674354 | 0.088256 | 0.942557 | | 6.60046 | -0.21058 | 0.843564 |
| CLEC1A | 2.389508 | -0.52577 | 0.999532 | | 2.90473 | -0.12949 | 0.942557 | | 2.130013 | 0.411273 | 0.803301 |
| EIF4E2 | 6.074276 | -0.10798 | 0.999532 | | 7.07498 | 0.088047 | 0.942557 | | 5.983268 | 0.21762 | 0.833685 |
| TKTL1 | 2.185659 | 0.245026 | 0.999532 | | 2.163059 | -0.14819 | 0.942557 | | 2.101493 | -0.37629 | 0.826705 |
| LYAR | 1.918858 | -0.43278 | 0.999532 | | 2.574084 | 0.13621 | 0.942582 | | 2.09247 | 0.589631 | 0.709825 |
| SNX11 | 3.353407 | -0.01155 | 0.999532 | | 3.546271 | -0.11342 | 0.942582 | | 3.182941 | -0.08915 | 0.962716 |
| JPH3 | 2.765302 | 0.162236 | 0.999532 | | 2.927566 | -0.14033 | 0.942582 | | 2.599586 | -0.2834 | 0.867976 |
| ZZEF1 | 4.760933 | 0.134927 | 0.999532 | | 5.208794 | 0.09431 | 0.942582 | | 4.67165 | -0.02259 | 0.991434 |
| ZNF350 | 2.840974 | 0.440872 | 0.999532 | | 2.639459 | -0.13354 | 0.942582 | | 2.623208 | -0.5567 | 0.666906 |
| STRIT1 | 2.066497 | 0.418738 | 0.999532 | | 2.142208 | 0.148012 | 0.942582 | | 2.11394 | -0.25424 | 0.88885 |
| KCTD19 | 2.041473 | 0.083936 | 0.999532 | | 2.095777 | -0.14855 | 0.942582 | | 1.892184 | -0.21527 | 0.914929 |
| PRPF6 | 7.056163 | -0.14152 | 0.999532 | | 7.948906 | -0.08607 | 0.942582 | | 6.483755 | 0.074445 | 0.958297 |
| AXL | 4.535052 | -0.05578 | 0.999532 | | 5.321938 | 0.112168 | 0.942582 | | 4.48366 | 0.191011 | 0.883116 |
| SGPP2 | 3.39969 | 0.159703 | 0.999532 | | 3.489824 | -0.1256 | 0.942582 | | 3.048796 | -0.26463 | 0.865065 |
| TAC1 | 2.379461 | 0.091591 | 0.999532 | | 2.613416 | 0.133583 | 0.942582 | | 2.310363 | 0.055479 | 0.982338 |
| STARD9 | 2.670593 | -0.24696 | 0.999532 | | 3.109344 | -0.1327 | 0.942582 | | 2.37842 | 0.134672 | 0.946765 |
| TNP2 | 1.900069 | 0.610089 | 0.999532 | | 1.896594 | 0.164255 | 0.942582 | | 2.040092 | -0.43016 | 0.809485 |
| NIPA1 | 2.296599 | -0.18829 | 0.999532 | | 2.847531 | -0.13124 | 0.942582 | | 2.219635 | 0.082769 | 0.972305 |
| RHBDD3 | 3.620134 | -0.22706 | 0.999532 | | 4.452479 | 0.106618 | 0.942582 | | 3.449403 | 0.33425 | 0.809994 |
| TSTD2 | 2.500935 | 0.133677 | 0.999532 | | 2.68349 | -0.13654 | 0.942582 | | 2.364885 | -0.24926 | 0.88885 |
| TTC19 | 7.166566 | 0.15229 | 0.999532 | | 7.820794 | 0.084776 | 0.942582 | | 7.225563 | -0.05594 | 0.970811 |
| VPREB1 | 1.936378 | 0.065514 | 0.999532 | | 2.079622 | -0.1495 | 0.942582 | | 1.812774 | -0.19672 | 0.923864 |
| OR1N1 | 2.316914 | 0.352642 | 0.999532 | | 2.207605 | -0.14861 | 0.942582 | | 2.298631 | -0.48298 | 0.760473 |
| SLC4A7 | 2.436001 | -0.04061 | 0.999532 | | 2.505582 | -0.1383 | 0.942582 | | 2.191883 | -0.08306 | 0.973356 |
| ZNF496 | 2.590763 | -0.0698 | 0.999532 | | 2.883019 | -0.12732 | 0.942582 | | 2.293242 | -0.03877 | 0.989278 |
| EIF2B2 | 3.57355 | 0.157029 | 0.999532 | | 3.832474 | 0.113682 | 0.942582 | | 3.551779 | -0.04225 | 0.986208 |
| BEX2 | 3.884609 | 0.433798 | 0.999532 | | 3.69651 | -0.11787 | 0.942582 | | 3.492359 | -0.53026 | 0.643361 |
| SCMH1 | 2.393304 | -0.14582 | 0.999532 | | 2.921694 | 0.129199 | 0.942582 | | 2.601304 | 0.30396 | 0.860036 |
| TRIM42 | 2.440812 | 0.66564 | 0.999532 | | 2.100018 | -0.15568 | 0.942582 | | 2.292138 | -0.80074 | 0.556502 |
| RRAD | 2.897058 | -0.40903 | 0.999532 | | 3.558339 | -0.12114 | 0.942582 | | 2.804975 | 0.299363 | 0.862823 |
| FAM13C | 2.159159 | -0.01348 | 0.999532 | | 2.457389 | -0.14611 | 0.942582 | | 2.184691 | -0.11738 | 0.960773 |
| LRRC34 | 2.520997 | 0.278636 | 0.999532 | | 2.458073 | -0.1401 | 0.942582 | | 2.337831 | -0.40082 | 0.796512 |
| FBXW2 | 3.426481 | -0.3334 | 0.999532 | | 4.345751 | 0.104973 | 0.942582 | | 3.559543 | 0.454516 | 0.706407 |
| BUD23 | 3.75979 | -0.06353 | 0.999532 | | 4.44422 | 0.102845 | 0.942582 | | 3.812828 | 0.184865 | 0.892646 |
| HKDC1 | 2.7735 | -0.20406 | 0.999532 | | 3.28085 | 0.121046 | 0.942582 | | 2.729073 | 0.3393 | 0.816618 |
| SPECC1L | 4.060141 | 0.449565 | 0.999532 | | 3.83826 | 0.111017 | 0.942582 | | 3.975388 | -0.32125 | 0.791576 |
| CSGALN | 3.437583 | -0.65111 | 0.999532 | | 4.507587 | -0.10852 | 0.942582 | | 3.140418 | 0.56124 | 0.637292 |
| ZNF441 | 2.908065 | 0.223662 | 0.999532 | | 2.780118 | -0.13365 | 0.942582 | | 2.681836 | -0.33724 | 0.827267 |
| MOB3A | 6.937133 | 0.048305 | 0.999532 | | 7.864607 | 0.083925 | 0.942582 | | 6.872843 | 0.053578 | 0.970811 |
| PTPRG | 8.5954 | -0.34665 | 0.999532 | | 10.40168 | -0.07337 | 0.942582 | | 8.098804 | 0.296374 | 0.717627 |
| SWT1 | 2.275384 | -0.11862 | 0.999532 | | 2.738715 | 0.130604 | 0.942582 | | 2.318041 | 0.268198 | 0.87591 |
| KLC2 | 3.656385 | 0.046002 | 0.999532 | | 3.994089 | -0.11756 | 0.942582 | | 3.272698 | -0.14599 | 0.926586 |
| ANKRD54 | 5.634233 | 0.25895 | 0.999532 | | 5.236467 | -0.0983 | 0.942588 | | 5.098151 | -0.35782 | 0.750389 |
| RHOJ | 3.060884 | -0.32596 | 0.999532 | | 4.088724 | 0.105485 | 0.942588 | | 2.949038 | 0.445446 | 0.723681 |
| CD274 | 3.783653 | -0.56679 | 0.999532 | | 5.312219 | -0.11533 | 0.942588 | | 3.853491 | 0.468174 | 0.718643 |
| ARSL | 2.567552 | 0.045204 | 0.999532 | | 2.645685 | -0.13982 | 0.942771 | | 2.229336 | -0.15978 | 0.93603 |
| FAM241A | 8.049325 | 0.008123 | 0.999532 | | 9.209406 | 0.077726 | 0.942851 | | 8.2039 | 0.099197 | 0.932589 |
| GOLGA1 | 3.884249 | -0.06793 | 0.999532 | | 4.678328 | -0.11303 | 0.942851 | | 3.698471 | -0.00379 | 0.998898 |
| SFT2D3 | 2.08649 | 0.275171 | 0.999532 | | 2.097205 | -0.14871 | 0.942851 | | 1.872421 | -0.40322 | 0.811123 |
| VPS33A | 3.905888 | -0.09313 | 0.999532 | | 4.218238 | -0.10662 | 0.942851 | | 3.483702 | 0.003269 | 0.999307 |
| NUP43 | 4.599307 | 0.357499 | 0.999532 | | 4.530734 | 0.099573 | 0.942851 | | 4.502924 | -0.24333 | 0.837256 |
| ANGPT1 | 2.976856 | -0.3536 | 0.999532 | | 3.450943 | -0.11823 | 0.942851 | | 2.689599 | 0.239289 | 0.88476 |
| KCNJ5 | 2.687061 | 0.287568 | 0.999532 | | 2.592706 | -0.1389 | 0.942851 | | 2.615441 | -0.4111 | 0.793131 |
| C5orf60 | 2.140516 | 0.467082 | 0.999532 | | 1.963057 | -0.15369 | 0.942851 | | 1.997153 | -0.60262 | 0.701419 |
| GRPR | 2.040807 | 0.470971 | 0.999532 | | 1.976773 | -0.14986 | 0.942851 | | 1.916901 | -0.59787 | 0.686214 |
| ANKRD65 | 2.69214 | -0.00124 | 0.999532 | | 3.170968 | 0.123014 | 0.942851 | | 2.678873 | 0.144036 | 0.938429 |
| ATE1 | 3.617127 | -0.18778 | 0.999532 | | 4.674659 | 0.102519 | 0.942851 | | 3.602833 | 0.31815 | 0.802421 |
| CHRM1 | 3.130242 | 0.160235 | 0.999532 | | 3.180277 | -0.12867 | 0.942851 | | 2.995694 | -0.2756 | 0.864245 |
| EBF1 | 8.698013 | -0.027 | 0.999532 | | 9.8231 | 0.080067 | 0.942851 | | 8.650679 | 0.126727 | 0.914687 |
| LEUTX | 2.052948 | 0.548697 | 0.999532 | | 1.749627 | -0.16496 | 0.942851 | | 1.905013 | -0.69686 | 0.660958 |
| SPTY2D1 | 2.424338 | -0.02531 | 0.999532 | | 2.593571 | -0.13613 | 0.942851 | | 2.242198 | -0.09706 | 0.966652 |
| ZNF546 | 1.85666 | 0.127345 | 0.999532 | | 2.127652 | 0.150465 | 0.942851 | | 1.987054 | 0.041621 | 0.990757 |
| SPANXA1 | 10.51475 | 0.113829 | 0.999532 | | 11.19718 | -0.09058 | 0.942851 | | 10.24965 | -0.19735 | 0.863134 |
| CLASP2 | 3.045829 | 0.09903 | 0.999532 | | 3.11645 | -0.12847 | 0.942851 | | 2.680218 | -0.2013 | 0.903339 |
| RPS6KA2 | 4.2841 | 0.160232 | 0.999532 | | 4.618042 | 0.099718 | 0.942851 | | 4.117008 | -0.04643 | 0.979089 |
| SMURF2 | 3.219617 | -0.04019 | 0.999532 | | 3.50342 | -0.11407 | 0.942851 | | 3.04613 | -0.05651 | 0.978282 |
| TMTC4 | 2.724525 | 0.057473 | 0.999532 | | 2.9928 | -0.12677 | 0.942851 | | 2.434837 | -0.16338 | 0.92697 |
| ANKRD50 | 5.059811 | 0.080856 | 0.999532 | | 5.724229 | 0.098448 | 0.942851 | | 5.117941 | 0.041362 | 0.979466 |
| FAM219A | 3.065415 | 0.249598 | 0.999532 | | 3.09122 | -0.12352 | 0.942851 | | 2.985662 | -0.35769 | 0.791576 |
| ZNF626 | 2.260756 | -0.12679 | 0.999532 | | 2.677344 | -0.12811 | 0.942851 | | 2.153356 | 0.022468 | 0.995112 |
| NIT1 | 3.955279 | -0.31662 | 0.999532 | | 4.548864 | -0.10462 | 0.942851 | | 3.517411 | 0.215777 | 0.883116 |
| CCDC92 | 3.825095 | -0.19789 | 0.999532 | | 4.610482 | 0.104825 | 0.942851 | | 3.585179 | 0.323547 | 0.792538 |
| CAMK2D | 8.456583 | -0.29841 | 0.999532 | | 10.79972 | 0.072184 | 0.942851 | | 8.113441 | 0.387989 | 0.623462 |
| RNPS1 | 6.191761 | -0.03425 | 0.999532 | | 7.322623 | 0.084568 | 0.942851 | | 5.899485 | 0.136437 | 0.903054 |
| AKIRIN2 | 5.576059 | 0.093415 | 0.999532 | | 5.513994 | -0.09262 | 0.942851 | | 5.085646 | -0.19455 | 0.888637 |
| SORBS1 | 3.038397 | -0.11538 | 0.999532 | | 3.451984 | -0.1171 | 0.942851 | | 2.765716 | 0.015572 | 0.996681 |
| PRSS2 | 2.304351 | 0.216882 | 0.999532 | | 2.257825 | -0.14749 | 0.942851 | | 2.091027 | -0.34984 | 0.846488 |
| SESN3 | 3.253977 | -0.01967 | 0.999532 | | 3.483508 | -0.11218 | 0.942851 | | 2.949145 | -0.07458 | 0.971113 |
| MIEN1 | 3.183611 | -0.02383 | 0.999532 | | 3.539492 | -0.11297 | 0.942851 | | 3.019373 | -0.06938 | 0.972758 |
| REG3A | 3.335169 | 0.35758 | 0.999532 | | 3.294035 | -0.14141 | 0.942851 | | 3.18079 | -0.46905 | 0.771685 |
| USP30 | 2.685343 | 0.806288 | 0.999532 | | 2.43385 | 0.141178 | 0.942851 | | 2.760828 | -0.64757 | 0.603483 |
| SEC61A2 | 2.411415 | 0.357623 | 0.999532 | | 2.333517 | 0.136247 | 0.942851 | | 2.421746 | -0.20176 | 0.910105 |
| FA2H | 2.505895 | 0.284617 | 0.999532 | | 2.708858 | -0.13501 | 0.942851 | | 2.32855 | -0.39719 | 0.791576 |
| RPP14 | 2.610169 | 0.438409 | 0.999532 | | 2.511058 | 0.135083 | 0.942851 | | 2.461941 | -0.28224 | 0.861119 |
| RPE65 | 2.660035 | -0.24637 | 0.999532 | | 3.311764 | -0.14871 | 0.942851 | | 2.543846 | 0.119159 | 0.959765 |
| FAM89A | 4.167165 | 0.082516 | 0.999532 | | 4.265568 | -0.10343 | 0.942851 | | 3.683916 | -0.16996 | 0.903054 |
| MSH4 | 2.061606 | -0.04522 | 0.999532 | | 2.239004 | -0.1466 | 0.942851 | | 1.833238 | -0.08258 | 0.974421 |
| NECTIN1 | 2.807883 | 0.103066 | 0.999532 | | 3.107314 | 0.121364 | 0.942851 | | 2.796074 | 0.032448 | 0.991159 |
| RIN1 | 3.521665 | -0.27665 | 0.999532 | | 3.968704 | -0.10727 | 0.942851 | | 3.131304 | 0.180671 | 0.906811 |
| TRIQK | 2.181208 | 0.264296 | 0.999532 | | 2.339968 | 0.137333 | 0.942851 | | 2.194843 | -0.10646 | 0.96129 |
| UBE2Q1 | 5.49454 | 0.069288 | 0.999532 | | 6.015565 | 0.092454 | 0.942851 | | 5.208039 | 0.038884 | 0.982809 |
| SMIM8 | 2.919756 | 0.367689 | 0.999532 | | 3.081035 | 0.129778 | 0.942851 | | 3.006408 | -0.22123 | 0.892225 |
| HOGA1 | 3.279856 | 0.50541 | 0.999532 | | 2.806593 | -0.12921 | 0.942851 | | 2.985144 | -0.61827 | 0.616301 |
| LRRC58 | 2.828626 | -0.29961 | 0.999532 | | 3.518416 | -0.11481 | 0.942931 | | 2.816766 | 0.205989 | 0.900848 |
| SCAP | 5.423461 | 0.099506 | 0.999532 | | 6.173646 | 0.091375 | 0.943312 | | 5.464115 | 0.012666 | 0.996681 |
| TSPAN15 | 4.167634 | -0.49754 | 0.999532 | | 5.410771 | 0.103563 | 0.943398 | | 4.009672 | 0.622006 | 0.552672 |
| XPR1 | 4.640415 | 0.195066 | 0.999532 | | 4.570333 | -0.10154 | 0.943515 | | 4.195002 | -0.27643 | 0.809845 |
| ANKRD13 | 3.705716 | 0.364665 | 0.999532 | | 3.694393 | 0.112887 | 0.944037 | | 3.608273 | -0.23338 | 0.868512 |
| ZFAND4 | 2.800201 | 0.113906 | 0.999532 | | 2.871764 | -0.12764 | 0.944039 | | 2.528373 | -0.22365 | 0.892869 |
| MPP1 | 3.013636 | -0.08772 | 0.999532 | | 3.131141 | -0.11998 | 0.944039 | | 2.682433 | -0.03778 | 0.989638 |
| C6 | 3.068098 | 0.03248 | 0.999532 | | 3.298253 | 0.112789 | 0.944039 | | 2.968325 | 0.096166 | 0.958991 |
| SMIM7 | 7.394468 | -0.08341 | 0.999532 | | 8.913366 | 0.07744 | 0.944212 | | 7.100479 | 0.186727 | 0.862823 |
| KCNE4 | 2.366076 | 0.447256 | 0.999532 | | 2.341891 | 0.136916 | 0.944277 | | 2.448829 | -0.28913 | 0.864245 |
| CROCC | 2.46721 | -0.14376 | 0.999532 | | 2.823559 | -0.13071 | 0.944277 | | 2.240516 | 0.030239 | 0.992286 |
| ACOX1 | 3.72913 | -0.38961 | 0.999532 | | 4.877433 | -0.09853 | 0.944277 | | 3.469646 | 0.319837 | 0.801572 |
| GLO1 | 4.482432 | -0.0094 | 0.999532 | | 5.064006 | 0.095473 | 0.944277 | | 4.38896 | 0.122918 | 0.931942 |
| ARHGAP | 2.509911 | -0.17574 | 0.999532 | | 2.858467 | -0.12645 | 0.944277 | | 2.358201 | 0.070376 | 0.976441 |
| CCSER2 | 6.287632 | -0.05757 | 0.999532 | | 6.988316 | -0.08442 | 0.944277 | | 5.970015 | -0.00558 | 0.997967 |
| WDR38 | 2.481256 | 0.505338 | 0.999532 | | 2.303148 | -0.15158 | 0.944277 | | 2.321136 | -0.63613 | 0.666492 |
| HSD17B4 | 5.739076 | -0.42173 | 0.999532 | | 7.330313 | -0.0804 | 0.944277 | | 5.281899 | 0.362595 | 0.712621 |
| ZNF599 | 2.354356 | 0.237007 | 0.999532 | | 2.354688 | -0.14127 | 0.944277 | | 2.225348 | -0.36218 | 0.827267 |
| YPEL1 | 3.749206 | 0.226359 | 0.999532 | | 3.756507 | -0.12628 | 0.944277 | | 3.316499 | -0.33329 | 0.813143 |
| ACSL3 | 6.457645 | -0.26892 | 0.999532 | | 7.689367 | 0.082634 | 0.944307 | | 6.002222 | 0.350218 | 0.723453 |
| RARB | 2.440592 | -0.25951 | 0.999532 | | 2.967291 | 0.124496 | 0.944307 | | 2.486645 | 0.386513 | 0.809845 |
| RAD51 | 3.025359 | -0.0552 | 0.999532 | | 3.243332 | -0.1299 | 0.944307 | | 2.815661 | -0.05135 | 0.984587 |
| ZNF44 | 2.754753 | 0.408608 | 0.999532 | | 2.530049 | -0.13627 | 0.944467 | | 2.496437 | -0.52543 | 0.699971 |
| GCSH | 3.229169 | -0.08365 | 0.999532 | | 3.887805 | -0.11452 | 0.94458 | | 3.252981 | 0.018512 | 0.996681 |
| MMP8 | 1.738098 | -0.20389 | 0.999532 | | 1.914936 | -0.14645 | 0.944749 | | 1.62365 | 0.072787 | 0.979089 |
| HSF2 | 2.9503 | 0.315104 | 0.999532 | | 2.769166 | -0.12905 | 0.944749 | | 2.717511 | -0.42873 | 0.760955 |
| MMP28 | 3.5453 | -0.091 | 0.999532 | | 4.089579 | -0.11652 | 0.944749 | | 3.253962 | -0.00123 | 0.999588 |
| NSUN2 | 4.194671 | 0.240869 | 0.999532 | | 4.311439 | 0.115616 | 0.944749 | | 3.956548 | -0.08121 | 0.963866 |
| DOP1A | 2.589194 | 0.530201 | 0.999532 | | 2.627589 | 0.137104 | 0.944749 | | 2.631532 | -0.37652 | 0.800503 |
| ZBTB5 | 2.160554 | 0.431849 | 0.999532 | | 2.127172 | 0.139391 | 0.944749 | | 2.210403 | -0.27345 | 0.878619 |
| FCHO1 | 2.051655 | 0.302904 | 0.999532 | | 2.167731 | 0.139411 | 0.944749 | | 2.069294 | -0.14595 | 0.94366 |
| HOMER2 | 2.614742 | 0.284638 | 0.999532 | | 2.601228 | -0.13187 | 0.944749 | | 2.443954 | -0.39818 | 0.793717 |
| KLHL15 | 2.405977 | 0.001471 | 0.999532 | | 2.591186 | -0.128 | 0.944749 | | 2.225024 | -0.11132 | 0.957663 |
| ZNF433 | 2.098685 | 0.473427 | 0.999532 | | 2.125025 | 0.145542 | 0.944749 | | 2.1591 | -0.31008 | 0.865065 |
| MC3R | 1.981881 | -0.15491 | 0.999532 | | 2.140687 | -0.142 | 0.944749 | | 1.804906 | 0.025129 | 0.995241 |
| ATMIN | 3.212423 | 0.382931 | 0.999532 | | 3.124647 | 0.121276 | 0.944749 | | 3.064702 | -0.24043 | 0.874653 |
| PUS1 | 2.500614 | 0.265107 | 0.999532 | | 2.521121 | 0.136249 | 0.944749 | | 2.532666 | -0.11549 | 0.956715 |
| PLEK2 | 1.987698 | 0.193912 | 0.999532 | | 2.00426 | -0.15702 | 0.944749 | | 1.719697 | -0.33128 | 0.865363 |
| VTI1B | 11.87411 | -0.11171 | 0.999532 | | 13.66081 | -0.06139 | 0.944938 | | 11.19211 | 0.072146 | 0.942931 |
| NCOA6 | 6.884627 | 0.083324 | 0.999532 | | 7.551152 | 0.08169 | 0.944938 | | 6.563687 | 0.016175 | 0.993618 |
| NRXN2 | 2.755607 | -0.16315 | 0.999532 | | 3.181246 | -0.12214 | 0.944938 | | 2.563673 | 0.057745 | 0.979239 |
| TIMM22 | 2.722142 | 0.327581 | 0.999532 | | 2.511168 | -0.13294 | 0.944938 | | 2.424707 | -0.44041 | 0.752562 |
| KIN | 2.663715 | 0.159744 | 0.999532 | | 2.801434 | 0.12647 | 0.944938 | | 2.567046 | -0.01466 | 0.996681 |
| CELA3B | 2.249448 | 0.459896 | 0.999532 | | 2.121968 | -0.14332 | 0.944938 | | 2.141014 | -0.58198 | 0.693099 |
| GFPT2 | 1.970941 | -0.03359 | 0.999532 | | 2.353229 | 0.139903 | 0.944938 | | 1.886919 | 0.182015 | 0.931827 |
| ZNF500 | 3.273587 | -0.20557 | 0.999532 | | 4.012968 | -0.11139 | 0.944938 | | 2.949626 | 0.118745 | 0.946512 |
| FAM43A | 5.858027 | 0.572707 | 0.999532 | | 5.302973 | -0.09737 | 0.944956 | | 5.625022 | -0.64325 | 0.458377 |
| PRPF4 | 3.772186 | 0.309611 | 0.999532 | | 3.814312 | 0.11124 | 0.944956 | | 3.581245 | -0.17834 | 0.90332 |
| SYNM | 4.552924 | -0.06586 | 0.999532 | | 5.275158 | -0.10088 | 0.944956 | | 4.404931 | -0.01716 | 0.995112 |
| NELFCD | 3.442739 | -0.08065 | 0.999532 | | 3.959361 | -0.10973 | 0.944956 | | 3.021511 | -0.02396 | 0.993435 |
| MAL2 | 8.341694 | 0.094607 | 0.999532 | | 8.94399 | 0.081478 | 0.944956 | | 8.090574 | -0.00802 | 0.996891 |
| TLR5 | 2.122154 | 0.416202 | 0.999532 | | 2.108416 | -0.14354 | 0.944956 | | 1.993279 | -0.5322 | 0.732486 |
| H2BC6 | 3.309619 | 0.171248 | 0.999532 | | 3.546595 | 0.114779 | 0.944956 | | 3.145144 | -0.03372 | 0.989646 |
| YTHDC2 | 2.384865 | -0.6105 | 0.999532 | | 3.11789 | -0.12395 | 0.944956 | | 2.133064 | 0.503232 | 0.736302 |
| OGFR | 9.099917 | -0.0611 | 0.999532 | | 10.00864 | -0.07178 | 0.944956 | | 8.323062 | 0.006601 | 0.996891 |
| PNPO | 4.866789 | 0.076635 | 0.999532 | | 5.548111 | 0.093501 | 0.944956 | | 4.663525 | 0.035796 | 0.985693 |
| ZNF304 | 3.415985 | 0.028505 | 0.999532 | | 3.897933 | 0.113209 | 0.944956 | | 3.560961 | 0.102648 | 0.956011 |
| ATG13 | 4.106197 | 0.035203 | 0.999532 | | 4.67959 | 0.097978 | 0.944999 | | 4.092638 | 0.080553 | 0.960773 |
| IQCB1 | 5.023927 | -0.02587 | 0.999532 | | 6.161976 | 0.093214 | 0.94505 | | 5.282557 | 0.12631 | 0.920527 |
| KLHDC8B | 7.485241 | 0.042648 | 0.999532 | | 8.429826 | 0.075681 | 0.945105 | | 7.333316 | 0.049969 | 0.972039 |
| EPHA7 | 2.747418 | 0.599767 | 0.999532 | | 2.579535 | 0.13261 | 0.94515 | | 2.724265 | -0.45094 | 0.749999 |
| PTGR2 | 4.553034 | 0.241533 | 0.999532 | | 4.368957 | -0.09651 | 0.945194 | | 4.243109 | -0.31921 | 0.77448 |
| SCYL3 | 2.957873 | -0.02153 | 0.999532 | | 3.023121 | -0.12646 | 0.945194 | | 2.767189 | -0.08659 | 0.971732 |
| TGFBR1 | 3.814916 | -0.2561 | 0.999532 | | 4.606323 | 0.098773 | 0.945194 | | 3.775127 | 0.372354 | 0.749206 |
| UTS2B | 1.977318 | 0.254452 | 0.999532 | | 1.976419 | -0.15614 | 0.945194 | | 1.939633 | -0.39503 | 0.836012 |
| NIPSNAP | 2.285117 | 0.109548 | 0.999532 | | 2.602256 | 0.130711 | 0.945194 | | 2.314629 | 0.039687 | 0.989437 |
| DYNLRB1 | 11.21986 | -0.16576 | 0.999532 | | 12.92189 | -0.06572 | 0.945222 | | 10.21163 | 0.11482 | 0.905952 |
| ANGPT4 | 2.167698 | 0.199619 | 0.999532 | | 2.323658 | -0.1429 | 0.945222 | | 2.07647 | -0.31875 | 0.863142 |
| ETV5 | 3.235285 | -0.08966 | 0.999532 | | 3.774116 | -0.11297 | 0.945371 | | 2.935241 | -0.00471 | 0.998586 |
| PFAS | 3.403265 | 0.179329 | 0.999532 | | 3.407528 | -0.11067 | 0.945371 | | 3.157171 | -0.27468 | 0.846786 |
| MTRNR2L | 2.94658 | 0.449228 | 0.999532 | | 3.066232 | 0.143819 | 0.945371 | | 2.966852 | -0.30586 | 0.843043 |
| CTIF | 5.022199 | 0.401177 | 0.999532 | | 5.125885 | 0.097917 | 0.945371 | | 4.978707 | -0.29047 | 0.784263 |
| TNXB | 2.122133 | -0.09554 | 0.999532 | | 2.112797 | -0.13938 | 0.945482 | | 1.9104 | -0.02587 | 0.994722 |
| APOD | 6.776187 | 0.34052 | 0.999532 | | 6.821544 | -0.10972 | 0.945482 | | 6.222632 | -0.44972 | 0.647218 |
| ROBO2 | 2.286057 | 0.486309 | 0.999532 | | 2.019893 | -0.13797 | 0.945482 | | 2.066444 | -0.60667 | 0.663009 |
| RALGAPA | 5.276416 | -0.10321 | 0.999532 | | 6.258268 | 0.088264 | 0.945482 | | 5.210366 | 0.209651 | 0.862006 |
| RMND5B | 2.345613 | 0.312568 | 0.999532 | | 2.331944 | -0.13626 | 0.945482 | | 2.084251 | -0.42553 | 0.786592 |
| C10orf12 | 2.390161 | 0.699446 | 0.999532 | | 1.88119 | -0.14847 | 0.945482 | | 2.238784 | -0.83635 | 0.542124 |
| SLC7A6O | 2.377672 | -0.06782 | 0.999532 | | 2.993241 | 0.121113 | 0.945482 | | 2.394417 | 0.208983 | 0.90275 |
| DTNBP1 | 2.293899 | 0.114584 | 0.999532 | | 2.765566 | 0.130353 | 0.94549 | | 2.372893 | 0.038018 | 0.991103 |
| ABL2 | 2.073432 | 0.164349 | 0.999532 | | 2.346536 | 0.136567 | 0.94549 | | 2.117198 | -0.01145 | 0.996891 |
| FSCN2 | 2.832214 | 0.418847 | 0.999532 | | 2.903632 | 0.133034 | 0.945686 | | 2.934985 | -0.28181 | 0.865065 |
| LEMD2 | 5.12919 | 0.210345 | 0.999532 | | 5.605145 | 0.092083 | 0.945691 | | 4.966125 | -0.09713 | 0.945129 |
| CAPN1 | 3.838382 | -0.07774 | 0.999532 | | 4.24411 | -0.10484 | 0.945691 | | 3.599691 | -0.00478 | 0.998522 |
| NOVA2 | 4.270543 | -0.17414 | 0.999532 | | 5.111942 | 0.097066 | 0.945691 | | 4.3931 | 0.285508 | 0.82977 |
| DNAJC11 | 4.85652 | -0.0707 | 0.999532 | | 5.511706 | -0.08937 | 0.945787 | | 4.43811 | 0.001549 | 0.999385 |
| CD177 | 2.536262 | -0.01178 | 0.999532 | | 2.892528 | -0.15012 | 0.945787 | | 2.424773 | -0.14494 | 0.945129 |
| ISOC1 | 2.65019 | 0.038715 | 0.999532 | | 3.201723 | 0.112508 | 0.945787 | | 2.713144 | 0.098231 | 0.960773 |
| KCNIP2 | 2.520936 | 0.361242 | 0.999532 | | 2.349573 | -0.13573 | 0.945787 | | 2.42722 | -0.48764 | 0.749222 |
| GEMIN6 | 2.808696 | 0.007743 | 0.999532 | | 3.12339 | -0.11683 | 0.945787 | | 2.654822 | -0.09859 | 0.961643 |
| NCAPH2 | 4.818329 | -0.13961 | 0.999532 | | 5.699877 | 0.09427 | 0.945787 | | 4.604414 | 0.255799 | 0.827267 |
| CCDC17 | 2.34624 | 0.311152 | 0.999532 | | 2.499429 | 0.136307 | 0.945787 | | 2.370959 | -0.1558 | 0.939759 |
| CLEC6A | 2.300577 | -0.18281 | 0.999532 | | 2.608335 | -0.13153 | 0.945787 | | 2.193227 | 0.070476 | 0.978229 |
| CNTNAP2 | 2.102263 | -0.18094 | 0.999532 | | 2.532239 | -0.13512 | 0.945787 | | 1.970857 | 0.074273 | 0.977473 |
| KLHDC4 | 2.449566 | 0.404198 | 0.999532 | | 2.471609 | 0.127023 | 0.945787 | | 2.423291 | -0.25955 | 0.87591 |
| PSD3 | 2.037754 | 0.088301 | 0.999532 | | 2.095062 | -0.14694 | 0.945787 | | 1.991999 | -0.2186 | 0.918694 |
| LLPH | 4.20369 | -0.05768 | 0.999532 | | 4.522749 | -0.10403 | 0.945787 | | 3.797663 | -0.02867 | 0.991134 |
| OXGR1 | 1.944011 | 0.417125 | 0.999532 | | 1.945472 | 0.150816 | 0.945787 | | 2.024246 | -0.24863 | 0.900848 |
| RGS10 | 7.145808 | -0.30144 | 0.999532 | | 8.915511 | 0.074986 | 0.945787 | | 6.903394 | 0.395968 | 0.630705 |
| SRPK1 | 3.186032 | 0.128296 | 0.999532 | | 3.510861 | 0.112565 | 0.945815 | | 3.365209 | -0.00264 | 0.999385 |
| SNRNP48 | 3.598243 | 0.324357 | 0.999532 | | 3.272549 | -0.12013 | 0.945989 | | 3.44067 | -0.43526 | 0.743576 |
| USP15 | 5.49957 | -0.15138 | 0.999532 | | 6.685062 | 0.084813 | 0.945989 | | 5.492211 | 0.254126 | 0.814483 |
| PWWP2A | 3.255729 | 0.114563 | 0.999532 | | 3.381774 | -0.1131 | 0.946024 | | 3.004086 | -0.21052 | 0.894612 |
| VPS28 | 7.974554 | -0.17841 | 0.999532 | | 10.04611 | 0.070055 | 0.946137 | | 7.569504 | 0.261369 | 0.7719 |
| CWF19L1 | 3.080276 | 0.596398 | 0.999532 | | 2.741763 | -0.12947 | 0.946196 | | 2.861372 | -0.70581 | 0.540342 |
| DLEU7 | 2.467761 | 0.764138 | 0.999532 | | 2.271317 | 0.139369 | 0.946196 | | 2.484876 | -0.60542 | 0.657467 |
| DEAF1 | 3.169528 | -0.18044 | 0.999532 | | 3.668978 | -0.10884 | 0.946265 | | 2.926682 | 0.090562 | 0.963309 |
| LDLRAD3 | 2.799542 | 0.050189 | 0.999532 | | 3.024618 | -0.11769 | 0.946265 | | 2.475536 | -0.14982 | 0.934701 |
| HDHD2 | 2.768571 | -0.30465 | 0.999532 | | 3.316321 | -0.11029 | 0.94636 | | 2.619382 | 0.213329 | 0.894612 |
| GMPS | 3.581462 | -0.01507 | 0.999532 | | 4.067221 | -0.10424 | 0.946385 | | 3.379908 | -0.07236 | 0.970498 |
| KCNK3 | 1.943881 | 0.223918 | 0.999532 | | 2.003768 | -0.14807 | 0.946385 | | 1.83163 | -0.35134 | 0.854633 |
| SH3KBP1 | 3.444936 | 0.024407 | 0.999532 | | 3.829987 | 0.106891 | 0.946385 | | 3.287489 | 0.095644 | 0.958191 |
| FOXE1 | 2.834353 | -0.01987 | 0.999532 | | 3.528312 | 0.118113 | 0.946385 | | 3.031584 | 0.165478 | 0.92697 |
| POLR3H | 3.550368 | 0.279116 | 0.999532 | | 3.292081 | -0.11187 | 0.946385 | | 3.276469 | -0.37898 | 0.776321 |
| DENND6B | 2.939563 | 0.278024 | 0.999532 | | 3.093087 | 0.115668 | 0.946385 | | 2.873699 | -0.14235 | 0.932589 |
| MYPOP | 2.194139 | -0.10692 | 0.999532 | | 2.687349 | 0.13581 | 0.946513 | | 2.122076 | 0.278868 | 0.88476 |
| SETD4 | 3.148722 | 0.604918 | 0.999532 | | 2.897762 | 0.119883 | 0.946598 | | 3.140302 | -0.46852 | 0.699971 |
| ZBTB3 | 2.359591 | -0.17219 | 0.999532 | | 2.748683 | -0.12716 | 0.946598 | | 2.235912 | 0.06567 | 0.978463 |
| IGIP | 3.231976 | 0.400751 | 0.999532 | | 3.287936 | 0.10965 | 0.946598 | | 3.249998 | -0.27407 | 0.836453 |
| AKAP8 | 5.384873 | 0.028375 | 0.999532 | | 5.679812 | -0.09046 | 0.946598 | | 4.877863 | -0.1023 | 0.943112 |
| VPS26A | 4.575384 | -0.35595 | 0.999532 | | 5.649881 | 0.087685 | 0.946598 | | 4.349621 | 0.446374 | 0.676565 |
| SPTBN4 | 1.902959 | -0.33058 | 0.999532 | | 2.206992 | -0.14062 | 0.946598 | | 1.800847 | 0.202583 | 0.929335 |
| NSMCE3 | 5.843721 | 0.122376 | 0.999532 | | 6.393408 | 0.086592 | 0.946598 | | 5.735694 | -0.01685 | 0.993753 |
| CENPS | 2.306862 | 0.221495 | 0.999532 | | 2.157752 | -0.13486 | 0.946598 | | 2.096631 | -0.34182 | 0.843043 |
| FGF5 | 2.427258 | 0.348374 | 0.999532 | | 2.281734 | -0.13462 | 0.946731 | | 2.317435 | -0.46166 | 0.791466 |
| ZNF250 | 2.716078 | 0.421115 | 0.999532 | | 2.694228 | 0.126498 | 0.946731 | | 2.580287 | -0.27097 | 0.864923 |
| ZNF274 | 3.981666 | -0.02713 | 0.999532 | | 4.262836 | -0.10261 | 0.946731 | | 3.639744 | -0.06012 | 0.974808 |
| MRPL34 | 3.734032 | 0.713236 | 0.999532 | | 3.079873 | 0.113714 | 0.946731 | | 3.636457 | -0.58043 | 0.602151 |
| VNN2 | 2.891545 | 0.211353 | 0.999532 | | 2.940633 | -0.12803 | 0.946731 | | 2.704148 | -0.31911 | 0.842754 |

| | | | | | | | | | | | |
|---|---|---|---|---|---|---|---|---|---|---|---|
| C10orf10 | 1.987588 | -0.40984 | 0.999532 | | 2.550588 | -0.13578 | 0.946731 | | 1.897921 | 0.299818 | 0.881837 |
| MBOAT7 | 3.118621 | -0.33336 | 0.999532 | | 3.822159 | -0.10288 | 0.946731 | | 2.834358 | 0.247366 | 0.867976 |
| NLRP4 | 2.008152 | 0.74027 | 0.999532 | | 1.760072 | -0.1552 | 0.946731 | | 1.862109 | -0.87361 | 0.533876 |
| KAZALD1 | 2.498572 | 0.260546 | 0.999532 | | 2.506367 | 0.130334 | 0.946731 | | 2.351362 | -0.11329 | 0.958191 |
| OR2A14 | 5.065933 | 0.424379 | 0.999532 | | 5.221768 | 0.110406 | 0.946731 | | 5.171263 | -0.30138 | 0.808547 |
| PM20D2 | 4.498245 | 0.257556 | 0.999532 | | 4.968209 | 0.097159 | 0.946731 | | 4.431203 | -0.13605 | 0.920486 |
| SST | 4.116522 | 1.362844 | 0.206846 | | 2.788404 | -0.13513 | 0.946731 | | 3.931822 | -1.456 | 0.107433 |
| DPM3 | 4.046373 | 0.072732 | 0.999532 | | 4.365313 | -0.10113 | 0.946731 | | 3.79953 | -0.15246 | 0.918694 |
| PGM2 | 4.645567 | -0.18255 | 0.999532 | | 5.815504 | 0.086525 | 0.946731 | | 4.520105 | 0.285765 | 0.79686 |
| CCNA2 | 2.307295 | 0.483009 | 0.999532 | | 2.271618 | 0.136949 | 0.946731 | | 2.3633 | -0.32934 | 0.846488 |
| ZNF460 | 6.540912 | 0.089771 | 0.999532 | | 7.262473 | 0.080461 | 0.946731 | | 6.355834 | 0.009012 | 0.996891 |
| PRSS55 | 2.022101 | 0.278682 | 0.999532 | | 2.076117 | -0.14145 | 0.946731 | | 1.870684 | -0.39223 | 0.824012 |
| CERS6 | 3.659111 | 0.226134 | 0.999532 | | 4.096131 | 0.109196 | 0.946731 | | 3.583995 | -0.09074 | 0.959765 |
| IKZF4 | 2.484104 | 0.523257 | 0.999532 | | 2.374512 | 0.132864 | 0.947077 | | 2.553924 | -0.37124 | 0.813438 |
| PLEKHM | 2.711772 | -0.73868 | 0.999532 | | 3.677156 | -0.10622 | 0.947084 | | 2.448495 | 0.650518 | 0.628515 |
| NEK6 | 3.871114 | 0.453659 | 0.999532 | | 3.818526 | 0.105546 | 0.947117 | | 3.844643 | -0.32884 | 0.792416 |
| GRIN2A | 2.037757 | -0.25156 | 0.999532 | | 2.437753 | 0.125882 | 0.947295 | | 2.047854 | 0.391223 | 0.816618 |
| PURG | 2.407512 | 0.858092 | 0.999532 | | 1.956827 | 0.146381 | 0.947295 | | 2.426251 | -0.6948 | 0.630705 |
| RBM39 | 17.19494 | 0.058941 | 0.999532 | | 18.09349 | -0.05685 | 0.947379 | | 15.49814 | -0.10461 | 0.90275 |
| RAB6C | 3.64717 | -0.11998 | 0.999532 | | 4.321851 | 0.113315 | 0.947543 | | 3.663157 | 0.256961 | 0.856551 |
| MCC | 3.660006 | 0.18236 | 0.999532 | | 3.965108 | 0.105249 | 0.947612 | | 3.680976 | -0.05889 | 0.974808 |
| FRS2 | 5.584562 | 0.116308 | 0.999532 | | 5.760113 | 0.088422 | 0.947617 | | 5.376362 | -0.02529 | 0.991273 |
| CCDC82 | 3.782652 | 0.628339 | 0.999532 | | 3.558794 | 0.116261 | 0.947617 | | 3.911106 | -0.50002 | 0.678541 |
| CDKL2 | 2.176968 | -0.31411 | 0.999532 | | 2.666477 | -0.12643 | 0.947617 | | 2.041623 | 0.210021 | 0.914929 |
| NBEA | 2.336944 | -0.63527 | 0.999532 | | 2.972003 | -0.11901 | 0.947617 | | 2.104907 | 0.5342 | 0.724411 |
| NLRP14 | 1.939694 | -0.03352 | 0.999532 | | 2.238316 | 0.132083 | 0.947617 | | 1.980348 | 0.185194 | 0.927668 |
| FAM114A | 3.628747 | 0.167909 | 0.999532 | | 3.604143 | -0.11013 | 0.947617 | | 3.264356 | -0.25746 | 0.859898 |
| GOT2 | 4.843028 | 0.058451 | 0.999532 | | 5.120707 | -0.1012 | 0.947617 | | 4.350792 | -0.13573 | 0.92697 |
| ESX1 | 2.476037 | -0.11908 | 0.999532 | | 2.886294 | -0.12105 | 0.947617 | | 2.262346 | 0.01625 | 0.996681 |
| C6orf120 | 3.743676 | 0.559353 | 0.999532 | | 3.812208 | 0.103207 | 0.947617 | | 3.713963 | -0.43154 | 0.700481 |
| LDOC1 | 7.846783 | 0.480997 | 0.999532 | | 7.232184 | 0.083721 | 0.947617 | | 7.59966 | -0.38702 | 0.666566 |
| DCLK2 | 3.990572 | -0.00176 | 0.999532 | | 4.19124 | -0.09799 | 0.947617 | | 3.687119 | -0.08171 | 0.963309 |
| STPG1 | 2.217839 | 0.274105 | 0.999532 | | 2.413542 | 0.134749 | 0.947617 | | 2.252601 | -0.1211 | 0.955834 |
| GNG4 | 2.296484 | -0.02066 | 0.999532 | | 2.547017 | -0.12851 | 0.947617 | | 2.13942 | -0.08939 | 0.97151 |
| TUBGCP | 4.344391 | -0.07032 | 0.999532 | | 5.077054 | 0.095885 | 0.947617 | | 4.434265 | 0.172019 | 0.903205 |
| CLC | 2.151784 | 0.877772 | 0.999532 | | 1.760852 | -0.15803 | 0.947617 | | 2.015875 | -1.01297 | 0.456464 |
| RGS7BP | 1.61426 | 0.051956 | 0.999532 | | 1.945881 | 0.14924 | 0.947617 | | 1.754704 | 0.119369 | 0.96556 |
| SPATA33 | 2.490864 | -0.04845 | 0.999532 | | 3.159996 | 0.124466 | 0.947617 | | 2.620667 | 0.206297 | 0.911581 |
| SH3BP4 | 4.501929 | -0.04541 | 0.999532 | | 4.711519 | -0.0917 | 0.947617 | | 4.136976 | -0.04285 | 0.983848 |
| MTG1 | 8.815508 | -0.13385 | 0.999532 | | 10.74609 | 0.064865 | 0.947617 | | 8.39608 | 0.2199 | 0.803337 |
| FAM110D | 3.359036 | 0.322649 | 0.999532 | | 3.452301 | 0.10954 | 0.947617 | | 3.265356 | -0.19343 | 0.896374 |
| ATP6V1B | 5.780297 | 0.288354 | 0.999532 | | 6.002437 | 0.08639 | 0.947617 | | 5.597212 | -0.18561 | 0.872561 |
| MGAM | 3.26936 | 0.007235 | 0.999532 | | 3.886521 | 0.110084 | 0.947617 | | 3.300386 | 0.11754 | 0.945129 |
| UTP11 | 4.22938 | 0.058967 | 0.999532 | | 4.47864 | -0.09826 | 0.947617 | | 3.709133 | -0.13191 | 0.929604 |
| SOAT1 | 3.155257 | -0.44902 | 0.999532 | | 4.207463 | 0.099058 | 0.947617 | | 3.26466 | 0.574397 | 0.632923 |
| TMEM184 | 3.586015 | 0.017724 | 0.999532 | | 4.016121 | -0.10288 | 0.947617 | | 3.32866 | -0.10005 | 0.955834 |
| KRTAP5- | 2.6635 | -0.00916 | 0.999532 | | 2.802167 | -0.12199 | 0.947617 | | 2.450377 | -0.09641 | 0.965649 |
| B3GALNT | 3.179944 | -0.15024 | 0.999532 | | 3.981472 | 0.11203 | 0.947617 | | 3.067948 | 0.282302 | 0.846373 |
| KIF6 | 2.008056 | 0.160774 | 0.999532 | | 2.131291 | 0.13501 | 0.947617 | | 2.027629 | -0.00937 | 0.997967 |
| TMEM178 | 3.673676 | -0.19953 | 0.999532 | | 4.477916 | 0.103839 | 0.947617 | | 3.482394 | 0.324803 | 0.810753 |
| CFAP94 | 1.832601 | 0.036739 | 0.999532 | | 1.797203 | -0.14793 | 0.947617 | | 1.652751 | -0.16871 | 0.944202 |
| MYL3 | 3.356902 | 0.614993 | 0.999532 | | 3.017313 | 0.11748 | 0.947617 | | 3.280406 | -0.48172 | 0.700381 |
| GSTO2 | 3.035959 | 0.311786 | 0.999532 | | 3.137949 | 0.112212 | 0.947617 | | 2.939211 | -0.18275 | 0.907163 |
| COL14A1 | 2.60514 | -0.59761 | 0.999532 | | 3.482645 | 0.114764 | 0.947617 | | 2.507967 | 0.733088 | 0.568812 |
| SERPINA | 1.76703 | 0.123407 | 0.999532 | | 1.829111 | -0.14346 | 0.947617 | | 1.698513 | -0.24785 | 0.903054 |
| PLEK | 2.260222 | 0.418186 | 0.999532 | | 2.20063 | 0.13861 | 0.947617 | | 2.240286 | -0.25931 | 0.892869 |
| SSTR5 | 2.024764 | -0.20903 | 0.999532 | | 2.345057 | -0.12805 | 0.947617 | | 1.757643 | 0.097582 | 0.969614 |
| ZNF875 | 3.017841 | -0.44824 | 0.999532 | | 3.694291 | 0.101772 | 0.947851 | | 3.000133 | 0.580091 | 0.649779 |
| KIAA089 | 2.32746 | 0.325237 | 0.999532 | | 2.359976 | -0.13692 | 0.947851 | | 2.353316 | -0.44785 | 0.780676 |
| DTWD2 | 2.467744 | -0.13567 | 0.999532 | | 2.962705 | 0.118437 | 0.947851 | | 2.440021 | 0.268562 | 0.87892 |
| LINGO3 | 2.071666 | 0.350384 | 0.999532 | | 2.026235 | 0.138009 | 0.947949 | | 2.131709 | -0.19969 | 0.920181 |
| HELZ | 3.702641 | 0.016626 | 0.999532 | | 4.013669 | 0.105331 | 0.947949 | | 3.418 | 0.115802 | 0.945033 |
| CYP27A1 | 2.770052 | 0.334114 | 0.999532 | | 2.884588 | 0.117646 | 0.947949 | | 2.74882 | -0.1985 | 0.903329 |
| DPH6 | 2.244751 | 0.00441 | 0.999532 | | 2.360732 | -0.12965 | 0.947949 | | 2.04971 | -0.11465 | 0.958889 |
| PLPP1 | 46.24615 | -0.2519 | 0.999532 | | 57.23631 | 0.05105 | 0.948571 | | 44.95009 | 0.324526 | 0.556502 |
| ZNRF3 | 6.269034 | 0.346016 | 0.999532 | | 6.343328 | 0.082351 | 0.948571 | | 6.030607 | -0.2441 | 0.816112 |
| SLC5A3 | 2.769946 | 0.408642 | 0.999532 | | 2.708741 | -0.12668 | 0.948571 | | 2.466487 | -0.51223 | 0.701691 |
| MAPK7 | 3.462875 | -0.33368 | 0.999532 | | 4.141133 | -0.12205 | 0.948842 | | 3.150034 | 0.228548 | 0.88885 |
| BTBD7 | 3.135326 | 0.063007 | 0.999532 | | 3.434898 | 0.108768 | 0.948842 | | 3.086525 | 0.065497 | 0.974421 |
| ZNF730 | 2.208261 | 0.034461 | 0.999532 | | 2.474654 | -0.12505 | 0.94899 | | 2.151956 | -0.13701 | 0.947126 |
| TPRKB | 5.838965 | -0.03293 | 0.999532 | | 6.797702 | 0.081097 | 0.94899 | | 5.67612 | 0.134664 | 0.910105 |
| ERV3-1 | 3.731526 | 0.319586 | 0.999532 | | 3.941957 | 0.104085 | 0.94899 | | 3.858214 | -0.19283 | 0.895807 |
| S100A4 | 15.04625 | -0.57653 | 0.999532 | | 19.45825 | -0.1022 | 0.94899 | | 13.8913 | 0.49738 | 0.552672 |
| FLT3 | 2.370643 | 0.904513 | 0.999532 | | 2.000177 | 0.138108 | 0.94899 | | 2.354887 | -0.74759 | 0.552672 |
| MTMR4 | 3.883146 | 0.345093 | 0.999532 | | 3.852117 | 0.106941 | 0.949456 | | 3.805516 | -0.21446 | 0.879947 |
| OR10K1 | 1.746343 | 0.282776 | 0.999532 | | 1.725955 | -0.15054 | 0.949456 | | 1.627901 | -0.41599 | 0.828643 |
| OLFM2 | 2.017358 | -0.03832 | 0.999532 | | 2.283485 | -0.12944 | 0.949567 | | 1.946013 | -0.07114 | 0.978004 |
| UEVLD | 3.295376 | 0.027402 | 0.999532 | | 3.328979 | -0.10972 | 0.949567 | | 3.092735 | -0.10332 | 0.956322 |
| WNT2B | 2.799622 | 0.006775 | 0.999532 | | 3.008568 | -0.11758 | 0.949567 | | 2.703443 | -0.10584 | 0.96129 |
| IGSF6 | 4.54991 | 0.58104 | 0.999532 | | 3.615602 | -0.10831 | 0.949652 | | 4.190485 | -0.66831 | 0.535662 |
| H2BC17 | 3.312617 | -0.27279 | 0.999532 | | 4.083458 | -0.10326 | 0.949753 | | 3.116949 | 0.194499 | 0.901399 |
| LRRC8A | 4.955828 | -0.06734 | 0.999532 | | 5.696925 | 0.087276 | 0.949753 | | 4.950283 | 0.175218 | 0.897958 |
| KRT80 | 3.0438 | 1.308215 | 0.484521 | | 2.030158 | 0.141371 | 0.949753 | | 3.049207 | -1.15284 | 0.280329 |
| PDLIM3 | 3.168108 | 0.349791 | 0.999532 | | 2.7211 | -0.12201 | 0.949753 | | 2.688744 | -0.45353 | 0.742183 |
| MROH1 | 4.191062 | -0.08484 | 0.999532 | | 4.71688 | -0.09233 | 0.949753 | | 3.755164 | 0.008589 | 0.996891 |
| SPATS2 | 2.150804 | -0.10484 | 0.999532 | | 2.454161 | -0.12542 | 0.949753 | | 2.064304 | -0.00433 | 0.999159 |
| RIC1 | 4.072025 | 0.153935 | 0.999532 | | 4.387073 | -0.09889 | 0.949815 | | 3.930782 | -0.22928 | 0.868181 |
| DNHD1 | 2.666272 | 0.285177 | 0.999532 | | 2.725848 | 0.119305 | 0.949815 | | 2.766762 | -0.1468 | 0.940266 |
| UNK | 3.539917 | 0.338089 | 0.999532 | | 3.43474 | -0.10922 | 0.949815 | | 3.220381 | -0.42897 | 0.723933 |
| NEIL2 | 3.088565 | 0.234732 | 0.999532 | | 3.067658 | -0.11101 | 0.949815 | | 2.777727 | -0.328 | 0.814483 |
| CTSS | 5.172116 | -0.37859 | 0.999532 | | 6.802216 | 0.119425 | 0.949815 | | 5.221691 | 0.516111 | 0.667704 |
| HPS1 | 3.241962 | 0.550809 | 0.999532 | | 3.085193 | 0.110925 | 0.949815 | | 3.119114 | -0.42174 | 0.732486 |
| PTGER4 | 7.11866 | 0.370977 | 0.999532 | | 6.844319 | 0.087862 | 0.949819 | | 6.739981 | -0.26272 | 0.799944 |
| CDC37L1 | 3.920475 | -0.45834 | 0.999532 | | 4.975165 | -0.09171 | 0.949819 | | 3.599376 | 0.385386 | 0.739496 |
| SLC51A | 2.681827 | 0.1517 | 0.999532 | | 3.026762 | -0.12211 | 0.949819 | | 2.545912 | -0.23452 | 0.893298 |
| YEATS2 | 5.467687 | -0.30834 | 0.999532 | | 6.53769 | -0.08328 | 0.949819 | | 4.948866 | 0.243022 | 0.836935 |
| CCNDBP | 6.34399 | 0.16155 | 0.999532 | | 6.689555 | 0.07653 | 0.949819 | | 6.263706 | -0.06676 | 0.962255 |
| GRIPAP1 | 3.59408 | -0.20943 | 0.999532 | | 4.400748 | 0.096444 | 0.949819 | | 3.727378 | 0.336955 | 0.800922 |
| PNMA3 | 2.181629 | 0.690433 | 0.999532 | | 1.946972 | 0.137893 | 0.949819 | | 2.199607 | -0.53721 | 0.717627 |
| GPT | 2.56227 | 0.51673 | 0.999532 | | 2.564889 | 0.121922 | 0.949819 | | 2.691585 | -0.37516 | 0.799487 |
| ZNF713 | 2.088418 | 0.233006 | 0.999532 | | 2.188419 | -0.13081 | 0.949819 | | 1.940352 | -0.34122 | 0.8445 |
| NPTXR | 3.35232 | 0.050773 | 0.999532 | | 3.461546 | -0.1045 | 0.949819 | | 3.070264 | -0.1375 | 0.935145 |
| MTF1 | 1.954622 | -0.18651 | 0.999532 | | 2.35091 | 0.123718 | 0.949819 | | 1.932048 | 0.329651 | 0.854633 |
| MBD3 | 9.52422 | -0.25319 | 0.999532 | | 11.32222 | -0.06411 | 0.949819 | | 8.723338 | 0.207506 | 0.813143 |
| MNT | 2.379091 | 0.029143 | 0.999532 | | 2.700405 | 0.119204 | 0.949819 | | 2.285494 | 0.107264 | 0.95999 |
| TMEM79 | 2.334215 | 0.139025 | 0.999532 | | 2.457766 | -0.12899 | 0.949819 | | 2.166257 | -0.24451 | 0.895246 |
| ARL14EP | 3.412733 | -0.24223 | 0.999532 | | 4.254174 | 0.100902 | 0.949819 | | 3.34873 | 0.361114 | 0.791466 |
| IQCE | 5.27145 | 0.091932 | 0.999532 | | 5.632498 | -0.08977 | 0.949819 | | 4.675909 | -0.16426 | 0.900376 |
| ZNF7 | 2.452922 | -0.06949 | 0.999532 | | 2.890112 | -0.1183 | 0.949819 | | 2.315013 | -0.02002 | 0.996471 |
| TAF13 | 3.725135 | 0.023149 | 0.999532 | | 4.039839 | 0.097633 | 0.949819 | | 3.541792 | 0.08902 | 0.960773 |
| TOX4 | 6.345441 | -0.01946 | 0.999532 | | 7.117285 | 0.081674 | 0.949819 | | 6.118571 | 0.124448 | 0.920805 |
| FBXL14 | 2.703565 | -0.16955 | 0.999532 | | 2.914461 | -0.12032 | 0.949819 | | 2.511202 | 0.051621 | 0.986366 |
| LRRC47 | 4.372396 | -0.21358 | 0.999532 | | 5.45468 | 0.086467 | 0.949819 | | 4.353948 | 0.319833 | 0.78258 |
| RAMP1 | 2.273351 | 0.266744 | 0.999532 | | 2.219706 | -0.12757 | 0.949819 | | 2.036766 | -0.37945 | 0.816589 |
| SYAP1 | 3.695646 | -0.09901 | 0.999532 | | 4.397141 | 0.095078 | 0.949819 | | 3.668539 | 0.213738 | 0.881316 |
| MMS19 | 5.450576 | -0.17581 | 0.999532 | | 6.412606 | -0.08402 | 0.949819 | | 5.039043 | 0.093673 | 0.948127 |
| LRRC40 | 3.062796 | 0.357268 | 0.999532 | | 2.898459 | -0.11238 | 0.949819 | | 2.754124 | -0.45175 | 0.720106 |
| SERPINF | 2.598746 | -0.07844 | 0.999532 | | 3.032215 | 0.110957 | 0.949819 | | 2.634138 | 0.206082 | 0.903339 |
| IQCK | 3.106347 | 0.305673 | 0.999532 | | 3.15755 | 0.110419 | 0.949819 | | 2.991757 | -0.17673 | 0.911581 |
| ZC3H6 | 3.985569 | -0.04924 | 0.999532 | | 4.520842 | -0.09322 | 0.949819 | | 3.747997 | -0.02399 | 0.992286 |
| TMEM158 | 6.344105 | 0.147343 | 0.999532 | | 6.914325 | 0.087449 | 0.949819 | | 5.974043 | -0.04918 | 0.977643 |
| KLF8 | 5.392084 | 0.112353 | 0.999532 | | 5.625311 | -0.09321 | 0.949819 | | 5.074121 | -0.18218 | 0.88476 |
| CACUL1 | 3.563676 | -0.02985 | 0.999532 | | 4.347271 | 0.094678 | 0.949819 | | 3.641637 | 0.151416 | 0.920181 |
| REEP6 | 2.540282 | -0.26234 | 0.999532 | | 3.265094 | 0.112762 | 0.949819 | | 2.526846 | 0.393628 | 0.796512 |
| SLC38A9 | 3.596268 | -0.13495 | 0.999532 | | 3.941977 | -0.09935 | 0.949819 | | 3.294746 | 0.050755 | 0.979967 |
| MFF | 7.414602 | -0.01628 | 0.999532 | | 8.734038 | 0.076282 | 0.949819 | | 7.112217 | 0.110433 | 0.92252 |
| S100A1 | 2.278659 | 0.056506 | 0.999532 | | 2.218544 | -0.1322 | 0.949819 | | 1.931917 | -0.17483 | 0.933842 |
| SLC39A7 | 5.087172 | -0.13515 | 0.999532 | | 6.210163 | 0.081216 | 0.949819 | | 5.100934 | 0.235089 | 0.836935 |
| HECW2 | 6.629346 | 0.030626 | 0.999532 | | 7.559169 | 0.078809 | 0.949819 | | 6.601104 | 0.069205 | 0.960773 |
| LVRN | 1.913363 | 0.705441 | 0.999532 | | 1.902591 | 0.143412 | 0.949819 | | 1.940511 | -0.53429 | 0.732864 |
| CXCR5 | 2.077878 | 0.802746 | 0.999532 | | 1.798889 | -0.14988 | 0.949819 | | 1.934134 | -0.92962 | 0.504121 |
| KLF3 | 11.2158 | -0.08677 | 0.999532 | | 13.15978 | 0.063737 | 0.949819 | | 11.16784 | 0.177295 | 0.826947 |
| MAP1S | 3.950516 | -0.05979 | 0.999532 | | 4.145626 | -0.09955 | 0.949819 | | 3.603705 | -0.01133 | 0.996891 |
| SFMBT2 | 2.040381 | 0.267515 | 0.999532 | | 2.163726 | -0.13209 | 0.949819 | | 1.911851 | -0.37653 | 0.824012 |
| MOB1B | 3.741078 | 0.038977 | 0.999532 | | 3.926504 | -0.1113 | 0.949819 | | 3.306742 | -0.11482 | 0.945962 |
| HLA-DPB | 16.82073 | -0.46929 | 0.999532 | | 22.44328 | 0.095244 | 0.949961 | | 16.63713 | 0.587645 | 0.470811 |
| IQCC | 1.691619 | 0.276191 | 0.999532 | | 1.870609 | 0.143241 | 0.949961 | | 1.765729 | -0.11572 | 0.963728 |
| GJC3 | 4.288989 | 0.383614 | 0.999532 | | 4.251213 | 0.097077 | 0.949961 | | 4.232547 | -0.26761 | 0.833604 |
| HFE | 2.70555 | -0.54764 | 0.999532 | | 3.450149 | -0.10834 | 0.949961 | | 2.432436 | 0.458769 | 0.751444 |
| SORCS2 | 2.041595 | -0.12046 | 0.999532 | | 2.202426 | -0.12498 | 0.950032 | | 1.819107 | 0.011685 | 0.996891 |
| LEFTY1 | 2.046365 | 0.41699 | 0.999532 | | 2.07484 | 0.136269 | 0.950166 | | 2.083225 | -0.26134 | 0.890746 |
| PLEKHG | 3.324 | 0.377016 | 0.999532 | | 3.460263 | 0.110413 | 0.950166 | | 3.218661 | -0.24723 | 0.864977 |
| ATP6V0B | 8.555416 | 0.199378 | 0.999532 | | 9.286489 | 0.070186 | 0.950215 | | 8.304469 | -0.10952 | 0.911296 |
| XPA | 2.728245 | -0.45296 | 0.999532 | | 3.68902 | 0.104204 | 0.950629 | | 2.64914 | 0.576059 | 0.65401 |
| LIFR | 8.375597 | -0.20529 | 0.999532 | | 9.898521 | -0.07328 | 0.950629 | | 7.669302 | 0.146871 | 0.887166 |
| MS4A6A | 2.757643 | 0.176934 | 0.999532 | | 2.769811 | -0.11899 | 0.950629 | | 2.555182 | -0.28009 | 0.869378 |
| EIF4E3 | 3.05591 | -0.38994 | 0.999532 | | 3.684279 | -0.10387 | 0.950629 | | 2.94999 | 0.294115 | 0.858647 |
| SETD1A | 3.476895 | -0.08053 | 0.999532 | | 3.616058 | -0.09968 | 0.950629 | | 3.253004 | -0.01559 | 0.996681 |
| KLHL14 | 2.219536 | -0.04042 | 0.999532 | | 2.49251 | -0.13314 | 0.950629 | | 2.063808 | -0.07201 | 0.978229 |
| MRPS25 | 3.718123 | -0.29174 | 0.999532 | | 4.869703 | 0.089879 | 0.950662 | | 3.765876 | 0.401109 | 0.722703 |
| SMN1 | 4.487481 | 0.32224 | 0.999532 | | 4.313999 | -0.09845 | 0.950662 | | 4.230059 | -0.40479 | 0.735774 |
| EIF1AY | 2.563208 | 0.015116 | 0.999532 | | 3.275616 | 0.139728 | 0.950662 | | 2.657448 | 0.14536 | 0.946512 |
| RHBDF2 | 2.316902 | -0.39606 | 0.999532 | | 3.061576 | -0.11812 | 0.950762 | | 2.15713 | 0.302981 | 0.864923 |
| ATL2 | 4.152307 | -0.22338 | 0.999532 | | 4.730445 | -0.09675 | 0.950762 | | 3.649651 | 0.154763 | 0.915533 |
| H2AC20 | 20.33983 | 0.171731 | 0.999532 | | 21.99915 | 0.068978 | 0.950762 | | 20.17659 | -0.09289 | 0.922437 |
| DHRS13 | 3.913829 | 0.610249 | 0.999532 | | 3.472767 | 0.119357 | 0.950762 | | 3.937981 | -0.50153 | 0.692762 |
| C11orf71 | 3.024143 | 0.307962 | 0.999532 | | 3.07109 | -0.11935 | 0.950909 | | 2.89106 | -0.40901 | 0.768974 |
| MICA | 6.202441 | 0.111163 | 0.999532 | | 6.886159 | 0.084156 | 0.950909 | | 6.115628 | -0.00528 | 0.997967 |
| INPP5J | 2.619289 | 0.161952 | 0.999532 | | 2.850241 | 0.115558 | 0.950909 | | 2.456897 | -0.02432 | 0.993618 |
| KLHL30 | 3.0971 | 0.226477 | 0.999532 | | 3.441742 | 0.121204 | 0.950925 | | 3.142824 | -0.08153 | 0.971732 |
| MYG1 | 4.192817 | 0.106458 | 0.999532 | | 4.500018 | 0.090553 | 0.950976 | | 4.046435 | -0.00191 | 0.999385 |
| URB2 | 2.151393 | 0.226525 | 0.999532 | | 1.908579 | -0.13568 | 0.951002 | | 2.034915 | -0.35674 | 0.856551 |
| ABLIM1 | 3.491572 | 0.001059 | 0.999532 | | 4.151207 | 0.093522 | 0.951033 | | 3.343919 | 0.105779 | 0.952785 |
| RANBP9 | 4.567617 | -0.27613 | 0.999532 | | 5.590637 | -0.08251 | 0.951033 | | 4.15383 | 0.212385 | 0.865065 |
| CA2 | 4.64954 | 0.48533 | 0.999532 | | 4.44903 | 0.098058 | 0.951033 | | 4.361205 | -0.36675 | 0.771685 |
| MFSD5 | 3.839973 | 0.526488 | 0.999532 | | 3.593268 | 0.104027 | 0.951033 | | 3.822485 | -0.39967 | 0.722703 |
| NR2C2AP | 2.466144 | 0.352084 | 0.999532 | | 2.494213 | 0.119617 | 0.951033 | | 2.42038 | -0.21415 | 0.900848 |
| HNRNPA | 5.216244 | 0.175677 | 0.999532 | | 5.441745 | 0.091384 | 0.951033 | | 5.053321 | -0.06872 | 0.967145 |
| MAIP1 | 2.414422 | 0.544174 | 0.999532 | | 2.183087 | 0.128294 | 0.951033 | | 2.251086 | -0.39825 | 0.79686 |
| TTC39C | 2.039496 | 0.892246 | 0.999532 | | 1.833523 | 0.142796 | 0.951033 | | 2.062522 | -0.73071 | 0.603483 |
| ADPRS | 7.394728 | -0.60906 | 0.999532 | | 9.98257 | -0.06736 | 0.951033 | | 6.767957 | 0.559627 | 0.49299 |
| TMEM138 | 3.896407 | 0.0099 | 0.999532 | | 4.47839 | -0.09103 | 0.951033 | | 3.675882 | -0.07475 | 0.96588 |
| PLIN3 | 2.898603 | -0.41287 | 0.999532 | | 3.837963 | 0.106013 | 0.951033 | | 2.756723 | 0.548584 | 0.673405 |
| TRMT10A | 2.358556 | 0.329437 | 0.999532 | | 2.493116 | 0.120693 | 0.951033 | | 2.309626 | -0.18653 | 0.917861 |
| IL1A | 2.20992 | 0.656977 | 0.999532 | | 1.97024 | 0.139386 | 0.951033 | | 2.211064 | -0.50507 | 0.750371 |
| CYP39A1 | 1.778358 | 0.618298 | 0.999532 | | 1.680291 | 0.145978 | 0.951033 | | 1.829147 | -0.45615 | 0.792688 |
| POLR3B | 2.373616 | 0.209815 | 0.999532 | | 2.587071 | 0.118953 | 0.951074 | | 2.508581 | -0.06518 | 0.978229 |
| BTBD3 | 3.443559 | -0.33586 | 0.999532 | | 3.895081 | -0.09572 | 0.951099 | | 3.070294 | 0.253105 | 0.862823 |
| PSIP1 | 8.777694 | 0.084721 | 0.999532 | | 9.690739 | 0.07092 | 0.951099 | | 8.547503 | 0.007594 | 0.996891 |
| OR8H3 | 2.309887 | 0.20287 | 0.999532 | | 2.300587 | -0.12889 | 0.951099 | | 2.123965 | -0.31325 | 0.862147 |
| LINS1 | 2.441545 | 0.297439 | 0.999532 | | 2.206543 | -0.12775 | 0.951353 | | 2.16798 | -0.4143 | 0.795802 |
| NDUFAF | 2.727987 | 0.783433 | 0.999532 | | 2.229007 | 0.123725 | 0.951519 | | 2.667803 | -0.64107 | 0.616301 |
| CTU1 | 3.720759 | 0.075458 | 0.999532 | | 4.211169 | 0.111623 | 0.951519 | | 3.815324 | 0.044544 | 0.987274 |
| NMUR1 | 2.275537 | -0.12626 | 0.999532 | | 2.661633 | -0.12029 | 0.951519 | | 2.152168 | 0.024937 | 0.994537 |
| PPDPF | 37.55042 | 0.105182 | 0.999532 | | 41.50275 | 0.051361 | 0.951519 | | 36.31899 | -0.03073 | 0.981929 |
| COX20 | 6.881685 | 0.135748 | 0.999532 | | 7.467683 | 0.080986 | 0.951519 | | 6.579829 | -0.02307 | 0.990309 |
| NR1D2 | 3.060505 | -0.63714 | 0.999532 | | 4.046974 | 0.096641 | 0.951519 | | 2.919448 | 0.753416 | 0.515762 |
| PGM3 | 2.963699 | -0.24912 | 0.999532 | | 3.489095 | -0.103 | 0.951519 | | 2.831793 | 0.163849 | 0.923223 |
| SECISBP | 2.905818 | -0.67926 | 0.999532 | | 3.851564 | -0.10062 | 0.951519 | | 2.684748 | 0.595183 | 0.660958 |
| HLX | 4.280741 | -0.02752 | 0.999532 | | 4.36945 | -0.09305 | 0.951519 | | 4.046855 | -0.03985 | 0.985299 |
| ZNF26 | 2.134653 | 0.131673 | 0.999532 | | 2.426735 | 0.131465 | 0.951519 | | 2.123591 | 0.023144 | 0.995781 |
| TEX10 | 2.139981 | 0.096735 | 0.999532 | | 2.347012 | 0.122941 | 0.951519 | | 2.09812 | 0.040672 | 0.989985 |
| NTHL1 | 3.239127 | 0.467366 | 0.999532 | | 3.043868 | 0.111047 | 0.951519 | | 3.019626 | -0.3323 | 0.806186 |
| HES1 | 5.250777 | -0.49827 | 0.999532 | | 6.615353 | -0.08152 | 0.951773 | | 4.762626 | 0.436716 | 0.660958 |
| ZNF354A | 2.558971 | -0.29083 | 0.999532 | | 3.019431 | -0.11212 | 0.951773 | | 2.36873 | 0.193894 | 0.91707 |
| ASB6 | 5.011355 | 0.122076 | 0.999532 | | 5.058191 | 0.084838 | 0.951773 | | 4.77487 | -0.04306 | 0.981692 |
| CCDC14 | 4.668125 | 0.670512 | 0.999532 | | 4.063161 | 0.097648 | 0.951798 | | 4.634659 | -0.54655 | 0.628895 |
| EPHB4 | 7.924694 | -0.37514 | 0.999532 | | 9.491 | -0.07737 | 0.951798 | | 7.043597 | 0.317036 | 0.73965 |
| SPAST | 2.9504 | 0.107996 | 0.999532 | | 2.997887 | -0.11235 | 0.951798 | | 2.809077 | -0.21087 | 0.903339 |
| RHOQ | 11.04072 | 0.146112 | 0.999532 | | 11.64222 | 0.06403 | 0.951798 | | 10.6057 | -0.07318 | 0.948415 |
| CTPS2 | 2.565956 | -0.52826 | 0.999532 | | 3.100021 | -0.10681 | 0.951798 | | 2.274434 | 0.42889 | 0.78258 |
| KRT36 | 3.351383 | 0.441478 | 0.999532 | | 3.157794 | -0.13122 | 0.951927 | | 3.149723 | -0.54995 | 0.701245 |
| NAA16 | 4.000037 | -0.28899 | 0.999532 | | 4.832887 | -0.09333 | 0.951941 | | 3.69509 | 0.217219 | 0.880375 |
| H4C3 | 6.013962 | 0.158501 | 0.999532 | | 6.117894 | -0.08644 | 0.951941 | | 5.454924 | -0.21694 | 0.849648 |
| SLC22A1 | 1.828522 | -0.13047 | 0.999532 | | 2.093102 | -0.13074 | 0.951941 | | 1.728897 | 0.019023 | 0.996681 |
| GMPPB | 4.209614 | -0.21561 | 0.999532 | | 5.412308 | 0.084287 | 0.951941 | | 4.365561 | 0.325832 | 0.781585 |
| CDKL5 | 2.131613 | 0.808198 | 0.999532 | | 1.790973 | 0.136042 | 0.951941 | | 2.115409 | -0.65634 | 0.649725 |
| PAXIP1 | 3.048558 | 0.448656 | 0.999532 | | 2.83564 | 0.109977 | 0.951941 | | 2.920167 | -0.3203 | 0.824012 |
| XPO6 | 4.553466 | 0.268233 | 0.999532 | | 4.72801 | -0.09269 | 0.951941 | | 4.274469 | -0.32579 | 0.795792 |
| MED13L | 6.510329 | 0.111756 | 0.999532 | | 7.683418 | 0.076658 | 0.951941 | | 6.585571 | -0.00324 | 0.998655 |
| HOXC13 | 2.434928 | 0.475959 | 0.999532 | | 2.084852 | -0.12914 | 0.951941 | | 2.257033 | -0.58821 | 0.686214 |
| LRRC53 | 1.867912 | 0.335604 | 0.999532 | | 1.862887 | 0.138128 | 0.951941 | | 1.901373 | -0.18191 | 0.936689 |
| SPATA31 | 2.620627 | 0.233084 | 0.999532 | | 3.02807 | 0.139811 | 0.951941 | | 2.810586 | -0.11006 | 0.963866 |
| MRPL37 | 3.206265 | 0.42408 | 0.999532 | | 3.284177 | 0.105698 | 0.951941 | | 3.194313 | -0.29391 | 0.836453 |
| GSPT2 | 2.523965 | 0.765272 | 0.999532 | | 2.08244 | -0.13248 | 0.951941 | | 2.435309 | -0.87708 | 0.491726 |
| BTG1 | 11.36925 | -0.50129 | 0.999532 | | 15.15313 | 0.073119 | 0.951941 | | 11.17008 | 0.590962 | 0.428819 |
| RNF150 | 2.533155 | 0.613513 | 0.999532 | | 2.350215 | 0.121937 | 0.951941 | | 2.513756 | -0.47348 | 0.734538 |
| RSBN1 | 3.734493 | 0.40508 | 0.999532 | | 3.466799 | -0.10326 | 0.951941 | | 3.641115 | -0.45802 | 0.706749 |
| COPE | 5.658476 | -0.50007 | 0.999532 | | 7.555134 | 0.074106 | 0.951941 | | 5.371419 | 0.594242 | 0.458377 |
| MAP3K4 | 2.383857 | 0.390731 | 0.999532 | | 2.495314 | 0.125042 | 0.951941 | | 2.371543 | -0.25494 | 0.881837 |
| BARD1 | 2.559687 | 0.054823 | 0.999532 | | 2.935304 | -0.11573 | 0.951941 | | 2.516049 | -0.15388 | 0.938604 |
| PSMG1 | 2.910614 | 0.046758 | 0.999532 | | 2.950414 | -0.12181 | 0.95198 | | 2.724872 | -0.12797 | 0.945129 |
| OTUB2 | 2.241716 | 0.30655 | 0.999532 | | 2.326739 | 0.126425 | 0.951997 | | 2.24557 | -0.16146 | 0.938429 |
| NDUFB3 | 6.902699 | 0.078383 | 0.999532 | | 7.880329 | 0.075856 | 0.951997 | | 6.693524 | 0.023672 | 0.991134 |
| KCNJ12 | 6.186307 | 0.402234 | 0.999532 | | 5.7494 | -0.08178 | 0.951997 | | 5.900605 | -0.46731 | 0.598052 |
| MOSPD2 | 3.281861 | -0.49674 | 0.999532 | | 4.358115 | 0.091887 | 0.951997 | | 3.321118 | 0.6107 | 0.598177 |
| ADGRD1 | 2.241146 | -0.11337 | 0.999532 | | 2.429209 | -0.12201 | 0.951997 | | 2.043476 | 0.008196 | 0.997967 |
| EVA1C | 1.835759 | -0.70603 | 0.999532 | | 2.425082 | 0.115398 | 0.952011 | | 1.792287 | 0.833673 | 0.575547 |
| ZFHX2 | 2.450429 | -0.08908 | 0.999532 | | 2.828406 | -0.11552 | 0.952011 | | 2.346298 | -0.00722 | 0.997967 |
| CBLN1 | 2.626717 | -0.46767 | 0.999532 | | 3.347476 | -0.11519 | 0.952011 | | 2.653134 | 0.354099 | 0.823571 |
| LEPROT | 12.92573 | -0.20046 | 0.999532 | | 15.90291 | 0.058131 | 0.952011 | | 12.79204 | 0.278806 | 0.682403 |
| AP2A2 | 8.026308 | 0.140868 | 0.999532 | | 8.46377 | -0.0726 | 0.952011 | | 7.297589 | -0.18509 | 0.863401 |
| HOXA3 | 3.825699 | 0.043066 | 0.999532 | | 4.846118 | 0.088451 | 0.952061 | | 3.978508 | 0.087945 | 0.958991 |
| KEAP1 | 2.571656 | -0.13691 | 0.999532 | | 2.948625 | -0.10875 | 0.952061 | | 2.505474 | 0.050423 | 0.985299 |
| SH2D1A | 2.430301 | 0.906546 | 0.999532 | | 2.002702 | 0.131799 | 0.952061 | | 2.393581 | -0.75792 | 0.542124 |
| C2CD2L | 3.220529 | 0.218227 | 0.999532 | | 3.301097 | -0.10579 | 0.952061 | | 3.088881 | -0.3054 | 0.833604 |
| DMRT2 | 2.323752 | -0.43215 | 0.999532 | | 2.917906 | -0.11321 | 0.952061 | | 2.191567 | 0.339922 | 0.846 |
| LIMA1 | 4.640663 | -0.69919 | 0.999532 | | 6.800474 | 0.076673 | 0.952061 | | 4.732473 | 0.794873 | 0.300261 |
| CBY2 | 2.273202 | 0.117859 | 0.999532 | | 2.473621 | -0.12157 | 0.952061 | | 2.199072 | -0.20909 | 0.914687 |
| NSF | 2.868486 | 0.1657 | 0.999532 | | 3.151322 | 0.1095 | 0.952061 | | 2.848166 | -0.03756 | 0.989148 |
| ZNF629 | 2.417952 | 0.357421 | 0.999532 | | 2.176418 | -0.12615 | 0.952108 | | 2.124391 | -0.46517 | 0.758124 |
| HHLA3 | 2.798365 | 0.675523 | 0.999532 | | 2.288766 | -0.12179 | 0.952139 | | 2.600988 | -0.78046 | 0.515762 |
| ZFYVE9 | 5.469153 | -0.13046 | 0.999532 | | 6.573457 | 0.081474 | 0.952204 | | 5.206624 | 0.23927 | 0.827267 |
| GNE | 2.732317 | 0.033441 | 0.999532 | | 2.851964 | -0.11052 | 0.952223 | | 2.547141 | -0.12869 | 0.947941 |
| ADGRE5 | 6.301688 | 0.102523 | 0.999532 | | 6.922578 | 0.085873 | 0.952267 | | 6.227257 | -0.00294 | 0.998898 |
| RAB39A | 3.267511 | 0.134743 | 0.999532 | | 3.548902 | 0.109829 | 0.952454 | | 3.092108 | 0.010533 | 0.996891 |
| ABITRAM | 6.24234 | -0.23792 | 0.999532 | | 7.2009 | -0.07931 | 0.952454 | | 5.742253 | 0.171934 | 0.891051 |
| PAN3 | 7.196229 | -0.18308 | 0.999532 | | 8.380209 | 0.069282 | 0.952493 | | 6.849382 | 0.259892 | 0.791466 |
| WDR3 | 2.614729 | 0.081691 | 0.999532 | | 2.699904 | 0.113454 | 0.952632 | | 2.522545 | 0.049066 | 0.985693 |
| ADAMTS | 2.176714 | 0.288145 | 0.999532 | | 2.306484 | 0.122748 | 0.952632 | | 2.202803 | -0.15358 | 0.941197 |
| CHMP5 | 6.755823 | 0.20365 | 0.999532 | | 7.121441 | 0.071637 | 0.952632 | | 6.581274 | -0.11448 | 0.918082 |
| SCEL | 1.944278 | 0.37472 | 0.999532 | | 1.904444 | -0.13326 | 0.952632 | | 1.944812 | -0.48966 | 0.76272 |
| HOXD12 | 2.25672 | -0.08876 | 0.999532 | | 2.598684 | -0.12614 | 0.952715 | | 2.044362 | -0.0184 | 0.996681 |
| HPS6 | 2.301573 | -0.22073 | 0.999532 | | 2.739693 | -0.11421 | 0.952715 | | 2.186297 | 0.132993 | 0.950342 |
| SCUBE2 | 2.504787 | -0.47145 | 0.999532 | | 3.110619 | -0.11166 | 0.952724 | | 2.260911 | 0.378979 | 0.816526 |
| TYRP1 | 2.188876 | 0.882609 | 0.999532 | | 1.78308 | -0.13939 | 0.953021 | | 2.053491 | -1.00211 | 0.420356 |
| NFXL1 | 3.408619 | 0.114835 | 0.999532 | | 3.721691 | 0.100136 | 0.953021 | | 3.264008 | 0.003594 | 0.999159 |
| CYYR1 | 3.594048 | 0.192529 | 0.999532 | | 3.795964 | -0.09631 | 0.953094 | | 3.412645 | -0.25543 | 0.861737 |
| DQX1 | 2.91542 | 0.071144 | 0.999532 | | 3.255275 | -0.12539 | 0.953177 | | 2.769738 | -0.17339 | 0.931649 |
| GABBR1 | 10.23964 | -0.1578 | 0.999532 | | 12.03105 | 0.061646 | 0.953195 | | 9.903527 | 0.231655 | 0.78258 |
| TLR1 | 3.620377 | 0.269679 | 0.999532 | | 3.800161 | -0.09549 | 0.953195 | | 3.424525 | -0.34473 | 0.77821 |
| FOXL1 | 1.993289 | 0.642793 | 0.999532 | | 1.705424 | -0.14113 | 0.953195 | | 1.862349 | -0.7862 | 0.617734 |
| CEP126 | 3.132954 | -0.37913 | 0.999532 | | 3.927623 | -0.1038 | 0.953304 | | 3.048725 | 0.302815 | 0.858767 |
| ZNF551 | 4.1122 | -0.10021 | 0.999532 | | 5.096508 | 0.091733 | 0.953619 | | 4.235443 | 0.209967 | 0.881316 |
| ITGA7 | 4.347379 | -0.13758 | 0.999532 | | 4.852659 | -0.09121 | 0.953619 | | 4.032504 | 0.05979 | 0.97502 |
| CWF19L2 | 3.372916 | -0.09548 | 0.999532 | | 4.126157 | 0.089717 | 0.953619 | | 3.446485 | 0.205156 | 0.88588 |
| NFATC2I | 4.53092 | -0.53883 | 0.999532 | | 5.583265 | -0.08069 | 0.953619 | | 4.067878 | 0.465076 | 0.673405 |
| SOX13 | 6.384261 | -0.33339 | 0.999532 | | 8.271089 | 0.066355 | 0.953619 | | 6.35549 | 0.418935 | 0.628895 |
| VCPIP1 | 2.288387 | -0.17601 | 0.999532 | | 2.760291 | 0.111955 | 0.953619 | | 2.178639 | 0.309016 | 0.859255 |
| MYH7B | 2.101196 | -0.0898 | 0.999532 | | 2.749419 | 0.120123 | 0.953619 | | 2.328637 | 0.224844 | 0.906811 |
| RWDD2A | 2.612782 | -0.02276 | 0.999532 | | 2.955533 | -0.10823 | 0.953619 | | 2.420671 | -0.06787 | 0.975753 |
| SLC39A6 | 5.100862 | -0.41375 | 0.999532 | | 7.013714 | 0.07028 | 0.953619 | | 5.017713 | 0.503635 | 0.568812 |
| PPIL2 | 2.531202 | 0.32721 | 0.999532 | | 2.398737 | -0.11788 | 0.953619 | | 2.316457 | -0.42743 | 0.776852 |
| CASR | 2.365356 | 0.089688 | 0.999532 | | 2.785166 | -0.11147 | 0.953619 | | 2.244615 | -0.15406 | 0.939107 |
| RCAN2 | 3.431626 | -0.30672 | 0.999532 | | 4.069552 | 0.096845 | 0.953619 | | 3.406932 | 0.410104 | 0.757037 |
| HAND1 | 2.473306 | 0.234722 | 0.999532 | | 2.462576 | -0.12659 | 0.953619 | | 2.290812 | -0.3493 | 0.827488 |
| TTLL7 | 2.748522 | 0.295965 | 0.999532 | | 2.896731 | 0.109771 | 0.953619 | | 2.852598 | -0.16265 | 0.928454 |
| RAP1GD | 2.842778 | 0.223945 | 0.999532 | | 2.626986 | -0.11182 | 0.953619 | | 2.681276 | -0.32599 | 0.849682 |
| PLEKHM | 6.608508 | 0.047894 | 0.999532 | | 7.135101 | -0.07336 | 0.953619 | | 5.981333 | -0.09839 | 0.93603 |
| ZNF618 | 4.570567 | -0.31651 | 0.999532 | | 5.60491 | -0.08273 | 0.953619 | | 4.44442 | 0.24978 | 0.842686 |
| ZNF524 | 4.811576 | 0.136288 | 0.999532 | | 5.06246 | -0.08644 | 0.953619 | | 4.558118 | -0.20424 | 0.872493 |
| GLCE | 2.796256 | 0.421584 | 0.999532 | | 2.882006 | 0.107666 | 0.953619 | | 2.861081 | -0.29306 | 0.84579 |
| TEKT3 | 2.1005 | 0.156171 | 0.999532 | | 2.148775 | -0.12816 | 0.953619 | | 2.016713 | -0.26876 | 0.889853 |
| MDFI | 2.490162 | 0.148301 | 0.999532 | | 2.442965 | -0.11463 | 0.953619 | | 2.283711 | -0.24993 | 0.88734 |
| TBC1D10 | 2.585029 | -0.32691 | 0.999532 | | 3.208308 | -0.10704 | 0.953619 | | 2.336744 | 0.243188 | 0.889853 |
| PBX3 | 3.099218 | 0.60319 | 0.999532 | | 2.835127 | 0.111843 | 0.953619 | | 3.088832 | -0.47585 | 0.706407 |
| NCS1 | 2.334439 | 0.298371 | 0.999532 | | 2.361339 | -0.11779 | 0.953619 | | 2.298253 | -0.39355 | 0.803301 |
| SPIDR | 4.825927 | 0.211429 | 0.999532 | | 4.923454 | -0.09034 | 0.953619 | | 4.486047 | -0.27871 | 0.814483 |
| SIM2 | 1.963375 | 0.626309 | 0.999532 | | 1.908807 | 0.131631 | 0.953619 | | 2.072012 | -0.48945 | 0.738316 |
| KNG1 | 6.004886 | 0.885275 | 0.999532 | | 5.023304 | 0.098969 | 0.953619 | | 5.660292 | -0.81436 | 0.636525 |
| BHLHB9 | 1.61809 | -0.32953 | 0.999532 | | 1.889144 | -0.13296 | 0.953619 | | 1.453126 | 0.213642 | 0.928522 |
| GPHB5 | 1.98647 | 0.398027 | 0.999532 | | 2.032273 | -0.13117 | 0.953619 | | 1.976466 | -0.50777 | 0.752562 |
| SNX13 | 6.26566 | 0.43603 | 0.999532 | | 5.811137 | 0.080007 | 0.953619 | | 6.050309 | -0.338 | 0.721294 |
| SNX18 | 5.102905 | -0.2635 | 0.999532 | | 6.077181 | -0.07891 | 0.953619 | | 4.828306 | 0.205845 | 0.864245 |
| HMGXB4 | 2.655087 | -0.33733 | 0.999532 | | 3.054255 | -0.10823 | 0.953619 | | 2.311787 | 0.239283 | 0.893262 |
| MRPL47 | 3.612948 | -0.38443 | 0.999532 | | 4.555699 | 0.089005 | 0.953619 | | 3.624929 | 0.50649 | 0.661553 |
| LETM1 | 4.246402 | 0.332485 | 0.999532 | | 3.967529 | -0.09039 | 0.953619 | | 3.920138 | -0.41093 | 0.718643 |
| MCMDC2 | 1.692579 | -0.11055 | 0.999532 | | 2.043672 | 0.126985 | 0.953619 | | 1.713498 | 0.256063 | 0.900848 |
| KMT5A | 2.843279 | -0.30678 | 0.999532 | | 3.649703 | 0.09731 | 0.953619 | | 2.868928 | 0.423638 | 0.739496 |
| CEP128 | 2.74058 | 0.175761 | 0.999532 | | 2.724083 | 0.1059 | 0.953619 | | 2.612403 | -0.05642 | 0.97928 |
| LCTL | 2.05391 | 0.249598 | 0.999532 | | 2.084682 | -0.13191 | 0.953619 | | 2.107575 | -0.37177 | 0.839811 |
| SNRPA | 6.57712 | 0.159091 | 0.999532 | | 6.844913 | 0.073784 | 0.953619 | | 6.234989 | -0.06804 | 0.961643 |
| NUTF2 | 5.043166 | -0.04278 | 0.999532 | | 5.904614 | 0.077363 | 0.953619 | | 4.940568 | 0.138571 | 0.911296 |
| TCP11L2 | 3.705001 | 0.16132 | 0.999532 | | 3.779032 | -0.09677 | 0.953619 | | 3.548783 | -0.23 | 0.868512 |
| GLIS2 | 10.85874 | 0.165733 | 0.999532 | | 11.70361 | 0.067152 | 0.953619 | | 10.47741 | -0.07168 | 0.952656 |
| DMTF1 | 3.399102 | -0.46313 | 0.999532 | | 4.358969 | -0.08956 | 0.953619 | | 3.066173 | 0.393087 | 0.757035 |
| CBX8 | 3.334193 | 0.434188 | 0.999532 | | 3.435885 | 0.108213 | 0.953619 | | 3.468273 | -0.29286 | 0.851785 |
| RAI14 | 5.376004 | 0.090097 | 0.999532 | | 5.957541 | 0.077238 | 0.953619 | | 5.159524 | 0.003704 | 0.998586 |
| IL2RB | 2.131957 | 0.125569 | 0.999532 | | 2.108842 | -0.12536 | 0.953619 | | 2.040627 | -0.23583 | 0.901033 |
| NEU2 | 2.138842 | 0.488847 | 0.999532 | | 2.048079 | -0.13504 | 0.953619 | | 2.127736 | -0.61172 | 0.699971 |
| OTOP3 | 1.990629 | 0.165281 | 0.999532 | | 2.012261 | -0.13195 | 0.953619 | | 1.920178 | -0.2764 | 0.88885 |
| C21orf91 | 3.261549 | -0.26119 | 0.999532 | | 3.87672 | -0.09482 | 0.953619 | | 3.040154 | 0.184496 | 0.906811 |
| DOCK10 | 3.105133 | 0.3357 | 0.999532 | | 3.178954 | 0.104803 | 0.953619 | | 3.074379 | -0.21256 | 0.889853 |
| ARL4C | 2.380278 | -0.35906 | 0.999532 | | 2.95634 | -0.11245 | 0.953619 | | 2.192659 | 0.265457 | 0.87892 |
| ZNF571 | 2.072804 | -0.2349 | 0.999532 | | 2.700089 | 0.112985 | 0.953619 | | 2.18124 | 0.365401 | 0.82512 |
| FAM168B | 3.101351 | 0.008058 | 0.999532 | | 3.515176 | 0.095339 | 0.953619 | | 2.88219 | 0.099959 | 0.955834 |
| IFTAP | 3.996802 | 0.14977 | 0.999532 | | 4.412983 | 0.087935 | 0.953619 | | 3.991857 | -0.04394 | 0.982124 |
| PRKAA2 | 2.787918 | 0.135701 | 0.999532 | | 2.941494 | -0.10805 | 0.953619 | | 2.392819 | -0.23299 | 0.884653 |
| SDK2 | 2.451529 | 0.126587 | 0.999532 | | 2.737799 | 0.109595 | 0.953619 | | 2.526116 | -0.00023 | 0.999949 |
| DNAJC2 | 2.929885 | 0.320379 | 0.999532 | | 3.032677 | 0.108728 | 0.953619 | | 2.807148 | -0.1904 | 0.905688 |
| CASP2 | 3.927443 | 0.076232 | 0.999532 | | 4.456939 | 0.091776 | 0.953619 | | 3.949622 | 0.035904 | 0.988079 |
| HYOU1 | 4.648665 | -0.08207 | 0.999532 | | 5.120555 | -0.08809 | 0.953619 | | 4.211452 | 0.013314 | 0.996681 |
| UTP14C | 2.80371 | 0.243617 | 0.999532 | | 2.966577 | 0.10578 | 0.953619 | | 2.781686 | -0.11913 | 0.947515 |
| DRG1 | 6.075133 | -0.31484 | 0.999532 | | 7.612014 | 0.074801 | 0.953619 | | 5.756749 | 0.405463 | 0.666566 |
| POLD4 | 10.00759 | 0.131992 | 0.999532 | | 10.09633 | -0.06437 | 0.953619 | | 9.392404 | -0.17692 | 0.843463 |
| MYBPC3 | 2.000992 | -0.01391 | 0.999532 | | 2.061573 | -0.1286 | 0.953619 | | 1.872572 | -0.11397 | 0.965649 |
| TCF19 | 2.121939 | -0.02434 | 0.999532 | | 2.084637 | -0.12181 | 0.953619 | | 1.983418 | -0.08931 | 0.972758 |
| JMJD4 | 3.083509 | 0.039695 | 0.999532 | | 3.164108 | -0.10868 | 0.953644 | | 2.782419 | -0.12639 | 0.945129 |
| SLC37A3 | 2.645152 | -0.69341 | 0.999532 | | 3.530614 | -0.1002 | 0.953644 | | 2.59339 | 0.611945 | 0.660958 |
| IDH3A | 4.148318 | 0.093629 | 0.999532 | | 4.808899 | 0.083349 | 0.953644 | | 4.27823 | 0.010093 | 0.996891 |
| COX11 | 4.996498 | 0.081254 | 0.999532 | | 5.589359 | 0.078445 | 0.953678 | | 4.747267 | 0.010599 | 0.996891 |
| KCTD3 | 3.80708 | -0.06722 | 0.999532 | | 4.347796 | 0.086479 | 0.9537 | | 3.788953 | 0.176143 | 0.899161 |
| ZNF426 | 2.683431 | 0.001733 | 0.999532 | | 2.853424 | -0.10917 | 0.9537 | | 2.398769 | -0.09417 | 0.965649 |
| PPIL3 | 2.787197 | 0.309602 | 0.999532 | | 2.607461 | -0.11094 | 0.953888 | | 2.66944 | -0.39604 | 0.790999 |
| HNRNPC | 11.04816 | 0.23559 | 0.999532 | | 11.59603 | 0.061266 | 0.953888 | | 11.06787 | -0.16262 | 0.858767 |
| RCC1L | 5.104317 | -0.04689 | 0.999532 | | 5.903397 | 0.07735 | 0.953888 | | 4.775278 | 0.142852 | 0.910105 |
| LSM12 | 5.609187 | -0.01418 | 0.999532 | | 6.582813 | 0.072232 | 0.953888 | | 5.393725 | 0.105476 | 0.934597 |
| MSH6 | 2.270094 | 0.144342 | 0.999532 | | 2.203848 | -0.11822 | 0.953888 | | 2.071605 | -0.24767 | 0.892869 |
| XRCC3 | 2.770474 | 0.148217 | 0.999532 | | 2.968178 | -0.1077 | 0.953888 | | 2.567968 | -0.23214 | 0.888003 |
| DROSHA | 2.795194 | -0.09688 | 0.999532 | | 3.169552 | 0.097695 | 0.953888 | | 2.634751 | 0.207401 | 0.897958 |
| CCDC18 | 2.466223 | 0.547999 | 0.999532 | | 2.433509 | 0.127229 | 0.953888 | | 2.482388 | -0.40092 | 0.803493 |
| TACC1 | 6.322576 | -0.70509 | 0.999532 | | 8.823124 | 0.07335 | 0.953888 | | 6.150835 | 0.804846 | 0.241605 |
| STXBP3 | 5.558501 | 0.000736 | 0.999532 | | 6.290128 | 0.074003 | 0.953888 | | 5.384263 | 0.090749 | 0.948127 |
| ANKRD1 | 10.3771 | -0.06064 | 0.999532 | | 12.30382 | -0.05615 | 0.953888 | | 9.965799 | 0.034814 | 0.978229 |
| CTPS1 | 2.733252 | 0.28419 | 0.999532 | | 2.759246 | 0.106495 | 0.953888 | | 2.626879 | -0.16108 | 0.926586 |
| AKNA | 2.750573 | 0.187021 | 0.999532 | | 3.097559 | 0.104143 | 0.953888 | | 2.845961 | -0.06733 | 0.974421 |
| IL1R2 | 2.237752 | -0.00439 | 0.999532 | | 2.524254 | -0.11268 | 0.953888 | | 2.143508 | -0.08901 | 0.970532 |
| CRTC3 | 3.216396 | -0.49513 | 0.999532 | | 3.798667 | -0.0908 | 0.953888 | | 2.959821 | 0.42535 | 0.750371 |
| ADAP2 | 2.932614 | -0.54665 | 0.999532 | | 3.648614 | -0.09396 | 0.953888 | | 2.643995 | 0.466131 | 0.735661 |
| MDFIC | 3.787704 | 0.113378 | 0.999532 | | 4.244671 | 0.094695 | 0.953888 | | 3.872583 | -0.00433 | 0.998586 |
| SUGCT | 2.449507 | -0.01861 | 0.999532 | | 2.903673 | 0.108451 | 0.953888 | | 2.362599 | 0.149471 | 0.939913 |
| MPPE1 | 4.36942 | 0.187989 | 0.999532 | | 4.395253 | 0.087592 | 0.953888 | | 4.252766 | -0.09097 | 0.958855 |
| NEURL4 | 2.59983 | 0.042492 | 0.999532 | | 2.733432 | -0.11956 | 0.953888 | | 2.482864 | -0.14043 | 0.946808 |
| CD200R1 | 1.974701 | 0.289476 | 0.999532 | | 2.253515 | 0.120629 | 0.953888 | | 2.093677 | -0.14692 | 0.944635 |
| DGKB | 2.729238 | 0.916155 | 0.999532 | | 2.036083 | -0.12202 | 0.953888 | | 2.582314 | -1.01854 | 0.339404 |
| KLHL31 | 2.094815 | 0.61003 | 0.999532 | | 1.940951 | 0.130562 | 0.953888 | | 2.088381 | -0.45982 | 0.790208 |
| ALMS1 | 5.259882 | 0.107537 | 0.999532 | | 5.261001 | -0.08165 | 0.953892 | | 4.718577 | -0.17397 | 0.892869 |
| LMO3 | 2.56764 | -0.04732 | 0.999532 | | 2.999728 | -0.10887 | 0.953892 | | 2.350075 | -0.03961 | 0.989148 |
| SNX32 | 2.283184 | -0.17066 | 0.999532 | | 2.663846 | -0.11514 | 0.953893 | | 2.1862 | 0.073724 | 0.977359 |
| H1-8 | 2.162964 | 0.252562 | 0.999532 | | 2.148106 | -0.1215 | 0.953964 | | 2.022956 | -0.35446 | 0.837633 |
| CIAO3 | 2.969633 | -0.32594 | 0.999532 | | 3.711101 | 0.100506 | 0.953964 | | 2.811694 | 0.455256 | 0.722703 |
| PORCN | 1.964854 | 0.047624 | 0.999532 | | 2.105542 | -0.12454 | 0.953964 | | 1.755482 | -0.15644 | 0.945129 |
| TRUB1 | 3.532505 | -0.16363 | 0.999532 | | 4.084803 | -0.0872 | 0.953964 | | 3.342715 | 0.094404 | 0.956322 |
| TOP3A | 4.178755 | -0.19137 | 0.999532 | | 4.775761 | -0.0857 | 0.953964 | | 3.773116 | 0.121035 | 0.940162 |
| CA12 | 11.7243 | 0.606954 | 0.999532 | | 10.86327 | 0.077273 | 0.953964 | | 11.14377 | -0.54526 | 0.588052 |
| BTN3A1 | 4.758564 | 0.290564 | 0.999532 | | 5.15368 | 0.083762 | 0.953964 | | 4.80734 | -0.17958 | 0.891464 |
| DMPK | 5.719052 | -0.0185 | 0.999532 | | 6.327163 | 0.078036 | 0.953964 | | 5.489131 | 0.10573 | 0.939913 |
| C8orf44 | 2.176014 | -0.04453 | 0.999532 | | 2.569947 | 0.110464 | 0.954012 | | 2.077559 | 0.171933 | 0.928522 |
| ADCY1 | 3.004903 | -0.0787 | 0.999532 | | 3.589644 | 0.100855 | 0.954094 | | 3.025351 | 0.199816 | 0.904721 |
| KLK10 | 1.886536 | -0.11174 | 0.999532 | | 2.192398 | -0.12255 | 0.954094 | | 1.82532 | 0.00846 | 0.997967 |
| CLDN14 | 2.948682 | 0.718642 | 0.999532 | | 2.538962 | -0.11738 | 0.954143 | | 2.743204 | -0.80625 | 0.515762 |
| HSD17B1 | 2.647505 | -0.46209 | 0.999532 | | 3.676789 | 0.095731 | 0.954244 | | 2.768944 | 0.581255 | 0.657538 |
| INTS11 | 5.832502 | 0.255676 | 0.999532 | | 6.273059 | 0.078762 | 0.954244 | | 5.650146 | -0.1582 | 0.891836 |
| ETV4 | 2.444913 | 0.360164 | 0.999532 | | 2.339223 | -0.11458 | 0.954488 | | 2.21855 | -0.45661 | 0.75311 |
| MAP2K5 | 2.938681 | -0.26766 | 0.999532 | | 3.389264 | -0.09558 | 0.954681 | | 2.620913 | 0.189052 | 0.908829 |
| ADAMTS | 2.459872 | 0.147797 | 0.999532 | | 2.630211 | 0.108836 | 0.954681 | | 2.384996 | -0.02137 | 0.995112 |
| TBC1D20 | 5.180081 | 0.281123 | 0.999532 | | 5.364997 | 0.08264 | 0.954876 | | 5.182656 | -0.18222 | 0.881837 |
| MAFB | 3.371377 | 0.231785 | 0.999532 | | 3.748228 | 0.093813 | 0.954876 | | 3.225594 | -0.12021 | 0.943627 |
| RAB3IL1 | 3.412658 | -0.43315 | 0.999532 | | 4.358936 | -0.08702 | 0.954876 | | 3.293506 | 0.367182 | 0.773849 |
| RRN3 | 4.982318 | 0.35005 | 0.999532 | | 4.918158 | 0.092493 | 0.954902 | | 4.919134 | -0.26306 | 0.85066 |
| AKAP7 | 2.491669 | -0.10916 | 0.999532 | | 2.788514 | -0.11025 | 0.954902 | | 2.380488 | 0.015945 | 0.996681 |
| PCNX1 | 4.799946 | -0.27399 | 0.999532 | | 5.905504 | 0.085174 | 0.954902 | | 4.543197 | 0.389004 | 0.723933 |
| CHI3L1 | 11.22248 | 0.080188 | 0.999532 | | 11.70173 | -0.10403 | 0.954902 | | 10.2808 | -0.15148 | 0.907968 |
| HOOK2 | 2.761031 | 0.400675 | 0.999532 | | 2.864031 | 0.104256 | 0.954902 | | 2.62711 | -0.28136 | 0.851785 |
| ZNF610 | 2.325042 | -0.25291 | 0.999532 | | 2.952163 | 0.110066 | 0.954902 | | 2.422087 | 0.382394 | 0.816618 |
| MRM2 | 4.310853 | -0.06056 | 0.999532 | | 4.844602 | -0.10051 | 0.954902 | | 4.00999 | -0.00836 | 0.997084 |
| SLC2A13 | 3.072411 | -0.26744 | 0.999532 | | 3.955203 | 0.095903 | 0.954902 | | 3.095143 | 0.381928 | 0.776283 |
| LZTS3 | 3.470883 | -0.27633 | 0.999532 | | 4.499237 | 0.092055 | 0.954902 | | 3.412328 | 0.374954 | 0.764149 |
| SHISA2 | 2.859975 | -0.014 | 0.999532 | | 3.185781 | 0.102839 | 0.954902 | | 2.723472 | 0.133066 | 0.94366 |
| SLC30A7 | 5.223262 | -0.28602 | 0.999532 | | 6.571288 | -0.07981 | 0.954902 | | 4.973721 | 0.210039 | 0.859426 |
| TUBA3D | 2.704678 | 0.106549 | 0.999532 | | 2.771757 | -0.1093 | 0.954902 | | 2.421039 | -0.20713 | 0.911296 |
| TMEM8B | 3.192019 | 0.213123 | 0.999532 | | 3.403105 | 0.098292 | 0.954902 | | 3.082376 | -0.09301 | 0.95999 |
| FRAT1 | 2.546315 | 0.057859 | 0.999532 | | 3.057025 | 0.119668 | 0.954902 | | 2.64793 | 0.06993 | 0.977473 |
| FAM24B | 2.431666 | 0.125633 | 0.999532 | | 2.662642 | 0.107341 | 0.954902 | | 2.373885 | -0.00387 | 0.999348 |
| GJC1 | 2.112483 | -0.19924 | 0.999532 | | 2.601198 | 0.111655 | 0.954902 | | 2.169315 | 0.327728 | 0.851503 |
| TNFRSF1 | 2.463768 | 0.007753 | 0.999532 | | 2.686319 | -0.11432 | 0.954902 | | 2.281749 | -0.1028 | 0.963866 |
| TENM2 | 2.420662 | -0.15962 | 0.999532 | | 2.644536 | -0.10865 | 0.954902 | | 2.156799 | 0.067102 | 0.978229 |
| EXT1 | 3.117492 | -0.39645 | 0.999532 | | 3.769962 | -0.09547 | 0.954902 | | 2.878118 | 0.319594 | 0.829716 |
| MBP | 3.870417 | 0.510164 | 0.999532 | | 3.423582 | -0.09497 | 0.954902 | | 3.567925 | -0.58718 | 0.568812 |
| PSTPIP2 | 2.295408 | -0.23228 | 0.999532 | | 2.78146 | -0.10567 | 0.954902 | | 2.131758 | 0.146301 | 0.941197 |
| FAM163B | 2.02721 | 0.342694 | 0.999532 | | 1.884329 | -0.12792 | 0.954902 | | 1.862926 | -0.45275 | 0.792416 |
| MR01 | 4.657435 | 0.460669 | 0.999532 | | 4.177098 | -0.08779 | 0.954902 | | 4.258435 | -0.53192 | 0.595502 |
| TMEM140 | 7.453073 | -0.1263 | 0.999532 | | 8.627738 | 0.064291 | 0.954902 | | 7.06638 | 0.202233 | 0.840357 |
| FAM91A1 | 5.707399 | 0.126848 | 0.999532 | | 6.172797 | 0.076714 | 0.954902 | | 5.808091 | -0.04162 | 0.979967 |
| NBEAL2 | 3.304258 | 0.332048 | 0.999532 | | 3.181072 | 0.098339 | 0.954902 | | 3.252486 | -0.21256 | 0.890225 |
| TRAF2 | 2.861686 | -0.40072 | 0.999532 | | 3.539837 | -0.09606 | 0.954902 | | 2.862995 | 0.32946 | 0.823571 |
| ZFP42 | 2.67298 | 0.17721 | 0.999532 | | 2.736651 | -0.11569 | 0.954902 | | 2.530288 | -0.2757 | 0.879538 |
| ST6GALN | 2.892331 | 0.476765 | 0.999532 | | 2.760887 | 0.111964 | 0.954902 | | 2.802346 | -0.34973 | 0.821967 |
| SCGB1A1 | 2.561659 | 0.295854 | 0.999532 | | 2.610501 | -0.11379 | 0.954902 | | 2.528039 | -0.39254 | 0.800239 |
| MRPL53 | 3.784776 | -0.41464 | 0.999532 | | 5.246084 | 0.079113 | 0.954902 | | 3.832354 | 0.518897 | 0.636525 |
| LGI1 | 2.383025 | 0.394388 | 0.999532 | | 2.196548 | -0.12271 | 0.954902 | | 2.304395 | -0.50161 | 0.752416 |
| TAS2R50 | 2.320991 | 0.27005 | 0.999532 | | 2.158499 | -0.11834 | 0.954902 | | 2.184194 | -0.37928 | 0.830822 |
| KCNJ10 | 1.841014 | 0.137619 | 0.999532 | | 1.968326 | -0.12468 | 0.954902 | | 1.796645 | -0.24136 | 0.903558 |
| LPAR6 | 5.083778 | -0.00164 | 0.999532 | | 5.684941 | 0.095952 | 0.954902 | | 4.971176 | 0.112787 | 0.944993 |
| PSPC1 | 5.599562 | 0.023455 | 0.999532 | | 6.454239 | 0.077324 | 0.954902 | | 5.576251 | 0.0748 | 0.95857 |
| WNT3A | 3.439812 | 0.315856 | 0.999532 | | 3.256352 | -0.10364 | 0.954934 | | 3.196767 | -0.40544 | 0.764366 |
| FLRT2 | 2.602865 | 0.144251 | 0.999532 | | 2.621481 | -0.10968 | 0.954934 | | 2.278117 | -0.23444 | 0.890746 |
| TSPAN11 | 2.300801 | -0.24259 | 0.999532 | | 2.575632 | -0.11256 | 0.954934 | | 2.107469 | 0.148715 | 0.945129 |
| ZNF705B | 2.080795 | 0.263787 | 0.999532 | | 2.095938 | 0.121371 | 0.955074 | | 2.218816 | -0.12115 | 0.95999 |
| PLEKHA | 5.618727 | -0.2773 | 0.999532 | | 6.836363 | 0.069675 | 0.955188 | | 5.529624 | 0.369182 | 0.699971 |
| STAC | 2.505172 | 0.657435 | 0.999532 | | 2.031584 | -0.12491 | 0.955188 | | 2.37684 | -0.75811 | 0.622065 |
| AGO3 | 3.193373 | -0.02739 | 0.999532 | | 3.230697 | -0.09631 | 0.955219 | | 2.948482 | -0.05439 | 0.979432 |
| IQCG | 1.742041 | 0.090241 | 0.999532 | | 1.961986 | 0.123915 | 0.955219 | | 1.776218 | 0.050898 | 0.987608 |
| THEM4 | 2.617323 | 0.222471 | 0.999532 | | 2.942124 | 0.109894 | 0.955219 | | 2.558289 | -0.10149 | 0.960282 |
| ZACN | 2.880059 | 0.959687 | 0.999532 | | 2.03133 | -0.12299 | 0.955219 | | 2.747709 | -1.07113 | 0.30352 |
| SAMD12 | 2.800626 | -0.12283 | 0.999532 | | 3.14702 | -0.10025 | 0.955219 | | 2.606048 | 0.041066 | 0.988436 |
| WIPF3 | 4.94102 | 0.149945 | 0.999532 | | 5.375764 | 0.082041 | 0.955322 | | 4.631445 | -0.0667 | 0.970811 |
| CENPP | 2.681709 | 0.333536 | 0.999532 | | 2.533865 | -0.10993 | 0.95543 | | 2.505363 | -0.42682 | 0.765852 |
| HBM | 2.946611 | 0.157727 | 0.999532 | | 3.251416 | 0.101975 | 0.955461 | | 2.834404 | -0.03643 | 0.989437 |
| RS1 | 1.782306 | 0.156621 | 0.999532 | | 1.961471 | 0.123565 | 0.955642 | | 1.736718 | -0.01532 | 0.996891 |
| H1-0 | 13.40189 | -0.05932 | 0.999532 | | 15.25715 | -0.05005 | 0.955723 | | 12.44587 | 0.027906 | 0.98506 |
| PYCARD | 3.197374 | -0.16396 | 0.999532 | | 3.417267 | -0.0941 | 0.956321 | | 2.99796 | 0.100378 | 0.959437 |
| TESK2 | 2.006648 | -0.13613 | 0.999532 | | 2.272826 | -0.11709 | 0.956341 | | 1.926103 | 0.037742 | 0.991341 |
| H2AC12 | 2.932817 | 0.415596 | 0.999532 | | 2.954699 | 0.104487 | 0.956356 | | 2.824238 | -0.29261 | 0.84579 |
| DSC3 | 3.359508 | 0.265602 | 0.999532 | | 3.349015 | -0.09695 | 0.956392 | | 3.138526 | -0.34152 | 0.803301 |
| TDRD3 | 3.103077 | 0.272853 | 0.999532 | | 3.171432 | -0.09834 | 0.956392 | | 2.881709 | -0.34531 | 0.812283 |
| AAR2 | 3.187231 | -0.09959 | 0.999532 | | 4.014781 | 0.095691 | 0.956392 | | 3.177136 | 0.215629 | 0.890219 |
| SPIN4 | 2.244027 | -0.14393 | 0.999532 | | 2.525157 | -0.11029 | 0.956392 | | 2.116384 | 0.051977 | 0.985693 |
| PGAM4 | 16.25323 | -0.0174 | 0.999532 | | 17.29017 | -0.05128 | 0.956392 | | 15.02719 | -0.02911 | 0.980921 |
| PDE9A | 2.283356 | -0.48067 | 0.999532 | | 2.860972 | 0.100232 | 0.956558 | | 2.188089 | 0.592485 | 0.679818 |
| LAT2 | 2.070773 | -0.2119 | 0.999532 | | 2.767432 | 0.106715 | 0.956558 | | 2.212121 | 0.341507 | 0.842696 |
| IHH | 2.321132 | 0.472892 | 0.999532 | | 2.190386 | 0.117509 | 0.956558 | | 2.223675 | -0.34201 | 0.838733 |
| MYO18A | 5.584407 | 0.202678 | 0.999532 | | 6.052964 | 0.075091 | 0.956558 | | 5.396782 | -0.10875 | 0.9402 |
| MILR1 | 1.873202 | 0.379471 | 0.999532 | | 1.804847 | 0.123681 | 0.956558 | | 1.834152 | -0.23999 | 0.902986 |
| THOC3 | 2.635098 | 0.268985 | 0.999532 | | 2.779378 | -0.11009 | 0.956558 | | 2.386809 | -0.35712 | 0.814402 |
| CPEB1 | 2.681887 | 0.308715 | 0.999532 | | 2.682495 | -0.10647 | 0.956558 | | 2.519051 | -0.39107 | 0.794815 |
| STRN3 | 3.177185 | -0.69472 | 0.999532 | | 4.293425 | 0.088269 | 0.956558 | | 3.002148 | 0.810961 | 0.442674 |
| TERF1 | 9.990911 | 0.12204 | 0.999532 | | 10.4093 | -0.06137 | 0.956558 | | 9.18312 | -0.15956 | 0.859898 |
| MSRB2 | 3.121384 | 0.134891 | 0.999532 | | 3.677112 | 0.090979 | 0.956558 | | 3.158169 | -0.01354 | 0.996681 |
| TTI1 | 3.408864 | 0.145121 | 0.999532 | | 3.77066 | 0.093692 | 0.956558 | | 3.370464 | -0.03314 | 0.989646 |
| KHDRBS | 3.411476 | -0.1179 | 0.999532 | | 3.804204 | 0.095815 | 0.956558 | | 3.106416 | 0.236231 | 0.88082 |
| C12orf60 | 2.036027 | 0.234819 | 0.999532 | | 2.187992 | -0.1205 | 0.956558 | | 1.965457 | -0.32894 | 0.861119 |
| SARNP | 5.590498 | 0.043387 | 0.999532 | | 6.039577 | -0.07355 | 0.956613 | | 5.107012 | -0.09374 | 0.946572 |
| ANHX | 2.297057 | 0.501106 | 0.999532 | | 2.213574 | -0.11994 | 0.956728 | | 2.220087 | -0.60028 | 0.67937 |
| IRF2BPL | 4.056831 | -0.555 | 0.999532 | | 5.338147 | -0.07628 | 0.956932 | | 3.668572 | 0.496794 | 0.65401 |
| ERBB3 | 3.388727 | 0.286034 | 0.999532 | | 3.335376 | -0.09871 | 0.956932 | | 3.027201 | -0.36161 | 0.789815 |
| PEX16 | 3.480072 | -0.23636 | 0.999532 | | 4.189102 | -0.08697 | 0.956932 | | 3.216963 | 0.166413 | 0.911296 |
| PEX2 | 1.985171 | -0.06113 | 0.999532 | | 2.155595 | 0.112748 | 0.956932 | | 1.891786 | 0.183199 | 0.929712 |
| DEPP1 | 4.199938 | -0.60697 | 0.999532 | | 5.504809 | -0.08485 | 0.956932 | | 3.769103 | 0.550995 | 0.619423 |
| CLIC2 | 3.851914 | -0.40213 | 0.999532 | | 4.747617 | 0.085913 | 0.956932 | | 3.675971 | 0.478333 | 0.705425 |
| CBLL1 | 2.275845 | 0.220554 | 0.999532 | | 2.272215 | -0.11434 | 0.956932 | | 2.181185 | -0.3175 | 0.859898 |
| RNF166 | 2.31884 | 0.032149 | 0.999532 | | 2.380706 | -0.11487 | 0.956932 | | 2.125721 | -0.13944 | 0.950895 |
| RIOK2 | 6.346068 | 0.281437 | 0.999532 | | 6.176832 | -0.08216 | 0.956932 | | 5.990099 | -0.34442 | 0.750371 |
| HEMK1 | 2.889595 | 0.278978 | 0.999532 | | 2.892919 | 0.099223 | 0.956932 | | 2.88941 | -0.16231 | 0.922922 |
| ZNF777 | 2.184864 | 0.158265 | 0.999532 | | 2.225264 | -0.11271 | 0.956932 | | 2.02435 | -0.25244 | 0.891464 |
| ASPH | 5.303314 | -0.48429 | 0.999532 | | 6.819752 | -0.07179 | 0.956932 | | 4.75458 | 0.439019 | 0.67937 |
| LCLAT1 | 3.501007 | 0.268224 | 0.999532 | | 3.617265 | 0.096682 | 0.957087 | | 3.501342 | -0.15451 | 0.924131 |
| TMEM132 | 3.104542 | 0.196378 | 0.999532 | | 3.089508 | -0.10045 | 0.957165 | | 3.013593 | -0.2738 | 0.864923 |
| ROPN1B | 2.733041 | 0.22214 | 0.999532 | | 2.772702 | -0.11478 | 0.957165 | | 2.546505 | -0.32503 | 0.84579 |
| RALBP1 | 3.955338 | 0.304509 | 0.999532 | | 4.178858 | 0.086214 | 0.957165 | | 4.088546 | -0.20152 | 0.880949 |

| | | | | | | | | | | | |
|---|---|---|---|---|---|---|---|---|---|---|---|
| SOX10 | 2.021224 | 0.206431 | 0.999532 | | 2.129167 | -0.12183 | 0.957165 | | 2.03363 | -0.30855 | 0.87591 |
| WFDC5 | 2.156047 | 0.948753 | 0.999532 | | 1.781306 | 0.1283 | 0.957165 | | 2.156172 | -0.80212 | 0.552672 |
| SPINDOC | 5.368484 | -0.00272 | 0.999532 | | 5.71425 | -0.07679 | 0.957165 | | 5.014135 | -0.06108 | 0.972221 |
| EGFL8 | 2.93145 | 0.370309 | 0.999532 | | 2.620068 | -0.11427 | 0.957165 | | 2.697896 | -0.458 | 0.760473 |
| CLASRP | 2.454275 | 0.602837 | 0.999532 | | 2.158511 | -0.11494 | 0.957165 | | 2.272119 | -0.70059 | 0.575703 |
| SMCP | 1.787237 | -0.38756 | 0.999532 | | 2.207902 | 0.117166 | 0.957165 | | 1.737569 | 0.517409 | 0.77448 |
| NEDD1 | 2.345372 | 0.007555 | 0.999532 | | 2.472682 | -0.10848 | 0.957208 | | 2.217513 | -0.10168 | 0.965876 |
| DUSP2 | 5.119856 | -0.02894 | 0.999532 | | 5.955111 | -0.08586 | 0.957411 | | 5.069582 | -0.03619 | 0.987491 |
| CD55 | 5.977086 | -0.05711 | 0.999532 | | 7.052583 | 0.084511 | 0.957411 | | 5.995577 | 0.178993 | 0.877943 |
| C20orf14 | 2.74474 | 0.68639 | 0.999532 | | 2.319012 | -0.11906 | 0.957411 | | 2.603458 | -0.79091 | 0.53177 |
| ANKRD3 | 6.711257 | -0.15988 | 0.999532 | | 8.176716 | -0.08312 | 0.957411 | | 6.482081 | 0.113782 | 0.942816 |
| MND1 | 2.114902 | 0.060371 | 0.999532 | | 2.4202 | 0.110657 | 0.957411 | | 2.220083 | 0.070939 | 0.977473 |
| ZNF317 | 4.34998 | 0.805622 | 0.999532 | | 3.74936 | 0.095572 | 0.957411 | | 4.363395 | -0.69065 | 0.465298 |
| TTL | 2.885123 | 0.301581 | 0.999532 | | 2.811197 | -0.10362 | 0.957411 | | 2.740065 | -0.38858 | 0.797544 |
| CACHD1 | 2.04102 | 0.307095 | 0.999532 | | 2.061299 | 0.118504 | 0.957411 | | 1.997798 | -0.17212 | 0.93603 |
| JRK | 2.673659 | 0.116882 | 0.999532 | | 2.809099 | -0.11078 | 0.957411 | | 2.593691 | -0.21305 | 0.906811 |
| FZD1 | 7.491205 | -0.41904 | 0.999532 | | 9.889617 | 0.066923 | 0.957411 | | 7.476626 | 0.507651 | 0.484369 |
| COX7B2 | 2.084245 | 0.349378 | 0.999532 | | 2.106996 | -0.1204 | 0.957411 | | 2.063964 | -0.44602 | 0.792538 |
| IFFO1 | 4.026941 | -0.1221 | 0.999532 | | 4.634864 | 0.082111 | 0.957411 | | 3.705106 | 0.210484 | 0.883565 |
| GNB5 | 2.529704 | 0.034878 | 0.999532 | | 2.72492 | -0.10292 | 0.957411 | | 2.420215 | -0.11793 | 0.955167 |
| RPL26L1 | 3.349039 | 0.492074 | 0.999532 | | 3.095945 | -0.10268 | 0.957411 | | 3.170137 | -0.57618 | 0.64142 |
| USF1 | 1.857669 | -0.22609 | 0.999532 | | 2.39184 | 0.111145 | 0.957411 | | 1.957643 | 0.352845 | 0.843564 |
| PFDN2 | 4.487185 | -0.15674 | 0.999532 | | 5.430198 | 0.074739 | 0.957411 | | 4.248861 | 0.247819 | 0.83987 |
| ATXN7L3 | 7.450282 | 0.320117 | 0.999532 | | 7.899944 | 0.065658 | 0.957411 | | 7.476613 | -0.23082 | 0.816259 |
| WDR61 | 4.483019 | -0.15495 | 0.999532 | | 5.415146 | 0.076416 | 0.957411 | | 4.307297 | 0.250371 | 0.836453 |
| APPBP2 | 3.598878 | -0.62882 | 0.999532 | | 4.712851 | 0.07996 | 0.957458 | | 3.46271 | 0.722755 | 0.49299 |
| DNAH5 | 2.667758 | 0.72908 | 0.999532 | | 1.986949 | -0.11802 | 0.957458 | | 2.548551 | -0.83512 | 0.540342 |
| YDJC | 1.942182 | -0.24021 | 0.999532 | | 2.232267 | -0.11242 | 0.957458 | | 1.864453 | 0.145926 | 0.948127 |
| CCSER1 | 2.437001 | 0.772045 | 0.999532 | | 2.133124 | -0.12011 | 0.957458 | | 2.364801 | -0.88052 | 0.475375 |
| RIPK4 | 1.6997 | -0.01303 | 0.999532 | | 2.012894 | 0.122922 | 0.957458 | | 1.722305 | 0.155333 | 0.948097 |
| EVI5 | 3.28782 | -0.13599 | 0.999532 | | 3.710728 | -0.08988 | 0.957458 | | 2.96857 | 0.059971 | 0.977473 |
| AKAP17A | 8.086605 | -0.16182 | 0.999532 | | 9.464719 | 0.059054 | 0.957458 | | 8.00361 | 0.242838 | 0.792539 |
| MARK4 | 3.903082 | -0.05417 | 0.999532 | | 4.666787 | 0.082002 | 0.957458 | | 4.001782 | 0.152749 | 0.914929 |
| VARS2 | 2.836846 | 0.735206 | 0.999532 | | 2.357495 | -0.10652 | 0.957458 | | 2.632557 | -0.82306 | 0.464413 |
| CD40 | 2.877825 | -0.22559 | 0.999532 | | 3.408422 | -0.09279 | 0.957458 | | 2.723559 | 0.149351 | 0.931155 |
| SYN2 | 2.385357 | 0.537426 | 0.999532 | | 2.127515 | -0.1155 | 0.957519 | | 2.33332 | -0.63848 | 0.65516 |
| PASK | 2.943695 | 0.752179 | 0.999532 | | 2.529915 | -0.10797 | 0.957519 | | 2.755232 | -0.83778 | 0.451774 |
| PLAU | 4.002091 | 0.039044 | 0.999532 | | 4.198254 | -0.08241 | 0.957622 | | 3.749815 | -0.09239 | 0.955834 |
| ITPR3 | 2.508361 | -0.69928 | 0.999532 | | 3.340238 | -0.09421 | 0.957622 | | 2.257389 | 0.625097 | 0.647218 |
| EML4 | 3.512107 | 0.127036 | 0.999532 | | 3.669351 | 0.090148 | 0.957622 | | 3.350416 | -0.01714 | 0.996109 |
| MRO | 3.342017 | 0.159445 | 0.999532 | | 3.357322 | -0.09641 | 0.957622 | | 3.073712 | -0.23958 | 0.881837 |
| ITIH4 | 2.803917 | 0.088279 | 0.999532 | | 3.273558 | 0.095199 | 0.957622 | | 2.868225 | 0.026568 | 0.992286 |
| FZD7 | 4.593493 | 0.073324 | 0.999532 | | 5.294463 | 0.078069 | 0.957622 | | 4.352569 | 0.024576 | 0.991434 |
| SPIN3 | 2.866582 | 0.305966 | 0.999532 | | 2.745751 | -0.10596 | 0.957622 | | 2.588702 | -0.39017 | 0.790999 |
| STAT6 | 13.16725 | 0.016656 | 0.999532 | | 14.51967 | 0.068773 | 0.957622 | | 12.71912 | 0.052566 | 0.971732 |
| STMN3 | 4.181948 | -0.13252 | 0.999532 | | 4.77365 | -0.08003 | 0.957622 | | 3.792895 | 0.068505 | 0.971683 |
| ABHD10 | 2.250655 | 0.186343 | 0.999532 | | 2.149486 | -0.11438 | 0.957622 | | 2.15391 | -0.28827 | 0.881837 |
| PRPS2 | 3.842931 | -0.0387 | 0.999532 | | 4.313144 | -0.08228 | 0.957622 | | 3.604274 | -0.02518 | 0.992286 |
| TMED6 | 2.941017 | 0.103152 | 0.999532 | | 3.253082 | 0.096145 | 0.957622 | | 2.932359 | 0.013603 | 0.996891 |
| GARS1 | 3.610159 | 0.612829 | 0.999532 | | 3.216294 | 0.097207 | 0.957622 | | 3.455397 | -0.50018 | 0.672452 |
| PITPNA | 2.83314 | -0.06788 | 0.999532 | | 3.313341 | 0.091284 | 0.957622 | | 2.818947 | 0.177679 | 0.911296 |
| MFSD11 | 2.83111 | -0.09796 | 0.999532 | | 2.960445 | -0.09745 | 0.957622 | | 2.458369 | 0.001903 | 0.999429 |
| ZBTB11 | 2.468612 | -0.25157 | 0.999532 | | 2.955825 | 0.095157 | 0.957693 | | 2.342576 | 0.359861 | 0.813143 |
| SETX | 5.239679 | -0.13781 | 0.999532 | | 5.887508 | -0.07929 | 0.957882 | | 4.809926 | 0.086535 | 0.952144 |
| PIGBOS1 | 5.195462 | 0.248614 | 0.999532 | | 5.546067 | -0.07528 | 0.957882 | | 5.083852 | -0.29011 | 0.796512 |
| TBC1D5 | 3.701002 | -0.61371 | 0.999532 | | 5.044493 | -0.07843 | 0.957882 | | 3.385319 | 0.559983 | 0.630705 |
| RCL1 | 2.876831 | 0.071581 | 0.999532 | | 3.365828 | 0.088834 | 0.957882 | | 2.843552 | 0.035559 | 0.989093 |
| CFAP251 | 3.090752 | 0.720282 | 0.999532 | | 2.597317 | -0.10579 | 0.957882 | | 2.99239 | -0.80933 | 0.474973 |
| DYRK4 | 2.235054 | 0.179505 | 0.999532 | | 2.203682 | -0.11287 | 0.957882 | | 2.1308 | -0.27728 | 0.881689 |
| THBS4 | 2.180622 | 0.134485 | 0.999532 | | 2.163465 | -0.11554 | 0.958047 | | 1.905641 | -0.23654 | 0.904576 |
| PACRG | 1.94249 | 0.068104 | 0.999532 | | 2.147479 | 0.111889 | 0.958047 | | 1.906897 | 0.061782 | 0.98128 |
| LMNTD1 | 2.135524 | -0.27302 | 0.999532 | | 2.485156 | -0.11437 | 0.958139 | | 1.866137 | 0.181757 | 0.933197 |
| USP36 | 5.394333 | 0.279682 | 0.999532 | | 5.495315 | 0.080129 | 0.958264 | | 5.239021 | -0.17537 | 0.892869 |
| HELZ2 | 4.371149 | 0.090179 | 0.999532 | | 4.819953 | 0.079464 | 0.958264 | | 4.126748 | 0.010932 | 0.996891 |
| TNFRSF1 | 3.665565 | 0.380816 | 0.999532 | | 3.539065 | 0.090271 | 0.958264 | | 3.435356 | -0.27089 | 0.840357 |
| SLC6A5 | 2.291883 | -0.29064 | 0.999532 | | 2.780182 | -0.10629 | 0.958264 | | 2.211061 | 0.204603 | 0.920103 |
| SMPDL3A | 3.407827 | -0.19762 | 0.999532 | | 4.167737 | 0.084698 | 0.958264 | | 3.413626 | 0.30219 | 0.818726 |
| PLEKHA | 3.063317 | -0.16516 | 0.999532 | | 3.64248 | 0.090219 | 0.958264 | | 2.900354 | 0.273193 | 0.858705 |
| SEC22B | 3.572832 | -0.16448 | 0.999532 | | 4.329331 | 0.082605 | 0.958264 | | 3.361383 | 0.267024 | 0.846125 |
| HMGCL | 3.100276 | 0.196475 | 0.999532 | | 3.215657 | 0.091179 | 0.958264 | | 3.040259 | -0.08519 | 0.965619 |
| TRIML1 | 2.243853 | 0.572153 | 0.999532 | | 2.279141 | 0.126138 | 0.958264 | | 2.193505 | -0.4146 | 0.810208 |
| SDF2 | 3.002283 | -0.27388 | 0.999532 | | 3.500911 | -0.09079 | 0.958264 | | 2.607598 | 0.203253 | 0.901033 |
| TXNL4B | 3.406513 | 0.43602 | 0.999532 | | 3.39909 | 0.093128 | 0.958264 | | 3.489409 | -0.32347 | 0.810753 |
| BEND7 | 3.593027 | 0.060325 | 0.999532 | | 4.255148 | -0.09107 | 0.958264 | | 3.508867 | -0.10623 | 0.954304 |
| FAM189B | 2.443142 | 0.062213 | 0.999532 | | 2.491137 | 0.105595 | 0.958264 | | 2.424117 | 0.04529 | 0.988853 |
| CAPN15 | 8.718119 | 0.126537 | 0.999532 | | 9.010811 | -0.06361 | 0.958264 | | 7.93198 | -0.16562 | 0.870663 |
| PLPP2 | 2.089508 | 0.175192 | 0.999532 | | 2.200136 | 0.110868 | 0.95832 | | 2.164521 | -0.05099 | 0.986447 |
| TP53INP2 | 8.631238 | -0.37029 | 0.999532 | | 11.31821 | 0.059754 | 0.958362 | | 8.552823 | 0.451476 | 0.53888 |
| TRMT44 | 4.49255 | 0.096326 | 0.999532 | | 5.097828 | -0.08141 | 0.958528 | | 4.436297 | -0.16241 | 0.90424 |
| UBE2V1 | 16.31145 | -0.23908 | 0.999532 | | 20.59873 | 0.045969 | 0.958528 | | 16.1229 | 0.312604 | 0.630705 |
| SSNA1 | 4.77704 | 0.010573 | 0.999532 | | 5.542627 | 0.075938 | 0.958712 | | 4.760854 | 0.089088 | 0.957779 |
| AIFM2 | 2.495162 | -0.32838 | 0.999532 | | 3.271362 | 0.095339 | 0.95894 | | 2.41526 | 0.442816 | 0.75311 |
| CNTN2 | 2.333536 | 0.270044 | 0.999532 | | 2.5784 | 0.10992 | 0.95894 | | 2.255464 | -0.13996 | 0.945456 |
| EDRF1 | 2.241846 | 0.09399 | 0.999532 | | 2.310362 | -0.10959 | 0.958952 | | 2.168387 | -0.18987 | 0.926513 |
| LRRC45 | 2.415461 | -0.11518 | 0.999532 | | 2.989426 | 0.095577 | 0.959061 | | 2.399846 | 0.228793 | 0.892869 |
| ZNF672 | 4.836154 | 0.309734 | 0.999532 | | 4.676914 | -0.08148 | 0.959061 | | 4.294726 | -0.36372 | 0.736481 |
| LEMD1 | 2.453435 | 0.574741 | 0.999532 | | 2.335129 | 0.112981 | 0.959061 | | 2.414285 | -0.44162 | 0.776574 |
| SLC13A2 | 2.873797 | 0.209964 | 0.999532 | | 2.904783 | 0.099483 | 0.959061 | | 2.827317 | -0.10717 | 0.959765 |
| PSMD5 | 4.530502 | 0.059515 | 0.999532 | | 4.872352 | 0.083464 | 0.959061 | | 4.269886 | 0.018454 | 0.995781 |
| CSNK1E | 8.797194 | -0.57946 | 0.999532 | | 11.98784 | 0.053735 | 0.959061 | | 8.38323 | 0.6504 | 0.287765 |
| RDH10 | 3.09584 | 0.424255 | 0.999532 | | 2.92997 | -0.10034 | 0.959061 | | 2.770883 | -0.51325 | 0.695561 |
| IDUA | 2.913486 | 0.65159 | 0.999532 | | 2.551122 | -0.10076 | 0.959061 | | 2.767713 | -0.73443 | 0.52058 |
| HLA-G | 9.825499 | 0.523813 | 0.999532 | | 9.272482 | 0.090621 | 0.959061 | | 9.588367 | -0.40643 | 0.699971 |
| TBC1D7 | 2.598774 | -0.13459 | 0.999532 | | 2.967981 | -0.09643 | 0.959061 | | 2.356356 | 0.056855 | 0.981715 |
| RSPRY1 | 5.292683 | 0.083239 | 0.999532 | | 5.892197 | 0.073915 | 0.959061 | | 5.139802 | 0.009152 | 0.996891 |
| MAGEH1 | 3.720531 | -0.55894 | 0.999532 | | 4.866156 | 0.072569 | 0.959061 | | 3.629489 | 0.642039 | 0.535079 |
| IST1 | 3.617936 | -0.17371 | 0.999532 | | 4.323685 | 0.080673 | 0.959061 | | 3.413346 | 0.27521 | 0.834092 |
| RASL12 | 3.080727 | 0.236838 | 0.999532 | | 3.157076 | -0.0964 | 0.959061 | | 3.044439 | -0.31011 | 0.839811 |
| RCE1 | 2.197976 | 0.157338 | 0.999532 | | 2.518884 | 0.098772 | 0.959061 | | 2.281939 | -0.0349 | 0.991159 |
| PITPNM2 | 2.221586 | -0.14171 | 0.999532 | | 2.562077 | 0.101782 | 0.959061 | | 2.173479 | 0.267647 | 0.881837 |
| AMOT | 5.889258 | 0.073039 | 0.999532 | | 6.22354 | -0.06922 | 0.959061 | | 5.532923 | -0.12563 | 0.923864 |
| PRIM2 | 2.741248 | 0.160884 | 0.999532 | | 2.960993 | -0.09846 | 0.959061 | | 2.666923 | -0.23477 | 0.892869 |
| BPNT2 | 3.18601 | -0.37875 | 0.999532 | | 3.998844 | 0.081054 | 0.959061 | | 3.057683 | 0.473031 | 0.69872 |
| EXOSC4 | 2.673678 | -0.19691 | 0.999532 | | 3.23197 | 0.089765 | 0.959061 | | 2.59365 | 0.305001 | 0.842754 |
| SGO2 | 2.088133 | 0.22644 | 0.999532 | | 1.960039 | -0.11548 | 0.959061 | | 1.935771 | -0.32682 | 0.862823 |
| CAV2 | 2.840909 | 0.304613 | 0.999532 | | 2.704849 | -0.10188 | 0.959061 | | 2.651315 | -0.38138 | 0.791576 |
| LARP1B | 2.930289 | -0.23667 | 0.999532 | | 3.459799 | 0.087073 | 0.959061 | | 2.845215 | 0.33606 | 0.82268 |
| UAP1 | 4.526315 | 0.395353 | 0.999532 | | 4.424498 | 0.077433 | 0.959061 | | 4.417148 | -0.30017 | 0.791576 |
| RGL2 | 5.067099 | -0.02183 | 0.999532 | | 5.666897 | 0.072804 | 0.959061 | | 4.733366 | 0.115865 | 0.93275 |
| RARA | 4.384858 | -0.34669 | 0.999532 | | 5.426762 | -0.07588 | 0.959061 | | 4.158374 | 0.289173 | 0.811123 |
| MYL4 | 2.03349 | -0.24781 | 0.999532 | | 2.351407 | 0.106354 | 0.959061 | | 1.981194 | 0.347248 | 0.859522 |
| METTL26 | 3.434661 | 0.260262 | 0.999532 | | 3.459507 | 0.088046 | 0.959061 | | 3.371555 | -0.15259 | 0.923864 |
| E4F1 | 4.533137 | 0.433559 | 0.999532 | | 4.514472 | -0.08089 | 0.959061 | | 4.374922 | -0.49455 | 0.628053 |
| PITPNM1 | 3.344805 | 0.069707 | 0.999532 | | 3.734124 | 0.084513 | 0.959061 | | 3.257315 | 0.029715 | 0.991159 |
| BRD8 | 3.613925 | -0.11261 | 0.999532 | | 3.990893 | -0.08261 | 0.959061 | | 3.393164 | 0.055494 | 0.978229 |
| SLC30A2 | 2.702958 | 0.197021 | 0.999532 | | 2.817465 | -0.09858 | 0.959061 | | 2.630976 | -0.27359 | 0.874251 |
| SCN4A | 2.653785 | 0.30374 | 0.999532 | | 2.606384 | -0.10347 | 0.959061 | | 2.434801 | -0.38652 | 0.795612 |
| CTAGE1 | 2.534236 | 0.130991 | 0.999532 | | 2.75025 | 0.102985 | 0.959061 | | 2.507271 | -0.01829 | 0.996681 |
| MOB3C | 3.521187 | -0.73933 | 0.999532 | | 4.337239 | -0.08168 | 0.959061 | | 3.108287 | 0.689152 | 0.556866 |
| RNF167 | 4.860146 | -0.24176 | 0.999532 | | 6.037923 | -0.07355 | 0.959061 | | 4.677442 | 0.194345 | 0.88476 |
| IL12RB1 | 1.849097 | -0.23858 | 0.999532 | | 2.277221 | 0.108792 | 0.959061 | | 1.934389 | 0.360709 | 0.856551 |
| PTPN18 | 5.554039 | 0.140384 | 0.999532 | | 5.557787 | -0.07308 | 0.959061 | | 5.159826 | -0.19527 | 0.870663 |
| ZNF431 | 2.563576 | 0.617446 | 0.999532 | | 2.541063 | 0.107445 | 0.959061 | | 2.661331 | -0.48888 | 0.722703 |
| ETV2 | 2.520341 | 0.134608 | 0.999532 | | 2.829546 | -0.10275 | 0.959061 | | 2.499214 | -0.21351 | 0.905952 |
| UGT2B11 | 2.142594 | 0.509548 | 0.999532 | | 2.207586 | 0.1108 | 0.959061 | | 2.057064 | -0.37504 | 0.816618 |
| AFAP1 | 3.215855 | 0.590834 | 0.999532 | | 3.139718 | 0.094083 | 0.959061 | | 3.184281 | -0.47617 | 0.692526 |
| PIP | 1.958103 | 0.126738 | 0.999532 | | 2.172922 | -0.11767 | 0.959061 | | 1.95556 | -0.22693 | 0.911296 |
| PKLR | 2.333684 | 0.239797 | 0.999532 | | 2.400669 | 0.108195 | 0.959061 | | 2.091858 | -0.10957 | 0.960773 |
| IRAK4 | 3.012056 | 0.201926 | 0.999532 | | 3.148576 | -0.09264 | 0.959061 | | 2.922713 | -0.27853 | 0.853018 |
| PPIH | 5.218082 | 0.464219 | 0.999532 | | 5.014369 | -0.07841 | 0.959061 | | 5.061338 | -0.52758 | 0.565022 |
| RIT2 | 2.422466 | 0.551777 | 0.999532 | | 2.066772 | -0.11193 | 0.959061 | | 2.341311 | -0.63694 | 0.666566 |
| C6orf58 | 2.037213 | 0.374136 | 0.999532 | | 1.855377 | -0.1185 | 0.959061 | | 1.862609 | -0.47748 | 0.776852 |
| ADCYAP1 | 1.973733 | -0.17064 | 0.999532 | | 2.364228 | -0.11038 | 0.959061 | | 1.865745 | 0.079254 | 0.976381 |
| AR | 4.476454 | -0.13361 | 0.999532 | | 5.290506 | -0.07244 | 0.959061 | | 4.24748 | 0.08113 | 0.95999 |
| CDH13 | 3.931818 | -0.27273 | 0.999532 | | 5.131254 | 0.086136 | 0.959061 | | 4.143191 | 0.369289 | 0.760473 |
| VDAC3 | 4.791512 | -0.07212 | 0.999532 | | 5.004944 | -0.07742 | 0.959061 | | 4.203102 | -0.00259 | 0.999385 |
| RNF145 | 5.788827 | -0.03327 | 0.999532 | | 6.789653 | 0.066043 | 0.959061 | | 5.844469 | 0.117448 | 0.922647 |
| NPC1 | 2.516282 | -0.13921 | 0.999532 | | 2.818913 | -0.09968 | 0.959061 | | 2.325871 | 0.059658 | 0.979996 |
| CPLANE1 | 3.440352 | 0.120446 | 0.999532 | | 3.962651 | 0.085293 | 0.959061 | | 3.378825 | -0.00355 | 0.999139 |
| SP140L | 5.201411 | -0.0106 | 0.999532 | | 5.928244 | -0.07835 | 0.959061 | | 4.890666 | -0.04777 | 0.978927 |
| ZC3H4 | 6.618748 | 0.081774 | 0.999532 | | 7.231219 | 0.066019 | 0.959061 | | 6.282812 | -0.00821 | 0.996891 |
| COL4A5 | 3.703771 | 0.105068 | 0.999532 | | 4.153108 | 0.082512 | 0.959061 | | 3.567573 | -0.00447 | 0.99858 |
| FHIP2B | 4.922403 | 0.12779 | 0.999532 | | 5.311398 | -0.07266 | 0.959061 | | 4.643287 | -0.17886 | 0.885712 |
| DOK7 | 4.141906 | -0.09436 | 0.999532 | | 5.030035 | -0.08137 | 0.959061 | | 4.020998 | 0.063964 | 0.974808 |
| UCK1 | 2.541183 | 0.294055 | 0.999532 | | 2.653394 | 0.096263 | 0.959061 | | 2.602344 | -0.17526 | 0.923266 |
| F11R | 3.593082 | -0.37428 | 0.999532 | | 4.864738 | 0.081459 | 0.959061 | | 3.746674 | 0.479688 | 0.694514 |
| SMARCD | 2.445324 | 0.449144 | 0.999532 | | 2.318646 | -0.10696 | 0.959061 | | 2.382043 | -0.54258 | 0.698562 |
| DHFR2 | 1.958079 | -0.00315 | 0.999532 | | 2.176604 | -0.10787 | 0.959061 | | 1.873856 | -0.08338 | 0.974421 |
| PRKAG2 | 2.767612 | 0.220034 | 0.999532 | | 2.838367 | 0.097754 | 0.959061 | | 2.659612 | -0.10679 | 0.958191 |
| CD163 | 2.889232 | -0.43554 | 0.999532 | | 3.830007 | 0.090817 | 0.959061 | | 2.767071 | 0.541807 | 0.686214 |
| TMEM200 | 5.089511 | 0.107991 | 0.999532 | | 5.428591 | -0.08368 | 0.959061 | | 4.59711 | -0.15326 | 0.923266 |
| CBFA2T3 | 2.666964 | -0.11603 | 0.999532 | | 3.095704 | -0.09936 | 0.959061 | | 2.492464 | 0.046325 | 0.987274 |
| ARV1 | 2.213898 | 0.297953 | 0.999532 | | 2.335513 | 0.105329 | 0.959061 | | 2.152691 | -0.17463 | 0.92697 |
| ANAPC2 | 4.124731 | 0.238005 | 0.999532 | | 4.079124 | -0.08359 | 0.959061 | | 3.583103 | -0.30662 | 0.805922 |
| RFC3 | 2.363652 | 0.260653 | 0.999532 | | 2.452718 | -0.10389 | 0.959061 | | 2.147879 | -0.34395 | 0.836935 |
| SPTB | 4.923663 | 0.484012 | 0.999532 | | 5.020707 | 0.078658 | 0.959061 | | 4.975302 | -0.38607 | 0.705046 |
| NSD3 | 5.035159 | -0.16045 | 0.999532 | | 5.932861 | -0.07275 | 0.959061 | | 4.840119 | 0.105825 | 0.941197 |
| SCFD2 | 4.40789 | 0.252156 | 0.999532 | | 4.250177 | -0.07946 | 0.959061 | | 4.091567 | -0.32178 | 0.791466 |
| COQ8B | 3.365754 | -0.11193 | 0.999532 | | 3.346781 | -0.08546 | 0.959061 | | 2.988061 | 0.004741 | 0.998586 |
| PIGX | 2.906855 | 0.648232 | 0.999532 | | 2.398904 | -0.10212 | 0.959061 | | 2.65668 | -0.73535 | 0.536289 |
| HSPBP1 | 2.220868 | -0.26281 | 0.999532 | | 2.74127 | -0.10342 | 0.959061 | | 2.211999 | 0.180096 | 0.931837 |
| COX7A2 | 6.935795 | -0.40564 | 0.999532 | | 8.928219 | 0.064035 | 0.959061 | | 6.515284 | 0.487219 | 0.612811 |
| PHOSPH | 4.588705 | 0.187528 | 0.999532 | | 4.605487 | -0.08077 | 0.959061 | | 4.296816 | -0.2593 | 0.859426 |
| LAMP5 | 2.042796 | 0.506423 | 0.999532 | | 1.889385 | 0.115535 | 0.959061 | | 2.044954 | -0.37796 | 0.829449 |
| ZHX1 | 2.953151 | -0.05053 | 0.999532 | | 3.204314 | -0.09352 | 0.959061 | | 2.599288 | -0.02678 | 0.992459 |
| LRRC49 | 1.950683 | 0.448325 | 0.999532 | | 1.940832 | 0.113287 | 0.959061 | | 1.909717 | -0.31648 | 0.859583 |
| CD19 | 2.002027 | -0.33812 | 0.999532 | | 2.408719 | -0.10832 | 0.959061 | | 1.90336 | 0.24807 | 0.903339 |
| LGSN | 2.309169 | 0.662683 | 0.999532 | | 2.027536 | -0.1217 | 0.959061 | | 2.355188 | -0.76052 | 0.628053 |
| TMEM86B | 4.481691 | -0.02459 | 0.999532 | | 5.069062 | -0.09921 | 0.959061 | | 4.330565 | -0.07756 | 0.971732 |
| MRS2 | 3.545819 | 0.661847 | 0.999532 | | 3.012537 | 0.10226 | 0.959061 | | 3.28466 | -0.54084 | 0.67937 |
| RNF149 | 5.788003 | -0.39552 | 0.999532 | | 7.348642 | 0.063262 | 0.959061 | | 5.414152 | 0.471178 | 0.598934 |
| FUCA1 | 2.526747 | 0.260914 | 0.999532 | | 2.799226 | 0.100722 | 0.959061 | | 2.533841 | -0.14426 | 0.938429 |
| BIRC3 | 2.369862 | 0.722746 | 0.999532 | | 2.393997 | 0.108036 | 0.959061 | | 2.476924 | -0.58607 | 0.666566 |
| NR2C2 | 6.125602 | -0.50971 | 0.999532 | | 8.152346 | -0.06151 | 0.959061 | | 5.772126 | 0.478514 | 0.603483 |
| TSPYL6 | 1.853584 | 0.186655 | 0.999532 | | 1.912919 | 0.117179 | 0.959061 | | 1.844154 | -0.05144 | 0.987274 |
| GRIP1 | 1.885266 | 0.053675 | 0.999532 | | 1.93467 | -0.11465 | 0.959061 | | 1.729944 | -0.15257 | 0.946765 |
| MEA1 | 2.761419 | -0.22907 | 0.999532 | | 3.561324 | 0.087801 | 0.959061 | | 2.621471 | 0.328132 | 0.82381 |
| WNT11 | 2.845333 | 0.068882 | 0.999532 | | 3.265906 | 0.10115 | 0.959061 | | 3.048379 | 0.039854 | 0.989638 |
| PDSS2 | 2.574119 | 0.31169 | 0.999532 | | 2.505371 | -0.10323 | 0.959061 | | 2.381993 | -0.39599 | 0.79686 |
| CRLF3 | 2.895201 | 0.22177 | 0.999532 | | 2.887054 | -0.0954 | 0.959061 | | 2.810801 | -0.30361 | 0.842754 |
| RNF144B | 4.218655 | -0.54579 | 0.999532 | | 5.283073 | -0.07833 | 0.959061 | | 3.969862 | 0.47828 | 0.69977 |
| LMBR1L | 2.951221 | 0.062633 | 0.999532 | | 3.083158 | -0.09319 | 0.959061 | | 2.828134 | -0.13789 | 0.941197 |
| PROS1 | 3.935576 | -0.19653 | 0.999532 | | 4.790157 | 0.075634 | 0.959061 | | 3.878937 | 0.289767 | 0.813127 |
| ATRAID | 3.204701 | -0.47624 | 0.999532 | | 4.050329 | 0.079411 | 0.959061 | | 3.04121 | 0.561371 | 0.647218 |
| ZDHHC8 | 7.377807 | 0.28679 | 0.999532 | | 7.47152 | 0.064615 | 0.959061 | | 6.975708 | -0.2034 | 0.836935 |
| FAM83G | 2.564455 | 0.11408 | 0.999532 | | 2.754323 | -0.09945 | 0.959061 | | 2.345386 | -0.18953 | 0.914929 |
| IL1RL2 | 2.097515 | 0.136238 | 0.999532 | | 2.280308 | 0.109224 | 0.959061 | | 2.041929 | -0.01662 | 0.996891 |
| TRAFD1 | 3.134724 | -0.15205 | 0.999532 | | 3.74674 | 0.085834 | 0.959061 | | 2.994407 | 0.251493 | 0.871957 |
| LPIN1 | 2.378316 | -0.24538 | 0.999532 | | 2.788299 | 0.091659 | 0.959085 | | 2.332734 | 0.356246 | 0.816618 |
| CCDC83 | 2.423912 | 0.567315 | 0.999532 | | 2.313071 | 0.112792 | 0.959085 | | 2.520472 | -0.44205 | 0.791502 |
| FAM219B | 4.073173 | -0.04977 | 0.999532 | | 4.829844 | -0.07569 | 0.959085 | | 3.809282 | -0.006 | 0.997967 |
| PIGP | 5.70674 | -0.28898 | 0.999532 | | 6.825429 | 0.064631 | 0.959085 | | 5.532985 | 0.365732 | 0.715437 |
| GBP2 | 6.920601 | -0.23045 | 0.999532 | | 8.120159 | -0.08707 | 0.959085 | | 6.517885 | 0.162423 | 0.892646 |
| UBOX5 | 2.384165 | 0.548897 | 0.999532 | | 1.954096 | -0.11533 | 0.959085 | | 2.230805 | -0.65114 | 0.673405 |
| ENPP3 | 2.342059 | 0.167243 | 0.999532 | | 2.425343 | -0.10797 | 0.959099 | | 2.222685 | -0.25762 | 0.893262 |
| OR6C65 | 2.191909 | 0.483725 | 0.999532 | | 2.132768 | 0.108866 | 0.959099 | | 2.129306 | -0.35948 | 0.8233 |
| TTC4 | 3.974728 | 0.381959 | 0.999532 | | 3.650111 | -0.08549 | 0.959099 | | 3.690912 | -0.44405 | 0.723933 |
| GPSM2 | 2.23673 | 0.645193 | 0.999532 | | 2.035065 | 0.113097 | 0.959099 | | 2.188002 | -0.51335 | 0.735469 |
| CDK20 | 2.800127 | 0.28501 | 0.999532 | | 2.912789 | 0.091604 | 0.959744 | | 2.725154 | -0.17504 | 0.914813 |
| GAPT | 2.035243 | 0.379794 | 0.999532 | | 2.122048 | 0.109143 | 0.959744 | | 1.952026 | -0.25014 | 0.892869 |
| PWP2 | 1.852214 | 0.291513 | 0.999532 | | 1.838616 | 0.116037 | 0.959744 | | 1.842414 | -0.15937 | 0.945129 |
| LRRC17 | 2.040283 | -0.20544 | 0.999532 | | 2.303867 | -0.10126 | 0.959815 | | 1.905949 | 0.120401 | 0.958698 |
| UBE2O | 3.135885 | -0.20095 | 0.999532 | | 3.569775 | -0.08862 | 0.959847 | | 2.858735 | 0.133012 | 0.941197 |
| XPO5 | 3.196668 | -0.22765 | 0.999532 | | 3.515124 | -0.08308 | 0.959847 | | 2.988202 | 0.160345 | 0.922679 |
| ZXDC | 4.115009 | 0.470537 | 0.999532 | | 3.711257 | -0.08159 | 0.959847 | | 3.788789 | -0.53768 | 0.623565 |
| KIRREL2 | 2.217359 | 0.112294 | 0.999532 | | 2.352195 | -0.1038 | 0.959847 | | 2.037408 | -0.19683 | 0.918194 |
| BMPR1A | 3.401813 | 0.137114 | 0.999532 | | 3.913109 | 0.083714 | 0.959847 | | 3.289754 | -0.03022 | 0.991159 |
| ZBTB7C | 4.129181 | 0.046746 | 0.999532 | | 4.422625 | -0.07622 | 0.959847 | | 3.775114 | -0.10552 | 0.946572 |
| IL10 | 2.047747 | 0.528731 | 0.999532 | | 1.871305 | -0.1142 | 0.959847 | | 1.93022 | -0.6258 | 0.680428 |
| ADNP2 | 3.671829 | -0.067 | 0.999532 | | 4.000653 | -0.08211 | 0.959847 | | 3.598152 | 0.003098 | 0.999385 |
| CEP55 | 3.509148 | 0.232115 | 0.999532 | | 3.296213 | -0.08641 | 0.959847 | | 3.302749 | -0.30619 | 0.82526 |
| ASL | 3.03892 | 0.167991 | 0.999532 | | 3.187343 | 0.093724 | 0.959847 | | 3.019807 | -0.06176 | 0.977473 |
| GTPBP8 | 3.155805 | -0.04461 | 0.999532 | | 3.517859 | -0.09278 | 0.959847 | | 2.917535 | -0.02758 | 0.992286 |
| ASAP1 | 5.237349 | -0.3753 | 0.999532 | | 6.366182 | -0.07114 | 0.960152 | | 4.988858 | 0.322821 | 0.776852 |
| PRR15 | 2.084402 | 0.503443 | 0.999532 | | 2.083093 | 0.107543 | 0.960152 | | 2.136659 | -0.37924 | 0.814133 |
| UNC5C | 2.449868 | 0.128654 | 0.999532 | | 2.506399 | -0.10346 | 0.960152 | | 2.42224 | -0.23095 | 0.903054 |
| TTC31 | 4.974156 | -0.04249 | 0.999532 | | 5.843074 | 0.071212 | 0.960152 | | 4.994744 | 0.136876 | 0.920487 |
| ZBTB33 | 2.361221 | 0.49807 | 0.999532 | | 2.080285 | 0.107248 | 0.960152 | | 2.284375 | -0.37613 | 0.814402 |
| HES4 | 4.72436 | 0.02393 | 0.999532 | | 5.005445 | -0.07454 | 0.960152 | | 4.463434 | -0.07657 | 0.96556 |
| KANK2 | 6.982528 | -0.16758 | 0.999532 | | 8.691659 | 0.068748 | 0.960152 | | 6.694076 | 0.246419 | 0.800239 |
| VLDLR | 2.705719 | -0.29972 | 0.999532 | | 3.194797 | 0.084794 | 0.960152 | | 2.621203 | 0.39836 | 0.783005 |
| VRK2 | 2.601165 | -0.14457 | 0.999532 | | 2.955894 | -0.09179 | 0.960152 | | 2.311108 | 0.071066 | 0.974588 |
| DDX52 | 3.788467 | 0.423757 | 0.999532 | | 3.406744 | -0.08555 | 0.960152 | | 3.432634 | -0.48997 | 0.66549 |
| SRPK3 | 2.057537 | 0.393155 | 0.999532 | | 1.89562 | -0.1107 | 0.960152 | | 1.890783 | -0.48514 | 0.761861 |
| RBM14 | 3.647259 | 0.140678 | 0.999532 | | 3.861803 | 0.084729 | 0.960152 | | 3.757748 | -0.0485 | 0.982187 |
| VPS9D1 | 2.428477 | 0.06002 | 0.999532 | | 2.791661 | 0.097132 | 0.960152 | | 2.434101 | 0.056262 | 0.982124 |
| SYTL3 | 2.347713 | 0.118726 | 0.999532 | | 2.593159 | -0.10623 | 0.960152 | | 2.283303 | -0.21488 | 0.906811 |
| EIPR1 | 2.011038 | 0.286834 | 0.999532 | | 2.006196 | -0.11261 | 0.960152 | | 1.888165 | -0.38101 | 0.829716 |
| PTPDC1 | 2.749343 | 0.295336 | 0.999532 | | 2.558518 | -0.09769 | 0.960152 | | 2.509715 | -0.37783 | 0.803942 |
| PIK3C2G | 2.211978 | 0.426669 | 0.999532 | | 2.094614 | -0.10737 | 0.960152 | | 2.151489 | -0.51886 | 0.728588 |
| CADPS2 | 3.450372 | 0.154063 | 0.999532 | | 3.668255 | -0.08543 | 0.960152 | | 3.256608 | -0.22209 | 0.87892 |
| ELANE | 1.715373 | -0.34294 | 0.999532 | | 2.2461 | 0.107095 | 0.960152 | | 1.830832 | 0.465828 | 0.799835 |
| ZNF692 | 4.233577 | -0.18145 | 0.999532 | | 4.988887 | -0.07537 | 0.960152 | | 4.171671 | 0.124946 | 0.938429 |
| BBS12 | 1.985476 | 0.398734 | 0.999532 | | 1.892968 | -0.11488 | 0.960152 | | 1.851432 | -0.49341 | 0.765852 |
| ZIK1 | 2.853729 | 0.470399 | 0.999532 | | 2.716496 | -0.10395 | 0.960306 | | 2.655532 | -0.55745 | 0.673659 |
| BAZ2A | 5.236548 | -0.0326 | 0.999532 | | 5.850625 | 0.07003 | 0.960354 | | 5.084109 | 0.127458 | 0.92463 |
| ACBD4 | 2.1106 | -0.55578 | 0.999532 | | 2.625194 | -0.095 | 0.960686 | | 1.902609 | 0.476092 | 0.77059 |
| LSMEM1 | 2.319278 | 0.002155 | 0.999532 | | 2.626215 | 0.101473 | 0.960735 | | 2.373441 | 0.122488 | 0.956011 |
| RANBP10 | 5.099712 | -0.0726 | 0.999532 | | 6.015382 | 0.072391 | 0.960735 | | 5.077738 | 0.160342 | 0.901813 |
| MLLT3 | 6.241976 | 0.068367 | 0.999532 | | 6.847135 | -0.06853 | 0.960735 | | 5.591037 | -0.1391 | 0.907163 |
| COLQ | 1.93281 | -0.01565 | 0.999532 | | 2.186791 | -0.10432 | 0.960736 | | 1.828789 | -0.0693 | 0.978312 |
| PHF13 | 4.825994 | -0.00781 | 0.999532 | | 5.616567 | 0.072178 | 0.960736 | | 4.722387 | 0.101533 | 0.946572 |
| DNTT | 2.14288 | 0.042746 | 0.999532 | | 2.301217 | -0.10819 | 0.960736 | | 2.08383 | -0.14228 | 0.948141 |
| NGLY1 | 3.139958 | 0.39831 | 0.999532 | | 2.89067 | -0.09373 | 0.960767 | | 3.001161 | -0.4824 | 0.709825 |
| EIF5B | 4.271854 | -0.06162 | 0.999532 | | 4.896783 | -0.07768 | 0.960767 | | 3.940142 | 0.009117 | 0.996891 |
| MAPKBP | 11.0475 | -0.08349 | 0.999532 | | 12.08475 | -0.06431 | 0.960767 | | 10.3122 | 0.017706 | 0.994537 |
| TFEC | 1.858773 | 0.103671 | 0.999532 | | 2.012061 | 0.108311 | 0.960798 | | 1.78485 | 0.021032 | 0.996681 |
| H2AW | 2.68291 | 0.012462 | 0.999532 | | 3.196158 | 0.091958 | 0.960955 | | 2.867681 | 0.096875 | 0.96556 |
| ACP2 | 3.372849 | 0.2047 | 0.999532 | | 3.346697 | -0.08743 | 0.960955 | | 2.946931 | -0.27262 | 0.852179 |
| ZNF623 | 4.434567 | -0.10538 | 0.999532 | | 4.975317 | -0.07048 | 0.960955 | | 4.040641 | 0.04847 | 0.978487 |
| GPBP1 | 2.76389 | -0.45013 | 0.999532 | | 3.53853 | 0.085025 | 0.960955 | | 2.583697 | 0.553948 | 0.668481 |
| DAAM2 | 13.83451 | -0.25759 | 0.999532 | | 16.72866 | -0.04873 | 0.960978 | | 13.14503 | 0.230315 | 0.7611 |
| INPP1 | 3.636303 | -0.1723 | 0.999532 | | 4.172682 | 0.084308 | 0.961006 | | 3.202394 | 0.302921 | 0.823066 |
| TAS2R16 | 1.840127 | 0.428749 | 0.999532 | | 1.917791 | 0.111554 | 0.961006 | | 1.774855 | -0.2985 | 0.877268 |
| SEC63 | 7.449911 | -0.37416 | 0.999532 | | 9.47751 | -0.05895 | 0.961006 | | 7.143079 | 0.33387 | 0.705187 |
| ZNF740 | 3.462179 | -0.11309 | 0.999532 | | 4.070135 | 0.077879 | 0.961006 | | 3.542745 | 0.215692 | 0.881637 |
| MTF2 | 2.671974 | 0.017196 | 0.999532 | | 2.673087 | -0.09456 | 0.961006 | | 2.384552 | -0.09652 | 0.965649 |
| DDB2 | 2.311264 | 0.182296 | 0.999532 | | 2.371128 | -0.0986 | 0.961006 | | 2.245856 | -0.26494 | 0.879579 |
| ZNF563 | 2.78448 | 0.396122 | 0.999532 | | 2.581814 | -0.09778 | 0.961006 | | 2.594171 | -0.47408 | 0.736481 |
| BEX1 | 2.156952 | 0.130621 | 0.999532 | | 2.312364 | -0.10349 | 0.961006 | | 2.051516 | -0.21127 | 0.912867 |
| P4HA2 | 5.479895 | 0.537019 | 0.999532 | | 4.646793 | 0.071506 | 0.961006 | | 5.21075 | -0.46777 | 0.647218 |
| COX6A1 | 23.44445 | -0.1842 | 0.999532 | | 28.7949 | 0.036746 | 0.961006 | | 22.6091 | 0.236711 | 0.666566 |
| RTN3 | 6.859265 | -0.07921 | 0.999532 | | 7.708847 | 0.059443 | 0.961006 | | 6.634866 | 0.154757 | 0.890225 |
| ZNF416 | 2.772833 | 0.681328 | 0.999532 | | 2.187167 | -0.10686 | 0.961006 | | 2.552387 | -0.7646 | 0.557909 |
| MCFD2 | 4.358448 | -0.39683 | 0.999532 | | 5.767356 | -0.06856 | 0.961006 | | 4.095302 | 0.356897 | 0.756297 |
| C17orf75 | 2.865885 | 0.511972 | 0.999532 | | 2.740852 | 0.088697 | 0.961006 | | 2.815984 | -0.40549 | 0.7494 |
| ASIC4 | 2.110825 | 0.043835 | 0.999532 | | 2.152272 | 0.10542 | 0.961006 | | 2.131453 | 0.048794 | 0.988853 |
| RXFP4 | 1.897251 | 0.679488 | 0.999532 | | 1.796368 | 0.11535 | 0.961006 | | 1.887172 | -0.54551 | 0.725338 |
| RASIP1 | 3.202598 | -0.22568 | 0.999532 | | 4.157104 | -0.08135 | 0.961006 | | 3.199467 | 0.195273 | 0.906811 |
| CD1C | 2.710811 | 0.189476 | 0.999532 | | 2.588299 | -0.0931 | 0.961006 | | 2.452186 | -0.28282 | 0.8691 |
| C15orf65 | 2.736967 | -0.09362 | 0.999532 | | 3.205171 | -0.09424 | 0.961006 | | 2.797121 | 0.018688 | 0.996681 |
| MAN2A2 | 3.883409 | 0.169785 | 0.999532 | | 4.254762 | 0.072772 | 0.961006 | | 3.840345 | -0.07577 | 0.965649 |
| TANGO2 | 6.454166 | -0.07292 | 0.999532 | | 7.560069 | 0.065568 | 0.961006 | | 6.086312 | 0.164467 | 0.889853 |
| EEFSEC | 1.929863 | 0.078389 | 0.999532 | | 2.081819 | 0.101397 | 0.961006 | | 1.810807 | 0.033838 | 0.992286 |
| ZNF688 | 2.75552 | 0.567748 | 0.999532 | | 2.547373 | 0.095465 | 0.961006 | | 2.702007 | -0.45516 | 0.727219 |
| CLDN3 | 2.066802 | -0.08736 | 0.999532 | | 2.27002 | -0.0987 | 0.961006 | | 1.962791 | 0.007458 | 0.997967 |
| DERL1 | 2.864632 | -0.03016 | 0.999532 | | 3.11374 | 0.086235 | 0.961006 | | 2.823574 | 0.136332 | 0.939913 |
| ALDH5A1 | 2.699951 | 0.502449 | 0.999532 | | 2.444515 | -0.09971 | 0.961006 | | 2.529701 | -0.5855 | 0.663633 |
| KCNS3 | 2.409514 | 0.881298 | 0.999532 | | 1.801869 | 0.112967 | 0.961006 | | 2.329709 | -0.74996 | 0.595157 |
| SNTB2 | 2.786685 | -0.32256 | 0.999532 | | 3.449117 | 0.085476 | 0.961006 | | 2.772003 | 0.429589 | 0.75311 |
| DIS3 | 2.570864 | -0.02219 | 0.999532 | | 3.095907 | 0.087127 | 0.961006 | | 2.532565 | 0.12766 | 0.945129 |
| SNCAIP | 2.40195 | -0.04232 | 0.999532 | | 2.930484 | 0.092309 | 0.961006 | | 2.556675 | 0.152748 | 0.939549 |
| GTF3C3 | 2.616362 | -0.31904 | 0.999532 | | 3.17847 | -0.08492 | 0.961006 | | 2.318222 | 0.253467 | 0.87813 |
| DNAJB7 | 1.998058 | 0.593737 | 0.999532 | | 1.723403 | -0.11451 | 0.961006 | | 1.936948 | -0.6901 | 0.658914 |
| COLGALT | 3.842476 | 0.027226 | 0.999532 | | 3.877173 | -0.07977 | 0.961006 | | 3.437665 | -0.08718 | 0.96129 |
| LYPLA1 | 4.54766 | -0.07321 | 0.999532 | | 5.207039 | -0.07119 | 0.961006 | | 4.345399 | 0.02215 | 0.992286 |
| FAM227A | 1.830725 | 0.145478 | 0.999532 | | 1.959437 | -0.10702 | 0.961006 | | 1.707197 | -0.23007 | 0.908891 |
| ARRDC2 | 2.614079 | -0.45976 | 0.999532 | | 3.273765 | -0.09962 | 0.961006 | | 2.26791 | 0.388446 | 0.815055 |
| DUX1 | 3.424805 | 0.129163 | 0.999532 | | 4.039678 | 0.082597 | 0.961006 | | 3.530145 | -0.02646 | 0.992286 |
| CCDC17 | 2.118055 | -0.05243 | 0.999532 | | 2.466247 | -0.09808 | 0.961006 | | 1.974593 | -0.0265 | 0.993618 |
| GGA1 | 2.530365 | 0.144848 | 0.999532 | | 2.601559 | -0.09454 | 0.961006 | | 2.350087 | -0.22132 | 0.896973 |
| CIAO1 | 3.04706 | -0.3095 | 0.999532 | | 3.722234 | 0.080106 | 0.961006 | | 2.880089 | 0.408085 | 0.752562 |
| ALKBH7 | 3.175097 | -0.47377 | 0.999532 | | 4.315291 | 0.074644 | 0.961006 | | 3.047155 | 0.569045 | 0.647218 |
| ABCA5 | 3.191 | 0.002438 | 0.999532 | | 3.67361 | -0.08318 | 0.961006 | | 2.885618 | -0.05828 | 0.978229 |
| EHBP1 | 2.900144 | 0.46472 | 0.999532 | | 2.590314 | 0.093427 | 0.961006 | | 2.788868 | -0.36571 | 0.803301 |
| MCEE | 3.600511 | 0.064496 | 0.999532 | | 3.739841 | 0.081306 | 0.961006 | | 3.431845 | 0.03222 | 0.989985 |
| DNAJC21 | 4.626918 | -0.002 | 0.999532 | | 5.088205 | -0.07153 | 0.961006 | | 4.318566 | -0.05056 | 0.977514 |
| STAU1 | 4.983775 | -0.41262 | 0.999532 | | 6.406652 | 0.062073 | 0.961006 | | 4.750457 | 0.490143 | 0.605785 |
| OGG1 | 3.304117 | 0.525046 | 0.999532 | | 3.208756 | 0.087729 | 0.961006 | | 3.208588 | -0.41559 | 0.743059 |
| SMARCB | 5.435235 | -0.30925 | 0.999532 | | 6.611302 | -0.06539 | 0.961006 | | 4.944182 | 0.268961 | 0.809832 |
| SNRPG | 16.36907 | -0.28044 | 0.999532 | | 20.25927 | 0.039865 | 0.961006 | | 15.813 | 0.339479 | 0.542124 |
| FBXO28 | 2.961196 | -0.18262 | 0.999532 | | 3.731395 | 0.081818 | 0.961006 | | 2.825354 | 0.285031 | 0.844931 |
| PHKG2 | 2.685846 | 0.267054 | 0.999532 | | 2.923217 | 0.091291 | 0.961006 | | 2.682807 | -0.15579 | 0.929335 |
| SIPA1 | 5.180331 | -0.2646 | 0.999532 | | 6.364936 | 0.067366 | 0.961006 | | 5.162574 | 0.359167 | 0.724386 |
| TMEM134 | 4.892821 | 0.226659 | 0.999532 | | 5.11835 | 0.069521 | 0.961006 | | 4.949514 | -0.13286 | 0.920486 |
| CAMK2G | 5.014977 | 0.043207 | 0.999532 | | 5.276828 | -0.07204 | 0.961006 | | 4.415712 | -0.10508 | 0.946958 |
| TBCC | 4.908706 | 0.090086 | 0.999532 | | 5.240314 | 0.069769 | 0.961006 | | 4.663689 | 0.001602 | 0.999385 |
| TC2N | 2.133952 | 0.235621 | 0.999532 | | 2.178554 | -0.10285 | 0.961006 | | 1.958208 | -0.32073 | 0.861119 |
| SNX15 | 3.518581 | 0.223936 | 0.999532 | | 3.728843 | -0.0823 | 0.961006 | | 3.257645 | -0.27845 | 0.850724 |
| FOXO3 | 8.075927 | -0.11817 | 0.999532 | | 9.802505 | 0.052893 | 0.961006 | | 7.609962 | 0.1863 | 0.842754 |
| APOBEC | 2.27253 | -0.71495 | 0.999532 | | 3.102913 | 0.085278 | 0.961006 | | 2.279616 | 0.815865 | 0.520848 |
| PIGO | 3.356722 | -0.54741 | 0.999532 | | 4.767571 | 0.075854 | 0.961006 | | 3.501801 | 0.639899 | 0.555736 |
| ZNF679 | 2.77575 | 0.442928 | 0.999532 | | 2.663593 | 0.096802 | 0.961006 | | 2.748475 | -0.32837 | 0.837256 |
| MSC | 3.447936 | -0.12528 | 0.999532 | | 4.168218 | 0.087353 | 0.961006 | | 3.521015 | 0.214598 | 0.901033 |
| MED20 | 3.580222 | 0.329745 | 0.999532 | | 3.767058 | 0.083427 | 0.961006 | | 3.719179 | -0.22289 | 0.885892 |
| SLC27A3 | 5.782668 | -0.30962 | 0.999532 | | 7.135521 | 0.063715 | 0.961006 | | 5.378731 | 0.390516 | 0.703463 |
| LYPD6B | 2.340729 | 0.26177 | 0.999532 | | 2.363794 | -0.1032 | 0.961006 | | 2.170924 | -0.34495 | 0.836935 |
| SMNDC1 | 3.381683 | -0.12678 | 0.999532 | | 3.929753 | -0.08225 | 0.961006 | | 3.048353 | 0.064138 | 0.974421 |
| TNFRSF1 | 3.309394 | 0.321547 | 0.999532 | | 3.549699 | 0.086762 | 0.961006 | | 3.182928 | -0.19641 | 0.901033 |
| KCNG1 | 2.39742 | 0.810083 | 0.999532 | | 1.8516 | -0.11255 | 0.961006 | | 2.243244 | -0.91009 | 0.487694 |
| C11orf65 | 2.464662 | 0.525136 | 0.999532 | | 2.366247 | -0.10091 | 0.961006 | | 2.390735 | -0.60463 | 0.660958 |
| CPEB4 | 6.901072 | -0.16699 | 0.999532 | | 8.342927 | 0.057006 | 0.961006 | | 6.336298 | 0.244397 | 0.801572 |
| LARS2 | 2.872221 | 0.364103 | 0.999532 | | 2.5921 | -0.09411 | 0.961006 | | 2.688826 | -0.44526 | 0.753585 |
| MCRS1 | 3.22302 | 0.209689 | 0.999532 | | 3.30785 | -0.08769 | 0.961006 | | 3.04208 | -0.27733 | 0.859426 |
| RNF216 | 5.31263 | 0.028171 | 0.999532 | | 5.834502 | 0.066229 | 0.961006 | | 5.254259 | 0.069214 | 0.965649 |
| SLC38A1 | 2.667182 | 0.187339 | 0.999532 | | 2.722788 | -0.09585 | 0.961006 | | 2.489224 | -0.26382 | 0.876781 |
| IGDCC3 | 2.807405 | 0.572009 | 0.999532 | | 2.594803 | 0.098203 | 0.961006 | | 2.850359 | -0.4597 | 0.750371 |
| ZNF596 | 3.223898 | 0.19848 | 0.999532 | | 3.517554 | -0.0854 | 0.961006 | | 3.041584 | -0.25481 | 0.864923 |
| TERB2 | 2.309126 | -0.17197 | 0.999532 | | 2.682032 | -0.09567 | 0.961006 | | 2.301041 | 0.09344 | 0.969504 |
| SMYD5 | 3.044987 | -0.24088 | 0.999532 | | 3.805977 | 0.081404 | 0.961006 | | 2.960942 | 0.339682 | 0.809832 |
| CDCA8 | 2.185589 | 0.161535 | 0.999532 | | 2.211115 | 0.100815 | 0.961006 | | 2.216431 | -0.04311 | 0.989234 |
| NDUFB7 | 15.68031 | 0.160867 | 0.999532 | | 16.9748 | 0.046635 | 0.961006 | | 15.21902 | -0.09596 | 0.90424 |
| MRPS14 | 3.858619 | -0.11636 | 0.999532 | | 4.384392 | -0.07891 | 0.961006 | | 3.386032 | 0.068816 | 0.972758 |
| SNRK | 6.007686 | 0.020449 | 0.999532 | | 6.71438 | 0.063467 | 0.961006 | | 5.845044 | 0.061335 | 0.971732 |
| SACS | 1.937606 | -0.51278 | 0.999532 | | 2.444662 | 0.09402 | 0.961006 | | 1.825771 | 0.618853 | 0.686214 |
| SFXN4 | 2.953064 | -0.04291 | 0.999532 | | 3.460163 | 0.08641 | 0.961013 | | 2.815856 | 0.150258 | 0.93222 |
| RNF123 | 3.230992 | 0.562502 | 0.999532 | | 3.053062 | 0.08462 | 0.961013 | | 3.283046 | -0.45777 | 0.702463 |
| PMS2 | 2.791186 | 0.087711 | 0.999532 | | 3.179175 | 0.086111 | 0.961013 | | 2.63387 | 0.014507 | 0.996681 |
| ATP6V1C | 2.137146 | 0.474259 | 0.999532 | | 1.961136 | -0.10384 | 0.961043 | | 1.960588 | -0.55903 | 0.704316 |
| SIX1 | 1.980697 | 0.294998 | 0.999532 | | 2.192655 | 0.104611 | 0.961043 | | 2.073977 | -0.17161 | 0.934572 |
| G3BP2 | 2.741717 | 0.023599 | 0.999532 | | 3.302178 | 0.085026 | 0.961043 | | 2.750488 | 0.080176 | 0.970457 |
| ZNF131 | 2.401015 | -0.32502 | 0.999532 | | 2.694746 | -0.08934 | 0.961043 | | 2.164108 | 0.246249 | 0.892869 |
| REV3L | 4.511117 | -0.25465 | 0.999532 | | 5.465658 | -0.06951 | 0.961043 | | 4.210179 | 0.205131 | 0.877487 |
| CEP104 | 2.655318 | 0.092408 | 0.999532 | | 2.615482 | -0.09636 | 0.961043 | | 2.446276 | -0.1665 | 0.931488 |
| NCKIPSD | 4.719386 | 0.26576 | 0.999532 | | 4.860916 | -0.07585 | 0.961043 | | 4.400548 | -0.32081 | 0.776359 |
| GP9 | 2.383389 | 0.132649 | 0.999532 | | 2.523142 | -0.09856 | 0.961043 | | 2.328245 | -0.21344 | 0.909431 |
| RICTOR | 4.973608 | -0.01734 | 0.999532 | | 5.415664 | -0.06749 | 0.961043 | | 4.779689 | -0.02857 | 0.990309 |
| MVB12A | 4.241027 | 0.329871 | 0.999532 | | 4.33497 | 0.073407 | 0.961115 | | 4.048131 | -0.23441 | 0.854218 |
| KLHDC7A | 2.045247 | 0.832187 | 0.999532 | | 1.687679 | 0.114281 | 0.961115 | | 1.994621 | -0.70061 | 0.644433 |
| MYH15 | 1.826011 | 0.207508 | 0.999532 | | 1.920274 | -0.10872 | 0.961115 | | 1.703558 | -0.29123 | 0.884653 |
| C2CD5 | 2.888374 | -0.47631 | 0.999532 | | 3.728032 | 0.080006 | 0.961115 | | 2.698478 | 0.568976 | 0.66232 |
| MTHFSD | 2.689175 | 0.298774 | 0.999532 | | 2.671193 | -0.09236 | 0.961115 | | 2.600217 | -0.37251 | 0.803301 |
| NTAN1 | 2.60928 | -0.00627 | 0.999532 | | 3.017385 | 0.087226 | 0.961115 | | 2.44535 | 0.10476 | 0.960773 |
| ACTR6 | 3.305471 | -0.03645 | 0.999532 | | 3.515417 | -0.07831 | 0.961115 | | 3.154838 | -0.02283 | 0.993618 |
| DGKD | 3.643819 | 0.088328 | 0.999532 | | 4.12071 | 0.081554 | 0.961115 | | 3.445433 | 0.000873 | 0.999635 |
| ENO3 | 2.378866 | 0.124197 | 0.999532 | | 2.523141 | -0.0943 | 0.961115 | | 2.16757 | -0.19736 | 0.913432 |
| AACS | 1.994976 | 0.823003 | 0.999532 | | 1.86035 | 0.109549 | 0.961115 | | 2.115649 | -0.6959 | 0.626234 |
| ADCY10 | 2.553818 | 0.389061 | 0.999532 | | 2.361561 | -0.09772 | 0.961115 | | 2.254856 | -0.47359 | 0.749222 |
| PLCD3 | 5.989613 | 0.394344 | 0.999532 | | 6.229949 | 0.063937 | 0.961115 | | 5.937953 | -0.3108 | 0.737109 |
| NXT1 | 3.334147 | -0.50983 | 0.999532 | | 4.275576 | -0.07355 | 0.961115 | | 3.021054 | 0.452213 | 0.717627 |
| ARHGAP | 1.821364 | -0.20562 | 0.999532 | | 2.094585 | -0.10114 | 0.961115 | | 1.753454 | 0.123279 | 0.960427 |
| C22orf23 | 2.192255 | 0.008692 | 0.999532 | | 2.449959 | -0.10046 | 0.961115 | | 2.009694 | -0.08966 | 0.972221 |
| SHMT1 | 4.420845 | 0.019121 | 0.999532 | | 4.86607 | -0.08238 | 0.961115 | | 3.958679 | -0.07763 | 0.963866 |
| MRPS9 | 3.430992 | -0.07773 | 0.999532 | | 3.916263 | 0.078264 | 0.961115 | | 3.264361 | 0.174225 | 0.909084 |
| TGM2 | 13.88819 | -0.64435 | 0.999532 | | 18.42799 | -0.06115 | 0.961155 | | 12.84148 | 0.604131 | 0.414073 |
| DOCK9 | 4.615777 | -0.28767 | 0.999532 | | 5.15457 | -0.06851 | 0.961237 | | 4.300262 | 0.250125 | 0.852487 |
| HYPK | 2.602447 | -0.54843 | 0.999532 | | 3.514948 | -0.08586 | 0.961237 | | 2.601122 | 0.481645 | 0.742839 |
| RPL10 | 2.442435 | 0.415739 | 0.999532 | | 2.57553 | 0.097927 | 0.961237 | | 2.424454 | -0.29058 | 0.864245 |
| ENTPD1 | 2.44838 | -0.05812 | 0.999532 | | 2.56041 | -0.09169 | 0.961316 | | 2.252912 | -0.02831 | 0.993111 |
| CCDC47 | 13.44514 | -0.00892 | 0.999532 | | 14.35029 | -0.0479 | 0.961316 | | 12.08673 | -0.02123 | 0.989146 |
| ABCA3 | 2.730547 | -0.01456 | 0.999532 | | 3.216141 | 0.088489 | 0.961316 | | 2.586816 | 0.121226 | 0.949067 |
| PTPN13 | 3.485259 | -0.05024 | 0.999532 | | 4.151092 | 0.075067 | 0.961345 | | 3.365715 | 0.144051 | 0.924131 |
| CENPVL3 | 2.148476 | 0.292126 | 0.999532 | | 2.11276 | -0.10594 | 0.961345 | | 1.983643 | -0.37959 | 0.82512 |
| RIC8B | 3.889453 | -0.04219 | 0.999532 | | 4.230118 | -0.07405 | 0.961345 | | 3.599606 | -0.01617 | 0.996681 |
| ERCC4 | 2.487622 | 0.185652 | 0.999532 | | 2.551857 | 0.093798 | 0.961345 | | 2.354621 | -0.0737 | 0.974421 |
| RCHY1 | 4.711881 | 0.476598 | 0.999532 | | 4.38248 | 0.073776 | 0.961345 | | 4.61458 | -0.37336 | 0.722703 |
| NUCKS1 | 12.0564 | -0.01319 | 0.999532 | | 12.84426 | -0.04729 | 0.961355 | | 11.37298 | -0.02851 | 0.98556 |
| ZNF710 | 3.701998 | 0.16287 | 0.999532 | | 4.128065 | 0.08423 | 0.961452 | | 3.623727 | -0.06048 | 0.97502 |
| IGFALS | 1.814268 | -0.08271 | 0.999532 | | 1.956169 | -0.10613 | 0.961659 | | 1.806787 | -0.01702 | 0.996891 |
| CASP7 | 2.072255 | 0.484135 | 0.999532 | | 1.908421 | 0.107446 | 0.961659 | | 2.016985 | -0.35722 | 0.83987 |
| GAL3ST4 | 3.167642 | -0.23429 | 0.999532 | | 3.638836 | -0.08178 | 0.961659 | | 3.008957 | 0.169652 | 0.924186 |
| GNG2 | 2.382506 | -0.0471 | 0.999532 | | 2.473235 | 0.091958 | 0.961659 | | 2.236839 | 0.13367 | 0.950616 |
| SLC30A8 | 2.251942 | 0.308874 | 0.999532 | | 2.021081 | -0.10912 | 0.961659 | | 2.086334 | -0.40365 | 0.834627 |
| RTBDN | 2.150145 | 0.042248 | 0.999532 | | 2.411266 | -0.09615 | 0.961659 | | 2.056923 | -0.11648 | 0.958798 |
| OASL | 2.2949 | 0.873153 | 0.999532 | | 1.927865 | -0.10919 | 0.961659 | | 2.213452 | -0.96262 | 0.43983 |
| CACTIN | 3.004799 | 0.153032 | 0.999532 | | 3.281477 | 0.083546 | 0.961756 | | 3.025813 | -0.05141 | 0.982124 |
| SMO | 1.982626 | 0.068692 | 0.999532 | | 2.124579 | -0.10083 | 0.961756 | | 1.791521 | -0.15411 | 0.945129 |
| CRB3 | 2.173758 | 0.4453 | 0.999532 | | 2.107074 | 0.099384 | 0.961756 | | 2.064136 | -0.32812 | 0.845709 |
| MICAL3 | 4.62141 | -0.14608 | 0.999532 | | 5.48886 | 0.068959 | 0.961808 | | 4.40465 | 0.234201 | 0.857377 |
| RSPH3 | 3.074334 | 0.136872 | 0.999532 | | 3.05139 | -0.08387 | 0.961835 | | 2.801474 | -0.204 | 0.897958 |
| POLR2H | 5.344272 | -0.13188 | 0.999532 | | 6.540235 | 0.063399 | 0.962013 | | 5.133173 | 0.223611 | 0.862823 |
| ARHGAP | 2.471809 | 0.144245 | 0.999532 | | 2.927862 | 0.093237 | 0.96208 | | 2.488285 | -0.01576 | 0.996681 |
| RPE | 3.64766 | 0.03138 | 0.999532 | | 3.931102 | -0.07619 | 0.962117 | | 3.40289 | -0.09082 | 0.95999 |
| ZNF19 | 2.350864 | 0.103591 | 0.999532 | | 2.659558 | -0.09692 | 0.962446 | | 2.278364 | -0.1757 | 0.929604 |
| STAT5B | 7.045885 | 0.202809 | 0.999532 | | 7.227062 | -0.06152 | 0.962446 | | 6.738953 | -0.24798 | 0.800239 |
| KTI12 | 2.15933 | -0.23392 | 0.999532 | | 2.620344 | 0.092719 | 0.962446 | | 2.006187 | 0.343344 | 0.848617 |
| RASEF | 2.561715 | 0.455445 | 0.999532 | | 2.637155 | 0.091057 | 0.962446 | | 2.666243 | -0.34293 | 0.816775 |
| USB1 | 2.75228 | 0.090025 | 0.999532 | | 2.78437 | -0.08624 | 0.962446 | | 2.623599 | -0.16038 | 0.927544 |
| EBPL | 2.810331 | 0.08846 | 0.999532 | | 3.084295 | 0.08391 | 0.962446 | | 2.76095 | 0.014235 | 0.996681 |
| ST3GAL1 | 11.80156 | 0.18881 | 0.999532 | | 12.15632 | 0.045417 | 0.962446 | | 11.44136 | -0.12659 | 0.88476 |
| HM13 | 4.851686 | -0.01703 | 0.999532 | | 5.315552 | -0.06424 | 0.962446 | | 4.482142 | -0.02903 | 0.989093 |
| TMEM43 | 7.646435 | -0.22539 | 0.999532 | | 9.544876 | 0.053021 | 0.962446 | | 7.474214 | 0.296337 | 0.732916 |
| USP20 | 3.501259 | 0.499949 | 0.999532 | | 3.115109 | 0.083189 | 0.962446 | | 3.386066 | -0.40611 | 0.75311 |
| NEDD4 | 3.566843 | 0.097073 | 0.999532 | | 3.830422 | 0.076446 | 0.962446 | | 3.379715 | -0.00109 | 0.999588 |
| ZNF790 | 2.115494 | -0.06789 | 0.999532 | | 2.224821 | 0.096732 | 0.962446 | | 2.009936 | 0.17294 | 0.936909 |
| PIK3C3 | 4.000927 | -0.02089 | 0.999532 | | 4.496331 | 0.072739 | 0.962446 | | 3.728241 | 0.113035 | 0.944296 |
| CSNKA2I | 1.905211 | 0.405216 | 0.999532 | | 2.056861 | 0.102348 | 0.962446 | | 2.023556 | -0.27887 | 0.881837 |
| TOP2A | 2.010378 | 0.251402 | 0.999532 | | 1.971792 | -0.10142 | 0.962446 | | 1.88373 | -0.33765 | 0.855173 |
| APRT | 3.259181 | 0.107572 | 0.999532 | | 3.494778 | 0.078333 | 0.962446 | | 3.09255 | -0.01331 | 0.996681 |
| RBM34 | 3.540672 | -0.05641 | 0.999532 | | 4.155759 | -0.07203 | 0.962703 | | 3.339917 | 0.015409 | 0.996681 |
| CLCF1 | 2.624313 | 0.197852 | 0.999532 | | 2.89857 | 0.093871 | 0.962838 | | 2.539918 | -0.07852 | 0.972758 |
| SERPINB | 2.050358 | 0.287233 | 0.999532 | | 1.90448 | -0.10463 | 0.963075 | | 1.944741 | -0.3769 | 0.839277 |
| MACO1 | 3.773672 | -0.39158 | 0.999532 | | 4.852168 | 0.070306 | 0.963075 | | 3.589804 | 0.482152 | 0.66232 |
| RAB40B | 2.170268 | -0.02482 | 0.999532 | | 2.36623 | -0.09684 | 0.963075 | | 1.925519 | -0.05348 | 0.985675 |
| ATP6V0A | 3.62561 | -0.06917 | 0.999532 | | 3.92357 | -0.07657 | 0.963075 | | 3.393096 | 0.003801 | 0.999047 |
| PRR3 | 2.756115 | 0.220859 | 0.999532 | | 2.956739 | 0.089929 | 0.963273 | | 2.752632 | -0.10631 | 0.96129 |
| ORM1 | 2.229034 | 0.892446 | 0.999532 | | 1.831715 | -0.11016 | 0.96338 | | 2.163984 | -0.98078 | 0.445661 |
| GPR65 | 2.63821 | 0.423187 | 0.999532 | | 2.550107 | -0.09567 | 0.96338 | | 2.640707 | -0.50177 | 0.732486 |
| TBCB | 7.349518 | 0.165515 | 0.999532 | | 8.035459 | 0.057061 | 0.96338 | | 7.326755 | -0.09534 | 0.938429 |
| ARMH3 | 2.351477 | 0.017416 | 0.999532 | | 2.577607 | -0.08956 | 0.96338 | | 2.173289 | -0.09005 | 0.968434 |
| FITM1 | 3.135511 | -0.07736 | 0.999532 | | 3.806399 | 0.085834 | 0.963394 | | 3.139268 | 0.181158 | 0.915533 |
| SMPD3 | 2.698125 | 0.644589 | 0.999532 | | 2.236111 | -0.09943 | 0.963394 | | 2.665382 | -0.72986 | 0.612811 |
| OR56A4 | 2.606765 | -0.00892 | 0.999532 | | 3.046854 | 0.084846 | 0.963526 | | 2.619818 | 0.109916 | 0.955834 |
| KDM4C | 3.651111 | 0.126507 | 0.999532 | | 3.789487 | 0.074163 | 0.963536 | | 3.544577 | -0.03138 | 0.989638 |
| POLD1 | 3.495218 | 0.065145 | 0.999532 | | 3.60112 | -0.07977 | 0.96365 | | 3.195681 | -0.12409 | 0.943248 |
| EOLA1 | 3.540767 | -0.0539 | 0.999532 | | 3.913884 | -0.07349 | 0.963798 | | 3.209808 | -0.00089 | 0.999635 |
| DPT | 2.115077 | 0.009977 | 0.999532 | | 2.39633 | 0.093267 | 0.964043 | | 2.111357 | 0.098449 | 0.968235 |
| IQUB | 1.725014 | 0.252237 | 0.999532 | | 1.910936 | -0.10572 | 0.964043 | | 1.693552 | -0.32822 | 0.867739 |
| GPR162 | 2.323578 | -0.06015 | 0.999532 | | 2.516089 | -0.09181 | 0.964268 | | 2.234719 | -0.01787 | 0.996681 |
| SOX8 | 2.105339 | -0.5024 | 0.999532 | | 2.828781 | -0.09243 | 0.964268 | | 1.989228 | 0.434966 | 0.803301 |
| PRPF40B | 4.84602 | 0.294758 | 0.999532 | | 4.872562 | 0.070208 | 0.964375 | | 4.883634 | -0.2089 | 0.876258 |
| SPPL2B | 4.086973 | -0.4671 | 0.999532 | | 5.301185 | 0.067497 | 0.964375 | | 3.789874 | 0.555861 | 0.61656 |
| TIMM8A | 4.531598 | 0.240453 | 0.999532 | | 4.786009 | 0.069426 | 0.964375 | | 4.586254 | -0.15862 | 0.90424 |
| REST | 4.414051 | 0.082765 | 0.999532 | | 4.862536 | 0.066751 | 0.964375 | | 4.157604 | 0.000598 | 0.999746 |
| MTAP | 4.240681 | 0.026249 | 0.999532 | | 4.235061 | -0.07539 | 0.964375 | | 3.720664 | -0.08252 | 0.965649 |
| MYO7A | 3.487944 | 0.584611 | 0.999532 | | 3.448254 | 0.081219 | 0.964375 | | 3.481714 | -0.48364 | 0.668481 |
| ICE2 | 3.639039 | 0.19568 | 0.999532 | | 3.8446 | 0.073694 | 0.964375 | | 3.612378 | -0.10435 | 0.95001 |
| CCT3 | 5.238083 | 0.103369 | 0.999532 | | 5.638486 | 0.062499 | 0.964375 | | 5.177143 | -0.02509 | 0.990309 |
| PLEKHN1 | 2.046392 | 0.369704 | 0.999532 | | 1.923634 | -0.1023 | 0.964375 | | 1.901809 | -0.45628 | 0.789815 |
| MED16 | 3.012542 | -0.16798 | 0.999532 | | 3.499434 | 0.075131 | 0.964375 | | 2.895378 | 0.25628 | 0.864245 |
| USP28 | 3.234316 | 0.285102 | 0.999532 | | 3.407479 | -0.08249 | 0.964375 | | 3.179017 | -0.34907 | 0.797942 |
| RIF1 | 3.231091 | -0.16378 | 0.999532 | | 3.672156 | 0.075218 | 0.964407 | | 3.159751 | 0.254818 | 0.864245 |
| OAS1 | 4.019499 | -0.08858 | 0.999532 | | 4.482594 | -0.06891 | 0.964407 | | 3.564942 | 0.028747 | 0.991341 |
| TRIM66 | 2.04652 | 0.375275 | 0.999532 | | 1.741273 | -0.1066 | 0.964407 | | 1.805546 | -0.46966 | 0.791961 |
| ATF2 | 4.479492 | -0.20369 | 0.999532 | | 5.208461 | 0.065957 | 0.964407 | | 4.39615 | 0.278894 | 0.836453 |
| CBWD2 | 20.36064 | -0.09746 | 0.999532 | | 23.6042 | -0.03556 | 0.964407 | | 19.24544 | 0.087846 | 0.913842 |
| MKX | 2.028927 | -0.29372 | 0.999532 | | 2.290068 | -0.09537 | 0.964407 | | 1.971104 | 0.206696 | 0.923864 |
| CHKB | 12.85243 | -0.15215 | 0.999532 | | 15.90919 | 0.042887 | 0.964407 | | 12.89606 | 0.222236 | 0.765852 |
| ACACB | 3.1014 | 0.076248 | 0.999532 | | 3.15907 | -0.07815 | 0.964407 | | 2.837812 | -0.13938 | 0.936557 |
| BAIAP2L | 1.788796 | -0.00638 | 0.999532 | | 2.038104 | 0.10266 | 0.964474 | | 1.72443 | 0.127599 | 0.959765 |
| USP53 | 4.547056 | 0.14772 | 0.999532 | | 5.254093 | 0.065908 | 0.964474 | | 4.618137 | -0.05297 | 0.976192 |
| ATP10A | 7.746382 | 0.009779 | 0.999532 | | 8.348071 | -0.0642 | 0.964485 | | 7.146855 | -0.05561 | 0.972758 |
| EDA | 3.096893 | 0.081808 | 0.999532 | | 3.520133 | 0.077532 | 0.964485 | | 3.130896 | 0.009717 | 0.996891 |
| PINLYP | 2.655143 | 0.64068 | 0.999532 | | 2.303814 | -0.09822 | 0.964503 | | 2.583903 | -0.72694 | 0.572297 |
| SNAP47 | 2.524898 | 0.117576 | 0.999532 | | 2.607472 | -0.08682 | 0.964503 | | 2.312293 | -0.18648 | 0.915533 |
| FLG | 4.29806 | 0.917363 | 0.999532 | | 3.506665 | -0.08073 | 0.964503 | | 4.022766 | -0.97879 | 0.269743 |
| CCDC127 | 3.286955 | -0.01478 | 0.999532 | | 3.908143 | 0.07236 | 0.964503 | | 3.246241 | 0.102224 | 0.951149 |
| GC | 2.084521 | 0.128955 | 0.999532 | | 2.042232 | -0.10175 | 0.964503 | | 1.958298 | -0.21765 | 0.921811 |
| MTCP1 | 2.279243 | -0.2727 | 0.999532 | | 2.839842 | 0.085275 | 0.964503 | | 2.247383 | 0.376275 | 0.813172 |
| PLXNC1 | 2.315625 | 0.467969 | 0.999532 | | 2.269668 | 0.092384 | 0.964503 | | 2.368409 | -0.35842 | 0.818118 |
| NPLOC4 | 3.219718 | -0.25839 | 0.999532 | | 3.873636 | 0.073523 | 0.964503 | | 3.233059 | 0.337544 | 0.810753 |
| EPS15L1 | 5.719325 | -0.21573 | 0.999532 | | 6.963797 | -0.05714 | 0.964503 | | 5.369837 | 0.180774 | 0.876781 |
| PLOD3 | 3.14846 | -0.35557 | 0.999532 | | 4.283348 | 0.068882 | 0.964503 | | 3.293067 | 0.44507 | 0.706407 |
| SYT3 | 2.267461 | -0.07467 | 0.999532 | | 2.582126 | -0.09006 | 0.964503 | | 2.209676 | 0.004738 | 0.998898 |
| TLE3 | 3.831944 | -0.00829 | 0.999532 | | 4.543921 | 0.068909 | 0.964503 | | 3.780874 | 0.092316 | 0.954875 |
| EIF6 | 5.868435 | -0.10326 | 0.999532 | | 6.884015 | 0.057746 | 0.964503 | | 5.708695 | 0.179321 | 0.877562 |
| DDX39A | 3.935089 | -0.21888 | 0.999532 | | 4.6169 | 0.067899 | 0.964503 | | 3.796289 | 0.297784 | 0.818918 |
| CHCHD5 | 4.27776 | -0.00824 | 0.999532 | | 4.806808 | -0.06557 | 0.964503 | | 3.899415 | -0.0327 | 0.988853 |
| BMS1 | 3.853211 | 0.008906 | 0.999532 | | 3.902471 | -0.07169 | 0.964503 | | 3.539436 | -0.0626 | 0.974421 |
| TTLL3 | 7.5929 | 0.309961 | 0.999532 | | 7.304225 | 0.057994 | 0.964503 | | 7.207862 | -0.22794 | 0.810208 |

| | | | | | | | | | | | |
|---|---|---|---|---|---|---|---|---|---|---|---|
| PRB3 | 2.070434 | 0.140786 | 0.999532 | | 2.199675 | 0.093011 | 0.964503 | | 1.964689 | -0.03102 | 0.992286 |
| GABRG1 | 1.981871 | -0.0181 | 0.999532 | | 2.270626 | -0.09829 | 0.964503 | | 1.854585 | -0.05932 | 0.984155 |
| FNBP4 | 5.020142 | -0.27411 | 0.999532 | | 6.060427 | 0.059906 | 0.964523 | | 4.742681 | 0.344199 | 0.746876 |
| ENTPD7 | 2.33479 | 0.255091 | 0.999532 | | 2.244729 | -0.09501 | 0.964587 | | 2.185934 | -0.33877 | 0.839477 |
| TRIM3 | 2.516669 | -0.04923 | 0.999532 | | 2.909613 | 0.081242 | 0.964587 | | 2.436733 | 0.148124 | 0.93603 |
| H2AC8 | 2.720882 | 0.101236 | 0.999532 | | 2.8588 | -0.08255 | 0.964587 | | 2.502665 | -0.16915 | 0.923864 |
| NME3 | 4.342067 | -0.37642 | 0.999532 | | 5.691473 | 0.06053 | 0.964587 | | 4.355389 | 0.457137 | 0.654075 |
| PCYT2 | 5.317453 | 0.390002 | 0.999532 | | 5.247109 | 0.065362 | 0.964587 | | 5.175762 | -0.30808 | 0.773057 |
| TBXA2R | 6.182329 | -0.02453 | 0.999532 | | 7.294707 | 0.056508 | 0.96462 | | 6.140778 | 0.106261 | 0.934665 |
| ANKRD30 | 15.26846 | -0.17836 | 0.999532 | | 18.59755 | 0.039769 | 0.964641 | | 15.03702 | 0.246881 | 0.723509 |
| MPHOSPH | 3.601303 | -0.03072 | 0.999532 | | 3.789028 | -0.07178 | 0.964752 | | 3.252072 | -0.03279 | 0.989967 |
| CRMP1 | 2.137703 | -0.24816 | 0.999532 | | 2.46838 | -0.08877 | 0.964752 | | 2.018922 | 0.172054 | 0.93603 |
| RBM4 | 5.239221 | -0.21217 | 0.999532 | | 6.399167 | 0.061465 | 0.964752 | | 4.875174 | 0.296176 | 0.78258 |
| PEX26 | 4.560964 | 0.065412 | 0.999532 | | 4.802518 | -0.06537 | 0.964752 | | 4.149221 | -0.11413 | 0.939549 |
| CSRNP2 | 2.511098 | 0.02099 | 0.999532 | | 2.613446 | -0.08468 | 0.964752 | | 2.338211 | -0.08899 | 0.969501 |
| COA5 | 5.351049 | 0.271596 | 0.999532 | | 5.061556 | -0.06541 | 0.964752 | | 5.008047 | -0.31448 | 0.78258 |
| HOMEZ | 2.60217 | 0.129682 | 0.999532 | | 2.774765 | 0.08559 | 0.964752 | | 2.527557 | -0.02843 | 0.992286 |
| NUDT2 | 2.5461 | -0.2202 | 0.999532 | | 2.878744 | 0.082905 | 0.964752 | | 2.378285 | 0.305399 | 0.859426 |
| SPG11 | 3.171023 | -0.19217 | 0.999532 | | 3.892152 | 0.073448 | 0.964873 | | 3.13659 | 0.282267 | 0.853068 |
| ZDHHC3 | 4.796766 | 0.143144 | 0.999532 | | 4.931612 | -0.06541 | 0.964873 | | 4.391219 | -0.19102 | 0.881837 |
| TPCN1 | 11.53189 | 0.040923 | 0.999532 | | 13.07864 | 0.046049 | 0.964873 | | 11.27619 | 0.030837 | 0.983281 |
| ZKSCAN | 3.16182 | 0.400888 | 0.999532 | | 3.078239 | 0.084261 | 0.96489 | | 3.029726 | -0.28926 | 0.839811 |
| C12orf43 | 2.994914 | 0.409752 | 0.999532 | | 2.906596 | 0.085147 | 0.964892 | | 3.018454 | -0.31533 | 0.837256 |
| PDCD1 | 4.198969 | 0.254116 | 0.999532 | | 4.212324 | 0.075015 | 0.964988 | | 3.927944 | -0.15654 | 0.917499 |
| PDZD9 | 2.626438 | 0.616536 | 0.999532 | | 2.140478 | -0.09239 | 0.964991 | | 2.419554 | -0.70106 | 0.617899 |
| SDCBP2 | 3.104124 | 0.355189 | 0.999532 | | 3.092735 | 0.084946 | 0.965093 | | 2.875531 | -0.24359 | 0.876781 |
| ALX3 | 2.198815 | 0.255939 | 0.999532 | | 2.386685 | 0.097203 | 0.965094 | | 2.273799 | -0.14143 | 0.948127 |
| TMEM22 | 2.16333 | 0.32574 | 0.999532 | | 1.979911 | -0.09522 | 0.965094 | | 2.003913 | -0.40741 | 0.806394 |
| FLI1 | 4.741659 | -0.26833 | 0.999532 | | 6.022381 | -0.06557 | 0.965094 | | 4.69249 | 0.238372 | 0.865068 |
| KCNA10 | 1.912007 | -0.04067 | 0.999532 | | 2.18947 | -0.1003 | 0.965094 | | 1.91942 | -0.0409 | 0.991159 |
| MORC4 | 2.061482 | -0.40879 | 0.999532 | | 2.479866 | -0.08689 | 0.965094 | | 1.968884 | 0.334576 | 0.858767 |
| HSD3B7 | 2.788668 | 0.105337 | 0.999532 | | 3.173633 | 0.087354 | 0.965094 | | 2.75889 | 0.010528 | 0.996891 |
| FAM207A | 3.221673 | 0.157851 | 0.999532 | | 3.468506 | 0.081388 | 0.965094 | | 3.068513 | -0.05322 | 0.979996 |
| RALA | 4.54216 | -0.08949 | 0.999532 | | 4.992871 | -0.06447 | 0.965094 | | 4.245501 | 0.034083 | 0.987765 |
| ZNF208 | 14.11843 | 0.03919 | 0.999532 | | 15.95565 | 0.054758 | 0.965094 | | 13.46545 | 0.052298 | 0.971732 |
| ANGPTL | 2.061556 | 0.079988 | 0.999532 | | 2.286609 | 0.089945 | 0.965094 | | 2.070316 | 0.028054 | 0.993435 |
| TRRAP | 4.062538 | 0.266853 | 0.999532 | | 4.170513 | 0.077656 | 0.965094 | | 4.117867 | -0.18212 | 0.894617 |
| CAMK2N | 3.075901 | -0.13723 | 0.999532 | | 3.759312 | 0.084865 | 0.965094 | | 3.165592 | 0.221114 | 0.896886 |
| AGPAT2 | 5.075321 | 0.33464 | 0.999532 | | 5.102704 | 0.072991 | 0.965094 | | 4.899251 | -0.23748 | 0.858705 |
| DKK4 | 2.270772 | 0.125115 | 0.999532 | | 2.818668 | 0.086084 | 0.965094 | | 2.40145 | -0.01389 | 0.996891 |
| B3GAT3 | 6.61725 | -0.12021 | 0.999532 | | 7.608859 | -0.05388 | 0.965094 | | 6.346597 | 0.086937 | 0.948665 |
| EIF4EBP | 3.586184 | 0.601765 | 0.999532 | | 3.218592 | 0.077692 | 0.965094 | | 3.597306 | -0.49998 | 0.658332 |
| RBM43 | 3.291672 | 0.29533 | 0.999532 | | 3.344033 | 0.079388 | 0.965094 | | 3.245606 | -0.19934 | 0.900848 |
| STX17 | 3.706958 | -0.74281 | 0.999532 | | 5.110377 | 0.062782 | 0.965115 | | 3.747016 | 0.822675 | 0.407399 |
| PTP4A1 | 5.583037 | -0.17884 | 0.999532 | | 6.083872 | -0.0652 | 0.965115 | | 5.106314 | 0.124691 | 0.93603 |
| TEAD3 | 4.743277 | 0.258233 | 0.999532 | | 4.655335 | -0.06805 | 0.96559 | | 4.232436 | -0.31325 | 0.799944 |
| PCCB | 2.273015 | 0.242012 | 0.999532 | | 2.277729 | 0.089059 | 0.965646 | | 2.235705 | -0.13978 | 0.945456 |
| FBXO32 | 3.173593 | -0.11573 | 0.999532 | | 3.72059 | -0.07686 | 0.965646 | | 3.246911 | 0.055553 | 0.979996 |
| TLL1 | 2.470906 | 0.410653 | 0.999532 | | 2.548548 | 0.09775 | 0.965646 | | 2.421601 | -0.28953 | 0.870965 |
| PTH2 | 3.636344 | 0.175101 | 0.999532 | | 3.683008 | -0.07503 | 0.965646 | | 3.417735 | -0.23232 | 0.876781 |
| LIAS | 4.186448 | 0.750136 | 0.999532 | | 3.274861 | -0.08196 | 0.965646 | | 3.868896 | -0.82554 | 0.422479 |
| ABRACL | 5.173812 | -0.29949 | 0.999532 | | 6.331298 | 0.062057 | 0.965646 | | 5.035718 | 0.389996 | 0.717138 |
| SLC31A2 | 5.693632 | -0.02985 | 0.999532 | | 6.45856 | 0.067 | 0.965656 | | 5.28859 | 0.137087 | 0.912963 |
| LCAT | 3.015085 | -0.09854 | 0.999532 | | 3.04482 | -0.07657 | 0.965656 | | 2.787065 | 0.015836 | 0.996681 |
| HDGFL2 | 4.197423 | 0.708498 | 0.999532 | | 3.737742 | 0.071511 | 0.965704 | | 4.07539 | -0.61855 | 0.515762 |
| CBX3 | 8.378812 | -0.1134 | 0.999532 | | 10.54092 | 0.049733 | 0.965704 | | 8.3793 | 0.195668 | 0.845814 |
| TMEM26 | 2.547677 | 0.378306 | 0.999532 | | 2.41736 | -0.08988 | 0.965704 | | 2.428614 | -0.45392 | 0.755977 |
| FAM222B | 5.707488 | 0.135434 | 0.999532 | | 5.952922 | 0.060399 | 0.965704 | | 5.440796 | -0.05826 | 0.971732 |
| CASK | 2.046368 | 0.182925 | 0.999532 | | 2.118463 | 0.09491 | 0.965759 | | 1.91818 | -0.07302 | 0.977635 |
| SOBP | 3.367333 | 0.056084 | 0.999532 | | 3.745015 | -0.0721 | 0.965759 | | 3.162475 | -0.10552 | 0.951656 |
| MEAK7 | 4.17158 | 0.344059 | 0.999532 | | 4.222705 | -0.07039 | 0.965759 | | 4.068796 | -0.39093 | 0.735774 |
| TMEM91 | 3.054733 | -0.00758 | 0.999532 | | 3.185291 | 0.075473 | 0.965917 | | 2.937281 | 0.080748 | 0.971732 |
| LRRN4 | 2.20152 | -0.07106 | 0.999532 | | 2.539961 | -0.08733 | 0.965917 | | 2.147718 | -0.0021 | 0.999429 |
| ARPC5L | 4.931346 | 0.163179 | 0.999532 | | 5.258113 | 0.062156 | 0.966016 | | 4.774212 | -0.08771 | 0.956011 |
| DMC1 | 3.112543 | 0.162128 | 0.999532 | | 3.34318 | 0.078479 | 0.966016 | | 3.239041 | -0.05672 | 0.978931 |
| DUXA | 3.040693 | 0.257588 | 0.999532 | | 3.245116 | -0.07998 | 0.966016 | | 2.90308 | -0.31481 | 0.831349 |
| DEFB123 | 2.244594 | 0.421138 | 0.999532 | | 2.095168 | -0.09687 | 0.966016 | | 2.101009 | -0.50228 | 0.75311 |
| CORO1A | 4.627961 | 0.013569 | 0.999532 | | 5.184933 | -0.07345 | 0.966016 | | 4.536594 | -0.08639 | 0.96169 |
| PYY | 3.724348 | 0.237997 | 0.999532 | | 3.957812 | -0.07079 | 0.966016 | | 3.491912 | -0.28419 | 0.827267 |
| UPF1 | 4.04505 | -0.38244 | 0.999532 | | 4.855589 | -0.06252 | 0.966081 | | 3.622648 | 0.328038 | 0.794815 |
| CRHR1 | 3.051434 | 0.244887 | 0.999532 | | 2.891995 | 0.083211 | 0.966106 | | 2.800042 | -0.15157 | 0.936911 |
| CNTNAP | 2.108257 | 0.819801 | 0.999532 | | 1.789649 | 0.101714 | 0.966106 | | 2.160197 | -0.69602 | 0.644433 |
| SLC2A6 | 2.287122 | 0.091519 | 0.999532 | | 2.663415 | 0.086689 | 0.966185 | | 2.496049 | 0.016276 | 0.996681 |
| IL11RA | 3.936957 | 0.125691 | 0.999532 | | 3.998802 | -0.07298 | 0.966272 | | 3.703917 | -0.17571 | 0.915533 |
| SALL1 | 3.617707 | 0.177306 | 0.999532 | | 3.721148 | 0.070502 | 0.966316 | | 3.422141 | -0.08983 | 0.959377 |
| TRIM2 | 2.431674 | 0.22881 | 0.999532 | | 2.666295 | 0.083343 | 0.966453 | | 2.414847 | -0.12437 | 0.948099 |
| INIP | 4.225917 | 0.085436 | 0.999532 | | 4.391429 | 0.06508 | 0.966636 | | 4.075403 | -0.00997 | 0.996891 |
| SIRPD | 2.025074 | 0.398424 | 0.999532 | | 1.935416 | -0.09687 | 0.966636 | | 1.906748 | -0.47562 | 0.766974 |
| GGT5 | 4.378172 | -0.63903 | 0.999532 | | 5.802368 | -0.06397 | 0.966655 | | 3.912771 | 0.597033 | 0.55738 |
| IL27RA | 3.260295 | -0.16522 | 0.999532 | | 3.987484 | 0.071102 | 0.967097 | | 3.327075 | 0.253554 | 0.864245 |
| RFWD3 | 2.282716 | -0.11948 | 0.999532 | | 2.55074 | -0.0876 | 0.967097 | | 2.157821 | 0.046132 | 0.98861 |
| C14orf17 | 2.117959 | 0.321719 | 0.999532 | | 2.179018 | 0.092363 | 0.967158 | | 2.121875 | -0.21438 | 0.910105 |
| HJV | 2.058256 | 0.378645 | 0.999532 | | 1.900727 | -0.09748 | 0.967166 | | 1.988295 | -0.45903 | 0.790999 |
| H2AZ1 | 10.00723 | -0.13079 | 0.999532 | | 11.60304 | -0.04351 | 0.967267 | | 9.40348 | 0.107843 | 0.913842 |
| SPR | 5.061197 | -0.31661 | 0.999532 | | 6.474265 | 0.057618 | 0.967518 | | 5.070146 | 0.397346 | 0.702501 |
| SUPV3L1 | 2.516449 | 0.114802 | 0.999532 | | 2.53021 | -0.08544 | 0.967551 | | 2.235865 | -0.19282 | 0.920716 |
| DYNC2I1 | 3.150871 | -0.07166 | 0.999532 | | 3.284439 | -0.07584 | 0.967551 | | 2.784894 | 0.007003 | 0.997967 |
| TAF1B | 3.794539 | 0.318294 | 0.999532 | | 3.752683 | 0.071387 | 0.967551 | | 3.655464 | -0.22679 | 0.867976 |
| SELENO | 2.60097 | 0.526743 | 0.999532 | | 2.457424 | -0.09111 | 0.967551 | | 2.640951 | -0.61076 | 0.662584 |
| PARP14 | 4.061736 | 0.024376 | 0.999532 | | 4.483187 | -0.07021 | 0.967551 | | 3.999735 | -0.07968 | 0.965649 |
| NDUFAF | 5.72955 | -0.01075 | 0.999532 | | 6.817975 | 0.059072 | 0.967551 | | 5.669475 | 0.093099 | 0.948127 |
| CLDN10 | 3.567557 | -0.14225 | 0.999532 | | 4.349548 | -0.08667 | 0.967575 | | 3.281476 | 0.090569 | 0.967145 |
| PSMB6 | 4.687401 | 0.182951 | 0.999532 | | 5.014252 | 0.063882 | 0.967575 | | 4.609646 | -0.10074 | 0.944993 |
| RRAS2 | 4.615928 | -0.37829 | 0.999532 | | 5.809062 | -0.06533 | 0.967578 | | 4.455137 | 0.333381 | 0.797147 |
| BMP3 | 2.213876 | 0.379368 | 0.999532 | | 2.137624 | 0.095174 | 0.967578 | | 2.221921 | -0.27279 | 0.888003 |
| ACHE | 3.004951 | -0.17246 | 0.999532 | | 3.540066 | -0.0783 | 0.967578 | | 2.756377 | 0.113776 | 0.954304 |
| MEX3C | 3.732391 | 0.133357 | 0.999532 | | 4.124922 | 0.067662 | 0.967746 | | 3.828973 | -0.0502 | 0.978931 |
| FIZ1 | 4.7601 | 0.002107 | 0.999532 | | 5.406533 | 0.061387 | 0.968251 | | 4.444516 | 0.077146 | 0.96129 |
| EIF1AD | 2.963469 | -0.47437 | 0.999532 | | 3.751687 | -0.07097 | 0.968445 | | 2.708573 | 0.420522 | 0.753748 |
| FAXDC2 | 4.080797 | 0.221328 | 0.999532 | | 4.563557 | 0.067252 | 0.968445 | | 4.006964 | -0.13305 | 0.92697 |
| CHRNA1 | 2.342332 | 0.085745 | 0.999532 | | 2.421937 | -0.08636 | 0.968588 | | 2.109128 | -0.14879 | 0.940744 |
| IMPG2 | 2.340361 | 0.586578 | 0.999532 | | 2.153415 | -0.09507 | 0.968588 | | 2.20277 | -0.65712 | 0.657538 |
| KDM5A | 3.878658 | -0.17157 | 0.999532 | | 4.608365 | 0.066284 | 0.968588 | | 3.813763 | 0.25515 | 0.848065 |
| EXOSC9 | 3.047982 | 0.646552 | 0.999532 | | 2.760302 | 0.078961 | 0.968627 | | 3.082652 | -0.55027 | 0.651111 |
| FBXL6 | 2.558469 | 0.216618 | 0.999532 | | 2.515957 | -0.08254 | 0.969003 | | 2.251955 | -0.28343 | 0.864268 |
| MEGF6 | 2.233777 | 0.134464 | 0.999532 | | 2.396655 | 0.084796 | 0.969008 | | 2.216219 | -0.02595 | 0.994192 |
| KIF24 | 2.311544 | -0.05585 | 0.999532 | | 2.409809 | -0.08924 | 0.9691 | | 2.099305 | -0.0209 | 0.996681 |
| FAM120A | 5.616392 | 0.11245 | 0.999532 | | 5.955959 | -0.06077 | 0.9691 | | 5.390461 | -0.15811 | 0.896886 |
| PIK3IP1 | 3.336654 | -0.15028 | 0.999532 | | 4.068292 | 0.074302 | 0.969165 | | 3.411971 | 0.238149 | 0.87813 |
| SLC12A7 | 3.619428 | -0.15935 | 0.999532 | | 3.811417 | -0.06738 | 0.969165 | | 3.334295 | 0.100764 | 0.955834 |
| H2AC19 | 14.0579 | -0.41844 | 0.999532 | | 17.56009 | -0.04317 | 0.969165 | | 12.95655 | 0.400206 | 0.536289 |
| FSTL4 | 2.458263 | 0.096203 | 0.999532 | | 2.731283 | 0.081231 | 0.969165 | | 2.373729 | 0.001843 | 0.999429 |
| PLEKHH | 5.845778 | -0.01269 | 0.999532 | | 6.684746 | 0.054778 | 0.969165 | | 5.512451 | 0.082865 | 0.954538 |
| ELP4 | 2.220605 | 0.176058 | 0.999532 | | 2.185847 | -0.08693 | 0.969165 | | 2.103227 | -0.24565 | 0.892869 |
| EML2 | 4.139336 | -0.23145 | 0.999532 | | 5.141944 | 0.058972 | 0.969165 | | 4.099731 | 0.307881 | 0.787104 |
| TRNAU1A | 2.341724 | -0.2656 | 0.999532 | | 3.012972 | 0.078048 | 0.969165 | | 2.444867 | 0.362833 | 0.820834 |
| SUV39H1 | 2.194851 | 0.637028 | 0.999532 | | 1.981252 | -0.09596 | 0.969165 | | 2.067229 | -0.71588 | 0.619775 |
| MMRN2 | 4.322701 | -0.62433 | 0.999532 | | 5.734902 | -0.0635 | 0.969165 | | 3.965909 | 0.576374 | 0.626811 |
| HPGDS | 3.754771 | 0.049821 | 0.999532 | | 4.016744 | -0.0796 | 0.969165 | | 3.659538 | -0.1375 | 0.943134 |
| FTO | 4.092026 | -0.27177 | 0.999532 | | 4.945137 | -0.06271 | 0.969165 | | 3.749852 | 0.226175 | 0.863134 |
| BOD1 | 4.47669 | -0.16583 | 0.999532 | | 5.294556 | 0.057959 | 0.969165 | | 4.189159 | 0.238076 | 0.846155 |
| ORC2 | 2.772242 | 0.235237 | 0.999532 | | 2.621132 | -0.08208 | 0.969165 | | 2.51863 | -0.29644 | 0.858647 |
| GMDS | 3.377353 | 0.175503 | 0.999532 | | 3.812039 | 0.06907 | 0.969165 | | 3.221821 | -0.09816 | 0.955834 |
| TIMM8B | 5.29928 | 0.169118 | 0.999532 | | 5.686172 | 0.062767 | 0.969214 | | 5.17251 | -0.08455 | 0.954538 |
| ASTE1 | 2.175605 | 0.260963 | 0.999532 | | 2.285941 | 0.084707 | 0.969214 | | 2.143942 | -0.15772 | 0.938429 |
| EXOC8 | 3.517726 | -0.25132 | 0.999532 | | 4.377831 | -0.06311 | 0.969214 | | 3.327692 | 0.211998 | 0.881837 |
| LRRFIP1 | 7.758717 | -0.07056 | 0.999532 | | 8.999215 | -0.04575 | 0.969214 | | 7.423918 | 0.045548 | 0.973754 |
| FAM189A | 4.890886 | 0.421587 | 0.999532 | | 4.705672 | 0.063343 | 0.969214 | | 4.830464 | -0.34441 | 0.743576 |
| LRRFIP2 | 4.111265 | 0.314672 | 0.999532 | | 4.083021 | 0.069089 | 0.969214 | | 3.903344 | -0.22681 | 0.872493 |
| ALDH1A1 | 7.393578 | -0.24334 | 0.999532 | | 8.835928 | 0.054007 | 0.969214 | | 7.017047 | 0.319056 | 0.728126 |
| NAIP | 4.88897 | 0.150933 | 0.999532 | | 4.740688 | -0.06362 | 0.969214 | | 4.467096 | -0.20612 | 0.881837 |
| FIP1L1 | 4.825205 | 0.243526 | 0.999532 | | 5.205841 | 0.061637 | 0.969214 | | 4.674813 | -0.15393 | 0.905952 |
| SPRED3 | 1.930988 | -0.25101 | 0.999532 | | 2.282205 | -0.08851 | 0.969214 | | 1.964305 | 0.179081 | 0.938429 |
| SLC43A3 | 2.904809 | 0.266167 | 0.999532 | | 2.818871 | -0.07949 | 0.969214 | | 2.705172 | -0.32172 | 0.821951 |
| ZNF835 | 2.267431 | 0.906383 | 0.999532 | | 1.825631 | -0.09694 | 0.969214 | | 2.204522 | -0.9837 | 0.437893 |
| ZNF513 | 2.939588 | 0.273386 | 0.999532 | | 2.86506 | -0.0769 | 0.969214 | | 2.695632 | -0.33024 | 0.821113 |
| METTL15 | 3.609718 | 0.508015 | 0.999532 | | 3.211915 | -0.07689 | 0.969214 | | 3.293664 | -0.56617 | 0.639848 |
| ADSS1 | 2.242218 | 0.324976 | 0.999532 | | 2.317725 | 0.084618 | 0.969214 | | 2.137385 | -0.22321 | 0.901033 |
| IMMT | 4.841438 | -0.12844 | 0.999532 | | 5.983852 | 0.059221 | 0.96933 | | 4.598014 | 0.212033 | 0.865065 |
| SLC19A1 | 2.942528 | 0.093531 | 0.999532 | | 3.232635 | -0.07532 | 0.96933 | | 2.79226 | -0.14102 | 0.940162 |
| RAB3D | 5.814154 | 0.361302 | 0.999532 | | 5.560048 | 0.060508 | 0.96933 | | 5.574357 | -0.28164 | 0.794731 |
| LRG1 | 2.470169 | 0.061035 | 0.999532 | | 2.56541 | -0.08714 | 0.96933 | | 2.209627 | -0.12742 | 0.953949 |
| SLF2 | 3.595372 | 0.934659 | 0.955244 | | 2.896717 | 0.07544 | 0.96933 | | 3.60993 | -0.84046 | 0.359441 |
| BEAN1 | 1.844624 | 0.21558 | 0.999532 | | 1.973147 | -0.09508 | 0.96933 | | 1.776413 | -0.28971 | 0.883679 |
| CFHR3 | 2.551565 | -0.20318 | 0.999532 | | 3.009667 | -0.07618 | 0.96933 | | 2.41446 | 0.145694 | 0.939913 |
| PRPSAP | 4.913375 | -0.18292 | 0.999532 | | 6.07362 | 0.05554 | 0.96933 | | 4.952513 | 0.258749 | 0.823571 |
| ODAD2 | 2.198709 | 0.319521 | 0.999532 | | 2.243293 | -0.08619 | 0.96933 | | 1.996264 | -0.38251 | 0.811123 |
| GUCA1A | 2.832907 | 0.096333 | 0.999532 | | 3.196114 | -0.08549 | 0.96933 | | 2.822199 | -0.16107 | 0.938429 |
| CD70 | 2.635623 | 0.484563 | 0.999532 | | 2.48812 | -0.09791 | 0.96933 | | 2.460672 | -0.56037 | 0.717627 |
| ARMCX1 | 3.669507 | -0.14313 | 0.999532 | | 4.172211 | -0.06542 | 0.96933 | | 3.500429 | 0.106788 | 0.94861 |
| LACTB | 3.20982 | -0.16414 | 0.999532 | | 3.609996 | -0.06858 | 0.96933 | | 3.076581 | 0.114196 | 0.948127 |
| FOXF1 | 2.687421 | 0.045694 | 0.999532 | | 2.980146 | -0.07854 | 0.96933 | | 2.635931 | -0.10194 | 0.962014 |
| LTB | 2.678552 | -0.09308 | 0.999532 | | 3.287813 | 0.0757 | 0.96933 | | 2.85604 | 0.183702 | 0.918694 |
| LHX2 | 3.214655 | 0.097806 | 0.999532 | | 3.576003 | -0.07388 | 0.96933 | | 3.127969 | -0.15114 | 0.92883 |
| KLF12 | 3.643668 | 0.055822 | 0.999532 | | 4.053971 | -0.06692 | 0.96933 | | 3.52299 | -0.10428 | 0.950616 |
| ELOA | 5.188014 | 0.112288 | 0.999532 | | 5.107502 | -0.06126 | 0.96933 | | 4.578494 | -0.17069 | 0.900848 |
| DOK2 | 2.343694 | -0.26486 | 0.999532 | | 2.725348 | -0.07739 | 0.96933 | | 2.101672 | 0.203827 | 0.909259 |
| EXOC3L | 3.691584 | -0.29387 | 0.999532 | | 4.623086 | -0.06519 | 0.96933 | | 3.595847 | 0.247398 | 0.861119 |
| BSND | 1.946181 | -0.1208 | 0.999532 | | 2.177902 | -0.08959 | 0.96933 | | 1.815629 | 0.049262 | 0.988714 |
| GNL3 | 3.580796 | 0.03086 | 0.999532 | | 3.975853 | -0.07012 | 0.96933 | | 3.340065 | -0.0915 | 0.958191 |
| NXN | 3.077464 | -0.11717 | 0.999532 | | 3.792291 | 0.068942 | 0.96933 | | 3.129476 | 0.20802 | 0.892869 |
| CALY | 2.473678 | 0.494693 | 0.999532 | | 2.326439 | 0.086924 | 0.96933 | | 2.444053 | -0.3878 | 0.805466 |
| ZNF205 | 2.033873 | 0.250152 | 0.999532 | | 2.073849 | -0.08746 | 0.969449 | | 1.947096 | -0.31911 | 0.861119 |
| PRSS50 | 3.02325 | 0.493655 | 0.999532 | | 2.873221 | -0.08652 | 0.96948 | | 2.837695 | -0.55889 | 0.673405 |
| GPAT3 | 2.303586 | -0.55381 | 0.999532 | | 2.881185 | -0.08121 | 0.96948 | | 1.992368 | 0.492137 | 0.755977 |
| ZNF253 | 3.105483 | 0.27858 | 0.999532 | | 3.210692 | 0.072949 | 0.969544 | | 2.956831 | -0.18829 | 0.903339 |
| ABHD3 | 3.742869 | 0.268159 | 0.999532 | | 3.924624 | 0.06533 | 0.969544 | | 3.687216 | -0.18506 | 0.892869 |
| SDHAF4 | 3.066373 | 0.439051 | 0.999532 | | 2.788436 | -0.07551 | 0.969839 | | 2.835147 | -0.49875 | 0.699971 |
| PRRG1 | 2.143655 | 0.078905 | 0.999532 | | 2.19142 | -0.08518 | 0.969839 | | 1.976794 | -0.14981 | 0.945129 |
| CX3CR1 | 3.657123 | 0.001473 | 0.999532 | | 4.096352 | 0.066665 | 0.969839 | | 3.492838 | 0.084377 | 0.961873 |
| RAP2C | 3.542567 | 0.075402 | 0.999532 | | 4.001576 | -0.07274 | 0.969839 | | 3.343214 | -0.14572 | 0.926957 |
| GMCL1 | 2.705871 | 0.258599 | 0.999532 | | 2.62042 | -0.0778 | 0.969839 | | 2.500705 | -0.3218 | 0.835554 |
| CCDC12 | 3.565925 | 0.347251 | 0.999532 | | 3.336149 | -0.06983 | 0.969839 | | 3.400232 | -0.402 | 0.737109 |
| DNASE1 | 2.415695 | 0.66006 | 0.999532 | | 2.308542 | -0.08655 | 0.969839 | | 2.390673 | -0.70122 | 0.616301 |
| CAV3 | 2.606866 | 0.505688 | 0.999532 | | 2.40118 | -0.0871 | 0.969839 | | 2.549424 | -0.57653 | 0.689943 |
| PDZRN3 | 2.192765 | 0.1909 | 0.999532 | | 2.310597 | -0.08449 | 0.969839 | | 2.157746 | -0.25421 | 0.88885 |
| AMIGO2 | 4.945186 | -0.401 | 0.999532 | | 6.027767 | -0.05822 | 0.969839 | | 4.431268 | 0.36588 | 0.737109 |
| ARHGEF | 6.770691 | -0.37974 | 0.999532 | | 8.374119 | -0.05224 | 0.969839 | | 6.320923 | 0.350428 | 0.706407 |
| RO60 | 3.997603 | -0.09458 | 0.999532 | | 4.551542 | -0.06256 | 0.970065 | | 3.613726 | 0.05048 | 0.978927 |
| HDAC8 | 3.170034 | 0.174024 | 0.999532 | | 3.307725 | -0.07033 | 0.970065 | | 2.968091 | -0.22597 | 0.881316 |
| ZNF283 | 2.530699 | -0.08767 | 0.999532 | | 2.867505 | -0.07776 | 0.970112 | | 2.263662 | 0.028579 | 0.992286 |
| CREB3L1 | 2.118501 | 0.081368 | 0.999532 | | 2.472088 | 0.08372 | 0.97015 | | 2.090142 | 0.023021 | 0.995256 |
| AGFG1 | 4.722488 | -0.22115 | 0.999532 | | 5.821301 | -0.05836 | 0.970411 | | 4.488706 | 0.189846 | 0.88885 |
| MIOS | 3.12728 | -0.21717 | 0.999532 | | 3.565584 | 0.069167 | 0.970459 | | 2.981897 | 0.294856 | 0.849608 |
| PSD4 | 2.670194 | 0.004488 | 0.999532 | | 2.778359 | -0.07577 | 0.970459 | | 2.557628 | -0.05918 | 0.978931 |
| BOLA1 | 3.203832 | 0.127022 | 0.999532 | | 3.228946 | -0.0729 | 0.970459 | | 2.942363 | -0.1875 | 0.909255 |
| FGD2 | 3.762959 | 0.296018 | 0.999532 | | 3.832667 | 0.068067 | 0.970459 | | 3.751867 | -0.21665 | 0.877268 |
| TASP1 | 2.325027 | -0.36796 | 0.999532 | | 3.081901 | 0.077101 | 0.97062 | | 2.396284 | 0.467019 | 0.765852 |
| ITPRIPL1 | 2.435666 | 0.049247 | 0.999532 | | 2.597897 | -0.0825 | 0.97062 | | 2.212635 | -0.1121 | 0.959593 |
| ANKFY1 | 2.869908 | 0.015155 | 0.999532 | | 3.299188 | 0.071185 | 0.97062 | | 2.905264 | 0.068266 | 0.974421 |
| INO80 | 2.431361 | -0.01319 | 0.999532 | | 2.886568 | 0.075458 | 0.97062 | | 2.352859 | 0.106078 | 0.958798 |
| ASNS | 3.5876 | -0.05199 | 0.999532 | | 4.137788 | -0.06403 | 0.97062 | | 3.358831 | 0.009822 | 0.996891 |
| NETO2 | 2.440628 | 0.139323 | 0.999532 | | 2.412001 | -0.0803 | 0.97062 | | 2.333215 | -0.20328 | 0.910105 |
| NACC1 | 3.2876 | -0.19684 | 0.999532 | | 4.301846 | 0.067458 | 0.97062 | | 3.326501 | 0.295499 | 0.839299 |
| SOX15 | 1.961571 | 0.368145 | 0.999532 | | 1.962383 | 0.093041 | 0.97062 | | 1.988257 | -0.25979 | 0.897302 |
| ZNF337 | 1.979032 | -0.28202 | 0.999532 | | 2.379276 | 0.082989 | 0.97062 | | 1.82078 | 0.385393 | 0.825181 |
| OR10G8 | 1.832166 | 0.463869 | 0.999532 | | 1.71957 | 0.094273 | 0.97062 | | 1.895307 | -0.35103 | 0.85585 |
| ANO3 | 1.855079 | -0.24217 | 0.999532 | | 2.136987 | -0.08615 | 0.97062 | | 1.734376 | 0.173954 | 0.939107 |
| KRT75 | 3.327239 | 0.066344 | 0.999532 | | 3.621106 | -0.07005 | 0.97062 | | 3.180764 | -0.11763 | 0.948426 |
| GOLPH3L | 3.484413 | -0.13445 | 0.999532 | | 3.838836 | -0.06927 | 0.97062 | | 3.333287 | 0.07351 | 0.972305 |
| DMRTC1 | 2.232572 | 0.143893 | 0.999532 | | 2.272254 | -0.0854 | 0.970658 | | 2.037496 | -0.21003 | 0.914692 |
| AKR7A2 | 2.458139 | -0.23601 | 0.999532 | | 2.923552 | -0.07754 | 0.970658 | | 2.27702 | 0.176142 | 0.929529 |
| PIGY | 5.296581 | 0.246789 | 0.999532 | | 5.664013 | 0.054985 | 0.970658 | | 5.288468 | -0.17409 | 0.881689 |
| ARL13B | 4.291192 | 0.058538 | 0.999532 | | 4.646225 | -0.06514 | 0.970658 | | 4.03095 | -0.10824 | 0.946512 |
| CENPX | 4.37367 | 0.074669 | 0.999532 | | 4.931581 | 0.059357 | 0.970658 | | 4.305022 | 0.000877 | 0.999635 |
| TRAF5 | 2.448084 | 0.401203 | 0.999532 | | 2.447884 | -0.08322 | 0.970658 | | 2.38912 | -0.46729 | 0.750922 |
| TAF12 | 4.041851 | 0.106289 | 0.999532 | | 4.660217 | 0.064532 | 0.970658 | | 4.004511 | -0.02353 | 0.992286 |
| CD80 | 3.263871 | 0.331915 | 0.999532 | | 3.298134 | -0.07644 | 0.970658 | | 3.210448 | -0.40446 | 0.764149 |
| ANP32A | 5.584252 | -0.17515 | 0.999532 | | 6.226137 | -0.05552 | 0.970658 | | 5.19919 | 0.132913 | 0.920259 |
| ATXN7L1 | 2.723324 | 0.304857 | 0.999532 | | 2.996523 | 0.074634 | 0.970658 | | 2.681026 | -0.19873 | 0.905952 |
| CFAP74 | 1.855022 | 0.067729 | 0.999532 | | 2.090317 | 0.088506 | 0.970658 | | 1.902596 | 0.039646 | 0.991159 |
| C3orf70 | 2.939979 | 0.065896 | 0.999532 | | 3.13646 | -0.07236 | 0.970658 | | 2.877783 | -0.1165 | 0.952656 |
| ANKRD1 | 6.480975 | -0.04781 | 0.999532 | | 7.288418 | 0.051144 | 0.970658 | | 6.386091 | 0.130278 | 0.909087 |
| TMX1 | 2.427232 | 0.163697 | 0.999532 | | 2.534297 | -0.07902 | 0.970658 | | 2.191252 | -0.22279 | 0.897096 |
| ID2 | 10.27887 | -0.40081 | 0.999532 | | 12.36277 | -0.04455 | 0.970658 | | 9.358399 | 0.35925 | 0.672452 |
| SV2C | 2.079259 | 0.183665 | 0.999532 | | 2.320873 | 0.084547 | 0.970658 | | 2.095251 | -0.0782 | 0.975284 |
| CCDC107 | 17.33264 | -0.01675 | 0.999532 | | 19.64637 | 0.038589 | 0.970658 | | 16.88773 | 0.074642 | 0.941197 |
| COMMD | 3.714052 | -0.13276 | 0.999532 | | 3.850088 | -0.06395 | 0.970658 | | 3.36862 | 0.049249 | 0.983749 |
| CLEC4M | 2.584313 | 0.469404 | 0.999532 | | 2.537561 | 0.078248 | 0.970658 | | 2.601794 | -0.37371 | 0.803301 |
| TRAPPC | 4.891771 | 0.209169 | 0.999532 | | 5.122476 | 0.055321 | 0.970658 | | 4.536249 | -0.13433 | 0.917861 |
| PLEKHJ1 | 4.330943 | 0.235045 | 0.999532 | | 4.304577 | -0.06105 | 0.970658 | | 3.889933 | -0.27317 | 0.825181 |
| FGB | 2.7959 | -0.28781 | 0.999532 | | 3.356293 | -0.0766 | 0.970658 | | 2.557098 | 0.234041 | 0.898369 |
| C12orf76 | 2.437879 | -0.00771 | 0.999532 | | 2.8114 | -0.07655 | 0.970658 | | 2.502692 | -0.04322 | 0.988853 |
| MAP3K1 | 3.728035 | 0.008525 | 0.999532 | | 4.251148 | 0.061192 | 0.970658 | | 3.728227 | 0.073025 | 0.969501 |
| IMP3 | 4.238072 | -0.21871 | 0.999532 | | 4.941951 | -0.06467 | 0.970658 | | 3.715728 | 0.180718 | 0.897096 |
| ZNF844 | 2.699909 | 0.179983 | 0.999532 | | 2.656987 | -0.07687 | 0.970658 | | 2.442253 | -0.24118 | 0.888003 |
| LRRC8B | 2.173256 | -0.12275 | 0.999532 | | 2.711228 | 0.086746 | 0.970658 | | 2.107178 | 0.23741 | 0.903339 |
| FCGR3A | 4.124205 | -0.23666 | 0.999532 | | 4.848399 | -0.06618 | 0.970658 | | 3.909053 | 0.187891 | 0.906811 |
| ZBTB41 | 4.155463 | -0.05506 | 0.999532 | | 4.735456 | 0.057301 | 0.970658 | | 4.217009 | 0.141861 | 0.920527 |
| SATB2 | 1.767097 | -0.20626 | 0.999532 | | 2.137657 | 0.086387 | 0.970658 | | 1.778935 | 0.309858 | 0.87764 |
| PDK3 | 2.719483 | -0.03552 | 0.999532 | | 3.153979 | -0.07507 | 0.970658 | | 2.426589 | -0.02239 | 0.994537 |
| OBSCN | 4.354239 | 0.236922 | 0.999532 | | 4.212585 | -0.0648 | 0.970658 | | 4.140802 | -0.28846 | 0.831749 |
| NFIX | 4.333429 | 0.011686 | 0.999532 | | 4.694512 | -0.06006 | 0.970658 | | 3.954285 | -0.05295 | 0.977473 |
| ZNF627 | 3.028199 | -0.16066 | 0.999532 | | 3.483046 | -0.06705 | 0.970658 | | 2.805821 | 0.111551 | 0.952783 |
| WASF1 | 2.154618 | 0.486783 | 0.999532 | | 2.090435 | 0.08865 | 0.970658 | | 2.208991 | -0.38451 | 0.823193 |
| ENSA | 7.609333 | -0.35894 | 0.999532 | | 9.79858 | 0.046018 | 0.970658 | | 7.190012 | 0.429788 | 0.609651 |
| GID4 | 4.754037 | 0.006834 | 0.999532 | | 5.218249 | -0.05808 | 0.970658 | | 4.424734 | -0.04889 | 0.978931 |
| ANKS1B | 1.748244 | 0.711193 | 0.999532 | | 1.575071 | 0.098747 | 0.970658 | | 1.679346 | -0.5952 | 0.721561 |
| CANT1 | 6.159316 | 0.18619 | 0.999532 | | 6.325093 | 0.054728 | 0.970658 | | 5.746353 | -0.11836 | 0.927668 |
| OR4A5 | 2.274765 | 0.626209 | 0.999532 | | 2.148224 | 0.086683 | 0.970658 | | 2.116348 | -0.52008 | 0.720106 |
| AGPAT3 | 2.572577 | -0.24171 | 0.999532 | | 3.245692 | 0.068271 | 0.970658 | | 2.527947 | 0.33267 | 0.827267 |
| ATRNL1 | 2.349796 | -0.55843 | 0.999532 | | 3.048097 | -0.07194 | 0.970658 | | 2.214352 | 0.504697 | 0.730921 |
| NOP2 | 3.038338 | -0.09061 | 0.999532 | | 3.529378 | -0.073 | 0.970658 | | 2.848085 | 0.039273 | 0.989278 |
| LYPD3 | 2.327756 | 0.438135 | 0.999532 | | 2.072199 | -0.08728 | 0.970658 | | 2.211149 | -0.51111 | 0.743745 |
| INVS | 3.615657 | 0.222128 | 0.999532 | | 3.412824 | -0.06845 | 0.970658 | | 3.301388 | -0.29191 | 0.839373 |
| SYT13 | 2.011431 | 0.253459 | 0.999532 | | 2.139592 | 0.083106 | 0.970658 | | 2.012262 | -0.15234 | 0.941197 |
| A1CF | 2.022838 | 0.014338 | 0.999532 | | 2.212064 | -0.08525 | 0.970658 | | 1.895036 | -0.08113 | 0.974421 |
| RPA1 | 5.341775 | 0.237559 | 0.999532 | | 5.603496 | 0.055859 | 0.970658 | | 5.269611 | -0.16548 | 0.897655 |
| KLRF1 | 1.846061 | 0.247015 | 0.999532 | | 2.033773 | 0.086357 | 0.970658 | | 1.89408 | -0.1409 | 0.948127 |
| NOL8 | 11.05848 | 0.094007 | 0.999532 | | 12.50886 | 0.052355 | 0.970658 | | 11.00524 | -0.00874 | 0.996891 |
| BBS4 | 1.936448 | -0.24226 | 0.999532 | | 2.169505 | -0.0842 | 0.970658 | | 1.755252 | 0.172312 | 0.940162 |
| SAR1A | 12.02317 | -0.08028 | 0.999532 | | 13.60983 | -0.03773 | 0.970658 | | 10.9341 | 0.056908 | 0.960773 |
| C10orf14 | 2.2988 | 0.242929 | 0.999532 | | 2.17341 | -0.09051 | 0.970658 | | 2.013421 | -0.30099 | 0.869666 |
| IL36A | 2.225104 | 0.464272 | 0.999532 | | 2.150745 | -0.08636 | 0.970658 | | 2.152089 | -0.52546 | 0.735469 |
| CAMKK1 | 2.983453 | 0.30993 | 0.999532 | | 2.999404 | -0.07436 | 0.970658 | | 2.826304 | -0.36661 | 0.792416 |
| MUC2 | 2.35153 | 0.723297 | 0.999532 | | 2.21407 | 0.083985 | 0.970658 | | 2.33273 | -0.61295 | 0.65401 |
| TSPAN5 | 2.204456 | -0.02254 | 0.999532 | | 2.463658 | 0.077662 | 0.970658 | | 2.027257 | 0.108193 | 0.9618 |
| MT1B | 1.931823 | 0.050934 | 0.999532 | | 2.194757 | -0.0885 | 0.970658 | | 1.838177 | -0.1209 | 0.960773 |
| WSCD2 | 2.448979 | 0.482083 | 0.999532 | | 2.606764 | 0.079497 | 0.970658 | | 2.438542 | -0.37787 | 0.80383 |
| MEMO1 | 3.716574 | 0.335267 | 0.999532 | | 3.557103 | -0.072 | 0.970658 | | 3.513621 | -0.38857 | 0.749206 |
| BCAR3 | 6.061316 | 0.033441 | 0.999532 | | 6.455617 | -0.05054 | 0.970658 | | 5.74162 | -0.06945 | 0.961567 |
| LTBR | 8.865057 | -0.11784 | 0.999532 | | 9.888826 | -0.04594 | 0.970658 | | 8.426469 | 0.084101 | 0.945456 |
| RPA4 | 2.193944 | 0.07983 | 0.999532 | | 2.332549 | -0.08563 | 0.970715 | | 1.944828 | -0.14667 | 0.946572 |
| RIN3 | 2.300019 | -0.15126 | 0.999532 | | 2.547997 | -0.07776 | 0.970749 | | 2.176763 | 0.095394 | 0.968434 |
| EPHA2 | 3.528082 | -0.91095 | 0.999532 | | 5.139393 | -0.05869 | 0.970749 | | 3.201523 | 0.871208 | 0.408315 |
| ELP6 | 3.276328 | 0.633288 | 0.999532 | | 3.002028 | 0.071989 | 0.970749 | | 3.241118 | -0.5445 | 0.646821 |
| RDH13 | 4.91793 | -0.10811 | 0.999532 | | 5.556801 | -0.05552 | 0.970761 | | 4.577457 | 0.076264 | 0.96588 |
| POP1 | 1.95692 | 0.246841 | 0.999532 | | 2.0175 | -0.08586 | 0.970774 | | 1.906309 | -0.31038 | 0.872808 |
| CD37 | 2.124306 | 0.294358 | 0.999532 | | 2.091928 | -0.0848 | 0.970774 | | 2.063482 | -0.36765 | 0.828869 |
| KRTAP8 | 2.506967 | 0.533731 | 0.999532 | | 2.356693 | 0.07969 | 0.970774 | | 2.441074 | -0.43583 | 0.761337 |
| THOC6 | 3.143919 | -0.24691 | 0.999532 | | 3.409791 | -0.06645 | 0.970774 | | 2.904831 | 0.207546 | 0.901813 |
| TRIP13 | 2.192567 | 0.005359 | 0.999532 | | 2.244437 | -0.08118 | 0.970774 | | 1.987205 | -0.07355 | 0.977473 |
| AMIGO3 | 3.012081 | -0.23764 | 0.999532 | | 3.581645 | -0.06723 | 0.970774 | | 2.798442 | 0.191404 | 0.909074 |
| GRAMD1 | 2.788497 | 0.653244 | 0.999532 | | 2.293823 | -0.0838 | 0.970774 | | 2.641899 | -0.73044 | 0.569849 |
| C1RL | 2.675007 | 0.097382 | 0.999532 | | 2.688363 | -0.07474 | 0.970774 | | 2.515164 | -0.15496 | 0.934701 |
| TRIM41 | 3.453127 | -0.13618 | 0.999532 | | 3.970143 | 0.063036 | 0.970774 | | 3.154235 | 0.213216 | 0.886495 |
| UBN1 | 4.201527 | 0.274391 | 0.999532 | | 4.569466 | 0.059328 | 0.970774 | | 4.02035 | -0.20524 | 0.881689 |
| ASZ1 | 2.199071 | 0.928806 | 0.999532 | | 1.819352 | 0.09306 | 0.970774 | | 2.229744 | -0.82268 | 0.536289 |
| UBE2R2 | 3.413734 | -0.06476 | 0.999532 | | 3.99149 | 0.062581 | 0.970774 | | 3.314907 | 0.146177 | 0.92403 |
| ERCC8 | 2.332009 | -0.08951 | 0.999532 | | 2.813001 | 0.07726 | 0.970774 | | 2.374052 | 0.180482 | 0.920805 |
| SNX16 | 2.003558 | -0.06833 | 0.999532 | | 2.323326 | 0.079033 | 0.970844 | | 2.05722 | 0.160883 | 0.939913 |
| DNAJC22 | 3.468672 | -0.21768 | 0.999532 | | 4.323315 | -0.0626 | 0.970844 | | 3.200374 | 0.188608 | 0.906811 |
| CDV3 | 6.159567 | 0.067656 | 0.999532 | | 6.535247 | -0.05346 | 0.970844 | | 5.633881 | -0.09977 | 0.94109 |
| FGF14 | 2.004364 | 0.228798 | 0.999532 | | 2.169617 | 0.087537 | 0.970844 | | 1.969931 | -0.12025 | 0.96106 |
| PUM3 | 3.050512 | -0.44987 | 0.999532 | | 4.156858 | 0.0633 | 0.970844 | | 3.049166 | 0.544867 | 0.668481 |
| VAMP4 | 4.917365 | 0.740765 | 0.999532 | | 4.054491 | -0.0634 | 0.970844 | | 4.629691 | -0.78396 | 0.34208 |
| SH3YL1 | 4.808207 | -0.23313 | 0.999532 | | 5.584884 | -0.05406 | 0.970844 | | 4.568007 | 0.191462 | 0.881837 |
| MYNN | 2.617642 | -0.03133 | 0.999532 | | 3.10493 | -0.07113 | 0.970844 | | 2.621717 | -0.01696 | 0.996681 |
| DNAH2 | 2.22093 | 0.514115 | 0.999532 | | 2.011135 | -0.08751 | 0.970844 | | 2.106577 | -0.58545 | 0.699971 |
| JOSD1 | 3.930837 | 0.37192 | 0.999532 | | 4.246719 | -0.0628 | 0.970844 | | 3.833303 | -0.38091 | 0.75311 |
| EGLN1 | 3.514128 | -0.12549 | 0.999532 | | 4.148251 | 0.05968 | 0.970844 | | 3.386048 | 0.201157 | 0.889041 |
| ACVR1B | 3.522272 | -0.05267 | 0.999532 | | 4.036255 | -0.06623 | 0.970844 | | 3.269912 | 0.003053 | 0.999385 |
| GLIPR2 | 3.641119 | -0.02843 | 0.999532 | | 3.860798 | -0.06893 | 0.970844 | | 3.558381 | 0.007048 | 0.997967 |
| ZRSR2 | 5.511592 | 0.214495 | 0.999532 | | 5.441536 | -0.05457 | 0.970844 | | 5.008425 | -0.25872 | 0.823193 |
| FAM110B | 2.792921 | -0.22522 | 0.999532 | | 3.167559 | -0.0675 | 0.970844 | | 2.505088 | 0.173367 | 0.920181 |
| RNASE10 | 2.091204 | 0.332454 | 0.999532 | | 2.156246 | -0.08375 | 0.970844 | | 1.996192 | -0.39464 | 0.810635 |
| TPRG1 | 2.188851 | 0.649016 | 0.999532 | | 1.887879 | -0.08685 | 0.970889 | | 2.103039 | -0.71937 | 0.60629 |
| RGS17 | 2.309807 | 0.269255 | 0.999532 | | 2.457803 | 0.079962 | 0.970998 | | 2.331641 | -0.17027 | 0.933313 |
| ENOX2 | 2.038993 | -0.08309 | 0.999532 | | 2.161323 | -0.08103 | 0.970998 | | 1.90992 | 0.018026 | 0.996681 |
| TRAPPC | 2.963669 | 0.4845 | 0.999532 | | 2.734344 | 0.075042 | 0.970998 | | 2.898753 | -0.39043 | 0.788106 |
| PTPRU | 1.951387 | -0.21811 | 0.999532 | | 2.412383 | -0.07783 | 0.970998 | | 1.907506 | 0.161726 | 0.940719 |
| CEP170B | 2.249327 | 0.262223 | 0.999532 | | 2.355566 | -0.07674 | 0.970998 | | 2.137533 | -0.31871 | 0.847837 |
| ELOVL4 | 2.34538 | 0.139499 | 0.999532 | | 2.426609 | -0.0832 | 0.970998 | | 2.170768 | -0.20321 | 0.918675 |
| DNAJB5 | 2.935995 | 0.465906 | 0.999532 | | 2.948132 | 0.073879 | 0.970998 | | 2.94472 | -0.37545 | 0.791576 |
| DALRD3 | 3.083322 | 0.353731 | 0.999532 | | 3.005201 | -0.07158 | 0.970998 | | 2.743089 | -0.395 | 0.7719 |
| TECPR2 | 3.521743 | -0.3972 | 0.999532 | | 4.578956 | -0.05969 | 0.970998 | | 3.471947 | 0.352462 | 0.779082 |
| CD200 | 4.186639 | 0.413417 | 0.999532 | | 3.816313 | -0.06495 | 0.970998 | | 3.854707 | -0.46156 | 0.692762 |
| MEPCE | 14.3569 | -0.35387 | 0.999532 | | 17.2789 | -0.03612 | 0.970998 | | 13.00611 | 0.338535 | 0.609581 |
| DDX1 | 6.212928 | -0.24263 | 0.999532 | | 7.639322 | 0.047654 | 0.970998 | | 5.940865 | 0.309079 | 0.75311 |
| ERI1 | 2.185992 | -0.02228 | 0.999532 | | 2.453033 | 0.075609 | 0.970998 | | 2.117335 | 0.114853 | 0.956889 |
| IRF2 | 4.083505 | 0.101979 | 0.999532 | | 4.136804 | -0.05942 | 0.970998 | | 3.759441 | -0.14685 | 0.920487 |
| ZNF33A | 2.319763 | -0.58203 | 0.999532 | | 2.939068 | -0.07158 | 0.970998 | | 2.140837 | 0.528075 | 0.728015 |
| NMT2 | 3.264537 | 0.094889 | 0.999532 | | 3.767525 | 0.061388 | 0.970998 | | 3.127934 | -0.01268 | 0.996867 |
| C1GALT1 | 5.675737 | 0.100423 | 0.999532 | | 6.170999 | 0.049703 | 0.970998 | | 5.406541 | -0.03469 | 0.985052 |
| SLC26A1 | 2.482544 | 0.106693 | 0.999532 | | 2.514868 | -0.07758 | 0.970998 | | 2.438787 | -0.16579 | 0.938624 |
| PDP2 | 3.345047 | -0.03237 | 0.999532 | | 3.641359 | 0.064652 | 0.970998 | | 3.140616 | 0.105573 | 0.955167 |
| OR10AC | 2.680549 | 0.109686 | 0.999532 | | 2.867323 | 0.073878 | 0.970998 | | 2.564211 | -0.01652 | 0.996681 |
| SLC22A1 | 3.972811 | -0.2024 | 0.999532 | | 4.829704 | 0.056417 | 0.970998 | | 3.820658 | 0.277279 | 0.819366 |
| FBXO24 | 4.2268 | 0.257224 | 0.999532 | | 4.253217 | -0.0595 | 0.970998 | | 4.094799 | -0.29477 | 0.814133 |
| SINHCAF | 5.173754 | 0.482043 | 0.999532 | | 4.743623 | 0.055996 | 0.970998 | | 5.041676 | -0.41497 | 0.675955 |
| CATIP | 2.503993 | 0.603991 | 0.999532 | | 2.166398 | 0.082867 | 0.970998 | | 2.350768 | -0.49937 | 0.730921 |
| SLC6A3 | 2.595311 | 0.559917 | 0.999532 | | 2.23804 | 0.082915 | 0.970998 | | 2.608756 | -0.40726 | 0.809832 |
| PTH | 1.859317 | -0.20498 | 0.999532 | | 2.224992 | -0.08125 | 0.970998 | | 1.721124 | 0.143714 | 0.950616 |
| ZNF235 | 3.620739 | 0.204983 | 0.999532 | | 3.71143 | -0.06424 | 0.970998 | | 3.529412 | -0.24749 | 0.864245 |
| LRRC4B | 3.72243 | 0.650009 | 0.999532 | | 3.082293 | -0.07032 | 0.970998 | | 3.547157 | -0.70255 | 0.515762 |
| CCR5 | 2.853529 | 0.006957 | 0.999532 | | 3.218189 | -0.07572 | 0.971001 | | 2.829077 | -0.07121 | 0.976663 |
| SPRYD7 | 2.478177 | 0.213387 | 0.999532 | | 2.269638 | -0.07764 | 0.971095 | | 2.270145 | -0.28704 | 0.874251 |
| RTL8B | 1.758811 | -0.30004 | 0.999532 | | 2.086275 | -0.08005 | 0.971095 | | 1.645012 | 0.23698 | 0.907968 |
| TP53BP2 | 2.893463 | -0.37262 | 0.999532 | | 3.522865 | -0.06645 | 0.971095 | | 2.705978 | 0.325864 | 0.833941 |
| RAB11FI | 3.413448 | -0.1267 | 0.999532 | | 3.994184 | -0.06501 | 0.971095 | | 3.099998 | 0.08913 | 0.96588 |
| SIGLEC8 | 1.553662 | -0.0139 | 0.999532 | | 1.876718 | 0.089609 | 0.971095 | | 1.647531 | 0.123355 | 0.963866 |
| ANKRD4 | 5.473113 | 0.192208 | 0.999532 | | 5.74494 | 0.051016 | 0.971095 | | 5.237954 | -0.12718 | 0.921791 |
| ITGB7 | 2.66697 | 0.07288 | 0.999532 | | 2.877118 | 0.075484 | 0.971095 | | 2.782789 | -0.00159 | 0.999463 |
| TMEM12 | 5.848048 | -0.03442 | 0.999532 | | 6.469672 | 0.052375 | 0.971204 | | 5.730071 | 0.086639 | 0.955834 |
| ACTRT2 | 2.992009 | 0.05008 | 0.999532 | | 3.354442 | -0.07211 | 0.971204 | | 2.859885 | -0.10056 | 0.959674 |
| ZNF468 | 3.563578 | 0.114662 | 0.999532 | | 4.001108 | 0.066668 | 0.97123 | | 3.442458 | -0.03472 | 0.988853 |
| SMARCA | 2.932458 | -0.06802 | 0.999532 | | 3.274384 | 0.07016 | 0.97123 | | 2.749283 | 0.106285 | 0.954538 |
| MRPL36 | 4.84641 | 0.234349 | 0.999532 | | 5.229179 | 0.057031 | 0.97123 | | 4.655022 | -0.158 | 0.906244 |
| ANKRD3 | 2.053454 | 0.510015 | 0.999532 | | 1.917067 | 0.084861 | 0.97123 | | 2.060911 | -0.40771 | 0.809832 |
| TOMM22 | 4.070075 | 0.068931 | 0.999532 | | 4.484615 | -0.05773 | 0.97123 | | 3.935337 | -0.10894 | 0.943533 |
| BAZ2B | 4.223655 | 0.025803 | 0.999532 | | 4.671079 | -0.05672 | 0.97123 | | 3.984051 | -0.06241 | 0.972305 |
| NYAP2 | 2.245473 | 0.173845 | 0.999532 | | 2.39914 | -0.08175 | 0.97129 | | 2.237722 | -0.24046 | 0.901033 |
| RPL23A | 4.557683 | -0.10709 | 0.999532 | | 5.802163 | 0.054443 | 0.971303 | | 4.495384 | 0.188139 | 0.886495 |
| RPAP1 | 2.248933 | -0.1136 | 0.999532 | | 2.570081 | -0.07549 | 0.971418 | | 2.22751 | 0.050705 | 0.985873 |
| FOXD1 | 2.582468 | 0.649412 | 0.999532 | | 2.375485 | 0.082608 | 0.971418 | | 2.658949 | -0.55349 | 0.707142 |
| SETDB2 | 2.209448 | 0.015962 | 0.999532 | | 2.361204 | 0.077198 | 0.971418 | | 2.145936 | 0.081386 | 0.974421 |
| DCAKD | 2.142079 | -0.09461 | 0.999532 | | 2.563939 | 0.073927 | 0.971418 | | 2.107768 | 0.185586 | 0.920527 |
| NENF | 4.555559 | -0.10842 | 0.999532 | | 5.086766 | -0.05338 | 0.971418 | | 4.289144 | 0.068218 | 0.968434 |
| C4orf51 | 1.920544 | -0.62733 | 0.999532 | | 2.394578 | -0.07366 | 0.971418 | | 1.784433 | 0.569655 | 0.722703 |
| C3orf35 | 2.141893 | -0.34311 | 0.999532 | | 2.555877 | -0.07425 | 0.971418 | | 2.117998 | 0.283761 | 0.881689 |
| ASB8 | 4.603895 | 0.015235 | 0.999532 | | 5.085134 | -0.05726 | 0.971471 | | 4.219817 | -0.04106 | 0.985299 |
| ANAPC1 | 4.09894 | -0.15058 | 0.999532 | | 4.71764 | 0.056312 | 0.971471 | | 3.930507 | 0.21861 | 0.87326 |
| HABP2 | 3.197281 | 0.226599 | 0.999532 | | 3.389684 | -0.06878 | 0.971476 | | 2.923896 | -0.27364 | 0.858899 |
| GNA12 | 3.399911 | -0.00035 | 0.999532 | | 3.675018 | -0.06292 | 0.971476 | | 3.266291 | -0.04739 | 0.983281 |
| HSD17B7 | 2.777526 | 0.115073 | 0.999532 | | 3.072134 | 0.068592 | 0.971533 | | 2.66571 | -0.02739 | 0.992286 |
| ABHD17B | 2.525802 | 0.374884 | 0.999532 | | 2.363184 | -0.07704 | 0.971561 | | 2.328608 | -0.43546 | 0.773057 |
| NIBAN2 | 6.349763 | -0.22028 | 0.999532 | | 7.096799 | -0.06049 | 0.971561 | | 5.929606 | 0.122947 | 0.939549 |
| SYNGR1 | 2.01789 | 0.343349 | 0.999532 | | 1.921808 | -0.08435 | 0.971653 | | 1.952975 | -0.407 | 0.814434 |
| EPPIN | 2.068186 | -0.22757 | 0.999532 | | 2.426384 | -0.08106 | 0.971653 | | 1.917993 | 0.166858 | 0.940266 |
| SNX2 | 6.916839 | -0.26773 | 0.999532 | | 8.34515 | 0.048637 | 0.971653 | | 6.640486 | 0.324089 | 0.746709 |
| SHBG | 2.246994 | 0.2449 | 0.999532 | | 2.283417 | -0.07719 | 0.971683 | | 2.126668 | -0.30471 | 0.862823 |
| CHUK | 3.893233 | -0.27648 | 0.999532 | | 4.454456 | -0.05841 | 0.971686 | | 3.562179 | 0.23368 | 0.876428 |
| ACAP2 | 4.530378 | -0.85719 | 0.999532 | | 6.524183 | 0.048314 | 0.971686 | | 4.480958 | 0.91317 | 0.274496 |
| TATDN1 | 4.282387 | 0.131447 | 0.999532 | | 4.519127 | -0.05761 | 0.971797 | | 4.252035 | -0.17165 | 0.903339 |
| OR5M3 | 2.593817 | 0.362993 | 0.999532 | | 2.442761 | -0.07956 | 0.97192 | | 2.464041 | -0.42003 | 0.794914 |
| CEP20 | 2.523938 | 0.193712 | 0.999532 | | 2.751515 | 0.072167 | 0.97192 | | 2.53311 | -0.10273 | 0.96129 |
| ZNF225 | 2.303714 | 0.935307 | 0.999532 | | 1.781177 | 0.086851 | 0.97192 | | 2.353905 | -0.83018 | 0.533876 |
| TMEM25 | 1.924742 | 0.612152 | 0.999532 | | 1.83126 | 0.08825 | 0.972088 | | 1.981936 | -0.50067 | 0.764702 |
| INO80B | 3.076127 | -0.59466 | 0.999532 | | 4.460189 | 0.057562 | 0.972088 | | 3.035388 | 0.663751 | 0.543211 |
| ABCD1 | 2.144492 | -0.15713 | 0.999532 | | 2.460512 | -0.07338 | 0.972088 | | 1.985217 | 0.100202 | 0.967145 |
| CENPC | 3.604018 | -0.22415 | 0.999532 | | 4.541014 | -0.06309 | 0.972088 | | 3.554362 | 0.17221 | 0.91024 |
| MMD | 2.858013 | -0.07577 | 0.999532 | | 3.292057 | 0.067334 | 0.972088 | | 2.825148 | 0.150147 | 0.938429 |
| AUTS2 | 3.244192 | 0.421927 | 0.999532 | | 3.133809 | 0.068519 | 0.972088 | | 3.10619 | -0.33685 | 0.810635 |
| HLA-DOB | 2.239623 | 0.16433 | 0.999532 | | 2.310906 | -0.07932 | 0.972088 | | 2.161303 | -0.22237 | 0.908595 |
| TMEM12 | 2.134075 | 0.392136 | 0.999532 | | 2.126016 | 0.080403 | 0.972088 | | 2.170597 | -0.30632 | 0.864384 |
| LAGE3 | 3.150118 | 0.063359 | 0.999532 | | 3.209817 | 0.064165 | 0.972088 | | 2.943996 | 0.015371 | 0.996681 |
| ANO10 | 4.076875 | -0.40694 | 0.999532 | | 5.055214 | -0.05684 | 0.972088 | | 3.768163 | 0.366986 | 0.765852 |
| FUT11 | 6.064844 | -0.16434 | 0.999532 | | 7.256962 | 0.045635 | 0.972295 | | 5.764872 | 0.226994 | 0.833237 |
| MOGS | 3.222939 | -0.19248 | 0.999532 | | 3.821836 | -0.05993 | 0.972352 | | 2.92149 | 0.15107 | 0.925486 |
| OR13C8 | 2.351222 | 0.836117 | 0.999532 | | 1.764784 | -0.08552 | 0.972352 | | 2.1638 | -0.90099 | 0.49299 |
| OPHN1 | 3.139554 | -0.01681 | 0.999532 | | 3.49535 | -0.0619 | 0.972352 | | 3.000803 | -0.02479 | 0.992839 |
| CST3 | 15.98935 | -0.09282 | 0.999532 | | 19.11175 | 0.050596 | 0.972352 | | 15.69806 | 0.156156 | 0.885078 |
| FAM81A | 3.1557 | 0.232039 | 0.999532 | | 3.188628 | -0.06832 | 0.972352 | | 2.880594 | -0.27241 | 0.861737 |
| LMOD3 | 2.263774 | 0.150046 | 0.999532 | | 2.394881 | -0.07513 | 0.972352 | | 2.228188 | -0.2093 | 0.907968 |
| SULT1C3 | 2.443899 | 0.397281 | 0.999532 | | 2.418197 | 0.079451 | 0.972352 | | 2.487148 | -0.30348 | 0.8691 |
| ALG12 | 3.484031 | -0.09348 | 0.999532 | | 4.012046 | 0.056117 | 0.972352 | | 3.411064 | 0.16626 | 0.908829 |
| ZNF404 | 1.819952 | 0.158375 | 0.999532 | | 2.021334 | 0.085255 | 0.972352 | | 1.856074 | -0.05641 | 0.986208 |
| RPSA | 59.50631 | 0.019088 | 0.999532 | | 66.67628 | 0.021632 | 0.972352 | | 57.20856 | 0.023514 | 0.978003 |
| FAM98C | 3.910289 | 0.3591 | 0.999532 | | 3.84427 | 0.059474 | 0.972352 | | 3.863343 | -0.28574 | 0.816618 |
| GDPD5 | 2.305033 | 0.213134 | 0.999532 | | 2.165746 | -0.0758 | 0.972352 | | 2.135419 | -0.27418 | 0.880949 |
| UBL4A | 4.066427 | 0.349102 | 0.999532 | | 3.759615 | -0.06056 | 0.972352 | | 3.779007 | -0.39128 | 0.745799 |
| RNF40 | 7.483805 | 0.134565 | 0.999532 | | 8.086537 | -0.04652 | 0.972352 | | 7.047296 | -0.158 | 0.888109 |
| ZNF10 | 3.2581 | 0.299636 | 0.999532 | | 3.704338 | 0.068207 | 0.972352 | | 3.414997 | -0.19149 | 0.908698 |
| IL17RC | 4.788693 | 0.311585 | 0.999532 | | 4.66319 | -0.05408 | 0.972352 | | 4.524657 | -0.34734 | 0.732486 |
| MAPT | 5.17844 | -0.29821 | 0.999532 | | 6.559287 | 0.054631 | 0.972352 | | 4.866 | 0.374199 | 0.722703 |
| POLB | 2.262507 | -0.5669 | 0.999532 | | 2.925865 | -0.06805 | 0.972352 | | 2.057846 | 0.516276 | 0.735469 |
| LACC1 | 1.947786 | 0.228017 | 0.999532 | | 1.884037 | 0.080908 | 0.972352 | | 1.937412 | -0.12937 | 0.956011 |
| ZNF644 | 3.339617 | -0.17818 | 0.999532 | | 3.52569 | -0.06209 | 0.972352 | | 2.974296 | 0.115621 | 0.952783 |
| RBM8A | 8.980917 | -0.33341 | 0.999532 | | 11.19428 | -0.03932 | 0.972352 | | 8.269771 | 0.320619 | 0.705425 |
| RCBTB1 | 6.062459 | -0.1065 | 0.999532 | | 7.244337 | 0.046676 | 0.972352 | | 6.036136 | 0.173605 | 0.884993 |
| HGSNAT | 5.621848 | -0.20994 | 0.999532 | | 7.19808 | 0.049548 | 0.972352 | | 5.595039 | 0.284293 | 0.791576 |
| TMPRSS | 2.38769 | 0.842117 | 0.999532 | | 2.064364 | 0.084718 | 0.972352 | | 2.212078 | -0.74444 | 0.581082 |
| UGT2B4 | 2.062683 | -0.13747 | 0.999532 | | 2.504121 | 0.076467 | 0.972352 | | 2.031578 | 0.231596 | 0.906811 |
| SNTB1 | 3.486352 | 0.068089 | 0.999532 | | 4.305894 | 0.059613 | 0.972352 | | 3.534622 | 0.042541 | 0.986324 |
| MAGEA4 | 3.221056 | 0.442204 | 0.999532 | | 2.92739 | -0.0735 | 0.972352 | | 3.067608 | -0.49962 | 0.706407 |
| THAP6 | 2.917266 | -0.36047 | 0.999532 | | 3.68991 | -0.05964 | 0.972352 | | 2.677939 | 0.318147 | 0.824012 |
| LAMA3 | 2.377896 | 0.526708 | 0.999532 | | 2.131253 | -0.07907 | 0.972352 | | 2.258829 | -0.58879 | 0.686214 |
| PNPLA2 | 13.20951 | -0.21873 | 0.999532 | | 16.40902 | 0.037181 | 0.972352 | | 12.46278 | 0.2795 | 0.717627 |
| PLD6 | 4.026427 | 0.301869 | 0.999532 | | 4.267834 | 0.06695 | 0.972352 | | 4.064485 | -0.20778 | 0.899732 |
| GNAQ | 7.618371 | 0.019605 | 0.999532 | | 8.701339 | 0.043756 | 0.972352 | | 7.478329 | 0.041135 | 0.977183 |
| TLL2 | 2.120462 | 0.248259 | 0.999532 | | 2.238271 | 0.075672 | 0.972352 | | 2.082271 | -0.15274 | 0.941197 |
| PFKM | 2.751458 | -0.01725 | 0.999532 | | 3.129741 | 0.064166 | 0.972352 | | 2.654942 | 0.096693 | 0.962597 |
| KRTCAP | 2.238958 | 0.606342 | 0.999532 | | 2.055527 | -0.08154 | 0.972352 | | 2.208907 | -0.66545 | 0.647632 |
| BYSL | 2.961952 | 0.045076 | 0.999532 | | 3.293278 | -0.06463 | 0.972352 | | 2.833104 | -0.09003 | 0.965338 |
| CCDC122 | 2.578032 | 0.636007 | 0.999532 | | 2.34708 | -0.0762 | 0.972352 | | 2.28272 | -0.6888 | 0.590442 |
| METTL3 | 6.451845 | -0.62156 | 0.999532 | | 8.965267 | 0.04622 | 0.972352 | | 6.134231 | 0.684859 | 0.32832 |
| TEX37 | 2.754196 | 0.472105 | 0.999532 | | 2.713461 | -0.07985 | 0.972352 | | 2.487802 | -0.5339 | 0.695449 |
| PRMT8 | 97.52916 | 0.240328 | 0.999532 | | 99.09791 | -0.04786 | 0.972352 | | 92.23489 | -0.25756 | 0.793131 |
| NEURL2 | 2.887014 | 0.314572 | 0.999532 | | 2.902961 | 0.069073 | 0.972352 | | 2.702895 | -0.22753 | 0.886495 |
| TCP11X2 | 1.87249 | -0.13501 | 0.999532 | | 2.144634 | -0.08014 | 0.972352 | | 1.753274 | 0.072998 | 0.97843 |
| ZDHHC2 | 2.949012 | -0.34516 | 0.999532 | | 3.677247 | -0.05913 | 0.972352 | | 2.722869 | 0.301047 | 0.83987 |
| DMD | 2.301087 | 0.28539 | 0.999532 | | 2.356145 | 0.075753 | 0.972352 | | 2.223281 | -0.18402 | 0.927544 |
| OR52M1 | 2.095141 | 0.480143 | 0.999532 | | 2.009702 | -0.08275 | 0.972352 | | 2.075883 | -0.5433 | 0.733618 |
| MRPS35 | 3.203082 | 0.098255 | 0.999532 | | 3.654367 | 0.063521 | 0.972352 | | 3.031499 | -0.01653 | 0.996681 |
| CCDC18 | 1.942638 | 0.776771 | 0.999532 | | 1.736202 | 0.088442 | 0.972352 | | 1.995891 | -0.67305 | 0.655324 |
| CYP2A6 | 6.651862 | 0.222858 | 0.999532 | | 6.571469 | -0.04915 | 0.972352 | | 6.321571 | -0.26976 | 0.81335 |
| KRT8 | 9.601752 | -0.0505 | 0.999532 | | 10.52335 | -0.04383 | 0.972429 | | 8.885006 | 0.027546 | 0.988379 |
| ASCC3 | 3.640495 | -0.27532 | 0.999532 | | 4.477865 | 0.054009 | 0.972429 | | 3.486342 | 0.347074 | 0.776283 |
| SPC25 | 2.783577 | 0.46983 | 0.999532 | | 2.634322 | -0.07104 | 0.972486 | | 2.484172 | -0.51868 | 0.70494 |
| ZNF780A | 3.266229 | -0.42113 | 0.999532 | | 4.006265 | -0.05832 | 0.972486 | | 3.120975 | 0.383478 | 0.782299 |
| PAPSS2 | 2.948662 | -0.2403 | 0.999532 | | 3.577857 | 0.059956 | 0.972486 | | 2.842629 | 0.316899 | 0.829716 |
| TRAF1 | 2.3035 | 0.277017 | 0.999532 | | 2.246382 | 0.074824 | 0.972486 | | 2.131953 | -0.18972 | 0.920487 |
| CLSTN3 | 3.876624 | -0.08497 | 0.999532 | | 4.319772 | -0.05695 | 0.972486 | | 3.558701 | 0.046149 | 0.982124 |
| ALB | 2.448405 | 0.364317 | 0.999532 | | 2.52806 | -0.07521 | 0.972486 | | 2.256097 | -0.41914 | 0.783005 |
| USP16 | 3.236087 | 0.291699 | 0.999532 | | 3.155763 | -0.06424 | 0.972486 | | 3.127444 | -0.33709 | 0.805825 |
| MGAT3 | 1.850245 | 0.211815 | 0.999532 | | 1.861533 | 0.079213 | 0.972486 | | 1.869677 | -0.11951 | 0.960773 |
| IVNS1AB | 4.870845 | 0.045183 | 0.999532 | | 5.591933 | -0.05305 | 0.972486 | | 4.312101 | -0.15033 | 0.914929 |
| AK5 | 2.846393 | -0.40198 | 0.999532 | | 3.919556 | 0.062017 | 0.972486 | | 2.894306 | 0.48035 | 0.714657 |
| FKBP14 | 3.139333 | 0.041665 | 0.999532 | | 3.182743 | -0.06708 | 0.972486 | | 2.845327 | -0.08617 | 0.969412 |
| ADCY7 | 3.88221 | -0.00398 | 0.999532 | | 4.414775 | -0.05582 | 0.972486 | | 3.659921 | -0.02903 | 0.991159 |
| PKN2 | 2.681444 | -0.14542 | 0.999532 | | 3.123257 | -0.06495 | 0.972486 | | 2.49105 | 0.098143 | 0.961643 |
| ZBTB43 | 2.145058 | -0.15688 | 0.999532 | | 2.622777 | 0.068343 | 0.972486 | | 2.240931 | 0.247543 | 0.891531 |
| PROM1 | 2.156018 | 0.402597 | 0.999532 | | 2.132227 | 0.076738 | 0.972486 | | 2.106632 | -0.30742 | 0.862823 |
| MRPL54 | 5.944993 | 0.027941 | 0.999532 | | 6.820988 | 0.048589 | 0.972486 | | 5.834734 | 0.03753 | 0.981069 |
| ANO7 | 2.288464 | 0.423682 | 0.999532 | | 2.189293 | -0.07587 | 0.9725 | | 2.212457 | -0.47939 | 0.752803 |
| COBL | 2.504775 | -0.02833 | 0.999532 | | 2.863149 | 0.069945 | 0.97254 | | 2.435337 | 0.110823 | 0.957624 |
| GSTM5 | 2.328326 | -0.21498 | 0.999532 | | 2.730122 | -0.07106 | 0.972576 | | 2.216691 | 0.162929 | 0.938429 |
| SLU7 | 2.58197 | 0.143381 | 0.999532 | | 2.780319 | 0.067805 | 0.972576 | | 2.453092 | -0.05964 | 0.978931 |
| FN1 | 6.367418 | -0.31908 | 0.999532 | | 8.144519 | 0.045794 | 0.972583 | | 6.295315 | 0.425409 | 0.758124 |
| SLC26A7 | 2.060391 | 0.638407 | 0.999532 | | 1.925369 | 0.080428 | 0.9728 | | 1.916177 | -0.54293 | 0.718643 |
| IL17RD | 3.443855 | 0.288176 | 0.999532 | | 3.3603 | 0.061499 | 0.9728 | | 3.433772 | -0.2092 | 0.892004 |
| NUP58 | 6.331513 | 0.248128 | 0.999532 | | 6.357202 | -0.05039 | 0.9728 | | 5.731419 | -0.26976 | 0.783005 |
| MVP | 7.317205 | -0.2122 | 0.999532 | | 8.819616 | -0.04906 | 0.9728 | | 7.02039 | 0.182567 | 0.877268 |
| ERAL1 | 2.470192 | 0.267448 | 0.999532 | | 2.478858 | 0.068473 | 0.972897 | | 2.371108 | -0.18316 | 0.918024 |
| SGK2 | 3.848628 | -0.12246 | 0.999532 | | 4.493668 | 0.057051 | 0.972984 | | 3.557632 | 0.180624 | 0.901033 |
| PLD4 | 3.730024 | -0.12068 | 0.999532 | | 4.465725 | 0.055923 | 0.972999 | | 3.715413 | 0.192956 | 0.892869 |
| BCL2L2 | 11.67588 | 0.034539 | 0.999532 | | 12.79646 | 0.038953 | 0.973085 | | 11.27072 | 0.027388 | 0.985569 |
| TMOD1 | 8.616783 | -0.39723 | 0.999532 | | 11.02846 | -0.03984 | 0.973329 | | 8.21809 | 0.376668 | 0.647581 |
| UBAC2 | 3.025155 | 0.37336 | 0.999532 | | 3.070164 | 0.061859 | 0.973329 | | 3.020257 | -0.29361 | 0.831749 |
| ELP3 | 4.004924 | 0.035796 | 0.999532 | | 4.529754 | -0.05473 | 0.973342 | | 3.881741 | -0.0713 | 0.969073 |
| SLAIN1 | 1.748527 | -0.29997 | 0.999532 | | 1.973101 | -0.07979 | 0.973494 | | 1.527767 | 0.236164 | 0.915533 |
| FGR | 2.60269 | -0.05935 | 0.999532 | | 3.112367 | 0.068587 | 0.973606 | | 2.822934 | 0.119713 | 0.952656 |
| GPSM1 | 5.89907 | 0.262734 | 0.999532 | | 5.99345 | -0.05418 | 0.973742 | | 5.397382 | -0.27925 | 0.791466 |
| TCERG1L | 1.746169 | 0.012871 | 0.999532 | | 2.095528 | 0.078126 | 0.973742 | | 1.75701 | 0.086191 | 0.974421 |
| SLC12A6 | 2.52617 | -0.06346 | 0.999532 | | 2.704098 | -0.06883 | 0.974771 | | 2.438106 | 0.008332 | 0.997967 |
| TEX44 | 2.344124 | 0.474213 | 0.999532 | | 2.143131 | 0.073232 | 0.974771 | | 2.267707 | -0.38604 | 0.807425 |
| ELAPOR | 2.306214 | -0.20186 | 0.999532 | | 2.754836 | 0.066385 | 0.974811 | | 2.211566 | 0.284276 | 0.870663 |
| ASB3 | 1.977192 | -0.39926 | 0.999532 | | 2.296956 | -0.07123 | 0.974883 | | 1.845816 | 0.346131 | 0.858705 |
| KLHL3 | 3.180013 | 0.34946 | 0.999532 | | 3.28251 | 0.066745 | 0.974891 | | 3.01625 | -0.26198 | 0.865065 |
| CDKN1B | 12.16431 | 0.107786 | 0.999532 | | 13.15519 | 0.036185 | 0.974894 | | 11.87136 | -0.04455 | 0.972221 |

| | | | | | | | | | | | |
|---|---|---|---|---|---|---|---|---|---|---|---|
| TRNT1 | 3.582332 | -0.34423 | 0.999532 | | 4.677399 | -0.05211 | 0.974894 | | 3.367104 | 0.310517 | 0.809832 |
| CCNYL1 | 3.64407 | -0.03809 | 0.999532 | | 4.198035 | -0.05604 | 0.974894 | | 3.409738 | 0.001054 | 0.99959 |
| FAM149B | 5.250849 | 0.151723 | 0.999532 | | 5.428083 | -0.05002 | 0.974894 | | 4.799836 | -0.18325 | 0.885712 |
| NECTIN3 | 3.434995 | 0.160926 | 0.999532 | | 3.39888 | -0.0585 | 0.974894 | | 3.226251 | -0.20498 | 0.892869 |
| RNASE9 | 2.127592 | 0.567002 | 0.999532 | | 2.071741 | 0.07753 | 0.974894 | | 2.11838 | -0.46847 | 0.776133 |
| HAT1 | 2.86534 | -0.74283 | 0.999532 | | 3.842036 | -0.05419 | 0.974894 | | 2.74898 | 0.706983 | 0.553283 |
| SMCO1 | 2.441951 | 0.208594 | 0.999532 | | 2.592545 | -0.07256 | 0.974894 | | 2.321783 | -0.25297 | 0.892869 |
| DPYSL4 | 2.042199 | -0.15874 | 0.999532 | | 2.287928 | -0.07289 | 0.974894 | | 1.916853 | 0.101528 | 0.969783 |
| DGKH | 4.594007 | -0.30045 | 0.999532 | | 5.340937 | -0.04869 | 0.974894 | | 4.24343 | 0.265568 | 0.830822 |
| CUL7 | 3.32847 | 0.139773 | 0.999532 | | 3.357934 | -0.05919 | 0.974894 | | 2.987274 | -0.18367 | 0.905952 |
| ELOF1 | 3.775308 | 0.089342 | 0.999532 | | 4.05926 | -0.05307 | 0.974894 | | 3.47847 | -0.12442 | 0.938429 |
| AQP11 | 2.433405 | 0.154504 | 0.999532 | | 2.648148 | -0.06564 | 0.974894 | | 2.352396 | -0.19973 | 0.908891 |
| FOXD2 | 6.873955 | 0.414597 | 0.999532 | | 6.637939 | -0.04399 | 0.974894 | | 6.504011 | -0.43663 | 0.598177 |
| SCOC | 7.18043 | 0.337542 | 0.999532 | | 6.737649 | -0.04339 | 0.974894 | | 6.690127 | -0.37241 | 0.682165 |
| GPR180 | 2.894927 | 0.250316 | 0.999532 | | 2.982433 | -0.06629 | 0.974894 | | 2.727118 | -0.29757 | 0.849021 |
| ASAP3 | 2.021583 | 0.266142 | 0.999532 | | 1.960074 | -0.07682 | 0.974894 | | 1.938392 | -0.32878 | 0.862823 |
| SEPTIN4 | 6.051992 | 0.061143 | 0.999532 | | 6.590962 | -0.05026 | 0.974894 | | 5.848704 | -0.09391 | 0.946311 |
| ABCE1 | 2.683125 | -0.05822 | 0.999532 | | 3.105375 | 0.063302 | 0.974894 | | 2.532287 | 0.140525 | 0.940246 |
| SLC25A3 | 2.899006 | 0.307103 | 0.999532 | | 2.749126 | 0.070497 | 0.974894 | | 2.68012 | -0.20996 | 0.905952 |
| AGER | 2.28264 | 0.306901 | 0.999532 | | 2.350217 | 0.072676 | 0.974894 | | 2.340778 | -0.21756 | 0.906811 |
| OSGEP | 5.531137 | -0.31295 | 0.999532 | | 7.253216 | 0.045542 | 0.974894 | | 5.548463 | 0.378024 | 0.699971 |
| ERBB2 | 5.506129 | 0.341227 | 0.999532 | | 5.23147 | -0.04908 | 0.974894 | | 5.005198 | -0.37502 | 0.707142 |
| KHDRBS1 | 6.408233 | -0.75368 | 0.893099 | | 8.974388 | 0.03764 | 0.974894 | | 6.28996 | 0.807981 | 0.222058 |
| SFSWAP | 7.734286 | 0.034579 | 0.999532 | | 8.309904 | -0.0398 | 0.974894 | | 7.180648 | -0.0572 | 0.966652 |
| LRSAM1 | 3.669746 | 0.175066 | 0.999532 | | 3.662575 | -0.05764 | 0.974894 | | 3.31865 | -0.21591 | 0.883785 |
| MSMO1 | 3.725379 | -0.36526 | 0.999532 | | 4.564728 | -0.05364 | 0.974894 | | 3.472661 | 0.331099 | 0.806394 |
| TNPO2 | 2.436407 | 0.348998 | 0.999532 | | 2.433793 | -0.07096 | 0.974894 | | 2.328193 | -0.40182 | 0.794251 |
| ZAN | 1.732183 | 0.069256 | 0.999532 | | 1.807374 | -0.07775 | 0.974894 | | 1.685213 | -0.12833 | 0.958191 |
| NEU4 | 2.064154 | -0.05619 | 0.999532 | | 2.330819 | -0.0759 | 0.974894 | | 1.981538 | -0.00084 | 0.999746 |
| VPS53 | 3.942446 | -0.17035 | 0.999532 | | 4.639851 | 0.050139 | 0.974894 | | 4.024499 | 0.239879 | 0.859092 |
| CDR2 | 3.460598 | 0.159482 | 0.999532 | | 3.537396 | -0.06348 | 0.974894 | | 3.178051 | -0.19654 | 0.899732 |
| PCSK7 | 5.608717 | -0.09241 | 0.999532 | | 6.590879 | 0.044821 | 0.974894 | | 5.507831 | 0.153875 | 0.898965 |
| GDPD1 | 2.084255 | 0.056633 | 0.999532 | | 2.274064 | 0.072971 | 0.974894 | | 2.069279 | 0.036394 | 0.991273 |
| LBHD2 | 1.82381 | 0.012928 | 0.999532 | | 1.92899 | -0.07643 | 0.974894 | | 1.65464 | -0.07606 | 0.978282 |
| CERS4 | 3.32108 | 0.319626 | 0.999532 | | 3.1861 | 0.05891 | 0.974894 | | 3.259401 | -0.2384 | 0.871713 |
| DENND1A | 3.236053 | 0.303186 | 0.999532 | | 3.19366 | 0.059947 | 0.974894 | | 3.125147 | -0.22347 | 0.881772 |
| RNF19A | 5.398877 | -0.43915 | 0.999532 | | 6.942969 | 0.047725 | 0.974894 | | 5.047614 | 0.512578 | 0.578798 |
| APIP | 3.5604 | -0.05645 | 0.999532 | | 3.956453 | -0.0568 | 0.974894 | | 3.291267 | 0.015269 | 0.996681 |
| ACTL6B | 2.093796 | 0.121702 | 0.999532 | | 2.332084 | -0.07299 | 0.974894 | | 2.140506 | -0.17743 | 0.929087 |
| SIGLEC7 | 2.356563 | 0.793029 | 0.999532 | | 2.132383 | 0.07786 | 0.974894 | | 2.447184 | -0.69265 | 0.630705 |
| FITM2 | 2.340474 | 0.264643 | 0.999532 | | 2.367913 | 0.071728 | 0.974894 | | 2.283979 | -0.17482 | 0.93042 |
| BRCA2 | 2.781331 | 0.104824 | 0.999532 | | 2.99811 | -0.06623 | 0.974894 | | 2.774476 | -0.15315 | 0.938429 |
| DUSP14 | 4.124914 | 0.193848 | 0.999532 | | 4.445739 | 0.055039 | 0.974894 | | 4.057814 | -0.12104 | 0.939107 |
| H4C1 | 6.275456 | 0.140915 | 0.999532 | | 6.746955 | -0.0453 | 0.974894 | | 5.708972 | -0.15795 | 0.895647 |
| ZNF574 | 3.205751 | -0.78886 | 0.999532 | | 4.204218 | -0.05276 | 0.974894 | | 2.960148 | 0.753315 | 0.515762 |
| LSM2 | 3.972007 | -0.00122 | 0.999532 | | 4.514442 | 0.051975 | 0.974952 | | 3.88942 | 0.070841 | 0.968235 |
| CSNK2A2 | 3.661495 | -0.13879 | 0.999532 | | 4.24738 | -0.05148 | 0.975077 | | 3.545667 | 0.110464 | 0.946512 |
| HSPD1 | 15.58854 | -0.13175 | 0.999532 | | 17.98323 | 0.03092 | 0.975164 | | 14.7138 | 0.185828 | 0.788446 |
| PTPN2 | 3.842572 | -0.05633 | 0.999532 | | 4.250943 | 0.053071 | 0.975164 | | 3.529746 | 0.122152 | 0.940266 |
| DNAH9 | 2.518257 | 0.336512 | 0.999532 | | 2.433065 | 0.070297 | 0.975164 | | 2.318195 | -0.24298 | 0.88885 |
| BECN1 | 4.911537 | -0.01752 | 0.999532 | | 5.341127 | -0.05035 | 0.975164 | | 4.378983 | -0.00575 | 0.997967 |
| RPL22L1 | 3.192949 | 0.065701 | 0.999532 | | 3.646784 | 0.059953 | 0.975242 | | 2.905889 | 0.032992 | 0.990309 |
| ARID3A | 2.350601 | 0.207495 | 0.999532 | | 2.396156 | -0.06806 | 0.975242 | | 2.219724 | -0.25704 | 0.883565 |
| HDDC3 | 2.821157 | -0.04748 | 0.999532 | | 3.180582 | 0.060982 | 0.975242 | | 2.629609 | 0.124738 | 0.947076 |
| EFCAB12 | 2.585531 | 0.28713 | 0.999532 | | 2.542356 | -0.06916 | 0.975242 | | 2.42449 | -0.33841 | 0.839373 |
| CHPT1 | 5.090513 | -0.3012 | 0.999532 | | 6.049746 | -0.0479 | 0.975242 | | 4.517921 | 0.276472 | 0.809832 |
| SLC2A11 | 2.973424 | 0.470416 | 0.999532 | | 2.883192 | 0.065834 | 0.975242 | | 3.017246 | -0.39111 | 0.791466 |
| COMMD3 | 4.831285 | 0.232249 | 0.999532 | | 5.10551 | 0.048764 | 0.975242 | | 4.774632 | -0.16326 | 0.903339 |
| ABCA1 | 2.828494 | -0.12443 | 0.999532 | | 3.332012 | 0.057904 | 0.975242 | | 2.593123 | 0.195961 | 0.90424 |
| CXCR4 | 4.636033 | -0.50265 | 0.999532 | | 6.441079 | 0.044002 | 0.975242 | | 4.602489 | 0.569774 | 0.536289 |
| SGMS2 | 1.767151 | -0.20391 | 0.999532 | | 2.200933 | 0.0738 | 0.975242 | | 1.758841 | 0.295575 | 0.881837 |
| WDR5B | 2.420754 | 0.336558 | 0.999532 | | 2.546438 | 0.069346 | 0.975242 | | 2.377875 | -0.248 | 0.886495 |
| MEGF8 | 8.10187 | -0.06524 | 0.999532 | | 8.998999 | -0.03845 | 0.9755 | | 7.703851 | 0.038783 | 0.978229 |
| ZNF185 | 3.75029 | 0.568983 | 0.999532 | | 3.259394 | 0.057517 | 0.9755 | | 3.60414 | -0.50075 | 0.660958 |
| ADGRL4 | 2.169907 | -0.52256 | 0.999532 | | 2.724779 | -0.06195 | 0.9755 | | 2.064157 | 0.474532 | 0.760473 |
| TRIM65 | 3.989391 | 0.009558 | 0.999532 | | 4.475215 | -0.05097 | 0.9755 | | 3.791044 | -0.03834 | 0.985693 |
| THBD | 3.282728 | -0.03747 | 0.999532 | | 4.032489 | 0.054297 | 0.9755 | | 3.254001 | 0.118296 | 0.946564 |
| CEACAM | 8.840727 | 0.032869 | 0.999532 | | 10.16979 | 0.043984 | 0.9755 | | 8.504368 | 0.034171 | 0.984624 |
| TTC9C | 6.499971 | 0.165501 | 0.999532 | | 6.807804 | -0.04759 | 0.975664 | | 6.215112 | -0.19559 | 0.864923 |
| KNSTRN | 2.500764 | 0.129026 | 0.999532 | | 2.50963 | -0.06893 | 0.975664 | | 2.336349 | -0.19112 | 0.919541 |
| FBLN7 | 2.611457 | 0.833394 | 0.999532 | | 2.179018 | 0.072578 | 0.97568 | | 2.605221 | -0.73992 | 0.562307 |
| RNASE4 | 2.808581 | 0.270301 | 0.999532 | | 2.723678 | 0.065498 | 0.97568 | | 2.533943 | -0.18586 | 0.914929 |
| GDAP2 | 2.082837 | -0.12457 | 0.999532 | | 2.466212 | 0.069767 | 0.97568 | | 1.834377 | 0.200223 | 0.920259 |
| SART1 | 4.135252 | -0.25605 | 0.999532 | | 5.124739 | -0.04885 | 0.97568 | | 4.071523 | 0.231115 | 0.865065 |
| GJB1 | 2.793539 | 0.565137 | 0.999532 | | 2.378037 | -0.06726 | 0.97568 | | 2.526436 | -0.61327 | 0.632923 |
| PRR20G | 1.996109 | 0.229855 | 0.999532 | | 2.058359 | -0.0739 | 0.975898 | | 2.014563 | -0.29259 | 0.878299 |
| NIPAL2 | 2.815694 | 0.008436 | 0.999532 | | 3.034276 | 0.062218 | 0.976021 | | 2.691637 | 0.059626 | 0.979466 |
| RNFT1 | 2.219823 | -0.12466 | 0.999532 | | 2.533137 | -0.06431 | 0.976021 | | 2.031705 | 0.077699 | 0.974421 |
| RNF11 | 12.61648 | -0.24113 | 0.999532 | | 15.23336 | 0.030772 | 0.976021 | | 12.06397 | 0.281576 | 0.706749 |
| RAP2B | 8.076057 | 0.070071 | 0.999532 | | 8.89988 | 0.039703 | 0.976024 | | 7.71693 | -0.00898 | 0.996891 |
| VSX2 | 2.740044 | 0.24038 | 0.999532 | | 2.870606 | -0.06426 | 0.976024 | | 2.720913 | -0.28504 | 0.864245 |
| SERHL2 | 3.121606 | 0.959696 | 0.999532 | | 2.393473 | 0.07216 | 0.976024 | | 3.264667 | -0.88418 | 0.464413 |
| C4orf33 | 2.538222 | 0.439094 | 0.999532 | | 2.444057 | -0.06829 | 0.976267 | | 2.431813 | -0.4877 | 0.736481 |
| NECAP2 | 6.802411 | 0.093454 | 0.999532 | | 7.280731 | 0.041656 | 0.976267 | | 6.594265 | -0.04029 | 0.979282 |
| ECSIT | 3.042383 | 0.126286 | 0.999532 | | 3.154164 | 0.057106 | 0.976267 | | 2.952732 | -0.04975 | 0.982124 |
| IGLL5 | 2.47801 | 0.273694 | 0.999532 | | 2.260519 | -0.07195 | 0.976267 | | 2.24974 | -0.35713 | 0.855088 |
| RILPL1 | 5.168892 | -0.02933 | 0.999532 | | 5.593242 | -0.0451 | 0.976267 | | 4.755142 | -0.0055 | 0.997967 |
| TTC23 | 2.935454 | -0.21739 | 0.999532 | | 3.416093 | -0.05665 | 0.976267 | | 2.650468 | 0.175616 | 0.916944 |
| ATP6V1C | 2.771009 | -0.11125 | 0.999532 | | 3.183688 | 0.057646 | 0.976267 | | 2.642185 | 0.187158 | 0.909074 |
| FOXA1 | 2.043537 | 0.335887 | 0.999532 | | 2.165307 | 0.076687 | 0.976267 | | 2.046545 | -0.23673 | 0.905952 |
| KRTAP7- | 2.360537 | 0.309732 | 0.999532 | | 2.34206 | -0.07029 | 0.976267 | | 2.281057 | -0.3613 | 0.827625 |
| SARM1 | 2.072144 | -0.26575 | 0.999532 | | 2.554396 | 0.063416 | 0.976267 | | 1.952564 | 0.34716 | 0.840357 |
| SPEM1 | 2.23927 | 0.597871 | 0.999532 | | 1.92777 | -0.0748 | 0.976342 | | 2.043431 | -0.65237 | 0.654161 |
| CHRNA2 | 1.571853 | -0.07757 | 0.999532 | | 1.834211 | 0.075137 | 0.976342 | | 1.611045 | 0.172473 | 0.942744 |
| ELN | 3.570044 | -0.13758 | 0.999532 | | 4.236995 | -0.0552 | 0.976378 | | 3.423266 | 0.109942 | 0.954538 |
| NOS3 | 4.536486 | -0.48608 | 0.999532 | | 5.757205 | -0.04619 | 0.976488 | | 4.143434 | 0.455845 | 0.67176 |
| GTF3C4 | 2.666425 | -0.1727 | 0.999532 | | 3.349704 | 0.057488 | 0.976576 | | 2.584664 | 0.248433 | 0.875951 |
| DEFB130 | 2.685007 | 0.204584 | 0.999532 | | 2.828719 | -0.06399 | 0.976634 | | 2.623484 | -0.25021 | 0.881837 |
| HECTD2 | 4.341241 | 0.213998 | 0.999532 | | 4.211992 | -0.05033 | 0.976721 | | 4.095044 | -0.2464 | 0.853068 |
| DUSP12 | 2.690306 | 0.228713 | 0.999532 | | 2.671558 | -0.06169 | 0.976886 | | 2.540955 | -0.27327 | 0.870247 |
| ARTN | 2.024114 | 0.106508 | 0.999532 | | 2.2687 | -0.07152 | 0.976886 | | 1.93901 | -0.15178 | 0.945902 |
| KLHL23 | 2.366177 | -0.08597 | 0.999532 | | 2.523814 | -0.06345 | 0.9769 | | 2.254558 | 0.030948 | 0.992286 |
| AKAP12 | 20.89667 | -0.04279 | 0.999532 | | 24.44847 | 0.02942 | 0.9769 | | 20.23842 | 0.081753 | 0.914687 |
| POP7 | 3.06306 | 0.130293 | 0.999532 | | 3.44229 | 0.061383 | 0.9769 | | 3.019911 | -0.04889 | 0.984624 |
| VAPB | 4.049328 | -0.23176 | 0.999532 | | 4.63556 | -0.04864 | 0.976938 | | 3.782812 | 0.192416 | 0.892869 |
| SOHLH2 | 1.9276 | 0.300234 | 0.999532 | | 2.009041 | -0.07274 | 0.976938 | | 1.941692 | -0.34772 | 0.853053 |
| BCAS1 | 1.527112 | 0.272654 | 0.999532 | | 1.683199 | 0.0813 | 0.976938 | | 1.636308 | -0.17053 | 0.945759 |
| DRAM2 | 2.185555 | 0.391997 | 0.999532 | | 2.226876 | 0.068862 | 0.976938 | | 2.102399 | -0.3038 | 0.861737 |
| PECR | 2.624816 | 0.290136 | 0.999532 | | 2.60711 | 0.06198 | 0.976938 | | 2.616936 | -0.20602 | 0.901393 |
| POU4F2 | 2.326272 | 0.319933 | 0.999532 | | 2.302009 | -0.0689 | 0.976938 | | 2.248973 | -0.38627 | 0.837927 |
| TMEM52B | 4.708251 | 0.286461 | 0.999532 | | 4.668636 | -0.06257 | 0.976984 | | 4.153053 | -0.34519 | 0.840285 |
| PDRG1 | 2.268588 | 0.391164 | 0.999532 | | 2.156667 | -0.06977 | 0.9771 | | 2.222521 | -0.44597 | 0.78258 |
| COLEC12 | 2.105176 | -0.64927 | 0.999532 | | 2.769398 | -0.06359 | 0.9771 | | 1.906812 | 0.604203 | 0.703463 |
| NSL1 | 3.884755 | -0.14652 | 0.999532 | | 4.38935 | -0.05261 | 0.9771 | | 3.675855 | 0.116297 | 0.941197 |
| LARGE2 | 2.700911 | 0.228012 | 0.999532 | | 2.918347 | 0.065065 | 0.977145 | | 2.754092 | -0.15898 | 0.932589 |
| CST5 | 2.200102 | 0.395561 | 0.999532 | | 2.219214 | 0.0711 | 0.977145 | | 2.172972 | -0.30697 | 0.861737 |
| DDX3Y | 2.942241 | -0.0292 | 0.999532 | | 3.524309 | 0.069555 | 0.977195 | | 2.783465 | 0.147566 | 0.96129 |
| DCPS | 2.689778 | 0.146301 | 0.999532 | | 3.063796 | 0.060276 | 0.977195 | | 2.690361 | -0.0708 | 0.974421 |
| YES1 | 7.612387 | -0.4831 | 0.999532 | | 9.824698 | -0.03558 | 0.977195 | | 7.1922 | 0.470043 | 0.563627 |
| PTPN3 | 2.33593 | 0.30162 | 0.999532 | | 2.528801 | 0.062645 | 0.977195 | | 2.330197 | -0.21109 | 0.90424 |
| DGAT2 | 2.629903 | 0.898844 | 0.999532 | | 2.010066 | -0.06978 | 0.977195 | | 2.505305 | -0.95412 | 0.387954 |
| SLC1A5 | 3.62289 | 0.378826 | 0.999532 | | 3.495545 | -0.0583 | 0.977195 | | 3.408539 | -0.41935 | 0.723453 |
| LEO1 | 3.325109 | -0.06447 | 0.999532 | | 3.998113 | -0.05642 | 0.977195 | | 3.222188 | 0.037993 | 0.988853 |
| NT5C1B | 2.051182 | 0.778726 | 0.999532 | | 1.603186 | -0.07862 | 0.977195 | | 1.92819 | -0.84255 | 0.556866 |
| AKAP10 | 4.225632 | -0.15866 | 0.999532 | | 4.91652 | -0.04643 | 0.977195 | | 4.026097 | 0.134099 | 0.924131 |
| LPA | 2.138634 | 0.406361 | 0.999532 | | 2.071905 | -0.07258 | 0.977221 | | 2.14108 | -0.46619 | 0.791576 |
| PRPF31 | 4.660617 | -0.50319 | 0.999532 | | 6.270197 | 0.041521 | 0.977505 | | 4.406293 | 0.561993 | 0.541754 |
| REXO2 | 4.459238 | -0.39238 | 0.999532 | | 5.932136 | 0.04467 | 0.977505 | | 4.434834 | 0.450544 | 0.664512 |
| MRFAP1 | 12.25309 | -0.05374 | 0.999532 | | 14.15503 | 0.03239 | 0.977505 | | 11.95089 | 0.110085 | 0.920259 |
| FUT7 | 2.447019 | 0.572737 | 0.999532 | | 2.14699 | -0.06644 | 0.977505 | | 2.311721 | -0.61948 | 0.652925 |
| PAICS | 3.102129 | -0.44619 | 0.999532 | | 4.406629 | 0.049566 | 0.977505 | | 3.228935 | 0.522465 | 0.664512 |
| TBPL1 | 3.278219 | -0.22747 | 0.999532 | | 4.158866 | 0.053974 | 0.977505 | | 3.153257 | 0.295793 | 0.836453 |
| MAGEF1 | 4.593651 | -0.30448 | 0.999532 | | 6.167901 | 0.044082 | 0.977505 | | 4.703823 | 0.3742 | 0.723933 |
| ANKRD13 | 4.987019 | -0.24945 | 0.999532 | | 6.043654 | -0.04469 | 0.977505 | | 4.519926 | 0.227676 | 0.858705 |
| ZNF701 | 2.018159 | -0.1808 | 0.999532 | | 2.322412 | 0.065306 | 0.977505 | | 1.9622 | 0.267167 | 0.88734 |
| PLA2G2C | 1.877563 | -0.04358 | 0.999532 | | 1.996684 | -0.0718 | 0.977505 | | 1.839527 | -0.01733 | 0.996891 |
| NMRK1 | 3.399616 | -0.02709 | 0.999532 | | 3.720452 | 0.052788 | 0.977505 | | 3.253934 | 0.096523 | 0.956322 |
| MRAP2 | 2.323354 | 0.733031 | 0.999532 | | 2.023666 | 0.070704 | 0.977505 | | 2.402231 | -0.64971 | 0.636625 |
| FAM163A | 3.00465 | 0.052504 | 0.999532 | | 3.552733 | -0.06023 | 0.977505 | | 3.069097 | -0.08905 | 0.967577 |
| MRPL28 | 5.336164 | -0.0103 | 0.999532 | | 5.749331 | 0.045444 | 0.977505 | | 5.077616 | 0.049028 | 0.978282 |
| GUCY2C | 2.294931 | 0.014036 | 0.999532 | | 2.436499 | -0.06489 | 0.977505 | | 2.138856 | -0.0599 | 0.981069 |
| IFT27 | 2.044299 | 0.182151 | 0.999532 | | 2.191768 | -0.06515 | 0.977505 | | 1.91662 | -0.22761 | 0.901033 |
| PABPN1L | 2.44817 | 0.479156 | 0.999532 | | 2.155976 | -0.06706 | 0.977505 | | 2.359958 | -0.52845 | 0.714657 |
| LRCH3 | 2.577719 | -0.50597 | 0.999532 | | 3.326939 | -0.05491 | 0.977505 | | 2.463927 | 0.468706 | 0.737651 |
| CMKLR1 | 3.793987 | -0.19141 | 0.999532 | | 4.546492 | -0.04953 | 0.977626 | | 3.569507 | 0.160958 | 0.914929 |
| FBXO36 | 2.072375 | -0.22456 | 0.999532 | | 2.510033 | 0.061031 | 0.977626 | | 1.93111 | 0.301148 | 0.864923 |
| STK39 | 2.582879 | 0.140778 | 0.999532 | | 2.961252 | 0.059133 | 0.977626 | | 2.54146 | -0.06243 | 0.978229 |
| PARG | 2.608578 | 0.298459 | 0.999532 | | 2.723288 | 0.061653 | 0.977626 | | 2.548821 | -0.22084 | 0.892869 |
| PTPRF | 4.570309 | 0.1137 | 0.999532 | | 4.950135 | 0.045087 | 0.977626 | | 4.280925 | -0.0508 | 0.977473 |
| SH2B2 | 2.164577 | -0.0264 | 0.999532 | | 2.312307 | -0.06454 | 0.977626 | | 2.019815 | -0.01944 | 0.996681 |
| FPGT | 4.856889 | 0.406321 | 0.999532 | | 4.350093 | -0.05008 | 0.977626 | | 4.419952 | -0.43227 | 0.668481 |
| EPHB6 | 1.859379 | 0.442136 | 0.999532 | | 1.774143 | 0.073538 | 0.977626 | | 1.844613 | -0.35145 | 0.857043 |
| CD248 | 9.289542 | -0.52504 | 0.999532 | | 11.76399 | -0.0432 | 0.977626 | | 8.757184 | 0.509555 | 0.511836 |
| PBK | 2.280104 | 0.168558 | 0.999532 | | 2.359909 | -0.0697 | 0.977626 | | 2.081254 | -0.21662 | 0.910105 |
| STIMATE | 4.349536 | -0.94414 | 0.727584 | | 6.404231 | 0.041202 | 0.977626 | | 4.350903 | 0.997439 | 0.21862 |
| GRWD1 | 3.568873 | 0.06014 | 0.999532 | | 3.933548 | 0.051111 | 0.977626 | | 3.616487 | 0.011242 | 0.996891 |
| AKR7L | 2.124289 | 0.280015 | 0.999532 | | 2.344378 | 0.070262 | 0.977626 | | 2.136654 | -0.18773 | 0.92697 |
| TRPV4 | 2.482071 | 0.29424 | 0.999532 | | 2.534815 | -0.06465 | 0.977626 | | 2.47244 | -0.33833 | 0.839373 |
| MRPL40 | 2.127778 | 0.032843 | 0.999532 | | 2.503138 | 0.06231 | 0.977626 | | 2.070653 | 0.047674 | 0.986679 |
| TSPAN3 | 5.905184 | -0.02871 | 0.999532 | | 6.492793 | -0.04555 | 0.977626 | | 5.36871 | -0.00563 | 0.997967 |
| TMEM175 | 3.960309 | 0.040046 | 0.999532 | | 3.974652 | -0.04897 | 0.977626 | | 3.653746 | -0.06653 | 0.972221 |
| SLC7A13 | 1.869064 | 0.356626 | 0.999532 | | 1.898853 | 0.07436 | 0.977626 | | 1.87952 | -0.26376 | 0.898369 |
| DLG4 | 4.894581 | -0.61639 | 0.999532 | | 6.507649 | -0.04283 | 0.977626 | | 4.614854 | 0.590875 | 0.532088 |
| KCNQ1 | 2.968001 | 0.155832 | 0.999532 | | 3.104985 | -0.05444 | 0.977626 | | 2.812732 | -0.19226 | 0.900583 |
| WFDC10A | 1.922452 | 0.109031 | 0.999532 | | 2.07168 | 0.072686 | 0.977626 | | 1.866671 | -0.0217 | 0.996681 |
| RPP21 | 2.567433 | -0.07769 | 0.999532 | | 2.952691 | -0.05763 | 0.977626 | | 2.272981 | 0.042849 | 0.987491 |
| DZIP1 | 2.04397 | -0.04692 | 0.999532 | | 2.436016 | 0.065182 | 0.977626 | | 2.093497 | 0.128344 | 0.953949 |
| MAP2K7 | 4.711402 | 0.140621 | 0.999532 | | 4.900752 | 0.046603 | 0.977626 | | 4.49153 | -0.07751 | 0.962682 |
| POLD3 | 2.308601 | 0.237968 | 0.999532 | | 2.460797 | 0.066005 | 0.977626 | | 2.167012 | -0.14877 | 0.941976 |
| ZNF114 | 2.646085 | 0.087412 | 0.999532 | | 2.808739 | -0.06024 | 0.977637 | | 2.653199 | -0.13275 | 0.946572 |
| HCAR1 | 2.900996 | 0.056742 | 0.999532 | | 3.140264 | -0.05879 | 0.977637 | | 2.650421 | -0.09791 | 0.961948 |
| MRPL19 | 4.362267 | 0.115575 | 0.999532 | | 4.758651 | 0.045989 | 0.977671 | | 4.296968 | -0.05245 | 0.976389 |
| CHMP7 | 4.327842 | -0.41107 | 0.999532 | | 5.592705 | 0.043322 | 0.977671 | | 4.075445 | 0.472515 | 0.647218 |
| MIEF1 | 4.524321 | -0.1276 | 0.999532 | | 5.135394 | -0.04665 | 0.977671 | | 4.309102 | 0.097891 | 0.948127 |
| KIAA2013 | 11.82565 | -0.47559 | 0.999532 | | 15.2945 | 0.027769 | 0.977671 | | 11.24985 | 0.517947 | 0.368714 |
| RTCB | 4.068783 | 0.317506 | 0.999532 | | 4.195535 | 0.048961 | 0.977671 | | 3.930052 | -0.24862 | 0.848153 |
| TWSG1 | 5.79399 | 0.256558 | 0.999532 | | 6.019323 | -0.04502 | 0.977671 | | 5.299096 | -0.28048 | 0.79686 |
| WDR81 | 4.135905 | 0.068563 | 0.999532 | | 4.102557 | -0.05111 | 0.977671 | | 3.721788 | -0.11297 | 0.946512 |
| OR5AS1 | 2.034086 | 0.097465 | 0.999532 | | 2.567463 | 0.068523 | 0.977671 | | 2.112112 | 0.021466 | 0.996681 |
| TTLL9 | 2.007348 | 0.241839 | 0.999532 | | 2.099638 | -0.06801 | 0.977671 | | 1.975635 | -0.28542 | 0.881689 |
| EREG | 2.792087 | 0.579235 | 0.999532 | | 2.836531 | 0.062345 | 0.977671 | | 2.834544 | -0.49575 | 0.709825 |
| CDKL1 | 4.500924 | 0.001606 | 0.999532 | | 4.74555 | -0.04557 | 0.977671 | | 4.204076 | -0.02921 | 0.989646 |
| RTN4RL1 | 2.114029 | 0.673961 | 0.999532 | | 1.683713 | 0.072731 | 0.977698 | | 2.086936 | -0.58149 | 0.717724 |
| PATL1 | 6.105158 | -0.41139 | 0.999532 | | 7.841851 | 0.038914 | 0.977698 | | 6.088691 | 0.471182 | 0.598589 |
| USP40 | 2.321968 | -0.2603 | 0.999532 | | 2.683286 | -0.0573 | 0.977698 | | 2.152711 | 0.217827 | 0.901813 |
| ECHDC1 | 4.645459 | -0.21289 | 0.999532 | | 5.615866 | -0.04289 | 0.977698 | | 4.200487 | 0.187152 | 0.881837 |
| MFHAS1 | 3.980801 | 0.001762 | 0.999532 | | 4.445759 | -0.04976 | 0.977698 | | 3.75395 | -0.03355 | 0.988853 |
| ZNF544 | 3.410615 | 0.04408 | 0.999532 | | 3.714465 | 0.050634 | 0.977698 | | 3.277886 | 0.022856 | 0.993334 |
| AGPAT4 | 2.088688 | 0.410867 | 0.999532 | | 2.014075 | 0.06716 | 0.977698 | | 2.014397 | -0.32693 | 0.853549 |
| CD200R1 | 2.737662 | 0.179011 | 0.999532 | | 2.837113 | -0.06491 | 0.977698 | | 2.520209 | -0.22271 | 0.901033 |
| B3GNT6 | 2.998707 | 0.303806 | 0.999532 | | 3.258693 | 0.059219 | 0.977698 | | 2.989384 | -0.222 | 0.891051 |
| SPATA5L | 2.955711 | 0.836238 | 0.999532 | | 2.430173 | 0.060659 | 0.977804 | | 2.925392 | -0.75678 | 0.507168 |
| ATP1B2 | 3.78435 | -0.38456 | 0.999532 | | 5.08184 | 0.049147 | 0.977804 | | 3.76362 | 0.456272 | 0.720106 |
| SFTPA2 | 1.747011 | -0.32852 | 0.999532 | | 2.345626 | 0.070444 | 0.977949 | | 1.767215 | 0.418447 | 0.831749 |
| VWCE | 3.088222 | 0.051972 | 0.999532 | | 3.303813 | -0.05576 | 0.977966 | | 2.878493 | -0.09312 | 0.963492 |
| STEAP3 | 2.184504 | 0.255951 | 0.999532 | | 2.466064 | 0.061621 | 0.978074 | | 2.252802 | -0.1678 | 0.929529 |
| ZG16B | 2.599289 | 0.262559 | 0.999532 | | 2.822319 | -0.06361 | 0.978152 | | 2.534476 | -0.30023 | 0.86257 |
| ACOT13 | 3.524988 | 0.14968 | 0.999532 | | 3.826654 | 0.056392 | 0.978159 | | 3.262379 | -0.05572 | 0.97843 |
| ITIH6 | 1.913639 | 0.1454 | 0.999532 | | 2.041632 | -0.0673 | 0.978228 | | 1.888346 | -0.1937 | 0.92697 |
| ZNF414 | 3.589332 | 0.618852 | 0.999532 | | 3.096275 | -0.05625 | 0.978228 | | 3.491399 | -0.66206 | 0.531732 |
| GFRA2 | 2.074718 | -0.04112 | 0.999532 | | 2.500899 | 0.0615 | 0.978435 | | 2.135446 | 0.123244 | 0.955834 |
| CNEP1R1 | 2.362983 | 0.544758 | 0.999532 | | 2.144349 | 0.06464 | 0.978435 | | 2.239041 | -0.46198 | 0.748224 |
| SLC1A3 | 2.894021 | 0.326201 | 0.999532 | | 2.987629 | 0.057981 | 0.978435 | | 2.841402 | -0.24794 | 0.87893 |
| PRPF38B | 5.362906 | -0.16572 | 0.999532 | | 6.53854 | 0.040323 | 0.978435 | | 5.31654 | 0.223398 | 0.837256 |
| RBM22 | 3.97553 | -0.33678 | 0.999532 | | 4.856275 | -0.04393 | 0.978435 | | 3.758651 | 0.30976 | 0.79686 |
| CCNT2 | 1.97571 | -0.24222 | 0.999532 | | 2.214146 | 0.062124 | 0.978435 | | 1.924612 | 0.318162 | 0.868512 |
| HSPE1 | 13.92735 | 0.278344 | 0.999532 | | 13.95171 | 0.029711 | 0.978435 | | 13.27005 | -0.2389 | 0.742864 |
| PYGL | 3.867027 | 0.015288 | 0.999532 | | 4.858981 | 0.051841 | 0.978435 | | 4.108601 | 0.069318 | 0.972758 |
| DSG4 | 2.668153 | 0.248691 | 0.999532 | | 2.53603 | -0.05994 | 0.978529 | | 2.576085 | -0.29161 | 0.862823 |
| CCDC136 | 2.286176 | 0.189515 | 0.999532 | | 2.298491 | 0.059828 | 0.978529 | | 2.159207 | -0.11926 | 0.955834 |
| CSMD1 | 2.233199 | 0.127769 | 0.999532 | | 2.409552 | -0.06619 | 0.978529 | | 2.222514 | -0.17769 | 0.938429 |
| PCNX4 | 3.803772 | 0.001945 | 0.999532 | | 4.314333 | 0.047992 | 0.978529 | | 3.50866 | 0.065059 | 0.972221 |
| MYORG | 2.586127 | -0.14243 | 0.999532 | | 2.838599 | -0.05854 | 0.978529 | | 2.430597 | 0.094047 | 0.969783 |
| ALDH8A1 | 2.149249 | 0.678563 | 0.999532 | | 1.910649 | -0.07134 | 0.978529 | | 2.052437 | -0.73117 | 0.6215 |
| CRTAC1 | 3.911551 | 0.162943 | 0.999532 | | 4.033511 | -0.0525 | 0.978529 | | 3.553118 | -0.18491 | 0.901033 |
| MRPL2 | 4.009635 | 0.21134 | 0.999532 | | 4.260856 | -0.05181 | 0.978529 | | 3.872406 | -0.24483 | 0.862432 |
| LRFN1 | 3.229588 | 0.24518 | 0.999532 | | 3.189187 | -0.05777 | 0.978529 | | 3.131384 | -0.2878 | 0.861737 |
| RDH16 | 2.369454 | -0.29672 | 0.999532 | | 3.061019 | -0.06029 | 0.978529 | | 2.326525 | 0.270263 | 0.885078 |
| KLHL7 | 2.241304 | -0.65579 | 0.999532 | | 2.914348 | -0.05539 | 0.978529 | | 2.107314 | 0.615675 | 0.674041 |
| SYNGR4 | 3.284301 | 0.468113 | 0.999532 | | 2.99999 | -0.0587 | 0.978529 | | 3.310449 | -0.51411 | 0.725827 |
| POLE | 2.334646 | 0.183065 | 0.999532 | | 2.347128 | -0.06165 | 0.978529 | | 2.171131 | -0.22729 | 0.900848 |
| TMUB2 | 3.7263 | 0.481599 | 0.999532 | | 3.410351 | -0.0519 | 0.978529 | | 3.499967 | -0.516 | 0.651427 |
| OCA2 | 2.042399 | 0.346573 | 0.999532 | | 1.941637 | 0.067559 | 0.978529 | | 2.026723 | -0.26093 | 0.892869 |
| NUDT22 | 2.821031 | -0.11387 | 0.999532 | | 3.385691 | 0.053579 | 0.978529 | | 2.680401 | 0.187818 | 0.909074 |
| TAF1A | 1.893793 | 0.070883 | 0.999532 | | 2.049196 | -0.06789 | 0.978529 | | 1.911089 | -0.12355 | 0.960773 |
| CA8 | 2.25251 | -0.26637 | 0.999532 | | 2.82361 | -0.05843 | 0.978529 | | 2.236877 | 0.228257 | 0.901033 |
| CD14 | 4.210385 | 0.033585 | 0.999532 | | 4.825018 | 0.04677 | 0.978529 | | 4.118181 | 0.031203 | 0.989278 |
| DGUOK | 3.406359 | 0.00427 | 0.999532 | | 3.687013 | 0.052461 | 0.978529 | | 3.27958 | 0.059485 | 0.977401 |
| FGFR3 | 3.374683 | 0.478597 | 0.999532 | | 3.027189 | -0.05323 | 0.978529 | | 3.310234 | -0.51737 | 0.660097 |
| PISD | 3.715985 | -0.07791 | 0.999532 | | 4.381565 | -0.04777 | 0.978529 | | 3.398168 | 0.056906 | 0.977473 |
| PRPS1 | 4.605738 | 0.224349 | 0.999532 | | 4.826532 | 0.043709 | 0.978529 | | 4.347678 | -0.16341 | 0.901033 |
| SLC35E3 | 2.318627 | 0.172274 | 0.999532 | | 2.365061 | -0.06387 | 0.978529 | | 2.34333 | -0.22372 | 0.905859 |
| ALPK3 | 2.502758 | 0.129085 | 0.999532 | | 2.688098 | -0.05729 | 0.978529 | | 2.452504 | -0.168 | 0.926957 |
| AHI1 | 4.210292 | -0.1926 | 0.999532 | | 5.029459 | 0.045739 | 0.978533 | | 3.995575 | 0.260542 | 0.837615 |
| SH2D3A | 2.339363 | 0.067425 | 0.999532 | | 2.74416 | -0.06306 | 0.978533 | | 2.217675 | -0.09937 | 0.967 |
| TIMP3 | 52.83882 | -0.43454 | 0.893099 | | 69.35272 | 0.024699 | 0.978533 | | 50.92364 | 0.498263 | 0.397004 |
| POLR2J | 5.745035 | 0.371936 | 0.999532 | | 5.483195 | 0.042202 | 0.97856 | | 5.336788 | -0.31314 | 0.753597 |
| MBD3L2 | 4.118464 | -0.11095 | 0.999532 | | 4.680624 | -0.05168 | 0.978845 | | 4.032809 | 0.060904 | 0.977183 |
| CDR2L | 5.01509 | 0.36808 | 0.999532 | | 5.087427 | 0.043877 | 0.978845 | | 4.967794 | -0.3045 | 0.783825 |
| MPEG1 | 4.478067 | -0.23689 | 0.999532 | | 5.21511 | 0.0516 | 0.978845 | | 4.281054 | 0.269773 | 0.86306 |
| RBM42 | 3.897397 | 0.043166 | 0.999532 | | 4.343435 | -0.04608 | 0.978845 | | 3.575336 | -0.07153 | 0.969412 |
| A1BG | 2.510557 | 0.633495 | 0.999532 | | 2.010622 | -0.06597 | 0.978845 | | 2.266913 | -0.69001 | 0.644433 |
| MYLK4 | 3.454779 | -0.20096 | 0.999532 | | 4.230017 | -0.0466 | 0.978845 | | 3.221889 | 0.175792 | 0.906811 |
| PSMG3 | 4.79973 | -0.17491 | 0.999532 | | 5.821969 | 0.042717 | 0.978845 | | 4.653738 | 0.239489 | 0.849608 |
| BUB1 | 2.066279 | -0.15769 | 0.999532 | | 2.249953 | -0.06486 | 0.978845 | | 1.977115 | 0.106502 | 0.969074 |
| ZNF491 | 2.004623 | 0.177285 | 0.999532 | | 2.100728 | 0.06859 | 0.978845 | | 1.945729 | -0.09039 | 0.973356 |
| ZC3HC1 | 2.980074 | -0.22286 | 0.999532 | | 3.21633 | -0.05398 | 0.97887 | | 2.781939 | 0.184001 | 0.915533 |
| MAPK8 | 4.181986 | -0.2034 | 0.999532 | | 4.921832 | 0.04717 | 0.978895 | | 4.049697 | 0.278088 | 0.832386 |
| CCDC91 | 6.594874 | 0.350285 | 0.999532 | | 6.634057 | 0.044401 | 0.978895 | | 6.305992 | -0.27688 | 0.791466 |
| C15orf61 | 4.381992 | -0.00239 | 0.999532 | | 4.962007 | -0.04802 | 0.978895 | | 4.271892 | -0.02585 | 0.992034 |
| HMG20A | 5.016397 | -0.09672 | 0.999532 | | 5.829324 | 0.041126 | 0.978895 | | 4.840456 | 0.154564 | 0.904886 |
| UCP2 | 3.140648 | -0.44606 | 0.999532 | | 3.949096 | 0.049311 | 0.978895 | | 3.125358 | 0.498777 | 0.699971 |
| HDGFL1 | 4.225955 | 0.539884 | 0.999532 | | 3.859544 | 0.049502 | 0.978895 | | 4.235981 | -0.47501 | 0.666492 |
| ASGR2 | 2.346478 | 0.684179 | 0.999532 | | 2.036602 | -0.06776 | 0.978895 | | 2.20228 | -0.73326 | 0.606305 |
| SHISA4 | 2.983206 | 0.586456 | 0.999532 | | 2.6865 | -0.05697 | 0.978895 | | 2.839511 | -0.62455 | 0.602151 |
| RASSF2 | 2.09699 | 0.114831 | 0.999532 | | 2.396415 | 0.061298 | 0.978895 | | 2.169154 | -0.03622 | 0.991159 |
| SUZ12 | 3.271429 | -0.21806 | 0.999532 | | 3.889227 | -0.04637 | 0.978895 | | 3.105044 | 0.191095 | 0.896369 |
| CELF3 | 1.945412 | -0.36339 | 0.999532 | | 2.353689 | -0.06343 | 0.978895 | | 1.823318 | 0.315925 | 0.876781 |
| HNRNPA | 40.55195 | 0.068072 | 0.999532 | | 45.29097 | 0.02657 | 0.978895 | | 39.06043 | -0.00474 | 0.996891 |
| GIMAP6 | 3.286119 | -0.38239 | 0.999532 | | 4.163039 | 0.046265 | 0.979069 | | 3.234496 | 0.444759 | 0.720526 |
| CEP164 | 3.546602 | -0.07845 | 0.999532 | | 3.897851 | 0.049953 | 0.979084 | | 3.175423 | 0.160922 | 0.915888 |
| GXYLT1 | 2.763272 | 0.079664 | 0.999532 | | 3.163086 | 0.052692 | 0.979118 | | 2.679213 | -0.00855 | 0.997084 |
| ASB1 | 3.171303 | 0.282651 | 0.999532 | | 3.114633 | -0.05489 | 0.979126 | | 3.081011 | -0.32565 | 0.821463 |
| EXOSC6 | 3.947235 | -0.34136 | 0.999532 | | 4.983696 | -0.04305 | 0.979134 | | 3.849034 | 0.318784 | 0.794815 |
| PPP2R5B | 3.872826 | 0.046424 | 0.999532 | | 4.235203 | 0.045906 | 0.979134 | | 3.802214 | 0.020043 | 0.994192 |
| BIRC2 | 4.577146 | -0.34273 | 0.999532 | | 5.485448 | -0.04321 | 0.979155 | | 4.366421 | 0.313374 | 0.801905 |
| DOK3 | 2.47108 | 0.190872 | 0.999532 | | 2.457616 | -0.06023 | 0.979155 | | 2.461092 | -0.24098 | 0.90016 |
| KRT72 | 1.967116 | -0.43192 | 0.999532 | | 2.29778 | -0.05952 | 0.979162 | | 1.814889 | 0.385588 | 0.836012 |
| TAAR2 | 2.500755 | 0.425516 | 0.999532 | | 2.357271 | -0.06406 | 0.979241 | | 2.43753 | -0.48121 | 0.750371 |
| CLDN15 | 2.740642 | 0.161932 | 0.999532 | | 2.885482 | 0.057058 | 0.979241 | | 2.620509 | -0.09397 | 0.965649 |
| CHAMP1 | 1.989613 | 0.563239 | 0.999532 | | 1.760642 | 0.067413 | 0.979241 | | 1.983776 | -0.47637 | 0.77094 |
| GP2 | 2.972556 | 0.393552 | 0.999532 | | 2.929018 | 0.056169 | 0.979241 | | 2.798811 | -0.31777 | 0.833604 |
| NXPH1 | 2.272431 | 0.728079 | 0.999532 | | 1.960887 | 0.065261 | 0.979241 | | 2.313153 | -0.65155 | 0.645492 |
| KPNA4 | 2.273436 | 0.093577 | 0.999532 | | 2.52581 | -0.06007 | 0.979241 | | 2.171882 | -0.13722 | 0.946512 |
| KCNIP4 | 2.147375 | 0.364406 | 0.999532 | | 2.084183 | -0.06766 | 0.979416 | | 2.229983 | -0.41297 | 0.816618 |
| INKA1 | 2.40715 | 0.218319 | 0.999532 | | 2.551387 | 0.059339 | 0.97944 | | 2.544979 | -0.14001 | 0.945902 |
| DCDC2C | 1.663794 | 0.200775 | 0.999532 | | 1.832809 | 0.067585 | 0.979997 | | 1.705691 | -0.11273 | 0.965649 |
| APPL2 | 2.719712 | -0.21598 | 0.999532 | | 3.100955 | 0.053437 | 0.980048 | | 2.647543 | 0.307919 | 0.8529 |
| HAUS7 | 5.105609 | 0.42915 | 0.999532 | | 4.939758 | -0.04422 | 0.980152 | | 4.780893 | -0.45261 | 0.647218 |
| FPGS | 3.466555 | 0.202926 | 0.999532 | | 3.666239 | 0.048243 | 0.980152 | | 3.389777 | -0.13727 | 0.928522 |
| VPS13C | 4.651809 | -0.14466 | 0.999532 | | 5.345645 | -0.04123 | 0.980152 | | 4.314215 | 0.122116 | 0.931155 |
| ANKRA2 | 3.72163 | 0.398596 | 0.999532 | | 3.770887 | 0.051719 | 0.980152 | | 3.631805 | -0.32753 | 0.79686 |
| CDC42SE | 3.424516 | -0.14531 | 0.999532 | | 4.013864 | 0.04468 | 0.980152 | | 3.405279 | 0.21117 | 0.881837 |
| CYP4F11 | 2.404362 | 0.374356 | 0.999532 | | 2.486483 | -0.06116 | 0.980152 | | 2.284588 | -0.41863 | 0.783005 |
| PLD1 | 2.396581 | -0.05114 | 0.999532 | | 3.00468 | -0.05681 | 0.980232 | | 2.580549 | 0.005614 | 0.99858 |
| RIPK2 | 2.40034 | 0.240115 | 0.999532 | | 2.5411 | -0.05646 | 0.980296 | | 2.242683 | -0.2753 | 0.871069 |
| FOXN3 | 8.012162 | -0.11866 | 0.999532 | | 9.359925 | -0.03539 | 0.980326 | | 7.628685 | 0.10367 | 0.929604 |
| KDM4D | 2.354404 | 0.504552 | 0.999532 | | 2.249636 | 0.061262 | 0.980326 | | 2.426303 | -0.4243 | 0.791466 |
| DNAJC30 | 3.354922 | -0.06846 | 0.999532 | | 3.922414 | 0.045453 | 0.980426 | | 3.281837 | 0.13323 | 0.934665 |
| CNTLN | 2.580843 | 0.111357 | 0.999532 | | 2.642019 | -0.05681 | 0.980426 | | 2.400235 | -0.15403 | 0.939759 |
| FUZ | 3.61132 | 0.111443 | 0.999532 | | 3.88049 | 0.047848 | 0.980426 | | 3.410034 | -0.04551 | 0.983281 |
| INPP4A | 3.159513 | -0.55656 | 0.999532 | | 4.202616 | -0.04763 | 0.980426 | | 2.934922 | 0.529792 | 0.679818 |
| DPF2 | 4.090289 | 0.021011 | 0.999532 | | 4.611113 | 0.047189 | 0.980426 | | 3.861209 | 0.049192 | 0.978723 |
| U2AF2 | 10.639 | 0.020291 | 0.999532 | | 11.23034 | -0.03374 | 0.980426 | | 9.775876 | -0.05509 | 0.972031 |
| TTC34 | 3.777067 | 0.974029 | 0.999532 | | 2.959675 | -0.05802 | 0.980426 | | 3.567487 | -1.01179 | 0.297366 |
| MYH14 | 2.951507 | 0.104801 | 0.999532 | | 3.073098 | -0.05052 | 0.980426 | | 2.7623 | -0.14289 | 0.93603 |
| ABCC6 | 3.081605 | 0.373614 | 0.999532 | | 3.160117 | -0.05151 | 0.980444 | | 2.856591 | -0.39827 | 0.760473 |
| NHLRC4 | 2.672556 | 0.26261 | 0.999532 | | 2.632017 | -0.05523 | 0.980444 | | 2.619651 | -0.30162 | 0.861119 |
| TYW5 | 2.26414 | -0.02072 | 0.999532 | | 2.607762 | 0.056686 | 0.980444 | | 2.219753 | 0.095638 | 0.967273 |
| MYOM2 | 3.008245 | 0.510399 | 0.999532 | | 2.605516 | -0.05645 | 0.980444 | | 2.793565 | -0.55178 | 0.682165 |
| THAP5 | 4.195708 | -0.08963 | 0.999532 | | 4.310875 | -0.04572 | 0.980444 | | 3.91668 | 0.082493 | 0.965649 |
| UTP23 | 2.309959 | -0.08583 | 0.999532 | | 2.338878 | -0.05704 | 0.980444 | | 2.130617 | 0.02768 | 0.994192 |
| OR4X1 | 2.409414 | 0.011843 | 0.999532 | | 2.926866 | -0.05707 | 0.980444 | | 2.380637 | -0.04941 | 0.985693 |
| AP1B1 | 2.66553 | -0.30766 | 0.999532 | | 3.336604 | 0.04838 | 0.980444 | | 2.634574 | 0.372318 | 0.79686 |
| TOP1 | 7.105913 | -0.24766 | 0.999532 | | 8.616672 | 0.033537 | 0.980444 | | 6.706131 | 0.298062 | 0.749588 |
| FOLR1 | 3.674646 | 0.24065 | 0.999532 | | 3.869607 | -0.04865 | 0.980444 | | 3.370932 | -0.26488 | 0.850539 |
| EBF4 | 2.620338 | 0.195336 | 0.999532 | | 2.720721 | 0.054988 | 0.980444 | | 2.505185 | -0.12406 | 0.949097 |
| GMFB | 4.578771 | -0.39609 | 0.999532 | | 5.587754 | -0.03889 | 0.980444 | | 4.167284 | 0.372838 | 0.730921 |
| CA10 | 3.953524 | 0.406878 | 0.999532 | | 4.209357 | -0.04814 | 0.980444 | | 3.951855 | -0.42448 | 0.718643 |
| SRP72 | 4.48224 | 0.132845 | 0.999532 | | 4.790051 | 0.040761 | 0.980444 | | 4.201158 | -0.07383 | 0.964251 |
| STUM | 2.613533 | 0.199544 | 0.999532 | | 2.593267 | -0.05783 | 0.980444 | | 2.574738 | -0.23733 | 0.898856 |
| PDAP1 | 10.30966 | -0.1345 | 0.999532 | | 12.58936 | 0.034711 | 0.980444 | | 10.20342 | 0.18911 | 0.852482 |
| SRARP | 1.926754 | 0.452882 | 0.999532 | | 1.834817 | -0.0643 | 0.980444 | | 1.798746 | -0.49875 | 0.757037 |
| FAP | 2.504861 | -0.07765 | 0.999532 | | 2.902291 | 0.054982 | 0.980444 | | 2.299282 | 0.152213 | 0.938429 |
| MALL | 2.995948 | 0.449281 | 0.999532 | | 2.977544 | 0.053567 | 0.980444 | | 3.03106 | -0.37778 | 0.777203 |
| DDHD1 | 1.850951 | -0.2566 | 0.999532 | | 2.198421 | 0.05835 | 0.980444 | | 1.739613 | 0.330154 | 0.863401 |
| KLHL21 | 4.102346 | 0.269501 | 0.999532 | | 3.982397 | -0.04645 | 0.980444 | | 3.744633 | -0.2919 | 0.814402 |
| JAK2 | 3.639573 | 0.398371 | 0.999532 | | 3.261041 | -0.04804 | 0.980444 | | 3.43973 | -0.42884 | 0.717627 |
| IFT46 | 2.651461 | 0.425225 | 0.999532 | | 2.603564 | 0.054599 | 0.980444 | | 2.517193 | -0.35383 | 0.813438 |
| IFT172 | 2.687743 | 0.07905 | 0.999532 | | 3.042509 | 0.051473 | 0.980444 | | 2.714773 | -0.00927 | 0.996891 |
| CCL14 | 2.80332 | 0.355438 | 0.999532 | | 2.792101 | -0.05558 | 0.980444 | | 2.636573 | -0.38868 | 0.791576 |
| RPTOR | 3.138017 | 0.224104 | 0.999532 | | 3.23168 | -0.04852 | 0.980444 | | 2.906295 | -0.25307 | 0.864923 |
| GALNT2 | 10.30948 | 0.286997 | 0.999532 | | 10.35336 | 0.031542 | 0.980444 | | 9.933068 | -0.23991 | 0.779082 |
| ADRA1D | 2.139142 | 0.333855 | 0.999532 | | 2.063582 | -0.06228 | 0.980444 | | 1.957663 | -0.37616 | 0.825181 |
| MFAP3 | 3.01003 | -0.2363 | 0.999532 | | 3.445444 | -0.05116 | 0.980444 | | 2.769665 | 0.204713 | 0.903054 |
| BARHL2 | 4.517258 | -0.34211 | 0.999532 | | 5.500451 | -0.04399 | 0.980444 | | 4.237194 | 0.311545 | 0.809498 |
| CENPBD1 | 2.087472 | -0.12493 | 0.999532 | | 2.527051 | -0.0568 | 0.980444 | | 2.108716 | 0.088358 | 0.972221 |
| ZNF646 | 1.837628 | -0.22455 | 0.999532 | | 2.222121 | 0.06129 | 0.980444 | | 1.831422 | 0.302345 | 0.87813 |
| GLE1 | 2.967301 | 0.139867 | 0.999532 | | 3.042822 | -0.05446 | 0.980444 | | 2.849983 | -0.17156 | 0.925487 |
| MRPL50 | 3.718117 | -0.18276 | 0.999532 | | 4.234112 | 0.043574 | 0.980444 | | 3.429566 | 0.236791 | 0.86595 |
| KCNC3 | 5.412539 | -0.07341 | 0.999532 | | 6.100277 | -0.03916 | 0.980444 | | 4.904051 | 0.036035 | 0.987608 |
| AKAP1 | 5.059354 | 0.228824 | 0.999532 | | 5.066561 | -0.04267 | 0.980444 | | 4.529081 | -0.2466 | 0.835554 |
| APOL2 | 5.324649 | 0.408169 | 0.999532 | | 5.241755 | 0.039328 | 0.980444 | | 5.233235 | -0.35089 | 0.717627 |
| MKRN1 | 5.264918 | 0.294083 | 0.999532 | | 5.398592 | 0.042548 | 0.980598 | | 5.135672 | -0.23343 | 0.867009 |
| SVOP | 2.419455 | 0.23953 | 0.999532 | | 2.518722 | 0.055577 | 0.980675 | | 2.399408 | -0.16896 | 0.92697 |
| MAGEB1 | 1.7154 | 0.248588 | 0.999532 | | 1.943555 | 0.065768 | 0.980852 | | 1.74632 | -0.15976 | 0.945456 |
| SEPHS2 | 4.342147 | -0.48823 | 0.999532 | | 5.522787 | 0.040288 | 0.980894 | | 4.06208 | 0.541438 | 0.625955 |
| CFAP97 | 2.725618 | -0.15967 | 0.999532 | | 3.092514 | 0.048753 | 0.980894 | | 2.575584 | 0.225391 | 0.88885 |
| ZDHHC24 | 2.236852 | 0.709863 | 0.999532 | | 1.801352 | 0.063484 | 0.980894 | | 2.177148 | -0.63022 | 0.68528 |
| ABRAXAS | 2.71943 | -0.28346 | 0.999532 | | 3.275882 | 0.049564 | 0.980913 | | 2.626496 | 0.346831 | 0.823231 |
| FOXK1 | 4.382342 | 0.330903 | 0.999532 | | 4.106336 | 0.045654 | 0.98098 | | 4.02481 | -0.27787 | 0.829716 |
| R3HDM1 | 2.502333 | 0.04358 | 0.999532 | | 2.61033 | -0.05368 | 0.98098 | | 2.221523 | -0.07494 | 0.974421 |
| DDX39B | 19.96859 | -0.06772 | 0.999532 | | 22.9344 | 0.02403 | 0.98098 | | 18.92708 | 0.107732 | 0.901033 |
| ERCC3 | 3.458606 | -0.11614 | 0.999532 | | 4.193564 | 0.044836 | 0.98098 | | 3.398591 | 0.177722 | 0.903339 |
| CCDC88A | 4.252151 | -0.0859 | 0.999532 | | 4.760766 | -0.04206 | 0.98098 | | 4.207399 | 0.063241 | 0.974421 |
| ATG9B | 2.395772 | -0.03931 | 0.999532 | | 2.696238 | -0.05368 | 0.981061 | | 2.304753 | 0.001487 | 0.999463 |
| SPATA18 | 2.841765 | -0.1395 | 0.999532 | | 3.374271 | -0.05255 | 0.981061 | | 2.65556 | 0.118656 | 0.954304 |
| GSG1L | 3.066863 | 0.392173 | 0.999532 | | 3.074575 | 0.058445 | 0.981155 | | 3.036382 | -0.33431 | 0.835554 |
| ZNF527 | 2.001065 | 0.065881 | 0.999532 | | 2.243889 | -0.06037 | 0.981155 | | 1.91018 | -0.1041 | 0.966652 |
| TADA1 | 2.570088 | -0.0221 | 0.999532 | | 2.676331 | -0.05164 | 0.981162 | | 2.358435 | -0.01581 | 0.996681 |
| MAD1L1 | 2.355398 | -0.62461 | 0.999532 | | 3.13809 | -0.04846 | 0.981162 | | 2.211852 | 0.593902 | 0.667066 |
| CHURC1 | 6.511403 | -0.02871 | 0.999532 | | 6.974578 | -0.03469 | 0.981162 | | 5.96649 | 0.006793 | 0.996891 |
| PKD2 | 4.366268 | -0.10508 | 0.999532 | | 5.304837 | 0.039684 | 0.981203 | | 4.256765 | 0.166956 | 0.901033 |
| EBAG9 | 3.353594 | 0.561848 | 0.999532 | | 2.990011 | 0.049956 | 0.981203 | | 3.10075 | -0.49388 | 0.682165 |
| DGKI | 1.853146 | 0.195853 | 0.999532 | | 1.981172 | -0.06095 | 0.981203 | | 1.843842 | -0.23897 | 0.903339 |
| EDC3 | 3.476426 | 0.437754 | 0.999532 | | 3.296873 | -0.04718 | 0.981203 | | 3.288317 | -0.46666 | 0.69872 |
| C15orf48 | 3.358678 | -0.26047 | 0.999532 | | 4.058388 | -0.04475 | 0.981203 | | 3.228202 | 0.234527 | 0.885892 |
| SRSF4 | 5.127264 | -0.1346 | 0.999532 | | 5.788866 | -0.03733 | 0.981203 | | 4.843831 | 0.11025 | 0.939929 |
| U2SURP | 4.144155 | 0.048644 | 0.999532 | | 4.612059 | 0.041415 | 0.981203 | | 4.067661 | 0.012208 | 0.996681 |
| ETS2 | 11.64775 | -0.24447 | 0.999532 | | 14.33586 | 0.029264 | 0.981203 | | 11.39733 | 0.298239 | 0.701803 |
| CTC1 | 3.265115 | 0.361912 | 0.999532 | | 3.233908 | 0.049534 | 0.981203 | | 3.016817 | -0.29042 | 0.837256 |
| CAP2 | 2.123218 | -0.03009 | 0.999532 | | 2.48026 | 0.055775 | 0.981203 | | 2.101859 | 0.102486 | 0.964728 |
| DENND2D | 1.932794 | 0.454866 | 0.999532 | | 1.895784 | 0.060269 | 0.981203 | | 1.877802 | -0.37671 | 0.8233 |
| SLC9A1 | 3.675737 | -0.05144 | 0.999532 | | 4.254356 | 0.043909 | 0.981203 | | 3.519711 | 0.112675 | 0.945129 |
| DPP9 | 1.77272 | -0.08678 | 0.999532 | | 1.896607 | 0.059337 | 0.981203 | | 1.699649 | 0.159395 | 0.945902 |
| SSX1 | 2.008896 | 0.220544 | 0.999532 | | 2.218048 | 0.061981 | 0.981203 | | 2.133551 | -0.1198 | 0.96556 |
| CALCRL | 3.557319 | -0.22789 | 0.999532 | | 4.184538 | -0.04361 | 0.981203 | | 3.314365 | 0.196534 | 0.897136 |
| ELMOD3 | 2.947112 | -0.00601 | 0.999532 | | 3.453993 | 0.04675 | 0.981203 | | 3.017131 | 0.073908 | 0.972221 |
| DHRS9 | 1.721093 | -0.16878 | 0.999532 | | 2.011012 | 0.059049 | 0.981203 | | 1.695328 | 0.244002 | 0.905952 |
| TNIP2 | 5.661574 | -0.28301 | 0.999532 | | 6.946524 | 0.034431 | 0.981203 | | 5.538998 | 0.338395 | 0.722703 |
| CRKL | 9.490709 | -0.18355 | 0.999532 | | 10.91508 | -0.02885 | 0.981203 | | 8.887554 | 0.180354 | 0.842686 |
| VPS33B | 2.734106 | -0.28479 | 0.999532 | | 3.323544 | -0.05179 | 0.981203 | | 2.576552 | 0.250586 | 0.883565 |
| DMWD | 9.095606 | -0.17771 | 0.999532 | | 10.80037 | -0.02759 | 0.981203 | | 8.456775 | 0.167635 | 0.859583 |
| LCE2C | 2.195456 | 0.360508 | 0.999532 | | 2.054737 | -0.05951 | 0.981203 | | 2.133826 | -0.40288 | 0.815713 |
| ZNF57 | 2.020683 | 0.492252 | 0.999532 | | 1.918574 | -0.06275 | 0.981203 | | 1.925873 | -0.53712 | 0.735469 |
| GLUL | 9.1604 | -0.07299 | 0.999532 | | 10.70132 | 0.042725 | 0.981203 | | 8.684438 | 0.167872 | 0.883116 |
| NCOA1 | 5.067816 | -0.3211 | 0.999532 | | 6.27252 | -0.03649 | 0.981203 | | 4.709097 | 0.30243 | 0.779804 |
| RAD50 | 3.11084 | 0.169583 | 0.999532 | | 3.474836 | 0.047871 | 0.981203 | | 2.988667 | -0.10244 | 0.955834 |
| CP | 1.89664 | 0.268185 | 0.999532 | | 1.906203 | -0.06141 | 0.981203 | | 1.741701 | -0.31213 | 0.871957 |
| UPK3BL1 | 19.59968 | -0.06196 | 0.999532 | | 22.2508 | -0.03024 | 0.981203 | | 18.92222 | 0.039086 | 0.973356 |
| COMMD2 | 2.872949 | 0.157101 | 0.999532 | | 3.117791 | -0.04796 | 0.981203 | | 2.858956 | -0.18577 | 0.905952 |
| PSG2 | 3.870411 | 0.260828 | 0.999532 | | 3.848253 | -0.04482 | 0.981203 | | 3.714978 | -0.28938 | 0.836453 |
| ZNF581 | 3.919739 | 0.350346 | 0.999532 | | 3.776882 | -0.05009 | 0.981203 | | 3.534815 | -0.38522 | 0.759211 |
| MKI67 | 1.631235 | -0.27411 | 0.999532 | | 1.862903 | -0.06075 | 0.981203 | | 1.510428 | 0.226098 | 0.920259 |
| PRKCH | 3.607449 | -0.24888 | 0.999532 | | 4.262273 | -0.04149 | 0.981291 | | 3.416188 | 0.223841 | 0.876781 |
| RSF1 | 13.18666 | 0.186368 | 0.999532 | | 13.72354 | -0.03025 | 0.981291 | | 13.06342 | -0.20508 | 0.834627 |
| CPSF2 | 3.239746 | 0.217007 | 0.999532 | | 3.12419 | 0.048338 | 0.981291 | | 2.993345 | -0.15415 | 0.927882 |
| C17orf97 | 3.268462 | 0.003873 | 0.999532 | | 3.596592 | -0.04633 | 0.981291 | | 3.042968 | -0.029 | 0.991273 |
| NAP1L2 | 3.726071 | 0.480047 | 0.999532 | | 3.264353 | -0.04861 | 0.981346 | | 3.495527 | -0.50697 | 0.672333 |
| ADCY2 | 2.081417 | 0.573877 | 0.999532 | | 1.940169 | 0.060254 | 0.981527 | | 2.03923 | -0.4961 | 0.747677 |
| ADGRB1 | 2.304525 | -0.50332 | 0.999532 | | 2.950215 | -0.04958 | 0.981581 | | 2.249135 | 0.470523 | 0.765012 |
| HCK | 4.046679 | -0.02371 | 0.999532 | | 4.155392 | -0.04574 | 0.981581 | | 3.70738 | -0.0222 | 0.993618 |
| GSTM3 | 2.115678 | 0.157756 | 0.999532 | | 2.365712 | 0.055337 | 0.981581 | | 1.998653 | -0.08473 | 0.972221 |
| TMEM63 | 3.296172 | 0.04869 | 0.999532 | | 3.751453 | 0.044035 | 0.981581 | | 3.23223 | 0.013465 | 0.996681 |
| PLCB4 | 2.434616 | 0.558026 | 0.999532 | | 2.340712 | -0.05644 | 0.981581 | | 2.305668 | -0.59494 | 0.663009 |
| TSHZ3 | 2.890003 | 0.734358 | 0.999532 | | 2.259895 | -0.05436 | 0.981581 | | 2.751124 | -0.77714 | 0.515762 |
| ELMO3 | 2.423652 | 0.182012 | 0.999532 | | 2.595237 | -0.05544 | 0.981581 | | 2.234982 | -0.21936 | 0.904476 |
| UNG | 3.989702 | -0.3443 | 0.999532 | | 4.801991 | 0.039673 | 0.981581 | | 3.847207 | 0.408936 | 0.72472 |
| RAB7B | 2.323551 | -0.06161 | 0.999532 | | 2.628865 | 0.051054 | 0.981581 | | 2.283462 | 0.130623 | 0.948127 |
| MYO10 | 3.161432 | -0.11528 | 0.999532 | | 3.585509 | -0.0446 | 0.981581 | | 3.063202 | 0.084943 | 0.965619 |
| LNX1 | 2.487456 | -0.19002 | 0.999532 | | 2.996549 | 0.049758 | 0.981581 | | 2.364001 | 0.259582 | 0.881837 |
| SCO1 | 4.633868 | 0.147429 | 0.999532 | | 5.093976 | 0.038576 | 0.981581 | | 4.516076 | -0.08257 | 0.960282 |
| SEMA4G | 3.308447 | 0.199036 | 0.999532 | | 3.434615 | 0.045676 | 0.981581 | | 3.26698 | -0.13718 | 0.934597 |
| FAM98B | 3.001848 | -0.31584 | 0.999532 | | 4.007808 | 0.042182 | 0.981581 | | 3.083535 | 0.380461 | 0.765852 |
| PSMA4 | 4.909777 | -0.14053 | 0.999532 | | 5.508747 | -0.04008 | 0.981581 | | 4.52443 | 0.116146 | 0.939628 |
| ADAM19 | 3.026096 | -0.05499 | 0.999532 | | 3.484666 | -0.04603 | 0.981581 | | 2.929712 | 0.030838 | 0.991273 |
| MROH7 | 1.896363 | 0.110043 | 0.999532 | | 1.944693 | -0.06138 | 0.981581 | | 1.812238 | -0.15745 | 0.946572 |
| TMPRSS | 2.360544 | 0.300478 | 0.999532 | | 2.209194 | -0.05693 | 0.981581 | | 2.318973 | -0.3427 | 0.847148 |
| VPS13B | 5.957618 | 0.246594 | 0.999532 | | 5.930946 | 0.036134 | 0.981581 | | 5.386262 | -0.20228 | 0.864612 |
| CRACDL | 2.039344 | -0.28932 | 0.999532 | | 2.45877 | 0.054372 | 0.981581 | | 1.974229 | 0.360327 | 0.845164 |
| KCNAB2 | 3.591363 | -0.1663 | 0.999532 | | 4.132584 | 0.042886 | 0.981581 | | 3.392052 | 0.224171 | 0.880949 |
| TMEM101 | 3.800937 | 0.566106 | 0.999532 | | 3.513123 | -0.04513 | 0.981615 | | 3.525809 | -0.5872 | 0.568257 |
| HLF | 2.848198 | -0.14032 | 0.999532 | | 3.365782 | -0.05005 | 0.981643 | | 2.691024 | 0.114056 | 0.956678 |
| PHGDH | 3.201289 | 0.35384 | 0.999532 | | 3.248461 | 0.049446 | 0.981729 | | 2.953793 | -0.27965 | 0.851503 |
| CREB3 | 5.11916 | 0.036509 | 0.999532 | | 5.564764 | -0.03738 | 0.981729 | | 4.814782 | -0.0545 | 0.974421 |
| EPC1 | 6.939399 | 0.098647 | 0.999532 | | 7.674695 | -0.03397 | 0.981794 | | 6.555157 | -0.1197 | 0.914501 |
| SRRM3 | 2.470468 | -0.08668 | 0.999532 | | 2.650657 | -0.05077 | 0.981807 | | 2.369452 | 0.044555 | 0.988853 |
| MARVELD | 2.243279 | 0.302066 | 0.999532 | | 2.387812 | 0.055327 | 0.981807 | | 2.294289 | -0.22633 | 0.901813 |
| MMP11 | 2.070264 | 0.309385 | 0.999532 | | 1.973754 | -0.05833 | 0.981807 | | 1.905202 | -0.35021 | 0.845164 |
| MACROH | 5.857262 | -0.08421 | 0.999532 | | 6.782762 | 0.035298 | 0.98183 | | 5.667365 | 0.135288 | 0.914774 |
| ARSH | 2.335824 | 0.693158 | 0.999532 | | 2.102801 | -0.05944 | 0.98183 | | 2.159776 | -0.72432 | 0.599054 |
| ANKDD1 | 2.232006 | -0.19596 | 0.999532 | | 2.583413 | -0.05193 | 0.98183 | | 2.090177 | 0.161743 | 0.938429 |
| FUT2 | 2.28628 | 0.665423 | 0.999532 | | 1.964906 | -0.06035 | 0.981994 | | 2.174881 | -0.70754 | 0.630705 |
| GSDMD | 4.81051 | 0.346606 | 0.999532 | | 4.345376 | -0.04164 | 0.98204 | | 4.440757 | -0.36563 | 0.75311 |
| LGI2 | 3.100896 | 0.025363 | 0.999532 | | 3.281996 | -0.04729 | 0.98204 | | 3.038562 | -0.03925 | 0.988853 |
| R3HCC1 | 3.164358 | -0.6606 | 0.999532 | | 4.466251 | 0.040555 | 0.98204 | | 3.007052 | 0.723162 | 0.530776 |
| LPIN2 | 4.235423 | -0.21951 | 0.999532 | | 5.379262 | 0.039824 | 0.98204 | | 4.111728 | 0.277153 | 0.823193 |
| SLC39A5 | 2.76606 | -0.29822 | 0.999532 | | 3.375262 | 0.048525 | 0.98204 | | 2.60805 | 0.360289 | 0.827267 |
| GAR1 | 2.979423 | -0.43642 | 0.999532 | | 3.946784 | 0.042645 | 0.98204 | | 2.951082 | 0.497277 | 0.699971 |
| BTC | 2.903698 | 0.436527 | 0.999532 | | 2.613135 | -0.05064 | 0.982144 | | 2.880213 | -0.47092 | 0.735774 |
| FAM117A | 2.864187 | -0.14221 | 0.999532 | | 3.080114 | -0.04547 | 0.982288 | | 2.694213 | 0.118682 | 0.952656 |
| RRM2B | 4.05353 | 0.232029 | 0.999532 | | 4.07935 | 0.042726 | 0.982369 | | 3.934388 | -0.15259 | 0.920103 |
| MYCT1 | 2.607927 | -0.11398 | 0.999532 | | 2.995683 | 0.0467 | 0.982369 | | 2.461672 | 0.177387 | 0.918024 |
| PCP2 | 2.982812 | 0.690199 | 0.999532 | | 2.401552 | 0.052883 | 0.982369 | | 2.746057 | -0.6132 | 0.647218 |
| KATNBL1 | 2.145112 | 0.255143 | 0.999532 | | 2.075984 | -0.05615 | 0.982369 | | 1.977248 | -0.2943 | 0.87591 |
| MTMR11 | 3.28851 | -0.11583 | 0.999532 | | 3.904213 | -0.04241 | 0.982369 | | 3.171996 | 0.090956 | 0.960705 |
| STARD8 | 5.23511 | -0.27806 | 0.999532 | | 6.360473 | 0.035883 | 0.982686 | | 4.880225 | 0.333911 | 0.750417 |
| RERE | 11.18131 | -0.03696 | 0.999532 | | 12.83077 | 0.024806 | 0.982865 | | 10.77609 | 0.083351 | 0.942911 |
| JADE3 | 2.227619 | 0.101471 | 0.999532 | | 2.507845 | 0.049323 | 0.98301 | | 2.253633 | -0.03218 | 0.991783 |
| MAP3K13 | 5.504934 | 0.427633 | 0.999532 | | 5.352874 | 0.037462 | 0.98301 | | 5.54684 | -0.3697 | 0.725408 |
| PCDH9 | 2.763031 | 0.32334 | 0.999532 | | 2.871055 | 0.047496 | 0.98301 | | 2.708857 | -0.25775 | 0.868465 |
| MAN1A2 | 2.286941 | 0.642106 | 0.999532 | | 1.882071 | -0.05621 | 0.98301 | | 2.117759 | -0.68739 | 0.632113 |
| LARGE1 | 6.280431 | 0.040326 | 0.999532 | | 6.765232 | -0.03859 | 0.98301 | | 5.79433 | -0.04604 | 0.978004 |
| POM121C | 8.657064 | -0.09924 | 0.999532 | | 10.19126 | -0.03036 | 0.98301 | | 8.285325 | 0.093789 | 0.938429 |
| UGT2B15 | 2.380403 | 0.371379 | 0.999532 | | 2.546233 | 0.055883 | 0.98301 | | 2.525037 | -0.30734 | 0.863808 |
| ST20 | 4.69496 | 0.302357 | 0.999532 | | 4.670737 | -0.03993 | 0.98301 | | 4.493889 | -0.32197 | 0.791466 |
| SLC4A5 | 2.972885 | 0.415401 | 0.999532 | | 3.025111 | -0.0488 | 0.983027 | | 3.003304 | -0.43946 | 0.749999 |
| RPUSD3 | 2.692659 | 0.010474 | 0.999532 | | 3.134605 | -0.04481 | 0.983031 | | 2.587502 | -0.03367 | 0.990309 |
| PACSIN3 | 2.566369 | 0.414369 | 0.999532 | | 2.545125 | 0.053805 | 0.983031 | | 2.460131 | -0.33755 | 0.834627 |
| FGF21 | 2.674432 | -0.24435 | 0.999532 | | 3.102275 | -0.04852 | 0.983031 | | 2.526578 | 0.211398 | 0.906811 |
| VWDE | 1.838845 | 0.110081 | 0.999532 | | 1.864022 | -0.0587 | 0.983082 | | 1.751463 | -0.15638 | 0.947126 |
| NAGLU | 3.231147 | -0.18119 | 0.999532 | | 4.195824 | 0.038787 | 0.983206 | | 3.188228 | 0.234952 | 0.865363 |
| LIN7A | 2.065875 | 0.401695 | 0.999532 | | 2.087593 | 0.056103 | 0.983206 | | 2.101481 | -0.33007 | 0.859522 |
| EPG5 | 4.566962 | 0.176513 | 0.999532 | | 4.54783 | -0.03724 | 0.983206 | | 4.162566 | -0.19755 | 0.877703 |
| MT1HL1 | 14.37179 | 0.011548 | 0.999532 | | 15.96318 | 0.033006 | 0.983206 | | 13.6834 | 0.03679 | 0.981739 |
| EXOSC5 | 2.193927 | 0.126957 | 0.999532 | | 2.330616 | -0.05137 | 0.983206 | | 2.059679 | -0.15913 | 0.939107 |
| IFI30 | 11.00439 | -0.18176 | 0.999532 | | 12.5595 | -0.04622 | 0.983206 | | 10.31222 | 0.154098 | 0.914929 |
| TGFBI | 5.317146 | -0.45853 | 0.999532 | | 6.702349 | -0.04797 | 0.983206 | | 5.067812 | 0.426881 | 0.71172 |
| SOWAHC | 2.166181 | -0.39355 | 0.999532 | | 2.631337 | -0.05048 | 0.983206 | | 2.042735 | 0.360606 | 0.842754 |
| HS2ST1 | 2.97383 | -0.0953 | 0.999532 | | 3.673691 | 0.042371 | 0.983206 | | 2.856145 | 0.163946 | 0.920655 |
| CCR7 | 2.706744 | 0.28065 | 0.999532 | | 2.960457 | -0.05931 | 0.983206 | | 2.59597 | -0.33515 | 0.843564 |
| MYBPH | 3.635814 | -0.3445 | 0.999532 | | 4.426866 | -0.04023 | 0.983206 | | 3.40207 | 0.323796 | 0.808457 |
| ZNF212 | 3.097367 | -0.05161 | 0.999532 | | 3.36028 | -0.04197 | 0.983206 | | 3.036131 | 0.028305 | 0.991783 |
| IGF2BP2 | 4.112384 | 0.066937 | 0.999532 | | 4.962972 | 0.038981 | 0.983206 | | 4.165362 | -0.00783 | 0.996891 |
| PSG11 | 4.385807 | 0.26919 | 0.999532 | | 4.248329 | -0.04244 | 0.983206 | | 4.036685 | -0.28533 | 0.82268 |
| PSAT1 | 5.436857 | 0.531445 | 0.999532 | | 4.685773 | -0.03901 | 0.983206 | | 5.068647 | -0.561 | 0.552788 |
| KRTAP13 | 2.146014 | 0.376845 | 0.999532 | | 1.997704 | -0.05667 | 0.983206 | | 2.02163 | -0.4188 | 0.810276 |
| KDM8 | 2.195826 | -0.05772 | 0.999532 | | 2.607275 | -0.05166 | 0.983206 | | 2.145704 | 0.027626 | 0.993618 |
| ZNF793 | 3.978987 | 0.319589 | 0.999532 | | 4.030095 | -0.04107 | 0.983377 | | 3.761318 | -0.34029 | 0.786052 |
| PKNOX1 | 4.599324 | -0.13696 | 0.999532 | | 5.588312 | -0.03817 | 0.983379 | | 4.372027 | 0.138052 | 0.930403 |

| | | | | | | | | | | | |
|---|---|---|---|---|---|---|---|---|---|---|---|
| C4BPB | 2.500986 | 0.314363 | 0.999532 | | 2.508673 | -0.05022 | 0.983604 | | 2.335025 | -0.34374 | 0.833604 |
| REG1A | 1.897401 | 0.332601 | 0.999532 | | 1.796615 | -0.0585 | 0.983721 | | 1.853111 | -0.37157 | 0.846155 |
| VRK1 | 1.979582 | 0.326767 | 0.999532 | | 1.957312 | -0.05436 | 0.98383 | | 1.818543 | -0.36507 | 0.836453 |
| TMEM203 | 3.378091 | 0.271462 | 0.999532 | | 3.456914 | -0.04308 | 0.983859 | | 3.377165 | -0.29696 | 0.825181 |
| CHST7 | 2.414585 | 0.291009 | 0.999532 | | 2.365158 | -0.05136 | 0.98388 | | 2.187095 | -0.32252 | 0.847164 |
| PELO | 7.216185 | 0.20631 | 0.999532 | | 7.264883 | 0.02979 | 0.983898 | | 6.901163 | -0.17233 | 0.878619 |
| ST6GALN | 2.821636 | 0.052939 | 0.999532 | | 2.924414 | -0.04396 | 0.983898 | | 2.755109 | -0.07372 | 0.974331 |
| ZNF891 | 3.410004 | 0.142964 | 0.999532 | | 3.536519 | -0.04318 | 0.983898 | | 3.231996 | -0.16629 | 0.917753 |
| ATF6 | 4.732421 | -0.41141 | 0.999532 | | 6.10108 | -0.03655 | 0.983898 | | 4.328643 | 0.411946 | 0.717627 |
| DYM | 4.773676 | -0.07013 | 0.999532 | | 5.430052 | -0.03379 | 0.983898 | | 4.391169 | 0.051698 | 0.976389 |
| ZNF839 | 2.587313 | 0.080425 | 0.999532 | | 2.711226 | -0.04467 | 0.983898 | | 2.451772 | -0.10904 | 0.957624 |
| SCRN1 | 3.007796 | -0.28431 | 0.999532 | | 3.795828 | -0.04153 | 0.983898 | | 2.87661 | 0.267658 | 0.86639 |
| RAB27A | 2.405559 | -0.52898 | 0.999532 | | 3.004141 | 0.043889 | 0.983898 | | 2.292951 | 0.583195 | 0.679452 |
| DAO | 2.782767 | -0.22304 | 0.999532 | | 3.129347 | -0.04868 | 0.983898 | | 2.478385 | 0.198628 | 0.914929 |
| CHIC1 | 3.051399 | -0.10423 | 0.999532 | | 3.700486 | -0.04176 | 0.983898 | | 2.979407 | 0.092398 | 0.964971 |
| PAFAH1B | 7.259549 | 0.038287 | 0.999532 | | 7.784738 | 0.029192 | 0.983898 | | 6.819131 | -0.00636 | 0.996891 |
| PTX4 | 2.154749 | 0.572613 | 0.999532 | | 2.075939 | -0.05505 | 0.983898 | | 2.152894 | -0.60711 | 0.678411 |
| PFKP | 3.872855 | 0.052775 | 0.999532 | | 4.167602 | 0.042356 | 0.983898 | | 3.646037 | 0.024152 | 0.992286 |
| ACE | 3.359036 | 0.202449 | 0.999532 | | 3.650205 | 0.043422 | 0.983898 | | 3.120199 | -0.13779 | 0.934701 |
| EDNRB | 5.105609 | 0.087065 | 0.999532 | | 5.306902 | -0.0368 | 0.983898 | | 4.774818 | -0.12051 | 0.936911 |
| PPY | 2.11728 | 0.295373 | 0.999532 | | 2.090074 | -0.05354 | 0.983898 | | 1.965379 | -0.33095 | 0.859898 |
| PRKD2 | 3.449699 | 0.146749 | 0.999532 | | 3.455394 | 0.040102 | 0.983898 | | 3.229681 | -0.10005 | 0.955834 |
| OAZ2 | 7.61651 | 0.081137 | 0.999532 | | 8.193981 | 0.030549 | 0.983898 | | 7.330589 | -0.02898 | 0.986714 |
| TAF10 | 4.510454 | -0.40083 | 0.999532 | | 5.925582 | -0.03351 | 0.983898 | | 4.187971 | 0.384308 | 0.721843 |
| BABAM1 | 6.173396 | 0.290441 | 0.999532 | | 6.267555 | 0.034259 | 0.983898 | | 5.87416 | -0.23583 | 0.824012 |
| NTF3 | 2.456038 | -0.06468 | 0.999532 | | 2.928368 | 0.046146 | 0.983898 | | 2.546145 | 0.136795 | 0.946512 |
| KHDC4 | 4.197378 | 0.312754 | 0.999532 | | 4.288939 | 0.037259 | 0.983898 | | 3.946951 | -0.25511 | 0.836453 |
| AGPS | 4.786856 | -0.15487 | 0.999532 | | 5.471461 | -0.03407 | 0.983898 | | 4.570032 | 0.142373 | 0.912963 |
| CREB3L4 | 2.636723 | 0.519055 | 0.999532 | | 2.544812 | 0.049258 | 0.983898 | | 2.573992 | -0.45082 | 0.747457 |
| TAGLN3 | 2.718343 | -0.03279 | 0.999532 | | 3.018426 | 0.0427 | 0.983898 | | 2.608439 | 0.088403 | 0.96588 |
| NDUFB11 | 3.928873 | -0.02965 | 0.999532 | | 4.357549 | -0.03763 | 0.983898 | | 3.758495 | 0.025897 | 0.992286 |
| CAMK1 | 3.93858 | 0.100439 | 0.999532 | | 4.328737 | 0.038336 | 0.983898 | | 3.778776 | -0.043 | 0.983211 |
| FAM214B | 2.660255 | -0.41276 | 0.999532 | | 3.190551 | -0.04367 | 0.983898 | | 2.549325 | 0.383522 | 0.805466 |
| UBQLN2 | 5.251241 | 0.49953 | 0.999532 | | 4.852936 | 0.036029 | 0.983898 | | 5.056351 | -0.44537 | 0.641721 |
| TARS2 | 2.624236 | 0.076423 | 0.999532 | | 2.840866 | -0.04627 | 0.983898 | | 2.39781 | -0.10741 | 0.958889 |
| PMP22 | 2.765466 | 0.418555 | 0.999532 | | 2.738839 | 0.046376 | 0.983898 | | 2.671722 | -0.35436 | 0.81294 |
| THAP7 | 4.281957 | 0.479486 | 0.999532 | | 3.965028 | -0.03952 | 0.983898 | | 4.012131 | -0.50058 | 0.644433 |
| LSM14B | 4.445975 | 0.095338 | 0.999532 | | 4.807989 | 0.036755 | 0.983898 | | 4.222446 | -0.03829 | 0.98558 |
| CSF2RA | 2.247979 | 0.308079 | 0.999532 | | 2.465222 | -0.05443 | 0.983898 | | 2.275507 | -0.32575 | 0.864245 |
| RWDD1 | 5.272745 | -0.29946 | 0.999532 | | 6.376931 | -0.03223 | 0.983898 | | 5.157971 | 0.284717 | 0.806797 |
| SLC30A6 | 3.039046 | 0.49683 | 0.999532 | | 2.693641 | -0.04857 | 0.983898 | | 2.733859 | -0.52045 | 0.678411 |
| TXNDC11 | 3.680733 | 0.039081 | 0.999532 | | 4.011658 | 0.040712 | 0.983898 | | 3.41651 | 0.02957 | 0.991159 |
| MAP2K4 | 3.965435 | 0.02277 | 0.999532 | | 4.418209 | 0.037747 | 0.983898 | | 3.82626 | 0.03262 | 0.988853 |
| EPB41L3 | 2.314874 | -0.21888 | 0.999532 | | 2.612844 | -0.0479 | 0.983898 | | 2.144003 | 0.186694 | 0.923834 |
| POLR3GL | 3.328529 | -0.19729 | 0.999532 | | 3.902938 | 0.039404 | 0.983898 | | 3.256465 | 0.256465 | 0.858767 |
| TJP2 | 3.020016 | 0.188005 | 0.999532 | | 3.281813 | 0.045714 | 0.983898 | | 3.122793 | -0.12919 | 0.945129 |
| PHACTR4 | 7.366295 | 0.174784 | 0.999532 | | 7.470269 | 0.03402 | 0.983898 | | 6.815617 | -0.11804 | 0.927544 |
| STAP2 | 1.924582 | 0.38596 | 0.999532 | | 1.962007 | 0.054892 | 0.983898 | | 1.823824 | -0.3105 | 0.866612 |
| MOV10 | 3.848648 | 0.076578 | 0.999532 | | 4.352901 | 0.038692 | 0.983898 | | 3.812891 | -0.02004 | 0.993618 |
| NFX1 | 3.295572 | 0.313567 | 0.999532 | | 3.312586 | -0.0421 | 0.983898 | | 3.045677 | -0.33965 | 0.792688 |
| TSN | 5.663537 | 0.210511 | 0.999532 | | 6.051507 | 0.035475 | 0.983898 | | 5.246065 | -0.15619 | 0.907568 |
| KIFC1 | 1.731092 | 0.075573 | 0.999532 | | 1.917548 | -0.05583 | 0.983898 | | 1.661745 | -0.11149 | 0.966652 |
| PALM2AK | 6.58479 | -0.09498 | 0.999532 | | 7.24342 | -0.03133 | 0.983898 | | 6.312436 | 0.085466 | 0.946765 |
| HNRNPCL | 2.590449 | 0.483331 | 0.999532 | | 2.57013 | -0.04751 | 0.983898 | | 2.503451 | -0.50924 | 0.705888 |
| RHOT1 | 2.864403 | 0.32702 | 0.999532 | | 2.771085 | -0.04609 | 0.983898 | | 2.657152 | -0.35587 | 0.811123 |
| SCYL1 | 6.791492 | -0.13149 | 0.999532 | | 8.447899 | -0.03017 | 0.983898 | | 6.535177 | 0.157151 | 0.895246 |
| ACAD8 | 4.47231 | 0.156443 | 0.999532 | | 4.563909 | 0.037938 | 0.983898 | | 4.22831 | -0.08381 | 0.959333 |
| CLP1 | 2.433606 | 0.127791 | 0.999532 | | 2.633519 | 0.048854 | 0.983898 | | 2.468449 | -0.06199 | 0.979282 |
| COX17 | 7.559237 | -0.13595 | 0.999532 | | 8.567854 | -0.02792 | 0.983898 | | 7.04125 | 0.120761 | 0.917499 |
| MYD88 | 3.413865 | 0.164845 | 0.999532 | | 3.467215 | -0.04151 | 0.983898 | | 3.085934 | -0.18402 | 0.903329 |
| SFI1 | 3.950932 | -0.26135 | 0.999532 | | 4.730427 | -0.03543 | 0.983898 | | 3.698085 | 0.243788 | 0.861815 |
| LOXL3 | 2.20136 | 0.093513 | 0.999532 | | 2.208372 | -0.05062 | 0.983898 | | 1.968874 | -0.12839 | 0.953948 |
| TNFRSF1 | 3.394571 | 0.241028 | 0.999532 | | 3.411476 | 0.039438 | 0.983898 | | 3.26756 | -0.18459 | 0.900583 |
| SLCO2A1 | 11.88731 | -0.53811 | 0.999532 | | 15.64327 | -0.02206 | 0.983898 | | 11.20733 | 0.533159 | 0.389782 |
| IL10RA | 4.649259 | -0.19045 | 0.999532 | | 5.638107 | 0.039132 | 0.983898 | | 4.450951 | 0.244629 | 0.874653 |
| TRIM25 | 5.476696 | 0.098499 | 0.999532 | | 5.680452 | -0.03308 | 0.983898 | | 5.182486 | -0.11265 | 0.930352 |
| SELENOI | 15.8777 | -0.09766 | 0.999532 | | 18.12679 | -0.02669 | 0.983898 | | 15.19285 | 0.090539 | 0.934354 |
| MAP3K7 | 5.998421 | 0.024294 | 0.999532 | | 6.593067 | 0.030026 | 0.983898 | | 5.668767 | 0.020283 | 0.992286 |
| IL22RA2 | 2.35675 | 0.581865 | 0.999532 | | 2.15555 | -0.05098 | 0.983898 | | 2.305911 | -0.61498 | 0.653492 |
| RCSD1 | 2.140422 | 0.002153 | 0.999532 | | 2.543717 | 0.050386 | 0.983898 | | 2.273057 | 0.064263 | 0.978927 |
| ATP5MC1 | 4.817504 | -0.10343 | 0.999532 | | 5.887115 | 0.035534 | 0.983898 | | 4.560672 | 0.146554 | 0.907968 |
| SLBP | 3.288128 | -0.40478 | 0.999532 | | 4.33134 | -0.03897 | 0.98395 | | 3.280751 | 0.378596 | 0.769058 |
| OCRL | 2.161711 | 0.366705 | 0.999532 | | 2.128836 | -0.04949 | 0.98395 | | 2.105107 | -0.39704 | 0.803337 |
| PIR | 2.386831 | -0.02207 | 0.999532 | | 2.605221 | 0.045365 | 0.98395 | | 2.371517 | 0.089022 | 0.970498 |
| BCL2L14 | 3.701484 | 0.311757 | 0.999532 | | 3.791189 | -0.04233 | 0.98395 | | 3.499616 | -0.3224 | 0.827267 |
| PIK3R3 | 6.410451 | -0.5429 | 0.999532 | | 8.484374 | -0.03216 | 0.98395 | | 5.948267 | 0.52667 | 0.531575 |
| TBC1D13 | 6.172392 | 0.108108 | 0.999532 | | 6.573072 | 0.031677 | 0.98413 | | 5.892071 | -0.05008 | 0.974421 |
| AJAP1 | 3.583848 | -0.46613 | 0.999532 | | 4.471183 | 0.037133 | 0.98413 | | 3.3013 | 0.515725 | 0.687954 |
| TBC1D16 | 3.414457 | -0.24677 | 0.999532 | | 4.437332 | 0.037032 | 0.98417 | | 3.386265 | 0.300643 | 0.82512 |
| CCER2 | 2.366275 | 0.435869 | 0.999532 | | 2.337044 | 0.049482 | 0.98417 | | 2.421981 | -0.36794 | 0.819175 |
| NUGGC | 2.523589 | 0.064189 | 0.999532 | | 2.874761 | -0.04634 | 0.984199 | | 2.458362 | -0.0846 | 0.972221 |
| SNUPN | 3.658945 | 0.481793 | 0.999532 | | 3.460945 | -0.03887 | 0.984199 | | 3.524077 | -0.49625 | 0.660958 |
| RNF26 | 4.251354 | 0.364028 | 0.999532 | | 4.076392 | -0.03687 | 0.984286 | | 3.996593 | -0.38342 | 0.735469 |
| CNOT10 | 2.445151 | 0.406324 | 0.999532 | | 2.038096 | -0.04918 | 0.984317 | | 2.240448 | -0.45962 | 0.764702 |
| ZNF311 | 2.087634 | 0.282113 | 0.999532 | | 2.08112 | -0.05177 | 0.984317 | | 1.929354 | -0.31537 | 0.865065 |
| CD58 | 3.775939 | -0.0581 | 0.999532 | | 4.202397 | -0.03578 | 0.984391 | | 3.626935 | 0.038506 | 0.986208 |
| CES1 | 2.499163 | 0.156314 | 0.999532 | | 2.699659 | -0.04807 | 0.984391 | | 2.451207 | -0.19095 | 0.918421 |
| HHEX | 2.710743 | 0.367826 | 0.999532 | | 2.698353 | 0.044032 | 0.984479 | | 2.618853 | -0.30591 | 0.839373 |
| ATF5 | 3.878371 | 0.367129 | 0.999532 | | 3.582259 | 0.039909 | 0.984542 | | 3.626632 | -0.31362 | 0.809832 |
| CSTF3 | 8.382218 | 0.078565 | 0.999532 | | 9.056786 | 0.027372 | 0.984542 | | 7.85633 | -0.03344 | 0.982781 |
| RSPO1 | 2.085747 | -0.16258 | 0.999532 | | 2.443591 | 0.047226 | 0.984637 | | 2.145989 | 0.230136 | 0.905952 |
| SENP2 | 3.557782 | -0.09935 | 0.999532 | | 4.154574 | 0.037581 | 0.984637 | | 3.44003 | 0.154584 | 0.923864 |
| CALCOCO | 6.209046 | -0.74798 | 0.999532 | | 8.414626 | -0.02783 | 0.984637 | | 5.602711 | 0.736054 | 0.304062 |
| UBE2I | 6.490755 | -0.10873 | 0.999532 | | 7.682087 | 0.028035 | 0.984637 | | 6.161485 | 0.147651 | 0.893828 |
| BCL2L1 | 10.99242 | -0.23354 | 0.999532 | | 12.82546 | 0.02459 | 0.984867 | | 10.32746 | 0.266846 | 0.748224 |
| L3MBTL3 | 2.195182 | 0.200547 | 0.999532 | | 2.398985 | -0.04776 | 0.984867 | | 2.124905 | -0.22835 | 0.900848 |
| CDH24 | 2.98181 | -0.19701 | 0.999532 | | 3.482713 | 0.042004 | 0.984883 | | 2.931529 | 0.253962 | 0.877703 |
| SPIRE2 | 2.061453 | 0.631845 | 0.999532 | | 1.779924 | -0.05292 | 0.984883 | | 1.952147 | -0.66834 | 0.652925 |
| AGA | 2.42736 | 0.188081 | 0.999532 | | 2.542333 | -0.0467 | 0.984883 | | 2.194863 | -0.2168 | 0.90332 |
| RBM38 | 4.370613 | -0.1006 | 0.999532 | | 5.223151 | 0.034867 | 0.984883 | | 4.156583 | 0.156353 | 0.909074 |
| PVALB | 2.001957 | -0.02089 | 0.999532 | | 2.224144 | -0.05101 | 0.984883 | | 1.875251 | -0.01132 | 0.996891 |
| EPYC | 2.331525 | 0.362551 | 0.999532 | | 2.319708 | 0.047026 | 0.984883 | | 2.301506 | -0.29701 | 0.862823 |
| VARS1 | 3.499181 | -0.1419 | 0.999532 | | 3.850805 | -0.03807 | 0.9849 | | 3.260811 | 0.116809 | 0.950372 |
| KLF15 | 2.817422 | 0.161656 | 0.999532 | | 3.086479 | 0.048659 | 0.9849 | | 2.590568 | -0.08153 | 0.972031 |
| TRIM13 | 2.360569 | 0.181529 | 0.999532 | | 2.451175 | -0.04595 | 0.98503 | | 2.282476 | -0.20881 | 0.907106 |
| SLC1A4 | 2.651581 | 0.230857 | 0.999532 | | 2.662801 | -0.04423 | 0.98503 | | 2.522734 | -0.24375 | 0.884754 |
| RRP1 | 3.44789 | 0.210203 | 0.999532 | | 3.406707 | -0.03963 | 0.98503 | | 3.277531 | -0.23665 | 0.875951 |
| NUDCD3 | 6.166522 | -0.03637 | 0.999532 | | 6.696813 | 0.028923 | 0.98503 | | 6.0055 | 0.073276 | 0.96129 |
| ABTB2 | 2.585176 | 0.118735 | 0.999532 | | 2.812869 | 0.0427 | 0.985075 | | 2.635707 | -0.05901 | 0.978927 |
| FBL | 4.952533 | -0.32034 | 0.999532 | | 5.710983 | -0.03209 | 0.985075 | | 4.484341 | 0.295643 | 0.800239 |
| RANBP2 | 21.01827 | -0.17194 | 0.999532 | | 24.32619 | -0.01731 | 0.985075 | | 19.67174 | 0.167764 | 0.795928 |
| PDS5B | 5.254794 | -0.0955 | 0.999532 | | 5.912309 | -0.03281 | 0.985168 | | 4.768027 | 0.089663 | 0.954304 |
| USP5 | 3.064826 | 0.25304 | 0.999532 | | 3.179122 | 0.040876 | 0.985168 | | 3.035038 | -0.19418 | 0.89916 |
| RANBP3 | 4.549017 | -0.54278 | 0.999532 | | 5.987083 | -0.0328 | 0.985168 | | 4.304875 | 0.528557 | 0.625071 |
| FIG4 | 2.744293 | -0.22735 | 0.999532 | | 2.897326 | -0.04131 | 0.985168 | | 2.531168 | 0.195426 | 0.91707 |
| SBSN | 2.565036 | 0.297192 | 0.999532 | | 2.767822 | 0.045765 | 0.985168 | | 2.546498 | -0.23112 | 0.892869 |
| PLXND1 | 13.50438 | -0.43185 | 0.999532 | | 17.25819 | -0.02533 | 0.985168 | | 12.72917 | 0.428314 | 0.638728 |
| BPIFB6 | 2.010571 | 0.455895 | 0.999532 | | 1.788283 | -0.05267 | 0.985168 | | 1.937285 | -0.49161 | 0.773849 |
| C1QTNF1 | 2.098131 | -0.01492 | 0.999532 | | 2.235 | -0.04652 | 0.985168 | | 2.060051 | -0.01149 | 0.996891 |
| ZNF606 | 2.100997 | -0.1064 | 0.999532 | | 2.322757 | 0.045839 | 0.985168 | | 2.06426 | 0.172082 | 0.935818 |
| SLC16A1 | 2.477648 | 0.223061 | 0.999532 | | 2.700005 | -0.0479 | 0.985168 | | 2.523288 | -0.24513 | 0.901033 |
| SPON2 | 7.371726 | -0.38533 | 0.999532 | | 9.590133 | -0.02984 | 0.985168 | | 6.941061 | 0.372497 | 0.705888 |
| XKR4 | 2.085367 | 0.191759 | 0.999532 | | 2.115834 | -0.04798 | 0.985168 | | 1.904199 | -0.2253 | 0.907968 |
| DGKG | 2.186747 | 0.530361 | 0.999532 | | 2.047755 | 0.04907 | 0.985168 | | 2.045921 | -0.46566 | 0.765852 |
| AP1G1 | 3.712363 | 0.294412 | 0.999532 | | 3.778839 | 0.036198 | 0.985168 | | 3.691434 | -0.23903 | 0.857945 |
| LENG9 | 2.961929 | 0.306016 | 0.999532 | | 3.125417 | -0.04195 | 0.985168 | | 2.944876 | -0.33528 | 0.809832 |
| C5orf24 | 4.393228 | -0.25191 | 0.999532 | | 5.293241 | -0.03273 | 0.985168 | | 4.280612 | 0.240855 | 0.845575 |
| ZC4H2 | 2.544411 | 0.259887 | 0.999532 | | 2.518992 | 0.043805 | 0.985168 | | 2.495327 | -0.19925 | 0.907626 |
| FBXO6 | 2.194982 | -0.05542 | 0.999532 | | 2.540729 | 0.045357 | 0.985168 | | 2.180891 | 0.119366 | 0.958191 |
| ESPNL | 1.866803 | 0.196478 | 0.999532 | | 2.037173 | 0.050243 | 0.985168 | | 1.944447 | -0.13071 | 0.955834 |
| C15orf40 | 3.400614 | 0.205739 | 0.999532 | | 3.45347 | 0.038765 | 0.985168 | | 3.202931 | -0.15588 | 0.922437 |
| PPRC1 | 2.478209 | -0.12037 | 0.999532 | | 2.704039 | -0.04374 | 0.985168 | | 2.170488 | 0.089767 | 0.970811 |
| DCTPP1 | 2.437037 | -0.0724 | 0.999532 | | 2.55684 | -0.04492 | 0.985168 | | 2.168748 | 0.046962 | 0.987274 |
| ANP32E | 3.719186 | -0.18057 | 0.999532 | | 4.59199 | -0.03679 | 0.985168 | | 3.490714 | 0.187117 | 0.908595 |
| RBM7 | 4.428136 | -0.57397 | 0.999532 | | 6.070864 | -0.02994 | 0.985168 | | 4.177434 | 0.564423 | 0.553283 |
| ZSCAN21 | 2.455635 | -0.49454 | 0.999532 | | 3.022661 | 0.04024 | 0.985168 | | 2.300923 | 0.546404 | 0.705046 |
| FOXD3 | 2.203761 | 0.623735 | 0.999532 | | 1.996849 | -0.05189 | 0.985474 | | 2.132943 | -0.65761 | 0.660097 |
| LBHD1 | 3.518679 | -0.16885 | 0.999532 | | 4.223108 | -0.03502 | 0.985474 | | 3.279781 | 0.149933 | 0.920716 |
| TRPM5 | 1.886651 | 0.1991 | 0.999532 | | 1.867344 | -0.0495 | 0.985474 | | 1.77732 | -0.23134 | 0.907882 |
| OR13D1 | 1.847365 | 0.479984 | 0.999532 | | 1.779675 | 0.054739 | 0.985474 | | 1.833394 | -0.40603 | 0.828126 |
| TRAF3IP3 | 2.709364 | 0.374013 | 0.999532 | | 2.651124 | 0.042572 | 0.985484 | | 2.771106 | -0.31759 | 0.835478 |
| ARHGAP | 3.391487 | -0.48864 | 0.999532 | | 4.630984 | 0.034116 | 0.985506 | | 3.453545 | 0.539682 | 0.64142 |
| SLC41A1 | 5.712857 | 0.063643 | 0.999532 | | 6.293019 | -0.02921 | 0.985545 | | 5.174252 | -0.07987 | 0.955834 |
| GGT7 | 2.968287 | 0.298786 | 0.999532 | | 3.091285 | -0.04095 | 0.985561 | | 2.746858 | -0.33106 | 0.816589 |
| RAB43 | 4.132099 | 0.075801 | 0.999532 | | 4.597758 | 0.033115 | 0.985694 | | 4.103401 | -0.0239 | 0.992286 |
| CD22 | 2.380328 | -0.08339 | 0.999532 | | 2.706881 | -0.04568 | 0.985771 | | 2.189841 | 0.062746 | 0.978931 |
| WNT8B | 1.827563 | 0.476484 | 0.999532 | | 1.772367 | 0.053872 | 0.985801 | | 1.781894 | -0.404 | 0.82977 |
| PPP3R1 | 5.332899 | -0.59541 | 0.999532 | | 7.242371 | -0.0309 | 0.985801 | | 4.876657 | 0.576495 | 0.515762 |
| SBNO2 | 2.83997 | -0.2295 | 0.999532 | | 3.288834 | -0.04084 | 0.985801 | | 2.645164 | 0.214029 | 0.894617 |
| ERO1B | 2.879347 | 0.456996 | 0.999532 | | 2.950995 | -0.04243 | 0.985801 | | 2.837428 | -0.46292 | 0.736481 |
| IRF2BP2 | 11.19941 | -0.57104 | 0.999532 | | 15.12405 | -0.02221 | 0.985801 | | 10.49014 | 0.57514 | 0.30043 |
| MCF2L | 1.792621 | -0.04725 | 0.999532 | | 2.030377 | 0.046084 | 0.985801 | | 1.700699 | 0.108445 | 0.965649 |
| LIMS2 | 9.806768 | -0.35501 | 0.999532 | | 12.28833 | -0.02674 | 0.985801 | | 9.331909 | 0.344784 | 0.66718 |
| PIK3CD | 4.636202 | 0.020183 | 0.999532 | | 5.337013 | 0.031017 | 0.985801 | | 4.573251 | 0.0316 | 0.989148 |
| ADAT3 | 5.40035 | 0.352228 | 0.999532 | | 5.46988 | -0.03346 | 0.985801 | | 5.250722 | -0.38678 | 0.737952 |
| ETV6 | 3.337345 | -0.08005 | 0.999532 | | 3.863991 | -0.03707 | 0.985801 | | 3.261734 | 0.062977 | 0.974421 |
| ZNF444 | 5.95103 | -0.15507 | 0.999532 | | 7.195437 | 0.028135 | 0.985801 | | 5.815713 | 0.208652 | 0.868998 |
| CDK13 | 5.409859 | -0.0804 | 0.999532 | | 6.06835 | 0.03154 | 0.985801 | | 5.003286 | 0.133991 | 0.920202 |
| OR2M2 | 2.301641 | 0.485409 | 0.999532 | | 2.366638 | 0.046147 | 0.985801 | | 2.278044 | -0.42532 | 0.784955 |
| SIAH1 | 6.768016 | 0.024357 | 0.999532 | | 7.616581 | -0.02834 | 0.985801 | | 6.666412 | -0.03534 | 0.983152 |
| BDKRB2 | 1.878431 | -0.17747 | 0.999532 | | 2.209652 | 0.045231 | 0.985801 | | 1.752613 | 0.233322 | 0.906168 |
| SLC12A1 | 5.463047 | 0.170248 | 0.999532 | | 5.861684 | -0.05228 | 0.985801 | | 5.07957 | -0.17776 | 0.943307 |
| CLBA1 | 2.72965 | 0.169001 | 0.999532 | | 2.859317 | 0.039367 | 0.985801 | | 2.639345 | -0.11282 | 0.955167 |
| HYAL1 | 4.277375 | -0.22602 | 0.999532 | | 5.046765 | -0.03459 | 0.985801 | | 3.910118 | 0.21256 | 0.881837 |
| TMSB15B | 1.817264 | 0.04832 | 0.999532 | | 1.927259 | -0.04833 | 0.985801 | | 1.721751 | -0.07921 | 0.976603 |
| SLC26A2 | 2.120723 | 0.00063 | 0.999532 | | 2.449728 | 0.043274 | 0.985801 | | 2.110042 | 0.062918 | 0.979239 |
| TBC1D2 | 2.991235 | 0.30382 | 0.999532 | | 2.757752 | -0.04103 | 0.985801 | | 2.887505 | -0.33668 | 0.830822 |
| PLD2 | 2.733896 | -0.18153 | 0.999532 | | 3.208098 | 0.037222 | 0.985801 | | 2.531916 | 0.233784 | 0.88476 |
| ZBTB12 | 2.194335 | 0.670237 | 0.999532 | | 1.81246 | 0.050916 | 0.985801 | | 2.098172 | -0.6028 | 0.701962 |
| AXDND1 | 2.516092 | 0.49888 | 0.999532 | | 2.319042 | -0.04678 | 0.985801 | | 2.368737 | -0.52853 | 0.717627 |
| KDM5D | 2.80204 | 0.023435 | 0.999532 | | 3.19818 | 0.062922 | 0.985801 | | 2.733681 | 0.048573 | 0.988853 |
| PACRGL | 3.593688 | 0.479141 | 0.999532 | | 3.449005 | -0.03992 | 0.985801 | | 3.482203 | -0.50108 | 0.679818 |
| DNAAF2 | 2.601634 | 0.09659 | 0.999532 | | 2.649926 | 0.040922 | 0.985801 | | 2.512343 | -0.0339 | 0.991159 |
| DNAL4 | 4.14377 | 0.322526 | 0.999532 | | 4.114497 | 0.034352 | 0.985801 | | 4.113214 | -0.27101 | 0.829158 |
| CDC42BP | 8.72267 | -0.0277 | 0.999532 | | 9.50191 | -0.02314 | 0.985801 | | 8.20311 | 0.020515 | 0.991103 |
| QPCT | 2.626351 | 0.093172 | 0.999532 | | 2.851628 | 0.040579 | 0.985801 | | 2.591675 | -0.03418 | 0.991159 |
| DPH5 | 2.478999 | -0.1853 | 0.999532 | | 3.001835 | -0.03944 | 0.985801 | | 2.503661 | 0.165246 | 0.92683 |
| IRX3 | 2.711361 | 0.260042 | 0.999532 | | 3.068326 | 0.040649 | 0.985801 | | 2.724541 | -0.18401 | 0.914929 |
| PIGM | 3.506589 | 0.147016 | 0.999532 | | 3.865609 | 0.036269 | 0.985801 | | 3.365319 | -0.08678 | 0.963866 |
| GRAMD1A | 4.016964 | -0.3423 | 0.999532 | | 4.961084 | -0.03322 | 0.985801 | | 3.974323 | 0.324192 | 0.806797 |
| SGCE | 5.807064 | -0.13311 | 0.999532 | | 6.467265 | -0.03127 | 0.985801 | | 5.326974 | 0.110682 | 0.940266 |
| UPP1 | 3.14492 | -0.71498 | 0.999532 | | 4.396202 | -0.03415 | 0.985801 | | 3.038525 | 0.699265 | 0.537919 |
| CCND3 | 4.690268 | -0.34841 | 0.999532 | | 5.763496 | 0.030332 | 0.985801 | | 4.611092 | 0.396852 | 0.717627 |
| CCR2 | 1.981762 | -0.24214 | 0.999532 | | 2.275657 | 0.047134 | 0.985801 | | 1.888472 | 0.299131 | 0.881837 |
| RBFA | 3.570926 | 0.080435 | 0.999532 | | 4.044052 | 0.034802 | 0.985801 | | 3.298615 | -0.02472 | 0.992286 |
| NBAS | 3.091522 | 0.329268 | 0.999532 | | 2.972759 | 0.0392 | 0.985801 | | 2.804612 | -0.28011 | 0.85585 |
| ANP32B | 8.802278 | 0.266906 | 0.999532 | | 9.11464 | 0.025036 | 0.985801 | | 8.550582 | -0.22107 | 0.80308 |
| MTREX | 3.731311 | -0.55205 | 0.999532 | | 4.805066 | 0.032879 | 0.985801 | | 3.749984 | 0.58635 | 0.652925 |
| NRARP | 2.973876 | 0.032989 | 0.999532 | | 3.314151 | 0.038436 | 0.985801 | | 2.97932 | 0.022164 | 0.994192 |
| MED26 | 3.362221 | 0.562126 | 0.999532 | | 3.101179 | 0.038527 | 0.985801 | | 3.480762 | -0.51297 | 0.662584 |
| CLDN25 | 2.081664 | 0.010306 | 0.999532 | | 2.246075 | 0.043343 | 0.985801 | | 2.048883 | 0.055287 | 0.984624 |
| CLEC12B | 2.656797 | 0.369055 | 0.999532 | | 2.569158 | -0.04346 | 0.985801 | | 2.647545 | -0.39533 | 0.789885 |
| MTIF3 | 4.956222 | 0.351937 | 0.999532 | | 4.86567 | 0.031757 | 0.985801 | | 4.813886 | -0.30811 | 0.783005 |
| SCAI | 1.941324 | 0.266696 | 0.999532 | | 1.896141 | -0.0486 | 0.985801 | | 1.880592 | -0.30078 | 0.877943 |
| RAB10 | 5.277117 | -0.13153 | 0.999532 | | 6.495867 | 0.02837 | 0.985801 | | 5.226166 | 0.185533 | 0.877703 |
| PLLP | 4.435274 | 0.67436 | 0.999532 | | 3.815854 | -0.03454 | 0.985801 | | 4.331836 | -0.67318 | 0.515762 |
| IFNA5 | 2.560466 | 0.846324 | 0.999532 | | 2.197574 | 0.050178 | 0.985801 | | 2.633778 | -0.80273 | 0.533876 |
| STK38L | 3.772689 | -0.42138 | 0.999532 | | 4.733146 | -0.03148 | 0.98584 | | 3.52149 | 0.406278 | 0.735469 |
| TMEM189 | 2.908793 | 0.287608 | 0.999532 | | 2.987622 | -0.04106 | 0.985882 | | 2.763116 | -0.3092 | 0.843043 |
| TRNP1 | 2.708273 | 0.084623 | 0.999532 | | 2.816673 | -0.04078 | 0.985882 | | 2.543561 | -0.10658 | 0.958422 |
| RWDD3 | 2.680428 | 0.759919 | 0.999532 | | 2.447647 | 0.043047 | 0.985882 | | 2.654096 | -0.69452 | 0.565022 |
| FAM171B | 2.828496 | 0.477553 | 0.999532 | | 2.703448 | -0.04012 | 0.985882 | | 2.676069 | -0.4989 | 0.699971 |
| ZCCHC3 | 3.585162 | -0.36455 | 0.999532 | | 4.360256 | 0.03206 | 0.985882 | | 3.328388 | 0.411708 | 0.733942 |
| ACOT2 | 3.843841 | 0.16624 | 0.999532 | | 3.983601 | 0.035519 | 0.985882 | | 3.668635 | -0.11699 | 0.945857 |
| TAF9B | 3.565049 | 0.241564 | 0.999532 | | 3.727174 | 0.034342 | 0.985882 | | 3.418169 | -0.1894 | 0.896891 |
| ZNF823 | 2.639298 | 0.044155 | 0.999532 | | 2.843662 | -0.04024 | 0.985882 | | 2.522074 | -0.06323 | 0.978229 |
| ANAPC15 | 3.731247 | 0.26815 | 0.999532 | | 3.634768 | -0.03514 | 0.985882 | | 3.582765 | -0.28471 | 0.845164 |
| DLG5 | 5.97827 | -0.14658 | 0.999532 | | 7.137522 | -0.02752 | 0.985882 | | 5.509708 | 0.153666 | 0.901033 |
| KDM3A | 3.262083 | -0.26353 | 0.999532 | | 4.143273 | 0.032764 | 0.985882 | | 3.287182 | 0.318202 | 0.810635 |
| OTUD7A | 2.939674 | -0.04659 | 0.999532 | | 3.492761 | 0.038096 | 0.985882 | | 2.764102 | 0.099969 | 0.958889 |
| AKR1C2 | 2.714411 | -0.14686 | 0.999532 | | 3.354648 | 0.043147 | 0.985949 | | 2.910785 | 0.197118 | 0.925804 |
| EHD1 | 5.141308 | 0.2336 | 0.999532 | | 5.252435 | 0.029099 | 0.985972 | | 4.864416 | -0.18623 | 0.877268 |
| TBC1D32 | 3.026497 | -0.61047 | 0.999532 | | 3.830492 | -0.03566 | 0.985972 | | 2.76067 | 0.582007 | 0.669902 |
| SURF2 | 2.73793 | 0.143738 | 0.999532 | | 3.318979 | -0.03994 | 0.985972 | | 2.817771 | -0.13158 | 0.946765 |
| LRWD1 | 3.53983 | -0.00929 | 0.999532 | | 3.924073 | 0.03277 | 0.985972 | | 3.335806 | 0.056832 | 0.977514 |
| FEN1 | 3.678053 | 0.042946 | 0.999532 | | 4.190598 | -0.03348 | 0.985972 | | 3.471602 | -0.053 | 0.97843 |
| STK10 | 3.380289 | -0.36728 | 0.999532 | | 3.987468 | -0.03429 | 0.985972 | | 3.028144 | 0.340397 | 0.811123 |
| SEMA3F | 3.208675 | -0.0679 | 0.999532 | | 3.703521 | 0.035063 | 0.985972 | | 3.212043 | 0.117246 | 0.948127 |
| IPCEF1 | 2.668483 | 0.56972 | 0.999532 | | 2.309669 | -0.04245 | 0.985972 | | 2.577442 | -0.59609 | 0.660097 |
| AK8 | 3.628034 | 0.105607 | 0.999532 | | 3.923739 | -0.0351 | 0.985972 | | 3.576946 | -0.12881 | 0.939549 |
| CSNK2B | 10.13996 | -0.2096 | 0.999532 | | 11.66524 | 0.021845 | 0.985972 | | 9.585928 | 0.235897 | 0.791576 |
| MED30 | 2.597557 | -0.18074 | 0.999532 | | 3.201044 | -0.03785 | 0.985972 | | 2.567077 | 0.16483 | 0.928522 |
| PATZ1 | 4.038753 | -0.21373 | 0.999532 | | 4.910861 | -0.03192 | 0.985972 | | 3.861345 | 0.201541 | 0.88885 |
| INTS12 | 2.432601 | 0.381655 | 0.999532 | | 2.510099 | 0.041765 | 0.985972 | | 2.408606 | -0.31689 | 0.844187 |
| LAMB4 | 1.948253 | -0.19664 | 0.999532 | | 2.326042 | -0.04323 | 0.985972 | | 1.922976 | 0.174999 | 0.938429 |
| DDX19B | 2.847923 | 0.201493 | 0.999532 | | 2.91634 | -0.03882 | 0.985972 | | 2.752392 | -0.22284 | 0.892869 |
| LRP11 | 5.020202 | 0.234543 | 0.999532 | | 4.938665 | -0.03053 | 0.985972 | | 4.565828 | -0.25144 | 0.835289 |
| STRBP | 3.313483 | 0.228949 | 0.999532 | | 3.28325 | -0.03581 | 0.985972 | | 3.113388 | -0.25002 | 0.864977 |
| FGF22 | 2.452021 | 0.157513 | 0.999532 | | 2.664989 | -0.04066 | 0.985972 | | 2.345769 | -0.17731 | 0.923864 |
| MIA2 | 3.898848 | 0.012629 | 0.999532 | | 4.470279 | -0.03322 | 0.985972 | | 3.632547 | -0.02251 | 0.992586 |
| MAPRE1 | 12.06592 | -0.39473 | 0.999532 | | 15.04108 | 0.019806 | 0.985972 | | 11.54579 | 0.432193 | 0.515762 |
| PEG10 | 2.122198 | 0.081051 | 0.999532 | | 2.448853 | 0.045104 | 0.985972 | | 2.173959 | -0.02073 | 0.996681 |
| FAM174A | 5.918012 | 0.18941 | 0.999532 | | 6.080427 | -0.02769 | 0.985972 | | 5.557938 | -0.1989 | 0.857377 |
| PSME2 | 6.958876 | -0.15304 | 0.999532 | | 8.266119 | 0.026406 | 0.985972 | | 6.675204 | 0.197648 | 0.859522 |
| GRK5 | 4.48736 | 0.211281 | 0.999532 | | 4.596898 | -0.03364 | 0.985972 | | 4.1453 | -0.22598 | 0.864923 |
| NOL12 | 3.984817 | 0.200676 | 0.999532 | | 4.238881 | 0.032265 | 0.985972 | | 3.800486 | -0.15028 | 0.917499 |
| PTPMT1 | 4.240628 | 0.264975 | 0.999532 | | 4.594864 | 0.030082 | 0.986014 | | 4.181936 | -0.20651 | 0.876107 |
| SPATA21 | 2.98834 | 0.594424 | 0.999532 | | 2.75064 | -0.0401 | 0.986144 | | 2.881865 | -0.61664 | 0.622543 |
| IFT43 | 3.241335 | -0.18681 | 0.999532 | | 4.063184 | 0.032574 | 0.986144 | | 3.173202 | 0.235899 | 0.869378 |
| PRDM13 | 2.3696 | 0.268904 | 0.999532 | | 2.676784 | 0.0405 | 0.986144 | | 2.494803 | -0.20878 | 0.905128 |
| FAM78B | 2.154152 | 0.031705 | 0.999532 | | 2.210241 | -0.04247 | 0.986144 | | 1.980264 | -0.06177 | 0.982188 |
| ZNF667 | 2.194577 | 0.591446 | 0.999532 | | 2.057447 | -0.04758 | 0.986144 | | 2.110995 | -0.62067 | 0.682485 |
| SLC9A6 | 3.108867 | 0.607611 | 0.999532 | | 2.708943 | 0.038459 | 0.986144 | | 2.999157 | -0.54946 | 0.662641 |
| LRRC1 | 2.477928 | 0.277625 | 0.999532 | | 2.566385 | 0.038592 | 0.986144 | | 2.397538 | -0.2217 | 0.894617 |
| TARBP1 | 2.695739 | -0.22251 | 0.999532 | | 3.223878 | 0.035625 | 0.986144 | | 2.62606 | 0.276418 | 0.864923 |
| SSPN | 3.098685 | -0.43739 | 0.999532 | | 3.807841 | 0.032558 | 0.986144 | | 2.94823 | 0.491918 | 0.694961 |
| C12orf75 | 2.112889 | 0.350066 | 0.999532 | | 2.061804 | 0.042764 | 0.986144 | | 1.98207 | -0.2962 | 0.877644 |
| NegProbe | 1.284381 | 0.419039 | 0.999532 | | 1.22145 | -0.05642 | 0.986144 | | 1.249335 | -0.45605 | 0.836453 |
| SPTSSA | 1.863047 | -0.25479 | 0.999532 | | 2.2959 | -0.04206 | 0.986144 | | 1.872684 | 0.233144 | 0.907882 |
| FOXA3 | 2.136633 | -0.22413 | 0.999532 | | 2.552511 | -0.04095 | 0.986144 | | 1.95187 | 0.202363 | 0.91993 |
| SLC26A6 | 3.500516 | 0.423226 | 0.999532 | | 3.494935 | -0.03692 | 0.986144 | | 3.344823 | -0.4345 | 0.736481 |
| PRKAR1B | 3.103408 | -0.16229 | 0.999532 | | 3.808983 | -0.03345 | 0.986144 | | 3.024946 | 0.152676 | 0.92697 |
| CAMK1G | 3.474518 | -0.27331 | 0.999532 | | 4.125456 | -0.03005 | 0.986144 | | 3.226812 | 0.254916 | 0.859426 |
| RFNG | 6.306691 | 0.24653 | 0.999532 | | 6.479842 | -0.02559 | 0.986144 | | 6.076468 | -0.25095 | 0.800724 |
| BOD1L1 | 2.625307 | 0.174305 | 0.999532 | | 2.83466 | -0.0401 | 0.986144 | | 2.545817 | -0.19875 | 0.907968 |
| MYOD1 | 2.330125 | 0.039492 | 0.999532 | | 2.643844 | -0.03844 | 0.986144 | | 2.287945 | -0.05874 | 0.979996 |
| TTI2 | 2.965232 | 0.522087 | 0.999532 | | 2.589914 | 0.041252 | 0.986144 | | 2.829687 | -0.46305 | 0.739496 |
| C12orf66 | 2.300742 | 0.077251 | 0.999532 | | 2.542541 | 0.039806 | 0.986144 | | 2.223875 | -0.01968 | 0.996681 |
| DISP2 | 2.372835 | 0.520729 | 0.999532 | | 2.065084 | -0.04536 | 0.986144 | | 2.27876 | -0.55175 | 0.741408 |
| PALMD | 3.914263 | 0.149082 | 0.999532 | | 4.484357 | -0.03447 | 0.986144 | | 4.058247 | -0.16548 | 0.914929 |
| WFDC2 | 4.36414 | 0.295757 | 0.999532 | | 4.506675 | -0.03785 | 0.986144 | | 4.017243 | -0.30271 | 0.835289 |
| HEATR1 | 2.763998 | 0.047995 | 0.999532 | | 3.035722 | 0.036149 | 0.986144 | | 2.655851 | 0.001914 | 0.999429 |
| CNTNAP1 | 3.318337 | 0.250392 | 0.999532 | | 3.404624 | 0.033646 | 0.986144 | | 3.230222 | -0.19876 | 0.892869 |
| ZNF451 | 2.895611 | 0.285673 | 0.999532 | | 2.918798 | 0.037666 | 0.986144 | | 2.727029 | -0.22935 | 0.883792 |
| MTURN | 3.666509 | 0.120221 | 0.999532 | | 4.042697 | -0.03205 | 0.986144 | | 3.434081 | -0.13513 | 0.936909 |
| NAALADL | 2.829743 | 0.607274 | 0.999532 | | 2.536094 | 0.039331 | 0.986144 | | 2.714194 | -0.54832 | 0.662818 |
| LYRM1 | 2.520303 | -0.0888 | 0.999532 | | 3.238411 | 0.036513 | 0.986144 | | 2.65203 | 0.153167 | 0.935145 |
| PLCXD2 | 1.952908 | 0.041603 | 0.999532 | | 2.161873 | -0.04419 | 0.986144 | | 1.999878 | -0.07148 | 0.97843 |
| PIGG | 3.481432 | -0.16612 | 0.999532 | | 3.973476 | -0.03121 | 0.986144 | | 3.297121 | 0.152802 | 0.920605 |
| CLVS1 | 2.196488 | 0.333665 | 0.999532 | | 2.194649 | 0.041691 | 0.986144 | | 2.214514 | -0.27471 | 0.874653 |
| ZNF302 | 3.607105 | 0.171329 | 0.999532 | | 3.924261 | -0.03404 | 0.986144 | | 3.629419 | -0.18906 | 0.903329 |
| RFTN2 | 2.996496 | 0.293463 | 0.999532 | | 3.093074 | -0.03885 | 0.986144 | | 2.9085 | -0.31349 | 0.836012 |
| SLC35E1 | 3.125367 | 0.310777 | 0.999532 | | 3.063874 | -0.03692 | 0.986144 | | 2.918334 | -0.33 | 0.808547 |
| TSTD1 | 3.511224 | 0.457508 | 0.999532 | | 3.169538 | -0.03623 | 0.986144 | | 3.265159 | -0.4799 | 0.694301 |
| HDAC3 | 3.792573 | -0.13462 | 0.999532 | | 4.577272 | -0.03136 | 0.986144 | | 3.817342 | 0.125266 | 0.939942 |
| UBAC1 | 4.972484 | -0.00496 | 0.999532 | | 5.669251 | -0.02923 | 0.986144 | | 4.825501 | -0.00712 | 0.996908 |
| TOR2A | 2.119381 | 0.65129 | 0.999532 | | 1.860345 | 0.043649 | 0.986206 | | 1.996118 | -0.59 | 0.678541 |
| THTPA | 3.814097 | 0.186703 | 0.999532 | | 4.098487 | 0.031033 | 0.986206 | | 3.640518 | -0.13769 | 0.924836 |
| B3GALT2 | 2.457802 | -0.03834 | 0.999532 | | 2.852873 | -0.04014 | 0.98625 | | 2.382554 | 0.020888 | 0.99666 |
| TMEM120 | 2.345775 | -0.18397 | 0.999532 | | 2.674574 | -0.03795 | 0.98625 | | 2.102847 | 0.163522 | 0.934172 |
| SH3BP5L | 3.403661 | -0.20752 | 0.999532 | | 4.111906 | 0.033381 | 0.986263 | | 3.215618 | 0.26027 | 0.864019 |
| OTUD3 | 2.00776 | -0.33071 | 0.999532 | | 2.54688 | 0.039191 | 0.986303 | | 1.955881 | 0.389138 | 0.823571 |
| NDUFAF6 | 2.694546 | -0.39941 | 0.999532 | | 3.308174 | -0.03343 | 0.986331 | | 2.489192 | 0.383911 | 0.791466 |
| ADGRG4 | 2.450979 | 0.3191 | 0.999532 | | 2.419041 | -0.04028 | 0.986343 | | 2.39403 | -0.3402 | 0.836012 |
| ZNF765 | 3.004075 | 0.124117 | 0.999532 | | 3.38084 | -0.03567 | 0.986343 | | 2.996743 | -0.14856 | 0.929529 |
| SIDT2 | 3.997612 | -0.40742 | 0.999532 | | 4.928672 | -0.02859 | 0.986507 | | 3.611398 | 0.395921 | 0.735469 |
| C3orf22 | 2.784139 | -0.04903 | 0.999532 | | 3.473932 | 0.034151 | 0.986507 | | 2.885555 | 0.119494 | 0.950616 |
| MED12 | 4.428681 | -0.15512 | 0.999532 | | 5.059381 | -0.03114 | 0.986507 | | 4.110834 | 0.144078 | 0.923864 |
| PTCH1 | 2.552892 | 0.307415 | 0.999532 | | 2.624817 | -0.04026 | 0.986507 | | 2.47977 | -0.32551 | 0.849128 |
| IL9R | 1.85571 | 0.216398 | 0.999532 | | 2.046813 | -0.04403 | 0.986507 | | 1.858112 | -0.23834 | 0.905952 |
| KCNK15 | 2.213068 | 0.051695 | 0.999532 | | 2.469932 | 0.0392 | 0.986507 | | 2.169957 | 0.006479 | 0.998401 |
| FBXO7 | 4.416191 | -0.02552 | 0.999532 | | 4.894507 | -0.0276 | 0.986507 | | 4.002037 | 0.013018 | 0.996681 |
| TAF1 | 4.873243 | -0.11455 | 0.999532 | | 5.571015 | -0.02609 | 0.986507 | | 4.574214 | 0.10297 | 0.943248 |
| POPDC2 | 2.610546 | -0.10235 | 0.999532 | | 3.123519 | 0.033895 | 0.986507 | | 2.620792 | 0.159157 | 0.929087 |
| FOXP1 | 6.106592 | -0.17506 | 0.999532 | | 6.745909 | -0.02464 | 0.986507 | | 5.623022 | 0.132795 | 0.92403 |
| ZFTA | 4.506225 | 0.002148 | 0.999532 | | 4.871497 | 0.029366 | 0.986507 | | 4.285335 | 0.049119 | 0.978282 |
| DHX57 | 2.128445 | 0.419033 | 0.999532 | | 2.137855 | 0.041005 | 0.986507 | | 2.148835 | -0.35926 | 0.831749 |
| SVBP | 2.958725 | -0.33001 | 0.999532 | | 3.620631 | -0.03455 | 0.986507 | | 2.896262 | 0.305441 | 0.839943 |
| TRIM26 | 4.388619 | 0.040169 | 0.999532 | | 4.964881 | 0.027489 | 0.986507 | | 4.250796 | 0.004737 | 0.99833 |
| OR2AT4 | 2.2426 | 0.473722 | 0.999532 | | 2.109577 | -0.04239 | 0.986507 | | 2.219684 | -0.50529 | 0.736481 |
| CCDC66 | 3.488028 | 0.055205 | 0.999532 | | 3.905237 | 0.031686 | 0.986507 | | 3.402503 | -0.00441 | 0.998586 |
| DMAC2L | 2.203552 | -0.19239 | 0.999532 | | 2.706473 | 0.035354 | 0.986507 | | 2.242506 | 0.24918 | 0.885078 |
| CEACAM | 3.013588 | 0.26768 | 0.999532 | | 2.769883 | -0.03791 | 0.986507 | | 2.718119 | -0.30146 | 0.861119 |
| OR5M8 | 2.30994 | 0.428984 | 0.999532 | | 2.368507 | 0.04221 | 0.986507 | | 2.292867 | -0.35826 | 0.849532 |
| ZFP14 | 2.310235 | 0.102295 | 0.999532 | | 2.598221 | 0.040187 | 0.986507 | | 2.272622 | -0.04432 | 0.988852 |
| H2BC14 | 1.653896 | 0.256677 | 0.999532 | | 1.768322 | 0.045551 | 0.986507 | | 1.585722 | -0.19364 | 0.931827 |
| ADAMTS | 2.183235 | -0.3259 | 0.999532 | | 2.783403 | -0.03793 | 0.986507 | | 2.140328 | 0.307874 | 0.865065 |
| ATP5MG | 15.91804 | 0.160681 | 0.999532 | | 16.50118 | -0.01803 | 0.986507 | | 15.1999 | -0.15653 | 0.862923 |
| TMEM41B | 5.600081 | -0.47455 | 0.999532 | | 7.353253 | 0.029726 | 0.986507 | | 5.658402 | 0.522076 | 0.663774 |
| CDC14B | 4.504058 | 0.099873 | 0.999532 | | 4.718664 | -0.02995 | 0.986544 | | 4.46158 | -0.11621 | 0.943463 |
| CYP4X1 | 4.409455 | -0.11758 | 0.999532 | | 5.229569 | -0.02853 | 0.986544 | | 4.2346 | 0.115729 | 0.941512 |
| PLD5 | 2.370083 | 0.423053 | 0.999532 | | 2.1571 | -0.0408 | 0.986544 | | 2.23622 | -0.44964 | 0.780456 |
| RAPGEF | 5.488597 | -0.13644 | 0.999532 | | 6.291946 | 0.025705 | 0.986639 | | 5.263646 | 0.169094 | 0.898369 |
| KRAS | 6.463659 | 0.035779 | 0.999532 | | 7.106 | 0.024202 | 0.986639 | | 5.940672 | 0.007688 | 0.996891 |
| RUNX2 | 3.276823 | 0.450838 | 0.999532 | | 3.050352 | -0.03601 | 0.986639 | | 2.995088 | -0.46677 | 0.713145 |
| MFSD6L | 4.927789 | -0.07946 | 0.999532 | | 5.751299 | -0.02992 | 0.986639 | | 4.75467 | 0.069976 | 0.970532 |
| CCNB1 | 2.225227 | 0.038023 | 0.999532 | | 2.501209 | -0.03771 | 0.986639 | | 2.207483 | -0.05848 | 0.982124 |
| C22orf31 | 2.337449 | 0.523626 | 0.999532 | | 2.200282 | -0.0414 | 0.986639 | | 2.180293 | -0.54307 | 0.717627 |
| GLB1L3 | 2.166056 | 0.472313 | 0.999532 | | 2.099881 | 0.043198 | 0.986639 | | 2.184862 | -0.40811 | 0.816589 |
| CPA4 | 1.834686 | -0.38128 | 0.999532 | | 2.367484 | 0.039081 | 0.986639 | | 1.866613 | 0.439923 | 0.805466 |
| TTLL12 | 2.719993 | 0.011369 | 0.999532 | | 2.994181 | -0.03501 | 0.986639 | | 2.583837 | -0.02923 | 0.992286 |
| GLI3 | 2.205003 | 0.115927 | 0.999532 | | 2.298826 | 0.039095 | 0.986639 | | 2.146899 | -0.05907 | 0.982124 |
| ZNF518B | 2.091996 | -0.13939 | 0.999532 | | 2.488317 | 0.036415 | 0.986639 | | 2.043499 | 0.193322 | 0.918694 |
| ZSCAN9 | 2.666297 | -0.15493 | 0.999532 | | 3.073575 | -0.0339 | 0.986639 | | 2.583875 | 0.141033 | 0.94109 |
| ZSCAN16 | 2.639631 | -0.15067 | 0.999532 | | 3.008133 | 0.035761 | 0.986639 | | 2.266697 | 0.208626 | 0.903567 |
| TMEM252 | 2.294757 | -0.02902 | 0.999532 | | 2.56261 | 0.038096 | 0.986639 | | 2.23782 | 0.080455 | 0.973659 |
| ZFP64 | 2.201106 | 0.219112 | 0.999532 | | 2.365797 | -0.03747 | 0.986639 | | 2.194282 | -0.23785 | 0.892225 |
| PPP2R5B | 3.861291 | 0.234403 | 0.999532 | | 3.738416 | -0.03075 | 0.986639 | | 3.606362 | -0.25753 | 0.858767 |
| B3GALT6 | 2.212989 | 0.16118 | 0.999532 | | 2.403978 | 0.039266 | 0.986639 | | 2.151337 | -0.10433 | 0.964728 |
| ZNHIT3 | 4.744048 | 0.171071 | 0.999532 | | 5.12795 | -0.02953 | 0.986639 | | 4.569537 | -0.18668 | 0.885892 |
| SH3BP5 | 7.366318 | -0.2047 | 0.999532 | | 8.656724 | 0.022707 | 0.986639 | | 6.826176 | 0.244042 | 0.809832 |
| ARL5B | 2.823908 | -0.12772 | 0.999532 | | 3.266228 | -0.03334 | 0.986639 | | 2.698454 | 0.112906 | 0.955834 |
| FHIP1A | 1.884226 | -0.13015 | 0.999532 | | 2.146858 | -0.03992 | 0.986639 | | 1.804804 | 0.107624 | 0.965649 |
| CCDC102 | 2.644875 | 0.098101 | 0.999532 | | 2.771393 | -0.0352 | 0.986639 | | 2.503201 | -0.11478 | 0.953949 |
| NCF2 | 2.163447 | -0.33937 | 0.999532 | | 2.462705 | -0.03877 | 0.986639 | | 2.099258 | 0.330658 | 0.859932 |
| TMEM33 | 4.385376 | 0.030152 | 0.999532 | | 4.951643 | 0.028635 | 0.986639 | | 4.231271 | 0.018094 | 0.994347 |
| ZSCAN25 | 2.458602 | -0.02434 | 0.999532 | | 2.729218 | -0.03525 | 0.986639 | | 2.312424 | 0.006886 | 0.997967 |
| CDH18 | 1.818714 | 0.719833 | 0.999532 | | 1.672693 | 0.047176 | 0.986639 | | 1.781554 | -0.65373 | 0.67937 |
| FAM111A | 6.108419 | 0.095542 | 0.999532 | | 6.606788 | 0.025699 | 0.986639 | | 5.777939 | -0.05161 | 0.974421 |
| OR52K2 | 1.935832 | 0.227653 | 0.999532 | | 2.017838 | -0.04237 | 0.986693 | | 1.86701 | -0.25111 | 0.900848 |
| FYTTD1 | 4.176404 | -0.10131 | 0.999532 | | 4.578363 | 0.027972 | 0.986847 | | 3.828895 | 0.139264 | 0.926586 |
| UVRAG | 5.76005 | -0.00612 | 0.999532 | | 6.649361 | 0.023575 | 0.986847 | | 5.698698 | 0.048205 | 0.974421 |
| WDR89 | 3.061164 | 0.192723 | 0.999532 | | 3.255838 | 0.034645 | 0.986847 | | 3.027045 | -0.14031 | 0.939913 |
| F2 | 2.314145 | -0.19515 | 0.999532 | | 2.48698 | -0.03757 | 0.986847 | | 2.13862 | 0.1652 | 0.940266 |
| IRGQ | 3.501999 | 0.29287 | 0.999532 | | 3.680542 | 0.032073 | 0.986847 | | 3.368473 | -0.24339 | 0.865065 |
| ZNF569 | 2.660922 | 0.293707 | 0.999532 | | 2.648969 | 0.035227 | 0.986872 | | 2.563299 | -0.24407 | 0.881837 |
| MTR | 2.671775 | 0.171457 | 0.999532 | | 3.097394 | 0.034951 | 0.986994 | | 2.7703 | -0.11634 | 0.952822 |
| TAP1 | 5.260206 | -0.37634 | 0.999532 | | 6.339576 | -0.02717 | 0.986994 | | 5.012734 | 0.389989 | 0.702463 |
| EBLN2 | 3.160593 | 0.652691 | 0.999532 | | 2.868345 | -0.03503 | 0.986994 | | 2.849949 | -0.67121 | 0.540342 |
| EIF2S1 | 4.979936 | 0.016737 | 0.999532 | | 5.667534 | 0.024715 | 0.986994 | | 4.786833 | 0.026527 | 0.99013 |
| CLDN16 | 2.748379 | 0.232863 | 0.999532 | | 3.044174 | 0.036468 | 0.986994 | | 2.663556 | -0.15058 | 0.939549 |
| AKAP13 | 3.421374 | -0.23798 | 0.999532 | | 3.958239 | 0.032449 | 0.987179 | | 3.135143 | 0.290243 | 0.855088 |
| TTYH2 | 3.393067 | 0.17361 | 0.999532 | | 3.670066 | -0.03235 | 0.987179 | | 3.47518 | -0.19139 | 0.903659 |
| APBA3 | 5.776065 | 0.3483 | 0.999532 | | 5.775285 | 0.026287 | 0.987179 | | 5.646184 | -0.30601 | 0.752562 |
| ZNF530 | 3.094437 | 0.241061 | 0.999532 | | 3.243434 | 0.03184 | 0.987179 | | 3.005706 | -0.1919 | 0.898369 |
| AP1M2 | 2.298162 | 0.344465 | 0.999532 | | 2.241041 | -0.03897 | 0.987179 | | 2.184438 | -0.36413 | 0.82512 |
| SUSD3 | 2.033563 | 0.291121 | 0.999532 | | 2.176197 | 0.037921 | 0.987179 | | 2.06033 | -0.23321 | 0.897958 |
| MCM8 | 1.912667 | -0.58886 | 0.999532 | | 2.350505 | 0.037726 | 0.987273 | | 1.716616 | 0.643237 | 0.702463 |
| BET1 | 4.021418 | 0.003044 | 0.999532 | | 4.549104 | -0.02735 | 0.987273 | | 3.805644 | -0.00847 | 0.996891 |
| COPS9 | 6.08496 | 0.178428 | 0.999532 | | 6.05907 | 0.02913 | 0.987344 | | 5.60524 | -0.0927 | 0.946958 |
| PLRG1 | 3.915232 | -0.01314 | 0.999532 | | 4.326973 | 0.029796 | 0.987344 | | 3.831596 | 0.058854 | 0.974808 |
| JHY | 4.366968 | 0.242606 | 0.999532 | | 4.511044 | -0.02976 | 0.987344 | | 3.981305 | -0.25386 | 0.842754 |
| RPH3A | 1.763494 | 0.462179 | 0.999532 | | 1.723598 | 0.044011 | 0.987344 | | 1.680237 | -0.39987 | 0.835554 |
| COL17A1 | 2.180817 | 0.426213 | 0.999532 | | 2.162919 | 0.039202 | 0.987344 | | 2.17768 | -0.36751 | 0.836935 |
| DOK5 | 1.931469 | -0.31056 | 0.999532 | | 2.28526 | 0.036474 | 0.987344 | | 1.881671 | 0.35957 | 0.84579 |
| OR2T3 | 2.554843 | 0.3347 | 0.999532 | | 2.617124 | 0.037604 | 0.987344 | | 2.566594 | -0.28404 | 0.87591 |
| CRY1 | 4.480153 | 0.480681 | 0.999532 | | 4.093881 | 0.029698 | 0.987344 | | 4.28975 | -0.43333 | 0.709825 |
| REELD1 | 2.354537 | -0.03481 | 0.999532 | | 2.518334 | -0.03689 | 0.987624 | | 2.184758 | 0.01923 | 0.996681 |
| COMTD1 | 2.647289 | 0.299551 | 0.999532 | | 2.763238 | -0.03488 | 0.987892 | | 2.539848 | -0.31153 | 0.843043 |
| GFOD2 | 2.188144 | 0.749116 | 0.999532 | | 1.974002 | 0.040528 | 0.987892 | | 2.058207 | -0.68741 | 0.628895 |
| TMEM168 | 2.590689 | -0.76115 | 0.999532 | | 3.671991 | 0.030419 | 0.987975 | | 2.558126 | 0.810372 | 0.515762 |
| CSTF2 | 2.934018 | -0.03829 | 0.999532 | | 3.325376 | -0.03143 | 0.988011 | | 2.787186 | 0.022715 | 0.994347 |
| RUSC1 | 6.653351 | -0.11964 | 0.999532 | | 7.788458 | -0.02165 | 0.988102 | | 6.41344 | 0.116111 | 0.919541 |
| APMAP | 5.952394 | -0.03877 | 0.999532 | | 6.771823 | 0.022265 | 0.988102 | | 5.662529 | 0.079126 | 0.952656 |
| OR1L1 | 1.84869 | 0.427239 | 0.999532 | | 1.861033 | 0.039912 | 0.988102 | | 1.874825 | -0.36793 | 0.836453 |
| ZNF862 | 2.072004 | 0.240165 | 0.999532 | | 2.098607 | -0.03784 | 0.988102 | | 1.938953 | -0.25997 | 0.88885 |
| DGKA | 2.199609 | -0.00608 | 0.999532 | | 2.564623 | 0.035539 | 0.988102 | | 2.212818 | 0.057642 | 0.982124 |
| SS18L2 | 2.472665 | 0.092772 | 0.999532 | | 2.735533 | 0.033835 | 0.988102 | | 2.532413 | -0.04105 | 0.988853 |
| MLEC | 5.888231 | 0.14446 | 0.999532 | | 6.527661 | 0.023913 | 0.988172 | | 5.712559 | -0.10495 | 0.93603 |
| PRICKLE | 2.623959 | 0.258883 | 0.999532 | | 2.74855 | 0.034263 | 0.988191 | | 2.51684 | -0.20528 | 0.904941 |
| TMEM135 | 2.364699 | 0.008969 | 0.999532 | | 2.482362 | -0.03459 | 0.988191 | | 2.139121 | -0.04828 | 0.987274 |
| OTUD1 | 3.323425 | 0.200616 | 0.999532 | | 3.338528 | -0.0303 | 0.988191 | | 3.020216 | -0.21335 | 0.888003 |
| KRT77 | 1.978655 | 0.272667 | 0.999532 | | 2.070769 | -0.04023 | 0.988191 | | 1.91323 | -0.29191 | 0.881689 |
| CCNF | 2.212406 | 0.679493 | 0.999532 | | 2.026112 | 0.038308 | 0.988191 | | 2.20112 | -0.62065 | 0.660097 |
| PDZD7 | 2.219533 | 0.540222 | 0.999532 | | 2.131247 | -0.03943 | 0.988191 | | 2.105936 | -0.56984 | 0.701199 |
| LRRC10B | 1.790693 | -0.15351 | 0.999532 | | 2.167738 | -0.03757 | 0.988385 | | 1.752206 | 0.13565 | 0.955834 |
| AIMP2 | 2.218474 | 0.092739 | 0.999532 | | 2.64976 | -0.03422 | 0.988385 | | 2.196762 | -0.0954 | 0.966652 |
| RAB36 | 3.674092 | 0.389464 | 0.999532 | | 3.621081 | -0.02954 | 0.988385 | | 3.488336 | -0.39991 | 0.747457 |
| GALNS | 2.576654 | 0.17244 | 0.999532 | | 2.471239 | 0.034339 | 0.988385 | | 2.458846 | -0.13415 | 0.94866 |
| JADE2 | 3.534865 | -0.31505 | 0.999532 | | 4.467547 | 0.028138 | 0.988385 | | 3.405529 | 0.363369 | 0.772912 |
| C6orf226 | 2.95347 | 0.542506 | 0.999532 | | 2.650967 | -0.0326 | 0.988385 | | 2.783425 | -0.55773 | 0.65401 |
| CHD6 | 3.352978 | -0.26509 | 0.999532 | | 4.024483 | 0.027553 | 0.988385 | | 3.281092 | 0.310972 | 0.820072 |
| FAM236C | 2.38106 | 0.537743 | 0.999532 | | 2.19755 | -0.03892 | 0.988385 | | 2.348433 | -0.55907 | 0.717627 |
| KLRC3 | 2.288175 | 0.608741 | 0.999532 | | 2.119211 | -0.03897 | 0.988385 | | 2.218935 | -0.63016 | 0.660097 |
| MYOZ1 | 3.449396 | 0.022433 | 0.999532 | | 3.87535 | 0.033236 | 0.988385 | | 3.328055 | 0.03517 | 0.989437 |
| PRRG2 | 2.824801 | 0.556442 | 0.999532 | | 2.58287 | 0.034275 | 0.988385 | | 2.792704 | -0.50682 | 0.718643 |
| EFS | 1.836402 | 0.234247 | 0.999532 | | 1.972524 | 0.038404 | 0.988385 | | 1.845875 | -0.17487 | 0.93603 |
| FRG1 | 8.316958 | 0.276295 | 0.999532 | | 8.459045 | -0.02296 | 0.988443 | | 8.100389 | -0.28912 | 0.771839 |
| CPSF1 | 6.678635 | -0.00846 | 0.999532 | | 7.712088 | -0.02126 | 0.988491 | | 6.368553 | 0.013544 | 0.995782 |
| MYEF2 | 4.013139 | 0.188458 | 0.999532 | | 4.037306 | -0.02792 | 0.988502 | | 3.789862 | -0.19618 | 0.896374 |
| STS | 2.842886 | 0.324227 | 0.999532 | | 2.876215 | -0.03074 | 0.988502 | | 2.708014 | -0.33675 | 0.811123 |
| CSNK1D | 6.234336 | 0.129757 | 0.999532 | | 6.732706 | 0.022125 | 0.988502 | | 5.754122 | -0.08511 | 0.949669 |
| ZNF284 | 2.232335 | 0.14504 | 0.999532 | | 2.506193 | 0.034351 | 0.988502 | | 2.364227 | -0.08561 | 0.972221 |
| ZNF227 | 2.993478 | 0.352081 | 0.999532 | | 3.03007 | -0.03561 | 0.988502 | | 2.812143 | -0.35923 | 0.814402 |
| ZNF410 | 2.593099 | -0.18334 | 0.999532 | | 3.132914 | 0.030895 | 0.988502 | | 2.580973 | 0.231457 | 0.887703 |
| XPOT | 6.033675 | 0.057187 | 0.999532 | | 6.691888 | -0.02221 | 0.988502 | | 5.655608 | -0.06101 | 0.970532 |
| HTR2B | 2.38048 | -0.33582 | 0.999532 | | 2.918246 | 0.031918 | 0.988502 | | 2.348616 | 0.387686 | 0.803301 |
| IFRD1 | 3.758002 | 0.214019 | 0.999532 | | 3.777128 | -0.02824 | 0.988502 | | 3.543855 | -0.22249 | 0.881316 |
| RND2 | 1.813708 | 0.156926 | 0.999532 | | 1.972789 | -0.0384 | 0.988502 | | 1.750167 | -0.17561 | 0.938429 |
| TSPYL4 | 4.916261 | 0.233289 | 0.999532 | | 4.893287 | -0.02636 | 0.988502 | | 4.424787 | -0.23013 | 0.854633 |
| TNNI3K | 2.592665 | 0.017323 | 0.999532 | | 2.858892 | -0.0344 | 0.988502 | | 2.619428 | -0.04764 | 0.987274 |
| CDCA7 | 2.198152 | -0.10199 | 0.999532 | | 2.412229 | -0.03436 | 0.988502 | | 2.006979 | 0.078504 | 0.97502 |
| CBY1 | 2.910984 | 0.276836 | 0.999532 | | 2.916017 | 0.032072 | 0.988502 | | 2.939572 | -0.2313 | 0.895361 |
| PSMD13 | 6.412052 | -0.28082 | 0.999532 | | 8.153638 | 0.019646 | 0.988502 | | 6.06366 | 0.322409 | 0.724411 |
| CT47B1 | 3.864654 | 0.243144 | 0.999532 | | 3.794602 | 0.028898 | 0.988502 | | 3.831083 | -0.20104 | 0.892869 |
| PCED1A | 3.898449 | -0.23023 | 0.999532 | | 4.387746 | -0.0269 | 0.988502 | | 3.540196 | 0.224468 | 0.87591 |
| PARVG | 2.309073 | -0.06795 | 0.999532 | | 2.673008 | -0.03214 | 0.988502 | | 2.214881 | 0.053826 | 0.983749 |
| PRMT7 | 2.795477 | -0.02343 | 0.999532 | | 3.363229 | 0.030292 | 0.988502 | | 2.81476 | 0.083511 | 0.970268 |
| CD79B | 5.554564 | 0.392909 | 0.999532 | | 5.280815 | -0.02415 | 0.988502 | | 5.439284 | -0.40109 | 0.672452 |
| IL15RA | 3.673059 | 0.333612 | 0.999532 | | 3.825773 | 0.027791 | 0.988502 | | 3.565081 | -0.28619 | 0.823193 |
| EOGT | 3.409276 | -0.34582 | 0.999532 | | 4.038995 | -0.0274 | 0.988502 | | 3.069087 | 0.329867 | 0.810753 |
| EXOC3L2 | 2.886252 | -0.02298 | 0.999532 | | 3.315568 | -0.03069 | 0.988502 | | 2.812045 | 0.01043 | 0.996891 |
| RABL6 | 5.610939 | 0.36264 | 0.999532 | | 5.17733 | 0.025556 | 0.988502 | | 5.2396 | -0.32388 | 0.754635 |
| GABRB3 | 2.026067 | 0.053091 | 0.999532 | | 2.163495 | 0.035691 | 0.988612 | | 1.993719 | 0.006728 | 0.998401 |
| CCNJ | 2.063738 | -0.1404 | 0.999532 | | 2.4775 | -0.03321 | 0.988729 | | 2.008339 | 0.128008 | 0.954304 |
| EFNA4 | 1.971309 | -0.30173 | 0.999532 | | 2.33654 | 0.033593 | 0.988729 | | 1.963343 | 0.347101 | 0.854633 |
| OR2A25 | 1.942744 | 0.370589 | 0.999532 | | 2.001311 | 0.038908 | 0.988729 | | 1.694671 | -0.31146 | 0.868512 |
| CCM2L | 4.022093 | -0.63609 | 0.999532 | | 5.356282 | -0.0244 | 0.988729 | | 3.661246 | 0.629429 | 0.533876 |
| TMEM222 | 2.244784 | 0.097126 | 0.999532 | | 2.382099 | 0.033627 | 0.988729 | | 2.210018 | -0.04964 | 0.986471 |
| NRTN | 2.474451 | 0.348457 | 0.999532 | | 2.657518 | 0.03168 | 0.988729 | | 2.487481 | -0.29013 | 0.858705 |
| DIS3L2 | 2.649373 | -0.42397 | 0.999532 | | 3.375862 | 0.027916 | 0.988729 | | 2.551057 | 0.468424 | 0.724411 |
| SELENOH | 4.556623 | -0.50404 | 0.999532 | | 5.58365 | -0.02223 | 0.988729 | | 4.339657 | 0.49965 | 0.634624 |
| ZNF285 | 2.867435 | 0.075046 | 0.999532 | | 3.252785 | 0.03074 | 0.988753 | | 2.908821 | -0.02689 | 0.992586 |
| ODF2L | 2.841086 | -0.31681 | 0.999532 | | 3.457177 | -0.02877 | 0.988753 | | 2.583795 | 0.304631 | 0.842696 |
| DAPK2 | 3.017788 | -0.71352 | 0.999532 | | 4.112367 | -0.0267 | 0.988763 | | 2.856603 | 0.703939 | 0.552725 |
| KRT222 | 2.345558 | 0.409692 | 0.999532 | | 2.414714 | -0.03568 | 0.988763 | | 2.417295 | -0.42794 | 0.791466 |
| KANSL2 | 3.745802 | -0.08515 | 0.999532 | | 4.275313 | -0.02854 | 0.988775 | | 3.465716 | 0.075481 | 0.972344 |
| TEX48 | 2.206201 | 0.700401 | 0.999532 | | 2.016423 | -0.03566 | 0.988775 | | 2.146715 | -0.71071 | 0.60629 |
| ZNF333 | 2.333953 | 0.1291 | 0.999532 | | 2.779723 | 0.031881 | 0.988775 | | 2.284323 | -0.07258 | 0.974421 |
| MVK | 1.941154 | 0.455566 | 0.999532 | | 1.892254 | -0.03683 | 0.988775 | | 1.870019 | -0.4714 | 0.7719 |
| NUP133 | 4.930238 | -0.24409 | 0.999532 | | 5.911202 | -0.02168 | 0.988775 | | 4.535018 | 0.239323 | 0.842754 |
| LRRC24 | 1.731817 | -0.07186 | 0.999532 | | 1.943269 | -0.03534 | 0.988775 | | 1.623445 | 0.05443 | 0.986208 |
| ASDURF | 4.410564 | 0.022561 | 0.999532 | | 5.056914 | -0.02479 | 0.988775 | | 4.345162 | -0.02953 | 0.989985 |
| COPS6 | 6.337729 | -0.08362 | 0.999532 | | 7.380405 | 0.0199 | 0.988775 | | 5.91139 | 0.121708 | 0.918942 |
| MRAP | 1.988429 | 0.398486 | 0.999532 | | 1.881401 | 0.036974 | 0.988775 | | 1.978451 | -0.34454 | 0.849885 |
| BCAT2 | 5.286249 | 0.052976 | 0.999532 | | 5.829162 | 0.022189 | 0.988775 | | 5.124163 | -0.01115 | 0.996681 |
| COL2A1 | 1.77063 | 0.047642 | 0.999532 | | 2.002019 | 0.037609 | 0.988775 | | 1.83471 | 0.009476 | 0.997967 |
| SEC23B | 3.680615 | -0.33136 | 0.999532 | | 4.789662 | 0.025069 | 0.988775 | | 3.62385 | 0.378314 | 0.760473 |
| VPS35L | 4.228047 | -0.14787 | 0.999532 | | 4.934781 | 0.023498 | 0.988775 | | 3.962585 | 0.188484 | 0.885078 |
| MRTFB | 4.39319 | 0.053525 | 0.999532 | | 4.783146 | -0.02399 | 0.988775 | | 4.26554 | -0.05641 | 0.974588 |
| NEU1 | 3.740132 | -0.00187 | 0.999532 | | 3.914793 | -0.0263 | 0.988775 | | 3.397397 | -0.01305 | 0.996681 |
| PNKP | 3.714987 | -0.35573 | 0.999532 | | 4.485642 | -0.02426 | 0.988775 | | 3.442754 | 0.348096 | 0.7719 |
| APH1B | 2.404007 | 0.04863 | 0.999532 | | 2.684233 | 0.030619 | 0.988775 | | 2.255301 | -0.00152 | 0.999463 |
| NOTO | 2.349172 | -0.2144 | 0.999532 | | 2.963277 | 0.03081 | 0.988775 | | 2.381791 | 0.263577 | 0.880577 |
| RABGGTA | 2.723787 | 0.02522 | 0.999532 | | 2.925133 | -0.03059 | 0.988775 | | 2.63041 | -0.04308 | 0.987274 |
| PIK3R1 | 3.912824 | 0.215825 | 0.999532 | | 4.18229 | 0.02609 | 0.988775 | | 3.705012 | -0.16947 | 0.904721 |
| UBE2L6 | 8.904762 | 0.121466 | 0.999532 | | 9.412706 | 0.019909 | 0.988775 | | 8.636373 | -0.06725 | 0.955834 |
| OGFOD3 | 2.991066 | 0.069456 | 0.999532 | | 3.210164 | -0.02863 | 0.98887 | | 2.902872 | -0.08641 | 0.965619 |
| PRLR | 2.40702 | 0.366928 | 0.999532 | | 2.232263 | -0.03346 | 0.98887 | | 2.227528 | -0.38272 | 0.80953 |
| HRAS | 4.132219 | 0.237463 | 0.999532 | | 4.273134 | -0.0249 | 0.98887 | | 4.034498 | -0.24423 | 0.849682 |
| TMEM39B | 2.470769 | 0.0133 | 0.999532 | | 2.677865 | -0.03098 | 0.98887 | | 2.376484 | -0.02575 | 0.993435 |
| TLCD5 | 2.616691 | 0.814119 | 0.999532 | | 2.172806 | -0.035 | 0.98887 | | 2.528423 | -0.83151 | 0.505783 |
| AHNAK2 | 2.676306 | 0.145355 | 0.999532 | | 2.79433 | -0.0318 | 0.98887 | | 2.690486 | -0.14981 | 0.941197 |
| MTRR | 3.12956 | -0.25019 | 0.999532 | | 3.773525 | -0.02716 | 0.98887 | | 3.060612 | 0.235738 | 0.881951 |
| NHSL1 | 2.810103 | 0.064131 | 0.999532 | | 2.990736 | 0.030051 | 0.988882 | | 2.634966 | -0.02219 | 0.995112 |
| PSME4 | 3.583328 | -0.24637 | 0.999532 | | 4.387102 | 0.025531 | 0.988916 | | 3.4547 | 0.291962 | 0.827267 |
| CHMP1A | 6.073377 | -0.29969 | 0.999532 | | 8.043588 | 0.018726 | 0.988916 | | 6.096003 | 0.351036 | 0.706749 |
| BAHCC1 | 2.789261 | -0.19529 | 0.999532 | | 3.119345 | 0.028388 | 0.988916 | | 2.593641 | 0.238259 | 0.88476 |
| NCLN | 2.98799 | -0.08071 | 0.999532 | | 3.416443 | -0.02817 | 0.988916 | | 2.873459 | 0.070129 | 0.974331 |
| OTUD5 | 6.582137 | -0.47839 | 0.999532 | | 8.62228 | -0.01933 | 0.988916 | | 6.132108 | 0.48413 | 0.563885 |

| | | | | | | | | | | | |
|---|---|---|---|---|---|---|---|---|---|---|---|
| BRPF3 | 4.181563 | 0.318062 | 0.999532 | | 4.059839 | -0.02463 | 0.988916 | | 3.819322 | -0.3246 | 0.791074 |
| NCDN | 2.988314 | -0.08381 | 0.999532 | | 3.453621 | 0.027 | 0.988916 | | 2.929537 | 0.128218 | 0.942744 |
| POMT2 | 2.829734 | 0.316838 | 0.999532 | | 2.826849 | -0.02899 | 0.988916 | | 2.646453 | -0.32717 | 0.819366 |
| CIP2A | 2.038723 | 0.172267 | 0.999532 | | 2.249549 | -0.03453 | 0.988916 | | 1.982764 | -0.18673 | 0.926586 |
| HOXA9 | 5.041334 | -0.30228 | 0.999532 | | 6.192473 | 0.021004 | 0.988974 | | 4.772973 | 0.342319 | 0.738861 |
| IPP | 2.184675 | -0.37041 | 0.999532 | | 2.82762 | 0.028826 | 0.988974 | | 2.137143 | 0.41915 | 0.790999 |
| TRIM21 | 4.465413 | 0.000934 | 0.999532 | | 5.148807 | 0.022862 | 0.988974 | | 4.352513 | 0.042849 | 0.981929 |
| SPA17 | 2.163802 | 0.217535 | 0.999532 | | 2.414594 | -0.03366 | 0.988974 | | 2.254079 | -0.23116 | 0.903054 |
| NCAPD3 | 2.732835 | 0.051088 | 0.999532 | | 2.931337 | 0.029495 | 0.988974 | | 2.611017 | -0.0016 | 0.999429 |
| ZNF557 | 1.954916 | 0.200372 | 0.999532 | | 2.228493 | -0.03443 | 0.988974 | | 1.877457 | -0.20836 | 0.91707 |
| ARMCX5 | 1.947996 | 0.10882 | 0.999532 | | 2.216959 | 0.033698 | 0.988974 | | 1.933793 | -0.05363 | 0.985693 |
| OR4C3 | 2.10435 | 0.38858 | 0.999532 | | 2.198911 | -0.03554 | 0.988974 | | 2.095876 | -0.40378 | 0.815713 |
| FMR1 | 5.69802 | 0.113517 | 0.999532 | | 6.002073 | -0.02189 | 0.988974 | | 5.44129 | -0.11806 | 0.928441 |
| RNF180 | 2.512778 | -0.17423 | 0.999532 | | 3.027868 | -0.03017 | 0.988974 | | 2.30381 | 0.166427 | 0.929549 |
| PIH1D1 | 3.787383 | -0.09433 | 0.999532 | | 4.401424 | 0.023061 | 0.988974 | | 3.54809 | 0.130677 | 0.936689 |
| NPHP4 | 2.381696 | 0.092983 | 0.999532 | | 2.780639 | -0.03152 | 0.988974 | | 2.331098 | -0.10252 | 0.964434 |
| CLTCL1 | 1.780252 | -0.26137 | 0.999532 | | 2.102359 | -0.0336 | 0.988974 | | 1.747849 | 0.245459 | 0.905952 |
| NAF1 | 3.415361 | 0.151222 | 0.999532 | | 3.647778 | 0.02893 | 0.988974 | | 3.351043 | -0.11289 | 0.953949 |
| CATSPEF | 2.694108 | 0.386225 | 0.999532 | | 2.547149 | -0.0316 | 0.988974 | | 2.589341 | -0.4046 | 0.799944 |
| ENDOG | 4.51487 | 0.297101 | 0.999532 | | 4.352256 | -0.02457 | 0.988974 | | 4.086802 | -0.30257 | 0.800239 |
| DRD4 | 5.224642 | 0.118797 | 0.999532 | | 5.790858 | -0.02435 | 0.988974 | | 5.00488 | -0.1223 | 0.931405 |
| VIRMA | 3.973948 | -0.04071 | 0.999532 | | 4.693922 | 0.024114 | 0.988974 | | 3.984961 | 0.08005 | 0.962716 |
| CFAP206 | 2.033562 | 0.299805 | 0.999532 | | 2.133674 | 0.034801 | 0.988974 | | 2.027994 | -0.24722 | 0.899022 |
| MXRA5 | 1.844542 | 0.057795 | 0.999532 | | 2.155059 | 0.033552 | 0.98904 | | 1.892766 | -0.0054 | 0.998655 |
| NKX2-3 | 1.803931 | -0.37512 | 0.999532 | | 2.545629 | 0.034424 | 0.98915 | | 1.939792 | 0.442086 | 0.817234 |
| CCDC88C | 3.866563 | 0.210618 | 0.999532 | | 3.850086 | 0.026294 | 0.989239 | | 3.828931 | -0.18881 | 0.901813 |
| SMIM4 | 3.367329 | 0.109756 | 0.999532 | | 3.686047 | -0.02642 | 0.989303 | | 3.368336 | -0.12013 | 0.945129 |
| COPZ2 | 2.868991 | -0.0932 | 0.999532 | | 3.619815 | 0.027064 | 0.989303 | | 3.000931 | 0.140812 | 0.938429 |
| DDX19A | 4.667642 | -0.15869 | 0.999532 | | 5.563593 | -0.02332 | 0.989303 | | 4.47789 | 0.159505 | 0.911296 |
| TNFAIP8L | 1.933071 | -0.27172 | 0.999532 | | 2.256598 | 0.033408 | 0.989311 | | 1.886072 | 0.319532 | 0.877943 |
| C14orf17 | 1.969696 | 0.397996 | 0.999532 | | 1.942663 | -0.03479 | 0.989311 | | 1.90713 | -0.41395 | 0.809832 |
| TMEM20 | 3.343562 | -0.24849 | 0.999532 | | 3.974532 | 0.025211 | 0.98937 | | 3.182286 | 0.296977 | 0.834867 |
| ATP8B2 | 3.775032 | -0.0285 | 0.999532 | | 4.218118 | -0.02493 | 0.989412 | | 3.842697 | 0.028608 | 0.991134 |
| MAK16 | 1.864374 | 0.20105 | 0.999532 | | 1.885055 | -0.03367 | 0.989452 | | 1.809227 | -0.21949 | 0.913432 |
| ZSCAN20 | 2.438334 | 0.178226 | 0.999532 | | 2.66058 | -0.0319 | 0.989452 | | 2.400997 | -0.18528 | 0.92697 |
| AOAH | 2.688122 | 0.54995 | 0.999532 | | 2.327776 | -0.03076 | 0.989461 | | 2.53396 | -0.57965 | 0.683846 |
| THOC7 | 3.514603 | -0.27777 | 0.999532 | | 4.007055 | 0.024164 | 0.989461 | | 3.195587 | 0.310707 | 0.82198 |
| C7orf26 | 4.403801 | 0.144472 | 0.999532 | | 4.657282 | 0.025399 | 0.989461 | | 4.016066 | -0.08116 | 0.965649 |
| RMDN3 | 4.085893 | 0.189895 | 0.999532 | | 4.31423 | 0.023474 | 0.989461 | | 4.038471 | -0.14972 | 0.914929 |
| FGFR2 | 2.397681 | 0.514084 | 0.999532 | | 2.159335 | -0.03297 | 0.989461 | | 2.239829 | -0.52948 | 0.722363 |
| RNASE7 | 3.408087 | 0.881953 | 0.999532 | | 2.79444 | 0.028824 | 0.989461 | | 3.405332 | -0.83341 | 0.415282 |
| LRFN4 | 3.708839 | 0.161691 | 0.999532 | | 3.880289 | 0.024378 | 0.989515 | | 3.374832 | -0.13375 | 0.935427 |
| RPP38 | 1.99865 | -0.2049 | 0.999532 | | 2.385002 | -0.03157 | 0.989649 | | 1.989198 | 0.192959 | 0.925336 |
| DUS2 | 2.046038 | 0.499783 | 0.999532 | | 2.075808 | 0.033205 | 0.989649 | | 2.098666 | -0.44793 | 0.779652 |
| NPFFR1 | 3.623337 | 0.656625 | 0.999532 | | 3.371141 | 0.027643 | 0.989649 | | 3.771883 | -0.60949 | 0.608189 |
| C16orf46 | 2.301421 | 0.259739 | 0.999532 | | 2.142411 | -0.0325 | 0.989699 | | 2.03759 | -0.27867 | 0.881837 |
| ATP13A2 | 3.067277 | 0.066431 | 0.999532 | | 3.580968 | 0.02609 | 0.989699 | | 2.971606 | -0.02291 | 0.993618 |
| PAK5 | 2.29375 | 0.167831 | 0.999532 | | 2.306284 | -0.03189 | 0.989699 | | 2.190051 | -0.18339 | 0.929613 |
| RAB34 | 6.365303 | 0.082925 | 0.999532 | | 6.897588 | 0.018478 | 0.989699 | | 6.159454 | -0.04657 | 0.974421 |
| NEK8 | 3.34304 | 0.47709 | 0.999532 | | 3.293123 | 0.026415 | 0.989745 | | 3.171848 | -0.43833 | 0.713076 |
| GABRR2 | 1.857716 | 0.273564 | 0.999532 | | 2.035209 | 0.034221 | 0.989745 | | 1.85089 | -0.21924 | 0.914929 |
| DMAC2 | 4.130651 | -0.09304 | 0.999532 | | 4.80873 | 0.022486 | 0.989745 | | 4.051323 | 0.133903 | 0.927544 |
| TRIM54 | 2.581009 | 0.319284 | 0.999532 | | 2.655754 | -0.0292 | 0.989745 | | 2.532085 | -0.32755 | 0.836935 |
| ARHGAP | 2.698436 | 0.381686 | 0.999532 | | 2.395757 | 0.029284 | 0.989745 | | 2.54191 | -0.33721 | 0.831349 |
| DHRS4 | 6.978239 | -0.24529 | 0.999532 | | 8.03375 | -0.0182 | 0.989832 | | 6.407688 | 0.233383 | 0.836453 |
| SMIM11A | 3.066625 | -0.41452 | 0.999532 | | 3.813745 | -0.02411 | 0.989832 | | 2.945945 | 0.412739 | 0.763806 |
| BCLAF3 | 2.822727 | 0.452704 | 0.999532 | | 2.820216 | 0.026663 | 0.989832 | | 2.634517 | -0.40817 | 0.757904 |
| IL6 | 2.041566 | 0.483157 | 0.999532 | | 1.881795 | -0.03229 | 0.989832 | | 2.021423 | -0.49759 | 0.754186 |
| SLX1A | 2.445523 | 0.400237 | 0.999532 | | 2.405489 | 0.027917 | 0.989832 | | 2.313136 | -0.35434 | 0.816618 |
| EAF1 | 2.177997 | -0.3445 | 0.999532 | | 2.494624 | -0.03059 | 0.989832 | | 1.906585 | 0.336919 | 0.859522 |
| TRPS1 | 2.329894 | 0.132617 | 0.999532 | | 2.578816 | 0.027891 | 0.989832 | | 2.274897 | -0.08663 | 0.969693 |
| PRKCI | 11.6943 | 0.19569 | 0.999532 | | 12.05033 | -0.01644 | 0.989832 | | 11.01082 | -0.18588 | 0.825181 |
| DISP1 | 2.624166 | 0.411339 | 0.999532 | | 2.826021 | 0.028434 | 0.989832 | | 2.608674 | -0.36236 | 0.806394 |
| STARD13 | 4.471772 | -0.23103 | 0.999532 | | 5.851495 | 0.019915 | 0.989832 | | 4.52483 | 0.273744 | 0.815899 |
| DOK4 | 4.194805 | 0.328259 | 0.999532 | | 4.223477 | -0.02173 | 0.989832 | | 3.897315 | -0.33428 | 0.776954 |
| APBB3 | 9.709271 | 0.13165 | 0.999532 | | 10.80096 | -0.01576 | 0.989832 | | 9.395992 | -0.13019 | 0.891051 |
| MRPS2 | 3.670407 | 0.152107 | 0.999532 | | 3.788422 | 0.022774 | 0.989832 | | 3.414258 | -0.11299 | 0.94366 |
| EXPH5 | 5.057858 | 0.285517 | 0.999532 | | 5.048046 | -0.02207 | 0.989832 | | 4.785744 | -0.28851 | 0.801572 |
| PPIP5K2 | 3.640136 | 0.206044 | 0.999532 | | 4.066821 | 0.023371 | 0.989832 | | 3.683083 | -0.16774 | 0.906902 |
| NOL7 | 5.722074 | 0.219085 | 0.999532 | | 6.142538 | -0.01974 | 0.989832 | | 5.396482 | -0.20737 | 0.873936 |
| NDUFAF7 | 3.124153 | -0.09181 | 0.999532 | | 3.637233 | 0.025279 | 0.989832 | | 3.052933 | 0.133945 | 0.939913 |
| DUSP18 | 3.717716 | 0.164435 | 0.999532 | | 4.345414 | 0.02531 | 0.989832 | | 3.801077 | -0.10391 | 0.954538 |
| PANK2 | 7.141933 | 0.056567 | 0.999532 | | 7.910271 | 0.017155 | 0.989832 | | 6.95186 | -0.02149 | 0.991103 |
| BLOC1S2 | 3.319958 | 0.003725 | 0.999532 | | 3.80493 | -0.02272 | 0.989832 | | 3.277845 | -0.00517 | 0.998401 |
| ACACA | 2.608095 | -0.17771 | 0.999532 | | 2.995557 | -0.02569 | 0.989832 | | 2.387571 | 0.165819 | 0.92697 |
| HORMAD | 2.56266 | 0.359731 | 0.999532 | | 2.483933 | -0.03235 | 0.989832 | | 2.450596 | -0.36926 | 0.816618 |
| C9orf116 | 1.887735 | -0.18711 | 0.999532 | | 2.340134 | 0.029991 | 0.989832 | | 1.861364 | 0.238729 | 0.909259 |
| HIP1R | 3.857341 | 0.430495 | 0.999532 | | 3.657422 | -0.0236 | 0.989832 | | 3.667139 | -0.43533 | 0.704316 |
| SLC25A2 | 4.030721 | 0.480564 | 0.999532 | | 3.798731 | -0.02374 | 0.989832 | | 3.979996 | -0.48966 | 0.657467 |
| UMPS | 2.746641 | -0.44004 | 0.999532 | | 3.53141 | -0.02449 | 0.989832 | | 2.572409 | 0.43327 | 0.75311 |
| RINT1 | 2.626573 | 0.145282 | 0.999532 | | 2.654759 | 0.029305 | 0.989832 | | 2.453212 | -0.11291 | 0.959437 |
| ITSN1 | 3.519822 | -0.1698 | 0.999532 | | 4.330069 | -0.02191 | 0.989832 | | 3.392849 | 0.16752 | 0.907626 |
| NXPH3 | 2.136001 | 0.324589 | 0.999532 | | 2.184557 | -0.03318 | 0.989832 | | 2.10623 | -0.33887 | 0.861119 |
| VBP1 | 2.962542 | -0.29666 | 0.999532 | | 3.480656 | -0.02448 | 0.989832 | | 2.694146 | 0.291063 | 0.845575 |
| SEC24C | 5.560367 | 0.011477 | 0.999532 | | 6.164986 | 0.018606 | 0.989832 | | 5.370553 | 0.026725 | 0.9894 |
| TMX3 | 12.03317 | -0.15978 | 0.999532 | | 14.60383 | 0.014311 | 0.989832 | | 11.98343 | 0.19626 | 0.814133 |
| ZNF76 | 3.619411 | -0.24799 | 0.999532 | | 4.445324 | 0.022544 | 0.989832 | | 3.43359 | 0.292496 | 0.839943 |
| PHETA2 | 3.587542 | 0.034806 | 0.999532 | | 4.221433 | 0.023241 | 0.989832 | | 3.542456 | 0.017942 | 0.995943 |
| SDAD1 | 3.179774 | -0.03554 | 0.999532 | | 3.812156 | 0.024505 | 0.989832 | | 3.266009 | 0.081373 | 0.967145 |
| PTER | 2.0349 | 0.450819 | 0.999532 | | 1.878879 | 0.030803 | 0.989832 | | 2.02808 | -0.40478 | 0.806797 |
| GNPTAB | 5.888507 | -0.06104 | 0.999532 | | 6.347795 | 0.01856 | 0.989832 | | 5.568627 | 0.098389 | 0.94378 |
| ZNF3 | 2.825424 | 0.055931 | 0.999532 | | 3.170156 | 0.025727 | 0.989832 | | 2.738485 | -0.01141 | 0.996891 |
| CHMP6 | 6.254151 | 0.021036 | 0.999532 | | 6.785037 | -0.01893 | 0.989832 | | 5.93267 | -0.02274 | 0.991159 |
| RAET1E | 2.053719 | 0.486569 | 0.999532 | | 1.898297 | -0.0317 | 0.989832 | | 2.043426 | -0.50123 | 0.750417 |
| KCNK12 | 2.08682 | 0.473284 | 0.999532 | | 1.926742 | 0.031016 | 0.989832 | | 2.081994 | -0.4229 | 0.799157 |
| KIF1C | 2.331761 | 0.254027 | 0.999532 | | 2.418953 | 0.028211 | 0.989832 | | 2.25127 | -0.20856 | 0.907144 |
| UFSP1 | 2.534716 | 0.43941 | 0.999532 | | 2.377954 | -0.02961 | 0.989832 | | 2.403858 | -0.44985 | 0.756633 |
| MPPED1 | 1.909067 | -0.01053 | 0.999532 | | 2.107506 | -0.03206 | 0.989832 | | 1.797226 | -0.00295 | 0.999385 |
| TSPY1 | 4.817897 | -0.96046 | 0.999532 | | 7.463431 | 0.044032 | 0.989832 | | 4.751991 | 1.045024 | 0.590692 |
| CCR6 | 2.31855 | 0.271108 | 0.999532 | | 2.343271 | -0.03038 | 0.989832 | | 2.244709 | -0.28282 | 0.876107 |
| ADGRF5 | 17.35143 | -0.40483 | 0.999532 | | 22.07924 | -0.01687 | 0.989832 | | 16.39432 | 0.40983 | 0.563461 |
| BBS10 | 2.052074 | -0.07544 | 0.999532 | | 2.292615 | 0.030723 | 0.989832 | | 2.026714 | 0.119629 | 0.960773 |
| EEPD1 | 2.41311 | 0.2123 | 0.999532 | | 2.642424 | 0.027903 | 0.989832 | | 2.446888 | -0.16671 | 0.92697 |
| ARHGEF | 1.963111 | 0.149824 | 0.999532 | | 2.226222 | -0.0314 | 0.989832 | | 1.891199 | -0.1563 | 0.943768 |
| ZNF322 | 3.090504 | -0.2889 | 0.999532 | | 3.91114 | -0.0244 | 0.989832 | | 2.97448 | 0.2837 | 0.849021 |
| TRMT13 | 3.748946 | -0.07965 | 0.999532 | | 4.21206 | -0.02351 | 0.989832 | | 3.55259 | 0.074852 | 0.972221 |
| TEAD4 | 3.44767 | -0.33517 | 0.999532 | | 4.437965 | -0.02204 | 0.989925 | | 3.455985 | 0.334096 | 0.801572 |
| KCMF1 | 6.666184 | -0.12042 | 0.999532 | | 7.903991 | 0.01756 | 0.989973 | | 6.408259 | 0.157832 | 0.890109 |
| FTSJ3 | 3.967808 | -0.06412 | 0.999532 | | 4.538797 | -0.02143 | 0.989973 | | 3.813965 | 0.05994 | 0.974421 |
| TMEM50B | 18.95598 | -0.17155 | 0.999532 | | 22.60247 | -0.01122 | 0.989973 | | 17.95896 | 0.18317 | 0.787995 |
| STON1 | 4.858326 | 0.473182 | 0.999532 | | 4.609344 | 0.023546 | 0.98999 | | 4.500721 | -0.42004 | 0.717627 |
| AGBL5 | 2.42559 | 0.072509 | 0.999532 | | 2.54117 | 0.02628 | 0.990177 | | 2.308309 | -0.03314 | 0.991273 |
| SAP130 | 2.536554 | 0.313053 | 0.999532 | | 2.531954 | 0.026822 | 0.990177 | | 2.457066 | -0.26826 | 0.872561 |
| CMTM8 | 3.505407 | -0.00018 | 0.99982 | | 3.880435 | -0.02385 | 0.990177 | | 3.249081 | -0.0022 | 0.999385 |
| ASB9 | 2.949184 | -0.30331 | 0.999532 | | 3.605299 | -0.02293 | 0.990177 | | 2.833134 | 0.300275 | 0.845724 |
| SMIM29 | 3.015304 | -0.07835 | 0.999532 | | 3.494977 | -0.02459 | 0.990177 | | 2.865675 | 0.073549 | 0.973754 |
| FMO4 | 2.966635 | 0.634777 | 0.999532 | | 2.68029 | 0.027083 | 0.990177 | | 2.813484 | -0.58215 | 0.647581 |
| LAPTM5 | 4.305165 | -0.59312 | 0.999532 | | 5.525281 | 0.023425 | 0.990177 | | 4.042913 | 0.595748 | 0.652925 |
| USP21 | 4.582054 | 0.250409 | 0.999532 | | 4.658452 | 0.02064 | 0.9904 | | 4.488792 | -0.20998 | 0.876258 |
| C1orf105 | 2.223955 | 0.604319 | 0.999532 | | 2.085402 | 0.030356 | 0.9904 | | 2.289229 | -0.54893 | 0.722703 |
| BNIP1 | 2.6546 | 0.05236 | 0.999532 | | 2.854701 | -0.02481 | 0.9904 | | 2.494384 | -0.05969 | 0.978282 |
| IFRD2 | 3.174424 | 0.034946 | 0.999532 | | 3.534409 | -0.02377 | 0.990515 | | 2.977952 | -0.04008 | 0.988043 |
| FHIP1B | 4.570004 | -0.26813 | 0.999532 | | 5.207965 | 0.019489 | 0.990593 | | 4.34395 | 0.292032 | 0.817471 |
| NRF1 | 2.386221 | 0.114763 | 0.999532 | | 2.530767 | 0.026946 | 0.990593 | | 2.390847 | -0.07592 | 0.974421 |
| SH3D19 | 7.05565 | -0.09069 | 0.999532 | | 7.691231 | -0.01958 | 0.990593 | | 6.64407 | 0.070486 | 0.964434 |
| KIF19 | 2.078261 | 0.674833 | 0.999532 | | 1.709318 | 0.032051 | 0.9906 | | 1.933389 | -0.62659 | 0.686214 |
| SLPI | 3.30251 | 0.766188 | 0.999532 | | 2.881556 | -0.02645 | 0.99065 | | 3.103905 | -0.75589 | 0.534789 |
| MOB3B | 2.013811 | 0.31411 | 0.999532 | | 1.971799 | -0.02745 | 0.990694 | | 1.93447 | -0.32449 | 0.858899 |
| EXD1 | 2.010849 | 0.410314 | 0.999532 | | 1.988318 | 0.027389 | 0.990694 | | 2.016036 | -0.36534 | 0.833604 |
| SLFN5 | 5.962018 | -0.29183 | 0.999532 | | 7.368043 | -0.01528 | 0.990694 | | 5.540724 | 0.296501 | 0.761337 |
| REPS2 | 3.495077 | 0.042922 | 0.999532 | | 3.675231 | -0.02384 | 0.990694 | | 3.060868 | -0.01527 | 0.996681 |
| TSNAX | 4.611085 | -0.12672 | 0.999532 | | 5.308505 | -0.01977 | 0.990694 | | 4.435446 | 0.125206 | 0.931837 |
| SDC1 | 2.175545 | 0.150748 | 0.999532 | | 2.364045 | -0.02674 | 0.990694 | | 2.069011 | -0.15887 | 0.939549 |
| SUGP1 | 3.544041 | 0.31869 | 0.999532 | | 3.235805 | 0.021717 | 0.990694 | | 3.32783 | -0.28432 | 0.843564 |
| STRN | 2.308545 | 0.073051 | 0.999532 | | 2.536484 | -0.02395 | 0.990694 | | 2.158228 | -0.07877 | 0.972758 |
| ETV3 | 8.945261 | -0.32661 | 0.999532 | | 10.91106 | -0.01561 | 0.990694 | | 8.539289 | 0.328465 | 0.717627 |
| NEK10 | 1.801912 | -0.19162 | 0.999532 | | 2.005182 | -0.02837 | 0.990694 | | 1.727558 | 0.181156 | 0.938429 |
| MT1E | 11.4639 | -0.48743 | 0.999532 | | 15.35251 | 0.016836 | 0.990694 | | 10.67756 | 0.567523 | 0.504121 |
| WDR44 | 2.698947 | 0.017585 | 0.999532 | | 2.968467 | 0.024695 | 0.990694 | | 2.50661 | 0.030845 | 0.991452 |
| KCNK4 | 5.796771 | -0.36047 | 0.999532 | | 7.292444 | 0.017407 | 0.990694 | | 5.739013 | 0.386427 | 0.723933 |
| CCDC103 | 2.079941 | 0.138746 | 0.999532 | | 2.208841 | 0.028621 | 0.990694 | | 2.053056 | -0.09094 | 0.972305 |
| CDC25C | 2.054826 | -0.07853 | 0.999532 | | 2.487695 | -0.02735 | 0.990694 | | 2.035918 | 0.070066 | 0.978927 |
| KLHL33 | 1.862655 | 0.516595 | 0.999532 | | 1.871716 | -0.03029 | 0.990694 | | 1.915524 | -0.52495 | 0.747677 |
| POC5 | 3.219588 | 0.325743 | 0.999532 | | 3.301768 | 0.022763 | 0.990694 | | 3.118645 | -0.28508 | 0.842754 |
| CYREN | 2.989751 | -0.12244 | 0.999532 | | 3.514584 | 0.021266 | 0.990694 | | 2.839105 | 0.161362 | 0.922437 |
| PWWP2B | 2.228058 | -0.03651 | 0.999532 | | 2.441314 | 0.025596 | 0.990694 | | 2.140905 | 0.077493 | 0.974421 |
| POLR1H | 2.754431 | 0.576048 | 0.999532 | | 2.508179 | -0.02538 | 0.990694 | | 2.579207 | -0.58161 | 0.663633 |
| CHAC1 | 2.706164 | 0.716144 | 0.999532 | | 2.157821 | -0.02621 | 0.990694 | | 2.572424 | -0.72538 | 0.566578 |
| DPYSL2 | 11.83711 | -0.30175 | 0.999532 | | 14.61676 | -0.0136 | 0.990694 | | 11.45996 | 0.304828 | 0.669465 |
| CHRNA3 | 2.096264 | 0.058414 | 0.999532 | | 2.155623 | -0.0278 | 0.990694 | | 1.998143 | -0.06991 | 0.978282 |
| FOXRED2 | 2.38947 | 0.110854 | 0.999532 | | 2.571751 | -0.02438 | 0.990694 | | 2.307985 | -0.11543 | 0.955834 |
| VPS4A | 5.687492 | -0.22322 | 0.999532 | | 6.807183 | -0.01642 | 0.990694 | | 5.345658 | 0.224995 | 0.836935 |
| ITGA9 | 2.258529 | -0.10586 | 0.999532 | | 2.654884 | -0.02608 | 0.990694 | | 2.301298 | 0.097492 | 0.96588 |
| PYGO1 | 1.929229 | 0.520066 | 0.999532 | | 1.895933 | -0.02995 | 0.990694 | | 1.979017 | -0.53046 | 0.736481 |
| MRPL45 | 3.016192 | -0.44783 | 0.999532 | | 4.086591 | 0.020674 | 0.990694 | | 2.916659 | 0.50296 | 0.702501 |
| TMEM268 | 3.051618 | -0.17196 | 0.999532 | | 3.564175 | -0.02311 | 0.990694 | | 2.746225 | 0.174058 | 0.919181 |
| MXI1 | 2.772657 | 0.077063 | 0.999532 | | 3.208994 | 0.02303 | 0.990694 | | 2.704158 | -0.03648 | 0.989437 |
| CILP2 | 2.011352 | 0.323646 | 0.999532 | | 2.02397 | 0.030489 | 0.990694 | | 1.97684 | -0.27749 | 0.889126 |
| PTEN | 4.593995 | -0.55244 | 0.999532 | | 5.873241 | 0.018091 | 0.990694 | | 4.309355 | 0.591313 | 0.540342 |
| EPSTI1 | 3.343741 | 0.031408 | 0.999532 | | 3.633597 | 0.021424 | 0.990694 | | 3.234647 | 0.009867 | 0.996891 |
| RIOK3 | 3.537444 | -0.02346 | 0.999532 | | 4.440785 | -0.02109 | 0.990694 | | 3.601673 | 0.075818 | 0.972221 |
| FGFR1OP | 3.878283 | 0.191379 | 0.999532 | | 4.063874 | -0.02034 | 0.990694 | | 3.676684 | -0.19414 | 0.889853 |
| PLAAT4 | 8.323559 | -0.18481 | 0.999532 | | 9.765234 | -0.01427 | 0.990694 | | 7.985568 | 0.191432 | 0.844931 |
| RABL2B | 4.706186 | 0.561601 | 0.999532 | | 4.264635 | 0.020602 | 0.990694 | | 4.500106 | -0.52654 | 0.636184 |
| CCNC | 4.57114 | -0.3851 | 0.999532 | | 5.971799 | -0.01737 | 0.990694 | | 4.231074 | 0.381354 | 0.723933 |
| ZCCHC12 | 2.281992 | 0.164525 | 0.999532 | | 2.597538 | 0.028278 | 0.990694 | | 2.461736 | -0.11409 | 0.963866 |
| C3orf85 | 2.258353 | 0.31158 | 0.999532 | | 2.187312 | 0.02719 | 0.990694 | | 2.191453 | -0.26168 | 0.885078 |
| TRMU | 2.419673 | 0.183064 | 0.999532 | | 2.690669 | -0.02413 | 0.990694 | | 2.239345 | -0.18328 | 0.918194 |
| JMJD8 | 7.055798 | 0.021551 | 0.999532 | | 8.017391 | -0.01763 | 0.990694 | | 6.416248 | -0.00947 | 0.996891 |
| FBXL16 | 2.857203 | 1.113046 | 0.950261 | | 2.070333 | -0.02784 | 0.990694 | | 2.809718 | -1.12019 | 0.282041 |
| DENND6A | 5.09608 | -0.62388 | 0.999532 | | 7.104546 | -0.01644 | 0.990694 | | 4.901174 | 0.628766 | 0.462548 |
| PSMG4 | 4.586609 | 0.185317 | 0.999532 | | 4.943743 | 0.019573 | 0.990694 | | 4.129049 | -0.13546 | 0.926506 |
| HROB | 2.392357 | 0.497826 | 0.999532 | | 2.417166 | -0.02878 | 0.990694 | | 2.400239 | -0.49126 | 0.760572 |
| ZNF200 | 2.523034 | 0.468549 | 0.999532 | | 2.408173 | 0.026501 | 0.990694 | | 2.494395 | -0.41608 | 0.782543 |
| TMEM125 | 1.915825 | 0.141255 | 0.999532 | | 2.016497 | 0.027969 | 0.990694 | | 1.91164 | -0.09647 | 0.970811 |
| PHLPP1 | 4.659006 | -0.28183 | 0.999532 | | 6.117731 | 0.017476 | 0.990694 | | 4.668427 | 0.32683 | 0.771839 |
| ABHD17A | 16.5115 | -0.02822 | 0.999532 | | 18.55707 | 0.010386 | 0.990694 | | 15.79709 | 0.058048 | 0.947322 |
| ZKSCAN3 | 4.342836 | 0.32821 | 0.999532 | | 4.346812 | 0.021601 | 0.990694 | | 4.116957 | -0.28658 | 0.830822 |
| WDR31 | 2.059627 | 0.333853 | 0.999532 | | 2.139413 | 0.0267 | 0.990694 | | 2.006911 | -0.287 | 0.870663 |
| ASCL3 | 2.70902 | 0.596093 | 0.999532 | | 2.499234 | -0.02585 | 0.990694 | | 2.534544 | -0.60156 | 0.652925 |
| RGS4 | 2.106609 | 0.072634 | 0.999532 | | 2.298031 | -0.029 | 0.990694 | | 2.032977 | -0.08325 | 0.974421 |
| POLR1G | 2.604408 | 0.11172 | 0.999532 | | 2.659619 | 0.026284 | 0.990694 | | 2.440862 | -0.07683 | 0.974421 |
| THUMPD2 | 2.672513 | 0.152368 | 0.999532 | | 2.907496 | 0.024053 | 0.990694 | | 2.724453 | -0.09741 | 0.96556 |
| OR13J1 | 2.49543 | 0.359247 | 0.999532 | | 2.728497 | -0.02697 | 0.990694 | | 2.475412 | -0.35453 | 0.836453 |
| KBTBD7 | 2.801612 | -0.31685 | 0.999532 | | 3.474019 | 0.021573 | 0.990694 | | 2.690826 | 0.357282 | 0.805825 |
| CYP4F12 | 3.707902 | 0.203776 | 0.999532 | | 3.937394 | 0.021632 | 0.990793 | | 3.45758 | -0.15859 | 0.917499 |
| DBI | 4.474613 | -0.24203 | 0.999532 | | 5.432098 | -0.01717 | 0.990793 | | 4.388368 | 0.244266 | 0.839373 |
| CIBAR1 | 4.054865 | -0.13509 | 0.999532 | | 4.942549 | -0.01868 | 0.990793 | | 3.892902 | 0.14076 | 0.923834 |
| TMEM41A | 1.98723 | 0.010865 | 0.999532 | | 2.195421 | -0.02648 | 0.990793 | | 1.94434 | -0.01842 | 0.996681 |
| BGLAP | 2.694967 | 0.07384 | 0.999532 | | 3.126625 | 0.02204 | 0.990793 | | 2.633672 | -0.03265 | 0.991103 |
| SMIM13 | 2.348475 | -0.00499 | 0.999532 | | 2.590461 | -0.02402 | 0.990793 | | 2.219108 | -0.00609 | 0.99853 |
| HAUS2 | 1.769587 | -0.22645 | 0.999532 | | 2.192918 | -0.02654 | 0.990816 | | 1.754711 | 0.220866 | 0.918082 |
| MOB2 | 3.972591 | 0.227831 | 0.999532 | | 4.092816 | -0.01923 | 0.990816 | | 3.77742 | -0.22943 | 0.861119 |
| CGRRF1 | 3.274529 | 0.384837 | 0.999532 | | 3.280368 | -0.02108 | 0.990816 | | 3.114947 | -0.38856 | 0.749206 |
| FDX2 | 2.119688 | -0.01913 | 0.999532 | | 2.279653 | 0.024472 | 0.990826 | | 1.98598 | 0.055985 | 0.98556 |
| TNPO1 | 7.911309 | -0.31264 | 0.999532 | | 9.407782 | 0.013308 | 0.990826 | | 7.361542 | 0.340858 | 0.701419 |
| TFDP2 | 2.634778 | -0.16334 | 0.999532 | | 3.039682 | 0.022225 | 0.990826 | | 2.576222 | 0.20112 | 0.910574 |
| PET117 | 3.890365 | 0.106355 | 0.999532 | | 3.995007 | 0.018469 | 0.990826 | | 3.608095 | -0.07979 | 0.965649 |
| ZMYND8 | 2.880299 | 0.109399 | 0.999532 | | 3.085746 | 0.022103 | 0.990826 | | 2.702608 | -0.07188 | 0.974421 |
| NUDT18 | 3.793409 | 0.26364 | 0.999532 | | 3.702752 | 0.020562 | 0.990826 | | 3.742658 | -0.23039 | 0.881689 |
| RHBDD1 | 3.737991 | 0.020379 | 0.999532 | | 4.325972 | 0.019945 | 0.990826 | | 3.525184 | -0.00217 | 0.999385 |
| LRIG1 | 3.28164 | 0.198652 | 0.999532 | | 3.312993 | 0.020771 | 0.990826 | | 3.035358 | -0.16222 | 0.920486 |
| ZNRF4 | 2.649088 | 0.030661 | 0.999532 | | 2.917564 | 0.023462 | 0.990826 | | 2.491833 | 0.016905 | 0.996681 |
| OR1S1 | 2.317646 | 0.265911 | 0.999532 | | 2.404933 | -0.02551 | 0.990826 | | 2.264994 | -0.27417 | 0.883588 |
| TLCD1 | 1.904248 | 0.48977 | 0.999532 | | 1.904389 | 0.027078 | 0.990826 | | 1.90254 | -0.443 | 0.790283 |
| CLCNKB | 3.593395 | 0.521979 | 0.999532 | | 3.419809 | -0.02174 | 0.990826 | | 3.321974 | -0.54354 | 0.694677 |
| ATP2A3 | 6.07256 | -0.1653 | 0.999532 | | 6.788837 | -0.01595 | 0.990826 | | 5.702371 | 0.139477 | 0.920487 |
| TMEM263 | 5.941288 | 0.112362 | 0.999532 | | 6.794832 | -0.01475 | 0.990826 | | 5.638366 | -0.09749 | 0.941197 |
| TTC27 | 3.297352 | 0.162781 | 0.999532 | | 3.392456 | 0.019796 | 0.990826 | | 3.189986 | -0.1195 | 0.943248 |
| ZDHHC14 | 2.416741 | -0.04093 | 0.999532 | | 2.671949 | 0.022114 | 0.990826 | | 2.340758 | 0.082735 | 0.971797 |
| CLIP2 | 3.189726 | 0.011932 | 0.999532 | | 3.514218 | 0.01913 | 0.990826 | | 2.99313 | 0.020856 | 0.994537 |
| FLVCR2 | 2.911627 | 0.139411 | 0.999532 | | 2.978346 | -0.0223 | 0.990826 | | 2.662281 | -0.144 | 0.939913 |
| RFC1 | 3.267953 | 0.317585 | 0.999532 | | 3.328817 | -0.02193 | 0.990826 | | 2.95081 | -0.33427 | 0.806797 |
| PAXBP1 | 3.380659 | -0.06726 | 0.999532 | | 3.796806 | 0.019575 | 0.990826 | | 3.270649 | 0.093045 | 0.96129 |
| NID1 | 6.403455 | -0.15556 | 0.999532 | | 7.444428 | 0.018374 | 0.990826 | | 6.002544 | 0.187236 | 0.892869 |
| TMEM185 | 3.150452 | 0.197522 | 0.999532 | | 3.281787 | 0.020131 | 0.990826 | | 2.958342 | -0.16061 | 0.916969 |
| EXOC7 | 8.913995 | -0.00584 | 0.999532 | | 10.11737 | -0.01608 | 0.990826 | | 8.464762 | 0.022686 | 0.992286 |
| FAM104A | 2.907764 | 0.194546 | 0.999532 | | 3.171059 | -0.02198 | 0.990826 | | 2.710184 | -0.19002 | 0.905952 |
| SAP25 | 4.661878 | 0.129098 | 0.999532 | | 4.819855 | 0.016961 | 0.990826 | | 4.559773 | -0.11151 | 0.94386 |
| ALPL | 3.048999 | -0.281 | 0.999532 | | 3.657176 | -0.02018 | 0.990826 | | 2.934075 | 0.278175 | 0.853068 |
| RXRB | 3.525585 | 0.060163 | 0.999532 | | 3.90074 | 0.018213 | 0.990826 | | 3.375734 | -0.0234 | 0.992459 |
| RAD54L2 | 3.237271 | 0.048394 | 0.999532 | | 3.769863 | 0.020078 | 0.990826 | | 3.234624 | -0.0079 | 0.997608 |
| TMEM129 | 4.62147 | -0.24054 | 0.999532 | | 5.777132 | -0.01571 | 0.990826 | | 4.365608 | 0.247327 | 0.836935 |
| ZDHHC17 | 2.128009 | -0.22574 | 0.999532 | | 2.586256 | -0.0228 | 0.990826 | | 1.978515 | 0.220733 | 0.905952 |
| LRIG2 | 3.27528 | -0.0846 | 0.999532 | | 3.609501 | 0.019699 | 0.990826 | | 3.22561 | 0.114103 | 0.952656 |
| ADGRL1 | 4.309801 | -0.02136 | 0.999532 | | 5.157561 | 0.017973 | 0.990826 | | 4.342316 | 0.056927 | 0.974433 |
| ARHGAP | 10.47311 | 0.219016 | 0.999532 | | 10.92716 | 0.013086 | 0.990826 | | 9.980071 | -0.18882 | 0.816775 |
| FOXM1 | 2.31881 | 0.39174 | 0.999532 | | 2.185533 | -0.02522 | 0.990826 | | 2.272432 | -0.40035 | 0.801572 |
| OR52D1 | 2.127938 | 0.400868 | 0.999532 | | 2.075875 | -0.02519 | 0.990826 | | 2.048019 | -0.40196 | 0.814126 |
| MYBPC1 | 2.06643 | 0.298004 | 0.999532 | | 2.130875 | -0.02638 | 0.990826 | | 2.005938 | -0.30282 | 0.872892 |
| USH1G | 2.589935 | 0.243559 | 0.999532 | | 2.742587 | -0.0255 | 0.990826 | | 2.425602 | -0.24247 | 0.890979 |
| ATG16L1 | 3.202887 | 0.293721 | 0.999532 | | 3.281173 | 0.019834 | 0.990826 | | 3.111348 | -0.25716 | 0.853549 |
| PLCB2 | 3.43504 | 0.642605 | 0.999532 | | 3.016988 | 0.021368 | 0.990826 | | 3.481282 | -0.59834 | 0.609581 |
| PLEKHA3 | 4.833482 | -0.02048 | 0.999532 | | 5.529718 | 0.017062 | 0.990826 | | 4.584544 | 0.053879 | 0.97652 |
| PPP1R9A | 2.782872 | 0.180875 | 0.999532 | | 2.972669 | -0.02153 | 0.990826 | | 2.710065 | -0.18442 | 0.913432 |
| MORN2 | 2.84148 | 0.028617 | 0.999532 | | 3.091249 | -0.02183 | 0.990826 | | 2.704843 | -0.03571 | 0.990309 |
| TBC1D25 | 3.381648 | -0.39618 | 0.999532 | | 4.472941 | -0.01865 | 0.990826 | | 3.398764 | 0.398991 | 0.750389 |
| CMSS1 | 2.377271 | -0.26462 | 0.999532 | | 2.989083 | -0.02145 | 0.990826 | | 2.38487 | 0.262087 | 0.879947 |
| ACOT7 | 2.301674 | 0.160344 | 0.999532 | | 2.390369 | 0.023933 | 0.990826 | | 2.304084 | -0.11942 | 0.955728 |
| BICRAL | 3.226118 | 0.107697 | 0.999532 | | 3.479407 | -0.0205 | 0.990826 | | 2.927488 | -0.11002 | 0.954538 |
| OSBPL3 | 3.601828 | 0.123694 | 0.999532 | | 3.802073 | 0.018808 | 0.990826 | | 3.483599 | -0.08687 | 0.96129 |
| JAZF1 | 5.437734 | 0.088498 | 0.999532 | | 5.91421 | -0.01707 | 0.990826 | | 5.362621 | -0.08461 | 0.955167 |
| AGRP | 3.592542 | 0.623512 | 0.999532 | | 3.3171 | 0.021313 | 0.990826 | | 3.500837 | -0.58093 | 0.623462 |
| TNFRSF2 | 3.702185 | -0.36007 | 0.999532 | | 4.619472 | -0.01778 | 0.991025 | | 3.492815 | 0.358762 | 0.788052 |
| CITED2 | 13.65331 | -0.16155 | 0.999532 | | 16.32628 | 0.015669 | 0.991025 | | 12.67381 | 0.224677 | 0.79686 |
| CPT2 | 2.35969 | 0.246487 | 0.999532 | | 2.601813 | 0.022707 | 0.991025 | | 2.465946 | -0.21356 | 0.904892 |
| SLC25A3 | 3.36656 | 0.011403 | 0.999532 | | 3.636537 | -0.01841 | 0.991025 | | 3.172334 | -0.0134 | 0.996681 |
| TPSAB1 | 2.989343 | 0.013177 | 0.999532 | | 3.299887 | -0.01982 | 0.991025 | | 2.682349 | -0.0109 | 0.996891 |
| RABL3 | 4.289047 | -0.10021 | 0.999532 | | 5.062025 | -0.01627 | 0.991025 | | 4.128181 | 0.104271 | 0.946512 |
| SAV1 | 2.948311 | -0.75158 | 0.999532 | | 3.757923 | 0.01761 | 0.991025 | | 2.726902 | 0.762992 | 0.533876 |
| KAAG1 | 2.045562 | 0.201779 | 0.999532 | | 2.298341 | 0.024399 | 0.991025 | | 1.993286 | -0.15093 | 0.945129 |
| OR3A3 | 2.131287 | 0.324451 | 0.999532 | | 2.288753 | -0.02472 | 0.991025 | | 2.092035 | -0.32659 | 0.858705 |
| TICAM2 | 2.637341 | 0.357723 | 0.999532 | | 2.42231 | 0.022448 | 0.991025 | | 2.440822 | -0.3168 | 0.844881 |
| HSPA1B | 3.829453 | -0.06977 | 0.999532 | | 4.217957 | 0.01746 | 0.991025 | | 3.49486 | 0.10384 | 0.948484 |
| DTWD1 | 4.659069 | 0.434667 | 0.999532 | | 4.598716 | 0.017618 | 0.991025 | | 4.424493 | -0.39206 | 0.720627 |
| TRAF6 | 3.078627 | -0.03707 | 0.999532 | | 3.545805 | 0.019469 | 0.991025 | | 2.923487 | 0.073616 | 0.972031 |
| HIRA | 3.89506 | 0.588759 | 0.999532 | | 3.636606 | -0.01915 | 0.991025 | | 3.717729 | -0.57922 | 0.595502 |
| JAM3 | 4.964449 | -0.34126 | 0.999532 | | 6.169192 | -0.01547 | 0.991025 | | 4.585391 | 0.345379 | 0.737952 |
| HMGXB3 | 2.383339 | -0.21834 | 0.999532 | | 2.911488 | -0.02053 | 0.991025 | | 2.179138 | 0.213735 | 0.90424 |
| OAZ3 | 1.861805 | 0.172596 | 0.999532 | | 2.035964 | 0.024853 | 0.991025 | | 1.840139 | -0.12655 | 0.958422 |
| FOXR1 | 2.394491 | 0.868384 | 0.999532 | | 1.877658 | -0.0249 | 0.991025 | | 2.320365 | -0.87582 | 0.506666 |
| CMTM6 | 4.43337 | -0.272 | 0.999532 | | 4.980378 | -0.01607 | 0.991025 | | 4.271117 | 0.263508 | 0.842754 |
| CD68 | 4.860989 | -0.27889 | 0.999532 | | 6.056433 | -0.0176 | 0.991025 | | 4.682035 | 0.277358 | 0.820072 |
| FHL3 | 4.362056 | -0.39529 | 0.999532 | | 5.390211 | -0.01612 | 0.991025 | | 4.109314 | 0.390833 | 0.737452 |
| E2F1 | 2.732027 | 0.436096 | 0.999532 | | 2.416096 | 0.023072 | 0.991025 | | 2.499435 | -0.39513 | 0.799944 |
| BLOC1S6 | 4.850922 | -0.14727 | 0.999532 | | 5.623388 | 0.016045 | 0.991025 | | 4.503792 | 0.188385 | 0.881384 |
| SLC9A8 | 3.210638 | 0.721882 | 0.999532 | | 2.632149 | 0.021213 | 0.991096 | | 3.222324 | -0.68876 | 0.557909 |
| FBXW4 | 6.400679 | -0.19484 | 0.999532 | | 7.500257 | -0.01397 | 0.991115 | | 5.921336 | 0.203521 | 0.852386 |
| CGAS | 3.095318 | 0.107042 | 0.999532 | | 3.152293 | 0.019041 | 0.991115 | | 2.960815 | -0.06528 | 0.975389 |
| UQCC3 | 2.682689 | 0.349801 | 0.999532 | | 2.786035 | 0.020765 | 0.991115 | | 2.584324 | -0.33017 | 0.836453 |
| MAPK1IP | 11.62429 | -0.21896 | 0.999532 | | 14.09419 | 0.010264 | 0.991115 | | 11.30664 | 0.250481 | 0.724363 |
| FUT1 | 1.817797 | 0.103992 | 0.999532 | | 2.086629 | -0.02394 | 0.991115 | | 1.807696 | -0.10958 | 0.964621 |
| OR9G9 | 2.724762 | -0.27056 | 0.999532 | | 3.295662 | -0.02037 | 0.991115 | | 2.536314 | 0.269867 | 0.87591 |
| ZNF248 | 4.170405 | 0.064841 | 0.999532 | | 4.503675 | 0.016734 | 0.991115 | | 3.94631 | -0.03243 | 0.98918 |
| WAPL | 4.133806 | -0.24623 | 0.999532 | | 5.028163 | -0.01583 | 0.991137 | | 3.818455 | 0.253109 | 0.844187 |
| CSTF2T | 3.066659 | -0.25739 | 0.999532 | | 3.967664 | -0.0185 | 0.991137 | | 2.961851 | 0.264113 | 0.865065 |
| P2RY14 | 2.978871 | 0.646925 | 0.999532 | | 2.576411 | 0.022238 | 0.991137 | | 2.843278 | -0.60335 | 0.647581 |
| THOC1 | 3.697958 | 0.115348 | 0.999532 | | 3.860594 | -0.01879 | 0.99134 | | 3.450833 | -0.11008 | 0.947126 |
| TMEM184 | 4.902705 | -0.09233 | 0.999532 | | 5.616648 | 0.016093 | 0.991388 | | 4.67348 | 0.123436 | 0.93603 |
| APOBEC | 5.302356 | 0.151713 | 0.999532 | | 5.635193 | -0.01721 | 0.991428 | | 5.216518 | -0.1551 | 0.913083 |
| MCTP2 | 2.164887 | 0.516353 | 0.999532 | | 1.971169 | 0.02428 | 0.991428 | | 2.149111 | -0.47181 | 0.765852 |
| NPAT | 2.812974 | 0.354166 | 0.999532 | | 2.718458 | -0.02021 | 0.991436 | | 2.524437 | -0.3529 | 0.80383 |
| HOXD8 | 4.606144 | 0.373917 | 0.999532 | | 4.672187 | 0.016272 | 0.991633 | | 4.196117 | -0.34879 | 0.75311 |
| CAMSAP | 2.581634 | -0.05394 | 0.999532 | | 2.907138 | -0.02062 | 0.991646 | | 2.457963 | 0.046637 | 0.987274 |
| TAS2R46 | 2.224125 | 0.46867 | 0.999532 | | 2.027227 | -0.02345 | 0.991758 | | 2.191976 | -0.4765 | 0.771685 |
| SRXN1 | 3.047658 | -0.09826 | 0.999532 | | 3.738046 | -0.01821 | 0.991758 | | 3.051181 | 0.103152 | 0.958191 |
| CLN8 | 3.296541 | 0.207604 | 0.999532 | | 3.417175 | 0.0175 | 0.991758 | | 3.134389 | -0.17254 | 0.907968 |
| CCDC50 | 4.173407 | 0.006561 | 0.999532 | | 4.443452 | -0.01602 | 0.991758 | | 3.929365 | -0.01725 | 0.995304 |
| PPTC7 | 4.431968 | -0.2121 | 0.999532 | | 5.249135 | -0.01546 | 0.992024 | | 4.026953 | 0.220451 | 0.864923 |
| ADCY4 | 5.414988 | -0.63073 | 0.999532 | | 7.329937 | 0.013392 | 0.992068 | | 5.238151 | 0.668292 | 0.413122 |
| KATNAL1 | 2.437902 | -0.14148 | 0.999532 | | 3.091011 | -0.01932 | 0.992068 | | 2.374218 | 0.145623 | 0.9402 |
| SLC25A4 | 2.324415 | 0.574262 | 0.999532 | | 2.21831 | 0.021712 | 0.992068 | | 2.23035 | -0.5322 | 0.706738 |
| WDSUB1 | 3.751884 | -0.175 | 0.999532 | | 4.386876 | -0.01623 | 0.992069 | | 3.492926 | 0.178977 | 0.902515 |
| SOGA3 | 2.171193 | 0.437569 | 0.999532 | | 2.048536 | 0.021492 | 0.992504 | | 2.113524 | -0.3998 | 0.813127 |
| PGBD3 | 2.204739 | 0.006254 | 0.999532 | | 2.546723 | -0.01873 | 0.992504 | | 2.182125 | -0.00663 | 0.998123 |
| NR1H4 | 3.399497 | -0.04086 | 0.999532 | | 3.650579 | -0.01609 | 0.992504 | | 3.128055 | 0.041007 | 0.986446 |
| TMEM72 | 3.758397 | 0.779184 | 0.999532 | | 3.426802 | 0.016573 | 0.992504 | | 3.561692 | -0.73197 | 0.49299 |
| UPF3B | 2.701667 | 0.118734 | 0.999532 | | 3.206056 | 0.017472 | 0.992504 | | 2.755708 | -0.07416 | 0.972568 |
| C5orf22 | 3.734199 | -0.1062 | 0.999532 | | 4.357349 | 0.015332 | 0.992504 | | 3.572444 | 0.138828 | 0.929613 |
| TRIM33 | 4.073376 | 0.049348 | 0.999532 | | 4.551488 | -0.0153 | 0.992504 | | 3.943054 | -0.03986 | 0.988811 |
| AATK | 1.945494 | 0.008597 | 0.999532 | | 2.217126 | 0.021446 | 0.992504 | | 1.999326 | 0.028384 | 0.993618 |
| SLC35B1 | 2.98872 | -0.22449 | 0.999532 | | 3.809628 | 0.016541 | 0.992504 | | 3.146808 | 0.264064 | 0.861119 |
| MCHR2 | 2.163992 | -0.00407 | 0.999532 | | 2.602171 | 0.01995 | 0.992504 | | 2.149272 | 0.052401 | 0.98556 |
| RNF215 | 2.315572 | 0.28007 | 0.999532 | | 2.477715 | 0.018818 | 0.992504 | | 2.329521 | -0.24069 | 0.886495 |
| ALDH1A3 | 2.018974 | -0.52836 | 0.999532 | | 2.432908 | -0.02021 | 0.992504 | | 1.845266 | 0.523998 | 0.753597 |
| ADPRHL1 | 2.113028 | -0.01739 | 0.999532 | | 2.26785 | -0.02089 | 0.992504 | | 2.016122 | 0.019873 | 0.996681 |
| BACE2 | 3.245005 | -0.22136 | 0.999532 | | 3.991859 | -0.01676 | 0.992504 | | 3.192242 | 0.222245 | 0.88476 |
| H2AZ2 | 9.6025 | -0.1298 | 0.999532 | | 10.9062 | 0.012269 | 0.992504 | | 9.079415 | 0.154386 | 0.892328 |
| ZFP91 | 5.406124 | -0.01837 | 0.999532 | | 6.177416 | 0.013421 | 0.992504 | | 5.135732 | 0.050455 | 0.974421 |
| ZNF383 | 13.85766 | -0.05041 | 0.999532 | | 15.97844 | -0.00986 | 0.992504 | | 13.45099 | 0.063315 | 0.951656 |
| RETREG1 | 7.463665 | 0.432783 | 0.999532 | | 6.8486 | 0.012789 | 0.992504 | | 7.07017 | -0.40598 | 0.657617 |
| P2RX7 | 2.444232 | -0.24609 | 0.999532 | | 3.009687 | -0.01977 | 0.992504 | | 2.350523 | 0.244475 | 0.892869 |
| NR2E1 | 2.812806 | 0.535856 | 0.999532 | | 2.572587 | -0.02065 | 0.992504 | | 2.794905 | -0.52348 | 0.728015 |
| ZNF174 | 3.949547 | -0.07833 | 0.999532 | | 4.596673 | 0.014949 | 0.992504 | | 3.94312 | 0.115647 | 0.942744 |
| ENAH | 4.040215 | -0.0459 | 0.999532 | | 4.625794 | -0.01522 | 0.992504 | | 4.004306 | 0.048465 | 0.978463 |
| ANKRD39 | 2.408042 | 0.128695 | 0.999532 | | 2.537766 | 0.020585 | 0.992504 | | 2.247205 | -0.09138 | 0.970496 |
| WBP4 | 3.804733 | -0.0766 | 0.999532 | | 4.159468 | 0.015572 | 0.992504 | | 3.42494 | 0.110531 | 0.946765 |
| KDM2A | 6.765326 | -0.42603 | 0.999532 | | 8.738173 | -0.01187 | 0.992504 | | 6.333816 | 0.435154 | 0.647218 |
| PGP | 4.751609 | -0.12441 | 0.999532 | | 5.744365 | -0.01353 | 0.992504 | | 4.628352 | 0.134893 | 0.920259 |
| KCNK1 | 2.06218 | -0.1454 | 0.999532 | | 2.334024 | -0.02086 | 0.992504 | | 1.83619 | 0.13959 | 0.951656 |
| FSTL5 | 1.836254 | 0.188894 | 0.999532 | | 1.939422 | -0.02302 | 0.992504 | | 1.719078 | -0.19292 | 0.932496 |
| SLC4A2 | 3.296194 | 0.254579 | 0.999532 | | 3.292571 | 0.017114 | 0.992504 | | 2.97699 | -0.21897 | 0.88476 |
| RBAK | 2.143025 | 0.341819 | 0.999532 | | 1.978388 | 0.021442 | 0.992504 | | 2.109546 | -0.29835 | 0.877582 |
| GOPC | 3.300359 | 0.353565 | 0.999532 | | 3.256782 | -0.0167 | 0.992504 | | 3.089218 | -0.35219 | 0.787644 |
| SF3B6 | 5.298712 | 0.06445 | 0.999532 | | 5.799489 | 0.014452 | 0.992504 | | 5.083541 | -0.02961 | 0.988853 |
| CISH | 2.225718 | 0.102678 | 0.999532 | | 2.388572 | -0.01984 | 0.992504 | | 2.133746 | -0.10387 | 0.964445 |
| PEX11B | 3.352306 | -0.05182 | 0.999532 | | 3.909237 | -0.01541 | 0.992573 | | 3.271069 | 0.055359 | 0.978229 |
| ASPA | 2.021218 | -0.01028 | 0.999532 | | 2.280212 | -0.01911 | 0.992584 | | 1.91382 | 0.009808 | 0.997153 |
| PDZD11 | 2.573844 | -0.55521 | 0.999532 | | 3.451102 | -0.01632 | 0.992584 | | 2.45285 | 0.556796 | 0.675955 |
| METTL24 | 2.598663 | 0.071503 | 0.999532 | | 2.737607 | -0.01872 | 0.992584 | | 2.299863 | -0.06649 | 0.977473 |
| DDX20 | 3.144332 | -0.47308 | 0.999532 | | 4.316024 | 0.015 | 0.992584 | | 3.195993 | 0.510979 | 0.670353 |
| SGPP1 | 3.012775 | 0.268631 | 0.999532 | | 3.078761 | 0.016411 | 0.992584 | | 2.88297 | -0.23493 | 0.87784 |
| FGL1 | 2.518918 | 0.735022 | 0.999532 | | 2.085971 | -0.02089 | 0.992584 | | 2.490887 | -0.74172 | 0.575547 |
| KSR1 | 3.939578 | -0.17968 | 0.999532 | | 4.907211 | -0.01411 | 0.992584 | | 3.910138 | 0.198253 | 0.892869 |
| SLC25A4 | 2.828165 | -0.18255 | 0.999532 | | 3.349347 | 0.017438 | 0.992584 | | 2.590224 | 0.222337 | 0.892869 |
| AFMID | 2.91357 | 0.613477 | 0.999532 | | 2.692584 | 0.018273 | 0.992584 | | 2.714931 | -0.5812 | 0.634624 |
| GFUS | 4.364412 | -0.07326 | 0.999532 | | 5.045032 | -0.01326 | 0.992584 | | 4.09964 | 0.078091 | 0.96129 |
| GRIN3B | 3.011347 | 0.17656 | 0.999532 | | 3.106105 | 0.018263 | 0.992584 | | 2.923188 | -0.14664 | 0.941197 |
| SLC16A6 | 2.752831 | 0.648638 | 0.999532 | | 2.281441 | -0.01957 | 0.992584 | | 2.64116 | -0.6529 | 0.629345 |
| ERI2 | 1.935344 | 0.508153 | 0.999532 | | 1.901908 | -0.02123 | 0.992584 | | 1.939246 | -0.51084 | 0.742171 |
| KIAA0586 | 2.374326 | -0.1348 | 0.999532 | | 2.582046 | 0.018337 | 0.992584 | | 2.200651 | 0.17858 | 0.926586 |
| GMFG | 6.420485 | -0.32842 | 0.999532 | | 7.948335 | 0.012537 | 0.992584 | | 6.192251 | 0.367145 | 0.692534 |
| FRAT2 | 5.390338 | 0.115838 | 0.999532 | | 5.966906 | 0.013684 | 0.992584 | | 5.148964 | -0.0793 | 0.958191 |
| DIMT1 | 2.962268 | -0.3124 | 0.999532 | | 3.417375 | 0.015817 | 0.992616 | | 2.781996 | 0.32025 | 0.842754 |
| GSTCD | 2.328053 | 0.009106 | 0.999532 | | 2.502047 | 0.01796 | 0.992616 | | 2.212086 | 0.017751 | 0.996681 |
| MARCHF | 2.982103 | -0.55896 | 0.999532 | | 4.071655 | 0.014405 | 0.992616 | | 2.854917 | 0.59284 | 0.628895 |
| SUSD1 | 2.451245 | 0.12689 | 0.999532 | | 2.512652 | -0.01971 | 0.992616 | | 2.242293 | -0.12175 | 0.956322 |
| GPATCH2 | 3.752174 | -0.12492 | 0.999532 | | 4.176568 | 0.014966 | 0.992616 | | 3.550539 | 0.157219 | 0.918024 |
| OR10T2 | 2.330048 | 0.158669 | 0.999532 | | 2.68425 | 0.019551 | 0.992616 | | 2.19816 | -0.10916 | 0.96129 |
| BEND6 | 2.251205 | -0.18607 | 0.999532 | | 2.705922 | 0.017801 | 0.992616 | | 2.186332 | 0.221377 | 0.901033 |
| RFPL2 | 1.807361 | -0.27551 | 0.999532 | | 2.403915 | 0.0202 | 0.992616 | | 1.95851 | 0.319467 | 0.877268 |
| ANKS6 | 3.18756 | 0.211558 | 0.999532 | | 3.254854 | 0.016477 | 0.992616 | | 2.970699 | -0.1763 | 0.910105 |
| FAM71B | 2.160543 | 0.233364 | 0.999532 | | 2.129607 | -0.01897 | 0.992617 | | 2.079183 | -0.23959 | 0.90016 |
| SRSF7 | 5.982997 | 0.284207 | 0.999532 | | 6.059196 | -0.01199 | 0.992617 | | 5.745225 | -0.2786 | 0.78258 |
| PALM3 | 2.486358 | 0.070898 | 0.999532 | | 2.706024 | -0.01849 | 0.992617 | | 2.417807 | -0.07429 | 0.974421 |
| ZNF561 | 3.506565 | -0.05179 | 0.999532 | | 3.85265 | 0.01497 | 0.992617 | | 3.314596 | 0.086973 | 0.962255 |
| DYRK3 | 3.077474 | 0.17699 | 0.999532 | | 3.103107 | 0.016234 | 0.992617 | | 2.925993 | -0.14495 | 0.934172 |
| RASSF3 | 1.939273 | -0.09888 | 0.999532 | | 2.204436 | 0.018771 | 0.992617 | | 1.881312 | 0.131652 | 0.955167 |
| H2BC13 | 2.58657 | -0.16109 | 0.999532 | | 3.079395 | 0.01756 | 0.992617 | | 2.453719 | 0.202444 | 0.910105 |
| STON2 | 4.062744 | -0.12708 | 0.999532 | | 4.652509 | 0.013116 | 0.992617 | | 3.925892 | 0.155019 | 0.914692 |
| MSH2 | 2.177245 | 0.232727 | 0.999532 | | 2.159275 | -0.01852 | 0.992617 | | 2.113109 | -0.23079 | 0.901033 |
| RPH3AL | 5.121515 | 0.491071 | 0.999532 | | 4.881322 | -0.01418 | 0.992617 | | 5.02792 | -0.4907 | 0.636625 |
| MBTPS2 | 2.294744 | 0.53358 | 0.999532 | | 1.940467 | -0.01989 | 0.992617 | | 2.107782 | -0.54881 | 0.722703 |
| INTS6 | 4.355316 | 0.043337 | 0.999532 | | 4.788159 | 0.015798 | 0.992617 | | 4.188244 | -0.00451 | 0.99858 |
| CAMSAP | 3.160221 | 0.297959 | 0.999532 | | 3.342329 | 0.016166 | 0.992617 | | 3.275586 | -0.26621 | 0.861737 |
| FBLIM1 | 2.61042 | -0.06044 | 0.999532 | | 3.087505 | -0.01578 | 0.992617 | | 2.544448 | 0.066726 | 0.975753 |
| SLC26A1 | 2.530747 | -0.03554 | 0.999532 | | 2.737067 | -0.01681 | 0.992617 | | 2.322507 | 0.034606 | 0.991159 |
| CD302 | 3.828164 | 0.009601 | 0.999532 | | 4.140716 | 0.013983 | 0.992617 | | 3.633205 | 0.02174 | 0.993435 |
| SNW1 | 7.612715 | -0.06506 | 0.999532 | | 8.2733 | -0.01163 | 0.992617 | | 6.837307 | 0.084575 | 0.946572 |
| RNF43 | 2.688411 | 0.118831 | 0.999532 | | 2.914688 | 0.017481 | 0.992617 | | 2.535236 | -0.08292 | 0.971683 |
| ADSL | 4.124725 | -0.21858 | 0.999532 | | 5.046291 | 0.013614 | 0.992617 | | 4.203845 | 0.241246 | 0.858767 |
| TMEM177 | 2.727563 | 0.149938 | 0.999532 | | 2.867836 | -0.0172 | 0.992617 | | 2.633791 | -0.14872 | 0.938429 |
| PARP1 | 5.065079 | -0.12332 | 0.999532 | | 5.809424 | 0.012798 | 0.992617 | | 4.604771 | 0.162996 | 0.903054 |
| DIO2 | 2.235099 | 0.612762 | 0.999532 | | 2.053579 | 0.019699 | 0.992758 | | 2.188682 | -0.57479 | 0.707142 |
| CXXC4 | 1.954108 | -0.52486 | 0.999532 | | 2.438175 | 0.017445 | 0.992758 | | 1.866176 | 0.550444 | 0.740093 |
| CCDC182 | 2.297867 | 0.277298 | 0.999532 | | 2.293593 | 0.018145 | 0.992758 | | 2.152912 | -0.24504 | 0.897096 |
| NME6 | 2.437295 | -0.09468 | 0.999532 | | 3.026002 | 0.016312 | 0.992758 | | 2.451317 | 0.13689 | 0.945759 |
| UBN2 | 3.399487 | -0.21396 | 0.999532 | | 4.210588 | 0.013886 | 0.992758 | | 3.230629 | 0.250282 | 0.865065 |
| MNAT1 | 2.215553 | -0.47479 | 0.999532 | | 2.81279 | -0.01675 | 0.992758 | | 2.129259 | 0.477101 | 0.75038 |
| KRTAP11 | 1.965069 | 0.211403 | 0.999532 | | 2.032384 | -0.01955 | 0.992758 | | 1.828711 | -0.21165 | 0.918694 |
| NFKBIE | 2.964592 | -0.04249 | 0.999532 | | 3.239491 | 0.015793 | 0.992758 | | 2.68449 | 0.057284 | 0.980581 |
| CDON | 1.931699 | 0.292099 | 0.999532 | | 2.010088 | 0.019048 | 0.992758 | | 1.884119 | -0.25407 | 0.894612 |
| IDH3B | 5.157066 | -0.14279 | 0.999532 | | 6.104572 | 0.011952 | 0.992758 | | 4.944292 | 0.173132 | 0.88885 |
| DSN1 | 1.743207 | -0.12987 | 0.999532 | | 2.04754 | 0.018494 | 0.992758 | | 1.705698 | 0.166647 | 0.9402 |
| SPDL1 | 3.405752 | -0.06671 | 0.999532 | | 3.817061 | -0.01492 | 0.992758 | | 3.237773 | 0.066926 | 0.974421 |
| STIM1 | 4.472284 | -0.21035 | 0.999532 | | 4.933198 | -0.01321 | 0.992758 | | 4.133756 | 0.20876 | 0.881637 |
| SHLD2 | 4.039113 | -0.00881 | 0.999532 | | 4.560233 | 0.013013 | 0.992778 | | 4.02147 | 0.040046 | 0.98556 |
| GCC2 | 8.042454 | -0.11525 | 0.999532 | | 9.85695 | 0.009754 | 0.992778 | | 7.904426 | 0.145501 | 0.88885 |
| OR12D2 | 2.140496 | 0.306723 | 0.999532 | | 2.368225 | -0.01849 | 0.992908 | | 2.124794 | -0.29714 | 0.871957 |
| KLHL22 | 3.190582 | -0.06805 | 0.999532 | | 3.746353 | 0.014305 | 0.993151 | | 3.011912 | 0.104495 | 0.955834 |
| IVD | 3.386877 | -0.00973 | 0.999532 | | 3.927267 | -0.01335 | 0.9933 | | 3.28038 | 0.018482 | 0.995304 |
| SPI1 | 5.571452 | -0.47524 | 0.999532 | | 6.845195 | -0.012 | 0.9933 | | 5.203604 | 0.474502 | 0.656225 |
| TAC3 | 2.088502 | 0.422742 | 0.999532 | | 1.984261 | -0.01878 | 0.9933 | | 2.061486 | -0.42851 | 0.79686 |
| PCCA | 2.820408 | -0.17666 | 0.999532 | | 3.266398 | 0.014246 | 0.9933 | | 2.666818 | 0.200831 | 0.904721 |
| MPZL2 | 2.937547 | 0.329602 | 0.999532 | | 2.995733 | 0.014893 | 0.9933 | | 2.795826 | -0.29695 | 0.835554 |
| TMEM50A | 5.01868 | -0.51407 | 0.999532 | | 6.518485 | 0.010477 | 0.9933 | | 4.803782 | 0.538863 | 0.572084 |
| PPP6R2 | 3.488698 | -0.25805 | 0.999532 | | 4.179152 | -0.01289 | 0.9933 | | 3.321043 | 0.26023 | 0.859522 |
| STT3B | 10.71181 | 0.000447 | 0.999532 | | 11.4775 | 0.009918 | 0.9933 | | 9.948482 | 0.024949 | 0.989148 |
| GAB1 | 3.174036 | 0.340445 | 0.999532 | | 3.161861 | 0.014754 | 0.9933 | | 3.045702 | -0.30938 | 0.824012 |
| SCARF1 | 4.239768 | -0.69029 | 0.999532 | | 5.947826 | 0.011845 | 0.9933 | | 4.111884 | 0.72276 | 0.485816 |
| RBIS | 6.435039 | -0.248 | 0.999532 | | 7.835452 | -0.01002 | 0.9933 | | 6.18394 | 0.256388 | 0.791576 |
| EPS15 | 6.088512 | -0.06954 | 0.999532 | | 6.707057 | -0.01027 | 0.9933 | | 5.577558 | 0.072464 | 0.960773 |
| TNNT1 | 2.000227 | -0.07137 | 0.999532 | | 2.417255 | 0.017054 | 0.993304 | | 2.069132 | 0.109867 | 0.964971 |
| SP110 | 6.933422 | 0.017886 | 0.999532 | | 8.003639 | -0.00976 | 0.99336 | | 6.738165 | -0.01065 | 0.996681 |
| CDC6 | 2.183045 | 0.313965 | 0.999532 | | 2.180945 | -0.01743 | 0.99336 | | 2.127 | -0.31301 | 0.862564 |
| APCDD1L | 2.335922 | -0.03174 | 0.999532 | | 2.720732 | -0.01603 | 0.99336 | | 2.443945 | 0.019453 | 0.996681 |
| TMEM14A | 4.275888 | -0.13822 | 0.999532 | | 4.985606 | -0.01158 | 0.99336 | | 4.096915 | 0.145864 | 0.914687 |
| UBE2D2 | 5.197487 | 0.06468 | 0.999532 | | 5.699734 | 0.011352 | 0.99336 | | 4.913312 | -0.03493 | 0.985675 |
| PTF1A | 2.342946 | 0.004341 | 0.999532 | | 2.343254 | -0.01586 | 0.99337 | | 2.187342 | -0.01498 | 0.996891 |
| GPX1 | 10.50159 | -0.24253 | 0.999532 | | 12.7484 | 0.009224 | 0.993425 | | 9.995431 | 0.267495 | 0.799127 |
| CALHM5 | 5.919035 | -0.0832 | 0.999532 | | 6.677804 | -0.01073 | 0.993425 | | 5.724136 | 0.086359 | 0.955834 |
| ZBED4 | 2.354687 | 0.654297 | 0.999532 | | 2.071387 | 0.017459 | 0.993475 | | 2.411267 | -0.62665 | 0.652925 |
| HS6ST2 | 2.48061 | 0.633793 | 0.999532 | | 2.016672 | -0.0171 | 0.993673 | | 2.248227 | -0.63707 | 0.660097 |
| TRMT12 | 3.60341 | 0.089807 | 0.999532 | | 3.967055 | -0.01341 | 0.993673 | | 3.48777 | -0.07835 | 0.972125 |
| DHX34 | 2.744161 | 0.22642 | 0.999532 | | 2.701793 | 0.015204 | 0.993673 | | 2.537592 | -0.19906 | 0.910105 |
| DLX3 | 1.806913 | 0.179239 | 0.999532 | | 2.017993 | -0.01816 | 0.993673 | | 1.849641 | -0.17277 | 0.941512 |
| TRPT1 | 4.652772 | -0.08444 | 0.999532 | | 5.594623 | 0.011207 | 0.993673 | | 4.515997 | 0.108571 | 0.940162 |
| CELF4 | 2.972735 | 0.550829 | 0.999532 | | 2.626118 | -0.01659 | 0.993673 | | 2.836279 | -0.55841 | 0.703463 |
| DNAJB6 | 7.505498 | -0.32083 | 0.999532 | | 9.056356 | 0.00893 | 0.993673 | | 7.054057 | 0.34027 | 0.705888 |
| SLFNL1 | 7.571694 | 0.282305 | 0.999532 | | 7.519922 | 0.009912 | 0.993673 | | 7.2859 | -0.26033 | 0.828971 |
| ADORA2A | 2.935512 | 0.31744 | 0.999532 | | 2.794569 | 0.014574 | 0.993673 | | 2.875016 | -0.28416 | 0.848012 |
| SSBP4 | 7.810256 | -0.36118 | 0.999532 | | 10.05786 | -0.0082 | 0.993713 | | 7.419264 | 0.371298 | 0.647218 |
| NUBP1 | 3.899695 | -0.11029 | 0.999532 | | 4.518867 | 0.011712 | 0.993713 | | 3.608941 | 0.139691 | 0.923864 |
| CYBB | 2.576704 | 0.032906 | 0.999532 | | 2.768888 | -0.0145 | 0.993713 | | 2.552808 | -0.03281 | 0.991273 |
| NKTR | 5.788412 | 0.007115 | 0.999532 | | 6.098097 | -0.01071 | 0.993713 | | 5.560986 | -0.0085 | 0.996891 |
| ARID1A | 3.684635 | 0.219399 | 0.999532 | | 3.952313 | -0.01215 | 0.993713 | | 3.588809 | -0.20992 | 0.880949 |
| CWC25 | 3.64023 | 0.512663 | 0.999532 | | 3.381329 | -0.01352 | 0.993713 | | 3.607364 | -0.51507 | 0.657538 |
| KCNS1 | 2.672826 | 0.007574 | 0.999532 | | 3.013639 | -0.0145 | 0.993713 | | 2.664207 | -0.00271 | 0.999385 |
| DENND10 | 2.273621 | 0.36667 | 0.999532 | | 2.231611 | 0.016742 | 0.993713 | | 2.220958 | -0.32962 | 0.851372 |
| ENTPD5 | 3.859763 | 0.040373 | 0.999532 | | 4.428392 | -0.0121 | 0.993713 | | 3.634075 | -0.03409 | 0.988853 |
| CHCHD2 | 8.378756 | -0.10592 | 0.999532 | | 9.513094 | 0.007936 | 0.993853 | | 8.030458 | 0.132292 | 0.893262 |
| CDH23 | 1.964636 | 0.113974 | 0.999532 | | 2.12407 | 0.016134 | 0.993886 | | 1.895674 | -0.0788 | 0.975473 |
| MGAT2 | 2.871305 | -0.14747 | 0.999532 | | 3.38764 | -0.01325 | 0.993886 | | 2.500732 | 0.149793 | 0.929645 |
| UBR2 | 4.534142 | -0.04072 | 0.999532 | | 4.899737 | -0.0112 | 0.99401 | | 4.224959 | 0.046893 | 0.979591 |
| WFDC12 | 2.138479 | 0.07641 | 0.999532 | | 2.328802 | 0.016014 | 0.994125 | | 2.061719 | -0.04172 | 0.989985 |
| WWC2 | 6.256415 | 0.192288 | 0.999532 | | 6.594802 | 0.010451 | 0.994125 | | 5.96833 | -0.16199 | 0.903339 |
| DOLK | 3.112648 | 0.157734 | 0.999532 | | 3.363189 | -0.01271 | 0.994125 | | 3.098777 | -0.15435 | 0.925336 |
| NCAPD2 | 2.089189 | -0.12011 | 0.999532 | | 2.554697 | 0.01481 | 0.994125 | | 2.116108 | 0.154868 | 0.941197 |
| SCN3B | 2.133173 | -0.10947 | 0.999532 | | 2.381121 | 0.014436 | 0.994125 | | 2.117123 | 0.140902 | 0.946765 |
| TM7SF2 | 3.313238 | 0.416471 | 0.999532 | | 3.269907 | -0.01402 | 0.994125 | | 3.235923 | -0.4105 | 0.761337 |
| PICK1 | 2.758296 | -0.28728 | 0.999532 | | 3.243047 | -0.01277 | 0.994125 | | 2.465904 | 0.287675 | 0.859426 |
| TENM3 | 2.123476 | 0.645394 | 0.999532 | | 1.87614 | -0.01753 | 0.994125 | | 2.015056 | -0.64509 | 0.676565 |
| LRP3 | 3.733048 | 0.285012 | 0.999532 | | 3.776367 | -0.01224 | 0.994125 | | 3.557727 | -0.27555 | 0.835289 |
| USP10 | 9.375883 | 0.024514 | 0.999532 | | 10.65095 | 0.007681 | 0.994125 | | 8.954037 | 0.001605 | 0.999385 |
| XRN1 | 4.927955 | -0.38299 | 0.999532 | | 6.23786 | 0.00963 | 0.994125 | | 4.670659 | 0.409639 | 0.686214 |
| CENATAC | 3.989153 | -0.45714 | 0.999532 | | 4.885371 | 0.01003 | 0.994206 | | 3.756439 | 0.483722 | 0.652925 |
| RNF121 | 5.933218 | -0.25511 | 0.999532 | | 7.103209 | -0.00891 | 0.994206 | | 5.499191 | 0.26062 | 0.810105 |
| TRMT9B | 1.83083 | 0.11331 | 0.999532 | | 1.918961 | 0.015884 | 0.994206 | | 1.769415 | -0.07998 | 0.976115 |
| YEATS4 | 3.01051 | 0.126248 | 0.999532 | | 3.163124 | -0.0129 | 0.994206 | | 2.763107 | -0.11517 | 0.950616 |
| ABI3 | 2.297782 | -0.38024 | 0.999532 | | 2.787339 | -0.01279 | 0.994206 | | 2.095406 | 0.37762 | 0.816372 |
| NCAPG2 | 3.228575 | -0.13299 | 0.999532 | | 3.685828 | 0.011922 | 0.994206 | | 3.153283 | 0.156512 | 0.924186 |
| ZNF276 | 4.449142 | 0.091262 | 0.999532 | | 4.931442 | -0.01094 | 0.994206 | | 4.434949 | -0.06908 | 0.954538 |
| ZNF316 | 4.372347 | 0.228803 | 0.999532 | | 4.668203 | 0.011131 | 0.994206 | | 4.364677 | -0.19647 | 0.886495 |
| PHC1 | 6.784107 | 0.194062 | 0.999532 | | 6.880869 | 0.008962 | 0.994206 | | 6.326946 | -0.16657 | 0.877582 |
| FILIP1 | 2.855427 | -0.02555 | 0.999532 | | 3.340258 | -0.01232 | 0.994206 | | 2.738236 | 0.035411 | 0.989536 |
| DEK | 6.464461 | 0.004711 | 0.999532 | | 7.347059 | -0.00875 | 0.994206 | | 6.00044 | 0.00324 | 0.998586 |
| BPI | 2.020983 | 0.675289 | 0.999532 | | 1.840264 | -0.01656 | 0.994206 | | 1.912212 | -0.67401 | 0.66232 |
| CTDSPL2 | 3.16105 | -0.25238 | 0.999532 | | 3.723875 | -0.01238 | 0.994206 | | 2.890824 | 0.261189 | 0.871713 |
| FCN1 | 3.432545 | -0.07068 | 0.999532 | | 3.899193 | -0.01267 | 0.994286 | | 3.309174 | 0.071477 | 0.972221 |
| OR4C46 | 2.982284 | -0.11015 | 0.999532 | | 3.383272 | 0.012565 | 0.994286 | | 2.999782 | 0.13906 | 0.943533 |
| RHBDD2 | 5.4966 | -0.15719 | 0.999532 | | 6.369407 | -0.00903 | 0.994295 | | 5.098567 | 0.163111 | 0.892869 |
| TSPO | 2.484315 | 0.069238 | 0.999532 | | 2.575461 | -0.01367 | 0.994315 | | 2.219143 | -0.05584 | 0.982187 |
| THSD4 | 1.673486 | -0.34353 | 0.999532 | | 2.226295 | 0.014839 | 0.994315 | | 1.792138 | 0.380093 | 0.848938 |
| PTGER3 | 3.606589 | -0.22508 | 0.999532 | | 4.38588 | 0.011546 | 0.994315 | | 3.267346 | 0.268368 | 0.849848 |
| NOP58 | 5.743816 | -0.18343 | 0.999532 | | 6.953691 | -0.00891 | 0.994315 | | 5.55632 | 0.198825 | 0.865363 |
| MYH11 | 3.333431 | -0.41373 | 0.999532 | | 4.311936 | 0.012161 | 0.994356 | | 3.284667 | 0.448554 | 0.735469 |
| BRPF1 | 2.973441 | 0.060264 | 0.999532 | | 3.338968 | 0.012406 | 0.994599 | | 2.936349 | -0.03163 | 0.991159 |
| FEM1C | 6.101026 | 0.156388 | 0.999532 | | 6.63955 | -0.00933 | 0.994728 | | 5.984784 | -0.15327 | 0.892869 |
| SKIV2L | 2.909032 | 0.437029 | 0.999532 | | 2.741498 | 0.01012 | 0.994737 | | 2.775554 | -0.40914 | 0.749999 |
| ZFYVE21 | 2.300572 | 0.382146 | 0.999532 | | 2.285998 | 0.013841 | 0.994737 | | 2.140904 | -0.35079 | 0.832317 |

| | | | | | | | | | | | |
|---|---|---|---|---|---|---|---|---|---|---|---|
| TMEM159 | 4.21525 | -0.56426 | 0.999532 | | 5.246135 | -0.00914 | 0.994737 | | 3.778432 | 0.57791 | 0.572084 |
| EIF2S2 | 8.139449 | 0.154089 | 0.999532 | | 8.300808 | -0.00701 | 0.994737 | | 7.809719 | -0.14712 | 0.893298 |
| LTC4S | 4.753489 | 0.110872 | 0.999532 | | 5.250226 | -0.00821 | 0.994737 | | 4.658371 | -0.09962 | 0.946512 |
| ITPR1 | 3.727742 | -0.04516 | 0.999532 | | 3.955516 | 0.009692 | 0.994737 | | 3.571689 | 0.084362 | 0.964728 |
| EMC6 | 2.20816 | -0.11029 | 0.999532 | | 2.556311 | -0.01265 | 0.994737 | | 2.162381 | 0.116879 | 0.958889 |
| CAPZA2 | 3.046231 | 0.126675 | 0.999532 | | 3.147659 | 0.011074 | 0.994737 | | 2.935647 | -0.09729 | 0.96106 |
| DCHS2 | 3.593761 | 0.602424 | 0.999532 | | 3.185074 | 0.011711 | 0.994737 | | 3.554694 | -0.58589 | 0.662584 |
| MFSD4A | 2.269291 | 0.332941 | 0.999532 | | 2.290944 | 0.013773 | 0.994737 | | 2.266125 | -0.30209 | 0.864245 |
| TXLNG | 3.378509 | 0.395884 | 0.999532 | | 3.241505 | 0.011323 | 0.994737 | | 3.206816 | -0.36727 | 0.788243 |
| ATXN7 | 5.054454 | -0.02035 | 0.999532 | | 5.946673 | 0.009324 | 0.994737 | | 4.890867 | 0.06351 | 0.972221 |
| SH3BGR | 6.185487 | -0.73433 | 0.999532 | | 8.645491 | -0.00882 | 0.994737 | | 6.026887 | 0.735199 | 0.510286 |
| RNASET2 | 4.118059 | 0.260437 | 0.999532 | | 4.195485 | 0.009278 | 0.994737 | | 3.993771 | -0.23327 | 0.862823 |
| ZCCHC8 | 2.941422 | 0.423892 | 0.999532 | | 2.707635 | 0.011724 | 0.994737 | | 2.786172 | -0.39691 | 0.766974 |
| CNTN4 | 2.809196 | 0.424017 | 0.999532 | | 2.778479 | -0.01186 | 0.994737 | | 2.839395 | -0.4156 | 0.791576 |
| PLCD1 | 5.097583 | 0.547706 | 0.999532 | | 4.250241 | -0.0093 | 0.994737 | | 4.789819 | -0.51432 | 0.630705 |
| TMEM25 | 4.28647 | -0.11211 | 0.999532 | | 5.053214 | 0.008544 | 0.994737 | | 4.082035 | 0.137992 | 0.918675 |
| MICAL1 | 3.376352 | -0.03563 | 0.999532 | | 3.921373 | 0.01001 | 0.994737 | | 3.312085 | 0.063576 | 0.974808 |
| CLCNKA | 4.552774 | 0.70094 | 0.999532 | | 3.981573 | 0.009643 | 0.994737 | | 4.282488 | -0.66255 | 0.51741 |
| RELL1 | 5.22524 | -0.06029 | 0.999532 | | 5.904579 | -0.00835 | 0.994737 | | 5.078005 | 0.066832 | 0.965649 |
| SMAD3 | 3.130236 | -0.16763 | 0.999532 | | 3.653278 | -0.01028 | 0.994737 | | 3.082076 | 0.167723 | 0.918421 |
| ALDH18A | 2.455364 | 0.287929 | 0.999532 | | 2.457341 | -0.01169 | 0.994737 | | 2.220136 | -0.27922 | 0.872435 |
| ESPL1 | 2.827763 | 0.380799 | 0.999532 | | 2.750419 | -0.012 | 0.994737 | | 2.731944 | -0.37866 | 0.801572 |
| KRT35 | 1.877047 | -0.23001 | 0.999532 | | 2.386141 | 0.01293 | 0.994737 | | 1.946219 | 0.271631 | 0.892869 |
| PRNP | 4.224083 | -0.28182 | 0.999532 | | 4.797771 | 0.009748 | 0.994737 | | 3.731794 | 0.28779 | 0.842754 |
| PLEKHH | 3.391 | -0.16216 | 0.999532 | | 3.976529 | -0.01101 | 0.994737 | | 3.227831 | 0.174985 | 0.91015 |
| TAF2 | 5.448794 | -0.15597 | 0.999532 | | 6.585836 | -0.00783 | 0.994737 | | 5.26242 | 0.171957 | 0.892869 |
| LTV1 | 2.142894 | -0.07901 | 0.999532 | | 2.564877 | 0.011833 | 0.994737 | | 2.003505 | 0.115602 | 0.959377 |
| CAMTA2 | 5.240567 | -0.38322 | 0.999532 | | 6.639174 | 0.007283 | 0.994737 | | 5.029672 | 0.406052 | 0.682485 |
| FOXP4 | 3.498733 | -0.08382 | 0.999532 | | 4.133867 | -0.0089 | 0.994737 | | 3.167767 | 0.096891 | 0.955834 |
| SLC25A2 | 3.679136 | -0.55272 | 0.999532 | | 4.987155 | -0.0101 | 0.994737 | | 3.749529 | 0.567958 | 0.623462 |
| C16orf78 | 2.220685 | -0.27145 | 0.999532 | | 2.657518 | 0.011497 | 0.994737 | | 2.169932 | 0.305071 | 0.863907 |
| EPC2 | 3.084355 | 0.00701 | 0.999532 | | 3.349801 | -0.00986 | 0.994737 | | 2.906898 | -0.00402 | 0.998898 |
| SLC35D3 | 2.35723 | 0.396793 | 0.999532 | | 2.30308 | -0.01182 | 0.994737 | | 2.290508 | -0.3886 | 0.809485 |
| LDLRAD1 | 3.395459 | 0.829618 | 0.999532 | | 2.786517 | -0.0133 | 0.994737 | | 3.134122 | -0.82172 | 0.455747 |
| RNF41 | 3.154994 | 0.47525 | 0.999532 | | 2.840594 | -0.01133 | 0.994737 | | 2.923804 | -0.47032 | 0.713778 |
| ZMPSTE2 | 2.675082 | -0.136 | 0.999532 | | 3.002653 | -0.01099 | 0.994737 | | 2.436572 | 0.141927 | 0.939942 |
| GPATCH | 2.291433 | 0.206307 | 0.999532 | | 2.241898 | 0.012469 | 0.994737 | | 2.161303 | -0.17228 | 0.92697 |
| RPAP3 | 2.559101 | 0.259443 | 0.999532 | | 2.625883 | -0.013 | 0.994737 | | 2.382508 | -0.25395 | 0.881316 |
| LIPE | 2.298314 | 0.035502 | 0.999532 | | 2.488551 | 0.011867 | 0.994737 | | 2.211288 | -0.00633 | 0.99853 |
| TTR | 3.072395 | -0.23578 | 0.999532 | | 3.551425 | -0.00984 | 0.994737 | | 2.981213 | 0.223048 | 0.90016 |
| COL11A1 | 2.791785 | 0.434535 | 0.999532 | | 2.648259 | -0.01133 | 0.994737 | | 2.613972 | -0.42473 | 0.765852 |
| ZNF391 | 5.211178 | -0.14196 | 0.999532 | | 6.234789 | 0.008625 | 0.994737 | | 5.000586 | 0.167119 | 0.892004 |
| EFNA3 | 2.332727 | -0.06512 | 0.999532 | | 2.627118 | -0.01147 | 0.994737 | | 2.224876 | 0.072745 | 0.975473 |
| NOS1 | 2.157309 | 0.170989 | 0.999532 | | 2.133718 | 0.012826 | 0.994737 | | 2.080402 | -0.14909 | 0.946765 |
| B3GALNT | 4.098937 | 0.366219 | 0.999532 | | 4.146232 | 0.009737 | 0.994737 | | 3.881822 | -0.32776 | 0.797942 |
| RETN | 2.579616 | 0.544842 | 0.999532 | | 2.538576 | -0.01284 | 0.994737 | | 2.554328 | -0.5286 | 0.706407 |
| SESTD1 | 2.390245 | 0.247028 | 0.999532 | | 2.521641 | -0.01244 | 0.994737 | | 2.324176 | -0.242 | 0.895609 |
| SLC35D1 | 3.214483 | 0.730826 | 0.999532 | | 2.58209 | -0.01325 | 0.994737 | | 3.044102 | -0.72477 | 0.540342 |
| TFAP2C | 2.899282 | 0.575912 | 0.999532 | | 2.746873 | 0.012743 | 0.994737 | | 2.718647 | -0.53971 | 0.666566 |
| MAD2L2 | 2.448568 | 0.49451 | 0.999532 | | 2.311207 | 0.011762 | 0.994737 | | 2.434365 | -0.46446 | 0.755301 |
| RAB9A | 4.22121 | 0.029389 | 0.999532 | | 4.49349 | -0.00892 | 0.994737 | | 3.988498 | -0.02227 | 0.992286 |
| HOXB2 | 3.594959 | 0.13711 | 0.999532 | | 3.863299 | 0.01015 | 0.994737 | | 3.446407 | -0.10875 | 0.947851 |
| DEFB107 | 2.191748 | 0.110991 | 0.999532 | | 2.304528 | -0.01197 | 0.994737 | | 2.145156 | -0.11116 | 0.963359 |
| ZWILCH | 2.642492 | 0.150148 | 0.999532 | | 2.971373 | 0.010988 | 0.994737 | | 2.594353 | -0.10896 | 0.956322 |
| MBNL3 | 4.235329 | 0.361071 | 0.999532 | | 4.053156 | -0.01041 | 0.994737 | | 4.006848 | -0.35297 | 0.759211 |
| ABCC3 | 2.989403 | 0.240745 | 0.999532 | | 3.054589 | -0.01195 | 0.994737 | | 2.874518 | -0.23632 | 0.880137 |
| KLHDC3 | 5.433883 | -0.10367 | 0.999532 | | 6.335935 | 0.008356 | 0.994737 | | 5.302618 | 0.130617 | 0.917592 |
| CISD1 | 3.730429 | 0.068596 | 0.999532 | | 4.071217 | 0.010692 | 0.994737 | | 3.539739 | -0.03787 | 0.987274 |
| ITPKA | 2.221446 | 0.680901 | 0.999532 | | 1.876154 | 0.014998 | 0.994737 | | 2.196415 | -0.65097 | 0.665188 |
| HDAC1 | 7.861385 | -0.05281 | 0.999532 | | 9.198363 | 0.007371 | 0.994737 | | 7.578452 | 0.079612 | 0.946572 |
| GK5 | 2.863357 | -0.00609 | 0.999532 | | 3.226979 | -0.01173 | 0.994737 | | 2.730118 | 0.013326 | 0.996891 |
| CSAD | 2.945519 | -0.15696 | 0.999532 | | 3.361589 | 0.009819 | 0.994737 | | 2.858423 | 0.183177 | 0.91158 |
| HCRTR1 | 2.278908 | 0.25762 | 0.999532 | | 2.25158 | -0.01274 | 0.994737 | | 2.321732 | -0.27171 | 0.885892 |
| H2BC10 | 2.421812 | 0.066182 | 0.999532 | | 2.751365 | 0.011768 | 0.994737 | | 2.392109 | -0.03431 | 0.991159 |
| NR5A2 | 2.452099 | 0.346147 | 0.999532 | | 2.486334 | -0.01175 | 0.994737 | | 2.438837 | -0.33922 | 0.842754 |
| ALOX15B | 1.863553 | 0.105733 | 0.999532 | | 2.113053 | -0.01368 | 0.994737 | | 1.932097 | -0.09921 | 0.971732 |
| TPD52L1 | 2.421505 | 0.551081 | 0.999532 | | 2.101247 | 0.013696 | 0.994737 | | 2.264285 | -0.52399 | 0.720106 |
| ALPP | 1.954537 | -0.1015 | 0.999532 | | 2.284241 | 0.013571 | 0.994737 | | 1.857637 | 0.137699 | 0.955167 |
| ALYREF | 5.817643 | 0.104796 | 0.999532 | | 6.258234 | 0.007208 | 0.99479 | | 5.570855 | -0.08229 | 0.955167 |
| H1-6 | 2.184429 | 0.347693 | 0.999532 | | 2.063417 | -0.01204 | 0.99479 | | 1.996873 | -0.34149 | 0.848095 |
| LRRC57 | 2.453545 | -0.24768 | 0.999532 | | 2.805904 | -0.01 | 0.99479 | | 2.333901 | 0.259728 | 0.880577 |
| ARSA | 3.268508 | 0.367098 | 0.999532 | | 3.201717 | 0.009512 | 0.99479 | | 3.158488 | -0.33749 | 0.80013 |
| BAGE3 | 8.073219 | 0.057266 | 0.999532 | | 9.333087 | 0.00637 | 0.994797 | | 7.918392 | -0.01883 | 0.992286 |
| MAPK8IP | 3.340562 | 0.169642 | 0.999532 | | 3.646213 | 0.009831 | 0.994797 | | 3.277808 | -0.14165 | 0.934572 |
| FAAP100 | 2.760129 | -0.01022 | 0.999532 | | 3.123132 | 0.009627 | 0.994797 | | 2.690513 | 0.037311 | 0.989148 |
| CALML3 | 7.229987 | 0.4589 | 0.999532 | | 7.391678 | 0.007296 | 0.994917 | | 7.136181 | -0.41647 | 0.643272 |
| CES2 | 4.481843 | 0.046342 | 0.999532 | | 4.668807 | -0.00806 | 0.994948 | | 4.080537 | -0.0495 | 0.978723 |
| SPATA6 | 5.38385 | -0.10199 | 0.999532 | | 6.141606 | -0.00791 | 0.995008 | | 5.009718 | 0.124623 | 0.922679 |
| NACA2 | 2.259898 | 0.221999 | 0.999532 | | 2.201916 | 0.011469 | 0.995141 | | 2.143522 | -0.20228 | 0.921 |
| NTSR1 | 2.271836 | 0.180067 | 0.999532 | | 2.573425 | 0.010916 | 0.995141 | | 2.25749 | -0.14278 | 0.946572 |
| VPS26C | 2.378593 | 0.44435 | 0.999532 | | 2.327018 | -0.0111 | 0.995141 | | 2.282506 | -0.43759 | 0.774044 |
| HMBOX1 | 5.84085 | 0.024725 | 0.999532 | | 6.453666 | 0.007205 | 0.995141 | | 5.537857 | 0.00374 | 0.998585 |
| FKBP3 | 7.662537 | 0.048988 | 0.999532 | | 8.196214 | 0.005885 | 0.995141 | | 7.009717 | -0.03989 | 0.978312 |
| MARCHF | 3.326949 | 0.332537 | 0.999532 | | 3.30819 | -0.00889 | 0.995141 | | 3.133206 | -0.32323 | 0.806797 |
| CRYBG1 | 3.120956 | 0.166523 | 0.999532 | | 3.553531 | 0.009327 | 0.995141 | | 3.07026 | -0.13173 | 0.940266 |
| TXNRD3 | 2.219066 | 0.470244 | 0.999532 | | 2.079159 | -0.01095 | 0.995141 | | 2.138264 | -0.46114 | 0.765012 |
| RMDN1 | 3.733655 | -0.23594 | 0.999532 | | 4.254336 | -0.00787 | 0.99518 | | 3.406561 | 0.240134 | 0.865363 |
| ABCA9 | 2.001197 | -0.08949 | 0.999532 | | 2.312719 | -0.01056 | 0.99518 | | 1.99124 | 0.098508 | 0.969783 |
| NBPF14 | 19.10575 | 0.063882 | 0.999532 | | 21.43287 | 0.004496 | 0.99518 | | 18.41017 | -0.03551 | 0.973754 |
| MBD6 | 2.444915 | 0.261257 | 0.999532 | | 2.562965 | 0.009532 | 0.99518 | | 2.356411 | -0.23271 | 0.88885 |
| GPRC5A | 5.954674 | 0.061313 | 0.999532 | | 6.782739 | 0.008429 | 0.99518 | | 5.621645 | -0.00538 | 0.997967 |
| TRIP11 | 8.975666 | -0.21742 | 0.999532 | | 10.97109 | 0.005945 | 0.995181 | | 8.635525 | 0.247125 | 0.791576 |
| GPN3 | 2.876311 | 0.151022 | 0.999532 | | 3.022201 | 0.009144 | 0.995181 | | 2.698529 | -0.12461 | 0.946572 |
| MROH2A | 1.877199 | 0.320256 | 0.999532 | | 1.745585 | 0.011558 | 0.995227 | | 1.844414 | -0.2973 | 0.884798 |
| PKMYT1 | 7.631962 | -0.02941 | 0.999532 | | 8.560831 | -0.00623 | 0.995227 | | 7.316614 | 0.038628 | 0.97843 |
| EEF2K | 3.259576 | -0.4147 | 0.999532 | | 4.387678 | 0.007801 | 0.995227 | | 3.229705 | 0.446042 | 0.718643 |
| LRRC8D | 2.542342 | -0.41843 | 0.999532 | | 3.199808 | 0.009291 | 0.995227 | | 2.280035 | 0.440071 | 0.767131 |
| GSS | 2.344917 | -0.28137 | 0.999532 | | 2.707648 | -0.00928 | 0.995227 | | 2.188818 | 0.289747 | 0.867009 |
| FANK1 | 2.237189 | 0.507791 | 0.999532 | | 2.001713 | 0.01098 | 0.995227 | | 2.155692 | -0.48068 | 0.761337 |
| NF1 | 3.799828 | -0.38666 | 0.999532 | | 4.936346 | 0.007161 | 0.995254 | | 3.666435 | 0.412528 | 0.717627 |
| TUBA4B | 2.295849 | 0.395594 | 0.999532 | | 2.386973 | -0.0101 | 0.99535 | | 2.276171 | -0.37941 | 0.816589 |
| TFCP2L1 | 3.965338 | 0.035059 | 0.999532 | | 4.418595 | 0.008136 | 0.995438 | | 3.473812 | -0.02319 | 0.993618 |
| TANGO6 | 2.760101 | 0.242859 | 0.999532 | | 2.918854 | -0.00891 | 0.995438 | | 2.685172 | -0.22991 | 0.889563 |
| ISYNA1 | 3.614277 | 0.175999 | 0.999532 | | 4.027935 | -0.00771 | 0.995438 | | 3.579852 | -0.16205 | 0.913432 |
| TGFB1I1 | 6.035822 | -0.28889 | 0.999532 | | 7.366444 | 0.006114 | 0.995438 | | 5.797566 | 0.310116 | 0.756501 |
| MTFR1L | 4.09109 | -0.154 | 0.999532 | | 4.574219 | 0.007045 | 0.995438 | | 3.779642 | 0.173158 | 0.901916 |
| OR56A1 | 2.231699 | 0.829695 | 0.999532 | | 1.914208 | 0.010991 | 0.995438 | | 2.177043 | -0.79969 | 0.557909 |
| FXN | 4.749331 | 0.277176 | 0.999532 | | 4.718415 | -0.00823 | 0.995438 | | 4.595559 | -0.27639 | 0.827429 |
| ADAM33 | 2.771137 | 0.50263 | 0.999532 | | 2.573904 | 0.009295 | 0.995438 | | 2.713698 | -0.4745 | 0.725814 |
| CBARP | 2.648717 | 0.232392 | 0.999532 | | 2.801927 | -0.00881 | 0.995467 | | 2.492232 | -0.22243 | 0.89689 |
| LRFN3 | 2.295765 | 0.62738 | 0.999532 | | 2.134311 | 0.010019 | 0.995467 | | 2.211836 | -0.59636 | 0.669356 |
| NAT9 | 3.412586 | -0.07525 | 0.999532 | | 3.794112 | 0.007569 | 0.995467 | | 3.102078 | 0.094028 | 0.960773 |
| CREBRF | 6.09927 | 0.303146 | 0.999532 | | 6.149516 | -0.00604 | 0.995467 | | 5.872785 | -0.28945 | 0.769793 |
| NAPSA | 6.174403 | 0.555357 | 0.999532 | | 5.280671 | -0.00674 | 0.995467 | | 5.82618 | -0.54281 | 0.519634 |
| PMF1 | 2.934002 | -0.09925 | 0.999532 | | 3.435804 | 0.007655 | 0.995531 | | 2.706295 | 0.123563 | 0.945129 |
| PBRM1 | 3.617333 | -0.08358 | 0.999532 | | 4.073452 | 0.007231 | 0.995531 | | 3.493074 | 0.108048 | 0.947322 |
| FBN1 | 3.320025 | -0.29853 | 0.999532 | | 4.337795 | -0.00859 | 0.995531 | | 3.313662 | 0.325699 | 0.798849 |
| HVCN1 | 2.687896 | -0.01175 | 0.999532 | | 2.913806 | -0.00826 | 0.995531 | | 2.579016 | 0.016176 | 0.996681 |
| COQ10A | 2.868523 | 0.180913 | 0.999532 | | 3.330181 | 0.008685 | 0.995531 | | 2.991444 | -0.15841 | 0.927433 |
| ERICH2 | 2.243016 | 0.929796 | 0.999532 | | 1.847617 | 0.010451 | 0.995531 | | 2.180454 | -0.89641 | 0.499025 |
| SLC18A1 | 1.966461 | 0.639358 | 0.999532 | | 1.7567 | 0.010218 | 0.995567 | | 1.943939 | -0.61134 | 0.682165 |
| TPK1 | 2.039532 | -0.25386 | 0.999532 | | 2.686633 | -0.0093 | 0.995567 | | 2.086133 | 0.266584 | 0.890355 |
| RAMP3 | 8.663431 | -0.392 | 0.999532 | | 11.35966 | 0.004756 | 0.995578 | | 8.318776 | 0.42594 | 0.602786 |
| LPXN | 3.216371 | 0.059484 | 0.999532 | | 3.893516 | -0.00711 | 0.995631 | | 3.112808 | -0.03219 | 0.990103 |
| PPL | 2.53634 | 0.293228 | 0.999532 | | 2.458869 | -0.00844 | 0.995717 | | 2.407035 | -0.2826 | 0.859898 |
| GRM1 | 1.805711 | 0.316062 | 0.999532 | | 1.790711 | 0.009959 | 0.996095 | | 1.795914 | -0.29103 | 0.88885 |
| SETSIP | 2.025618 | 0.188317 | 0.999532 | | 1.983754 | -0.00931 | 0.996095 | | 2.015113 | -0.18242 | 0.931749 |
| ABTB1 | 6.766667 | 0.258162 | 0.999532 | | 6.910772 | 0.005448 | 0.996396 | | 6.429696 | -0.23374 | 0.806394 |
| ZDHHC1 | 3.984516 | -0.18184 | 0.999532 | | 5.05677 | -0.00594 | 0.996396 | | 3.927623 | 0.199912 | 0.882613 |
| ZNF783 | 4.216151 | -0.08337 | 0.999532 | | 4.776694 | 0.006135 | 0.996396 | | 4.254426 | 0.094811 | 0.955834 |
| C9orf85 | 3.359034 | 0.011714 | 0.999532 | | 3.746648 | 0.00692 | 0.996396 | | 3.109697 | 0.014932 | 0.996681 |
| ACBD3 | 5.056012 | -0.06053 | 0.999532 | | 5.796077 | -0.00568 | 0.996451 | | 4.81084 | 0.073355 | 0.963359 |
| FAM180B | 1.946238 | 0.358786 | 0.999532 | | 1.878989 | -0.00932 | 0.996486 | | 1.829441 | -0.35205 | 0.856551 |
| PPP1R3C | 1.821207 | 0.556897 | 0.999532 | | 1.61865 | -0.0095 | 0.996598 | | 1.757715 | -0.55166 | 0.738316 |
| ANKRD2 | 4.536056 | 0.124664 | 0.999532 | | 4.992247 | 0.005849 | 0.996598 | | 4.473411 | -0.09652 | 0.949669 |
| EEF1AKN | 3.399587 | 0.252389 | 0.999532 | | 3.476497 | -0.00672 | 0.996598 | | 3.128732 | -0.24037 | 0.865363 |
| TUBB1 | 2.142798 | 0.53559 | 0.999532 | | 2.030469 | -0.00907 | 0.996598 | | 2.134339 | -0.53205 | 0.735469 |
| FBXO22 | 2.071828 | -0.21995 | 0.999532 | | 2.377983 | -0.00795 | 0.996598 | | 2.057781 | 0.230553 | 0.906811 |
| FAM189A | 1.95573 | 0.371493 | 0.999532 | | 1.866801 | -0.00912 | 0.99663 | | 1.871806 | -0.36278 | 0.849682 |
| ZNF639 | 3.623878 | 0.278738 | 0.999532 | | 3.51626 | 0.00673 | 0.996633 | | 3.579367 | -0.24284 | 0.867976 |
| SLIT2 | 3.256773 | 0.55569 | 0.999532 | | 3.034591 | 0.007072 | 0.996642 | | 2.9694 | -0.52618 | 0.655993 |
| APC | 2.234249 | -0.24847 | 0.999532 | | 2.743486 | -0.00748 | 0.996642 | | 2.264012 | 0.253704 | 0.88885 |
| PAN2 | 3.049526 | 0.022753 | 0.999532 | | 3.350606 | -0.00683 | 0.996646 | | 3.06666 | -0.0119 | 0.996891 |
| CNTD1 | 1.936458 | 0.460196 | 0.999532 | | 1.930851 | -0.00893 | 0.996646 | | 1.972726 | -0.45451 | 0.792416 |
| MAN2B1 | 3.018487 | -0.22794 | 0.999532 | | 3.252613 | -0.00696 | 0.996646 | | 2.788859 | 0.207915 | 0.907626 |
| ZDHHC15 | 2.171296 | 0.534122 | 0.999532 | | 2.006565 | 0.008084 | 0.996661 | | 2.055227 | -0.50725 | 0.730921 |
| BRD7 | 6.22152 | -0.17953 | 0.999532 | | 7.329202 | -0.00468 | 0.996754 | | 5.665699 | 0.198491 | 0.859898 |
| PES1 | 3.793067 | -0.15206 | 0.999532 | | 4.662947 | -0.00545 | 0.996797 | | 3.626053 | 0.166142 | 0.908698 |
| OSGIN2 | 3.721895 | 0.067612 | 0.999532 | | 4.148998 | 0.005804 | 0.996797 | | 3.560475 | -0.04231 | 0.984624 |
| TEFM | 2.449225 | 0.615346 | 0.999532 | | 2.038919 | 0.007794 | 0.996797 | | 2.328056 | -0.58845 | 0.678158 |
| ZNF25 | 3.425096 | 0.069915 | 0.999532 | | 3.616277 | 0.006143 | 0.996797 | | 3.283727 | -0.05102 | 0.979996 |
| ACSS3 | 2.385325 | -0.09785 | 0.999532 | | 2.50497 | 0.007312 | 0.996797 | | 2.296618 | 0.129103 | 0.956011 |
| TSPOAP1 | 4.155418 | -0.06145 | 0.999532 | | 4.768777 | -0.00551 | 0.996797 | | 3.882913 | 0.076544 | 0.965649 |
| DR1 | 4.587357 | -0.22695 | 0.999532 | | 5.615994 | -0.00508 | 0.996797 | | 4.323906 | 0.246586 | 0.84579 |
| CELF2 | 1.994768 | 0.160677 | 0.999532 | | 1.979782 | 0.006767 | 0.996973 | | 1.880813 | -0.14542 | 0.946958 |
| IL18BP | 5.365864 | -0.39016 | 0.999532 | | 6.599154 | -0.00425 | 0.996973 | | 5.117918 | 0.407736 | 0.690673 |
| ADH1A | 2.303821 | 0.989871 | 0.999532 | | 1.825391 | 0.007339 | 0.996973 | | 2.231355 | -0.96196 | 0.413122 |
| PAK1 | 3.665725 | -0.08039 | 0.999532 | | 4.114978 | 0.004632 | 0.996973 | | 3.472467 | 0.100855 | 0.955167 |
| CYFIP2 | 4.377258 | -0.04071 | 0.999532 | | 5.143734 | 0.00525 | 0.996973 | | 4.171064 | 0.070742 | 0.96743 |
| NDUFAF | 3.858023 | 0.067827 | 0.999532 | | 4.137467 | -0.00545 | 0.996973 | | 3.544155 | -0.05679 | 0.976783 |
| CEMIP2 | 4.125145 | -0.29298 | 0.999532 | | 5.031223 | -0.00435 | 0.996973 | | 3.861605 | 0.307266 | 0.806394 |
| KATNIP | 2.903547 | 0.127371 | 0.999532 | | 3.115085 | 0.006373 | 0.996973 | | 2.83326 | -0.10048 | 0.95999 |
| NXT2 | 2.075427 | 0.058015 | 0.999532 | | 2.2051 | 0.005941 | 0.996973 | | 1.943942 | -0.03399 | 0.991434 |
| MUC5AC | 2.812405 | 0.242511 | 0.999532 | | 2.996772 | -0.0064 | 0.996973 | | 2.878244 | -0.21996 | 0.89901 |
| PTPN4 | 2.994083 | -0.12143 | 0.999532 | | 3.555767 | -0.00522 | 0.996973 | | 2.972518 | 0.135359 | 0.941197 |
| SERTAD3 | 3.575291 | -0.27348 | 0.999532 | | 4.380474 | -0.00474 | 0.996973 | | 3.206251 | 0.285644 | 0.836453 |
| CPSF4 | 2.481277 | 0.505946 | 0.999532 | | 2.307781 | -0.00677 | 0.996973 | | 2.388555 | -0.49545 | 0.719612 |
| PPP2R3A | 3.434429 | -0.44653 | 0.999532 | | 4.231934 | -0.00537 | 0.996973 | | 3.158191 | 0.472998 | 0.722703 |
| SFXN5 | 3.237634 | 0.183986 | 0.999532 | | 3.285173 | -0.0051 | 0.996973 | | 3.077528 | -0.16981 | 0.914692 |
| WBP1 | 8.529878 | -0.10128 | 0.999532 | | 9.780977 | -0.00339 | 0.996973 | | 8.252655 | 0.11877 | 0.903339 |
| CHTF18 | 1.790254 | 0.015095 | 0.999532 | | 2.093463 | 0.007358 | 0.996973 | | 1.707335 | 0.016877 | 0.996891 |
| CLSTN2 | 2.03126 | -0.09909 | 0.999532 | | 2.145771 | -0.00727 | 0.996973 | | 1.843176 | 0.102019 | 0.969691 |
| HMCES | 3.136715 | -0.45988 | 0.999532 | | 4.171623 | -0.00496 | 0.996973 | | 3.092846 | 0.474127 | 0.699971 |
| PRKCE | 4.510478 | 0.21173 | 0.999532 | | 4.752407 | -0.00519 | 0.996973 | | 4.429317 | -0.20102 | 0.87764 |
| PSMA1 | 8.555521 | -0.14228 | 0.999532 | | 9.749331 | -0.0034 | 0.996973 | | 8.243404 | 0.167245 | 0.877268 |
| PAG1 | 4.772468 | -0.29562 | 0.999532 | | 6.150993 | -0.0049 | 0.996973 | | 4.482185 | 0.322619 | 0.78258 |
| SAFB2 | 14.68496 | 0.330621 | 0.999532 | | 14.37626 | 0.003127 | 0.996973 | | 13.76502 | -0.31566 | 0.707745 |
| MARF1 | 4.495684 | -0.05636 | 0.999532 | | 5.230132 | 0.004259 | 0.996973 | | 4.158817 | 0.074175 | 0.964728 |
| MIX23 | 4.059075 | 0.159189 | 0.999532 | | 4.317713 | -0.00536 | 0.996973 | | 3.906353 | -0.14613 | 0.920487 |
| MAPK14 | 6.892536 | 0.017192 | 0.999532 | | 7.578927 | 0.003685 | 0.996973 | | 6.536245 | 0.007357 | 0.996891 |
| UBE4A | 4.774233 | -0.38056 | 0.999532 | | 6.398752 | -0.00401 | 0.996973 | | 4.478739 | 0.396947 | 0.702908 |
| SASH1 | 5.302219 | -0.0677 | 0.999532 | | 6.199618 | 0.00431 | 0.996973 | | 4.978662 | 0.094616 | 0.946512 |
| TTF2 | 2.033824 | -0.18354 | 0.999532 | | 2.541919 | -0.00682 | 0.996973 | | 2.126471 | 0.192404 | 0.928522 |
| SLC30A1 | 4.204797 | -0.15884 | 0.999532 | | 4.85075 | 0.004519 | 0.996973 | | 4.059046 | 0.173647 | 0.906811 |
| PRX | 4.621743 | 0.485389 | 0.999532 | | 4.352151 | 0.005082 | 0.996973 | | 4.412504 | -0.46152 | 0.686214 |
| MTIF2 | 3.584156 | -0.0544 | 0.999532 | | 4.202902 | 0.005638 | 0.996973 | | 3.424755 | 0.079915 | 0.965649 |
| CITED1 | 1.93952 | -0.16009 | 0.999532 | | 2.317029 | -0.00657 | 0.996973 | | 1.92575 | 0.173272 | 0.938429 |
| IGHM | 2.644404 | -0.04454 | 0.999532 | | 2.829641 | -0.00601 | 0.996973 | | 2.499009 | 0.052791 | 0.983749 |
| SCNN1A | 5.855768 | 0.356816 | 0.999532 | | 5.790238 | 0.006112 | 0.996973 | | 5.465884 | -0.31361 | 0.800239 |
| SERPINA | 2.01654 | 0.300544 | 0.999532 | | 2.164106 | 0.006409 | 0.996973 | | 2.03502 | -0.27541 | 0.881837 |
| ZNF195 | 2.503569 | -0.31053 | 0.999532 | | 2.891015 | -0.00536 | 0.996973 | | 2.469726 | 0.304775 | 0.870663 |
| CEP85 | 2.359118 | -0.00926 | 0.999532 | | 2.503898 | 0.005893 | 0.996973 | | 2.175039 | 0.025671 | 0.994192 |
| INHBA | 2.011552 | -0.30946 | 0.999532 | | 2.430475 | -0.00682 | 0.996973 | | 2.034025 | 0.317644 | 0.870694 |
| VIPAS39 | 3.252881 | 0.027416 | 0.999532 | | 3.605367 | -0.0053 | 0.996973 | | 3.165845 | -0.01344 | 0.996891 |
| FKBP7 | 3.437107 | 0.034492 | 0.999532 | | 3.561116 | -0.00504 | 0.996973 | | 3.257398 | -0.01955 | 0.995112 |
| CYB561D | 2.935617 | 0.089767 | 0.999532 | | 3.225677 | -0.00496 | 0.996973 | | 2.850497 | -0.07471 | 0.972221 |
| MAML1 | 4.960226 | 0.063774 | 0.999532 | | 5.012845 | 0.004272 | 0.996973 | | 4.520434 | -0.06507 | 0.972145 |
| UBE3D | 2.169636 | 0.140572 | 0.999532 | | 2.330844 | 0.007364 | 0.996973 | | 2.079752 | -0.1167 | 0.959426 |
| CLIP4 | 3.60769 | -0.11413 | 0.999532 | | 4.216672 | -0.00437 | 0.997068 | | 3.274802 | 0.11976 | 0.941298 |
| GFOD1 | 2.284622 | -0.39747 | 0.999532 | | 2.849514 | -0.00521 | 0.997068 | | 2.226106 | 0.410082 | 0.809832 |
| FABP3 | 3.926554 | 0.203797 | 0.999532 | | 4.283335 | 0.004423 | 0.997068 | | 3.793903 | -0.17626 | 0.901033 |
| TNFRSF9 | 2.161307 | 0.89788 | 0.999532 | | 1.795349 | -0.00647 | 0.997068 | | 2.099286 | -0.88622 | 0.515762 |
| ZNF417 | 19.32749 | 0.259584 | 0.999532 | | 19.86613 | -0.00242 | 0.997068 | | 18.4608 | -0.241 | 0.695449 |
| ABCA13 | 2.131342 | 0.370202 | 0.999532 | | 2.059269 | -0.00577 | 0.997068 | | 2.081682 | -0.35958 | 0.836453 |
| DTNB | 3.105568 | 0.420195 | 0.999532 | | 3.062769 | -0.00521 | 0.997068 | | 2.968418 | -0.40402 | 0.755206 |
| C1orf162 | 4.817075 | 0.041842 | 0.999532 | | 5.364383 | 0.004565 | 0.997068 | | 4.651414 | -0.00887 | 0.996891 |
| SLC9A5 | 2.221069 | 0.23081 | 0.999532 | | 2.314362 | 0.005617 | 0.997068 | | 2.17964 | -0.20702 | 0.908891 |
| INMT | 8.008269 | -0.38184 | 0.999532 | | 10.33854 | -0.00297 | 0.997288 | | 7.788357 | 0.404145 | 0.644433 |
| TAFA4 | 3.276095 | -0.48764 | 0.999532 | | 4.206862 | 0.004571 | 0.997288 | | 3.194392 | 0.505651 | 0.720106 |
| DNAAF1 | 2.745894 | 0.148314 | 0.999532 | | 3.04814 | -0.00489 | 0.997524 | | 2.82118 | -0.14019 | 0.94389 |
| ATG4C | 3.052581 | -0.63101 | 0.999532 | | 3.759179 | -0.00399 | 0.997524 | | 2.842155 | 0.622751 | 0.647218 |
| IMPACT | 4.25752 | -0.12996 | 0.999532 | | 4.999032 | -0.0035 | 0.997556 | | 3.846954 | 0.141068 | 0.920103 |
| ZBTB9 | 3.093628 | -0.31033 | 0.999532 | | 3.918022 | 0.004249 | 0.997556 | | 3.161129 | 0.329595 | 0.827267 |
| FBXO25 | 5.474716 | -0.45018 | 0.999532 | | 7.121616 | -0.00309 | 0.997556 | | 5.214958 | 0.465571 | 0.618451 |
| PLAC8 | 3.082372 | 0.238513 | 0.999532 | | 3.020015 | -0.00451 | 0.997556 | | 2.989489 | -0.23513 | 0.889271 |
| LSM5 | 3.364747 | -0.23446 | 0.999532 | | 4.075514 | -0.00382 | 0.997623 | | 3.2886 | 0.250196 | 0.865065 |
| TATDN2 | 3.942071 | 0.24221 | 0.999532 | | 4.024968 | 0.004034 | 0.997632 | | 3.741684 | -0.22003 | 0.87892 |
| LRRC27 | 3.247654 | 0.205163 | 0.999532 | | 3.333048 | 0.003828 | 0.997632 | | 3.178466 | -0.17978 | 0.906902 |
| ARRB2 | 5.008123 | -0.20223 | 0.999532 | | 5.553831 | 0.003421 | 0.997632 | | 4.732922 | 0.195509 | 0.89356 |
| FAT4 | 2.186525 | 0.371986 | 0.999532 | | 2.285392 | 0.004969 | 0.997632 | | 2.258344 | -0.34948 | 0.836453 |
| SH3RF2 | 2.333915 | 0.052981 | 0.999532 | | 2.571401 | -0.00466 | 0.997632 | | 2.225907 | -0.03841 | 0.990309 |
| ZBTB10 | 4.810731 | -0.2194 | 0.999532 | | 5.702882 | 0.003182 | 0.997632 | | 4.37782 | 0.240948 | 0.847755 |
| WDR82 | 8.036434 | -0.15325 | 0.999532 | | 9.124492 | 0.002515 | 0.997632 | | 7.789304 | 0.170792 | 0.880375 |
| PLEKHA1 | 4.03422 | -0.08982 | 0.999532 | | 4.344464 | -0.00366 | 0.997632 | | 3.664554 | 0.0848 | 0.965649 |
| ZNF852 | 2.526081 | 0.226932 | 0.999532 | | 2.679337 | 0.004237 | 0.997632 | | 2.539999 | -0.20819 | 0.900797 |
| H2AC21 | 7.600567 | 0.148344 | 0.999532 | | 7.908527 | -0.00256 | 0.997718 | | 7.204891 | -0.13046 | 0.90539 |
| RORA | 3.457285 | 0.088947 | 0.999532 | | 3.762031 | -0.0034 | 0.997718 | | 3.370835 | -0.07389 | 0.970532 |
| TBC1D21 | 3.341769 | 0.654202 | 0.999532 | | 3.049454 | -0.00392 | 0.997718 | | 3.342926 | -0.64263 | 0.598589 |
| HSD17B1 | 2.581881 | 0.246508 | 0.999532 | | 2.533455 | -0.00411 | 0.997718 | | 2.403673 | -0.23728 | 0.892869 |
| SMIM23 | 2.56108 | 0.169983 | 0.999532 | | 2.588918 | 0.004171 | 0.997718 | | 2.502094 | -0.14787 | 0.940266 |
| LITAF | 7.62313 | -0.08784 | 0.999532 | | 8.921394 | -0.0027 | 0.997718 | | 7.295674 | 0.105092 | 0.934701 |
| OR2F1 | 2.002189 | 0.014054 | 0.999532 | | 2.176949 | 0.004421 | 0.997718 | | 1.853664 | 0.006694 | 0.998401 |
| C18orf54 | 2.733249 | -0.05247 | 0.999532 | | 3.1352 | 0.003674 | 0.997718 | | 2.464052 | 0.054658 | 0.982609 |
| GPRC5B | 6.976198 | -0.22685 | 0.999532 | | 8.343952 | -0.00272 | 0.997718 | | 6.587642 | 0.245461 | 0.821953 |
| GATC | 4.679696 | 0.587559 | 0.999532 | | 4.257678 | -0.00304 | 0.997718 | | 4.448081 | -0.56398 | 0.620976 |
| PAAF1 | 2.760898 | 0.215633 | 0.999532 | | 2.820045 | -0.00382 | 0.997754 | | 2.501861 | -0.20118 | 0.903339 |
| PRDM5 | 3.351071 | 0.139423 | 0.999532 | | 3.653833 | 0.003215 | 0.997844 | | 3.129572 | -0.11245 | 0.947322 |
| CNIH4 | 5.421747 | 0.09637 | 0.999532 | | 5.813983 | -0.00265 | 0.997899 | | 4.913202 | -0.07401 | 0.963359 |
| SNRPF | 7.813466 | -0.08822 | 0.999532 | | 9.319883 | 0.00225 | 0.997899 | | 7.518933 | 0.121768 | 0.919003 |
| ANGEL1 | 2.971386 | -0.19474 | 0.999532 | | 3.238573 | -0.0032 | 0.997899 | | 2.789293 | 0.208417 | 0.897958 |
| TENT2 | 3.880619 | -0.10932 | 0.999532 | | 4.50163 | -0.00277 | 0.998022 | | 3.766067 | 0.124793 | 0.934572 |
| SCHIP1 | 4.130582 | -0.03165 | 0.999532 | | 4.674755 | 0.002786 | 0.998022 | | 3.914704 | 0.055898 | 0.977746 |
| AFG1L | 1.831793 | 0.184731 | 0.999532 | | 2.021621 | 0.003949 | 0.998022 | | 1.717354 | -0.1606 | 0.941197 |
| FANCI | 2.569521 | 0.475048 | 0.999532 | | 2.410986 | 0.003503 | 0.998022 | | 2.534055 | -0.45821 | 0.75038 |
| PSMD9 | 2.135922 | 0.171555 | 0.999532 | | 2.310398 | -0.00358 | 0.998022 | | 2.116259 | -0.15841 | 0.939913 |
| PACC1 | 2.746119 | -0.04154 | 0.999532 | | 2.96314 | 0.003186 | 0.998022 | | 2.589851 | 0.070247 | 0.97502 |
| SAAL1 | 3.586608 | 0.154457 | 0.999532 | | 3.798424 | -0.00277 | 0.998022 | | 3.398982 | -0.13964 | 0.926586 |
| RAB27B | 2.057933 | 0.179457 | 0.999532 | | 2.330849 | -0.00385 | 0.998022 | | 2.081148 | -0.16011 | 0.943059 |
| SAMD9L | 3.993887 | -0.07668 | 0.999532 | | 4.264117 | -0.00275 | 0.998022 | | 3.910967 | 0.092216 | 0.960773 |
| ADGRA2 | 3.907559 | -0.18932 | 0.999532 | | 4.50964 | -0.0029 | 0.998022 | | 3.701983 | 0.217566 | 0.883116 |
| SPAG7 | 4.31414 | 0.472727 | 0.999532 | | 4.029397 | 0.002689 | 0.998237 | | 4.022785 | -0.44746 | 0.664325 |
| PITHD1 | 3.948504 | -0.08965 | 0.999532 | | 4.415972 | -0.00246 | 0.998411 | | 3.748779 | 0.106503 | 0.948127 |
| ARAF | 5.854322 | -0.01037 | 0.999532 | | 6.888156 | 0.001942 | 0.998456 | | 5.538774 | 0.033673 | 0.985693 |
| PROSER | 3.372406 | 0.105274 | 0.999532 | | 3.513418 | -0.00267 | 0.998464 | | 3.010592 | -0.08422 | 0.965649 |
| KCNJ13 | 1.999265 | -0.08021 | 0.999532 | | 2.204276 | -0.00322 | 0.998506 | | 1.885246 | 0.094234 | 0.972221 |
| IRAG2 | 2.128351 | 0.948769 | 0.999532 | | 1.676615 | 0.003618 | 0.998506 | | 2.130836 | -0.92752 | 0.515762 |
| JAML | 2.561869 | 0.093306 | 0.999532 | | 2.685517 | -0.00284 | 0.998571 | | 2.476086 | -0.08548 | 0.972221 |
| STIM2 | 4.345003 | 0.005096 | 0.999532 | | 5.073043 | -0.00202 | 0.998707 | | 4.329704 | 0.003547 | 0.998655 |
| PIBF1 | 1.902943 | 0.254703 | 0.999532 | | 2.021434 | -0.00307 | 0.998707 | | 1.9314 | -0.24082 | 0.902626 |
| SPOUT1 | 4.502762 | 0.370589 | 0.999532 | | 4.386148 | 0.002077 | 0.998733 | | 4.105116 | -0.34655 | 0.752523 |
| ZSWIM3 | 2.128958 | 0.043482 | 0.999532 | | 2.371234 | -0.00263 | 0.998859 | | 2.075521 | -0.02421 | 0.995112 |
| L1TD1 | 1.98987 | 0.355172 | 0.999532 | | 2.137571 | 0.002492 | 0.999251 | | 2.107687 | -0.3291 | 0.860036 |
| FAM180A | 2.854442 | -0.171 | 0.999532 | | 3.392749 | 0.002355 | 0.999251 | | 2.641414 | 0.206393 | 0.904644 |
| PPIP5K1 | 2.271867 | -0.17898 | 0.999532 | | 2.765506 | 0.002138 | 0.999251 | | 2.31448 | 0.198788 | 0.914761 |
| TMPRSS4 | 2.071982 | 0.135546 | 0.999532 | | 2.306288 | -0.00253 | 0.999251 | | 1.965196 | -0.11495 | 0.96129 |
| GPR108 | 5.980718 | 0.086908 | 0.999532 | | 6.672442 | 0.001356 | 0.999251 | | 5.734341 | -0.06767 | 0.962752 |
| PRKD1 | 3.561701 | -0.5038 | 0.999532 | | 4.574305 | 0.001824 | 0.999251 | | 3.360267 | 0.525846 | 0.663089 |
| SLC15A4 | 3.467249 | 0.173571 | 0.999532 | | 3.562236 | -0.00189 | 0.999251 | | 3.269465 | -0.16033 | 0.92167 |
| TMEM64 | 2.347446 | 0.23152 | 0.999532 | | 2.45147 | 0.002268 | 0.999251 | | 2.441943 | -0.21161 | 0.908829 |
| AHDC1 | 8.952114 | 0.115558 | 0.999532 | | 9.723883 | 0.001009 | 0.999367 | | 8.645386 | -0.09198 | 0.938429 |
| TNFSF4 | 1.928056 | 0.363113 | 0.999532 | | 2.00226 | 0.002277 | 0.999367 | | 1.847183 | -0.33965 | 0.858767 |
| C19orf44 | 2.040849 | 0.312091 | 0.999532 | | 1.96805 | 0.001902 | 0.999367 | | 1.967389 | -0.29106 | 0.87764 |
| ZSCAN29 | 3.700576 | 0.178165 | 0.999532 | | 3.930794 | -0.0013 | 0.999367 | | 3.564679 | -0.16277 | 0.908891 |
| TEF | 3.764695 | 0.340222 | 0.999532 | | 3.719683 | -0.00131 | 0.999367 | | 3.28241 | -0.30877 | 0.816112 |
| LRFN2 | 2.533095 | 0.02375 | 0.999532 | | 2.918169 | -0.00149 | 0.999367 | | 2.378291 | 0.000622 | 0.999759 |
| SLC37A2 | 1.922627 | 0.202153 | 0.999532 | | 2.144384 | 0.002047 | 0.999367 | | 1.843547 | -0.17812 | 0.93452 |
| DDX51 | 3.414682 | 0.296337 | 0.999532 | | 3.875661 | 0.001496 | 0.999367 | | 3.394784 | -0.26983 | 0.856551 |
| INTS13 | 2.05431 | -0.24621 | 0.999532 | | 2.368064 | -0.00203 | 0.999367 | | 1.987153 | 0.267116 | 0.892004 |
| PGS1 | 2.459833 | -0.46283 | 0.999532 | | 2.961269 | 0.001526 | 0.999367 | | 2.17099 | 0.465217 | 0.760138 |
| TUBD1 | 2.736713 | 0.113543 | 0.999532 | | 2.877169 | -0.00142 | 0.999367 | | 2.606345 | -0.09534 | 0.963866 |
| MAPKAP | 12.68861 | -0.4286 | 0.999532 | | 15.97267 | 0.000864 | 0.999367 | | 11.92642 | 0.448754 | 0.479755 |
| INO80C | 2.74154 | 0.201276 | 0.999532 | | 2.875454 | 0.001566 | 0.999367 | | 2.641737 | -0.18191 | 0.915533 |
| TAP2 | 7.514216 | -0.01785 | 0.999532 | | 8.504289 | 0.001164 | 0.999367 | | 7.320061 | 0.034422 | 0.984588 |
| ANKRD13 | 3.783439 | -0.33163 | 0.999532 | | 4.60306 | -0.00142 | 0.999367 | | 3.793409 | 0.340951 | 0.796512 |
| PRRT2 | 4.343071 | -0.00135 | 0.999532 | | 4.871024 | -0.00116 | 0.999367 | | 4.151229 | 0.018564 | 0.994537 |
| CSN2 | 2.077808 | 0.148916 | 0.999532 | | 2.270324 | -0.00222 | 0.999367 | | 1.950595 | -0.12963 | 0.955167 |
| DEFB131 | 2.986355 | -0.09218 | 0.999532 | | 3.568968 | -0.00115 | 0.999369 | | 2.904803 | 0.116236 | 0.953803 |
| POFUT1 | 3.828313 | -0.09411 | 0.999532 | | 4.823296 | 0.000931 | 0.999369 | | 3.687269 | 0.126265 | 0.938429 |
| PGAP3 | 5.246052 | 0.545434 | 0.999532 | | 4.619082 | 0.000943 | 0.999369 | | 4.857519 | -0.52704 | 0.560424 |
| DLG3 | 2.273803 | 0.040235 | 0.999532 | | 2.691872 | -0.00125 | 0.999369 | | 2.207842 | -0.01391 | 0.996891 |
| TGIF2 | 4.800858 | -0.25609 | 0.999532 | | 5.977776 | 0.000726 | 0.999369 | | 4.68861 | 0.276518 | 0.809832 |
| PLTP | 4.455328 | -0.08759 | 0.999532 | | 4.927848 | -0.00109 | 0.999369 | | 4.137081 | 0.113629 | 0.941197 |
| CABIN1 | 5.149364 | -0.30048 | 0.999532 | | 6.405349 | 0.000769 | 0.999369 | | 4.832519 | 0.31741 | 0.757037 |
| OTULIN | 2.348032 | 0.15718 | 0.999532 | | 2.565712 | 0.001275 | 0.999369 | | 2.284917 | -0.13381 | 0.947322 |
| PAM16 | 3.870423 | 0.523373 | 0.999532 | | 3.802157 | 0.001024 | 0.999369 | | 3.680608 | -0.49255 | 0.68597 |
| ARHGEF | 2.195863 | 0.49539 | 0.999532 | | 2.248023 | 0.001457 | 0.999369 | | 2.232195 | -0.45775 | 0.783005 |
| GSKIP | 2.818087 | 0.151493 | 0.999532 | | 2.826202 | -0.00107 | 0.999369 | | 2.649595 | -0.13201 | 0.943463 |
| TMEM208 | 4.334047 | 0.1277 | 0.999532 | | 4.648743 | -0.00086 | 0.999369 | | 4.187575 | -0.10941 | 0.942743 |
| PDE4B | 1.971298 | 0.18488 | 0.999532 | | 1.993962 | -0.00118 | 0.999369 | | 1.871987 | -0.17501 | 0.938429 |
| PDXP | 2.390043 | -0.22233 | 0.999532 | | 3.006738 | 0.001025 | 0.999396 | | 2.411518 | 0.230439 | 0.898369 |
| NFKB2 | 5.566516 | -0.43593 | 0.999532 | | 7.303911 | 0.000602 | 0.999441 | | 5.415446 | 0.455343 | 0.628688 |
| NUMB | 6.294992 | -0.09648 | 0.999532 | | 7.166354 | -0.00057 | 0.999471 | | 5.949786 | 0.111119 | 0.93222 |
| ICAM5 | 2.035232 | -0.154 | 0.999532 | | 2.274018 | 0.000695 | 0.999573 | | 2.034812 | 0.172244 | 0.936689 |
| LDHAL6B | 2.222675 | 0.042821 | 0.999532 | | 2.571784 | -0.00056 | 0.999573 | | 2.273587 | -0.02933 | 0.992459 |
| KCTD9 | 7.797384 | 0.200224 | 0.999532 | | 8.335385 | -0.00028 | 0.999573 | | 7.694907 | -0.17608 | 0.881771 |
| STC1 | 1.776115 | -0.14973 | 0.999532 | | 2.027363 | 0.000637 | 0.999573 | | 1.704996 | 0.168886 | 0.940266 |
| CYTH4 | 2.060613 | 0.544333 | 0.999532 | | 1.951906 | 0.00046 | 0.999573 | | 2.076779 | -0.52552 | 0.723453 |
| CCDC74A | 3.426553 | 0.119719 | 0.999532 | | 3.625729 | 0.000654 | 0.999573 | | 3.246351 | -0.10205 | 0.955834 |
| KRT12 | 2.591162 | 0.346656 | 0.999532 | | 2.631112 | -0.00054 | 0.999573 | | 2.631319 | -0.33257 | 0.834045 |
| STX8 | 6.27364 | -0.15284 | 0.999532 | | 7.240696 | 0.000427 | 0.999573 | | 5.862139 | 0.170266 | 0.882935 |
| PCYT1A | 3.5332 | -0.31715 | 0.999532 | | 4.390773 | 0.00049 | 0.999573 | | 3.45304 | 0.331646 | 0.799944 |
| RAB32 | 2.349615 | 0.042885 | 0.999532 | | 2.66463 | -0.00044 | 0.999573 | | 2.238702 | -0.02733 | 0.993618 |
| WDR5 | 4.717992 | 0.324375 | 0.999532 | | 4.543341 | 0.000493 | 0.999573 | | 4.445831 | -0.30594 | 0.816618 |
| EGFL7 | 25.31855 | -0.27888 | 0.999532 | | 31.33883 | 0.000254 | 0.999573 | | 24.57643 | 0.306209 | 0.699971 |
| BRMS1L | 2.049312 | 0.122104 | 0.999532 | | 2.291803 | 0.000815 | 0.999573 | | 2.073825 | -0.09435 | 0.971092 |
| ATR | 3.138126 | 0.120036 | 0.999532 | | 3.218624 | 0.000255 | 0.999752 | | 2.996126 | -0.09671 | 0.959413 |
| GEMIN4 | 2.738238 | 0.24304 | 0.999532 | | 2.732604 | 0.000251 | 0.999754 | | 2.723921 | -0.23057 | 0.89433 |
| NMNAT1 | 2.908107 | -0.11359 | 0.999532 | | 3.416503 | 0.000145 | 0.999807 | | 2.677484 | 0.133772 | 0.941197 |
| NTN1 | 1.810983 | 0.146887 | 0.999532 | | 1.985576 | -0.00015 | 0.999807 | | 1.697599 | -0.12659 | 0.958284 |
| EVPLL | 2.172135 | 0.48858 | 0.999532 | | 2.046863 | -0.00018 | 0.999807 | | 2.014986 | -0.46881 | 0.765012 |
| MADD | 3.662877 | -0.10284 | 0.999532 | | 4.17673 | -7.78E-05 | 0.999807 | | 3.346203 | 0.122918 | 0.940266 |
