## Supplemental Table 2 for "Glomerular spatial transcriptomics of IgA nephropathy according to the presence of mesangial proliferation"

| Comparison | Direction | Category | ID | Name | Source | p-value | q-value Bc | q-value FC | q-value FD | Hit Count | Hit Count | Hit in Query List |
|---|---|---|---|---|---|---|---|---|---|---|---|---|
| M1/Control | Up | GO: Biological Proces | GO:00486 | anatomica | | 2.37E-10 | 7.55E-07 | 7.55E-07 | 6.53E-06 | 27 | 1890 | TPM1,CFH,FMNL3,CNBP,BMP2,MYOF,THBS1,MYH10,ATP5IF1,THY1,MYL9,THSD7A,TCF21,CD9,IGFBP7,PTPRM,SULF1,CDIPT,LRP2,DAG1,MACF1,ACTN1,ITGA3,ITGAV,ITGB1,ITGB3,TNFRSF21 |
| M1/Control | Up | GO: Biological Proces | GO:00082 | negative r | | 1.92E-09 | 6.12E-06 | 2.36E-06 | 2.04E-05 | 19 | 989 | TPM1,BMP2,THBS1,ATP5IF1,NDRG1,THY1,CD9,IGFBP7,PTPRM,SULF1,PARK7,FRZB,EFEMP2,APOE,NDRG2,ITGB1,TNFRSF21,ATP5F1A,PLXNB1 |
| M1/Control | Up | GO: Biological Proces | GO:00420 | wound he | | 2.22E-09 | 7.07E-06 | 2.36E-06 | 2.04E-05 | 15 | 575 | TPM1,CFH,MYOF,THBS1,MYH10,MYL9,CD9,DSP,DAG1,MACF1,CD151,ACTN1,APOE,ITGB1,ITGB3 |
| M1/Control | Up | GO: Molecular Functi | GO:00508 | cell adhesi | | 9.09E-10 | 3.95E-07 | 3.95E-07 | 2.62E-06 | 16 | 600 | THBS1,NDRG1,THY1,CD9,PTPRM,DSP,PARK7,PRDX1,MACF1,CD151,ACTN1,BCAM,ITGA3,ITGAV,ITGB1,ITGB3 |
| M1/Control | Up | GO: Biological Proces | GO:00096 | response t | | 5.68E-09 | 1.81E-05 | 3.62E-06 | 3.13E-05 | 17 | 825 | TPM1,CFH,MYOF,THBS1,MYH10,KLK6,THY1,MYL9,CD9,DSP,DAG1,MACF1,CD151,ACTN1,APOE,ITGB1,ITGB3 |
| M1/Control | Up | GO: Biological Proces | GO:00071 | cell-matri | | 4.56E-09 | 1.46E-05 | 3.62E-06 | 3.13E-05 | 11 | 273 | THBS1,THY1,EFEMP2,DAG1,MACF1,ACTN1,BCAM,ITGA3,ITGAV,ITGB1,ITGB3 |
| M1/Control | Up | GO: Biological Proces | GO:00071 | cell adhesi | | 6.83E-09 | 2.18E-05 | 3.63E-06 | 3.14E-05 | 24 | 1742 | TPM1,CFH,BMP2,THBS1,MYH10,THY1,MYL9,CD9,IGFBP7,PTPRM,DSP,SULF1,EFEMP2,DAG1,MACF1,CD151,ACTN1,BCAM,ITGA3,ITGAV,ITGB1,ITGB3,TNFRSF21,PLXNB1 |
| M1/Control | Up | GO: Cellular Compon | GO:00099 | cell surfac | | 5.90E-09 | 2.28E-06 | 2.28E-06 | 1.49E-05 | 20 | 1178 | SORT1,CFH,ATP5PF,BMP2,THBS1,ATP5IF1,THY1,CD9,BST2,SULF1,LRP2,DAG1,CD151,BCAM,ITGA3,APOE,ITGAV,ITGB1,ITGB3,ATP5F1A |
| M1/Control | Up | GO: Biological Proces | GO:00015 | blood ves | | 2.29E-08 | 7.30E-05 | 9.45E-06 | 8.17E-05 | 19 | 1152 | CFH,FMNL3,THBS1,MYH10,ATP5IF1,THY1,THSD7A,TCF21,IGFBP7,PTPRM,SULF1,EFEMP2,LRP2,DAG1,APOE,ITGAV,ITGB1,ITGB3,TNFRSF21 |
| M1/Control | Up | GO: Biological Proces | GO:00485 | blood ves | | 2.37E-08 | 7.56E-05 | 9.45E-06 | 8.17E-05 | 18 | 1029 | CFH,FMNL3,THBS1,ATP5IF1,THY1,THSD7A,TCF21,IGFBP7,PTPRM,SULF1,EFEMP2,LRP2,DAG1,APOE,ITGAV,ITGB1,ITGB3,TNFRSF21 |
| M1/Control | Up | GO: Biological Proces | GO:00019 | vasculatur | | 7.27E-08 | 2.32E-04 | 2.58E-05 | 2.23E-04 | 19 | 1239 | CFH,FMNL3,THBS1,MYH10,ATP5IF1,THY1,THSD7A,TCF21,IGFBP7,PTPRM,SULF1,EFEMP2,LRP2,DAG1,APOE,ITGAV,ITGB1,ITGB3,TNFRSF21 |
| M1/Control | Up | GO: Biological Proces | GO:00303 | regulation | | 8.25E-08 | 2.63E-04 | 2.63E-05 | 2.27E-04 | 19 | 1249 | TPM1,BMP2,THBS1,THY1,CD9,PTPRM,BST2,SULF1,PARK7,DAG1,MACF1,CD151,ITGA3,APOE,ITGAV,ITGB1,ITGB3,ATP5F1A,PLXNB1 |
| M1/Control | Up | GO: Biological Proces | GO:00009 | cell morph | | 1.16E-07 | 3.69E-04 | 3.34E-05 | 2.89E-04 | 20 | 1415 | TPM1,FMNL3,S100A6,MYH10,THY1,THSD7A,PTPRM,GAS2,LRP2,DAG1,MACF1,APLP2,ACTN1,APOE,ITGAV,MYL12B,ITGB1,ITGB3,CDC42EP3,PLXNB1 |
| M1/Control | Up | GO: Biological Proces | GO:00486 | regulation | | 1.26E-07 | 4.01E-04 | 3.34E-05 | 2.89E-04 | 9 | 223 | TPM1,BMP2,THBS1,SULF1,PARK7,EFEMP2,APOE,NDRG2,ITGB3 |
| M1/Control | Up | GO: Biological Proces | GO:00343 | cell junctic | | 1.40E-07 | 4.46E-04 | 3.43E-05 | 2.97E-04 | 16 | 904 | THBS1,MYH10,NDRG1,THY1,CD9,DSP,DAG1,MACF1,ACTN1,ITGA3,APOE,ITGAV,NDRG2,ITGB1,ITGB3,PLXNB1 |
| M1/Control | Up | GO: Biological Proces | GO:00610 | muscle str | | 1.75E-07 | 5.57E-04 | 3.98E-05 | 3.44E-04 | 17 | 1044 | TPM1,SORT1,CFH,BMP2,MYOF,MYH10,MYL6,MYL9,TCF21,CD9,DSP,EFEMP2,LRP2,DAG1,ACTN1,SETD3,ITGB1 |
| M1/Control | Up | GO: Biological Proces | GO:20001 | regulation | | 1.98E-07 | 6.31E-04 | 4.05E-05 | 3.50E-04 | 19 | 1321 | TPM1,BMP2,THBS1,THY1,CD9,PTPRM,BST2,SULF1,PARK7,DAG1,MACF1,CD151,ITGA3,APOE,ITGAV,ITGB1,ITGB3,ATP5F1A,PLXNB1 |
| M1/Control | Up | GO: Biological Proces | GO:00486 | smooth m | | 2.03E-07 | 6.49E-04 | 4.05E-05 | 3.50E-04 | 9 | 236 | TPM1,BMP2,THBS1,SULF1,PARK7,EFEMP2,APOE,NDRG2,ITGB3 |
| M1/Control | Up | GO: Biological Proces | GO:00019 | endothelia | | 2.51E-07 | 8.02E-04 | 4.72E-05 | 4.08E-04 | 9 | 242 | BMP2,THBS1,ATP5IF1,PTPRM,SULF1,APOE,ITGAV,ITGB3,ATP5F1A |
| M1/Control | Up | GO: Biological Proces | GO:00083 | regulation | | 3.03E-07 | 9.67E-04 | 5.37E-05 | 4.64E-04 | 8 | 179 | TPM1,FMNL3,MYH10,GAS2,DAG1,MYL12B,CDC42EP3,PLXNB1 |
| M1/Control | Up | GO: Cellular Compon | GO:00451 | membrane | | 8.74E-08 | 3.38E-05 | 1.16E-05 | 7.55E-05 | 12 | 450 | MYOF,THY1,BST2,SULF1,PARK7,LRP2,DAG1,LAMP2,ITGB1,LDHB,ATP5F1A,VDAC2 |
| M1/Control | Up | GO: Cellular Compon | GO:00988 | membrane | | 8.96E-08 | 3.47E-05 | 1.16E-05 | 7.55E-05 | 12 | 451 | MYOF,THY1,BST2,SULF1,PARK7,LRP2,DAG1,LAMP2,ITGB1,LDHB,ATP5F1A,VDAC2 |
| M1/Control | Up | GO: Biological Proces | GO:00723 | circulatory | | 3.46E-07 | 1.10E-03 | 5.81E-05 | 5.02E-04 | 23 | 1977 | TPM1,CFH,FMNL3,BMP2,THBS1,MYH10,ATP5IF1,THY1,THSD7A,TCF21,IGFBP7,PTPRM,DSP,SULF1,EFEMP2,LRP2,DAG1,ITGA3,APOE,ITGAV,ITGB1,ITGB3,TNFRSF21 |
| M1/Control | Up | GO: Biological Proces | GO:00400 | regulation | | 3.88E-07 | 1.24E-03 | 5.89E-05 | 5.09E-04 | 19 | 1380 | TPM1,BMP2,THBS1,THY1,CD9,PTPRM,BST2,SULF1,PARK7,DAG1,MACF1,CD151,ITGA3,APOE,ITGAV,ITGB1,ITGB3,ATP5F1A,PLXNB1 |
| M1/Control | Up | GO: Biological Proces | GO:00330 | muscle cel | | 3.85E-07 | 1.23E-03 | 5.89E-05 | 5.09E-04 | 10 | 334 | TPM1,BMP2,THBS1,MYH10,SULF1,PARK7,EFEMP2,APOE,NDRG2,ITGB3 |
| M1/Control | Up | GO: Biological Proces | GO:00315 | cell-substr | | 4.41E-07 | 1.41E-03 | 6.39E-05 | 5.52E-04 | 11 | 428 | THBS1,THY1,EFEMP2,DAG1,MACF1,ACTN1,BCAM,ITGA3,ITGAV,ITGB1,ITGB3 |
| M1/Control | Up | GO: Biological Proces | GO:00015 | angiogene | | 5.33E-07 | 1.70E-03 | 7.39E-05 | 6.39E-04 | 15 | 873 | CFH,FMNL3,THBS1,ATP5IF1,THY1,THSD7A,TCF21,IGFBP7,PTPRM,SULF1,DAG1,ITGAV,ITGB1,ITGB3,TNFRSF21 |
| M1/Control | Up | GO: Cellular Compon | GO:00306 | cytoplasm | | 5.60E-07 | 2.17E-04 | 5.42E-05 | 3.54E-04 | 18 | 1266 | SORT1,MYOF,LAPTM4A,NDRG1,CD9,DSP,BST2,LRP2,APLP2,LAMP2,ACTN1,APOE,ITM2B,ITGAV,RAB2A,ITGB1,ITGB3,TMED3 |
| M1/Control | Up | GO: Molecular Functi | GO:00508 | extracellul | | 5.59E-09 | 2.43E-06 | 1.21E-06 | 8.07E-06 | 7 | 65 | THBS1,DAG1,BCAM,ITGA3,ITGAV,ITGB1,ITGB3 |
| M1/Control | Up | GO: Molecular Functi | GO:00051 | integrin bi | | 1.97E-08 | 8.54E-06 | 2.85E-06 | 1.89E-05 | 9 | 171 | THBS1,THY1,CD9,CD151,ACTN1,ITGA3,ITGAV,ITGB1,ITGB3 |
| M1/Control | Up | GO: Biological Proces | GO:00329 | cellular co | | 6.10E-07 | 1.95E-03 | 8.11E-05 | 7.01E-04 | 17 | 1141 | TPM1,S100A6,MYH10,THY1,MYL9,THSD7A,CD9,PTPRM,LRP2,DAG1,MACF1,APLP2,ACTN1,APOE,ITGB1,ITGB3,PLXNB1 |
| M1/Control | Up | GO: Biological Proces | GO:00426 | muscle cel | | 7.20E-07 | 2.30E-03 | 9.10E-05 | 7.87E-04 | 13 | 658 | TPM1,SORT1,CFH,BMP2,MYOF,MYH10,MYL9,CD9,EFEMP2,DAG1,ACTN1,SETD3,ITGB1 |
| M1/Control | Up | GO: Biological Proces | GO:00019 | negative r | | 7.42E-07 | 2.37E-03 | 9.10E-05 | 7.87E-04 | 6 | 86 | THBS1,ATP5IF1,PTPRM,SULF1,APOE,ATP5F1A |
| M1/Control | Up | GO: Biological Proces | GO:00352 | tube morp | | 9.81E-07 | 3.13E-03 | 1.12E-04 | 9.66E-04 | 19 | 1467 | CFH,FMNL3,BMP2,THBS1,ATP5IF1,THY1,THSD7A,TCF21,IGFBP7,PTPRM,SULF1,EFEMP2,LRP2,DAG1,APOE,ITGAV,ITGB1,ITGB3,TNFRSF21 |
| M1/Control | Up | GO: Biological Proces | GO:00486 | negative r | | 9.72E-07 | 3.10E-03 | 1.12E-04 | 9.66E-04 | 6 | 90 | TPM1,BMP2,PARK7,EFEMP2,APOE,NDRG2 |
| M1/Control | Up | GO: Cellular Compon | GO:00125 | vesicle me | | 7.28E-07 | 2.82E-04 | 5.63E-05 | 3.68E-04 | 18 | 1289 | SORT1,MYOF,LAPTM4A,NDRG1,CD9,DSP,BST2,LRP2,APLP2,LAMP2,ACTN1,APOE,ITM2B,ITGAV,RAB2A,ITGB1,ITGB3,TMED3 |
| M1/M0 | Up | GO: Biological Proces | GO:00019 | vasculatur | | 1.14E-14 | 2.42E-11 | 2.28E-11 | 1.88E-10 | 16 | 1239 | COL4A1,HSPG2,FLNA,COL18A1,TCF4,GJA5,HEG1,CTNNB1,IGFBP7,RCAN1,ZFP36L1,ECM1,ENG,ITGB1,VCAM1,MSN |
| M1/M0 | Up | GO: Biological Proces | GO:00485 | blood ves | | 2.14E-14 | 4.57E-11 | 2.28E-11 | 1.88E-10 | 15 | 1029 | COL4A1,HSPG2,FLNA,COL18A1,TCF4,GJA5,HEG1,CTNNB1,IGFBP7,RCAN1,ZFP36L1,ECM1,ENG,ITGB1,MSN |
| M1/Control | Up | GO: Molecular Functi | GO:00432 | laminin bi | | 1.39E-07 | 6.03E-05 | 1.51E-05 | 1.00E-04 | 5 | 32 | THBS1,DAG1,BCAM,ITGA3,ITGB1 |
| M1/Control | Up | GO: Biological Proces | GO:00019 | regulation | | 1.06E-06 | 3.36E-03 | 1.16E-04 | 1.00E-03 | 8 | 211 | BMP2,THBS1,ATP5IF1,PTPRM,SULF1,APOE,ITGB3,ATP5F1A |
| M1/Control | Up | GO: Biological Proces | GO:00226 | regulation | | 1.19E-06 | 3.78E-03 | 1.26E-04 | 1.09E-03 | 10 | 378 | TPM1,FMNL3,MYH10,GAS2,DAG1,MACF1,MYL12B,ITGB3,CDC42EP3,PLXNB1 |
| M1/Control | Up | GO: Biological Proces | GO:00108 | positive re | | 1.49E-06 | 4.75E-03 | 1.53E-04 | 1.32E-03 | 7 | 153 | THBS1,THY1,EFEMP2,DAG1,ITGA3,ITGB1,ITGB3 |
| M1/Control | Up | GO: Biological Proces | GO:00400 | locomotio | | 1.70E-06 | 5.41E-03 | 1.69E-04 | 1.46E-03 | 21 | 1834 | TPM1,BMP2,THBS1,MYH10,THY1,CD9,PTPRM,BST2,SULF1,PARK7,LRP2,DAG1,MACF1,CD151,ITGA3,APOE,ITGAV,ITGB1,ITGB3,ATP5F1A,PLXNB1 |
| M1/Control | Up | GO: Biological Proces | GO:00300 | actin filam | | 2.01E-06 | 6.42E-03 | 1.95E-04 | 1.68E-03 | 15 | 971 | TPM1,FMNL3,MYH10,MYL6,MYL9,THSD7A,DSP,BST2,GAS2,ACTN1,CGNL1,SETD3,ITGB1,ITGB3,CDC42EP3 |
| M1/Control | Up | GO: Biological Proces | GO:00323 | negative r | | 2.29E-06 | 7.29E-03 | 2.15E-04 | 1.86E-03 | 5 | 58 | ATP5PF,THBS1,APOE,ITGAV,ITGB3 |
| M1/Control | Up | GO: Biological Proces | GO:00352 | tube deve | | 2.52E-06 | 8.05E-03 | 2.30E-04 | 1.99E-03 | 21 | 1880 | CFH,FMNL3,BMP2,THBS1,MYH10,ATP5IF1,THY1,THSD7A,TCF21,IGFBP7,PTPRM,SULF1,EFEMP2,LRP2,DAG1,ITGA3,APOE,ITGAV,ITGB1,ITGB3,TNFRSF21 |
| M1/Control | Up | GO: Biological Proces | GO:00226 | regulation | | 3.08E-06 | 9.81E-03 | 2.72E-04 | 2.35E-03 | 17 | 1284 | TPM1,FMNL3,BMP2,THBS1,MYH10,THY1,PTPRM,SULF1,GAS2,DAG1,MACF1,APOE,MYL12B,ITGB1,ITGB3,CDC42EP3,PLXNB1 |
| M1/Control | Up | GO: Biological Proces | GO:00070 | cell-substr | | 3.15E-06 | 1.01E-02 | 2.72E-04 | 2.35E-03 | 6 | 110 | THBS1,THY1,MACF1,ACTN1,ITGAV,ITGB3 |
| M1/Control | Up | GO: Biological Proces | GO:01501 | cell-substr | | 4.30E-06 | 1.37E-02 | 3.60E-04 | 3.12E-03 | 6 | 116 | THBS1,THY1,MACF1,ACTN1,ITGAV,ITGB3 |
| M1/Control | Up | GO: Biological Proces | GO:00615 | axon deve | | 4.71E-06 | 1.50E-02 | 3.85E-04 | 3.33E-03 | 13 | 780 | S100A6,MYH10,THY1,THSD7A,PTPRM,LRP2,DAG1,MACF1,APLP2,APOE,ITGB1,TNFRSF21,PLXNB1 |
| M1/Control | Up | GO: Biological Proces | GO:00108 | regulation | | 4.84E-06 | 1.54E-02 | 3.86E-04 | 3.34E-03 | 8 | 259 | THBS1,THY1,EFEMP2,DAG1,MACF1,ITGA3,ITGB1,ITGB3 |
| M1/Control | Up | GO: Biological Proces | GO:00009 | cell morph | | 7.44E-06 | 2.37E-02 | 5.38E-04 | 4.65E-03 | 15 | 1081 | S100A6,MYH10,THY1,THSD7A,PTPRM,LRP2,DAG1,MACF1,APLP2,ACTN1,APOE,ITGAV,ITGB1,ITGB3,PLXNB1 |
| M1/Control | Up | GO: Cellular Compon | GO:00098 | external si | | 1.82E-06 | 7.04E-04 | 1.13E-04 | 7.39E-04 | 12 | 599 | CFH,THBS1,THY1,CD9,LRP2,DAG1,BCAM,ITGA3,APOE,ITGAV,ITGB1,ITGB3 |
| M1/M0 | Up | GO: Biological Proces | GO:00015 | blood ves | | 1.12E-13 | 2.38E-10 | 7.94E-11 | 6.54E-10 | 15 | 1152 | COL4A1,HSPG2,FLNA,COL18A1,TCF4,GJA5,HEG1,CTNNB1,IGFBP7,RCAN1,ZFP36L1,ECM1,ENG,ITGB1,MSN |
| M1/M0 | Up | GO: Biological Proces | GO:00352 | tube morp | | 3.74E-12 | 7.96E-09 | 1.99E-09 | 1.64E-08 | 15 | 1467 | COL4A1,HSPG2,FLNA,COL18A1,TCF4,GJA5,HEG1,CTNNB1,IGFBP7,RCAN1,ZFP36L1,ECM1,ENG,ITGB1,MSN |
| M1/M0 | Up | GO: Biological Proces | GO:00723 | circulatory | | 1.56E-11 | 3.32E-08 | 6.64E-09 | 5.47E-08 | 16 | 1977 | COL4A1,HSPG2,FLNA,COL18A1,TCF4,GJA5,HEG1,CTNNB1,IGFBP7,RCAN1,ZFP36L1,ECM1,ENG,ITGB1,VCAM1,MSN |
| M1/M0 | Up | GO: Biological Proces | GO:00015 | angiogene | | 4.92E-11 | 1.05E-07 | 1.75E-08 | 1.44E-07 | 12 | 873 | COL4A1,HSPG2,FLNA,COL18A1,TCF4,GJA5,CTNNB1,IGFBP7,RCAN1,ECM1,ENG,ITGB1 |
| M1/Control | Up | GO: Molecular Functi | GO:00019 | fibronectin | | 5.03E-07 | 2.18E-04 | 3.97E-05 | 2.64E-04 | 5 | 41 | THBS1,ITGA3,ITGAV,ITGB1,ITGB3 |
| M1/Control | Up | GO: Biological Proces | GO:00106 | epithelial | | 6.93E-06 | 2.21E-02 | 5.38E-04 | 4.65E-03 | 10 | 461 | THBS1,THSD7A,PTPRM,MACF1,ITGA3,APOE,ITGAV,ITGB1,ITGB3,ATP5F1A |
| M1/Control | Up | GO: Biological Proces | GO:00901 | epithelium | | 7.76E-06 | 2.47E-02 | 5.38E-04 | 4.65E-03 | 10 | 467 | THBS1,THSD7A,PTPRM,MACF1,ITGA3,APOE,ITGAV,ITGB1,ITGB3,ATP5F1A |
| M1/Control | Up | GO: Biological Proces | GO:00109 | cellular co | | 7.42E-06 | 2.37E-02 | 5.38E-04 | 4.65E-03 | 7 | 195 | TPM1,MYH10,MYL9,CD9,DAG1,ACTN1,ITGB1 |
| M1/Control | Up | GO: Biological Proces | GO:00341 | homotypic | | 7.25E-06 | 2.31E-02 | 5.38E-04 | 4.65E-03 | 6 | 127 | CFH,MYL9,CD9,DSP,ACTN1,ITGB3 |
| M1/Control | Up | GO: Cellular Compon | GO:00312 | cell leadin | | 2.05E-06 | 7.92E-04 | 1.13E-04 | 7.39E-04 | 11 | 499 | TPM1,S100A6,MYH10,THY1,PTPRM,DAG1,MACF1,ACTN1,ITGAV,ITGB1,ITGB3 |
| M1/Control | Up | GO: Cellular Compon | GO:00985 | side of me | | 2.41E-06 | 9.34E-04 | 1.17E-04 | 7.63E-04 | 14 | 853 | CFH,S100A6,THBS1,MYH10,THY1,CD9,LRP2,DAG1,BCAM,ITGA3,APOE,ITGAV,ITGB1,ITGB3 |
| M1/Control | Up | GO: Cellular Compon | GO:00164 | myosin co | | 2.75E-06 | 1.07E-03 | 1.18E-04 | 7.74E-04 | 5 | 60 | MYH10,MYL6,MYL9,CGNL1,MYL12B |
| M1/Control | Up | GO: Cellular Compon | GO:00164 | myosin II | | 3.24E-06 | 1.25E-03 | 1.25E-04 | 8.20E-04 | 4 | 28 | MYH10,MYL6,MYL9,MYL12B |
| M1/Control | Up | GO: Cellular Compon | GO:00620 | collagen-c | | 4.44E-06 | 1.72E-03 | 1.56E-04 | 1.02E-03 | 11 | 541 | S100A6,THBS1,IGFBP7,SULF1,EFEMP2,DAG1,CD151,BCAM,APOE,ITGB1,ITIH5 |
| M1/M0 | Up | GO: Biological Proces | GO:00352 | tube deve | | 1.31E-10 | 2.79E-07 | 3.98E-08 | 3.28E-07 | 15 | 1880 | COL4A1,HSPG2,FLNA,COL18A1,TCF4,GJA5,HEG1,CTNNB1,IGFBP7,RCAN1,ZFP36L1,ECM1,ENG,ITGB1,MSN |
| M1/Control | Up | GO: Biological Proces | GO:00341 | heterotypi | | 7.68E-06 | 2.45E-02 | 5.38E-04 | 4.65E-03 | 5 | 74 | THY1,DSP,ITGAV,ITGB1,ITGB3 |
| M1/Control | Up | GO: Biological Proces | GO:00074 | axonogen | | 9.25E-06 | 2.95E-02 | 5.95E-04 | 5.14E-03 | 12 | 705 | S100A6,MYH10,THY1,THSD7A,PTPRM,LRP2,DAG1,MACF1,APLP2,APOE,ITGB1,PLXNB1 |
| M1/Control | Up | GO: Biological Proces | GO:00901 | tissue mig | | 9.33E-06 | 2.98E-02 | 5.95E-04 | 5.14E-03 | 10 | 477 | THBS1,THSD7A,PTPRM,MACF1,ITGA3,APOE,ITGAV,ITGB1,ITGB3,ATP5F1A |
| M1/Control | Up | GO: Biological Proces | GO:00550 | muscle cel | | 8.87E-06 | 2.83E-02 | 5.95E-04 | 5.14E-03 | 9 | 373 | TPM1,BMP2,MYOF,MYH10,MYL9,EFEMP2,DAG1,ACTN1,ITGB1 |
| M1/Control | Up | GO: Biological Proces | GO:00336 | regulation | | 9.01E-06 | 2.87E-02 | 5.95E-04 | 5.14E-03 | 4 | 36 | BMP2,ITGAV,ITGB3,PLXNB1 |
| M1/Control | Up | GO: Biological Proces | GO:19059 | negative r | | 9.95E-06 | 3.17E-02 | 6.22E-04 | 5.38E-03 | 5 | 78 | ATP5PF,THBS1,APOE,ITGAV,ITGB3 |
| M1/Control | Up | GO: Biological Proces | GO:00301 | regulation | | 1.02E-05 | 3.26E-02 | 6.26E-04 | 5.41E-03 | 14 | 971 | TPM1,BMP2,THBS1,THY1,CD9,EFEMP2,DAG1,MACF1,ITGA3,ITGAV,ITGB1,ITGB3,TNFRSF21,PLXNB1 |
| M1/Control | Up | GO: Biological Proces | GO:19907 | cellular de | | 1.12E-05 | 3.57E-02 | 6.74E-04 | 5.82E-03 | 6 | 137 | ESD,PARK7,SELENOF,PRDX1,AKR1A1,APOE |
| M1/Control | Up | GO: Biological Proces | GO:00070 | cytoskelet | | 1.16E-05 | 3.69E-02 | 6.83E-04 | 5.91E-03 | 19 | 1736 | TPM1,FMNL3,MYH10,THY1,MYL9,THSD7A,DSP,BST2,GAS2,DAG1,MACF1,ACTN1,CGNL1,APOE,SETD3,ITGB1,ITGB3,CDC42EP3,PLXNB1 |
| M1/Control | Up | GO: Biological Proces | GO:00511 | striated m | | 1.18E-05 | 3.76E-02 | 6.83E-04 | 5.91E-03 | 10 | 490 | TPM1,SORT1,BMP2,MYOF,MYH10,MYL9,CD9,DAG1,ACTN1,ITGB1 |
| M1/Control | Up | GO: Biological Proces | GO:00100 | glial cell d | | 1.22E-05 | 3.88E-02 | 6.92E-04 | 5.99E-03 | 9 | 388 | S100A6,BMP2,NDRG1,CD9,NFE2L1,DAG1,APOE,ITGB1,TNFRSF21 |
| M1/Control | Up | GO: Cellular Compon | GO:00083 | integrin co | | 5.63E-06 | 2.18E-03 | 1.82E-04 | 1.19E-03 | 4 | 32 | ITGA3,ITGAV,ITGB1,ITGB3 |
| M1/Control | Up | GO: Cellular Compon | GO:00059 | adherens j | | 1.02E-05 | 3.94E-03 | 2.90E-04 | 1.90E-03 | 7 | 204 | NDRG1,PTPRM,DSP,PARK7,DAG1,ACTN1,ITGB1 |
| M1/Control | Up | GO: Cellular Compon | GO:00017 | ruffle | | 1.05E-05 | 4.06E-03 | 2.90E-04 | 1.90E-03 | 7 | 205 | TPM1,S100A6,MACF1,ACTN1,ITGAV,ITGB1,ITGB3 |
| M1/Control | Up | GO: Cellular Compon | GO:00301 | secretory | | 1.28E-05 | 4.96E-03 | 3.05E-04 | 2.00E-03 | 14 | 987 | THBS1,KLK6,CD9,DSP,BST2,APLP2,LAMP2,ACTN1,ITGAV,RAB2A,ITGB1,ITGB3,GNS,VDAC2 |
| M1/Control | Up | GO: Molecular Functi | GO:00199 | C-X3-C ch | | 5.49E-07 | 2.38E-04 | 3.97E-05 | 2.64E-04 | 3 | 5 | ITGAV,ITGB1,ITGB3 |
| M1/Control | Up | GO: Biological Proces | GO:00099 | negative r | | 1.33E-05 | 4.24E-02 | 7.07E-04 | 6.11E-03 | 18 | 1591 | BMP2,THBS1,THY1,TCF21,SLC25A6,SULF1,PARK7,FRZB,LRP2,DAG1,CGNL1,ITGA3,APOE,ITGAV,NDRG2,ITGB1,PTTG1IP,VDAC2 |
| M1/Control | Up | GO: Biological Proces | GO:00487 | tissue mor | | 1.29E-05 | 4.11E-02 | 7.07E-04 | 6.11E-03 | 15 | 1132 | TPM1,BMP2,TCF21,IGFBP7,DSP,SULF1,FRZB,EFEMP2,LRP2,DAG1,MACF1,CD151,ITGA3,ITGB1,ITGB3 |
| M1/Control | Up | GO: Biological Proces | GO:00511 | negative r | | 1.33E-05 | 4.23E-02 | 7.07E-04 | 6.11E-03 | 13 | 859 | TPM1,THBS1,THY1,SLC25A6,PARK7,LAMP2,CGNL1,ITGA3,APOE,ITGAV,ITGB1,ITGB3,VDAC2 |
| M1/Control | Up | GO: Biological Proces | GO:00170 | protein im | | 1.32E-05 | 4.21E-02 | 7.07E-04 | 6.11E-03 | 3 | 13 | LRP2,LAMP2,APOE |
| M1/Control | Up | GO: Biological Proces | GO:00300 | actin cytos | | 1.54E-05 | 4.90E-02 | 7.96E-04 | 6.88E-03 | 13 | 871 | TPM1,FMNL3,MYH10,MYL9,THSD7A,BST2,GAS2,ACTN1,CGNL1,SETD3,ITGB1,ITGB3,CDC42EP3 |
| M1/Control | Up | GO: Biological Proces | GO:00019 | regulation | | 1.55E-05 | 4.93E-02 | 7.96E-04 | 6.88E-03 | 6 | 145 | THBS1,THY1,EFEMP2,DAG1,MACF1,ITGB3 |
| M1/Control | Up | GO: Biological Proces | GO:00420 | gliogenesi | | 1.61E-05 | 5.13E-02 | 8.14E-04 | 7.04E-03 | 10 | 508 | S100A6,BMP2,NDRG1,CD9,NFE2L1,LRP2,DAG1,APOE,ITGB1,TNFRSF21 |
| M1/Control | Up | GO: Biological Proces | GO:00972 | cellular re | | 1.95E-05 | 6.21E-02 | 9.70E-04 | 8.39E-03 | 6 | 151 | ESD,PARK7,SELENOF,PRDX1,AKR1A1,APOE |
| M1/Control | Up | GO: Biological Proces | GO:00336 | positive re | | 2.09E-05 | 6.67E-02 | 1.03E-03 | 8.87E-03 | 3 | 15 | BMP2,ITGAV,ITGB3 |
| M1/Control | Up | GO: Biological Proces | GO:00336 | osteoblast | | 2.22E-05 | 7.09E-02 | 1.07E-03 | 9.28E-03 | 4 | 45 | BMP2,ITGAV,ITGB3,PLXNB1 |
| M1/Control | Up | GO: Biological Proces | GO:00303 | negative r | | 2.59E-05 | 8.26E-02 | 1.23E-03 | 1.07E-02 | 9 | 427 | TPM1,THBS1,THY1,PTPRM,BST2,SULF1,PARK7,DAG1,APOE |
| M1/Control | Up | GO: Biological Proces | GO:00720 | nephron d | | 2.84E-05 | 9.06E-02 | 1.31E-03 | 1.13E-02 | 7 | 240 | CFH,BMP2,THSD7A,TCF21,SULF1,ITGA3,ITGB3 |
| M1/Control | Up | GO: Cellular Compon | GO:00346 | integrin al | | 1.34E-05 | 5.19E-03 | 3.05E-04 | 2.00E-03 | 2 | 2 | ITGA3,ITGB1 |
| M1/Control | Up | GO: Cellular Compon | GO:00346 | integrin al | | 1.34E-05 | 5.19E-03 | 3.05E-04 | 2.00E-03 | 2 | 2 | ITGB1,ITGB3 |
| M1/Control | Up | GO: Cellular Compon | GO:00432 | myelin she | | 1.76E-05 | 6.81E-03 | 3.78E-04 | 2.47E-03 | 7 | 222 | NDRG1,THY1,PRDX1,ITGB1,LDHB,ATP5F1A,VDAC2 |
| M1/Control | Up | GO: Cellular Compon | GO:00988 | plasma me | | 2.47E-05 | 9.54E-03 | 4.15E-04 | 2.71E-03 | 17 | 1498 | TPM1,SORT1,MYOF,MYH10,THY1,SLC22A8,CD9,DSP,BST2,LRP2,DAG1,MACF1,AKR1A1,ITGA3,ITGAV,ITGB1,ITGB3 |
| M1/Control | Up | GO: Cellular Compon | GO:00300 | lamellipod | | 2.21E-05 | 8.55E-03 | 4.15E-04 | 2.71E-03 | 7 | 230 | MYH10,PTPRM,DAG1,ACTN1,ITGAV,ITGB1,ITGB3 |
| M1/M0 | Up | GO: Biological Proces | GO:00486 | anatomica | | 2.15E-09 | 4.57E-06 | 5.71E-07 | 4.71E-06 | 14 | 1890 | COL4A1,HSPG2,FLNA,COL18A1,TCF4,GJA5,CD81,CTNNB1,IGFBP7,RCAN1,ECM1,ENG,ITGB1,VCAM1 |
| M1/M0 | Up | GO: Biological Proces | GO:00071 | cell adhesi | | 1.25E-07 | 2.66E-04 | 2.95E-05 | 2.43E-04 | 12 | 1742 | HSPG2,FLNA,COL18A1,CD81,CTNNB1,IGFBP7,ZFP36L1,ENG,ITGB1,VCAM1,LRRC32,MSN |
| M1/M0 | Up | GO: Biological Proces | GO:00031 | endothelia | | 1.71E-07 | 3.64E-04 | 3.64E-05 | 3.00E-04 | 6 | 224 | COL18A1,GJA5,HEG1,CTNNB1,ENG,MSN |
| M1/M0 | Up | GO: Biological Proces | GO:00714 | cellular re | | 3.29E-07 | 7.01E-04 | 6.38E-05 | 5.26E-04 | 12 | 1903 | COL4A1,FLNA,GJA5,CTNNB1,IGFBP7,ZFP36L1,ENG,ITGB1,SGK1,VCAM1,LRRC32,PGRMC2 |
| M1/Control | Up | GO: Molecular Functi | GO:00448 | protein-co | | 1.46E-06 | 6.35E-04 | 9.08E-05 | 6.04E-04 | 21 | 1726 | TPM1,SORT1,FMNL3,ATP5PF,THBS1,MYH10,ATP5IF1,THY1,CD9,NFE2L1,PARK7,GAS2,LRP2,MACF1,CD151,ACTN1,ITGA3,APOE,ITGAV,ITGB1,ITGB3 |
| M1/Control | Up | GO: Molecular Functi | GO:00080 | cytoskelet | | 4.79E-06 | 2.08E-03 | 2.08E-04 | 1.38E-03 | 16 | 1123 | TPM1,FMNL3,S100A6,GLYAT,MYH10,NDRG1,MYL9,GAS2,DAG1,MACF1,ACTN1,APOE,SETD3,MYL12B,ITGB1,CDC42EP3 |
| M1/Control | Up | GO: Biological Proces | GO:00480 | focal adhe | | 2.88E-05 | 9.18E-02 | 1.31E-03 | 1.13E-02 | 5 | 97 | THBS1,THY1,MACF1,ACTN1,ITGAV |
| M1/Control | Up | GO: Biological Proces | GO:00705 | platelet ac | | 2.88E-05 | 9.18E-02 | 1.31E-03 | 1.13E-02 | 5 | 97 | CFH,MYL9,CD9,ACTN1,ITGB3 |
| M1/Control | Up | GO: Biological Proces | GO:19025 | negative r | | 3.03E-05 | 9.66E-02 | 1.36E-03 | 1.18E-02 | 11 | 667 | BMP2,THBS1,PARK7,DAG1,CGNL1,ITGA3,APOE,NDRG2,ITGB1,PTTG1IP,VDAC2 |
| M1/Control | Up | GO: Biological Proces | GO:00301 | platelet ac | | 3.32E-05 | 1.06E-01 | 1.47E-03 | 1.27E-02 | 6 | 166 | CFH,MYL9,CD9,ACTN1,APOE,ITGB3 |
| M1/Control | Up | GO: Biological Proces | GO:20001 | negative r | | 3.51E-05 | 1.12E-01 | 1.54E-03 | 1.33E-02 | 9 | 444 | TPM1,THBS1,THY1,PTPRM,BST2,SULF1,PARK7,DAG1,APOE |
| M1/Control | Up | GO: Biological Proces | GO:00075 | muscle or | | 3.92E-05 | 1.25E-01 | 1.68E-03 | 1.46E-02 | 10 | 564 | TPM1,BMP2,MYL6,TCF21,DSP,EFEMP2,LRP2,DAG1,ACTN1,ITGB1 |
| M1/Control | Up | GO: Biological Proces | GO:19047 | negative r | | 3.96E-05 | 1.26E-01 | 1.68E-03 | 1.46E-02 | 4 | 52 | TPM1,PARK7,EFEMP2,NDRG2 |
| M1/Control | Up | GO: Biological Proces | GO:00488 | cell projec | | 4.15E-05 | 1.32E-01 | 1.74E-03 | 1.51E-02 | 13 | 958 | S100A6,MYH10,THY1,THSD7A,PTPRM,LRP2,DAG1,MACF1,APLP2,APOE,ITGB1,ITGB3,PLXNB1 |
| M1/Control | Up | GO: Biological Proces | GO:19011 | carbohydr | | 4.33E-05 | 1.38E-01 | 1.79E-03 | 1.55E-02 | 15 | 1256 | CFH,GLYAT,ATP5PF,BMP2,ATP5IF1,NFE2L1,SULF1,PARK7,DAD1,AKR1A1,ITM2B,GNS,ATP5F1A,NDUFS5,ITIH5 |
| M1/Control | Up | GO: Biological Proces | GO:01100 | cellular de | | 4.41E-05 | 1.41E-01 | 1.80E-03 | 1.56E-02 | 3 | 19 | ESD,PARK7,AKR1A1 |
| M1/Control | Up | GO: Biological Proces | GO:00605 | muscle tis | | 4.52E-05 | 1.44E-01 | 1.82E-03 | 1.58E-02 | 11 | 697 | TPM1,BMP2,MYH10,MYL6,TCF21,DSP,EFEMP2,LRP2,DAG1,ACTN1,ITGB1 |
| M1/Control | Up | GO: Cellular Compon | GO:00975 | contractile | | 2.26E-05 | 8.76E-03 | 4.15E-04 | 2.71E-03 | 5 | 92 | TPM1,MYH10,MYL9,ACTN1,MYL12B |
| M1/Control | Up | GO: Cellular Compon | GO:00017 | stress fibe | | 2.26E-05 | 8.76E-03 | 4.15E-04 | 2.71E-03 | 5 | 92 | TPM1,MYH10,MYL9,ACTN1,MYL12B |
| M1/Control | Up | GO: Cellular Compon | GO:00310 | platelet al | | 2.38E-05 | 9.23E-03 | 4.15E-04 | 2.71E-03 | 5 | 93 | THBS1,CD9,APLP2,ACTN1,ITGB3 |
| M1/Control | Up | GO: Cellular Compon | GO:00059 | focal adhe | | 2.81E-05 | 1.09E-02 | 4.53E-04 | 2.96E-03 | 9 | 430 | THY1,CD9,DAG1,CD151,ACTN1,ITGA3,ITGAV,ITGB1,ITGB3 |
| M1/Control | Up | GO: Biological Proces | GO:00106 | negative r | | 4.65E-05 | 1.48E-01 | 1.83E-03 | 1.58E-02 | 18 | 1747 | BMP2,THBS1,THY1,TCF21,SLC25A6,SULF1,PARK7,FRZB,LRP2,DAG1,CGNL1,ITGA3,APOE,ITGAV,NDRG2,ITGB1,PTTG1IP,VDAC2 |
| M1/Control | Up | GO: Biological Proces | GO:00435 | endothelia | | 4.60E-05 | 1.47E-01 | 1.83E-03 | 1.58E-02 | 8 | 354 | THBS1,THSD7A,PTPRM,APOE,ITGAV,ITGB1,ITGB3,ATP5F1A |
| M1/Control | Up | GO: Biological Proces | GO:00230 | negative r | | 4.79E-05 | 1.53E-01 | 1.86E-03 | 1.61E-02 | 18 | 1751 | BMP2,THBS1,THY1,TCF21,SLC25A6,SULF1,PARK7,FRZB,LRP2,DAG1,CGNL1,ITGA3,APOE,ITGAV,NDRG2,ITGB1,PTTG1IP,VDAC2 |
| M1/Control | Up | GO: Biological Proces | GO:00988 | cell-cell ac | | 4.87E-05 | 1.55E-01 | 1.87E-03 | 1.62E-02 | 14 | 1118 | CFH,BMP2,THBS1,THY1,MYL9,CD9,PTPRM,DSP,ACTN1,ITGA3,ITGAV,ITGB1,ITGB3,TNFRSF21 |
| M1/Control | Up | GO: Biological Proces | GO:00604 | muscle tis | | 5.04E-05 | 1.61E-01 | 1.91E-03 | 1.66E-02 | 5 | 109 | TPM1,BMP2,DSP,EFEMP2,LRP2 |
| M1/Control | Up | GO: Biological Proces | GO:00016 | ameboidal | | 5.27E-05 | 1.68E-01 | 1.98E-03 | 1.71E-02 | 11 | 709 | BMP2,THBS1,THSD7A,PTPRM,MACF1,ITGA3,APOE,ITGAV,ITGB1,ITGB3,ATP5F1A |
| M1/Control | Up | GO: Biological Proces | GO:00329 | cell part m | | 5.40E-05 | 1.72E-01 | 2.00E-03 | 1.73E-02 | 13 | 983 | S100A6,MYH10,THY1,THSD7A,PTPRM,LRP2,DAG1,MACF1,APLP2,APOE,ITGB1,ITGB3,PLXNB1 |
| M1/Control | Up | GO: Biological Proces | GO:00987 | detoxificat | | 5.73E-05 | 1.83E-01 | 2.10E-03 | 1.81E-02 | 6 | 183 | ESD,PARK7,SELENOF,PRDX1,AKR1A1,APOE |
| M1/Control | Up | GO: Biological Proces | GO:00974 | supramole | | 5.93E-05 | 1.89E-01 | 2.15E-03 | 1.86E-02 | 13 | 992 | TPM1,FMNL3,MYH10,MYL9,DSP,PARK7,EFEMP2,GAS2,ACTN1,CGNL1,APOE,ITGB1,CDC42EP3 |
| M1/Control | Up | GO: Biological Proces | GO:00461 | aldehyde | | 6.01E-05 | 1.92E-01 | 2.16E-03 | 1.86E-02 | 3 | 21 | ESD,PARK7,AKR1A1 |
| M1/Control | Up | GO: Biological Proces | GO:00511 | positive re | | 6.58E-05 | 2.10E-01 | 2.28E-03 | 1.97E-02 | 15 | 1303 | TPM1,BMP2,ATP5IF1,THY1,PARK7,EFEMP2,LRP2,DAG1,MACF1,CD151,ITGA3,APOE,ITGB1,CDC42EP3,PLXNB1 |
| M1/Control | Up | GO: Biological Proces | GO:00400 | negative r | | 6.52E-05 | 2.08E-01 | 2.28E-03 | 1.97E-02 | 9 | 481 | TPM1,THBS1,THY1,PTPRM,BST2,SULF1,PARK7,DAG1,APOE |
| M1/Control | Up | GO: Biological Proces | GO:00070 | plasma me | | 6.46E-05 | 2.06E-01 | 2.28E-03 | 1.97E-02 | 6 | 187 | MYOF,MYH10,NDRG1,CD9,DAG1,ITGA3 |
| M1/Control | Up | GO: Biological Proces | GO:00096 | response t | | 7.15E-05 | 2.28E-01 | 2.42E-03 | 2.09E-02 | 8 | 377 | GLYAT,SLC22A8,ESD,PARK7,SELENOF,PRDX1,AKR1A1,APOE |
| M1/Control | Up | GO: Biological Proces | GO:00075 | blood coa | | 7.20E-05 | 2.30E-01 | 2.42E-03 | 2.09E-02 | 7 | 278 | CFH,THBS1,MYL9,CD9,ACTN1,APOE,ITGB3 |
| M1/Control | Up | GO: Biological Proces | GO:00486 | muscle or | | 7.07E-05 | 2.25E-01 | 2.42E-03 | 2.09E-02 | 5 | 117 | TPM1,BMP2,DSP,EFEMP2,LRP2 |
| M1/Control | Up | GO: Cellular Compon | GO:00310 | extracellul | | 3.63E-05 | 1.41E-02 | 4.55E-04 | 2.97E-03 | 11 | 678 | S100A6,THBS1,IGFBP7,SULF1,EFEMP2,DAG1,CD151,BCAM,APOE,ITGB1,ITIH5 |
| M1/Control | Up | GO: Cellular Compon | GO:00303 | external e | | 3.73E-05 | 1.44E-02 | 4.55E-04 | 2.97E-03 | 11 | 680 | S100A6,THBS1,IGFBP7,SULF1,EFEMP2,DAG1,CD151,BCAM,APOE,ITGB1,ITIH5 |
| M1/M0 | Up | GO: Biological Proces | GO:00604 | epithelium | | 5.05E-07 | 1.08E-03 | 8.96E-05 | 7.38E-04 | 12 | 1979 | COL4A1,FLNA,COL18A1,GJA5,HEG1,CTNNB1,IGFBP7,ZFP36L1,ENG,ITGB1,VCAM1,MSN |
| M1/Control | Up | GO: Molecular Functi | GO:00055 | insulin-like | | 4.52E-06 | 1.96E-03 | 2.08E-04 | 1.38E-03 | 4 | 29 | IGFBP7,LRP2,ITGAV,ITGB3 |
| M1/Control | Up | GO: Molecular Functi | GO:00519 | peroxiredo | | 4.56E-06 | 1.98E-03 | 2.08E-04 | 1.38E-03 | 3 | 9 | PARK7,SELENOF,PRDX1 |
| M1/Control | Up | GO: Biological Proces | GO:00017 | formation | | 7.47E-05 | 2.38E-01 | 2.48E-03 | 2.15E-02 | 6 | 192 | BMP2,MACF1,ITGA3,ITGAV,ITGB1,ITGB3 |
| M1/Control | Up | GO: Biological Proces | GO:00017 | mesoderm | | 7.66E-05 | 2.44E-01 | 2.52E-03 | 2.18E-02 | 5 | 119 | BMP2,MACF1,ITGA3,ITGB1,ITGB3 |
| M1/Control | Up | GO: Biological Proces | GO:00486 | cell morph | | 8.28E-05 | 2.64E-01 | 2.67E-03 | 2.31E-02 | 12 | 882 | S100A6,MYH10,THY1,THSD7A,PTPRM,LRP2,DAG1,MACF1,APLP2,APOE,ITGB1,PLXNB1 |
| M1/Control | Up | GO: Biological Proces | GO:00075 | hemostasi | | 8.23E-05 | 2.63E-01 | 2.67E-03 | 2.31E-02 | 7 | 284 | CFH,THBS1,MYL9,CD9,ACTN1,APOE,ITGB3 |
| M1/Control | Up | GO: Biological Proces | GO:00508 | coagulatio | | 8.41E-05 | 2.68E-01 | 2.68E-03 | 2.32E-02 | 7 | 285 | CFH,THBS1,MYL9,CD9,ACTN1,APOE,ITGB3 |
| M1/Control | Up | GO: Biological Proces | GO:00483 | mesoderm | | 8.99E-05 | 2.87E-01 | 2.84E-03 | 2.45E-02 | 4 | 64 | BMP2,ITGA3,ITGB1,ITGB3 |
| M1/Control | Up | GO: Biological Proces | GO:00713 | cellular re | | 9.08E-05 | 2.90E-01 | 2.84E-03 | 2.45E-02 | 3 | 24 | NFE2L1,DAG1,APOE |
| M1/Control | Up | GO: Biological Proces | GO:00301 | extracellul | | 9.72E-05 | 3.10E-01 | 2.98E-03 | 2.58E-02 | 8 | 394 | BMP2,SULF1,EFEMP2,GAS2,DAG1,APLP2,ITGB1,ITGB3 |
| M1/Control | Up | GO: Biological Proces | GO:00328 | glomerulu | | 9.67E-05 | 3.09E-01 | 2.98E-03 | 2.58E-02 | 5 | 125 | CFH,THSD7A,TCF21,SULF1,ITGB3 |
| M1/Control | Up | GO: Biological Proces | GO:00430 | extracellul | | 9.89E-05 | 3.15E-01 | 3.00E-03 | 2.60E-02 | 8 | 395 | BMP2,SULF1,EFEMP2,GAS2,DAG1,APLP2,ITGB1,ITGB3 |
| M1/Control | Up | GO: Biological Proces | GO:00452 | external e | | 1.04E-04 | 3.32E-01 | 3.14E-03 | 2.71E-02 | 8 | 398 | BMP2,SULF1,EFEMP2,GAS2,DAG1,APLP2,ITGB1,ITGB3 |
| M1/Control | Up | GO: Biological Proces | GO:00302 | myofibril a | | 1.08E-04 | 3.45E-01 | 3.22E-03 | 2.79E-02 | 5 | 128 | TPM1,MYH10,MYL9,ACTN1,ITGB1 |
| M1/Control | Up | GO: Biological Proces | GO:00400 | growth | | 1.11E-04 | 3.54E-01 | 3.28E-03 | 2.84E-02 | 16 | 1528 | THBS1,MYH10,KLK6,THSD7A,CD9,IGFBP7,BST2,FRZB,GAS2,DAG1,MACF1,APOE,ITGAV,ITGB1,PLXNB1,VDAC2 |
| M1/Control | Up | GO: Biological Proces | GO:00483 | mesoderm | | 1.12E-04 | 3.58E-01 | 3.28E-03 | 2.84E-02 | 5 | 129 | BMP2,MACF1,ITGA3,ITGB1,ITGB3 |
| M1/Control | Up | GO: Biological Proces | GO:00343 | cell junctic | | 1.21E-04 | 3.87E-01 | 3.52E-03 | 3.04E-02 | 9 | 522 | THBS1,THY1,CD9,MACF1,ACTN1,ITGAV,ITGB1,ITGB3,PLXNB1 |
| M1/Control | Up | GO: Biological Proces | GO:00107 | positive re | | 1.30E-04 | 4.15E-01 | 3.74E-03 | 3.23E-02 | 3 | 27 | THBS1,ITGB1,ITGB3 |
| M1/Control | Up | GO: Biological Proces | GO:00507 | negative r | | 1.32E-04 | 4.20E-01 | 3.75E-03 | 3.25E-02 | 2 | 5 | ITGAV,ITGB3 |
| M1/Control | Up | GO: Biological Proces | GO:00488 | neuron pr | | 1.39E-04 | 4.44E-01 | 3.93E-03 | 3.40E-02 | 12 | 932 | S100A6,MYH10,THY1,THSD7A,PTPRM,LRP2,DAG1,MACF1,APLP2,APOE,ITGB1,PLXNB1 |
| M1/Control | Up | GO: Biological Proces | GO:00149 | myotube | | 1.43E-04 | 4.55E-01 | 3.95E-03 | 3.42E-02 | 6 | 216 | SORT1,MYOF,CD9,DAG1,ACTN1,ITGB1 |
| M1/Control | Up | GO: Biological Proces | GO:00019 | positive re | | 1.42E-04 | 4.54E-01 | 3.95E-03 | 3.42E-02 | 4 | 72 | THY1,EFEMP2,DAG1,ITGB3 |
| M1/Control | Up | GO: Biological Proces | GO:19030 | regulation | | 1.50E-04 | 4.79E-01 | 4.13E-03 | 3.57E-02 | 4 | 73 | BMP2,EFEMP2,DAG1,ITGB3 |
| M1/Control | Up | GO: Biological Proces | GO:00311 | neuron pr | | 1.55E-04 | 4.95E-01 | 4.18E-03 | 3.61E-02 | 15 | 1407 | S100A6,MYH10,KLK6,THY1,THSD7A,PTPRM,LRP2,DAG1,MACF1,APLP2,ITGA3,APOE,ITGB1,TNFRSF21,PLXNB1 |
| M1/Control | Up | GO: Cellular Compon | GO:00300 | cell-substr | | 3.49E-05 | 1.35E-02 | 4.55E-04 | 2.97E-03 | 9 | 442 | THY1,CD9,DAG1,CD151,ACTN1,ITGA3,ITGAV,ITGB1,ITGB3 |
| M1/Control | Up | GO: Cellular Compon | GO:00306 | secretory | | 3.06E-05 | 1.18E-02 | 4.55E-04 | 2.97E-03 | 8 | 333 | CD9,DSP,BST2,APLP2,LAMP2,ACTN1,ITGAV,ITGB3 |
| M1/Control | Up | GO: Cellular Compon | GO:00324 | actin filam | | 3.72E-05 | 1.44E-02 | 4.55E-04 | 2.97E-03 | 5 | 102 | TPM1,MYH10,MYL9,ACTN1,MYL12B |
| M1/Control | Up | GO: Cellular Compon | GO:00426 | actomyosi | | 3.90E-05 | 1.51E-02 | 4.55E-04 | 2.97E-03 | 5 | 103 | TPM1,MYH10,MYL9,ACTN1,MYL12B |
| M1/Control | Up | GO: Cellular Compon | GO:00310 | platelet al | | 3.14E-05 | 1.21E-02 | 4.55E-04 | 2.97E-03 | 3 | 17 | CD9,APLP2,ITGB3 |
| M1/Control | Up | GO: Cellular Compon | GO:00358 | alphav-bet | | 4.00E-05 | 1.55E-02 | 4.55E-04 | 2.97E-03 | 2 | 3 | ITGAV,ITGB3 |
| M1/Control | Up | GO: Cellular Compon | GO:00358 | alphav-bet | | 4.00E-05 | 1.55E-02 | 4.55E-04 | 2.97E-03 | 2 | 3 | ITGAV,ITGB3 |
| M1/M0 | Up | GO: Molecular Functi | GO:00520 | extracellul | | 2.29E-09 | 4.79E-07 | 4.79E-07 | 2.84E-06 | 7 | 195 | COL4A1,HSPG2,COL18A1,IGFBP7,ECM1,ENG,BGN |
| M1/M0 | Up | GO: Molecular Functi | GO:00051 | structural | | 4.86E-07 | 1.02E-04 | 5.08E-05 | 3.01E-04 | 9 | 892 | COL4A1,HSPG2,COL18A1,IGFBP7,ECM1,ENG,H4C5,BGN,MSN |
| M1/M0 | Up | GO: Biological Proces | GO:00015 | branching | | 6.04E-07 | 1.29E-03 | 9.90E-05 | 8.16E-04 | 4 | 59 | COL4A1,CTNNB1,IGFBP7,ENG |
| M1/M0 | Up | GO: Biological Proces | GO:00018 | endothelia | | 1.31E-06 | 2.79E-03 | 1.99E-04 | 1.64E-03 | 3 | 19 | COL18A1,HEG1,ENG |
| M1/M0 | Up | GO: Biological Proces | GO:00308 | epithelial | | 1.41E-06 | 3.00E-03 | 2.00E-04 | 1.65E-03 | 9 | 1051 | COL4A1,FLNA,COL18A1,HEG1,CTNNB1,ZFP36L1,ENG,ITGB1,MSN |
| M1/M0 | Up | GO: Biological Proces | GO:00750 | heart deve | | 1.93E-06 | 4.12E-03 | 2.58E-04 | 2.12E-03 | 9 | 1092 | HSPG2,FLNA,GJA5,HEG1,CTNNB1,ZFP36L1,ENG,ITGB1,VCAM1 |
| M1/M0 | Up | GO: Biological Proces | GO:00988 | cell-cell ac | | 2.35E-06 | 5.01E-03 | 2.90E-04 | 2.39E-03 | 9 | 1118 | HSPG2,FLNA,CD81,CTNNB1,ZFP36L1,ITGB1,VCAM1,LRRC32,MSN |
| M1/M0 | Up | GO: Biological Proces | GO:00454 | endothelia | | 2.45E-06 | 5.22E-03 | 2.90E-04 | 2.39E-03 | 5 | 193 | COL18A1,HEG1,CTNNB1,ENG,MSN |
| M1/M0 | Up | GO: Biological Proces | GO:00020 | epithelial | | 2.73E-06 | 5.81E-03 | 3.06E-04 | 2.52E-03 | 6 | 360 | FLNA,COL18A1,HEG1,ENG,ITGB1,MSN |
| M1/M0 | Up | GO: Biological Proces | GO:00487 | cardiac m | | 3.56E-06 | 7.58E-03 | 3.79E-04 | 3.12E-03 | 6 | 377 | HSPG2,GJA5,HEG1,CTNNB1,ENG,ITGB1 |
| M1/M0 | Up | GO: Biological Proces | GO:00147 | striated m | | 4.93E-06 | 1.05E-02 | 4.77E-04 | 3.93E-03 | 6 | 399 | HSPG2,GJA5,HEG1,CTNNB1,ENG,ITGB1 |
| M1/M0 | Up | GO: Biological Proces | GO:00018 | endothelia | | 4.84E-06 | 1.03E-02 | 4.77E-04 | 3.93E-03 | 4 | 99 | COL18A1,HEG1,ENG,MSN |
| M1/M0 | Up | GO: Biological Proces | GO:00226 | regulation | | 7.33E-06 | 1.56E-02 | 6.79E-04 | 5.59E-03 | 9 | 1284 | HSPG2,FLNA,TCF4,CTNNB1,ECM1,ENG,ITGB1,SGK1,MSN |
| M1/M0 | Up | GO: Cellular Compon | GO:00310 | extracellul | | 3.15E-08 | 7.27E-06 | 3.73E-06 | 2.25E-05 | 9 | 678 | COL4A1,S100A6,HSPG2,COL18A1,IGFBP7,ECM1,ITGB1,LRRC32,BGN |
| M1/M0 | Up | GO: Cellular Compon | GO:00303 | external e | | 3.23E-08 | 7.46E-06 | 3.73E-06 | 2.25E-05 | 9 | 680 | COL4A1,S100A6,HSPG2,COL18A1,IGFBP7,ECM1,ITGB1,LRRC32,BGN |
| M1/M0 | Up | GO: Cellular Compon | GO:00701 | anchoring | | 1.52E-05 | 3.50E-03 | 3.50E-04 | 2.11E-03 | 9 | 1419 | HSPG2,FLNA,GJA5,CD81,HEG1,CTNNB1,ENG,ITGB1,MSN |
| M1/Control | Up | GO: Biological Proces | GO:00606 | alcohol m | | 1.56E-04 | 4.97E-01 | 4.18E-03 | 3.61E-02 | 8 | 422 | BMP2,MYOF,NFE2L1,PARK7,LRP2,APLP2,AKR1A1,APOE |
| M1/Control | Up | GO: Biological Proces | GO:00550 | striated m | | 1.54E-04 | 4.91E-01 | 4.18E-03 | 3.61E-02 | 5 | 138 | TPM1,MYH10,MYL9,ACTN1,ITGB1 |
| M1/Control | Up | GO: Biological Proces | GO:00722 | integrin-m | | 1.59E-04 | 5.08E-01 | 4.24E-03 | 3.66E-02 | 5 | 139 | THY1,ITGA3,ITGAV,ITGB1,ITGB3 |
| M1/Control | Up | GO: Biological Proces | GO:00363 | cellular re | | 1.62E-04 | 5.16E-01 | 4.26E-03 | 3.69E-02 | 3 | 29 | NFE2L1,DAG1,APOE |
| M1/Control | Up | GO: Biological Proces | GO:01200 | plasma me | | 1.66E-04 | 5.31E-01 | 4.35E-03 | 3.76E-02 | 12 | 950 | S100A6,MYH10,THY1,THSD7A,PTPRM,LRP2,DAG1,MACF1,APLP2,APOE,ITGB1,PLXNB1 |
| M1/Control | Up | GO: Biological Proces | GO:00506 | negative r | | 1.87E-04 | 5.95E-01 | 4.84E-03 | 4.18E-02 | 6 | 227 | THBS1,ATP5IF1,PTPRM,SULF1,APOE,ATP5F1A |
| M1/Control | Up | GO: Biological Proces | GO:00107 | positive re | | 2.07E-04 | 6.60E-01 | 5.32E-03 | 4.60E-02 | 8 | 440 | BMP2,LRP2,DAG1,MACF1,APOE,ITGB1,ITGB3,PLXNB1 |
| M1/Control | Up | GO: Biological Proces | GO:00513 | negative r | | 2.14E-04 | 6.81E-01 | 5.45E-03 | 4.71E-02 | 8 | 442 | SORT1,THBS1,ATP5IF1,BST2,PARK7,LRP2,APLP2,ITIH5 |
| M1/Control | Up | GO: Biological Proces | GO:00991 | postsynap | | 2.25E-04 | 7.18E-01 | 5.70E-03 | 4.93E-02 | 6 | 235 | MYH10,NDRG1,ACTN1,ITGA3,APOE,NDRG2 |
| M1/Control | Up | GO: Biological Proces | GO:00608 | artery dev | | 2.27E-04 | 7.25E-01 | 5.71E-03 | 4.93E-02 | 5 | 150 | MYH10,TCF21,EFEMP2,LRP2,APOE |
| M1/Control | Up | GO: Biological Proces | GO:00017 | cell activat | | 2.40E-04 | 7.64E-01 | 5.88E-03 | 5.08E-02 | 15 | 1464 | CFH,THBS1,NDRG1,THY1,MYL9,CD9,BST2,PRDX1,CD151,ACTN1,APOE,ITGAV,ITGB1,ITGB3,TNFRSF21 |
| M1/Control | Up | GO: Biological Proces | GO:01100 | cellular re | | 2.39E-04 | 7.62E-01 | 5.88E-03 | 5.08E-02 | 3 | 33 | ESD,PARK7,AKR1A1 |
| M1/Control | Up | GO: Biological Proces | GO:19030 | positive re | | 2.39E-04 | 7.62E-01 | 5.88E-03 | 5.08E-02 | 3 | 33 | BMP2,EFEMP2,DAG1 |
| M1/Control | Up | GO: Biological Proces | GO:00610 | membrane | | 2.79E-04 | 8.91E-01 | 6.68E-03 | 5.77E-02 | 12 | 1005 | MYOF,THBS1,MYH10,ATP5IF1,NDRG1,CD9,SLC25A6,DAG1,ITGA3,APOE,ITGB1,VDAC2 |
| M1/Control | Up | GO: Biological Proces | GO:00602 | regulation | | 2.81E-04 | 8.96E-01 | 6.68E-03 | 5.77E-02 | 10 | 717 | BMP2,THY1,FRZB,LRP2,DAG1,MACF1,APOE,ITGB1,ITGB3,PLXNB1 |
| M1/Control | Up | GO: Biological Proces | GO:00507 | positive re | | 2.82E-04 | 9.00E-01 | 6.68E-03 | 5.77E-02 | 7 | 347 | BMP2,LRP2,DAG1,MACF1,APOE,ITGB1,PLXNB1 |
| M1/Control | Up | GO: Biological Proces | GO:00608 | cellular alc | | 2.83E-04 | 9.01E-01 | 6.68E-03 | 5.77E-02 | 4 | 86 | BMP2,ESD,PARK7,AKR1A1 |
| M1/Control | Up | GO: Biological Proces | GO:00380 | apolipopro | | 2.76E-04 | 8.79E-01 | 6.68E-03 | 5.77E-02 | 2 | 7 | ITGAV,ITGB3 |
| M1/Control | Up | GO: Biological Proces | GO:00457 | positive re | | 2.95E-04 | 9.39E-01 | 6.91E-03 | 5.97E-02 | 9 | 588 | TPM1,THBS1,THY1,EFEMP2,DAG1,ITGA3,ITGAV,ITGB1,ITGB3 |
| M1/Control | Up | GO: Biological Proces | GO:00485 | developm | | 3.01E-04 | 9.58E-01 | 7.00E-03 | 6.05E-02 | 13 | 1168 | THBS1,MYH10,KLK6,THSD7A,CD9,GAS2,DAG1,MACF1,APOE,ITGAV,ITGB1,PLXNB1,VDAC2 |
| M1/Control | Up | GO: Biological Proces | GO:00486 | neuron de | | 3.08E-04 | 9.82E-01 | 7.11E-03 | 6.15E-02 | 16 | 1670 | CFH,S100A6,MYH10,KLK6,THY1,THSD7A,PTPRM,LRP2,DAG1,MACF1,APLP2,ITGA3,APOE,ITGB1,TNFRSF21,PLXNB1 |
| M1/Control | Up | GO: Biological Proces | GO:00717 | response t | | 3.10E-04 | 9.90E-01 | 7.12E-03 | 6.16E-02 | 3 | 36 | THBS1,ITGAV,ATP5F1A |
| M1/Control | Up | GO: Biological Proces | GO:00508 | synapse o | | 3.13E-04 | 9.99E-01 | 7.14E-03 | 6.17E-02 | 9 | 593 | MYH10,NDRG1,DAG1,ACTN1,ITGA3,APOE,NDRG2,ITGB1,PLXNB1 |
| M1/Control | Up | GO: Biological Proces | GO:00550 | cardiac m | | 3.22E-04 | 1.00E+00 | 7.29E-03 | 6.30E-02 | 4 | 89 | TPM1,BMP2,DSP,LRP2 |
| M1/Control | Up | GO: Biological Proces | GO:00106 | regulation | | 3.30E-04 | 1.00E+00 | 7.40E-03 | 6.40E-02 | 7 | 356 | THBS1,PTPRM,MACF1,ITGA3,APOE,ITGB3,ATP5F1A |
| M1/Control | Up | GO: Cellular Compon | GO:00346 | integrin al | | 4.00E-05 | 1.55E-02 | 4.55E-04 | 2.97E-03 | 2 | 3 | ITGAV,ITGB3 |
| M1/Control | Up | GO: Cellular Compon | GO:00156 | actin cytos | | 4.29E-05 | 1.66E-02 | 4.74E-04 | 3.10E-03 | 10 | 568 | TPM1,MYH10,MYL6,MYL9,GAS2,MACF1,ACTN1,CGNL1,MYL12B,CDC42EP3 |
| M1/Control | Up | GO: Cellular Compon | GO:00325 | ruffle mem | | 4.48E-05 | 1.73E-02 | 4.82E-04 | 3.15E-03 | 5 | 106 | TPM1,MACF1,ITGAV,ITGB1,ITGB3 |
| M1/Control | Up | GO: Cellular Compon | GO:00986 | protein co | | 4.66E-05 | 1.80E-02 | 4.87E-04 | 3.18E-03 | 4 | 54 | ITGA3,ITGAV,ITGB1,ITGB3 |
| M1/Control | Up | GO: Cellular Compon | GO:00315 | filopodium | | 6.07E-05 | 2.35E-02 | 6.18E-04 | 4.04E-03 | 3 | 21 | ITGA3,ITGAV,ITGB3 |
| M1/M0 | Up | GO: Biological Proces | GO:00713 | cellular re | | 7.99E-06 | 1.70E-02 | 7.10E-04 | 5.85E-03 | 8 | 963 | TCF4,GJA5,CTNNB1,ZFP36L1,ENG,ITGB1,VCAM1,LRRC32 |
| M1/M0 | Up | GO: Biological Proces | GO:00605 | muscle tis | | 9.93E-06 | 2.12E-02 | 8.14E-04 | 6.71E-03 | 7 | 697 | HSPG2,GJA5,HEG1,CTNNB1,RCAN1,ENG,ITGB1 |
| M1/M0 | Up | GO: Biological Proces | GO:00452 | cell-cell ju | | 9.71E-06 | 2.07E-02 | 8.14E-04 | 6.71E-03 | 5 | 256 | FLNA,GJA5,HEG1,CTNNB1,ITGB1 |
| M1/M0 | Up | GO: Biological Proces | GO:00708 | response t | | 1.10E-05 | 2.35E-02 | 8.69E-04 | 7.16E-03 | 8 | 1006 | TCF4,GJA5,CTNNB1,ZFP36L1,ENG,ITGB1,VCAM1,LRRC32 |
| M1/M0 | Up | GO: Biological Proces | GO:00457 | regulation | | 1.17E-05 | 2.49E-02 | 8.88E-04 | 7.32E-03 | 6 | 464 | HSPG2,TCF4,CTNNB1,ECM1,ENG,ITGB1 |
| M1/M0 | Up | GO: Biological Proces | GO:19013 | regulation | | 1.35E-05 | 2.87E-02 | 8.98E-04 | 7.40E-03 | 6 | 476 | HSPG2,TCF4,CTNNB1,ECM1,ENG,ITGB1 |
| M1/M0 | Up | GO: Biological Proces | GO:00032 | cardiac atr | | 1.31E-05 | 2.80E-02 | 8.98E-04 | 7.40E-03 | 3 | 40 | GJA5,HEG1,ENG |
| M1/M0 | Up | GO: Biological Proces | GO:00226 | membrane | | 1.28E-05 | 2.73E-02 | 8.98E-04 | 7.40E-03 | 2 | 5 | VCAM1,MSN |
| M1/M0 | Up | GO: Biological Proces | GO:00219 | formation | | 1.28E-05 | 2.73E-02 | 8.98E-04 | 7.40E-03 | 2 | 5 | FLNA,ITGB1 |
| M1/M0 | Up | GO: Biological Proces | GO:00610 | muscle str | | 1.44E-05 | 3.08E-02 | 9.32E-04 | 7.68E-03 | 8 | 1044 | HSPG2,CD81,HEG1,CTNNB1,RCAN1,ZFP36L1,ENG,ITGB1 |
| M1/M0 | Up | GO: Biological Proces | GO:00009 | cell morph | | 1.61E-05 | 3.43E-02 | 9.90E-04 | 8.16E-03 | 9 | 1415 | S100A6,FLNA,COL18A1,HEG1,CTNNB1,ENG,ITGB1,SGK1,MSN |
| M1/M0 | Up | GO: Biological Proces | GO:00509 | leukocyte | | 1.63E-05 | 3.47E-02 | 9.90E-04 | 8.16E-03 | 6 | 492 | CD81,ECM1,ENG,ITGB1,VCAM1,MSN |
| M1/M0 | Up | GO: Biological Proces | GO:00400 | locomotio | | 1.77E-05 | 3.77E-02 | 1.05E-03 | 8.64E-03 | 10 | 1834 | FLNA,COL18A1,CD81,RCAN1,ECM1,ENG,ITGB1,SGK1,VCAM1,MSN |
| M1/M0 | Up | GO: Biological Proces | GO:00009 | cell morph | | 1.86E-05 | 3.96E-02 | 1.07E-03 | 8.82E-03 | 8 | 1081 | S100A6,FLNA,COL18A1,HEG1,CTNNB1,ENG,ITGB1,SGK1 |
| M1/M0 | Up | GO: Biological Proces | GO:00017 | cell activat | | 2.12E-05 | 4.51E-02 | 1.16E-03 | 9.53E-03 | 9 | 1464 | FLNA,TCF4,CD81,CTNNB1,ZFP36L1,ITGB1,VCAM1,LRRC32,MSN |
| M1/M0 | Up | GO: Biological Proces | GO:00420 | T cell proli | | 2.09E-05 | 4.45E-02 | 1.16E-03 | 9.53E-03 | 5 | 300 | CD81,CTNNB1,VCAM1,LRRC32,MSN |
| M1/M0 | Up | GO: Biological Proces | GO:00032 | cardiac atr | | 2.43E-05 | 5.18E-02 | 1.29E-03 | 1.07E-02 | 3 | 49 | GJA5,HEG1,ENG |
| M1/M0 | Up | GO: Biological Proces | GO:00487 | tissue mor | | 2.60E-05 | 5.53E-02 | 1.35E-03 | 1.11E-02 | 8 | 1132 | COL4A1,FLNA,HEG1,CTNNB1,IGFBP7,ENG,ITGB1,MSN |
| M1/M0 | Up | GO: Biological Proces | GO:00220 | central ne | | 2.69E-05 | 5.73E-02 | 1.37E-03 | 1.13E-02 | 2 | 7 | CTNNB1,ENG |
| M1/M0 | Up | GO: Cellular Compon | GO:00620 | collagen-c | | 9.66E-08 | 2.23E-05 | 7.44E-06 | 4.48E-05 | 8 | 541 | COL4A1,S100A6,HSPG2,COL18A1,IGFBP7,ECM1,ITGB1,BGN |
| M1/Control | Up | GO: Molecular Functi | GO:00319 | insulin-like | | 1.54E-05 | 6.66E-03 | 6.06E-04 | 4.03E-03 | 3 | 13 | LRP2,ITGAV,ITGB3 |
| M1/Control | Up | GO: Molecular Functi | GO:00083 | structural | | 2.47E-05 | 1.07E-02 | 8.93E-04 | 5.94E-03 | 4 | 44 | TPM1,MYL6,MYL9,DAG1 |
| M1/Control | Up | GO: Biological Proces | GO:00485 | embryonic | | 3.45E-04 | 1.00E+00 | 7.70E-03 | 6.66E-02 | 13 | 1185 | CNBP,BMP2,MYH10,TCF21,SULF1,FRZB,LRP2,DAG1,MACF1,ITGA3,ITGAV,ITGB1,ITGB3 |
| M1/Control | Up | GO: Biological Proces | GO:00336 | cell adhes | | 3.51E-04 | 1.00E+00 | 7.77E-03 | 6.72E-02 | 4 | 91 | ITGA3,ITGAV,ITGB1,ITGB3 |
| M1/Control | Up | GO: Biological Proces | GO:00988 | neurotran | | 3.94E-04 | 1.00E+00 | 8.67E-03 | 7.49E-02 | 3 | 39 | PARK7,ITGB1,ITGB3 |
| M1/Control | Up | GO: Biological Proces | GO:20000 | regulation | | 4.52E-04 | 1.00E+00 | 9.84E-03 | 8.51E-02 | 17 | 1908 | BMP2,THBS1,NDRG1,THY1,PTPRM,NFE2L1,SULF1,H4C5,FRZB,LRP2,DAG1,MACF1,APOE,ITGB1,ITGB3,TNFRSF21,PLXNB1 |
| M1/Control | Up | GO: Biological Proces | GO:00098 | animal org | | 4.54E-04 | 1.00E+00 | 9.84E-03 | 8.51E-02 | 16 | 1729 | TPM1,CFH,CNBP,BMP2,THBS1,THY1,TCF21,PTPRM,DSP,SULF1,CDIPT,FRZB,EFEMP2,LRP2,DAG1,ITGAV |
| M1/Control | Up | GO: Biological Proces | GO:00550 | response t | | 4.57E-04 | 1.00E+00 | 9.85E-03 | 8.52E-02 | 3 | 41 | CD9,APOE,ITGB1 |
| M1/Control | Up | GO: Biological Proces | GO:00506 | epithelial | | 4.70E-04 | 1.00E+00 | 9.93E-03 | 8.58E-02 | 9 | 627 | BMP2,THBS1,ATP5IF1,PTPRM,SULF1,APOE,ITGAV,ITGB3,ATP5F1A |
| M1/Control | Up | GO: Biological Proces | GO:19055 | negative r | | 4.70E-04 | 1.00E+00 | 9.93E-03 | 8.58E-02 | 2 | 9 | ITGAV,ITGB3 |
| M1/Control | Up | GO: Biological Proces | GO:00192 | lactate bio | | 4.70E-04 | 1.00E+00 | 9.93E-03 | 8.58E-02 | 2 | 9 | PARK7,LDHB |
| M1/Control | Up | GO: Biological Proces | GO:00973 | mitochonc | | 5.27E-04 | 1.00E+00 | 1.11E-02 | 9.55E-02 | 3 | 43 | ATP5IF1,SLC25A6,VDAC2 |
| M1/Control | Up | GO: Biological Proces | GO:00430 | negative r | | 5.51E-04 | 1.00E+00 | 1.15E-02 | 9.93E-02 | 11 | 929 | SORT1,BMP2,THBS1,ATP5IF1,THY1,BST2,PARK7,LRP2,APLP2,APOE,ITIH5 |
| M1/Control | Up | GO: Biological Proces | GO:00707 | response t | | 5.64E-04 | 1.00E+00 | 1.17E-02 | 1.01E-01 | 3 | 44 | NFE2L1,DAG1,APOE |
| M1/Control | Up | GO: Biological Proces | GO:00094 | response t | | 5.73E-04 | 1.00E+00 | 1.18E-02 | 1.02E-01 | 10 | 785 | SORT1,GLYAT,THBS1,PTPRM,PARK7,DAD1,LRP2,ITGA3,ITGB1,ITGB3 |
| M1/Control | Up | GO: Biological Proces | GO:00519 | regulation | | 5.76E-04 | 1.00E+00 | 1.18E-02 | 1.02E-01 | 9 | 645 | BMP2,THY1,LRP2,DAG1,MACF1,APOE,ITGB1,TNFRSF21,PLXNB1 |
| M1/Control | Up | GO: Biological Proces | GO:19016 | organic hy | | 5.89E-04 | 1.00E+00 | 1.18E-02 | 1.02E-01 | 9 | 647 | BMP2,MYOF,NFE2L1,PARK7,LRP2,APLP2,AKR1A1,APOE,LDHB |
| M1/Control | Up | GO: Biological Proces | GO:00704 | monocyte | | 5.86E-04 | 1.00E+00 | 1.18E-02 | 1.02E-01 | 2 | 10 | CFH,THBS1 |
| M1/Control | Up | GO: Biological Proces | GO:00032 | ventricular | | 5.86E-04 | 1.00E+00 | 1.18E-02 | 1.02E-01 | 2 | 10 | DSP,LRP2 |
| M1/Control | Up | GO: Biological Proces | GO:00301 | regulation | | 5.93E-04 | 1.00E+00 | 1.18E-02 | 1.02E-01 | 6 | 282 | LRP2,CD151,APOE,ITGAV,ITGB1,ITGB3 |
| M1/Control | Up | GO: Biological Proces | GO:00070 | actin filam | | 6.06E-04 | 1.00E+00 | 1.20E-02 | 1.04E-01 | 8 | 517 | TPM1,FMNL3,MYH10,GAS2,ACTN1,CGNL1,ITGB1,CDC42EP3 |
| M1/Control | Up | GO: Cellular Compon | GO:00358 | alphav-bet | | 7.98E-05 | 3.09E-02 | 7.92E-04 | 5.17E-03 | 2 | 4 | ITGAV,ITGB3 |
| M1/Control | Up | GO: Cellular Compon | GO:00301 | filopodium | | 9.82E-05 | 3.80E-02 | 9.50E-04 | 6.21E-03 | 5 | 125 | DAG1,ITGA3,ITGAV,ITGB1,ITGB3 |
| M1/Control | Up | GO: Cellular Compon | GO:00312 | leading ed | | 1.21E-04 | 4.69E-02 | 1.14E-03 | 7.48E-03 | 6 | 209 | TPM1,THY1,MACF1,ITGAV,ITGB1,ITGB3 |
| M1/Control | Up | GO: Cellular Compon | GO:00995 | secretory | | 1.43E-04 | 5.52E-02 | 1.32E-03 | 8.60E-03 | 14 | 1232 | THBS1,KLK6,CD9,DSP,BST2,APLP2,LAMP2,ACTN1,ITGAV,RAB2A,ITGB1,ITGB3,GNS,VDAC2 |
| M1/Control | Up | GO: Cellular Compon | GO:00312 | cell projec | | 1.76E-04 | 6.79E-02 | 1.58E-03 | 1.03E-02 | 8 | 428 | TPM1,THY1,LRP2,MACF1,ITGA3,ITGAV,ITGB1,ITGB3 |
| M1/Control | Up | GO: Cellular Compon | GO:00057 | endoplasm | | 2.33E-04 | 9.00E-02 | 2.05E-03 | 1.34E-02 | 7 | 335 | THBS1,IGFBP7,ESD,SELENOF,DAG1,APLP2,APOE |
| M1/Control | Up | GO: Cellular Compon | GO:00059 | cell-cell ju | | 3.10E-04 | 1.20E-01 | 2.66E-03 | 1.74E-02 | 9 | 590 | NDRG1,PTPRM,DSP,PARK7,DAG1,ACTN1,CGNL1,ITGB1,ITGB3 |
| M1/Control | Up | GO: Cellular Compon | GO:00995 | supramole | | 3.22E-04 | 1.25E-01 | 2.71E-03 | 1.77E-02 | 13 | 1172 | TPM1,GLYAT,MYH10,NDRG1,MYL9,DSP,EFEMP2,GAS2,DAG1,MACF1,ACTN1,APOE,MYL12B |
| M1/Control | Up | GO: Cellular Compon | GO:00452 | synapse | | 3.50E-04 | 1.36E-01 | 2.77E-03 | 1.81E-02 | 16 | 1683 | MYH10,NDRG1,SLC12A4,PARK7,GLS,DAG1,MACF1,AKR1A1,ACTN1,ITGA3,APOE,RAB2A,NDRG2,ITGB1,ITGB3,VDAC2 |
| M1/Control | Up | GO: Cellular Compon | GO:00990 | supramole | | 3.46E-04 | 1.34E-01 | 2.77E-03 | 1.81E-02 | 13 | 1181 | TPM1,GLYAT,MYH10,NDRG1,MYL9,DSP,EFEMP2,GAS2,DAG1,MACF1,ACTN1,APOE,MYL12B |
| M1/Control | Down | GO: Biological Proces | GO:00339 | response t | | 1.60E-05 | 1.84E-02 | 2.31E-03 | 1.76E-02 | 7 | 1567 | EGR1,FOS,ZFP36,DUSP1,REN,CEBPB,NR4A1 |
| M1/Control | Down | GO: Biological Proces | GO:00429 | regulation | | 5.61E-05 | 6.48E-02 | 4.11E-03 | 3.13E-02 | 7 | 1899 | EGR1,ZFP36,ATF3,DUSP1,CEBPB,NR4A1,USP17L12 |
| M1/Control | Down | GO: Biological Proces | GO:00714 | cellular re | | 5.69E-05 | 6.57E-02 | 4.11E-03 | 3.13E-02 | 7 | 1903 | EGR1,FOS,ZFP36,DUSP1,REN,CEBPB,NR4A1 |
| M1/Control | Down | GO: Biological Proces | GO:00430 | regulation | | 6.53E-05 | 7.55E-02 | 4.44E-03 | 3.39E-02 | 7 | 1944 | EGR1,ZFP36,ATF3,DUSP1,CEBPB,NR4A1,USP17L12 |
| M1/M0 | Up | GO: Molecular Functi | GO:00508 | cell adhes | | 4.74E-06 | 9.91E-04 | 3.30E-04 | 1.96E-03 | 7 | 600 | HSPG2,FLNA,CD81,CTNNB1,ITGB1,VCAM1,MSN |
| M1/M0 | Up | GO: Molecular Functi | GO:00198 | growth fac | | 4.41E-05 | 9.21E-03 | 2.30E-03 | 1.36E-02 | 4 | 167 | COL4A1,IGFBP7,ENG,LRRC32 |
| M1/M0 | Up | GO: Biological Proces | GO:00033 | epithelial | | 2.91E-05 | 6.20E-02 | 1.44E-03 | 1.19E-02 | 3 | 52 | COL18A1,HEG1,ENG |
| M1/M0 | Up | GO: Biological Proces | GO:00604 | bone deve | | 3.34E-05 | 7.12E-02 | 1.58E-03 | 1.31E-02 | 5 | 331 | HSPG2,FLNA,ECM1,ENG,BGN |
| M1/M0 | Up | GO: Biological Proces | GO:00015 | vasculoge | | 3.31E-05 | 7.04E-02 | 1.58E-03 | 1.31E-02 | 4 | 161 | HEG1,CTNNB1,ZFP36L1,ENG |
| M1/M0 | Up | GO: Biological Proces | GO:00075 | muscle or | | 3.51E-05 | 7.48E-02 | 1.63E-03 | 1.34E-02 | 6 | 564 | HSPG2,HEG1,CTNNB1,RCAN1,ENG,ITGB1 |
| M1/M0 | Up | GO: Biological Proces | GO:00339 | response t | | 3.64E-05 | 7.76E-02 | 1.65E-03 | 1.36E-02 | 9 | 1567 | CTNNB1,IGFBP7,ZFP36L1,ENG,ITGB1,SGK1,VCAM1,PGRMC2,MSN |
| M1/M0 | Up | GO: Biological Proces | GO:00015 | skeletal sy | | 3.94E-05 | 8.40E-02 | 1.75E-03 | 1.44E-02 | 7 | 863 | HSPG2,FLNA,GJA5,CTNNB1,ECM1,ENG,BGN |
| M1/M0 | Up | GO: Biological Proces | GO:00603 | regulation | | 4.24E-05 | 9.03E-02 | 1.84E-03 | 1.52E-02 | 8 | 1212 | FLNA,GJA5,CD81,CTNNB1,ZFP36L1,ITGB1,SGK1,MSN |
| M1/M0 | Up | GO: Biological Proces | GO:00140 | response t | | 4.92E-05 | 1.05E-01 | 1.87E-03 | 1.54E-02 | 9 | 1628 | FLNA,CTNNB1,IGFBP7,ZFP36L1,ENG,ITGB1,SGK1,PGRMC2,MSN |
| M1/M0 | Up | GO: Biological Proces | GO:00082 | positive re | | 4.88E-05 | 1.04E-01 | 1.87E-03 | 1.54E-02 | 8 | 1236 | S100A6,FLNA,COL18A1,CD81,CTNNB1,ECM1,ITGB1,VCAM1 |
| M1/M0 | Up | GO: Biological Proces | GO:00714 | cellular re | | 4.56E-05 | 9.72E-02 | 1.87E-03 | 1.54E-02 | 7 | 883 | FLNA,CTNNB1,ZFP36L1,ITGB1,SGK1,PGRMC2,MSN |
| M1/M0 | Up | GO: Biological Proces | GO:00032 | cardiac ch | | 4.68E-05 | 9.98E-02 | 1.87E-03 | 1.54E-02 | 4 | 176 | GJA5,HEG1,CTNNB1,ENG |
| M1/M0 | Up | GO: Biological Proces | GO:00032 | cardiac ve | | 4.68E-05 | 9.98E-02 | 1.87E-03 | 1.54E-02 | 4 | 176 | GJA5,HEG1,CTNNB1,ENG |
| M1/M0 | Up | GO: Biological Proces | GO:00070 | cell-cell ju | | 4.79E-05 | 1.02E-01 | 1.87E-03 | 1.54E-02 | 4 | 177 | GJA5,HEG1,CTNNB1,ITGB1 |
| M1/M0 | Up | GO: Biological Proces | GO:00973 | establishm | | 4.61E-05 | 9.82E-02 | 1.87E-03 | 1.54E-02 | 2 | 9 | FLNA,ITGB1 |
| M1/M0 | Up | GO: Biological Proces | GO:00303 | regulation | | 5.25E-05 | 1.12E-01 | 1.93E-03 | 1.59E-02 | 8 | 1249 | FLNA,COL18A1,CD81,ECM1,ENG,ITGB1,SGK1,MSN |
| M1/M0 | Up | GO: Biological Proces | GO:00550 | ventricular | | 5.18E-05 | 1.10E-01 | 1.93E-03 | 1.59E-02 | 3 | 63 | HEG1,CTNNB1,ENG |
| M1/M0 | Up | GO: Cellular Compon | GO:00059 | focal adhe | | 3.76E-07 | 8.69E-05 | 2.09E-05 | 1.26E-04 | 7 | 430 | HSPG2,FLNA,CD81,CTNNB1,ENG,ITGB1,MSN |
| M1/M0 | Up | GO: Cellular Compon | GO:00300 | cell-substr | | 4.53E-07 | 1.05E-04 | 2.09E-05 | 1.26E-04 | 7 | 442 | HSPG2,FLNA,CD81,CTNNB1,ENG,ITGB1,MSN |
| M1/M0 | Up | GO: Cellular Compon | GO:00059 | cell-cell ju | | 3.11E-06 | 7.18E-04 | 1.20E-04 | 7.21E-04 | 7 | 590 | HSPG2,FLNA,GJA5,HEG1,CTNNB1,ITGB1,MSN |
| M1/M0 | Up | GO: Cellular Compon | GO:00099 | cell surfac | | 2.61E-04 | 6.02E-02 | 3.35E-03 | 2.01E-02 | 7 | 1178 | HEG1,ENG,ITGB1,VCAM1,LRRC32,BGN,MSN |
| M1/M0 | Up | GO: Cellular Compon | GO:00588 | integral cc | | 4.27E-03 | 9.85E-01 | 2.29E-02 | 1.38E-01 | 7 | 1893 | HSPG2,FLNA,GJA5,CD81,ITGB1,VCAM1,LRRC32 |
| M1/M0 | Up | GO: Cellular Compon | GO:00312 | intrinsic cc | | 5.66E-03 | 1.00E+00 | 2.64E-02 | 1.59E-01 | 7 | 1992 | HSPG2,FLNA,GJA5,CD81,ITGB1,VCAM1,LRRC32 |
| M1/Control | Up | GO: Molecular Functi | GO:00198 | growth fac | | 4.55E-05 | 1.97E-02 | 1.52E-03 | 1.01E-02 | 6 | 167 | SORT1,THBS1,IGFBP7,LRP2,ITGAV,ITGB3 |
| M1/Control | Up | GO: Molecular Functi | GO:00055 | glycosami | | 8.60E-05 | 3.73E-02 | 2.24E-03 | 1.49E-02 | 7 | 272 | CFH,THBS1,SULF1,EFEMP2,APLP2,APOE,GNS |
| M1/Control | Up | GO: Molecular Functi | GO:00020 | protease b | | 7.33E-05 | 3.18E-02 | 2.24E-03 | 1.49E-02 | 6 | 182 | PARK7,ITGA3,ITGAV,ITGB1,ITGB3,ATP5F1A |
| M1/Control | Up | GO: Molecular Functi | GO:00435 | angiostatin | | 8.77E-05 | 3.81E-02 | 2.24E-03 | 1.49E-02 | 2 | 4 | ATP5IF1,ATP5F1A |
| M1/Control | Up | GO: Molecular Functi | GO:00844 | N-acetylgl | | 8.77E-05 | 3.81E-02 | 2.24E-03 | 1.49E-02 | 2 | 4 | SULF1,GNS |
| M1/Control | Up | GO: Molecular Functi | GO:00051 | structural | | 1.49E-04 | 6.45E-02 | 2.85E-03 | 1.90E-02 | 12 | 892 | TPM1,THBS1,MYL6,MYL9,IGFBP7,DSP,H4C5,EFEMP2,DAG1,MACF1,ACTN1,APOE |
| M1/Control | Up | GO: Biological Proces | GO:20012 | negative r | | 6.27E-04 | 1.00E+00 | 1.23E-02 | 1.06E-01 | 6 | 285 | THBS1,SLC25A6,PARK7,ITGAV,PTTG1IP,VDAC2 |
| M1/Control | Up | GO: Biological Proces | GO:00310 | actomyosi | | 6.27E-04 | 1.00E+00 | 1.23E-02 | 1.06E-01 | 6 | 285 | TPM1,MYH10,MYL9,ACTN1,CGNL1,ITGB1 |
| M1/Control | Up | GO: Biological Proces | GO:00986 | import int | | 6.39E-04 | 1.00E+00 | 1.23E-02 | 1.06E-01 | 6 | 286 | THBS1,SLC12A4,PARK7,LRP2,ITGB1,ITGB3 |
| M1/Control | Up | GO: Biological Proces | GO:00109 | negative r | | 6.39E-04 | 1.00E+00 | 1.23E-02 | 1.06E-01 | 6 | 286 | THBS1,BST2,PARK7,LRP2,APLP2,ITIH5 |
| M1/Control | Up | GO: Biological Proces | GO:00717 | basement | | 6.43E-04 | 1.00E+00 | 1.23E-02 | 1.06E-01 | 3 | 46 | GAS2,DAG1,ITGB1 |
| M1/Control | Up | GO: Biological Proces | GO:00714 | cellular re | | 6.43E-04 | 1.00E+00 | 1.23E-02 | 1.06E-01 | 3 | 46 | CD9,APOE,ITGB1 |
| M1/Control | Up | GO: Biological Proces | GO:00506 | regulation | | 6.62E-04 | 1.00E+00 | 1.26E-02 | 1.09E-01 | 8 | 524 | BMP2,THBS1,ATP5IF1,PTPRM,SULF1,APOE,ITGB3,ATP5F1A |
| M1/Control | Up | GO: Biological Proces | GO:00102 | endomem | | 6.79E-04 | 1.00E+00 | 1.26E-02 | 1.09E-01 | 9 | 660 | MYOF,MYH10,NDRG1,CD9,CDIPT,DAG1,ITGA3,RAB2A,TMED3 |
| M1/Control | Up | GO: Biological Proces | GO:00018 | kidney de | | 6.79E-04 | 1.00E+00 | 1.26E-02 | 1.09E-01 | 8 | 526 | CFH,BMP2,THSD7A,TCF21,SULF1,LRP2,ITGA3,ITGB3 |
| M1/Control | Up | GO: Biological Proces | GO:00323 | regulation | | 6.71E-04 | 1.00E+00 | 1.26E-02 | 1.09E-01 | 5 | 190 | ATP5PF,THBS1,APOE,ITGAV,ITGB3 |
| M1/Control | Up | GO: Biological Proces | GO:19047 | regulation | | 6.71E-04 | 1.00E+00 | 1.26E-02 | 1.09E-01 | 4 | 108 | TPM1,PARK7,EFEMP2,NDRG2 |
| M1/Control | Up | GO: Biological Proces | GO:20001 | negative r | | 7.15E-04 | 1.00E+00 | 1.32E-02 | 1.14E-01 | 2 | 11 | ATP5PF,THBS1 |
| M1/Control | Up | GO: Biological Proces | GO:00440 | regulation | | 7.29E-04 | 1.00E+00 | 1.34E-02 | 1.16E-01 | 10 | 810 | TPM1,ATP5PF,MYL9,DSP,GLS,DAG1,APOE,SETD3,TNFRSF21,VDAC2 |
| M1/Control | Up | GO: Biological Proces | GO:00020 | sprouting | | 7.38E-04 | 1.00E+00 | 1.34E-02 | 1.16E-01 | 6 | 294 | FMNL3,THBS1,THSD7A,IGFBP7,ITGB1,TNFRSF21 |
| M1/Control | Up | GO: Biological Proces | GO:00507 | regulation | | 7.58E-04 | 1.00E+00 | 1.37E-02 | 1.19E-01 | 8 | 535 | BMP2,THY1,LRP2,DAG1,MACF1,APOE,ITGB1,PLXNB1 |
| M1/Control | Up | GO: Biological Proces | GO:00519 | positive re | | 7.63E-04 | 1.00E+00 | 1.37E-02 | 1.19E-01 | 7 | 410 | BMP2,LRP2,DAG1,MACF1,APOE,ITGB1,PLXNB1 |
| M1/Control | Up | GO: Biological Proces | GO:01046 | negative r | | 7.78E-04 | 1.00E+00 | 1.38E-02 | 1.19E-01 | 6 | 297 | THBS1,BST2,PARK7,LRP2,APLP2,ITIH5 |
| M1/Control | Up | GO: Biological Proces | GO:00363 | response t | | 7.74E-04 | 1.00E+00 | 1.38E-02 | 1.19E-01 | 3 | 49 | NFE2L1,DAG1,APOE |
| M1/Control | Up | GO: Biological Proces | GO:19021 | positive re | | 7.74E-04 | 1.00E+00 | 1.38E-02 | 1.19E-01 | 3 | 49 | ATP5IF1,SLC25A6,VDAC2 |
| M1/Control | Up | GO: Biological Proces | GO:00720 | renal syste | | 7.95E-04 | 1.00E+00 | 1.39E-02 | 1.20E-01 | 8 | 539 | CFH,BMP2,THSD7A,TCF21,SULF1,LRP2,ITGA3,ITGB3 |
| M1/Control | Up | GO: Biological Proces | GO:00988 | cellular ox | | 7.95E-04 | 1.00E+00 | 1.39E-02 | 1.20E-01 | 4 | 113 | PARK7,SELENOF,PRDX1,APOE |
| M1/Control | Up | GO: Biological Proces | GO:19908 | vascular a | | 7.95E-04 | 1.00E+00 | 1.39E-02 | 1.20E-01 | 4 | 113 | TPM1,PARK7,EFEMP2,NDRG2 |
| M1/Control | Up | GO: Biological Proces | GO:00510 | actin filam | | 8.08E-04 | 1.00E+00 | 1.40E-02 | 1.21E-01 | 5 | 198 | TPM1,MYH10,ACTN1,CGNL1,ITGB1 |
| M1/Control | Up | GO: Biological Proces | GO:00015 | neurotran | | 8.21E-04 | 1.00E+00 | 1.42E-02 | 1.22E-01 | 3 | 50 | PARK7,ITGB1,ITGB3 |
| M1/Control | Up | GO: Biological Proces | GO:00486 | axon exter | | 8.45E-04 | 1.00E+00 | 1.45E-02 | 1.25E-01 | 5 | 200 | THSD7A,MACF1,APOE,ITGB1,PLXNB1 |
| M1/Control | Up | GO: Biological Proces | GO:00069 | muscle co | | 8.54E-04 | 1.00E+00 | 1.46E-02 | 1.26E-01 | 7 | 418 | TPM1,MYOF,MYL6,MYL9,DSP,SULF1,SETD3 |
| M1/Control | Up | GO: Biological Proces | GO:19026 | mitochonc | | 8.70E-04 | 1.00E+00 | 1.48E-02 | 1.28E-01 | 3 | 51 | ATP5IF1,SLC25A6,VDAC2 |
| M1/Control | Up | GO: Biological Proces | GO:00073 | gastrulatic | | 9.03E-04 | 1.00E+00 | 1.52E-02 | 1.32E-01 | 7 | 422 | BMP2,DAG1,MACF1,ITGA3,ITGAV,ITGB1,ITGB3 |
| M1/Control | Up | GO: Biological Proces | GO:00342 | ion transm | | 9.20E-04 | 1.00E+00 | 1.54E-02 | 1.33E-01 | 13 | 1315 | S100A6,ATP5PF,THY1,SLC22A8,SLC12A4,SLC25A6,PARK7,LRP2,ITGAV,ITGB1,ITGB3,ATP5F1A,VDAC2 |
| M1/Control | Up | GO: Biological Proces | GO:00196 | organophc | | 9.15E-04 | 1.00E+00 | 1.54E-02 | 1.33E-01 | 12 | 1148 | CFH,GLYAT,ATP5PF,ATP5IF1,SERINC1,NFE2L1,PARK7,CDIPT,AKR1A1,LDHB,ATP5F1A,NDUFS5 |
| M1/Control | Up | GO: Biological Proces | GO:00615 | actin filam | | 9.44E-04 | 1.00E+00 | 1.57E-02 | 1.36E-01 | 5 | 205 | TPM1,MYH10,ACTN1,CGNL1,ITGB1 |
| M1/Control | Up | GO: Biological Proces | GO:00107 | regulation | | 9.74E-04 | 1.00E+00 | 1.61E-02 | 1.39E-01 | 3 | 53 | THBS1,ITGB1,ITGB3 |
| M1/Control | Up | GO: Biological Proces | GO:00072 | small GTP | | 9.94E-04 | 1.00E+00 | 1.64E-02 | 1.41E-01 | 8 | 558 | PARK7,CGNL1,ITGA3,APOE,ITGAV,RAB2A,ITGB1,CDC42EP3 |
| M1/Control | Up | GO: Biological Proces | GO:00332 | T-tubule o | | 1.01E-03 | 1.00E+00 | 1.65E-02 | 1.43E-01 | 2 | 13 | MYOF,DAG1 |
| M1/Control | Up | GO: Biological Proces | GO:00303 | positive re | | 1.02E-03 | 1.00E+00 | 1.66E-02 | 1.43E-01 | 9 | 699 | BMP2,THBS1,THY1,CD151,ITGA3,ITGAV,ITGB1,ITGB3,ATP5F1A |
| M1/Control | Up | GO: Biological Proces | GO:00357 | positive re | | 1.03E-03 | 1.00E+00 | 1.67E-02 | 1.44E-01 | 3 | 54 | ATP5IF1,SLC25A6,VDAC2 |
| M1/Control | Up | GO: Biological Proces | GO:00030 | muscle sys | | 1.08E-03 | 1.00E+00 | 1.74E-02 | 1.50E-01 | 8 | 565 | TPM1,MYOF,MYL6,MYL9,DSP,SULF1,DAG1,SETD3 |
| M1/Control | Up | GO: Biological Proces | GO:00030 | circulatory | | 1.09E-03 | 1.00E+00 | 1.75E-02 | 1.51E-01 | 10 | 854 | TPM1,ATP5PF,MYOF,SLC22A8,PTPRM,DSP,SULF1,LRP2,APOE,VDAC2 |
| M1/Control | Up | GO: Biological Proces | GO:00513 | regulation | | 1.14E-03 | 1.00E+00 | 1.82E-02 | 1.58E-01 | 12 | 1178 | SORT1,BMP2,THBS1,ATP5IF1,THY1,BST2,PARK7,LRP2,APLP2,ITGB1,PLXNB1,ITIH5 |
| M1/Control | Up | GO: Biological Proces | GO:00107 | negative r | | 1.17E-03 | 1.00E+00 | 1.86E-02 | 1.61E-01 | 2 | 14 | ITGAV,ITGB3 |
| M1/Control | Up | GO: Biological Proces | GO:00971 | apoptotic | | 1.19E-03 | 1.00E+00 | 1.88E-02 | 1.63E-01 | 9 | 715 | SORT1,THBS1,ATP5IF1,SLC25A6,PARK7,ITM2B,ITGAV,PTTG1IP,VDAC2 |
| M1/Control | Up | GO: Biological Proces | GO:00432 | apoptotic | | 1.20E-03 | 1.00E+00 | 1.89E-02 | 1.63E-01 | 3 | 57 | THBS1,ITGAV,ITGB3 |
| M1/Control | Up | GO: Biological Proces | GO:00068 | receptor-m | | 1.24E-03 | 1.00E+00 | 1.93E-02 | 1.67E-01 | 6 | 325 | CD9,LRP2,APOE,ITGAV,ITGB1,ITGB3 |
| M1/Control | Up | GO: Cellular Compon | GO:00059 | brush bor | | 3.39E-04 | 1.31E-01 | 2.77E-03 | 1.81E-02 | 5 | 163 | MYH10,MYL6,LRP2,ACTN1,MYL12B |
| M1/Control | Up | GO: Cellular Compon | GO:00701 | anchoring | | 6.04E-04 | 2.34E-01 | 4.67E-03 | 3.05E-02 | 14 | 1419 | NDRG1,THY1,CD9,PTPRM,DSP,PARK7,DAG1,CD151,ACTN1,CGNL1,ITGA3,ITGAV,ITGB1,ITGB3 |
| M1/Control | Down | GO: Biological Proces | GO:00328 | cellular re | | 6.99E-06 | 8.07E-03 | 1.38E-03 | 1.05E-02 | 6 | 866 | EGR1,FOS,ZFP36,DUSP1,REN,NR4A1 |
| M1/Control | Down | GO: Biological Proces | GO:00096 | response t | | 5.46E-05 | 6.31E-02 | 4.11E-03 | 3.13E-02 | 6 | 1242 | FOS,ZFP36,REN,IGKC,CEBPB,NR4A1 |
| M1/Control | Down | GO: Biological Proces | GO:00097 | response t | | 9.47E-05 | 1.09E-01 | 5.96E-03 | 4.55E-02 | 6 | 1370 | EGR1,FOS,ZFP36,DUSP1,REN,NR4A1 |
| M1/Control | Down | GO: Biological Proces | GO:00102 | response t | | 2.43E-04 | 2.80E-01 | 1.30E-02 | 9.89E-02 | 6 | 1623 | EGR1,FOS,DUSP1,REN,CEBPB,NR4A1 |
| M1/Control | Down | GO: Biological Proces | GO:00140 | response t | | 2.47E-04 | 2.85E-01 | 1.30E-02 | 9.89E-02 | 6 | 1628 | EGR1,FOS,ZFP36,DUSP1,REN,CEBPB |
| M1/Control | Down | GO: Biological Proces | GO:19016 | response t | | 3.53E-04 | 4.08E-01 | 1.63E-02 | 1.25E-01 | 6 | 1738 | EGR1,FOS,DUSP1,REN,CEBPB,NR4A1 |
| M1/Control | Down | GO: Biological Proces | GO:00459 | negative r | | 4.19E-04 | 4.84E-01 | 1.75E-02 | 1.34E-01 | 6 | 1793 | EGR1,RASD1,ZFP36,ATF3,DUSP1,CEBPB |
| M1/Control | Down | GO: Biological Proces | GO:00026 | regulation | | 4.56E-04 | 5.27E-01 | 1.75E-02 | 1.34E-01 | 6 | 1821 | FOS,ZFP36,DUSP1,IGKC,CEBPB,USP17L12 |
| M1/Control | Down | GO: Biological Proces | GO:00105 | negative r | | 4.94E-04 | 5.71E-01 | 1.75E-02 | 1.34E-01 | 6 | 1848 | EGR1,RASD1,ZFP36,ATF3,DUSP1,CEBPB |
| M1/Control | Down | GO: Biological Proces | GO:00313 | negative r | | 6.46E-04 | 7.47E-01 | 1.75E-02 | 1.34E-01 | 6 | 1942 | EGR1,RASD1,ZFP36,ATF3,DUSP1,CEBPB |
| M1/Control | Down | GO: Biological Proces | GO:00098 | negative r | | 7.41E-04 | 8.57E-01 | 1.86E-02 | 1.42E-01 | 6 | 1992 | EGR1,RASD1,ZFP36,ATF3,DUSP1,CEBPB |
| M1/M0 | Up | GO: Molecular Functi | GO:00508 | extracellul | | 6.35E-05 | 1.33E-02 | 2.65E-03 | 1.57E-02 | 3 | 65 | ECM1,ITGB1,BGN |
| M1/M0 | Up | GO: Biological Proces | GO:00607 | chorio-alla | | 5.75E-05 | 1.23E-01 | 2.08E-03 | 1.71E-02 | 2 | 10 | ZFP36L1,VCAM1 |
| M1/M0 | Up | GO: Biological Proces | GO:00506 | epithelial | | 6.33E-05 | 1.35E-01 | 2.25E-03 | 1.85E-02 | 6 | 627 | HSPG2,COL18A1,CTNNB1,ZFP36L1,ECM1,ENG |
| M1/M0 | Up | GO: Biological Proces | GO:00510 | negative r | | 6.95E-05 | 1.48E-01 | 2.43E-03 | 2.00E-02 | 8 | 1299 | HSPG2,TCF4,CTNNB1,RCAN1,ZFP36L1,ECM1,ITGB1,H4C5 |
| M1/M0 | Up | GO: Biological Proces | GO:20001 | regulation | | 7.82E-05 | 1.67E-01 | 2.60E-03 | 2.15E-02 | 8 | 1321 | FLNA,COL18A1,CD81,ECM1,ENG,ITGB1,SGK1,MSN |
| M1/M0 | Up | GO: Biological Proces | GO:00020 | morphoge | | 8.07E-05 | 1.72E-01 | 2.60E-03 | 2.15E-02 | 7 | 966 | COL4A1,FLNA,CTNNB1,IGFBP7,ENG,ITGB1,MSN |
| M1/M0 | Up | GO: Biological Proces | GO:00301 | extracellul | | 7.65E-05 | 1.63E-01 | 2.60E-03 | 2.15E-02 | 5 | 394 | COL4A1,HSPG2,COL18A1,ENG,ITGB1 |
| M1/M0 | Up | GO: Biological Proces | GO:00430 | extracellul | | 7.74E-05 | 1.65E-01 | 2.60E-03 | 2.15E-02 | 5 | 395 | COL4A1,HSPG2,COL18A1,ENG,ITGB1 |
| M1/M0 | Up | GO: Biological Proces | GO:00452 | external e | | 8.02E-05 | 1.71E-01 | 2.60E-03 | 2.15E-02 | 5 | 398 | COL4A1,HSPG2,COL18A1,ENG,ITGB1 |
| M1/M0 | Up | GO: Biological Proces | GO:20001 | positive re | | 8.73E-05 | 1.86E-01 | 2.74E-03 | 2.25E-02 | 3 | 75 | FLNA,CTNNB1,ITGB1 |
| M1/M0 | Up | GO: Biological Proces | GO:00032 | ventricular | | 8.73E-05 | 1.86E-01 | 2.74E-03 | 2.25E-02 | 3 | 75 | HEG1,CTNNB1,ENG |
| M1/M0 | Up | GO: Biological Proces | GO:00466 | lymphocyt | | 9.55E-05 | 2.04E-01 | 2.95E-03 | 2.43E-02 | 5 | 413 | CD81,CTNNB1,VCAM1,LRRC32,MSN |
| M1/M0 | Up | GO: Biological Proces | GO:00096 | response t | | 1.02E-04 | 2.17E-01 | 3.01E-03 | 2.48E-02 | 9 | 1786 | COL18A1,CTNNB1,IGFBP7,RCAN1,ZFP36L1,ENG,ITGB1,SGK1,VCAM1 |
| M1/M0 | Up | GO: Biological Proces | GO:00097 | response t | | 1.01E-04 | 2.16E-01 | 3.01E-03 | 2.48E-02 | 8 | 1370 | CTNNB1,IGFBP7,RCAN1,ZFP36L1,ENG,ITGB1,SGK1,PGRMC2 |
| M1/M0 | Up | GO: Biological Proces | GO:00487 | branching | | 1.00E-04 | 2.13E-01 | 3.01E-03 | 2.48E-02 | 4 | 214 | COL4A1,CTNNB1,IGFBP7,ENG |
| M1/M0 | Up | GO: Biological Proces | GO:00329 | mononucl | | 1.03E-04 | 2.20E-01 | 3.02E-03 | 2.49E-02 | 5 | 420 | CD81,CTNNB1,VCAM1,LRRC32,MSN |
| M1/M0 | Up | GO: Biological Proces | GO:00400 | regulation | | 1.06E-04 | 2.27E-01 | 3.07E-03 | 2.53E-02 | 8 | 1380 | FLNA,COL18A1,CD81,ECM1,ENG,ITGB1,SGK1,MSN |
| M1/M0 | Up | GO: Biological Proces | GO:00421 | T cell activ | | 1.20E-04 | 2.56E-01 | 3.41E-03 | 2.81E-02 | 6 | 704 | CD81,CTNNB1,ZFP36L1,VCAM1,LRRC32,MSN |
| M1/M0 | Up | GO: Biological Proces | GO:00219 | hindbrain | | 1.34E-04 | 2.85E-01 | 3.75E-03 | 3.09E-02 | 2 | 15 | FLNA,ITGB1 |
| M1/M0 | Up | GO: Biological Proces | GO:00466 | lymphocyt | | 1.43E-04 | 3.04E-01 | 3.95E-03 | 3.26E-02 | 7 | 1058 | CD81,CTNNB1,ZFP36L1,ITGB1,VCAM1,LRRC32,MSN |
| M1/M0 | Up | GO: Biological Proces | GO:00550 | cardiac m | | 1.45E-04 | 3.10E-01 | 3.97E-03 | 3.27E-02 | 3 | 89 | HEG1,CTNNB1,ENG |
| M1/M0 | Up | GO: Biological Proces | GO:00706 | leukocyte | | 1.57E-04 | 3.34E-01 | 4.23E-03 | 3.48E-02 | 5 | 459 | CD81,CTNNB1,VCAM1,LRRC32,MSN |
| M1/M0 | Up | GO: Biological Proces | GO:00328 | regulation | | 1.61E-04 | 3.44E-01 | 4.24E-03 | 3.49E-02 | 7 | 1079 | FLNA,GJA5,CD81,CTNNB1,ITGB1,SGK1,MSN |
| M1/M0 | Up | GO: Biological Proces | GO:00019 | endothelia | | 1.61E-04 | 3.42E-01 | 4.24E-03 | 3.49E-02 | 4 | 242 | HSPG2,COL18A1,ECM1,ENG |
| M1/M0 | Up | GO: Biological Proces | GO:00032 | cardiac ch | | 1.63E-04 | 3.48E-01 | 4.24E-03 | 3.49E-02 | 4 | 243 | GJA5,HEG1,CTNNB1,ENG |
| M1/M0 | Up | GO: Biological Proces | GO:20000 | regulation | | 1.70E-04 | 3.61E-01 | 4.35E-03 | 3.59E-02 | 9 | 1908 | HSPG2,TCF4,CTNNB1,ZFP36L1,ECM1,ENG,ITGB1,SGK1,H4C5 |
| M1/M0 | Up | GO: Biological Proces | GO:00614 | connective | | 1.73E-04 | 3.69E-01 | 4.39E-03 | 3.62E-02 | 5 | 469 | HSPG2,CTNNB1,ECM1,PGRMC2,BGN |
| M1/M0 | Up | GO: Biological Proces | GO:00030 | heart mor | | 1.77E-04 | 3.76E-01 | 4.43E-03 | 3.65E-02 | 5 | 471 | FLNA,GJA5,HEG1,CTNNB1,ENG |
| M1/M0 | Up | GO: Biological Proces | GO:00421 | regulation | | 1.82E-04 | 3.87E-01 | 4.45E-03 | 3.67E-02 | 4 | 250 | CD81,CTNNB1,VCAM1,LRRC32 |
| M1/M0 | Up | GO: Biological Proces | GO:00032 | cardiac ve | | 1.82E-04 | 3.87E-01 | 4.45E-03 | 3.67E-02 | 3 | 96 | HEG1,CTNNB1,ENG |
| M1/M0 | Up | GO: Biological Proces | GO:00611 | morphoge | | 1.87E-04 | 3.99E-01 | 4.54E-03 | 3.74E-02 | 4 | 252 | COL4A1,CTNNB1,IGFBP7,ENG |
| M1/M0 | Up | GO: Biological Proces | GO:00017 | immunolo | | 1.95E-04 | 4.15E-01 | 4.61E-03 | 3.80E-02 | 2 | 18 | CD81,MSN |
| M1/M0 | Up | GO: Biological Proces | GO:00488 | venous bl | | 1.95E-04 | 4.15E-01 | 4.61E-03 | 3.80E-02 | 2 | 18 | HEG1,ENG |
| M1/M0 | Up | GO: Biological Proces | GO:00512 | positive re | | 2.00E-04 | 4.27E-01 | 4.68E-03 | 3.86E-02 | 9 | 1950 | GJA5,CD81,HEG1,CTNNB1,ZFP36L1,ECM1,ENG,ITGB1,SGK1 |
| M1/M0 | Up | GO: Cellular Compon | GO:00987 | plasma me | | 2.16E-04 | 4.99E-02 | 2.93E-03 | 1.77E-02 | 6 | 792 | HSPG2,FLNA,GJA5,CTNNB1,ITGB1,VCAM1 |
| M1/M0 | Up | GO: Cellular Compon | GO:00985 | side of me | | 3.22E-04 | 7.45E-02 | 3.72E-03 | 2.24E-02 | 6 | 853 | S100A6,HEG1,ENG,ITGB1,VCAM1,MSN |
| M1/M0 | Up | GO: Cellular Compon | GO:00484 | perinuclea | | 5.57E-04 | 1.29E-01 | 5.36E-03 | 3.23E-02 | 6 | 945 | S100A6,FLNA,CTNNB1,ITGB1,SGK1,MSN |
| M1/M0 | Up | GO: Cellular Compon | GO:00987 | membrane | | 5.24E-03 | 1.00E+00 | 2.57E-02 | 1.55E-01 | 6 | 1465 | HSPG2,FLNA,GJA5,CTNNB1,ITGB1,VCAM1 |
| M1/M0 | Up | GO: Cellular Compon | GO:00985 | plasma me | | 5.84E-03 | 1.00E+00 | 2.65E-02 | 1.59E-01 | 6 | 1498 | CD81,CTNNB1,ENG,ITGB1,VCAM1,MSN |
| M1/Control | Up | GO: Molecular Functi | GO:00037 | actin bind | | 1.23E-04 | 5.33E-02 | 2.85E-03 | 1.90E-02 | 9 | 497 | TPM1,FMNL3,MYH10,GAS2,DAG1,MACF1,ACTN1,SETD3,ITGB1 |
| M1/Control | Up | GO: Molecular Functi | GO:00050 | protein kir | | 1.46E-04 | 6.35E-02 | 2.85E-03 | 1.90E-02 | 4 | 69 | SLC22A8,DSP,ITGAV,ITGB3 |
| M1/Control | Up | GO: Molecular Functi | GO:00171 | fibroblast | | 1.51E-04 | 6.55E-02 | 2.85E-03 | 1.90E-02 | 3 | 27 | THBS1,ITGAV,ITGB3 |
| M1/Control | Up | GO: Molecular Functi | GO:00551 | cytoskelet | | 1.46E-04 | 6.33E-02 | 2.85E-03 | 1.90E-02 | 2 | 5 | ACTN1,CDC42EP3 |
| M1/Control | Up | GO: Biological Proces | GO:00902 | regulation | | 1.26E-03 | 1.00E+00 | 1.96E-02 | 1.69E-01 | 7 | 447 | BMP2,MYOF,THBS1,SULF1,LRP2,ITGA3,ITGB3 |
| M1/Control | Up | GO: Biological Proces | GO:19021 | regulation | | 1.27E-03 | 1.00E+00 | 1.96E-02 | 1.69E-01 | 3 | 58 | ATP5IF1,SLC25A6,VDAC2 |
| M1/Control | Up | GO: Biological Proces | GO:00600 | retina dev | | 1.28E-03 | 1.00E+00 | 1.97E-02 | 1.70E-01 | 7 | 448 | CFH,S100A6,MYH10,THY1,PTPRM,CDIPT,ITGB1 |
| M1/Control | Up | GO: Biological Proces | GO:00425 | myelinatio | | 1.29E-03 | 1.00E+00 | 1.98E-02 | 1.71E-01 | 5 | 220 | KLK6,NDRG1,CD9,DAG1,TNFRSF21 |
| M1/Control | Up | GO: Biological Proces | GO:00435 | blood ves | | 1.35E-03 | 1.00E+00 | 2.05E-02 | 1.77E-01 | 5 | 222 | THBS1,THSD7A,APOE,ITGB1,ATP5F1A |
| M1/Control | Up | GO: Biological Proces | GO:00300 | muscle fila | | 1.35E-03 | 1.00E+00 | 2.05E-02 | 1.77E-01 | 2 | 15 | TPM1,MYL6 |
| M1/Control | Up | GO: Biological Proces | GO:00083 | axon ensh | | 1.37E-03 | 1.00E+00 | 2.05E-02 | 1.78E-01 | 5 | 223 | KLK6,NDRG1,CD9,DAG1,TNFRSF21 |
| M1/Control | Up | GO: Biological Proces | GO:19059 | regulation | | 1.37E-03 | 1.00E+00 | 2.05E-02 | 1.78E-01 | 5 | 223 | ATP5PF,THBS1,APOE,ITGAV,ITGB3 |
| M1/Control | Up | GO: Biological Proces | GO:00072 | ensheathin | | 1.37E-03 | 1.00E+00 | 2.05E-02 | 1.78E-01 | 5 | 223 | KLK6,NDRG1,CD9,DAG1,TNFRSF21 |
| M1/Control | Up | GO: Biological Proces | GO:20001 | positive re | | 1.39E-03 | 1.00E+00 | 2.07E-02 | 1.79E-01 | 9 | 731 | BMP2,THBS1,THY1,CD151,ITGA3,ITGAV,ITGB1,ITGB3,ATP5F1A |
| M1/Control | Up | GO: Biological Proces | GO:00068 | anion tran | | 1.40E-03 | 1.00E+00 | 2.08E-02 | 1.80E-01 | 8 | 589 | ATP5PF,SLC22A8,SLC12A4,SLC25A6,NFE2L1,LRP2,ITGB1,VDAC2 |
| M1/Control | Up | GO: Biological Proces | GO:00016 | urogenital | | 1.43E-03 | 1.00E+00 | 2.12E-02 | 1.83E-01 | 8 | 591 | CFH,BMP2,THSD7A,TCF21,SULF1,LRP2,ITGA3,ITGB3 |

| | | | | | | | | | | |
|---|---|---|---|---|---|---|---|---|---|---|
| M1/Control | Up | GO: Biological Proces | GO:006144connective | 1.66E-03 | 1.00E+00 | 2.37E-02 | 2.05E-01 | 7 | 469 | CFH,CNBP,BMP2,THBS1,SULF1,FRZB,ITGB3 |
| M1/Control | Up | GO: Biological Proces | GO:000741central ne | 1.68E-03 | 1.00E+00 | 2.39E-02 | 2.07E-01 | 15 | 1766 | ATP5PF,BMP2,MYH10,KLK6,CD9,NFE2L1,SULF1,CDIPT,LRP2,DAG1,APLP2,APOE,NDRG2,ITGB1,TNFRSF21 |
| M1/Control | Up | GO: Biological Proces | GO:004001positive re | 1.69E-03 | 1.00E+00 | 2.39E-02 | 2.07E-01 | 9 | 752 | BMP2,THBS1,THY1,CD151,ITGA3,ITGAV,ITGB1,ITGB3,ATP5F1A |
| M1/Control | Up | GO: Biological Proces | GO:00518regulation | 1.70E-03 | 1.00E+00 | 2.39E-02 | 2.07E-01 | 5 | 234 | THBS1,PARK7,LRP2,DAG1,ITGB1 |
| M1/Control | Up | GO: Biological Proces | GO:19029negative n | 1.74E-03 | 1.00E+00 | 2.45E-02 | 2.12E-01 | 2 | 17 | BMP2,APOE |
| M1/Control | Up | GO: Biological Proces | GO:00458positive re | 1.76E-03 | 1.00E+00 | 2.45E-02 | 2.12E-01 | 4 | 140 | LRP2,CD151,APOE,ITGB1 |
| M1/Control | Up | GO: Biological Proces | GO:190571positive re | 1.76E-03 | 1.00E+00 | 2.45E-02 | 2.12E-01 | 3 | 65 | ATP5IF1,SLC25A6,VDAC2 |
| M1/Control | Up | GO: Biological Proces | GO:00074mesoderm | 1.79E-03 | 1.00E+00 | 2.49E-02 | 2.15E-01 | 5 | 237 | BMP2,MACF1,ITGA3,ITGB1,ITGB3 |
| M1/Control | Up | GO: Biological Proces | GO:007126cellular re | 1.85E-03 | 1.00E+00 | 2.56E-02 | 2.21E-01 | 4 | 142 | BMP2,DAG1,ITGB1,ITGB3 |
| M1/Control | Up | GO: Biological Proces | GO:00604epithelium | 1.92E-03 | 1.00E+00 | 2.63E-02 | 2.28E-01 | 16 | 1979 | CFH,BMP2,MYH10,THSD7A,TCF21,IGFBP7,DSP,NFE2L1,SULF1,FRZB,LRP2,DAG1,CD151,ITGAV,ITGB1,TNFRSF21 |
| M1/Control | Up | GO: Biological Proces | GO:00510negative n | 1.92E-03 | 1.00E+00 | 2.63E-02 | 2.28E-01 | 3 | 67 | CGNL1,ITGA3,ITGB1 |
| M1/Control | Up | GO: Biological Proces | GO:00332actin-myo | 1.96E-03 | 1.00E+00 | 2.65E-02 | 2.29E-01 | 2 | 18 | TPM1,MYL6 |
| M1/Control | Up | GO: Biological Proces | GO:00019suckling b | 1.96E-03 | 1.00E+00 | 2.65E-02 | 2.29E-01 | 2 | 18 | GLS,APLP2 |
| M1/Control | Up | GO: Biological Proces | GO:00032cardiac ch | 2.00E-03 | 1.00E+00 | 2.68E-02 | 2.32E-01 | 5 | 243 | TPM1,BMP2,MYH10,DSP,LRP2 |
| M1/Control | Up | GO: Biological Proces | GO:00602camera-ty | 2.00E-03 | 1.00E+00 | 2.68E-02 | 2.32E-01 | 3 | 68 | CFH,THY1,CDIPT |
| M1/Control | Up | GO: Biological Proces | GO:00075myoblast f | 2.00E-03 | 1.00E+00 | 2.68E-02 | 2.32E-01 | 3 | 68 | MYOF,CD9,ITGB1 |
| M1/Control | Up | GO: Biological Proces | GO:00100response t | 2.06E-03 | 1.00E+00 | 2.73E-02 | 2.36E-01 | 10 | 930 | CFH,THBS1,NDRG1,CD9,NFE2L1,PARK7,PRDX1,APOE,ITGAV,ATP5F1A |
| M1/Control | Up | GO: Biological Proces | GO:00602anatomica | 2.05E-03 | 1.00E+00 | 2.73E-02 | 2.36E-01 | 7 | 487 | SULF1,CDIPT,PRDX1,LAMP2,ITGAV,ITGB1,ITGB3 |
| M1/Control | Up | GO: Biological Proces | GO:00068endocytos | 2.07E-03 | 1.00E+00 | 2.75E-02 | 2.37E-01 | 9 | 775 | SORT1,THBS1,CD9,LRP2,CD151,APOE,ITGAV,ITGB1,ITGB3 |
| M1/Control | Up | GO: Biological Proces | GO:00095fertilizatio | 2.15E-03 | 1.00E+00 | 2.83E-02 | 2.45E-01 | 5 | 247 | CD9,PARK7,ITGA3,ITGAV,VDAC2 |
| M1/Control | Up | GO: Biological Proces | GO:00512cartilage d | 2.16E-03 | 1.00E+00 | 2.84E-02 | 2.45E-01 | 6 | 363 | CFH,CNBP,BMP2,THBS1,SULF1,FRZB |
| M1/Control | Up | GO: Biological Proces | GO:00427proton mo | 2.18E-03 | 1.00E+00 | 2.84E-02 | 2.45E-01 | 3 | 70 | ATP5PF,ATP5F1A,NDUFS5 |
| M1/Control | Up | GO: Biological Proces | GO:00507regulation | 2.18E-03 | 1.00E+00 | 2.84E-02 | 2.45E-01 | 2 | 19 | ITGAV,ITGB3 |
| M1/Control | Up | GO: Biological Proces | GO:00518positive re | 2.21E-03 | 1.00E+00 | 2.86E-02 | 2.47E-01 | 4 | 149 | THBS1,PARK7,LRP2,ITGB1 |
| M1/Control | Up | GO: Biological Proces | GO:00300actin filam | 2.26E-03 | 1.00E+00 | 2.90E-02 | 2.51E-01 | 4 | 150 | TPM1,MYH10,MYL6,DSP |
| M1/Control | Up | GO: Biological Proces | GO:00518regulation | 2.27E-03 | 1.00E+00 | 2.90E-02 | 2.51E-01 | 3 | 71 | THBS1,THY1,MACF1 |
| M1/Control | Up | GO: Cellular Compon | GO:00989glutamate | 6.85E-04 | 2.65E-01 | 5.20E-03 | 3.40E-02 | 8 | 525 | MYH10,NDRG1,DAG1,ACTN1,APOE,NDRG2,ITGB1,ITGB3 |
| M1/Control | Up | GO: Cellular Compon | GO:00424melanosor | 7.78E-04 | 3.01E-01 | 5.68E-03 | 3.72E-02 | 4 | 112 | PRDX1,RAB2A,ITGB1,ITGB3 |
| M1/Control | Up | GO: Cellular Compon | GO:00487pigment g | 7.78E-04 | 3.01E-01 | 5.68E-03 | 3.72E-02 | 4 | 112 | PRDX1,RAB2A,ITGB1,ITGB3 |
| M1/Control | Up | GO: Cellular Compon | GO:00987membrane | 8.26E-04 | 3.20E-01 | 5.92E-03 | 3.87E-02 | 14 | 1465 | ATP5PF,BMP2,SLC25A6,DAD1,DAG1,LAMP2,ITGA3,ITGAV,ITGB1,ITGB3,ATP5F1A,NDUFS5,VDAC2,TMED3 |
| M1/Control | Up | GO: Cellular Compon | GO:00059fascia adh | 1.02E-03 | 3.93E-01 | 7.14E-03 | 4.67E-02 | 2 | 13 | DSP,ACTN1 |
| M1/Control | Up | GO: Cellular Compon | GO:00057lysosomal | 1.08E-03 | 4.19E-01 | 7.35E-03 | 4.81E-02 | 7 | 434 | SORT1,LAPTM4A,SLC12A4,BST2,LRP2,LAMP2,RAB2A |
| M1/Control | Up | GO: Cellular Compon | GO:00988lytic vacuo | 1.08E-03 | 4.19E-01 | 7.35E-03 | 4.81E-02 | 7 | 434 | SORT1,LAPTM4A,SLC12A4,BST2,LRP2,LAMP2,RAB2A |
| M1/Control | Up | GO: Cellular Compon | GO:00451apical part | 1.15E-03 | 4.45E-01 | 7.68E-03 | 5.02E-02 | 8 | 569 | THY1,SLC22A8,CD9,BST2,LRP2,AKR1A1,MYL12B,ITGB3 |
| M1/Control | Up | GO: Cellular Compon | GO:00056basement | 1.43E-03 | 5.54E-01 | 9.40E-03 | 6.14E-02 | 4 | 132 | EFEMP2,DAG1,CD151,ITGB1 |
| M1/Control | Up | GO: Cellular Compon | GO:00163apical plas | 1.59E-03 | 6.15E-01 | 1.03E-02 | 6.70E-02 | 7 | 464 | THY1,SLC22A8,CD9,BST2,LRP2,AKR1A1,ITGB3 |
| M1/Control | Up | GO: Cellular Compon | GO:00988cluster of | 1.66E-03 | 6.41E-01 | 1.05E-02 | 6.87E-02 | 5 | 232 | MYH10,MYL6,LRP2,ACTN1,MYL12B |
| M1/Control | Up | GO: Cellular Compon | GO:00057vacuolar n | 1.90E-03 | 7.37E-01 | 1.19E-02 | 7.77E-02 | 7 | 479 | SORT1,LAPTM4A,SLC12A4,BST2,LRP2,LAMP2,RAB2A |
| M1/Control | Up | GO: Cellular Compon | GO:00304growth co | 2.18E-03 | 8.42E-01 | 1.34E-02 | 8.74E-02 | 5 | 247 | MYH10,THY1,LRP2,ITGA3,NDRG2 |
| M1/Control | Up | GO: Cellular Compon | GO:00147intercalate | 2.29E-03 | 8.85E-01 | 1.38E-02 | 9.04E-02 | 3 | 71 | DSP,ACTN1,ITGB1 |
| M1/Control | Up | GO: Cellular Compon | GO:00484perinuclea | 2.37E-03 | 9.17E-01 | 1.41E-02 | 9.22E-02 | 10 | 945 | SORT1,S100A6,NDRG1,PTPRM,PARK7,LAMP2,ITGA3,RAB2A,NDRG2,ITGB1 |
| M1/Control | Up | GO: Cellular Compon | GO:00057Golgi appa | 2.44E-03 | 9.45E-01 | 1.42E-02 | 9.27E-02 | 15 | 1828 | SORT1,FMNL3,LAPTM4A,BST2,SULF1,CDIPT,LRP2,DAG1,MACF1,LAMP2,APOE,ITM2B,RAB2A,NDRG2,TMED3 |
| M1/Control | Up | GO: Cellular Compon | GO:00304site of pol | 2.46E-03 | 9.50E-01 | 1.42E-02 | 9.27E-02 | 5 | 254 | MYH10,THY1,LRP2,ITGA3,NDRG2 |
| M1/Control | Up | GO: Cellular Compon | GO:00057mitochonc | 2.68E-03 | 1.00E+00 | 1.53E-02 | 9.97E-02 | 2 | 21 | ATP5PF,ATP5F1A |
| M1/Control | Up | GO: Cellular Compon | GO:00306clathrin-co | 2.88E-03 | 1.00E+00 | 1.62E-02 | 1.06E-01 | 3 | 77 | CD9,LRP2,APOE |
| M1/Control | Down | GO: Molecular Functi | GO:00122DNA-bind | 2.99E-05 | 2.84E-03 | 7.40E-04 | 3.80E-03 | 5 | 569 | EGR1,FOS,ATF3,CEBPB,NR4A1 |
| M1/Control | Down | GO: Molecular Functi | GO:00121DNA-bind | 3.12E-05 | 2.96E-03 | 7.40E-04 | 3.80E-03 | 5 | 574 | EGR1,FOS,ATF3,CEBPB,NR4A1 |
| M1/Control | Down | GO: Molecular Functi | GO:00093RNA polyr | 1.35E-03 | 1.28E-01 | 1.54E-02 | 7.89E-02 | 5 | 1286 | EGR1,FOS,ATF3,CEBPB,NR4A1 |
| M1/Control | Down | GO: Molecular Functi | GO:00009cis-regulat | 1.46E-03 | 1.38E-01 | 1.54E-02 | 7.89E-02 | 5 | 1308 | EGR1,FOS,ATF3,CEBPB,NR4A1 |
| M1/Control | Down | GO: Molecular Functi | GO:00098DNA-bind | 2.15E-03 | 2.04E-01 | 1.86E-02 | 9.53E-02 | 5 | 1427 | EGR1,FOS,ATF3,CEBPB,NR4A1 |
| M1/Control | Down | GO: Molecular Functi | GO:00037DNA-bind | 2.55E-03 | 2.42E-01 | 1.87E-02 | 9.59E-02 | 5 | 1483 | EGR1,FOS,ATF3,CEBPB,NR4A1 |
| M1/Control | Down | GO: Molecular Functi | GO:00093RNA polyr | 2.65E-03 | 2.52E-01 | 1.87E-02 | 9.59E-02 | 5 | 1496 | EGR1,FOS,ATF3,CEBPB,NR4A1 |
| M1/Control | Down | GO: Molecular Functi | GO:00097transcripti | 3.55E-03 | 3.37E-01 | 2.01E-02 | 1.03E-01 | 5 | 1599 | EGR1,FOS,ATF3,CEBPB,NR4A1 |
| M1/Control | Down | GO: Molecular Functi | GO:00010transcripti | 3.60E-03 | 3.42E-01 | 2.01E-02 | 1.03E-01 | 5 | 1604 | EGR1,FOS,ATF3,CEBPB,NR4A1 |
| M1/Control | Down | GO: Molecular Functi | GO:19908sequence- | 4.16E-03 | 3.95E-01 | 2.19E-02 | 1.13E-01 | 5 | 1658 | EGR1,FOS,ATF3,CEBPB,NR4A1 |
| M1/Control | Down | GO: Molecular Functi | GO:00435sequence- | 5.58E-03 | 5.30E-01 | 2.53E-02 | 1.30E-01 | 5 | 1775 | EGR1,FOS,ATF3,CEBPB,NR4A1 |
| M1/Control | Down | GO: Molecular Functi | GO:00036double-str | 5.60E-03 | 5.32E-01 | 2.53E-02 | 1.30E-01 | 5 | 1776 | EGR1,FOS,ATF3,CEBPB,NR4A1 |
| M1/Control | Down | GO: Biological Proces | GO:00324response t | 2.07E-05 | 2.40E-02 | 2.66E-03 | 2.03E-02 | 5 | 597 | FOS,ZFP36,REN,CEBPB,NR4A1 |
| M1/Control | Down | GO: Biological Proces | GO:00022response t | 2.57E-05 | 2.97E-02 | 2.70E-03 | 2.06E-02 | 5 | 624 | FOS,ZFP36,REN,CEBPB,NR4A1 |
| M1/Control | Down | GO: Biological Proces | GO:00109positive re | 9.80E-05 | 1.13E-01 | 5.96E-03 | 4.55E-02 | 5 | 826 | EGR1,FOS,ATF3,DUSP1,NR4A1 |
| M1/Control | Down | GO: Biological Proces | GO:00713cellular re | 5.54E-04 | 6.40E-01 | 1.75E-02 | 1.34E-01 | 5 | 1196 | EGR1,FOS,ZFP36,DUSP1,CEBPB |
| M1/Control | Down | GO: Biological Proces | GO:00340response t | 9.61E-04 | 1.00E+00 | 1.98E-02 | 1.51E-01 | 5 | 1349 | EGR1,FOS,ZFP36,DUSP1,CEBPB |
| M1/Control | Down | GO: Biological Proces | GO:00459positive re | 1.24E-03 | 1.00E+00 | 2.13E-02 | 1.63E-01 | 5 | 1427 | EGR1,FOS,ATF3,CEBPB,NR4A1 |
| M1/Control | Down | GO: Biological Proces | GO:00458negative n | 1.54E-03 | 1.00E+00 | 2.41E-02 | 1.84E-01 | 5 | 1497 | EGR1,RASD1,ZFP36,ATF3,CEBPB |
| M1/Control | Down | GO: Biological Proces | GO:19035negative n | 1.55E-03 | 1.00E+00 | 2.41E-02 | 1.84E-01 | 5 | 1500 | EGR1,RASD1,ZFP36,ATF3,CEBPB |
| M1/Control | Down | GO: Biological Proces | GO:19026negative n | 1.57E-03 | 1.00E+00 | 2.41E-02 | 1.84E-01 | 5 | 1504 | EGR1,RASD1,ZFP36,ATF3,CEBPB |
| M1/Control | Down | GO: Biological Proces | GO:00300hemopoie | 1.59E-03 | 1.00E+00 | 2.41E-02 | 1.84E-01 | 5 | 1507 | EGR1,FOS,ZFP36,IGKC,CEBPB |
| M1/Control | Down | GO: Biological Proces | GO:00485hematopo | 1.88E-03 | 1.00E+00 | 2.56E-02 | 1.95E-01 | 5 | 1565 | EGR1,FOS,ZFP36,IGKC,CEBPB |
| M1/Control | Down | GO: Biological Proces | GO:00025immune s | 2.29E-03 | 1.00E+00 | 2.87E-02 | 2.19E-01 | 5 | 1636 | EGR1,FOS,ZFP36,IGKC,CEBPB |
| M1/Control | Down | GO: Biological Proces | GO:00512negative n | 2.35E-03 | 1.00E+00 | 2.87E-02 | 2.19E-01 | 5 | 1645 | EGR1,RASD1,ZFP36,ATF3,CEBPB |
| M1/Control | Down | GO: Biological Proces | GO:19017cellular re | 3.63E-03 | 1.00E+00 | 3.56E-02 | 2.72E-01 | 5 | 1815 | EGR1,FOS,ZFP36,CEBPB,NR4A1 |
| M1/Control | Down | GO: Biological Proces | GO:19035positive re | 4.43E-03 | 1.00E+00 | 3.73E-02 | 2.85E-01 | 5 | 1900 | EGR1,FOS,ATF3,CEBPB,NR4A1 |
| M1/Control | Down | GO: Biological Proces | GO:00458positive re | 4.43E-03 | 1.00E+00 | 3.73E-02 | 2.85E-01 | 5 | 1900 | EGR1,FOS,ATF3,CEBPB,NR4A1 |
| M1/Control | Down | GO: Biological Proces | GO:19026positive re | 4.49E-03 | 1.00E+00 | 3.73E-02 | 2.85E-01 | 5 | 1906 | EGR1,FOS,ATF3,CEBPB,NR4A1 |
| M1/Control | Down | GO: Biological Proces | GO:00508nervous sy | 4.88E-03 | 1.00E+00 | 3.81E-02 | 2.91E-01 | 5 | 1943 | EGR1,TAS1R3,FOS,CEBPB,NR4A1 |
| M1/Control | Down | GO: Cellular Compon | GO:00056transcripti | 4.99E-04 | 3.35E-02 | 1.49E-02 | 7.15E-02 | 4 | 590 | FOS,ATF3,CEBPB,NR4A1 |
| M1/M0 | Up | GO: Molecular Functi | GO:00051signaling r | 1.52E-04 | 3.19E-02 | 5.31E-03 | 3.14E-02 | 9 | 1813 | HSPG2,FLNA,CD81,CTNNB1,ECM1,ENG,ITGB1,VCAM1,MSN |
| M1/M0 | Up | GO: Molecular Functi | GO:00080protein C- | 2.00E-04 | 4.18E-02 | 5.97E-03 | 3.54E-02 | 4 | 247 | HSPG2,TCF4,CTNNB1,ECM1 |
| M1/M0 | Up | GO: Molecular Functi | GO:00300extracellul | 3.45E-04 | 7.21E-02 | 9.02E-03 | 5.34E-02 | 2 | 23 | HSPG2,BGN |
| M1/M0 | Up | GO: Biological Proces | GO:00400growth | 2.17E-04 | 4.61E-01 | 4.68E-03 | 3.86E-02 | 8 | 1528 | CD81,HEG1,CTNNB1,IGFBP7,ZFP36L1,ECM1,ITGB1,SGK1 |
| M1/M0 | Up | GO: Biological Proces | GO:00604mesenchy | 2.02E-04 | 4.31E-01 | 4.68E-03 | 3.86E-02 | 5 | 485 | FLNA,COL18A1,CTNNB1,ZFP36L1,ENG |
| M1/M0 | Up | GO: Biological Proces | GO:00715leukocyte | 2.16E-04 | 4.61E-01 | 4.68E-03 | 3.86E-02 | 5 | 492 | CD81,ITGB1,VCAM1,LRRC32,MSN |
| M1/M0 | Up | GO: Biological Proces | GO:00031outflow tr | 2.05E-04 | 4.37E-01 | 4.68E-03 | 3.86E-02 | 3 | 100 | GJA5,CTNNB1,ENG |
| M1/M0 | Up | GO: Biological Proces | GO:00488artery mor | 2.17E-04 | 4.63E-01 | 4.68E-03 | 3.86E-02 | 3 | 102 | GJA5,CTNNB1,ENG |
| M1/M0 | Up | GO: Biological Proces | GO:00860atrial card | 2.17E-04 | 4.63E-01 | 4.68E-03 | 3.86E-02 | 2 | 19 | FLNA,GJA5 |
| M1/M0 | Up | GO: Biological Proces | GO:00860atrial card | 2.17E-04 | 4.63E-01 | 4.68E-03 | 3.86E-02 | 2 | 19 | FLNA,GJA5 |
| M1/M0 | Up | GO: Biological Proces | GO:00860atrial card | 2.17E-04 | 4.63E-01 | 4.68E-03 | 3.86E-02 | 2 | 19 | FLNA,GJA5 |
| M1/M0 | Up | GO: Biological Proces | GO:00350cardiocyte | 2.37E-04 | 5.05E-01 | 5.00E-03 | 4.12E-02 | 4 | 268 | CTNNB1,ENG,ITGB1,VCAM1 |
| M1/M0 | Up | GO: Biological Proces | GO:00511smooth m | 2.37E-04 | 5.05E-01 | 5.00E-03 | 4.12E-02 | 3 | 105 | CTNNB1,RCAN1,ENG |
| M1/M0 | Up | GO: Biological Proces | GO:00032ventricular | 2.41E-04 | 5.14E-01 | 5.04E-03 | 4.15E-02 | 2 | 20 | HEG1,ENG |
| M1/M0 | Up | GO: Biological Proces | GO:00487tissue rem | 2.44E-04 | 5.20E-01 | 5.05E-03 | 4.16E-02 | 4 | 270 | FLNA,GJA5,CTNNB1,BGN |
| M1/M0 | Up | GO: Biological Proces | GO:00017morphoge | 2.65E-04 | 5.65E-01 | 5.38E-03 | 4.44E-02 | 4 | 276 | COL4A1,CTNNB1,IGFBP7,ENG |
| M1/M0 | Up | GO: Biological Proces | GO:00604muscle tis | 2.65E-04 | 5.64E-01 | 5.38E-03 | 4.44E-02 | 3 | 109 | HEG1,CTNNB1,ENG |
| M1/M0 | Up | GO: Biological Proces | GO:00485embryonic | 2.88E-04 | 6.13E-01 | 5.72E-03 | 4.71E-02 | 7 | 1185 | HSPG2,GJA5,CTNNB1,ZFP36L1,ENG,ITGB1,VCAM1 |
| M1/M0 | Up | GO: Biological Proces | GO:00096response t | 2.84E-04 | 6.06E-01 | 5.72E-03 | 4.71E-02 | 6 | 825 | FLNA,CD81,ZFP36L1,ENG,ITGB1,LRRC32 |
| M1/M0 | Up | GO: Biological Proces | GO:00506regulation | 2.90E-04 | 6.17E-01 | 5.72E-03 | 4.71E-02 | 5 | 524 | COL18A1,CTNNB1,ZFP36L1,ECM1,ENG |
| M1/M0 | Up | GO: Biological Proces | GO:00455positive re | 3.09E-04 | 6.59E-01 | 6.04E-03 | 4.98E-02 | 7 | 1199 | FLNA,TCF4,CTNNB1,ZFP36L1,ENG,ITGB1,SGK1 |
| M1/M0 | Up | GO: Biological Proces | GO:00102response t | 3.28E-04 | 6.99E-01 | 6.29E-03 | 5.19E-02 | 8 | 1623 | COL4A1,FLNA,CD81,CTNNB1,ZFP36L1,ITGB1,SGK1,VCAM1 |
| M1/M0 | Up | GO: Biological Proces | GO:00486muscle org | 3.26E-04 | 6.94E-01 | 6.29E-03 | 5.19E-02 | 3 | 117 | HEG1,CTNNB1,ENG |
| M1/M0 | Up | GO: Biological Proces | GO:00020sprouting | 3.37E-04 | 7.19E-01 | 6.42E-03 | 5.29E-02 | 4 | 294 | IGFBP7,RCAN1,ENG,ITGB1 |
| M1/M0 | Up | GO: Biological Proces | GO:20001regulation | 3.49E-04 | 7.45E-01 | 6.59E-03 | 5.43E-02 | 2 | 24 | CTNNB1,ENG |
| M1/M0 | Up | GO: Biological Proces | GO:00613cell prolife | 4.11E-04 | 8.76E-01 | 7.68E-03 | 6.33E-02 | 2 | 26 | CTNNB1,ENG |
| M1/M0 | Up | GO: Biological Proces | GO:00508regulation | 4.15E-04 | 8.83E-01 | 7.68E-03 | 6.33E-02 | 6 | 885 | FLNA,CD81,CTNNB1,ZFP36L1,VCAM1,LRRC32 |
| M1/M0 | Up | GO: Biological Proces | GO:20001regulation | 4.44E-04 | 9.45E-01 | 8.08E-03 | 6.66E-02 | 3 | 130 | FLNA,CTNNB1,ITGB1 |
| M1/M0 | Up | GO: Biological Proces | GO:00350negative n | 4.43E-04 | 9.45E-01 | 8.08E-03 | 6.66E-02 | 2 | 27 | HEG1,ITGB1 |
| M1/M0 | Up | GO: Biological Proces | GO:00453leukocyte | 4.54E-04 | 9.67E-01 | 8.20E-03 | 6.76E-02 | 7 | 1277 | CD81,CTNNB1,ZFP36L1,ITGB1,VCAM1,LRRC32,MSN |
| M1/M0 | Up | GO: Biological Proces | GO:00343cell junctic | 4.64E-04 | 9.90E-01 | 8.25E-03 | 6.80E-02 | 6 | 904 | COL4A1,FLNA,GJA5,HEG1,CTNNB1,ITGB1 |
| M1/M0 | Up | GO: Biological Proces | GO:00506regulation | 4.65E-04 | 9.90E-01 | 8.25E-03 | 6.80E-02 | 4 | 320 | CD81,CTNNB1,VCAM1,LRRC32 |
| M1/M0 | Up | GO: Biological Proces | GO:00485response t | 4.84E-04 | 1.00E+00 | 8.31E-03 | 6.85E-02 | 5 | 586 | IGFBP7,ZFP36L1,ENG,SGK1,PGRMC2 |
| M1/M0 | Up | GO: Biological Proces | GO:00605epithelial t | 4.88E-04 | 1.00E+00 | 8.31E-03 | 6.85E-02 | 5 | 587 | COL4A1,CTNNB1,IGFBP7,ENG,MSN |
| M1/M0 | Up | GO: Biological Proces | GO:00329regulation | 4.87E-04 | 1.00E+00 | 8.31E-03 | 6.85E-02 | 4 | 324 | CD81,CTNNB1,VCAM1,LRRC32 |
| M1/M0 | Up | GO: Biological Proces | GO:00032cardiac se | 4.85E-04 | 1.00E+00 | 8.31E-03 | 6.85E-02 | 3 | 134 | GJA5,HEG1,ENG |
| M1/M0 | Up | GO: Biological Proces | GO:00607labyrinthin | 4.77E-04 | 1.00E+00 | 8.31E-03 | 6.85E-02 | 2 | 28 | ZFP36L1,VCAM1 |
| M1/M0 | Up | GO: Biological Proces | GO:00098animal org | 5.04E-04 | 1.00E+00 | 8.52E-03 | 7.03E-02 | 8 | 1729 | HSPG2,FLNA,COL18A1,GJA5,HEG1,CTNNB1,ENG,MSN |
| M1/M0 | Up | GO: Biological Proces | GO:00714cellular re | 5.12E-04 | 1.00E+00 | 8.59E-03 | 7.08E-02 | 2 | 29 | CD81,ITGB1 |
| M1/M0 | Up | GO: Cellular Compon | GO:00163apical plas | 1.57E-04 | 3.63E-02 | 2.39E-03 | 1.44E-02 | 5 | 464 | CD81,CTNNB1,ENG,VCAM1,MSN |
| M1/M0 | Up | GO: Cellular Compon | GO:00451apical part | 4.04E-04 | 9.33E-02 | 4.24E-03 | 2.55E-02 | 5 | 569 | CD81,CTNNB1,ENG,VCAM1,MSN |
| M1/Control | Up | GO: Molecular Functi | GO:00700fibrinogen | 1.46E-04 | 6.33E-02 | 2.85E-03 | 1.90E-02 | 2 | 5 | THBS1,ITGB3 |
| M1/Control | Up | GO: Molecular Functi | GO:00055calcium io | 1.86E-04 | 8.05E-02 | 3.36E-03 | 2.23E-02 | 11 | 777 | S100A6,THBS1,MYL6,MYL9,SULF1,EFEMP2,LRP2,DAG1,MACF1,ACTN1,MYL12B |
| M1/Control | Up | GO: Molecular Functi | GO:00469protein dir | 2.07E-04 | 8.99E-02 | 3.60E-03 | 2.39E-02 | 14 | 1218 | TPM1,S100A6,BMP2,ATP5IF1,TCF21,NFE2L1,BST2,H4C5,PARK7,EFEMP2,ACTN1,ITGA3,APOE,ITGB1 |
| M1/Control | Up | GO: Molecular Functi | GO:00381neuregulin | 2.18E-04 | 9.47E-02 | 3.64E-03 | 2.42E-02 | 2 | 6 | ITGAV,ITGB3 |
| M1/Control | Up | GO: Molecular Functi | GO:00513alpha-acti | 2.53E-04 | 1.10E-01 | 4.06E-03 | 2.70E-02 | 3 | 32 | DAG1,ACTN1,ITGB1 |
| M1/Control | Up | GO: Molecular Functi | GO:00199chemokine | 3.03E-04 | 1.32E-01 | 4.70E-03 | 3.12E-02 | 3 | 34 | ITGAV,ITGB1,ITGB3 |
| M1/Control | Up | GO: Biological Proces | GO:00901regulation | 2.27E-03 | 1.00E+00 | 2.90E-02 | 2.51E-01 | 3 | 71 | THBS1,THY1,MACF1 |
| M1/Control | Up | GO: Biological Proces | GO:00611morphoge | 2.34E-03 | 1.00E+00 | 2.99E-02 | 2.58E-01 | 5 | 252 | BMP2,TCF21,IGFBP7,SULF1,DAG1 |
| M1/Control | Up | GO: Biological Proces | GO:00301negative n | 2.45E-03 | 1.00E+00 | 3.12E-02 | 2.69E-01 | 3 | 73 | THBS1,CD9,APOE |
| M1/Control | Up | GO: Biological Proces | GO:00305regulation | 2.49E-03 | 1.00E+00 | 3.15E-02 | 2.72E-01 | 4 | 154 | BMP2,SULF1,LRP2,ITGA3 |
| M1/Control | Up | GO: Biological Proces | GO:00460ATP metab | 2.51E-03 | 1.00E+00 | 3.16E-02 | 2.73E-01 | 5 | 256 | CFH,ATP5PF,ATP5IF1,ATP5F1A,NDUFS5 |
| M1/Control | Up | GO: Biological Proces | GO:00452sarcomere | 2.55E-03 | 1.00E+00 | 3.18E-02 | 2.75E-01 | 3 | 74 | TPM1,ACTN1,ITGB1 |
| M1/Control | Up | GO: Biological Proces | GO:00469regulation | 2.55E-03 | 1.00E+00 | 3.18E-02 | 2.75E-01 | 3 | 74 | ATP5IF1,SLC25A6,VDAC2 |
| M1/Control | Up | GO: Biological Proces | GO:19000negative n | 2.55E-03 | 1.00E+00 | 3.18E-02 | 2.75E-01 | 3 | 74 | THBS1,CD9,APOE |
| M1/Control | Up | GO: Biological Proces | GO:00510negative n | 2.61E-03 | 1.00E+00 | 3.20E-02 | 2.76E-01 | 12 | 1299 | SORT1,BMP2,THBS1,THY1,PTPRM,SULF1,H4C5,FRZB,APOE,ITGAV,ITGB1,ITGB3 |
| M1/Control | Up | GO: Biological Proces | GO:00713cellular re | 2.65E-03 | 1.00E+00 | 3.20E-02 | 2.76E-01 | 10 | 963 | SORT1,BMP2,MYOF,THBS1,SULF1,LRP2,ITGA3,APOE,ITGB1,ITGB3 |
| M1/Control | Up | GO: Biological Proces | GO:00487cardiac mu | 2.61E-03 | 1.00E+00 | 3.20E-02 | 2.76E-01 | 6 | 377 | TPM1,BMP2,MYH10,DSP,LRP2,ITGB1 |
| M1/Control | Up | GO: Biological Proces | GO:00190viral life cy | 2.65E-03 | 1.00E+00 | 3.20E-02 | 2.76E-01 | 6 | 378 | BST2,DAG1,APOE,ITGAV,ITGB1,ITGB3 |
| M1/Control | Up | GO: Biological Proces | GO:00971extrinsic a | 2.64E-03 | 1.00E+00 | 3.20E-02 | 2.76E-01 | 5 | 259 | SORT1,THBS1,PARK7,ITM2B,ITGAV |
| M1/Control | Up | GO: Biological Proces | GO:00107fibroblast | 2.65E-03 | 1.00E+00 | 3.20E-02 | 2.76E-01 | 3 | 75 | THBS1,ITGB1,ITGB3 |
| M1/Control | Up | GO: Biological Proces | GO:00159long-chain | 2.65E-03 | 1.00E+00 | 3.20E-02 | 2.76E-01 | 3 | 75 | ATP5PF,THBS1,APOE |
| M1/Control | Up | GO: Biological Proces | GO:00032ventricular | 2.65E-03 | 1.00E+00 | 3.20E-02 | 2.76E-01 | 3 | 75 | TPM1,DSP,LRP2 |
| M1/Control | Up | GO: Biological Proces | GO:00704leukocyte | 2.67E-03 | 1.00E+00 | 3.20E-02 | 2.76E-01 | 2 | 21 | CFH,THBS1 |
| M1/Control | Up | GO: Biological Proces | GO:190514lysosomal | 2.67E-03 | 1.00E+00 | 3.20E-02 | 2.76E-01 | 2 | 21 | LRP2,LAMP2 |
| M1/Control | Up | GO: Biological Proces | GO:01501regulation | 2.75E-03 | 1.00E+00 | 3.29E-02 | 2.84E-01 | 3 | 76 | THBS1,THY1,MACF1 |
| M1/Control | Up | GO: Biological Proces | GO:00508regulation | 2.77E-03 | 1.00E+00 | 3.30E-02 | 2.85E-01 | 7 | 514 | CFH,THBS1,MYL9,CD9,ACTN1,APOE,ITGB3 |
| M1/Control | Up | GO: Biological Proces | GO:00481fibroblast | 2.79E-03 | 1.00E+00 | 3.31E-02 | 2.86E-01 | 4 | 159 | S100A6,THY1,PRDX1,ITGB3 |
| M1/Control | Up | GO: Biological Proces | GO:00429regulation | 3.40E-03 | 1.00E+00 | 3.32E-02 | 2.87E-01 | 15 | 1899 | SORT1,BMP2,THBS1,THY1,SLC25A6,PARK7,FRZB,DAD1,LRP2,ACTN1,APOE,ITGAV,ITGB1,PTTG1IP,VDAC2 |
| M1/Control | Up | GO: Biological Proces | GO:00512negative n | 3.20E-03 | 1.00E+00 | 3.32E-02 | 2.87E-01 | 12 | 1332 | BMP2,THBS1,THY1,BST2,PARK7,LRP2,APLP2,APOE,ITM2B,ITGAV,ITGB3,ITIH5 |
| M1/Control | Up | GO: Biological Proces | GO:00605negative n | 3.76E-03 | 1.00E+00 | 3.32E-02 | 2.87E-01 | 12 | 1359 | THBS1,THY1,SLC25A6,PARK7,DAD1,LRP2,APOE,ITGAV,ITGB1,ITGB3,PTTG1IP,VDAC2 |
| M1/Control | Up | GO: Biological Proces | GO:00440regulation | 2.97E-03 | 1.00E+00 | 3.32E-02 | 2.87E-01 | 11 | 1146 | TPM1,THBS1,THY1,PARK7,DAG1,MACF1,LAMP2,CGNL1,APOE,CDC42EP3,VDAC2 |
| M1/Control | Up | GO: Biological Proces | GO:00430negative n | 3.41E-03 | 1.00E+00 | 3.32E-02 | 2.87E-01 | 11 | 1167 | THBS1,THY1,SLC25A6,PARK7,DAD1,LRP2,APOE,ITGAV,ITGB1,PTTG1IP,VDAC2 |
| M1/Control | Up | GO: Biological Proces | GO:00708response t | 3.62E-03 | 1.00E+00 | 3.32E-02 | 2.87E-01 | 10 | 1006 | SORT1,BMP2,MYOF,THBS1,SULF1,LRP2,ITGA3,APOE,ITGB1,ITGB3 |
| M1/Control | Up | GO: Biological Proces | GO:00091nucleotide | 3.54E-03 | 1.00E+00 | 3.32E-02 | 2.87E-01 | 8 | 684 | CFH,GLYAT,ATP5PF,ATP5IF1,PARK7,LDHB,ATP5F1A,NDUFS5 |
| M1/Control | Up | GO: Biological Proces | GO:00067nucleoside | 3.77E-03 | 1.00E+00 | 3.32E-02 | 2.87E-01 | 8 | 691 | CFH,GLYAT,ATP5PF,ATP5IF1,PARK7,LDHB,ATP5F1A,NDUFS5 |
| M1/Control | Up | GO: Biological Proces | GO:00310regenerati | 3.60E-03 | 1.00E+00 | 3.32E-02 | 2.87E-01 | 7 | 539 | KLK6,THY1,CD9,LRP2,DAG1,APOE,VDAC2 |
| M1/Control | Up | GO: Biological Proces | GO:00158organic ac | 2.97E-03 | 1.00E+00 | 3.32E-02 | 2.87E-01 | 6 | 387 | ATP5PF,THBS1,NFE2L1,LRP2,APOE,ITGB1 |
| M1/Control | Up | GO: Biological Proces | GO:00082steroid me | 3.01E-03 | 1.00E+00 | 3.32E-02 | 2.87E-01 | 6 | 388 | BMP2,IGFBP7,NFE2L1,LRP2,APLP2,APOE |
| M1/Control | Up | GO: Biological Proces | GO:00067sulfur com | 3.29E-03 | 1.00E+00 | 3.32E-02 | 2.87E-01 | 6 | 395 | GLYAT,NFE2L1,SULF1,PARK7,AKR1A1,GNS |
| M1/Control | Up | GO: Cellular Compon | GO:00452proton-tra | 2.94E-03 | 1.00E+00 | 1.63E-02 | 1.06E-01 | 2 | 22 | ATP5PF,ATP5F1A |
| M1/Control | Up | GO: Cellular Compon | GO:00057lysosome | 3.17E-03 | 1.00E+00 | 1.68E-02 | 1.10E-01 | 9 | 823 | SORT1,LAPTM4A,SLC12A4,BST2,LRP2,LAMP2,APOE,RAB2A,GNS |
| M1/Control | Up | GO: Cellular Compon | GO:00003lytic vacuo | 3.17E-03 | 1.00E+00 | 1.68E-02 | 1.10E-01 | 9 | 823 | SORT1,LAPTM4A,SLC12A4,BST2,LRP2,LAMP2,APOE,RAB2A,GNS |
| M1/Control | Up | GO: Cellular Compon | GO:00988inner mito | 3.15E-03 | 1.00E+00 | 1.68E-02 | 1.10E-01 | 4 | 164 | ATP5PF,SLC25A6,ATP5F1A,NDUFS5 |
| M1/Control | Up | GO: Cellular Compon | GO:00312lamellipod | 3.21E-03 | 1.00E+00 | 1.68E-02 | 1.10E-01 | 2 | 23 | ITGAV,ITGB3 |
| M1/Control | Up | GO: Cellular Compon | GO:00300myofibril | 3.56E-03 | 1.00E+00 | 1.84E-02 | 1.20E-01 | 5 | 277 | TPM1,MYL9,DAG1,ACTN1,MYL12B |
| M1/Control | Up | GO: Cellular Compon | GO:00304smooth m | 3.69E-03 | 1.00E+00 | 1.86E-02 | 1.21E-01 | 1 | 1 | ACTN1 |
| M1/Control | Up | GO: Cellular Compon | GO:00617chaperone | 3.69E-03 | 1.00E+00 | 1.86E-02 | 1.21E-01 | 1 | 1 | LAMP2 |
| M1/Control | Up | GO: Cellular Compon | GO:00990supramole | 4.05E-03 | 1.00E+00 | 2.01E-02 | 1.31E-01 | 13 | 1549 | TPM1,GLYAT,MYH10,NDRG1,MYL9,DSP,EFEMP2,GAS2,DAG1,MACF1,ACTN1,APOE,MYL12B |
| M1/Control | Up | GO: Cellular Compon | GO:00988actin-base | 4.14E-03 | 1.00E+00 | 2.03E-02 | 1.33E-01 | 5 | 287 | DAG1,ITGA3,ITGAV,ITGB1,ITGB3 |
| M1/Control | Up | GO: Cellular Compon | GO:00432contractile | 4.52E-03 | 1.00E+00 | 2.18E-02 | 1.43E-01 | 5 | 293 | TPM1,MYL9,DAG1,ACTN1,MYL12B |
| M1/Control | Up | GO: Cellular Compon | GO:00442cell-cell co | 5.05E-03 | 1.00E+00 | 2.41E-02 | 1.58E-01 | 3 | 94 | DSP,ACTN1,ITGB1 |
| M1/Control | Down | GO: Biological Proces | GO:00359skeletal m | 2.64E-07 | 3.05E-04 | 3.05E-04 | 2.33E-03 | 4 | 93 | EGR1,FOS,ATF3,NR4A1 |
| M1/Control | Down | GO: Biological Proces | GO:00515response t | 1.84E-06 | 2.13E-03 | 1.06E-03 | 8.12E-03 | 4 | 151 | EGR1,FOS,DUSP1,REN |
| M1/Control | Down | GO: Biological Proces | GO:00466response t | 6.09E-06 | 7.04E-03 | 1.38E-03 | 1.05E-02 | 4 | 204 | EGR1,FOS,DUSP1,REN |
| M1/Control | Down | GO: Biological Proces | GO:00140response t | 1.08E-05 | 1.25E-02 | 1.79E-03 | 1.36E-02 | 4 | 236 | EGR1,FOS,DUSP1,REN |
| M1/Control | Down | GO: Biological Proces | GO:00075skeletal m | 4.08E-05 | 4.72E-02 | 3.93E-03 | 3.00E-02 | 4 | 331 | EGR1,FOS,ATF3,NR4A1 |
| M1/Control | Down | GO: Biological Proces | GO:00605skeletal m | 5.19E-05 | 6.00E-02 | 4.11E-03 | 3.13E-02 | 4 | 352 | EGR1,FOS,ATF3,NR4A1 |
| M1/Control | Down | GO: Biological Proces | GO:00075muscle org | 3.19E-04 | 3.69E-01 | 1.54E-02 | 1.17E-01 | 4 | 564 | EGR1,FOS,ATF3,NR4A1 |
| M1/Control | Down | GO: Biological Proces | GO:00485response t | 3.69E-04 | 4.27E-01 | 1.64E-02 | 1.25E-01 | 4 | 586 | EGR1,FOS,ZFP36,DUSP1 |
| M1/Control | Down | GO: Biological Proces | GO:19031mononucl | 4.55E-04 | 5.26E-01 | 1.75E-02 | 1.34E-01 | 4 | 619 | EGR1,FOS,IGKC,CEBPB |
| M1/Control | Down | GO: Biological Proces | GO:00605muscle tis | 7.12E-04 | 8.23E-01 | 1.84E-02 | 1.41E-01 | 4 | 697 | EGR1,FOS,ATF3,NR4A1 |
| M1/Control | Down | GO: Biological Proces | GO:00430positive re | 8.34E-04 | 9.64E-01 | 1.90E-02 | 1.45E-01 | 4 | 727 | EGR1,ATF3,DUSP1,NR4A1 |
| M1/Control | Down | GO: Biological Proces | GO:00430positive re | 8.96E-04 | 1.00E+00 | 1.90E-02 | 1.45E-01 | 4 | 741 | EGR1,ATF3,DUSP1,NR4A1 |
| M1/Control | Down | GO: Biological Proces | GO:00316response t | 1.17E-03 | 1.00E+00 | 2.13E-02 | 1.63E-01 | 4 | 796 | EGR1,FOS,ZFP36,ATF3 |
| M1/Control | Down | GO: Biological Proces | GO:00025leukocyte | 1.28E-03 | 1.00E+00 | 2.15E-02 | 1.64E-01 | 4 | 816 | EGR1,FOS,IGKC,CEBPB |
| M1/Control | Down | GO: Biological Proces | GO:00099response t | 1.40E-03 | 1.00E+00 | 2.31E-02 | 1.76E-01 | 4 | 835 | EGR1,FOS,ZFP36,ATF3 |
| M1/Control | Down | GO: Biological Proces | GO:00430negative n | 2.07E-03 | 1.00E+00 | 2.73E-02 | 2.08E-01 | 4 | 929 | ZFP36,DUSP1,NR4A1,USP17L12 |
| M1/Control | Down | GO: Biological Proces | GO:00713cellular re | 2.27E-03 | 1.00E+00 | 2.87E-02 | 2.19E-01 | 4 | 952 | EGR1,FOS,ZFP36,CEBPB |
| M1/Control | Down | GO: Biological Proces | GO:00713cellular re | 2.37E-03 | 1.00E+00 | 2.87E-02 | 2.19E-01 | 4 | 963 | EGR1,FOS,ZFP36,NR4A1 |
| M1/Control | Down | GO: Biological Proces | GO:00714cellular re | 2.41E-03 | 1.00E+00 | 2.87E-02 | 2.19E-01 | 4 | 968 | EGR1,FOS,CEBPB,NR4A1 |
| M1/Control | Down | GO: Biological Proces | GO:00001MAPK cas | 2.47E-03 | 1.00E+00 | 2.87E-02 | 2.19E-01 | 4 | 974 | ZFP36,ATF3,DUSP1,REN |
| M1/Control | Down | GO: Biological Proces | GO:00708response t | 2.78E-03 | 1.00E+00 | 3.10E-02 | 2.36E-01 | 4 | 1006 | EGR1,FOS,ZFP36,NR4A1 |
| M1/Control | Down | GO: Biological Proces | GO:00076behavior | 2.79E-03 | 1.00E+00 | 3.10E-02 | 2.36E-01 | 4 | 1007 | EGR1,FOS,REN,CEBPB |
| M1/Control | Down | GO: Biological Proces | GO:00610muscle str | 3.18E-03 | 1.00E+00 | 3.27E-02 | 2.50E-01 | 4 | 1044 | EGR1,FOS,ATF3,NR4A1 |
| M1/Control | Down | GO: Biological Proces | GO:19016cellular re | 3.20E-03 | 1.00E+00 | 3.27E-02 | 2.50E-01 | 4 | 1046 | EGR1,FOS,CEBPB,NR4A1 |
| M1/Control | Down | GO: Biological Proces | GO:00001negative n | 4.14E-03 | 1.00E+00 | 3.73E-02 | 2.85E-01 | 4 | 1123 | EGR1,ZFP36,ATF3,CEBPB |
| M1/Control | Down | GO: Biological Proces | GO:00321regulation | 6.61E-03 | 1.00E+00 | 4.41E-02 | 3.37E-01 | 4 | 1280 | ZFP36,DUSP1,CEBPB,USP17L12 |
| M1/Control | Down | GO: Biological Proces | GO:00512negative n | 7.61E-03 | 1.00E+00 | 4.88E-02 | 3.73E-01 | 4 | 1332 | ZFP36,DUSP1,NR4A1,USP17L12 |
| M1/Control | Down | GO: Cellular Compon | GO:00905RNA polyr | 6.16E-04 | 4.13E-02 | 1.49E-02 | 7.15E-02 | 3 | 259 | FOS,ATF3,CEBPB |
| M1/M0 | Up | GO: Molecular Functi | GO:00504transformi | 4.77E-04 | 9.98E-02 | 1.11E-02 | 6.57E-02 | 2 | 27 | ENG,LRRC32 |
| M1/M0 | Up | GO: Molecular Functi | GO:00432laminin bi | 6.72E-04 | 1.41E-01 | 1.41E-02 | 8.32E-02 | 2 | 32 | ECM1,ITGB1 |
| M1/M0 | Up | GO: Biological Proces | GO:19016response t | 5.22E-04 | 1.00E+00 | 8.69E-03 | 7.16E-02 | 8 | 1738 | COL4A1,FLNA,CD81,CTNNB1,ZFP36L1,ITGB1,SGK1,VCAM1 |
| M1/M0 | Up | GO: Biological Proces | GO:00017in utero e | 5.39E-04 | 1.00E+00 | 8.85E-03 | 7.30E-02 | 5 | 600 | HEG1,CTNNB1,ZFP36L1,ITGB1,VCAM1 |
| M1/M0 | Up | GO: Biological Proces | GO:00996regulation | 5.48E-04 | 1.00E+00 | 8.85E-03 | 7.30E-02 | 2 | 30 | FLNA,GJA5 |
| M1/M0 | Up | GO: Biological Proces | GO:00215cell migrat | 5.48E-04 | 1.00E+00 | 8.85E-03 | 7.30E-02 | 2 | 30 | FLNA,ITGB1 |
| M1/M0 | Up | GO: Biological Proces | GO:00600pericardiu | 5.48E-04 | 1.00E+00 | 8.85E-03 | 7.30E-02 | 2 | 30 | HEG1,VCAM1 |
| M1/M0 | Up | GO: Biological Proces | GO:00712cellular re | 5.74E-04 | 1.00E+00 | 9.20E-03 | 7.58E-02 | 3 | 142 | CTNNB1,ENG,ITGB1 |
| M1/M0 | Up | GO: Biological Proces | GO:00512negative n | 5.86E-04 | 1.00E+00 | 9.31E-03 | 7.68E-02 | 7 | 1332 | HSPG2,FLNA,HEG1,CTNNB1,ZFP36L1,ECM1,ENG |
| M1/M0 | Up | GO: Biological Proces | GO:00510positive re | 5.95E-04 | 1.00E+00 | 9.37E-03 | 7.72E-02 | 8 | 1772 | FLNA,TCF4,CTNNB1,ZFP36L1,ECM1,ENG,ITGB1,SGK1 |
| M1/M0 | Up | GO: Biological Proces | GO:00456negative n | 5.98E-04 | 1.00E+00 | 9.37E-03 | 7.72E-02 | 3 | 144 | CTNNB1,ZFP36L1,H4C5 |
| M1/M0 | Up | GO: Biological Proces | GO:00454myoblast d | 6.10E-04 | 1.00E+00 | 9.49E-03 | 7.82E-02 | 3 | 145 | CTNNB1,ZFP36L1,ITGB1 |
| M1/M0 | Up | GO: Biological Proces | GO:00715cellular re | 6.23E-04 | 1.00E+00 | 9.63E-03 | 7.93E-02 | 4 | 346 | ZFP36L1,ENG,ITGB1,LRRC32 |
| M1/M0 | Up | GO: Biological Proces | GO:00714cellular re | 6.67E-04 | 1.00E+00 | 9.69E-03 | 7.99E-02 | 6 | 968 | COL4A1,FLNA,CTNNB1,ZFP36L1,SGK1,VCAM1 |
| M1/M0 | Up | GO: Biological Proces | GO:00706regulation | 6.65E-04 | 1.00E+00 | 9.69E-03 | 7.99E-02 | 4 | 352 | CD81,CTNNB1,VCAM1,LRRC32 |
| M1/M0 | Up | GO: Biological Proces | GO:00319response t | 6.72E-04 | 1.00E+00 | 9.69E-03 | 7.99E-02 | 4 | 353 | IGFBP7,ZFP36L1,ENG,SGK1 |
| M1/M0 | Up | GO: Biological Proces | GO:00020chondrocy | 6.35E-04 | 1.00E+00 | 9.69E-03 | 7.99E-02 | 3 | 147 | HSPG2,CTNNB1,ECM1 |
| M1/M0 | Up | GO: Biological Proces | GO:00074endoderm | 6.47E-04 | 1.00E+00 | 9.69E-03 | 7.99E-02 | 3 | 148 | CTNNB1,ZFP36L1,MSN |
| M1/M0 | Up | GO: Biological Proces | GO:00608artery dev | 6.73E-04 | 1.00E+00 | 9.69E-03 | 7.99E-02 | 3 | 150 | GJA5,CTNNB1,ENG |
| M1/M0 | Up | GO: Biological Proces | GO:00031morphoge | 6.64E-04 | 1.00E+00 | 9.69E-03 | 7.99E-02 | 2 | 33 | CTNNB1,MSN |
| M1/M0 | Up | GO: Biological Proces | GO:00611endothelia | 6.64E-04 | 1.00E+00 | 9.69E-03 | 7.99E-02 | 2 | 33 | CTNNB1,MSN |
| M1/M0 | Up | GO: Biological Proces | GO:00606embryonic | 6.64E-04 | 1.00E+00 | 9.69E-03 | 7.99E-02 | 2 | 33 | ZFP36L1,VCAM1 |
| M1/M0 | Up | GO: Biological Proces | GO:00600Sertoli cell | 6.64E-04 | 1.00E+00 | 9.69E-03 | 7.99E-02 | 2 | 33 | FLNA,ITGB1 |
| M1/M0 | Up | GO: Biological Proces | GO:00715response t | 6.79E-04 | 1.00E+00 | 9.71E-03 | 8.01E-02 | 4 | 354 | ZFP36L1,ENG,ITGB1,LRRC32 |
| M1/M0 | Up | GO: Biological Proces | GO:19017cellular re | 6.99E-04 | 1.00E+00 | 9.92E-03 | 8.18E-02 | 8 | 1815 | COL4A1,FLNA,CTNNB1,ZFP36L1,ITGB1,SGK1,VCAM1,MSN |
| M1/M0 | Up | GO: Biological Proces | GO:00020positive re | 7.05E-04 | 1.00E+00 | 9.95E-03 | 8.20E-02 | 2 | 34 | CTNNB1,ITGB1 |
| M1/M0 | Up | GO: Biological Proces | GO:00026regulation | 7.14E-04 | 1.00E+00 | 1.00E-02 | 8.25E-02 | 8 | 1821 | CD81,CTNNB1,ZFP36L1,ECM1,H4C5,VCAM1,LRRC32,MSN |
| M1/M0 | Up | GO: Biological Proces | GO:00082negative n | 7.47E-04 | 1.00E+00 | 1.03E-02 | 8.47E-02 | 6 | 989 | COL18A1,CTNNB1,IGFBP7,ENG,ITGB1,LRRC32 |
| M1/M0 | Up | GO: Biological Proces | GO:00512cartilage d | 7.46E-04 | 1.00E+00 | 1.03E-02 | 8.47E-02 | 4 | 363 | HSPG2,CTNNB1,ECM1,BGN |
| M1/M0 | Up | GO: Biological Proces | GO:00608venous bl | 7.47E-04 | 1.00E+00 | 1.03E-02 | 8.47E-02 | 2 | 35 | HEG1,ENG |
| M1/M0 | Up | GO: Biological Proces | GO:00509leukocyte | 7.91E-04 | 1.00E+00 | 1.08E-02 | 8.90E-02 | 2 | 36 | ITGB1,VCAM1 |
| M1/M0 | Up | GO: Biological Proces | GO:00485embryonic | 8.13E-04 | 1.00E+00 | 1.10E-02 | 9.09E-02 | 6 | 1005 | HSPG2,GJA5,CTNNB1,ZFP36L1,ENG,VCAM1 |
| M1/M0 | Up | GO: Biological Proces | GO:00426muscle cel | 8.19E-04 | 1.00E+00 | 1.10E-02 | 9.10E-02 | 5 | 658 | CD81,CTNNB1,RCAN1,ENG,ITGB1 |
| M1/M0 | Up | GO: Biological Proces | GO:00860cell-cell si | 8.35E-04 | 1.00E+00 | 1.12E-02 | 9.23E-02 | 2 | 37 | FLNA,GJA5 |
| M1/M0 | Up | GO: Biological Proces | GO:00487mesenchy | 8.51E-04 | 1.00E+00 | 1.13E-02 | 9.34E-02 | 4 | 376 | FLNA,COL18A1,CTNNB1,ENG |
| M1/M0 | Up | GO: Biological Proces | GO:00300myeloid ce | 8.59E-04 | 1.00E+00 | 1.14E-02 | 9.37E-02 | 5 | 665 | FLNA,CD81,CTNNB1,ZFP36L1,H4C5 |
| M1/M0 | Up | GO: Biological Proces | GO:00028regulation | 8.81E-04 | 1.00E+00 | 1.16E-02 | 9.55E-02 | 2 | 38 | CD81,ECM1 |
| M1/M0 | Up | GO: Biological Proces | GO:00096response t | 9.75E-04 | 1.00E+00 | 1.27E-02 | 1.05E-01 | 4 | 390 | CTNNB1,RCAN1,ENG,ITGB1 |
| M1/M0 | Up | GO: Biological Proces | GO:00603regulation | 9.76E-04 | 1.00E+00 | 1.27E-02 | 1.05E-01 | 2 | 40 | FLNA,GJA5 |
| M1/M0 | Up | GO: Biological Proces | GO:19016cellular re | 1.00E-03 | 1.00E+00 | 1.29E-02 | 1.07E-01 | 6 | 1046 | COL4A1,FLNA,CTNNB1,ZFP36L1,SGK1,VCAM1 |
| M1/M0 | Up | GO: Biological Proces | GO:00069inflammat | 1.01E-03 | 1.00E+00 | 1.30E-02 | 1.07E-01 | 6 | 1048 | HSPG2,CD81,ECM1,ITGB1,SGK1,VCAM1 |
| M1/M0 | Up | GO: Biological Proces | GO:00550response t | 1.03E-03 | 1.00E+00 | 1.31E-02 | 1.08E-01 | 2 | 41 | CD81,ITGB1 |
| M1/M0 | Up | GO: Biological Proces | GO:00300metal ion | 1.04E-03 | 1.00E+00 | 1.32E-02 | 1.09E-01 | 6 | 1053 | FLNA,GJA5,CTNNB1,ITGB1,SGK1,PGRMC2 |
| M1/M0 | Up | GO: Biological Proces | GO:00512regulation | 1.08E-03 | 1.00E+00 | 1.35E-02 | 1.11E-01 | 5 | 700 | CD81,CTNNB1,ZFP36L1,VCAM1,LRRC32 |
| M1/M0 | Up | GO: Biological Proces | GO:00160cell growt | 1.08E-03 | 1.00E+00 | 1.35E-02 | 1.11E-01 | 5 | 700 | CD81,CTNNB1,IGFBP7,ITGB1,SGK1 |
| M1/M0 | Up | GO: Biological Proces | GO:00996cardiac mu | 1.08E-03 | 1.00E+00 | 1.35E-02 | 1.11E-01 | 2 | 42 | FLNA,GJA5 |
| M1/M0 | Up | GO: Biological Proces | GO:00016ameboidal | 1.14E-03 | 1.00E+00 | 1.35E-02 | 1.11E-01 | 5 | 709 | HSPG2,COL18A1,GJA5,ENG,ITGB1 |
| M1/M0 | Up | GO: Biological Proces | GO:00600Sertoli cell | 1.13E-03 | 1.00E+00 | 1.35E-02 | 1.11E-01 | 2 | 43 | FLNA,ITGB1 |
| M1/M0 | Up | GO: Biological Proces | GO:19056positive re | 1.16E-03 | 1.00E+00 | 1.35E-02 | 1.11E-01 | 1 | 1 | CD81 |
| M1/M0 | Up | GO: Biological Proces | GO:00431regulation | 1.16E-03 | 1.00E+00 | 1.35E-02 | 1.11E-01 | 1 | 1 | CD81 |
| M1/M0 | Up | GO: Biological Proces | GO:00431positive re | 1.16E-03 | 1.00E+00 | 1.35E-02 | 1.11E-01 | 1 | 1 | CD81 |
| M1/M0 | Up | GO: Biological Proces | GO:19900vasomotio | 1.16E-03 | 1.00E+00 | 1.35E-02 | 1.11E-01 | 1 | 1 | GJA5 |
| M1/M0 | Up | GO: Biological Proces | GO:00860Purkinje m | 1.16E-03 | 1.00E+00 | 1.35E-02 | 1.11E-01 | 1 | 1 | GJA5 |
| M1/M0 | Up | GO: Cellular Compon | GO:00056basement | 1.46E-05 | 3.36E-03 | 3.50E-04 | 2.11E-03 | 4 | 132 | COL4A1,HSPG2,COL18A1,ITGB1 |
| M1/M0 | Up | GO: Cellular Compon | GO:00423sarcolemm | 6.06E-05 | 1.40E-02 | 1.17E-03 | 7.03E-03 | 4 | 190 | ITGB1,VCAM1,BGN,MSN |
| M1/M0 | Up | GO: Cellular Compon | GO:00988actin-base | 2.96E-04 | 6.84E-02 | 3.60E-03 | 2.17E-02 | 4 | 287 | CTNNB1,ITGB1,VCAM1,MSN |
| M1/M0 | Up | GO: Cellular Compon | GO:00098external si | 4.47E-03 | 1.00E+00 | 2.35E-02 | 1.41E-01 | 4 | 599 | HEG1,ENG,ITGB1,VCAM1 |
| M1/Control | Up | GO: Molecular Functi | GO:00510actin filam | 3.30E-04 | 1.43E-01 | 4.94E-03 | 3.28E-02 | 6 | 240 | TPM1,FMNL3,MYH10,GAS2,MACF1,ACTN1 |
| M1/Control | Up | GO: Molecular Functi | GO:00199cytokine b | 3.97E-04 | 1.72E-01 | 5.49E-03 | 3.65E-02 | 5 | 161 | THBS1,PARK7,ITGAV,ITGB1,ITGB3 |
| M1/Control | Up | GO: Molecular Functi | GO:01408thioredoxi | 4.05E-04 | 1.76E-01 | 5.49E-03 | 3.65E-02 | 2 | 8 | SELENOF,PRDX1 |
| M1/Control | Up | GO: Molecular Functi | GO:00083thioredoxi | 4.05E-04 | 1.76E-01 | 5.49E-03 | 3.65E-02 | 2 | 8 | SELENOF,PRDX1 |
| M1/Control | Up | GO: Molecular Functi | GO:00016virus recep | 5.40E-04 | 2.35E-01 | 7.05E-03 | 4.69E-02 | 4 | 97 | DAG1,ITGAV,ITGB1,ITGB3 |
| M1/Control | Up | GO: Molecular Functi | GO:01402exogenous | 5.62E-04 | 2.44E-01 | 7.05E-03 | 4.69E-02 | 4 | 98 | DAG1,ITGAV,ITGB1,ITGB3 |
| M1/Control | Up | GO: Molecular Functi | GO:00428actinin bin | 5.68E-04 | 2.47E-01 | 7.05E-03 | 4.69E-02 | 3 | 42 | DAG1,ACTN1,ITGB1 |
| M1/Control | Up | GO: Molecular Functi | GO:00162antioxidan | 6.78E-04 | 2.94E-01 | 8.17E-03 | 5.43E-02 | 4 | 103 | PARK7,SELENOF,PRDX1,APOE |
| M1/Control | Up | GO: Molecular Functi | GO:00199protein dc | 7.14E-04 | 3.10E-01 | 8.37E-03 | 5.57E-02 | 11 | 911 | TPM1,BMP2,NFE2L1,H4C5,LRP2,DAG1,LAMP2,ACTN1,ITGA3,ITGB1,SH3BGRL2 |
| M1/Control | Up | GO: Molecular Functi | GO:00082heparin bi | 9.89E-04 | 4.29E-01 | 1.13E-02 | 7.51E-02 | 5 | 197 | CFH,THBS1,EFEMP2,APLP2,APOE |
| M1/Control | Up | GO: Molecular Functi | GO:00433proteoglyc | 1.13E-03 | 4.88E-01 | 1.22E-02 | 8.12E-02 | 3 | 53 | CFH,THBS1,APOE |
| M1/Control | Up | GO: Molecular Functi | GO:00171vinculin bi | 1.11E-03 | 4.84E-01 | 1.22E-02 | 8.12E-02 | 2 | 13 | DAG1,ACTN1 |
| M1/Control | Up | GO: Molecular Functi | GO:19016sulfur com | 1.70E-03 | 7.38E-01 | 1.80E-02 | 1.20E-01 | 6 | 329 | CFH,THBS1,EFEMP2,APLP2,APOE,GNS |
| M1/Control | Up | GO: Molecular Functi | GO:00150coreceptor | 1.77E-03 | 7.69E-01 | 1.83E-02 | 1.22E-01 | 3 | 62 | ITGAV,ITGB1,ITGB3 |
| M1/Control | Up | GO: Molecular Functi | GO:00084sulfuric es | 1.92E-03 | 8.35E-01 | 1.90E-02 | 1.26E-01 | 2 | 17 | SULF1,GNS |
| M1/Control | Up | GO: Biological Proces | GO:00018tissue hor | 3.29E-03 | 1.00E+00 | 3.32E-02 | 2.87E-01 | 6 | 395 | SULF1,CDIPT,PRDX1,ITGAV,ITGB1,ITGB3 |
| M1/Control | Up | GO: Biological Proces | GO:00147striated m | 3.45E-03 | 1.00E+00 | 3.32E-02 | 2.87E-01 | 6 | 399 | TPM1,BMP2,MYH10,DSP,LRP2,ITGB1 |
| M1/Control | Up | GO: Biological Proces | GO:00072Ras protei | 3.58E-03 | 1.00E+00 | 3.32E-02 | 2.87E-01 | 6 | 402 | PARK7,ITGA3,APOE,RAB2A,ITGB1,CDC42EP3 |
| M1/Control | Up | GO: Biological Proces | GO:00301regulation | 3.76E-03 | 1.00E+00 | 3.32E-02 | 2.87E-01 | 6 | 406 | BMP2,SULF1,FRZB,MACF1,ITGA3,APOE |
| M1/Control | Up | GO: Biological Proces | GO:00605developm | 3.76E-03 | 1.00E+00 | 3.32E-02 | 2.87E-01 | 6 | 406 | THSD7A,MACF1,APOE,ITGAV,ITGB1,PLXNB1 |
| M1/Control | Up | GO: Biological Proces | GO:19901neuron pr | 2.82E-03 | 1.00E+00 | 3.32E-02 | 2.87E-01 | 5 | 263 | THSD7A,MACF1,APOE,ITGB1,PLXNB1 |
| M1/Control | Up | GO: Biological Proces | GO:00434protein kir | 3.10E-03 | 1.00E+00 | 3.32E-02 | 2.87E-01 | 5 | 269 | THBS1,PARK7,LRP2,DAG1,ITGB1 |
| M1/Control | Up | GO: Biological Proces | GO:00516maintenan | 3.10E-03 | 1.00E+00 | 3.32E-02 | 2.87E-01 | 5 | 269 | THY1,PARK7,APOE,ITGAV,ITGB3 |
| M1/Control | Up | GO: Biological Proces | GO:00017morphoge | 3.46E-03 | 1.00E+00 | 3.32E-02 | 2.87E-01 | 5 | 276 | BMP2,TCF21,IGFBP7,SULF1,DAG1 |
| M1/Control | Up | GO: Biological Proces | GO:00105regulation | 3.73E-03 | 1.00E+00 | 3.32E-02 | 2.87E-01 | 5 | 281 | THBS1,PTPRM,APOE,ITGB3,ATP5F1A |
| M1/Control | Up | GO: Biological Proces | GO:00072Rho prote | 2.98E-03 | 1.00E+00 | 3.32E-02 | 2.87E-01 | 4 | 162 | ITGA3,APOE,ITGB1,CDC42EP3 |
| M1/Control | Up | GO: Biological Proces | GO:00148response t | 3.19E-03 | 1.00E+00 | 3.32E-02 | 2.87E-01 | 4 | 165 | ATP5PF,ITGB1,ITGB3,ATP5F1A |
| M1/Control | Up | GO: Biological Proces | GO:00061oxidative p | 3.70E-03 | 1.00E+00 | 3.32E-02 | 2.87E-01 | 4 | 172 | ATP5PF,PARK7,ATP5F1A,NDUFS5 |
| M1/Control | Up | GO: Biological Proces | GO:00610regulation | 3.77E-03 | 1.00E+00 | 3.32E-02 | 2.87E-01 | 4 | 173 | THBS1,CD9,APOE,ITGB1 |
| M1/Control | Up | GO: Biological Proces | GO:00508negative n | 2.86E-03 | 1.00E+00 | 3.32E-02 | 2.87E-01 | 3 | 77 | THBS1,CD9,APOE |
| M1/Control | Up | GO: Biological Proces | GO:00159proton mo | 2.96E-03 | 1.00E+00 | 3.32E-02 | 2.87E-01 | 3 | 78 | ATP5PF,ATP5F1A,NDUFS5 |
| M1/Control | Up | GO: Biological Proces | GO:00480vascular e | 3.07E-03 | 1.00E+00 | 3.32E-02 | 2.87E-01 | 3 | 79 | MYOF,SULF1,ITGB3 |
| M1/Control | Up | GO: Biological Proces | GO:00515regulation | 2.93E-03 | 1.00E+00 | 3.32E-02 | 2.87E-01 | 2 | 22 | ITGB1,ITGB3 |
| M1/Control | Up | GO: Biological Proces | GO:00108negative n | 3.48E-03 | 1.00E+00 | 3.32E-02 | 2.87E-01 | 2 | 24 | ITGAV,ITGB3 |
| M1/Control | Up | GO: Biological Proces | GO:00550ventricular | 3.48E-03 | 1.00E+00 | 3.32E-02 | 2.87E-01 | 2 | 24 | BMP2,MYH10 |
| M1/Control | Up | GO: Biological Proces | GO:19026tertiary alc | 3.77E-03 | 1.00E+00 | 3.32E-02 | 2.87E-01 | 2 | 25 | BMP2,AKR1A1 |
| M1/Control | Up | GO: Biological Proces | GO:00060lactate me | 3.77E-03 | 1.00E+00 | 3.32E-02 | 2.87E-01 | 2 | 25 | PARK7,LDHB |
| M1/Control | Up | GO: Biological Proces | GO:00364cellular re | 3.68E-03 | 1.00E+00 | 3.32E-02 | 2.87E-01 | 1 | 1 | PARK7 |
| M1/Control | Up | GO: Biological Proces | GO:00703negative n | 3.68E-03 | 1.00E+00 | 3.32E-02 | 2.87E-01 | 1 | 1 | FRZB |
| M1/Control | Up | GO: Biological Proces | GO:01060guanine d | 3.68E-03 | 1.00E+00 | 3.32E-02 | 2.87E-01 | 1 | 1 | PARK7 |
| M1/Control | Up | GO: Biological Proces | GO:01060guanine d | 3.68E-03 | 1.00E+00 | 3.32E-02 | 2.87E-01 | 1 | 1 | PARK7 |
| M1/Control | Up | GO: Biological Proces | GO:01060guanine d | 3.68E-03 | 1.00E+00 | 3.32E-02 | 2.87E-01 | 1 | 1 | PARK7 |
| M1/Control | Up | GO: Biological Proces | GO:19030regulation | 3.68E-03 | 1.00E+00 | 3.32E-02 | 2.87E-01 | 1 | 1 | APOE |
| M1/Control | Up | GO: Biological Proces | GO:19030negative n | 3.68E-03 | 1.00E+00 | 3.32E-02 | 2.87E-01 | 1 | 1 | APOE |
| M1/Control | Up | GO: Biological Proces | GO:19030positive re | 3.68E-03 | 1.00E+00 | 3.32E-02 | 2.87E-01 | 1 | 1 | APOE |
| M1/Control | Up | GO: Biological Proces | GO:01502negative n | 3.68E-03 | 1.00E+00 | 3.32E-02 | 2.87E-01 | 1 | 1 | APOE |
| M1/Control | Up | GO: Biological Proces | GO:01502regulation | 3.68E-03 | 1.00E+00 | 3.32E-02 | 2.87E-01 | 1 | 1 | APOE |
| M1/Control | Up | GO: Biological Proces | GO:01502positive re | 3.68E-03 | 1.00E+00 | 3.32E-02 | 2.87E-01 | 1 | 1 | APOE |
| M1/Control | Up | GO: Biological Proces | GO:19007positive re | 3.68E-03 | 1.00E+00 | 3.32E-02 | 2.87E-01 | 1 | 1 | ITGB3 |
| M1/Control | Up | GO: Biological Proces | GO:19007regulation | 3.68E-03 | 1.00E+00 | 3.32E-02 | 2.87E-01 | 1 | 1 | ITGB3 |
| M1/Control | Up | GO: Biological Proces | GO:00365protein de | 3.68E-03 | 1.00E+00 | 3.32E-02 | 2.87E-01 | 1 | 1 | PARK7 |
| M1/Control | Up | GO: Biological Proces | GO:00365glutathion | 3.68E-03 | 1.00E+00 | 3.32E-02 | 2.87E-01 | 1 | 1 | PARK7 |
| M1/Control | Up | GO: Biological Proces | GO:00365peptidyl-c | 3.68E-03 | 1.00E+00 | 3.32E-02 | 2.87E-01 | 1 | 1 | PARK7 |
| M1/Control | Up | GO: Biological Proces | GO:00365peptidyl-a | 3.68E-03 | 1.00E+00 | 3.32E-02 | 2.87E-01 | 1 | 1 | PARK7 |
| M1/Control | Up | GO: Biological Proces | GO:00365peptidyl-ly | 3.68E-03 | 1.00E+00 | 3.32E-02 | 2.87E-01 | 1 | 1 | PARK7 |
| M1/Control | Up | GO: Biological Proces | GO:00365protein de | 3.68E-03 | 1.00E+00 | 3.32E-02 | 2.87E-01 | 1 | 1 | PARK7 |
| M1/Control | Up | GO: Biological Proces | GO:20002positive re | 3.68E-03 | 1.00E+00 | 3.32E-02 | 2.87E-01 | 1 | 1 | PARK7 |
| M1/Control | Up | GO: Biological Proces | GO:19017benzoyl-C | 3.68E-03 | 1.00E+00 | 3.32E-02 | 2.87E-01 | 1 | 1 | GLYAT |
| M1/Control | Up | GO: Biological Proces | GO:00907cellular re | 3.68E-03 | 1.00E+00 | 3.32E-02 | 2.87E-01 | 1 | 1 | APOE |
| M1/Control | Up | GO: Biological Proces | GO:00608positive re | 3.68E-03 | 1.00E+00 | 3.32E-02 | 2.87E-01 | 1 | 1 | BMP2 |
| M1/Control | Up | GO: Biological Proces | GO:00082biolumine | 3.68E-03 | 1.00E+00 | 3.32E-02 | 2.87E-01 | 1 | 1 | SETD3 |
| M1/Control | Up | GO: Biological Proces | GO:00510negative n | 3.68E-03 | 1.00E+00 | 3.32E-02 | 2.87E-01 | 1 | 1 | BMP2 |
| M1/Control | Up | GO: Biological Proces | GO:19029negative n | 3.68E-03 | 1.00E+00 | 3.32E-02 | 2.87E-01 | 1 | 1 | APOE |
| M1/Control | Up | GO: Biological Proces | GO:00462glycolate b | 3.68E-03 | 1.00E+00 | 3.32E-02 | 2.87E-01 | 1 | 1 | PARK7 |
| M1/Control | Up | GO: Biological Proces | GO:00850entry into | 3.68E-03 | 1.00E+00 | 3.32E-02 | 2.87E-01 | 1 | 1 | ITGAV |
| M1/Control | Up | GO: Biological Proces | GO:19016negative n | 3.68E-03 | 1.00E+00 | 3.32E-02 | 2.87E-01 | 1 | 1 | APOE |
| M1/Control | Up | GO: Biological Proces | GO:20004positive re | 3.68E-03 | 1.00E+00 | 3.32E-02 | 2.87E-01 | 1 | 1 | ITGB3 |
| M1/Control | Up | GO: Biological Proces | GO:00428D-glucuro | 3.68E-03 | 1.00E+00 | 3.32E-02 | 2.87E-01 | 1 | 1 | AKR1A1 |
| M1/Control | Up | GO: Biological Proces | GO:00428D-glucuro | 3.68E-03 | 1.00E+00 | 3.32E-02 | 2.87E-01 | 1 | 1 | AKR1A1 |
| M1/Control | Up | GO: Biological Proces | GO:19033regulation | 3.68E-03 | 1.00E+00 | 3.32E-02 | 2.87E-01 | 1 | 1 | NFE2L1 |
| M1/Control | Up | GO: Biological Proces | GO:00027regulation | 3.68E-03 | 1.00E+00 | 3.32E-02 | 2.87E-01 | 1 | 1 | BST2 |
| M1/Control | Up | GO: Biological Proces | GO:00027negative n | 3.68E-03 | 1.00E+00 | 3.32E-02 | 2.87E-01 | 1 | 1 | BST2 |
| M1/Control | Up | GO: Biological Proces | GO:19058regulation | 3.68E-03 | 1.00E+00 | 3.32E-02 | 2.87E-01 | 1 | 1 | APOE |
| M1/Control | Up | GO: Biological Proces | GO:19058positive re | 3.68E-03 | 1.00E+00 | 3.32E-02 | 2.87E-01 | 1 | 1 | APOE |
| M1/Control | Up | GO: Biological Proces | GO:19058regulation | 3.68E-03 | 1.00E+00 | 3.32E-02 | 2.87E-01 | 1 | 1 | APOE |
| M1/Control | Up | GO: Biological Proces | GO:19058positive re | 3.68E-03 | 1.00E+00 | 3.32E-02 | 2.87E-01 | 1 | 1 | APOE |
| M1/Control | Up | GO: Biological Proces | GO:19032positive re | 3.68E-03 | 1.00E+00 | 3.32E-02 | 2.87E-01 | 1 | 1 | PARK7 |
| M1/Control | Up | GO: Biological Proces | GO:19058regulation | 3.68E-03 | 1.00E+00 | 3.32E-02 | 2.87E-01 | 1 | 1 | APOE |
| M1/Control | Up | GO: Biological Proces | GO:00718intermedia | 3.68E-03 | 1.00E+00 | 3.32E-02 | 2.87E-01 | 1 | 1 | APOE |
| M1/Control | Up | GO: Biological Proces | GO:19031regulation | 3.68E-03 | 1.00E+00 | 3.32E-02 | 2.87E-01 | 1 | 1 | PARK7 |
| M1/Control | Up | GO: Biological Proces | GO:19031positive re | 3.68E-03 | 1.00E+00 | 3.32E-02 | 2.87E-01 | 1 | 1 | PARK7 |
| M1/Control | Up | GO: Biological Proces | GO:19031positive re | 3.68E-03 | 1.00E+00 | 3.32E-02 | 2.87E-01 | 1 | 1 | PARK7 |
| M1/Control | Up | GO: Biological Proces | GO:19031regulation | 3.68E-03 | 1.00E+00 | 3.32E-02 | 2.87E-01 | 1 | 1 | PARK7 |
| M1/Control | Up | GO: Biological Proces | GO:19031positive re | 3.68E-03 | 1.00E+00 | 3.32E-02 | 2.87E-01 | 1 | 1 | PARK7 |
| M1/Control | Up | GO: Biological Proces | GO:19031regulation | 3.68E-03 | 1.00E+00 | 3.32E-02 | 2.87E-01 | 1 | 1 | PARK7 |
| M1/Control | Up | GO: Biological Proces | GO:19031regulation | 3.68E-03 | 1.00E+00 | 3.32E-02 | 2.87E-01 | 1 | 1 | PARK7 |
| M1/Control | Up | GO: Cellular Compon | GO:00319organelle | 6.09E-03 | 1.00E+00 | 2.66E-02 | 1.74E-01 | 12 | 1439 | SORT1,S100A6,ATP5PF,MYOF,SLC25A6,PARK7,APOE,LDHB,ATP5F1A,SH3BGRL2,NDUFS5,VDAC2 |
| M1/Control | Up | GO: Cellular Compon | GO:00319envelope | 6.09E-03 | 1.00E+00 | 2.66E-02 | 1.74E-01 | 12 | 1439 | SORT1,S100A6,ATP5PF,MYOF,SLC25A6,PARK7,APOE,LDHB,ATP5F1A,SH3BGRL2,NDUFS5,VDAC2 |
| M1/Control | Up | GO: Cellular Compon | GO:00304axon | 5.72E-03 | 1.00E+00 | 2.66E-02 | 1.74E-01 | 9 | 901 | CFH,MYH10,THY1,PARK7,LRP2,DAG1,ITGA3,NDRG2,TNFRSF21 |
| M1/Control | Up | GO: Cellular Compon | GO:00057vacuole | 6.53E-03 | 1.00E+00 | 2.66E-02 | 1.74E-01 | 9 | 920 | SORT1,LAPTM4A,SLC12A4,BST2,LRP2,LAMP2,APOE,RAB2A,GNS |
| M1/Control | Up | GO: Cellular Compon | GO:00432receptor c | 6.31E-03 | 1.00E+00 | 2.66E-02 | 1.74E-01 | 7 | 596 | BMP2,LRP2,ITGA3,ITGAV,ITGB1,ITGB3,PLXNB1 |
| M1/Control | Up | GO: Cellular Compon | GO:00306endocytic | 6.81E-03 | 1.00E+00 | 2.66E-02 | 1.74E-01 | 4 | 204 | CD9,LRP2,LAMP2,APOE |
| M1/Control | Up | GO: Cellular Compon | GO:00453clathrin-co | 5.67E-03 | 1.00E+00 | 2.66E-02 | 1.74E-01 | 3 | 98 | CD9,LRP2,APOE |
| M1/Control | Up | GO: Cellular Compon | GO:00300desmosom | 5.80E-03 | 1.00E+00 | 2.66E-02 | 1.74E-01 | 2 | 31 | DSP,ACTN1 |
| M1/Control | Up | GO: Cellular Compon | GO:00057mitochonc | 7.34E-03 | 1.00E+00 | 2.66E-02 | 1.74E-01 | 1 | 2 | ATP5F1A |
| M1/Control | Down | GO: Molecular Functi | GO:00009RNA polyr | 5.51E-07 | 5.23E-05 | 5.23E-05 | 2.69E-04 | 3 | 24 | EGR1,FOS,CEBPB |
| M1/Control | Down | GO: Molecular Functi | GO:00010core prom | 7.05E-06 | 6.70E-04 | 3.35E-04 | 1.72E-03 | 3 | 55 | EGR1,FOS,CEBPB |
| M1/Control | Down | GO: Molecular Functi | GO:00469protein he | 2.89E-03 | 2.75E-01 | 1.87E-02 | 9.59E-02 | 3 | 423 | ATF3,CEBPB,NR4A1 |
| M1/Control | Down | GO: Molecular Functi | GO:00616RNA polyr | 2.95E-03 | 2.80E-01 | 1.87E-02 | 9.59E-02 | 3 | 426 | FOS,CEBPB,NR4A1 |
| M1/Control | Down | GO: Molecular Functi | GO:01402DNA-bind | 6.76E-03 | 6.42E-01 | 2.92E-02 | 1.50E-01 | 3 | 573 | FOS,CEBPB,NR4A1 |
| M1/Control | Down | GO: Molecular Functi | GO:00036chromatin | 1.36E-02 | 1.00E+00 | 4.46E-02 | 2.29E-01 | 3 | 740 | EGR1,FOS,CEBPB |
| M1/Control | Down | GO: Molecular Functi | GO:00081transcripti | 1.36E-02 | 1.00E+00 | 4.46E-02 | 2.29E-01 | 3 | 740 | FOS,CEBPB,NR4A1 |
| M1/Control | Down | GO: Biological Proces | GO:00436regulation | 4.97E-06 | 5.74E-03 | 1.38E-03 | 1.05E-02 | 3 | 55 | EGR1,ATF3,CEBPB |
| M1/Control | Down | GO: Biological Proces | GO:00436regulation | 7.14E-06 | 8.26E-03 | 1.38E-03 | 1.05E-02 | 3 | 62 | EGR1,ATF3,CEBPB |
| M1/Control | Down | GO: Biological Proces | GO:00425response t | 6.52E-04 | 7.53E-01 | 1.75E-02 | 1.34E-01 | 3 | 283 | FOS,ZFP36,ATF3 |
| M1/Control | Down | GO: Biological Proces | GO:00454fat cell dif | 7.57E-04 | 8.75E-01 | 1.86E-02 | 1.42E-01 | 3 | 298 | ZFP36,CEBPB,NR4A1 |
| M1/Control | Down | GO: Biological Proces | GO:00456regulation | 7.94E-04 | 9.18E-01 | 1.90E-02 | 1.45E-01 | 3 | 303 | FOS,ZFP36,CEBPB |
| M1/Control | Down | GO: Biological Proces | GO:00513response t | 9.05E-04 | 1.00E+00 | 1.90E-02 | 1.45E-01 | 3 | 317 | FOS,ZFP36,DUSP1 |
| M1/Control | Down | GO: Biological Proces | GO:00319response t | 1.24E-03 | 1.00E+00 | 2.13E-02 | 1.63E-01 | 3 | 353 | FOS,ZFP36,DUSP1 |
| M1/Control | Down | GO: Biological Proces | GO:00076learning o | 1.50E-03 | 1.00E+00 | 2.41E-02 | 1.84E-01 | 3 | 378 | EGR1,FOS,CEBPB |
| M1/Control | Down | GO: Biological Proces | GO:00508cognition | 2.10E-03 | 1.00E+00 | 2.73E-02 | 2.08E-01 | 3 | 425 | EGR1,FOS,CEBPB |
| M1/Control | Down | GO: Biological Proces | GO:00713cellular re | 2.51E-03 | 1.00E+00 | 2.87E-02 | 2.19E-01 | 3 | 452 | EGR1,FOS,NR4A1 |
| M1/Control | Down | GO: Biological Proces | GO:19012regulation | 2.87E-03 | 1.00E+00 | 3.16E-02 | 2.41E-01 | 3 | 474 | EGR1,FOS,CEBPB |
| M1/Control | Down | GO: Biological Proces | GO:00714cellular re | 3.17E-03 | 1.00E+00 | 3.27E-02 | 2.50E-01 | 3 | 491 | EGR1,FOS,ATF3 |
| M1/Control | Down | GO: Biological Proces | GO:00709neuron de | 3.95E-03 | 1.00E+00 | 3.71E-02 | 2.83E-01 | 3 | 531 | EGR1,FOS,CEBPB |
| M1/Control | Down | GO: Biological Proces | GO:19016cellular re | 4.06E-03 | 1.00E+00 | 3.73E-02 | 2.85E-01 | 3 | 536 | EGR1,FOS,NR4A1 |
| M1/Control | Down | GO: Biological Proces | GO:19037regulation | 4.52E-03 | 1.00E+00 | 3.73E-02 | 2.85E-01 | 3 | 557 | FOS,ZFP36,CEBPB |
| M1/Control | Down | GO: Biological Proces | GO:00506epithelial c | 6.29E-03 | 1.00E+00 | 4.25E-02 | 3.25E-01 | 3 | 627 | ZFP36,CEBPB,NR4A1 |
| M1/Control | Down | GO: Biological Proces | GO:00093response t | 6.83E-03 | 1.00E+00 | 4.49E-02 | 3.43E-01 | 3 | 646 | EGR1,FOS,DUSP1 |
| M1/Control | Down | GO: Biological Proces | GO:00300myeloid ce | 7.40E-03 | 1.00E+00 | 4.78E-02 | 3.65E-01 | 3 | 665 | FOS,ZFP36,CEBPB |
| M1/Control | Down | GO: Biological Proces | GO:00434response t | 7.77E-03 | 1.00E+00 | 4.97E-02 | 3.79E-01 | 3 | 677 | EGR1,FOS,NR4A1 |
| M1/Control | Down | GO: Cellular Compon | GO:19037sweet tast | 1.34E-03 | 8.96E-02 | 1.49E-02 | 7.15E-02 | 1 | 2 | TAS1R3 |
| M1/M0 | Up | GO: Molecular Functi | GO:00199cytokine b | 9.20E-04 | 1.92E-01 | 1.48E-02 | 8.76E-02 | 3 | 161 | ENG,ITGB1,LRRC32 |
| M1/M0 | Up | GO: Molecular Functi | GO:00051integrin bi | 1.10E-03 | 2.29E-01 | 1.48E-02 | 8.76E-02 | 3 | 171 | CD81,ITGB1,VCAM1 |
| M1/M0 | Up | GO: Molecular Functi | GO:00300extracellul | 1.16E-03 | 2.42E-01 | 1.48E-02 | 8.76E-02 | 2 | 42 | COL4A1,COL18A1 |
| M1/M0 | Up | GO: Molecular Functi | GO:00977disordered | 1.16E-03 | 2.42E-01 | 1.48E-02 | 8.76E-02 | 2 | 42 | GJA5,CTNNB1 |
| M1/M0 | Up | GO: Molecular Functi | GO:00860gap juncti | 1.20E-03 | 2.52E-01 | 1.48E-02 | 8.76E-02 | 1 | 1 | GJA5 |
| M1/M0 | Up | GO: Biological Proces | GO:00860atrial card | 1.16E-03 | 1.00E+00 | 1.35E-02 | 1.11E-01 | 1 | 1 | GJA5 |
| M1/M0 | Up | GO: Biological Proces | GO:00989regulation | 1.16E-03 | 1.00E+00 | 1.35E-02 | 1.11E-01 | 1 | 1 | GJA5 |
| M1/M0 | Up | GO: Biological Proces | GO:00363transformi | 1.16E-03 | 1.00E+00 | 1.35E-02 | 1.11E-01 | 1 | 1 | LRRC32 |
| M1/M0 | Up | GO: Biological Proces | GO:00995synaptic m | 1.16E-03 | 1.00E+00 | 1.35E-02 | 1.11E-01 | 1 | 1 | ITGB1 |
| M1/M0 | Up | GO: Biological Proces | GO:19013regulation | 1.16E-03 | 1.00E+00 | 1.35E-02 | 1.11E-01 | 1 | 1 | LRRC32 |
| M1/M0 | Up | GO: Biological Proces | GO:00300hemopoie | 1.22E-03 | 1.00E+00 | 1.41E-02 | 1.17E-01 | 7 | 1507 | FLNA,CD81,CTNNB1,ZFP36L1,ITGB1,H4C5,VCAM1 |
| M1/M0 | Up | GO: Biological Proces | GO:00421B cell activ | 1.23E-03 | 1.00E+00 | 1.41E-02 | 1.17E-01 | 4 | 415 | CD81,ZFP36L1,ITGB1,VCAM1 |
| M1/M0 | Up | GO: Biological Proces | GO:00613heart trab | 1.24E-03 | 1.00E+00 | 1.42E-02 | 1.17E-01 | 2 | 45 | HEG1,ENG |
| M1/M0 | Up | GO: Biological Proces | GO:00717basement | 1.29E-03 | 1.00E+00 | 1.46E-02 | 1.21E-01 | 2 | 46 | COL4A1,ITGB1 |
| M1/M0 | Up | GO: Biological Proces | GO:00714cellular re | 1.29E-03 | 1.00E+00 | 1.46E-02 | 1.21E-01 | 2 | 46 | CD81,ITGB1 |
| M1/M0 | Up | GO: Biological Proces | GO:00165negative n | 1.31E-03 | 1.00E+00 | 1.47E-02 | 1.21E-01 | 3 | 189 | HSPG2,TCF4,CTNNB1 |
| M1/M0 | Up | GO: Biological Proces | GO:00030renal syste | 1.31E-03 | 1.00E+00 | 1.47E-02 | 1.21E-01 | 3 | 189 | GJA5,CTNNB1,SGK1 |
| M1/M0 | Up | GO: Biological Proces | GO:00315cell-substr | 1.37E-03 | 1.00E+00 | 1.53E-02 | 1.26E-01 | 4 | 428 | FLNA,CTNNB1,ITGB1,VCAM1 |
| M1/M0 | Up | GO: Biological Proces | GO:20001negative n | 1.37E-03 | 1.00E+00 | 1.53E-02 | 1.26E-01 | 3 | 192 | HSPG2,TCF4,CTNNB1 |
| M1/M0 | Up | GO: Biological Proces | GO:00430camera-ty | 1.42E-03 | 1.00E+00 | 1.56E-02 | 1.28E-01 | 5 | 744 | COL4A1,S100A6,TCF4,CTNNB1,ITGB1 |
| M1/M0 | Up | GO: Biological Proces | GO:19013negative n | 1.42E-03 | 1.00E+00 | 1.56E-02 | 1.28E-01 | 3 | 194 | HSPG2,TCF4,CTNNB1 |
| M1/M0 | Up | GO: Biological Proces | GO:00420type 2 imm | 1.46E-03 | 1.00E+00 | 1.60E-02 | 1.32E-01 | 2 | 49 | CD81,ECM1 |
| M1/M0 | Up | GO: Biological Proces | GO:00485hematopo | 1.52E-03 | 1.00E+00 | 1.65E-02 | 1.36E-01 | 7 | 1565 | FLNA,CD81,CTNNB1,ZFP36L1,ITGB1,H4C5,VCAM1 |
| M1/M0 | Up | GO: Biological Proces | GO:00107positive re | 1.52E-03 | 1.00E+00 | 1.65E-02 | 1.36E-01 | 4 | 440 | FLNA,CTNNB1,ITGB1,SGK1 |
| M1/M0 | Up | GO: Biological Proces | GO:00301B cell diffe | 1.54E-03 | 1.00E+00 | 1.66E-02 | 1.37E-01 | 3 | 200 | ZFP36L1,ITGB1,VCAM1 |
| M1/M0 | Up | GO: Biological Proces | GO:00450early endo | 1.58E-03 | 1.00E+00 | 1.69E-02 | 1.39E-01 | 2 | 51 | FLNA,MSN |
| M1/M0 | Up | GO: Biological Proces | GO:00511regulation | 1.58E-03 | 1.00E+00 | 1.69E-02 | 1.39E-01 | 2 | 51 | RCAN1,ENG |
| M1/M0 | Up | GO: Biological Proces | GO:00431positive re | 1.63E-03 | 1.00E+00 | 1.73E-02 | 1.43E-01 | 3 | 204 | FLNA,CTNNB1,ECM1 |
| M1/M0 | Up | GO: Biological Proces | GO:19026regulation | 1.65E-03 | 1.00E+00 | 1.74E-02 | 1.43E-01 | 2 | 52 | CTNNB1,ITGB1 |
| M1/M0 | Up | GO: Biological Proces | GO:00018epithelial t | 1.68E-03 | 1.00E+00 | 1.76E-02 | 1.45E-01 | 3 | 206 | FLNA,CTNNB1,ENG |
| M1/M0 | Up | GO: Biological Proces | GO:00031atrioventri | 1.71E-03 | 1.00E+00 | 1.79E-02 | 1.47E-01 | 2 | 53 | GJA5,ITGB1 |
| M1/M0 | Up | GO: Biological Proces | GO:00485developm | 1.77E-03 | 1.00E+00 | 1.83E-02 | 1.51E-01 | 6 | 1168 | CD81,HEG1,CTNNB1,ZFP36L1,ECM1,ITGB1 |
| M1/M0 | Up | GO: Biological Proces | GO:00860cardiac mu | 1.77E-03 | 1.00E+00 | 1.83E-02 | 1.51E-01 | 2 | 54 | FLNA,GJA5 |
| M1/M0 | Up | GO: Biological Proces | GO:00989vesicle-me | 1.77E-03 | 1.00E+00 | 1.83E-02 | 1.51E-01 | 2 | 54 | FLNA,MSN |
| M1/M0 | Up | GO: Biological Proces | GO:00106epithelial c | 1.81E-03 | 1.00E+00 | 1.84E-02 | 1.52E-01 | 4 | 461 | HSPG2,COL18A1,GJA5,ITGB1 |
| M1/M0 | Up | GO: Biological Proces | GO:00019regulation | 1.80E-03 | 1.00E+00 | 1.84E-02 | 1.52E-01 | 3 | 211 | COL18A1,ECM1,ENG |
| M1/M0 | Up | GO: Biological Proces | GO:00905epiboly inv | 1.84E-03 | 1.00E+00 | 1.86E-02 | 1.53E-01 | 2 | 55 | FLNA,ITGB1 |
| M1/M0 | Up | GO: Biological Proces | GO:00443wound hea | 1.84E-03 | 1.00E+00 | 1.86E-02 | 1.53E-01 | 2 | 55 | FLNA,ITGB1 |
| M1/M0 | Up | GO: Biological Proces | GO:00508regulation | 1.88E-03 | 1.00E+00 | 1.89E-02 | 1.56E-01 | 4 | 466 | CD81,CTNNB1,VCAM1,LRRC32 |
| M1/M0 | Up | GO: Biological Proces | GO:00901epithelium | 1.89E-03 | 1.00E+00 | 1.89E-02 | 1.56E-01 | 4 | 467 | HSPG2,COL18A1,GJA5,ITGB1 |
| M1/M0 | Up | GO: Biological Proces | GO:00860membrane | 1.91E-03 | 1.00E+00 | 1.89E-02 | 1.56E-01 | 2 | 56 | FLNA,GJA5 |
| M1/M0 | Up | GO: Biological Proces | GO:00030glomerula | 1.91E-03 | 1.00E+00 | 1.89E-02 | 1.56E-01 | 2 | 56 | GJA5,CTNNB1 |
| M1/M0 | Up | GO: Biological Proces | GO:00149myotube d | 1.92E-03 | 1.00E+00 | 1.90E-02 | 1.56E-01 | 3 | 216 | CD81,RCAN1,ITGB1 |
| M1/M0 | Up | GO: Biological Proces | GO:00074sensory or | 1.95E-03 | 1.00E+00 | 1.91E-02 | 1.58E-01 | 6 | 1190 | COL4A1,S100A6,TCF4,CTNNB1,IGFBP7,ITGB1 |
| M1/M0 | Up | GO: Biological Proces | GO:00025immune s | 1.97E-03 | 1.00E+00 | 1.93E-02 | 1.59E-01 | 7 | 1636 | FLNA,CD81,CTNNB1,ZFP36L1,ITGB1,H4C5,VCAM1 |
| M1/M0 | Up | GO: Biological Proces | GO:00488stem cell d | 2.01E-03 | 1.00E+00 | 1.96E-02 | 1.62E-01 | 4 | 475 | COL18A1,CTNNB1,ENG,ITGB1 |

| | | | | | | | | | | | |
|---|---|---|---|---|---|---|---|---|---|---|---|
| M1/M0 | Up | GO: Biological Proces | GO:19025 | positive re | 2.21E-03 | 1.00E+00 | 1.97E-02 | 1.62E-01 | 6 | 1220 | FLNA,CD81,CTNNB1,ECM1,ENG,ITGB1 |
| M1/M0 | Up | GO: Biological Proces | GO:00025 | leukocyte | 2.13E-03 | 1.00E+00 | 1.97E-02 | 1.62E-01 | 5 | 816 | CD81,CTNNB1,ZFP36L1,ITGB1,VCAM1 |
| M1/M0 | Up | GO: Biological Proces | GO:00026 | regulation | 2.19E-03 | 1.00E+00 | 1.97E-02 | 1.62E-01 | 5 | 821 | CD81,CTNNB1,ZFP36L1,VCAM1,LRRC32 |
| M1/M0 | Up | GO: Biological Proces | GO:00901 | tissue mig | 2.04E-03 | 1.00E+00 | 1.97E-02 | 1.62E-01 | 4 | 477 | HSPG2,COL18A1,GJA5,ITGB1 |
| M1/M0 | Up | GO: Biological Proces | GO:07121 | cellular re | 2.06E-03 | 1.00E+00 | 1.97E-02 | 1.62E-01 | 4 | 478 | CTNNB1,ZFP36L1,ENG,ITGB1 |
| M1/M0 | Up | GO: Biological Proces | GO:01040 | cellular re | 2.06E-03 | 1.00E+00 | 1.97E-02 | 1.62E-01 | 4 | 478 | CTNNB1,ZFP36L1,ENG,ITGB1 |
| M1/M0 | Up | GO: Biological Proces | GO:00714 | cellular re | 2.27E-03 | 1.00E+00 | 1.97E-02 | 1.62E-01 | 4 | 491 | CTNNB1,ENG,ITGB1,VCAM1 |
| M1/M0 | Up | GO: Biological Proces | GO:00435 | blood ves | 2.08E-03 | 1.00E+00 | 1.97E-02 | 1.62E-01 | 3 | 222 | HSPG2,COL18A1,ITGB1 |
| M1/M0 | Up | GO: Biological Proces | GO:00107 | positive re | 2.04E-03 | 1.00E+00 | 1.97E-02 | 1.62E-01 | 2 | 58 | CTNNB1,ENG |
| M1/M0 | Up | GO: Biological Proces | GO:00465 | negative n | 2.18E-03 | 1.00E+00 | 1.97E-02 | 1.62E-01 | 2 | 60 | HEG1,ITGB1 |
| M1/M0 | Up | GO: Biological Proces | GO:00358 | vascular a | 2.26E-03 | 1.00E+00 | 1.97E-02 | 1.62E-01 | 2 | 61 | CTNNB1,ENG |
| M1/M0 | Up | GO: Biological Proces | GO:00031 | pulmonary | 2.32E-03 | 1.00E+00 | 1.97E-02 | 1.62E-01 | 1 | 2 | GJA5 |
| M1/M0 | Up | GO: Biological Proces | GO:00106 | positive re | 2.32E-03 | 1.00E+00 | 1.97E-02 | 1.62E-01 | 1 | 2 | GJA5 |
| M1/M0 | Up | GO: Biological Proces | GO:00359 | foramen o | 2.32E-03 | 1.00E+00 | 1.97E-02 | 1.62E-01 | 1 | 2 | GJA5 |
| M1/M0 | Up | GO: Biological Proces | GO:00106 | regulation | 2.32E-03 | 1.00E+00 | 1.97E-02 | 1.62E-01 | 1 | 2 | GJA5 |
| M1/M0 | Up | GO: Biological Proces | GO:00609 | cardiac ne | 2.32E-03 | 1.00E+00 | 1.97E-02 | 1.62E-01 | 1 | 2 | VCAM1 |
| M1/M0 | Up | GO: Biological Proces | GO:00609 | epicardium | 2.32E-03 | 1.00E+00 | 1.97E-02 | 1.62E-01 | 1 | 2 | CTNNB1 |
| M1/M0 | Up | GO: Biological Proces | GO:00704 | detection | 2.32E-03 | 1.00E+00 | 1.97E-02 | 1.62E-01 | 1 | 2 | ENG |
| M1/M0 | Up | GO: Biological Proces | GO:20000 | positive re | 2.32E-03 | 1.00E+00 | 1.97E-02 | 1.62E-01 | 1 | 2 | CTNNB1 |
| M1/M0 | Up | GO: Biological Proces | GO:19050 | regulation | 2.32E-03 | 1.00E+00 | 1.97E-02 | 1.62E-01 | 1 | 2 | FLNA |
| M1/M0 | Up | GO: Biological Proces | GO:00459 | negative n | 2.32E-03 | 1.00E+00 | 1.97E-02 | 1.62E-01 | 1 | 2 | CTNNB1 |
| M1/M0 | Up | GO: Biological Proces | GO:00860 | bundle of | 2.32E-03 | 1.00E+00 | 1.97E-02 | 1.62E-01 | 1 | 2 | GJA5 |
| M1/M0 | Up | GO: Biological Proces | GO:00860 | SA node c | 2.32E-03 | 1.00E+00 | 1.97E-02 | 1.62E-01 | 1 | 2 | GJA5 |
| M1/M0 | Up | GO: Biological Proces | GO:00989 | regulation | 2.32E-03 | 1.00E+00 | 1.97E-02 | 1.62E-01 | 1 | 2 | GJA5 |
| M1/M0 | Up | GO: Biological Proces | GO:00486 | myoblast f | 2.32E-03 | 1.00E+00 | 1.97E-02 | 1.62E-01 | 1 | 2 | ITGB1 |
| M1/M0 | Up | GO: Biological Proces | GO:00613 | canonical | 2.32E-03 | 1.00E+00 | 1.97E-02 | 1.62E-01 | 1 | 2 | CTNNB1 |
| M1/M0 | Up | GO: Biological Proces | GO:00030 | lymph circ | 2.32E-03 | 1.00E+00 | 1.97E-02 | 1.62E-01 | 1 | 2 | HEG1 |
| M1/M0 | Up | GO: Biological Proces | GO:01400 | cell-cell ac | 2.32E-03 | 1.00E+00 | 1.97E-02 | 1.62E-01 | 1 | 2 | VCAM1 |
| M1/M0 | Up | GO: Biological Proces | GO:00720 | renal oute | 2.32E-03 | 1.00E+00 | 1.97E-02 | 1.62E-01 | 1 | 2 | CTNNB1 |
| M1/M0 | Up | GO: Biological Proces | GO:00443 | canonical | 2.32E-03 | 1.00E+00 | 1.97E-02 | 1.62E-01 | 1 | 2 | CTNNB1 |
| M1/M0 | Up | GO: Biological Proces | GO:00519 | positive re | 2.32E-03 | 1.00E+00 | 1.97E-02 | 1.62E-01 | 1 | 2 | ITGB1 |
| M1/M0 | Up | GO: Cellular Compon | GO:00147 | intercalate | 7.19E-05 | 1.66E-02 | 1.28E-03 | 7.70E-03 | 3 | 71 | GJA5,CTNNB1,ITGB1 |
| M1/M0 | Up | GO: Cellular Compon | GO:00442 | cell-cell cc | 1.66E-04 | 3.83E-02 | 2.39E-03 | 1.44E-02 | 3 | 94 | GJA5,CTNNB1,ITGB1 |
| M1/M0 | Up | GO: Cellular Compon | GO:00301 | filopodium | 3.84E-04 | 8.87E-02 | 4.22E-03 | 2.54E-02 | 3 | 125 | ITGB1,VCAM1,MSN |
| M1/M0 | Up | GO: Cellular Compon | GO:00059 | microvillus | 4.40E-04 | 1.02E-01 | 4.42E-03 | 2.66E-02 | 3 | 131 | CTNNB1,VCAM1,MSN |
| M1/M0 | Up | GO: Cellular Compon | GO:00059 | adherens j | 1.59E-03 | 3.67E-01 | 1.31E-02 | 7.89E-02 | 3 | 204 | CTNNB1,ITGB1,MSN |
| M1/M0 | Up | GO: Cellular Compon | GO:00163 | basolatera | 4.99E-03 | 1.00E+00 | 2.51E-02 | 1.51E-01 | 3 | 306 | CD81,CTNNB1,MSN |
| M1/M0 | Up | GO: Cellular Compon | GO:00057 | endoplasm | 6.41E-03 | 1.00E+00 | 2.79E-02 | 1.68E-01 | 3 | 335 | COL4A1,COL18A1,IGFBP7 |
| M1/M0 | Up | GO: Cellular Compon | GO:00099 | basal plas | 6.41E-03 | 1.00E+00 | 2.79E-02 | 1.68E-01 | 3 | 335 | CD81,CTNNB1,MSN |
| M1/M0 | Up | GO: Cellular Compon | GO:00451 | basal part | 7.87E-03 | 1.00E+00 | 3.19E-02 | 1.92E-01 | 3 | 361 | CD81,CTNNB1,MSN |
| M1/M0 | Up | GO: Cellular Compon | GO:00312 | cell projec | 1.25E-02 | 1.00E+00 | 4.33E-02 | 2.60E-01 | 3 | 428 | CTNNB1,ITGB1,MSN |
| M1/Control | Up | GO: Molecular Functi | GO:00320 | myosin he | 1.92E-03 | 8.35E-01 | 1.90E-02 | 1.26E-01 | 2 | 17 | MYL9,MYL12B |
| M1/Control | Up | GO: Molecular Functi | GO:00452 | cadherin b | 2.04E-03 | 8.84E-01 | 1.96E-02 | 1.31E-01 | 6 | 341 | NDRG1,PTPRM,PARK7,PRDX1,MACF1,ITGB1 |
| M1/Control | Up | GO: Molecular Functi | GO:00469 | proton-tra | 2.16E-03 | 9.37E-01 | 2.04E-02 | 1.36E-01 | 2 | 18 | ATP5PF,ATP5F1A |
| M1/Control | Up | GO: Molecular Functi | GO:00046 | peroxidase | 2.31E-03 | 1.00E+00 | 2.13E-02 | 1.42E-01 | 3 | 68 | PARK7,SELENOF,PRDX1 |
| M1/Control | Up | GO: Molecular Functi | GO:00166 | oxidoredu | 2.51E-03 | 1.00E+00 | 2.27E-02 | 1.51E-01 | 3 | 70 | PARK7,SELENOF,PRDX1 |
| M1/Control | Up | GO: Molecular Functi | GO:00051 | signaling r | 3.55E-03 | 1.00E+00 | 2.55E-02 | 1.69E-01 | 15 | 1813 | BMP2,THBS1,THY1,CD9,PARK7,LRP2,CD151,ACTN1,ITGA3,APOE,ITGAV,ITGB1,ITGB3,ATP5F1A,PLXNB1 |
| M1/Control | Up | GO: Molecular Functi | GO:00986 | cell adhesi | 2.94E-03 | 1.00E+00 | 2.55E-02 | 1.69E-01 | 3 | 74 | DSP,BCAM,ITGB1 |
| M1/Control | Up | GO: Molecular Functi | GO:00055 | collagen b | 3.79E-03 | 1.00E+00 | 2.55E-02 | 1.69E-01 | 3 | 81 | THBS1,ITGA3,ITGB1 |
| M1/Control | Up | GO: Molecular Functi | GO:00364 | L-dopa de | 3.87E-03 | 1.00E+00 | 2.55E-02 | 1.69E-01 | 1 | 1 | PARK7 |
| M1/Control | Up | GO: Molecular Functi | GO:00364 | tyrosine 3- | 3.87E-03 | 1.00E+00 | 2.55E-02 | 1.69E-01 | 1 | 1 | PARK7 |
| M1/Control | Up | GO: Molecular Functi | GO:00187 | 5-formylgl | 3.87E-03 | 1.00E+00 | 2.55E-02 | 1.69E-01 | 1 | 1 | ESD |
| M1/Control | Up | GO: Molecular Functi | GO:00166 | oxidoredu | 3.87E-03 | 1.00E+00 | 2.55E-02 | 1.69E-01 | 1 | 1 | AKR1A1 |
| M1/Control | Up | GO: Molecular Functi | GO:19900 | methylglyo | 3.87E-03 | 1.00E+00 | 2.55E-02 | 1.69E-01 | 1 | 1 | AKR1A1 |
| M1/Control | Up | GO: Molecular Functi | GO:00197 | mevaldate | 3.87E-03 | 1.00E+00 | 2.55E-02 | 1.69E-01 | 1 | 1 | AKR1A1 |
| M1/Control | Up | GO: Molecular Functi | GO:00044 | mevaldate | 3.87E-03 | 1.00E+00 | 2.55E-02 | 1.69E-01 | 1 | 1 | AKR1A1 |
| M1/Control | Up | GO: Molecular Functi | GO:00469 | metal chel | 3.87E-03 | 1.00E+00 | 2.55E-02 | 1.69E-01 | 1 | 1 | APOE |
| M1/Control | Up | GO: Molecular Functi | GO:00303 | neurotensi | 3.87E-03 | 1.00E+00 | 2.55E-02 | 1.69E-01 | 1 | 1 | SORT1 |
| M1/Control | Up | GO: Molecular Functi | GO:00479 | glucurono | 3.87E-03 | 1.00E+00 | 2.55E-02 | 1.69E-01 | 1 | 1 | AKR1A1 |
| M1/Control | Up | GO: Molecular Functi | GO:00479 | glycine N- | 3.87E-03 | 1.00E+00 | 2.55E-02 | 1.69E-01 | 1 | 1 | GLYAT |
| M1/Control | Up | GO: Molecular Functi | GO:19904 | glyoxalase | 3.87E-03 | 1.00E+00 | 2.55E-02 | 1.69E-01 | 1 | 1 | PARK7 |
| M1/Control | Up | GO: Molecular Functi | GO:00479 | L-glucuror | 3.87E-03 | 1.00E+00 | 2.55E-02 | 1.69E-01 | 1 | 1 | AKR1A1 |
| M1/Control | Up | GO: Molecular Functi | GO:00365 | protein de | 3.87E-03 | 1.00E+00 | 2.55E-02 | 1.69E-01 | 1 | 1 | PARK7 |
| M1/Control | Up | GO: Molecular Functi | GO:00038 | CDP-diacy | 3.87E-03 | 1.00E+00 | 2.55E-02 | 1.69E-01 | 1 | 1 | CDIPT |
| M1/Control | Up | GO: Molecular Functi | GO:00152 | proton ch | 4.16E-03 | 1.00E+00 | 2.62E-02 | 1.74E-01 | 2 | 25 | ATP5PF,ATP5F1A |
| M1/Control | Up | GO: Biological Proces | GO:19031 | glyoxal cat | 3.68E-03 | 1.00E+00 | 3.32E-02 | 2.87E-01 | 1 | 1 | PARK7 |
| M1/Control | Up | GO: Biological Proces | GO:19031 | L-dopa me | 3.68E-03 | 1.00E+00 | 3.32E-02 | 2.87E-01 | 1 | 1 | PARK7 |
| M1/Control | Up | GO: Biological Proces | GO:19031 | L-dopa bio | 3.68E-03 | 1.00E+00 | 3.32E-02 | 2.87E-01 | 1 | 1 | PARK7 |
| M1/Control | Up | GO: Biological Proces | GO:19031 | glyoxal me | 3.68E-03 | 1.00E+00 | 3.32E-02 | 2.87E-01 | 1 | 1 | PARK7 |
| M1/Control | Up | GO: Biological Proces | GO:20011 | negative n | 3.68E-03 | 1.00E+00 | 3.32E-02 | 2.87E-01 | 1 | 1 | APOE |
| M1/Control | Up | GO: Biological Proces | GO:01400 | cellular de | 3.68E-03 | 1.00E+00 | 3.32E-02 | 2.87E-01 | 1 | 1 | PARK7 |
| M1/Control | Up | GO: Biological Proces | GO:00030 | positive re | 3.68E-03 | 1.00E+00 | 3.32E-02 | 2.87E-01 | 1 | 1 | TPM1 |
| M1/Control | Up | GO: Biological Proces | GO:00995 | synaptic m | 3.68E-03 | 1.00E+00 | 3.32E-02 | 2.87E-01 | 1 | 1 | ITGB1 |
| M1/Control | Up | GO: Biological Proces | GO:01400 | positive re | 3.68E-03 | 1.00E+00 | 3.32E-02 | 2.87E-01 | 1 | 1 | LRP2 |
| M1/Control | Up | GO: Biological Proces | GO:19044 | response t | 3.68E-03 | 1.00E+00 | 3.32E-02 | 2.87E-01 | 1 | 1 | APOE |
| M1/Control | Up | GO: Biological Proces | GO:00719 | G-quadrup | 3.68E-03 | 1.00E+00 | 3.32E-02 | 2.87E-01 | 1 | 1 | CNBP |
| M1/Control | Up | GO: Biological Proces | GO:00488 | chemical h | 3.90E-03 | 1.00E+00 | 3.40E-02 | 2.94E-01 | 12 | 1365 | CFH,CNBP,THY1,SLC12A4,NFE2L1,PARK7,GLS,APLP2,AKR1A1,APOE,ITGAV,ITGB3 |
| M1/Control | Up | GO: Biological Proces | GO:00092 | purine ribo | 3.91E-03 | 1.00E+00 | 3.40E-02 | 2.94E-01 | 5 | 284 | CFH,ATP5PF,ATP5IF1,ATP5F1A,NDUFS5 |
| M1/Control | Up | GO: Biological Proces | GO:00431 | receptor c | 3.90E-03 | 1.00E+00 | 3.40E-02 | 2.94E-01 | 3 | 86 | THY1,DAG1,APOE |
| M1/Control | Up | GO: Biological Proces | GO:00313 | negative n | 3.93E-03 | 1.00E+00 | 3.42E-02 | 2.95E-01 | 4 | 175 | TPM1,PARK7,LAMP2,VDAC2 |
| M1/Control | Up | GO: Biological Proces | GO:00074 | sensory or | 3.95E-03 | 1.00E+00 | 3.43E-02 | 2.96E-01 | 11 | 1190 | CFH,S100A6,BMP2,MYH10,THY1,IGFBP7,PTPRM,SULF1,CDIPT,FRZB,ITGB1 |
| M1/Control | Up | GO: Biological Proces | GO:00097 | response t | 4.02E-03 | 1.00E+00 | 3.46E-02 | 2.99E-01 | 12 | 1370 | SORT1,THBS1,TCF21,IGFBP7,PARK7,LRP2,DAG1,ITGA3,APOE,ITGB1,ITGB3,ATP5F1A |
| M1/Control | Up | GO: Biological Proces | GO:00160 | cell growt | 4.07E-03 | 1.00E+00 | 3.46E-02 | 2.99E-01 | 8 | 700 | THSD7A,IGFBP7,BST2,FRZB,MACF1,APOE,ITGB1,PLXNB1 |
| M1/Control | Up | GO: Biological Proces | GO:00032 | cardiac ch | 4.01E-03 | 1.00E+00 | 3.46E-02 | 2.99E-01 | 4 | 176 | TPM1,BMP2,DSP,LRP2 |
| M1/Control | Up | GO: Biological Proces | GO:01501 | transport a | 4.03E-03 | 1.00E+00 | 3.46E-02 | 2.99E-01 | 3 | 87 | SLC22A8,LRP2,APOE |
| M1/Control | Up | GO: Biological Proces | GO:00721 | cell prolife | 4.08E-03 | 1.00E+00 | 3.46E-02 | 2.99E-01 | 2 | 26 | BMP2,ITGB3 |
| M1/Control | Up | GO: Biological Proces | GO:00070 | protein ca | 4.08E-03 | 1.00E+00 | 3.46E-02 | 2.99E-01 | 2 | 26 | LRP2,LAMP2 |
| M1/Control | Up | GO: Biological Proces | GO:00031 | heart field | 4.08E-03 | 1.00E+00 | 3.46E-02 | 2.99E-01 | 2 | 26 | BMP2,LRP2 |
| M1/Control | Up | GO: Biological Proces | GO:00073 | fusion of s | 4.08E-03 | 1.00E+00 | 3.46E-02 | 2.99E-01 | 2 | 26 | CD9,ITGA3 |
| M1/Control | Up | GO: Biological Proces | GO:00430 | negative n | 4.16E-03 | 1.00E+00 | 3.50E-02 | 3.03E-01 | 11 | 1198 | THBS1,THY1,SLC25A6,PARK7,DAD1,LRP2,APOE,ITGAV,ITGB1,PTTG1IP,VDAC2 |
| M1/Control | Up | GO: Biological Proces | GO:00035 | aorta deve | 4.16E-03 | 1.00E+00 | 3.50E-02 | 3.03E-01 | 3 | 88 | MYH10,EFEMP2,LRP2 |
| M1/Control | Up | GO: Biological Proces | GO:00102 | vascular tr | 4.16E-03 | 1.00E+00 | 3.50E-02 | 3.03E-01 | 3 | 88 | SLC22A8,LRP2,APOE |
| M1/Control | Up | GO: Biological Proces | GO:00455 | positive re | 4.19E-03 | 1.00E+00 | 3.50E-02 | 3.03E-01 | 11 | 1199 | BMP2,FRZB,EFEMP2,LRP2,DAG1,MACF1,APOE,SETD3,ITGB1,ITGB3,PLXNB1 |
| M1/Control | Up | GO: Biological Proces | GO:00487 | oligodend | 4.18E-03 | 1.00E+00 | 3.50E-02 | 3.03E-01 | 4 | 178 | CD9,DAG1,APOE,TNFRSF21 |
| M1/Control | Up | GO: Biological Proces | GO:00430 | regulation | 4.24E-03 | 1.00E+00 | 3.54E-02 | 3.06E-01 | 15 | 1944 | SORT1,BMP2,THBS1,THY1,SLC25A6,PARK7,FRZB,DAD1,LRP2,ACTN1,APOE,ITGAV,ITGB1,PTTG1IP,VDAC2 |
| M1/Control | Up | GO: Biological Proces | GO:00091 | purine nuc | 4.27E-03 | 1.00E+00 | 3.55E-02 | 3.07E-01 | 5 | 290 | CFH,ATP5PF,ATP5IF1,ATP5F1A,NDUFS5 |
| M1/Control | Up | GO: Biological Proces | GO:00091 | ribonuclec | 4.33E-03 | 1.00E+00 | 3.60E-02 | 3.11E-01 | 5 | 291 | CFH,ATP5PF,ATP5IF1,ATP5F1A,NDUFS5 |
| M1/Control | Up | GO: Biological Proces | GO:01501 | protein loc | 4.39E-03 | 1.00E+00 | 3.63E-02 | 3.14E-01 | 2 | 27 | DSP,CGNL1 |
| M1/Control | Up | GO: Biological Proces | GO:00350 | negative n | 4.39E-03 | 1.00E+00 | 3.63E-02 | 3.14E-01 | 2 | 27 | ITGA3,ITGB1 |
| M1/Control | Up | GO: Biological Proces | GO:00986 | anion tran | 4.46E-03 | 1.00E+00 | 3.67E-02 | 3.17E-01 | 5 | 293 | SLC12A4,SLC25A6,LRP2,ITGB1,VDAC2 |
| M1/Control | Up | GO: Biological Proces | GO:00458 | negative n | 4.54E-03 | 1.00E+00 | 3.73E-02 | 3.22E-01 | 6 | 422 | THBS1,BST2,PARK7,LRP2,APLP2,ITIH5 |
| M1/Control | Up | GO: Biological Proces | GO:01402 | cell-cell fu | 4.71E-03 | 1.00E+00 | 3.85E-02 | 3.33E-01 | 3 | 92 | MYOF,CD9,ITGB1 |
| M1/Control | Up | GO: Biological Proces | GO:00007 | syncytium | 4.71E-03 | 1.00E+00 | 3.85E-02 | 3.33E-01 | 3 | 92 | MYOF,CD9,ITGB1 |
| M1/Control | Up | GO: Biological Proces | GO:00717 | cellular re | 4.72E-03 | 1.00E+00 | 3.85E-02 | 3.33E-01 | 2 | 28 | THBS1,ATP5F1A |
| M1/Control | Up | GO: Biological Proces | GO:00325 | response t | 4.79E-03 | 1.00E+00 | 3.89E-02 | 3.37E-01 | 4 | 185 | BMP2,IGFBP7,LRP2,APOE |
| M1/Control | Up | GO: Biological Proces | GO:00301 | regulation | 4.81E-03 | 1.00E+00 | 3.91E-02 | 3.38E-01 | 9 | 880 | THBS1,ATP5IF1,NFE2L1,BST2,PARK7,LRP2,APLP2,APOE,ITIH5 |
| M1/Control | Up | GO: Biological Proces | GO:00069 | inflammat | 4.83E-03 | 1.00E+00 | 3.91E-02 | 3.38E-01 | 10 | 1048 | CFH,CNBP,BMP2,THBS1,ATP5IF1,NFE2L1,PARK7,APOE,ITGAV,ITGB1 |
| M1/Control | Up | GO: Biological Proces | GO:00467 | viral entry | 4.88E-03 | 1.00E+00 | 3.92E-02 | 3.39E-01 | 4 | 186 | DAG1,ITGAV,ITGB1,ITGB3 |
| M1/Control | Up | GO: Biological Proces | GO:00600 | retina mor | 4.88E-03 | 1.00E+00 | 3.92E-02 | 3.39E-01 | 4 | 186 | CFH,THY1,PTPRM,CDIPT |
| M1/Control | Up | GO: Biological Proces | GO:00086 | extrinsic a | 4.86E-03 | 1.00E+00 | 3.92E-02 | 3.39E-01 | 3 | 93 | SORT1,THBS1,PARK7 |
| M1/Control | Up | GO: Biological Proces | GO:00160 | Wnt signa | 4.98E-03 | 1.00E+00 | 3.98E-02 | 3.44E-01 | 7 | 572 | BMP2,SULF1,FRZB,MACF1,ITGA3,APOE,NDRG2 |
| M1/Control | Up | GO: Biological Proces | GO:00034 | neural reti | 4.97E-03 | 1.00E+00 | 3.98E-02 | 3.44E-01 | 4 | 187 | CFH,THY1,PTPRM,CDIPT |
| M1/Control | Up | GO: Biological Proces | GO:00140 | positive re | 5.00E-03 | 1.00E+00 | 3.99E-02 | 3.45E-01 | 3 | 94 | BMP2,LRP2,DAG1 |
| M1/Control | Up | GO: Biological Proces | GO:00066 | protein ta | 5.06E-03 | 1.00E+00 | 4.01E-02 | 3.47E-01 | 2 | 29 | SORT1,LAMP2 |
| M1/Control | Up | GO: Biological Proces | GO:00714 | cellular re | 5.06E-03 | 1.00E+00 | 4.01E-02 | 3.47E-01 | 2 | 29 | CD9,ITGB1 |
| M1/Control | Up | GO: Biological Proces | GO:01987 | cell-cell sig | 5.07E-03 | 1.00E+00 | 4.01E-02 | 3.47E-01 | 7 | 574 | BMP2,SULF1,FRZB,MACF1,ITGA3,APOE,NDRG2 |
| M1/Control | Up | GO: Biological Proces | GO:00030 | renal syste | 5.16E-03 | 1.00E+00 | 4.06E-02 | 3.51E-01 | 4 | 189 | THSD7A,SULF1,LRP2,ITGA3 |
| M1/Control | Up | GO: Cellular Compon | GO:00711 | alpha9-be | 7.34E-03 | 1.00E+00 | 2.66E-02 | 1.74E-01 | 1 | 2 | ITGB3 |
| M1/Control | Up | GO: Cellular Compon | GO:00976 | intrinsic cc | 7.34E-03 | 1.00E+00 | 2.66E-02 | 1.74E-01 | 1 | 2 | LAMP2 |
| M1/Control | Up | GO: Cellular Compon | GO:00988 | podocyte | 7.34E-03 | 1.00E+00 | 2.66E-02 | 1.74E-01 | 1 | 2 | THSD7A |
| M1/Control | Up | GO: Cellular Compon | GO:00976 | integral cc | 7.34E-03 | 1.00E+00 | 2.66E-02 | 1.74E-01 | 1 | 2 | LAMP2 |
| M1/Control | Up | GO: Cellular Compon | GO:00346 | integrin al | 7.34E-03 | 1.00E+00 | 2.66E-02 | 1.74E-01 | 1 | 2 | ITGB1 |
| M1/Control | Up | GO: Cellular Compon | GO:00906 | outer dens | 7.34E-03 | 1.00E+00 | 2.66E-02 | 1.74E-01 | 1 | 2 | ACTN1 |
| M1/Control | Up | GO: Cellular Compon | GO:00906 | inner dens | 7.34E-03 | 1.00E+00 | 2.66E-02 | 1.74E-01 | 1 | 2 | ACTN1 |
| M1/Control | Up | GO: Cellular Compon | GO:00346 | integrin al | 7.34E-03 | 1.00E+00 | 2.66E-02 | 1.74E-01 | 1 | 2 | ITGB1 |
| M1/Control | Up | GO: Cellular Compon | GO:00346 | integrin al | 7.34E-03 | 1.00E+00 | 2.66E-02 | 1.74E-01 | 1 | 2 | ITGAV |
| M1/Control | Up | GO: Cellular Compon | GO:00346 | integrin al | 7.34E-03 | 1.00E+00 | 2.66E-02 | 1.74E-01 | 1 | 2 | ITGB1 |
| M1/Control | Up | GO: Cellular Compon | GO:00346 | integrin al | 7.34E-03 | 1.00E+00 | 2.66E-02 | 1.74E-01 | 1 | 2 | ITGB1 |
| M1/Control | Up | GO: Cellular Compon | GO:19908 | lysosomal | 7.34E-03 | 1.00E+00 | 2.66E-02 | 1.74E-01 | 1 | 2 | LAMP2 |
| M1/Control | Down | GO: Molecular Functi | GO:00352 | nuclear glu | 8.62E-05 | 8.18E-03 | 1.64E-03 | 8.41E-03 | 2 | 20 | CEBPB,NR4A1 |
| M1/Control | Down | GO: Molecular Functi | GO:00350 | histone ac | 1.83E-04 | 1.74E-02 | 2.90E-03 | 1.49E-02 | 2 | 29 | EGR1,CEBPB |
| M1/Control | Down | GO: Molecular Functi | GO:00169 | nuclear re | 7.68E-03 | 7.30E-01 | 3.04E-02 | 1.56E-01 | 2 | 191 | CEBPB,NR4A1 |
| M1/Control | Down | GO: Biological Proces | GO:19904 | positive re | 2.39E-05 | 2.76E-02 | 2.70E-03 | 2.06E-02 | 2 | 12 | ATF3,CEBPB |
| M1/Control | Down | GO: Biological Proces | GO:00360 | positive re | 1.36E-04 | 1.57E-01 | 7.87E-03 | 6.00E-02 | 2 | 28 | ATF3,CEBPB |
| M1/Control | Down | GO: Biological Proces | GO:01404 | integrated | 2.94E-04 | 3.40E-01 | 1.48E-02 | 1.13E-01 | 2 | 41 | FOS,CEBPB |
| M1/Control | Down | GO: Biological Proces | GO:19028 | positive re | 4.55E-04 | 5.26E-01 | 1.75E-02 | 1.34E-01 | 2 | 51 | EGR1,FOS |
| M1/Control | Down | GO: Biological Proces | GO:00359 | response t | 4.73E-04 | 5.47E-01 | 1.75E-02 | 1.34E-01 | 2 | 52 | FOS,REN |
| M1/Control | Down | GO: Biological Proces | GO:00713 | cellular re | 6.09E-04 | 7.04E-01 | 1.75E-02 | 1.34E-01 | 2 | 59 | FOS,ZFP36 |
| M1/Control | Down | GO: Biological Proces | GO:20006 | positive re | 6.30E-04 | 7.28E-01 | 1.75E-02 | 1.34E-01 | 2 | 60 | EGR1,FOS |
| M1/Control | Down | GO: Biological Proces | GO:00019 | response t | 7.17E-04 | 8.29E-01 | 1.84E-02 | 1.41E-01 | 2 | 64 | EGR1,NR4A1 |
| M1/Control | Down | GO: Biological Proces | GO:19028 | regulation | 8.33E-04 | 9.62E-01 | 1.90E-02 | 1.45E-01 | 2 | 69 | EGR1,FOS |
| M1/Control | Down | GO: Biological Proces | GO:00616 | miRNA tra | 8.57E-04 | 9.90E-01 | 1.90E-02 | 1.45E-01 | 2 | 70 | EGR1,FOS |
| M1/Control | Down | GO: Biological Proces | GO:00708 | response t | 8.81E-04 | 1.00E+00 | 1.90E-02 | 1.45E-01 | 2 | 71 | FOS,ZFP36 |
| M1/Control | Down | GO: Biological Proces | GO:00380 | p38MAPK | 9.06E-04 | 1.00E+00 | 1.90E-02 | 1.45E-01 | 2 | 72 | ZFP36,DUSP1 |
| M1/Control | Down | GO: Biological Proces | GO:00456 | positive re | 1.15E-03 | 1.00E+00 | 2.13E-02 | 1.63E-01 | 2 | 81 | ZFP36,CEBPB |
| M1/Control | Down | GO: Biological Proces | GO:00516 | response t | 1.17E-03 | 1.00E+00 | 2.13E-02 | 1.63E-01 | 2 | 82 | EGR1,NR4A1 |
| M1/Control | Down | GO: Biological Proces | GO:20006 | regulation | 1.23E-03 | 1.00E+00 | 2.13E-02 | 1.63E-01 | 2 | 84 | EGR1,FOS |
| M1/Control | Down | GO: Biological Proces | GO:00140 | response t | 1.44E-03 | 1.00E+00 | 2.35E-02 | 1.79E-01 | 2 | 91 | EGR1,NR4A1 |
| M1/Control | Down | GO: Biological Proces | GO:00703 | negative n | 1.67E-03 | 1.00E+00 | 2.47E-02 | 1.89E-01 | 2 | 98 | ATF3,DUSP1 |
| M1/Control | Down | GO: Biological Proces | GO:00105 | miRNA me | 1.67E-03 | 1.00E+00 | 2.47E-02 | 1.89E-01 | 2 | 98 | EGR1,FOS |
| M1/Control | Down | GO: Biological Proces | GO:00063 | DNA alkyl | 2.10E-03 | 1.00E+00 | 2.73E-02 | 2.08E-01 | 2 | 110 | FOS,CEBPB |
| M1/Control | Down | GO: Biological Proces | GO:00063 | DNA meth | 2.10E-03 | 1.00E+00 | 2.73E-02 | 2.08E-01 | 2 | 110 | FOS,CEBPB |
| M1/Control | Down | GO: Biological Proces | GO:00456 | regulation | 2.25E-03 | 1.00E+00 | 2.87E-02 | 2.19E-01 | 2 | 114 | FOS,CEBPB |
| M1/Control | Down | GO: Biological Proces | GO:00602 | hematopo | 2.53E-03 | 1.00E+00 | 2.87E-02 | 2.19E-01 | 2 | 121 | FOS,ZFP36 |
| M1/Control | Down | GO: Biological Proces | GO:00447 | DNA meth | 3.18E-03 | 1.00E+00 | 3.27E-02 | 2.50E-01 | 2 | 136 | FOS,CEBPB |
| M1/Control | Down | GO: Biological Proces | GO:19012 | positive re | 3.51E-03 | 1.00E+00 | 3.56E-02 | 2.72E-01 | 2 | 143 | EGR1,FOS |
| M1/Control | Down | GO: Biological Proces | GO:00063 | DNA mod | 4.21E-03 | 1.00E+00 | 3.73E-02 | 2.85E-01 | 2 | 157 | FOS,CEBPB |
| M1/Control | Down | GO: Biological Proces | GO:00443 | cellular re | 4.37E-03 | 1.00E+00 | 3.73E-02 | 2.85E-01 | 2 | 160 | ZFP36,NR4A1 |
| M1/Control | Down | GO: Biological Proces | GO:00030 | osteoclast | 4.48E-03 | 1.00E+00 | 3.73E-02 | 2.85E-01 | 2 | 162 | FOS,CEBPB |
| M1/Control | Down | GO: Biological Proces | GO:00076 | feeding be | 4.75E-03 | 1.00E+00 | 3.81E-02 | 2.91E-01 | 2 | 167 | FOS,REN |
| M1/Control | Down | GO: Biological Proces | GO:00717 | response t | 4.81E-03 | 1.00E+00 | 3.81E-02 | 2.91E-01 | 2 | 168 | ZFP36,NR4A1 |
| M1/Control | Down | GO: Biological Proces | GO:00713 | cellular re | 4.92E-03 | 1.00E+00 | 3.81E-02 | 2.91E-01 | 2 | 170 | EGR1,CEBPB |
| M1/Control | Down | GO: Biological Proces | GO:00455 | regulation | 4.98E-03 | 1.00E+00 | 3.81E-02 | 2.91E-01 | 2 | 171 | ZFP36,CEBPB |
| M1/Control | Down | GO: Biological Proces | GO:00076 | memory | 5.56E-03 | 1.00E+00 | 4.05E-02 | 3.09E-01 | 2 | 181 | EGR1,CEBPB |
| M1/Control | Down | GO: Biological Proces | GO:00027 | regulation | 6.11E-03 | 1.00E+00 | 4.25E-02 | 3.25E-01 | 2 | 190 | FOS,CEBPB |
| M1/Control | Down | GO: Biological Proces | GO:00016 | temperatu | 6.29E-03 | 1.00E+00 | 4.25E-02 | 3.25E-01 | 2 | 193 | EGR1,CEBPB |
| M1/Control | Down | GO: Biological Proces | GO:00515 | response t | 6.48E-03 | 1.00E+00 | 4.36E-02 | 3.33E-01 | 2 | 196 | FOS,DUSP1 |
| M1/Control | Down | GO: Biological Proces | GO:00705 | response t | 7.34E-03 | 1.00E+00 | 4.77E-02 | 3.64E-01 | 2 | 209 | EGR1,CEBPB |
| M1/M0 | Up | GO: Molecular Functi | GO:00860 | gap juncti | 1.20E-03 | 2.52E-01 | 1.48E-02 | 8.76E-02 | 1 | 1 | GJA5 |
| M1/M0 | Up | GO: Molecular Functi | GO:00860 | gap juncti | 1.20E-03 | 2.52E-01 | 1.48E-02 | 8.76E-02 | 1 | 1 | GJA5 |
| M1/M0 | Up | GO: Molecular Functi | GO:00020 | protease b | 1.31E-03 | 2.74E-01 | 1.52E-02 | 9.01E-02 | 3 | 182 | HSPG2,ECM1,ITGB1 |
| M1/M0 | Up | GO: Molecular Functi | GO:00154 | potassium | 2.35E-03 | 4.91E-01 | 2.50E-02 | 1.48E-01 | 2 | 60 | FLNA,SGK1 |
| M1/M0 | Up | GO: Molecular Functi | GO:00150 | corecepto | 2.51E-03 | 5.24E-01 | 2.50E-02 | 1.48E-01 | 2 | 62 | ENG,ITGB1 |
| M1/M0 | Up | GO: Molecular Functi | GO:00860 | gap juncti | 2.40E-03 | 5.02E-01 | 2.50E-02 | 1.48E-01 | 1 | 2 | GJA5 |
| M1/M0 | Up | GO: Molecular Functi | GO:00469 | protein di | 2.65E-03 | 5.53E-01 | 2.52E-02 | 1.49E-01 | 6 | 1218 | S100A6,FLNA,TCF4,ENG,ITGB1,H4C5 |
| M1/M0 | Up | GO: Molecular Functi | GO:00199 | protein kin | 3.07E-03 | 6.41E-01 | 2.79E-02 | 1.65E-01 | 5 | 855 | FLNA,CTNNB1,ITGB1,SGK1,MSN |
| M1/M0 | Up | GO: Molecular Functi | GO:00434 | 3-phospho | 3.60E-03 | 7.53E-01 | 3.14E-02 | 1.86E-01 | 1 | 3 | SGK1 |
| M1/M0 | Up | GO: Molecular Functi | GO:00199 | kinase bin | 4.86E-03 | 1.00E+00 | 3.66E-02 | 2.17E-01 | 5 | 952 | FLNA,CTNNB1,ITGB1,SGK1,MSN |
| M1/M0 | Up | GO: Molecular Functi | GO:00463 | SMAD bin | 4.98E-03 | 1.00E+00 | 3.66E-02 | 2.17E-01 | 2 | 88 | FLNA,CTNNB1 |
| M1/M0 | Up | GO: Biological Proces | GO:00519 | positive re | 2.32E-03 | 1.00E+00 | 1.97E-02 | 1.62E-01 | 1 | 2 | ITGB1 |
| M1/M0 | Up | GO: Biological Proces | GO:00617 | leukocyte | 2.33E-03 | 1.00E+00 | 1.97E-02 | 1.62E-01 | 2 | 62 | ITGB1,VCAM1 |
| M1/M0 | Up | GO: Biological Proces | GO:00323 | positive re | 2.39E-03 | 1.00E+00 | 2.01E-02 | 1.66E-01 | 3 | 233 | FLNA,CD81,MSN |
| M1/M0 | Up | GO: Biological Proces | GO:00972 | renal filtra | 2.41E-03 | 1.00E+00 | 2.02E-02 | 1.66E-01 | 2 | 63 | GJA5,CTNNB1 |
| M1/M0 | Up | GO: Biological Proces | GO:00457 | positive re | 2.45E-03 | 1.00E+00 | 2.04E-02 | 1.68E-01 | 3 | 235 | ECM1,ENG,ITGB1 |
| M1/M0 | Up | GO: Biological Proces | GO:19040 | positive re | 2.47E-03 | 1.00E+00 | 2.05E-02 | 1.69E-01 | 3 | 236 | ECM1,ENG,ITGB1 |
| M1/M0 | Up | GO: Biological Proces | GO:00350 | embryonic | 2.47E-03 | 1.00E+00 | 2.05E-02 | 1.69E-01 | 3 | 236 | GJA5,CTNNB1,ENG |
| M1/M0 | Up | GO: Biological Proces | GO:00607 | labyrinthin | 2.48E-03 | 1.00E+00 | 2.05E-02 | 1.69E-01 | 2 | 64 | ZFP36L1,VCAM1 |
| M1/M0 | Up | GO: Biological Proces | GO:00071 | enzyme-lin | 2.50E-03 | 1.00E+00 | 2.06E-02 | 1.70E-01 | 6 | 1250 | COL4A1,TCF4,CTNNB1,ENG,ITGB1,LRRC32 |
| M1/M0 | Up | GO: Biological Proces | GO:00430 | chordate e | 2.57E-03 | 1.00E+00 | 2.08E-02 | 1.71E-01 | 6 | 1257 | HSPG2,HEG1,CTNNB1,ZFP36L1,ITGB1,VCAM1 |
| M1/M0 | Up | GO: Biological Proces | GO:00016 | eye develo | 2.55E-03 | 1.00E+00 | 2.08E-02 | 1.71E-01 | 5 | 850 | COL4A1,S100A6,TCF4,CTNNB1,ITGB1 |
| M1/M0 | Up | GO: Biological Proces | GO:00613 | neural pre | 2.56E-03 | 1.00E+00 | 2.08E-02 | 1.71E-01 | 3 | 239 | FLNA,CTNNB1,ITGB1 |
| M1/M0 | Up | GO: Biological Proces | GO:00860 | cell comm | 2.56E-03 | 1.00E+00 | 2.08E-02 | 1.71E-01 | 2 | 65 | FLNA,GJA5 |
| M1/M0 | Up | GO: Biological Proces | GO:00613 | trabecula | 2.56E-03 | 1.00E+00 | 2.08E-02 | 1.71E-01 | 2 | 65 | HEG1,ENG |
| M1/M0 | Up | GO: Biological Proces | GO:01500 | visual syst | 2.59E-03 | 1.00E+00 | 2.08E-02 | 1.71E-01 | 5 | 853 | COL4A1,S100A6,TCF4,CTNNB1,ITGB1 |
| M1/M0 | Up | GO: Biological Proces | GO:00030 | circulatory | 2.60E-03 | 1.00E+00 | 2.08E-02 | 1.72E-01 | 5 | 854 | FLNA,GJA5,HEG1,ENG,SGK1 |
| M1/M0 | Up | GO: Biological Proces | GO:00508 | regulation | 2.68E-03 | 1.00E+00 | 2.14E-02 | 1.76E-01 | 4 | 514 | FLNA,GJA5,HEG1,LRRC32 |
| M1/M0 | Up | GO: Biological Proces | GO:00721 | mesenchy | 2.72E-03 | 1.00E+00 | 2.15E-02 | 1.77E-01 | 2 | 67 | CTNNB1,ENG |
| M1/M0 | Up | GO: Biological Proces | GO:00510 | negative n | 2.72E-03 | 1.00E+00 | 2.15E-02 | 1.77E-01 | 2 | 67 | HEG1,ITGB1 |
| M1/M0 | Up | GO: Biological Proces | GO:00328 | cellular re | 2.76E-03 | 1.00E+00 | 2.18E-02 | 1.80E-01 | 5 | 866 | CTNNB1,IGFBP7,ZFP36L1,SGK1,PGRMC2 |
| M1/M0 | Up | GO: Biological Proces | GO:00075 | myoblast f | 2.80E-03 | 1.00E+00 | 2.20E-02 | 1.81E-01 | 2 | 68 | CD81,ITGB1 |
| M1/M0 | Up | GO: Biological Proces | GO:00097 | embryo de | 2.83E-03 | 1.00E+00 | 2.21E-02 | 1.82E-01 | 6 | 1281 | HSPG2,HEG1,CTNNB1,ZFP36L1,ITGB1,VCAM1 |
| M1/M0 | Up | GO: Biological Proces | GO:00343 | cell junctic | 2.84E-03 | 1.00E+00 | 2.21E-02 | 1.82E-01 | 4 | 522 | GJA5,HEG1,CTNNB1,ITGB1 |
| M1/M0 | Up | GO: Biological Proces | GO:00082 | regulation | 2.82E-03 | 1.00E+00 | 2.21E-02 | 1.82E-01 | 3 | 247 | GJA5,ENG,SGK1 |
| M1/M0 | Up | GO: Biological Proces | GO:00466 | male sex c | 2.91E-03 | 1.00E+00 | 2.26E-02 | 1.86E-01 | 3 | 250 | FLNA,CTNNB1,ITGB1 |
| M1/M0 | Up | GO: Biological Proces | GO:00973 | response t | 3.04E-03 | 1.00E+00 | 2.34E-02 | 1.93E-01 | 4 | 532 | CTNNB1,IGFBP7,SGK1,VCAM1 |
| M1/M0 | Up | GO: Biological Proces | GO:19016 | cellular re | 3.12E-03 | 1.00E+00 | 2.39E-02 | 1.97E-01 | 4 | 536 | CTNNB1,ZFP36L1,SGK1,VCAM1 |
| M1/M0 | Up | GO: Biological Proces | GO:00071 | transformi | 3.11E-03 | 1.00E+00 | 2.39E-02 | 1.97E-01 | 3 | 256 | ENG,ITGB1,LRRC32 |
| M1/M0 | Up | GO: Biological Proces | GO:00302 | megakaryo | 3.13E-03 | 1.00E+00 | 2.39E-02 | 1.97E-01 | 2 | 72 | FLNA,H4C5 |
| M1/M0 | Up | GO: Biological Proces | GO:00109 | regulation | 3.31E-03 | 1.00E+00 | 2.42E-02 | 2.00E-01 | 4 | 545 | FLNA,CTNNB1,ITGB1,SGK1 |
| M1/M0 | Up | GO: Biological Proces | GO:00031 | heart valv | 3.30E-03 | 1.00E+00 | 2.42E-02 | 2.00E-01 | 2 | 74 | GJA5,ITGB1 |
| M1/M0 | Up | GO: Biological Proces | GO:00341 | heterotypi | 3.30E-03 | 1.00E+00 | 2.42E-02 | 2.00E-01 | 2 | 74 | ITGB1,VCAM1 |
| M1/M0 | Up | GO: Biological Proces | GO:00107 | regulation | 3.39E-03 | 1.00E+00 | 2.42E-02 | 2.00E-01 | 2 | 75 | ENG,ITGB1 |
| M1/M0 | Up | GO: Biological Proces | GO:00085 | visual lear | 3.48E-03 | 1.00E+00 | 2.42E-02 | 2.00E-01 | 2 | 76 | ITGB1,SGK1 |
| M1/M0 | Up | GO: Biological Proces | GO:00974 | cellular re | 3.47E-03 | 1.00E+00 | 2.42E-02 | 2.00E-01 | 1 | 3 | ZFP36L1 |
| M1/M0 | Up | GO: Biological Proces | GO:00360 | embryonic | 3.47E-03 | 1.00E+00 | 2.42E-02 | 2.00E-01 | 1 | 3 | CTNNB1 |
| M1/M0 | Up | GO: Biological Proces | GO:00610 | positive re | 3.47E-03 | 1.00E+00 | 2.42E-02 | 2.00E-01 | 1 | 3 | CTNNB1 |
| M1/M0 | Up | GO: Biological Proces | GO:19056 | regulation | 3.47E-03 | 1.00E+00 | 2.42E-02 | 2.00E-01 | 1 | 3 | CD81 |
| M1/M0 | Up | GO: Biological Proces | GO:00106 | cell comm | 3.47E-03 | 1.00E+00 | 2.42E-02 | 2.00E-01 | 1 | 3 | GJA5 |
| M1/M0 | Up | GO: Biological Proces | GO:20000 | positive re | 3.47E-03 | 1.00E+00 | 2.42E-02 | 2.00E-01 | 1 | 3 | CTNNB1 |
| M1/M0 | Up | GO: Biological Proces | GO:19015 | response t | 3.47E-03 | 1.00E+00 | 2.42E-02 | 2.00E-01 | 1 | 3 | ZFP36L1 |
| M1/M0 | Up | GO: Biological Proces | GO:19042 | regulation | 3.47E-03 | 1.00E+00 | 2.42E-02 | 2.00E-01 | 1 | 3 | SERINC1 |
| M1/M0 | Up | GO: Biological Proces | GO:19042 | positive re | 3.47E-03 | 1.00E+00 | 2.42E-02 | 2.00E-01 | 1 | 3 | SERINC1 |
| M1/M0 | Up | GO: Biological Proces | GO:00365 | astrocyte-c | 3.47E-03 | 1.00E+00 | 2.42E-02 | 2.00E-01 | 1 | 3 | CTNNB1 |
| M1/M0 | Up | GO: Biological Proces | GO:19019 | regulation | 3.47E-03 | 1.00E+00 | 2.42E-02 | 2.00E-01 | 1 | 3 | CTNNB1 |
| M1/M0 | Up | GO: Biological Proces | GO:00109 | positive re | 3.47E-03 | 1.00E+00 | 2.42E-02 | 2.00E-01 | 1 | 3 | CTNNB1 |
| M1/M0 | Up | GO: Biological Proces | GO:19045 | positive re | 3.47E-03 | 1.00E+00 | 2.42E-02 | 2.00E-01 | 1 | 3 | ZFP36L1 |
| M1/M0 | Up | GO: Biological Proces | GO:00309 | regulation | 3.47E-03 | 1.00E+00 | 2.42E-02 | 2.00E-01 | 1 | 3 | CTNNB1 |
| M1/M0 | Up | GO: Biological Proces | GO:00607 | positive re | 3.47E-03 | 1.00E+00 | 2.42E-02 | 2.00E-01 | 1 | 3 | CTNNB1 |
| M1/M0 | Up | GO: Biological Proces | GO:00357 | CD4-posit | 3.47E-03 | 1.00E+00 | 2.42E-02 | 2.00E-01 | 1 | 3 | CD81 |
| M1/M0 | Up | GO: Biological Proces | GO:00604 | lung induc | 3.47E-03 | 1.00E+00 | 2.42E-02 | 2.00E-01 | 1 | 3 | CTNNB1 |
| M1/M0 | Up | GO: Biological Proces | GO:00720 | renal inne | 3.47E-03 | 1.00E+00 | 2.42E-02 | 2.00E-01 | 1 | 3 | CTNNB1 |
| M1/M0 | Up | GO: Biological Proces | GO:00907 | adaptive i | 3.47E-03 | 1.00E+00 | 2.42E-02 | 2.00E-01 | 1 | 3 | CD81 |
| M1/M0 | Up | GO: Biological Proces | GO:00443 | canonical | 3.47E-03 | 1.00E+00 | 2.42E-02 | 2.00E-01 | 1 | 3 | CTNNB1 |
| M1/M0 | Up | GO: Biological Proces | GO:00604 | foregut re | 3.47E-03 | 1.00E+00 | 2.42E-02 | 2.00E-01 | 1 | 3 | CTNNB1 |
| M1/M0 | Up | GO: Biological Proces | GO:00604 | lung field | 3.47E-03 | 1.00E+00 | 2.42E-02 | 2.00E-01 | 1 | 3 | CTNNB1 |
| M1/M0 | Up | GO: Biological Proces | GO:00434 | protein kin | 3.58E-03 | 1.00E+00 | 2.49E-02 | 2.05E-01 | 3 | 269 | ZFP36L1,ENG,ITGB1 |
| M1/M0 | Up | GO: Biological Proces | GO:00300 | lymphocyt | 3.60E-03 | 1.00E+00 | 2.49E-02 | 2.05E-01 | 4 | 558 | CTNNB1,ZFP36L1,ITGB1,VCAM1 |
| M1/M0 | Up | GO: Biological Proces | GO:00074 | brain deve | 3.67E-03 | 1.00E+00 | 2.53E-02 | 2.08E-01 | 6 | 1349 | COL4A1,HSPG2,FLNA,TCF4,CTNNB1,ITGB1 |
| M1/M0 | Up | GO: Biological Proces | GO:00455 | negative n | 3.70E-03 | 1.00E+00 | 2.55E-02 | 2.10E-01 | 5 | 927 | CTNNB1,RCAN1,ZFP36L1,ITGB1,H4C5 |
| M1/M0 | Up | GO: Biological Proces | GO:00071 | cell-matrix | 3.73E-03 | 1.00E+00 | 2.56E-02 | 2.11E-01 | 3 | 273 | CTNNB1,ITGB1,VCAM1 |
| M1/M0 | Up | GO: Biological Proces | GO:00068 | ion transp | 3.76E-03 | 1.00E+00 | 2.56E-02 | 2.11E-01 | 7 | 1832 | S100A6,FLNA,GJA5,CTNNB1,ITGB1,SGK1,PGRMC2 |
| M1/M0 | Up | GO: Biological Proces | GO:19038 | positive re | 3.77E-03 | 1.00E+00 | 2.56E-02 | 2.11E-01 | 4 | 565 | FLNA,CD81,ITGB1,MSN |
| M1/M0 | Up | GO: Biological Proces | GO:00019 | lymph ves | 3.85E-03 | 1.00E+00 | 2.61E-02 | 2.15E-01 | 2 | 80 | HEG1,VCAM1 |
| M1/M0 | Up | GO: Biological Proces | GO:00506 | positive re | 3.93E-03 | 1.00E+00 | 2.66E-02 | 2.19E-01 | 3 | 278 | COL18A1,CTNNB1,ECM1 |
| M1/M0 | Up | GO: Biological Proces | GO:00431 | regulation | 3.97E-03 | 1.00E+00 | 2.67E-02 | 2.20E-01 | 3 | 279 | FLNA,CTNNB1,ECM1 |
| M1/M0 | Up | GO: Biological Proces | GO:00488 | sensory sy | 4.02E-03 | 1.00E+00 | 2.69E-02 | 2.22E-01 | 5 | 945 | COL4A1,S100A6,TCF4,CTNNB1,ITGB1 |
| M1/M0 | Up | GO: Biological Proces | GO:00420 | wound he | 4.01E-03 | 1.00E+00 | 2.69E-02 | 2.22E-01 | 4 | 575 | FLNA,ENG,ITGB1,LRRC32 |
| M1/M0 | Up | GO: Biological Proces | GO:00068 | potassium | 4.01E-03 | 1.00E+00 | 2.69E-02 | 2.22E-01 | 3 | 280 | FLNA,GJA5,ITGB1 |
| M1/M0 | Up | GO: Biological Proces | GO:00352 | organ grov | 4.09E-03 | 1.00E+00 | 2.72E-02 | 2.24E-01 | 3 | 282 | HEG1,CTNNB1,ECM1 |
| M1/M0 | Up | GO: Biological Proces | GO:00708 | bicellular t | 4.13E-03 | 1.00E+00 | 2.74E-02 | 2.26E-01 | 2 | 83 | CTNNB1,ITGB1 |
| M1/M0 | Up | GO: Biological Proces | GO:00713 | cellular re | 4.15E-03 | 1.00E+00 | 2.75E-02 | 2.26E-01 | 5 | 952 | ZFP36L1,ITGB1,SGK1,PGRMC2,MSN |
| M1/M0 | Up | GO: Biological Proces | GO:00026 | regulation | 4.17E-03 | 1.00E+00 | 2.75E-02 | 2.27E-01 | 3 | 284 | CD81,ECM1,MSN |
| M1/M0 | Up | GO: Biological Proces | GO:00429 | regulation | 4.59E-03 | 1.00E+00 | 2.76E-02 | 2.27E-01 | 7 | 1899 | FLNA,COL18A1,CTNNB1,ZFP36L1,ENG,ITGB1,SGK1 |
| M1/M0 | Up | GO: Biological Proces | GO:00301 | regulation | 4.52E-03 | 1.00E+00 | 2.76E-02 | 2.27E-01 | 5 | 971 | FLNA,CD81,ITGB1,VCAM1,LRRC32 |
| M1/M0 | Up | GO: Biological Proces | GO:00457 | positive re | 4.34E-03 | 1.00E+00 | 2.76E-02 | 2.27E-01 | 4 | 588 | FLNA,CD81,ITGB1,VCAM1 |
| M1/M0 | Up | GO: Biological Proces | GO:00508 | synapse o | 4.48E-03 | 1.00E+00 | 2.76E-02 | 2.27E-01 | 4 | 593 | COL4A1,FLNA,CTNNB1,ITGB1 |
| M1/M0 | Up | GO: Biological Proces | GO:00860 | cardiac mu | 4.23E-03 | 1.00E+00 | 2.76E-02 | 2.27E-01 | 2 | 84 | FLNA,GJA5 |
| M1/M0 | Up | GO: Biological Proces | GO:00343 | protein loc | 4.23E-03 | 1.00E+00 | 2.76E-02 | 2.27E-01 | 2 | 84 | FLNA,CTNNB1 |
| M1/M0 | Up | GO: Biological Proces | GO:00989 | regulation | 4.23E-03 | 1.00E+00 | 2.76E-02 | 2.27E-01 | 2 | 84 | FLNA,GJA5 |
| M1/M0 | Up | GO: Biological Proces | GO:00359 | cellular re | 4.33E-03 | 1.00E+00 | 2.76E-02 | 2.27E-01 | 2 | 85 | TCF4,VCAM1 |
| M1/M0 | Up | GO: Biological Proces | GO:00860 | cardiac mu | 4.33E-03 | 1.00E+00 | 2.76E-02 | 2.27E-01 | 2 | 85 | FLNA,GJA5 |
| M1/M0 | Up | GO: Biological Proces | GO:20004 | regulation | 4.33E-03 | 1.00E+00 | 2.76E-02 | 2.27E-01 | 2 | 85 | ECM1,MSN |
| M1/M0 | Up | GO: Biological Proces | GO:00431 | receptor c | 4.43E-03 | 1.00E+00 | 2.76E-02 | 2.27E-01 | 2 | 86 | FLNA,CD81 |
| M1/M0 | Up | GO: Biological Proces | GO:00726 | T cell migr | 4.53E-03 | 1.00E+00 | 2.76E-02 | 2.27E-01 | 2 | 87 | ECM1,MSN |
| M1/M0 | Up | GO: Biological Proces | GO:00019 | blood ves | 4.53E-03 | 1.00E+00 | 2.76E-02 | 2.27E-01 | 2 | 87 | FLNA,BGN |
| M1/M0 | Up | GO: Biological Proces | GO:00451 | cellular ex | 4.63E-03 | 1.00E+00 | 2.76E-02 | 2.27E-01 | 2 | 88 | ITGB1,VCAM1 |
| M1/M0 | Up | GO: Biological Proces | GO:01500 | glial cell-n | 4.63E-03 | 1.00E+00 | 2.76E-02 | 2.27E-01 | 1 | 4 | CTNNB1 |
| M1/M0 | Up | GO: Biological Proces | GO:00704 | T cell aggr | 4.63E-03 | 1.00E+00 | 2.76E-02 | 2.27E-01 | 1 | 4 | MSN |
| M1/M0 | Up | GO: Biological Proces | GO:19042 | positive re | 4.63E-03 | 1.00E+00 | 2.76E-02 | 2.27E-01 | 1 | 4 | SERINC1 |
| M1/M0 | Up | GO: Biological Proces | GO:00032 | atrial vent | 4.63E-03 | 1.00E+00 | 2.76E-02 | 2.27E-01 | 1 | 4 | GJA5 |
| M1/M0 | Up | GO: Biological Proces | GO:00611 | fungiform | 4.63E-03 | 1.00E+00 | 2.76E-02 | 2.27E-01 | 1 | 4 | CTNNB1 |
| M1/M0 | Up | GO: Biological Proces | GO:00032 | cell migrat | 4.63E-03 | 1.00E+00 | 2.76E-02 | 2.27E-01 | 1 | 4 | ENG |
| M1/M0 | Up | GO: Biological Proces | GO:00071 | calcium-in | 4.63E-03 | 1.00E+00 | 2.76E-02 | 2.27E-01 | 1 | 4 | ITGB1 |
| M1/M0 | Up | GO: Biological Proces | GO:01501 | reactive gl | 4.63E-03 | 1.00E+00 | 2.76E-02 | 2.27E-01 | 1 | 4 | ITGB1 |
| M1/M0 | Up | GO: Biological Proces | GO:00334 | negative n | 4.63E-03 | 1.00E+00 | 2.76E-02 | 2.27E-01 | 1 | 4 | CTNNB1 |
| M1/M0 | Up | GO: Biological Proces | GO:00109 | regulation | 4.63E-03 | 1.00E+00 | 2.76E-02 | 2.27E-01 | 1 | 4 | CTNNB1 |
| M1/M0 | Up | GO: Biological Proces | GO:00020 | positive re | 4.63E-03 | 1.00E+00 | 2.76E-02 | 2.27E-01 | 1 | 4 | ITGB1 |
| M1/M0 | Up | GO: Biological Proces | GO:19049 | canonical | 4.63E-03 | 1.00E+00 | 2.76E-02 | 2.27E-01 | 1 | 4 | CTNNB1 |
| M1/M0 | Up | GO: Biological Proces | GO:00989 | regulation | 4.63E-03 | 1.00E+00 | 2.76E-02 | 2.27E-01 | 1 | 4 | GJA5 |
| M1/M0 | Up | GO: Biological Proces | GO:00434 | glucocorti | 4.63E-03 | 1.00E+00 | 2.76E-02 | 2.27E-01 | 1 | 4 | SGK1 |
| M1/M0 | Up | GO: Biological Proces | GO:00860 | AV node c | 4.63E-03 | 1.00E+00 | 2.76E-02 | 2.27E-01 | 1 | 4 | GJA5 |
| M1/M0 | Up | GO: Biological Proces | GO:00860 | Purkinje m | 4.63E-03 | 1.00E+00 | 2.76E-02 | 2.27E-01 | 1 | 4 | GJA5 |
| M1/M0 | Up | GO: Biological Proces | GO:20002 | negative n | 4.63E-03 | 1.00E+00 | 2.76E-02 | 2.27E-01 | 1 | 4 | HEG1 |
| M1/M0 | Up | GO: Biological Proces | GO:19053 | regulation | 4.63E-03 | 1.00E+00 | 2.76E-02 | 2.27E-01 | 1 | 4 | ENG |
| M1/M0 | Up | GO: Biological Proces | GO:00074 | glial cell fa | 4.63E-03 | 1.00E+00 | 2.76E-02 | 2.27E-01 | 1 | 4 | CTNNB1 |
| M1/M0 | Up | GO: Biological Proces | GO:00519 | regulation | 4.63E-03 | 1.00E+00 | 2.76E-02 | 2.27E-01 | 1 | 4 | ITGB1 |
| M1/M0 | Up | GO: Biological Proces | GO:00519 | regulation | 4.63E-03 | 1.00E+00 | 2.76E-02 | 2.27E-01 | 1 | 4 | ITGB1 |
| M1/M0 | Up | GO: Biological Proces | GO:00068 | cation tran | 4.66E-03 | 1.00E+00 | 2.77E-02 | 2.28E-01 | 6 | 1416 | FLNA,GJA5,CTNNB1,ITGB1,SGK1,PGRMC2 |
| M1/M0 | Up | GO: Biological Proces | GO:00603 | head deve | 4.72E-03 | 1.00E+00 | 2.80E-02 | 2.30E-01 | 6 | 1420 | COL4A1,HSPG2,FLNA,TCF4,CTNNB1,ITGB1 |
| M1/M0 | Up | GO: Biological Proces | GO:00099 | cell-cell re | 4.74E-03 | 1.00E+00 | 2.80E-02 | 2.30E-01 | 2 | 89 | CD81,MSN |
| M1/M0 | Up | GO: Biological Proces | GO:00484 | autonomic | 4.84E-03 | 1.00E+00 | 2.83E-02 | 2.33E-01 | 2 | 90 | CTNNB1,VCAM1 |
| M1/M0 | Up | GO: Biological Proces | GO:00032 | ventricular | 4.84E-03 | 1.00E+00 | 2.83E-02 | 2.33E-01 | 2 | 90 | GJA5,HEG1 |
| M1/M0 | Up | GO: Biological Proces | GO:01201 | tight junct | 4.84E-03 | 1.00E+00 | 2.83E-02 | 2.33E-01 | 2 | 90 | CTNNB1,ITGB1 |
| M1/M0 | Up | GO: Biological Proces | GO:00604 | cardiac se | 4.84E-03 | 1.00E+00 | 2.83E-02 | 2.33E-01 | 2 | 90 | GJA5,ENG |
| M1/M0 | Up | GO: Biological Proces | GO:00456 | regulation | 4.99E-03 | 1.00E+00 | 2.91E-02 | 2.40E-01 | 3 | 303 | CTNNB1,ZFP36L1,H4C5 |
| M1/M0 | Up | GO: Biological Proces | GO:01402 | cell-cell fu | 5.05E-03 | 1.00E+00 | 2.91E-02 | 2.40E-01 | 2 | 92 | CD81,ITGB1 |
| M1/M0 | Up | GO: Biological Proces | GO:00432 | apical junc | 5.05E-03 | 1.00E+00 | 2.91E-02 | 2.40E-01 | 2 | 92 | CTNNB1,ITGB1 |
| M1/M0 | Up | GO: Biological Proces | GO:00074 | neuroblast | 5.05E-03 | 1.00E+00 | 2.91E-02 | 2.40E-01 | 2 | 92 | CTNNB1,ITGB1 |
| M1/M0 | Up | GO: Biological Proces | GO:00007 | syncytium | 5.05E-03 | 1.00E+00 | 2.91E-02 | 2.40E-01 | 2 | 92 | CD81,ITGB1 |
| M1/M0 | Up | GO: Biological Proces | GO:00713 | cellular re | 5.08E-03 | 1.00E+00 | 2.92E-02 | 2.41E-01 | 3 | 305 | ZFP36L1,SGK1,PGRMC2 |
| M1/M0 | Up | GO: Biological Proces | GO:19031 | mononucl | 5.21E-03 | 1.00E+00 | 2.98E-02 | 2.46E-01 | 4 | 619 | CTNNB1,ZFP36L1,ITGB1,VCAM1 |
| M1/M0 | Up | GO: Biological Proces | GO:00430 | regulation | 5.23E-03 | 1.00E+00 | 2.99E-02 | 2.46E-01 | 7 | 1944 | FLNA,COL18A1,CTNNB1,ZFP36L1,ENG,ITGB1,SGK1 |
| M1/M0 | Up | GO: Biological Proces | GO:00106 | positive re | 5.58E-03 | 1.00E+00 | 3.11E-02 | 2.56E-01 | 6 | 1469 | CD81,HEG1,CTNNB1,ENG,LRRC32,MSN |
| M1/M0 | Up | GO: Biological Proces | GO:00512 | negative n | 5.63E-03 | 1.00E+00 | 3.11E-02 | 2.56E-01 | 6 | 1472 | HSPG2,TCF4,GJA5,CTNNB1,ECM1,LRRC32 |
| M1/M0 | Up | GO: Biological Proces | GO:00513 | response t | 5.66E-03 | 1.00E+00 | 3.11E-02 | 2.56E-01 | 3 | 317 | IGFBP7,ZFP36L1,SGK1 |
| M1/M0 | Up | GO: Biological Proces | GO:01201 | tight junct | 5.49E-03 | 1.00E+00 | 3.11E-02 | 2.56E-01 | 2 | 96 | CTNNB1,ITGB1 |
| M1/M0 | Up | GO: Cellular Compon | GO:00703 | beta-cater | 3.78E-06 | 8.73E-04 | 1.25E-04 | 7.51E-04 | 2 | 3 | TCF4,CTNNB1 |
| M1/M0 | Up | GO: Cellular Compon | GO:00716 | catenin-TC | 7.55E-06 | 1.74E-03 | 2.18E-04 | 1.31E-03 | 2 | 4 | TCF4,CTNNB1 |
| M1/M0 | Up | GO: Cellular Compon | GO:00986 | basement | 5.64E-05 | 1.30E-02 | 1.17E-03 | 7.03E-03 | 2 | 10 | COL4A1,COL18A1 |
| M1/M0 | Up | GO: Cellular Compon | GO:19909 | beta-cater | 1.14E-04 | 2.63E-02 | 1.88E-03 | 1.13E-02 | 2 | 14 | TCF4,CTNNB1 |
| M1/M0 | Up | GO: Cellular Compon | GO:00315 | microvillus | 1.38E-03 | 3.18E-01 | 1.18E-02 | 7.09E-02 | 2 | 48 | CTNNB1,MSN |
| M1/M0 | Up | GO: Cellular Compon | GO:00431 | dendritic s | 3.15E-03 | 7.28E-01 | 1.89E-02 | 1.14E-01 | 2 | 73 | FLNA,CTNNB1 |
| M1/M0 | Up | GO: Cellular Compon | GO:00055 | collagen tr | 5.49E-03 | 1.00E+00 | 2.64E-02 | 1.59E-01 | 2 | 97 | COL4A1,COL18A1 |
| M1/M0 | Up | GO: Cellular Compon | GO:00432 | lysosomal | 6.52E-03 | 1.00E+00 | 2.79E-02 | 1.68E-01 | 2 | 106 | HSPG2,BGN |
| M1/M0 | Up | GO: Cellular Compon | GO:00057 | Golgi lum | 7.00E-03 | 1.00E+00 | 2.94E-02 | 1.77E-01 | 2 | 110 | HSPG2,BGN |
| M1/M0 | Up | GO: Cellular Compon | GO:00986 | Schaffer c | 7.76E-03 | 1.00E+00 | 3.19E-02 | 1.92E-01 | 2 | 116 | CTNNB1,ITGB1 |
| M1/M0 | Up | GO: Cellular Compon | GO:00725 | blood mic | 1.21E-02 | 1.00E+00 | 4.29E-02 | 2.58E-01 | 2 | 146 | ENG,MSN |
| M1/M0 | Up | GO: Cellular Compon | GO:00300 | Z disc | 1.45E-02 | 1.00E+00 | 4.75E-02 | 2.86E-01 | 2 | 161 | FLNA,CTNNB1 |
| M1/Control | Up | GO: Molecular Functi | GO:00327 | GTPase ac | 4.16E-03 | 1.00E+00 | 2.62E-02 | 1.74E-01 | 2 | 25 | FMNL3,PLXNB1 |
| M1/Control | Up | GO: Molecular Functi | GO:00018 | opsonin b | 4.16E-03 | 1.00E+00 | 2.62E-02 | 1.74E-01 | 2 | 25 | CFH,ITGAV |
| M1/Control | Up | GO: Molecular Functi | GO:00504 | transformi | 4.84E-03 | 1.00E+00 | 3.00E-02 | 2.00E-01 | 2 | 27 | THBS1,ITGAV |
| M1/Control | Up | GO: Molecular Functi | GO:00469 | protein he | 5.84E-03 | 1.00E+00 | 3.57E-02 | 2.38E-01 | 6 | 423 | TPM1,BMP2,NFE2L1,H4C5,ITGA3,ITGB1 |
| M1/Control | Up | GO: Molecular Functi | GO:00433 | heparan s | 5.96E-03 | 1.00E+00 | 3.59E-02 | 2.39E-01 | 2 | 30 | CFH,APOE |
| M1/Control | Up | GO: Molecular Functi | GO:00431 | alcohol bi | 7.60E-03 | 1.00E+00 | 3.89E-02 | 2.58E-01 | 3 | 104 | NFE2L1,CDIPT,VDAC2 |
| M1/Control | Up | GO: Molecular Functi | GO:01403 | solute:anic | 6.76E-03 | 1.00E+00 | 3.89E-02 | 2.58E-01 | 2 | 32 | SLC22A8,SLC25A6 |
| M1/Control | Up | GO: Molecular Functi | GO:00506 | nuclear an | 7.61E-03 | 1.00E+00 | 3.89E-02 | 2.58E-01 | 2 | 34 | TCF21,PARK7 |
| M1/Control | Up | GO: Molecular Functi | GO:00507 | low-densit | 7.61E-03 | 1.00E+00 | 3.89E-02 | 2.58E-01 | 2 | 34 | LRP2,APOE |
| M1/Control | Up | GO: Molecular Functi | GO:00718 | lipoprotein | 7.61E-03 | 1.00E+00 | 3.89E-02 | 2.58E-01 | 2 | 34 | THBS1,APOE |
| M1/Control | Up | GO: Molecular Functi | GO:00718 | protein-lip | 7.61E-03 | 1.00E+00 | 3.89E-02 | 2.58E-01 | 2 | 34 | THBS1,APOE |
| M1/Control | Up | GO: Molecular Functi | GO:01402 | mitochond | 7.70E-03 | 1.00E+00 | 3.89E-02 | 2.58E-01 | 1 | 2 | ATP5IF1 |
| M1/Control | Up | GO: Molecular Functi | GO:00364 | L-dopa de | 7.70E-03 | 1.00E+00 | 3.89E-02 | 2.58E-01 | 1 | 2 | PARK7 |
| M1/Control | Up | GO: Molecular Functi | GO:19900 | glycerol d | 7.70E-03 | 1.00E+00 | 3.89E-02 | 2.58E-01 | 1 | 2 | AKR1A1 |
| M1/Control | Up | GO: Molecular Functi | GO:00040 | aromatic-L | 7.70E-03 | 1.00E+00 | 3.89E-02 | 2.58E-01 | 1 | 2 | PARK7 |
| M1/Control | Up | GO: Molecular Functi | GO:00479 | glycerol d | 7.70E-03 | 1.00E+00 | 3.89E-02 | 2.58E-01 | 1 | 2 | AKR1A1 |
| M1/Control | Up | GO: Molecular Functi | GO:00050 | laminin re | 7.70E-03 | 1.00E+00 | 3.89E-02 | 2.58E-01 | 1 | 2 | BCAM |
| M1/Control | Up | GO: Molecular Functi | GO:00104 | nerve grov | 7.70E-03 | 1.00E+00 | 3.89E-02 | 2.58E-01 | 1 | 2 | SORT1 |
| M1/Control | Up | GO: Molecular Functi | GO:00045 | tyrosine 3- | 7.70E-03 | 1.00E+00 | 3.89E-02 | 2.58E-01 | 1 | 2 | PARK7 |
| M1/Control | Up | GO: Molecular Functi | GO:00015 | amyloid-b | 8.86E-03 | 1.00E+00 | 4.42E-02 | 2.94E-01 | 3 | 110 | LRP2,APOE,ITM2B |
| M1/Control | Up | GO: Molecular Functi | GO:00430 | gamma-tu | 9.92E-03 | 1.00E+00 | 4.79E-02 | 3.18E-01 | 2 | 39 | GLYAT,NDRG1 |
| M1/Control | Up | GO: Molecular Functi | GO:00703 | lipoprotein | 9.92E-03 | 1.00E+00 | 4.79E-02 | 3.18E-01 | 2 | 39 | LRP2,APOE |
| M1/Control | Up | GO: Molecular Functi | GO:00001 | microfilam | 9.92E-03 | 1.00E+00 | 4.79E-02 | 3.18E-01 | 2 | 39 | MYH10,MYL6 |
| M1/Control | Up | GO: Biological Proces | GO:00301 | regulation | 5.15E-03 | 1.00E+00 | 4.06E-02 | 3.51E-01 | 3 | 95 | THBS1,CD9,APOE |
| M1/Control | Up | GO: Biological Proces | GO:19029 | regulation | 5.31E-03 | 1.00E+00 | 4.15E-02 | 3.59E-01 | 6 | 436 | TPM1,PARK7,EFEMP2,CGNL1,APOE,CDC42EP3 |
| M1/Control | Up | GO: Biological Proces | GO:00032 | cardiac ve | 5.31E-03 | 1.00E+00 | 4.15E-02 | 3.59E-01 | 3 | 96 | TPM1,DSP,LRP2 |
| M1/Control | Up | GO: Biological Proces | GO:00069 | syncytium | 5.31E-03 | 1.00E+00 | 4.15E-02 | 3.59E-01 | 3 | 96 | MYOF,CD9,ITGB1 |
| M1/Control | Up | GO: Biological Proces | GO:00970 | dendritic s | 5.40E-03 | 1.00E+00 | 4.20E-02 | 3.63E-01 | 2 | 30 | ITGA3,APOE |
| M1/Control | Up | GO: Biological Proces | GO:00450 | plasma me | 5.40E-03 | 1.00E+00 | 4.20E-02 | 3.63E-01 | 2 | 30 | CD9,ITGA3 |
| M1/Control | Up | GO: Biological Proces | GO:00157 | organic an | 5.42E-03 | 1.00E+00 | 4.21E-02 | 3.64E-01 | 6 | 438 | ATP5PF,SLC22A8,SLC25A6,NFE2L1,LRP2,ITGB1 |
| M1/Control | Up | GO: Biological Proces | GO:00601 | maternal p | 5.46E-03 | 1.00E+00 | 4.22E-02 | 3.65E-01 | 3 | 97 | PARK7,ITGA3,ITGB1 |
| M1/Control | Up | GO: Biological Proces | GO:19000 | regulation | 5.46E-03 | 1.00E+00 | 4.22E-02 | 3.65E-01 | 3 | 97 | THBS1,CD9,APOE |
| M1/Control | Up | GO: Biological Proces | GO:00905 | regulation | 5.62E-03 | 1.00E+00 | 4.33E-02 | 3.74E-01 | 3 | 98 | ATP5IF1,SLC25A6,VDAC2 |
| M1/Control | Up | GO: Biological Proces | GO:00444 | entry into | 5.66E-03 | 1.00E+00 | 4.34E-02 | 3.75E-01 | 4 | 194 | DAG1,ITGAV,ITGB1,ITGB3 |
| M1/Control | Up | GO: Biological Proces | GO:00073 | single fert | 5.66E-03 | 1.00E+00 | 4.34E-02 | 3.75E-01 | 4 | 194 | CD9,PARK7,ITGA3,VDAC2 |
| M1/Control | Up | GO: Biological Proces | GO:00469 | intracellula | 5.70E-03 | 1.00E+00 | 4.35E-02 | 3.76E-01 | 14 | 1811 | SORT1,MYH10,ATP5IF1,SLC25A6,PARK7,LRP2,MACF1,LAMP2,APOE,RAB2A,ITGB1,PTTG1IP,VDAC2,TMED3 |
| M1/Control | Up | GO: Biological Proces | GO:00091 | nucleoside | 5.72E-03 | 1.00E+00 | 4.35E-02 | 3.76E-01 | 5 | 311 | CFH,ATP5PF,ATP5IF1,ATP5F1A,NDUFS5 |
| M1/Control | Up | GO: Biological Proces | GO:00199 | lipid stora | 5.78E-03 | 1.00E+00 | 4.35E-02 | 3.76E-01 | 3 | 99 | APOE,ITGAV,ITGB3 |
| M1/Control | Up | GO: Biological Proces | GO:00488 | neural nuc | 5.78E-03 | 1.00E+00 | 4.35E-02 | 3.76E-01 | 3 | 99 | ATP5PF,BMP2,NDRG2 |
| M1/Control | Up | GO: Biological Proces | GO:00439 | cellular co | 5.78E-03 | 1.00E+00 | 4.35E-02 | 3.76E-01 | 3 | 99 | CDIPT,ITGA3,APOE |
| M1/Control | Up | GO: Biological Proces | GO:00607 | regulation | 5.76E-03 | 1.00E+00 | 4.35E-02 | 3.76E-01 | 2 | 31 | TCF21,PARK7 |
| M1/Control | Up | GO: Biological Proces | GO:00328 | negative n | 5.76E-03 | 1.00E+00 | 4.35E-02 | 3.76E-01 | 2 | 31 | ATP5PF,THBS1 |
| M1/Control | Up | GO: Biological Proces | GO:00108 | negative n | 5.76E-03 | 1.00E+00 | 4.35E-02 | 3.76E-01 | 2 | 31 | BMP2,APOE |
| M1/Control | Up | GO: Biological Proces | GO:00328 | regulation | 5.94E-03 | 1.00E+00 | 4.46E-02 | 3.86E-01 | 3 | 100 | ATP5PF,THBS1,ITGB1 |
| M1/Control | Up | GO: Biological Proces | GO:00620 | regulation | 6.10E-03 | 1.00E+00 | 4.54E-02 | 3.92E-01 | 6 | 449 | BMP2,ATP5IF1,NFE2L1,PARK7,AKR1A1,APOE |
| M1/Control | Up | GO: Biological Proces | GO:00508 | regulation | 6.11E-03 | 1.00E+00 | 4.54E-02 | 3.92E-01 | 3 | 101 | THBS1,CD9,APOE |
| M1/Control | Up | GO: Biological Proces | GO:00429 | amyloid p | 6.11E-03 | 1.00E+00 | 4.54E-02 | 3.92E-01 | 3 | 101 | KLK6,APOE,ITM2B |
| M1/Control | Up | GO: Biological Proces | GO:00105 | negative n | 6.11E-03 | 1.00E+00 | 4.54E-02 | 3.92E-01 | 3 | 101 | THBS1,PTPRM,APOE |
| M1/Control | Up | GO: Biological Proces | GO:19010 | regulation | 6.13E-03 | 1.00E+00 | 4.54E-02 | 3.92E-01 | 2 | 32 | ATP5IF1,SLC25A6 |
| M1/Control | Up | GO: Biological Proces | GO:19021 | cellular re | 6.13E-03 | 1.00E+00 | 4.54E-02 | 3.92E-01 | 2 | 32 | THBS1,ATP5F1A |
| M1/Control | Up | GO: Biological Proces | GO:00703 | ERK1 and | 6.23E-03 | 1.00E+00 | 4.60E-02 | 3.98E-01 | 6 | 451 | BMP2,APOE,ITGAV,NDRG2,ITGB1,ITGB3 |
| M1/Control | Up | GO: Biological Proces | GO:00003 | response t | 6.27E-03 | 1.00E+00 | 4.60E-02 | 3.98E-01 | 5 | 318 | TPM1,CFH,PARK7,PRDX1,APOE |
| M1/Control | Up | GO: Biological Proces | GO:00350 | regulation | 6.28E-03 | 1.00E+00 | 4.60E-02 | 3.98E-01 | 3 | 102 | ITGA3,APOE,ITGB1 |
| M1/Control | Up | GO: Biological Proces | GO:00488 | artery mor | 6.28E-03 | 1.00E+00 | 4.60E-02 | 3.98E-01 | 3 | 102 | EFEMP2,LRP2,APOE |
| M1/Control | Up | GO: Biological Proces | GO:00068 | ion transp | 6.30E-03 | 1.00E+00 | 4.61E-02 | 3.99E-01 | 14 | 1832 | S100A6,ATP5PF,THY1,SLC22A8,SLC12A4,SLC25A6,NFE2L1,PARK7,LRP2,ITGAV,ITGB1,ITGB3,ATP5F1A,VDAC2 |
| M1/Control | Up | GO: Biological Proces | GO:00400 | negative n | 6.51E-03 | 1.00E+00 | 4.73E-02 | 4.09E-01 | 2 | 33 | THBS1,SULF1 |
| M1/Control | Up | GO: Biological Proces | GO:00439 | ossificatio | 6.51E-03 | 1.00E+00 | 4.73E-02 | 4.09E-01 | 2 | 33 | BMP2,PLXNB1 |
| M1/Control | Up | GO: Biological Proces | GO:00550 | ventricular | 6.51E-03 | 1.00E+00 | 4.73E-02 | 4.09E-01 | 2 | 33 | BMP2,MYH10 |
| M1/Control | Up | GO: Biological Proces | GO:00080 | blood circ | 7.09E-03 | 1.00E+00 | 4.76E-02 | 4.11E-01 | 8 | 769 | TPM1,ATP5PF,MYOF,PTPRM,DSP,SULF1,APOE,VDAC2 |
| M1/Control | Up | GO: Biological Proces | GO:00550 | nucleobas | 7.25E-03 | 1.00E+00 | 4.76E-02 | 4.11E-01 | 8 | 772 | CFH,GLYAT,ATP5PF,ATP5IF1,PARK7,LDHB,ATP5F1A,NDUFS5 |
| M1/Control | Up | GO: Biological Proces | GO:20012 | regulation | 6.84E-03 | 1.00E+00 | 4.76E-02 | 4.11E-01 | 6 | 460 | THBS1,SLC25A6,PARK7,ITGAV,PTTG1IP,VDAC2 |
| M1/Control | Up | GO: Biological Proces | GO:00329 | regulation | 7.28E-03 | 1.00E+00 | 4.76E-02 | 4.11E-01 | 6 | 466 | TPM1,DSP,BST2,CGNL1,ITGB3,CDC42EP3 |
| M1/Control | Up | GO: Biological Proces | GO:00486 | reproducti | 7.28E-03 | 1.00E+00 | 4.76E-02 | 4.11E-01 | 6 | 466 | TCF21,SULF1,GAS2,LRP2,ITGAV,ITGB1 |
| M1/Control | Up | GO: Biological Proces | GO:19029 | positive re | 6.74E-03 | 1.00E+00 | 4.76E-02 | 4.11E-01 | 4 | 204 | TPM1,EFEMP2,APOE,CDC42EP3 |
| M1/Control | Up | GO: Biological Proces | GO:00508 | regulation | 6.62E-03 | 1.00E+00 | 4.76E-02 | 4.11E-01 | 3 | 104 | BMP2,IGFBP7,APOE |
| M1/Control | Up | GO: Biological Proces | GO:00610 | negative n | 6.62E-03 | 1.00E+00 | 4.76E-02 | 4.11E-01 | 3 | 104 | THBS1,CD9,APOE |
| M1/Control | Up | GO: Biological Proces | GO:00456 | regulation | 6.62E-03 | 1.00E+00 | 4.76E-02 | 4.11E-01 | 3 | 104 | BMP2,DAG1,TNFRSF21 |
| M1/Control | Up | GO: Biological Proces | GO:00107 | regulation | 6.90E-03 | 1.00E+00 | 4.76E-02 | 4.11E-01 | 2 | 34 | ITGAV,ITGB3 |
| M1/Control | Up | GO: Biological Proces | GO:00017 | plasma me | 6.90E-03 | 1.00E+00 | 4.76E-02 | 4.11E-01 | 2 | 34 | MYOF,MYH10 |
| M1/Control | Up | GO: Biological Proces | GO:00720 | glomerulu | 6.90E-03 | 1.00E+00 | 4.76E-02 | 4.11E-01 | 2 | 34 | TCF21,ITGB3 |
| M1/Control | Up | GO: Biological Proces | GO:00459 | negative n | 6.90E-03 | 1.00E+00 | 4.76E-02 | 4.11E-01 | 2 | 34 | BMP2,APOE |
| M1/Control | Up | GO: Biological Proces | GO:00600 | angiogene | 7.31E-03 | 1.00E+00 | 4.76E-02 | 4.11E-01 | 2 | 35 | DAG1,ITGB3 |
| M1/Control | Up | GO: Biological Proces | GO:00105 | negative n | 7.31E-03 | 1.00E+00 | 4.76E-02 | 4.11E-01 | 2 | 35 | CD9,APOE |
| M1/Control | Up | GO: Biological Proces | GO:00457 | positive re | 7.31E-03 | 1.00E+00 | 4.76E-02 | 4.11E-01 | 2 | 35 | KLK6,ITGB3 |
| M1/Control | Up | GO: Biological Proces | GO:00364 | neuron int | 7.32E-03 | 1.00E+00 | 4.76E-02 | 4.11E-01 | 1 | 2 | PARK7 |
| M1/Control | Up | GO: Biological Proces | GO:19056 | positive re | 7.32E-03 | 1.00E+00 | 4.76E-02 | 4.11E-01 | 1 | 2 | EFEMP2 |
| M1/Control | Up | GO: Biological Proces | GO:20001 | negative n | 7.32E-03 | 1.00E+00 | 4.76E-02 | 4.11E-01 | 1 | 2 | PARK7 |
| M1/Control | Up | GO: Biological Proces | GO:00200 | endocytic | 7.32E-03 | 1.00E+00 | 4.76E-02 | 4.11E-01 | 1 | 2 | LRP2 |
| M1/Control | Up | GO: Biological Proces | GO:00023 | plasmacyt | 7.32E-03 | 1.00E+00 | 4.76E-02 | 4.11E-01 | 1 | 2 | BST2 |
| M1/Control | Up | GO: Biological Proces | GO:00423 | keratan su | 7.32E-03 | 1.00E+00 | 4.76E-02 | 4.11E-01 | 1 | 2 | GNS |
| M1/Control | Up | GO: Biological Proces | GO:00365 | protein de | 7.32E-03 | 1.00E+00 | 4.76E-02 | 4.11E-01 | 1 | 2 | PARK7 |
| M1/Control | Up | GO: Biological Proces | GO:00717 | endoplasm | 7.32E-03 | 1.00E+00 | 4.76E-02 | 4.11E-01 | 1 | 2 | CDIPT |
| M1/Control | Up | GO: Biological Proces | GO:00510 | regulation | 7.32E-03 | 1.00E+00 | 4.76E-02 | 4.11E-01 | 1 | 2 | BMP2 |
| M1/Control | Up | GO: Biological Proces | GO:00972 | cellular re | 7.32E-03 | 1.00E+00 | 4.76E-02 | 4.11E-01 | 1 | 2 | PARK7 |
| M1/Control | Up | GO: Biological Proces | GO:00015 | initiation c | 7.32E-03 | 1.00E+00 | 4.76E-02 | 4.11E-01 | 1 | 2 | GAS2 |
| M1/Control | Up | GO: Biological Proces | GO:00148 | esophagu | 7.32E-03 | 1.00E+00 | 4.76E-02 | 4.11E-01 | 1 | 2 | SULF1 |
| M1/Control | Up | GO: Biological Proces | GO:00709 | detection | 7.32E-03 | 1.00E+00 | 4.76E-02 | 4.11E-01 | 1 | 2 | PARK7 |
| M1/Control | Up | GO: Biological Proces | GO:20002 | positive re | 7.32E-03 | 1.00E+00 | 4.76E-02 | 4.11E-01 | 1 | 2 | PARK7 |
| M1/Control | Up | GO: Biological Proces | GO:19052 | negative n | 7.32E-03 | 1.00E+00 | 4.76E-02 | 4.11E-01 | 1 | 2 | PARK7 |
| M1/Control | Up | GO: Biological Proces | GO:19052 | regulation | 7.32E-03 | 1.00E+00 | 4.76E-02 | 4.11E-01 | 1 | 2 | PARK7 |
| M1/Control | Up | GO: Biological Proces | GO:00519 | positive re | 7.32E-03 | 1.00E+00 | 4.76E-02 | 4.11E-01 | 1 | 2 | ITGB1 |
| M1/Control | Up | GO: Biological Proces | GO:00519 | positive re | 7.32E-03 | 1.00E+00 | 4.76E-02 | 4.11E-01 | 1 | 2 | ITGB1 |
| M1/Control | Up | GO: Biological Proces | GO:19029 | positive re | 7.32E-03 | 1.00E+00 | 4.76E-02 | 4.11E-01 | 1 | 2 | APOE |
| M1/Control | Up | GO: Biological Proces | GO:00000 | mitotic me | 7.32E-03 | 1.00E+00 | 4.76E-02 | 4.11E-01 | 1 | 2 | NFE2L1 |
| M1/Control | Up | GO: Biological Proces | GO:00000 | mitotic M | 7.32E-03 | 1.00E+00 | 4.76E-02 | 4.11E-01 | 1 | 2 | NFE2L1 |
| M1/Control | Up | GO: Biological Proces | GO:19016 | positive re | 7.32E-03 | 1.00E+00 | 4.76E-02 | 4.11E-01 | 1 | 2 | APOE |
| M1/Control | Up | GO: Biological Proces | GO:19002 | semaphori | 7.32E-03 | 1.00E+00 | 4.76E-02 | 4.11E-01 | 1 | 2 | PLXNB1 |
| M1/Control | Up | GO: Biological Proces | GO:19033 | negative n | 7.32E-03 | 1.00E+00 | 4.76E-02 | 4.11E-01 | 1 | 2 | PARK7 |
| M1/Control | Up | GO: Biological Proces | GO:19033 | regulation | 7.32E-03 | 1.00E+00 | 4.76E-02 | 4.11E-01 | 1 | 2 | PARK7 |
| M1/Control | Up | GO: Biological Proces | GO:00611 | pancreatic | 7.32E-03 | 1.00E+00 | 4.76E-02 | 4.11E-01 | 1 | 2 | BMP2 |
| M1/Control | Up | GO: Biological Proces | GO:00611 | pancreas f | 7.32E-03 | 1.00E+00 | 4.76E-02 | 4.11E-01 | 1 | 2 | BMP2 |
| M1/Control | Up | GO: Biological Proces | GO:00027 | negative n | 7.32E-03 | 1.00E+00 | 4.76E-02 | 4.11E-01 | 1 | 2 | BST2 |
| M1/Control | Up | GO: Biological Proces | GO:20005 | negative n | 7.32E-03 | 1.00E+00 | 4.76E-02 | 4.11E-01 | 1 | 2 | ITGAV |

| | | | | | | | | | | |
|---|---|---|---|---|---|---|---|---|---|---|
| M1/Control | Up | GO: Biological Proces | GO:00108 lipid trans | 7.32E-03 | 1.00E+00 | 4.76E-02 | 4.11E-01 | 1 | 2 | APOE |
| M1/Control | Up | GO: Biological Proces | GO:00486 myoblast f | 7.32E-03 | 1.00E+00 | 4.76E-02 | 4.11E-01 | 1 | 2 | ITGB1 |
| M1/Control | Up | GO: Biological Proces | GO:00163 calcium-de | 7.32E-03 | 1.00E+00 | 4.76E-02 | 4.11E-01 | 1 | 2 | DAG1 |
| M1/Control | Up | GO: Biological Proces | GO:19030 negative n | 7.32E-03 | 1.00E+00 | 4.76E-02 | 4.11E-01 | 1 | 2 | PARK7 |
| M1/Control | Up | GO: Biological Proces | GO:00323 negative n | 7.32E-03 | 1.00E+00 | 4.76E-02 | 4.11E-01 | 1 | 2 | ATP5PF |
| M1/Control | Up | GO: Biological Proces | GO:19030 regulation | 7.32E-03 | 1.00E+00 | 4.76E-02 | 4.11E-01 | 1 | 2 | PARK7 |
| M1/Control | Up | GO: Biological Proces | GO:19030 negative n | 7.32E-03 | 1.00E+00 | 4.76E-02 | 4.11E-01 | 1 | 2 | PARK7 |
| M1/Control | Up | GO: Biological Proces | GO:00486 animal org | 7.35E-03 | 1.00E+00 | 4.76E-02 | 4.12E-01 | 3 | 108 | BMP2,SULF1,LRP2 |
| M1/Control | Up | GO: Biological Proces | GO:00603 bone deve | 7.40E-03 | 1.00E+00 | 4.78E-02 | 4.14E-01 | 5 | 331 | CFH,BMP2,THBS1,SULF1,PLXNB1 |
| M1/Control | Up | GO: Biological Proces | GO:00902 regulation | 7.49E-03 | 1.00E+00 | 4.83E-02 | 4.18E-01 | 5 | 332 | TPM1,MYL9,DSP,DAG1,SETD3 |
| M1/Control | Up | GO: Biological Proces | GO:00488 sensory sy | 7.58E-03 | 1.00E+00 | 4.88E-02 | 4.22E-01 | 9 | 945 | CFH,S100A6,MYH10,THY1,CD9,PTPRM,SULF1,CDIPT,ITGB1 |
| M1/Control | Up | GO: Biological Proces | GO:00614 reproducti | 7.65E-03 | 1.00E+00 | 4.92E-02 | 4.25E-01 | 6 | 471 | TCF21,SULF1,GAS2,LRP2,ITGAV,ITGB1 |
| M1/Control | Up | GO: Biological Proces | GO:00614 kidney vas | 7.72E-03 | 1.00E+00 | 4.92E-02 | 4.26E-01 | 2 | 36 | TCF21,ITGB3 |
| M1/Control | Up | GO: Biological Proces | GO:00614 renal syste | 7.72E-03 | 1.00E+00 | 4.92E-02 | 4.26E-01 | 2 | 36 | TCF21,ITGB3 |
| M1/Control | Up | GO: Biological Proces | GO:00104 positive re | 7.72E-03 | 1.00E+00 | 4.92E-02 | 4.26E-01 | 2 | 36 | TPM1,ATP5PF |
| M1/Control | Up | GO: Biological Proces | GO:00031 regulation | 7.72E-03 | 1.00E+00 | 4.92E-02 | 4.26E-01 | 2 | 36 | BMP2,SULF1 |
| M1/Control | Up | GO: Cellular Compon | GO:00346 integrin al | 7.34E-03 | 1.00E+00 | 2.66E-02 | 1.74E-01 | 1 | 2 | ITGB1 |
| M1/Control | Up | GO: Cellular Compon | GO:00346 integrin al | 7.34E-03 | 1.00E+00 | 2.66E-02 | 1.74E-01 | 1 | 2 | ITGB1 |
| M1/Control | Up | GO: Cellular Compon | GO:00452 proton-tra | 7.34E-03 | 1.00E+00 | 2.66E-02 | 1.74E-01 | 1 | 2 | ATP5F1A |
| M1/Control | Up | GO: Cellular Compon | GO:00346 integrin al | 7.34E-03 | 1.00E+00 | 2.66E-02 | 1.74E-01 | 1 | 2 | ITGAV |
| M1/Control | Up | GO: Cellular Compon | GO:00346 integrin al | 7.34E-03 | 1.00E+00 | 2.66E-02 | 1.74E-01 | 1 | 2 | ITGAV |
| M1/Control | Up | GO: Cellular Compon | GO:00306 coated ves | 7.66E-03 | 1.00E+00 | 2.74E-02 | 1.79E-01 | 4 | 211 | CD9,LRP2,APOE,TMED3 |
| M1/Control | Up | GO: Cellular Compon | GO:00312 anchored | 8.56E-03 | 1.00E+00 | 3.04E-02 | 1.99E-01 | 4 | 218 | THY1,BST2,XPNPEP2,RAB2A |
| M1/Control | Up | GO: Cellular Compon | GO:00057 Golgi-asso | 8.80E-03 | 1.00E+00 | 3.10E-02 | 2.02E-01 | 3 | 115 | SORT1,ITM2B,TMED3 |
| M1/Control | Up | GO: Cellular Compon | GO:00301 coated ves | 9.96E-03 | 1.00E+00 | 3.47E-02 | 2.27E-01 | 5 | 355 | SORT1,CD9,LRP2,APOE,TMED3 |
| M1/Control | Up | GO: Cellular Compon | GO:00988 plasma me | 1.04E-02 | 1.00E+00 | 3.57E-02 | 2.34E-01 | 5 | 359 | BMP2,ITGA3,ITGAV,ITGB1,ITGB3 |
| M1/Control | Up | GO: Cellular Compon | GO:00301 clathrin-co | 1.04E-02 | 1.00E+00 | 3.57E-02 | 2.34E-01 | 4 | 231 | SORT1,CD9,LRP2,APOE |
| M1/Control | Up | GO: Cellular Compon | GO:00975 myosin II | 1.10E-02 | 1.00E+00 | 3.61E-02 | 2.36E-01 | 1 | 3 | MYH10 |
| M1/Control | Up | GO: Cellular Compon | GO:00710 alphav-bet | 1.10E-02 | 1.00E+00 | 3.61E-02 | 2.36E-01 | 1 | 3 | ITGB3 |
| M1/Control | Up | GO: Cellular Compon | GO:00346 integrin al | 1.10E-02 | 1.00E+00 | 3.61E-02 | 2.36E-01 | 1 | 3 | ITGB1 |
| M1/Control | Up | GO: Cellular Compon | GO:00346 integrin al | 1.10E-02 | 1.00E+00 | 3.61E-02 | 2.36E-01 | 1 | 3 | ITGB1 |
| M1/Control | Up | GO: Cellular Compon | GO:00343 discoidal h | 1.10E-02 | 1.00E+00 | 3.61E-02 | 2.36E-01 | 1 | 3 | APOE |
| M1/Control | Up | GO: Cellular Compon | GO:00057 mitochonc | 1.16E-02 | 1.00E+00 | 3.78E-02 | 2.47E-01 | 6 | 514 | GLYAT,PARK7,GLS,PRDX1,ATP5F1A,VDAC2 |
| M1/Control | Up | GO: Cellular Compon | GO:00301 endocytic | 1.28E-02 | 1.00E+00 | 4.13E-02 | 2.70E-01 | 5 | 378 | CD9,LRP2,LAMP2,APOE,ITGAV |
| M1/Control | Up | GO: Cellular Compon | GO:00430 neuron pr | 1.29E-02 | 1.00E+00 | 4.14E-02 | 2.71E-01 | 13 | 1787 | SORT1,CFH,MYH10,THY1,PARK7,LRP2,DAG1,ACTN1,ITGA3,APOE,NDRG2,ITGB1,TNFRSF21 |
| M1/Control | Up | GO: Cellular Compon | GO:00059 cell cortex | 1.32E-02 | 1.00E+00 | 4.19E-02 | 2.74E-01 | 5 | 381 | MYH10,MYL9,MACF1,ACTN1,MYL12B |
| M1/Control | Up | GO: Cellular Compon | GO:00315 microvillus | 1.35E-02 | 1.00E+00 | 4.25E-02 | 2.78E-01 | 2 | 48 | ITGAV,ITGB3 |
| M1/Control | Up | GO: Cellular Compon | GO:00300 sarcomere | 1.40E-02 | 1.00E+00 | 4.37E-02 | 2.85E-01 | 4 | 252 | TPM1,MYL9,ACTN1,MYL12B |
| M1/Control | Up | GO: Cellular Compon | GO:00058 muscle thi | 1.46E-02 | 1.00E+00 | 4.50E-02 | 2.94E-01 | 1 | 4 | TPM1 |
| M1/Control | Up | GO: Cellular Compon | GO:19908 growth co | 1.46E-02 | 1.00E+00 | 4.50E-02 | 2.94E-01 | 1 | 4 | ITGA3 |
| M1/Control | Up | GO: Cellular Compon | GO:00058 actin filam | 1.55E-02 | 1.00E+00 | 4.73E-02 | 3.09E-01 | 3 | 142 | TPM1,GAS2,ACTN1 |
| M1/Control | Up | GO: Cellular Compon | GO:00306 clathrin-co | 1.58E-02 | 1.00E+00 | 4.78E-02 | 3.12E-01 | 3 | 143 | CD9,LRP2,APOE |
| M1/Control | Up | GO: Cellular Compon | GO:00995 polymeric | 1.63E-02 | 1.00E+00 | 4.89E-02 | 3.20E-01 | 8 | 889 | TPM1,GLYAT,NDRG1,DSP,GAS2,MACF1,ACTN1,APOE |
| M1/Control | Down | GO: Molecular Functi | GO:00330 sweet tast | 1.40E-03 | 1.33E-01 | 1.54E-02 | 7.89E-02 | 1 | 2 | TAS1R3 |
| M1/Control | Down | GO: Molecular Functi | GO:00447 hemi-meth | 2.10E-03 | 2.00E-01 | 1.86E-02 | 9.53E-02 | 1 | 3 | EGR1 |
| M1/Control | Down | GO: Molecular Functi | GO:00103 double-str | 4.90E-03 | 4.66E-01 | 2.45E-02 | 1.26E-01 | 1 | 7 | EGR1 |
| M1/Control | Down | GO: Molecular Functi | GO:00083 protein tyr | 7.69E-03 | 7.31E-01 | 3.04E-02 | 1.56E-01 | 1 | 11 | DUSP1 |
| M1/Control | Down | GO: Molecular Functi | GO:00170 MAP kinas | 9.78E-03 | 9.29E-01 | 3.72E-02 | 1.91E-01 | 1 | 14 | DUSP1 |
| M1/Control | Down | GO: Molecular Functi | GO:00051 insulin-like | 1.19E-02 | 1.00E+00 | 4.33E-02 | 2.23E-01 | 1 | 17 | REN |
| M1/Control | Down | GO: Molecular Functi | GO:00335 MAP kinas | 1.33E-02 | 1.00E+00 | 4.46E-02 | 2.29E-01 | 1 | 19 | DUSP1 |
| M1/Control | Down | GO: Biological Proces | GO:00723 positive re | 6.28E-04 | 7.26E-01 | 1.75E-02 | 1.34E-01 | 1 | 1 | EGR1 |
| M1/Control | Down | GO: Biological Proces | GO:19018 positive re | 6.28E-04 | 7.26E-01 | 1.75E-02 | 1.34E-01 | 1 | 1 | EGR1 |
| M1/Control | Down | GO: Biological Proces | GO:00148 neurotrans | 6.28E-04 | 7.26E-01 | 1.75E-02 | 1.34E-01 | 1 | 1 | NR4A1 |
| M1/Control | Down | GO: Biological Proces | GO:19018 positive re | 6.28E-04 | 7.26E-01 | 1.75E-02 | 1.34E-01 | 1 | 1 | ZFP36 |
| M1/Control | Down | GO: Biological Proces | GO:19018 regulation | 6.28E-04 | 7.26E-01 | 1.75E-02 | 1.34E-01 | 1 | 1 | ZFP36 |
| M1/Control | Down | GO: Biological Proces | GO:19042 negative n | 6.28E-04 | 7.26E-01 | 1.75E-02 | 1.34E-01 | 1 | 1 | ZFP36 |
| M1/Control | Down | GO: Biological Proces | GO:00723 regulation | 1.25E-03 | 1.00E+00 | 2.13E-02 | 1.63E-01 | 1 | 2 | EGR1 |
| M1/Control | Down | GO: Biological Proces | GO:00015 detection | 1.25E-03 | 1.00E+00 | 2.13E-02 | 1.63E-01 | 1 | 2 | TAS1R3 |
| M1/Control | Down | GO: Biological Proces | GO:19042 regulation | 1.25E-03 | 1.00E+00 | 2.13E-02 | 1.63E-01 | 1 | 2 | ZFP36 |
| M1/Control | Down | GO: Biological Proces | GO:00613 regulation | 1.25E-03 | 1.00E+00 | 2.13E-02 | 1.63E-01 | 1 | 2 | ATF3 |
| M1/Control | Down | GO: Biological Proces | GO:00715 response t | 1.25E-03 | 1.00E+00 | 2.13E-02 | 1.63E-01 | 1 | 2 | EGR1 |
| M1/Control | Down | GO: Biological Proces | GO:00715 cellular res | 1.25E-03 | 1.00E+00 | 2.13E-02 | 1.63E-01 | 1 | 2 | EGR1 |
| M1/Control | Down | GO: Biological Proces | GO:20001 positive re | 1.88E-03 | 1.00E+00 | 2.56E-02 | 1.95E-01 | 1 | 3 | CEBPB |
| M1/Control | Down | GO: Biological Proces | GO:19045 positive re | 1.88E-03 | 1.00E+00 | 2.56E-02 | 1.95E-01 | 1 | 3 | ZFP36 |
| M1/Control | Down | GO: Biological Proces | GO:00987 cellular res | 1.88E-03 | 1.00E+00 | 2.56E-02 | 1.95E-01 | 1 | 3 | EGR1 |
| M1/Control | Down | GO: Biological Proces | GO:00987 response t | 1.88E-03 | 1.00E+00 | 2.56E-02 | 1.95E-01 | 1 | 3 | EGR1 |
| M1/Control | Down | GO: Biological Proces | GO:00148 regulation | 1.88E-03 | 1.00E+00 | 2.56E-02 | 1.95E-01 | 1 | 3 | NR4A1 |
| M1/Control | Down | GO: Biological Proces | GO:19039 positive re | 1.88E-03 | 1.00E+00 | 2.56E-02 | 1.95E-01 | 1 | 3 | ATF3 |
| M1/Control | Down | GO: Biological Proces | GO:00020 renin-angi | 2.51E-03 | 1.00E+00 | 2.87E-02 | 2.19E-01 | 1 | 4 | REN |
| M1/Control | Down | GO: Biological Proces | GO:19013 regulation | 2.51E-03 | 1.00E+00 | 2.87E-02 | 2.19E-01 | 1 | 4 | CEBPB |
| M1/Control | Down | GO: Biological Proces | GO:00600 circadian t | 2.51E-03 | 1.00E+00 | 2.87E-02 | 2.19E-01 | 1 | 4 | EGR1 |
| M1/Control | Down | GO: Biological Proces | GO:00722 metaneph | 2.51E-03 | 1.00E+00 | 2.87E-02 | 2.19E-01 | 1 | 4 | EGR1 |
| M1/Control | Down | GO: Biological Proces | GO:20001 regulation | 3.13E-03 | 1.00E+00 | 3.27E-02 | 2.50E-01 | 1 | 5 | EGR1 |
| M1/Control | Down | GO: Biological Proces | GO:19045 regulation | 3.13E-03 | 1.00E+00 | 3.27E-02 | 2.50E-01 | 1 | 5 | ZFP36 |
| M1/Control | Down | GO: Biological Proces | GO:00722 metaneph | 3.13E-03 | 1.00E+00 | 3.27E-02 | 2.50E-01 | 1 | 5 | EGR1 |
| M1/Control | Down | GO: Biological Proces | GO:00715 cellular res | 3.13E-03 | 1.00E+00 | 3.27E-02 | 2.50E-01 | 1 | 5 | EGR1 |
| M1/Control | Down | GO: Biological Proces | GO:20001 regulation | 3.76E-03 | 1.00E+00 | 3.56E-02 | 2.72E-01 | 1 | 6 | CEBPB |
| M1/Control | Down | GO: Biological Proces | GO:00357 T-helper 1 | 3.76E-03 | 1.00E+00 | 3.56E-02 | 2.72E-01 | 1 | 6 | CEBPB |
| M1/Control | Down | GO: Biological Proces | GO:00713 cellular res | 3.76E-03 | 1.00E+00 | 3.56E-02 | 2.72E-01 | 1 | 6 | NR4A1 |
| M1/Control | Down | GO: Biological Proces | GO:19031 regulation | 3.76E-03 | 1.00E+00 | 3.56E-02 | 2.72E-01 | 1 | 6 | ATF3 |
| M1/Control | Down | GO: Biological Proces | GO:00434 response t | 3.76E-03 | 1.00E+00 | 3.56E-02 | 2.72E-01 | 1 | 6 | NR4A1 |
| M1/Control | Down | GO: Biological Proces | GO:00310 deadenyla | 3.76E-03 | 1.00E+00 | 3.56E-02 | 2.72E-01 | 1 | 6 | ZFP36 |
| M1/Control | Down | GO: Biological Proces | GO:00080 establishm | 3.76E-03 | 1.00E+00 | 3.56E-02 | 2.72E-01 | 1 | 6 | REN |
| M1/Control | Down | GO: Biological Proces | GO:20008 regulation | 4.38E-03 | 1.00E+00 | 3.73E-02 | 2.85E-01 | 1 | 7 | REN |
| M1/Control | Down | GO: Biological Proces | GO:20008 regulation | 4.38E-03 | 1.00E+00 | 3.73E-02 | 2.85E-01 | 1 | 7 | REN |
| M1/Control | Down | GO: Biological Proces | GO:19018 regulation | 4.38E-03 | 1.00E+00 | 3.73E-02 | 2.85E-01 | 1 | 7 | EGR1 |
| M1/Control | Down | GO: Biological Proces | GO:00509 sensory pe | 4.38E-03 | 1.00E+00 | 3.73E-02 | 2.85E-01 | 1 | 7 | TAS1R3 |
| M1/Control | Down | GO: Biological Proces | GO:00509 sensory pe | 4.38E-03 | 1.00E+00 | 3.73E-02 | 2.85E-01 | 1 | 7 | TAS1R3 |
| M1/Control | Down | GO: Biological Proces | GO:00310 nuclear-tra | 4.38E-03 | 1.00E+00 | 3.73E-02 | 2.85E-01 | 1 | 7 | ZFP36 |
| M1/Control | Down | GO: Biological Proces | GO:00016 conditione | 4.38E-03 | 1.00E+00 | 3.73E-02 | 2.85E-01 | 1 | 7 | FOS |
| M1/Control | Down | GO: Biological Proces | GO:00900 negative n | 4.38E-03 | 1.00E+00 | 3.73E-02 | 2.85E-01 | 1 | 7 | DUSP1 |
| M1/Control | Down | GO: Biological Proces | GO:00721 positive re | 5.01E-03 | 1.00E+00 | 3.81E-02 | 2.91E-01 | 1 | 8 | EGR1 |
| M1/Control | Down | GO: Biological Proces | GO:19021 regulation | 5.01E-03 | 1.00E+00 | 3.81E-02 | 2.91E-01 | 1 | 8 | ZFP36 |
| M1/Control | Down | GO: Biological Proces | GO:00359 aldosteror | 5.01E-03 | 1.00E+00 | 3.81E-02 | 2.91E-01 | 1 | 8 | REN |
| M1/Control | Down | GO: Biological Proces | GO:00359 mineraloco | 5.01E-03 | 1.00E+00 | 3.81E-02 | 2.91E-01 | 1 | 8 | REN |
| M1/Control | Down | GO: Biological Proces | GO:00703 response t | 5.01E-03 | 1.00E+00 | 3.81E-02 | 2.91E-01 | 1 | 8 | REN |
| M1/Control | Down | GO: Biological Proces | GO:00972 keratinocy | 5.01E-03 | 1.00E+00 | 3.81E-02 | 2.91E-01 | 1 | 8 | ZFP36 |
| M1/Control | Down | GO: Biological Proces | GO:00715 response t | 5.01E-03 | 1.00E+00 | 3.81E-02 | 2.91E-01 | 1 | 8 | EGR1 |
| M1/Control | Down | GO: Biological Proces | GO:00020 angiotensi | 5.63E-03 | 1.00E+00 | 4.05E-02 | 3.09E-01 | 1 | 9 | REN |
| M1/Control | Down | GO: Biological Proces | GO:19029 positive re | 5.63E-03 | 1.00E+00 | 4.05E-02 | 3.09E-01 | 1 | 9 | EGR1 |
| M1/Control | Down | GO: Biological Proces | GO:00344 response t | 5.63E-03 | 1.00E+00 | 4.05E-02 | 3.09E-01 | 1 | 9 | EGR1 |
| M1/Control | Down | GO: Biological Proces | GO:00024 granuloma | 5.63E-03 | 1.00E+00 | 4.05E-02 | 3.09E-01 | 1 | 9 | CEBPB |
| M1/Control | Down | GO: Biological Proces | GO:00718 response t | 5.63E-03 | 1.00E+00 | 4.05E-02 | 3.09E-01 | 1 | 9 | EGR1 |
| M1/Control | Down | GO: Biological Proces | GO:00067 progestero | 5.63E-03 | 1.00E+00 | 4.05E-02 | 3.09E-01 | 1 | 9 | EGR1 |
| M1/Control | Down | GO: Biological Proces | GO:00722 metaneph | 5.63E-03 | 1.00E+00 | 4.05E-02 | 3.09E-01 | 1 | 9 | EGR1 |
| M1/Control | Down | GO: Biological Proces | GO:19037 negative n | 5.63E-03 | 1.00E+00 | 4.05E-02 | 3.09E-01 | 1 | 9 | DUSP1 |
| M1/Control | Down | GO: Biological Proces | GO:00020 regulation | 6.26E-03 | 1.00E+00 | 4.25E-02 | 3.25E-01 | 1 | 10 | REN |
| M1/Control | Down | GO: Biological Proces | GO:00714 cellular res | 6.26E-03 | 1.00E+00 | 4.25E-02 | 3.25E-01 | 1 | 10 | EGR1 |
| M1/Control | Down | GO: Biological Proces | GO:00109 regulation | 6.26E-03 | 1.00E+00 | 4.25E-02 | 3.25E-01 | 1 | 10 | CEBPB |
| M1/Control | Down | GO: Biological Proces | GO:00718 odontobla | 6.26E-03 | 1.00E+00 | 4.25E-02 | 3.25E-01 | 1 | 10 | CEBPB |
| M1/Control | Down | GO: Biological Proces | GO:19020 negative n | 6.26E-03 | 1.00E+00 | 4.25E-02 | 3.25E-01 | 1 | 10 | ZFP36 |
| M1/Control | Down | GO: Biological Proces | GO:00713 cellular res | 6.26E-03 | 1.00E+00 | 4.25E-02 | 3.25E-01 | 1 | 10 | EGR1 |
| M1/Control | Down | GO: Biological Proces | GO:19017 positive re | 6.26E-03 | 1.00E+00 | 4.25E-02 | 3.25E-01 | 1 | 10 | EGR1 |
| M1/Control | Down | GO: Biological Proces | GO:19906 cellular res | 6.88E-03 | 1.00E+00 | 4.49E-02 | 3.43E-01 | 1 | 11 | FOS |
| M1/Control | Down | GO: Biological Proces | GO:00722 cell prolife | 6.88E-03 | 1.00E+00 | 4.49E-02 | 3.43E-01 | 1 | 11 | EGR1 |
| M1/Control | Down | GO: Biological Proces | GO:00443 sodium-de | 6.88E-03 | 1.00E+00 | 4.49E-02 | 3.43E-01 | 1 | 11 | CEBPB |
| M1/Control | Down | GO: Cellular Compon | GO:19037 taste recep | 1.34E-03 | 8.96E-02 | 1.49E-02 | 7.15E-02 | 1 | 2 | TAS1R3 |
| M1/Control | Down | GO: Cellular Compon | GO:19906 C/EBP com | 1.34E-03 | 8.96E-02 | 1.49E-02 | 7.15E-02 | 1 | 2 | CEBPB |
| M1/Control | Down | GO: Cellular Compon | GO:19906 CHOP-ATF | 1.34E-03 | 8.96E-02 | 1.49E-02 | 7.15E-02 | 1 | 2 | ATF3 |
| M1/Control | Down | GO: Cellular Compon | GO:00007 chromatin | 1.70E-03 | 1.14E-01 | 1.62E-02 | 7.78E-02 | 5 | 1419 | EGR1,FOS,ATF3,CEBPB,NR4A1 |
| M1/Control | Down | GO: Cellular Compon | GO:00364 CHOP-C/E | 2.01E-03 | 1.34E-01 | 1.68E-02 | 8.04E-02 | 1 | 3 | CEBPB |
| M1/Control | Down | GO: Cellular Compon | GO:00359 transcripti | 3.34E-03 | 2.24E-01 | 2.44E-02 | 1.17E-01 | 1 | 5 | FOS |
| M1/Control | Down | GO: Cellular Compon | GO:00717 IgD immu | 4.01E-03 | 2.69E-01 | 2.44E-02 | 1.17E-01 | 1 | 6 | IGKC |
| M1/Control | Down | GO: Cellular Compon | GO:00717 IgE immu | 4.01E-03 | 2.69E-01 | 2.44E-02 | 1.17E-01 | 1 | 6 | IGKC |
| M1/Control | Down | GO: Cellular Compon | GO:00705 RISC-loadi | 7.33E-03 | 4.91E-01 | 4.10E-02 | 1.96E-01 | 1 | 11 | ZFP36 |
| M1/Control | Down | GO: Cellular Compon | GO:00717 IgG immu | 8.66E-03 | 5.80E-01 | 4.15E-02 | 1.99E-01 | 1 | 13 | IGKC |
| M1/Control | Down | GO: Cellular Compon | GO:00717 IgM immu | 8.66E-03 | 5.80E-01 | 4.15E-02 | 1.99E-01 | 1 | 13 | IGKC |
| M1/Control | Down | GO: Cellular Compon | GO:00717 IgA immu | 9.99E-03 | 6.69E-01 | 4.46E-02 | 2.14E-01 | 1 | 15 | IGKC |
| M1/Control | Down | GO: Cellular Compon | GO:00300 CCR4-NOT | 1.07E-02 | 7.14E-01 | 4.46E-02 | 2.14E-01 | 1 | 16 | ZFP36 |
| M0/Control | Up | GO: Biological Proces | GO:00722 metaneph | 1.45E-04 | 2.09E-02 | 4.47E-03 | 2.48E-02 | 1 | 3 | TCF21 |
| M0/Control | Up | GO: Biological Proces | GO:00722 metaneph | 1.45E-04 | 2.09E-02 | 4.47E-03 | 2.48E-02 | 1 | 3 | TCF21 |
| M0/Control | Up | GO: Biological Proces | GO:00722 metaneph | 1.45E-04 | 2.09E-02 | 4.47E-03 | 2.48E-02 | 1 | 3 | TCF21 |
| M0/Control | Up | GO: Biological Proces | GO:00721 metaneph | 2.41E-04 | 3.48E-02 | 4.47E-03 | 2.48E-02 | 1 | 5 | TCF21 |
| M0/Control | Up | GO: Biological Proces | GO:00604 bronchiole | 2.90E-04 | 4.17E-02 | 4.47E-03 | 2.48E-02 | 1 | 6 | TCF21 |
| M0/Control | Up | GO: Biological Proces | GO:00147 branchiom | 2.90E-04 | 4.17E-02 | 4.47E-03 | 2.48E-02 | 1 | 6 | TCF21 |
| M0/Control | Up | GO: Biological Proces | GO:00721 glomerulu | 3.38E-04 | 4.87E-02 | 4.47E-03 | 2.48E-02 | 1 | 7 | TCF21 |
| M0/Control | Up | GO: Biological Proces | GO:00721 glomerula | 3.38E-04 | 4.87E-02 | 4.47E-03 | 2.48E-02 | 1 | 7 | TCF21 |
| M0/Control | Up | GO: Biological Proces | GO:20010 mesenchy | 3.86E-04 | 5.56E-02 | 4.47E-03 | 2.48E-02 | 1 | 8 | TCF21 |
| M0/Control | Up | GO: Biological Proces | GO:00721 mesenchy | 3.86E-04 | 5.56E-02 | 4.47E-03 | 2.48E-02 | 1 | 8 | TCF21 |
| M0/Control | Up | GO: Biological Proces | GO:00722 metaneph | 4.34E-04 | 6.26E-02 | 4.47E-03 | 2.48E-02 | 1 | 9 | TCF21 |
| M0/Control | Up | GO: Biological Proces | GO:00604 lung vascu | 4.34E-04 | 6.26E-02 | 4.47E-03 | 2.48E-02 | 1 | 9 | TCF21 |
| M0/Control | Up | GO: Biological Proces | GO:00614 renal syste | 4.34E-04 | 6.26E-02 | 4.47E-03 | 2.48E-02 | 1 | 9 | TCF21 |
| M0/Control | Up | GO: Biological Proces | GO:00614 kidney vas | 4.34E-04 | 6.26E-02 | 4.47E-03 | 2.48E-02 | 1 | 9 | TCF21 |
| M0/Control | Up | GO: Biological Proces | GO:00605 diaphragm | 6.27E-04 | 9.04E-02 | 6.02E-03 | 3.34E-02 | 1 | 13 | TCF21 |
| M0/Control | Up | GO: Biological Proces | GO:00720 metaneph | 7.24E-04 | 1.04E-01 | 6.52E-03 | 3.62E-02 | 1 | 15 | TCF21 |
| M0/Control | Up | GO: Biological Proces | GO:00607 negative n | 8.21E-04 | 1.18E-01 | 6.56E-03 | 3.64E-02 | 1 | 17 | TCF21 |
| M0/Control | Up | GO: Biological Proces | GO:00722 metaneph | 8.21E-04 | 1.18E-01 | 6.56E-03 | 3.64E-02 | 1 | 17 | TCF21 |
| M0/Control | Up | GO: Biological Proces | GO:00720 kidney me | 9.17E-04 | 1.32E-01 | 6.95E-03 | 3.86E-02 | 1 | 19 | TCF21 |
| M0/Control | Up | GO: Biological Proces | GO:00485 embryonic | 1.11E-03 | 1.60E-01 | 7.99E-03 | 4.44E-02 | 1 | 23 | TCF21 |
| M0/Control | Up | GO: Biological Proces | GO:00722 cell differe | 1.26E-03 | 1.81E-01 | 8.61E-03 | 4.78E-02 | 1 | 26 | TCF21 |
| M0/Control | Up | GO: Biological Proces | GO:00721 glomerulu | 1.40E-03 | 2.02E-01 | 9.16E-03 | 5.09E-02 | 1 | 29 | TCF21 |
| M0/Control | Up | GO: Biological Proces | GO:00607 regulation | 1.50E-03 | 2.15E-01 | 9.37E-03 | 5.20E-02 | 1 | 31 | TCF21 |
| M0/Control | Up | GO: Biological Proces | GO:00720 glomerulu | 1.64E-03 | 2.36E-01 | 9.62E-03 | 5.34E-02 | 1 | 34 | TCF21 |
| M0/Control | Up | GO: Biological Proces | GO:00614 kidney vas | 1.74E-03 | 2.50E-01 | 9.62E-03 | 5.34E-02 | 1 | 36 | TCF21 |
| M0/Control | Up | GO: Biological Proces | GO:00614 renal syste | 1.74E-03 | 2.50E-01 | 9.62E-03 | 5.34E-02 | 1 | 36 | TCF21 |
| M0/Control | Up | GO: Biological Proces | GO:00331 negative n | 1.98E-03 | 2.85E-01 | 9.96E-03 | 5.53E-02 | 1 | 41 | TCF21 |
| M0/Control | Up | GO: Biological Proces | GO:00485 embryonic | 2.03E-03 | 2.92E-01 | 9.96E-03 | 5.53E-02 | 1 | 42 | TCF21 |
| M0/Control | Up | GO: Biological Proces | GO:00722 metaneph | 2.08E-03 | 2.99E-01 | 9.96E-03 | 5.53E-02 | 1 | 43 | TCF21 |
| M0/Control | Up | GO: Biological Proces | GO:00600 Sertoli cell | 2.08E-03 | 2.99E-01 | 9.96E-03 | 5.53E-02 | 1 | 43 | TCF21 |
| M0/Control | Up | GO: Biological Proces | GO:00075 sex determ | 2.22E-03 | 3.20E-01 | 1.03E-02 | 5.72E-02 | 1 | 46 | TCF21 |
| M0/Control | Up | GO: Biological Proces | GO:00485 spleen dev | 2.32E-03 | 3.34E-01 | 1.03E-02 | 5.73E-02 | 1 | 48 | TCF21 |
| M0/Control | Up | GO: Biological Proces | GO:00305 androgen | 2.37E-03 | 3.41E-01 | 1.03E-02 | 5.73E-02 | 1 | 49 | TCF21 |
| M0/Control | Up | GO: Biological Proces | GO:00016 branching | 3.33E-03 | 4.80E-01 | 1.41E-02 | 7.82E-02 | 1 | 69 | TCF21 |
| M0/Control | Up | GO: Biological Proces | GO:00604 lung morp | 3.57E-03 | 5.14E-01 | 1.41E-02 | 7.82E-02 | 1 | 74 | TCF21 |
| M0/Control | Up | GO: Biological Proces | GO:00310 regulation | 3.62E-03 | 5.21E-01 | 1.41E-02 | 7.82E-02 | 1 | 75 | TCF21 |
| M0/Control | Up | GO: Biological Proces | GO:00482 lung alveo | 3.62E-03 | 5.21E-01 | 1.41E-02 | 7.82E-02 | 1 | 75 | TCF21 |
| M0/Control | Up | GO: Biological Proces | GO:00606 ureteric bu | 3.72E-03 | 5.35E-01 | 1.41E-02 | 7.82E-02 | 1 | 77 | TCF21 |
| M0/Control | Up | GO: Biological Proces | GO:00721 mesoneph | 3.91E-03 | 5.63E-01 | 1.44E-02 | 8.01E-02 | 1 | 81 | TCF21 |
| M0/Control | Up | GO: Biological Proces | GO:00485 digestive t | 4.34E-03 | 6.26E-01 | 1.48E-02 | 8.23E-02 | 1 | 90 | TCF21 |
| M0/Control | Up | GO: Biological Proces | GO:00720 nephron t | 4.39E-03 | 6.32E-01 | 1.48E-02 | 8.23E-02 | 1 | 91 | TCF21 |
| M0/Control | Up | GO: Biological Proces | GO:00331 regulation | 4.44E-03 | 6.39E-01 | 1.48E-02 | 8.23E-02 | 1 | 92 | TCF21 |
| M0/Control | Up | GO: Biological Proces | GO:00720 nephron e | 4.49E-03 | 6.46E-01 | 1.48E-02 | 8.23E-02 | 1 | 93 | TCF21 |
| M0/Control | Up | GO: Biological Proces | GO:00903 regulation | 4.59E-03 | 6.60E-01 | 1.48E-02 | 8.23E-02 | 1 | 95 | TCF21 |
| M0/Control | Up | GO: Biological Proces | GO:00613 renal tubu | 4.63E-03 | 6.67E-01 | 1.48E-02 | 8.23E-02 | 1 | 96 | TCF21 |
| M0/Control | Up | GO: Biological Proces | GO:00610 cell differe | 4.88E-03 | 7.02E-01 | 1.52E-02 | 8.44E-02 | 1 | 101 | TCF21 |
| M0/Control | Up | GO: Biological Proces | GO:00016 metaneph | 4.97E-03 | 7.16E-01 | 1.52E-02 | 8.44E-02 | 1 | 103 | TCF21 |
| M0/Control | Up | GO: Biological Proces | GO:00720 nephron m | 5.07E-03 | 7.30E-01 | 1.52E-02 | 8.44E-02 | 1 | 105 | TCF21 |
| M0/Control | Up | GO: Biological Proces | GO:00600 roof of mo | 5.45E-03 | 7.85E-01 | 1.58E-02 | 8.75E-02 | 1 | 113 | TCF21 |
| M0/Control | Up | GO: Biological Proces | GO:00016 ureteric bu | 5.50E-03 | 7.92E-01 | 1.58E-02 | 8.75E-02 | 1 | 114 | TCF21 |
| M0/Control | Up | GO: Biological Proces | GO:00721 mesoneph | 5.70E-03 | 8.20E-01 | 1.58E-02 | 8.75E-02 | 1 | 118 | TCF21 |
| M0/Control | Up | GO: Biological Proces | GO:00721 mesoneph | 5.70E-03 | 8.20E-01 | 1.58E-02 | 8.75E-02 | 1 | 118 | TCF21 |
| M0/Control | Up | GO: Biological Proces | GO:00018 mesoneph | 5.94E-03 | 8.55E-01 | 1.58E-02 | 8.77E-02 | 1 | 123 | TCF21 |
| M0/Control | Up | GO: Biological Proces | GO:00328 glomerulu | 6.03E-03 | 8.69E-01 | 1.58E-02 | 8.77E-02 | 1 | 125 | TCF21 |
| M0/Control | Up | GO: Biological Proces | GO:00720 nephron t | 6.03E-03 | 8.69E-01 | 1.58E-02 | 8.77E-02 | 1 | 125 | TCF21 |
| M0/Control | Up | GO: Biological Proces | GO:00613 renal tubu | 6.27E-03 | 9.04E-01 | 1.61E-02 | 8.96E-02 | 1 | 130 | TCF21 |
| M0/Control | Up | GO: Biological Proces | GO:00305 intracellula | 6.47E-03 | 9.31E-01 | 1.63E-02 | 9.05E-02 | 1 | 134 | TCF21 |
| M0/Control | Up | GO: Biological Proces | GO:00165 histone de | 6.56E-03 | 9.45E-01 | 1.63E-02 | 9.05E-02 | 1 | 136 | TCF21 |
| M0/Control | Up | GO: Biological Proces | GO:00609 kidney mo | 7.05E-03 | 1.00E+00 | 1.72E-02 | 9.55E-02 | 1 | 146 | TCF21 |
| M0/Control | Up | GO: Biological Proces | GO:00608 artery dev | 7.24E-03 | 1.00E+00 | 1.74E-02 | 9.64E-02 | 1 | 150 | TCF21 |
| M0/Control | Up | GO: Biological Proces | GO:00434 steroid ho | 7.67E-03 | 1.00E+00 | 1.78E-02 | 9.89E-02 | 1 | 159 | TCF21 |
| M0/Control | Up | GO: Biological Proces | GO:00064 protein de | 7.67E-03 | 1.00E+00 | 1.78E-02 | 9.89E-02 | 1 | 159 | TCF21 |
| M0/Control | Up | GO: Biological Proces | GO:00356 protein de | 8.21E-03 | 1.00E+00 | 1.87E-02 | 1.04E-01 | 1 | 170 | TCF21 |
| M0/Control | Up | GO: Biological Proces | GO:00987 macromol | 8.40E-03 | 1.00E+00 | 1.87E-02 | 1.04E-01 | 1 | 174 | TCF21 |
| M0/Control | Up | GO: Biological Proces | GO:00720 nephron e | 8.45E-03 | 1.00E+00 | 1.87E-02 | 1.04E-01 | 1 | 175 | TCF21 |
| M0/Control | Up | GO: Molecular Functi | GO:00506 nuclear an | 1.70E-03 | 3.92E-02 | 1.88E-02 | 7.03E-02 | 1 | 34 | TCF21 |
| M0/Control | Up | GO: Molecular Functi | GO:00434 bHLH tran | 1.75E-03 | 4.03E-02 | 1.88E-02 | 7.03E-02 | 1 | 35 | TCF21 |
| M0/Control | Up | GO: Molecular Functi | GO:00708 E-box binc | 2.46E-03 | 5.65E-02 | 1.88E-02 | 7.03E-02 | 1 | 49 | TCF21 |
| M0/Control | Up | GO: Biological Proces | GO:00487 branching | 1.03E-02 | 1.00E+00 | 2.18E-02 | 1.21E-01 | 1 | 214 | TCF21 |
| M0/Control | Up | GO: Biological Proces | GO:00085 male gona | 1.05E-02 | 1.00E+00 | 2.18E-02 | 1.21E-01 | 1 | 218 | TCF21 |
| M0/Control | Up | GO: Biological Proces | GO:00465 developme | 1.06E-02 | 1.00E+00 | 2.18E-02 | 1.21E-01 | 1 | 219 | TCF21 |
| M0/Control | Up | GO: Biological Proces | GO:00097 hormone- | 1.06E-02 | 1.00E+00 | 2.18E-02 | 1.21E-01 | 1 | 219 | TCF21 |
| M0/Control | Up | GO: Biological Proces | GO:00720 kidney epi | 1.06E-02 | 1.00E+00 | 2.18E-02 | 1.21E-01 | 1 | 220 | TCF21 |
| M0/Control | Up | GO: Biological Proces | GO:00720 nephron d | 1.16E-02 | 1.00E+00 | 2.35E-02 | 1.30E-01 | 1 | 240 | TCF21 |
| M0/Control | Up | GO: Biological Proces | GO:00310 regulation | 1.18E-02 | 1.00E+00 | 2.36E-02 | 1.31E-01 | 1 | 244 | TCF21 |
| M0/Control | Up | GO: Biological Proces | GO:00466 male sex c | 1.21E-02 | 1.00E+00 | 2.37E-02 | 1.31E-01 | 1 | 250 | TCF21 |
| M0/Control | Up | GO: Biological Proces | GO:00611 morphoge | 1.22E-02 | 1.00E+00 | 2.37E-02 | 1.31E-01 | 1 | 252 | TCF21 |
| M0/Control | Up | GO: Biological Proces | GO:00485 digestive t | 1.26E-02 | 1.00E+00 | 2.42E-02 | 1.34E-01 | 1 | 261 | TCF21 |
| M0/Control | Up | GO: Biological Proces | GO:00017 morphoge | 1.33E-02 | 1.00E+00 | 2.52E-02 | 1.40E-01 | 1 | 276 | TCF21 |
| M0/Control | Up | GO: Biological Proces | GO:00303 lung devel | 1.37E-02 | 1.00E+00 | 2.55E-02 | 1.42E-01 | 1 | 283 | TCF21 |
| M0/Control | Up | GO: Biological Proces | GO:00303 respiratory | 1.39E-02 | 1.00E+00 | 2.56E-02 | 1.42E-01 | 1 | 287 | TCF21 |
| M0/Control | Up | GO: Biological Proces | GO:00713 cellular res | 1.47E-02 | 1.00E+00 | 2.68E-02 | 1.49E-01 | 1 | 305 | TCF21 |
| M0/Control | Up | GO: Biological Proces | GO:00551 digestive s | 1.50E-02 | 1.00E+00 | 2.70E-02 | 1.50E-01 | 1 | 311 | TCF21 |
| M0/Control | Up | GO: Biological Proces | GO:00605 respiratory | 1.56E-02 | 1.00E+00 | 2.77E-02 | 1.54E-01 | 1 | 324 | TCF21 |
| M0/Control | Up | GO: Biological Proces | GO:00075 skeletal m | 1.60E-02 | 1.00E+00 | 2.77E-02 | 1.54E-01 | 1 | 331 | TCF21 |
| M0/Control | Up | GO: Biological Proces | GO:00305 intracellula | 1.60E-02 | 1.00E+00 | 2.77E-02 | 1.54E-01 | 1 | 331 | TCF21 |
| M0/Control | Up | GO: Biological Proces | GO:00605 skeletal m | 1.70E-02 | 1.00E+00 | 2.91E-02 | 1.62E-01 | 1 | 352 | TCF21 |
| M0/Control | Up | GO: Biological Proces | GO:00084 gonad dev | 1.80E-02 | 1.00E+00 | 3.02E-02 | 1.68E-01 | 1 | 372 | TCF21 |
| M0/Control | Up | GO: Biological Proces | GO:00487 mesenchy | 1.82E-02 | 1.00E+00 | 3.02E-02 | 1.68E-01 | 1 | 376 | TCF21 |
| M0/Control | Up | GO: Biological Proces | GO:00451 developme | 1.82E-02 | 1.00E+00 | 3.02E-02 | 1.68E-01 | 1 | 378 | TCF21 |
| M0/Control | Up | GO: Biological Proces | GO:00075 sex differe | 2.15E-02 | 1.00E+00 | 3.51E-02 | 1.95E-01 | 1 | 445 | TCF21 |
| M0/Control | Up | GO: Biological Proces | GO:00486 reproducti | 2.25E-02 | 1.00E+00 | 3.64E-02 | 2.02E-01 | 1 | 466 | TCF21 |
| M0/Control | Up | GO: Biological Proces | GO:00614 reproducti | 2.27E-02 | 1.00E+00 | 3.64E-02 | 2.02E-01 | 1 | 471 | TCF21 |
| M0/Control | Up | GO: Biological Proces | GO:00604 mesenchy | 2.34E-02 | 1.00E+00 | 3.70E-02 | 2.06E-01 | 1 | 485 | TCF21 |
| M0/Control | Up | GO: Biological Proces | GO:00018 kidney dev | 2.54E-02 | 1.00E+00 | 3.97E-02 | 2.21E-01 | 1 | 526 | TCF21 |
| M0/Control | Up | GO: Biological Proces | GO:00720 renal syste | 2.60E-02 | 1.00E+00 | 4.03E-02 | 2.24E-01 | 1 | 539 | TCF21 |
| M0/Control | Up | GO: Biological Proces | GO:00165 histone m | 2.72E-02 | 1.00E+00 | 4.13E-02 | 2.29E-01 | 1 | 563 | TCF21 |
| M0/Control | Up | GO: Biological Proces | GO:00075 muscle org | 2.72E-02 | 1.00E+00 | 4.13E-02 | 2.29E-01 | 1 | 564 | TCF21 |
| M0/Control | Up | GO: Biological Proces | GO:00485 response t | 2.83E-02 | 1.00E+00 | 4.19E-02 | 2.33E-01 | 1 | 586 | TCF21 |
| M0/Control | Up | GO: Biological Proces | GO:00605 epithelial t | 2.83E-02 | 1.00E+00 | 4.19E-02 | 2.33E-01 | 1 | 587 | TCF21 |
| M0/Control | Up | GO: Biological Proces | GO:00016 urogenital | 2.85E-02 | 1.00E+00 | 4.19E-02 | 2.33E-01 | 1 | 591 | TCF21 |
| M0/Control | Up | GO: Molecular Functi | GO:00428 histone de | 7.41E-03 | 1.71E-01 | 4.26E-02 | 1.59E-01 | 1 | 148 | TCF21 |
| M0/Control | Up | GO: Molecular Functi | GO:00169 nuclear re | 9.57E-03 | 2.20E-01 | 4.40E-02 | 1.64E-01 | 1 | 191 | TCF21 |
| M0/Control | Up | GO: Biological Proces | GO:00485 embryonic | 3.18E-02 | 1.00E+00 | 4.62E-02 | 2.56E-01 | 1 | 658 | TCF21 |
| M0/Control | Up | GO: Biological Proces | GO:00605 muscle tis | 3.36E-02 | 1.00E+00 | 4.84E-02 | 2.69E-01 | 1 | 697 | TCF21 |
| M1/M0 | Up | GO: Molecular Functi | GO:00016 virus recep | 6.02E-03 | 1.00E+00 | 3.66E-02 | 2.17E-01 | 2 | 97 | CD81,ITGB1 |
| M1/M0 | Up | GO: Molecular Functi | GO:01402 exogenous | 6.14E-03 | 1.00E+00 | 3.66E-02 | 2.17E-01 | 2 | 98 | CD81,ITGB1 |
| M1/M0 | Up | GO: Molecular Functi | GO:00860 gap juncti | 4.80E-03 | 1.00E+00 | 3.66E-02 | 2.17E-01 | 1 | 4 | GJA5 |
| M1/M0 | Up | GO: Molecular Functi | GO:00085 calcium-de | 6.00E-03 | 1.00E+00 | 3.66E-02 | 2.17E-01 | 1 | 5 | RCAN1 |
| M1/M0 | Up | GO: Molecular Functi | GO:00860 gap juncti | 6.00E-03 | 1.00E+00 | 3.66E-02 | 2.17E-01 | 1 | 5 | GJA5 |
| M1/M0 | Up | GO: Molecular Functi | GO:00199 C-X3-C ch | 6.00E-03 | 1.00E+00 | 3.66E-02 | 2.17E-01 | 1 | 5 | ITGB1 |
| M1/M0 | Up | GO: Molecular Functi | GO:00318 mu-type o | 6.00E-03 | 1.00E+00 | 3.66E-02 | 2.17E-01 | 1 | 5 | FLNA |
| M1/M0 | Up | GO: Molecular Functi | GO:00051 interleukin | 6.00E-03 | 1.00E+00 | 3.66E-02 | 2.17E-01 | 1 | 5 | ECM1 |
| M1/M0 | Up | GO: Molecular Functi | GO:00986 collagen b | 6.00E-03 | 1.00E+00 | 3.66E-02 | 2.17E-01 | 1 | 5 | ITGB1 |
| M1/M0 | Up | GO: Molecular Functi | GO:00452 cadherin b | 7.66E-03 | 1.00E+00 | 3.98E-02 | 2.36E-01 | 3 | 341 | FLNA,CTNNB1,ITGB1 |
| M1/M0 | Up | GO: Molecular Functi | GO:00080 beta-cater | 7.54E-03 | 1.00E+00 | 3.98E-02 | 2.36E-01 | 2 | 109 | TCF4,CTNNB1 |
| M1/M0 | Up | GO: Molecular Functi | GO:00010 TFIIB-class | 7.19E-03 | 1.00E+00 | 3.98E-02 | 2.36E-01 | 1 | 6 | TCF4 |
| M1/M0 | Up | GO: Molecular Functi | GO:00712 connexin b | 8.39E-03 | 1.00E+00 | 3.98E-02 | 2.36E-01 | 1 | 7 | GJA5 |
| M1/M0 | Up | GO: Molecular Functi | GO:00349 Fc-gamma | 8.39E-03 | 1.00E+00 | 3.98E-02 | 2.36E-01 | 1 | 7 | FLNA |
| M1/M0 | Up | GO: Molecular Functi | GO:00422 MHC class | 8.39E-03 | 1.00E+00 | 3.98E-02 | 2.36E-01 | 1 | 7 | CD81 |
| M1/M0 | Up | GO: Molecular Functi | GO:00081 primary ar | 8.39E-03 | 1.00E+00 | 3.98E-02 | 2.36E-01 | 1 | 7 | VCAM1 |
| M1/M0 | Up | GO: Molecular Functi | GO:00055 galactose | 8.39E-03 | 1.00E+00 | 3.98E-02 | 2.36E-01 | 1 | 7 | ENG |
| M1/M0 | Up | GO: Molecular Functi | GO:00986 cell-matrix | 8.39E-03 | 1.00E+00 | 3.98E-02 | 2.36E-01 | 1 | 7 | ITGB1 |
| M1/M0 | Up | GO: Molecular Functi | GO:00080 cytoskelet | 9.69E-03 | 1.00E+00 | 4.50E-02 | 2.67E-01 | 5 | 1123 | S100A6,FLNA,ITGB1,SGK1,MSN |
| M1/M0 | Up | GO: Molecular Functi | GO:19037 gap juncti | 1.08E-02 | 1.00E+00 | 4.89E-02 | 2.90E-01 | 1 | 9 | GJA5 |
| M1/M0 | Up | GO: Biological Proces | GO:00613 renal tubu | 5.49E-03 | 1.00E+00 | 3.11E-02 | 2.56E-01 | 2 | 96 | COL4A1,CTNNB1 |
| M1/M0 | Up | GO: Biological Proces | GO:00069 syncytium | 5.49E-03 | 1.00E+00 | 3.11E-02 | 2.56E-01 | 2 | 96 | CD81,ITGB1 |
| M1/M0 | Up | GO: Biological Proces | GO:00031 heart valv | 5.71E-03 | 1.00E+00 | 3.11E-02 | 2.56E-01 | 2 | 98 | GJA5,ITGB1 |
| M1/M0 | Up | GO: Biological Proces | GO:00716 cellular res | 5.78E-03 | 1.00E+00 | 3.11E-02 | 2.56E-01 | 1 | 5 | CTNNB1 |
| M1/M0 | Up | GO: Biological Proces | GO:00329 regulation | 5.78E-03 | 1.00E+00 | 3.11E-02 | 2.56E-01 | 1 | 5 | LRRC32 |
| M1/M0 | Up | GO: Biological Proces | GO:00329 transformi | 5.78E-03 | 1.00E+00 | 3.11E-02 | 2.56E-01 | 1 | 5 | LRRC32 |
| M1/M0 | Up | GO: Biological Proces | GO:00609 cardiac ce | 5.78E-03 | 1.00E+00 | 3.11E-02 | 2.56E-01 | 1 | 5 | ITGB1 |
| M1/M0 | Up | GO: Biological Proces | GO:00904 acinar cell | 5.78E-03 | 1.00E+00 | 3.11E-02 | 2.56E-01 | 1 | 5 | CTNNB1 |
| M1/M0 | Up | GO: Biological Proces | GO:19050 positive re | 5.78E-03 | 1.00E+00 | 3.11E-02 | 2.56E-01 | 1 | 5 | ENG |
| M1/M0 | Up | GO: Biological Proces | GO:19001 regulation | 5.78E-03 | 1.00E+00 | 3.11E-02 | 2.56E-01 | 1 | 5 | GJA5 |
| M1/M0 | Up | GO: Biological Proces | GO:19045 regulation | 5.78E-03 | 1.00E+00 | 3.11E-02 | 2.56E-01 | 1 | 5 | ZFP36L1 |
| M1/M0 | Up | GO: Biological Proces | GO:00989 regulation | 5.78E-03 | 1.00E+00 | 3.11E-02 | 2.56E-01 | 1 | 5 | GJA5 |
| M1/M0 | Up | GO: Biological Proces | GO:19027 positive re | 5.78E-03 | 1.00E+00 | 3.11E-02 | 2.56E-01 | 1 | 5 | CTNNB1 |
| M1/M0 | Up | GO: Biological Proces | GO:00613 cell prolife | 5.78E-03 | 1.00E+00 | 3.11E-02 | 2.56E-01 | 1 | 5 | CTNNB1 |
| M1/M0 | Up | GO: Biological Proces | GO:00604 primary lu | 5.78E-03 | 1.00E+00 | 3.11E-02 | 2.56E-01 | 1 | 5 | CTNNB1 |
| M1/M0 | Up | GO: Biological Proces | GO:00030 detection | 5.78E-03 | 1.00E+00 | 3.11E-02 | 2.56E-01 | 1 | 5 | ENG |
| M1/M0 | Up | GO: Biological Proces | GO:20011 positive re | 5.78E-03 | 1.00E+00 | 3.11E-02 | 2.56E-01 | 1 | 5 | CD81 |
| M1/M0 | Up | GO: Biological Proces | GO:00706 regulation | 5.78E-03 | 1.00E+00 | 3.11E-02 | 2.56E-01 | 1 | 5 | CTNNB1 |
| M1/M0 | Up | GO: Biological Proces | GO:19052 atrioventri | 5.78E-03 | 1.00E+00 | 3.11E-02 | 2.56E-01 | 1 | 5 | ENG |
| M1/M0 | Up | GO: Biological Proces | GO:00724 I-kappaB k | 5.91E-03 | 1.00E+00 | 3.17E-02 | 2.61E-01 | 3 | 322 | FLNA,CTNNB1,ECM1 |
| M1/M0 | Up | GO: Biological Proces | GO:19030 positive re | 5.94E-03 | 1.00E+00 | 3.18E-02 | 2.62E-01 | 2 | 100 | FLNA,ITGB1 |
| M1/M0 | Up | GO: Biological Proces | GO:00068 receptor-m | 6.06E-03 | 1.00E+00 | 3.23E-02 | 2.66E-01 | 3 | 325 | HSPG2,CD81,ITGB1 |
| M1/M0 | Up | GO: Biological Proces | GO:00309 hindbrain | 6.06E-03 | 1.00E+00 | 3.23E-02 | 2.66E-01 | 3 | 325 | FLNA,CTNNB1,ITGB1 |
| M1/M0 | Up | GO: Biological Proces | GO:00350 regulation | 6.17E-03 | 1.00E+00 | 3.28E-02 | 2.70E-01 | 2 | 102 | HEG1,ITGB1 |
| M1/M0 | Up | GO: Biological Proces | GO:00076 visual beh | 6.29E-03 | 1.00E+00 | 3.33E-02 | 2.75E-01 | 2 | 103 | ITGB1,SGK1 |
| M1/M0 | Up | GO: Biological Proces | GO:00075 skeletal m | 6.38E-03 | 1.00E+00 | 3.37E-02 | 2.78E-01 | 3 | 331 | HSPG2,CTNNB1,RCAN1 |
| M1/M0 | Up | GO: Biological Proces | GO:00060 proteoglyc | 6.53E-03 | 1.00E+00 | 3.44E-02 | 2.84E-01 | 2 | 105 | CTNNB1,BGN |
| M1/M0 | Up | GO: Biological Proces | GO:00025 myeloid le | 6.59E-03 | 1.00E+00 | 3.45E-02 | 2.84E-01 | 3 | 335 | CD81,CTNNB1,ZFP36L1 |
| M1/M0 | Up | GO: Biological Proces | GO:00726 protein loc | 6.65E-03 | 1.00E+00 | 3.45E-02 | 2.84E-01 | 3 | 336 | FLNA,CD81,ITGB1 |
| M1/M0 | Up | GO: Biological Proces | GO:00613 cardiac co | 6.77E-03 | 1.00E+00 | 3.45E-02 | 2.84E-01 | 2 | 107 | FLNA,GJA5 |
| M1/M0 | Up | GO: Biological Proces | GO:00360 limb joint | 6.93E-03 | 1.00E+00 | 3.45E-02 | 2.84E-01 | 1 | 6 | CTNNB1 |
| M1/M0 | Up | GO: Biological Proces | GO:00031 negative n | 6.93E-03 | 1.00E+00 | 3.45E-02 | 2.84E-01 | 1 | 6 | GJA5 |
| M1/M0 | Up | GO: Biological Proces | GO:19043 positive re | 6.93E-03 | 1.00E+00 | 3.45E-02 | 2.84E-01 | 1 | 6 | CD81 |
| M1/M0 | Up | GO: Biological Proces | GO:19047 positive re | 6.93E-03 | 1.00E+00 | 3.45E-02 | 2.84E-01 | 1 | 6 | CTNNB1 |
| M1/M0 | Up | GO: Biological Proces | GO:00609 mesenchy | 6.93E-03 | 1.00E+00 | 3.45E-02 | 2.84E-01 | 1 | 6 | CTNNB1 |
| M1/M0 | Up | GO: Biological Proces | GO:20006 negative n | 6.93E-03 | 1.00E+00 | 3.45E-02 | 2.84E-01 | 1 | 6 | CTNNB1 |
| M1/M0 | Up | GO: Biological Proces | GO:20000 regulation | 6.93E-03 | 1.00E+00 | 3.45E-02 | 2.84E-01 | 1 | 6 | CTNNB1 |
| M1/M0 | Up | GO: Biological Proces | GO:19042 regulation | 6.93E-03 | 1.00E+00 | 3.45E-02 | 2.84E-01 | 1 | 6 | SERINC1 |
| M1/M0 | Up | GO: Biological Proces | GO:00032 septum pr | 6.93E-03 | 1.00E+00 | 3.45E-02 | 2.84E-01 | 1 | 6 | GJA5 |
| M1/M0 | Up | GO: Biological Proces | GO:00611 fungiform | 6.93E-03 | 1.00E+00 | 3.45E-02 | 2.84E-01 | 1 | 6 | CTNNB1 |
| M1/M0 | Up | GO: Biological Proces | GO:00071 adenylate | 6.93E-03 | 1.00E+00 | 3.45E-02 | 2.84E-01 | 1 | 6 | FLNA |
| M1/M0 | Up | GO: Biological Proces | GO:00518 regulation | 6.93E-03 | 1.00E+00 | 3.45E-02 | 2.84E-01 | 1 | 6 | CTNNB1 |
| M1/M0 | Up | GO: Biological Proces | GO:19058 epididymis | 6.93E-03 | 1.00E+00 | 3.45E-02 | 2.84E-01 | 1 | 6 | CTNNB1 |
| M1/M0 | Up | GO: Biological Proces | GO:00860 Purkinje m | 6.93E-03 | 1.00E+00 | 3.45E-02 | 2.84E-01 | 1 | 6 | GJA5 |
| M1/M0 | Up | GO: Biological Proces | GO:00860 Purkinje m | 6.93E-03 | 1.00E+00 | 3.45E-02 | 2.84E-01 | 1 | 6 | GJA5 |
| M1/M0 | Up | GO: Biological Proces | GO:00607 hair follicl | 6.93E-03 | 1.00E+00 | 3.45E-02 | 2.84E-01 | 1 | 6 | CTNNB1 |
| M1/M0 | Up | GO: Biological Proces | GO:00600 oviduct de | 6.93E-03 | 1.00E+00 | 3.45E-02 | 2.84E-01 | 1 | 6 | CTNNB1 |
| M1/M0 | Up | GO: Biological Proces | GO:19008 regulation | 6.93E-03 | 1.00E+00 | 3.45E-02 | 2.84E-01 | 1 | 6 | GJA5 |
| M1/M0 | Up | GO: Biological Proces | GO:00443 canonical | 6.93E-03 | 1.00E+00 | 3.45E-02 | 2.84E-01 | 1 | 6 | CTNNB1 |
| M1/M0 | Up | GO: Biological Proces | GO:00519 glutamate | 6.93E-03 | 1.00E+00 | 3.45E-02 | 2.84E-01 | 1 | 6 | ITGB1 |
| M1/M0 | Up | GO: Biological Proces | GO:00902 positive re | 6.93E-03 | 1.00E+00 | 3.45E-02 | 2.84E-01 | 1 | 6 | HEG1 |
| M1/M0 | Up | GO: Biological Proces | GO:00507 positive re | 7.26E-03 | 1.00E+00 | 3.59E-02 | 2.96E-01 | 3 | 347 | CTNNB1,ITGB1,SGK1 |
| M1/M0 | Up | GO: Biological Proces | GO:00107 positive re | 7.27E-03 | 1.00E+00 | 3.59E-02 | 2.96E-01 | 2 | 111 | FLNA,SGK1 |
| M1/M0 | Up | GO: Biological Proces | GO:00224 membrane | 7.27E-03 | 1.00E+00 | 3.59E-02 | 2.96E-01 | 2 | 111 | VCAM1,MSN |
| M1/M0 | Up | GO: Biological Proces | GO:19013 regulation | 7.52E-03 | 1.00E+00 | 3.71E-02 | 3.06E-01 | 2 | 113 | FLNA,ITGB1 |
| M1/M0 | Up | GO: Biological Proces | GO:00605 skeletal m | 7.55E-03 | 1.00E+00 | 3.72E-02 | 3.06E-01 | 3 | 352 | HSPG2,CTNNB1,RCAN1 |
| M1/M0 | Up | GO: Biological Proces | GO:00435 endothelia | 7.67E-03 | 1.00E+00 | 3.77E-02 | 3.10E-01 | 3 | 354 | HSPG2,COL18A1,ITGB1 |
| M1/M0 | Up | GO: Biological Proces | GO:00716 response t | 8.08E-03 | 1.00E+00 | 3.78E-02 | 3.11E-01 | 1 | 7 | CTNNB1 |
| M1/M0 | Up | GO: Biological Proces | GO:00610 regulation | 8.08E-03 | 1.00E+00 | 3.78E-02 | 3.11E-01 | 1 | 7 | CTNNB1 |
| M1/M0 | Up | GO: Biological Proces | GO:00359 detection | 8.08E-03 | 1.00E+00 | 3.78E-02 | 3.11E-01 | 1 | 7 | CTNNB1 |
| M1/M0 | Up | GO: Biological Proces | GO:00726 osteoclast | 8.08E-03 | 1.00E+00 | 3.78E-02 | 3.11E-01 | 1 | 7 | CD81 |
| M1/M0 | Up | GO: Biological Proces | GO:00611 fungiform | 8.08E-03 | 1.00E+00 | 3.78E-02 | 3.11E-01 | 1 | 7 | CTNNB1 |
| M1/M0 | Up | GO: Biological Proces | GO:19050 regulation | 8.08E-03 | 1.00E+00 | 3.78E-02 | 3.11E-01 | 1 | 7 | FLNA |
| M1/M0 | Up | GO: Biological Proces | GO:00989 membrane | 8.08E-03 | 1.00E+00 | 3.78E-02 | 3.11E-01 | 1 | 7 | FLNA |
| M1/M0 | Up | GO: Biological Proces | GO:19023 protein loc | 8.08E-03 | 1.00E+00 | 3.78E-02 | 3.11E-01 | 1 | 7 | FLNA |
| M1/M0 | Up | GO: Biological Proces | GO:00860 bundle of | 8.08E-03 | 1.00E+00 | 3.78E-02 | 3.11E-01 | 1 | 7 | GJA5 |
| M1/M0 | Up | GO: Biological Proces | GO:00860 bundle of | 8.08E-03 | 1.00E+00 | 3.78E-02 | 3.11E-01 | 1 | 7 | GJA5 |
| M1/M0 | Up | GO: Biological Proces | GO:00989 regulation | 8.08E-03 | 1.00E+00 | 3.78E-02 | 3.11E-01 | 1 | 7 | GJA5 |
| M1/M0 | Up | GO: Biological Proces | GO:01500 regulation | 8.08E-03 | 1.00E+00 | 3.78E-02 | 3.11E-01 | 1 | 7 | ITGB1 |
| M1/M0 | Up | GO: Biological Proces | GO:00486 myoblast f | 8.08E-03 | 1.00E+00 | 3.78E-02 | 3.11E-01 | 1 | 7 | ITGB1 |
| M1/M0 | Up | GO: Biological Proces | GO:00198 peptide cr | 8.08E-03 | 1.00E+00 | 3.78E-02 | 3.11E-01 | 1 | 7 | BGN |
| M1/M0 | Up | GO: Biological Proces | GO:00604 trachea fo | 8.08E-03 | 1.00E+00 | 3.78E-02 | 3.11E-01 | 1 | 7 | CTNNB1 |
| M1/M0 | Up | GO: Biological Proces | GO:00613 retinal blo | 8.08E-03 | 1.00E+00 | 3.78E-02 | 3.11E-01 | 1 | 7 | COL4A1 |
| M1/M0 | Up | GO: Biological Proces | GO:00715 lymphocyt | 8.08E-03 | 1.00E+00 | 3.78E-02 | 3.11E-01 | 1 | 7 | MSN |
| M1/M0 | Up | GO: Biological Proces | GO:00515 positive re | 8.08E-03 | 1.00E+00 | 3.78E-02 | 3.11E-01 | 1 | 7 | ITGB1 |
| M1/M0 | Up | GO: Biological Proces | GO:00149 myoblast f | 8.08E-03 | 1.00E+00 | 3.78E-02 | 3.11E-01 | 1 | 7 | CD81 |
| M1/M0 | Up | GO: Biological Proces | GO:00443 canonical | 8.08E-03 | 1.00E+00 | 3.78E-02 | 3.11E-01 | 1 | 7 | CTNNB1 |
| M1/M0 | Up | GO: Biological Proces | GO:00443 canonical | 8.08E-03 | 1.00E+00 | 3.78E-02 | 3.11E-01 | 1 | 7 | CTNNB1 |
| M1/M0 | Up | GO: Biological Proces | GO:00310 nuclear-tra | 8.08E-03 | 1.00E+00 | 3.78E-02 | 3.11E-01 | 1 | 7 | ZFP36L1 |
| M1/M0 | Up | GO: Biological Proces | GO:00017 neural cre | 8.18E-03 | 1.00E+00 | 3.81E-02 | 3.14E-01 | 2 | 118 | COL18A1,ENG |
| M1/M0 | Up | GO: Biological Proces | GO:00702 actin-med | 8.18E-03 | 1.00E+00 | 3.81E-02 | 3.14E-01 | 2 | 118 | FLNA,GJA5 |
| M1/M0 | Up | GO: Biological Proces | GO:00099 negative n | 8.20E-03 | 1.00E+00 | 3.81E-02 | 3.14E-01 | 6 | 1591 | HEG1,CTNNB1,RCAN1,ECM1,ENG,ITGB1 |
| M1/M0 | Up | GO: Biological Proces | GO:00316 cellular res | 8.28E-03 | 1.00E+00 | 3.84E-02 | 3.16E-01 | 3 | 364 | CTNNB1,ITGB1,VCAM1 |
| M1/M0 | Up | GO: Biological Proces | GO:00018 embryonic | 8.31E-03 | 1.00E+00 | 3.84E-02 | 3.17E-01 | 2 | 119 | ZFP36L1,VCAM1 |
| M1/M0 | Up | GO: Biological Proces | GO:00107 regulation | 8.45E-03 | 1.00E+00 | 3.90E-02 | 3.21E-01 | 2 | 120 | CTNNB1,ENG |
| M1/M0 | Up | GO: Biological Proces | GO:00904 mesenchy | 8.58E-03 | 1.00E+00 | 3.95E-02 | 3.26E-01 | 2 | 121 | COL18A1,ENG |
| M1/M0 | Up | GO: Biological Proces | GO:00602 regulation | 8.70E-03 | 1.00E+00 | 4.00E-02 | 3.29E-01 | 4 | 717 | FLNA,CTNNB1,ITGB1,SGK1 |
| M1/M0 | Up | GO: Biological Proces | GO:00098 regulation | 9.17E-03 | 1.00E+00 | 4.05E-02 | 3.34E-01 | 5 | 1150 | FLNA,CD81,ZFP36L1,ITGB1,MSN |
| M1/M0 | Up | GO: Biological Proces | GO:00550 muscle cel | 8.85E-03 | 1.00E+00 | 4.05E-02 | 3.34E-01 | 3 | 373 | RCAN1,ENG,ITGB1 |
| M1/M0 | Up | GO: Biological Proces | GO:00076 learning o | 9.18E-03 | 1.00E+00 | 4.05E-02 | 3.34E-01 | 3 | 378 | RCAN1,ITGB1,SGK1 |
| M1/M0 | Up | GO: Biological Proces | GO:00226 regulation | 9.18E-03 | 1.00E+00 | 4.05E-02 | 3.34E-01 | 3 | 378 | FLNA,SGK1,MSN |
| M1/M0 | Up | GO: Biological Proces | GO:00330 T cell diffe | 9.14E-03 | 1.00E+00 | 4.05E-02 | 3.34E-01 | 2 | 125 | CTNNB1,ZFP36L1 |
| M1/M0 | Up | GO: Biological Proces | GO:00720 regulation | 9.14E-03 | 1.00E+00 | 4.05E-02 | 3.34E-01 | 2 | 125 | CTNNB1,ZFP36L1 |
| M1/M0 | Up | GO: Biological Proces | GO:19021 regulation | 9.23E-03 | 1.00E+00 | 4.05E-02 | 3.34E-01 | 1 | 8 | ZFP36L1 |
| M1/M0 | Up | GO: Biological Proces | GO:19047 regulation | 9.23E-03 | 1.00E+00 | 4.05E-02 | 3.34E-01 | 1 | 8 | CTNNB1 |
| M1/M0 | Up | GO: Biological Proces | GO:20006 positive re | 9.23E-03 | 1.00E+00 | 4.05E-02 | 3.34E-01 | 1 | 8 | MSN |
| M1/M0 | Up | GO: Biological Proces | GO:00342 macropha | 9.23E-03 | 1.00E+00 | 4.05E-02 | 3.34E-01 | 1 | 8 | CD81 |
| M1/M0 | Up | GO: Biological Proces | GO:00905 endocardi | 9.23E-03 | 1.00E+00 | 4.05E-02 | 3.34E-01 | 1 | 8 | ENG |
| M1/M0 | Up | GO: Biological Proces | GO:19050 regulation | 9.23E-03 | 1.00E+00 | 4.05E-02 | 3.34E-01 | 1 | 8 | ENG |
| M1/M0 | Up | GO: Biological Proces | GO:00619 articular ca | 9.23E-03 | 1.00E+00 | 4.05E-02 | 3.34E-01 | 1 | 8 | BGN |
| M1/M0 | Up | GO: Biological Proces | GO:00514 response t | 9.23E-03 | 1.00E+00 | 4.05E-02 | 3.34E-01 | 1 | 8 | IGFBP7 |
| M1/M0 | Up | GO: Biological Proces | GO:00603 regulation | 9.23E-03 | 1.00E+00 | 4.05E-02 | 3.34E-01 | 1 | 8 | FLNA |
| M1/M0 | Up | GO: Biological Proces | GO:00603 regulation | 9.23E-03 | 1.00E+00 | 4.05E-02 | 3.34E-01 | 1 | 8 | GJA5 |
| M1/M0 | Up | GO: Biological Proces | GO:00468 receptor-m | 9.23E-03 | 1.00E+00 | 4.05E-02 | 3.34E-01 | 1 | 8 | CD81 |
| M1/M0 | Up | GO: Biological Proces | GO:20002 regulation | 9.23E-03 | 1.00E+00 | 4.05E-02 | 3.34E-01 | 1 | 8 | HEG1 |
| M1/M0 | Up | GO: Biological Proces | GO:20011 regulation | 9.23E-03 | 1.00E+00 | 4.05E-02 | 3.34E-01 | 1 | 8 | CD81 |
| M1/M0 | Up | GO: Biological Proces | GO:00720 renal vesic | 9.23E-03 | 1.00E+00 | 4.05E-02 | 3.34E-01 | 1 | 8 | CTNNB1 |
| M1/M0 | Up | GO: Biological Proces | GO:00972 keratinocy | 9.23E-03 | 1.00E+00 | 4.05E-02 | 3.34E-01 | 1 | 8 | ZFP36L1 |
| M1/M0 | Up | GO: Biological Proces | GO:00550 atrial card | 9.23E-03 | 1.00E+00 | 4.05E-02 | 3.34E-01 | 1 | 8 | ENG |
| M1/M0 | Up | GO: Biological Proces | GO:00713 cellular res | 9.28E-03 | 1.00E+00 | 4.06E-02 | 3.35E-01 | 2 | 126 | ZFP36L1,SGK1 |
| M1/M0 | Up | GO: Biological Proces | GO:00432 positive re | 9.51E-03 | 1.00E+00 | 4.15E-02 | 3.42E-01 | 3 | 383 | FLNA,ITGB1,SGK1 |
| M1/M0 | Up | GO: Biological Proces | GO:19054 positive re | 9.56E-03 | 1.00E+00 | 4.17E-02 | 3.43E-01 | 2 | 128 | CD81,ITGB1 |
| M1/M0 | Up | GO: Biological Proces | GO:19024 protein loc | 9.71E-03 | 1.00E+00 | 4.20E-02 | 3.47E-01 | 2 | 129 | FLNA,HEG1 |
| M1/M0 | Up | GO: Biological Proces | GO:00019 positive re | 9.71E-03 | 1.00E+00 | 4.20E-02 | 3.47E-01 | 2 | 129 | COL18A1,ECM1 |
| M1/M0 | Up | GO: Biological Proces | GO:00083 associative | 9.71E-03 | 1.00E+00 | 4.20E-02 | 3.47E-01 | 2 | 129 | ITGB1,SGK1 |
| M1/M0 | Up | GO: Biological Proces | GO:00100 glial cell d | 9.85E-03 | 1.00E+00 | 4.24E-02 | 3.50E-01 | 3 | 388 | S100A6,CTNNB1,ITGB1 |
| M1/M0 | Up | GO: Biological Proces | GO:00613 renal tubu | 9.85E-03 | 1.00E+00 | 4.24E-02 | 3.50E-01 | 2 | 130 | COL4A1,CTNNB1 |
| M1/M0 | Up | GO: Biological Proces | GO:00030 regulation | 9.85E-03 | 1.00E+00 | 4.24E-02 | 3.50E-01 | 2 | 130 | GJA5,ENG |
| M1/M0 | Up | GO: Biological Proces | GO:00002 microtubu | 9.97E-03 | 1.00E+00 | 4.29E-02 | 3.53E-01 | 4 | 746 | FLNA,CTNNB1,ITGB1,SGK1 |
| M1/M0 | Up | GO: Biological Proces | GO:00905 epiboly | 1.00E-02 | 1.00E+00 | 4.29E-02 | 3.53E-01 | 2 | 131 | FLNA,ITGB1 |
| M1/M0 | Up | GO: Biological Proces | GO:00510 positive re | 1.03E-02 | 1.00E+00 | 4.30E-02 | 3.55E-01 | 5 | 1182 | FLNA,CD81,ITGB1,SGK1,MSN |
| M1/M0 | Up | GO: Biological Proces | GO:00098 determina | 1.03E-02 | 1.00E+00 | 4.30E-02 | 3.55E-01 | 3 | 394 | FLNA,CTNNB1,ENG |
| M1/M0 | Up | GO: Biological Proces | GO:00097 specificatio | 1.03E-02 | 1.00E+00 | 4.30E-02 | 3.55E-01 | 3 | 394 | FLNA,CTNNB1,ENG |
| M1/M0 | Up | GO: Biological Proces | GO:00164 negative n | 1.04E-02 | 1.00E+00 | 4.30E-02 | 3.55E-01 | 1 | 9 | FLNA |
| M1/M0 | Up | GO: Biological Proces | GO:00107 regulation | 1.04E-02 | 1.00E+00 | 4.30E-02 | 3.55E-01 | 1 | 9 | ITGB1 |
| M1/M0 | Up | GO: Biological Proces | GO:19029 regulation | 1.04E-02 | 1.00E+00 | 4.30E-02 | 3.55E-01 | 1 | 9 | MSN |
| M1/M0 | Up | GO: Biological Proces | GO:19029 positive re | 1.04E-02 | 1.00E+00 | 4.30E-02 | 3.55E-01 | 1 | 9 | MSN |
| M1/M0 | Up | GO: Biological Proces | GO:00615 sympathet | 1.04E-02 | 1.00E+00 | 4.30E-02 | 3.55E-01 | 1 | 9 | CTNNB1 |
| M1/M0 | Up | GO: Biological Proces | GO:00620 positive re | 1.04E-02 | 1.00E+00 | 4.30E-02 | 3.55E-01 | 1 | 9 | ENG |
| M1/M0 | Up | GO: Biological Proces | GO:19019 regulation | 1.04E-02 | 1.00E+00 | 4.30E-02 | 3.55E-01 | 1 | 9 | ITGB1 |
| M1/M0 | Up | GO: Biological Proces | GO:19050 cardiac jel | 1.04E-02 | 1.00E+00 | 4.30E-02 | 3.55E-01 | 1 | 9 | ENG |
| M1/M0 | Up | GO: Biological Proces | GO:00033 proepicard | 1.04E-02 | 1.00E+00 | 4.30E-02 | 3.55E-01 | 1 | 9 | ZFP36L1 |
| M1/M0 | Up | GO: Biological Proces | GO:00033 septum tra | 1.04E-02 | 1.00E+00 | 4.30E-02 | 3.55E-01 | 1 | 9 | ZFP36L1 |
| M1/M0 | Up | GO: Biological Proces | GO:00033 regulation | 1.04E-02 | 1.00E+00 | 4.30E-02 | 3.55E-01 | 1 | 9 | CTNNB1 |
| M1/M0 | Up | GO: Biological Proces | GO:00020 renin secr | 1.04E-02 | 1.00E+00 | 4.30E-02 | 3.55E-01 | 1 | 9 | GJA5 |
| M1/M0 | Up | GO: Biological Proces | GO:00020 regulation | 1.04E-02 | 1.00E+00 | 4.30E-02 | 3.55E-01 | 1 | 9 | ITGB1 |
| M1/M0 | Up | GO: Biological Proces | GO:19013 regulation | 1.04E-02 | 1.00E+00 | 4.30E-02 | 3.55E-01 | 1 | 9 | LRRC32 |
| M1/M0 | Up | GO: Biological Proces | GO:00485 oocyte dev | 1.04E-02 | 1.00E+00 | 4.31E-02 | 3.55E-01 | 2 | 134 | CTNNB1,PGRMC2 |
| M1/M0 | Up | GO: Biological Proces | GO:00107 regulation | 1.04E-02 | 1.00E+00 | 4.31E-02 | 3.55E-01 | 2 | 134 | FLNA,SGK1 |
| M1/M0 | Up | GO: Biological Proces | GO:00726 lymphocyt | 1.07E-02 | 1.00E+00 | 4.42E-02 | 3.64E-01 | 2 | 136 | ECM1,MSN |
| M1/M0 | Up | GO: Biological Proces | GO:00456 positive re | 1.07E-02 | 1.00E+00 | 4.42E-02 | 3.64E-01 | 2 | 136 | TCF4,ITGB1 |
| M1/M0 | Up | GO: Biological Proces | GO:00481 regulation | 1.09E-02 | 1.00E+00 | 4.47E-02 | 3.69E-01 | 2 | 137 | S100A6,CTNNB1 |
| M1/M0 | Up | GO: Biological Proces | GO:00432 regulation | 1.10E-02 | 1.00E+00 | 4.53E-02 | 3.73E-01 | 2 | 138 | FLNA,ITGB1 |
| M1/M0 | Up | GO: Biological Proces | GO:00080 blood circ | 1.11E-02 | 1.00E+00 | 4.53E-02 | 3.73E-01 | 4 | 769 | FLNA,GJA5,ENG,SGK1 |
| M1/M0 | Up | GO: Biological Proces | GO:19907 protein loc | 1.11E-02 | 1.00E+00 | 4.55E-02 | 3.75E-01 | 3 | 406 | FLNA,CD81,ITGB1 |
| M1/M0 | Up | GO: Biological Proces | GO:00722 integrin-m | 1.12E-02 | 1.00E+00 | 4.56E-02 | 3.76E-01 | 2 | 139 | FLNA,ITGB1 |
| M1/M0 | Up | GO: Biological Proces | GO:00421 regulation | 1.13E-02 | 1.00E+00 | 4.57E-02 | 3.77E-01 | 3 | 408 | FLNA,CD81,MSN |
| M1/M0 | Up | GO: Biological Proces | GO:00519 positive re | 1.14E-02 | 1.00E+00 | 4.57E-02 | 3.77E-01 | 3 | 410 | CTNNB1,ITGB1,SGK1 |
| M1/M0 | Up | GO: Biological Proces | GO:00713 cellular res | 1.15E-02 | 1.00E+00 | 4.57E-02 | 3.77E-01 | 2 | 141 | ZFP36L1,SGK1 |
| M1/M0 | Up | GO: Biological Proces | GO:00099 oocyte dif | 1.15E-02 | 1.00E+00 | 4.57E-02 | 3.77E-01 | 2 | 141 | CTNNB1,PGRMC2 |
| M1/M0 | Up | GO: Biological Proces | GO:00488 regulation | 1.15E-02 | 1.00E+00 | 4.57E-02 | 3.77E-01 | 1 | 10 | CTNNB1 |
| M1/M0 | Up | GO: Biological Proces | GO:00609 coronary a | 1.15E-02 | 1.00E+00 | 4.57E-02 | 3.77E-01 | 1 | 10 | CTNNB1 |
| M1/M0 | Up | GO: Biological Proces | GO:00032 ventricular | 1.15E-02 | 1.00E+00 | 4.57E-02 | 3.77E-01 | 1 | 10 | CTNNB1 |
| M1/M0 | Up | GO: Biological Proces | GO:00726 multinucle | 1.15E-02 | 1.00E+00 | 4.57E-02 | 3.77E-01 | 1 | 10 | CD81 |
| M1/M0 | Up | GO: Biological Proces | GO:00508 positive re | 1.15E-02 | 1.00E+00 | 4.57E-02 | 3.77E-01 | 1 | 10 | CD81 |
| M1/M0 | Up | GO: Biological Proces | GO:20005 positive re | 1.15E-02 | 1.00E+00 | 4.57E-02 | 3.77E-01 | 1 | 10 | CD81 |
| M1/M0 | Up | GO: Biological Proces | GO:00603 regulation | 1.15E-02 | 1.00E+00 | 4.57E-02 | 3.77E-01 | 1 | 10 | GJA5 |
| M1/M0 | Up | GO: Biological Proces | GO:00019 renal resp | 1.15E-02 | 1.00E+00 | 4.57E-02 | 3.77E-01 | 1 | 10 | GJA5 |
| M1/M0 | Up | GO: Biological Proces | GO:00989 regulation | 1.15E-02 | 1.00E+00 | 4.57E-02 | 3.77E-01 | 1 | 10 | FLNA |
| M1/M0 | Up | GO: Biological Proces | GO:00519 amino aci | 1.15E-02 | 1.00E+00 | 4.57E-02 | 3.77E-01 | 1 | 10 | ITGB1 |
| M1/M0 | Up | GO: Biological Proces | GO:19016 response t | 1.19E-02 | 1.00E+00 | 4.69E-02 | 3.87E-01 | 4 | 785 | CTNNB1,ZFP36L1,SGK1,VCAM1 |
| M1/M0 | Up | GO: Biological Proces | GO:00094 response t | 1.19E-02 | 1.00E+00 | 4.69E-02 | 3.87E-01 | 4 | 785 | COL18A1,CTNNB1,ENG,ITGB1 |
| M1/M0 | Up | GO: Biological Proces | GO:00329 collagen m | 1.20E-02 | 1.00E+00 | 4.73E-02 | 3.90E-01 | 2 | 144 | ENG,ITGB1 |
| M1/M0 | Up | GO: Biological Proces | GO:00020 lens devel | 1.21E-02 | 1.00E+00 | 4.78E-02 | 3.94E-01 | 2 | 145 | TCF4,CTNNB1 |
| M1/M0 | Up | GO: Biological Proces | GO:00716 regulation | 1.23E-02 | 1.00E+00 | 4.83E-02 | 3.98E-01 | 2 | 146 | ECM1,MSN |
| M1/M0 | Up | GO: Biological Proces | GO:00323 regulation | 1.24E-02 | 1.00E+00 | 4.85E-02 | 4.00E-01 | 3 | 422 | FLNA,CD81,MSN |
| M1/M0 | Up | GO: Biological Proces | GO:00508 cognition | 1.26E-02 | 1.00E+00 | 4.87E-02 | 4.01E-01 | 3 | 425 | RCAN1,ITGB1,SGK1 |

| | | | | | | | | | | |
|---|---|---|---|---|---|---|---|---|---|---|
| M1/M0 | Up | GO: Biological Proces | GO:00996 atrial card | 1.27E-02 | 1.00E+00 | 4.87E-02 | 4.01E-01 | 1 | 11 | FLNA |
| M1/M0 | Up | GO: Biological Proces | GO:06105 visceral se | 1.27E-02 | 1.00E+00 | 4.87E-02 | 4.01E-01 | 1 | 11 | VCAM1 |
| M1/M0 | Up | GO: Biological Proces | GO:01906 virion atta | 1.27E-02 | 1.00E+00 | 4.87E-02 | 4.01E-01 | 1 | 11 | CD81 |
| M1/M0 | Up | GO: Biological Proces | GO:00032 regulation | 1.27E-02 | 1.00E+00 | 4.87E-02 | 4.01E-01 | 1 | 11 | CTNNB1 |
| M1/M0 | Up | GO: Biological Proces | GO:00325 protein tra | 1.27E-02 | 1.00E+00 | 4.87E-02 | 4.01E-01 | 1 | 11 | ITGB1 |
| M1/M0 | Up | GO: Biological Proces | GO:00607 regulation | 1.27E-02 | 1.00E+00 | 4.87E-02 | 4.01E-01 | 1 | 11 | CTNNB1 |
| M1/M0 | Up | GO: Biological Proces | GO:00604 lung-assoc | 1.27E-02 | 1.00E+00 | 4.87E-02 | 4.01E-01 | 1 | 11 | CTNNB1 |
| M1/M0 | Up | GO: Biological Proces | GO:01588 heme tran | 1.27E-02 | 1.00E+00 | 4.87E-02 | 4.01E-01 | 1 | 11 | PGRMC2 |
| M1/M0 | Up | GO: Biological Proces | GO:00359 dorsal aor | 1.27E-02 | 1.00E+00 | 4.87E-02 | 4.01E-01 | 1 | 11 | ENG |
| M1/M0 | Up | GO: Biological Proces | GO:01490 myotube c | 1.27E-02 | 1.00E+00 | 4.87E-02 | 4.01E-01 | 1 | 11 | CD81 |
| M1/M0 | Up | GO: Biological Proces | GO:00323 negative r | 1.27E-02 | 1.00E+00 | 4.87E-02 | 4.01E-01 | 1 | 11 | CTNNB1 |
| M1/M0 | Up | GO: Biological Proces | GO:01064 negative r | 1.27E-02 | 1.00E+00 | 4.88E-02 | 4.02E-01 | 6 | 1747 | HEG1,CTNNB1,RCAN1,ECM1,ENG,ITGB1 |
| M1/M0 | Up | GO: Biological Proces | GO:00303 embryonic | 1.28E-02 | 1.00E+00 | 4.88E-02 | 4.02E-01 | 2 | 149 | GJA5,CTNNB1 |
| M1/M0 | Up | GO: Biological Proces | GO:05185 positive re | 1.28E-02 | 1.00E+00 | 4.88E-02 | 4.02E-01 | 2 | 149 | ENG,ITGB1 |
| M1/M0 | Up | GO: Biological Proces | GO:02305 negative r | 1.29E-02 | 1.00E+00 | 4.91E-02 | 4.05E-01 | 6 | 1751 | HEG1,CTNNB1,RCAN1,ECM1,ENG,ITGB1 |
| M1/M0 | Up | GO: Biological Proces | GO:03004 actin filam | 1.29E-02 | 1.00E+00 | 4.93E-02 | 4.06E-01 | 2 | 150 | FLNA,GJA5 |
| M1/M0 | Up | GO: Biological Proces | GO:05501 cardiac mu | 1.31E-02 | 1.00E+00 | 4.98E-02 | 4.10E-01 | 2 | 151 | HEG1,CTNNB1 |
| M1/M0 | Up | GO: Cellular Compon | GO:00309 collagen ty | 1.15E-03 | 2.65E-01 | 1.02E-02 | 6.14E-02 | 1 | 1 | COL18A1 |
| M1/M0 | Up | GO: Cellular Compon | GO:07106 alpha9-bet | 1.15E-03 | 2.65E-01 | 1.02E-02 | 6.14E-02 | 1 | 1 | VCAM1 |
| M1/M0 | Up | GO: Cellular Compon | GO:03466 integrin al | 2.29E-03 | 5.29E-01 | 1.43E-02 | 8.62E-02 | 1 | 2 | ITGB1 |
| M1/M0 | Up | GO: Cellular Compon | GO:03466 integrin al | 2.29E-03 | 5.29E-01 | 1.43E-02 | 8.62E-02 | 1 | 2 | ITGB1 |
| M1/M0 | Up | GO: Cellular Compon | GO:03468 integrin al | 2.29E-03 | 5.29E-01 | 1.43E-02 | 8.62E-02 | 1 | 2 | ITGB1 |
| M1/M0 | Up | GO: Cellular Compon | GO:03468 integrin al | 2.29E-03 | 5.29E-01 | 1.43E-02 | 8.62E-02 | 1 | 2 | ITGB1 |
| M1/M0 | Up | GO: Cellular Compon | GO:19907 beta-cater | 2.29E-03 | 5.29E-01 | 1.43E-02 | 8.62E-02 | 1 | 2 | CTNNB1 |
| M1/M0 | Up | GO: Cellular Compon | GO:03461 integrin al | 2.29E-03 | 5.29E-01 | 1.43E-02 | 8.62E-02 | 1 | 2 | ITGB1 |
| M1/M0 | Up | GO: Cellular Compon | GO:03461 integrin al | 2.29E-03 | 5.29E-01 | 1.43E-02 | 8.62E-02 | 1 | 2 | ITGB1 |
| M1/M0 | Up | GO: Cellular Compon | GO:03461 integrin al | 2.29E-03 | 5.29E-01 | 1.43E-02 | 8.62E-02 | 1 | 2 | ITGB1 |
| M1/M0 | Up | GO: Cellular Compon | GO:03461 integrin al | 2.29E-03 | 5.29E-01 | 1.43E-02 | 8.62E-02 | 1 | 2 | ITGB1 |
| M1/M0 | Up | GO: Cellular Compon | GO:03475 Scrib-APC- | 3.44E-03 | 7.94E-01 | 1.89E-02 | 1.14E-01 | 1 | 3 | CTNNB1 |
| M1/M0 | Up | GO: Cellular Compon | GO:03466 integrin al | 3.44E-03 | 7.94E-01 | 1.89E-02 | 1.14E-01 | 1 | 3 | ITGB1 |
| M1/M0 | Up | GO: Cellular Compon | GO:03466 integrin al | 3.44E-03 | 7.94E-01 | 1.89E-02 | 1.14E-01 | 1 | 3 | ITGB1 |
| M1/M0 | Up | GO: Cellular Compon | GO:07256 endothelia | 3.44E-03 | 7.94E-01 | 1.89E-02 | 1.14E-01 | 1 | 3 | ENG |
| M1/M0 | Up | GO: Cellular Compon | GO:03152 Myb comp | 4.58E-03 | 1.00E+00 | 2.35E-02 | 1.42E-01 | 1 | 4 | FLNA |
| M1/M0 | Up | GO: Cellular Compon | GO:19907 glycoprote | 5.72E-03 | 1.00E+00 | 2.64E-02 | 1.59E-01 | 1 | 5 | FLNA |
| M1/M0 | Up | GO: Cellular Compon | GO:00558 collagen ty | 8.00E-03 | 1.00E+00 | 3.19E-02 | 1.92E-01 | 1 | 7 | COL4A1 |
| M1/M0 | Up | GO: Cellular Compon | GO:03574 myelin she | 1.03E-02 | 1.00E+00 | 3.89E-02 | 2.34E-01 | 1 | 9 | ITGB1 |
| M1/M0 | Up | GO: Cellular Compon | GO:09864 network-fo | 1.03E-02 | 1.00E+00 | 3.89E-02 | 2.34E-01 | 1 | 9 | COL4A1 |
| M1/M0 | Up | GO: Cellular Compon | GO:09864 collagen n | 1.03E-02 | 1.00E+00 | 3.89E-02 | 2.34E-01 | 1 | 9 | COL4A1 |
| M1/M0 | Up | GO: Cellular Compon | GO:09719 tetraspani | 1.14E-02 | 1.00E+00 | 4.12E-02 | 2.48E-01 | 1 | 10 | CD81 |
| M1/M0 | Up | GO: Cellular Compon | GO:05128 cell tip | 1.14E-02 | 1.00E+00 | 4.12E-02 | 2.48E-01 | 1 | 10 | MSN |
| M1/M0 | Up | GO: Cellular Compon | GO:01660 flotillin co | 1.14E-02 | 1.00E+00 | 4.12E-02 | 2.48E-01 | 1 | 10 | CTNNB1 |
| M1/M0 | Up | GO: Cellular Compon | GO:03005 hemidesm | 1.25E-02 | 1.00E+00 | 4.33E-02 | 2.60E-01 | 1 | 11 | ITGB1 |
| M1/M0 | Up | GO: Cellular Compon | GO:00591 zonula adh | 1.37E-02 | 1.00E+00 | 4.65E-02 | 2.80E-01 | 1 | 12 | CTNNB1 |
| M1/M0 | Up | GO: Cellular Compon | GO:00193 uropod | 1.48E-02 | 1.00E+00 | 4.75E-02 | 2.86E-01 | 1 | 13 | MSN |
| M1/M0 | Up | GO: Cellular Compon | GO:03125 cell trailing | 1.48E-02 | 1.00E+00 | 4.75E-02 | 2.86E-01 | 1 | 13 | MSN |
| M1/M0 | Up | GO: Cellular Compon | GO:00591 fascia adh | 1.48E-02 | 1.00E+00 | 4.75E-02 | 2.86E-01 | 1 | 13 | CTNNB1 |
| M1/M0 | Up | GO: Cellular Compon | GO:03081 beta-cater | 1.59E-02 | 1.00E+00 | 4.91E-02 | 2.96E-01 | 1 | 14 | CTNNB1 |
| M1/M0 | Up | GO: Cellular Compon | GO:03108 platelet de | 1.59E-02 | 1.00E+00 | 4.91E-02 | 2.96E-01 | 1 | 14 | ECM1 |
| M1/M0 | Up | GO: Cellular Compon | GO:06018 cell pole | 1.59E-02 | 1.00E+00 | 4.91E-02 | 2.96E-01 | 1 | 14 | MSN |
| M0/Control | Down | GO: Biological Proces | GO:03405 response t | 3.69E-08 | 4.02E-05 | 2.42E-05 | 1.83E-04 | 7 | 1349 | FOS,ZFP36,KLF4,DUSP1,EGR1,KLF2,JUNB |
| M0/Control | Down | GO: Biological Proces | GO:01407 response t | 1.36E-07 | 1.48E-04 | 4.54E-05 | 3.44E-04 | 7 | 1628 | FOS,ZFP36,KLF4,DUSP1,EGR1,KLF2,JUNB |
| M0/Control | Down | GO: Biological Proces | GO:07145 cellular res | 4.02E-07 | 4.37E-04 | 5.47E-05 | 4.14E-04 | 7 | 1903 | FOS,ZFP36,KLF4,DUSP1,EGR1,KLF2,JUNB |
| M0/Control | Down | GO: Biological Proces | GO:01003 response t | 2.09E-07 | 2.27E-04 | 4.54E-05 | 3.44E-04 | 6 | 930 | FOS,KLF4,DUSP1,RHOB,KLF2,JUNB |
| M0/Control | Down | GO: Biological Proces | GO:07134 cellular res | 9.25E-07 | 1.01E-03 | 6.59E-05 | 4.99E-04 | 6 | 1196 | FOS,ZFP36,KLF4,DUSP1,EGR1,KLF2 |
| M0/Control | Down | GO: Biological Proces | GO:03009 hemopoie | 3.61E-06 | 3.93E-03 | 1.57E-04 | 1.19E-03 | 6 | 1507 | FOS,ZFP36,KLF4,EGR1,KLF2,JUNB |
| M0/Control | Down | GO: Biological Proces | GO:04853 hematopo | 4.51E-06 | 4.91E-03 | 1.76E-04 | 1.33E-03 | 6 | 1565 | FOS,ZFP36,KLF4,EGR1,KLF2,JUNB |
| M0/Control | Down | GO: Biological Proces | GO:03399 response t | 4.54E-06 | 4.95E-03 | 1.76E-04 | 1.33E-03 | 6 | 1567 | FOS,ZFP36,KLF4,DUSP1,EGR1,JUNB |
| M0/Control | Down | GO: Biological Proces | GO:01024 response t | 5.58E-06 | 6.08E-03 | 1.92E-04 | 1.46E-03 | 6 | 1623 | FOS,KLF4,DUSP1,EGR1,KLF2,JUNB |
| M0/Control | Down | GO: Biological Proces | GO:00252 immune sy | 5.85E-06 | 6.37E-03 | 1.93E-04 | 1.46E-03 | 6 | 1636 | FOS,ZFP36,KLF4,EGR1,KLF2,JUNB |
| M0/Control | Down | GO: Biological Proces | GO:19016 response t | 8.34E-06 | 9.08E-03 | 2.46E-04 | 1.86E-03 | 6 | 1738 | FOS,KLF4,DUSP1,EGR1,KLF2,JUNB |
| M0/Control | Down | GO: Biological Proces | GO:19017 cellular res | 1.08E-05 | 1.17E-02 | 3.08E-04 | 2.33E-03 | 6 | 1815 | FOS,ZFP36,KLF4,RHOB,EGR1,KLF2 |
| M0/Control | Down | GO: Biological Proces | GO:20002 regulation | 1.44E-05 | 1.57E-02 | 4.02E-04 | 3.04E-03 | 6 | 1908 | FOS,ZFP36,KLF4,RHOB,KLF2,JUNB |
| M0/Control | Down | GO: Molecular Functi | GO:00097 RNA polyr | 5.23E-05 | 3.82E-03 | 8.29E-04 | 4.04E-03 | 5 | 1286 | FOS,KLF4,EGR1,KLF2,JUNB |
| M0/Control | Down | GO: Molecular Functi | GO:00098 cis-regulat | 5.68E-05 | 4.15E-03 | 8.29E-04 | 4.04E-03 | 5 | 1308 | FOS,KLF4,EGR1,KLF2,JUNB |
| M0/Control | Down | GO: Molecular Functi | GO:00098 DNA-bind | 8.65E-05 | 6.31E-03 | 9.90E-04 | 4.83E-03 | 5 | 1427 | FOS,KLF4,EGR1,KLF2,JUNB |
| M0/Control | Down | GO: Molecular Functi | GO:00370 DNA-bind | 1.04E-04 | 7.60E-03 | 9.90E-04 | 4.83E-03 | 5 | 1483 | FOS,KLF4,EGR1,KLF2,JUNB |
| M0/Control | Down | GO: Molecular Functi | GO:00097 RNA polyr | 1.09E-04 | 7.92E-03 | 9.90E-04 | 4.83E-03 | 5 | 1496 | FOS,KLF4,EGR1,KLF2,JUNB |
| M0/Control | Down | GO: Molecular Functi | GO:00097 transcripti | 1.49E-04 | 1.09E-02 | 1.11E-03 | 5.40E-03 | 5 | 1599 | FOS,KLF4,EGR1,KLF2,JUNB |
| M0/Control | Down | GO: Molecular Functi | GO:00106 transcripti | 1.52E-04 | 1.11E-02 | 1.11E-03 | 5.40E-03 | 5 | 1604 | FOS,KLF4,EGR1,KLF2,JUNB |
| M0/Control | Down | GO: Molecular Functi | GO:19908 sequence- | 1.78E-04 | 1.30E-02 | 1.18E-03 | 5.75E-03 | 5 | 1658 | FOS,KLF4,EGR1,KLF2,JUNB |
| M0/Control | Down | GO: Molecular Functi | GO:04356 sequence- | 2.46E-04 | 1.80E-02 | 1.29E-03 | 6.27E-03 | 5 | 1775 | FOS,KLF4,EGR1,KLF2,JUNB |
| M0/Control | Down | GO: Molecular Functi | GO:00036 double-str | 2.47E-04 | 1.80E-02 | 1.29E-03 | 6.27E-03 | 5 | 1776 | FOS,KLF4,EGR1,KLF2,JUNB |
| M0/Control | Down | GO: Biological Proces | GO:00030 response t | 4.45E-08 | 4.85E-05 | 2.42E-05 | 1.83E-04 | 5 | 318 | FOS,KLF4,DUSP1,RHOB,KLF2 |
| M0/Control | Down | GO: Biological Proces | GO:19016 response t | 2.86E-07 | 3.11E-04 | 4.79E-05 | 3.63E-04 | 5 | 462 | FOS,KLF4,DUSP1,KLF2,JUNB |
| M0/Control | Down | GO: Biological Proces | GO:06211 cellular res | 3.08E-07 | 3.35E-04 | 4.79E-05 | 3.63E-04 | 5 | 469 | FOS,KLF4,RHOB,EGR1,KLF2 |
| M0/Control | Down | GO: Biological Proces | GO:09730 response t | 5.75E-07 | 6.27E-04 | 6.59E-05 | 4.99E-04 | 5 | 532 | FOS,KLF4,EGR1,KLF2,JUNB |
| M0/Control | Down | GO: Biological Proces | GO:04854 response t | 9.28E-07 | 1.01E-03 | 6.59E-05 | 4.99E-04 | 5 | 586 | FOS,ZFP36,DUSP1,EGR1,JUNB |
| M0/Control | Down | GO: Biological Proces | GO:00069 response t | 9.68E-07 | 1.05E-03 | 6.59E-05 | 4.99E-04 | 5 | 591 | FOS,KLF4,DUSP1,RHOB,KLF2 |
| M0/Control | Down | GO: Biological Proces | GO:00093 response t | 1.50E-06 | 1.64E-03 | 8.61E-05 | 6.52E-04 | 5 | 646 | FOS,DUSP1,RHOB,EGR1,JUNB |
| M0/Control | Down | GO: Biological Proces | GO:19016 response t | 3.92E-06 | 4.27E-03 | 1.64E-04 | 1.24E-03 | 5 | 785 | FOS,KLF4,EGR1,KLF2,JUNB |
| M0/Control | Down | GO: Biological Proces | GO:03281 cellular res | 6.36E-06 | 6.92E-03 | 2.04E-04 | 1.54E-03 | 5 | 866 | FOS,ZFP36,DUSP1,EGR1,JUNB |
| M0/Control | Down | GO: Biological Proces | GO:00015 blood vess | 2.56E-05 | 2.79E-02 | 6.64E-04 | 5.03E-03 | 5 | 1152 | KLF4,RHOB,EGR1,KLF2,JUNB |
| M0/Control | Down | GO: Biological Proces | GO:00019 vasculatur | 3.65E-05 | 3.97E-02 | 9.03E-04 | 6.84E-03 | 5 | 1239 | KLF4,RHOB,EGR1,KLF2,JUNB |
| M0/Control | Down | GO: Biological Proces | GO:00097 response t | 5.94E-05 | 6.46E-02 | 1.35E-03 | 1.02E-02 | 5 | 1370 | FOS,ZFP36,DUSP1,EGR1,JUNB |
| M0/Control | Down | GO: Biological Proces | GO:04459 positive re | 7.23E-05 | 7.87E-02 | 1.51E-03 | 1.15E-02 | 5 | 1427 | FOS,KLF4,EGR1,KLF2,JUNB |
| M0/Control | Down | GO: Biological Proces | GO:05105 positive re | 2.04E-04 | 2.23E-01 | 3.48E-03 | 2.63E-02 | 5 | 1772 | FOS,ZFP36,KLF4,RHOB,JUNB |
| M0/Control | Down | GO: Biological Proces | GO:00096 response t | 2.12E-04 | 2.31E-01 | 3.54E-03 | 2.68E-02 | 5 | 1786 | FOS,DUSP1,RHOB,EGR1,JUNB |
| M0/Control | Down | GO: Biological Proces | GO:04459 negative r | 2.16E-04 | 2.36E-01 | 3.54E-03 | 2.68E-02 | 5 | 1793 | ZFP36,KLF4,DUSP1,EGR1,KLF2 |
| M0/Control | Down | GO: Biological Proces | GO:01055 negative r | 2.50E-04 | 2.72E-01 | 3.78E-03 | 2.86E-02 | 5 | 1848 | ZFP36,KLF4,DUSP1,EGR1,KLF2 |
| M0/Control | Down | GO: Biological Proces | GO:04864 anatomica | 2.78E-04 | 3.03E-01 | 3.79E-03 | 2.87E-02 | 5 | 1890 | KLF4,DUSP1,RHOB,KLF2,JUNB |
| M0/Control | Down | GO: Biological Proces | GO:04298 regulation | 2.84E-04 | 3.10E-01 | 3.79E-03 | 2.87E-02 | 5 | 1899 | ZFP36,KLF4,DUSP1,RHOB,EGR1 |
| M0/Control | Down | GO: Biological Proces | GO:19035 positive re | 2.85E-04 | 3.10E-01 | 3.79E-03 | 2.87E-02 | 5 | 1900 | FOS,KLF4,EGR1,KLF2,JUNB |
| M0/Control | Down | GO: Biological Proces | GO:04458 positive re | 2.85E-04 | 3.10E-01 | 3.79E-03 | 2.87E-02 | 5 | 1900 | FOS,KLF4,EGR1,KLF2,JUNB |
| M0/Control | Down | GO: Biological Proces | GO:19026 positive re | 2.89E-04 | 3.15E-01 | 3.80E-03 | 2.87E-02 | 5 | 1906 | FOS,KLF4,EGR1,KLF2,JUNB |
| M0/Control | Down | GO: Biological Proces | GO:00313 negative r | 3.16E-04 | 3.44E-01 | 4.00E-03 | 3.03E-02 | 5 | 1942 | ZFP36,KLF4,DUSP1,EGR1,KLF2 |
| M0/Control | Down | GO: Biological Proces | GO:04306 regulation | 3.18E-04 | 3.46E-01 | 4.00E-03 | 3.03E-02 | 5 | 1944 | ZFP36,KLF4,DUSP1,RHOB,EGR1 |
| M0/Control | Down | GO: Biological Proces | GO:07235 circulatory | 3.44E-04 | 3.75E-01 | 4.08E-03 | 3.09E-02 | 5 | 1977 | KLF4,RHOB,EGR1,KLF2,JUNB |
| M0/Control | Down | GO: Biological Proces | GO:00098 negative r | 3.57E-04 | 3.89E-01 | 4.08E-03 | 3.09E-02 | 5 | 1992 | ZFP36,KLF4,DUSP1,EGR1,KLF2 |
| M0/Control | Down | GO: Cellular Compon | GO:00007 chromatin | 6.70E-05 | 2.48E-03 | 1.24E-03 | 5.21E-03 | 5 | 1419 | FOS,KLF4,EGR1,KLF2,JUNB |
| M0/Control | Down | GO: Molecular Functi | GO:00122 DNA-bind | 4.17E-05 | 3.05E-03 | 8.29E-04 | 4.04E-03 | 4 | 569 | FOS,KLF4,EGR1,JUNB |
| M0/Control | Down | GO: Molecular Functi | GO:00121 DNA-bind | 4.32E-05 | 3.15E-03 | 8.29E-04 | 4.04E-03 | 4 | 574 | FOS,KLF4,EGR1,JUNB |
| M0/Control | Down | GO: Biological Proces | GO:00515 response t | 1.86E-07 | 2.02E-04 | 4.54E-05 | 3.44E-04 | 4 | 151 | FOS,DUSP1,EGR1,JUNB |
| M0/Control | Down | GO: Biological Proces | GO:04666 response t | 6.19E-07 | 6.74E-04 | 6.59E-05 | 4.99E-04 | 4 | 204 | FOS,DUSP1,EGR1,JUNB |
| M0/Control | Down | GO: Biological Proces | GO:03461 cellular res | 8.84E-07 | 9.63E-04 | 6.59E-05 | 4.99E-04 | 4 | 223 | FOS,KLF4,RHOB,KLF2 |
| M0/Control | Down | GO: Biological Proces | GO:04254 response t | 9.32E-07 | 1.02E-03 | 6.59E-05 | 4.99E-04 | 4 | 226 | KLF4,DUSP1,RHOB,KLF2 |
| M0/Control | Down | GO: Biological Proces | GO:01407 response t | 1.11E-06 | 1.21E-03 | 7.10E-05 | 5.37E-04 | 4 | 236 | FOS,DUSP1,EGR1,JUNB |
| M0/Control | Down | GO: Biological Proces | GO:05138 response t | 3.59E-06 | 3.91E-03 | 1.57E-04 | 1.19E-03 | 4 | 317 | FOS,ZFP36,DUSP1,JUNB |
| M0/Control | Down | GO: Biological Proces | GO:03319 response t | 5.49E-06 | 5.98E-03 | 1.92E-04 | 1.46E-03 | 4 | 353 | FOS,ZFP36,DUSP1,JUNB |
| M0/Control | Down | GO: Biological Proces | GO:03455 cellular res | 7.67E-06 | 8.35E-03 | 2.32E-04 | 1.76E-03 | 4 | 384 | FOS,KLF4,RHOB,KLF2 |
| M0/Control | Down | GO: Biological Proces | GO:19016 cellular res | 2.85E-05 | 3.11E-02 | 7.23E-04 | 5.47E-03 | 4 | 536 | FOS,KLF4,EGR1,KLF2 |
| M0/Control | Down | GO: Biological Proces | GO:03005 myeloid ce | 6.64E-05 | 7.23E-02 | 1.42E-03 | 1.07E-02 | 4 | 665 | FOS,ZFP36,KLF2,JUNB |
| M0/Control | Down | GO: Biological Proces | GO:00109 positive re | 1.54E-04 | 1.68E-01 | 2.85E-03 | 2.16E-02 | 4 | 826 | FOS,DUSP1,RHOB,EGR1 |
| M0/Control | Down | GO: Biological Proces | GO:07140 cellular res | 2.00E-04 | 2.18E-01 | 3.45E-03 | 2.62E-02 | 4 | 883 | ZFP36,KLF4,EGR1,KLF2 |
| M0/Control | Down | GO: Biological Proces | GO:07135 cellular res | 2.67E-04 | 2.91E-01 | 3.79E-03 | 2.87E-02 | 4 | 952 | FOS,ZFP36,KLF4,EGR1 |
| M0/Control | Down | GO: Biological Proces | GO:07136 cellular res | 2.79E-04 | 3.04E-01 | 3.79E-03 | 2.87E-02 | 4 | 963 | FOS,ZFP36,KLF4,EGR1 |
| M0/Control | Down | GO: Biological Proces | GO:07141 cellular res | 2.85E-04 | 3.10E-01 | 3.79E-03 | 2.87E-02 | 4 | 968 | FOS,KLF4,EGR1,KLF2 |
| M0/Control | Down | GO: Biological Proces | GO:00018 regulation | 3.11E-04 | 3.38E-01 | 4.00E-03 | 3.03E-02 | 4 | 990 | ZFP36,KLF4,EGR1,KLF2 |
| M0/Control | Down | GO: Biological Proces | GO:07084 response t | 3.30E-04 | 3.60E-01 | 4.04E-03 | 3.06E-02 | 4 | 1006 | FOS,ZFP36,KLF4,EGR1 |
| M0/Control | Down | GO: Biological Proces | GO:04851 blood vess | 3.60E-04 | 3.93E-01 | 4.08E-03 | 3.09E-02 | 4 | 1029 | KLF4,RHOB,KLF2,JUNB |
| M0/Control | Down | GO: Biological Proces | GO:19016 cellular res | 3.84E-04 | 4.18E-01 | 4.08E-03 | 3.09E-02 | 4 | 1046 | FOS,KLF4,EGR1,KLF2 |
| M0/Control | Down | GO: Biological Proces | GO:00181 cytokine p | 4.04E-04 | 4.40E-01 | 4.20E-03 | 3.18E-02 | 4 | 1060 | ZFP36,KLF4,EGR1,KLF2 |
| M0/Control | Down | GO: Biological Proces | GO:05133 regulation | 4.05E-04 | 4.42E-01 | 4.20E-03 | 3.18E-02 | 4 | 1061 | ZFP36,KLF4,DUSP1,EGR1 |
| M0/Control | Down | GO: Biological Proces | GO:00012 negative r | 5.04E-04 | 5.49E-01 | 5.05E-03 | 3.82E-02 | 4 | 1123 | ZFP36,KLF4,EGR1,KLF2 |
| M0/Control | Down | GO: Biological Proces | GO:04856 embryonic | 6.19E-04 | 6.74E-01 | 5.67E-03 | 4.29E-02 | 4 | 1185 | KLF4,DUSP1,EGR1,KLF2 |
| M0/Control | Down | GO: Biological Proces | GO:03303 regulation | 7.56E-04 | 8.23E-01 | 6.39E-03 | 4.83E-02 | 4 | 1249 | KLF4,DUSP1,RHOB,EGR1 |
| M0/Control | Down | GO: Biological Proces | GO:04300 chordate e | 7.75E-04 | 8.44E-01 | 6.39E-03 | 4.83E-02 | 4 | 1257 | KLF4,EGR1,KLF2,JUNB |
| M0/Control | Down | GO: Biological Proces | GO:00097 embryo de | 8.32E-04 | 9.06E-01 | 6.76E-03 | 5.12E-02 | 4 | 1281 | KLF4,EGR1,KLF2,JUNB |
| M0/Control | Down | GO: Biological Proces | GO:20001 regulation | 9.35E-04 | 1.00E+00 | 7.16E-03 | 5.42E-02 | 4 | 1321 | KLF4,DUSP1,RHOB,EGR1 |
| M0/Control | Down | GO: Biological Proces | GO:05172 regulation | 1.07E-03 | 1.00E+00 | 7.86E-03 | 5.95E-02 | 4 | 1368 | KLF4,DUSP1,RHOB,JUNB |
| M0/Control | Down | GO: Biological Proces | GO:04400 regulation | 1.10E-03 | 1.00E+00 | 8.03E-03 | 6.08E-02 | 4 | 1380 | KLF4,DUSP1,RHOB,EGR1 |
| M0/Control | Down | GO: Biological Proces | GO:03523 tube morp | 1.39E-03 | 1.00E+00 | 9.29E-03 | 7.03E-02 | 4 | 1467 | KLF4,RHOB,KLF2,JUNB |
| M0/Control | Down | GO: Biological Proces | GO:04458 negative r | 1.50E-03 | 1.00E+00 | 9.77E-03 | 7.40E-02 | 4 | 1497 | ZFP36,KLF4,EGR1,KLF2 |
| M0/Control | Down | GO: Biological Proces | GO:19035 negative r | 1.51E-03 | 1.00E+00 | 9.77E-03 | 7.40E-02 | 4 | 1500 | ZFP36,KLF4,EGR1,KLF2 |
| M0/Control | Down | GO: Biological Proces | GO:19026 negative r | 1.53E-03 | 1.00E+00 | 9.77E-03 | 7.40E-02 | 4 | 1504 | ZFP36,KLF4,EGR1,KLF2 |
| M0/Control | Down | GO: Biological Proces | GO:05125 negative r | 2.14E-03 | 1.00E+00 | 1.24E-02 | 9.35E-02 | 4 | 1645 | ZFP36,KLF4,EGR1,KLF2 |
| M0/Control | Down | GO: Biological Proces | GO:00026 regulation | 3.12E-03 | 1.00E+00 | 1.53E-02 | 1.16E-01 | 4 | 1821 | FOS,ZFP36,DUSP1,JUNB |
| M0/Control | Down | GO: Biological Proces | GO:04001 locomotio | 3.20E-03 | 1.00E+00 | 1.55E-02 | 1.17E-01 | 4 | 1834 | KLF4,DUSP1,RHOB,EGR1 |
| M0/Control | Down | GO: Biological Proces | GO:03529 tube deve | 3.51E-03 | 1.00E+00 | 1.61E-02 | 1.22E-01 | 4 | 1880 | KLF4,RHOB,KLF2,JUNB |
| M0/Control | Down | GO: Biological Proces | GO:05124 positive re | 4.01E-03 | 1.00E+00 | 1.75E-02 | 1.32E-01 | 4 | 1950 | FOS,KLF4,RHOB,EGR1 |
| M0/Control | Down | GO: Biological Proces | GO:06604 epithelium | 4.24E-03 | 1.00E+00 | 1.79E-02 | 1.35E-01 | 4 | 1979 | ZFP36,KLF4,RHOB,KLF2 |
| M0/Control | Down | GO: Molecular Functi | GO:00036 chromatin | 2.47E-03 | 1.80E-01 | 9.02E-03 | 4.40E-02 | 3 | 740 | FOS,KLF4,EGR1 |
| M0/Control | Down | GO: Molecular Functi | GO:00081 transcripti | 2.47E-03 | 1.80E-01 | 9.02E-03 | 4.40E-02 | 3 | 740 | FOS,KLF4,JUNB |
| M0/Control | Down | GO: Molecular Functi | GO:00199 protein kin | 3.73E-03 | 2.72E-01 | 1.24E-02 | 6.03E-02 | 3 | 855 | ZFP36,DUSP1,RHOB |
| M0/Control | Down | GO: Molecular Functi | GO:00199 kinase bin | 5.05E-03 | 3.69E-01 | 1.54E-02 | 7.49E-02 | 3 | 952 | ZFP36,DUSP1,RHOB |
| M0/Control | Down | GO: Biological Proces | GO:19028 positive re | 7.80E-07 | 8.49E-04 | 6.59E-05 | 4.99E-04 | 3 | 51 | FOS,KLF4,EGR1 |
| M0/Control | Down | GO: Biological Proces | GO:20006 positive re | 1.28E-06 | 1.39E-03 | 7.74E-05 | 5.86E-04 | 3 | 60 | FOS,KLF4,EGR1 |
| M0/Control | Down | GO: Biological Proces | GO:19028 regulation | 1.96E-06 | 2.13E-03 | 1.01E-04 | 7.66E-04 | 3 | 69 | FOS,KLF4,EGR1 |
| M0/Control | Down | GO: Biological Proces | GO:00161 miRNA tra | 2.04E-06 | 2.23E-03 | 1.01E-04 | 7.66E-04 | 3 | 70 | FOS,KLF4,EGR1 |
| M0/Control | Down | GO: Biological Proces | GO:20006 regulation | 3.55E-06 | 3.86E-03 | 1.57E-04 | 1.19E-03 | 3 | 84 | FOS,KLF4,EGR1 |
| M0/Control | Down | GO: Biological Proces | GO:01058 miRNA me | 5.65E-06 | 6.15E-03 | 1.92E-04 | 1.46E-03 | 3 | 98 | FOS,KLF4,EGR1 |
| M0/Control | Down | GO: Biological Proces | GO:00703 cellular res | 2.28E-05 | 2.48E-02 | 6.21E-04 | 4.70E-03 | 3 | 156 | KLF4,RHOB,KLF2 |
| M0/Control | Down | GO: Biological Proces | GO:00170 formation | 4.24E-05 | 4.62E-02 | 1.03E-03 | 7.77E-03 | 3 | 192 | KLF4,DUSP1,KLF2 |
| M0/Control | Down | GO: Biological Proces | GO:00515 response t | 4.51E-05 | 4.91E-02 | 1.07E-03 | 8.08E-03 | 3 | 196 | FOS,DUSP1,JUNB |
| M0/Control | Down | GO: Biological Proces | GO:00973 cellular res | 6.44E-05 | 7.02E-02 | 1.42E-03 | 1.07E-02 | 3 | 221 | FOS,KLF4,KLF2 |
| M0/Control | Down | GO: Biological Proces | GO:07145 cellular res | 6.53E-05 | 7.11E-02 | 1.42E-03 | 1.07E-02 | 3 | 222 | FOS,EGR1,JUNB |
| M0/Control | Down | GO: Biological Proces | GO:19016 cellular res | 9.29E-05 | 1.01E-01 | 1.86E-03 | 1.41E-02 | 3 | 250 | FOS,KLF4,KLF2 |
| M0/Control | Down | GO: Biological Proces | GO:03625 cellular res | 9.40E-05 | 1.02E-01 | 1.86E-03 | 1.41E-02 | 3 | 251 | FOS,EGR1,JUNB |
| M0/Control | Down | GO: Biological Proces | GO:07145 cellular res | 1.18E-04 | 1.29E-01 | 2.26E-03 | 1.71E-02 | 3 | 271 | FOS,EGR1,JUNB |
| M0/Control | Down | GO: Biological Proces | GO:07135 cellular res | 1.69E-04 | 1.84E-01 | 3.07E-03 | 2.32E-02 | 3 | 306 | FOS,ZFP36,KLF2 |
| M0/Control | Down | GO: Biological Proces | GO:00300 muscle cel | 2.19E-04 | 2.38E-01 | 3.54E-03 | 2.68E-02 | 3 | 334 | FOS,KLF4,EGR1 |
| M0/Control | Down | GO: Biological Proces | GO:03461 response t | 2.25E-04 | 2.45E-01 | 3.55E-03 | 2.69E-02 | 3 | 337 | FOS,ZFP36,KLF2 |
| M0/Control | Down | GO: Biological Proces | GO:00961 response t | 3.45E-04 | 3.76E-01 | 4.08E-03 | 3.09E-02 | 3 | 390 | FOS,EGR1,JUNB |
| M0/Control | Down | GO: Biological Proces | GO:00736 gastrulatic | 4.35E-04 | 4.74E-01 | 4.47E-03 | 3.39E-02 | 3 | 422 | KLF4,DUSP1,KLF2 |
| M0/Control | Down | GO: Biological Proces | GO:00303 negative r | 4.51E-04 | 4.91E-01 | 4.59E-03 | 3.47E-02 | 3 | 427 | KLF4,DUSP1,RHOB |
| M0/Control | Down | GO: Biological Proces | GO:20001 negative r | 5.05E-04 | 5.50E-01 | 5.05E-03 | 3.82E-02 | 3 | 444 | KLF4,DUSP1,RHOB |
| M0/Control | Down | GO: Biological Proces | GO:00941 response t | 5.22E-04 | 5.68E-01 | 5.08E-03 | 3.84E-02 | 3 | 449 | FOS,DUSP1,JUNB |
| M0/Control | Down | GO: Biological Proces | GO:00181 negative r | 5.22E-04 | 5.68E-01 | 5.08E-03 | 3.84E-02 | 3 | 449 | ZFP36,KLF4,KLF2 |
| M0/Control | Down | GO: Biological Proces | GO:04578 regulation | 5.75E-04 | 6.26E-01 | 5.44E-03 | 4.12E-02 | 3 | 464 | KLF4,RHOB,KLF2 |
| M0/Control | Down | GO: Biological Proces | GO:19013 regulation | 6.19E-04 | 6.74E-01 | 5.67E-03 | 4.29E-02 | 3 | 476 | KLF4,RHOB,KLF2 |
| M0/Control | Down | GO: Biological Proces | GO:04001 negative r | 6.38E-04 | 6.95E-01 | 5.77E-03 | 4.37E-02 | 3 | 481 | KLF4,DUSP1,RHOB |
| M0/Control | Down | GO: Biological Proces | GO:04578 negative r | 6.46E-04 | 7.04E-01 | 5.77E-03 | 4.37E-02 | 3 | 483 | KLF4,DUSP1,RHOB |
| M0/Control | Down | GO: Biological Proces | GO:00166 response t | 7.66E-04 | 8.34E-01 | 6.39E-03 | 4.83E-02 | 3 | 512 | FOS,EGR1,JUNB |
| M0/Control | Down | GO: Biological Proces | GO:00321 negative r | 8.80E-04 | 9.58E-01 | 7.07E-03 | 5.35E-02 | 3 | 537 | ZFP36,KLF4,DUSP1 |
| M0/Control | Down | GO: Biological Proces | GO:03105 regenerati | 8.89E-04 | 9.68E-01 | 7.07E-03 | 5.35E-02 | 3 | 539 | FOS,KLF4,JUNB |
| M0/Control | Down | GO: Biological Proces | GO:03625 response t | 9.28E-04 | 1.00E+00 | 7.16E-03 | 5.42E-02 | 3 | 547 | FOS,EGR1,JUNB |
| M0/Control | Down | GO: Biological Proces | GO:19037 regulation | 9.78E-04 | 1.00E+00 | 7.40E-03 | 5.60E-02 | 3 | 557 | FOS,ZFP36,JUNB |
| M0/Control | Down | GO: Biological Proces | GO:00751 muscle org | 1.01E-03 | 1.00E+00 | 7.62E-03 | 5.77E-02 | 3 | 564 | FOS,EGR1,KLF2 |
| M0/Control | Down | GO: Biological Proces | GO:01003 response t | 1.12E-03 | 1.00E+00 | 8.03E-03 | 6.08E-02 | 3 | 584 | FOS,DUSP1,JUNB |
| M0/Control | Down | GO: Biological Proces | GO:00170 in utero e | 1.21E-03 | 1.00E+00 | 8.36E-03 | 6.33E-02 | 3 | 600 | KLF4,KLF2,JUNB |
| M0/Control | Down | GO: Biological Proces | GO:19031 mononucl | 1.33E-03 | 1.00E+00 | 8.98E-03 | 6.80E-02 | 3 | 619 | FOS,EGR1,JUNB |
| M0/Control | Down | GO: Biological Proces | GO:07048 response t | 1.37E-03 | 1.00E+00 | 9.22E-03 | 6.98E-02 | 3 | 626 | FOS,EGR1,JUNB |
| M0/Control | Down | GO: Biological Proces | GO:03466 ncRNA me | 1.66E-03 | 1.00E+00 | 1.04E-02 | 7.84E-02 | 3 | 668 | FOS,KLF4,EGR1 |
| M0/Control | Down | GO: Biological Proces | GO:04343 response t | 1.72E-03 | 1.00E+00 | 1.07E-02 | 8.10E-02 | 3 | 677 | FOS,EGR1,JUNB |
| M0/Control | Down | GO: Biological Proces | GO:06055 muscle tis | 1.87E-03 | 1.00E+00 | 1.14E-02 | 8.60E-02 | 3 | 697 | FOS,EGR1,KLF2 |
| M0/Control | Down | GO: Biological Proces | GO:04306 positive re | 2.11E-03 | 1.00E+00 | 1.23E-02 | 9.31E-02 | 3 | 727 | DUSP1,RHOB,EGR1 |
| M0/Control | Down | GO: Biological Proces | GO:04306 positive re | 2.23E-03 | 1.00E+00 | 1.27E-02 | 9.64E-02 | 3 | 741 | DUSP1,RHOB,EGR1 |
| M0/Control | Down | GO: Biological Proces | GO:00941 response t | 2.63E-03 | 1.00E+00 | 1.42E-02 | 1.08E-01 | 3 | 785 | FOS,EGR1,JUNB |
| M0/Control | Down | GO: Biological Proces | GO:03166 response t | 2.74E-03 | 1.00E+00 | 1.43E-02 | 1.08E-01 | 3 | 796 | FOS,ZFP36,EGR1 |
| M0/Control | Down | GO: Biological Proces | GO:00025 leukocyte | 2.94E-03 | 1.00E+00 | 1.52E-02 | 1.15E-01 | 3 | 816 | FOS,EGR1,JUNB |
| M0/Control | Down | GO: Biological Proces | GO:00961 response t | 3.03E-03 | 1.00E+00 | 1.53E-02 | 1.16E-01 | 3 | 825 | ZFP36,KLF4,EGR1 |
| M0/Control | Down | GO: Biological Proces | GO:00099 response t | 3.14E-03 | 1.00E+00 | 1.53E-02 | 1.16E-01 | 3 | 835 | FOS,ZFP36,EGR1 |
| M0/Control | Down | GO: Biological Proces | GO:00015 angiogene | 3.56E-03 | 1.00E+00 | 1.62E-02 | 1.23E-01 | 3 | 873 | KLF4,RHOB,KLF2 |
| M0/Control | Down | GO: Biological Proces | GO:04354 regulation | 4.16E-03 | 1.00E+00 | 1.79E-02 | 1.35E-01 | 3 | 922 | KLF4,DUSP1,EGR1 |
| M0/Control | Down | GO: Biological Proces | GO:19030 mitotic cel | 4.16E-03 | 1.00E+00 | 1.79E-02 | 1.35E-01 | 3 | 922 | KLF4,DUSP1,RHOB |
| M0/Control | Down | GO: Biological Proces | GO:04308 negative r | 4.25E-03 | 1.00E+00 | 1.79E-02 | 1.35E-01 | 3 | 929 | ZFP36,KLF4,DUSP1 |
| M0/Control | Down | GO: Biological Proces | GO:00001 MAPK cas | 4.85E-03 | 1.00E+00 | 2.01E-02 | 1.52E-01 | 3 | 974 | ZFP36,KLF4,DUSP1 |
| M0/Control | Down | GO: Biological Proces | GO:06106 muscle str | 5.90E-03 | 1.00E+00 | 2.21E-02 | 1.67E-01 | 3 | 1044 | FOS,EGR1,KLF2 |
| M0/Control | Down | GO: Biological Proces | GO:00069 inflammat | 5.97E-03 | 1.00E+00 | 2.23E-02 | 1.68E-01 | 3 | 1048 | FOS,ZFP36,KLF4 |
| M0/Control | Down | GO: Biological Proces | GO:03085 epithelial c | 6.01E-03 | 1.00E+00 | 2.23E-02 | 1.69E-01 | 3 | 1051 | ZFP36,KLF4,KLF2 |
| M0/Control | Down | GO: Biological Proces | GO:00002 mitotic cel | 6.99E-03 | 1.00E+00 | 2.41E-02 | 1.83E-01 | 3 | 1109 | KLF4,DUSP1,RHOB |
| M0/Control | Down | GO: Biological Proces | GO:04872 tissue mor | 7.40E-03 | 1.00E+00 | 2.50E-02 | 1.90E-01 | 3 | 1132 | KLF4,RHOB,KLF2 |
| M0/Control | Down | GO: Biological Proces | GO:04856 developme | 8.08E-03 | 1.00E+00 | 2.68E-02 | 2.03E-01 | 3 | 1168 | FOS,KLF2,JUNB |
| M0/Control | Down | GO: Biological Proces | GO:04555 positive re | 8.69E-03 | 1.00E+00 | 2.82E-02 | 2.13E-01 | 3 | 1199 | FOS,ZFP36,JUNB |
| M0/Control | Down | GO: Biological Proces | GO:00062 DNA meta | 8.73E-03 | 1.00E+00 | 2.82E-02 | 2.14E-01 | 3 | 1201 | FOS,KLF4,DUSP1 |
| M0/Control | Down | GO: Biological Proces | GO:03210 regulation | 1.04E-02 | 1.00E+00 | 3.12E-02 | 2.36E-01 | 3 | 1280 | ZFP36,KLF4,DUSP1 |
| M0/Control | Down | GO: Biological Proces | GO:02260 regulation | 1.05E-02 | 1.00E+00 | 3.12E-02 | 2.36E-01 | 3 | 1284 | KLF4,RHOB,KLF2 |
| M0/Control | Down | GO: Biological Proces | GO:05105 negative r | 1.09E-02 | 1.00E+00 | 3.13E-02 | 2.37E-01 | 3 | 1299 | ZFP36,KLF4,KLF2 |
| M0/Control | Down | GO: Biological Proces | GO:05124 negative r | 1.16E-02 | 1.00E+00 | 3.29E-02 | 2.49E-01 | 3 | 1332 | ZFP36,KLF4,DUSP1 |
| M0/Control | Down | GO: Biological Proces | GO:04405 negative r | 1.34E-02 | 1.00E+00 | 3.58E-02 | 2.71E-01 | 3 | 1401 | ZFP36,KLF4,DUSP1 |
| M0/Control | Down | GO: Biological Proces | GO:00106 positive re | 1.52E-02 | 1.00E+00 | 3.88E-02 | 2.93E-01 | 3 | 1469 | ZFP36,KLF4,EGR1 |
| M0/Control | Down | GO: Biological Proces | GO:05124 negative r | 1.53E-02 | 1.00E+00 | 3.89E-02 | 2.94E-01 | 3 | 1472 | ZFP36,KLF4,KLF2 |
| M0/Control | Down | GO: Biological Proces | GO:00224 cell cycle p | 1.58E-02 | 1.00E+00 | 3.95E-02 | 2.99E-01 | 3 | 1490 | KLF4,DUSP1,RHOB |
| M0/Control | Down | GO: Biological Proces | GO:04232 regulation | 1.59E-02 | 1.00E+00 | 3.97E-02 | 3.01E-01 | 3 | 1495 | KLF4,DUSP1,EGR1 |
| M0/Control | Down | GO: Biological Proces | GO:04000 growth | 1.69E-02 | 1.00E+00 | 4.08E-02 | 3.09E-01 | 3 | 1528 | FOS,KLF2,JUNB |
| M0/Control | Down | GO: Biological Proces | GO:00099 negative r | 1.89E-02 | 1.00E+00 | 4.43E-02 | 3.35E-01 | 3 | 1591 | KLF4,DUSP1,EGR1 |
| M0/Control | Down | GO: Biological Proces | GO:00192 regulation | 2.20E-02 | 1.00E+00 | 4.84E-02 | 3.66E-01 | 3 | 1684 | KLF4,DUSP1,EGR1 |
| M0/Control | Down | GO: Biological Proces | GO:05111 regulation | 2.20E-02 | 1.00E+00 | 4.84E-02 | 3.66E-01 | 3 | 1685 | KLF4,DUSP1,EGR1 |
| M0/Control | Down | GO: Cellular Compon | GO:00056 transcripti | 1.12E-03 | 4.15E-02 | 1.38E-02 | 5.82E-02 | 3 | 590 | FOS,KLF4,JUNB |
| M0/Control | Down | GO: Molecular Functi | GO:00097 RNA polyr | 3.86E-05 | 2.82E-03 | 8.29E-04 | 4.04E-03 | 2 | 24 | FOS,EGR1 |
| M0/Control | Down | GO: Molecular Functi | GO:00104 core prom | 2.07E-04 | 1.51E-02 | 1.26E-03 | 6.12E-03 | 2 | 55 | FOS,EGR1 |
| M0/Control | Down | GO: Molecular Functi | GO:19908 promoter- | 4.48E-04 | 3.27E-02 | 2.18E-03 | 1.06E-02 | 2 | 81 | KLF4,EGR1 |
| M0/Control | Down | GO: Molecular Functi | GO:00122 transcripti | 1.24E-03 | 9.04E-02 | 5.31E-03 | 2.59E-02 | 2 | 135 | FOS,KLF4 |
| M0/Control | Down | GO: Molecular Functi | GO:00616 RNA polyr | 1.17E-02 | 8.53E-01 | 2.67E-02 | 1.30E-01 | 2 | 426 | FOS,KLF4 |
| M0/Control | Down | GO: Molecular Functi | GO:01402 DNA-bind | 2.05E-02 | 1.00E+00 | 3.94E-02 | 1.92E-01 | 2 | 573 | FOS,KLF4 |
| M0/Control | Down | GO: Biological Proces | GO:07146 cellular res | 1.96E-06 | 2.13E-03 | 1.01E-04 | 7.66E-04 | 2 | 6 | KLF4,KLF2 |
| M0/Control | Down | GO: Biological Proces | GO:07145 cellular res | 4.69E-06 | 5.11E-03 | 1.76E-04 | 1.33E-03 | 2 | 9 | KLF4,KLF2 |
| M0/Control | Down | GO: Biological Proces | GO:04688 response t | 7.16E-06 | 7.80E-03 | 2.23E-04 | 1.69E-03 | 2 | 11 | KLF4,KLF2 |
| M0/Control | Down | GO: Biological Proces | GO:03461 response t | 2.47E-05 | 2.69E-02 | 6.56E-04 | 4.97E-03 | 2 | 20 | KLF4,KLF2 |
| M0/Control | Down | GO: Biological Proces | GO:07145 cellular res | 5.64E-05 | 6.15E-02 | 1.31E-03 | 9.90E-03 | 2 | 30 | KLF4,KLF2 |
| M0/Control | Down | GO: Biological Proces | GO:00171 mesoderm | 8.16E-05 | 8.89E-02 | 1.68E-03 | 1.27E-02 | 2 | 36 | KLF4,KLF2 |
| M0/Control | Down | GO: Biological Proces | GO:01404 integrated | 1.06E-04 | 1.16E-01 | 2.06E-03 | 1.56E-02 | 2 | 41 | FOS,JUNB |
| M0/Control | Down | GO: Biological Proces | GO:05141 response t | 1.28E-04 | 1.40E-01 | 2.41E-03 | 1.82E-02 | 2 | 45 | FOS,JUNB |
| M0/Control | Down | GO: Biological Proces | GO:03446 response t | 1.78E-04 | 1.94E-01 | 3.18E-03 | 2.41E-02 | 2 | 53 | KLF4,KLF2 |
| M0/Control | Down | GO: Biological Proces | GO:04436 regulation | 1.92E-04 | 2.09E-01 | 3.37E-03 | 2.55E-02 | 2 | 55 | EGR1,KLF2 |
| M0/Control | Down | GO: Biological Proces | GO:07136 cellular res | 2.21E-04 | 2.40E-01 | 3.54E-03 | 2.68E-02 | 2 | 59 | FOS,ZFP36 |
| M0/Control | Down | GO: Biological Proces | GO:06007 cell fate co | 2.36E-04 | 2.57E-01 | 3.67E-03 | 2.78E-02 | 2 | 61 | KLF4,KLF2 |
| M0/Control | Down | GO: Biological Proces | GO:04436 regulation | 2.44E-04 | 2.66E-01 | 3.74E-03 | 2.83E-02 | 2 | 62 | EGR1,KLF2 |
| M0/Control | Down | GO: Biological Proces | GO:04833 mesoderm | 2.60E-04 | 2.83E-01 | 3.79E-03 | 2.87E-02 | 2 | 64 | KLF4,KLF2 |
| M0/Control | Down | GO: Biological Proces | GO:04542 positive re | 2.68E-04 | 2.92E-01 | 3.79E-03 | 2.87E-02 | 2 | 65 | KLF4,KLF2 |
| M0/Control | Down | GO: Biological Proces | GO:19044 positive re | 2.85E-04 | 3.10E-01 | 3.79E-03 | 2.87E-02 | 2 | 67 | KLF4,KLF2 |
| M0/Control | Down | GO: Biological Proces | GO:07084 response t | 3.20E-04 | 3.48E-01 | 4.00E-03 | 3.03E-02 | 2 | 71 | FOS,ZFP36 |
| M0/Control | Down | GO: Biological Proces | GO:03800 p38MAPK | 3.29E-04 | 3.58E-01 | 4.04E-03 | 3.06E-02 | 2 | 72 | ZFP36,DUSP1 |
| M0/Control | Down | GO: Biological Proces | GO:05138 response t | 3.76E-04 | 4.10E-01 | 4.08E-03 | 3.09E-02 | 2 | 77 | FOS,JUNB |
| M0/Control | Down | GO: Biological Proces | GO:03251 response t | 3.76E-04 | 4.10E-01 | 4.08E-03 | 3.09E-02 | 2 | 77 | FOS,JUNB |
| M0/Control | Down | GO: Biological Proces | GO:07141 cellular res | 5.14E-04 | 5.59E-01 | 5.08E-03 | 3.84E-02 | 2 | 90 | RHOB,EGR1 |
| M0/Control | Down | GO: Biological Proces | GO:00359 skeletal m | 5.48E-04 | 5.97E-01 | 5.28E-03 | 4.00E-02 | 2 | 93 | FOS,EGR1 |
| M0/Control | Down | GO: Biological Proces | GO:19036 regulation | 5.60E-04 | 6.10E-01 | 5.35E-03 | 4.05E-02 | 2 | 94 | KLF4,KLF2 |
| M0/Control | Down | GO: Biological Proces | GO:04542 regulation | 5.96E-04 | 6.49E-01 | 5.60E-03 | 4.24E-02 | 2 | 97 | KLF4,KLF2 |
| M0/Control | Down | GO: Biological Proces | GO:00703 negative r | 6.09E-04 | 6.63E-01 | 5.67E-03 | 4.29E-02 | 2 | 98 | KLF4,DUSP1 |
| M0/Control | Down | GO: Biological Proces | GO:07121 cellular res | 6.46E-04 | 7.04E-01 | 5.77E-03 | 4.37E-02 | 2 | 101 | FOS,JUNB |
| M0/Control | Down | GO: Biological Proces | GO:08016 regulation | 6.72E-04 | 7.32E-01 | 5.95E-03 | 4.50E-02 | 2 | 103 | KLF4,KLF2 |
| M0/Control | Down | GO: Biological Proces | GO:00068 nitric oxid | 7.80E-04 | 8.49E-01 | 6.39E-03 | 4.83E-02 | 2 | 111 | KLF4,KLF2 |
| M0/Control | Down | GO: Biological Proces | GO:00170 mesoderm | 8.95E-04 | 9.75E-01 | 7.07E-03 | 5.35E-02 | 2 | 119 | KLF4,KLF2 |
| M0/Control | Down | GO: Biological Proces | GO:04620 nitric oxid | 8.95E-04 | 9.75E-01 | 7.07E-03 | 5.35E-02 | 2 | 119 | KLF4,KLF2 |
| M0/Control | Down | GO: Biological Proces | GO:20010 reactive ni | 9.26E-04 | 1.00E+00 | 7.16E-03 | 5.42E-02 | 2 | 121 | KLF4,KLF2 |
| M0/Control | Down | GO: Biological Proces | GO:06021 hematopo | 9.26E-04 | 1.00E+00 | 7.16E-03 | 5.42E-02 | 2 | 121 | FOS,ZFP36 |
| M0/Control | Down | GO: Biological Proces | GO:00326 regulation | 9.41E-04 | 1.00E+00 | 7.16E-03 | 5.42E-02 | 2 | 122 | KLF4,EGR1 |
| M0/Control | Down | GO: Biological Proces | GO:00326 chemokine | 1.04E-03 | 1.00E+00 | 7.72E-03 | 5.84E-02 | 2 | 128 | KLF4,EGR1 |
| M0/Control | Down | GO: Biological Proces | GO:04833 mesoderm | 1.05E-03 | 1.00E+00 | 7.79E-03 | 5.89E-02 | 2 | 129 | KLF4,KLF2 |
| M0/Control | Down | GO: Biological Proces | GO:19012 positive re | 1.29E-03 | 1.00E+00 | 8.83E-03 | 6.68E-02 | 2 | 143 | FOS,EGR1 |
| M0/Control | Down | GO: Biological Proces | GO:20002 regulation | 1.33E-03 | 1.00E+00 | 8.98E-03 | 6.80E-02 | 2 | 145 | KLF4,DUSP1 |
| M0/Control | Down | GO: Biological Proces | GO:00301 osteoclast | 1.65E-03 | 1.00E+00 | 1.04E-02 | 7.84E-02 | 2 | 162 | FOS,JUNB |
| M0/Control | Down | GO: Biological Proces | GO:07134 cellular res | 1.81E-03 | 1.00E+00 | 1.12E-02 | 8.50E-02 | 2 | 170 | EGR1,KLF2 |
| M0/Control | Down | GO: Biological Proces | GO:00018 blastocyst | 2.10E-03 | 1.00E+00 | 1.23E-02 | 9.30E-02 | 2 | 183 | KLF4,JUNB |
| M0/Control | Down | GO: Biological Proces | GO:03252 response t | 2.14E-03 | 1.00E+00 | 1.24E-02 | 9.35E-02 | 2 | 185 | KLF4,DUSP1 |
| M0/Control | Down | GO: Biological Proces | GO:00165 negative r | 2.24E-03 | 1.00E+00 | 1.27E-02 | 9.64E-02 | 2 | 189 | KLF4,KLF2 |
| M0/Control | Down | GO: Biological Proces | GO:20001 negative r | 2.31E-03 | 1.00E+00 | 1.27E-02 | 9.64E-02 | 2 | 192 | KLF4,KLF2 |
| M0/Control | Down | GO: Biological Proces | GO:19013 negative r | 2.35E-03 | 1.00E+00 | 1.29E-02 | 9.75E-02 | 2 | 194 | KLF4,KLF2 |
| M0/Control | Down | GO: Biological Proces | GO:00102 response t | 2.55E-03 | 1.00E+00 | 1.39E-02 | 1.05E-01 | 2 | 202 | RHOB,EGR1 |
| M0/Control | Down | GO: Biological Proces | GO:07055 response t | 2.72E-03 | 1.00E+00 | 1.43E-02 | 1.08E-01 | 2 | 209 | EGR1,KLF2 |
| M0/Control | Down | GO: Biological Proces | GO:05072 negative r | 3.04E-03 | 1.00E+00 | 1.53E-02 | 1.16E-01 | 2 | 221 | ZFP36,KLF4 |
| M0/Control | Down | GO: Biological Proces | GO:19019 negative r | 3.07E-03 | 1.00E+00 | 1.53E-02 | 1.16E-01 | 2 | 222 | KLF4,DUSP1 |
| M0/Control | Down | GO: Biological Proces | GO:04866 regulation | 3.09E-03 | 1.00E+00 | 1.53E-02 | 1.16E-01 | 2 | 223 | KLF4,EGR1 |
| M0/Control | Down | GO: Biological Proces | GO:07141 cellular res | 3.15E-03 | 1.00E+00 | 1.53E-02 | 1.16E-01 | 2 | 225 | RHOB,EGR1 |
| M0/Control | Down | GO: Biological Proces | GO:07189 DNA biosy | 3.15E-03 | 1.00E+00 | 1.53E-02 | 1.16E-01 | 2 | 225 | KLF4,DUSP1 |
| M0/Control | Down | GO: Biological Proces | GO:04340 negative r | 3.29E-03 | 1.00E+00 | 1.58E-02 | 1.20E-01 | 2 | 230 | KLF4,DUSP1 |
| M0/Control | Down | GO: Biological Proces | GO:04578 positive re | 3.43E-03 | 1.00E+00 | 1.60E-02 | 1.21E-01 | 2 | 235 | KLF4,RHOB |
| M0/Control | Down | GO: Biological Proces | GO:19040 positive re | 3.46E-03 | 1.00E+00 | 1.60E-02 | 1.21E-01 | 2 | 236 | KLF4,RHOB |
| M0/Control | Down | GO: Biological Proces | GO:04866 smooth m | 3.46E-03 | 1.00E+00 | 1.60E-02 | 1.21E-01 | 2 | 236 | KLF4,EGR1 |
| M0/Control | Down | GO: Biological Proces | GO:00749 mesoderm | 3.49E-03 | 1.00E+00 | 1.60E-02 | 1.21E-01 | 2 | 237 | KLF4,KLF2 |
| M0/Control | Down | GO: Biological Proces | GO:03021 erythrocyt | 3.78E-03 | 1.00E+00 | 1.69E-02 | 1.28E-01 | 2 | 247 | ZFP36,KLF2 |
| M0/Control | Down | GO: Biological Proces | GO:04224 tissue rege | 4.02E-03 | 1.00E+00 | 1.75E-02 | 1.32E-01 | 2 | 255 | FOS,JUNB |
| M0/Control | Down | GO: Biological Proces | GO:03410 erythrocyt | 4.18E-03 | 1.00E+00 | 1.79E-02 | 1.35E-01 | 2 | 260 | ZFP36,KLF2 |
| M0/Control | Down | GO: Biological Proces | GO:07124 cellular res | 4.83E-03 | 1.00E+00 | 2.01E-02 | 1.52E-01 | 2 | 280 | FOS,JUNB |
| M0/Control | Down | GO: Biological Proces | GO:00105 regulation | 4.86E-03 | 1.00E+00 | 2.01E-02 | 1.52E-01 | 2 | 281 | KLF4,RHOB |
| M0/Control | Down | GO: Biological Proces | GO:04255 response t | 4.93E-03 | 1.00E+00 | 2.01E-02 | 1.52E-01 | 2 | 283 | FOS,ZFP36 |
| M0/Control | Down | GO: Biological Proces | GO:04593 negative r | 5.14E-03 | 1.00E+00 | 2.06E-02 | 1.56E-01 | 2 | 289 | KLF4,DUSP1 |
| M0/Control | Down | GO: Biological Proces | GO:00204 sprouting | 5.31E-03 | 1.00E+00 | 2.10E-02 | 1.59E-01 | 2 | 294 | KLF4,KLF2 |
| M0/Control | Down | GO: Biological Proces | GO:04541 response t | 5.38E-03 | 1.00E+00 | 2.10E-02 | 1.59E-01 | 2 | 296 | FOS,EGR1 |
| M0/Control | Down | GO: Biological Proces | GO:04544 fat cell dif | 5.45E-03 | 1.00E+00 | 2.11E-02 | 1.60E-01 | 2 | 298 | ZFP36,KLF4 |
| M0/Control | Down | GO: Biological Proces | GO:05140 stress-acti | 5.63E-03 | 1.00E+00 | 2.17E-02 | 1.64E-01 | 2 | 303 | ZFP36,DUSP1 |
| M0/Control | Down | GO: Biological Proces | GO:04563 regulation | 5.63E-03 | 1.00E+00 | 2.17E-02 | 1.64E-01 | 2 | 303 | FOS,ZFP36 |
| M0/Control | Down | GO: Biological Proces | GO:07138 cellular res | 5.70E-03 | 1.00E+00 | 2.18E-02 | 1.65E-01 | 2 | 305 | ZFP36,EGR1 |
| M0/Control | Down | GO: Biological Proces | GO:00224 hematopo | 5.85E-03 | 1.00E+00 | 2.20E-02 | 1.66E-01 | 2 | 309 | FOS,ZFP36 |
| M0/Control | Down | GO: Biological Proces | GO:03105 stress-acti | 6.00E-03 | 1.00E+00 | 2.23E-02 | 1.69E-01 | 2 | 313 | ZFP36,DUSP1 |
| M0/Control | Down | GO: Biological Proces | GO:19019 negative r | 6.15E-03 | 1.00E+00 | 2.26E-02 | 1.71E-01 | 2 | 317 | KLF4,DUSP1 |
| M0/Control | Down | GO: Biological Proces | GO:00226 myeloid ce | 6.34E-03 | 1.00E+00 | 2.32E-02 | 1.75E-01 | 2 | 322 | ZFP36,KLF2 |
| M0/Control | Down | GO: Biological Proces | GO:05134 negative r | 6.61E-03 | 1.00E+00 | 2.34E-02 | 1.77E-01 | 2 | 329 | ZFP36,DUSP1 |
| M0/Control | Down | GO: Biological Proces | GO:00099 epidermal | 6.69E-03 | 1.00E+00 | 2.35E-02 | 1.78E-01 | 2 | 331 | ZFP36,KLF4 |
| M0/Control | Down | GO: Biological Proces | GO:00075 skeletal m | 6.69E-03 | 1.00E+00 | 2.35E-02 | 1.78E-01 | 2 | 331 | FOS,EGR1 |
| M0/Control | Down | GO: Biological Proces | GO:00025 myeloid le | 6.84E-03 | 1.00E+00 | 2.40E-02 | 1.81E-01 | 2 | 335 | FOS,JUNB |
| M0/Control | Down | GO: Biological Proces | GO:00756 female pre | 7.00E-03 | 1.00E+00 | 2.41E-02 | 1.83E-01 | 2 | 339 | FOS,JUNB |
| M0/Control | Down | GO: Biological Proces | GO:00314 negative r | 7.12E-03 | 1.00E+00 | 2.45E-02 | 1.85E-01 | 2 | 342 | ZFP36,KLF4 |
| M0/Control | Down | GO: Biological Proces | GO:06055 skeletal m | 7.53E-03 | 1.00E+00 | 2.54E-02 | 1.92E-01 | 2 | 352 | FOS,EGR1 |
| M0/Control | Down | GO: Biological Proces | GO:04354 endothelia | 7.61E-03 | 1.00E+00 | 2.56E-02 | 1.94E-01 | 2 | 354 | KLF4,RHOB |
| M0/Control | Down | GO: Biological Proces | GO:00106 regulation | 7.70E-03 | 1.00E+00 | 2.56E-02 | 1.94E-01 | 2 | 356 | KLF4,RHOB |
| M0/Control | Down | GO: Biological Proces | GO:07124 cellular res | 8.12E-03 | 1.00E+00 | 2.68E-02 | 2.03E-01 | 2 | 366 | FOS,JUNB |
| M0/Control | Down | GO: Biological Proces | GO:04470 multi-orga | 8.29E-03 | 1.00E+00 | 2.73E-02 | 2.07E-01 | 2 | 370 | FOS,JUNB |
| M0/Control | Down | GO: Biological Proces | GO:06000 canonical | 8.38E-03 | 1.00E+00 | 2.75E-02 | 2.08E-01 | 2 | 372 | KLF4,EGR1 |
| M0/Control | Down | GO: Biological Proces | GO:00761 learning o | 8.64E-03 | 1.00E+00 | 2.81E-02 | 2.13E-01 | 2 | 378 | FOS,EGR1 |
| M0/Control | Down | GO: Biological Proces | GO:00109 negative r | 8.86E-03 | 1.00E+00 | 2.82E-02 | 2.14E-01 | 2 | 383 | KLF4,DUSP1 |
| M0/Control | Down | GO: Biological Proces | GO:04470 multi-mult | 8.86E-03 | 1.00E+00 | 2.82E-02 | 2.14E-01 | 2 | 383 | FOS,JUNB |
| M0/Control | Down | GO: Biological Proces | GO:04358 skin devel | 9.04E-03 | 1.00E+00 | 2.87E-02 | 2.17E-01 | 2 | 387 | ZFP36,KLF4 |
| M0/Control | Down | GO: Biological Proces | GO:03021 T cell diffe | 9.41E-03 | 1.00E+00 | 2.95E-02 | 2.24E-01 | 2 | 395 | EGR1,JUNB |
| M0/Control | Down | GO: Biological Proces | GO:04516 cell fate co | 9.45E-03 | 1.00E+00 | 2.96E-02 | 2.24E-01 | 2 | 396 | KLF4,KLF2 |
| M0/Control | Down | GO: Biological Proces | GO:19019 regulation | 9.87E-03 | 1.00E+00 | 3.03E-02 | 2.30E-01 | 2 | 405 | KLF4,DUSP1 |
| M0/Control | Down | GO: Biological Proces | GO:00328 response t | 9.92E-03 | 1.00E+00 | 3.03E-02 | 2.30E-01 | 2 | 406 | FOS,EGR1 |
| M0/Control | Down | GO: Biological Proces | GO:00071 negative r | 1.01E-02 | 1.00E+00 | 3.06E-02 | 2.32E-01 | 2 | 410 | KLF4,DUSP1 |
| M0/Control | Down | GO: Biological Proces | GO:00703 regulation | 1.04E-02 | 1.00E+00 | 3.12E-02 | 2.36E-01 | 2 | 417 | KLF4,DUSP1 |
| M0/Control | Down | GO: Biological Proces | GO:05086 cognition | 1.08E-02 | 1.00E+00 | 3.13E-02 | 2.37E-01 | 2 | 425 | FOS,EGR1 |
| M0/Control | Down | GO: Biological Proces | GO:19021 regulation | 1.15E-02 | 1.00E+00 | 3.26E-02 | 2.47E-01 | 2 | 438 | FOS,JUNB |
| M0/Control | Down | GO: Biological Proces | GO:00703 ERK1 and | 1.21E-02 | 1.00E+00 | 3.42E-02 | 2.59E-01 | 2 | 451 | KLF4,DUSP1 |
| M0/Control | Down | GO: Biological Proces | GO:07131 cellular res | 1.22E-02 | 1.00E+00 | 3.42E-02 | 2.59E-01 | 2 | 452 | FOS,EGR1 |
| M0/Control | Down | GO: Biological Proces | GO:00106 epithelial c | 1.27E-02 | 1.00E+00 | 3.51E-02 | 2.66E-01 | 2 | 461 | KLF4,RHOB |
| M0/Control | Down | GO: Biological Proces | GO:09001 epithelium | 1.30E-02 | 1.00E+00 | 3.55E-02 | 2.69E-01 | 2 | 467 | KLF4,RHOB |
| M0/Control | Down | GO: Biological Proces | GO:19012 regulation | 1.34E-02 | 1.00E+00 | 3.58E-02 | 2.71E-01 | 2 | 474 | FOS,EGR1 |
| M0/Control | Down | GO: Biological Proces | GO:04888 stem cell c | 1.34E-02 | 1.00E+00 | 3.58E-02 | 2.71E-01 | 2 | 475 | FOS,ZFP36 |
| M0/Control | Down | GO: Biological Proces | GO:09001 tissue mig | 1.35E-02 | 1.00E+00 | 3.59E-02 | 2.71E-01 | 2 | 477 | KLF4,RHOB |
| M0/Control | Down | GO: Biological Proces | GO:07121 cellular res | 1.36E-02 | 1.00E+00 | 3.59E-02 | 2.71E-01 | 2 | 478 | RHOB,EGR1 |
| M0/Control | Down | GO: Biological Proces | GO:01040 cellular res | 1.36E-02 | 1.00E+00 | 3.59E-02 | 2.71E-01 | 2 | 478 | RHOB,EGR1 |
| M0/Control | Down | GO: Biological Proces | GO:07146 cellular res | 1.43E-02 | 1.00E+00 | 3.70E-02 | 2.80E-01 | 2 | 491 | FOS,EGR1 |
| M0/Control | Down | GO: Biological Proces | GO:00085 epidermis | 1.48E-02 | 1.00E+00 | 3.81E-02 | 2.89E-01 | 2 | 500 | ZFP36,KLF4 |
| M0/Control | Down | GO: Biological Proces | GO:00071 transmem | 1.60E-02 | 1.00E+00 | 3.97E-02 | 3.01E-01 | 2 | 521 | FOS,EGR1 |
| M0/Control | Down | GO: Biological Proces | GO:05072 regulation | 1.63E-02 | 1.00E+00 | 4.02E-02 | 3.04E-01 | 2 | 526 | ZFP36,KLF4 |
| M0/Control | Down | GO: Biological Proces | GO:04887 homeosta | 1.65E-02 | 1.00E+00 | 4.04E-02 | 3.06E-01 | 2 | 530 | ZFP36,KLF2 |
| M0/Control | Down | GO: Biological Proces | GO:07009 neuron de | 1.66E-02 | 1.00E+00 | 4.05E-02 | 3.06E-01 | 2 | 531 | FOS,EGR1 |
| M0/Control | Down | GO: Biological Proces | GO:04471 mitotic cel | 1.69E-02 | 1.00E+00 | 4.08E-02 | 3.09E-01 | 2 | 536 | KLF4,DUSP1 |
| M0/Control | Down | GO: Biological Proces | GO:19019 regulation | 1.72E-02 | 1.00E+00 | 4.12E-02 | 3.12E-01 | 2 | 542 | KLF4,DUSP1 |
| M0/Control | Down | GO: Biological Proces | GO:00300 lymphocyt | 1.82E-02 | 1.00E+00 | 4.32E-02 | 3.27E-01 | 2 | 558 | EGR1,JUNB |
| M0/Control | Down | GO: Biological Proces | GO:00160 Wnt signa | 1.91E-02 | 1.00E+00 | 4.47E-02 | 3.38E-01 | 2 | 572 | KLF4,EGR1 |
| M0/Control | Down | GO: Biological Proces | GO:01987 cell-cell si | 1.92E-02 | 1.00E+00 | 4.47E-02 | 3.38E-01 | 2 | 574 | KLF4,EGR1 |
| M0/Control | Down | GO: Biological Proces | GO:00324 response t | 2.07E-02 | 1.00E+00 | 4.65E-02 | 3.52E-01 | 2 | 597 | FOS,ZFP36 |
| M0/Control | Down | GO: Biological Proces | GO:00734 regulation | 2.18E-02 | 1.00E+00 | 4.81E-02 | 3.64E-01 | 2 | 614 | KLF4,DUSP1 |
| M0/Control | Down | GO: Biological Proces | GO:00022 response t | 2.25E-02 | 1.00E+00 | 4.90E-02 | 3.71E-01 | 2 | 624 | FOS,ZFP36 |
| M0/Control | Down | GO: Cellular Compon | GO:00359 transcripti | 1.28E-06 | 4.73E-05 | 4.73E-05 | 1.99E-04 | 2 | 5 | FOS,JUNB |
| M0/Control | Down | GO: Cellular Compon | GO:00005 RNA polyr | 4.07E-03 | 1.50E-01 | 3.11E-02 | 1.31E-01 | 2 | 259 | FOS,JUNB |
| M0/Control | Down | GO: Molecular Functi | GO:04472 hemi-meth | 1.20E-03 | 8.77E-02 | 5.31E-03 | 2.59E-02 | 1 | 3 | EGR1 |
| M0/Control | Down | GO: Molecular Functi | GO:00101 RNA polyr | 1.60E-03 | 1.17E-01 | 6.50E-03 | 3.17E-02 | 1 | 4 | KLF4 |
| M0/Control | Down | GO: Molecular Functi | GO:00103 double-str | 2.80E-03 | 2.05E-01 | 9.74E-03 | 4.75E-02 | 1 | 7 | EGR1 |
| M0/Control | Down | GO: Molecular Functi | GO:00083 protein ty | 4.40E-03 | 3.21E-01 | 1.40E-02 | 6.81E-02 | 1 | 11 | DUSP1 |
| M0/Control | Down | GO: Molecular Functi | GO:00170 MAP kina | 5.60E-03 | 4.09E-01 | 1.64E-02 | 7.97E-02 | 1 | 14 | DUSP1 |
| M0/Control | Down | GO: Molecular Functi | GO:00335 MAP kina | 7.59E-03 | 5.54E-01 | 2.13E-02 | 1.04E-01 | 1 | 19 | DUSP1 |
| M0/Control | Down | GO: Molecular Functi | GO:03501 phosphati | 8.78E-03 | 6.41E-01 | 2.38E-02 | 1.16E-01 | 1 | 22 | KLF4 |
| M0/Control | Down | GO: Molecular Functi | GO:00199 C-C chemo | 9.98E-03 | 7.28E-01 | 2.51E-02 | 1.22E-01 | 1 | 25 | ZFP36 |
| M0/Control | Down | GO: Molecular Functi | GO:00704 R-SMAD b | 9.98E-03 | 7.28E-01 | 2.51E-02 | 1.22E-01 | 1 | 25 | FOS |
| M0/Control | Down | GO: Molecular Functi | GO:03559 mRNA 3'-U | 1.04E-02 | 7.57E-01 | 2.52E-02 | 1.23E-01 | 1 | 26 | ZFP36 |
| M0/Control | Down | GO: Molecular Functi | GO:00350 histone ac | 1.16E-02 | 8.44E-01 | 2.67E-02 | 1.30E-01 | 1 | 29 | EGR1 |
| M0/Control | Down | GO: Molecular Functi | GO:00350 phosphati | 1.28E-02 | 9.31E-01 | 2.82E-02 | 1.38E-01 | 1 | 32 | KLF4 |
| M0/Control | Down | GO: Molecular Functi | GO:00199 chemokine | 1.36E-02 | 9.89E-01 | 2.91E-02 | 1.42E-01 | 1 | 34 | ZFP36 |
| M0/Control | Down | GO: Molecular Functi | GO:05101 mitogen-a | 1.39E-02 | 1.00E+00 | 2.91E-02 | 1.42E-01 | 1 | 35 | DUSP1 |
| M0/Control | Down | GO: Molecular Functi | GO:00718 14-3-3 pro | 1.63E-02 | 1.00E+00 | 3.31E-02 | 1.61E-01 | 1 | 41 | ZFP36 |
| M0/Control | Down | GO: Molecular Functi | GO:00081 protein ty | 1.95E-02 | 1.00E+00 | 3.84E-02 | 1.87E-01 | 1 | 49 | DUSP1 |
| M0/Control | Down | GO: Molecular Functi | GO:00700 RNA polyr | 2.50E-02 | 1.00E+00 | 4.68E-02 | 2.28E-01 | 1 | 63 | ZFP36 |
| M0/Control | Down | GO: Biological Proces | GO:19049 negative r | 3.86E-04 | 4.21E-01 | 4.08E-03 | 3.09E-02 | 1 | 1 | KLF4 |
| M0/Control | Down | GO: Biological Proces | GO:00723 positive re | 3.86E-04 | 4.21E-01 | 4.08E-03 | 3.09E-02 | 1 | 1 | EGR1 |
| M0/Control | Down | GO: Biological Proces | GO:19018 positive re | 3.86E-04 | 4.21E-01 | 4.08E-03 | 3.09E-02 | 1 | 1 | EGR1 |
| M0/Control | Down | GO: Biological Proces | GO:19018 positive re | 3.86E-04 | 4.21E-01 | 4.08E-03 | 3.09E-02 | 1 | 1 | ZFP36 |
| M0/Control | Down | GO: Biological Proces | GO:19018 regulation | 3.86E-04 | 4.21E-01 | 4.08E-03 | 3.09E-02 | 1 | 1 | ZFP36 |
| M0/Control | Down | GO: Biological Proces | GO:00727 response t | 3.86E-04 | 4.21E-01 | 4.08E-03 | 3.09E-02 | 1 | 1 | JUNB |
| M0/Control | Down | GO: Biological Proces | GO:19042 negative r | 3.86E-04 | 4.21E-01 | 4.08E-03 | 3.09E-02 | 1 | 1 | ZFP36 |
| M0/Control | Down | GO: Biological Proces | GO:06144 endocardi | 7.72E-04 | 8.41E-01 | 6.39E-03 | 4.83E-02 | 1 | 2 | KLF2 |
| M0/Control | Down | GO: Biological Proces | GO:00723 regulation | 7.72E-04 | 8.41E-01 | 6.39E-03 | 4.83E-02 | 1 | 2 | EGR1 |
| M0/Control | Down | GO: Biological Proces | GO:19042 regulation | 7.72E-04 | 8.41E-01 | 6.39E-03 | 4.83E-02 | 1 | 2 | ZFP36 |
| M0/Control | Down | GO: Biological Proces | GO:01474 negative r | 7.72E-04 | 8.41E-01 | 6.39E-03 | 4.83E-02 | 1 | 2 | KLF4 |
| M0/Control | Down | GO: Biological Proces | GO:07150 response t | 7.72E-04 | 8.41E-01 | 6.39E-03 | 4.83E-02 | 1 | 2 | EGR1 |
| M0/Control | Down | GO: Biological Proces | GO:07150 cellular res | 7.72E-04 | 8.41E-01 | 6.39E-03 | 4.83E-02 | 1 | 2 | EGR1 |
| M0/Control | Down | GO: Biological Proces | GO:06144 endocardi | 1.16E-03 | 1.00E+00 | 8.03E-03 | 6.08E-02 | 1 | 3 | KLF2 |
| M0/Control | Down | GO: Biological Proces | GO:00750 mesoderm | 1.16E-03 | 1.00E+00 | 8.03E-03 | 6.08E-02 | 1 | 3 | KLF4 |
| M0/Control | Down | GO: Biological Proces | GO:19045 positive re | 1.16E-03 | 1.00E+00 | 8.03E-03 | 6.08E-02 | 1 | 3 | ZFP36 |
| M0/Control | Down | GO: Biological Proces | GO:09875 cellular res | 1.16E-03 | 1.00E+00 | 8.03E-03 | 6.08E-02 | 1 | 3 | EGR1 |
| M0/Control | Down | GO: Biological Proces | GO:09875 response t | 1.16E-03 | 1.00E+00 | 8.03E-03 | 6.08E-02 | 1 | 3 | EGR1 |
| M0/Control | Down | GO: Biological Proces | GO:09753 cellular str | 1.16E-03 | 1.00E+00 | 8.03E-03 | 6.08E-02 | 1 | 3 | KLF2 |
| M0/Control | Down | GO: Biological Proces | GO:09753 stress resp | 1.16E-03 | 1.00E+00 | 8.03E-03 | 6.08E-02 | 1 | 3 | KLF2 |
| M0/Control | Down | GO: Biological Proces | GO:00171 ectoderma | 1.54E-03 | 1.00E+00 | 9.77E-03 | 7.40E-02 | 1 | 4 | KLF2 |
| M0/Control | Down | GO: Biological Proces | GO:01202 regulation | 1.54E-03 | 1.00E+00 | 9.77E-03 | 7.40E-02 | 1 | 4 | KLF4 |
| M0/Control | Down | GO: Biological Proces | GO:04266 regulation | 1.54E-03 | 1.00E+00 | 9.77E-03 | 7.40E-02 | 1 | 4 | KLF2 |
| M0/Control | Down | GO: Biological Proces | GO:06000 circadian t | 1.54E-03 | 1.00E+00 | 9.77E-03 | 7.40E-02 | 1 | 4 | EGR1 |
| M0/Control | Down | GO: Biological Proces | GO:07226 metaneph | 1.54E-03 | 1.00E+00 | 9.77E-03 | 7.40E-02 | 1 | 4 | EGR1 |
| M0/Control | Down | GO: Biological Proces | GO:04878 hatching g | 1.54E-03 | 1.00E+00 | 9.77E-03 | 7.40E-02 | 1 | 4 | KLF2 |
| M0/Control | Down | GO: Biological Proces | GO:19049 regulation | 1.93E-03 | 1.00E+00 | 1.14E-02 | 8.60E-02 | 1 | 5 | KLF4 |
| M0/Control | Down | GO: Biological Proces | GO:20001 regulation | 1.93E-03 | 1.00E+00 | 1.14E-02 | 8.60E-02 | 1 | 5 | EGR1 |
| M0/Control | Down | GO: Biological Proces | GO:19045 regulation | 1.93E-03 | 1.00E+00 | 1.14E-02 | 8.60E-02 | 1 | 5 | ZFP36 |
| M0/Control | Down | GO: Biological Proces | GO:07222 metaneph | 1.93E-03 | 1.00E+00 | 1.14E-02 | 8.60E-02 | 1 | 5 | EGR1 |
| M0/Control | Down | GO: Biological Proces | GO:04266 negative r | 1.93E-03 | 1.00E+00 | 1.14E-02 | 8.60E-02 | 1 | 5 | KLF2 |
| M0/Control | Down | GO: Biological Proces | GO:19057 negative r | 1.93E-03 | 1.00E+00 | 1.14E-02 | 8.60E-02 | 1 | 5 | KLF2 |
| M0/Control | Down | GO: Biological Proces | GO:01473 regulation | 1.93E-03 | 1.00E+00 | 1.14E-02 | 8.60E-02 | 1 | 5 | KLF4 |
| M0/Control | Down | GO: Biological Proces | GO:07150 cellular res | 1.93E-03 | 1.00E+00 | 1.14E-02 | 8.60E-02 | 1 | 5 | EGR1 |
| M0/Control | Down | GO: Biological Proces | GO:19047 positive re | 2.32E-03 | 1.00E+00 | 1.27E-02 | 9.64E-02 | 1 | 6 | KLF4 |
| M0/Control | Down | GO: Biological Proces | GO:04698 positive re | 2.32E-03 | 1.00E+00 | 1.27E-02 | 9.64E-02 | 1 | 6 | KLF4 |
| M0/Control | Down | GO: Biological Proces | GO:01490 muscle hy | 2.32E-03 | 1.00E+00 | 1.27E-02 | 9.64E-02 | 1 | 6 | KLF4 |
| M0/Control | Down | GO: Biological Proces | GO:03108 deadenyla | 2.32E-03 | 1.00E+00 | 1.27E-02 | 9.64E-02 | 1 | 6 | ZFP36 |
| M0/Control | Down | GO: Biological Proces | GO:06175 leukocyte | 2.32E-03 | 1.00E+00 | 1.27E-02 | 9.64E-02 | 1 | 6 | KLF4 |
| M0/Control | Down | GO: Biological Proces | GO:20003 negative r | 2.32E-03 | 1.00E+00 | 1.27E-02 | 9.64E-02 | 1 | 6 | KLF4 |
| M0/Control | Down | GO: Biological Proces | GO:03516 post-embr | 2.70E-03 | 1.00E+00 | 1.42E-02 | 1.08E-01 | 1 | 7 | KLF4 |
| M0/Control | Down | GO: Biological Proces | GO:19018 regulation | 2.70E-03 | 1.00E+00 | 1.42E-02 | 1.08E-01 | 1 | 7 | EGR1 |
| M0/Control | Down | GO: Biological Proces | GO:04838 positive re | 2.70E-03 | 1.00E+00 | 1.42E-02 | 1.08E-01 | 1 | 7 | KLF2 |
| M0/Control | Down | GO: Biological Proces | GO:03108 nuclear-tra | 2.70E-03 | 1.00E+00 | 1.42E-02 | 1.08E-01 | 1 | 7 | ZFP36 |
| M0/Control | Down | GO: Biological Proces | GO:00166 condition | 2.70E-03 | 1.00E+00 | 1.42E-02 | 1.08E-01 | 1 | 7 | FOS |
| M0/Control | Down | GO: Biological Proces | GO:00900 negative r | 2.70E-03 | 1.00E+00 | 1.42E-02 | 1.08E-01 | 1 | 7 | DUSP1 |
| M0/Control | Down | GO: Biological Proces | GO:07212 positive re | 3.09E-03 | 1.00E+00 | 1.53E-02 | 1.16E-01 | 1 | 8 | EGR1 |
| M0/Control | Down | GO: Biological Proces | GO:19021 regulation | 3.09E-03 | 1.00E+00 | 1.53E-02 | 1.16E-01 | 1 | 8 | ZFP36 |
| M0/Control | Down | GO: Biological Proces | GO:19047 regulation | 3.09E-03 | 1.00E+00 | 1.53E-02 | 1.16E-01 | 1 | 8 | KLF4 |
| M0/Control | Down | GO: Biological Proces | GO:00171 ectoderma | 3.09E-03 | 1.00E+00 | 1.53E-02 | 1.16E-01 | 1 | 8 | KLF2 |
| M0/Control | Down | GO: Biological Proces | GO:09728 keratinocy | 3.09E-03 | 1.00E+00 | 1.53E-02 | 1.16E-01 | 1 | 8 | ZFP36 |
| M0/Control | Down | GO: Biological Proces | GO:07150 response t | 3.09E-03 | 1.00E+00 | 1.53E-02 | 1.16E-01 | 1 | 8 | EGR1 |
| M0/Control | Down | GO: Biological Proces | GO:19029 positive re | 3.47E-03 | 1.00E+00 | 1.60E-02 | 1.21E-01 | 1 | 9 | EGR1 |
| M0/Control | Down | GO: Biological Proces | GO:03446 response t | 3.47E-03 | 1.00E+00 | 1.60E-02 | 1.21E-01 | 1 | 9 | EGR1 |
| M0/Control | Down | GO: Biological Proces | GO:07181 response t | 3.47E-03 | 1.00E+00 | 1.60E-02 | 1.21E-01 | 1 | 9 | EGR1 |
| M0/Control | Down | GO: Biological Proces | GO:00067 progester | 3.47E-03 | 1.00E+00 | 1.60E-02 | 1.21E-01 | 1 | 9 | EGR1 |
| M0/Control | Down | GO: Biological Proces | GO:07223 metaneph | 3.47E-03 | 1.00E+00 | 1.60E-02 | 1.21E-01 | 1 | 9 | EGR1 |
| M0/Control | Down | GO: Biological Proces | GO:19037 negative r | 3.47E-03 | 1.00E+00 | 1.60E-02 | 1.21E-01 | 1 | 9 | DUSP1 |

| | | | | | | | | | | |
|---|---|---|---|---|---|---|---|---|---|---|
| M0/Control | Down | GO: Biological Proces | GO:004698regulation | 3.47E-03 | 1.00E+00 | 1.60E-02 | 1.21E-01 | 1 | 9 | KLF4 |
| M0/Control | Down | GO: Biological Proces | GO:006015embryonic | 3.86E-03 | 1.00E+00 | 1.69E-02 | 1.28E-01 | 1 | 10 | JUNB |
| M0/Control | Down | GO: Biological Proces | GO:007145cellular re | 3.86E-03 | 1.00E+00 | 1.69E-02 | 1.28E-01 | 1 | 10 | EGR1 |
| M0/Control | Down | GO: Biological Proces | GO:000170ectoderm | 3.86E-03 | 1.00E+00 | 1.69E-02 | 1.28E-01 | 1 | 10 | KLF2 |
| M0/Control | Down | GO: Biological Proces | GO:006050type I pne | 3.86E-03 | 1.00E+00 | 1.69E-02 | 1.28E-01 | 1 | 10 | KLF2 |
| M0/Control | Down | GO: Biological Proces | GO:000322ventricular | 3.86E-03 | 1.00E+00 | 1.69E-02 | 1.28E-01 | 1 | 10 | KLF2 |
| M0/Control | Down | GO: Biological Proces | GO:004266regulation | 3.86E-03 | 1.00E+00 | 1.69E-02 | 1.28E-01 | 1 | 10 | KLF2 |
| M0/Control | Down | GO: Biological Proces | GO:190203negative n | 3.86E-03 | 1.00E+00 | 1.69E-02 | 1.28E-01 | 1 | 10 | ZFP36 |
| M0/Control | Down | GO: Biological Proces | GO:007131cellular re | 3.86E-03 | 1.00E+00 | 1.69E-02 | 1.28E-01 | 1 | 10 | EGR1 |
| M0/Control | Down | GO: Biological Proces | GO:190172positive re | 3.86E-03 | 1.00E+00 | 1.69E-02 | 1.28E-01 | 1 | 10 | EGR1 |
| M0/Control | Down | GO: Biological Proces | GO:199064cellular re | 4.24E-03 | 1.00E+00 | 1.79E-02 | 1.35E-01 | 1 | 11 | FOS |
| M0/Control | Down | GO: Biological Proces | GO:007220cell prolife | 4.24E-03 | 1.00E+00 | 1.79E-02 | 1.35E-01 | 1 | 11 | EGR1 |
| M0/Control | Down | GO: Biological Proces | GO:003101post-emb | 4.24E-03 | 1.00E+00 | 1.79E-02 | 1.35E-01 | 1 | 11 | KLF4 |
| M0/Control | Down | GO: Biological Proces | GO:003411negative n | 4.63E-03 | 1.00E+00 | 1.94E-02 | 1.47E-01 | 1 | 12 | KLF4 |
| M0/Control | Down | GO: Biological Proces | GO:190499negative n | 5.01E-03 | 1.00E+00 | 2.01E-02 | 1.52E-01 | 1 | 13 | KLF4 |
| M0/Control | Down | GO: Biological Proces | GO:009701cellular re | 5.01E-03 | 1.00E+00 | 2.01E-02 | 1.52E-01 | 1 | 13 | ZFP36 |
| M0/Control | Down | GO: Biological Proces | GO:007058regulation | 5.01E-03 | 1.00E+00 | 2.01E-02 | 1.52E-01 | 1 | 13 | KLF4 |
| M0/Control | Down | GO: Biological Proces | GO:190462cellular re | 5.01E-03 | 1.00E+00 | 2.01E-02 | 1.52E-01 | 1 | 13 | FOS |
| M0/Control | Down | GO: Biological Proces | GO:190462response t | 5.01E-03 | 1.00E+00 | 2.01E-02 | 1.52E-01 | 1 | 13 | FOS |
| M0/Control | Down | GO: Biological Proces | GO:000235defense re | 5.01E-03 | 1.00E+00 | 2.01E-02 | 1.52E-01 | 1 | 13 | KLF4 |
| M0/Control | Down | GO: Biological Proces | GO:000215osteoclast | 5.01E-03 | 1.00E+00 | 2.01E-02 | 1.52E-01 | 1 | 13 | JUNB |
| M0/Control | Down | GO: Biological Proces | GO:007212regulation | 5.39E-03 | 1.00E+00 | 2.10E-02 | 1.59E-01 | 1 | 14 | EGR1 |
| M0/Control | Down | GO: Biological Proces | GO:006141regulation | 5.39E-03 | 1.00E+00 | 2.10E-02 | 1.59E-01 | 1 | 14 | EGR1 |
| M0/Control | Down | GO: Biological Proces | GO:009701response t | 5.39E-03 | 1.00E+00 | 2.10E-02 | 1.59E-01 | 1 | 14 | ZFP36 |
| M0/Control | Down | GO: Biological Proces | GO:001066ectoderm | 5.39E-03 | 1.00E+00 | 2.10E-02 | 1.59E-01 | 1 | 14 | KLF2 |
| M0/Control | Down | GO: Biological Proces | GO:006021positive re | 5.39E-03 | 1.00E+00 | 2.10E-02 | 1.59E-01 | 1 | 14 | ZFP36 |
| M0/Control | Down | GO: Biological Proces | GO:190571regulation | 5.39E-03 | 1.00E+00 | 2.10E-02 | 1.59E-01 | 1 | 14 | KLF2 |
| M0/Control | Down | GO: Biological Proces | GO:007211glomerula | 5.78E-03 | 1.00E+00 | 2.18E-02 | 1.65E-01 | 1 | 15 | EGR1 |
| M0/Control | Down | GO: Biological Proces | GO:199063response t | 5.78E-03 | 1.00E+00 | 2.18E-02 | 1.65E-01 | 1 | 15 | FOS |
| M0/Control | Down | GO: Biological Proces | GO:007026peptidyl-s | 5.78E-03 | 1.00E+00 | 2.18E-02 | 1.65E-01 | 1 | 15 | DUSP1 |
| M0/Control | Down | GO: Biological Proces | GO:004564negative n | 5.78E-03 | 1.00E+00 | 2.18E-02 | 1.65E-01 | 1 | 15 | ZFP36 |
| M0/Control | Down | GO: Biological Proces | GO:001474negative n | 5.78E-03 | 1.00E+00 | 2.18E-02 | 1.65E-01 | 1 | 15 | KLF4 |
| M0/Control | Down | GO: Biological Proces | GO:190015positive re | 6.16E-03 | 1.00E+00 | 2.26E-02 | 1.71E-01 | 1 | 16 | ZFP36 |
| M0/Control | Down | GO: Biological Proces | GO:011015methylgua | 6.16E-03 | 1.00E+00 | 2.26E-02 | 1.71E-01 | 1 | 16 | ZFP36 |
| M0/Control | Down | GO: Biological Proces | GO:012012response t | 6.55E-03 | 1.00E+00 | 2.32E-02 | 1.76E-01 | 1 | 17 | FOS |
| M0/Control | Down | GO: Biological Proces | GO:190294regulation | 6.55E-03 | 1.00E+00 | 2.32E-02 | 1.76E-01 | 1 | 17 | EGR1 |
| M0/Control | Down | GO: Biological Proces | GO:003422cellular re | 6.55E-03 | 1.00E+00 | 2.32E-02 | 1.76E-01 | 1 | 17 | FOS |
| M0/Control | Down | GO: Biological Proces | GO:007058cell-cell ac | 6.55E-03 | 1.00E+00 | 2.32E-02 | 1.76E-01 | 1 | 17 | KLF4 |
| M0/Control | Down | GO: Biological Proces | GO:007222metaneph | 6.55E-03 | 1.00E+00 | 2.32E-02 | 1.76E-01 | 1 | 17 | EGR1 |
| M0/Control | Down | GO: Biological Proces | GO:007131cellular re | 6.55E-03 | 1.00E+00 | 2.32E-02 | 1.76E-01 | 1 | 17 | FOS |
| M0/Control | Down | GO: Biological Proces | GO:006021regulation | 6.55E-03 | 1.00E+00 | 2.32E-02 | 1.76E-01 | 1 | 17 | ZFP36 |
| M0/Control | Down | GO: Biological Proces | GO:0061113'-UTR-me | 6.55E-03 | 1.00E+00 | 2.32E-02 | 1.76E-01 | 1 | 17 | ZFP36 |
| M0/Control | Down | GO: Biological Proces | GO:000829intracellula | 6.55E-03 | 1.00E+00 | 2.32E-02 | 1.76E-01 | 1 | 17 | ZFP36 |
| M0/Control | Down | GO: Biological Proces | GO:003625cellular re | 6.93E-03 | 1.00E+00 | 2.40E-02 | 1.82E-01 | 1 | 18 | EGR1 |
| M0/Control | Down | GO: Biological Proces | GO:003289negative n | 6.93E-03 | 1.00E+00 | 2.40E-02 | 1.82E-01 | 1 | 18 | ZFP36 |
| M0/Control | Down | GO: Biological Proces | GO:190172regulation | 6.93E-03 | 1.00E+00 | 2.40E-02 | 1.82E-01 | 1 | 18 | EGR1 |
| M0/Control | Down | GO: Biological Proces | GO:000999negative n | 7.31E-03 | 1.00E+00 | 2.48E-02 | 1.88E-01 | 1 | 19 | KLF2 |
| M0/Control | Down | GO: Biological Proces | GO:006102cardiac m | 7.31E-03 | 1.00E+00 | 2.48E-02 | 1.88E-01 | 1 | 19 | FOS |
| M0/Control | Down | GO: Biological Proces | GO:011015RNA deca | 7.31E-03 | 1.00E+00 | 2.48E-02 | 1.88E-01 | 1 | 19 | ZFP36 |
| M0/Control | Down | GO: Biological Proces | GO:003591peptidyl-t | 7.31E-03 | 1.00E+00 | 2.48E-02 | 1.88E-01 | 1 | 19 | DUSP1 |
| M0/Control | Down | GO: Biological Proces | GO:007210glomerula | 7.70E-03 | 1.00E+00 | 2.56E-02 | 1.94E-01 | 1 | 20 | EGR1 |
| M0/Control | Down | GO: Biological Proces | GO:004688positive re | 7.70E-03 | 1.00E+00 | 2.56E-02 | 1.94E-01 | 1 | 20 | EGR1 |
| M0/Control | Down | GO: Biological Proces | GO:190153negative n | 8.08E-03 | 1.00E+00 | 2.68E-02 | 2.03E-01 | 1 | 21 | ZFP36 |
| M0/Control | Down | GO: Biological Proces | GO:190350regulation | 8.46E-03 | 1.00E+00 | 2.76E-02 | 2.09E-01 | 1 | 22 | DUSP1 |
| M0/Control | Down | GO: Biological Proces | GO:009026regulation | 8.46E-03 | 1.00E+00 | 2.76E-02 | 2.09E-01 | 1 | 22 | DUSP1 |
| M0/Control | Down | GO: Biological Proces | GO:007110response t | 8.85E-03 | 1.00E+00 | 2.82E-02 | 2.14E-01 | 1 | 23 | FOS |
| M0/Control | Down | GO: Biological Proces | GO:000962response t | 8.85E-03 | 1.00E+00 | 2.82E-02 | 2.14E-01 | 1 | 23 | FOS |
| M0/Control | Down | GO: Biological Proces | GO:001056regulation | 8.85E-03 | 1.00E+00 | 2.82E-02 | 2.14E-01 | 1 | 23 | EGR1 |
| M0/Control | Down | GO: Biological Proces | GO:007256chemokine | 9.23E-03 | 1.00E+00 | 2.91E-02 | 2.20E-01 | 1 | 24 | KLF4 |
| M0/Control | Down | GO: Biological Proces | GO:200034regulation | 9.23E-03 | 1.00E+00 | 2.91E-02 | 2.20E-01 | 1 | 24 | KLF4 |
| M0/Control | Down | GO: Biological Proces | GO:009023regulation | 9.23E-03 | 1.00E+00 | 2.91E-02 | 2.20E-01 | 1 | 24 | DUSP1 |
| M0/Control | Down | GO: Biological Proces | GO:000750mesoderm | 9.61E-03 | 1.00E+00 | 2.97E-02 | 2.25E-01 | 1 | 25 | KLF2 |
| M0/Control | Down | GO: Biological Proces | GO:001406negative n | 9.61E-03 | 1.00E+00 | 2.97E-02 | 2.25E-01 | 1 | 25 | KLF4 |
| M0/Control | Down | GO: Biological Proces | GO:001485skeletal m | 9.61E-03 | 1.00E+00 | 2.97E-02 | 2.25E-01 | 1 | 25 | FOS |
| M0/Control | Down | GO: Biological Proces | GO:004678regulation | 9.61E-03 | 1.00E+00 | 2.97E-02 | 2.25E-01 | 1 | 25 | ZFP36 |
| M0/Control | Down | GO: Biological Proces | GO:007211cell prolife | 1.00E-02 | 1.00E+00 | 3.03E-02 | 2.30E-01 | 1 | 26 | EGR1 |
| M0/Control | Down | GO: Biological Proces | GO:007161negative n | 1.00E-02 | 1.00E+00 | 3.03E-02 | 2.30E-01 | 1 | 26 | DUSP1 |
| M0/Control | Down | GO: Biological Proces | GO:004244progester | 1.00E-02 | 1.00E+00 | 3.03E-02 | 2.30E-01 | 1 | 26 | EGR1 |
| M0/Control | Down | GO: Biological Proces | GO:003235positive re | 1.00E-02 | 1.00E+00 | 3.03E-02 | 2.30E-01 | 1 | 26 | EGR1 |
| M0/Control | Down | GO: Biological Proces | GO:200031regulation | 1.00E-02 | 1.00E+00 | 3.03E-02 | 2.30E-01 | 1 | 26 | JUNB |
| M0/Control | Down | GO: Biological Proces | GO:005144negative n | 1.04E-02 | 1.00E+00 | 3.11E-02 | 2.36E-01 | 1 | 27 | DUSP1 |
| M0/Control | Down | GO: Biological Proces | GO:200063positive re | 1.04E-02 | 1.00E+00 | 3.11E-02 | 2.36E-01 | 1 | 27 | ZFP36 |
| M0/Control | Down | GO: Biological Proces | GO:004541locomotor | 1.04E-02 | 1.00E+00 | 3.11E-02 | 2.36E-01 | 1 | 27 | EGR1 |
| M0/Control | Down | GO: Biological Proces | GO:003600positive re | 1.08E-02 | 1.00E+00 | 3.12E-02 | 2.36E-01 | 1 | 28 | KLF2 |
| M0/Control | Down | GO: Biological Proces | GO:190031positive re | 1.08E-02 | 1.00E+00 | 3.12E-02 | 2.36E-01 | 1 | 28 | ZFP36 |
| M0/Control | Down | GO: Biological Proces | GO:190015regulation | 1.08E-02 | 1.00E+00 | 3.12E-02 | 2.36E-01 | 1 | 28 | ZFP36 |
| M0/Control | Down | GO: Biological Proces | GO:006014positive re | 1.08E-02 | 1.00E+00 | 3.12E-02 | 2.36E-01 | 1 | 28 | ZFP36 |
| M0/Control | Down | GO: Biological Proces | GO:004254hemoglob | 1.08E-02 | 1.00E+00 | 3.12E-02 | 2.36E-01 | 1 | 28 | KLF4 |
| M0/Control | Down | GO: Biological Proces | GO:200061positive re | 1.08E-02 | 1.00E+00 | 3.12E-02 | 2.36E-01 | 1 | 28 | KLF4 |
| M0/Control | Down | GO: Biological Proces | GO:000670C21-steroi | 1.08E-02 | 1.00E+00 | 3.12E-02 | 2.36E-01 | 1 | 28 | EGR1 |
| M0/Control | Down | GO: Biological Proces | GO:004838regulation | 1.08E-02 | 1.00E+00 | 3.12E-02 | 2.36E-01 | 1 | 28 | KLF2 |
| M0/Control | Down | GO: Biological Proces | GO:000182trophecto | 1.08E-02 | 1.00E+00 | 3.12E-02 | 2.36E-01 | 1 | 28 | JUNB |
| M0/Control | Down | GO: Biological Proces | GO:003411regulation | 1.08E-02 | 1.00E+00 | 3.12E-02 | 2.36E-01 | 1 | 28 | KLF4 |
| M0/Control | Down | GO: Biological Proces | GO:001045negative n | 1.11E-02 | 1.00E+00 | 3.18E-02 | 2.41E-01 | 1 | 29 | KLF2 |
| M0/Control | Down | GO: Biological Proces | GO:006071labyrinthin | 1.11E-02 | 1.00E+00 | 3.18E-02 | 2.41E-01 | 1 | 29 | JUNB |
| M0/Control | Down | GO: Biological Proces | GO:004856post-embr | 1.11E-02 | 1.00E+00 | 3.18E-02 | 2.41E-01 | 1 | 29 | KLF4 |
| M0/Control | Down | GO: Biological Proces | GO:000332regulation | 1.11E-02 | 1.00E+00 | 3.18E-02 | 2.41E-01 | 1 | 29 | EGR1 |
| M0/Control | Down | GO: Biological Proces | GO:000268negative n | 1.15E-02 | 1.00E+00 | 3.27E-02 | 2.48E-01 | 1 | 30 | DUSP1 |
| M0/Control | Down | GO: Biological Proces | GO:0070933'-UTR-me | 1.19E-02 | 1.00E+00 | 3.36E-02 | 2.54E-01 | 1 | 31 | ZFP36 |
| M0/Control | Down | GO: Biological Proces | GO:007049interleukin | 1.23E-02 | 1.00E+00 | 3.44E-02 | 2.61E-01 | 1 | 32 | EGR1 |
| M0/Control | Down | GO: Biological Proces | GO:000315morphoge | 1.27E-02 | 1.00E+00 | 3.51E-02 | 2.66E-01 | 1 | 33 | RHOB |
| M0/Control | Down | GO: Biological Proces | GO:003599response t | 1.27E-02 | 1.00E+00 | 3.51E-02 | 2.66E-01 | 1 | 33 | FOS |
| M0/Control | Down | GO: Biological Proces | GO:006115endothelia | 1.27E-02 | 1.00E+00 | 3.51E-02 | 2.66E-01 | 1 | 33 | RHOB |
| M0/Control | Down | GO: Biological Proces | GO:007148cellular re | 1.31E-02 | 1.00E+00 | 3.55E-02 | 2.69E-01 | 1 | 34 | EGR1 |
| M0/Control | Down | GO: Biological Proces | GO:190361negative n | 1.31E-02 | 1.00E+00 | 3.55E-02 | 2.69E-01 | 1 | 34 | KLF2 |
| M0/Control | Down | GO: Biological Proces | GO:190361positive re | 1.31E-02 | 1.00E+00 | 3.55E-02 | 2.69E-01 | 1 | 34 | KLF4 |
| M0/Control | Down | GO: Biological Proces | GO:007131cellular re | 1.31E-02 | 1.00E+00 | 3.55E-02 | 2.69E-01 | 1 | 34 | EGR1 |
| M0/Control | Down | GO: Biological Proces | GO:007201glomerulu | 1.31E-02 | 1.00E+00 | 3.55E-02 | 2.69E-01 | 1 | 34 | EGR1 |
| M0/Control | Down | GO: Biological Proces | GO:006005embryonic | 1.31E-02 | 1.00E+00 | 3.55E-02 | 2.69E-01 | 1 | 34 | EGR1 |
| M0/Control | Down | GO: Biological Proces | GO:007123cellular re | 1.34E-02 | 1.00E+00 | 3.58E-02 | 2.71E-01 | 1 | 35 | EGR1 |
| M0/Control | Down | GO: Biological Proces | GO:003268negative n | 1.34E-02 | 1.00E+00 | 3.58E-02 | 2.71E-01 | 1 | 35 | KLF4 |
| M0/Control | Down | GO: Biological Proces | GO:005191positive re | 1.34E-02 | 1.00E+00 | 3.58E-02 | 2.71E-01 | 1 | 35 | KLF4 |
| M0/Control | Down | GO: Biological Proces | GO:004484estrous cy | 1.34E-02 | 1.00E+00 | 3.58E-02 | 2.71E-01 | 1 | 35 | EGR1 |
| M0/Control | Down | GO: Biological Proces | GO:012025olefinic co | 1.34E-02 | 1.00E+00 | 3.58E-02 | 2.71E-01 | 1 | 35 | EGR1 |
| M0/Control | Down | GO: Biological Proces | GO:009002regulation | 1.34E-02 | 1.00E+00 | 3.58E-02 | 2.71E-01 | 1 | 35 | DUSP1 |
| M0/Control | Down | GO: Biological Proces | GO:006144kidney vas | 1.38E-02 | 1.00E+00 | 3.63E-02 | 2.75E-01 | 1 | 36 | EGR1 |
| M0/Control | Down | GO: Biological Proces | GO:000028nuclear-tr | 1.38E-02 | 1.00E+00 | 3.63E-02 | 2.75E-01 | 1 | 36 | ZFP36 |
| M0/Control | Down | GO: Biological Proces | GO:006143renal syste | 1.38E-02 | 1.00E+00 | 3.63E-02 | 2.75E-01 | 1 | 36 | EGR1 |
| M0/Control | Down | GO: Biological Proces | GO:006143establishm | 1.42E-02 | 1.00E+00 | 3.69E-02 | 2.79E-01 | 1 | 37 | KLF4 |
| M0/Control | Down | GO: Biological Proces | GO:002002hemoglob | 1.42E-02 | 1.00E+00 | 3.69E-02 | 2.79E-01 | 1 | 37 | KLF4 |
| M0/Control | Down | GO: Biological Proces | GO:003533peptidyl-t | 1.42E-02 | 1.00E+00 | 3.69E-02 | 2.79E-01 | 1 | 37 | DUSP1 |
| M0/Control | Down | GO: Biological Proces | GO:004324erythrocyt | 1.42E-02 | 1.00E+00 | 3.69E-02 | 2.79E-01 | 1 | 37 | KLF2 |
| M0/Control | Down | GO: Biological Proces | GO:007252T-helper 1 | 1.46E-02 | 1.00E+00 | 3.77E-02 | 2.86E-01 | 1 | 38 | JUNB |
| M0/Control | Down | GO: Biological Proces | GO:003271negative n | 1.50E-02 | 1.00E+00 | 3.83E-02 | 2.90E-01 | 1 | 39 | KLF4 |
| M0/Control | Down | GO: Biological Proces | GO:004688regulation | 1.50E-02 | 1.00E+00 | 3.83E-02 | 2.90E-01 | 1 | 39 | EGR1 |
| M0/Control | Down | GO: Biological Proces | GO:004669decidualiz | 1.50E-02 | 1.00E+00 | 3.83E-02 | 2.90E-01 | 1 | 39 | JUNB |
| M0/Control | Down | GO: Biological Proces | GO:004816regulation | 1.50E-02 | 1.00E+00 | 3.83E-02 | 2.90E-01 | 1 | 39 | EGR1 |
| M0/Control | Down | GO: Biological Proces | GO:200073negative n | 1.53E-02 | 1.00E+00 | 3.90E-02 | 2.95E-01 | 1 | 40 | ZFP36 |
| M0/Control | Down | GO: Biological Proces | GO:000318atrioventri | 1.57E-02 | 1.00E+00 | 3.94E-02 | 2.98E-01 | 1 | 41 | KLF2 |
| M0/Control | Down | GO: Biological Proces | GO:006041lung cell d | 1.57E-02 | 1.00E+00 | 3.94E-02 | 2.98E-01 | 1 | 41 | KLF2 |
| M0/Control | Down | GO: Biological Proces | GO:006048lung epith | 1.57E-02 | 1.00E+00 | 3.94E-02 | 2.98E-01 | 1 | 41 | KLF2 |
| M0/Control | Down | GO: Biological Proces | GO:004561positive re | 1.57E-02 | 1.00E+00 | 3.94E-02 | 2.98E-01 | 1 | 41 | FOS |
| M0/Control | Down | GO: Biological Proces | GO:003235response t | 1.57E-02 | 1.00E+00 | 3.94E-02 | 2.98E-01 | 1 | 41 | EGR1 |
| M0/Control | Down | GO: Biological Proces | GO:200031regulation | 1.57E-02 | 1.00E+00 | 3.94E-02 | 2.98E-01 | 1 | 41 | JUNB |
| M0/Control | Down | GO: Biological Proces | GO:004561regulation | 1.61E-02 | 1.00E+00 | 3.99E-02 | 3.02E-01 | 1 | 42 | ZFP36 |
| M0/Control | Down | GO: Biological Proces | GO:000739ectoderm | 1.61E-02 | 1.00E+00 | 3.99E-02 | 3.02E-01 | 1 | 42 | KLF2 |
| M0/Control | Down | GO: Biological Proces | GO:007221metaneph | 1.65E-02 | 1.00E+00 | 4.04E-02 | 3.06E-01 | 1 | 43 | EGR1 |
| M0/Control | Down | GO: Biological Proces | GO:006061placenta b | 1.65E-02 | 1.00E+00 | 4.04E-02 | 3.06E-01 | 1 | 43 | JUNB |
| M0/Control | Down | GO: Biological Proces | GO:009005negative n | 1.65E-02 | 1.00E+00 | 4.04E-02 | 3.06E-01 | 1 | 43 | KLF4 |
| M0/Control | Down | GO: Biological Proces | GO:190499regulation | 1.69E-02 | 1.00E+00 | 4.08E-02 | 3.09E-01 | 1 | 44 | KLF4 |
| M0/Control | Down | GO: Biological Proces | GO:003270negative n | 1.69E-02 | 1.00E+00 | 4.08E-02 | 3.09E-01 | 1 | 44 | ZFP36 |
| M0/Control | Down | GO: Biological Proces | GO:003356regulation | 1.69E-02 | 1.00E+00 | 4.08E-02 | 3.09E-01 | 1 | 44 | KLF4 |
| M0/Control | Down | GO: Biological Proces | GO:007168organism | 1.73E-02 | 1.00E+00 | 4.12E-02 | 3.12E-01 | 1 | 45 | KLF2 |
| M0/Control | Down | GO: Biological Proces | GO:003518hatching | 1.73E-02 | 1.00E+00 | 4.12E-02 | 3.12E-01 | 1 | 45 | KLF2 |
| M0/Control | Down | GO: Biological Proces | GO:007131cellular re | 1.73E-02 | 1.00E+00 | 4.12E-02 | 3.12E-01 | 1 | 45 | EGR1 |
| M0/Control | Down | GO: Biological Proces | GO:003368osteoblast | 1.73E-02 | 1.00E+00 | 4.12E-02 | 3.12E-01 | 1 | 45 | JUNB |
| M0/Control | Down | GO: Biological Proces | GO:190203regulation | 1.80E-02 | 1.00E+00 | 4.28E-02 | 3.24E-01 | 1 | 47 | ZFP36 |
| M0/Control | Down | GO: Biological Proces | GO:004873epidermis | 1.80E-02 | 1.00E+00 | 4.28E-02 | 3.24E-01 | 1 | 47 | KLF4 |
| M0/Control | Down | GO: Biological Proces | GO:200021negative n | 1.84E-02 | 1.00E+00 | 4.35E-02 | 3.30E-01 | 1 | 48 | DUSP1 |
| M0/Control | Down | GO: Biological Proces | GO:005145myoblast | 1.88E-02 | 1.00E+00 | 4.41E-02 | 3.34E-01 | 1 | 49 | FOS |
| M0/Control | Down | GO: Biological Proces | GO:012011steroid ho | 1.88E-02 | 1.00E+00 | 4.41E-02 | 3.34E-01 | 1 | 49 | EGR1 |
| M0/Control | Down | GO: Biological Proces | GO:004265regulation | 1.88E-02 | 1.00E+00 | 4.41E-02 | 3.34E-01 | 1 | 49 | KLF2 |
| M0/Control | Down | GO: Biological Proces | GO:004861regulation | 1.92E-02 | 1.00E+00 | 4.47E-02 | 3.38E-01 | 1 | 50 | KLF4 |
| M0/Control | Down | GO: Biological Proces | GO:000820C21-steroi | 1.92E-02 | 1.00E+00 | 4.47E-02 | 3.38E-01 | 1 | 50 | EGR1 |
| M0/Control | Down | GO: Biological Proces | GO:000709mitotic spi | 1.95E-02 | 1.00E+00 | 4.52E-02 | 3.42E-01 | 1 | 51 | DUSP1 |
| M0/Control | Down | GO: Biological Proces | GO:000761long-term | 1.95E-02 | 1.00E+00 | 4.52E-02 | 3.42E-01 | 1 | 51 | EGR1 |
| M0/Control | Down | GO: Biological Proces | GO:007111spindle as | 1.95E-02 | 1.00E+00 | 4.52E-02 | 3.42E-01 | 1 | 51 | DUSP1 |
| M0/Control | Down | GO: Biological Proces | GO:004838retinoic ac | 1.99E-02 | 1.00E+00 | 4.57E-02 | 3.46E-01 | 1 | 52 | KLF2 |
| M0/Control | Down | GO: Biological Proces | GO:003590response t | 1.99E-02 | 1.00E+00 | 4.57E-02 | 3.46E-01 | 1 | 52 | FOS |
| M0/Control | Down | GO: Biological Proces | GO:000189maternal p | 1.99E-02 | 1.00E+00 | 4.57E-02 | 3.46E-01 | 1 | 52 | JUNB |
| M0/Control | Down | GO: Biological Proces | GO:000311atrioventri | 2.03E-02 | 1.00E+00 | 4.60E-02 | 3.48E-01 | 1 | 53 | KLF2 |
| M0/Control | Down | GO: Biological Proces | GO:007051regulation | 2.03E-02 | 1.00E+00 | 4.60E-02 | 3.48E-01 | 1 | 53 | KLF4 |
| M0/Control | Down | GO: Biological Proces | GO:005509response t | 2.03E-02 | 1.00E+00 | 4.60E-02 | 3.48E-01 | 1 | 53 | EGR1 |
| M0/Control | Down | GO: Biological Proces | GO:007111mitotic spi | 2.03E-02 | 1.00E+00 | 4.60E-02 | 3.48E-01 | 1 | 53 | DUSP1 |
| M0/Control | Down | GO: Biological Proces | GO:004584negative n | 2.03E-02 | 1.00E+00 | 4.60E-02 | 3.48E-01 | 1 | 53 | DUSP1 |
| M0/Control | Down | GO: Biological Proces | GO:004562regulation | 2.03E-02 | 1.00E+00 | 4.60E-02 | 3.48E-01 | 1 | 53 | JUNB |
| M0/Control | Down | GO: Biological Proces | GO:005191regulation | 2.07E-02 | 1.00E+00 | 4.65E-02 | 3.52E-01 | 1 | 54 | KLF4 |
| M0/Control | Down | GO: Biological Proces | GO:001083regulation | 2.07E-02 | 1.00E+00 | 4.65E-02 | 3.52E-01 | 1 | 54 | ZFP36 |
| M0/Control | Down | GO: Biological Proces | GO:190191regulation | 2.07E-02 | 1.00E+00 | 4.65E-02 | 3.52E-01 | 1 | 54 | DUSP1 |
| M0/Control | Down | GO: Biological Proces | GO:190210negative n | 2.10E-02 | 1.00E+00 | 4.73E-02 | 3.58E-01 | 1 | 55 | DUSP1 |
| M0/Control | Down | GO: Biological Proces | GO:007121cellular re | 2.14E-02 | 1.00E+00 | 4.74E-02 | 3.59E-01 | 1 | 56 | FOS |
| M0/Control | Down | GO: Biological Proces | GO:007252T-helper 1 | 2.14E-02 | 1.00E+00 | 4.74E-02 | 3.59E-01 | 1 | 56 | JUNB |
| M0/Control | Down | GO: Biological Proces | GO:200081negative n | 2.14E-02 | 1.00E+00 | 4.74E-02 | 3.59E-01 | 1 | 56 | DUSP1 |
| M0/Control | Down | GO: Biological Proces | GO:003151spindle ch | 2.14E-02 | 1.00E+00 | 4.74E-02 | 3.59E-01 | 1 | 56 | DUSP1 |
| M0/Control | Down | GO: Biological Proces | GO:007131cellular re | 2.14E-02 | 1.00E+00 | 4.74E-02 | 3.59E-01 | 1 | 56 | EGR1 |
| M0/Control | Down | GO: Biological Proces | GO:003304negative n | 2.14E-02 | 1.00E+00 | 4.74E-02 | 3.59E-01 | 1 | 56 | DUSP1 |
| M0/Control | Down | GO: Biological Proces | GO:003304negative n | 2.14E-02 | 1.00E+00 | 4.74E-02 | 3.59E-01 | 1 | 56 | DUSP1 |
| M0/Control | Down | GO: Biological Proces | GO:004218ketone bio | 2.18E-02 | 1.00E+00 | 4.81E-02 | 3.64E-01 | 1 | 57 | EGR1 |
| M0/Control | Down | GO: Biological Proces | GO:190581negative n | 2.22E-02 | 1.00E+00 | 4.85E-02 | 3.67E-01 | 1 | 58 | DUSP1 |
| M0/Control | Down | GO: Biological Proces | GO:190074regulation | 2.22E-02 | 1.00E+00 | 4.85E-02 | 3.67E-01 | 1 | 58 | DUSP1 |
| M0/Control | Down | GO: Biological Proces | GO:005198negative n | 2.26E-02 | 1.00E+00 | 4.90E-02 | 3.71E-01 | 1 | 59 | DUSP1 |
| M0/Control | Down | GO: Biological Proces | GO:004368post-trans | 2.26E-02 | 1.00E+00 | 4.90E-02 | 3.71E-01 | 1 | 59 | EGR1 |
| M0/Control | Down | GO: Biological Proces | GO:000311endocardi | 2.29E-02 | 1.00E+00 | 4.94E-02 | 3.74E-01 | 1 | 60 | KLF2 |
| M0/Control | Down | GO: Biological Proces | GO:000170cell fate d | 2.29E-02 | 1.00E+00 | 4.94E-02 | 3.74E-01 | 1 | 60 | KLF4 |
| M0/Control | Down | GO: Biological Proces | GO:00609regulation | 2.29E-02 | 1.00E+00 | 4.94E-02 | 3.74E-01 | 1 | 60 | ZFP36 |
| M0/Control | Down | GO: Biological Proces | GO:200061regulation | 2.29E-02 | 1.00E+00 | 4.94E-02 | 3.74E-01 | 1 | 60 | KLF4 |
| M0/Control | Down | GO: Biological Proces | GO:006042lung epith | 2.29E-02 | 1.00E+00 | 4.94E-02 | 3.74E-01 | 1 | 60 | KLF2 |
| M0/Control | Down | GO: Biological Proces | GO:003625response t | 2.33E-02 | 1.00E+00 | 4.99E-02 | 3.78E-01 | 1 | 61 | EGR1 |
| M0/Control | Down | GO: Biological Proces | GO:001401response t | 2.33E-02 | 1.00E+00 | 4.99E-02 | 3.78E-01 | 1 | 61 | EGR1 |
| M0/Control | Down | GO: Biological Proces | GO:003304regulation | 2.33E-02 | 1.00E+00 | 4.99E-02 | 3.78E-01 | 1 | 61 | DUSP1 |
| M0/Control | Down | GO: Cellular Compon | GO:007053RISC-loadi | 4.20E-03 | 1.55E-01 | 3.11E-02 | 1.31E-01 | 1 | 11 | ZFP36 |
| M0/Control | Down | GO: Cellular Compon | GO:003001CCR4-NOT | 6.10E-03 | 2.26E-01 | 3.76E-02 | 1.58E-01 | 1 | 16 | ZFP36 |
| M0/Control | Down | GO: Cellular Compon | GO:000011exosome ( | 9.14E-03 | 3.38E-01 | 4.40E-02 | 1.85E-01 | 1 | 24 | ZFP36 |
| M0/Control | Down | GO: Cellular Compon | GO:190535exoribonu | 9.52E-03 | 3.52E-01 | 4.40E-02 | 1.85E-01 | 1 | 25 | ZFP36 |
